# Small-quantity lipid-based nutrient supplements for children age 6-24 months: a systematic review and individual participant data meta-analysis of effects on developmental outcomes and effect modifiers

**DOI:** 10.1101/2021.02.15.21251423

**Authors:** Elizabeth L. Prado, Charles D. Arnold, K. Ryan Wessells, Christine P. Stewart, Souheila Abbeddou, Seth Adu-Afarwuah, Benjamin F. Arnold, Ulla Ashorn, Per Ashorn, Elodie Becquey, Kenneth H. Brown, Jaya Chandna, Parul Christian, Holly N. Dentz, Sherlie J. L. Dulience, Lia C. H. Fernald, Emanuela Galasso, Lotta Hallamaa, Sonja Y. Hess, Lieven Huybregts, Lora L. Iannotti, Elizabeth Yakes Jimenez, Patricia Kohl, Anna Lartey, Agnes Le Port, Stephen P. Luby, Kenneth Maleta, Andrew Matchado, Susana L. Matias, Malay K. Mridha, Robert Ntozini, Clair Null, Maku E. Ocansey, Sarker Masud Parvez, John Phuka, Amy J. Pickering, Andrew J. Prendergast, Abu Ahmed Shamim, Zakia Siddiqui, Fahmida Tofail, Ann M. Weber, Lee S. F. Wu, Kathryn G. Dewey

**Author notes:** Correspondence: Address correspondence and reprint requests to Elizabeth Prado, Department of Nutrition, University of California, One Shields Ave., Davis, CA 95616. Registry and registry number*: Registered at www.crd.york.ac.uk/PROSPERO as CRD42020159971. Data described in the manuscript, code book, and analytic code will not be made available because they are compiled from 14 different trials, and access is under the control of the investigators of each of those trials. Parul Christian is a member of the Journal’s Editorial Board.

## Abstract

**Background:** Small-quantity lipid-based nutrient supplements (SQ-LNS) reduce child stunting and provide many of the fatty acids and micronutrients that are necessary for rapid brain development that occurs during infancy and early childhood. Positive effects of SQ-LNS on developmental outcomes have been found in some trials, but not others.

**Objectives:** Our objectives were to generate pooled estimates of the effect of SQ-LNS, compared to control groups that received no intervention or an intervention without any nutritional supplement, on developmental outcomes (language, social-emotional, motor, and executive function), and to identify study-level and individual-level modifiers of these effects.

**Methods:** We conducted a two-stage meta-analysis of individual participant data from 14 intervention versus control group comparisons in 13 randomized trials of SQ-LNS provided to infants and young children age 6 to 24 months in 9 low- or middle-income countries (total n=30,024). We generated study-specific estimates of SQ-LNS vs. control groups (including main effects and subgroup estimates for individual-level effect modifiers) and pooled the estimates using fixed-effects models. We used random effects meta-regression to examine potential study-level effect modifiers.

**Results:** In 11-13 intervention versus control group comparisons (n=23,588-24,561), SQ-LNS increased mean language (mean difference: 0.07 standard deviations; 95% CI: 0.04, 0.10), social-emotional (0.08; 0.05, 0.11), and motor scores (0.08; 0.05, 0.11) and reduced the prevalence of children in the lowest decile of these scores by 17% (prevalence ratio: 0.83, 95% CI 0.76, 0.91), 19% (0.81; 0.74, 0.90), and 16% (0.84; 0.77, 0.92), respectively. SQ-LNS also increased the prevalence of children walking without support at 12 months by 9% (1.09; 1.05, 1.14). Effects of SQ-LNS on language, social-emotional, and motor outcomes were larger among study populations with a higher burden (≥ 35%) of child stunting at 18 months (mean difference 0.11-0.13 SD; 8-9 comparisons) than in populations with lower stunting burden (estimates near zero). At the individual level, greater effects of SQ-LNS were found on language among children who were acutely malnourished (mean difference: 0.31) at baseline; on language (0.12), motor (0.11), and executive function (0.06) among children in households with lower socio-economic status; and on motor development among later-born children (0.11), children of older mothers (0.10), and children of mothers with lower education (0.11).

**Conclusions:** SQ-LNS provided daily to children in the range of 6-24 months of age can be expected to result in modest, but potentially important, developmental gains, particularly in populations with high child stunting burden. Certain groups of children who experience higher risk environments, such as those from poor households or with poor baseline nutritional status, have greater potential to benefit from SQ-LNS in developmental outcomes. This study was registered at www.crd.york.ac.uk/PROSPERO as CRD42020159971.

## Introduction

Brain development occurs rapidly in utero and during the first few years after birth, laying the foundation of the neural structures that underlie children’s development of cognitive skills, such as language and executive function, as well as social-emotional and motor skills (1). Adequate availability of nutrients, such as iron, iodine, zinc, B-vitamins, and essential fatty acids, are necessary for the neurodevelopmental processes that occur during this period, such as myelination, synaptogenesis, and axon and dendrite growth (2). Inadequate dietary intake during this foundational period could lead to lasting structural and functional neurodevelopmental deficits (3). From age 6 to 24 months, children are at particular risk of inadequate dietary intake of these nutrients as they transition from exclusive breastfeeding to joining family meals, called the complementary feeding period (4). Small-quantity lipid-based nutrient supplements (SQ-LNS) were designed to fill this gap between the needs and dietary intakes of key nutrients experienced by many children during this time period, for prevention of undernutrition in low- and middle-income countries (LMICs). SQ-LNS are typically made from vegetable oil, peanut paste, milk powder, and sugar, with added vitamins and minerals, thus providing many of the micronutrients and fatty acids that are necessary for brain development (5). SQ-LNS provide ∼120 kcal/d, while other LNS products (medium- and large-quantity) provide more energy and are designed for treatment of moderate and severe acute malnutrition.

Two previous systematic reviews and meta-analyses have addressed the effects of LNS provided during the complementary feeding period on developmental outcomes. (6, 7). In a 2019 Cochrane review by Das et al., the authors provided a narrative review of effects on these outcomes, but were not able to generate pooled estimates due to differences between studies in measurement and reporting of developmental outcomes (6). The other meta-analysis by Tam et al. generated pooled estimates of the effects of LNS using published data from studies on various developmental outcomes, including a total of fewer than 3,600 children (7). SQ-LNS had significant positive effects on mean language scores (effect size 0.13 standard deviations; SD; 5 studies), social-emotional scores (0.12 SD; 5 studies), and motor scores (0.13 SD; 6 studies), and no effect on executive function (3 studies) or on the prevalence of children standing or walking without support at age 12 months (4 studies), although heterogeneity across trials was moderate to substantial.

Here, we report an individual participant data (IPD) meta-analysis (8) of SQ-LNS provided during the complementary feeding period, which adds to the current evidence-base in several ways. First, we included a larger number of trials (13 trials) and children (30,024) than previous meta-analyses. Second, we analyzed IPD, rather than aggregate data from published reports, which enabled harmonization of the calculation of developmental outcomes across trials. Third, we examined study-level and individual-level factors that may modify the effect of SQ-LNS on developmental outcomes. Identifying characteristics of children and populations who experience greater benefits from SQ-LNS, or are more likely to respond to the intervention, may be useful to inform public health programs and policies. Our first objective was to generate pooled estimates of the effect of randomized controlled trials of SQ-LNS provided to infants and young children in the range of age 6 to 24 months, compared to children who received no intervention or an intervention without any nutritional supplement, on developmental outcomes. The other two objectives were to identify study-level modifiers (Objective 2) and individual-level modifiers (Objective 3) of these effects.

## Methods

The protocol for this IPD meta-analysis was registered as PROSPERO CRD42020159971 (9). The statistical analysis plan was posted to Open Science Framework prior to analysis (10), and the results are reported according to PRISMA-IPD guidelines (11). The analyses were approved by the institutional review board of the University of California, Davis. All individual trial protocols were approved by appropriate institutional ethics committees. The methods are presented in detail in a companion paper published in the same journal issue (12) and summarized here.

### Inclusion and exclusion criteria for this IPD meta-analysis

We included prospective randomized controlled trials of SQ-LNS provided to children in the range of age 6-24 months that met the inclusion criteria listed in Dewey et al. (12). In addition to those criteria, for the analyses presented here, we only included trials that measured at least one developmental outcome of interest, described below.

### Search methods and identification of studies

We identified studies cited in a recent systematic review and meta-analysis of child LNS (6) and through database searches, as described in Dewey et al. (12).

### Data collection

We invited all principal investigators of eligible studies to participate in this IPD meta-analysis. We provided a data dictionary listing definitions of variables requested for pooled analysis. For further details, see Dewey et al. (12). The variables requested for this IPD meta-analysis were (1) intervention group, as determined by each trial design, (2) randomization cluster, if cluster-randomized, (3) child sex, (4) child age at developmental assessment, (5) whether each motor milestone had been attained by the child at the time of assessment, (6) continuous unstandardized developmental outcome scores of interest measured at baseline (prior to child supplementation) and post-supplementation, as available, calculated according to the established method for the tool used in each study, and (7) indicator variables for potential effect modifiers, as pre-specified in the analysis plan. Study-level effect modifiers included variables reflecting sample characteristics and study design (Box 1). Individual-level effect modifiers included maternal, child, and household characteristics (Box 1).

#### Box 1.

Potential effect modifiers^1^

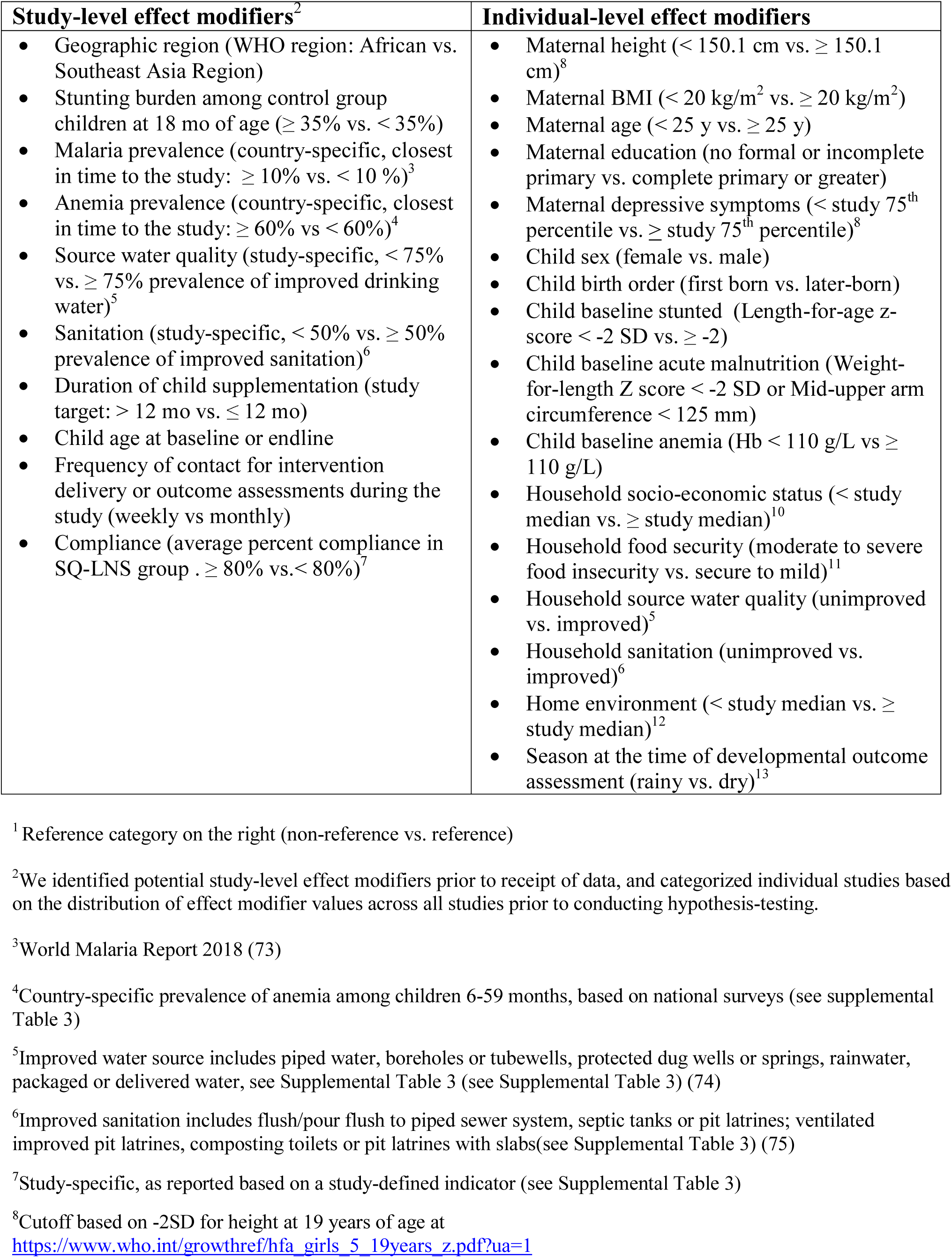
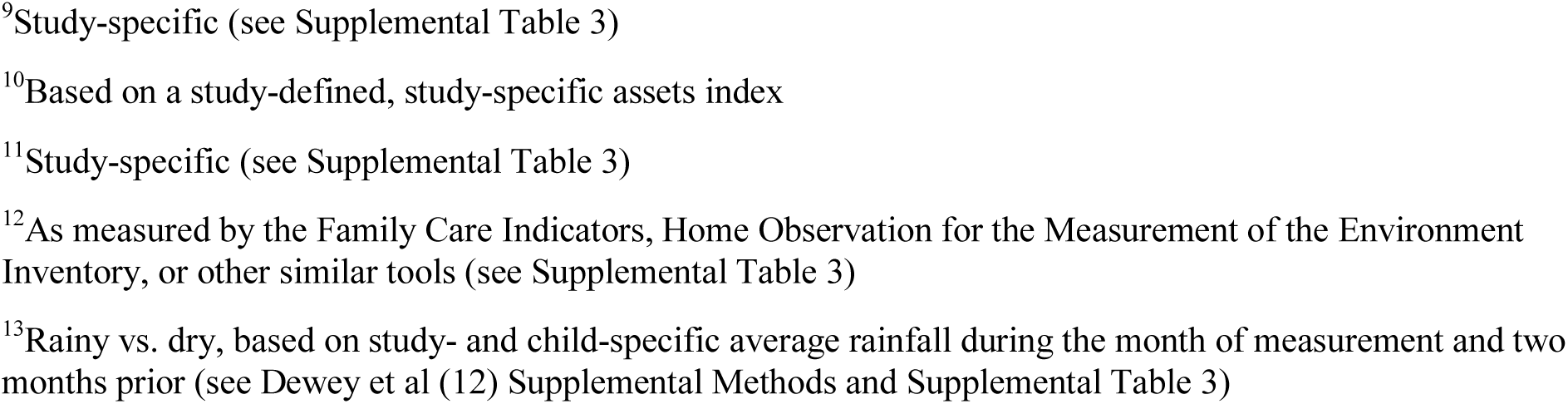

| Study-level effect modifiers<sup>2</sup> | Individual-level effect modifiers |
| --- | --- |
| • Geographic region (WHO region: African vs. Southeast Asia Region)<br>• Stunting burden among control group children at 18 mo of age ($\geq 35\%$ vs. $< 35\%$)<br>• Malaria prevalence (country-specific, closest in time to the study: $\geq 10\%$ vs. $< 10\%$)<sup>3</sup><br>• Anemia prevalence (country-specific, closest in time to the study: $\geq 60\%$ vs. $< 60\%$)<sup>4</sup><br>• Source water quality (study-specific, $< 75\%$ vs. $\geq 75\%$ prevalence of improved drinking water)<sup>5</sup><br>• Sanitation (study-specific, $< 50\%$ vs. $\geq 50\%$ prevalence of improved sanitation)<sup>6</sup><br>• Duration of child supplementation (study target: $> 12$ mo vs. $\leq 12$ mo)<br>• Child age at baseline or endline<br>• Frequency of contact for intervention delivery or outcome assessments during the study (weekly vs monthly)<br>• Compliance (average percent compliance in SQ-LNS group . $\geq 80\%$ vs. $< 80\%$)<sup>7</sup> | • Maternal height ($< 150.1$ cm vs. $\geq 150.1$ cm)<sup>8</sup><br>• Maternal BMI ($< 20$ kg/m<sup>2</sup> vs. $\geq 20$ kg/m<sup>2</sup>)<br>• Maternal age ($< 25$ y vs. $\geq 25$ y)<br>• Maternal education (no formal or incomplete primary vs. complete primary or greater)<br>• Maternal depressive symptoms ($<$ study 75<sup>th</sup> percentile vs. ≥ study 75<sup>th</sup> percentile)<sup>8</sup><br>• Child sex (female vs. male)<br>• Child birth order (first born vs. later-born)<br>• Child baseline stunted (Length-for-age z-score $< -2$ SD vs. $\geq -2$)<br>• Child baseline acute malnutrition (Weight-for-length Z score $< -2$ SD or Mid-upper arm circumference $< 125$ mm)<br>• Child baseline anemia (Hb $< 110$ g/L vs $\geq 110$ g/L)<br>• Household socio-economic status ($<$ study median vs. ≥ study median)<sup>10</sup><br>• Household food security (moderate to severe food insecurity vs. secure to mild)<sup>11</sup><br>• Household source water quality (unimproved vs. improved)<sup>5</sup><br>• Household sanitation (unimproved vs. improved)<sup>6</sup><br>• Home environment ($<$ study median vs. ≥ study median)<sup>12</sup><br>• Season at the time of developmental outcome assessment (rainy vs. dry)<sup>13</sup> |

<sup>1</sup> Reference category on the right (non-reference vs. reference)
<sup>2</sup>We identified potential study-level effect modifiers prior to receipt of data, and categorized individual studies based on the distribution of effect modifier values across all studies prior to conducting hypothesis-testing.
<sup>3</sup>World Malaria Report 2018 (73)
<sup>4</sup>Country-specific prevalence of anemia among children 6-59 months, based on national surveys (see supplemental Table 3)
<sup>5</sup>Improved water source includes piped water, boreholes or tubewells, protected dug wells or springs, rainwater, packaged or delivered water, see Supplemental Table 3 (see Supplemental Table 3) (74)
<sup>6</sup>Improved sanitation includes flush/pour flush to piped sewer system, septic tanks or pit latrines; ventilated improved pit latrines, composting toilets or pit latrines with slabs(see Supplemental Table 3) (75)
<sup>7</sup>Study-specific, as reported based on a study-defined indicator (see Supplemental Table 3)
<sup>8</sup>Cutoff based on -2SD for height at 19 years of age at https://www.who.int/growthref/hfa_girls_5_19years_z.pdf?ua=1
<sup>9</sup>Study-specific (see Supplemental Table 3) <sup>10</sup>Based on a study-defined, study-specific assets index <sup>11</sup>Study-specific (see Supplemental Table 3) <sup>12</sup>As measured by the Family Care Indicators, Home Observation for the Measurement of the Environment Inventory, or other similar tools (see Supplemental Table 3) <sup>13</sup>Rainy vs. dry, based on study- and child-specific average rainfall during the month of measurement and two months prior (see Dewey et al (12) Supplemental Methods and Supplemental Table 3)

### IPD integrity

We checked data for completeness by ensuring that the study sample sizes in our pooled dataset were the same as in study protocols and publications. We also checked summary statistics, such as means and standard deviations (SD), in our dataset against published values for each trial to ensure consistency. Implausible values were inspected for errors and truncated to 5 or −5 SD from the mean z-score (≤ 0.2% of values for each outcome, with the highest percentage for motor, gross and fine motor scores: 0.18-0.20%).

### Assessment of risk of bias and quality of evidence in each study and across studies

Independent reviewers (KRW, CDA, ELP) assessed risk of bias in each trial against the following criteria: random sequence generation and allocation concealment (selection bias), blinding of participants and personnel (performance bias), blinding of outcome assessment (detection bias), incomplete outcome data (attrition bias), selective reporting (reporting bias), and other sources of bias (13). Any discrepancies were resolved by discussion or consultation with the core working group, as needed. To assess risk of bias across studies, for each outcome that was measured in a sub-set of studies, we compared study-level maternal education (% completed primary) and child 18-month stunting burden to check for substantial differences between trials included and trials excluded. The same reviewers also assessed the quality of evidence for each outcome across all studies based on the five GRADE criteria: risk of bias, inconsistency of effect, imprecision, indirectness, and publication bias (14).

### Specification of outcomes and effect measures

The following primary and secondary outcomes were pre-specified in the analysis plan. Primary outcomes were language, motor, and social-emotional z-scores reported on a continuous scale; whether the child was in the lowest decile of continuous language z-scores, motor z-scores, and social-emotional z-scores; and walking without support at 12 months. Secondary outcomes were continuous gross and fine motor z-scores if these were reported separately, and continuous executive function z-scores and whether the child was in the lowest decile of executive function z-scores, plus whether the child had achieved four motor milestones at 12 months (crawling, standing with support, standing without support and walking with support) and five motor milestones at 18 months (the same four plus walking without support).

Z-scores were standardized within each study by regressing the unstandardized developmental score on child age and sex and calculating the standardized residuals. This approach is analogous to calculating length-for-age z-score (LAZ) in that the score represents deviations from the mean score for a given child’s age and sex in units of standard deviation. However, developmental outcome z-scores were calculated in reference to each within-study distribution, rather than an external standard. For example, a female child with a language z-score of −1 scored one SD below the mean of other female children of the same age in her study sample.

Lowest decile of each z-score was also defined for each study based on the within-study distribution. Given that most of the developmental assessment tools used in these studies do not have validated cut-offs to identify children at risk for delayed neurodevelopment, we used the lowest decile of scores as a proxy for children who may be at risk of experiencing developmental delay. We thus considered the lowest decile of scores to be an adverse developmental outcome. We selected the lowest decile as the cut-off because it is sufficiently low to capture poor development in populations that as a whole may lag behind populations with no environmental constraints on achieving developmental potential, and high enough to allow adequate power to detect group differences.

If any study used multiple tools or scores to assess the same domain at endline (e.g., language), we selected the tool or score that was used in the greatest number of other studies included in this IPD meta-analysis. For social-emotional scores, if any study reported a social-emotional difficulties score for which a higher score indicated greater problems, we reversed those scores so that for all scores, a higher score represented greater social-emotional competence. For the milestone assessment, we only used reports or observations of the child’s ability on the day of assessment, not retrospective reports of age of milestone achievement, as the latter is subject to potential recall inaccuracy. We used milestone data collected within one month of the target age (12 or 18 months). If both observation and parent report data existed at the same time point, we used observation data.

The principal measure of effect for continuous outcomes was the mean difference between intervention and comparison groups at endline, defined as the principal post-intervention time point as reported for trials with infrequent child assessment or at an age closest to the end of the supplementation period for trials with monthly child assessment. The principal measure of effect for binary outcomes was prevalence ratio at endline or at targeted age of milestone assessment (12 or 18 months). We also estimated prevalence differences as secondary assessments of binary outcomes. Prevalence ratios quantify the relative difference in proportions between groups, while prevalence differences are the difference in absolute percentage points and are less consistent than prevalence ratios (13).

The treatment and comparisons of interest were provision of children with SQ-LNS (<∼125 kcal/d, with or without co-interventions), compared to children who received no intervention or an intervention without any type of LNS or other child nutritional supplement (herein labeled “control”). Examples of other types of interventions that have been delivered with or without LNS are water, sanitation, and hygiene (WASH) interventions or child morbidity monitoring and treatment. In several trials, child LNS has been delivered to children whose mothers received maternal LNS during pregnancy and postpartum. Given that maternal supplementation may have an additive effect, we originally planned to include trial arms that provided both maternal and child LNS in a sensitivity analysis only (i.e., the all-trials analysis). However, to maximize study inclusion and participant sample size, we decided post-hoc that the results of the all-trials analysis would be presented as the principal findings if the following criteria were met, as determined for each outcome: if the main effects did not differ between the child-LNS-only analysis (excluding maternal plus child LNS arms) and the all-trials analysis (including maternal plus child LNS arms) by more than 20% for continuous outcomes or by 0.05 for prevalence ratios. Two additional sensitivity analyses were also conducted, as described below.

### Synthesis methods and exploration of variation in effects

We conducted three types of analyses, corresponding to the three objectives, to investigate: 1) full sample main effects of the intervention, 2) effect modification by study-level characteristics, and 3) effect modification by individual-level characteristics. We used a two-stage approach for all analyses. This approach is preferred when incorporating cluster-randomized trials as it allows intra-cluster correlations to be study specific (8). All analyses followed a complete case intention-to-treat framework (15).

In the first stage, we estimated intervention versus control group effects (mean differences or prevalence ratios) within each individual study. Given that continuous outcomes represented deviations from the study sample mean score for a given child’s age and sex in units of SD, these first stage individual study estimates represent mean differences between SQ-LNS and control groups in units of SD. For longitudinal study designs that provided baseline developmental assessment data, we adjusted for baseline score when estimating intervention effect. For cluster-randomized trials, we used robust standard errors to account for participant dependence within clusters.

In the second stage, first stage estimates were pooled using inverse-variance weighted fixed effects. We also conducted a sensitivity analysis calculating pooled estimates using inverse variance weighted random effects (16, 17). If there were fewer than three comparisons to include in a pooled estimate then the pooled estimate was not generated (e.g., if fewer than 3 comparisons were represented within a study-level effect modification category). This was the case for most of the milestones, therefore we did not examine study-level or individual-level effect modification for any of the 9 individual milestones specified as secondary outcomes.

For Objective 1, we pooled the first stage estimates to generate a pooled point estimate, 95% confidence interval, and corresponding p-value. For Objective 2, we used a bivariate random effects meta-regression to test the association of study-level characteristics with study intervention effect estimates. For Objective 3, we first estimated the parameter corresponding to the interaction term of the effect modifier and the intervention for each study (18). We then generated pooled intervention effect estimates within each category of the effect modifier to determine how the intervention effect in one subgroup differed from the intervention effect in the specified reference subgroup. For further details, see Dewey et al. (12).

Heterogeneity of effect estimates was assessed using I^2^ and Tau^2^ statistics, within strata when relevant (19). We used a p-value of < 0.05 for main effects and a p-for-interaction < 0.10 for effect modification. We did not adjust for multiple hypothesis testing because developmental outcomes are inter-related and the effect modification analyses are inherently exploratory (20).

### Sensitivity analyses

Two sensitivity analyses were conducted in addition to those described above (the child-LNS only analysis, all-trials analysis, and fixed and random effects models). First, we excluded passive control arms, defined as groups of participants who received no intervention and had no contact with project staff between enrollment and endline. Second, we separated comparisons within trials that included multi-component interventions to attempt to isolate the effect of SQ-LNS. For example, if a trial provided a water intervention to one group, a water plus sanitation intervention to a second group, and a water and sanitation plus SQ-LNS intervention to a third group, this sensitivity analysis would only compare the water and sanitation plus SQ-LNS arm to the water and sanitation arm. The all-trials analysis and child-LNS-only analysis would include both groups that did not receive SQ-LNS in the comparison group, while the sensitivity analysis would exclude the group that received the water intervention only. Behavior change communication and other messaging promoting recommended infant and young child feeding practices were not considered additional components. **Supplemental Table 1** lists all trial arms and specifies which comparisons were made in each sensitivity analysis.

## Results

### Literature search and trial characteristics

Of the 1466 publications identified through the search strategy and review of other meta-analyses and systematic reviews, 90 titles and abstracts were identified as relevant. Based on review of the full texts, 14 trials met the inclusion criteria and IPD were requested (21–35). Investigators for one trial were unable to participate (35). In that trial, only fine and gross motor outcomes were reported, therefore we examined pooled main effects on these two outcomes both without and with this trial, by calculating *Hedges’ g* (36) based on endline values extracted from the published report. For all other analyses, 13 trials were included in the IPD meta-analysis for developmental outcomes (21, 25, 29, 32, 37–46) (**Figure 1**). One trial, SHINE in Zimbabwe (34, 45), contributed two comparisons because it was designed a priori to report results separately for HIV-exposed and HIV-unexposed children. Thus, 14 SQ-LNS versus control group comparisons from 13 trials were analyzed. Of the 14 comparisons, language outcomes were reported for 13 comparisons, motor outcomes for 12, social-emotional outcomes for 11, executive function for 7, and various motor milestones for 7-10 comparisons.

**Figure 1.**
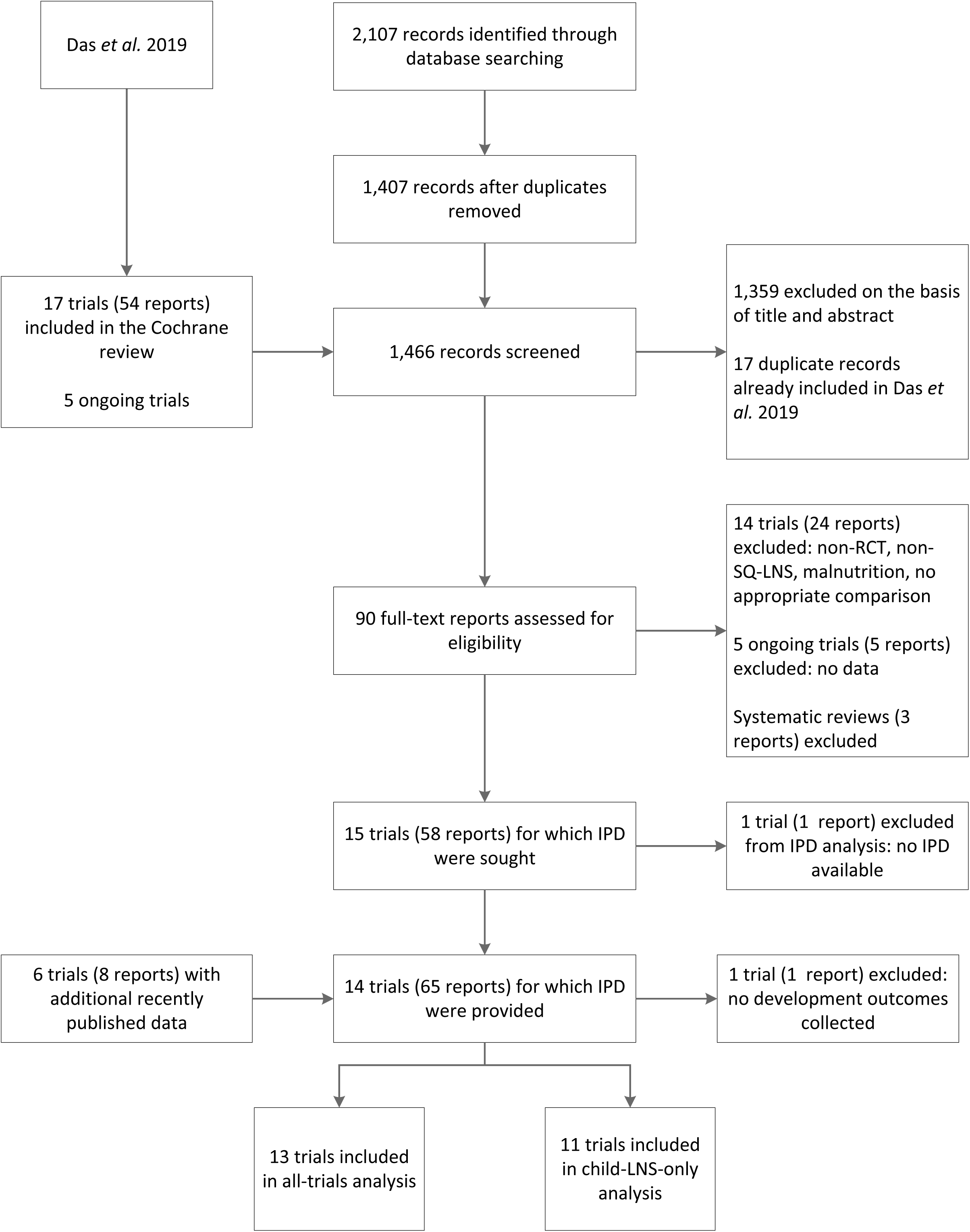
Study inclusion flow diagram

The included trials were conducted in Bangladesh (3 trials), Burkina Faso (1 trial), Ghana (2 trials), Haiti (1 trial), Kenya (1 trial), Madagascar (1 trial), Malawi (2 trials), Mali (1 trial), and Zimbabwe (1 trial). Child SQ-LNS was given starting at age 6 months in 11 trials, 6-11 months in 2 trials, and 9 months in 1 trial (**Table 1)**. Duration of supplementation ranged from 6 to 18 months. Four trials (22, 26, 29, 30) included intervention arms that provided SQ-LNS to mothers during pregnancy and/or the first 6 months postpartum. The majority of trials provided a peanut- and milk-based SQ-LNS providing approximately 120 kcal/d and 1 recommended daily allowance of most micronutrients (for further details see **Supplemental Table 2**). Two trials targeted a sub-sample of children for developmental assessment (methods described in Supplemental Table 1).

**Table 1.** Characteristics of trials included in the individual participant data analysis

| Country | Reference | Trial name | Infant SQ-LNS Supplement |  | Maternal LNS Supplement | Participants (n) |  |
| --- | --- | --- | --- | --- | --- | --- | --- |
|  |  |  | Age at start (mo) | Duration (mo) |  | Any growth, developmental, or biomarker outcome | Any developmental outcome |
| Bangladesh | Christian 2015 (21) | JiVitA-4 | 6 | 12 | N | 4568 | 4558 |
| Bangladesh | Dewey 2017 (22, 37) | RDNS | 6 | 18 | Y/N | 2567 | 2565 |
| Bangladesh | Luby 2018 (23, 38) | WASH B-B | 6 | 18 | N | 4824 | 4572 |
| Burkina Faso | Hess 2015 (24, 39) | iLiNS-Zinc | 9 | 9 | N | 2647 | 1176 |
| Ghana | Adu-Afarwuah 2007 (25) | GHANA | 6 | 12 | Y | 194 | 194 |
| Ghana | Adu-Afarwuah 2016 (26, 40) | iLiNS-DYAD-G | 6 | 6 | N | 1113 | 1103 |
| Haiti | Iannotti 2014 (27, 41) | HAITI | 6-11 | 3-6 | N | 322 | 322 |
| Kenya | Null 2018 (28, 42) | WASH B-K | 6 | 18 | N | 6815 | 6756 |
| Madagascar | Galasso 2019 (29) | MAHAY | 6-11 | 6-12 | Y/N | 3438 | 3217 |
| Malawi | Ashorn 2015 (30, 43) | iLiNS-DYAD-M | 6 | 12 | Y | 675 | 674 |
| Malawi | Maleta 2015 (31, 44) | iLiNS-Dose | 6 | 12 | N | 999 | 999 |
| Mali | Huybregts 2019 (32) | PROMIS | 6 | 18 | N | 1927<sup>1</sup> | 1927 |
| Zimbabwe | Humphrey 2019 (33, 45) | SHINE | 6 | 12 | N | 4347 | 1961 |
|  | Prendergast 2019 (34, 46) |  |  |  |  |  |  |

<sup>1</sup>Developmental assessments were conducted only in the cross-sectional sample, not the longitudinal sample, therefore the longitudinal sample was not included in the participant count.

The most commonly used developmental assessment tools were the MacArthur-Bates Communicative Development Inventory (CDI) vocabulary checklist to assess language (6 trials; Supplemental Table 1) and the A not B task to assess executive function (6 trials; all trials that measured executive function used this same task). Other tools were the Developmental Milestones Checklist (DMC; 3 trials), Extended Ages and Stages Questionnaire (EASQ; 2 trials), ASQ-Inventory (ASQI; 1 trial), Kilifi Development Inventory (KDI; 3 trials), Malawi Developmental Assessment Tool (MDAT; 1 trial), and Bayley Scales of Infant Development-III (BSID; 1 trial). All endline assessments were conducted when the children were ages 12 to 24 months. In this age range, all of these tools assess similar developmental skills and many items overlap between the tools. Parent-report was used to assess social-emotional development in all studies and to assess language in all studies except one. Direct child assessment was used to assess executive function in all studies. Motor development was assessed by parent report in 6 studies and direct child assessment in 6 studies.

All potential study-level and individual-level effect modifiers showed substantial variation between trials (**Supplemental Tables 3** and **4**). For example, at the study level, 8 study sites had a high burden of stunting (≥ 35% at 18 mo) and 5 had lower rates of stunting (< 35% at 18 mo). Study-specific prevalence of improved water quality ranged from 27% to 100%, and prevalence of improved sanitation ranged from 0% to 97%. Frequency of contact during the study was weekly in 7 trials and monthly in 6 trials. Average estimated reported compliance with SQ-LNS consumption was categorized as high (≥ 80%) in 7 trials and lower than that in the other trials.

### Main effects of SQ-LNS on developmental outcomes

Results from the child-LNS-only and all-trials analyses were similar for all outcomes (**Supplemental Figures 2A-G**). Therefore, results from the all-trials analyses, inclusive of maternal plus child LNS trials and arms, are presented in **Table 2**. SQ-LNS had a significant positive effect on all primary developmental outcomes, with effect sizes 0.07 to 0.08 SD in mean language, social-emotional, and motor scores (Table 2; **Supplemental Figures 3A-AB**), and relative reductions in the percentage of children in the lowest decile of these scores ranging from 16% to 19% (Table 2; **Figures 2, 3,** and **4**). For the prevalence of children walking without support at 12 months, there was a relative increase of 9% (4 percentage point difference; Table 2; **Figure 5**). In the JiVitA-4 trial, milestone data were collected using monthly surveillance rather than data collection at a single time point, the method used in all other trials. This trial also contributed 30% of the total sample size. Therefore, we conducted a sensitivity analysis excluding the data from this trial and found a similar estimate of an increase of 13% (95% CI 1.07-1.20) in the prevalence of children walking without support at 12 months.

**Figure 2.**
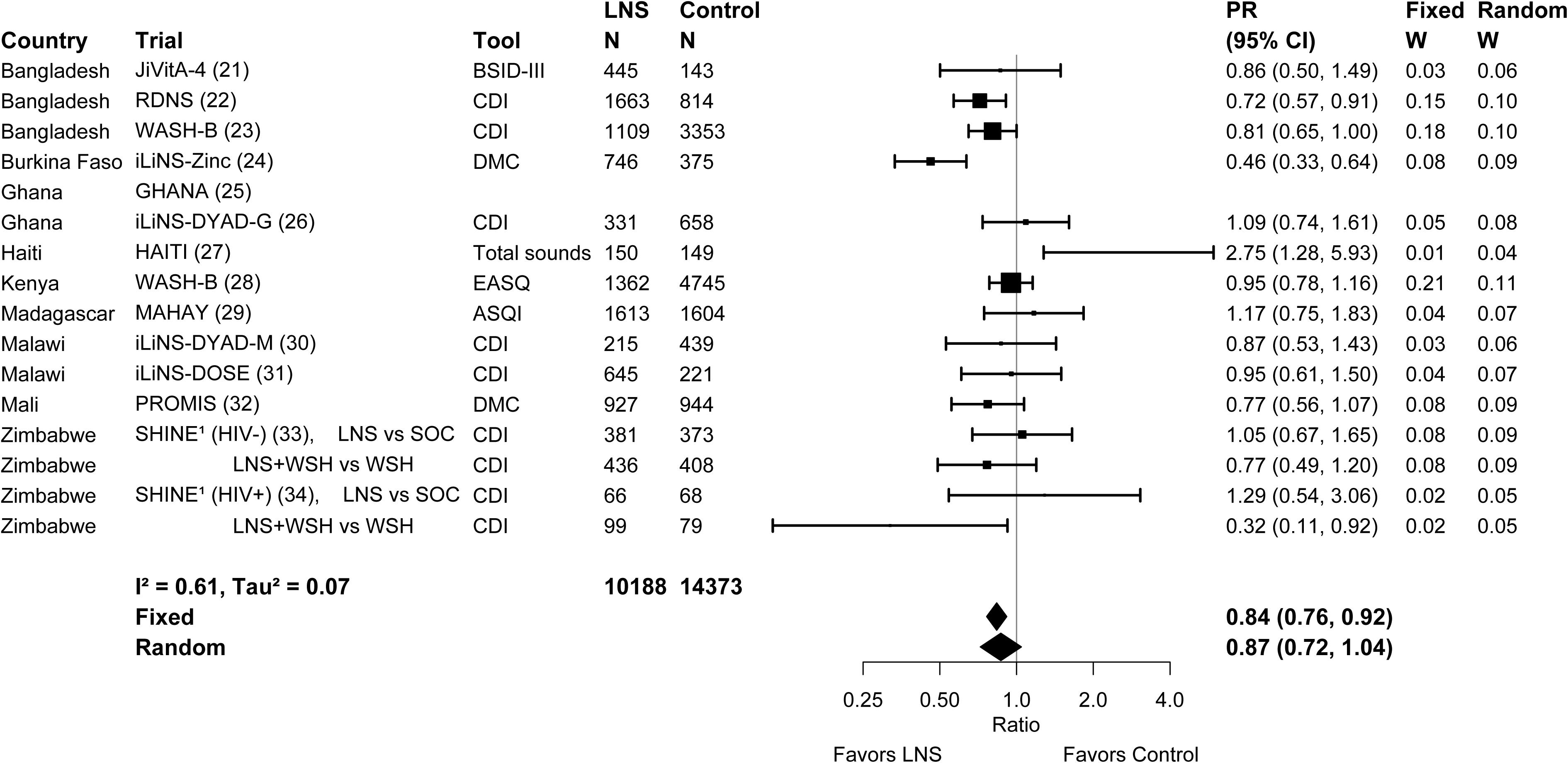
Forest plot of effect of SQ-LNS on prevalence of children in the lowest decile of language scores

**Figure 3.**
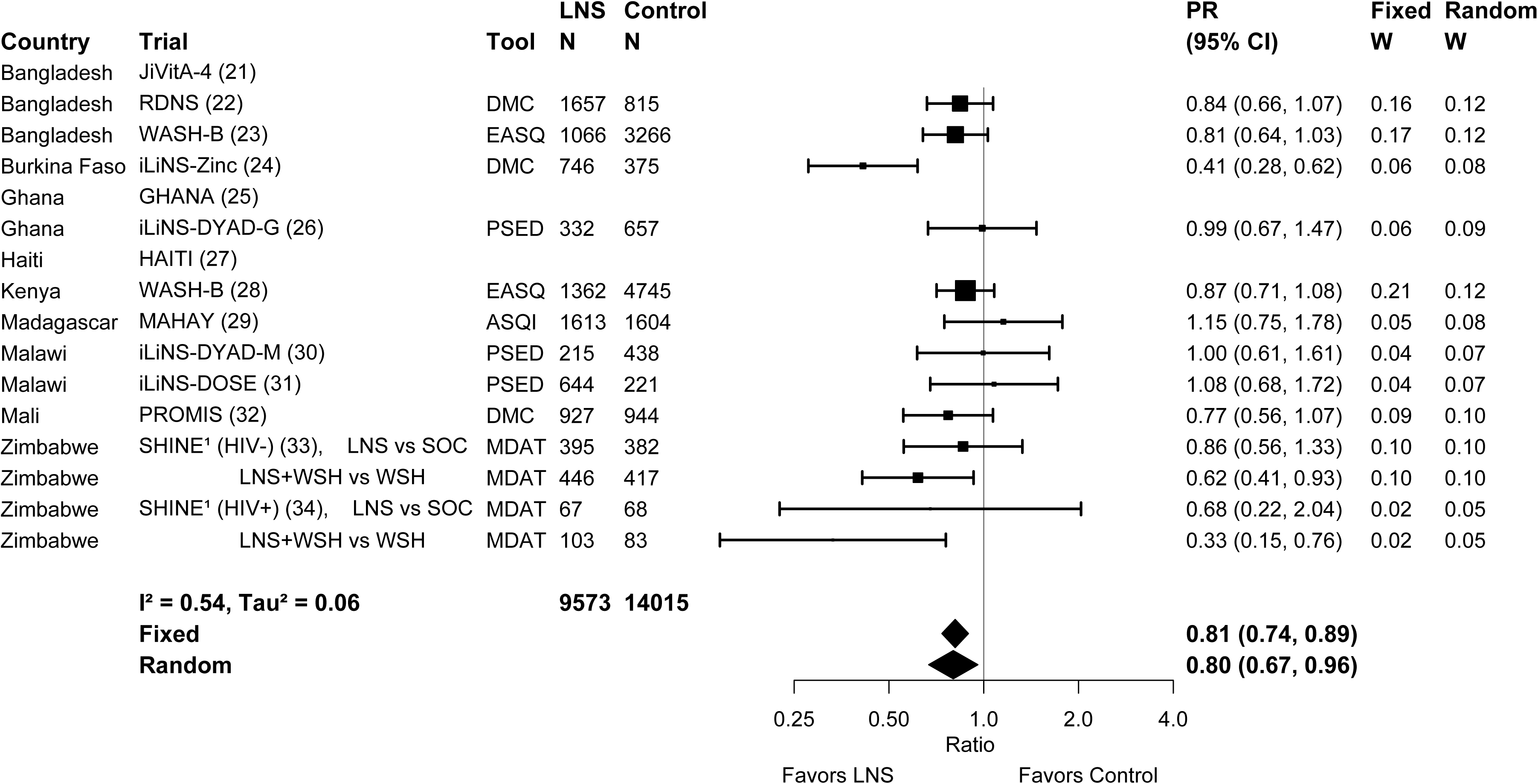
Forest plot of effect of SQ-LNS on prevalence of children in the lowest decile of social-emotional scores

**Figure 4.**
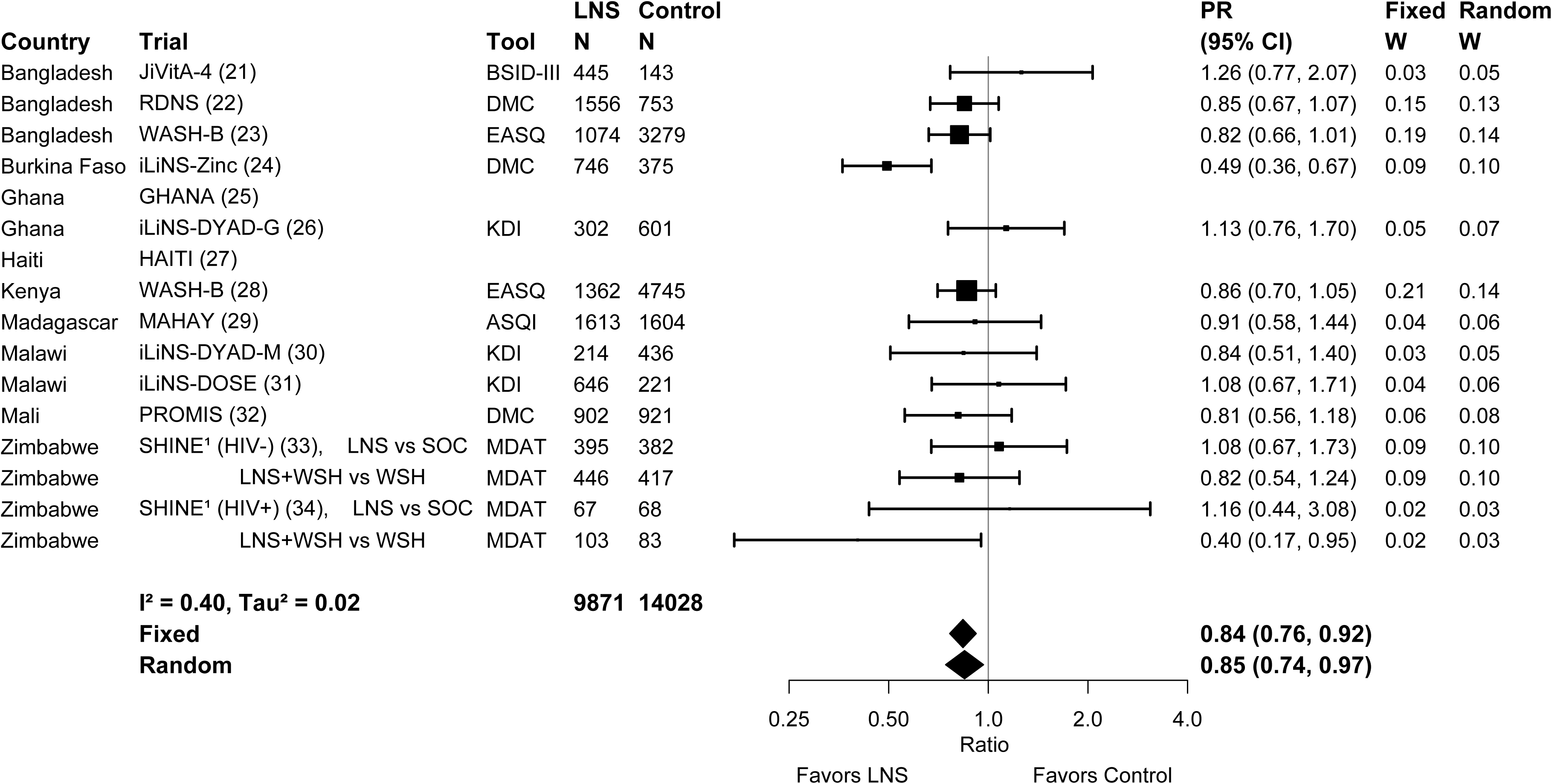
Forest plot of effect of SQ-LNS on prevalence of children in the lowest decile of motor scores

**Figure 5.**
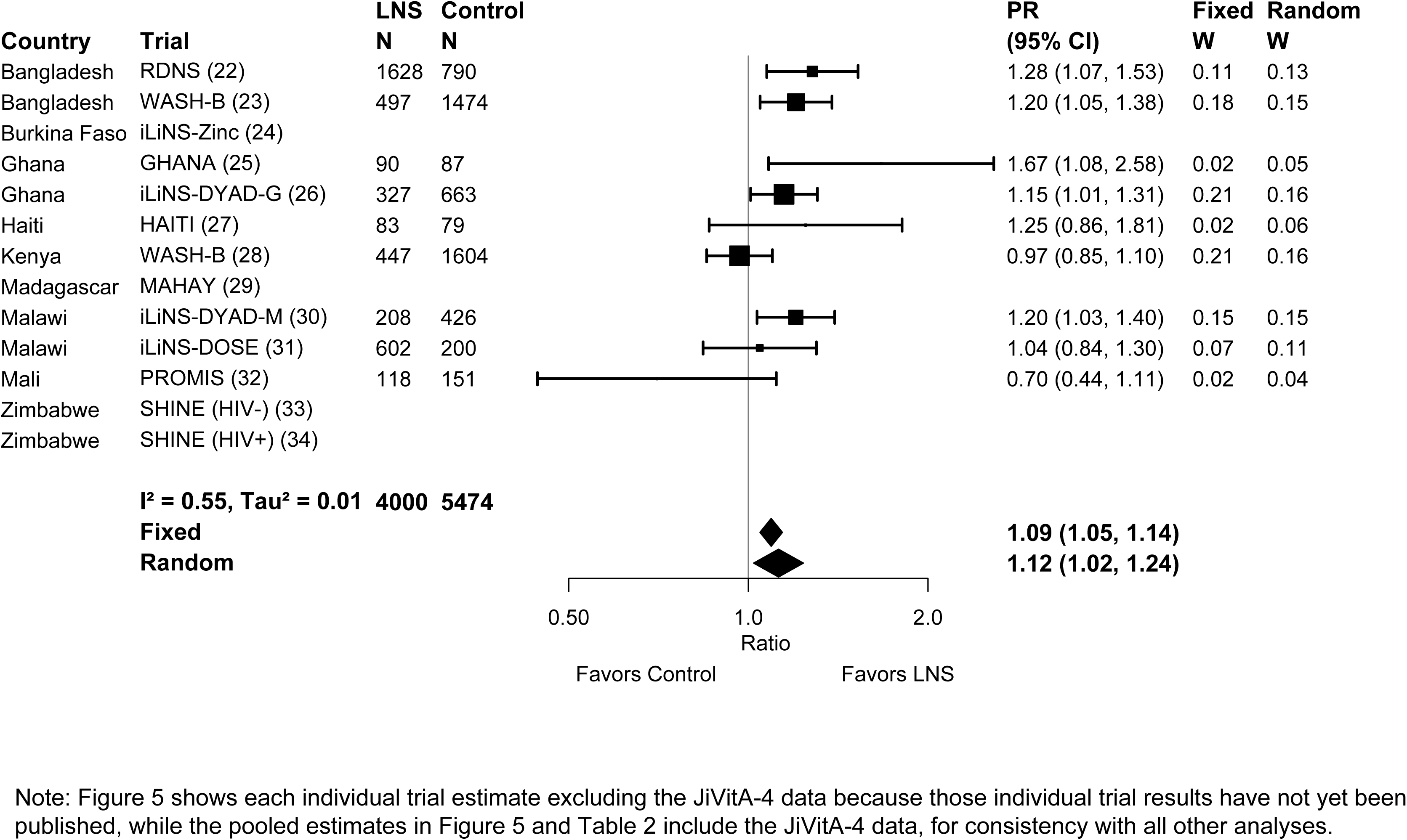
Forest plot of effect of SQ-LNS on prevalence of children walking without support at age 12 months

**Table 2.**
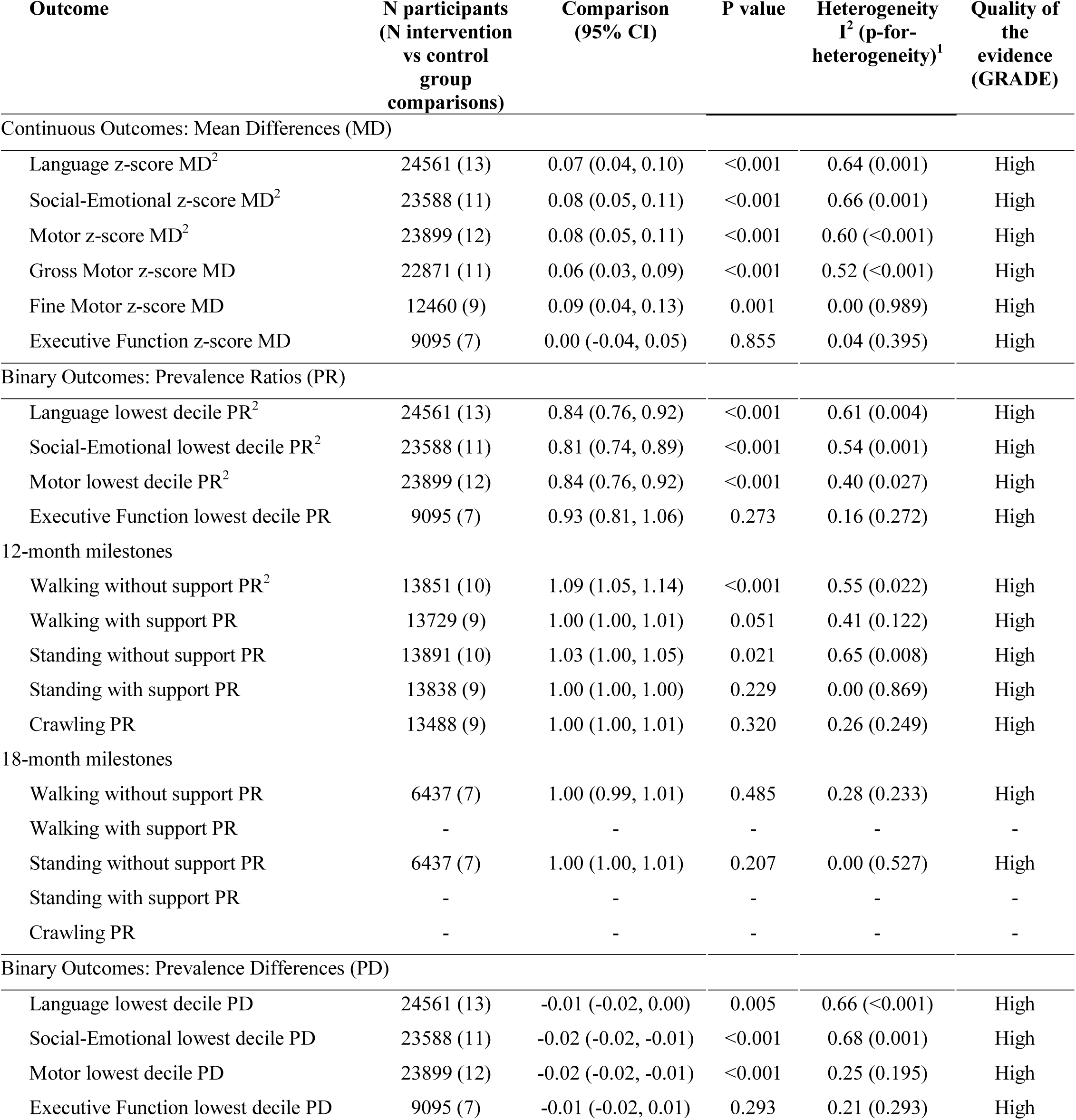
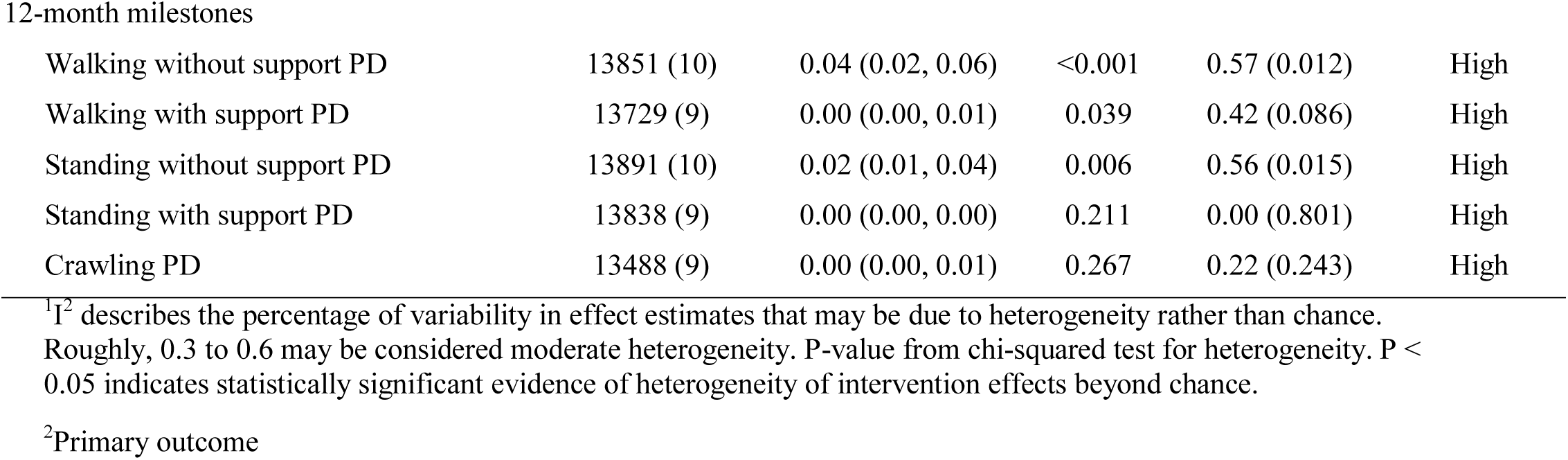
Pooled estimates of the effect of randomized controlled trials of SQ-LNS provided to infants and young children age 6 to 24 months, compared to children who received no intervention or an intervention without any nutritional supplement, on developmental outcomes

| Outcome | N participants<br>(N intervention<br>vs control<br>group<br>comparisons) | Comparison<br>(95% CI) | P value | Heterogeneity<br>I<sup>2</sup> (p-for-<br>heterogeneity)<sup>1</sup> | Quality of<br>the<br>evidence<br>(GRADE) |
| --- | --- | --- | --- | --- | --- |
| Continuous Outcomes: Mean Differences (MD) |  |  |  |  |  |
| Language z-score MD<sup>2</sup> | 24561 (13) | 0.07 (0.04, 0.10) | <0.001 | 0.64 (0.001) | High |
| Social-Emotional z-score MD<sup>2</sup> | 23588 (11) | 0.08 (0.05, 0.11) | <0.001 | 0.66 (0.001) | High |
| Motor z-score MD<sup>2</sup> | 23899 (12) | 0.08 (0.05, 0.11) | <0.001 | 0.60 (<0.001) | High |
| Gross Motor z-score MD | 22871 (11) | 0.06 (0.03, 0.09) | <0.001 | 0.52 (<0.001) | High |
| Fine Motor z-score MD | 12460 (9) | 0.09 (0.04, 0.13) | 0.001 | 0.00 (0.989) | High |
| Executive Function z-score MD | 9095 (7) | 0.00 (-0.04, 0.05) | 0.855 | 0.04 (0.395) | High |
| Binary Outcomes: Prevalence Ratios (PR) |  |  |  |  |  |
| Language lowest decile PR<sup>2</sup> | 24561 (13) | 0.84 (0.76, 0.92) | <0.001 | 0.61 (0.004) | High |
| Social-Emotional lowest decile PR<sup>2</sup> | 23588 (11) | 0.81 (0.74, 0.89) | <0.001 | 0.54 (0.001) | High |
| Motor lowest decile PR<sup>2</sup> | 23899 (12) | 0.84 (0.76, 0.92) | <0.001 | 0.40 (0.027) | High |
| Executive Function lowest decile PR | 9095 (7) | 0.93 (0.81, 1.06) | 0.273 | 0.16 (0.272) | High |
| 12-month milestones |  |  |  |  |  |
| Walking without support PR<sup>2</sup> | 13851 (10) | 1.09 (1.05, 1.14) | <0.001 | 0.55 (0.022) | High |
| Walking with support PR | 13729 (9) | 1.00 (1.00, 1.01) | 0.051 | 0.41 (0.122) | High |
| Standing without support PR | 13891 (10) | 1.03 (1.00, 1.05) | 0.021 | 0.65 (0.008) | High |
| Standing with support PR | 13838 (9) | 1.00 (1.00, 1.00) | 0.229 | 0.00 (0.869) | High |
| Crawling PR | 13488 (9) | 1.00 (1.00, 1.01) | 0.320 | 0.26 (0.249) | High |
| 18-month milestones |  |  |  |  |  |
| Walking without support PR | 6437 (7) | 1.00 (0.99, 1.01) | 0.485 | 0.28 (0.233) | High |
| Walking with support PR | - | - | - | - | - |
| Standing without support PR | 6437 (7) | 1.00 (1.00, 1.01) | 0.207 | 0.00 (0.527) | High |
| Standing with support PR | - | - | - | - | - |
| Crawling PR | - | - | - | - | - |
| Binary Outcomes: Prevalence Differences (PD) |  |  |  |  |  |
| Language lowest decile PD | 24561 (13) | -0.01 (-0.02, 0.00) | 0.005 | 0.66 (<0.001) | High |
| Social-Emotional lowest decile PD | 23588 (11) | -0.02 (-0.02, -0.01) | <0.001 | 0.68 (0.001) | High |
| Motor lowest decile PD | 23899 (12) | -0.02 (-0.02, -0.01) | <0.001 | 0.25 (0.195) | High |
| Executive Function lowest decile PD | 9095 (7) | -0.01 (-0.02, 0.01) | 0.293 | 0.21 (0.293) | High |
| 12-month milestones |  |  |  |  |  |
| Walking without support PD | 13851 (10) | 0.04 (0.02, 0.06) | <0.001 | 0.57 (0.012) | High |
| Walking with support PD | 13729 (9) | 0.00 (0.00, 0.01) | 0.039 | 0.42 (0.086) | High |
| Standing without support PD | 13891 (10) | 0.02 (0.01, 0.04) | 0.006 | 0.56 (0.015) | High |
| Standing with support PD | 13838 (9) | 0.00 (0.00, 0.00) | 0.211 | 0.00 (0.801) | High |
| Crawling PD | 13488 (9) | 0.00 (0.00, 0.01) | 0.267 | 0.22 (0.243) | High |

<sup>1</sup>I<sup>2</sup> describes the percentage of variability in effect estimates that may be due to heterogeneity rather than chance. Roughly, 0.3 to 0.6 may be considered moderate heterogeneity. P-value from chi-squared test for heterogeneity. P < 0.05 indicates statistically significant evidence of heterogeneity of intervention effects beyond chance.
<sup>2</sup>Primary outcome

For the secondary outcomes, SQ-LNS had a significant positive effect on mean gross and fine motor scores of 0.06 and 0.09 SD, respectively, but no significant effect on executive function mean or percentage in the lowest decile. Including the effect estimates from the published report by Smuts et al. (35), for which IPD were not available, results were similar (gross motor: 0.06, 95% CI 0.03-0.09, fine motor: 0.09, 95% CI 0.04-0.13). For consistency with other analyses, Table 2 reports the estimates excluding this trial by Smuts et al. Including the JiVitA-4 data, SQ-LNS increased the prevalence of children standing without support at age 12 months by 3%, and no significant effects were found on any other milestones examined (Table 2). Excluding the JiVitA-4 data, SQ-LNS increased the prevalence of children standing without support at age 12 months by 6% (95% CI 1.02-1.08). Pooled estimates were not generated for 3 of the 5 18-month milestones due to lack of variance, because almost all children had attained the milestones by this age (see **Supplemental Table 5** for the percentage of children in the control arms who attained each milestone at 12 and 18 months in each trial).

Supplemental Figures 2A-G show the results of all eight analyses, that is, fixed and random effects models for each of the (1) all-trials analysis, (2) child-LNS-only analysis, (3) sensitivity analysis excluding passive arms, and (4) sensitivity analysis separating multi-component arms to compare only pairs of arms that included the same non-nutrition components. Results were similar regardless of whether fixed effects or random effects models were used, although confidence intervals were wider for the latter, as expected. Results were also similar in the sensitivity analyses. For example, across the eight analyses, effect sizes on language scores ranged from 0.05 to 0.09 and reductions in the percentage of children in the lowest decile of language ranged from 11% to 20%.

### Risk of bias and quality of evidence

In general, we rated individual trials as having low risk of bias, except for the lack of blinding of participants due to the nature of the intervention. Because of the latter, outcome assessment was not blinded when development was assessed by parent report (language and social-emotional outcomes in most trials and motor outcomes in half of the trials; **Supplemental Tables 6A-M** and **Supplemental Figure 1**). In analyses that included a subset of studies, there was a mix of high and low child stunting burden and maternal educational levels among both the included studies and the excluded studies, with the following exceptions. For social-emotional outcomes, all of the four excluded studies were among the 7 studies in the category for higher maternal education (>50% completed primary). For motor outcomes, all of the three excluded studies were in the category for higher maternal education.

For all developmental outcomes, we rated the overall quality of evidence as high. All included studies were randomized controlled trials, therefore GRADE ratings started as high and we did not downgrade the quality of the evidence based on the following five criteria. (1) Heterogeneity across trials was low to moderate (I^2^ 0.00 to 0.60) for 20 outcomes and substantial (I^2^ 0.61 to 0.68) for 5 outcomes (Table 2) therefore, inconsistency was not considered high enough to downgrade the quality of the evidence. (2) Precision was rated as high because all but 2 trials had sample sizes > 600. (3) Directness was high because all trials were directly aimed at evaluating SQ-LNS. (4) Funnel plots revealed no indication of publication bias across studies. (5) We did not consider risk of bias in individual studies high enough to downgrade the quality of the evidence. As described above, the main potential source of bias was the lack of participant blinding and therefore the lack of blinding of outcome assessment when development was assessed by parent report. Parent-report methods were used for language and social-emotional outcomes in most trials and motor outcomes in half of the trials. To explore this potential bias, we calculated pooled effect sizes for motor outcomes stratified by parent report versus directly observed assessments and found that effects of SQ-LNS were larger among studies that used parent-report (0.13, 95% CI 0.02-0.23, 6 comparisons; vs. 0.07, 95% CI −0.01-0.15, 6 comparisons for direct child observation). However, 3 studies included in this IPD meta-analysis used direct observation for at least a sub-group of items or children to check the validity of the parent-report assessments and found similar intervention effects on observed motor and language outcomes compared to the corresponding parent-report outcomes (28, 43, 45). This suggests that reporting bias did not account for the effects of SQ-LNS, at least in those 3 trials. Given this evidence and given that the pooled effect size on observed motor outcomes (0.07) was in the same range as all primary outcome pooled effect sizes (0.06 to 0.08), we did not consider that this risk of bias was high enough to downgrade our confidence in the accuracy of the pooled estimates.

### Effect modification by study-level characteristics

Study-level effect modification results were consistent across all fixed and random effects analyses and across all sensitivity analyses (data not shown, available on request). The results presented below refer to the fixed effects all-trials analysis. For some outcomes, we were unable to generate pooled estimates for effect modification by certain potential study-level effect modifiers because fewer than 3 comparisons were categorized into one of the study-level effect modification categories (e.g., social-emotional development by geographic region). We were unable to examine potential effect modification by child age at baseline because there was insufficient heterogeneity in this aspect of study design: most of the trials began supplementation at 6 months of age.

The study-level stunting burden significantly modified the effect of SQ-LNS on language, social-emotional, motor, and gross motor development. Among studies with higher 18-month stunting burden in the control group (≥ 35%), effects on these developmental scores ranged from 0.08 to 0.13 SD, while effect sizes among studies with lower stunting burden (< 35%) were 0.01 SD (**Table 3**; **Figure 6**). There was also a greater reduction in the prevalence of children in the lowest decile of language scores among studies with higher stunting burden (**Table 4**).

**Figure 6.**
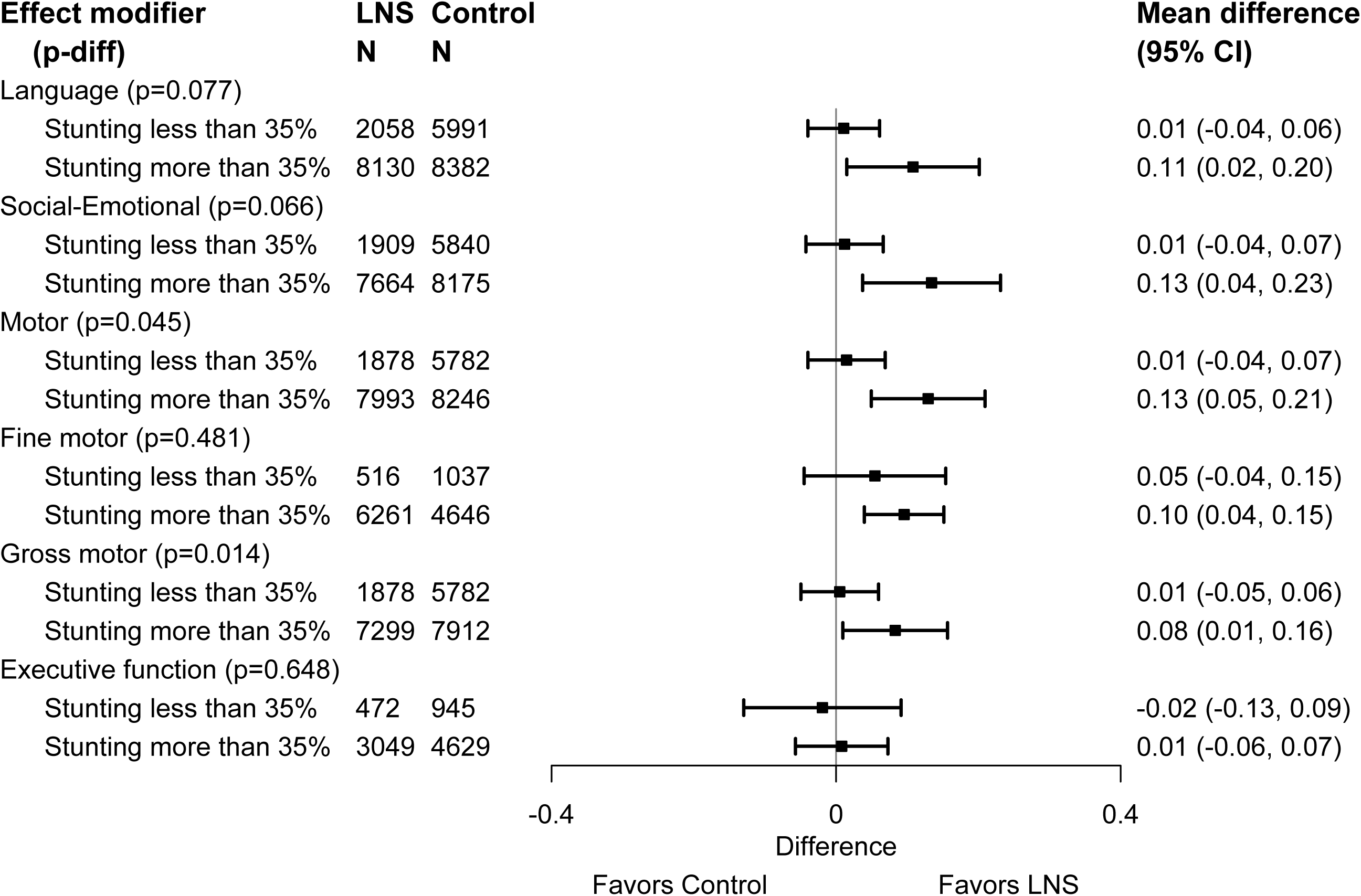
Pooled effects of SQ-LNS on all continuous developmental outcomes stratified by study-level stunting burden of children at age 18 months in control groups

**Table 3.** Study-level effect modifiers of effects of SQ-LNS provided to infants and young children age 6 to 24 months on continuous developmental outcomes

|  | Language |  | Social-Emotional |  | Executive Function |  | Motor |  | Fine Motor |  | Gross Motor |  |
| --- | --- | --- | --- | --- | --- | --- | --- | --- | --- | --- | --- | --- |
|  | N<br>(comp)<sup>1</sup> | Pooled MD<sup>2</sup><br>(95% CI) | N<br>(comp) | Pooled MD<br>(95% CI) | N<br>(comp) | Pooled MD<br>(95% CI) | N<br>(comp) | Pooled MD<br>(95% CI) | N<br>(comp) | Pooled MD<br>(95% CI) | N<br>(comp) | Pooled MD<br>(95% CI) |
| Geographic region |  | P-Diff:<sup>3</sup><br>0.978 |  | P-Diff:<br>- |  | P-Diff:<br>- |  | P-Diff:<br>0.591 |  | P-Diff:<br>- |  | P-Diff:<br>0.929 |
| Africa | 16735<br>(9) | 0.08<br>(-0.01, 0.17) | 16784<br>(9) | 0.10<br>(0.00, 0.20) | 3872<br>(5) | -0.02<br>(-0.09, 0.05) | 16649<br>(9) | 0.11<br>(0.03, 0.20) | 9427<br>(7) | 0.08<br>(0.02, 0.13) | 15602<br>(8) | 0.06<br>(-0.01, 0.14) |
| Southeast Asia | 7527 (3) | 0.08<br>(-0.04, 0.21) | 6804<br>(2) | - | 5223<br>(2) | - | 7250<br>(3) | 0.09<br>(0.04, 0.14) | 3033<br>(2) | - | 7269<br>(3) | 0.05<br>(-0.07, 0.16) |
| 18-mo stunting burden |  | P-Diff:<br>0.077 |  | P-Diff:<br>0.066 |  | P-Diff:<br>- |  | P-Diff:<br>0.045 |  | P-Diff:<br>- |  | P-Diff:<br>0.014 |
| < 35% | 8049 (4) | 0.01<br>(-0.04, 0.06) | 7749<br>(3) | 0.01<br>(-0.04, 0.07) | 1417<br>(2) | - | 7660<br>(3) | 0.01<br>(-0.04, 0.07) | 1553<br>(2) | - | 7660<br>(3) | 0.01<br>(-0.05, 0.06) |
| ≥ 35% | 16512<br>(9) | 0.11<br>(0.02, 0.20) | 15839<br>(8) | 0.13<br>(0.04, 0.23) | 7678<br>(5) | 0.01<br>(-0.06, 0.07) | 16239<br>(9) | 0.13<br>(0.05, 0.21) | 10907<br>(7) | 0.10<br>(0.04, 0.15) | 15211<br>(8) | 0.08<br>(0.01, 0.16) |
| Malaria prevalence |  | P-Diff:<br>0.365 |  | P-Diff:<br>0.863 |  | P-Diff:<br>0.242 |  | P-Diff:<br>0.557 |  | P-Diff:<br>0.626 |  | P-Diff:<br>0.387 |
| < 10% | 19060<br>(8) | 0.05<br>(-0.03, 0.13) | 18089<br>(6) | 0.09<br>(-0.01, 0.20) | 7039<br>(4) | 0.02<br>(-0.04, 0.08) | 18535<br>(7) | 0.08<br>(0.02, 0.14) | 8211<br>(5) | 0.10<br>(0.03, 0.16) | 18554<br>(7) | 0.08<br>(-0.01, 0.17) |
| ≥ 10% | 5501 (5) | 0.11<br>(-0.02, 0.25) | 5499<br>(5) | 0.11<br>(-0.02, 0.23) | 2056<br>(3) | -0.04<br>(-0.14, 0.05) | 5364<br>(5) | 0.12<br>(-0.01, 0.26) | 4249<br>(4) | 0.07<br>(-0.01, 0.15) | 4317<br>(4) | 0.04<br>(-0.02, 0.10) |
| Anemia prevalence |  | P-Diff:<br>0.530 |  | P-Diff:<br>0.496 |  | P-Diff:<br>- |  | P-Diff:<br>0.269 |  | P-Diff:<br>0.928 |  | P-Diff:<br>0.558 |
| < 60% | 19750<br>(8) | 0.06<br>(-0.01, 0.13) | 19078<br>(7) | 0.08<br>(-0.01, 0.17) | 7905<br>(5) | 0.02<br>(-0.03, 0.07) | 19438<br>(8) | 0.07<br>(0.02, 0.13) | 9114<br>(6) | 0.08<br>(0.03, 0.14) | 19457<br>(8) | 0.07<br>(0.00, 0.15) |
| ≥ 60% | 4811 (5) | 0.10<br>(-0.06, 0.26) | 4510<br>(4) | 0.13<br>(-0.02, 0.29) | 1190<br>(2) | - | 4461<br>(4) | 0.15<br>(-0.02, 0.32) | 3346<br>(3) | 0.09<br>(-0.01, 0.19) | 3414<br>(3) | 0.05<br>(-0.02, 0.12) |
| Source water quality |  | P-Diff:<br>0.536 |  | P-Diff:<br>0.681 |  | P-Diff:<br>- |  | P-Diff:<br>0.530 |  | P-Diff:<br>0.234 |  | P-Diff:<br>0.777 |
| < 75% improved | 9819 (6) | 0.09<br>(-0.04, 0.22) | 9843<br>(6) | 0.11<br>(-0.02, 0.24) | 847<br>(2) | - | 9795<br>(6) | 0.12<br>(-0.01, 0.24) | 5958<br>(4) | 0.01<br>(-0.09, 0.11) | 8748<br>(5) | 0.06<br>(0.01, 0.12) |
| ≥ 75% improved | 7520 (7) | 0.05<br>(-0.03, 0.14) | 6606<br>(5) | 0.08<br>(0.00, 0.16) | 4495<br>(5) | 0.02<br>(-0.06, 0.09) | 6944<br>(6) | 0.08<br>(0.02, 0.14) | 5453<br>(5) | 0.09<br>(0.02, 0.15) | 6963<br>(6) | 0.04<br>(-0.04, 0.12) |
| Sanitation |  | P-Diff:<br>0.991 |  | P-Diff:<br>0.660 |  | P-Diff:<br>**0.012** |  | P-Diff:<br>0.963 |  | P-Diff:<br>0.364 |  | P-Diff:<br>0.276 |
| < 50% improved | 9468 (7) | 0.07<br>(-0.04, 0.18) | 9490<br>(7) | 0.08<br>(-0.03, 0.19) | **2037**<br>**(4)** | **-0.09**<br>**(-0.18, 0.01)** | 9489<br>(7) | 0.09<br>(-0.02, 0.19) | 5646<br>(5) | 0.04<br>(-0.04, 0.12) | 8368<br>(6) | 0.03<br>(-0.03, 0.09) |
| ≥ 50% improved | 7871 (6) | 0.07<br>(-0.03, 0.17) | 6959<br>(4) | 0.11<br>(0.02, 0.20) | **3305**<br>**(3)** | **0.06**<br>**(-0.01, 0.13)** | 7250<br>(5) | 0.10<br>(0.03, 0.17) | 5765<br>(4) | 0.09<br>(0.02, 0.16) | 7343<br>(5) | 0.07<br>(-0.02, 0.16) |
| Supplement duration |  | P-Diff:<br>0.524 |  | P-Diff:<br>0.702 |  | P-Diff:<br>- |  | P-Diff:<br>0.621 |  | P-Diff:<br>- |  | P-Diff:<br>0.643 |
| ≤ 12 mo | 9644 (9) | 0.05<br>(-0.05, 0.16) | 8806<br>(7) | 0.11<br>(-0.01, 0.24) | 3872<br>(5) | -0.02<br>(-0.09, 0.05) | 9307<br>(8) | 0.11<br>(0.01, 0.22) | 8186<br>(7) | 0.08<br>(0.02, 0.14) | 8186<br>(7) | 0.05<br>(-0.05, 0.15) |
| > 12 mo | 14917<br>(4) | 0.09<br>(0.02, 0.16) | 14782<br>(4) | 0.07<br>(0.01, 0.13) | 5223<br>(2) | - | 14592<br>(4) | 0.07<br>(0.02, 0.13) | 4274<br>(2) | - | 14685<br>(4) | 0.07<br>(0.03, 0.11) |
| Frequency of contact |  | P-Diff:<br>0.853 |  | P-Diff:<br>0.897 |  | P-Diff:<br>0.192 |  | P-Diff:<br>0.932 |  | P-Diff:<br>0.791 |  | P-Diff:<br>0.150 |
| Weekly | 8680 (6) | 0.08<br>(-0.04, 0.20) | 7960<br>(5) | 0.10<br>(-0.02, 0.23) | 6480<br>(4) | -0.02<br>(-0.07, 0.04) | 8482<br>(6) | 0.10<br>(-0.02, 0.21) | 3008<br>(4) | 0.08<br>(0.00, 0.15) | 7361<br>(5) | 0.02<br>(-0.05, 0.09) |
| Monthly | 15881<br>(7) | 0.07<br>(-0.03, 0.16) | 15628<br>(6) | 0.09<br>(-0.01, 0.20) | 2615<br>(3) | 0.04<br>(-0.05, 0.13) | 15417<br>(6) | 0.09<br>(0.03, 0.16) | 9452<br>(5) | 0.09<br>(0.03, 0.15) | 15510<br>(6) | 0.10<br>(0.02, 0.17) |
| Average SQ-LNS compliance |  | P-Diff:<br>0.918 |  | P-Diff:<br>0.863 |  | P-Diff:<br>- |  | P-Diff:<br>0.898 |  | P-Diff:<br>- |  | P-Diff:<br>0.525 |
| < 80% | 6290 (6) | 0.07<br>(0.01, 0.13) | 6339<br>(6) | 0.10<br>(0.01, 0.20) | 3872<br>(5) | -0.02<br>(-0.09, 0.05) | 6204<br>(6) | 0.09<br>(0.02, 0.16) | 6210<br>(6) | 0.08<br>(0.02, 0.14) | 6278<br>(6) | 0.08<br>(-0.02, 0.18) |
| ≥ 80% | 15054<br>(6) | 0.07<br>(0.01, 0.13) | 14032<br>(4) | 0.10<br>(0.01, 0.20) | 5223<br>(2) | - | 14478<br>(5) | 0.09<br>(0.02, 0.16) | 3033<br>(2) | - | 13376<br>(4) | 0.08<br>(-0.02, 0.18) |

<sup>1</sup>N: number of individual participants; comp: number of intervention versus control group comparisons
<sup>2</sup>MD: mean difference
<sup>3</sup>P-Diff: P-value for the difference in effects of SQ-LNS between the two levels of the effect modifier.

**Table 4.** Study-level effect modifiers of effects of SQ-LNS provided, infants and young children age 6 to 24 months on binary developmental outcomes

|  | Language lowest decile |  | Social-Emotional lowest decile |  | Executive Function lowest decile |  | Motor lowest decile |  | Walking without support at 12 months |  |
| --- | --- | --- | --- | --- | --- | --- | --- | --- | --- | --- |
|  | N (comp)<sup>1</sup> | Pooled PR<sup>2</sup> (95% CI) | N (comp) | Pooled PR (95% CI) | N (comp) | Pooled PR (95% CI) | N (comp) | Pooled PR (95% CI) | N (comp) | Pooled PR (95% CI) |
| Geographic region |  | P-Diff:<sup>3</sup> 0.624 |  | P-Diff: - |  | P-Diff: - |  | P-Diff: 0.610 |  | P-Diff: 0.525 |
| Africa | 16735 (9) | 0.84 (0.70, 1.02) | 16784 (9) | 0.79 (0.63, 1.00) | 3872 (5) | 0.97 (0.72, 1.31) | 16649 (9) | 0.83 (0.70, 0.98) | 4923 (6) | 1.09 (0.91, 1.30) |
| Southeast Asia | 7527 (3) | 0.77 (0.66, 0.90) | 6804 (2) | - | 5223 (2) | - | 7250 (3) | 0.88 (0.72, 1.08) | 8766 (3) | 1.15 (1.02, 1.29) |
| 18-mo stunting burden |  | P-Diff: 0.046 |  | P-Diff: 0.228 |  | P-Diff: - |  | P-Diff: 0.466 |  | P-Diff: 0.711 |
| < 35% | 8049 (4) | 1.16 (0.73, 1.83) | 7749 (3) | 0.91 (0.77, 1.08) | 1417 (2) | - | 7660 (3) | 0.90 (0.76, 1.07) | 4014 (5) | 1.15 (1.00, 1.33) |
| ≥ 35% | 16512 (9) | 0.78 (0.66, 0.92) | 15839 (8) | 0.75 (0.59, 0.95) | 7678 (5) | 1.00 (0.78, 1.27) | 16239 (9) | 0.83 (0.70, 0.98) | 9837 (5) | 1.09 (0.94, 1.27) |
| Malaria prevalence |  | P-Diff: 0.322 |  | P-Diff: 0.902 |  | P-Diff: 0.990 |  | P-Diff: 0.491 |  | P-Diff: 0.768 |
| < 10% | 19060 (8) | 0.93 (0.73, 1.18) | 18089 (6) | 0.81 (0.67, 0.98) | 7039 (4) | 0.94 (0.75, 1.16) | 18535 (7) | 0.87 (0.78, 0.97) | 10979 (5) | 1.11 (1.00, 1.23) |
| ≥ 10% | 5501 (5) | 0.78 (0.58, 1.05) | 5499 (5) | 0.80 (0.56, 1.12) | 2056 (3) | 0.95 (0.64, 1.41) | 5364 (5) | 0.82 (0.60, 1.10) | 2872 (5) | 1.12 (0.90, 1.41) |
| Anemia prevalence |  | P-Diff: 0.644 |  | P-Diff: 0.534 |  | P-Diff: - |  | P-Diff: 0.093 |  | P-Diff: 0.720 |
| < 60% | 19750 (8) | 0.87 (0.78, 0.97) | 19078 (7) | 0.83 (0.71, 0.98) | 7905 (5) | 0.91 (0.77, 1.07) | 19438 (8) | 0.88 (0.79, 0.98) | 11807 (5) | 1.11 (1.01, 1.22) |
| ≥ 60% | 4811 (5) | 0.90 (0.53, 1.55) | 4510 (4) | 0.76 (0.49, 1.15) | 1190 (2) | - | 4461 (4) | 0.75 (0.54, 1.05) | 2044 (5) | 1.14 (0.89, 1.45) |
| Source water quality |  | P-Diff: 0.737 |  | P-Diff: 0.394 |  | P-Diff: - |  | P-Diff: 0.303 |  | P-Diff: - |
| < 75% improved | 9819 (6) | 0.85 (0.63, 1.15) | 9708 (5) | 0.77 (0.55, 1.06) | 847 (2) | - | 9795 (6) | 0.82 (0.64, 1.05) | 1177 (2) | - |
| ≥ 75% improved | 7520<br>(7) | 0.93<br>(0.68, 1.26) | 6606<br>(5) | 0.87<br>(0.74, 1.02) | 4495<br>(5) | 0.85<br>(0.69, 1.03) | 6944<br>(6) | 0.93<br>(0.80, 1.08) | 10284<br>(8) | 1.14<br>(1.07, 1.23) |
| Sanitation |  | P-Diff:<br>0.994 |  | P-Diff:<br>0.888 |  | P-Diff:<br>0.286 |  | P-Diff:<br>0.599 |  | P-Diff:<br>0.631 |
| < 50% improved | 9468<br>(7) | 0.88<br>(0.68, 1.14) | 9355<br>(6) | 0.84<br>(0.62, 1.14) | 2037<br>(4) | 0.95<br>(0.72, 1.27) | 9489<br>(7) | 0.85<br>(0.68, 1.08) | 2344 (3) | 1.10<br>(0.98, 1.24) |
| ≥ 50% improved | 7871<br>(6) | 0.91<br>(0.63, 1.33) | 6959<br>(4) | 0.81<br>(0.70, 0.95) | 3305<br>(3) | 0.79<br>(0.64, 0.97) | 7250<br>(5) | 0.90<br>(0.77, 1.05) | 9117 (7) | 1.15<br>(1.00, 1.33) |
| Supplement duration |  | P-Diff:<br>0.545 |  | P-Diff:<br>0.731 |  | P-Diff:<br>- |  | P-Diff:<br>0.839 |  | P-Diff:<br>0.618 |
| ≤ 12 mo | 9644<br>(9) | 0.92<br>(0.69, 1.23) | 8806<br>(7) | 0.78<br>(0.57, 1.06) | 3872<br>(5) | 0.97<br>(0.72, 1.31) | 9307<br>(8) | 0.87<br>(0.69, 1.08) | 7142 (6) | 1.13<br>(1.03, 1.23) |
| > 12 mo | 14917<br>(4) | 0.82<br>(0.73, 0.93) | 14782<br>(4) | 0.83<br>(0.74, 0.94) | 5223<br>(2) | - | 14592<br>(4) | 0.84<br>(0.74, 0.94) | 6709 (4) | 1.06<br>(0.85, 1.34) |
| Frequency of contact |  | P-Diff:<br>0.370 |  | P-Diff:<br>0.991 |  | P-Diff:<br>0.724 |  | P-Diff:<br>0.977 |  | P-Diff:<br>0.433 |
| Weekly | 8680<br>(6) | 0.79<br>(0.62, 1.02) | 7960<br>(5) | 0.81<br>(0.57, 1.13) | 6480<br>(4) | 0.94<br>(0.77, 1.16) | 8482<br>(6) | 0.87<br>(0.66, 1.15) | 8951 (6) | 1.14<br>(1.05, 1.23) |
| Monthly | 15881<br>(7) | 0.95<br>(0.71, 1.26) | 15628<br>(6) | 0.81<br>(0.66, 0.98) | 2615<br>(3) | 0.96<br>(0.66, 1.41) | 15417<br>(6) | 0.86<br>(0.76, 0.97) | 4900 (4) | 1.05<br>(0.83, 1.34) |
| Average SQ-LNS compliance |  | P-Diff:<br>0.616 |  | P-Diff:<br>0.575 |  | P-Diff:<br>- |  | P-Diff:<br>0.330 |  | P-Diff:<br>0.700 |
| < 80% | 6290<br>(6) | 0.88<br>(0.75, 1.04) | 6339<br>(6) | 0.82<br>(0.65, 1.02) | 3872<br>(5) | 0.97<br>(0.72, 1.31) | 6204<br>(6) | 0.92<br>(0.78, 1.09) | 2695 (4) | 1.08<br>(0.90, 1.29) |
| ≥ 80% | 15054<br>(6) | 0.88<br>(0.75, 1.04) | 14032<br>(4) | 0.82<br>(0.65, 1.02) | 5223<br>(2) | - | 14478<br>(5) | 0.92<br>(0.78, 1.09) | 11156<br>(6) | 1.08<br>(0.90, 1.29) |

<sup>1</sup>N: number of individual participants; comp: number of intervention versus control group comparisons
<sup>2</sup>PR: prevalence ratio
<sup>3</sup>P-Diff: P-value for the difference in effects of SQ-LNS between the two levels of the effect modifier.

Anemia prevalence among children age 6-59 months modified the effect of SQ-LNS on the prevalence of children in the lowest decile of motor scores. In countries with ≥ 60% child anemia prevalence, SQ-LNS reduced this adverse motor outcome by 25%, compared to a 12% reduction in countries with < 60% anemia prevalence (Table 4).

Study-level sanitation (< 50% vs. ≥ 50% prevalence of improved sanitation in the study sample) modified the effect of SQ-LNS on executive function (Table 3). Among the 3 comparisons with higher prevalence of improved sanitation, the pooled effect of SQ-LNS on executive function was 0.06 SD (95% CI −0.01, 0.13), while among the 4 comparisons with a lower prevalence of improved sanitation, the pooled effect size was −0.09 SD (95% CI −0.18, 0.01). No other study-level characteristics significantly modified effects of SQ-LNS on any other developmental outcome (**Supplemental Figures 4A-P**).

### Effect modification by individual-level characteristics

Individual-level effect modification results were consistent across fixed and random effects models and across all sensitivity analyses (data not shown; available upon request). The results presented below refer to the fixed effects all-trials analysis. The following individual-level characteristics did not significantly modify the effect of SQ-LNS on any developmental outcome: indicators of household food insecurity, water quality, sanitation, and home environment; maternal BMI and depressive symptoms; child sex and season at outcome assessment (**Tables 5** and **6; Supplemental Figures 5** and **6**).

Household socio-economic status (SES; above or below the study median) modified the effect of SQ-LNS on mean language, motor, and executive function scores (Table 5; **Figure 7**). Effects of SQ-LNS on these scores were larger among children in low-SES households (0.06 to 0.12 SD) compared to high-SES households (−0.04 to 0.05 SD). For the percentage of children in the lowest decile of scores, there was no significant effect modification by household SES with regard to prevalence ratios. However, for language there was a greater percentage point reduction in low scores among children in the low-SES group (3 percentage points) than in the high-SES group (1 percentage point) (Supplemental Figure 6C).

**Figure 7.**
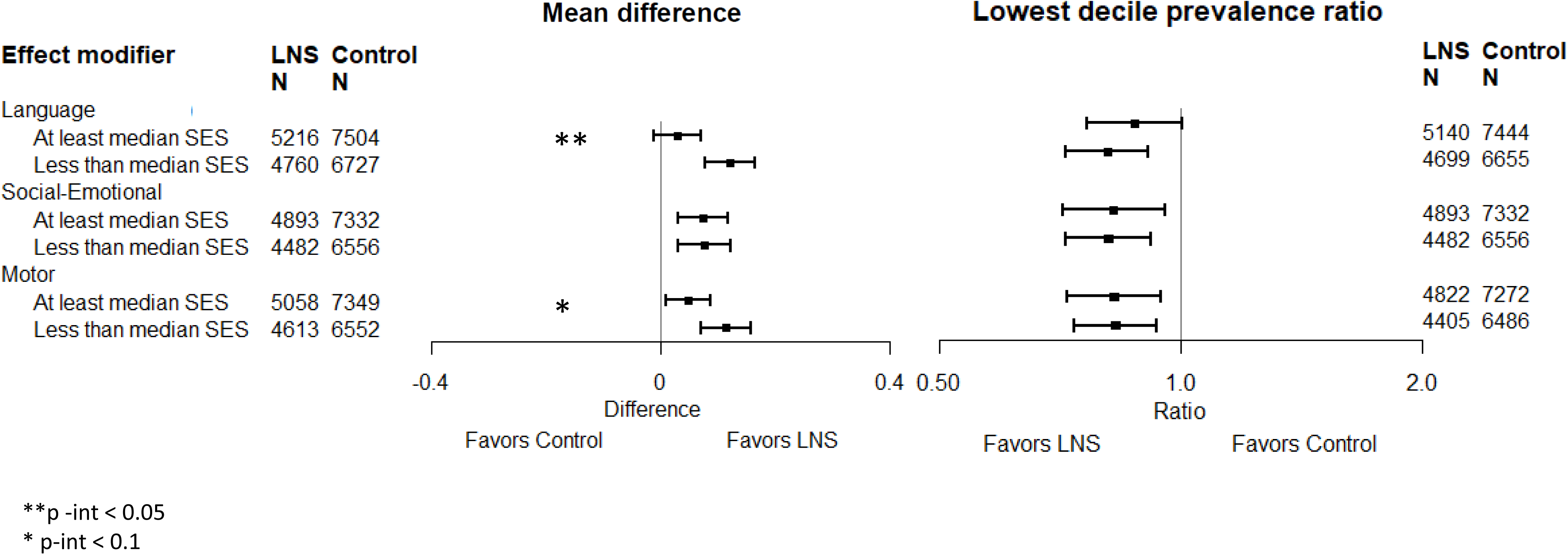
Pooled effects of SQ-LNS on 6 primary developmental outcomes stratified by individual-level household socio-economic status

**Table 5.**
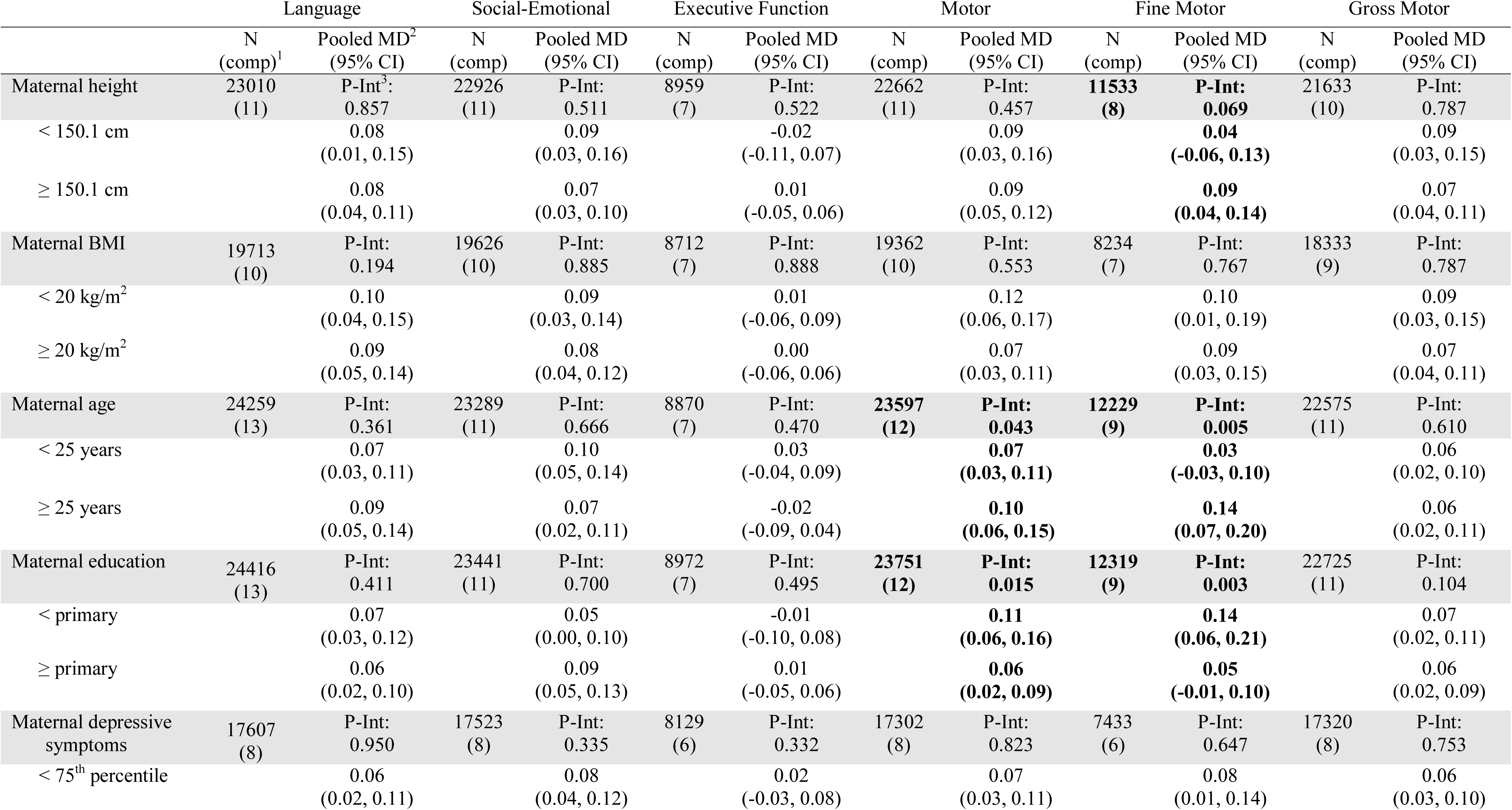
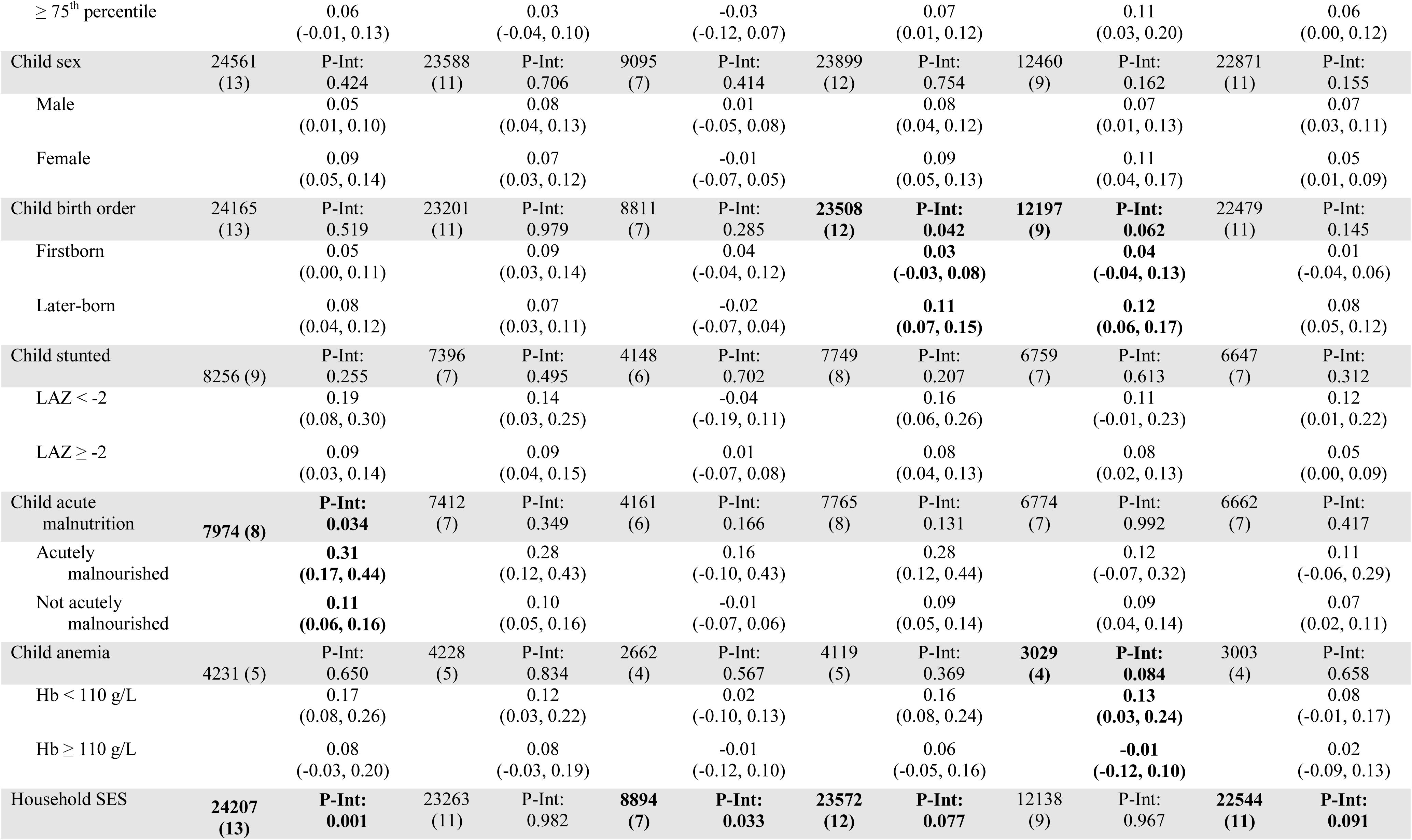
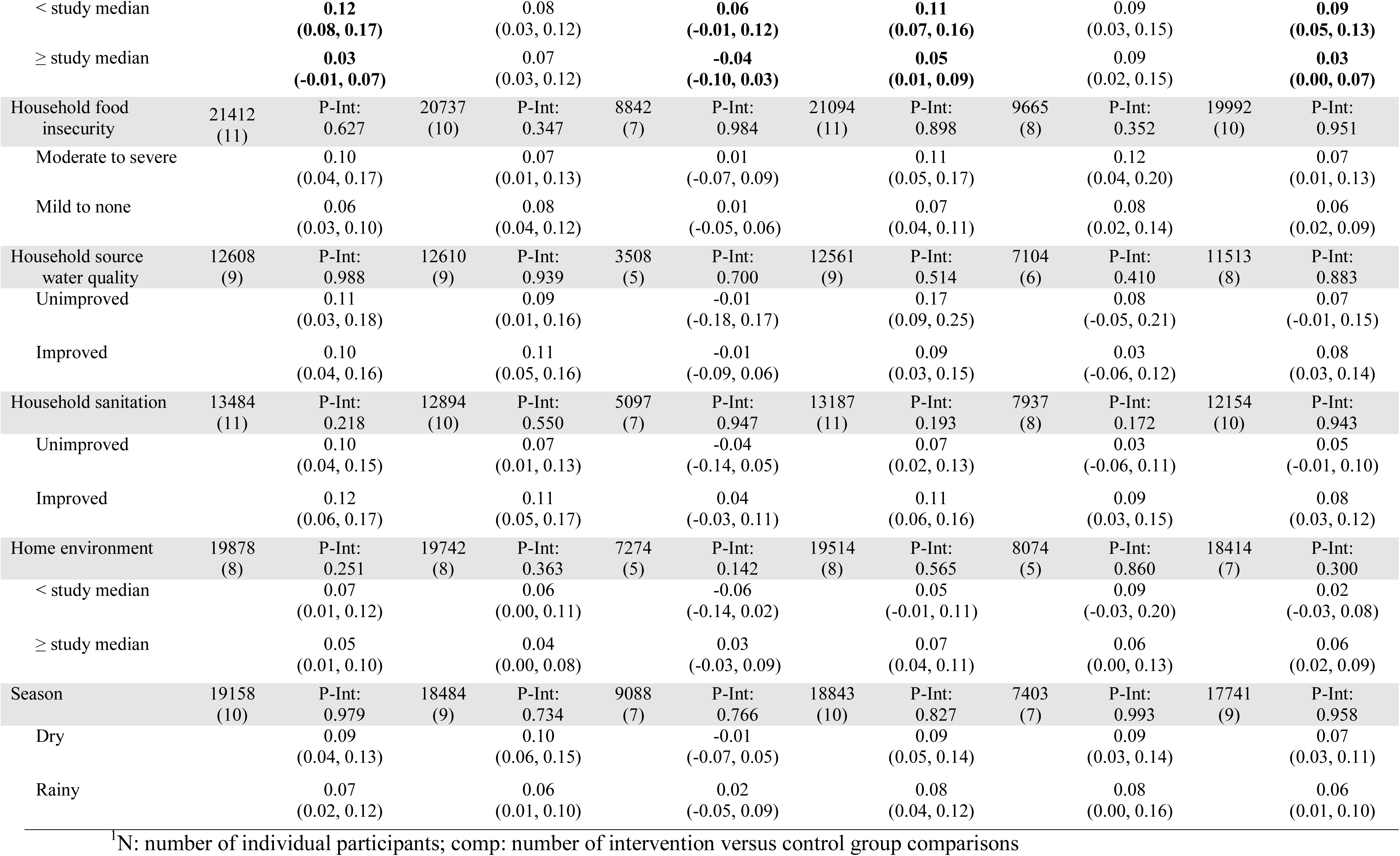
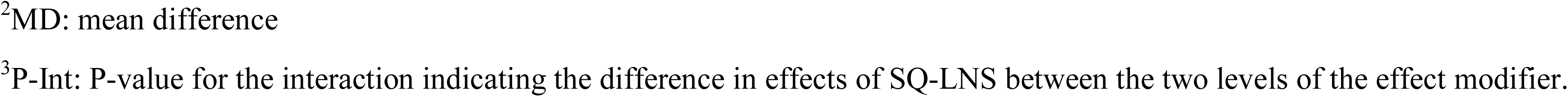
Individual-level effect modifiers of effects of SQ-LNS provided to infants and young children age 6 to 24 months on continuous developmental outcomes

Child baseline acute malnutrition (WLZ < −2 SD or MUAC < 125 mm) modified the effect of SQ-LNS on mean language scores (Table 5; **Figure 8**). The effect of SQ-LNS on mean language score was significantly larger among children who were malnourished when they began receiving SQ-LNS (0.30 SD) compared to those who were not (0.11 SD). Social-emotional and motor scores showed a similar pattern of greater effect sizes among acutely malnourished children (0.27 SD for both outcomes vs 0.09-0.10 SD among children who were not malnourished), however, the interaction tests were not statistically significant.

**Figure 8.**
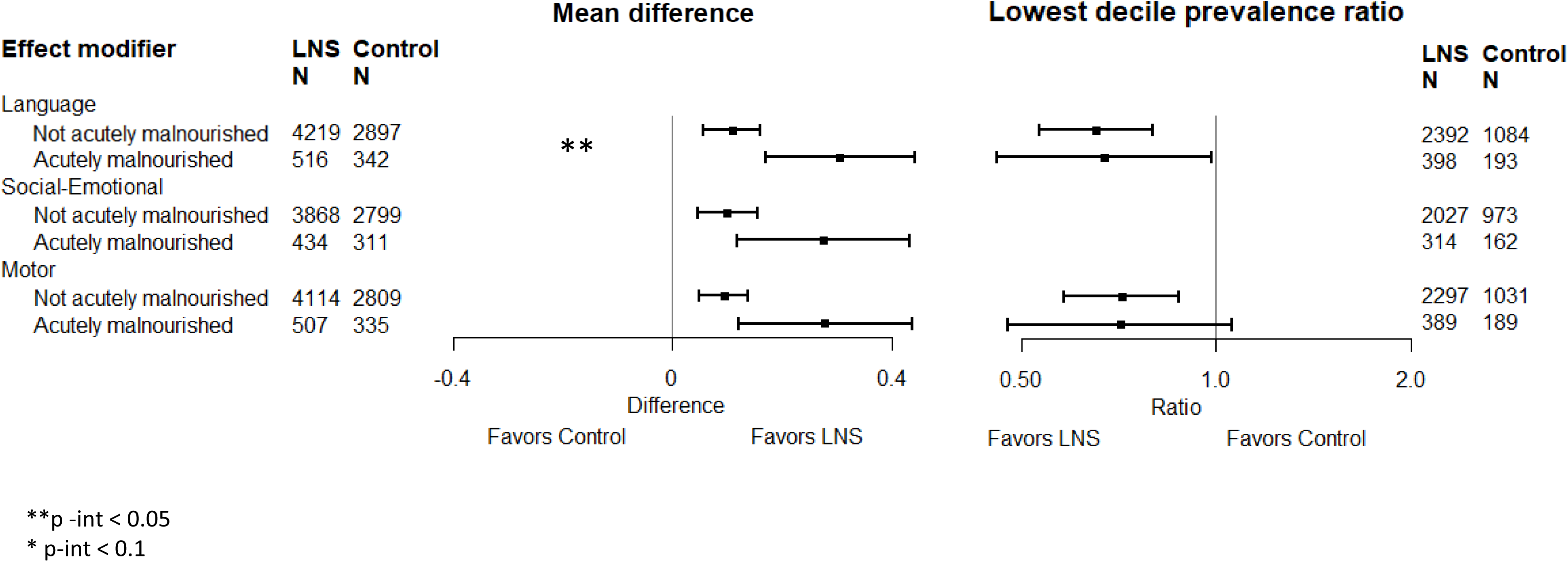
Pooled effects of SQ-LNS on 6 primary developmental outcomes stratified by individual-level child baseline acute malnutrition

Child baseline stunting (LAZ < −2 SD) modified the effect of SQ-LNS on the prevalence difference of children in the lowest decile of language scores. There was a 7 percentage point reduction of children in the lowest decile of language scores among children who were stunted when they began receiving SQ-LNS (95% CI −0.11, −0.04) compared to a 3 percentage point reduction (95% CI −0.01, −0.04; Supplemental Figure 5C8) among those who were not stunted. Social-emotional and motor scores showed similar trends of greater effect sizes among stunted children (**Figure 9**), however, no interactions between baseline stunting and intervention group were significant for these or any other outcomes.

**Figure 9.**
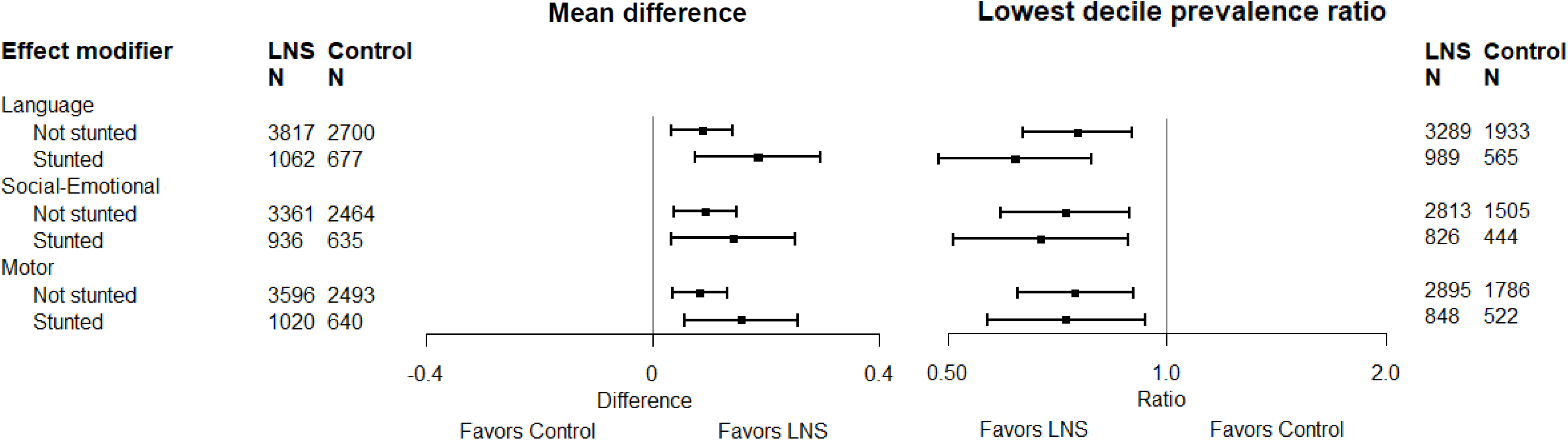
Pooled effects of SQ-LNS on 6 primary developmental outcomes stratified by individual-level child baseline stunting

Maternal education, maternal age, and child birth order modified the effects of SQ-LNS on mean motor and fine motor scores (Table 5). Greater effects of SQ-LNS on these scores were found among children of mothers with lower education (0.11 to 0.14 SD) than those with higher education (0.05 to 0.06 SD), among children of older mothers (0.10 to 0.14 SD) than younger mothers (0.03 to 0.07 SD), and among later-born children (i.e., those born after the first born child; 0.11 to 0.12 SD) than first born children (0.03 to 0.04 SD). Maternal education also modified the effects of SQ-LNS with respect to the prevalence difference for scoring in the lowest decile of motor scores, with greater reductions among children of mothers with lower education (Supplemental Figure 5I4). Birth order also modified the effect of SQ-LNS on this adverse motor outcome, with a greater reduction among later-born children (24% reduction versus 1% increase among first born children) and on walking without support at 12 months, with a greater positive effect among later-born children (16% increase vs. 5% increase among first born children; Table 6). Maternal age also modified the effect of SQ-LNS on the prevalence of children in the lowest decile of executive function scores. In contrast to the pattern for motor scores, for executive function greater effects with respect to both the prevalence ratio and the prevalence difference were found among children of younger mothers than older mothers (Table 6; Supplemental Figures 5M3 and 5N3). Maternal education, maternal age, and birth order did not modify the effects of SQ-LNS on any other developmental outcomes.

**Table 6.**
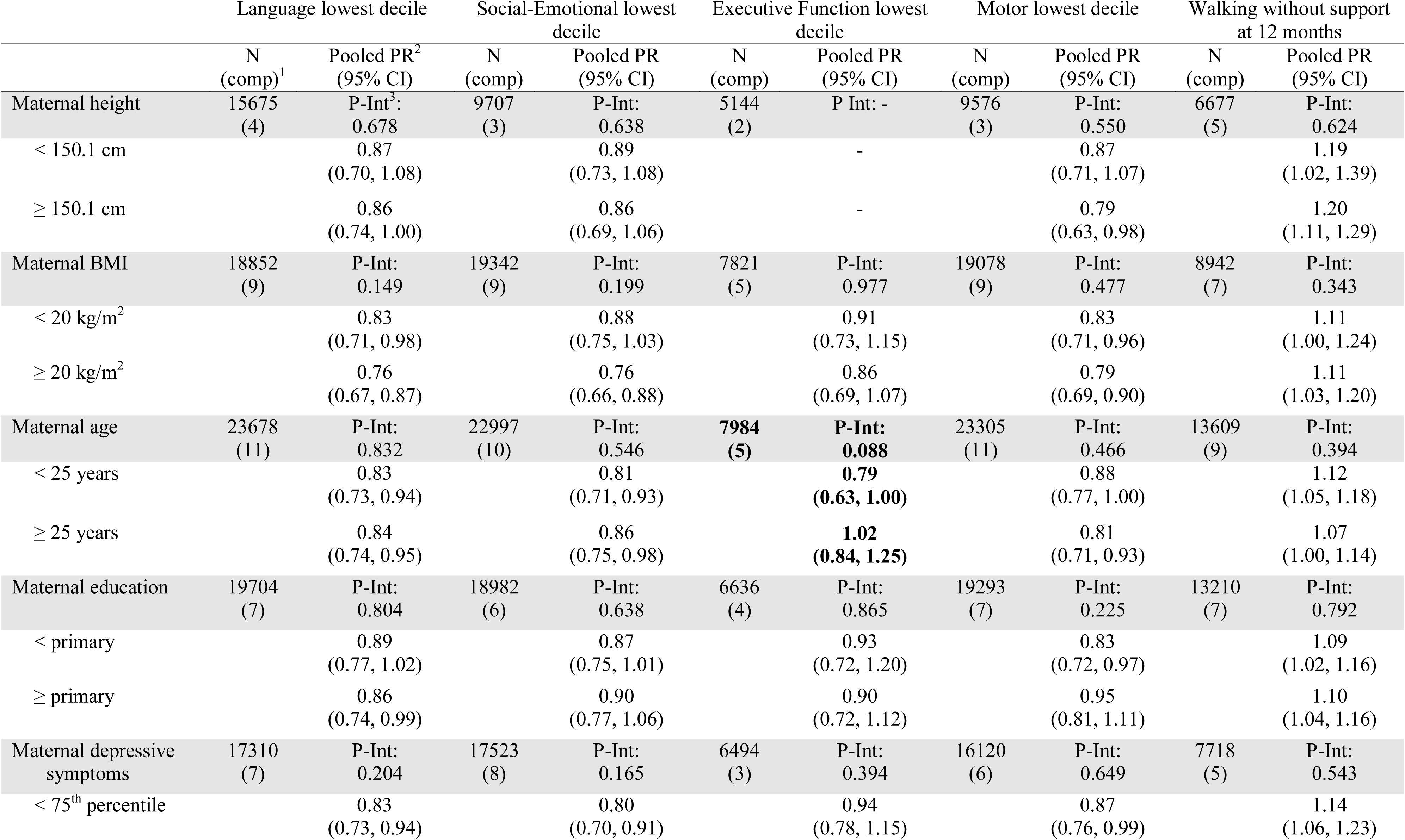
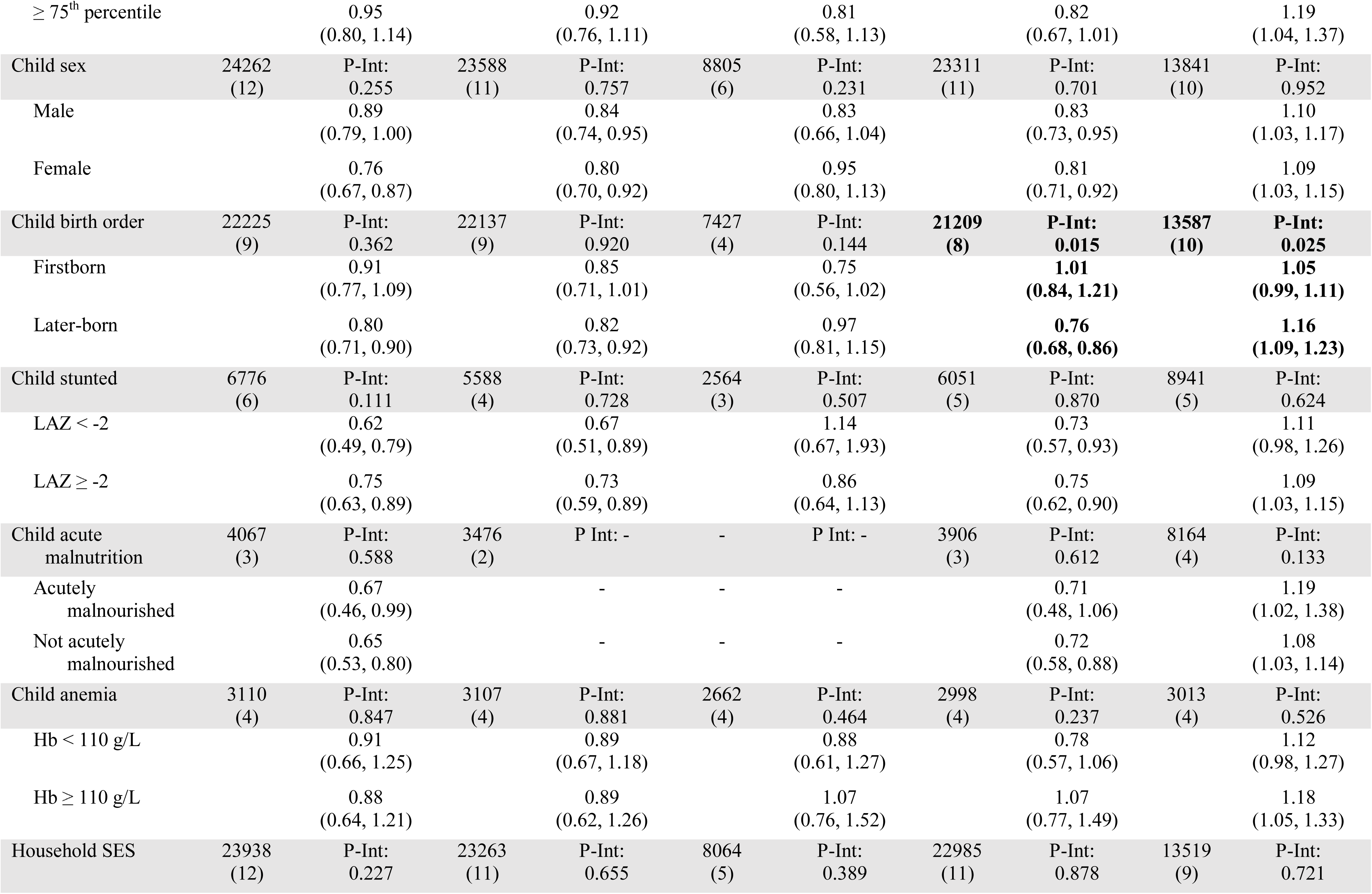
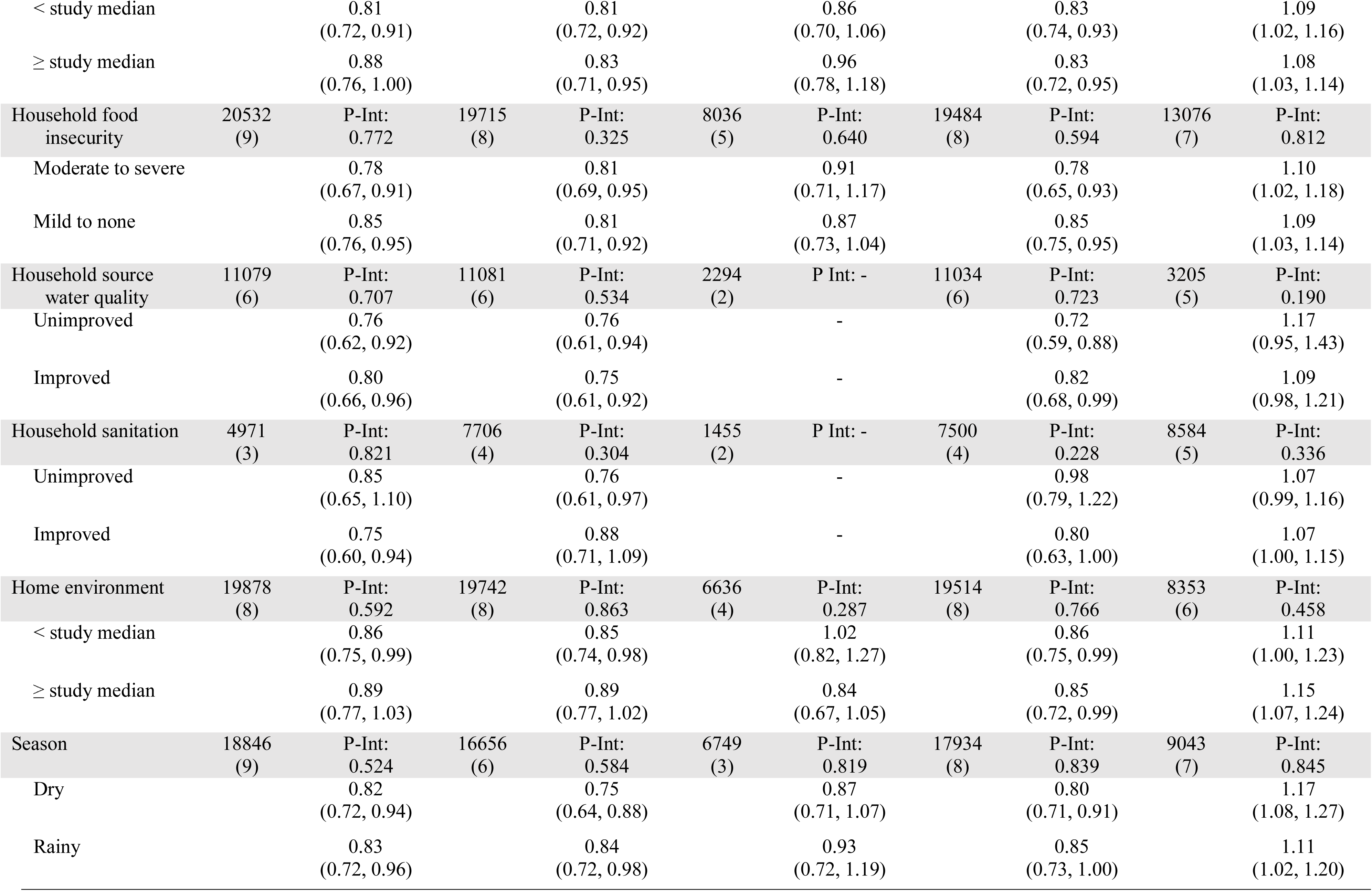
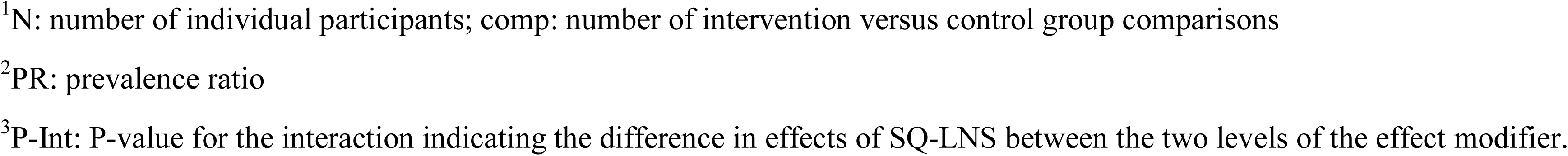
Individual-level effect modifiers of effects of SQ-LNS provided to infants and young children age 6 to 24 months on binary developmental outcomes

|  | Language lowest decile |  | Social-Emotional lowest decile |  | Executive Function lowest decile |  | Motor lowest decile |  | Walking without support at 12 months |  |
| --- | --- | --- | --- | --- | --- | --- | --- | --- | --- | --- |
|  | N<br>(comp)<sup>1</sup> | Pooled PR<sup>2</sup><br>(95% CI) | N<br>(comp) | Pooled PR<br>(95% CI) | N<br>(comp) | Pooled PR<br>(95% CI) | N<br>(comp) | Pooled PR<br>(95% CI) | N<br>(comp) | Pooled PR<br>(95% CI) |
| Maternal height | 15675<br>(4) | P-Int<sup>3</sup>:<br>0.678 | 9707<br>(3) | P-Int:<br>0.638 | 5144<br>(2) | P Int: - | 9576<br>(3) | P-Int:<br>0.550 | 6677<br>(5) | P-Int:<br>0.624 |
| < 150.1 cm |  | 0.87<br>(0.70, 1.08) |  | 0.89<br>(0.73, 1.08) |  | - |  | 0.87<br>(0.71, 1.07) |  | 1.19<br>(1.02, 1.39) |
| ≥ 150.1 cm |  | 0.86<br>(0.74, 1.00) |  | 0.86<br>(0.69, 1.06) |  | - |  | 0.79<br>(0.63, 0.98) |  | 1.20<br>(1.11, 1.29) |
| Maternal BMI | 18852<br>(9) | P-Int:<br>0.149 | 19342<br>(9) | P-Int:<br>0.199 | 7821<br>(5) | P-Int:<br>0.977 | 19078<br>(9) | P-Int:<br>0.477 | 8942<br>(7) | P-Int:<br>0.343 |
| < 20 kg/m<sup>2</sup> |  | 0.83<br>(0.71, 0.98) |  | 0.88<br>(0.75, 1.03) |  | 0.91<br>(0.73, 1.15) |  | 0.83<br>(0.71, 0.96) |  | 1.11<br>(1.00, 1.24) |
| ≥ 20 kg/m<sup>2</sup> |  | 0.76<br>(0.67, 0.87) |  | 0.76<br>(0.66, 0.88) |  | 0.86<br>(0.69, 1.07) |  | 0.79<br>(0.69, 0.90) |  | 1.11<br>(1.03, 1.20) |
| Maternal age | 23678<br>(11) | P-Int:<br>0.832 | 22997<br>(10) | P-Int:<br>0.546 | **7984**<br>**(5)** | **P-Int:**<br>**0.088** | 23305<br>(11) | P-Int:<br>0.466 | 13609<br>(9) | P-Int:<br>0.394 |
| < 25 years |  | 0.83<br>(0.73, 0.94) |  | 0.81<br>(0.71, 0.93) |  | **0.79**<br>**(0.63, 1.00)** |  | 0.88<br>(0.77, 1.00) |  | 1.12<br>(1.05, 1.18) |
| ≥ 25 years |  | 0.84<br>(0.74, 0.95) |  | 0.86<br>(0.75, 0.98) |  | **1.02**<br>**(0.84, 1.25)** |  | 0.81<br>(0.71, 0.93) |  | 1.07<br>(1.00, 1.14) |
| Maternal education | 19704<br>(7) | P-Int:<br>0.804 | 18982<br>(6) | P-Int:<br>0.638 | 6636<br>(4) | P-Int:<br>0.865 | 19293<br>(7) | P-Int:<br>0.225 | 13210<br>(7) | P-Int:<br>0.792 |
| < primary |  | 0.89<br>(0.77, 1.02) |  | 0.87<br>(0.75, 1.01) |  | 0.93<br>(0.72, 1.20) |  | 0.83<br>(0.72, 0.97) |  | 1.09<br>(1.02, 1.16) |
| ≥ primary |  | 0.86<br>(0.74, 0.99) |  | 0.90<br>(0.77, 1.06) |  | 0.90<br>(0.72, 1.12) |  | 0.95<br>(0.81, 1.11) |  | 1.10<br>(1.04, 1.16) |
| Maternal depressive symptoms | 17310<br>(7) | P-Int:<br>0.204 | 17523<br>(8) | P-Int:<br>0.165 | 6494<br>(3) | P-Int:<br>0.394 | 16120<br>(6) | P-Int:<br>0.649 | 7718<br>(5) | P-Int:<br>0.543 |
| < 75<sup>th</sup> percentile |  | 0.83<br>(0.73, 0.94) |  | 0.80<br>(0.70, 0.91) |  | 0.94<br>(0.78, 1.15) |  | 0.87<br>(0.76, 0.99) |  | 1.14<br>(1.06, 1.23) |
| $\geq 75^{\text{th}}$ percentile | | 0.95<br>(0.80, 1.14) | | 0.92<br>(0.76, 1.11) | | 0.81<br>(0.58, 1.13) | | 0.82<br>(0.67, 1.01) | | 1.19<br>(1.04, 1.37) |
| Child sex | 24262<br>(12) | P-Int:<br>0.255 | 23588<br>(11) | P-Int:<br>0.757 | 8805<br>(6) | P-Int:<br>0.231 | 23311<br>(11) | P-Int:<br>0.701 | 13841<br>(10) | P-Int:<br>0.952 |
| Male |  | 0.89<br>(0.79, 1.00) |  | 0.84<br>(0.74, 0.95) |  | 0.83<br>(0.66, 1.04) |  | 0.83<br>(0.73, 0.95) |  | 1.10<br>(1.03, 1.17) |
| Female |  | 0.76<br>(0.67, 0.87) |  | 0.80<br>(0.70, 0.92) |  | 0.95<br>(0.80, 1.13) |  | 0.81<br>(0.71, 0.92) |  | 1.09<br>(1.03, 1.15) |
| Child birth order | 22225<br>(9) | P-Int:<br>0.362 | 22137<br>(9) | P-Int:<br>0.920 | 7427<br>(4) | P-Int:<br>0.144 | **21209**<br>**(8)** | **P-Int:**<br>**0.015** | **13587**<br>**(10)** | **P-Int:**<br>**0.025** |
| Firstborn |  | 0.91<br>(0.77, 1.09) |  | 0.85<br>(0.71, 1.01) |  | 0.75<br>(0.56, 1.02) |  | **1.01**<br>**(0.84, 1.21)** |  | **1.05**<br>**(0.99, 1.11)** |
| Later-born |  | 0.80<br>(0.71, 0.90) |  | 0.82<br>(0.73, 0.92) |  | 0.97<br>(0.81, 1.15) |  | **0.76**<br>**(0.68, 0.86)** |  | **1.16**<br>**(1.09, 1.23)** |
| Child stunted | 6776<br>(6) | P-Int:<br>0.111 | 5588<br>(4) | P-Int:<br>0.728 | 2564<br>(3) | P-Int:<br>0.507 | 6051<br>(5) | P-Int:<br>0.870 | 8941<br>(5) | P-Int:<br>0.624 |
| LAZ < -2 |  | 0.62<br>(0.49, 0.79) |  | 0.67<br>(0.51, 0.89) |  | 1.14<br>(0.67, 1.93) |  | 0.73<br>(0.57, 0.93) |  | 1.11<br>(0.98, 1.26) |
| LAZ ≥ -2 | | 0.75<br>(0.63, 0.89) | | 0.73<br>(0.59, 0.89) | | 0.86<br>(0.64, 1.13) | | 0.75<br>(0.62, 0.90) | | 1.09<br>(1.03, 1.15) |
| Child acute malnutrition | 4067<br>(3) | P-Int:<br>0.588 | 3476<br>(2) | P Int: - | - | P Int: - | 3906<br>(3) | P-Int:<br>0.612 | 8164<br>(4) | P-Int:<br>0.133 |
| Acutely malnourished |  | 0.67<br>(0.46, 0.99) |  | - | - | - |  | 0.71<br>(0.48, 1.06) |  | 1.19<br>(1.02, 1.38) |
| Not acutely malnourished |  | 0.65<br>(0.53, 0.80) |  | - | - | - |  | 0.72<br>(0.58, 0.88) |  | 1.08<br>(1.03, 1.14) |
| Child anemia | 3110<br>(4) | P-Int:<br>0.847 | 3107<br>(4) | P-Int:<br>0.881 | 2662<br>(4) | P-Int:<br>0.464 | 2998<br>(4) | P-Int:<br>0.237 | 3013<br>(4) | P-Int:<br>0.526 |
| Hb < 110 g/L |  | 0.91<br>(0.66, 1.25) |  | 0.89<br>(0.67, 1.18) |  | 0.88<br>(0.61, 1.27) |  | 0.78<br>(0.57, 1.06) |  | 1.12<br>(0.98, 1.27) |
| Hb ≥ 110 g/L | | 0.88<br>(0.64, 1.21) | | 0.89<br>(0.62, 1.26) | | 1.07<br>(0.76, 1.52) | | 1.07<br>(0.77, 1.49) | | 1.18<br>(1.05, 1.33) |
| Household SES | 23938<br>(12) | P-Int:<br>0.227 | 23263<br>(11) | P-Int:<br>0.655 | 8064<br>(5) | P-Int:<br>0.389 | 22985<br>(11) | P-Int:<br>0.878 | 13519<br>(9) | P-Int:<br>0.721 |
| < study median |  | 0.81<br>(0.72, 0.91) |  | 0.81<br>(0.72, 0.92) |  | 0.86<br>(0.70, 1.06) |  | 0.83<br>(0.74, 0.93) |  | 1.09<br>(1.02, 1.16) |
| ≥ study median |  | 0.88<br>(0.76, 1.00) |  | 0.83<br>(0.71, 0.95) |  | 0.96<br>(0.78, 1.18) |  | 0.83<br>(0.72, 0.95) |  | 1.08<br>(1.03, 1.14) |
| Household food insecurity | 20532<br>(9) | P-Int:<br>0.772 | 19715<br>(8) | P-Int:<br>0.325 | 8036<br>(5) | P-Int:<br>0.640 | 19484<br>(8) | P-Int:<br>0.594 | 13076<br>(7) | P-Int:<br>0.812 |
| Moderate to severe |  | 0.78<br>(0.67, 0.91) |  | 0.81<br>(0.69, 0.95) |  | 0.91<br>(0.71, 1.17) |  | 0.78<br>(0.65, 0.93) |  | 1.10<br>(1.02, 1.18) |
| Mild to none |  | 0.85<br>(0.76, 0.95) |  | 0.81<br>(0.71, 0.92) |  | 0.87<br>(0.73, 1.04) |  | 0.85<br>(0.75, 0.95) |  | 1.09<br>(1.03, 1.14) |
| Household source water quality | 11079<br>(6) | P-Int:<br>0.707 | 11081<br>(6) | P-Int:<br>0.534 | 2294<br>(2) | P Int: - | 11034<br>(6) | P-Int:<br>0.723 | 3205<br>(5) | P-Int:<br>0.190 |
| Unimproved |  | 0.76<br>(0.62, 0.92) |  | 0.76<br>(0.61, 0.94) |  | - |  | 0.72<br>(0.59, 0.88) |  | 1.17<br>(0.95, 1.43) |
| Improved |  | 0.80<br>(0.66, 0.96) |  | 0.75<br>(0.61, 0.92) |  | - |  | 0.82<br>(0.68, 0.99) |  | 1.09<br>(0.98, 1.21) |
| Household sanitation | 4971<br>(3) | P-Int:<br>0.821 | 7706<br>(4) | P-Int:<br>0.304 | 1455<br>(2) | P Int: - | 7500<br>(4) | P-Int:<br>0.228 | 8584<br>(5) | P-Int:<br>0.336 |
| Unimproved |  | 0.85<br>(0.65, 1.10) |  | 0.76<br>(0.61, 0.97) |  | - |  | 0.98<br>(0.79, 1.22) |  | 1.07<br>(0.99, 1.16) |
| Improved |  | 0.75<br>(0.60, 0.94) |  | 0.88<br>(0.71, 1.09) |  | - |  | 0.80<br>(0.63, 1.00) |  | 1.07<br>(1.00, 1.15) |
| Home environment | 19878<br>(8) | P-Int:<br>0.592 | 19742<br>(8) | P-Int:<br>0.863 | 6636<br>(4) | P-Int:<br>0.287 | 19514<br>(8) | P-Int:<br>0.766 | 8353<br>(6) | P-Int:<br>0.458 |
| < study median |  | 0.86<br>(0.75, 0.99) |  | 0.85<br>(0.74, 0.98) |  | 1.02<br>(0.82, 1.27) |  | 0.86<br>(0.75, 0.99) |  | 1.11<br>(1.00, 1.23) |
| ≥ study median |  | 0.89<br>(0.77, 1.03) |  | 0.89<br>(0.77, 1.02) |  | 0.84<br>(0.67, 1.05) |  | 0.85<br>(0.72, 0.99) |  | 1.15<br>(1.07, 1.24) |
| Season | 18846<br>(9) | P-Int:<br>0.524 | 16656<br>(6) | P-Int:<br>0.584 | 6749<br>(3) | P-Int:<br>0.819 | 17934<br>(8) | P-Int:<br>0.839 | 9043<br>(7) | P-Int:<br>0.845 |
| Dry |  | 0.82<br>(0.72, 0.94) |  | 0.75<br>(0.64, 0.88) |  | 0.87<br>(0.71, 1.07) |  | 0.80<br>(0.71, 0.91) |  | 1.17<br>(1.08, 1.27) |
| Rainy |  | 0.83<br>(0.72, 0.96) |  | 0.84<br>(0.72, 0.98) |  | 0.93<br>(0.72, 1.19) |  | 0.85<br>(0.73, 1.00) |  | 1.11<br>(1.02, 1.20) |

<sup>1</sup>N: number of individual participants; comp: number of intervention versus control group comparisons
<sup>2</sup>PR: prevalence ratio
<sup>3</sup>P-Int: P-value for the interaction indicating the difference in effects of SQ-LNS between the two levels of the effect modifier.

Maternal height and child baseline anemia modified the effect of SQ-LNS on mean fine motor scores, but no other outcomes. There were greater effects of SQ-LNS on fine motor scores among children of taller mothers (0.09 SD) than children of shorter mothers (0.04 SD) and children who were anemic at baseline (0.13 SD) than children who were not (−0.01 SD; Table 5).

## Discussion

In this IPD meta-analysis of 13 randomized controlled trials in 9 countries with a total sample size of over 30,000 children, SQ-LNS provided to infants and young children 6 to 24 months of age increased mean language, social-emotional, and motor scores by 0.06 to 0.09 SD and led to a relative reduction of 16% to 19% in adverse developmental outcomes (1-2 percentage point difference). The quality of the evidence for all outcomes was high. The effects of SQ-LNS on developmental outcomes did not significantly differ by most study-level characteristics including region, malaria prevalence, water quality, supplementation duration, frequency of contact, or average compliance with SQ-LNS, indicating that these aspects of context and program delivery did not explain differences in effect sizes across these 13 trials. However, effects of SQ-LNS on language, social-emotional, and motor development were larger among study populations with a higher stunting burden and effects on motor development were larger in sites with higher prevalence of child anemia. At the individual level, greater effects of SQ-LNS were found on language among children who were stunted or acutely malnourished when they started receiving SQ-LNS; on language, motor, and executive function among households with lower SES; and on motor and fine motor development among later-born children, children of older mothers, and children of mothers with lower education. Children of taller mothers and children who were anemic at baseline also showed greater effects on fine motor scores, while children of younger mothers showed greater effects on executive function scores.

### Main effects

Our findings of significant positive effects of SQ-LNS on developmental outcomes in the range of 0.06 to 0.09 SD, which is equivalent to about 1-1.5 IQ points, are consistent with a previous meta-analysis of SQ-LNS by Tam et al. (7), which reported slightly larger effect sizes of 0.12 to 0.13 SD (∼1.8-2 IQ points). Effect sizes in the range of 0.06 to 0.09 SD are probably more accurate for a general population, as our analysis included a larger number of trials (13 vs 6) and children (∼30,000 vs ∼3,500). However, we found effect sizes closer to their estimates in sites with higher stunting burden (0.11 to 0.13 SD), and the trials included in the Tam et al. meta-analysis tended to have a high stunting burden. These effect sizes are also consistent with previous meta-analyses of child nutritional supplementation trials in LMICs. A meta-analysis by Larson et al. of 23 nutritional supplementation randomized controlled trials among children age 0-2 years indicated an effect size of 0.08 SD for cognitive/mental development (47). Another meta-analysis by Prado et al. of effects of interventions from pregnancy to 5 years on linear growth and development showed an effect of child nutritional supplementation of 0.13 SD on language (13 studies), 0.09 SD on motor (27 studies), and 0.09 SD on social-emotional development (13 studies) (48).

These previous meta-analyses pooled effects across studies that provided a wide range of different types of nutritional supplements, including single micronutrients, as well as multiple micronutrients with and without macronutrients. Few previous meta-analyses of specific types of nutritional supplements have been able to calculate pooled effects on developmental outcomes, due to small numbers of studies and differences in measurement and reporting of outcomes in this domain. For example, a 2020 Cochrane review of home fortification of children’s foods with multiple micronutrient powders was not able to calculate pooled effects on developmental outcomes (49). Therefore, it is difficult to compare the effects of SQ-LNS with the effects of other types of nutritional supplements. However, in Prado et al. (48), pooled effect sizes for language, social-emotional, and motor scores were similar in trials that provided multiple micronutrients with macronutrients (0.09 to 0.10 SD; 10-17 trials) and in trials that provided multiple micronutrients without macronutrients (0.08 to 0.11 SD; 5-11 trials). This suggests that the multiple micronutrients in SQ-LNS may be key ingredients for effects on developmental outcomes. However, further research is needed to understand how SQ-LNS compares to other types of nutritional supplements designed to fill nutrient gaps in children’s diets, such as micronutrient powders and fortified blended foods, with regard to effects on child development. One difference between SQ-LNS and other products that contain both macro- and micronutrients is that SQ-LNS contain substantial amounts of essential fatty acids, important for brain development, while other energy-containing supplements may not.

Our study is the first meta-analysis to report effects of child nutritional supplementation on reducing the prevalence of children in the lowest decile of developmental scores. We used the lowest decile of scores as a proxy for children who may be at risk for developmental delay. Typically, distributions of developmental assessment scores in LMIC settings have a left tail that is larger than the right tail, comprised of children who score substantially lower than their age- and sex-matched peers and may be developmentally delayed. In most of the studies in this IPD meta-analysis (in 9/13 comparisons for language, 10/12 for motor, and 8/11 for social-emotional scores), a greater proportion of children scored < 2 SD below the mean (overall ∼3%) than > 2 SD above the mean (< 1%). The finding of significant reductions in the lowest decile of scores shows that SQ-LNS not only shifts the mean of the distribution, but also improves outcomes among children in the lower tail of the distribution who may be at particular risk of developmental delay. Attaining developmental skills that are appropriate for the child’s age is likely to facilitate further advances in development because many skills developed at later stages build on those that were learned previously. For example, acquiring the skill of walking without support is a catalyst for change in multiple domains of development (50). Both the reduction in the percentage of children in the lowest decile of scores and the increase in the percentage of children walking without support with SQ-LNS could be important for supporting healthier developmental trajectories in the population. The percentage of children walking without support in the SQ-LNS groups (45% overall vs 39% in control groups) is closer to the prevalence in the World Health Organization Multi-Center Growth Reference study, which showed that in a healthy group of children, 50% were walking without support at age 12 months (51). Further research is needed to understand the longer-term effects of SQ-LNS on developmental outcomes and whether the observed positive effects at 12-24 months are sustained to later childhood. Two of the trials in this IPD meta-analysis conducted follow-up assessments at age 3-6 years. One follow-up study in Bangladesh found significantly higher composite cognitive scores (+0.13 SD) in the SQ-LNS group (52), while the other follow-up study in Ghana showed reduced social-emotional difficulties (−0.12 SD) in the SQ-LNS group, with greater effects among children with lower quality home environments (−0.22 SD) (53).

Among the 6 trials (7 comparisons) that assessed executive function, no overall effects of SQ-LNS were found. Executive function is the cognitive control of attention, self-regulation, and emotion, including the ability to plan and monitor actions, self-regulate actions and emotions, focus and sustain attention, and maintain information in short-term memory (54). These skills and the neural structures that underlie them (the prefrontal cortex and other connected cortical and sub-cortical structures) experience their peak rate of development in later childhood and adolescence (55). Therefore, 6-24 months of age may be too early for nutritional supplementation to have a measurable effect on the development of executive function.

### Study-level effect modification

The child stunting burden in the study sample was the most consistent effect modifier across developmental outcomes (5 of the 8 developmental outcomes with sufficient data to analyze this effect modifier). Stunted growth has been consistently associated with poor developmental outcomes (56), therefore stunted children may have greater potential to benefit from SQ-LNS. Within the control groups in this IPD meta-analysis, stunted children scored significantly lower than non-stunted children in language and motor scores in all trials, and in social-emotional scores in all trials except one. Recent evidence suggests that stunted growth does not cause delayed neurodevelopment, but instead is a sensitive marker of an environment that constrains growth and development through partly overlapping causes (48, 57). High child anemia prevalence, which also modified effects of SQ-LNS on motor outcomes, may also be a marker of a high-risk environment in which children have greater potential to benefit from SQ-LNS. The findings from this IPD meta-analysis show that inadequate dietary intake is one of the shared causes of faltering in both linear growth and development, as well as anemia, and that SQ-LNS has positive effects on all of these outcomes (12, 58). However, patterns of effect modification were different; greater effects on linear growth were not found in studies with a higher stunting burden. Linear growth may be less malleable to recovery through intervention after early growth restriction (e.g., in utero and from birth to 6 months of age), while development may be more responsive to post-natal intervention because brain plasticity continues throughout childhood (1).

Apart from stunting and anemia prevalence, we did not find significant effect modification by other study-level characteristics. In all study-level sub-groups, effect sizes were consistently in the expected direction favoring SQ-LNS groups. This suggests that effects are evident across a range of contexts from low to high malaria prevalence, water quality, supplementation duration, frequency of contact and average compliance with SQ-LNS. However, we had limited power to detect significant associations between study-level effect modifiers and effect sizes (maximum n=13 intervention versus control group comparisons). For example, the relative reduction in the prevalence of children in the lowest decile of language scores was 22% among studies with higher malaria prevalence versus 7% among studies with lower malaria prevalence. With additional studies and thus power, it is possible that such differences could reach significance. One exception was that the effect of SQ-LNS on executive function significantly differed between studies with a low versus high prevalence of improved sanitation. However, given the lack of overall effect of SQ-LNS on executive function and given that confidence intervals around the executive function estimates in both sub-groups (high and low prevalence of improved sanitation) included zero, it is likely that there was no true effect on executive function in either sub-group.

### Individual-level effect modification

Just as the study-level effect modification analysis showed that populations in higher risk environments, as indicated by higher prevalence of child stunting and anemia, had greater potential to benefit from SQ-LNS in developmental outcomes, the individual-level effect modification analysis consistently showed that certain sub-groups of children who may be in higher risk circumstances had a greater potential to benefit from SQ-LNS, including children who were stunted, anemic, acutely malnourished, in low-SES households, later-born, and whose mothers were older and less educated. As previously discussed for stunted children, in our pooled control group data, children in all of these categories, except children of older mothers, had lower language and motor scores and therefore had greater room for improvement in developmental skills. Iron-deficiency anemia, acute malnutrition, low SES, and low maternal education are associated with poor development (59, 60). Later-born children may have access to fewer household and caregiving resources because they are competing with older siblings, which may negatively affect their development. Although it is not clear why children of older mothers might be at higher risk, older mothers tended to be less educated and their children tended to be later-born, thus these sub-groups overlapped (46% of older mothers completed primary school, while 59% of younger mothers completed primary school; 84% of older mothers’ children were later born, while 46% of younger mothers’ children were later born; 75% of lower educated mothers’ children were later born, while 56% of higher educated mothers’ children were later born).

While the findings generally show that children in higher risk environments have greater potential to benefit in developmental outcomes from SQ-LNS, it is somewhat surprising that indicators of maternal depressive symptoms and the home environment did not modify effects of SQ-LNS in this IPD meta-analysis. Similar to stunted and malnourished children, children of mothers with depressive symptoms and children in low quality home environments also tend to have poor developmental scores (61, 62). Three SQ-LNS trials have found that children in lower quality home environments show greater benefits of SQ-LNS on developmental outcomes (37, 39, 53). One of these was a follow-up study at age 4-6 years that was not included in this IPD meta-analysis. However, findings from this IPD meta-analysis suggest that effects of SQ-LNS on development are generalizable regardless of maternal depressive symptoms and home environment, at least across the range represented in these studies. Similarly, lack of effect modification based on child sex, household food insecurity, water quality, sanitation, maternal BMI, and season suggest that effects on developmental outcomes are generalizable across these characteristics.

### Strengths and limitations of this IPD meta-analysis

This IPD meta-analysis had many strengths. A substantial number of high-quality trials that provided similar types of SQ-LNS products to children age 6-24 months were included. Investigators from all but one of the eligible studies participated and the sample size was very large. The availability of IPD allowed harmonization of calculation of developmental outcomes across trials, and enabled incorporation of cluster-randomized trials using robust standard errors and allowing for study-specific intra-cluster correlations. The 13 study sites were highly diverse in terms of sample characteristics and study designs, which provided heterogeneity for exploration of study-level effect modifiers. The consistency of findings across fixed and random effects models and sensitivity analyses strengthens confidence in the conclusions.

This IPD meta-analysis also had limitations. Although we were able to harmonize the calculation of developmental assessment scores across trials, different tools were used in different trials and we did not have an external reference to standardize scores. Thus, although all developmental scores were calculated in units of SD based on the within-study distribution, if SDs varied across studies, the point value of 1 SD could be larger in one trial than another, and interpretation of effect sizes would be different across trials. For example, among the 6 studies that used a 10-item A-not-B task to assess executive function, SDs ranged from 1.4 to 2.5 and among the 5 studies that used a 100-word vocabulary checklist to assess language, SDs ranged from 18.9 to 23.5. Ongoing efforts to develop a standardized scale of developmental scores will greatly improve future meta-analyses of developmental outcomes (63). Future research should also use developmental assessments directly observed by blinded assessors to address the potential risk of bias when parent-report developmental assessment tools are used in trials in which the intervention precludes participant blinding.

Another limitation was the limited diversity of geographical region. The majority of studies were conducted in Sub-Saharan Africa, with only one country representing the WHO Southeast Asia Region (Bangladesh), and one country representing Latin America and the Caribbean (Haiti). In addition, not all trials were included in all analyses because some trials did not measure some outcomes (e.g. executive function) or effect modifiers (e.g. baseline child stunting, maternal depressive symptoms, home environment). For study-level effect modifiers, statistical power was constrained by the limited number of trials. For individual-level effect modifiers, although we made every effort to standardize definitions and cutoffs for potential effect modifiers, there was variation across trials in the methods used to collect information on certain characteristics, such as household food insecurity and socioeconomic status. We examined multiple effect modifiers and numerous outcomes, so several of the significant p-for-interaction values are likely due to chance. As stated in Methods, we did not adjust for multiple hypothesis testing because developmental outcomes are inter-related and the effect modification analyses are inherently exploratory.

### Programmatic implications

Our findings suggest that if policy-makers and program planners implement SQ-LNS distribution to children age 6-24 months, they can expect modest, but potentially important, developmental gains among children in the population, particularly in areas with high child stunting burden. If the goal of a policy or program is to target not only developmental outcomes, but also child growth, iron deficiency, anemia, and mortality, then SQ-LNS should be considered. To our knowledge, SQ-LNS is the only nutrition intervention that has been documented to have positive effects on all of these outcomes (12, 58, 64).

However, if the primary goal of a policy or program is to improve developmental outcomes, investment is needed not only in nutrition but also other aspects of nurturing care, especially responsive care and learning opportunities (65). For all developmental domains, interventions that promote responsive care and learning opportunities have effect sizes 4-5 times larger (equivalent to about 5-7 IQ points) than those for nutritional supplementation alone (48). Integration of SQ-LNS with such programs should be considered. One advantage of integrating nutrition with caregiving interventions could be incentivizing participation in parenting groups or home visits through provision of SQ-LNS, thereby increasing coverage (66). Another advantage of integration is building on existing contact points between community front-line workers and families with young children, thus potentially reducing implementation costs (67). The cost of SQ-LNS is estimated at $0.07-0.14 per child per day not including distribution costs, depending on scale and location of production (68, 69). Further research is needed on the costs of programs promoting responsive care and learning opportunities targeting young children in LMICs and integration of such programs with nutrition programs (70).

Although we found that certain groups of children in higher risk environments showed greater benefits in developmental outcomes from SQ-LNS, such as children from low SES households, we also found that children from the lower risk groups (e.g. those from higher SES households) showed positive effects. We recommend that decisions regarding targeting specific communities or households be based on the wider body of evidence on all outcomes, including nutritional status and growth, not only developmental outcomes.

Our findings also have implications for programs designed for community management of acute malnutrition (CMAM). Many CMAM programs provide large-quantity LNS (∼1000-1500 kcal/d) to children who meet the cut-offs for severe acute malnutrition (WLZ < −3 or MUAC < 115 mm), according to WHO guidelines (71). Medium-quantity LNS (∼250-500 kcal/d) is typically used to treat moderate acute malnutrition (MAM; −3 < WLZ < −2 SD or 115 < MUAC < 125 mm), however, the coverage of MAM treatment is low. Most evidence for the efficacy of MAM treatment has focused on child survival and recovery, rather than developmental outcomes (72). We found that provision of SQ-LNS to children who experienced MAM at baseline increased developmental scores by 0.3 SD, which is the largest effect size observed in any sub-group, equivalent to about 5 IQ points, and more than three times larger than the overall effect of SQ-LNS. This suggests that these children have high potential to benefit from LNS distribution programs in developmental outcomes, and that investment in such programs will advance not only child survival, but also fulfillment of developmental potential among the most vulnerable.

SQ-LNS can fill nutrient gaps in children’s diets in key nutrients that are necessary for brain development. Given that provision of SQ-LNS has been documented to positively affect not only nutritional status and growth, but also child survival and development, it is one of the few interventions that can address multiple pillars of the United Nations’ Global Strategy for Women’s Children’s and Adolescents’ Health (2016–2030), which targets three pillars of survival (ending preventable deaths), thriving (ensuring health and well-being), and transformation (expanding enabling environments).

## Supporting information

PRISMA Checklist

## Data Availability

Data described in the manuscript, code book, and analytic code will not be made available because they are compiled from 14 different trials, and access is under the control of the investigators of each of those trials.

## Abbreviations

ASQI: Ages and Stages Questionnaire Inventory
BMI: body mass index
BSID: Bayley Scales of Infant Development
CDI: MacArthur-Bates Communicative Development Inventory
CMAM: Community management of acute malnutrition
DMC: Developmental Milestones Checklist
EASQ: Extended Ages and Stages Questionnaire
Hb: hemoglobin
IPD: individual participant data
KDI: Kilifi Developmental Inventory
LAZ: length-for-age z-score
LMIC: low- and middle-income country
LNS: lipid-based nutrient supplements
MAM: moderate acute malnutrition
MDAT: Malawi Developmental Assessment Tool
MUAC: mid-upper arm circumference
PSED: Profile of Social and Emotional Development
SD: standard deviations
SES: socio-economic status
SQ-LNS: small-quantity lipid-based nutrient supplements
WASH: water, sanitation, and hygiene
WLZ: weight-for-length z-score

## Acknowledgments

We thank all of the co-investigators, collaborators, study teams, participants and local communities involved in the trials included in these analyses. These trials benefitted from the contributions of many partner organizations, including: icddr,b (JiVitA-4, Rang-Din Nutrition Study and WASH Benefits trial in Bangladesh); the World Food Program (JiVitA-4 trial in Bangladesh); the Health District of Dandé and the relevant local health-care authorities (iLiNS-ZINC trial in Burkina Faso); AfricSanté and Helen Keller International (PROMIS trials in Burkina Faso and Mali); Ministry of Public Health and Population (Haiti trial); Innovations for Poverty Action and the Kenya Medical Research Institute (WASH-Benefits trial in Kenya); Unité Programme National de Nutrition Communautaire, Government of Madagascar, and World Bank Health and Nutrition and Population Global Practice (MAHAY trial in Madagascar); the Ministry of Health and Child Care in Harare, Chirumanzu and Shurugwi districts, and Midlands Province (SHINE trial in Zimbabwe); the International Lipid-based Nutrient Supplements Project Steering Committee (iLiNS Project trials); and Nutriset (for development of SQ-LNS). We thank Emily Smith and Julian Higgins for advice on IPD analysis methods.

## Author contributions

The authors’ responsibilities were as follows—ELP: drafted the manuscript with input from KGD, KRW, CDA, CPS and other coauthors; KRW, CDA, KGD, ELP and CPS: wrote the statistical analysis plan; BFA, PA, EB, LH and JHH: reviewed, contributed to, and approved the statistical analysis plan; KRW and CDA: compiled the data; CDA: conducted the data analysis; and all authors: read, contributed to, and approved the final manuscript.

## Supplemental Tables

Supplemental Table 1. Detailed characteristics of each trial included in analysis

Supplemental Table 2. Amount of SQ-LNS provided (g/day) and nutrient value (per daily ration)

Supplemental Table 3. Descriptive information on potential study-level effect modifiers by trial

Supplemental Table 4. Descriptive information on potential individual-level effect modifiers by trial

Supplemental Table 5. Prevalence of motor milestones acquired at age 12 and 18 months by trial

Supplemental Table 6. Risk of bias assessment in each trial

## Supplemental Figures

Supplemental Figure 1. Summary risk of bias as a percentage of all included studies for the effects of SQ-LNS on developmental outcomes

Supplemental Figure 2. Sensitivity analyses of main effects of SQ-LNS on developmental outcomes

Supplemental Figure 3. Forest plots for all main effects of SQ-LNS on developmental outcomes

Supplemental Figure 4. Forest plots for effects of SQ-LNS on developmental outcomes stratified by study-level effect modifiers

Supplemental Figure 5. Forest plots for effects of SQ-LNS on developmental outcomes stratified by individual-level maternal and child effect modifiers

Supplemental Figure 6. Forest plots for effects of SQ-LNS on developmental outcomes stratified by all individual-level household effect modifiers

## References

1. Couperus JW, Nelson CA. Early brain development and plasticity. Edtion ed. In: McCartney K, Phillips D, eds. The Blackwell Handbook of Early Childhood Development. Malden, MA: Blackwell Publsihing, 2006:85–105.

2. Prado EL, Dewey KG. Nutrition and brain development in early life. Nutrition Reviews 2014;72(4):267–84. doi: 10.1111/nure.12102.

3. Barks A, Hall AM, Tran PV, Georgieff MK. Iron as a model nutrient for understanding the nutritional origins of neuropsychiatric disease. Pediatr Res 2019;85(2):176–82. doi: 10.1038/s41390-018-0204-8.

4. Dewey KG. The challenge of meeting nutrient needs of infants and young children during the period of complementary feeding: an evolutionary perspective. The Journal of nutrition 2013;143(12):2050–4. doi: 10.3945/jn.113.182527.

5. Arimond M, Zeilani M, Jungjohann S, Brown KH, Ashorn P, Allen LH, Dewey KG. Considerations in developing lipid-based nutrient supplements for prevention of undernutrition: experience from the International Lipid-Based Nutrient Supplements (iLiNS) Project. Maternal & Child Nutrition 2013. doi: 10.1111/mcn.12049.

6. Das JK, Salam RA, Hadi YB, Sadiq Sheikh S, Bhutta AZ, Weise Prinzo Z, Bhutta ZA. Preventive lipid-based nutrient supplements given with complementary foods to infants and young children 6 to 23 months of age for health, nutrition, and developmental outcomes. Cochrane Database Syst Rev 2019;5:CD012611. doi: 10.1002/14651858.CD012611.pub3.

7. Tam E, Keats EC, Rind F, Das JK, Bhutta AZA. Micronutrient Supplementation and Fortification Interventions on Health and Development Outcomes among Children Under-Five in Low- and Middle-Income Countries: A Systematic Review and Meta-Analysis. Nutrients 2020;12(2). doi: 10.3390/nu12020289.

8. Burke DL, Ensor J, Riley RD. Meta-analysis using individual participant data: one-stage and two-stage approaches, and why they may differ. Stat Med 2017;36(5):855–75. doi: 10.1002/sim.7141.

9. Wessells R, Dewey K, Stewart C, Arnold C, Prado E. Internet: https://www.crd.york.ac.uk/prospero/display_record.php?ID=CRD42020159971.

10. Wessells R, Stewart C, Arnold C, Dewey K, Prado E. Internet: https://osf.io/ymsfu.

11. Stewart LA, Clarke M, Rovers M, Riley RD, Simmonds M, Stewart G, Tierney JF, Group P-ID. Preferred Reporting Items for Systematic Review and Meta-Analyses of individual participant data: the PRISMA-IPD Statement. JAMA 2015;313(16):1657–65. doi: 10.1001/jama.2015.3656.

12. Dewey KG, Wessells KR, Arnold CD, Prado EL, Abbeddou S, Adu-Afarwuah S, Ali H, Arnold BF, Ashorn P, Ashorn U, et al. Characteristics that modify the effect of small-quantity lipid-based nutrient supplementation on child growth: an individual participant data meta-analysis of randomized controlled trials. Am J Clin Nutr In Preparation.

13. Higgins J, Green S. Cochrane Handbook for Systematic Reviews for Interventions, Version 5.1.0. : The Cochrane Collaboration, 2011.

14. Guyatt GH, Oxman AD, Sultan S, Glasziou P, Akl EA, Alonso-Coello P, Atkins D, Kunz R, Brozek J, Montori V, et al. GRADE guidelines: 9. Rating up the quality of evidence. J Clin Epidemiol 2011;64(12):1311–6. doi: 10.1016/j.jclinepi.2011.06.004.

15. Johnston BC, Guyatt GH. Best (but oft-forgotten) practices: intention-to-treat, treatment adherence, and missing participant outcome data in the nutrition literature. The American Journal of Clinical Nutrition 2016;104(5):1197–201. doi: 10.3945/ajcn.115.123315.

16. Veroniki AA, Jackson D, Viechtbauer W, Bender R, Bowden J, Knapp G, Kuss O, Higgins JP, Langan D, Salanti G. Methods to estimate the between-study variance and its uncertainty in meta-analysis. Res Synth Methods 2016;7(1):55–79. doi: 10.1002/jrsm.1164.

17. Paule RC, Mandel J. 1982. Consensus values and weighting factors. National Institute of Standards and Technology.

18. Fisher DJ, Copas AJ, Tierney JF, Parmar MK. A critical review of methods for the assessment of patient-level interactions in individual participant data meta-analysis of randomized trials, and guidance for practitioners. J Clin Epidemiol 2011;64(9):949–67. doi: 10.1016/j.jclinepi.2010.11.016.

19. Higgins JP, Thompson SG, Deeks JJ, Altman DG. Measuring inconsistency in meta-analyses. BMJ (Clinical research ed 2003;327(7414):557–60. doi: 10.1136/bmj.327.7414.557.

20. Streiner DL. Best (but oft-forgotten) practices: the multiple problems of multiplicity-whether and how to correct for many statistical tests. Am J Clin Nutr 2015;102(4):721–8. doi: 10.3945/ajcn.115.113548.

21. Christian P, Shaikh S, Shamim AA, Mehra S, Wu L, Mitra M, Ali H, Merrill RD, Choudhury N, Parveen M, et al. Effect of fortified complementary food supplementation on child growth in rural Bangladesh: a cluster-randomized trial. Int J Epidemiol 2015;44(6):1862–76. doi: 10.1093/ije/dyv155.

22. Dewey KG, Mridha MK, Matias SL, Arnold CD, Cummins JR, Khan MS, Maalouf-Manasseh Z, Siddiqui Z, Ullah MB, Vosti SA. Lipid-based nutrient supplementation in the first 1000 d improves child growth in Bangladesh: a cluster-randomized effectiveness trial. Am J Clin Nutr 2017;105(4):944–57. doi: 10.3945/ajcn.116.147942.

23. Luby SP, Rahman M, Arnold BF, Unicomb L, Ashraf S, Winch PJ, Stewart CP, Begum F, Hussain F, Benjamin-Chung J, et al. Effects of water quality, sanitation, handwashing, and nutritional interventions on diarrhoea and child growth in rural Bangladesh: a cluster randomised controlled trial. Lancet Glob Health 2018;6(3):e302–e15. doi: 10.1016/S2214-109X(17)30490-4.

24. Hess SY, Abbeddou S, Jimenez EY, Somé JW, Vosti SA, Ouédraogo ZP, Guissou RM, Ouédraogo J-B, Brown KH. Small-quantity lipid-based nutrient supplements, regardless of their zinc content, increase growth and reduce the prevalence of stunting and wasting in young Burkinabe children: a cluster-randomized trial. PLoS One 2015;10(3):e0122242. doi: 10.1371/journal.pone.0122242.

25. Adu-Afarwuah S, Lartey A, Brown KH, Zlotkin S, Briend A, Dewey KG. Randomized comparison of 3 types of micronutrient supplements for home fortification of complementary foods in Ghana: effects on growth and motor development. Am J Clin Nutr 2007;86(2):412–20.

26. Adu-Afarwuah S, Lartey A, Okronipa H, Ashorn P, Peerson JM, Arimond M, Ashorn U, Zeilani M, Vosti S, Dewey KG. Small-quantity, lipid-based nutrient supplements provided to women during pregnancy and 6 mo postpartum and to their infants from 6 mo of age increase the mean attained length of 18-mo-old children in semi-urban Ghana: a randomized controlled trial. Am J Clin Nutr 2016;104(3):797–808. doi: 10.3945/ajcn.116.134692.

27. Iannotti LL, Dulience SJ, Green J, Joseph S, Francois J, Antenor ML, Lesorogol C, Mounce J, Nickerson NM. Linear growth increased in young children in an urban slum of Haiti: a randomized controlled trial of a lipid-based nutrient supplement. Am J Clin Nutr 2014;99(1):198–208. doi: 10.3945/ajcn.113.063883.

28. Null C, Stewart CP, Pickering AJ, Dentz HN, Arnold BF, Arnold CD, Benjamin-Chung J, Clasen T, Dewey KG, Fernald LCH, et al. Effects of water quality, sanitation, handwashing, and nutritional interventions on diarrhoea and child growth in rural Kenya: a cluster-randomised controlled trial. Lancet Glob Health 2018;6(3):e316–e29. doi: 10.1016/S2214-109X(18)30005-6.

29. Galasso E, Weber AM, Stewart CP, Ratsifandrihamanana L, Fernald LCH. Effects of nutritional supplementation and home visiting on growth and development in young children in Madagascar: a cluster-randomised controlled trial. Lancet Glob Health 2019;7(9):e1257–e68. doi: 10.1016/s2214-109x(19)30317-1.

30. Ashorn P, Alho L, Ashorn U, Cheung YB, Dewey KG, Gondwe A, Harjunmaa U, Lartey A, Phiri N, Phiri TE, et al. Supplementation of Maternal Diets during Pregnancy and for 6 Months Postpartum and Infant Diets Thereafter with Small-Quantity Lipid-Based Nutrient Supplements Does Not Promote Child Growth by 18 Months of Age in Rural Malawi: A Randomized Controlled Trial. J Nutr 2015;145(6):1345–53. doi: 10.3945/jn.114.207225.

31. Maleta KM, Phuka J, Alho L, Cheung YB, Dewey KG, Ashorn U, Phiri N, Phiri TE, Vosti SA, Zeilani M, et al. Provision of 10-40 g/d Lipid-Based Nutrient Supplements from 6 to 18 Months of Age Does Not Prevent Linear Growth Faltering in Malawi. J Nutr 2015;145(8):1909–15. doi: 10.3945/jn.114.208181.

32. Huybregts L, Le Port A, Becquey E, Zongrone A, Barba FM, Rawat R, Leroy JL, Ruel MT. Impact on child acute malnutrition of integrating small-quantity lipid-based nutrient supplements into community-level screening for acute malnutrition: A cluster-randomized controlled trial in Mali. PLoS Med 2019;16(8):e1002892. doi: 10.1371/journal.pmed.1002892.

33. Humphrey JH, Mbuya MNN, Ntozini R, Moulton LH, Stoltzfus RJ, Tavengwa NV, Mutasa K, Majo F, Mutasa B, Mangwadu G, et al. Independent and combined effects of improved water, sanitation, and hygiene, and improved complementary feeding, on child stunting and anaemia in rural Zimbabwe: a cluster-randomised trial. Lancet Glob Health 2019;7(1):e132–e47. doi: 10.1016/S2214-109X(18)30374-7.

34. Prendergast AJ, Chasekwa B, Evans C, Mutasa K, Mbuya MNN, Stoltzfus RJ, Smith LE, Majo FD, Tavengwa NV, Mutasa B, et al. Independent and combined effects of improved water, sanitation, and hygiene, and improved complementary feeding, on stunting and anaemia among HIV-exposed children in rural Zimbabwe: a cluster-randomised controlled trial. Lancet Child Adolesc Health 2019;3(2):77–90. doi: 10.1016/S2352-4642(18)30340-7.

35. Smuts CM, Matsungo TM, Malan L, Kruger HS, Rothman M, Kvalsvig JD, Covic N, Joosten K, Osendarp SJM, Bruins MJ, et al. Effect of small-quantity lipid-based nutrient supplements on growth, psychomotor development, iron status, and morbidity among 6- to 12-mo-old infants in South Africa: a randomized controlled trial. Am J Clin Nutr 2019;109(1):55–68. doi: 10.1093/ajcn/nqy282.

36. Hedges LV, Olkin I. Statistical methods for meta-analysis. San Diego, CA: Academic Press, 1985. 37.

37. Matias SL, Mridha MK, Tofail F, Arnold CD, Khan MS, Siddiqui Z, Ullah MB, Dewey KG. Home fortification during the first 1000 d improves child development in Bangladesh: a cluster-randomized effectiveness trial. Am J Clin Nutr 2017;105(4):958–69. doi: 10.3945/ajcn.116.150318.

38. Tofail F, Fernald LCH, Das KK, Rahman M, Ahmed T, Jannat KK, Unicomb L, Arnold BF, Ashraf S, Winch PJ, et al. Effect of water quality, sanitation, hand washing, and nutritional interventions on child development in rural Bangladesh (WASH Benefits Bangladesh): a cluster-randomised controlled trial. The Lancet Child & Adolescent Health 2018;2(4):255–68. doi: https://doi.org/10.1016/S2352-4642(18)30031-2.

39. Prado EL, Abbeddou S, Yakes Jimenez E, Some JW, Ouedraogo ZP, Vosti SA, Dewey KG, Brown KH, Hess SY, Ouedraogo JB. Lipid-Based Nutrient Supplements Plus Malaria and Diarrhea Treatment Increase Infant Development Scores in a Cluster-Randomized Trial in Burkina Faso. J Nutr 2016. doi: 10.3945/jn.115.225524.

40. Prado EL, Adu-Afarwuah S, Lartey A, Ocansey M, Ashorn P, Vosti SA, Dewey KG. Effects of pre- and post-natal lipid-based nutrient supplements on infant development in a randomized trial in Ghana. Early Human Development 2016;99:43–51. doi: http://dx.doi.org/10.1016/j.earlhumdev.2016.05.011.

41. Iannotti L, Jean Louis Dulience S, Wolff P, Cox K, Lesorogol C, Kohl P. Nutrition factors predict earlier acquisition of motor and language milestones among young children in Haiti. Acta Paediatr 2016;105(9):e406–11. doi: 10.1111/apa.13483.

42. Stewart CP, Kariger P, Fernald L, Pickering AJ, Arnold CD, Arnold BF, Hubbard AE, Dentz HN, Lin A, Meerkerk TJ, et al. Effects of water quality, sanitation, handwashing, and nutritional interventions on child development in rural Kenya (WASH Benefits Kenya): a cluster-randomised controlled trial. Lancet Child Adolesc Health 2018;2(4):269–80. doi: 10.1016/S2352-4642(18)30025-7.

43. Prado EL, Maleta K, Ashorn P, Ashorn U, Vosti SA, Sadalaki J, Dewey KG. Effects of maternal and child lipid-based nutrient supplements on infant development: a randomized trial in Malawi. Am J Clin Nutr 2016;103(3):784–93. doi: 10.3945/ajcn.115.114579.

44. Prado EL, Phuka J, Maleta K, Ashorn P, Ashorn U, Vosti SA, Dewey KG. Provision of Lipid-Based Nutrient Supplements from Age 6 to 18 Months Does Not Affect Infant Development Scores in a Randomized Trial in Malawi. Matern Child Health J 2016;20(10):2199–208. doi: 10.1007/s10995-016-2061-6.

45. Gladstone MJ, Chandna J, Kandawasvika G, Ntozini R, Majo FD, Tavengwa NV, Mbuya MNN, Mangwadu GT, Chigumira A, Chasokela CM, et al. Independent and combined effects of improved water, sanitation, and hygiene (WASH) and improved complementary feeding on early neurodevelopment among children born to HIV-negative mothers in rural Zimbabwe: Substudy of a cluster-randomized trial. PLoS Med 2019;16(3):e1002766. doi: 10.1371/journal.pmed.1002766.

46. Chandna J, Ntozini R, Evans C, Kandawasvika G, Chasekwa B, Majo F, Mutasa K, Tavengwa N, Mutasa B, Mbuya M, et al. Effects of improved complementary feeding and improved water, sanitation and hygiene on early child development among HIV-exposed children: substudy of a cluster randomised trial in rural Zimbabwe. BMJ Glob Health 2020;5(1):e001718. doi: 10.1136/bmjgh-2019-001718.

47. Larson LM, Yousafzai AK. A meta-analysis of nutrition interventions on mental development of children under-two in low- and middle-income countries. Matern Child Nutr 2017;13(1). doi: 10.1111/mcn.12229.

48. Prado EL, Larson LM, Cox K, Bettencourt K, Kubes JN, Shankar AH. Do effects of early life interventions on linear growth correspond to effects on neurobehavioural development? A systematic review and meta-analysis. Lancet Glob Health 2019;7(10):e1398–e413. doi: 10.1016/S2214-109X(19)30361-4.

49. Suchdev PS, Jefferds MED, Ota E, da Silva Lopes K, De-Regil LM. Home fortification of foods with multiple micronutrient powders for health and nutrition in children under two years of age. Cochrane Database Syst Rev 2020;2:CD008959. doi: 10.1002/14651858.CD008959.pub3.

50. Adolph K, Robinson SR. The road to walking: What learning to walk tells us about development. Edtion ed. In: Zelazo PD, ed. Oxford handbook of developmental psychology. New York: Oxford University Press, 2013.

51. WHO Multicentre Growth Reference Study Group. WHO Motor Development Study: Windows of achievement for six gross motor development milestones. Acta Paediatrica Supplement 2006;450:86–95.

52. Dewey KG, Mridha MK, Matias SL, Arnold CD, Young RT. Long-term effects of the Rang-Din Nutrition Study interventions on maternal and child outcomes. Washington, DC:. Washington, DC: FHI 360/Food and Nutrition Technical Assistance Project (FANTA), 2017.

53. Ocansey ME, Adu-Afarwuah S, Kumordzie SM, Okronipa H, Young RR, Tamakloe SM, Oaks BM, Dewey KG, Prado EL. Prenatal and postnatal lipid-based nutrient supplementation and cognitive, social-emotional, and motor function in preschool-aged children in Ghana: a follow-up of a randomized controlled trial. Am J Clin Nutr 2019;109(2):322–34. doi: 10.1093/ajcn/nqy303.

54. Diamond A. Executive functions. Annu Rev Psychol 2013;64:135-68. doi: 10.1146/annurev-psych-113011-143750.

55. Bunge SA, Crone EA. Neural correlates of the development of cognitive control. Edtion ed. In: Rumsey JM, Ernst M, eds. Neuroimaging in developmental clinical neuroscience. Cambridge: Cambridge University Press, 2009.

56. Sudfeld CR, McCoy DC, Danaei G, Fink G, Ezzati M, Andrews KG, Fawzi WW. Linear growth and child development in low- and middle-income countries: a meta-analysis. Pediatrics 2015;135(5):e1266–75. doi: 10.1542/peds.2014-3111.

57. Leroy JL, Frongillo EA. Perspective: What Does Stunting Really Mean? A Critical Review of the Evidence. Adv Nutr 2019;10(2):196–204. doi: 10.1093/advances/nmy101.

58. Wessells KR, Arnold CD, Stewart CP, Prado EL, Abbeddou S, Adu-Afarwuah S, Arnold BF, Ashorn P, Ashorn U, Becquey E, et al. Characteristics that modify the effect of small-quantity lipid-based nutrient supplementation on child anemia and micronutrient status: an individual participant data meta-analysis of randomized controlled trials. Am J Clin Nutr In Preparation.

59. Grantham-McGregor S. A review of studies of the effect of severe malnutrition on mental development. Journal of Nutrition 1995;125(8 Suppl):2233S–8S.

60. Prado EL, Abbeddou S, Adu-Afarwuah S, Arimond M, Ashorn P, Ashorn U, Bendabenda J, Brown KH, Hess SY, Kortekangas E, et al. Predictors and pathways of language and motor development in four prospective cohorts of young children in Ghana, Malawi, and Burkina Faso. J Child Psychol Psychiatry 2017;58(11):1264–75. doi: 10.1111/jcpp.12751.

61. Kingston D, Tough S, Whitfield H. Prenatal and postpartum maternal psychological distress and infant development: a systematic review. Child Psychiatry Hum Dev 2012;43(5):683–714. doi: 10.1007/s10578-012-0291-4.

62. Bradley RH. Constructing and Adapting Causal and Formative Measures of Family Settings: The HOME Inventory as Illustration. J Fam Theor Rev 2015;7(4):381–414. doi: 10.1111/jftr.12108.

63. Weber AM, Rubio-Codina M, Walker SP, van Buuren S, Eekhout I, Grantham-McGregor SM, Araujo MC, Chang SM, Fernald LC, Hamadani JD, et al. The D-score: a metric for interpreting the early development of infants and toddlers across global settings. BMJ Glob Health 2019;4(6):e001724. doi: 10.1136/bmjgh-2019-001724.

64. Stewart CP, Wessells KR, Arnold CD, Huybregts L, Ashorn P, Becquey E, Humphrey JH, Dewey KG. Lipid-based nutrient supplements and all-cause mortality in children 6-24 months of age: a meta-analysis of randomized controlled trials. Am J Clin Nutr 2020;111(1):207–18. doi: 10.1093/ajcn/nqz262.

65. Black MM, Walker SP, Fernald LCH, Andersen CT, DiGirolamo AM, Lu C, McCoy DC, Fink G, Shawar YR, Shiffman J, et al. Early childhood development coming of age: science through the life course. Lancet 2017;389(10064):77–90. doi: 10.1016/S0140-6736(16)31389-7.

66. Matias SL, Harding KL, Muniruzzaman M, Mridha MK, Ullah B, Vosti SA, Dewey KG. Process evaluation of the Rang-Din Nutrition Study: Final Report. Washington, DC: USAID FANTA III, 2017.

67. Grantham-McGregor SM, Fernald LC, Kagawa RM, Walker S. Effects of integrated child development and nutrition interventions on child development and nutritional status. Annals of the New York Academy of Sciences 2014;1308:11–32. doi: 10.1111/nyas.12284.

68. Adams KP, Vosti SA, Ayifah E, Phiri TE, Adu-Afarwuah S, Maleta K, Ashorn U, Arimond M, Dewey KG. Willingness to pay for small-quantity lipid-based nutrient supplements for women and children: Evidence from Ghana and Malawi. Maternal & child nutrition 2018;14(2):e12518. doi: 10.1111/mcn.12518.

69. Humber J, Vosti S, Cummins J, Mridha M, Matias S, Dewey K. The Rang-Din Nutrition Study in rural Bangladesh: the costs and cost-effectiveness of programmatic interventions to improve linear growth at birth and 18 months, and the costs of these interventions at 24 months. Washington, DC: FHI 360/FANTA. 2017.

70. Gustafsson-Wright E, Boggild-Jones I. Measuring the cost of investing in early childhood interventions and applications of a standardized costing tool. Ann N Y Acad Sci 2018;1419(1):74–89. doi: 10.1111/nyas.13679.

71. WHO (World Health Organization). Internet: http://www.who.int/nutrition/publications/guidelines/updates_management_SAM_infantandc hildren/en/.

72. Gera T, Pena-Rosas JP, Boy-Mena E, Sachdev HS. Lipid based nutrient supplements (LNS) for treatment of children (6 months to 59 months) with moderate acute malnutrition (MAM): A systematic review. PLoS One 2017;12(9):e0182096-e. doi: 10.1371/journal.pone.0182096.

73. World Malaria Report 2018. Geneva: World Health Organization; 2018. Available at: https://www.who.int/malaria/publications/world-malaria-report-2018/report/en/ Accessed on: 26 August 2019.

74. WHO, UNICEF Joint Monitoring Programme. Drinking Water. Available at http://washdata.org/monitoring/drinking-water. Accessed on: 26 August 2019.

75. WHO, UNICEF Joint Monitoring Programme. Sanitation. Available at http://washdata.org/monitoring/sanitation. Accessed on: 26 August 2019.

