## Supplemental Tables 1-6 for "Small-quantity lipid-based nutrient supplements for children age 6-24 months: a systematic review and individual participant data meta-analysis of effects on developmental outcomes and effect modifiers"

**Supplemental Table 1:** Description of each study included in the analysis, outcomes assessed, and the analytic contrasts in which they were included[1]

| Country | Author | Trial Name | Years of trial | Intervention Groups | Trial design | Infant Supplementation | | Selection of Developmental Sub-sample | Developmental Assessment Tools Used | | | | Analysis Contrasts | | | |
|---|---|---|---|---|---|---|---|---|---|---|---|---|---|---|---|---|
| | | | | | | Age at start | Duration | | Language | Social-Emotional | Motor | Executive Function | All-trials analysis | Child-only-LNS analysis | Separation of multi-component interventions | Exclusion of passive control arms |
| Bangladesh | Christian 2015 (21) | JiVitA-4 | 2012-2014 | Plumpy'Doz[2] + IYCF counseling | cluster RCT; longitudinal follow-up | 6 mo | 12 mo | Full sample | BSID-III | | BSID-III | | LNS | LNS | LNS | LNS |
| | | | | Chickpea based LNS + IYCF counseling | | | | | | | | | LNS | LNS | LNS | LNS |
| | | | | Rice-lentil LNS + IYCF counseling | | | | | | | | | LNS | LNS | LNS | LNS |
|  |  |  |  | Wheat-soy blend (++) + IYCF counseling |  |  |  |  |  |  |  |  | ----- | ----- | ----- | ----- |
| | | | | IYCF counseling only (Control) | | | | | | | | | control | control | control | control |
| Bangladesh | Dewey 2017 (22, 37) | RDNS | 2011-2015 | LNS-LNS: maternal SQ-LNS during pregnancy and 6 mo post-partum, child SQ-LNS 6-24 mo | cluster RCT; longitudinal follow-up | 6 mo | 18 mo | Full sample | CDI | DMC | DMC | A not B task | LNS | ----- | ----- | ----- |
| | | | | IFA-LNS: maternal IFA during pregnancy and 3 mo post-partum, child SQ-LNS 6-24 mo | | | | | | | | | LNS | LNS | LNS | LNS |
|  |  |  |  | IFA-MNP: maternal IFA during pregnancy and 3 mo post-partum, child MNP 6-24 mo |  |  |  |  |  |  |  |  | ----- | ----- | ----- | ----- |
| | | | | IFA-Control: maternal IFA during pregnancy and 3 mo post-partum, no child supplementation | | | | | | | | | control | control | control | control |
| Bangladesh | Luby 2018 (23, 38) | WASH-B | 2012-2015 | Nutrition: SQ-LNS + IYCF counseling | cluster RCT; cross-sectional surveys | 6 mo | 18 mo | Full sample | CDI | EASQ | EASQ | A not B task | LNS | LNS | LNS | LNS |
|  |  |  |  | Water: family received chlorine for drinking water |  |  |  |  |  |  |  |  | control | control | ----- | control |
|  |  |  |  | Sanitation: family received upgraded latrine, sani-scoop and child potty |  |  |  |  |  |  |  |  | control | control | ----- | control |
|  |  |  |  | Handwashing: family received handwashing stations with soap |  |  |  |  |  |  |  |  | control | control | ----- | control |
| | | | | WASH: family received all water, sanitation and hygiene interventions | | | | | | | | | control | control | control-WSH | control |
| | | | | WASH + nutrition: all water, sanitation, hygiene and nutrition interventions | | | | | | | | | LNS | LNS | LNS-WSH | LNS |
|  |  |  |  | Passive control (no intervention) |  |  |  |  |  |  |  |  | control | control | control | ----- |
| Burkina Faso | Hess 2015 (24, 39) | iLiNS-Zinc | 2010-2012 | LNS-Zn0: SQ-LNS containing 0 mg/d Zn and placebo tablet[3] | cluster RCT; longitudinal follow-up | 9 mo | 9 mo | A subsample of 446 children from the passive control group and 980 children from 3 of the 4 SQ-LNS groups (LNS-Zn0, LNS-Zn10, and LNS-TabZn5) were randomly selected for motor, language, and personal-social assessment at 18 mo of age. This selection was accomplished through a specialized SAS program (SAS Institute), which randomly assigned children to be assessed or not assessed within treatment group and time block. Children from 3 of 4 SQ-LNS groups were chosen because constraints on personnel resources precluded the assessment of all children. Thus, we targeted the minimum dose of zinc (LNS-Zn0), the maximum dose of zinc (LNS-Zn10), the positive control group (LNS-TabZn5), and the passive control group. The middle dose of zinc (LNS-Zn5) was not included in the developmental assessment subsample (DS). | DMC | DMC | DMC |  | LNS | LNS | LNS | ----- |
|  |  |  |  | LNS-Zn5: SQ-LNS containing 5 mg/d Zn and placebo tablet |  |  |  |  |  |  |  |  | LNS | LNS | LNS | ----- |
|  |  |  |  | LNS-Zn10: SQ-LNS containing 10 mg/d Zn and placebo tablet |  |  |  |  |  |  |  |  | LNS | LNS | LNS | ----- |
|  |  |  |  | LNS-TabZn5: SQ-LNS containing 0 mg/d Zn and Zn tablet containing 5 mg/d Zn |  |  |  |  |  |  |  |  | LNS | LNS | LNS | ----- |
|  |  |  |  | Passive control (no intervention) |  |  |  |  |  |  |  |  | control | control | control | ----- |
| Ghana | Adu Afarwuah 2007 (25) |  | 2004-2005 | SQ-LNS | RCT; longitudinal follow-up | 6 mo | 6 mo | Full sample |  |  |  |  | LNS | LNS | LNS | ----- |
|  |  |  |  | MNF |  |  |  |  |  |  |  |  | ----- | ----- | ----- | ----- |
|  |  |  |  | Nutritabs (MMN) |  |  |  |  |  |  |  |  | ----- | ----- | ----- | ----- |
|  |  |  |  | Passive control (no intervention) |  |  |  |  |  |  |  |  | control | control | control | ----- |
| Ghana | Adu Afarwuah 2016 (26, 40) | iLiNS-DYAD | 2009-2014 | LNS: maternal SQ-LNS during pregnancy and 6 mo post-partum, child SQ-LNS 6-18 mo | RCT; longitudinal follow-up | 6 mo | 12 mo | Full sample | CDI | PSED | KDI | A not B task | LNS | ----- | ----- | ----- |
|  |  |  |  | MMN: maternal MMN during pregnancy and 6 mo post-partum, no child supplementation |  |  |  |  |  |  |  |  | control | ----- | ----- | ----- |
|  |  |  |  | IFA: maternal IFA during pregnancy and placebo for 6 mo post-partum, no child supplementation |  |  |  |  |  |  |  |  | control | ----- | ----- | ----- |
| Haiti | Iannotti 2014[4] (27, 41) | | 2011-2012 | SQ-LNS for 6 mo | RCT; longitudinal follow-up | 6 mo - 11 mo | 6 mo | Full sample | Total vowel and consonant sounds | | | | LNS | LNS | LNS | LNS |
| | | | | Active control (standard of care) | | | | | | | | | control | control | control | control |
| Kenya | Null 2018 (28, 42) | WASH-B | 2012-2016 | Nutrition: SQ-LNS + IYCF counseling | cluster RCT; cross-sectional surveys | 6 mo | 18 mo | Full sample | EASQ | EASQ | EASQ | | LNS | LNS | LNS | LNS |
|  |  |  |  | Water: family received chlorine for drinking water |  |  |  |  |  |  |  |  | control | control | ----- | control |
|  |  |  |  | Sanitation: family received upgraded latrine, sani-scoop and child potty |  |  |  |  |  |  |  |  | control | control | ----- | control |
|  |  |  |  | Handwashing: family received handwashing stations with soap |  |  |  |  |  |  |  |  | control | control | ----- | control |
| | | | | WASH: family received all water, sanitation and hygiene interventions | | | | | | | | | control | control | control-WSH | control |

| | | | | | | | | | | | | | | | | |
|---|---|---|---|---|---|---|---|---|---|---|---|---|---|---|---|---|
| | | | | WASH + nutrition: all water, sanitation, hygiene and nutrition interventions | | | | | | | | | LNS | LNS | LNS-WSH | LNS |
|  |  |  |  | Passive control (no intervention) |  |  |  |  |  |  |  |  | control | control | control | ----- |
| | | | | Active control (visits to measure MUAC) | | | | | | | | | control | control | control | control |
| Madagascar | Galasso 2019 (29) | MAHAY | 2014-2016 | T4: early child stimulation + IYCF counseling | cluster RCT; longitudinal follow-up | 6-11 mo | 6-12 mo | Full sample | ASQI | ASQI | ASQI |  | ----- | ----- | ----- | ----- |
|  |  |  |  | T3: maternal SQ-LNS during pregnancy and 6 mo post-partum, child SQ-LNS 6-18 mo + IYCF counseling |  |  |  |  |  |  |  |  | LNS | ----- | ----- | ----- |
| | | | | T2: child SQ-LNS 6-18 mo + IYCF counseling | | | | | | | | | LNS | LNS | LNS | LNS |
| | | | | T1: IYCF counseling | | | | | | | | | control | control | control | control |
| | | | | T0: Control (standard of care) | | | | | | | | | control | control | control | control |
| Malawi | Ashorn 2015 (30, 43) | iLiNS-DYAD | 2011-2014 | LNS: maternal SQ-LNS during pregnancy and 6 mo post-partum, child SQ-LNS 6-18 mo | RCT; longitudinal follow-up | 6 mo | 12 mo | Full sample | CDI | PSED | KDI | A not B task | LNS | ----- | ----- | ----- |
|  |  |  |  | MMN: maternal MMN during pregnancy and 6 mo post-partum, no child supplementation |  |  |  |  |  |  |  |  | control | ----- | ----- | ----- |
|  |  |  |  | IFA: maternal IFA during pregnancy and placebo for 6 mo post-partum, no child supplementation |  |  |  |  |  |  |  |  | control | ----- | ----- | ----- |
| Malawi | Maleta 2015[5] (31, 44) | iLiNS-DOSE | 2009-2012 | SQ-LNS containing milk (10 g/d) | RCT; longitudinal follow-up | 6 mo | 12 mo | Full sample | CDI | PSED | KDI | A not B task | LNS | LNS | LNS | LNS |
| | | | | SQ-LNS containing milk (20 g/d) | | | | | | | | | LNS | LNS | LNS | LNS |
| | | | | SQ-LNS without milk (20 g/d) | | | | | | | | | LNS | LNS | LNS | LNS |
|  |  |  |  | MQ-LNS containing milk (40 g/d) |  |  |  |  |  |  |  |  | ----- | ----- | ----- | ----- |
|  |  |  |  | MQ-LNS without milk (40 g/d) |  |  |  |  |  |  |  |  | ----- | ----- | ----- | ----- |
| | | | | Active control | | | | | | | | | control | control | control | control |
| Mali | Huybregts 2019 (32) | PROMIS | 2015-2017 | SQ-LNS + IYCF counseling | cluster RCT; longitudinal follow-up and cross-sectional surveys | 6 mo | 18 mo | Full sample (Cross-sectional survey only) | DMC | DMC | DMC | | LNS | LNS | LNS | LNS |
| | | | | Active control (standard of care) + IYCF counseling | | | | | | | | | control | control | control | control |
| Zimbabwe | Humphrey 2019 (33, 45) | SHINE[6] | 2013-2017 | IYCF: child SQ-LNS + IYCF counseling | cluster RCT; longitudinal follow-up | 6 mo | 12 mo | Children were eligible for the ECD substudy if they had the trial primary outcomes (linear growth and hemoglobin) measured at 18 months of age, and turned 2 years of age (allowable range 102–112 weeks) between March 2016 and April 30, 2017. | CDI | MDAT | MDAT | A not B task | LNS | LNS | LNS | LNS |
| | Prendergast 2019 (34, 46) | | | WASH : family received ventilated improved pit latrine, handwashing stations, soap, chlorine, child play space | | | | | | | | | control | control | control-WSH | control |
| | | | | WASH and IYCF: child SQ-LNS + IYCF counseling, family received ventilated improved pit latrine, handwashing stations, soap, chlorine, child play space | | | | | | | | | LNS | LNS | LNS-WSH | LNS |
| | | | | Active control (standard of care) | | | | | | | | | control | control | control | control |

[1]IFA, iron-folic acid; IYCF: infant and young child feeding; LNS, lipid-based nutrient supplement; MMN: multiple micronutrients; MNP: multiple micronutrient powder; ORS, oral rehydration solution; RUCF, ready-to-use complementary food; SQ-LNS, small quantity lipid-based nutrient supplement; WASH, water sanitation and hygiene; WSB, wheat soy blend; RCT: randomized controlled trial; BSID: Bayley Scales of Infant Development; CDI: MacArthur-Bates Communicative Development Inventory; DMC: Developmental Milestones Checklist; EASQ: Extended Ages and Stages Questionnaire; ASQI: Ages and Stages Questionnaire Inventory; KDI: Kilifi Developmental Inventory; MDAT: Malawi Developmental Assessment Tool; PSED: Profile of Social and Emotional Development. Minimal IYCF defined as providing minimal counseling on IYCF other than reinforcing the normal IYCF messages alread promoted in that setting. Expanded IYCF defined as providing expanded counseling on IYCF that went beyond the usual messaging.

[2]All supplements were isocaloric, children 6-12 mo received 125 kcal/d, 12-18 mo received 250 kcal/d

[3]All children in the four intervention groups received ORS for diarrhea and treatment for malaria

[4]Trial also included a 3 mo duration intervention arm which is excluded from these analyses as there is no comprable control arm available

[5]Trial is cited as Kumwenda 2014 in Das et al. (Cochrane Database of Systematic Reviews 2019)

[6]Trial was designed *a priori* to present results separately for HIV exposed and un-exposed children; thus considered as two comparisons in all analyses and the presentation of results

**Supplemental Table 2. Amount of LNS provided (g/day) and nutrient value (per daily ration)**

| | Nutributter[1] | iLiNS Project formulation[2,3] | Revised iLiNS Project formulation[4] | Plumpy'Doz (6-11 mo)[5] | Plumpy'Doz (12-18 mo)[5] | Rice-lentil LNS (6-11 mo)[5] | Rice-lentil LNS (12-18 mo)[5] | Chickpea LNS (6-11 mo)[5] | Chickpea LNS (12-18 mo)[5] |
|---|---|---|---|---|---|---|---|---|---|
| Ration (g/day) | 20 | 20 | 20 | 23.2 | 46.4 | 25.7 | 51.4 | 23.6 | 47.2 |
| Total energy (kcal) | 108 | 118 | 118 | 123.4 | 246.8 | 133.9 | 267.8 | 128.6 | 257.2 |
| Protein (g) | 2.56 | 2.6 | 2.6 | 2.9 | 5.9 | 2.8 | 5.7 | 3.5 | 7.1 |
| Fat (g) | 7.08 | 9.6 | 9.6 | 7.9 | 15.8 | 6.9 | 13.9 | 6.6 | 13.2 |
| Linoleic acid (g) | 1.29 | 4.46 | 4.46 | | | | | | |
| α-Linolenic acid (g) | 0.29 | 0.58 | 0.58 | | | | | | |
| Vitamin A (µg RE) | 400 | 400 | 400 | 200 | 400 | 117.7 | 235.4 | 118.5 | 236.9 |
| Vitamin C (mg) | 30 | 30 | 30 | | | | | | |
| Vitamin B1 (mg) | 0.3 | 0.3 | 0.5 | 0.3 | 0.5 | 0.3 | 0.6 | 0.3 | 0.6 |
| Vitamin B2 (mg) | 0.4 | 0.4 | 0.5 | 0.3 | 0.5 | 0.2 | 0.5 | 0.2 | 0.5 |
| Niacin (mg) | 4 | 4 | 6 | 2.8 | 5.6 | 2.6 | 5.1 | 2.4 | 4.7 |
| Folic acid (µg) | 80 | 80 | 150 | 80 | 160.1 | 100 | 199.9 | 118.2 | 236.5 |
| Pantothenic acid (mg) | 1.8 | 1.8 | 2 | 1 | 2 | 1.1 | 2.2 | 1 | 2 |
| Vitamin B6 (mg) | 0.3 | 0.3 | 0.5 | 0.3 | 0.5 | 0.3 | 0.6 | 0.3 | 0.6 |
| Vitamin B12 (µg) | 0.5 | 0.5 | 0.9 | 0.4 | 0.8 | 0.5 | 0.9 | 0.5 | 0.9 |
| Vitamin D (IU) 1 IU = 0.025 ug | 0 | 200 | 200 | | | 226.2 | 452.3 | 226.6 | 453.1 |
| Vitamin E (mg) | 0 | 6 | 6 | 3 | 6 | 4.9 | 9.8 | 4.7 | 9.4 |
| Vitamin K (µg) | 0 | 30 | 30 | | | 11.3 | 22.6 | 11.3 | 22.7 |
| Iron (mg) | 9 | 6 | 9 | 4.5 | 9 | 3.3 | 6.7 | 3.5 | 7.1 |
| Zinc (mg) | 4 | 8 | 8 | 2 | 4 | 2.4 | 4.8 | 2.5 | 5.1 |
| Copper (mg) | 0.2 | 0.34 | 0.34 | 0.2 | 0.3 | 0.2 | 0.4 | 0.3 | 0.5 |
| Calcium (mg) | 100 | 280 | 280 | 193.5 | 387 | 208.2 | 416.3 | 219.5 | 439 |
| Phosphorus (mg) | 82 | 190 | 190 | 137.6 | 275.2 | 61.7 | 123.4 | 72.9 | 145.8 |
| Potassium (mg) | 152 | 200 | 200 | 155 | 310 | 206.6 | 413.3 | 220.7 | 441.3 |
| Magnesium (mg) | 16 | 40 | 40 | 29.9 | 59.9 | 41.6 | 83.3 | 47.7 | 95.3 |
| Selenium (µg) | 10 | 20 | 20 | 8.6 | 17.2 | 7.5 | 14.9 | 7.6 | 15.1 |
| Iodine (µg) | 90 | 90 | 90 | 27.6 | 55.2 | 33.7 | 67.3 | 33.7 | 67.5 |
| Manganese (mg) | 0.08 | 1.2 | 1.2 | 0.1 | 0.1 | 0.5 | 0.9 | 0.4 | 0.9 |

[1]Provided by Adu-Afarwuah 2007 (26), Iannotti 2014 (27)

[2]Provided by Hess 2015 (24), Adu-Afarwuah 2016 (26), Galasso 2019 (29), Ashorn 2015 (30), Maleta 2015 (31), Huybregts 2019 (32), Humphrey 2019 (33), Prendergast 2019 (34). iLiNS (International Lipid-based Nutrient Supplements) Project formulation described in Arimond et al. 2013.

[3]Hess 2015 (24) provided 0-10 mg zinc/d in the SQ-LNS product, plus a 5 mg/d zinc supplement in one intervention arm. Maleta 2015 provided 10-40 g/d of LNS, with and without milk powder, varying by intervention arm (the 40 g arms were not included in this IPD analysis); the micronutrient composition of the LNS was identical across intervention arms, but there were differences in total kcal, protein, fat, linoleic acid and α-linolenic acid.

[4]Provided by Dewey 2017 (22), Luby 2018 (23), Null 2018 (28).

[5]Provided by Christian 2015 (21). Product and quantities differed by intervention arm and age.

**Supplemental Table 3:** Descriptive information on potential study-level effect modifiers, by trial

| Country | Author | Region | Stunting prevalence at 18 mo (control) (%) | Malaria Prevalence (%) | Anemia Prevalence (%) | Water quality (% improved) | Santiation (% improved) | Duration of supplementation | Intensity of visitation | Average SQ-LNS compliance (%) | Compliance definition (in the SQ-LNS group) |
|---|---|---|---|---|---|---|---|---|---|---|---|
| Bangladesh | Christian 2015 (21) | SEAR | 44.2[b] | 0.2[a] | 51.3[a] | 100.0[a] | 77.0[a] | ≤ 12 mo | Weekly | 93.0[a] | % of total intended SQ-LNS consumed (quantity * day) |
| Bangladesh | Dewey 2017 (22) | SEAR | 35.2[b] | 0.2[a] | 51.3[a] | 100.0[a] | 71.1[a] | > 12 mo | Monthly | 97.4[a] | % reporting "high adherence" (≥ 4 days/week) |
| Bangladesh | Luby 2018 (23) | SEAR | 43.4[b] | 0.1[a] | 51.3[a] | 88.5[a] | 94.7[a] | > 12 mo | Weekly | 93.0[a] | Number of sachets consumed in 14 days prior to annual survey/14 |
| Burkina Faso | Hess 2015 (24) | AFR | 39.4[b] | 59.1[b] | 87.8[b] | 26.7[b] | 2.3[b] | ≤ 12 mo | Weekly | 96.8[a] | % of days SQ-LNS reported consumed |
| Ghana | Adu Afarwuah 2007 (25) | AFR | 7.3[a] | 37.4[b] | 76.1[b] | 91.9[a] | 91.5[a] | ≤ 12 mo | Weekly | 88.2[a] | % of days SQ-LNS reported consumed |
| Ghana | Adu Afarwuah 2016 (26) | AFR | 13.0[a] | 36.8[b] | 57.0[a] | 98.4[a] | 97.3[a] | ≤ 12 mo | Weekly | 73.5[b] | % of days SQ-LNS reported consumed |
| Haiti | Iannotti 2014 (27) | AMR | 13.4[a] | 0.9[a] | 65.0[b] | 98.7[a] | 94.0[a] | ≤ 12 mo | Monthly | 97.0[a] | Reported consuming all of the monthly supply of the SQ-LNS during the supplementation period |
| Kenya | Null 2018 (28) | AFR | 32.2[a] | 8.5[a] | 36.3[a] | 68.0[b] | 15.8[b] | > 12 mo | Monthly | 115.0[a] | Number of sachets consumed in 14 days prior to annual survey/14 |
| Madagascar | Galasso 2019 (29) | AFR | 63.0[b] | 5.5[a] | 51.0[a] | 26.9[b] | 0.0[b] | ≤ 12 mo | Monthly | - | Data unavailable. |
| Malawi | Ashorn 2015 (30) | AFR | 34.7[a] | 26.8[b] | 62.5[b] | 91.7[a] | 9.2[b] | ≤ 12 mo | Weekly | 77.1[b] | % of days SQ-LNS reported consumed |
| Malawi | Maleta 2015 (31) | AFR | 46.5[b] | 30.3[b] | 62.5[b] | 91.6[a] | 2.9[b] | ≤ 12 mo | Weekly | 71.6[b] | % of days SQ-LNS reported consumed (considering missed delivery visits) |
| Mali | Huybregts 2019[1] (33) | AFR | 36.4[b] | 39.1[b] | 81.8[b] | 50.8[a] | 75.3[a] | > 12 mo | Monthly | 47.0[b] | caregiver reported receiving SQ-LNS in the previous month |
| Zimbabwe | Humphrey 2019 (33); Prendergast 20 | AFR | 37.2[b] | 9.0[a] | 36.8[a] | 63.6[b] | 34.0[b] | ≤ 12 mo | Monthly | 73.5[b] | received > 11 (80% of expected) deliveries * consumed SQ-LNS in past 24 h (at 12 month visit) |

[1]Study-level effect modifier categorization based on longitudinal cohort to be consistent with the analyses on growth outcomes

[2]Study-level effect modifier categorization was the same for both HIV exposed and un-exposed children

Superscripts a and b designate the two categories, as described in Notes below.

**Abbreviations**: SQ-LNS, small-quantity lipid-based nutrient supplements

**Notes:**

Geograhic region based on WHO regions

Stunting was defined as length-for-age Z score < -2 SD. Stunting prevalence was based on study-specific data at 18 months (when available) in the control arms. Stunting was assessed at 12 months for Adu-Afarwuah 2007 and Iannotti 2014, and at ~25 months of age for Null 2018. These 3 studies were then categorized based on expected changes in stunting prevalence with age. Trials were categorized as (a) low/moderate burden when stunting was < 35% and (b) high burden when stunting was >35% .

Malaria prevalence: Data extracted from Annex, Data table F: Population at risk and restimated malaria cases and deaths, 2010-2017 (wmr2018-annex-table-f.xls); Point estimate (presumed and confirmed malaria cases), divided by population at risk, per 100 persons. Trials were categorized as (a) low burden when malaria was < 10% and (b) high burden when malaria was >10%.

Anemia prevalence: Data extracted from DHS and MICS. Trials were categorized as (a) moderate when anemia was below 60% prevalence and (b) high when anemia was above 60% prevalence.

Water quality: Data based on sample prevalences of improved source water quality. Study-level water quality was considered improved if the main source of drinking water for > 75% of participants was improved; study-level water quality was considered unimproved if the main source of drinking water for < 75% of participants was improved. "Improved water sources are those that have the potential to deliver safe water by nature of their design and construction, and include: piped water, boreholes or tubewells, protected dug wells, protected springs, rainwater and packaged or delivered water". Unimproved water sources include: water from an unprotected dug well or unprotected spring, or surface water (e.g., river, dam, lake, pond, stream, canal or irrigation canal). (washdata.org/monitoring/drinking-water).

Sanitation: Data based on sample prevalences of improved sanitation. Study-level sanitation was considered improved if sanitation services for > 50% of participants were improved; study-level sanitation was considered unimproved if sanitation services for < 50% of participants were improved. "Improved sanitation facilities are those designed to hygienically separate excreta from human contact, and include: flush/pour flush to piped sewer system, septic tanks or pit latrines; ventilated improved pit latrines, composting toilets or pit latrines with slabs". Unimproved sanitation services include: use of pit latrines without a slab or platform, hanging latrines or bucket latrines, or open defecation. (https://washdata.org/monitoring/sanitation)

Compliance: Data extracted from publication; study-specific definitions of compliance are noted in the table. Trials were categorized as (a) high compliance when complinace was > 80% or (b) low compliance when compliance was < 80% compliance.

**References**

World Malaria Report 2018. Geneva: World Health Organization; 2018. Available at: https://www.who.int/malaria/publications/world-malaria-report-2018/report/en/ Accessed on: 26 August 2019

**Supplementary Table 4:** Descriptive information on potential individual-level effect modifiers, by trial

| Country | Author | Maternal height < 150.1 cm (%) | Maternal BMI < 20 kg/m$^2$ (%) | Maternal age < 25 y (%) | Maternal education, completed primary (%) | Maternal depression, below 75th (%) | Child sex, male (%) | Child birth order, first born (%) | Child baseline LAZ < - 2 (%) | Child baseline acute malnutrition, WLZ < -2 or MUAC < 125 mm (%) | Child baseline anemia, hb < 110 g/L (%) | Moderate to severe food insecurity (%) | SES index, below median (%) | Improved source water quality (%) | Improved sanitation access (%) | Home environment, below median (%) | Season at endline, dry (%) |
|---|---|---|---|---|---|---|---|---|---|---|---|---|---|---|---|---|---|
| Bangladesh | Christian 2015 (21) | | | 57.4 | 63.4 | | 50.0 | 79.0* | 24.6 | 19.2 | | 29.9[a] | 49.7 | 100.0 | 77.0 | | 54.4 |
| Bangladesh | Dewey 2017 (22) | 45.5 | 55.5 | 73.0 | 74.2 | 66.1[a] | 50.2 | 40.4 | 22.9 | 7.5 | 60.5 | 37.2[b] | 49.7 | 100.0 | 71.0 | 28.3[b] | 44.4 |
| Bangladesh | Luby 2018 (23) | 45.6 | 54.2 | 56.1 | 71.2 | 74.7[d] | 49.9 | 33.6 | | | | 21.9[b] | 49.4 | 88.4 | 94.9 | 48.9[b] | 55.2 |
| Burkina Faso | Hess 2015 (24) | 2.3 | 36.8 | 41.4 | 3.9 | | 50.3 | 21.9 | 22.6 | 26.6 | 91.7 | 50.4[b] | 44.5 | 27.0 | 2.4 | 39.9[a] | 70.8 |
| Ghana | Adu Afarwuah 2007 (25) | 4.3 | 9.2 | 29.0 | 88.3 | | 52.1 | 40.0 | | | | | 46.3 | 91.9 | 91.5 | | 57.7 |
| Ghana | Adu Afarwuah 2016 (26) | 5.3 | 15.4 | 38.3 | 78.7 | 70.1[a] | 48.5 | 33.2 | 9.9 | 8.5 | 35.2 | 31.0[b] | 49.9 | 98.4 | 97.3 | 37.3[a] | 58.8 |
| Haiti | Iannotti 2014 (27) | | | 30.0 | 84.8 | | 43.5 | 37.0* | 10.0 | 2.2 | | | 49.5 | 98.7 | 94.0 | | |
| Kenya | Null 2018 (28) | 4.0 | 21.8 | 44.5 | 47.6 | 74.0[e] | 48.2 | 21.4 | | | | 10.3[b] | 43.4 | 68.0 | 15.8 | 43.0[b] | 22.5 |
| Madagascar | Galasso 2019 (29) | 36.0 | | 45.5 | 23.5 | 71.5[f] | 49.2 | 28.3 | | | | 28.4[b] | 49.4 | 26.9 | 0.0 | 47.7[c] | 99.9 |
| Malawi | Ashorn 2015 (30) | 13.4 | 39.9 | 50.1 | 15.8 | 74.2[a] | 47.2 | 20.4 | 23.4 | 8.3 | 65.8 | 71.0[b] | 46.3 | 91.7 | 9.2 | 34.7[a] | 71.1 |
| Malawi | Maleta 2015 (31) | 17.0 | 25.7 | 45.5 | 23.7 | | 50.9 | 23.6 | 30.4 | 5.9 | 63.7 | 73.9[b] | 49.0 | 91.7 | 2.7 | 41.3[a] | 59.2 |
| Mali | Huybregts 2019 (cross-sectional) (32) | 3.1 | 27.4 | 47.4 | 10.5 | | 52.2 | 14.3 | | | | | 49.8 | 59.7 | 74.9 | | |
| Zimbabwe | Humphrey 2019 (HIV-) (33) | 3.5 | 14.2 | 44.2 | 96.5 | 75.2[c] | 49.5 | 25.1* | 18.2 | 5.6 | | 19.5[c] | 50.5 | 63.9 | 34.4 | | 99.8 |
| Zimbabwe | Prendergast 2019 (HIV+) (34) | 5.1 | 16.8 | 18.8 | 94.3 | 78.8[c] | 50.8 | 15.0* | 24.4 | 8.1 | | 25.8[c] | 45.5 | 60.9 | 32.8 | | 100.0 |

**Abbreviations**: BMI, body-mass index; LAZ, length-for-age z-score; WLZ, weight-for-length z-score; MUACZ, mid-upper arm circumference z-score; WAZ, weight-for-age z-score; HCZ, head circumference z-score; SES, socio-economic status.

**Notes:**

Maternal depression scales used: a) Edinburgh Postnatal Depression Scale (EPDS) at 6 mo postpartum; b) EPDS at 2 mo postpartum; c) EPDS during pregnancy at enrollment; d) Center for Epidemiological Studies Depression (CESD) Scale at 12 mo postpartum; e) Patient Health Questionnaire (modified) at 24 mo postpartum; f) CESD Scale during pregnancy at enrollment

Baseline anthropometry is measured at enrollment into the study or start of supplementation if supplementation did not begin at enrollment.

Food security scales used: a) Food Access Survey Tool; b) Household Food Insecurity Access Scale (HFAIS); c) Coping Strategy Index

Water and sanitation references - as for study-level effect modifiers

Home environment assessed by the Family Care Indictors tool at: a) 18 mo of age; b) 24 mo of age; c) endline survey

Season is defined at time of outcome assessment as a dichotomous "Rainy" vs "Dry" category based on child-specific average rainfall during the month of measurement and 2 months prior.

*Data on birth order were not available for all children. Consequently, first-born vs later-born status was estimated based on the number of children under 5 years old in the household.

**Supplemental Table 5:** Prevalence of milestone attainment at 12 and 18 months by trial among control arms

| | | 12-month milestones assessed | | | | | 18-month milestones assessed | | | | |
|---|---|---|---|---|---|---|---|---|---|---|---|
| **Country** | **Author** | **Walk without support** | **Walk with support** | **Stand without support** | **Stand with support** | **Crawl** | **Walk without support** | **Walk with support** | **Stand without support** | **Stand with support** | **Crawl** |
| Bangladesh | Christian 2015 (21) | 736/1363 (54.0) | 1408/1418 (99.3) | 1120/1384 (80.9) | 1396/1409 (99.1) | 1357/1369 (99.1) | | | | | |
| Bangladesh | Dewey 2017 (22) | 202/790 (25.6) | 579/746 (77.6) | 351/767 (45.8) | 764/783 (97.6) | 507/742 (68.3) | 796/816 (97.5) | 811/816 (99.4) | 802/816 (98.3) | 813/816 (99.6) | 766/771 (99.4) |
| Bangladesh | Luby 2018 (23) | 375/1474 (25.4) | 1183/1470 (80.5) | 709/1474 (48.1) | 1391/1472 (94.5) | 1117/1480 (75.5) | | | | | |
| Burkina Faso | Hess 2015 (24) | | | | | | 304/342 (88.9) | 339/342 (99.1) | 328/342 (95.9) | 341/342 (99.7) | 339/342 (99.1) |
| Ghana | Adu Afarwuah 2007 (25) | 22/87 (25.3) | 59/87 (67.8) | 43/87 (49.4) | 80/87 (92.0) | | | | | | |
| Ghana | Adu Afarwuah 2016 (26) | 311/663 (46.9) | 654/662 (98.8) | 499/654 (76.3) | 669/670 (99.9) | 654/665 (98.3) | 688/693 (99.3) | 694/694 (100.0) | 690/693 (99.6) | 694/694 (100) | 694/694 (100) |
| Haiti | Iannotti 2014 (27) | 29/79 (36.7) | | 67/108 (62.0) | | 71/79 (89.9) | 59/60 (98.3) | 59/60 (98.3) | 59/60 (98.3) | 59/60 (98.3) | 59/60 (98.3) |
| Kenya | Null 2018 (28) | 646/1604 (40.3) | 1455/1604 (90.7) | 1168/1604 (72.8) | 1498/1604 (93.4) | 1555/1604 (96.9) | | | | | |
| Madagascar | Galasso 2019 (29) | | | | | | | | | | |
| Malawi | Ashorn 2015 (30) | 208/426 (48.8) | 385/426 (90.4) | 353/426 (82.9) | 411/426 (96.5) | 420/426 (98.6) | 423/441 (95.9) | 431/441 (97.7) | 434/441 (98.4) | 430/441 (97.5) | 435/441 (98.6) |
| Malawi | Maleta 2015 (31) | 69/200 (34.5) | 139/200 (69.5) | 151/200 (75.5) | 192/200 (96.0) | 189/200 (94.5) | 207/222 (93.2) | 219/222 (98.6) | 218/222 (98.2) | 220/222 (99.1) | 222/222 (100.0) |
| Mali | Huybregts 2019 (cross-sectional) (32) | 51/151 (33.8) | 112/151 (74.2) | 94/151 (62.3) | 136/151 (90.1) | 142/151 (94.0) | 90/106 (84.9) | 105/106 (99.1) | 101/106 (95.3) | 104/106 (98.1) | 104/106 (98.1) |
| Zimbabwe | Humphrey 2019 (HIV-) (33) | | | | | | | | | | |
| Zimbabwe | Prendergast 2019 (HIV+) (34) | | | | | | | | | | |

**Notes:**

Values are n/total (%)

IPD development

Assessment of Risk of Bias

Supplemental Table 6A: Risk of bias assessment in each trial

| Country | Author | Random sequence generation | Allocation concealment | Blinding participants | Outcome assessment[1] | Incomplete outcome | Selective reporting | Other |
|---|---|---|---|---|---|---|---|---|
| Bangladesh | Christian 2015 (21) | low | low | high | high | low | low | low |
| Bangladesh | Dewey 2017 (22, 37) | low | low | high | high (language, SE)<br>low (motor, EF) | low | low | low |
| Bangladesh | Luby 2018 (23,38) | low | low | high | high | low | low | low |
| Burkina Faso | Hess 2015 (24, 39) | low | low | high | high | high | low | low |
| Ghana | Adu Afarwuah 2007 (25) | low | low | high | low | low | low | low |
| Ghana | Adu Afarwuah 2016 (26, 40) | low | low | high | high (language, SE)<br>low (motor, EF) | low | low | low |
| Haiti | Iannotti 2014 (27, 41) | unclear | low | high | high | low | low | low |
| Kenya | Null 2018 (28, 42) | low | low | high | high | low | low | low |
| Madagascar | Galasso 2019 (29) | low | low | high | high | low | low | low |
| Malawi | Ashorn 2015 (30, 43) | low | low | high | high (language, SE)<br>low (motor, EF) | low | low | low |
| Malawi | Maleta 2015 (31, 44) | low | low | high | high (language, SE)<br>low (motor, EF) | low | low | low |
| Mali | Huybregts 2019 (32) | low | low | high | high | low | low | low |
| Zimbabwe | Humphrey 2019 (33, 45 | low | low | high | high | low | low | low |
| Zimbabwe | Prendergast 2019 (34. 46) | low | low | high | high | low | low | low |

1. Due to the nature of the intervention, blinding of participants was not possible. We considered developmental outcome assessment to be at low risk of bias only when it was clearly specified that data collectors who performed the developmental assessments were not aware of group allocation, and it would be unlikely that they could easily become aware of group allocation (i.e. observation of interventions materials in study communities, non-intervention passive control arms, etc.). We considered all parent-report assessments to be high risk due to lack of participant blinding in all trials. EF: executive function. SE: social-emotional.

Supplemental Table 6B. Risk of bias assessment in Adu-Afarwuah 2007

| Bias | Authors' judgement | Support for judgement |
|---|---|---|
| Random sequence generation (selection bias) | Low risk | **Quote:** "we randomly selected ~75% of the total number of eligible infants to enter the intervention trial. This was done on a weekly basis, when infants were 5 mo of age, by entering the identification numbers of the eligible infants in a dataset, and using an SAS data step (ranuni [1] le 0.75) to select those for the intervention…the NI infants were randomly selected from the pool of initially eligible infants"<br>**Comment:** adequately done |
| Allocation concealment (selection bias) | Low risk | **Quote:** "the infants were randomly assigned (with the use of opaque envelopes with group designation) to receive SP, NT, or NB until 12 mo of age"<br>**Comment:** adequately done |
| Blinding of participants and personnel (performance bias) | High risk | **Quote**: "NT was provided to the mothers in plastic bags, and the NB (20 g/d) was provided in foil packs with screw caps"<br>**Comment:** not adequate |
| Blinding of outcome assessment (detection bias) | Low risk | **Quote:** " A fieldworker who had previously been trained for the World Health Organization (WHO) Multicenter Growth Reference Study (MGRS) completed head circumference, weight, and length measurements at 6, 9, and 12 mo and assessed 4 gross motor milestones (standing with assistance, walking with assistance, standing independently, and walking independently) at 12 mo using protocols described for the MGRS"<br>**Personal communication with investigator (SAA):** "All anthropometric measurements were carried out by dedicated anthropometrists separate from the field workers who delivered the supplements in the homes, and the anthropometrists had no knowledge of the group assignments. At both 6 and 12 months of age, all children were brought to the laboratory where the anthropometric measurements were conducted; in this case, it was neither possible for the anthropometrists to determine group assignments within the intervention groups, nor was it likely that the anthropometrists could remember or know which children belonged to the intervention versus non-intervention groups."<br>**Comment:** adequately done |
| Incomplete outcome data (attrition bias) | Low risk | **Attrition**: NB group = 90/103; Control group = 87/97<br>**Comment:** Minimal attrition and reasons given for loss to follow-up |
| Selective reporting (reporting bias) | Low risk | **Comment**: The trial was registered at clinicaltrials.gov (NCT00379158); outcomes described in the methods section reported in the results section |
| Other bias | Low risk | **Comment**: no other potential sources of bias reported |

Supplemental Table 6C. Risk of bias assessment in Adu-Afarwuah 2016

| **Bias** | **Authors' judgement** | **Support for judgement** |
|---|---|---|
| Random sequence generation (selection bias) | Low risk | **Quote**: "The study statistician at University of California, Davis developed group allocations with the use of a computer generated (SAS version 9.3; SAS Institute) randomization scheme in blocks of 9"<br>**Comment:** adequately done |
| Allocation concealment (selection bias) | Low risk | **Quote**: "At each enrollment, the study nurse offered sealed, opaque envelopes bearing group allocations, 9 envelopes at a time, and the woman picked one to reveal the allocation"<br>**Comment:** adequately done |
| Blinding of participants and personnel (performance bias) | High risk | **Quote:** "It was not possible to blind study workers and participants to the capsules (IFA and MMN supplements) compared with the LNS supplements because of their different appearances" |
| Blinding of outcome assessment (detection bias) | Language: High risk<br>Social-emotional: High risk<br>Motor: Low risk<br>Executive function: Low risk | **Quote**: " Strengths of the study were that participants were allocated randomly to intervention groups, data collectors and analysts were blind to intervention group, there was a low rate of attrition, the developmental assessment tools had been developed in Africa and were suitable for the local context, and data collectors were rigorously trained and demonstrated high inter-rater agreement and inter-tester reliability."<br>**Comment:** parent-report assessments high risk, direct child assessments low risk |
| Incomplete outcome data (attrition bias) | Low risk | **Attrition:** LNS group = 358/397, Control group = 731/800<br>**Comment:** Minimal attrition and reasons given for loss to follow-up |
| Selective reporting (reporting bias) | Low risk | **Comment**: The trial was registered at clinicaltrials.gov (NCT00970866); SAP available online; outcomes described in the methods section reported in the results section |
| Other bias | Low risk | **Comment**: no other potential sources of bias reported |

Supplemental Table 6D. Risk of bias assessment in Ashorn 2015

| **Bias** | **Authors' judgement** | **Support for judgement** |
|---|---|---|
| Random sequence generation (selection bias) | Low risk | **Quote:** "Researcher not involved with the trial created individual randomisation slips (in blocks of 9)"<br>**Comment**: Additional details on randomization provided in Ashorn *et al.* AJCN 2015. |
| Allocation concealment (selection bias) | Low risk | **Quote**: "…packed them in sealed, numbered, opaque randomization envelopes that were stored in numerical order…Eligible pregnant women were requested to choose 1 of the top 6 envelopes in the stack, and the contents of the envelope indicated her participant number and group allocation""<br>**Comment:** adequately done |
| Blinding of participants and personnel (performance bias) | High risk | **Quote:** "The IFA and MMN interventions were provided with double-masked procedures…For the LNS group, we used single-masked procedures; that is field workers who delivered the supplements knew which mothers were receiving LNS, and the participants were advised not to disclose information about their supplements to anyone other than an iLiNS team member"<br>**Comment:** not done |
| Blinding of outcome assessment (detection bias) | Language: High risk<br>Social-emotional: High risk<br>Motor: Low risk<br>Executive function: Low risk | **Quote:** "data collectors who conducted observational developmental assessments were blind to intervention groups, the developmental assessment tools had been developed in Africa and were suitable for the local context, and data collectors were rigorously trained and demonstrated high inter-rater agreement and inter-tester reliability."<br>**Comment:** parent-report assessments high risk, direct child assessments low risk |
| Incomplete outcome data (attrition bias) | Low risk | **Attrition:** LNS group = 221/222, Control 451/456<br>**Comment:** Minimal attrition and reasons given for loss to follow-up |
| Selective reporting (reporting bias) | Low risk | **Comment**: The trial was registered at clinicaltrials.gov ( NCT01239693 ); SAP available online; outcomes described in the methods section reported in the results section |
| Other bias | Low risk | **Comment:** no other potential sources of bias reported |

Supplemental Table 6E. Risk of bias assessment in Dewey 2007

| **Bias** | **Authors' judgement** | **Support for judgement** |
|---|---|---|
| Random sequence generation (selection bias) | Low risk | **Quote:** "For the randomization, the study statistician at UCD first stratified all 64 clusters in the 11 unions by subdistrict and union and then randomly assigned each cluster to 1 of the 4 arms (each containing 16 clusters)"<br>**Comment:** adequately done |
| Allocation concealment (selection bias) | Low risk | **Quote:** "For the randomization, the study statistician at UCD first stratified all 64 clusters in the 11 unions by subdistrict and union and then randomly assigned each cluster to 1 of the 4 arms (each containing 16 clusters)"<br>**Comment**: central randomization of a cluster-randomized trial |
| Blinding of participants and personnel (performance bias) | High risk | **Comment**: participant blinding not possible due to the nature of the intervention (LNS, MNP, Control) |
| Blinding of outcome assessment (detection bias) | Language: High risk<br>Social-emotional: High risk<br>Motor: Low risk<br>Executive function: Low risk | **Quote:** "The trial was a researcher-blind, longitudinal, cluster randomized effectiveness trial"<br>**Comment:** parent-report assessments high risk, direct child assessments low risk |
| Incomplete outcome data (attrition bias) | Low risk | **Attrition**: LNS = 1712/1804; Control = 839/899<br>**Comment:** reasons given for loss to follow-up; missing outcome data balanced in numbers across intervention groups |
| Selective reporting (reporting bias) | Low risk | **Comment:** The trial was registered at ClinicalTrials.gov ( NCT01715038 ); outcomes described in the methods section reported in the results section |
| Other bias | Low risk | **Comment:** no other potential sources of bias reported |

IPD development
Assessment of Risk of Bias

Supplemental Table 6F. Risk of bias assessment in Hess 2015

| Bias | Authors' judgement | Support for judgement |
|---|---|---|
| Random sequence generation (selection bias) | Low risk | **Quote:** "computer-generated an assignment within strata to participate in the intervention cohort ... The same statistician, who was blinded to the intervention, generated a random allocation sequence at the level of the concession for the enrollment of eligible infants in the IC"<br>**Comment:** adequately done |
| Allocation concealment (selection bias) | Low risk | **Quote: "**The same statistician, who was blinded to the intervention, generated a random allocation sequence at the level of the concession for the enrollment of eligible infants in the IC"<br>**Comment:** central randomization of a cluster-randomized trial |
| Blinding of participants and personnel (performance bias) | High risk | **Quote:** "The trial was partially masked, as all participants, field staff and researchers remained blinded to the four intervention groups until data analyses were completed, but were aware which communities were assigned to IC and NIC"<br>**Comment:** IC and NIC non-blinded |
| Blinding of outcome assessment (detection bias) | High risk | **Quote:** "The trial was partially masked, as all participants, field staff and researchers remained blinded to the four intervention groups until data analyses were completed, but were aware which communities were assigned to IC and NIC."<br>**Comment:** IC and NIC non-blinded, all developmental assessments parent-report |
| Incomplete outcome data (attrition bias) | High risk | **Attrition:** LNS group = 746/980; NIC group = 375/446<br>**Comment:** Differential loss to follow-up because participants in the LNS group were excluded if they were absent for more than 3 weeks during the intervention period. |
| Selective reporting (reporting bias) | Low risk | **Comment**: Protocol attached as a supplement in the study paper; registered at ClinicalTrials.gov as NCT 00944281; outcomes described in the methods section reported in the results section |
| Other bias | Low risk | **Comment:** no other potential sources of bias reported |

IPD development

Assessment of Risk of Bias

Supplemental Table 6G. Risk of bias assessment in Humphrey 2019 and Prendergast 2019

| **Bias** | **Authors' judgement** | **Support for judgement** |
|---|---|---|
| Random sequence generation (selection bias) | Low risk | **Quote:** "clusters were allocated (1:1:1:1) to one of four treatment groups"; "the study's statistician used a constrained randomization technique to identify 500 allocation schemes…From these, 10 allocations were randomly selected. The final allocation was selected at a public randomization event attended by elected representatives"<br>**Comments:** Additional details available in Supplementary Materials (Appendix) and at https://osf.io/w93hy and in (SHINE Trial Team, Clin Infect Dis, 2015). |
| Allocation concealment (selection bias) | Low risk | **Quote:** "the study's statistician used a constrained randomization technique to identify 500 allocation schemes…From these, 10 allocations were randomly selected. The final allocation was selected at a public randomization event attended by elected representatives"<br>**Comments:** Additional details available at https://osf.io/w93hy and in (SHINE Trial Team, Clin Infect Dis, 2015). |
| Blinding of participants and personnel (performance bias) | High risk | **Quote**: "masking of participants and fieldworkers was not possible because of the obvious visual differences between interventions"<br>**Comment:** not done |
| Blinding of outcome assessment (detection bias) | High risk | **Quote**: "masking of participants and fieldworkers was not possible because of the obvious visual differences between interventions, but investigators were blinded to treatment groups until the final analysis of each pre-specified outcome."<br>**Comment:** not done |
| Incomplete outcome data (attrition bias) | Low risk | **Attrition:** LNS group = 1011/2400; Control group = 950/2327<br>**Comment:** Subsample selected for developmental assessment; attrition similar across all arms with reasons given |
| Selective reporting (reporting bias) | Low risk | **Comment:** trial registered as NCT01824940 at ClinicalTrials.gov, published protocol (SHINE Trial Team, Clin Infect Dis 2015), research and statistical analysis plan available at https://osf.io/w93hy ; outcomes described in the methods section reported in the results section |
| Other bias | Low risk | **Comment**: no other potential sources of bias reported |

IPD development

Assessment of Risk of Bias

Supplemental Table 6H. Risk of bias assessment in Huybregts 2019

| Bias | Authors' judgement | Support for judgement |
|---|---|---|
| Random sequence generation (selection bias) | Low risk | **Quote**: "we applied stratified random allocation of the HC catchment areas to control and intervention study groups"; "we first stratified the [health centers] by hierarchical clustering"; "random allocation to control or intervention groups was conducted within each stratum during a community ceremony…forty-eight identical pieces of paper with either 'control' or 'intervention' were mixed in a bag…each [health center] director drew one piece of paper"<br>**Comment:** adequately done |
| Allocation concealment (selection bias) | Low risk | "random allocation to control or intervention groups was conducted within each stratum during a community ceremony…forty-eight identical pieces of paper with either 'control' or 'intervention' were mixed in a bag…each [health center] director drew one piece of paper"<br>**Comment**: adequately done |
| Blinding of participants and personnel (performance bias) | High risk | **Quote**: "non-masked, community-based, trial"<br>**Comment**: blinding of participants who received no intervention was not possible |
| Blinding of outcome assessment (detection bias) | High risk | **Quote:** "We used a two-arm, cluster-randomized, non-blinded effectiveness trial"<br>**Comment**: not done, cluster randomized trial at level of HC |
| Incomplete outcome data (attrition bias) | Low risk | **Attrition:** LNS group = 942/1156; Control group = 956/1161[1]<br>**Comment**: reasons provided for loss to follow-up; missing outcome data balanced in numbers across intervention groups |
| Selective reporting (reporting bias) | Low risk | **Comment**: trial registered as NCT02323815 at ClinicalTrials.gov, published protocol (Huybregts BMC Public Health 2017); outcomes described in the methods section reported in the results section; data made available to IPD investigators. |
| Other bias | Low risk | **Comment**: no other potential sources of bias reported |

[1]The developmental outcomes for this trial are not yet published, therefore we used the denominator from the published paper reporting the growth outcomes and the numerator based on the developmental data provided to us for the IPD meta-analysis.

IPD development

Assessment of Risk of Bias

Supplemental Table 6I. Risk of bias assessment in Iannotti 2014

| **Bias** | **Authors' judgement** | **Support for judgement** |
|---|---|---|
| Random sequence generation (selection bias) | Unclear risk | **Quote:** "Random assignment was carried out through an allocation-concealment mechanism whereby sealed paper forms that masked group assignments were drawn from a container by mothers by using a simple random assignment ratio of 1:1:1 for group assignments"<br>**Comment:** randomization procedure not specified |
| Allocation concealment (selection bias) | Low risk | **Quote:** "Random assignment was carried out through an allocation-concealment mechanism whereby sealed paper forms that masked group assignments were drawn from a container by mothers by using a simple random assignment ratio of 1:1:1 for group assignments"<br>**Comment:** adequately done |
| Blinding of participants and personnel (performance bias) | High risk | **Comment**: participant blinding not possible due to the nature of the intervention (LNS, Control) |
| Blinding of outcome assessment (detection bias) | High risk | **Quote:** "The study team was comprised of one study coordinator and 3 enumerators who participated in a 1-wk training session at the beginning of the trial and another refresher training midway through covering the protocol of anthropometric measures, survey administration, and ethics."<br>**Comment:** same enumerators conducting anthropometric assessments as providing supplements |
| Incomplete outcome data (attrition bias) | Low risk | **Attrition:** control group = 150/191; 6-month LNS group = 153/202<br>**Comment:** reasons for loss-to-follow-up mentioned |
| Selective reporting (reporting bias) | Low risk | **Comment**: Trial registered at ClinicalTrials.gov (NCT01552512); outcomes described in the methodology section reported in the results section |
| Other bias | Low risk | **Comment**: no other potential sources of bias reported |

IPD development

Assessment of Risk of Bias

Supplemental Table 6J. Risk of bias assessment in Luby 2018

| **Bias** | **Authors' judgement** | **Support for judgement** |
| --- | --- | --- |
| Random sequence generation (selection bias) | Low risk | **Quote**: Clusters were randomly allocated to treatment using a random number generator by a coinvestigator at University of California, Berkeley (BFA). Each of the eight geographically adjacent clusters was block randomized to the double-sized control arm or one of the six interventions…Geographical matching ensured that arms were balanced across locations and time of measurement."<br>**Comment**: adequately done |
| Allocation concealment (selection bias) | Low risk | **Quote** "Clusters were randomly allocated to treatment using a random number generator by a coinvestigator at University of California, Berkeley (BFA)."<br>**Comment**: central randomization of a cluster-randomized trial |
| Blinding of participants and personnel (performance bias) | High risk | **Comment:** Not done due to the nature of the intervention |
| Blinding of outcome assessment (detection bias) | High risk | **Quote: "**Interventions included distinct visible components so neither participants nor data collectors were masked to intervention assignment, although the data collection and intervention teams were different individuals". "Outcome and adherence was assessed by a team of university graduates who were not involved in the delivery or promotion of interventions."<br>**Comment**: passive control arm; blinding not possible due to the nature of the intervention |
| Incomplete outcome data (attrition bias) | Low risk | **Attrition:** LNS group = 1141/1395; Control group = 3431/4187<br>**Comment:** attrition similar across all arms with reasons given |
| Selective reporting (reporting bias) | Low risk | **Comment** The trial was registered at clinicaltrials.gov (NCT 01590095); SAP and trial protocol available, and published (Arnold BMJ Open 2013); outcomes described in the methods section reported in the results section |
| Other bias | Low risk | **Comment:** no other potential sources of bias reported |

IPD development
Assessment of Risk of Bias

Supplemental Table 6K. Risk of bias assessment in Maleta 2015

| Bias | Authors' judgement | Support for judgement |
|---|---|---|
| Random sequence generation (selection bias) | Low risk | **Quote:** "We used block randomization and a set of opaque envelopes to assign participants to the intervention groups. The randomization list and envelopes were prepared by a study statistician not involved in trial implementation"<br>**Comment:** adequately done |
| Allocation concealment (selection bias) | Low risk | **Quote:** "We used block randomization and a set of opaque envelopes to assign participants to the intervention groups."<br>**Comment:** adequately done |
| Blinding of participants and personnel (performance bias) | High risk | **Comment**: participant blinding not possible due to the nature of the intervention (LNS, Control) |
| Blinding of outcome assessment (detection bias) | Language: High risk<br>Social-emotional: High risk<br>Motor: Low risk<br>Executive function: Low risk | **Quote:** "…and the code was not disclosed to the researchers or to those assessing the outcomes until all data had been entered and verified in a database." "For the LNS group, we used single-masked procedures (i.e., fieldworkers who delivered the supplements knew which children were receiving LNSs, but those who performed the anthropometric measurements or assessed other outcomes were not aware of group allocation)."<br>**Comment:** parent-report assessments high risk, direct child assessments low risk |
| Incomplete outcome data (attrition bias) | Low risk | **Attrition:** LNS = 1231/1612; control group = 248/320<br>**Comment**: similar levels of attrition across groups, reasons for dropout provided |
| Selective reporting (reporting bias) | Low risk | **Comment**: The trial was registered at clinicaltrials.gov ( NCT00945698); SAP and trial protocol available online; outcomes described in the methods section reported in the results section |
| Other bias | Low risk | **Comment:** no other potential sources of bias reported |

IPD development

Assessment of Risk of Bias

Supplemental Table 6L. Risk of bias assessment in Null 2018

| **Bias** | **Authors' judgement** | **Support for judgement** |
|---|---|---|
| Random sequence generation (selection bias) | Low risk | **Quote:** "Clusters were randomly allocated to treatment at the University of California, Berkeley using a random number generator with reproducible seed"<br>**Comment:** adequately done |
| Allocation concealment (selection bias) | Low risk | **Quote:** "Clusters were randomly allocated to treatment at the University of California, Berkeley using a random number generator with reproducible seed"<br>**Comment**: central randomization of a cluster-randomized trial |
| Blinding of participants and personnel (performance bias) | High risk | **Quote:** "Masking participants was not possible"<br>**Comment:** blinding of participants was not possible due to the nature of the intervention |
| Blinding of outcome assessment (detection bias) | High risk | **Quote**: "The health promoters and staff who delivered the interventions were not involved in data collection, but the data collection team could have inferred treatment status if they saw intervention materials in study communities."<br>**Comment:** blinding not possible due to the nature of the intervention |
| Incomplete outcome data (attrition bias) | Low risk | **Comment:** attrition similar across all seven arms with reasons given |
| Selective reporting (reporting bias) | Low risk | **Comment: T**rial registered as<br>NCT01704105 at ClincialTrials.gov. SAP and trial protocol available online, and published (Arnold BMJ Open 2013); outcomes described in the methods section reported in the results section |
| Other bias | Low risk | **Comment:** no other potential sources of bias reported |

IPD development

Assessment of Risk of Bias

Supplemental Table 6M. Risk of bias assessment in Galasso 2019

| Bias | Authors' judgement | Support for judgement |
|---|---|---|
| Random sequence generation (selection bias) | Low risk | **Quote: "**a random generator was used to block-randomise five sites per intervention group per region"… "An up-to-date registry of government-programme eligible women and children… was used as a sampling frame to select households eligible for enrolment in the trial. 30 households were randomly sampled per site, stratified by children's age at baseline"<br>**Comment:** adequately done |
| Allocation concealment (selection bias) | Low risk | **Comment:** central randomization of a cluster-randomized trial |
| Blinding of participants and personnel (performance bias) | High risk | **Quote:** "Due to the nature of the interventions, masking of participants and community health workers was not possible."<br>**Comment:** not done |
| Blinding of outcome assessment (detection bias) | High risk | **Quote:** "Due to the nature of the interventions, masking of participants and community health workers was not possible. Data analysts were not blinded to intervention group assignment due to differences in survey information"<br>**Comment:** not done |
| Incomplete outcome data (attrition bias) | Low risk | **Quote:** "Mothers or children who died before the final assessment were not replaced. Children who had permanently moved outside the programme site catchment area before final assessment were replaced with a randomly drawn child from the site within the same age range. Children and their households who returned to the site between the baseline and final assessment were re-interviewed."<br>**Comment**: similar levels of attrition across groups, reasons for dropout provided |
| Selective reporting (reporting bias) | Low risk | **Quote:** "This trial has been registered with the ISRCTN registry, number ISRCTN14393738."<br>**Comment:** published protocol (Fernald BMC Public Health 2016); outcomes described in the methods section reported in the results section |
| Other bias | Low risk | **Comment:** no other potential sources of bias reported |

References

21. Christian P, Shaikh S, Shamim AA, Mehra S, Wu L, Mitra M, Ali H, Merrill RD, Choudhury N, Parveen M, et al. Effect of fortified complementary food supplementation on child growth in rural Bangladesh: a cluster-randomized trial. Int J Epidemiol 2015;44(6):1862-76. doi: 10.1093/ije/dyv155.
22. Dewey KG, Mridha MK, Matias SL, Arnold CD, Cummins JR, Khan MS, Maalouf-Manasseh Z, Siddiqui Z, Ullah MB, Vosti SA. Lipid-based nutrient supplementation in the first 1000 d improves child growth in Bangladesh: a cluster-randomized effectiveness trial. Am J Clin Nutr 2017;105(4):944-57. doi: 10.3945/ajcn.116.147942.
23. Luby SP, Rahman M, Arnold BF, Unicomb L, Ashraf S, Winch PJ, Stewart CP, Begum F, Hussain F, Benjamin-Chung J, et al. Effects of water quality, sanitation, handwashing, and nutritional interventions on diarrhoea and child growth in rural Bangladesh: a cluster randomised controlled trial. Lancet Glob Health 2018;6(3):e302-e15. doi: 10.1016/S2214-109X(17)30490-4.
24. Hess SY, Abbeddou S, Jimenez EY, Somé JW, Vosti SA, Ouédraogo ZP, Guissou RM, Ouédraogo J-B, Brown KH. Small-quantity lipid-based nutrient supplements, regardless of their zinc content, increase growth and reduce the prevalence of stunting and wasting in young Burkinabe children: a cluster-randomized trial. PLoS One 2015;10(3):e0122242. doi: 10.1371/journal.pone.0122242.
25. Adu-Afarwuah S, Lartey A, Brown KH, Zlotkin S, Briend A, Dewey KG. Randomized comparison of 3 types of micronutrient supplements for home fortification of complementary foods in Ghana: effects on growth and motor development. Am J Clin Nutr 2007;86(2):412-20.
26. Adu-Afarwuah S, Lartey A, Okronipa H, Ashorn P, Peerson JM, Arimond M, Ashorn U, Zeilani M, Vosti S, Dewey KG. Small-quantity, lipid-based nutrient supplements provided to women during pregnancy and 6 mo postpartum and to their infants from 6 mo of age increase the mean attained length of 18-mo-old children in semi-urban Ghana: a randomized controlled trial. Am J Clin Nutr 2016;104(3):797-808. doi: 10.3945/ajcn.116.134692.
27. Iannotti LL, Dulience SJ, Green J, Joseph S, Francois J, Antenor ML, Lesorogol C, Mounce J, Nickerson NM. Linear growth increased in young children in an urban slum of Haiti: a randomized controlled trial of a lipid-based nutrient supplement. Am J Clin Nutr 2014;99(1):198-208. doi: 10.3945/ajcn.113.063883.
28. Null C, Stewart CP, Pickering AJ, Dentz HN, Arnold BF, Arnold CD, Benjamin-Chung J, Clasen T, Dewey KG, Fernald LCH, et al. Effects of water quality, sanitation, handwashing, and nutritional interventions on diarrhoea and child growth in rural Kenya: a cluster-randomised controlled trial. Lancet Glob Health 2018;6(3):e316-e29. doi: 10.1016/S2214-109X(18)30005-6.
29. Galasso E, Weber AM, Stewart CP, Ratsifandrihamanana L, Fernald LCH. Effects of nutritional supplementation and home visiting on growth and development in young children in Madagascar: a cluster-randomised controlled trial. Lancet Glob Health 2019;7(9):e1257-e68. doi: 10.1016/s2214-109x(19)30317-1.
30. Ashorn P, Alho L, Ashorn U, Cheung YB, Dewey KG, Gondwe A, Harjunmaa U, Lartey A, Phiri N, Phiri TE, et al. Supplementation of Maternal Diets during Pregnancy and for 6 Months Postpartum and Infant Diets Thereafter with Small-Quantity Lipid-Based Nutrient Supplements Does Not Promote Child Growth by 18 Months of Age in Rural Malawi: A Randomized Controlled Trial. J Nutr 2015;145(6):1345-53. doi: 10.3945/jn.114.207225.
31. Maleta KM, Phuka J, Alho L, Cheung YB, Dewey KG, Ashorn U, Phiri N, Phiri TE, Vosti SA, Zeilani M, et al. Provision of 10-40 g/d Lipid-Based Nutrient Supplements from 6 to 18 Months of Age Does Not Prevent Linear Growth Faltering in Malawi. J Nutr 2015;145(8):1909-15. doi: 10.3945/jn.114.208181.
32. Huybregts L, Le Port A, Becquey E, Zongrone A, Barba FM, Rawat R, Leroy JL, Ruel MT. Impact on child acute malnutrition of integrating small-quantity lipid-based nutrient supplements into community-level screening for acute malnutrition: A cluster-randomized controlled trial in Mali. PLoS Med 2019;16(8):e1002892. doi: 10.1371/journal.pmed.1002892.
33. Humphrey JH, Mbuya MNN, Ntozini R, Moulton LH, Stoltzfus RJ, Tavengwa NV, Mutasa K, Majo F, Mutasa B, Mangwadu G, et al. Independent and combined effects of improved water, sanitation, and hygiene, and improved complementary feeding, on child stunting and anaemia in rural Zimbabwe: a cluster-randomised trial. Lancet Glob Health 2019;7(1):e132-e47. doi: 10.1016/S2214-109X(18)30374-7.
34. Prendergast AJ, Chasekwa B, Evans C, Mutasa K, Mbuya MNN, Stoltzfus RJ, Smith LE, Majo FD, Tavengwa NV, Mutasa B, et al. Independent and combined effects of improved water, sanitation, and hygiene, and improved complementary feeding, on stunting and anaemia among HIV-exposed children in rural Zimbabwe: a cluster-randomised controlled trial. Lancet Child Adolesc Health 2019;3(2):77-90. doi: 10.1016/S2352-4642(18)30340-7.
35. Smuts CM, Matsungo TM, Malan L, Kruger HS, Rothman M, Kvalsvig JD, Covic N, Joosten K, Osendarp SJM, Bruins MJ, et al. Effect of small-quantity lipid-based nutrient supplements on growth, psychomotor development, iron status, and morbidity among 6- to 12-mo-old infants in South Africa: a randomized controlled trial. Am J Clin Nutr 2019;109(1):55-68. doi: 10.1093/ajcn/nqy282.
36. Hedges LV, Olkin I. Statistical methods for meta-analysis. San Diego, CA: Academic Press, 1985.

IPD development

Assessment of Risk of Bias


37. Matias SL, Mridha MK, Tofail F, Arnold CD, Khan MS, Siddiqui Z, Ullah MB, Dewey KG. Home fortification during the first 1000 d improves child development in Bangladesh: a cluster-randomized effectiveness trial. Am J Clin Nutr 2017;105(4):958-69. doi: 10.3945/ajcn.116.150318.
38. Tofail F, Fernald LCH, Das KK, Rahman M, Ahmed T, Jannat KK, Unicomb L, Arnold BF, Ashraf S, Winch PJ, et al. Effect of water quality, sanitation, hand washing, and nutritional interventions on child development in rural Bangladesh (WASH Benefits Bangladesh): a cluster-randomised controlled trial. The Lancet Child & Adolescent Health 2018;2(4):255-68. doi: https://doi.org/10.1016/S2352-4642(18)30031-2.
39. Prado EL, Abbeddou S, Yakes Jimenez E, Some JW, Ouedraogo ZP, Vosti SA, Dewey KG, Brown KH, Hess SY, Ouedraogo JB. Lipid-Based Nutrient Supplements Plus Malaria and Diarrhea Treatment Increase Infant Development Scores in a Cluster-Randomized Trial in Burkina Faso. J Nutr 2016. doi: 10.3945/jn.115.225524.
40. Prado EL, Adu-Afarwuah S, Lartey A, Ocansey M, Ashorn P, Vosti SA, Dewey KG. Effects of pre- and post-natal lipid-based nutrient supplements on infant development in a randomized trial in Ghana. Early Human Development 2016;99:43-51. doi: http://dx.doi.org/10.1016/j.earlhumdev.2016.05.011.
41. Iannotti L, Jean Louis Dulience S, Wolff P, Cox K, Lesorogol C, Kohl P. Nutrition factors predict earlier acquisition of motor and language milestones among young children in Haiti. Acta Paediatr 2016;105(9):e406-11. doi: 10.1111/apa.13483.
42. Stewart CP, Kariger P, Fernald L, Pickering AJ, Arnold CD, Arnold BF, Hubbard AE, Dentz HN, Lin A, Meerkerk TJ, et al. Effects of water quality, sanitation, handwashing, and nutritional interventions on child development in rural Kenya (WASH Benefits Kenya): a cluster-randomised controlled trial. Lancet Child Adolesc Health 2018;2(4):269-80. doi: 10.1016/S2352-4642(18)30025-7.
43. Prado EL, Maleta K, Ashorn P, Ashorn U, Vosti SA, Sadalaki J, Dewey KG. Effects of maternal and child lipid-based nutrient supplements on infant development: a randomized trial in Malawi. Am J Clin Nutr 2016;103(3):784-93. doi: 10.3945/ajcn.115.114579.
44. Prado EL, Phuka J, Maleta K, Ashorn P, Ashorn U, Vosti SA, Dewey KG. Provision of Lipid-Based Nutrient Supplements from Age 6 to 18 Months Does Not Affect Infant Development Scores in a Randomized Trial in Malawi. Matern Child Health J 2016;20(10):2199-208. doi: 10.1007/s10995-016-2061-6.
45. Gladstone MJ, Chandna J, Kandawasvika G, Ntozini R, Majo FD, Tavengwa NV, Mbuya MNN, Mangwadu GT, Chigumira A, Chasokela CM, et al. Independent and combined effects of improved water, sanitation, and hygiene (WASH) and improved complementary feeding on early neurodevelopment among children born to HIV-negative mothers in rural Zimbabwe: Substudy of a cluster-randomized trial. PLoS Med 2019;16(3):e1002766. doi: 10.1371/journal.pmed.1002766.
46. Chandna J, Ntozini R, Evans C, Kandawasvika G, Chasekwa B, Majo F, Mutasa K, Tavengwa N, Mutasa B, Mbuya M, et al. Effects of improved complementary feeding and improved water, sanitation and hygiene on early child development among HIV-exposed children: substudy of a cluster randomised trial in rural Zimbabwe. BMJ Glob Health 2020;5(1):e001718. doi: 10.1136/bmjgh-2019-001718.
