## Supplemental Figures 1-6 for "Small-quantity lipid-based nutrient supplements for children age 6-24 months: a systematic review and individual participant data meta-analysis of effects on developmental outcomes and effect modifiers"

Supplemental Figure 1: Summary risk of bias as a percentage of all included studies for the effects of SQ-LNS on developmental outcomes

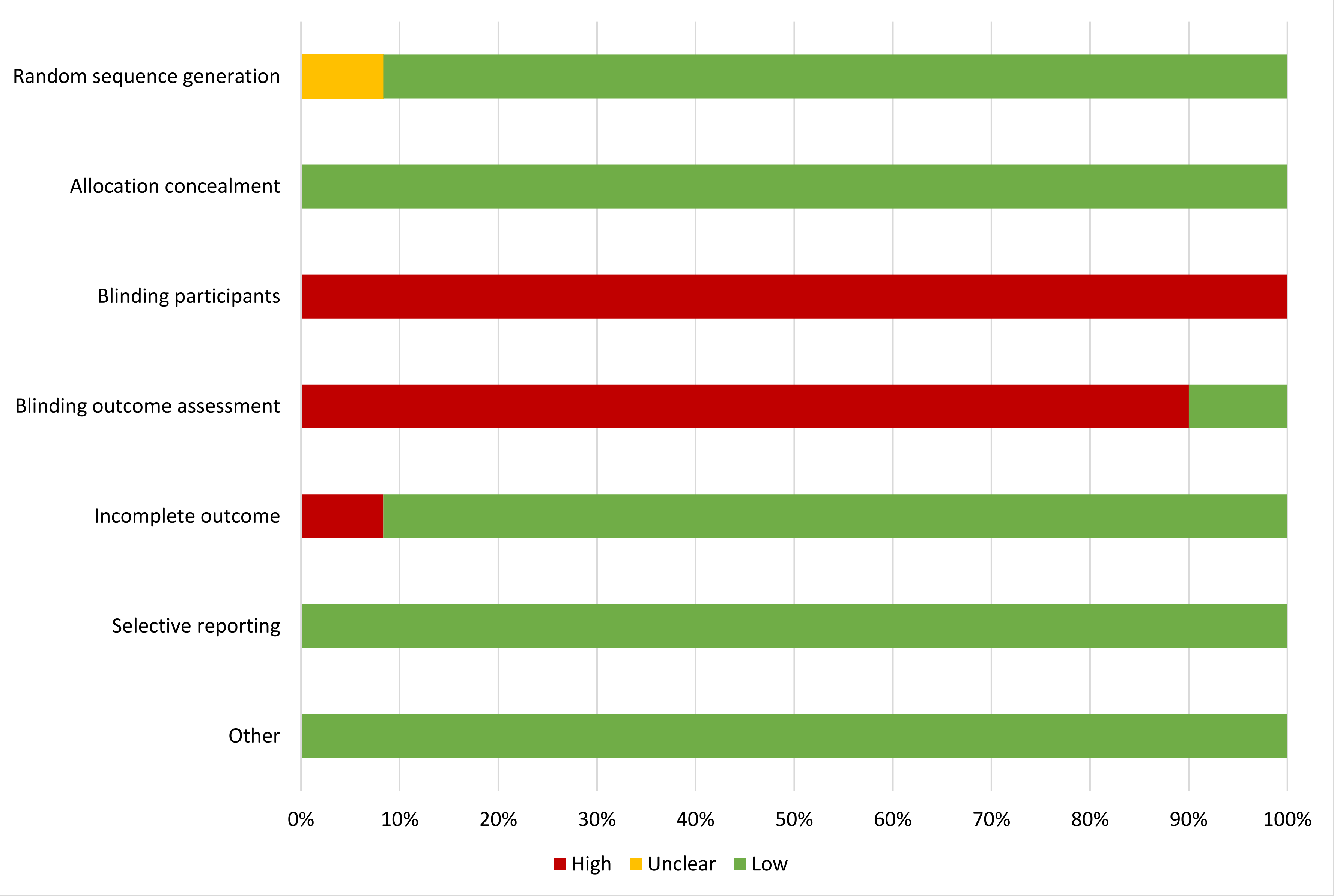

### Supplemental figure 2: Sensitivity analyses of main effects of SQ-LNS on developmental outcomes

#### Contents

**Supplemental figure 2A: Mean differences for continuous outcomes** **2**

**Supplemental figure 2B: Prevalence ratios for dichotomous outcomes** **3**

**Supplemental figure 2C: Prevalence differences for dichotomous outcomes** **4**

**Supplemental figure 2D: Prevalence ratios for 12 mo milestone outcomes** **5**

**Supplemental figure 2E: Prevalence differences for 12 mo milestone outcomes** **6**

**Supplemental figure 2F: Prevalence ratios for 18 mo milestone outcomes** **7**

**Supplemental figure 2G: Prevalence differences for 18 mo milestone outcomes** **8**

These figures show the pooled estimates of intervention effects by different pooling methods and different sensitivity analyses. For continuous outcomes, the intervention effect is measured by the difference in mean of the LNS group minus control. For dichotomous outcomes analyzed via prevalence ratios, the effect estimate is the prevalence in the LNS group divided by the prevalence in the control group. For dichotomous outcomes analyzed via prevalence differences, the effect estimate is the prevalence in the LNS group minus the prevalence in the control group. The labels on the left y-axis indicate which outcome is assessed. The different columns correspond to sensitivity analyses in which intervention group categorization differs. All-trial analysis includes all trials; Child-LNS-only excludes trial arms that provided both maternal and child LNS; Multi-component analysis separates comparisons within trials that included multi-component interventions, so that the SQ-LNS vs. no SQ-LNS comparisons were conducted separately between pairs of arms that included the same non-nutrition components (e.g. SQ-LNS+WASH vs. WASH; SQ-LNS vs. Control); Passive arms excluded analysis excludes passive control arms.

#### Supplemental figure 2A: Mean differences for continuous outcomes

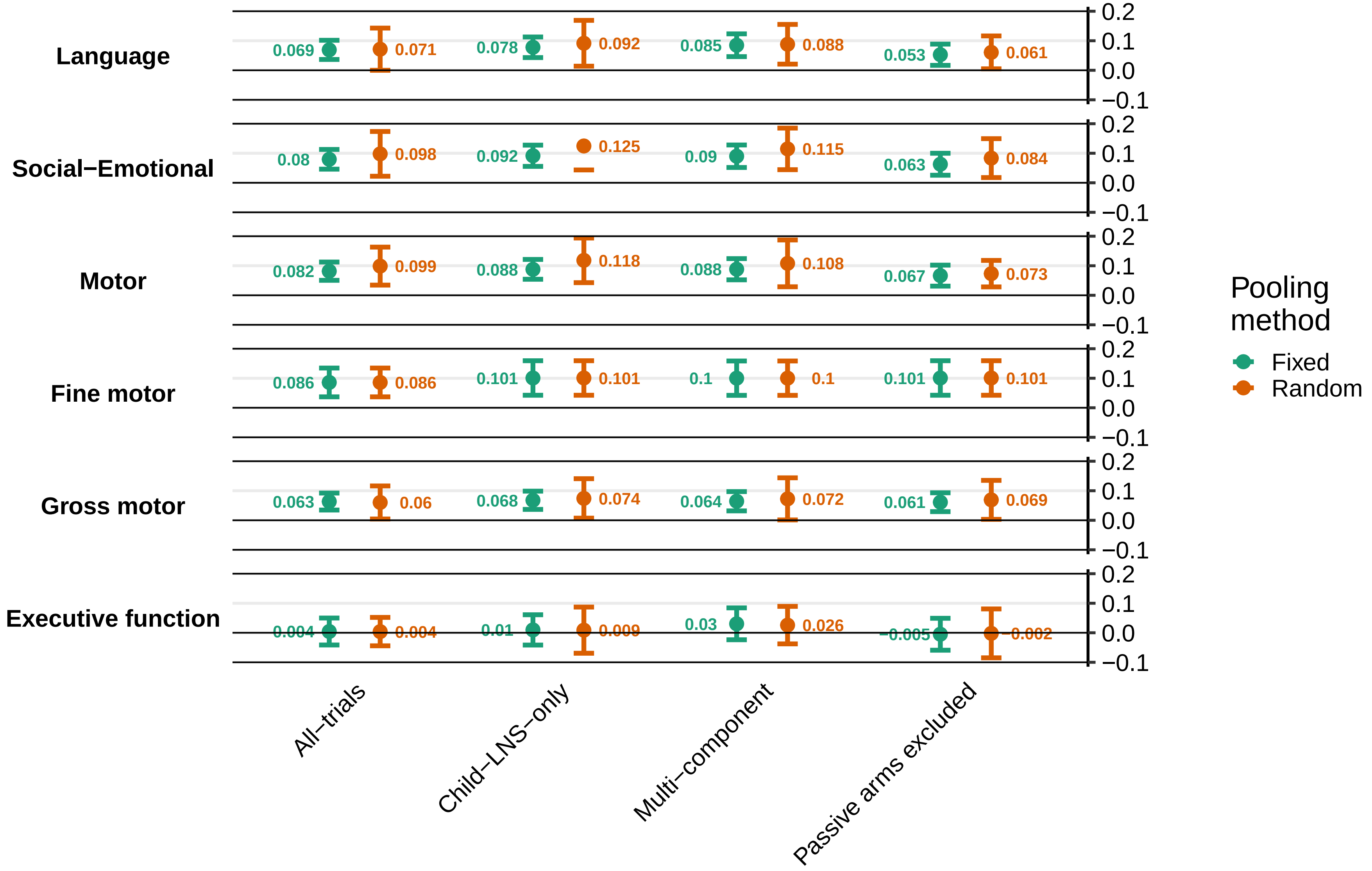

#### Supplemental figure 2B: Prevalence ratios for dichotomous outcomes

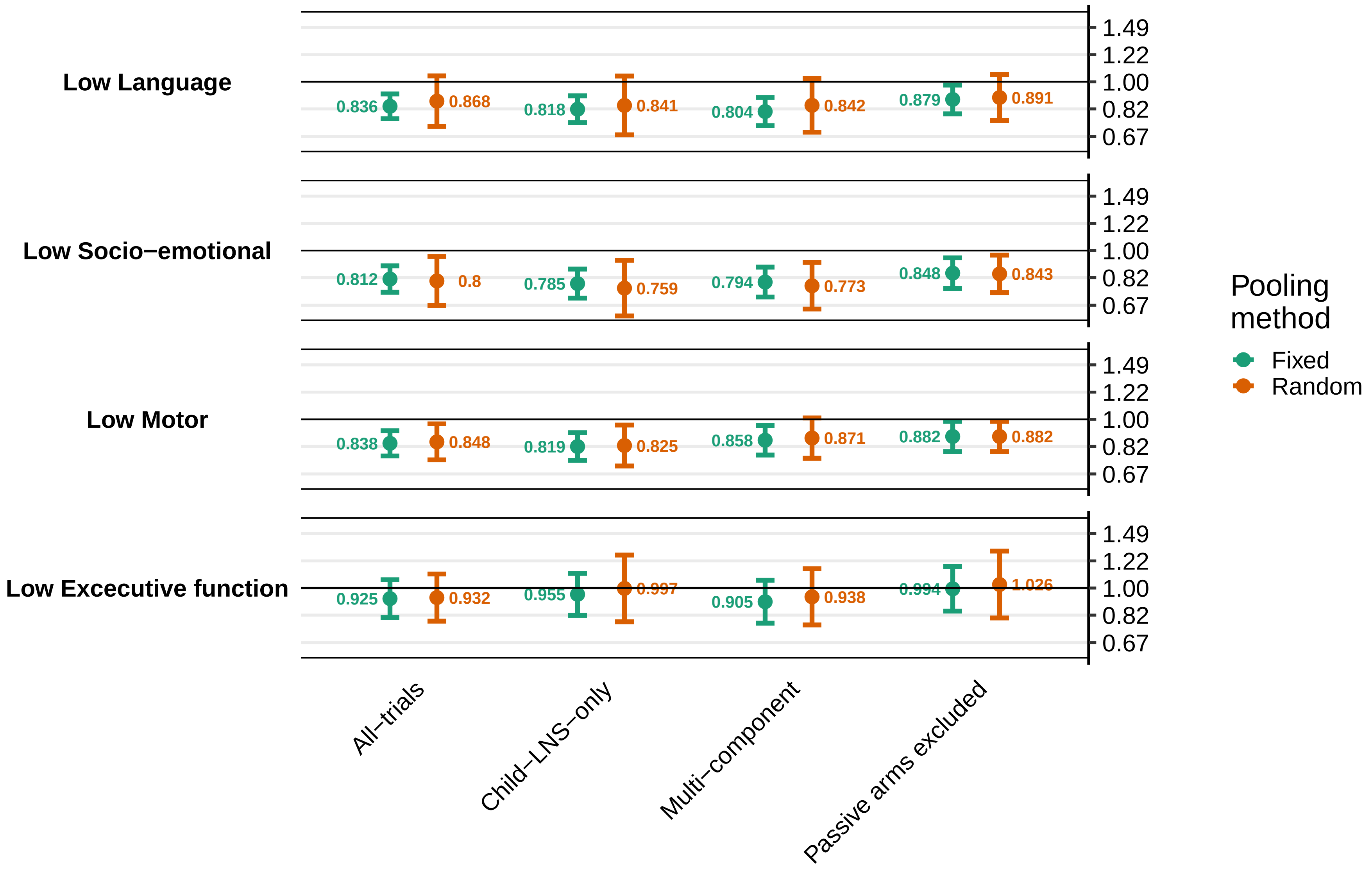

#### Supplemental figure 2C: Prevalence differences for dichotomous outcomes

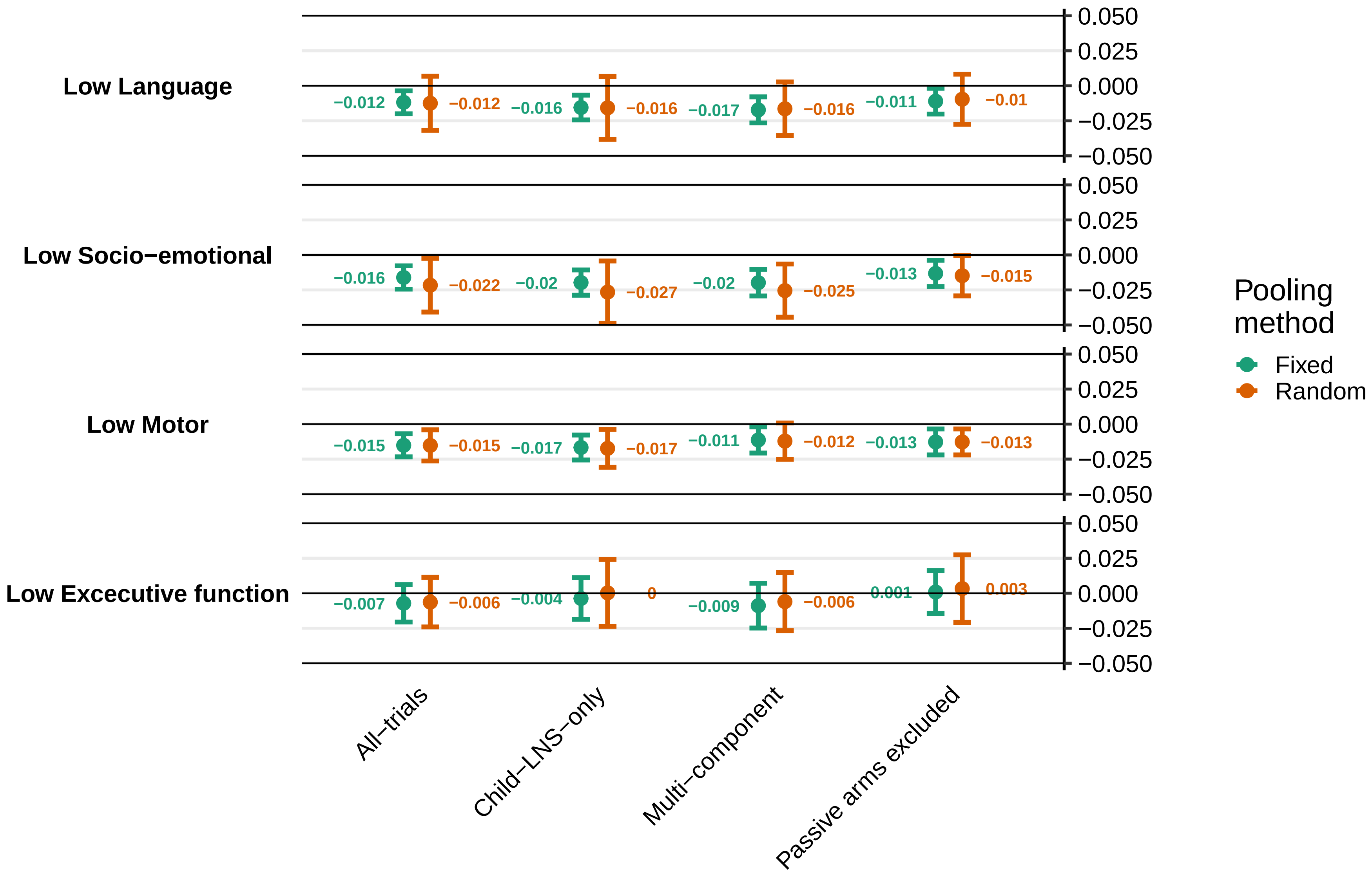

#### Supplemental figure 2D: Prevalence ratios for 12 mo milestone outcomes

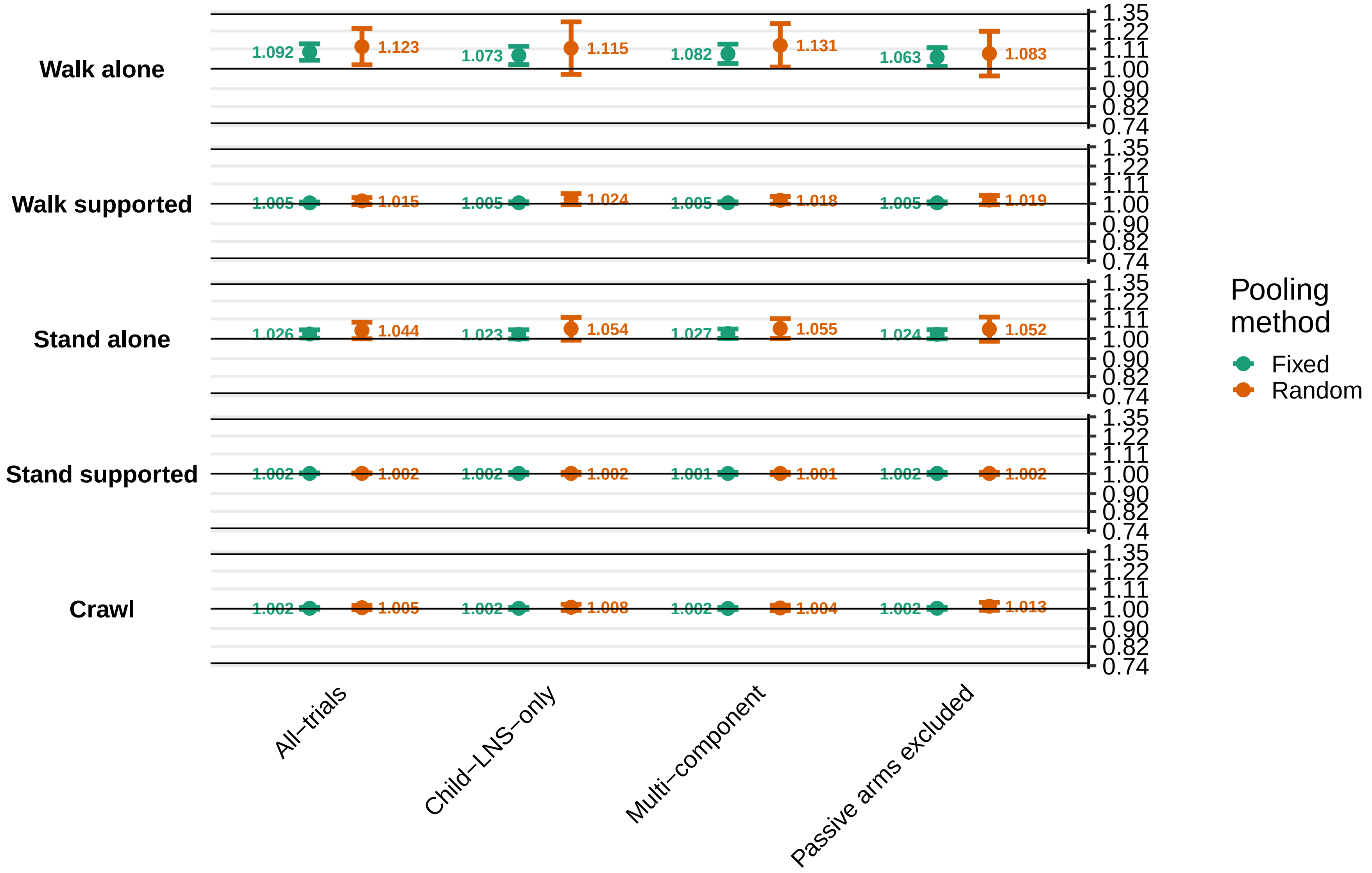

#### Supplemental figure 2E: Prevalence differences for 12 mo milestone outcomes

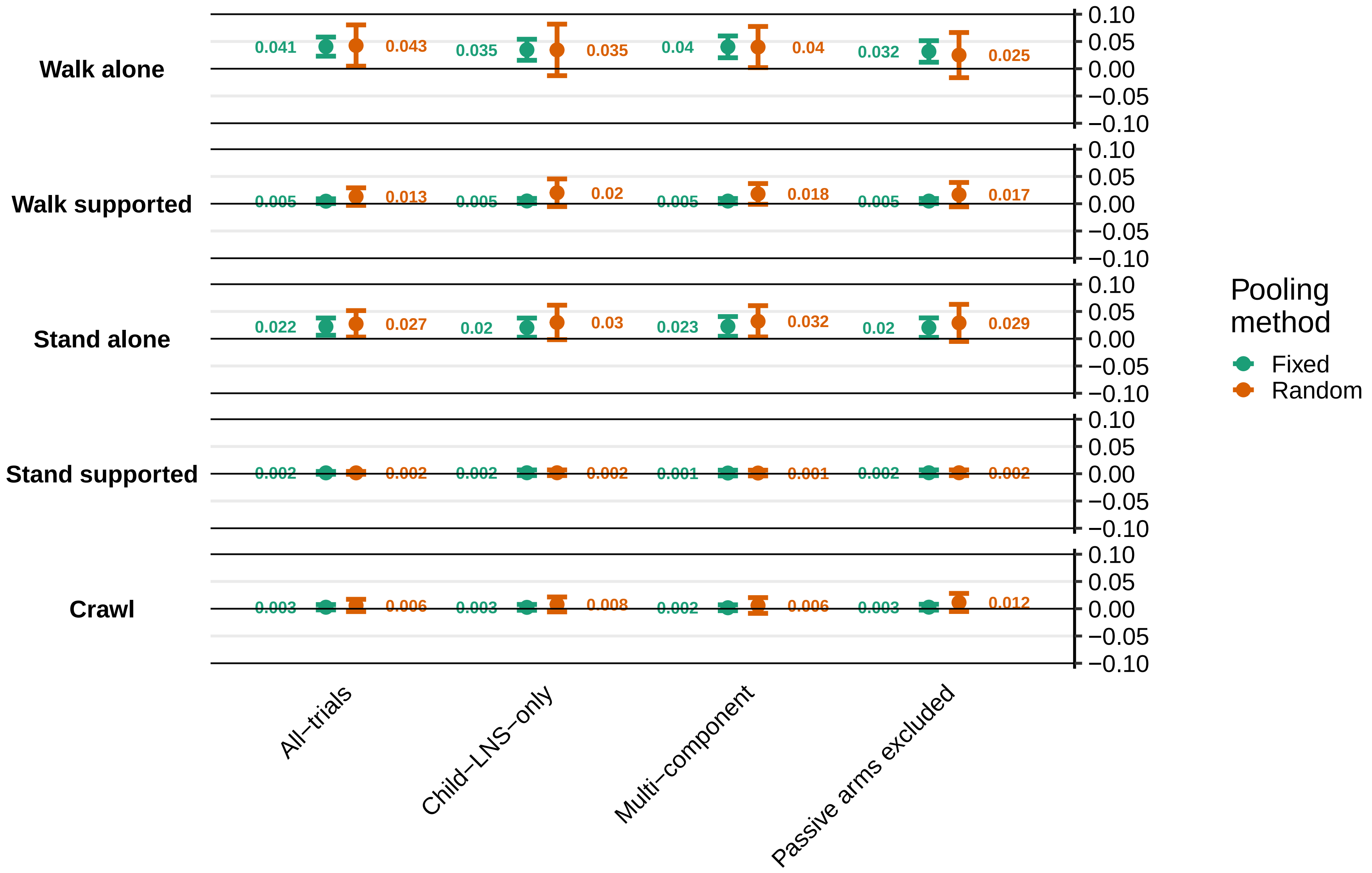

#### Supplemental figure 2F: Prevalence ratios for 18 mo milestone outcomes

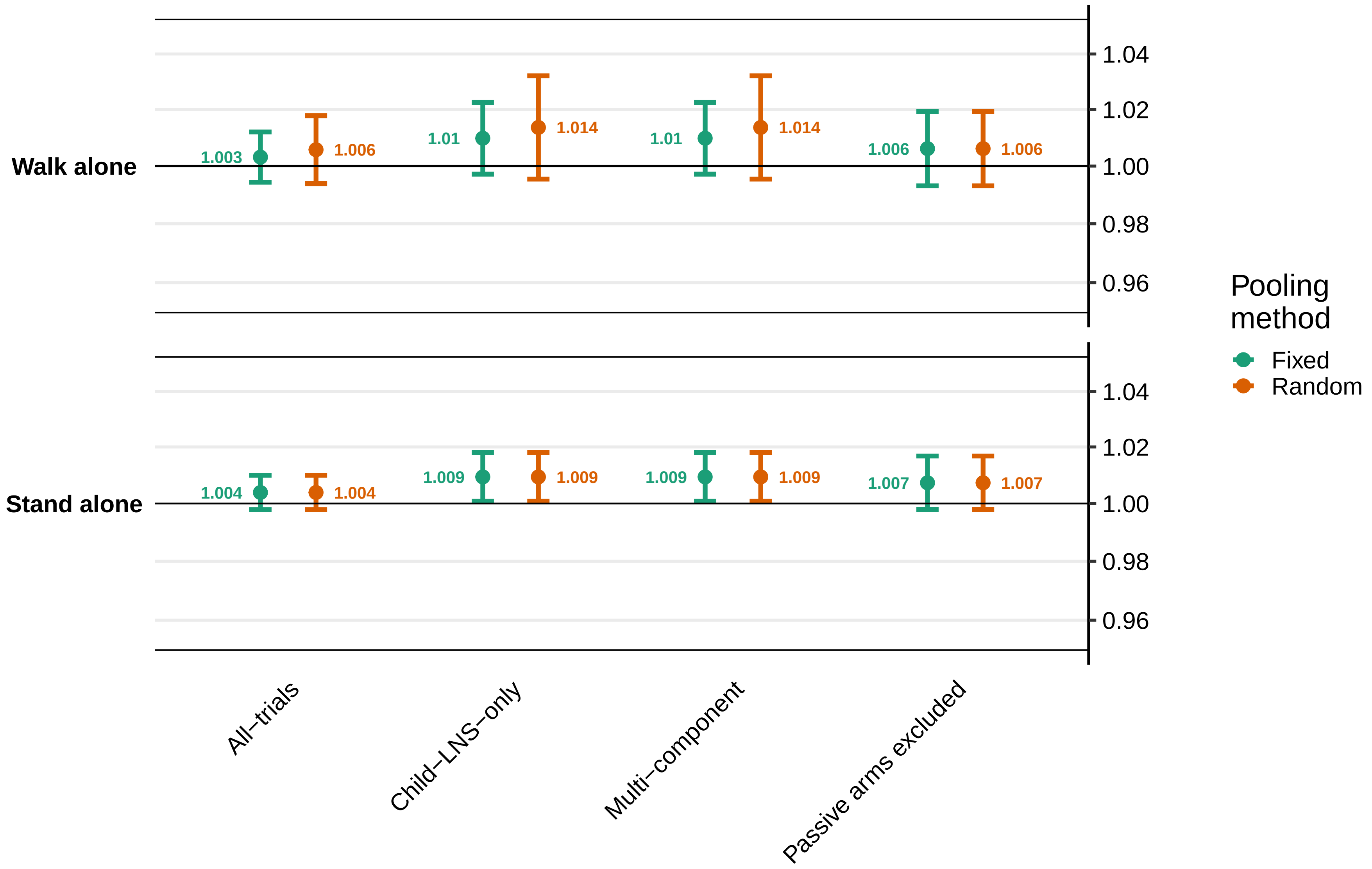

#### Supplemental figure 2G: Prevalence differences for 18 mo milestone outcomes

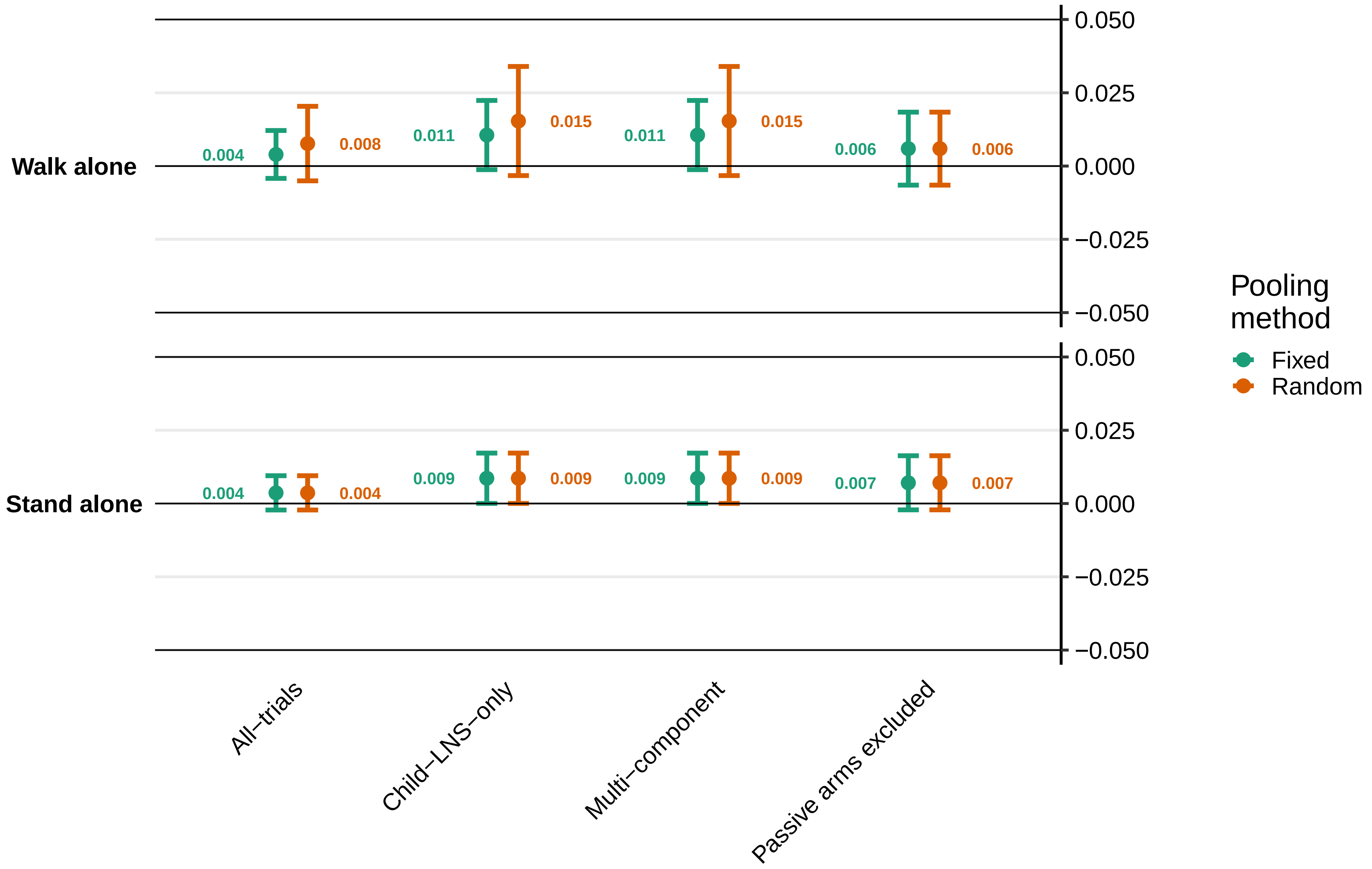

### Supplemental figure 3: Forest plots for all main effects of SQ-LNS on developmental outcomes

#### Contents

**Supplemental figure 3A: Mean difference in language z-score** **3**

**Supplemental figure 3B: Language lowest decile prevalence ratio** **4**

**Supplemental figure 3C: Language lowest decile prevalence difference** **5**

**Supplemental figure 3D: Mean difference in social-emotional z-score** **6**

**Supplemental figure 3E: Social-emotional lowest decile prevalence ratio** **7**

**Supplemental figure 3F: Social-emotional lowest decile prevalence difference** **8**

**Supplemental figure 3G: Mean difference in motor z-score** **9**

**Supplemental figure 3H: Motor lowest decile prevalence ratio** **10**

**Supplemental figure 3I: Motor lowest decile prevalence difference** **11**

**Supplemental figure 3J: Mean difference in gross motor z-score** **12**

**Supplemental figure 3K: Mean difference in fine motor z-score** **13**

**Supplemental figure 3L: Mean difference in executive function z-score** **14**

**Supplemental figure 3M: Executive function lowest decile prevalence ratio** **15**

**Supplemental figure 3N: Executive function lowest decile prevalence difference** **16**

**Supplemental figure 3O: 12-mo walking without support prevalence ratio** **17**

**Supplemental figure 3P: 12-mo walking without support prevalence difference** **18**

**Supplemental figure 3Q: 12-mo walking with support prevalence ratio** **19**

**Supplemental figure 3R: 12-mo walking with support prevalence difference** **20**

**Supplemental figure 3S: 12-mo standing without support prevalence ratio** **21**

**Supplemental figure 3T: 12-mo standing without support prevalence difference** **22**

**Supplemental figure 3U: 12-mo standing with support prevalence ratio** **23**

**Supplemental figure 3V: 12-mo standing with support prevalence difference** **24**

**Supplemental figure 3W: 12-mo crawling prevalence ratio** **25**

**Supplemental figure 3X: 12-mo crawling prevalence difference** **26**

**Supplemental figure 3Y: 18-mo walking without support prevalence ratio** **27**

**Supplemental figure 3Z: 18-mo walking without support prevalence difference** **28**

**Supplemental figure 3AA: 18-mo walking with support prevalence ratio** **29**

**Supplemental figure 3AB: 18-mo walking with support prevalence difference** **30**

These figures are forest plots showing the study-level estimates of intervention effect with the pooled estimate in the bottom summary rows. For continuous outcomes the intervention effect is measured by the difference in mean of the LNS group minus control. For dichotomous outcomes analyzed via prevalence ratios the effect estimate is the prevalence in the LNS group divided by the prevalence in the control group. For dichotomous outcomes analyzed via prevalence differences the effect estimate is the prevalence in the LNS group minus the prevalence in the control group. The labels on the left y-axis correspond to trial level information. The values on the right indicate the study level effect estimate, confidence interval, and weighting for deriving the pooled estimate.

Motor milestone figures show individual trial estimates excluding the JiVitA-4 data because those individual trial results have not yet been published. The pooled estimate in each figure includes the JiVitA-4 data, for consistency with all other analyses.

Figures showing individual trial estimates for the SHINE trial are split by comparison to reflect the cross-over design. For calculating the pooled estimates shown in these figures, the trial is analyzed with LNS intervention arms combined and non-LNS intervention arms combined.

### Supplemental figure 3A: Mean difference in language z-score

| Country | Trial | Tool | LNS N | Control N | MD (95% CI) | Fixed W | Random W |
|---|---|---|---|---|---|---|---|
| Bangladesh | JiVitA−4 (21) | BSID−III | 445 | 143 | −0.08 (−0.29, 0.12) | 0.03 | 0.06 |
| Bangladesh | RDNS (22) | CDI | 1663 | 814 | 0.17 (0.07, 0.26) | 0.11 | 0.09 |
| Bangladesh | WASH−B (23) | CDI | 1109 | 3353 | 0.09 (0.02, 0.17) | 0.20 | 0.10 |
| Burkina Faso | iLiNS−Zinc (24) | DMC | 746 | 375 | 0.37 (0.21, 0.53) | 0.04 | 0.07 |
| Ghana | GHANA (25) | | | | | | |
| Ghana | iLiNS−DYADG (26) | CDI | 331 | 658 | 0.01 (−0.12, 0.14) | 0.06 | 0.08 |
| Haiti | HAITI (27) | Total sounds | 150 | 149 | −0.13 (−0.35, 0.10) | 0.02 | 0.05 |
| Kenya | WASH−B (28) | EASQ | 1362 | 4745 | 0.02 (−0.04, 0.08) | 0.30 | 0.11 |
| Madagascar | MAHAY (29) | ASQI | 1613 | 1604 | −0.11 (−0.30, 0.07) | 0.03 | 0.07 |
| Malawi | iLiNS−DYADM (30) | CDI | 215 | 439 | 0.01 (−0.15, 0.18) | 0.04 | 0.07 |
| Malawi | iLiNS−DOSE (31) | CDI | 645 | 221 | 0.05 (−0.11, 0.20) | 0.05 | 0.08 |
| Mali | PROMIS CS (32) | DMC | 927 | 944 | 0.15 (−0.02, 0.31) | 0.04 | 0.07 |
| Zimbabwe | SHINE[1] (HIV−) (33), LNS vs SOC | CDI | 381 | 373 | 0.03 (−0.15, 0.21) | 0.07 | 0.09 |
| Zimbabwe | LNS+WSH vs WSH | CDI | 436 | 408 | 0.13 (−0.02, 0.29) | 0.07 | 0.09 |
| Zimbabwe | SHINE[1] (HIV+) (34), LNS vs SOC | CDI | 66 | 68 | 0.06 (−0.29, 0.40) | 0.02 | 0.05 |
| Zimbabwe | LNS+WSH vs WSH | CDI | 99 | 79 | 0.37 (0.07, 0.67) | 0.02 | 0.05 |
| | **I² = 0.64, Tau² = 0.01** | | **10188** | **14373** | | | |
| | **Fixed** | | | | **0.07 (0.04, 0.10)** | | |
| | **Random** | | | | **0.07 (0.00, 0.14)** | | |

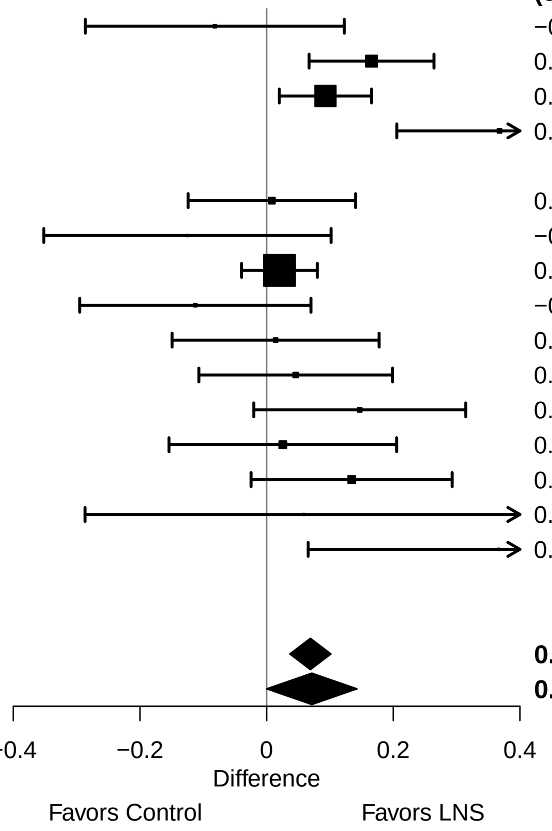

### Supplemental figure 3B: Language lowest decile prevalence ratio

| Country | Trial | Tool | LNS N | Control N | PR (95% CI) | Fixed W | Random W |
|---|---|---|---|---|---|---|---|
| Bangladesh | JiVitA−4 (21) | BSID−III | 445 | 143 | 0.86 (0.50, 1.49) | 0.03 | 0.06 |
| Bangladesh | RDNS (22) | CDI | 1663 | 814 | 0.72 (0.57, 0.91) | 0.15 | 0.10 |
| Bangladesh | WASH−B (23) | CDI | 1109 | 3353 | 0.81 (0.65, 1.00) | 0.18 | 0.10 |
| Burkina Faso | iLiNS−Zinc (24) | DMC | 746 | 375 | 0.46 (0.33, 0.64) | 0.08 | 0.09 |
| Ghana | GHANA (25) | | | | | | |
| Ghana | iLiNS−DYADG (26) | CDI | 331 | 658 | 1.09 (0.74, 1.61) | 0.05 | 0.08 |
| Haiti | HAITI (27) | Total sounds | 150 | 149 | 2.75 (1.28, 5.93) | 0.01 | 0.04 |
| Kenya | WASH−B (28) | EASQ | 1362 | 4745 | 0.95 (0.78, 1.16) | 0.21 | 0.11 |
| Madagascar | MAHAY (29) | ASQI | 1613 | 1604 | 1.17 (0.75, 1.83) | 0.04 | 0.07 |
| Malawi | iLiNS−DYADM (30) | CDI | 215 | 439 | 0.87 (0.53, 1.43) | 0.03 | 0.06 |
| Malawi | iLiNS−DOSE (31) | CDI | 645 | 221 | 0.95 (0.61, 1.50) | 0.04 | 0.07 |
| Mali | PROMIS CS (32) | DMC | 927 | 944 | 0.77 (0.56, 1.07) | 0.08 | 0.09 |
| Zimbabwe | SHINE[1] (HIV−) (33), LNS vs SOC | CDI | 381 | 373 | 1.05 (0.67, 1.65) | 0.08 | 0.09 |
| Zimbabwe | LNS+WSH vs WSH | CDI | 436 | 408 | 0.77 (0.49, 1.20) | 0.08 | 0.09 |
| Zimbabwe | SHINE[1] (HIV+) (34), LNS vs SOC | CDI | 66 | 68 | 1.29 (0.54, 3.06) | 0.02 | 0.05 |
| Zimbabwe | LNS+WSH vs WSH | CDI | 99 | 79 | 0.32 (0.11, 0.92) | 0.02 | 0.05 |
| | **$I^2$ = 0.61, $Tau^2$ = 0.07** | | **10188** | **14373** | | | |
| | **Fixed** | | | | **0.84 (0.76, 0.92)** | | |
| | **Random** | | | | **0.87 (0.72, 1.04)** | | |

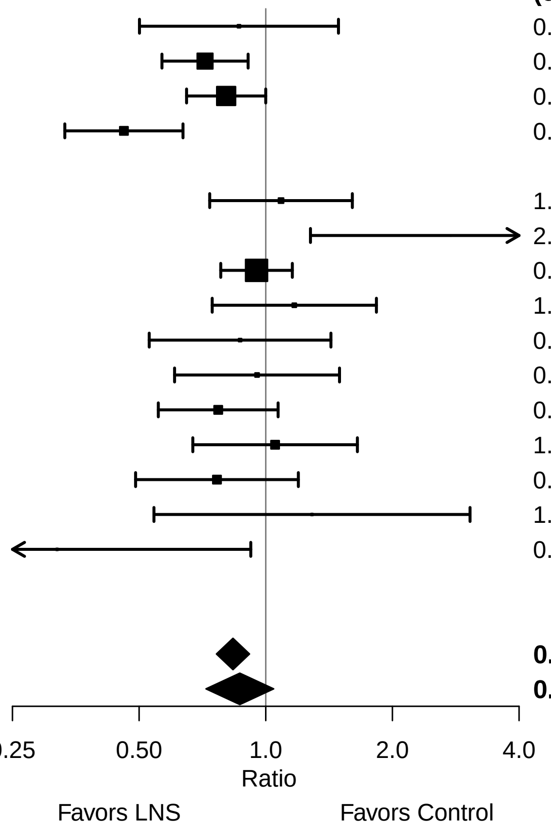

### Supplemental figure 3C: Language lowest decile prevalence difference

| Country | Trial | Tool | LNS N | Control N | PD (95% CI) | Fixed W | Random W |
|---|---|---|---|---|---|---|---|
| Bangladesh | JiVitA−4 (21) | BSID−III | 445 | 143 | −0.02 (−0.07, 0.04) | 0.02 | 0.05 |
| Bangladesh | RDNS (22) | CDI | 1663 | 814 | −0.04 (−0.06, −0.01) | 0.10 | 0.09 |
| Bangladesh | WASH−B (23) | CDI | 1109 | 3353 | −0.02 (−0.04, 0.00) | 0.18 | 0.10 |
| Burkina Faso | iLiNS−Zinc (24) | DMC | 746 | 375 | −0.08 (−0.13, −0.04) | 0.04 | 0.07 |
| Ghana | GHANA (25) | | | | | | |
| Ghana | iLiNS−DYADG (26) | CDI | 331 | 658 | 0.01 (−0.03, 0.05) | 0.04 | 0.07 |
| Haiti | HAITI (27) | Total sounds | 150 | 149 | 0.10 (0.03, 0.16) | 0.02 | 0.05 |
| Kenya | WASH−B (28) | EASQ | 1362 | 4745 | 0.00 (−0.02, 0.01) | 0.21 | 0.10 |
| Madagascar | MAHAY (29) | ASQI | 1613 | 1604 | 0.02 (−0.01, 0.04) | 0.16 | 0.10 |
| Malawi | iLiNS−DYADM (30) | CDI | 215 | 439 | −0.01 (−0.06, 0.03) | 0.03 | 0.07 |
| Malawi | iLiNS−DOSE (31) | CDI | 645 | 221 | 0.00 (−0.05, 0.04) | 0.03 | 0.07 |
| Mali | PROMIS CS (32) | DMC | 927 | 944 | −0.03 (−0.05, 0.00) | 0.09 | 0.09 |
| Zimbabwe | SHINE¹ (HIV−) (33), LNS vs SOC | CDI | 381 | 373 | 0.01 (−0.04, 0.05) | 0.08 | 0.09 |
| Zimbabwe | LNS+WSH vs WSH | CDI | 436 | 408 | −0.03 (−0.06, 0.01) | 0.08 | 0.09 |
| Zimbabwe | SHINE¹ (HIV+) (34), LNS vs SOC | CDI | 66 | 68 | 0.03 (−0.08, 0.15) | 0.01 | 0.05 |
| Zimbabwe | LNS+WSH vs WSH | CDI | 99 | 79 | −0.09 (−0.17, 0.00) | 0.01 | 0.05 |
| | **$I^2$ = 0.66, $Tau^2$ = 0.00** | | **10188** | **14373** | | | |
| | **Fixed** | | | | **−0.01 (−0.02, 0.00)** | | |
| | **Random** | | | | **−0.01 (−0.03, 0.01)** | | |

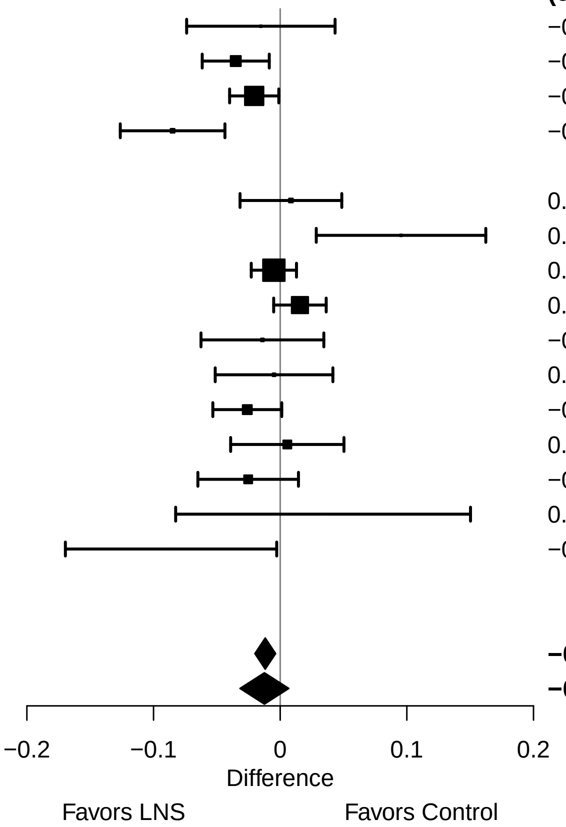

#### Supplemental figure 3D: Mean difference in social-emotional z-score

| Country | Trial | Tool | LNS N | Control N | MD (95% CI) | Fixed W | Random W |
|---|---|---|---|---|---|---|---|
| Bangladesh | JiVitA−4 (21) | | | | | | |
| Bangladesh | RDNS (22) | DMC | 1657 | 815 | 0.06 (−0.04, 0.16) | 0.12 | 0.11 |
| Bangladesh | WASH−B (23) | EASQ | 1066 | 3266 | 0.12 (0.05, 0.20) | 0.21 | 0.11 |
| Burkina Faso | iLiNS−Zinc (24) | DMC | 746 | 375 | 0.36 (0.18, 0.53) | 0.04 | 0.08 |
| Ghana | GHANA (25) | | | | | | |
| Ghana | iLiNS−DYADG (26) | PSED | 332 | 657 | 0.01 (−0.12, 0.14) | 0.06 | 0.09 |
| Haiti | HAITI (27) | | | | | | |
| Kenya | WASH−B (28) | EASQ | 1362 | 4745 | 0.01 (−0.05, 0.08) | 0.27 | 0.12 |
| Madagascar | MAHAY (29) | ASQI | 1613 | 1604 | −0.07 (−0.26, 0.11) | 0.03 | 0.07 |
| Malawi | iLiNS−DYADM (30) | PSED | 215 | 438 | 0.01 (−0.15, 0.18) | 0.04 | 0.08 |
| Malawi | iLiNS−DOSE (31) | PSED | 644 | 221 | 0.03 (−0.12, 0.18) | 0.05 | 0.08 |
| Mali | PROMIS CS (32) | DMC | 927 | 944 | 0.15 (−0.02, 0.31) | 0.04 | 0.08 |
| Zimbabwe | SHINE[1] (HIV−) (33), LNS vs SOC | MDAT | 395 | 382 | 0.07 (−0.07, 0.21) | 0.11 | 0.11 |
| Zimbabwe | LNS+WSH vs WSH | MDAT | 446 | 417 | 0.17 (0.02, 0.31) | 0.11 | 0.11 |
| Zimbabwe | SHINE[1] (HIV+) (34), LNS vs SOC | MDAT | 67 | 68 | 0.24 (0.00, 0.48) | 0.03 | 0.07 |
| Zimbabwe | LNS+WSH vs WSH | MDAT | 103 | 83 | 0.44 (0.14, 0.73) | 0.03 | 0.07 |
| | **$I^2$ = 0.66, $Tau^2$ = 0.01** | | **9573** | **14015** | | | |
| | **Fixed** | | | | **0.08 (0.05, 0.11)** | | |
| | **Random** | | | | **0.10 (0.02, 0.17)** | | |

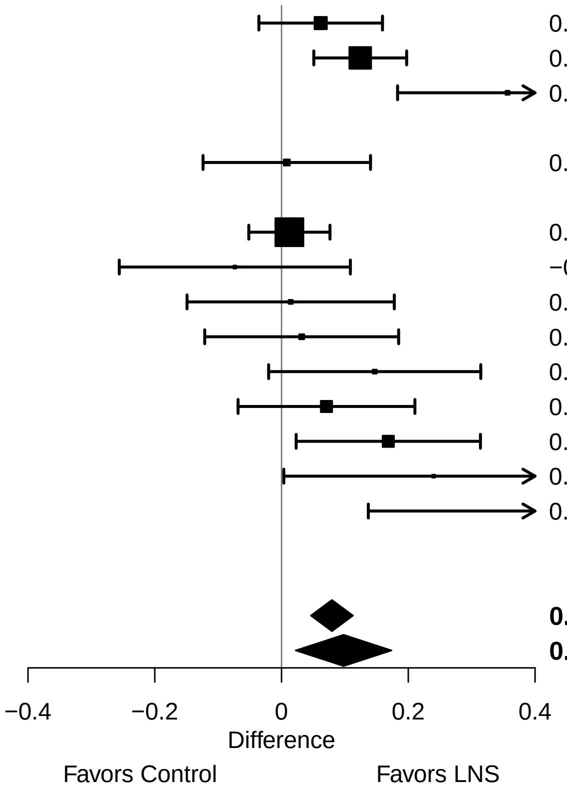

#### Supplemental figure 3E: Social-emotional lowest decile prevalence ratio

| Country | Trial | Tool | LNS N | Control N | PR (95% CI) | Fixed W | Random W |
|---|---|---|---|---|---|---|---|
| Bangladesh | JiVitA−4 (21) | | | | | | |
| Bangladesh | RDNS (22) | DMC | 1657 | 815 | 0.84 (0.66, 1.07) | 0.16 | 0.12 |
| Bangladesh | WASH−B (23) | EASQ | 1066 | 3266 | 0.81 (0.64, 1.03) | 0.17 | 0.12 |
| Burkina Faso | iLiNS−Zinc (24) | DMC | 746 | 375 | 0.41 (0.28, 0.62) | 0.06 | 0.08 |
| Ghana | GHANA (25) | | | | | | |
| Ghana | iLiNS−DYADG (26) | PSED | 332 | 657 | 0.99 (0.67, 1.47) | 0.06 | 0.09 |
| Haiti | HAITI (27) | | | | | | |
| Kenya | WASH−B (28) | EASQ | 1362 | 4745 | 0.87 (0.71, 1.08) | 0.21 | 0.12 |
| Madagascar | MAHAY (29) | ASQI | 1613 | 1604 | 1.15 (0.75, 1.78) | 0.05 | 0.08 |
| Malawi | iLiNS−DYADM (30) | PSED | 215 | 438 | 1.00 (0.61, 1.61) | 0.04 | 0.07 |
| Malawi | iLiNS−DOSE (31) | PSED | 644 | 221 | 1.08 (0.68, 1.72) | 0.04 | 0.07 |
| Mali | PROMIS CS (32) | DMC | 927 | 944 | 0.77 (0.56, 1.07) | 0.09 | 0.10 |
| Zimbabwe | SHINE[1] (HIV−) (33), LNS vs SOC | MDAT | 395 | 382 | 0.86 (0.56, 1.33) | 0.10 | 0.10 |
| Zimbabwe | LNS+WSH vs WSH | MDAT | 446 | 417 | 0.62 (0.41, 0.93) | 0.10 | 0.10 |
| Zimbabwe | SHINE[1] (HIV+) (34), LNS vs SOC | MDAT | 67 | 68 | 0.68 (0.22, 2.04) | 0.02 | 0.05 |
| Zimbabwe | LNS+WSH vs WSH | MDAT | 103 | 83 | 0.33 (0.15, 0.76) | 0.02 | 0.05 |
| | **$I^2$ = 0.54, $Tau^2$ = 0.06** | | **9573** | **14015** | | | |
| | **Fixed** | | | | **0.81 (0.74, 0.89)** | | |
| | **Random** | | | | **0.80 (0.67, 0.96)** | | |

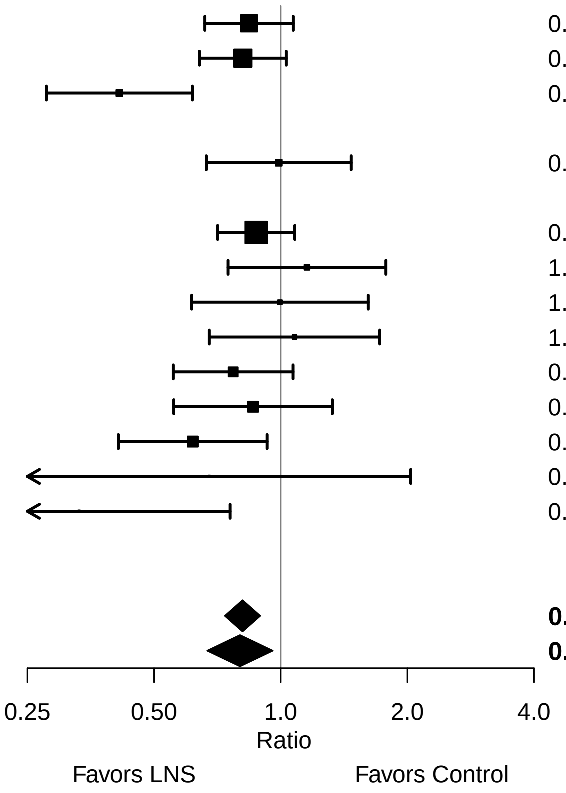

### Supplemental figure 3F: Social-emotional lowest decile prevalence difference

| Country | Trial | Tool | LNS N | Control N | PD (95% CI) | Fixed W | Random W |
|---|---|---|---|---|---|---|---|
| Bangladesh | JiVitA−4 (21) | | | | | | |
| Bangladesh | RDNS (22) | DMC | 1657 | 815 | −0.02 (−0.04, 0.01) | 0.10 | 0.10 |
| Bangladesh | WASH−B (23) | EASQ | 1066 | 3266 | −0.02 (−0.04, 0.00) | 0.18 | 0.11 |
| Burkina Faso | iLiNS−Zinc (24) | DMC | 746 | 375 | −0.10 (−0.14, −0.06) | 0.04 | 0.08 |
| Ghana | GHANA (25) | | | | | | |
| Ghana | iLiNS−DYADG (26) | PSED | 332 | 657 | 0.00 (−0.04, 0.04) | 0.04 | 0.08 |
| Haiti | HAITI (27) | | | | | | |
| Kenya | WASH−B (28) | EASQ | 1362 | 4745 | −0.01 (−0.03, 0.00) | 0.23 | 0.12 |
| Madagascar | MAHAY (29) | ASQI | 1613 | 1604 | 0.01 (−0.01, 0.04) | 0.16 | 0.11 |
| Malawi | iLiNS−DYADM (30) | PSED | 215 | 438 | 0.00 (−0.05, 0.05) | 0.03 | 0.07 |
| Malawi | iLiNS−DOSE (31) | PSED | 644 | 221 | 0.01 (−0.04, 0.05) | 0.03 | 0.07 |
| Mali | PROMIS CS (32) | DMC | 927 | 944 | −0.03 (−0.05, 0.00) | 0.09 | 0.10 |
| Zimbabwe | SHINE[1] (HIV−) (33), LNS vs SOC | MDAT | 395 | 382 | −0.01 (−0.05, 0.03) | 0.08 | 0.10 |
| Zimbabwe | LNS+WSH vs WSH | MDAT | 446 | 417 | −0.05 (−0.10, −0.01) | 0.08 | 0.10 |
| Zimbabwe | SHINE[1] (HIV+) (34), LNS vs SOC | MDAT | 67 | 68 | −0.03 (−0.12, 0.06) | 0.01 | 0.05 |
| Zimbabwe | LNS+WSH vs WSH | MDAT | 103 | 83 | −0.14 (−0.24, −0.04) | 0.01 | 0.05 |
| | **$I^2$ = 0.68, $Tau^2$ = 0.00** | | **9573** | **14015** | | | |
| | **Fixed** | | | | **−0.02 (−0.02, −0.01)** | | |
| | **Random** | | | | **−0.02 (−0.04, 0.00)** | | |

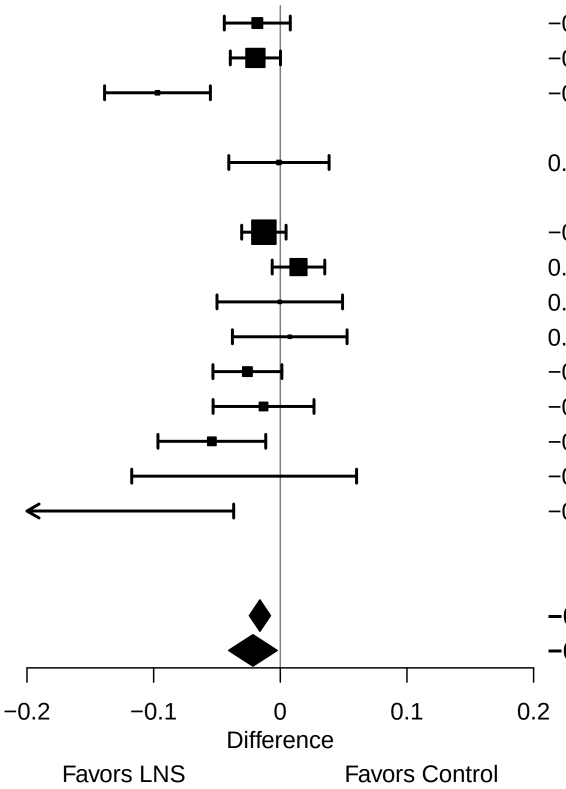

### Supplemental figure 3G: Mean difference in motor z-score

| Country | Trial | Tool | LNS N | Control N | MD (95% CI) | Fixed W | Random W |
|---|---|---|---|---|---|---|---|
| Bangladesh | JiVitA−4 (21) | BSID−III | 445 | 143 | −0.02 (−0.17, 0.14) | 0.04 | 0.07 |
| Bangladesh | RDNS (22) | DMC | 1556 | 753 | 0.10 (0.03, 0.17) | 0.19 | 0.11 |
| Bangladesh | WASH−B (23) | EASQ | 1074 | 3279 | 0.10 (0.04, 0.17) | 0.21 | 0.12 |
| Burkina Faso | iLiNS−Zinc (24) | DMC | 746 | 375 | 0.40 (0.24, 0.57) | 0.04 | 0.07 |
| Ghana | GHANA (25) | | | | | | |
| Ghana | iLiNS−DYADG (26) | KDI | 302 | 601 | 0.03 (−0.11, 0.16) | 0.05 | 0.08 |
| Haiti | HAITI (27) | | | | | | |
| Kenya | WASH−B (28) | EASQ | 1362 | 4745 | 0.01 (−0.05, 0.07) | 0.23 | 0.12 |
| Madagascar | MAHAY (29) | ASQI | 1613 | 1604 | 0.07 (−0.13, 0.27) | 0.02 | 0.06 |
| Malawi | iLiNS−DYADM (30) | KDI | 214 | 436 | 0.03 (−0.13, 0.18) | 0.04 | 0.07 |
| Malawi | iLiNS−DOSE (31) | KDI | 646 | 221 | 0.05 (−0.10, 0.20) | 0.04 | 0.08 |
| Mali | PROMIS CS (32) | DMC | 902 | 921 | 0.13 (−0.04, 0.30) | 0.03 | 0.07 |
| Zimbabwe | SHINE[1] (HIV−) (33), LNS vs SOC | MDAT | 395 | 382 | 0.00 (−0.16, 0.15) | 0.08 | 0.10 |
| Zimbabwe | LNS+WSH vs WSH | MDAT | 446 | 417 | 0.20 (0.05, 0.35) | 0.08 | 0.10 |
| Zimbabwe | SHINE[1] (HIV+) (34), LNS vs SOC | MDAT | 67 | 68 | 0.04 (−0.30, 0.39) | 0.02 | 0.05 |
| Zimbabwe | LNS+WSH vs WSH | MDAT | 103 | 83 | 0.49 (0.23, 0.74) | 0.02 | 0.05 |
| | **$I^2$ = 0.60, $Tau^2$ = 0.01** | | **9871** | **14028** | | | |
| | **Fixed** | | | | **0.08 (0.05, 0.11)** | | |
| | **Random** | | | | **0.10 (0.03, 0.16)** | | |

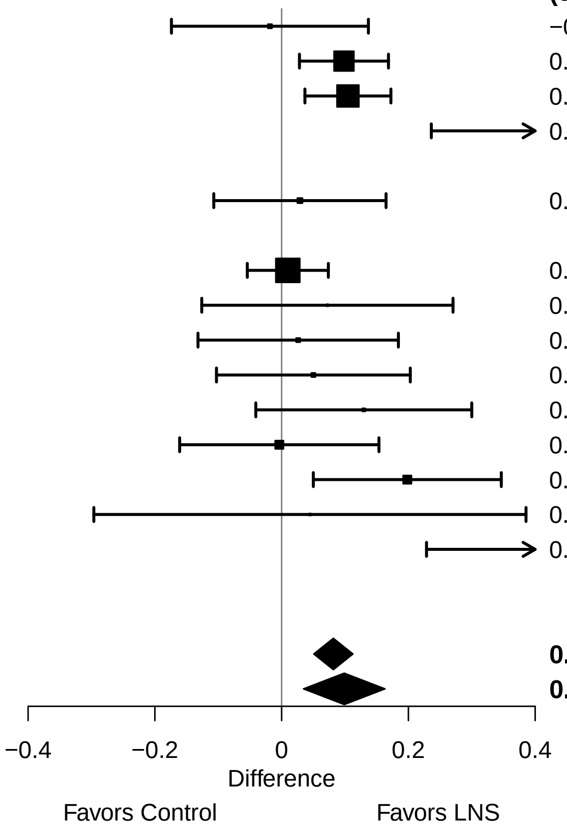

### Supplemental figure 3H: Motor lowest decile prevalence ratio

| Country | Trial | Tool | LNS N | Control N | PR (95% CI) | Fixed W | Random W |
|---|---|---|---|---|---|---|---|
| Bangladesh | JiVitA−4 (21) | BSID−III | 445 | 143 | 1.26 (0.77, 2.07) | 0.03 | 0.05 |
| Bangladesh | RDNS (22) | DMC | 1556 | 753 | 0.85 (0.67, 1.07) | 0.15 | 0.13 |
| Bangladesh | WASH−B (23) | EASQ | 1074 | 3279 | 0.82 (0.66, 1.01) | 0.19 | 0.14 |
| Burkina Faso | iLiNS−Zinc (24) | DMC | 746 | 375 | 0.49 (0.36, 0.67) | 0.09 | 0.10 |
| Ghana | GHANA (25) | | | | | | |
| Ghana | iLiNS−DYADG (26) | KDI | 302 | 601 | 1.13 (0.76, 1.70) | 0.05 | 0.07 |
| Haiti | HAITI (27) | | | | | | |
| Kenya | WASH−B (28) | EASQ | 1362 | 4745 | 0.86 (0.70, 1.05) | 0.21 | 0.14 |
| Madagascar | MAHAY (29) | ASQI | 1613 | 1604 | 0.91 (0.58, 1.44) | 0.04 | 0.06 |
| Malawi | iLiNS−DYADM (30) | KDI | 214 | 436 | 0.84 (0.51, 1.40) | 0.03 | 0.05 |
| Malawi | iLiNS−DOSE (31) | KDI | 646 | 221 | 1.08 (0.67, 1.71) | 0.04 | 0.06 |
| Mali | PROMIS CS (32) | DMC | 902 | 921 | 0.81 (0.56, 1.18) | 0.06 | 0.08 |
| Zimbabwe | SHINE[1] (HIV−) (33), LNS vs SOC | MDAT | 395 | 382 | 1.08 (0.67, 1.73) | 0.09 | 0.10 |
| Zimbabwe | LNS+WSH vs WSH | MDAT | 446 | 417 | 0.82 (0.54, 1.24) | 0.09 | 0.10 |
| Zimbabwe | SHINE[1] (HIV+) (34), LNS vs SOC | MDAT | 67 | 68 | 1.16 (0.44, 3.08) | 0.02 | 0.03 |
| Zimbabwe | LNS+WSH vs WSH | MDAT | 103 | 83 | 0.40 (0.17, 0.95) | 0.02 | 0.03 |
| | **$I^2$ = 0.40, $Tau^2$ = 0.02** | | **9871** | **14028** | | | |
| | **Fixed** | | | | **0.84 (0.76, 0.92)** | | |
| | **Random** | | | | **0.85 (0.74, 0.97)** | | |

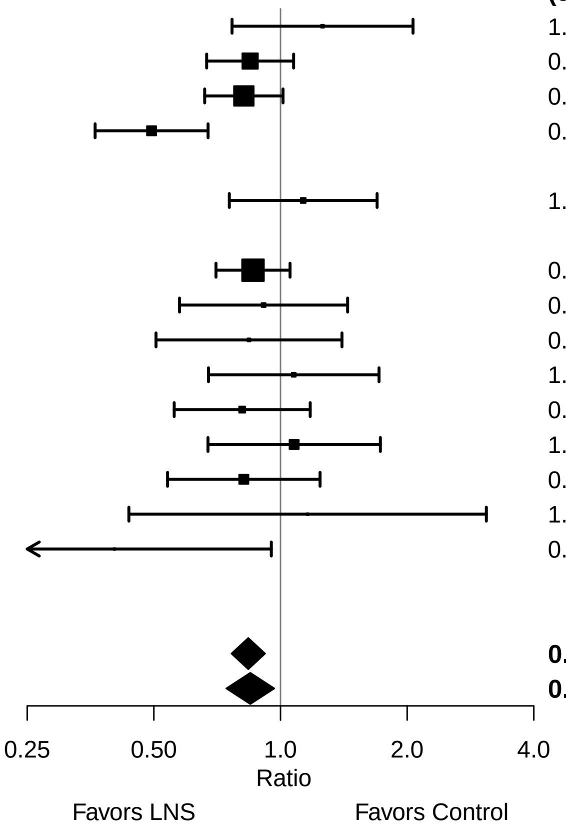

### Supplemental figure 3I: Motor lowest decile prevalence difference

| Country | Trial | Tool | LNS N | Control N | PD (95% CI) | Fixed W | Random W |
|---|---|---|---|---|---|---|---|
| Bangladesh | JiVitA−4 (21) | BSID−III | 445 | 143 | 0.02 (−0.03, 0.08) | 0.02 | 0.04 |
| Bangladesh | RDNS (22) | DMC | 1556 | 753 | −0.02 (−0.04, 0.01) | 0.09 | 0.10 |
| Bangladesh | WASH−B (23) | EASQ | 1074 | 3279 | −0.02 (−0.04, 0.00) | 0.17 | 0.14 |
| Burkina Faso | iLiNS−Zinc (24) | DMC | 746 | 375 | −0.08 (−0.12, −0.04) | 0.04 | 0.06 |
| Ghana | GHANA (25) | | | | | | |
| Ghana | iLiNS−DYADG (26) | KDI | 302 | 601 | 0.01 (−0.03, 0.06) | 0.04 | 0.05 |
| Haiti | HAITI (27) | | | | | | |
| Kenya | WASH−B (28) | EASQ | 1362 | 4745 | −0.01 (−0.03, 0.00) | 0.22 | 0.16 |
| Madagascar | MAHAY (29) | ASQI | 1613 | 1604 | −0.01 (−0.03, 0.01) | 0.16 | 0.14 |
| Malawi | iLiNS−DYADM (30) | KDI | 214 | 436 | −0.02 (−0.06, 0.03) | 0.03 | 0.04 |
| Malawi | iLiNS−DOSE (31) | KDI | 646 | 221 | 0.01 (−0.04, 0.05) | 0.03 | 0.05 |
| Mali | PROMIS CS (32) | DMC | 902 | 921 | −0.02 (−0.05, 0.01) | 0.09 | 0.10 |
| Zimbabwe | SHINE[1] (HIV−) (33), LNS vs SOC | MDAT | 395 | 382 | 0.01 (−0.03, 0.05) | 0.08 | 0.09 |
| Zimbabwe | LNS+WSH vs WSH | MDAT | 446 | 417 | −0.02 (−0.06, 0.02) | 0.08 | 0.09 |
| Zimbabwe | SHINE[1] (HIV+) (34), LNS vs SOC | MDAT | 67 | 68 | 0.02 (−0.09, 0.12) | 0.02 | 0.02 |
| Zimbabwe | LNS+WSH vs WSH | MDAT | 103 | 83 | −0.09 (−0.17, 0.00) | 0.02 | 0.02 |
| | **$I^2$ = 0.25, $Tau^2$ = 0.00** | | **9871** | **14028** | | | |
| | **Fixed** | | | | **−0.02 (−0.02, −0.01)** | | |
| | **Random** | | | | **−0.02 (−0.03, 0.00)** | | |

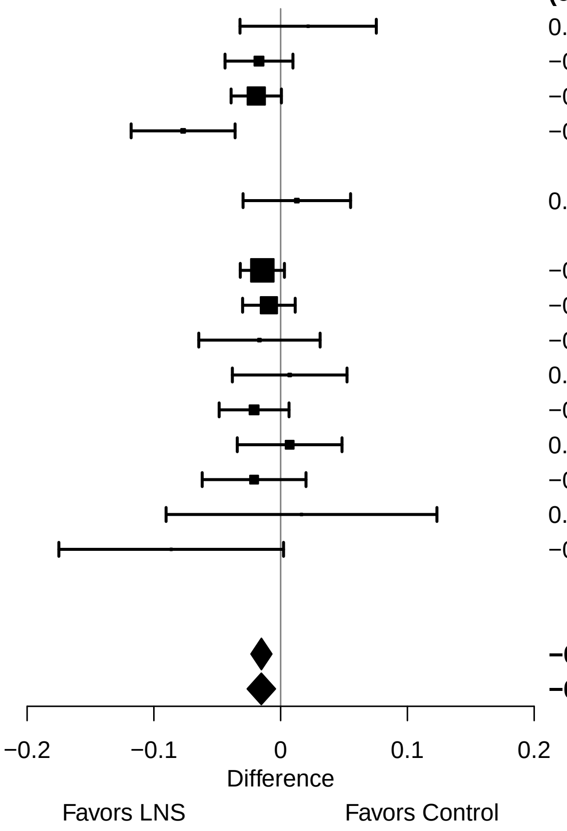

### Supplemental figure 3J: Mean difference in gross motor z-score

| Country | Trial | Tool | LNS N | Control N | MD (95% CI) | Fixed W | Random W |
|---|---|---|---|---|---|---|---|
| Bangladesh | JiVitA−4 (21) | BSID−III | 445 | 143 | −0.10 (−0.25, 0.05) | 0.04 | 0.07 |
| Bangladesh | RDNS (22) | DMC | 1570 | 758 | 0.08 (0.02, 0.15) | 0.22 | 0.13 |
| Bangladesh | WASH−B (23) | EASQ | 1074 | 3279 | 0.10 (0.04, 0.17) | 0.18 | 0.12 |
| Burkina Faso | iLiNS−Zinc (24) | DMC | | | | | |
| Ghana | GHANA (25) | | | | | | |
| Ghana | iLiNS−DYADG (26) | KDI | 302 | 601 | 0.01 (−0.13, 0.15) | 0.04 | 0.08 |
| Haiti | HAITI (27) | | | | | | |
| Kenya | WASH−B (28) | EASQ | 1362 | 4745 | 0.01 (−0.05, 0.07) | 0.20 | 0.13 |
| Madagascar | MAHAY (29) | ASQI | 1613 | 1604 | 0.08 (−0.08, 0.23) | 0.04 | 0.07 |
| Malawi | iLiNS−DYADM (30) | KDI | 214 | 436 | −0.03 (−0.19, 0.13) | 0.03 | 0.07 |
| Malawi | iLiNS−DOSE (31) | KDI | 646 | 221 | 0.00 (−0.15, 0.16) | 0.04 | 0.07 |
| Mali | PROMIS CS (32) | DMC | 940 | 957 | 0.09 (0.00, 0.17) | 0.13 | 0.11 |
| Zimbabwe | SHINE¹ (HIV−) (33), LNS vs SOC | MDAT | 395 | 382 | 0.03 (−0.13, 0.18) | 0.07 | 0.09 |
| Zimbabwe | LNS+WSH vs WSH | MDAT | 446 | 417 | 0.16 (0.00, 0.32) | 0.07 | 0.09 |
| Zimbabwe | SHINE¹ (HIV+) (34), LNS vs SOC | MDAT | 67 | 68 | 0.14 (−0.14, 0.42) | 0.02 | 0.06 |
| Zimbabwe | LNS+WSH vs WSH | MDAT | 103 | 83 | 0.48 (0.24, 0.72) | 0.02 | 0.06 |
| | **I² = 0.52, Tau² = 0.01** | | **9177** | **13694** | | | |
| | **Fixed** | | | | **0.06 (0.03, 0.09)** | | |
| | **Random** | | | | **0.06 (0.00, 0.12)** | | |

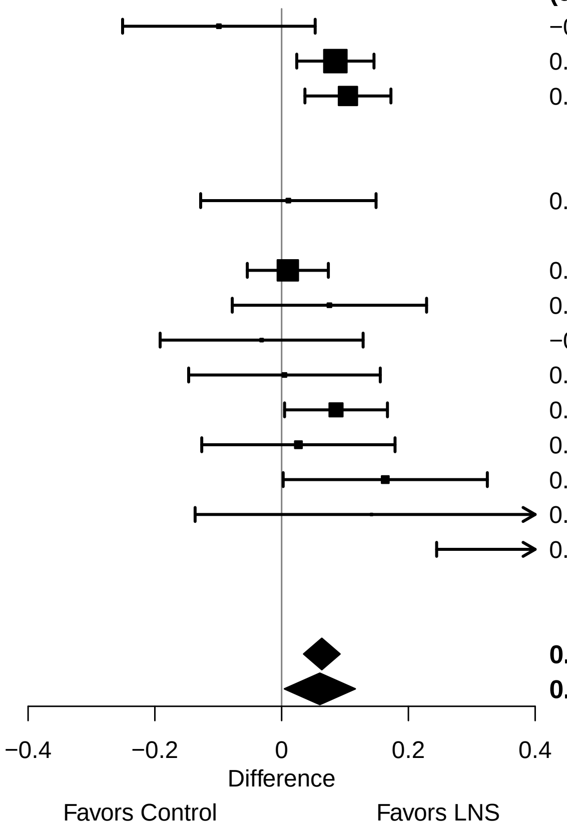

#### Supplemental figure 3K: Mean difference in fine motor z-score

| Country | Trial | Tool | LNS N | Control N | MD (95% CI) | Fixed W | Random W |
|---|---|---|---|---|---|---|---|
| Bangladesh | JiVitA−4 (21) | BSID−III | 445 | 143 | 0.14 (−0.05, 0.34) | 0.06 | 0.06 |
| Bangladesh | RDNS (22) | DMC | 1641 | 804 | 0.10 (−0.01, 0.20) | 0.21 | 0.21 |
| Bangladesh | WASH−B (23) | EASQ | | | | | |
| Burkina Faso | iLiNS−Zinc (24) | DMC | | | | | |
| Ghana | GHANA (25) | | | | | | |
| Ghana | iLiNS−DYADG (26) | KDI | 302 | 601 | 0.04 (−0.09, 0.17) | 0.14 | 0.14 |
| Haiti | HAITI (27) | | | | | | |
| Kenya | WASH−B (28) | EASQ | | | | | |
| Madagascar | MAHAY (29) | ASQI | 1613 | 1604 | 0.05 (−0.17, 0.26) | 0.05 | 0.05 |
| Malawi | iLiNS−DYADM (30) | KDI | 214 | 436 | 0.08 (−0.08, 0.24) | 0.10 | 0.10 |
| Malawi | iLiNS−DOSE (31) | KDI | 646 | 221 | 0.09 (−0.06, 0.24) | 0.10 | 0.10 |
| Mali | PROMIS CS (32) | DMC | 905 | 924 | 0.11 (−0.11, 0.32) | 0.05 | 0.05 |
| Zimbabwe | SHINE[1] (HIV−) (33), LNS vs SOC | MDAT | 395 | 382 | −0.03 (−0.18, 0.11) | 0.23 | 0.23 |
| Zimbabwe | LNS+WSH vs WSH | MDAT | 446 | 417 | 0.17 (0.03, 0.31) | 0.23 | 0.23 |
| Zimbabwe | SHINE[1] (HIV+) (34), LNS vs SOC | MDAT | 67 | 68 | −0.04 (−0.40, 0.31) | 0.05 | 0.05 |
| Zimbabwe | LNS+WSH vs WSH | MDAT | 103 | 83 | 0.31 (0.03, 0.59) | 0.05 | 0.05 |
| | **$I^2$ = 0.00, $Tau^2$ = 0.00** | | **6777** | **5683** | | | |
| | **Fixed** | | | | **0.09 (0.04, 0.13)** | | |
| | **Random** | | | | **0.09 (0.04, 0.13)** | | |

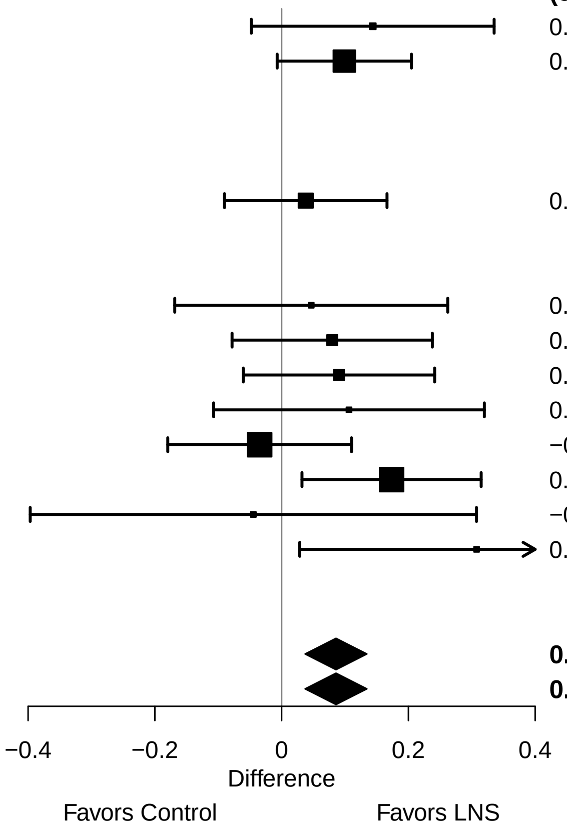

#### Supplemental figure 3L: Mean difference in executive function z-score

| Country | Trial | Tool | LNS N | Control N | MD (95% CI) | Fixed W | Random W |
|---|---|---|---|---|---|---|---|
| Bangladesh | JiVitA−4 (21) | | | | | | |
| Bangladesh | RDNS (22) | A not B task | 533 | 266 | 0.10 (−0.02, 0.22) | 0.14 | 0.15 |
| Bangladesh | WASH−B (23) | A not B task | 1097 | 3327 | 0.00 (−0.07, 0.07) | 0.43 | 0.40 |
| Burkina Faso | iLiNS−Zinc (24) | | | | | | |
| Ghana | GHANA (25) | | | | | | |
| Ghana | iLiNS−DYADG (26) | A not B task | 290 | 576 | 0.02 (−0.12, 0.17) | 0.10 | 0.11 |
| Haiti | HAITI (27) | | | | | | |
| Kenya | WASH−B (28) | | | | | | |
| Madagascar | MAHAY (29) | | | | | | |
| Malawi | iLiNS−DYADM (30) | A not B task | 182 | 369 | −0.09 (−0.27, 0.09) | 0.07 | 0.07 |
| Malawi | iLiNS−DOSE (31) | A not B task | 475 | 164 | −0.11 (−0.29, 0.07) | 0.07 | 0.07 |
| Mali | PROMIS CS (32) | | | | | | |
| Zimbabwe | SHINE[1] (HIV−) (33), LNS vs SOC | A not B task | 370 | 357 | −0.05 (−0.21, 0.10) | 0.15 | 0.16 |
| Zimbabwe | LNS+WSH vs WSH | A not B task | 417 | 382 | 0.12 (−0.05, 0.29) | 0.15 | 0.16 |
| Zimbabwe | SHINE[1] (HIV+) (34), LNS vs SOC | A not B task | 63 | 57 | −0.16 (−0.49, 0.16) | 0.04 | 0.05 |
| Zimbabwe | LNS+WSH vs WSH | A not B task | 94 | 76 | −0.05 (−0.35, 0.25) | 0.04 | 0.05 |
| | **$I^2$ = 0.04, $Tau^2$ = 0.00** | | **3521** | **5574** | | | |
| | **Fixed** | | | | **0.00 (−0.04, 0.05)** | | |
| | **Random** | | | | **0.00 (−0.04, 0.05)** | | |

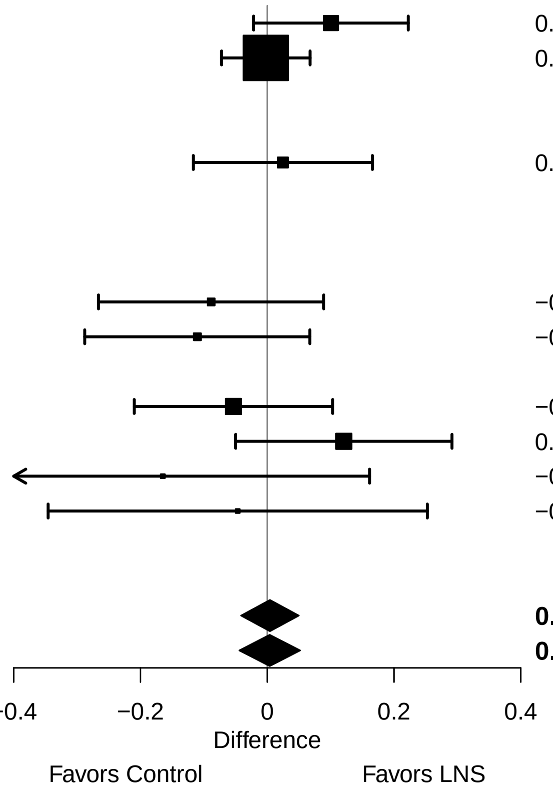

#### Supplemental figure 3M: Executive function lowest decile prevalence ratio

| Country | Trial | Tool | LNS N | Control N | PR (95% CI) | Fixed W | Random W |
|---|---|---|---|---|---|---|---|
| Bangladesh | JiVitA−4 (21) | | | | | | |
| Bangladesh | RDNS (22) | A not B task | 533 | 266 | 0.88 (0.60, 1.29) | 0.13 | 0.15 |
| Bangladesh | WASH−B (23) | A not B task | 1097 | 3327 | 0.95 (0.77, 1.18) | 0.41 | 0.31 |
| Burkina Faso | iLiNS−Zinc (24) | | | | | | |
| Ghana | GHANA (25) | | | | | | |
| Ghana | iLiNS−DYADG (26) | A not B task | 290 | 576 | 0.78 (0.50, 1.20) | 0.10 | 0.12 |
| Haiti | HAITI (27) | | | | | | |
| Kenya | WASH−B (28) | | | | | | |
| Madagascar | MAHAY (29) | | | | | | |
| Malawi | iLiNS−DYADM (30) | A not B task | 182 | 369 | 0.81 (0.47, 1.41) | 0.06 | 0.08 |
| Malawi | iLiNS−DOSE (31) | A not B task | 475 | 164 | 1.50 (0.82, 2.73) | 0.05 | 0.07 |
| Mali | PROMIS CS (32) | | | | | | |
| Zimbabwe | SHINE[1] (HIV−) (33), LNS vs SOC | A not B task | 370 | 357 | 0.82 (0.53, 1.26) | 0.20 | 0.20 |
| Zimbabwe | LNS+WSH vs WSH | A not B task | 417 | 382 | 0.78 (0.50, 1.21) | 0.20 | 0.20 |
| Zimbabwe | SHINE[1] (HIV+) (34), LNS vs SOC | A not B task | 63 | 57 | 1.09 (0.37, 3.22) | 0.05 | 0.06 |
| Zimbabwe | LNS+WSH vs WSH | A not B task | 94 | 76 | 2.10 (0.92, 4.82) | 0.05 | 0.06 |
| | **$I^2$ = 0.16, $Tau^2$ = 0.01** | | **3521** | **5574** | | | |
| | **Fixed** | | | | **0.93 (0.81, 1.06)** | | |
| | **Random** | | | | **0.93 (0.78, 1.11)** | | |

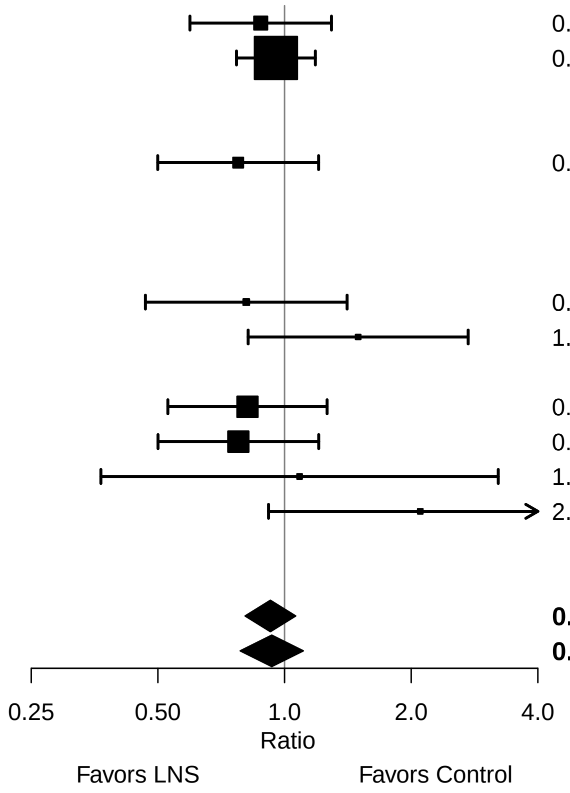

### Supplemental figure 3N: Executive function lowest decile prevalence difference

| Country | Trial | Tool | LNS N | Control N | PD (95% CI) | Fixed W | Random W |
|---|---|---|---|---|---|---|---|
| Bangladesh | JiVitA−4 (21) | | | | | | |
| Bangladesh | RDNS (22) | A not B task | 533 | 266 | −0.01 (−0.06, 0.03) | 0.09 | 0.12 |
| Bangladesh | WASH−B (23) | A not B task | 1097 | 3327 | 0.00 (−0.02, 0.02) | 0.44 | 0.30 |
| Burkina Faso | iLiNS−Zinc (24) | | | | | | |
| Ghana | GHANA (25) | | | | | | |
| Ghana | iLiNS−DYADG (26) | A not B task | 290 | 576 | −0.02 (−0.07, 0.02) | 0.10 | 0.13 |
| Haiti | HAITI (27) | | | | | | |
| Kenya | WASH−B (28) | | | | | | |
| Madagascar | MAHAY (29) | | | | | | |
| Malawi | iLiNS−DYADM (30) | A not B task | 182 | 369 | −0.02 (−0.07, 0.03) | 0.07 | 0.09 |
| Malawi | iLiNS−DOSE (31) | A not B task | 475 | 164 | 0.04 (−0.01, 0.09) | 0.08 | 0.10 |
| Mali | PROMIS CS (32) | | | | | | |
| Zimbabwe | SHINE¹ (HIV−) (33), LNS vs SOC | A not B task | 370 | 357 | −0.02 (−0.06, 0.02) | 0.19 | 0.20 |
| Zimbabwe | LNS+WSH vs WSH | A not B task | 417 | 382 | −0.03 (−0.07, 0.02) | 0.19 | 0.20 |
| Zimbabwe | SHINE¹ (HIV+) (34), LNS vs SOC | A not B task | 63 | 57 | 0.01 (−0.10, 0.11) | 0.04 | 0.06 |
| Zimbabwe | LNS+WSH vs WSH | A not B task | 94 | 76 | 0.07 (−0.02, 0.16) | 0.04 | 0.06 |
| | **$I^2$ = 0.21, $Tau^2$ = 0.00** | | **3521** | **5574** | | | |
| | **Fixed** | | | | **−0.01 (−0.02, 0.01)** | | |
| | **Random** | | | | **−0.01 (−0.02, 0.01)** | | |

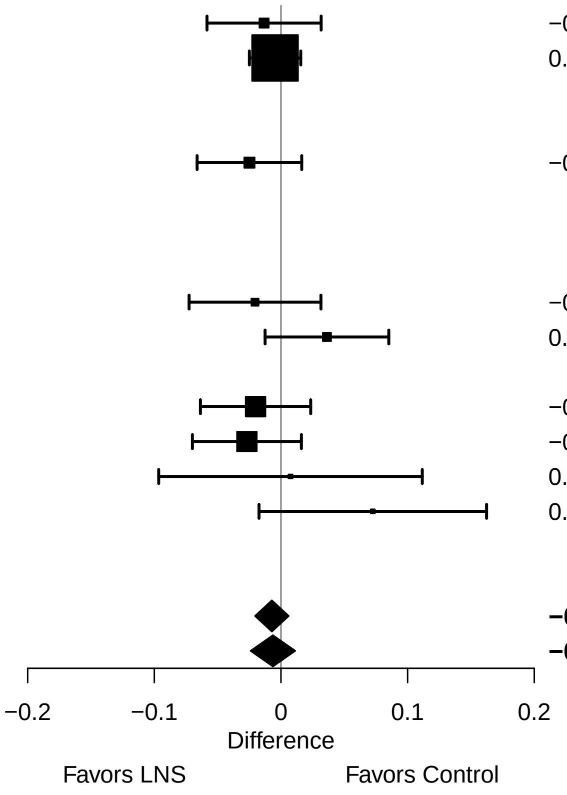

### Supplemental figure 3O: 12-mo walking without support prevalence ratio

| Country | Trial | LNS N | Control N | PR (95% CI) | Fixed W | Random W |
|---|---|---|---|---|---|---|
| Bangladesh | RDNS (22) | 1628 | 790 | 1.28 (1.07, 1.53) | 0.11 | 0.13 |
| Bangladesh | WASH−B (23) | 497 | 1474 | 1.20 (1.05, 1.38) | 0.18 | 0.15 |
| Burkina Faso | iLiNS−Zinc (24) | | | | | |
| Ghana | GHANA (25) | 90 | 87 | 1.67 (1.08, 2.58) | 0.02 | 0.05 |
| Ghana | iLiNS−DYADG (26) | 327 | 663 | 1.15 (1.01, 1.31) | 0.21 | 0.16 |
| Haiti | HAITI (27) | 83 | 79 | 1.25 (0.86, 1.81) | 0.02 | 0.06 |
| Kenya | WASH−B (28) | 447 | 1604 | 0.97 (0.85, 1.10) | 0.21 | 0.16 |
| Madagascar | MAHAY (29) | | | | | |
| Malawi | iLiNS−DYADM (30) | 208 | 426 | 1.20 (1.03, 1.40) | 0.15 | 0.15 |
| Malawi | iLiNS−DOSE (31) | 602 | 200 | 1.04 (0.84, 1.30) | 0.07 | 0.11 |
| Mali | PROMIS CS (32) | 118 | 151 | 0.70 (0.44, 1.11) | 0.02 | 0.04 |
| Zimbabwe | SHINE (HIV−) (33) | | | | | |
| Zimbabwe | SHINE (HIV+) (34) | | | | | |
| | **$I^2$ = 0.55, $Tau^2$ = 0.01** | **4000** | **5474** | | | |
| | **Fixed** | | | **1.09 (1.05, 1.14)** | | |
| | **Random** | | | **1.12 (1.02, 1.24)** | | |

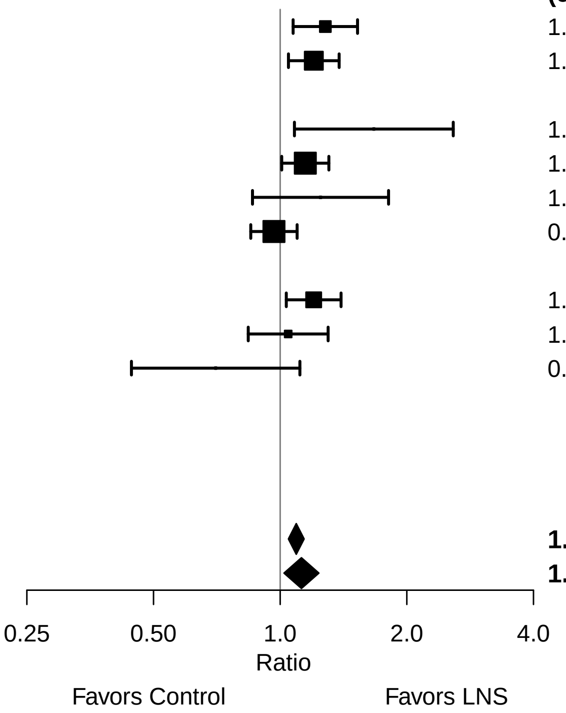

### Supplemental figure 3P: 12-mo walking without support prevalence difference

| Country | Trial | LNS N | Control N | PD (95% CI) | Fixed W | Random W |
|---|---|---|---|---|---|---|
| Bangladesh | RDNS (22) | 1628 | 790 | 0.07 (0.03, 0.11) | 0.30 | 0.16 |
| Bangladesh | WASH−B (23) | 497 | 1474 | 0.05 (0.01, 0.10) | 0.21 | 0.15 |
| Burkina Faso | iLiNS−Zinc (24) | | | | | |
| Ghana | GHANA (25) | 90 | 87 | 0.17 (0.03, 0.31) | 0.02 | 0.06 |
| Ghana | iLiNS−DYADG (26) | 327 | 663 | 0.07 (0.00, 0.14) | 0.10 | 0.13 |
| Haiti | HAITI (27) | 83 | 79 | 0.09 (−0.06, 0.24) | 0.02 | 0.05 |
| Kenya | WASH−B (28) | 447 | 1604 | −0.01 (−0.06, 0.04) | 0.17 | 0.15 |
| Madagascar | MAHAY (29) | | | | | |
| Malawi | iLiNS−DYADM (30) | 208 | 426 | 0.10 (0.02, 0.18) | 0.07 | 0.11 |
| Malawi | iLiNS−DOSE (31) | 602 | 200 | 0.02 (−0.06, 0.09) | 0.08 | 0.11 |
| Mali | PROMIS CS (32) | 118 | 151 | −0.10 (−0.21, 0.01) | 0.04 | 0.08 |
| Zimbabwe | SHINE (HIV−) (33) | | | | | |
| Zimbabwe | SHINE (HIV+) (34) | | | | | |
| | **I² = 0.57, Tau² = 0.00** | **4000** | **5474** | | | |
| | **Fixed** | | | **0.04 (0.02, 0.06)** | | |
| | **Random** | | | **0.04 (0.00, 0.08)** | | |

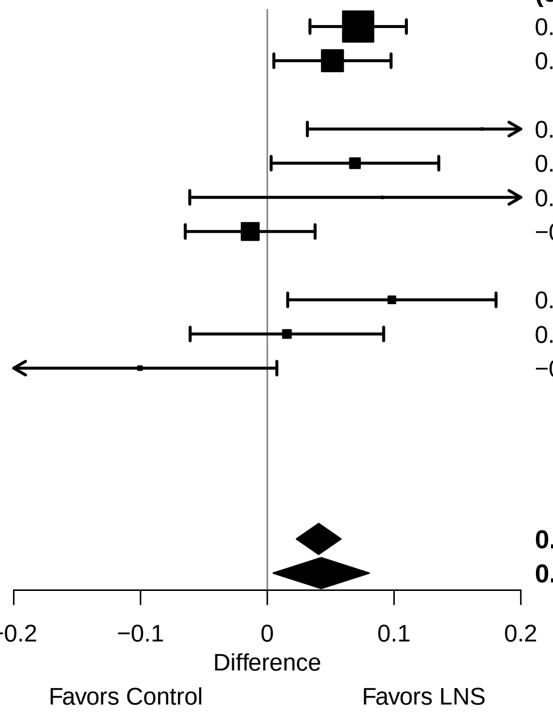

#### Supplemental figure 3Q: 12-mo walking with support prevalence ratio

| Country | Trial | LNS N | Control N | PR (95% CI) | Fixed W | Random W |
|---|---|---|---|---|---|---|
| Bangladesh | RDNS (22) | 1546 | 746 | 1.06 (1.01, 1.11) | 0.07 | 0.14 |
| Bangladesh | WASH−B (23) | 496 | 1470 | 0.99 (0.94, 1.05) | 0.05 | 0.12 |
| Burkina Faso | iLiNS−Zinc (24) | | | | | |
| Ghana | GHANA (25) | 90 | 87 | 1.16 (0.97, 1.39) | 0.00 | 0.01 |
| Ghana | iLiNS−DYADG (26) | 327 | 662 | 1.00 (0.99, 1.01) | 0.64 | 0.34 |
| Haiti | HAITI (27) | | | | | |
| Kenya | WASH−B (28) | 447 | 1604 | 1.03 (1.00, 1.06) | 0.16 | 0.23 |
| Madagascar | MAHAY (29) | | | | | |
| Malawi | iLiNS−DYADM (30) | 208 | 426 | 1.02 (0.97, 1.07) | 0.05 | 0.12 |
| Malawi | iLiNS−DOSE (31) | 602 | 200 | 1.05 (0.95, 1.16) | 0.01 | 0.04 |
| Mali | PROMIS CS (32) | 118 | 151 | 0.93 (0.76, 1.13) | 0.00 | 0.01 |
| Zimbabwe | SHINE (HIV−) (33) | | | | | |
| Zimbabwe | SHINE (HIV+) (34) | | | | | |
| | **$I^2$ = 0.41, $Tau^2$ = 0.00** | **3834** | **5346** | | | |
| | **Fixed** | | | **1.00 (1.00, 1.01)** | | |
| | **Random** | | | **1.01 (1.00, 1.03)** | | |

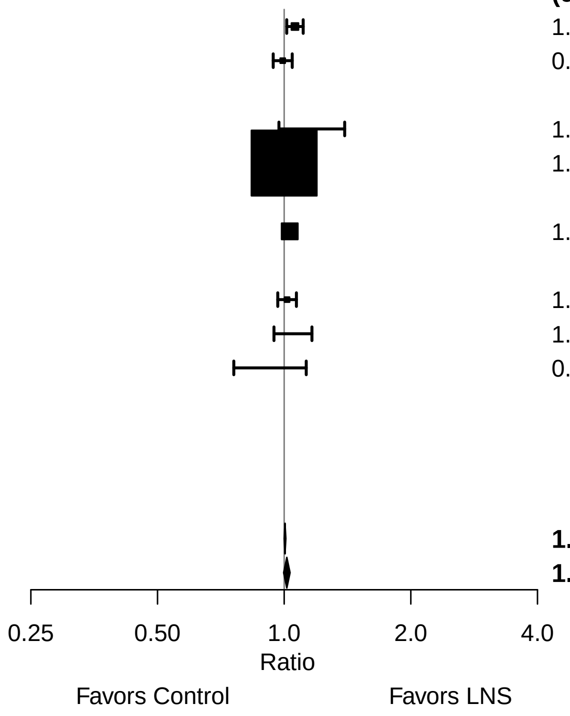

#### Supplemental figure 3R: 12-mo walking with support prevalence difference

| Country | Trial | LNS N | Control N | PD (95% CI) | Fixed W | Random W |
|---|---|---|---|---|---|---|
| Bangladesh | RDNS (22) | 1546 | 746 | 0.05 (0.01, 0.08) | 0.10 | 0.16 |
| Bangladesh | WASH−B (23) | 496 | 1470 | −0.01 (−0.05, 0.03) | 0.07 | 0.13 |
| Burkina Faso | iLiNS−Zinc (24) | | | | | |
| Ghana | GHANA (25) | 90 | 87 | 0.11 (−0.02, 0.24) | 0.01 | 0.02 |
| Ghana | iLiNS−DYADG (26) | 327 | 662 | 0.00 (−0.01, 0.01) | 0.57 | 0.29 |
| Haiti | HAITI (27) | | | | | |
| Kenya | WASH−B (28) | 447 | 1604 | 0.03 (0.00, 0.05) | 0.17 | 0.21 |
| Madagascar | MAHAY (29) | | | | | |
| Malawi | iLiNS−DYADM (30) | 208 | 426 | 0.01 (−0.03, 0.06) | 0.06 | 0.11 |
| Malawi | iLiNS−DOSE (31) | 602 | 200 | 0.03 (−0.04, 0.11) | 0.02 | 0.06 |
| Mali | PROMIS CS (32) | 118 | 151 | −0.06 (−0.16, 0.05) | 0.01 | 0.03 |
| Zimbabwe | SHINE (HIV−) (33) | | | | | |
| Zimbabwe | SHINE (HIV+) (34) | | | | | |
| | **$I^2$ = 0.42, $Tau^2$ = 0.00** | **3834** | **5346** | | | |
| | **Fixed** | | | **0.00 (0.00, 0.01)** | | |
| | **Random** | | | **0.01 (0.00, 0.03)** | | |

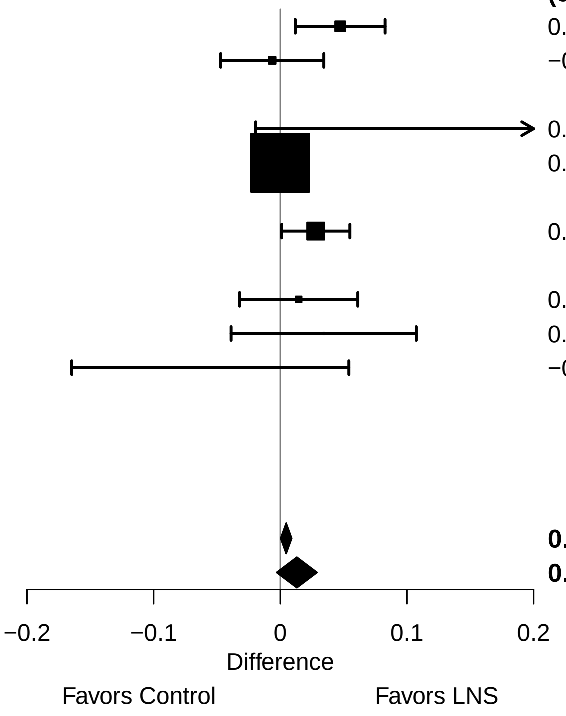

#### Supplemental figure 3S: 12-mo standing without support prevalence ratio

| Country | Trial | LNS N | Control N | PR (95% CI) | Fixed W | Random W |
|---|---|---|---|---|---|---|
| Bangladesh | RDNS (22) | 1550 | 767 | 1.22 (1.13, 1.33) | 0.14 | 0.14 |
| Bangladesh | WASH−B (23) | 499 | 1474 | 1.07 (0.96, 1.18) | 0.08 | 0.11 |
| Burkina Faso | iLiNS−Zinc (24) | | | | | |
| Ghana | GHANA (25) | 90 | 87 | 1.17 (0.89, 1.54) | 0.01 | 0.03 |
| Ghana | iLiNS−DYADG (26) | 323 | 654 | 0.99 (0.92, 1.07) | 0.16 | 0.15 |
| Haiti | HAITI (27) | 132 | 108 | 1.01 (0.83, 1.23) | 0.02 | 0.05 |
| Kenya | WASH−B (28) | 447 | 1604 | 1.00 (0.95, 1.06) | 0.26 | 0.18 |
| Madagascar | MAHAY (29) | | | | | |
| Malawi | iLiNS−DYADM (30) | 208 | 426 | 1.06 (0.99, 1.13) | 0.20 | 0.16 |
| Malawi | iLiNS−DOSE (31) | 602 | 200 | 1.04 (0.95, 1.14) | 0.11 | 0.13 |
| Mali | PROMIS CS (32) | 118 | 151 | 0.95 (0.78, 1.16) | 0.02 | 0.05 |
| Zimbabwe | SHINE (HIV−) (33) | | | | | |
| Zimbabwe | SHINE (HIV+) (34) | | | | | |
| | **$I^2$ = 0.65, $Tau^2$ = 0.00** | **3969** | **5471** | | | |
| | **Fixed** | | | **1.03 (1.00, 1.05)** | | |
| | **Random** | | | **1.04 (1.00, 1.09)** | | |

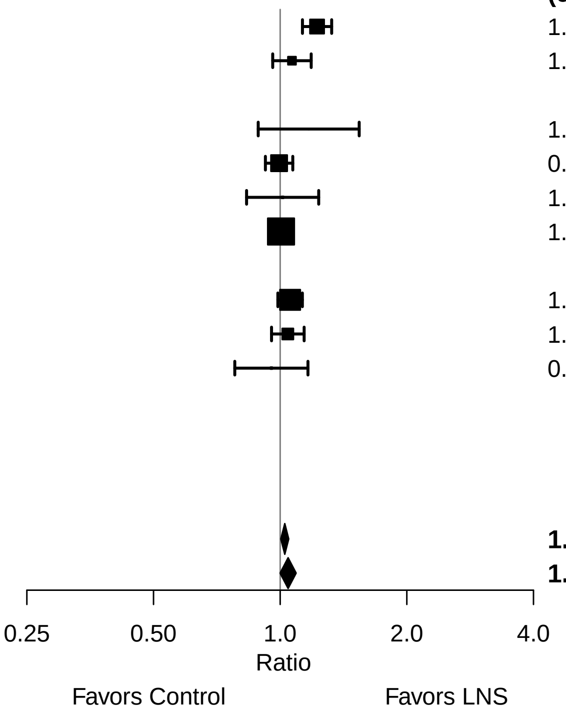

#### Supplemental figure 3T: 12-mo standing without support prevalence difference

| Country | Trial | LNS N | Control N | PD (95% CI) | Fixed W | Random W |
|---|---|---|---|---|---|---|
| Bangladesh | RDNS (22) | 1550 | 767 | 0.10 (0.06, 0.15) | 0.22 | 0.18 |
| Bangladesh | WASH−B (23) | 499 | 1474 | 0.03 (−0.02, 0.08) | 0.16 | 0.15 |
| Burkina Faso | iLiNS−Zinc (24) | | | | | |
| Ghana | GHANA (25) | 90 | 87 | 0.08 (−0.06, 0.23) | 0.02 | 0.03 |
| Ghana | iLiNS−DYADG (26) | 323 | 654 | 0.00 (−0.06, 0.05) | 0.13 | 0.14 |
| Haiti | HAITI (27) | 132 | 108 | 0.01 (−0.12, 0.13) | 0.03 | 0.04 |
| Kenya | WASH−B (28) | 447 | 1604 | 0.00 (−0.04, 0.05) | 0.19 | 0.17 |
| Madagascar | MAHAY (29) | | | | | |
| Malawi | iLiNS−DYADM (30) | 208 | 426 | 0.05 (−0.01, 0.10) | 0.13 | 0.13 |
| Malawi | iLiNS−DOSE (31) | 602 | 200 | 0.03 (−0.04, 0.10) | 0.09 | 0.11 |
| Mali | PROMIS CS (32) | 118 | 151 | −0.03 (−0.15, 0.09) | 0.03 | 0.05 |
| Zimbabwe | SHINE (HIV−) (33) | | | | | |
| Zimbabwe | SHINE (HIV+) (34) | | | | | |
| | **I² = 0.56, Tau² = 0.00** | **3969** | **5471** | | | |
| | **Fixed** | | | **0.02 (0.01, 0.04)** | | |
| | **Random** | | | **0.03 (0.00, 0.05)** | | |

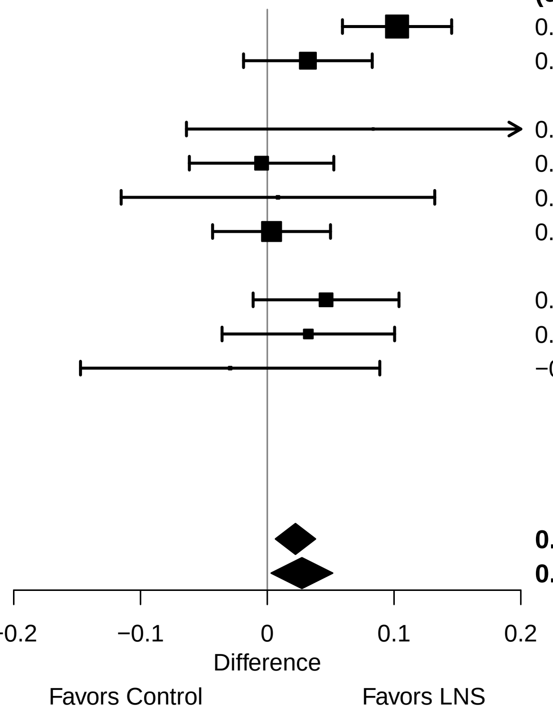

### Supplemental figure 3U: 12-mo standing with support prevalence ratio

| Country | Trial | LNS N | Control N | PR (95% CI) | Fixed W | Random W |
|---|---|---|---|---|---|---|
| Bangladesh | RDNS (22) | 1624 | 783 | 1.00 (0.99, 1.02) | 0.05 | 0.05 |
| Bangladesh | WASH−B (23) | 497 | 1472 | 1.00 (0.98, 1.02) | 0.02 | 0.02 |
| Burkina Faso | iLiNS−Zinc (24) | | | | | |
| Ghana | GHANA (25) | 90 | 87 | 1.01 (0.93, 1.10) | 0.00 | 0.00 |
| Ghana | iLiNS−DYADG (26) | 329 | 670 | 1.00 (1.00, 1.00) | 0.91 | 0.91 |
| Haiti | HAITI (27) | | | | | |
| Kenya | WASH−B (28) | 447 | 1604 | 1.02 (0.99, 1.05) | 0.01 | 0.01 |
| Madagascar | MAHAY (29) | | | | | |
| Malawi | iLiNS−DYADM (30) | 208 | 426 | 1.01 (0.98, 1.04) | 0.01 | 0.01 |
| Malawi | iLiNS−DOSE (31) | 602 | 200 | 1.02 (0.99, 1.05) | 0.01 | 0.01 |
| Mali | PROMIS CS (32) | 118 | 151 | 1.03 (0.94, 1.12) | 0.00 | 0.00 |
| Zimbabwe | SHINE (HIV−) (33) | | | | | |
| Zimbabwe | SHINE (HIV+) (34) | | | | | |
| | **I² = 0.00, Tau² = 0.00** | **3915** | **5393** | | | |
| | **Fixed** | | | **1.00 (1.00, 1.00)** | | |
| | **Random** | | | **1.00 (1.00, 1.00)** | | |

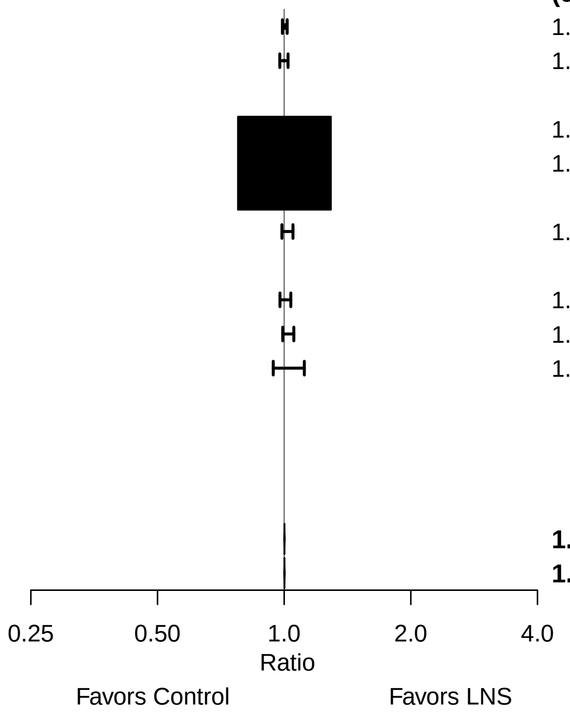

#### Supplemental figure 3V: 12-mo standing with support prevalence difference

| Country | Trial | LNS N | Control N | PD (95% CI) | Fixed W | Random W |
|---|---|---|---|---|---|---|
| Bangladesh | RDNS (22) | 1624 | 783 | 0.00 (−0.01, 0.02) | 0.05 | 0.05 |
| Bangladesh | WASH−B (23) | 497 | 1472 | 0.00 (−0.02, 0.02) | 0.01 | 0.01 |
| Burkina Faso | iLiNS−Zinc (24) | | | | | |
| Ghana | GHANA (25) | 90 | 87 | 0.01 (−0.06, 0.09) | 0.00 | 0.00 |
| Ghana | iLiNS−DYADG (26) | 329 | 670 | 0.00 (0.00, 0.00) | 0.90 | 0.90 |
| Haiti | HAITI (27) | | | | | |
| Kenya | WASH−B (28) | 447 | 1604 | 0.02 (−0.01, 0.04) | 0.01 | 0.01 |
| Madagascar | MAHAY (29) | | | | | |
| Malawi | iLiNS−DYADM (30) | 208 | 426 | 0.01 (−0.02, 0.04) | 0.01 | 0.01 |
| Malawi | iLiNS−DOSE (31) | 602 | 200 | 0.02 (−0.01, 0.05) | 0.01 | 0.01 |
| Mali | PROMIS CS (32) | 118 | 151 | 0.02 (−0.04, 0.09) | 0.00 | 0.00 |
| Zimbabwe | SHINE (HIV−) (33) | | | | | |
| Zimbabwe | SHINE (HIV+) (34) | | | | | |
| | **I² = 0.00, Tau² = 0.00** | **3915** | **5393** | | | |
| | **Fixed** | | | **0.00 (0.00, 0.00)** | | |
| | **Random** | | | **0.00 (0.00, 0.00)** | | |

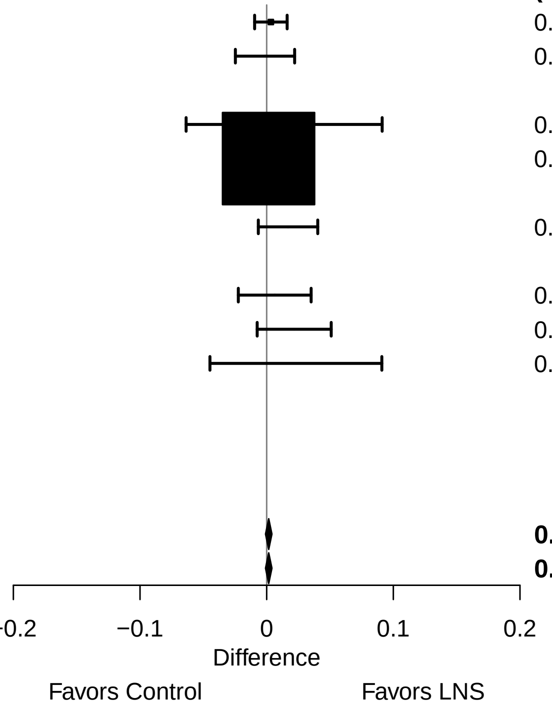

#### Supplemental figure 3W: 12-mo crawling prevalence ratio

| Country | Trial | LNS N | Control N | PR (95% CI) | Fixed W | Random W |
|---|---|---|---|---|---|---|
| Bangladesh | RDNS (22) | 1494 | 742 | 1.08 (1.02, 1.14) | 0.03 | 0.05 |
| Bangladesh | WASH−B (23) | 500 | 1480 | 1.03 (0.97, 1.08) | 0.03 | 0.05 |
| Burkina Faso | iLiNS−Zinc (24) | | | | | |
| Ghana | GHANA (25) | | | | | |
| Ghana | iLiNS−DYADG (26) | 328 | 665 | 1.01 (0.99, 1.02) | 0.43 | 0.30 |
| Haiti | HAITI (27) | 83 | 79 | 1.03 (0.94, 1.14) | 0.01 | 0.02 |
| Kenya | WASH−B (28) | 447 | 1604 | 1.01 (0.99, 1.02) | 0.28 | 0.26 |
| Madagascar | MAHAY (29) | | | | | |
| Malawi | iLiNS−DYADM (30) | 208 | 426 | 0.99 (0.97, 1.01) | 0.15 | 0.18 |
| Malawi | iLiNS−DOSE (31) | 602 | 200 | 0.99 (0.95, 1.03) | 0.06 | 0.09 |
| Mali | PROMIS CS (32) | 118 | 151 | 1.01 (0.95, 1.07) | 0.02 | 0.05 |
| Zimbabwe | SHINE (HIV−) (33) | | | | | |
| Zimbabwe | SHINE (HIV+) (34) | | | | | |
| | **$I^2$ = 0.26, $Tau^2$ = 0.00** | **3780** | **5347** | | | |
| | **Fixed** | | | **1.00 (1.00, 1.01)** | | |
| | **Random** | | | **1.01 (0.99, 1.02)** | | |

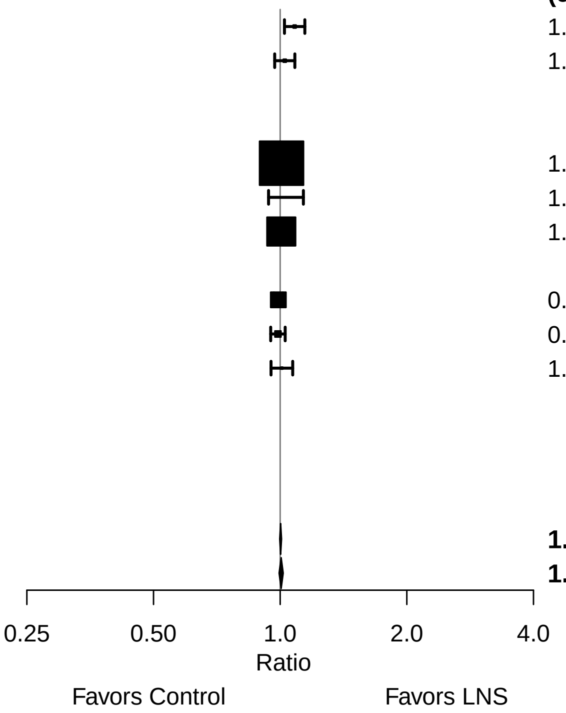

#### Supplemental figure 3X: 12-mo crawling prevalence difference

| Country | Trial | LNS N | Control N | PD (95% CI) | Fixed W | Random W |
|---|---|---|---|---|---|---|
| Bangladesh | RDNS (22) | 1494 | 742 | 0.06 (0.02, 0.10) | 0.05 | 0.08 |
| Bangladesh | WASH−B (23) | 500 | 1480 | 0.02 (−0.02, 0.06) | 0.04 | 0.08 |
| Burkina Faso | iLiNS−Zinc (24) | | | | | |
| Ghana | GHANA (25) | | | | | |
| Ghana | iLiNS−DYADG (26) | 328 | 665 | 0.01 (−0.01, 0.02) | 0.39 | 0.27 |
| Haiti | HAITI (27) | 83 | 79 | 0.03 (−0.06, 0.12) | 0.01 | 0.02 |
| Kenya | WASH−B (28) | 447 | 1604 | 0.01 (−0.01, 0.02) | 0.28 | 0.24 |
| Madagascar | MAHAY (29) | | | | | |
| Malawi | iLiNS−DYADM (30) | 208 | 426 | −0.01 (−0.03, 0.01) | 0.14 | 0.17 |
| Malawi | iLiNS−DOSE (31) | 602 | 200 | −0.01 (−0.05, 0.03) | 0.06 | 0.09 |
| Mali | PROMIS CS (32) | 118 | 151 | 0.01 (−0.05, 0.06) | 0.03 | 0.05 |
| Zimbabwe | SHINE (HIV−) (33) | | | | | |
| Zimbabwe | SHINE (HIV+) (34) | | | | | |
| | **I² = 0.22, Tau² = 0.00** | **3780** | **5347** | | | |
| | **Fixed** | | | **0.00 (0.00, 0.01)** | | |
| | **Random** | | | **0.01 (−0.01, 0.02)** | | |

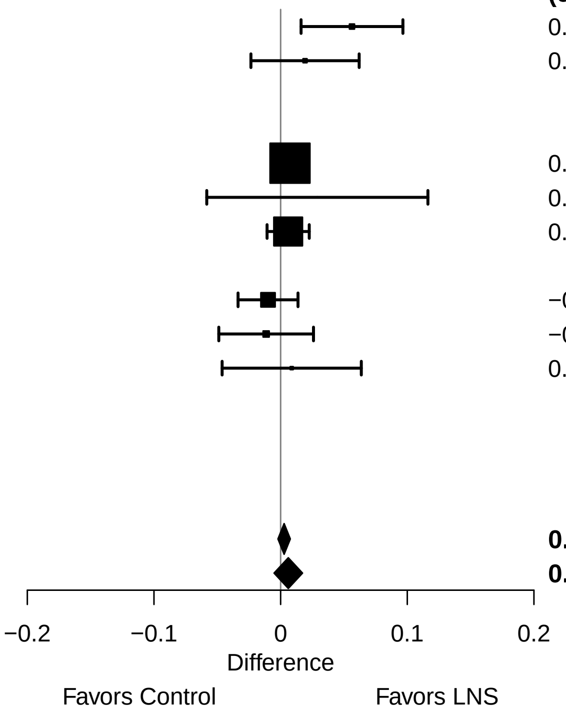

#### Supplemental figure 3Y: 18-mo walking without support prevalence ratio

| Country | Trial | LNS N | Control N | PR (95% CI) | Fixed W | Random W |
|---|---|---|---|---|---|---|
| Bangladesh | RDNS (22) | 1654 | 816 | 1.00 (0.99, 1.02) | 0.38 | 0.32 |
| Bangladesh | WASH−B (23) | | | | | |
| Burkina Faso | iLiNS−Zinc (24) | 763 | 342 | 1.06 (1.01, 1.11) | 0.03 | 0.06 |
| Ghana | GHANA (25) | | | | | |
| Ghana | iLiNS−DYADG (26) | 348 | 693 | 0.99 (0.98, 1.01) | 0.38 | 0.32 |
| Haiti | HAITI (27) | 43 | 60 | 1.02 (0.98, 1.05) | 0.07 | 0.11 |
| Kenya | WASH−B (28) | | | | | |
| Madagascar | MAHAY (29) | | | | | |
| Malawi | iLiNS−DYADM (30) | 219 | 441 | 1.01 (0.98, 1.04) | 0.08 | 0.12 |
| Malawi | iLiNS−DOSE (31) | 619 | 222 | 1.00 (0.96, 1.04) | 0.05 | 0.07 |
| Mali | PROMIS CS (32) | 111 | 106 | 1.04 (0.93, 1.16) | 0.01 | 0.01 |
| Zimbabwe | SHINE (HIV−) (33) | | | | | |
| Zimbabwe | SHINE (HIV+) (34) | | | | | |
| | **$I^2$ = 0.28, $Tau^2$ = 0.00** | **3757** | **2680** | | | |
| | **Fixed** | | | **1.00 (0.99, 1.01)** | | |
| | **Random** | | | **1.01 (0.99, 1.02)** | | |

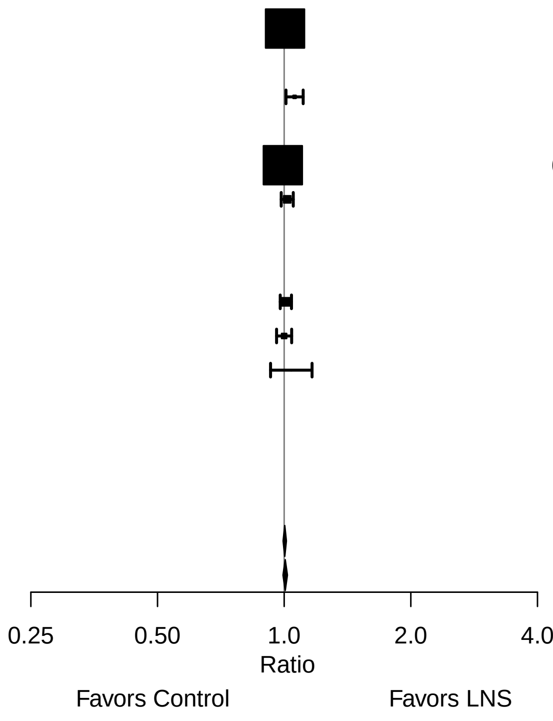

### Supplemental figure 3Z: 18-mo walking without support prevalence difference

| Country | Trial | LNS N | Control N | PD (95% CI) | Fixed W | Random W |
|---|---|---|---|---|---|---|
| Bangladesh | RDNS (22) | 1654 | 816 | 0.00 (−0.01, 0.02) | 0.42 | 0.29 |
| Bangladesh | WASH−B (23) | | | | | |
| Burkina Faso | iLiNS−Zinc (24) | 763 | 342 | 0.05 (0.01, 0.09) | 0.05 | 0.09 |
| Ghana | GHANA (25) | | | | | |
| Ghana | iLiNS−DYADG (26) | 348 | 693 | −0.01 (−0.02, 0.01) | 0.34 | 0.27 |
| Haiti | HAITI (27) | 43 | 60 | 0.02 (−0.02, 0.05) | 0.06 | 0.11 |
| Kenya | WASH−B (28) | | | | | |
| Madagascar | MAHAY (29) | | | | | |
| Malawi | iLiNS−DYADM (30) | 219 | 441 | 0.01 (−0.02, 0.04) | 0.08 | 0.13 |
| Malawi | iLiNS−DOSE (31) | 619 | 222 | 0.00 (−0.04, 0.04) | 0.04 | 0.09 |
| Mali | PROMIS CS (32) | 111 | 106 | 0.03 (−0.06, 0.12) | 0.01 | 0.02 |
| Zimbabwe | SHINE (HIV−) (33) | | | | | |
| Zimbabwe | SHINE (HIV+) (34) | | | | | |
| | **I² = 0.40, Tau² = 0.00** | **3757** | **2680** | | | |
| | **Fixed** | | | **0.00 (0.00, 0.01)** | | |
| | **Random** | | | **0.01 (−0.01, 0.02)** | | |

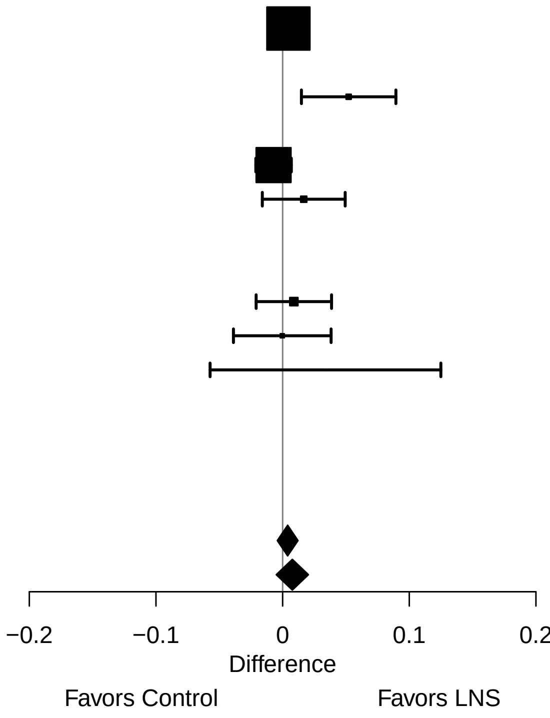

### Supplemental figure 3AA: 18-mo walking with support prevalence ratio

| Country | Trial | LNS N | Control N | PR (95% CI) | Fixed W | Random W |
|---|---|---|---|---|---|---|
| Bangladesh | RDNS (22) | 1654 | 816 | 1.01 (1.00, 1.02) | 0.28 | 0.28 |
| Bangladesh | WASH−B (23) | | | | | |
| Burkina Faso | iLiNS−Zinc (24) | 763 | 342 | 1.02 (1.00, 1.04) | 0.09 | 0.09 |
| Ghana | GHANA (25) | | | | | |
| Ghana | iLiNS−DYADG (26) | 348 | 693 | 1.00 (0.99, 1.01) | 0.41 | 0.41 |
| Haiti | HAITI (27) | 43 | 60 | 1.02 (0.98, 1.05) | 0.03 | 0.03 |
| Kenya | WASH−B (28) | | | | | |
| Madagascar | MAHAY (29) | | | | | |
| Malawi | iLiNS−DYADM (30) | 219 | 441 | 1.00 (0.98, 1.02) | 0.09 | 0.09 |
| Malawi | iLiNS−DOSE (31) | 619 | 222 | 1.00 (0.98, 1.02) | 0.08 | 0.08 |
| Mali | PROMIS CS (32) | 111 | 106 | 1.02 (0.97, 1.07) | 0.02 | 0.02 |
| Zimbabwe | SHINE (HIV−) (33) | | | | | |
| Zimbabwe | SHINE (HIV+) (34) | | | | | |
| | **$I^2$ = 0.00, $Tau^2$ = 0.00** | **3757** | **2680** | | | |
| | **Fixed** | | | **1.00 (1.00, 1.01)** | | |
| | **Random** | | | **1.00 (1.00, 1.01)** | | |

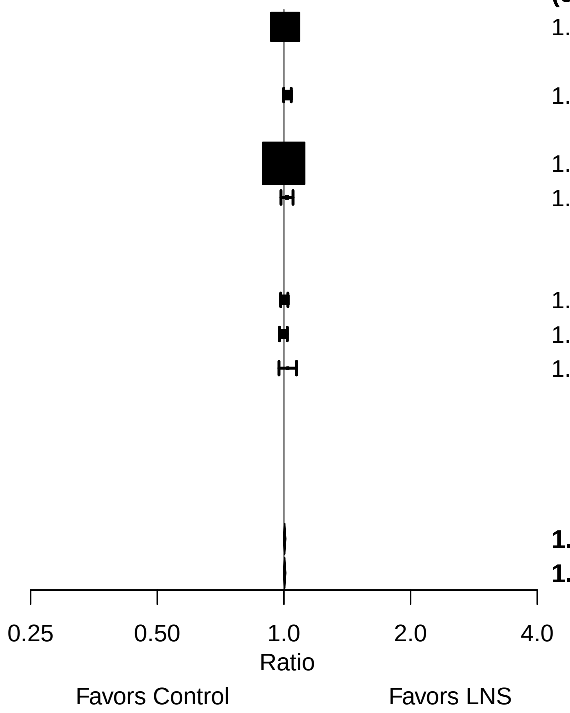

### Supplemental figure 3AB: 18-mo walking with support prevalence difference

| Country | Trial | LNS N | Control N | PD (95% CI) | Fixed W | Random W |
|---|---|---|---|---|---|---|
| Bangladesh | RDNS (22) | 1654 | 816 | 0.01 (0.00, 0.02) | 0.33 | 0.33 |
| Bangladesh | WASH−B (23) | | | | | |
| Burkina Faso | iLiNS−Zinc (24) | 763 | 342 | 0.02 (0.00, 0.04) | 0.06 | 0.06 |
| Ghana | GHANA (25) | | | | | |
| Ghana | iLiNS−DYADG (26) | 348 | 693 | 0.00 (−0.01, 0.01) | 0.39 | 0.39 |
| Haiti | HAITI (27) | 43 | 60 | 0.02 (−0.02, 0.05) | 0.03 | 0.03 |
| Kenya | WASH−B (28) | | | | | |
| Madagascar | MAHAY (29) | | | | | |
| Malawi | iLiNS−DYADM (30) | 219 | 441 | 0.00 (−0.02, 0.02) | 0.09 | 0.09 |
| Malawi | iLiNS−DOSE (31) | 619 | 222 | 0.00 (−0.02, 0.02) | 0.08 | 0.08 |
| Mali | PROMIS CS (32) | 111 | 106 | 0.02 (−0.03, 0.07) | 0.01 | 0.01 |
| Zimbabwe | SHINE (HIV−) (33) | | | | | |
| Zimbabwe | SHINE (HIV+) (34) | | | | | |
| | **I² = 0.00, Tau² = 0.00** | **3757** | **2680** | | | |
| | **Fixed** | | | **0.00 (0.00, 0.01)** | | |
| | **Random** | | | **0.00 (0.00, 0.01)** | | |

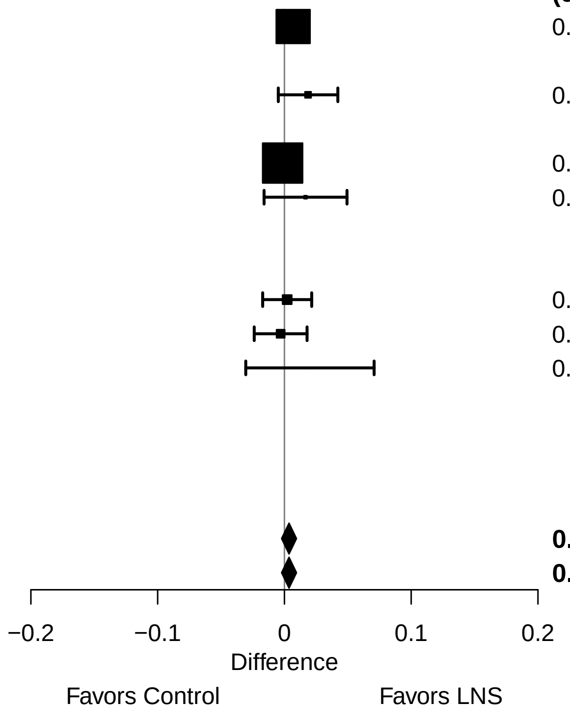

### Supplemental figure 4: Forest plots for effects of SQ-LNS on developmental outcomes stratified by study-level effect modifiers

These figures are forest plots showing the study-level effect modification of intervention effects. Each figure shows the study-level estimates along with the corresponding pooled estimate grouped by study-level effect modifier category. For dichotomous outcomes analyzed via prevalence ratios, the effect estimate is the prevalence in the LNS group divided by the prevalence in the control group. For dichotomous outcomes analyzed via prevalence differences, the effect estimate is the prevalence in the LNS group minus the prevalence in the control group. The labels on the left y-axis correspond to trial level information. The values on the right indicate the study level effect estimate, confidence interval, and weighting for deriving the pooled estimates.

Motor milestone figures show individual trial estimates excluding the JiVitA-4 data because those individual trial results have not yet been published. The pooled estimate in each figure includes the JiVitA-4 data, for consistency with all other analyses.

Figures showing individual trial estimates for the SHINE trial are split by comparison to reflect the cross-over design. For calculating the pooled estimates shown in these figures, the trial is analyzed with LNS intervention arms combined and non-LNS intervention arms combined. Except for when sanitation or water quality are the effect modifier then the LNS +WSH vs WSH comparison is excluded.

### Supplemental figure 4A: Mean difference in language z-score

#### 4A1: Stratified by Geographic region

**Geographic region**
**(p−diff = 0.978)**

| Country | Trial | | Tool | N | N | MD (95% CI) | W |
|---|---|---|---|---|---|---|---|
| **Geographic region − SEAR** | | | | | | | |
| Bangladesh | JiVitA−4 (21) | | BSID−III | 445 | 143 | −0.08 (−0.29, 0.12) | 0.22 |
| Bangladesh | RDNS (22) | | CDI | 1663 | 814 | 0.17 (0.07, 0.26) | 0.37 |
| Bangladesh | WASH−B (23) | | CDI | 1109 | 3353 | 0.09 (0.02, 0.17) | 0.41 |
| **I² = 0.58, Tau² = 0.01** | | | | **3217** | **4310** | **0.08 (−0.04, 0.21)** | |
| **Geographic region − AFR** | | | | | | | |
| Burkina Faso | iLiNS−Zinc (24) | | DMC | 746 | 375 | 0.37 (0.21, 0.53) | 0.10 |
| Ghana | GHANA (25) | | | | | | |
| Ghana | iLiNS−DYADG (26) | | CDI | 331 | 658 | 0.01 (−0.12, 0.14) | 0.12 |
| Kenya | WASH−B (28) | | EASQ | 1362 | 4745 | 0.02 (−0.04, 0.08) | 0.16 |
| Madagascar | MAHAY (29) | | ASQI | 1613 | 1604 | −0.11 (−0.30, 0.07) | 0.09 |
| Malawi | iLiNS−DYADM (30) | | CDI | 215 | 439 | 0.01 (−0.15, 0.18) | 0.10 |
| Malawi | iLiNS−DOSE (31) | | CDI | 645 | 221 | 0.05 (−0.11, 0.20) | 0.11 |
| Mali | PROMIS CS (32) | | DMC | 927 | 944 | 0.15 (−0.02, 0.31) | 0.10 |
| Zimbabwe | SHINE¹ (HIV−) (33), | LNS vs SOC | CDI | 381 | 373 | 0.03 (−0.15, 0.21) | 0.13 |
| Zimbabwe | | LNS+WSH vs WSH | CDI | 436 | 408 | 0.13 (−0.02, 0.29) | 0.13 |
| Zimbabwe | SHINE¹ (HIV+) (34), | LNS vs SOC | CDI | 66 | 68 | 0.06 (−0.29, 0.40) | 0.08 |
| Zimbabwe | | LNS+WSH vs WSH | CDI | 99 | 79 | 0.37 (0.07, 0.67) | 0.08 |
| **I² = 0.66, Tau² = 0.01** | | | | **6821** | **9914** | **0.08 (−0.01, 0.17)** | |

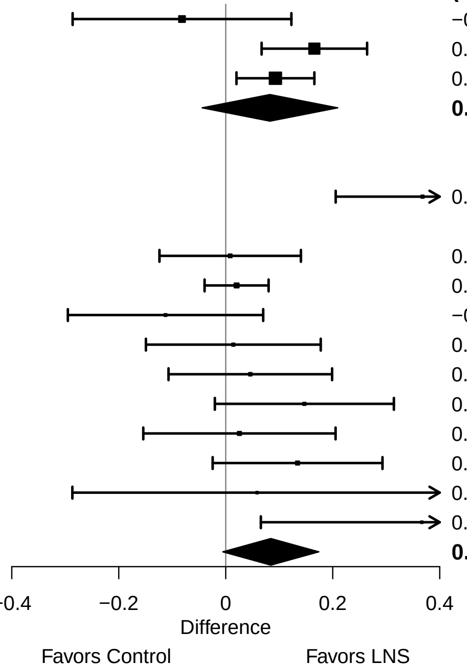

#### Supplemental figure 4A: Mean difference in language z-score

### 4A2: Stratified by Stunting burden

**Stunting burden**
**(p−diff = 0.077)**

| Country | Trial | | Tool | N | N | MD (95% CI) | W |
|---|---|---|---|---|---|---|---|
| **Stunting burden − Less than 35%** | | | | | | | |
| Ghana | GHANA (25) | | | | | | |
| Ghana | iLiNS−DYADG (26) | | CDI | 331 | 658 | 0.01 (−0.12, 0.14) | 0.26 |
| Haiti | HAITI (27) | | Total sounds | 150 | 149 | −0.13 (−0.35, 0.10) | 0.17 |
| Kenya | WASH−B (28) | | EASQ | 1362 | 4745 | 0.02 (−0.04, 0.08) | 0.34 |
| Malawi | iLiNS−DYADM (30) | | CDI | 215 | 439 | 0.01 (−0.15, 0.18) | 0.23 |
| **I² = 0.00, Tau² = 0.00** | | | | **2058** | **5991** | **0.01 (−0.04, 0.06)** | |
| **Stunting burden − More than 35%** | | | | | | | |
| Bangladesh | JiVitA−4 (21) | | BSID−III | 445 | 143 | −0.08 (−0.29, 0.12) | 0.09 |
| Bangladesh | RDNS (22) | | CDI | 1663 | 814 | 0.17 (0.07, 0.26) | 0.14 |
| Bangladesh | WASH−B (23) | | CDI | 1109 | 3353 | 0.09 (0.02, 0.17) | 0.15 |
| Burkina Faso | iLiNS−Zinc (24) | | DMC | 746 | 375 | 0.37 (0.21, 0.53) | 0.11 |
| Madagascar | MAHAY (29) | | ASQI | 1613 | 1604 | −0.11 (−0.30, 0.07) | 0.10 |
| Malawi | iLiNS−DOSE (31) | | CDI | 645 | 221 | 0.05 (−0.11, 0.20) | 0.11 |
| Mali | PROMIS CS (32) | | DMC | 927 | 944 | 0.15 (−0.02, 0.31) | 0.10 |
| Zimbabwe | SHINE¹ (HIV−) (33), | LNS vs SOC | CDI | 381 | 373 | 0.03 (−0.15, 0.21) | 0.13 |
| Zimbabwe | | LNS+WSH vs WSH | CDI | 436 | 408 | 0.13 (−0.02, 0.29) | 0.13 |
| Zimbabwe | SHINE¹ (HIV+) (34), | LNS vs SOC | CDI | 66 | 68 | 0.06 (−0.29, 0.40) | 0.08 |
| Zimbabwe | | LNS+WSH vs WSH | CDI | 99 | 79 | 0.37 (0.07, 0.67) | 0.08 |
| **I² = 0.65, Tau² = 0.01** | | | | **8130** | **8382** | **0.11 (0.02, 0.20)** | |

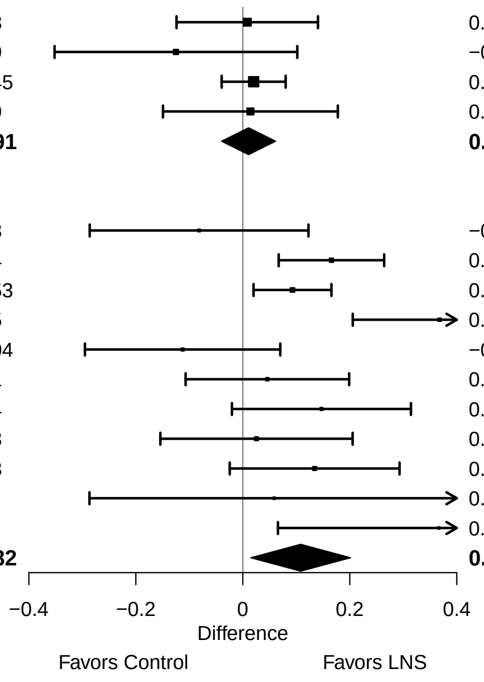

#### Supplemental figure 4A: Mean difference in language z-score

### 4A3: Stratified by Malaria prevalence

**Malaria prevalence**
**(p−diff = 0.365)**

| Country | Trial | Tool | N | N | MD (95% CI) | W |
|---|---|---|---|---|---|---|
| **Malaria prevalence − Less than 10%** | | | | | | |
| Bangladesh | JiVitA−4 (21) | BSID−III | 445 | 143 | −0.08 (−0.29, 0.12) | 0.10 |
| Bangladesh | RDNS (22) | CDI | 1663 | 814 | 0.17 (0.07, 0.26) | 0.15 |
| Bangladesh | WASH−B (23) | CDI | 1109 | 3353 | 0.09 (0.02, 0.17) | 0.17 |
| Haiti | HAITI (27) | Total sounds | 150 | 149 | −0.13 (−0.35, 0.10) | 0.09 |
| Kenya | WASH−B (28) | EASQ | 1362 | 4745 | 0.02 (−0.04, 0.08) | 0.17 |
| Madagascar | MAHAY (29) | ASQI | 1613 | 1604 | −0.11 (−0.30, 0.07) | 0.11 |
| Zimbabwe | SHINE[1] (HIV−) (33), LNS vs SOC | CDI | 381 | 373 | 0.03 (−0.15, 0.21) | 0.14 |
| Zimbabwe | LNS+WSH vs WSH | CDI | 436 | 408 | 0.13 (−0.02, 0.29) | 0.14 |
| Zimbabwe | SHINE[1] (HIV+) (34), LNS vs SOC | CDI | 66 | 68 | 0.06 (−0.29, 0.40) | 0.09 |
| Zimbabwe | LNS+WSH vs WSH | CDI | 99 | 79 | 0.37 (0.07, 0.67) | 0.09 |
| **$I^2$ = 0.60, $Tau^2$ = 0.01** | | | **7324** | **11736** | **0.05 (−0.03, 0.13)** | |
| **Malaria prevalence − At least 10%** | | | | | | |
| Burkina Faso | iLiNS−Zinc (24) | DMC | 746 | 375 | 0.37 (0.21, 0.53) | 0.19 |
| Ghana | GHANA (25) | | | | | |
| Ghana | iLiNS−DYADG (26) | CDI | 331 | 658 | 0.01 (−0.12, 0.14) | 0.22 |
| Malawi | iLiNS−DYADM (30) | CDI | 215 | 439 | 0.01 (−0.15, 0.18) | 0.19 |
| Malawi | iLiNS−DOSE (31) | CDI | 645 | 221 | 0.05 (−0.11, 0.20) | 0.20 |
| Mali | PROMIS CS (32) | DMC | 927 | 944 | 0.15 (−0.02, 0.31) | 0.19 |
| **$I^2$ = 0.72, $Tau^2$ = 0.02** | | | **2864** | **2637** | **0.11 (−0.02, 0.25)** | |

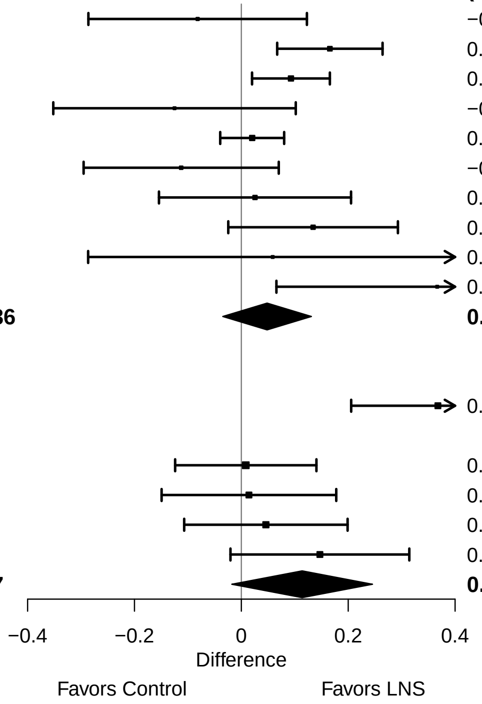

#### Supplemental figure 4A: Mean difference in language z-score

### 4A4: Stratified by Anemia prevalence

**Anemia prevalence**
**(p−diff = 0.530)**

| Country | Trial | Tool | N | N | MD (95% CI) | W |
|---|---|---|---|---|---|---|
| **Anemia prevalence − High** | | | | | | |
| Burkina Faso | iLiNS−Zinc (24) | DMC | 746 | 375 | 0.37 (0.21, 0.53) | 0.21 |
| Ghana | GHANA (25) | | | | | |
| Haiti | HAITI (27) | Total sounds | 150 | 149 | −0.13 (−0.35, 0.10) | 0.16 |
| Malawi | iLiNS−DYADM (30) | CDI | 215 | 439 | 0.01 (−0.15, 0.18) | 0.21 |
| Malawi | iLiNS−DOSE (31) | CDI | 645 | 221 | 0.05 (−0.11, 0.20) | 0.22 |
| Mali | PROMIS CS (32) | DMC | 927 | 944 | 0.15 (−0.02, 0.31) | 0.21 |
| **I² = 0.75, Tau² = 0.02** | | | **2683** | **2128** | **0.10 (−0.06, 0.26)** | |
| **Anemia prevalence − Moderate** | | | | | | |
| Bangladesh | JiVitA−4 (21) | BSID−III | 445 | 143 | −0.08 (−0.29, 0.12) | 0.09 |
| Bangladesh | RDNS (22) | CDI | 1663 | 814 | 0.17 (0.07, 0.26) | 0.15 |
| Bangladesh | WASH−B (23) | CDI | 1109 | 3353 | 0.09 (0.02, 0.17) | 0.16 |
| Ghana | iLiNS−DYADG (26) | CDI | 331 | 658 | 0.01 (−0.12, 0.14) | 0.13 |
| Kenya | WASH−B (28) | EASQ | 1362 | 4745 | 0.02 (−0.04, 0.08) | 0.16 |
| Madagascar | MAHAY (29) | ASQI | 1613 | 1604 | −0.11 (−0.30, 0.07) | 0.10 |
| Zimbabwe | SHINE¹ (HIV−) (33), LNS vs SOC | CDI | 381 | 373 | 0.03 (−0.15, 0.21) | 0.13 |
| Zimbabwe | LNS+WSH vs WSH | CDI | 436 | 408 | 0.13 (−0.02, 0.29) | 0.13 |
| Zimbabwe | SHINE¹ (HIV+) (34), LNS vs SOC | CDI | 66 | 68 | 0.06 (−0.29, 0.40) | 0.08 |
| Zimbabwe | LNS+WSH vs WSH | CDI | 99 | 79 | 0.37 (0.07, 0.67) | 0.08 |
| **I² = 0.55, Tau² = 0.01** | | | **7505** | **12245** | **0.06 (−0.01, 0.13)** | |

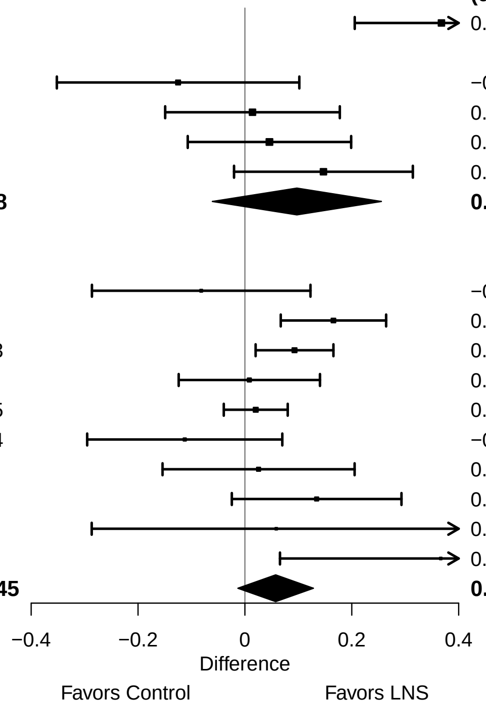

#### Supplemental figure 4A: Mean difference in language z-score

### 4A5: Stratified by Source water quality

**Source water quality**
**(p−diff = 0.536)**

| Country | Trial | Tool | N | N | MD (95% CI) | W |
|---|---|---|---|---|---|---|
| **Source water quality − Improved** | | | | | | |
| Bangladesh | JiVitA−4 (21) | BSID−III | 445 | 143 | −0.08 (−0.29, 0.12) | 0.11 |
| Bangladesh | RDNS (22) | CDI | 1663 | 814 | 0.17 (0.07, 0.26) | 0.18 |
| Bangladesh | WASH−B (23) | CDI | 548 | 1099 | 0.17 (0.06, 0.29) | 0.17 |
| Ghana | GHANA (25) | | | | | |
| Ghana | iLiNS−DYADG (26) | CDI | 331 | 658 | 0.01 (−0.12, 0.14) | 0.16 |
| Haiti | HAITI (27) | Total sounds | 150 | 149 | −0.13 (−0.35, 0.10) | 0.10 |
| Malawi | iLiNS−DYADM (30) | CDI | 215 | 439 | 0.01 (−0.15, 0.18) | 0.14 |
| Malawi | iLiNS−DOSE (31) | CDI | 645 | 221 | 0.05 (−0.11, 0.20) | 0.14 |
| **I² = 0.54, Tau² = 0.01** | | | **3997** | **3523** | **0.05 (−0.03, 0.14)** | |
| **Source water quality − Unimproved** | | | | | | |
| Burkina Faso | iLiNS−Zinc (24) | DMC | 746 | 375 | 0.37 (0.21, 0.53) | 0.18 |
| Kenya | WASH−B (28) | EASQ | 649 | 2073 | 0.05 (−0.05, 0.15) | 0.24 |
| Madagascar | MAHAY (29) | ASQI | 1613 | 1604 | −0.11 (−0.30, 0.07) | 0.16 |
| Mali | PROMIS CS (32) | DMC | 927 | 944 | 0.15 (−0.02, 0.31) | 0.18 |
| Zimbabwe | SHINE (HIV−) (33) | CDI | 381 | 373 | 0.03 (−0.15, 0.21) | 0.17 |
| Zimbabwe | SHINE (HIV+) (34) | CDI | 66 | 68 | 0.06 (−0.29, 0.40) | 0.08 |
| **I² = 0.72, Tau² = 0.02** | | | **4382** | **5437** | **0.09 (−0.04, 0.22)** | |

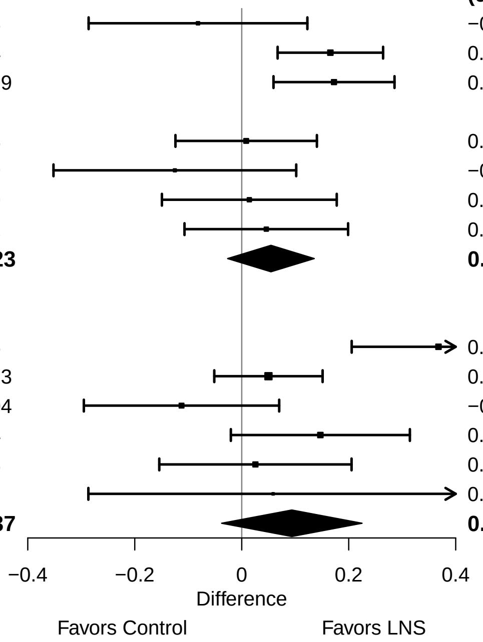

#### Supplemental figure 4A: Mean difference in language z-score

### 4A6: Stratified by Sanitation

**Sanitation**
**(p−diff = 0.991)**

| Country | Trial | Tool | N | N | MD (95% CI) | W |
|---|---|---|---|---|---|---|
| **Sanitation − Improved** | | | | | | |
| Bangladesh | JiVitA−4 (21) | BSID−III | 445 | 143 | −0.08 (−0.29, 0.12) | 0.13 |
| Bangladesh | RDNS (22) | CDI | 1663 | 814 | 0.17 (0.07, 0.26) | 0.21 |
| Bangladesh | WASH−B (23) | CDI | 548 | 1099 | 0.17 (0.06, 0.29) | 0.20 |
| Ghana | GHANA (25) | | | | | |
| Ghana | iLiNS−DYADG (26) | CDI | 331 | 658 | 0.01 (−0.12, 0.14) | 0.18 |
| Haiti | HAITI (27) | Total sounds | 150 | 149 | −0.13 (−0.35, 0.10) | 0.12 |
| Mali | PROMIS CS (32) | DMC | 927 | 944 | 0.15 (−0.02, 0.31) | 0.16 |
| **I² = 0.59, Tau² = 0.01** | | | **4064** | **3807** | **0.07 (−0.03, 0.17)** | |
| **Sanitation − Unimproved** | | | | | | |
| Burkina Faso | iLiNS−Zinc (24) | DMC | 746 | 375 | 0.37 (0.21, 0.53) | 0.15 |
| Kenya | WASH−B (28) | EASQ | 649 | 2073 | 0.05 (−0.05, 0.15) | 0.20 |
| Madagascar | MAHAY (29) | ASQI | 1613 | 1604 | −0.11 (−0.30, 0.07) | 0.14 |
| Malawi | iLiNS−DYADM (30) | CDI | 215 | 439 | 0.01 (−0.15, 0.18) | 0.15 |
| Malawi | iLiNS−DOSE (31) | CDI | 645 | 221 | 0.05 (−0.11, 0.20) | 0.16 |
| Zimbabwe | SHINE (HIV−) (33) | CDI | 381 | 373 | 0.03 (−0.15, 0.21) | 0.14 |
| Zimbabwe | SHINE (HIV+) (34) | CDI | 66 | 68 | 0.06 (−0.29, 0.40) | 0.06 |
| **I² = 0.66, Tau² = 0.01** | | | **4315** | **5153** | **0.07 (−0.04, 0.18)** | |

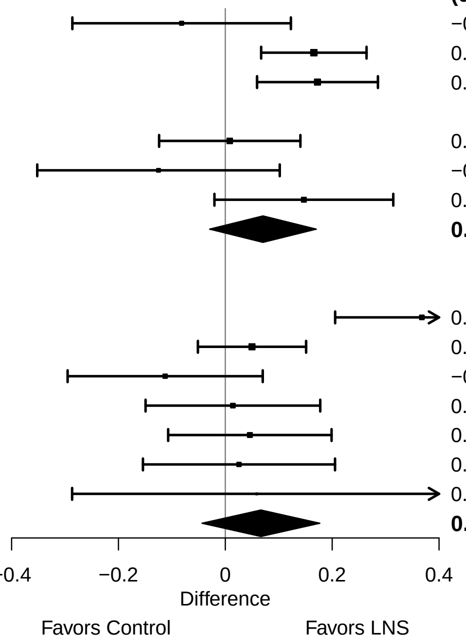

#### Supplemental figure 4A: Mean difference in language z-score

### 4A7: Stratified by Supplement duration

**Supplement duration**
**(p−diff = 0.524)**

| Country | Trial | Tool | N | N | MD (95% CI) | W |
|---|---|---|---|---|---|---|
| **Supplement duration − 12m or less** | | | | | | |
| Bangladesh | JiVitA−4 (21) | BSID−III | 445 | 143 | −0.08 (−0.29, 0.12) | 0.10 |
| Burkina Faso | iLiNS−Zinc (24) | DMC | 746 | 375 | 0.37 (0.21, 0.53) | 0.12 |
| Ghana | GHANA (25) | | | | | |
| Ghana | iLiNS−DYADG (26) | CDI | 331 | 658 | 0.01 (−0.12, 0.14) | 0.13 |
| Haiti | HAITI (27) | Total sounds | 150 | 149 | −0.13 (−0.35, 0.10) | 0.09 |
| Madagascar | MAHAY (29) | ASQI | 1613 | 1604 | −0.11 (−0.30, 0.07) | 0.11 |
| Malawi | iLiNS−DYADM (30) | CDI | 215 | 439 | 0.01 (−0.15, 0.18) | 0.12 |
| Malawi | iLiNS−DOSE (31) | CDI | 645 | 221 | 0.05 (−0.11, 0.20) | 0.12 |
| Zimbabwe | SHINE[1] (HIV−) (33), LNS vs SOC | CDI | 381 | 373 | 0.03 (−0.15, 0.21) | 0.14 |
| Zimbabwe | LNS+WSH vs WSH | CDI | 436 | 408 | 0.13 (−0.02, 0.29) | 0.14 |
| Zimbabwe | SHINE[1] (HIV+) (34), LNS vs SOC | CDI | 66 | 68 | 0.06 (−0.29, 0.40) | 0.09 |
| Zimbabwe | LNS+WSH vs WSH | CDI | 99 | 79 | 0.37 (0.07, 0.67) | 0.09 |
| **$I^2$ = 0.68, $Tau^2$ = 0.02** | | | **5127** | **4517** | **0.05 (−0.05, 0.16)** | |
| **Supplement duration − > 12m** | | | | | | |
| Bangladesh | RDNS (22) | CDI | 1663 | 814 | 0.17 (0.07, 0.26) | 0.25 |
| Bangladesh | WASH−B (23) | CDI | 1109 | 3353 | 0.09 (0.02, 0.17) | 0.27 |
| Kenya | WASH−B (28) | EASQ | 1362 | 4745 | 0.02 (−0.04, 0.08) | 0.28 |
| Mali | PROMIS CS (32) | DMC | 927 | 944 | 0.15 (−0.02, 0.31) | 0.19 |
| **$I^2$ = 0.59, $Tau^2$ = 0.00** | | | **5061** | **9856** | **0.09 (0.02, 0.16)** | |

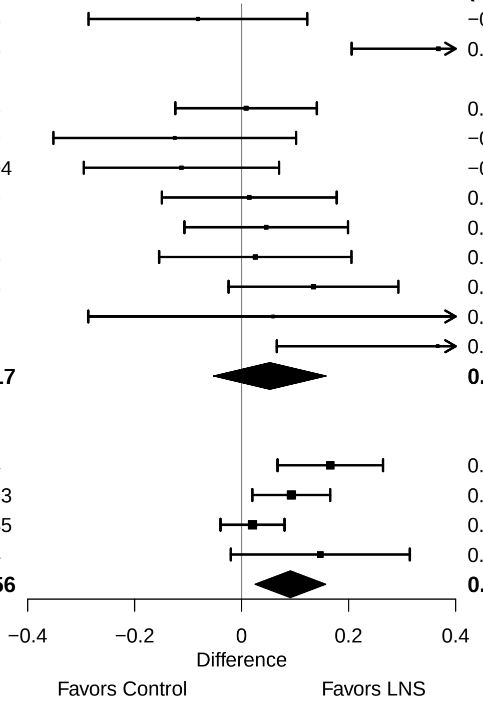

#### Supplemental figure 4A: Mean difference in language z-score

### 4A8: Stratified by Frequency of contact

**Frequency of contact**
**(p−diff = 0.853)**

| Country | Trial | Tool | N | N | MD (95% CI) | W |
|---|---|---|---|---|---|---|
| **Frequency of contact − Monthly** | | | | | | |
| Bangladesh | RDNS (22) | CDI | 1663 | 814 | 0.17 (0.07, 0.26) | 0.18 |
| Haiti | HAITI (27) | Total sounds | 150 | 149 | −0.13 (−0.35, 0.10) | 0.10 |
| Kenya | WASH−B (28) | EASQ | 1362 | 4745 | 0.02 (−0.04, 0.08) | 0.20 |
| Madagascar | MAHAY (29) | ASQI | 1613 | 1604 | −0.11 (−0.30, 0.07) | 0.12 |
| Mali | PROMIS CS (32) | DMC | 927 | 944 | 0.15 (−0.02, 0.31) | 0.13 |
| Zimbabwe | SHINE[1] (HIV−) (33), LNS vs SOC | CDI | 381 | 373 | 0.03 (−0.15, 0.21) | 0.16 |
| Zimbabwe | LNS+WSH vs WSH | CDI | 436 | 408 | 0.13 (−0.02, 0.29) | 0.16 |
| Zimbabwe | SHINE[1] (HIV+) (34), LNS vs SOC | CDI | 66 | 68 | 0.06 (−0.29, 0.40) | 0.10 |
| Zimbabwe | LNS+WSH vs WSH | CDI | 99 | 79 | 0.37 (0.07, 0.67) | 0.10 |
| **$I^2$ = 0.62, $Tau^2$ = 0.01** | | | **6697** | **9184** | **0.07 (−0.03, 0.16)** | |
| **Frequency of contact − Weekly** | | | | | | |
| Bangladesh | JiVitA−4 (21) | BSID−III | 445 | 143 | −0.08 (−0.29, 0.12) | 0.13 |
| Bangladesh | WASH−B (23) | CDI | 1109 | 3353 | 0.09 (0.02, 0.17) | 0.22 |
| Burkina Faso | iLiNS−Zinc (24) | DMC | 746 | 375 | 0.37 (0.21, 0.53) | 0.16 |
| Ghana | GHANA (25) | | | | | |
| Ghana | iLiNS−DYADG (26) | CDI | 331 | 658 | 0.01 (−0.12, 0.14) | 0.18 |
| Malawi | iLiNS−DYADM (30) | CDI | 215 | 439 | 0.01 (−0.15, 0.18) | 0.15 |
| Malawi | iLiNS−DOSE (31) | CDI | 645 | 221 | 0.05 (−0.11, 0.20) | 0.16 |
| **$I^2$ = 0.70, $Tau^2$ = 0.02** | | | **3491** | **5189** | **0.08 (−0.04, 0.20)** | |

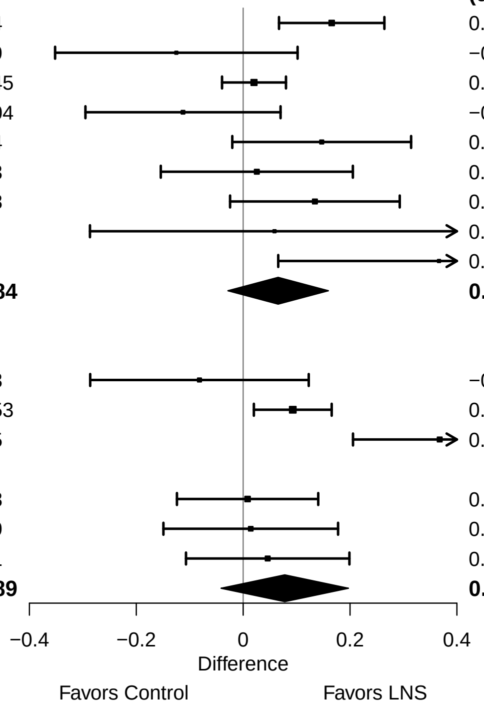

#### Supplemental figure 4A: Mean difference in language z-score

### 4A9: Stratified by Average SQ-LNS compliance

**Average SQ−LNS compliance**
**(p−diff = 0.918)**

**Average SQ−LNS compliance − Low**

| Country | Trial | Tool | N | N | MD (95% CI) | W |
|---|---|---|---|---|---|---|
| Ghana | iLiNS−DYADG (26) | CDI | 331 | 658 | 0.01 (−0.12, 0.14) | 0.19 |
| Malawi | iLiNS−DYADM (30) | CDI | 215 | 439 | 0.01 (−0.15, 0.18) | 0.16 |
| Malawi | iLiNS−DOSE (31) | CDI | 645 | 221 | 0.05 (−0.11, 0.20) | 0.17 |
| Mali | PROMIS CS (32) | DMC | 927 | 944 | 0.15 (−0.02, 0.31) | 0.16 |
| Zimbabwe | SHINE¹ (HIV−) (33), LNS vs SOC | CDI | 381 | 373 | 0.03 (−0.15, 0.21) | 0.20 |
| Zimbabwe | LNS+WSH vs WSH | CDI | 436 | 408 | 0.13 (−0.02, 0.29) | 0.20 |
| Zimbabwe | SHINE¹ (HIV+) (34), LNS vs SOC | CDI | 66 | 68 | 0.06 (−0.29, 0.40) | 0.12 |
| Zimbabwe | LNS+WSH vs WSH | CDI | 99 | 79 | 0.37 (0.07, 0.67) | 0.12 |
| **I² = 0.00, Tau² = 0.00** | | | **3100** | **3190** | **0.07 (0.01, 0.13)** | |

**Average SQ−LNS compliance − High**

| Country | Trial | Tool | N | N | MD (95% CI) | W |
|---|---|---|---|---|---|---|
| Bangladesh | JiVitA−4 (21) | BSID−III | 445 | 143 | −0.08 (−0.29, 0.12) | 0.12 |
| Bangladesh | RDNS (22) | CDI | 1663 | 814 | 0.17 (0.07, 0.26) | 0.20 |
| Bangladesh | WASH−B (23) | CDI | 1109 | 3353 | 0.09 (0.02, 0.17) | 0.21 |
| Burkina Faso | iLiNS−Zinc (24) | DMC | 746 | 375 | 0.37 (0.21, 0.53) | 0.15 |
| Ghana | GHANA (25) | | | | | |
| Haiti | HAITI (27) | Total sounds | 150 | 149 | −0.13 (−0.35, 0.10) | 0.10 |
| Kenya | WASH−B (28) | EASQ | 1362 | 4745 | 0.02 (−0.04, 0.08) | 0.22 |
| **I² = 0.80, Tau² = 0.02** | | | **5475** | **9579** | **0.08 (−0.05, 0.22)** | |

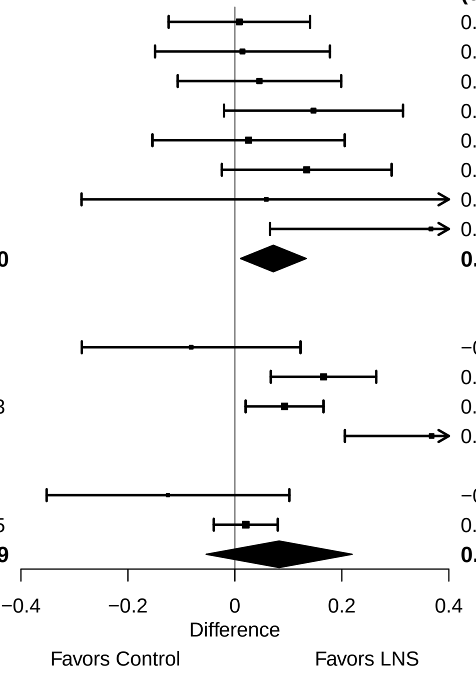

### Supplemental figure 4B: Language lowest decile prevalence ratio

#### 4B1: Stratified by Geographic region

**Geographic region**
**(p−diff = 0.624)**

| Country | Trial | Tool | N | N | PR (95% CI) | W |
|---|---|---|---|---|---|---|
| **Geographic region − SEAR** | | | | | | |
| Bangladesh | JiVitA−4 (21) | BSID−III | 445 | 143 | 0.86 (0.50, 1.49) | 0.16 |
| Bangladesh | RDNS (22) | CDI | 1663 | 814 | 0.72 (0.57, 0.91) | 0.41 |
| Bangladesh | WASH−B (23) | CDI | 1109 | 3353 | 0.81 (0.65, 1.00) | 0.43 |
| **I² = 0.00, Tau² = 0.00** | | | **3217** | **4310** | **0.77 (0.66, 0.90)** | |
| **Geographic region − AFR** | | | | | | |
| Burkina Faso | iLiNS−Zinc (24) | DMC | 746 | 375 | 0.46 (0.33, 0.64) | 0.13 |
| Ghana | GHANA (25) | | | | | |
| Ghana | iLiNS−DYADG (26) | CDI | 331 | 658 | 1.09 (0.74, 1.61) | 0.11 |
| Kenya | WASH−B (28) | EASQ | 1362 | 4745 | 0.95 (0.78, 1.16) | 0.20 |
| Madagascar | MAHAY (29) | ASQI | 1613 | 1604 | 1.17 (0.75, 1.83) | 0.09 |
| Malawi | iLiNS−DYADM (30) | CDI | 215 | 439 | 0.87 (0.53, 1.43) | 0.08 |
| Malawi | iLiNS−DOSE (31) | CDI | 645 | 221 | 0.95 (0.61, 1.50) | 0.09 |
| Mali | PROMIS CS (32) | DMC | 927 | 944 | 0.77 (0.56, 1.07) | 0.13 |
| Zimbabwe | SHINE¹ (HIV−) (33), LNS vs SOC | CDI | 381 | 373 | 1.05 (0.67, 1.65) | 0.13 |
| Zimbabwe | LNS+WSH vs WSH | CDI | 436 | 408 | 0.77 (0.49, 1.20) | 0.13 |
| Zimbabwe | SHINE¹ (HIV+) (34), LNS vs SOC | CDI | 66 | 68 | 1.29 (0.54, 3.06) | 0.05 |
| Zimbabwe | LNS+WSH vs WSH | CDI | 99 | 79 | 0.32 (0.11, 0.92) | 0.05 |
| **I² = 0.59, Tau² = 0.04** | | | **6821** | **9914** | **0.84 (0.70, 1.02)** | |

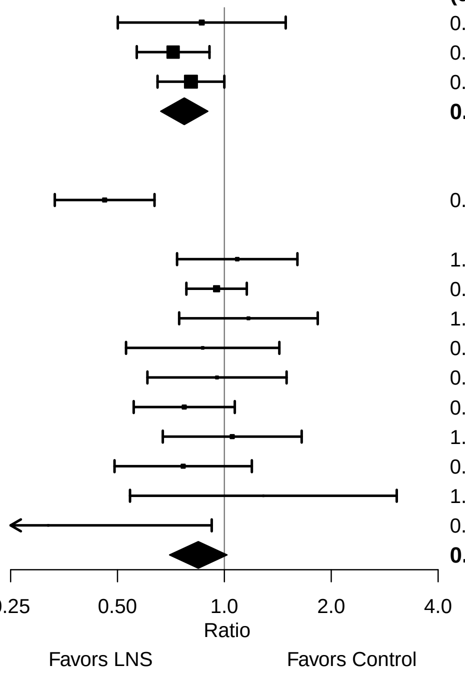

#### Supplemental figure 4B: Language lowest decile prevalence ratio

#### 4B2: Stratified by Stunting burden

**Stunting burden**
**(p−diff = 0.046)**

**Stunting burden − Less than 35%**

| Country | Trial | Tool | N | N | PR (95% CI) | W |
|---|---|---|---|---|---|---|
| Ghana | GHANA (25) | | | | | |
| Ghana | iLiNS−DYADG (26) | CDI | 331 | 658 | 1.09 (0.74, 1.61) | 0.27 |
| Haiti | HAITI (27) | Total sounds | 150 | 149 | 2.75 (1.28, 5.93) | 0.14 |
| Kenya | WASH−B (28) | EASQ | 1362 | 4745 | 0.95 (0.78, 1.16) | 0.37 |
| Malawi | iLiNS−DYADM (30) | CDI | 215 | 439 | 0.87 (0.53, 1.43) | 0.22 |
| **I² = 0.59, Tau² = 0.16** | | | **2058** | **5991** | **1.16 (0.73, 1.83)** | |

**Stunting burden − More than 35%**

| Country | Trial | Tool | N | N | PR (95% CI) | W |
|---|---|---|---|---|---|---|
| Bangladesh | JiVitA−4 (21) | BSID−III | 445 | 143 | 0.86 (0.50, 1.49) | 0.08 |
| Bangladesh | RDNS (22) | CDI | 1663 | 814 | 0.72 (0.57, 0.91) | 0.14 |
| Bangladesh | WASH−B (23) | CDI | 1109 | 3353 | 0.81 (0.65, 1.00) | 0.14 |
| Burkina Faso | iLiNS−Zinc (24) | DMC | 746 | 375 | 0.46 (0.33, 0.64) | 0.12 |
| Madagascar | MAHAY (29) | ASQI | 1613 | 1604 | 1.17 (0.75, 1.83) | 0.10 |
| Malawi | iLiNS−DOSE (31) | CDI | 645 | 221 | 0.95 (0.61, 1.50) | 0.10 |
| Mali | PROMIS CS (32) | DMC | 927 | 944 | 0.77 (0.56, 1.07) | 0.12 |
| Zimbabwe | SHINE¹ (HIV−) (33), LNS vs SOC | CDI | 381 | 373 | 1.05 (0.67, 1.65) | 0.12 |
| Zimbabwe | LNS+WSH vs WSH | CDI | 436 | 408 | 0.77 (0.49, 1.20) | 0.12 |
| Zimbabwe | SHINE¹ (HIV+) (34), LNS vs SOC | CDI | 66 | 68 | 1.29 (0.54, 3.06) | 0.06 |
| Zimbabwe | LNS+WSH vs WSH | CDI | 99 | 79 | 0.32 (0.11, 0.92) | 0.06 |
| **I² = 0.49, Tau² = 0.03** | | | **8130** | **8382** | **0.78 (0.66, 0.92)** | |

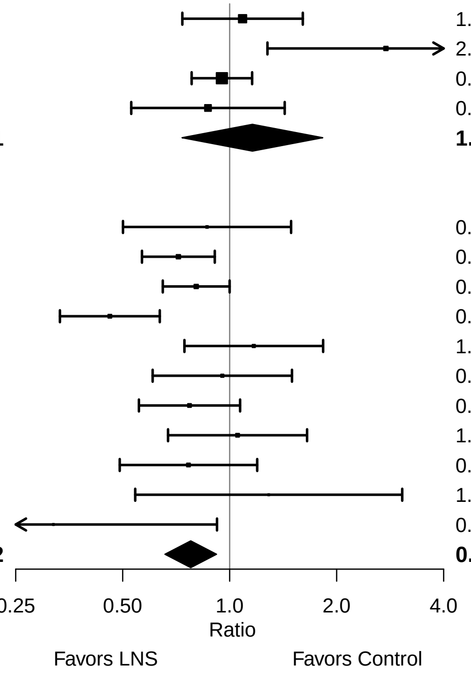

#### Supplemental figure 4B: Language lowest decile prevalence ratio

#### 4B3: Stratified by Malaria prevalence

**Malaria prevalence**
**(p−diff = 0.322)**

| Country | Trial | Tool | N | N | PR (95% CI) | W |
|---|---|---|---|---|---|---|
| **Malaria prevalence − Less than 10%** | | | | | | |
| Bangladesh | JiVitA−4 (21) | BSID−III | 445 | 143 | 0.86 (0.50, 1.49) | 0.10 |
| Bangladesh | RDNS (22) | CDI | 1663 | 814 | 0.72 (0.57, 0.91) | 0.16 |
| Bangladesh | WASH−B (23) | CDI | 1109 | 3353 | 0.81 (0.65, 1.00) | 0.17 |
| Haiti | HAITI (27) | Total sounds | 150 | 149 | 2.75 (1.28, 5.93) | 0.06 |
| Kenya | WASH−B (28) | EASQ | 1362 | 4745 | 0.95 (0.78, 1.16) | 0.17 |
| Madagascar | MAHAY (29) | ASQI | 1613 | 1604 | 1.17 (0.75, 1.83) | 0.11 |
| Zimbabwe | SHINE[1] (HIV−) (33), LNS vs SOC | CDI | 381 | 373 | 1.05 (0.67, 1.65) | 0.14 |
| Zimbabwe | LNS+WSH vs WSH | CDI | 436 | 408 | 0.77 (0.49, 1.20) | 0.14 |
| Zimbabwe | SHINE[1] (HIV+) (34), LNS vs SOC | CDI | 66 | 68 | 1.29 (0.54, 3.06) | 0.07 |
| Zimbabwe | LNS+WSH vs WSH | CDI | 99 | 79 | 0.32 (0.11, 0.92) | 0.07 |
| **$I^2$ = 0.52, $Tau^2$ = 0.08** | | | **7324** | **11736** | **0.93 (0.73, 1.18)** | |
| **Malaria prevalence − At least 10%** | | | | | | |
| Burkina Faso | iLiNS−Zinc (24) | DMC | 746 | 375 | 0.46 (0.33, 0.64) | 0.23 |
| Ghana | GHANA (25) | | | | | |
| Ghana | iLiNS−DYADG (26) | CDI | 331 | 658 | 1.09 (0.74, 1.61) | 0.20 |
| Malawi | iLiNS−DYADM (30) | CDI | 215 | 439 | 0.87 (0.53, 1.43) | 0.17 |
| Malawi | iLiNS−DOSE (31) | CDI | 645 | 221 | 0.95 (0.61, 1.50) | 0.18 |
| Mali | PROMIS CS (32) | DMC | 927 | 944 | 0.77 (0.56, 1.07) | 0.22 |
| **$I^2$ = 0.71, $Tau^2$ = 0.08** | | | **2864** | **2637** | **0.78 (0.58, 1.05)** | |

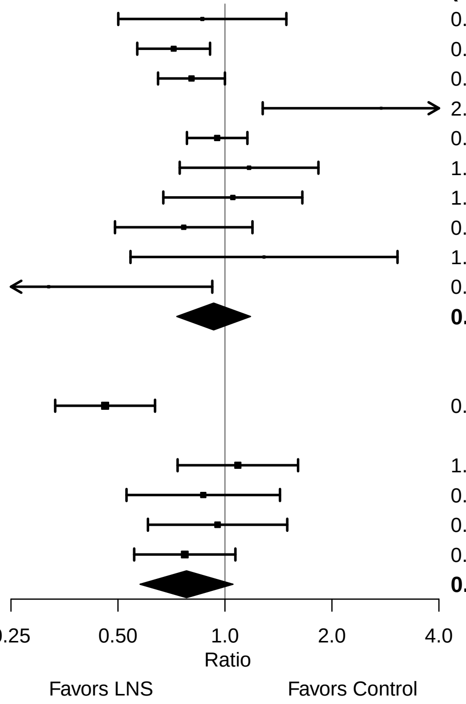

#### Supplemental figure 4B: Language lowest decile prevalence ratio

### 4B4: Stratified by Anemia prevalence

**Anemia prevalence**
**(p−diff = 0.644)**

| Country | Trial | Tool | N | N | PR (95% CI) | W |
|---|---|---|---|---|---|---|
| **Anemia prevalence − High** | | | | | | |
| Burkina Faso | iLiNS−Zinc (24) | DMC | 746 | 375 | 0.46 (0.33, 0.64) | 0.25 |
| Ghana | GHANA (25) | | | | | |
| Haiti | HAITI (27) | Total sounds | 150 | 149 | 2.75 (1.28, 5.93) | 0.11 |
| Malawi | iLiNS−DYADM (30) | CDI | 215 | 439 | 0.87 (0.53, 1.43) | 0.18 |
| Malawi | iLiNS−DOSE (31) | CDI | 645 | 221 | 0.95 (0.61, 1.50) | 0.20 |
| Mali | PROMIS CS (32) | DMC | 927 | 944 | 0.77 (0.56, 1.07) | 0.25 |
| **I² = 0.81, Tau² = 0.32** | | | **2683** | **2128** | **0.90 (0.53, 1.55)** | |
| **Anemia prevalence − Moderate** | | | | | | |
| Bangladesh | JiVitA−4 (21) | BSID−III | 445 | 143 | 0.86 (0.50, 1.49) | 0.09 |
| Bangladesh | RDNS (22) | CDI | 1663 | 814 | 0.72 (0.57, 0.91) | 0.15 |
| Bangladesh | WASH−B (23) | CDI | 1109 | 3353 | 0.81 (0.65, 1.00) | 0.16 |
| Ghana | iLiNS−DYADG (26) | CDI | 331 | 658 | 1.09 (0.74, 1.61) | 0.12 |
| Kenya | WASH−B (28) | EASQ | 1362 | 4745 | 0.95 (0.78, 1.16) | 0.16 |
| Madagascar | MAHAY (29) | ASQI | 1613 | 1604 | 1.17 (0.75, 1.83) | 0.11 |
| Zimbabwe | SHINE¹ (HIV−) (33), LNS vs SOC | CDI | 381 | 373 | 1.05 (0.67, 1.65) | 0.14 |
| Zimbabwe | LNS+WSH vs WSH | CDI | 436 | 408 | 0.77 (0.49, 1.20) | 0.14 |
| Zimbabwe | SHINE¹ (HIV+) (34), LNS vs SOC | CDI | 66 | 68 | 1.29 (0.54, 3.06) | 0.07 |
| Zimbabwe | LNS+WSH vs WSH | CDI | 99 | 79 | 0.32 (0.11, 0.92) | 0.07 |
| **I² = 0.03, Tau² = 0.00** | | | **7505** | **12245** | **0.87 (0.78, 0.97)** | |

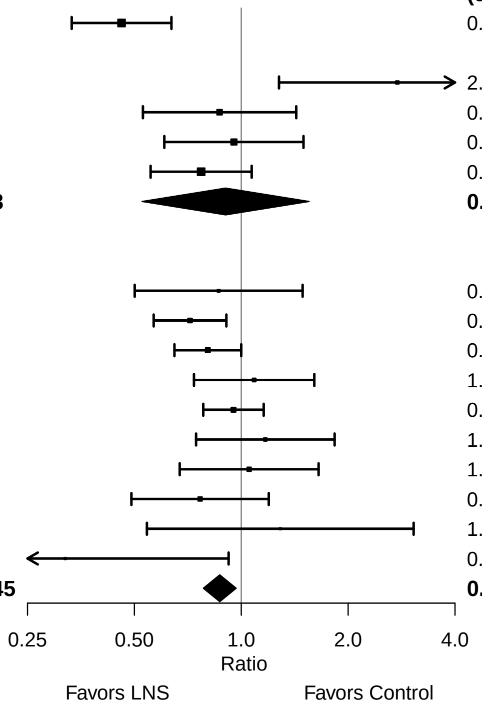

#### Supplemental figure 4B: Language lowest decile prevalence ratio

### 4B5: Stratified by Source water quality

**Source water quality**
**(p−diff = 0.737)**

**Source water quality − Improved**

| Country | Trial | Tool | N | N | PR (95% CI) | W |
|---|---|---|---|---|---|---|
| Bangladesh | JiVitA−4 (21) | BSID−III | 445 | 143 | 0.86 (0.50, 1.49) | 0.12 |
| Bangladesh | RDNS (22) | CDI | 1663 | 814 | 0.72 (0.57, 0.91) | 0.19 |
| Bangladesh | WASH−B (23) | CDI | 548 | 1099 | 0.64 (0.46, 0.89) | 0.17 |
| Ghana | GHANA (25) | | | | | |
| Ghana | iLiNS−DYADG (26) | CDI | 331 | 658 | 1.09 (0.74, 1.61) | 0.16 |
| Haiti | HAITI (27) | Total sounds | 150 | 149 | 2.75 (1.28, 5.93) | 0.08 |
| Malawi | iLiNS−DYADM (30) | CDI | 215 | 439 | 0.87 (0.53, 1.43) | 0.13 |
| Malawi | iLiNS−DOSE (31) | CDI | 645 | 221 | 0.95 (0.61, 1.50) | 0.14 |
| **I² = 0.61, Tau² = 0.12** | | | **3997** | **3523** | **0.93 (0.68, 1.26)** | |

**Source water quality − Unimproved**

| Country | Trial | Tool | N | N | PR (95% CI) | W |
|---|---|---|---|---|---|---|
| Burkina Faso | iLiNS−Zinc (24) | DMC | 746 | 375 | 0.46 (0.33, 0.64) | 0.20 |
| Kenya | WASH−B (28) | EASQ | 649 | 2073 | 0.96 (0.72, 1.28) | 0.21 |
| Madagascar | MAHAY (29) | ASQI | 1613 | 1604 | 1.17 (0.75, 1.83) | 0.16 |
| Mali | PROMIS CS (32) | DMC | 927 | 944 | 0.77 (0.56, 1.07) | 0.19 |
| Zimbabwe | SHINE (HIV−) (33) | CDI | 381 | 373 | 1.05 (0.67, 1.65) | 0.16 |
| Zimbabwe | SHINE (HIV+) (34) | CDI | 66 | 68 | 1.29 (0.54, 3.06) | 0.08 |
| **I² = 0.72, Tau² = 0.09** | | | **4382** | **5437** | **0.85 (0.63, 1.15)** | |

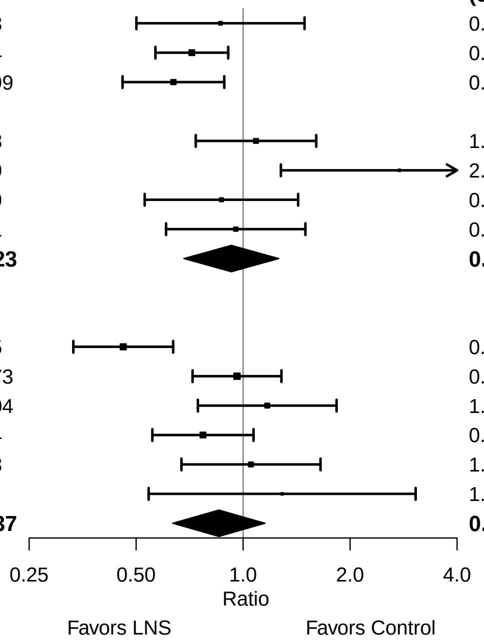

#### Supplemental figure 4B: Language lowest decile prevalence ratio

### 4B6: Stratified by Sanitation

**Sanitation**
**(p−diff = 0.994)**

| Country | Trial | Tool | N | N | PR (95% CI) | W |
|---|---|---|---|---|---|---|
| **Sanitation − Improved** | | | | | | |
| Bangladesh | JiVitA−4 (21) | BSID−III | 445 | 143 | 0.86 (0.50, 1.49) | 0.14 |
| Bangladesh | RDNS (22) | CDI | 1663 | 814 | 0.72 (0.57, 0.91) | 0.22 |
| Bangladesh | WASH−B (23) | CDI | 548 | 1099 | 0.64 (0.46, 0.89) | 0.19 |
| Ghana | GHANA (25) | | | | | |
| Ghana | iLiNS−DYADG (26) | CDI | 331 | 658 | 1.09 (0.74, 1.61) | 0.17 |
| Haiti | HAITI (27) | Total sounds | 150 | 149 | 2.75 (1.28, 5.93) | 0.09 |
| Mali | PROMIS CS (32) | DMC | 927 | 944 | 0.77 (0.56, 1.07) | 0.19 |
| **I² = 0.67, Tau² = 0.17** | | | **4064** | **3807** | **0.91 (0.63, 1.33)** | |
| **Sanitation − Unimproved** | | | | | | |
| Burkina Faso | iLiNS−Zinc (24) | DMC | 746 | 375 | 0.46 (0.33, 0.64) | 0.18 |
| Kenya | WASH−B (28) | EASQ | 649 | 2073 | 0.96 (0.72, 1.28) | 0.18 |
| Madagascar | MAHAY (29) | ASQI | 1613 | 1604 | 1.17 (0.75, 1.83) | 0.14 |
| Malawi | iLiNS−DYADM (30) | CDI | 215 | 439 | 0.87 (0.53, 1.43) | 0.13 |
| Malawi | iLiNS−DOSE (31) | CDI | 645 | 221 | 0.95 (0.61, 1.50) | 0.14 |
| Zimbabwe | SHINE (HIV−) (33) | CDI | 381 | 373 | 1.05 (0.67, 1.65) | 0.14 |
| Zimbabwe | SHINE (HIV+) (34) | CDI | 66 | 68 | 1.29 (0.54, 3.06) | 0.07 |
| **I² = 0.67, Tau² = 0.07** | | | **4315** | **5153** | **0.88 (0.68, 1.14)** | |

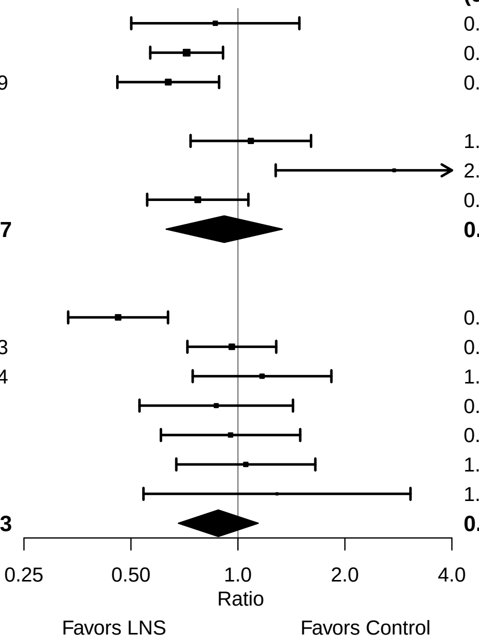

#### Supplemental figure 4B: Language lowest decile prevalence ratio

### 4B7: Stratified by Supplement duration

**Supplement duration**
**(p−diff = 0.545)**

| Country | Trial | Tool | N | N | PR (95% CI) | W |
|---|---|---|---|---|---|---|
| **Supplement duration − 12m or less** | | | | | | |
| Bangladesh | JiVitA−4 (21) | BSID−III | 445 | 143 | 0.86 (0.50, 1.49) | 0.10 |
| Burkina Faso | iLiNS−Zinc (24) | DMC | 746 | 375 | 0.46 (0.33, 0.64) | 0.15 |
| Ghana | GHANA (25) | | | | | |
| Ghana | iLiNS−DYADG (26) | CDI | 331 | 658 | 1.09 (0.74, 1.61) | 0.13 |
| Haiti | HAITI (27) | Total sounds | 150 | 149 | 2.75 (1.28, 5.93) | 0.07 |
| Madagascar | MAHAY (29) | ASQI | 1613 | 1604 | 1.17 (0.75, 1.83) | 0.12 |
| Malawi | iLiNS−DYADM (30) | CDI | 215 | 439 | 0.87 (0.53, 1.43) | 0.11 |
| Malawi | iLiNS−DOSE (31) | CDI | 645 | 221 | 0.95 (0.61, 1.50) | 0.12 |
| Zimbabwe | SHINE[1] (HIV−) (33), LNS vs SOC | CDI | 381 | 373 | 1.05 (0.67, 1.65) | 0.15 |
| Zimbabwe | LNS+WSH vs WSH | CDI | 436 | 408 | 0.77 (0.49, 1.20) | 0.15 |
| Zimbabwe | SHINE[1] (HIV+) (34), LNS vs SOC | CDI | 66 | 68 | 1.29 (0.54, 3.06) | 0.08 |
| Zimbabwe | LNS+WSH vs WSH | CDI | 99 | 79 | 0.32 (0.11, 0.92) | 0.08 |
| **$I^2$ = 0.70, $Tau^2$ = 0.13** | | | **5127** | **4517** | **0.92 (0.69, 1.23)** | |
| **Supplement duration − > 12m** | | | | | | |
| Bangladesh | RDNS (22) | CDI | 1663 | 814 | 0.72 (0.57, 0.91) | 0.25 |
| Bangladesh | WASH−B (23) | CDI | 1109 | 3353 | 0.81 (0.65, 1.00) | 0.26 |
| Kenya | WASH−B (28) | EASQ | 1362 | 4745 | 0.95 (0.78, 1.16) | 0.27 |
| Mali | PROMIS CS (32) | DMC | 927 | 944 | 0.77 (0.56, 1.07) | 0.22 |
| **$I^2$ = 0.16, $Tau^2$ = 0.00** | | | **5061** | **9856** | **0.82 (0.73, 0.93)** | |

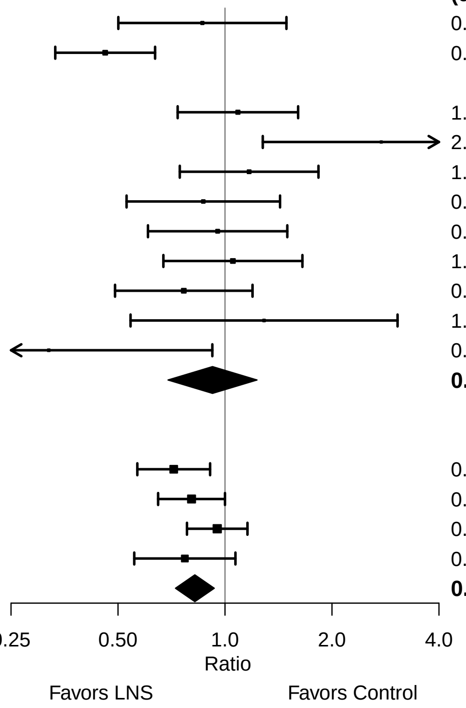

#### Supplemental figure 4B: Language lowest decile prevalence ratio

##### 4B8: Stratified by Frequency of contact

**Frequency of contact**
**(p−diff = 0.370)**

| Country | Trial | Tool | N | N | PR (95% CI) | W |
|---|---|---|---|---|---|---|
| **Frequency of contact − Monthly** | | | | | | |
| Bangladesh | RDNS (22) | CDI | 1663 | 814 | 0.72 (0.57, 0.91) | 0.19 |
| Haiti | HAITI (27) | Total sounds | 150 | 149 | 2.75 (1.28, 5.93) | 0.07 |
| Kenya | WASH−B (28) | EASQ | 1362 | 4745 | 0.95 (0.78, 1.16) | 0.20 |
| Madagascar | MAHAY (29) | ASQI | 1613 | 1604 | 1.17 (0.75, 1.83) | 0.13 |
| Mali | PROMIS CS (32) | DMC | 927 | 944 | 0.77 (0.56, 1.07) | 0.16 |
| Zimbabwe | SHINE[1] (HIV−) (33), LNS vs SOC | CDI | 381 | 373 | 1.05 (0.67, 1.65) | 0.16 |
| Zimbabwe | LNS+WSH vs WSH | CDI | 436 | 408 | 0.77 (0.49, 1.20) | 0.16 |
| Zimbabwe | SHINE[1] (HIV+) (34), LNS vs SOC | CDI | 66 | 68 | 1.29 (0.54, 3.06) | 0.08 |
| Zimbabwe | LNS+WSH vs WSH | CDI | 99 | 79 | 0.32 (0.11, 0.92) | 0.08 |
| **I² = 0.59, Tau² = 0.11** | | | **6697** | **9184** | **0.95 (0.71, 1.26)** | |
| **Frequency of contact − Weekly** | | | | | | |
| Bangladesh | JiVitA−4 (21) | BSID−III | 445 | 143 | 0.86 (0.50, 1.49) | 0.13 |
| Bangladesh | WASH−B (23) | CDI | 1109 | 3353 | 0.81 (0.65, 1.00) | 0.22 |
| Burkina Faso | iLiNS−Zinc (24) | DMC | 746 | 375 | 0.46 (0.33, 0.64) | 0.19 |
| Ghana | GHANA (25) | | | | | |
| Ghana | iLiNS−DYADG (26) | CDI | 331 | 658 | 1.09 (0.74, 1.61) | 0.17 |
| Malawi | iLiNS−DYADM (30) | CDI | 215 | 439 | 0.87 (0.53, 1.43) | 0.14 |
| Malawi | iLiNS−DOSE (31) | CDI | 645 | 221 | 0.95 (0.61, 1.50) | 0.15 |
| **I² = 0.65, Tau² = 0.06** | | | **3491** | **5189** | **0.79 (0.62, 1.02)** | |

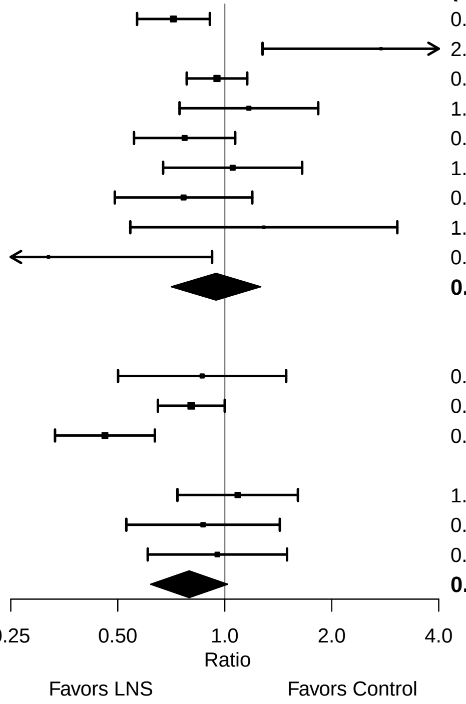

##### Supplemental figure 4B: Language lowest decile prevalence ratio

#### 4B9: Stratified by Average SQ-LNS compliance

**Average SQ−LNS compliance**
**(p−diff = 0.616)**

| Country | Trial | Tool | N | N | PR (95% CI) | W |
|---|---|---|---|---|---|---|
| **Average SQ−LNS compliance − Low** | | | | | | |
| Ghana | iLiNS−DYADG (26) | CDI | 331 | 658 | 1.09 (0.74, 1.61) | 0.18 |
| Malawi | iLiNS−DYADM (30) | CDI | 215 | 439 | 0.87 (0.53, 1.43) | 0.15 |
| Malawi | iLiNS−DOSE (31) | CDI | 645 | 221 | 0.95 (0.61, 1.50) | 0.16 |
| Mali | PROMIS CS (32) | DMC | 927 | 944 | 0.77 (0.56, 1.07) | 0.20 |
| Zimbabwe | SHINE[1] (HIV−) (33), LNS vs SOC | CDI | 381 | 373 | 1.05 (0.67, 1.65) | 0.20 |
| Zimbabwe | LNS+WSH vs WSH | CDI | 436 | 408 | 0.77 (0.49, 1.20) | 0.20 |
| Zimbabwe | SHINE[1] (HIV+) (34), LNS vs SOC | CDI | 66 | 68 | 1.29 (0.54, 3.06) | 0.11 |
| Zimbabwe | LNS+WSH vs WSH | CDI | 99 | 79 | 0.32 (0.11, 0.92) | 0.11 |
| **I² = 0.00, Tau² = 0.00** | | | **3100** | **3190** | **0.88 (0.75, 1.04)** | |
| **Average SQ−LNS compliance − High** | | | | | | |
| Bangladesh | JiVitA−4 (21) | BSID−III | 445 | 143 | 0.86 (0.50, 1.49) | 0.12 |
| Bangladesh | RDNS (22) | CDI | 1663 | 814 | 0.72 (0.57, 0.91) | 0.20 |
| Bangladesh | WASH−B (23) | CDI | 1109 | 3353 | 0.81 (0.65, 1.00) | 0.21 |
| Burkina Faso | iLiNS−Zinc (24) | DMC | 746 | 375 | 0.46 (0.33, 0.64) | 0.18 |
| Ghana | GHANA (25) | | | | | |
| Haiti | HAITI (27) | Total sounds | 150 | 149 | 2.75 (1.28, 5.93) | 0.08 |
| Kenya | WASH−B (28) | EASQ | 1362 | 4745 | 0.95 (0.78, 1.16) | 0.21 |
| **I² = 0.80, Tau² = 0.25** | | | **5475** | **9579** | **0.86 (0.56, 1.33)** | |

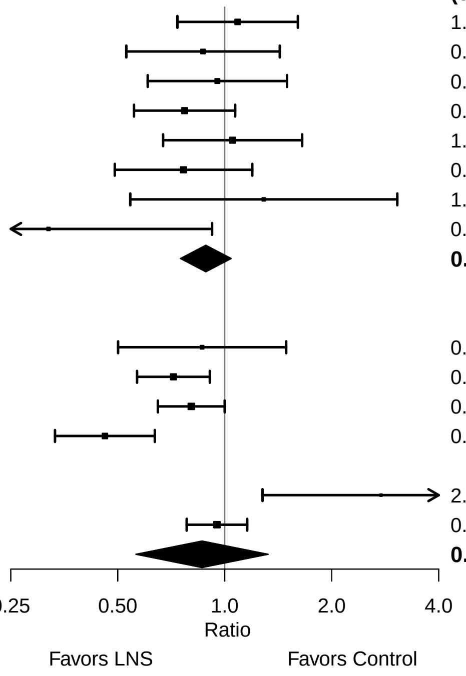

### Supplemental figure 4C: Language lowest decile prevalence difference

#### 4C1: Stratified by Geographic region

**Geographic region**
**(p−diff = 0.484)**

| Country | Trial | Tool | N | N | PD (95% CI) | W |
|---|---|---|---|---|---|---|
| **Geographic region − SEAR** | | | | | | |
| Bangladesh | JiVitA−4 (21) | BSID−III | 445 | 143 | −0.02 (−0.07, 0.04) | 0.16 |
| Bangladesh | RDNS (22) | CDI | 1663 | 814 | −0.04 (−0.06, −0.01) | 0.38 |
| Bangladesh | WASH−B (23) | CDI | 1109 | 3353 | −0.02 (−0.04, 0.00) | 0.46 |
| **$I^2$ = 0.00, $Tau^2$ = 0.00** | | | **3217** | **4310** | **−0.02 (−0.04, −0.01)** | |
| **Geographic region − AFR** | | | | | | |
| Burkina Faso | iLiNS−Zinc (24) | DMC | 746 | 375 | −0.08 (−0.13, −0.04) | 0.09 |
| Ghana | GHANA (25) | | | | | |
| Ghana | iLiNS−DYADG (26) | CDI | 331 | 658 | 0.01 (−0.03, 0.05) | 0.10 |
| Kenya | WASH−B (28) | EASQ | 1362 | 4745 | 0.00 (−0.02, 0.01) | 0.17 |
| Madagascar | MAHAY (29) | ASQI | 1613 | 1604 | 0.02 (−0.01, 0.04) | 0.16 |
| Malawi | iLiNS−DYADM (30) | CDI | 215 | 439 | −0.01 (−0.06, 0.03) | 0.08 |
| Malawi | iLiNS−DOSE (31) | CDI | 645 | 221 | 0.00 (−0.05, 0.04) | 0.08 |
| Mali | PROMIS CS (32) | DMC | 927 | 944 | −0.03 (−0.05, 0.00) | 0.14 |
| Zimbabwe | SHINE[1] (HIV−) (33), LNS vs SOC | CDI | 381 | 373 | 0.01 (−0.04, 0.05) | 0.13 |
| Zimbabwe | LNS+WSH vs WSH | CDI | 436 | 408 | −0.03 (−0.06, 0.01) | 0.13 |
| Zimbabwe | SHINE[1] (HIV+) (34), LNS vs SOC | CDI | 66 | 68 | 0.03 (−0.08, 0.15) | 0.05 |
| Zimbabwe | LNS+WSH vs WSH | CDI | 99 | 79 | −0.09 (−0.17, 0.00) | 0.05 |
| **$I^2$ = 0.63, $Tau^2$ = 0.00** | | | **6821** | **9914** | **−0.01 (−0.03, 0.00)** | |

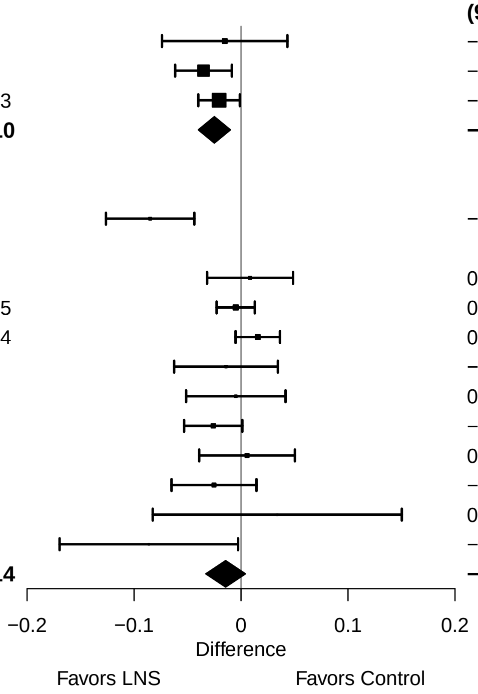

#### Supplemental figure 4C: Language lowest decile prevalence difference

### 4C2: Stratified by Stunting burden

**Stunting burden**
**(p−diff = 0.073)**

| Country | Trial | Tool | N | N | PD (95% CI) | W |
|---|---|---|---|---|---|---|
| **Stunting burden − Less than 35%** | | | | | | |
| Ghana | GHANA (25) | | | | | |
| Ghana | iLiNS−DYADG (26) | CDI | 331 | 658 | 0.01 (−0.03, 0.05) | 0.26 |
| Haiti | HAITI (27) | Total sounds | 150 | 149 | 0.10 (0.03, 0.16) | 0.16 |
| Kenya | WASH−B (28) | EASQ | 1362 | 4745 | 0.00 (−0.02, 0.01) | 0.35 |
| Malawi | iLiNS−DYADM (30) | CDI | 215 | 439 | −0.01 (−0.06, 0.03) | 0.23 |
| **I² = 0.65, Tau² = 0.00** | | | **2058** | **5991** | **0.02 (−0.03, 0.06)** | |
| **Stunting burden − More than 35%** | | | | | | |
| Bangladesh | JiVitA−4 (21) | BSID−III | 445 | 143 | −0.02 (−0.07, 0.04) | 0.08 |
| Bangladesh | RDNS (22) | CDI | 1663 | 814 | −0.04 (−0.06, −0.01) | 0.13 |
| Bangladesh | WASH−B (23) | CDI | 1109 | 3353 | −0.02 (−0.04, 0.00) | 0.14 |
| Burkina Faso | iLiNS−Zinc (24) | DMC | 746 | 375 | −0.08 (−0.13, −0.04) | 0.10 |
| Madagascar | MAHAY (29) | ASQI | 1613 | 1604 | 0.02 (−0.01, 0.04) | 0.14 |
| Malawi | iLiNS−DOSE (31) | CDI | 645 | 221 | 0.00 (−0.05, 0.04) | 0.10 |
| Mali | PROMIS CS (32) | DMC | 927 | 944 | −0.03 (−0.05, 0.00) | 0.13 |
| Zimbabwe | SHINE¹ (HIV−) (33), LNS vs SOC | CDI | 381 | 373 | 0.01 (−0.04, 0.05) | 0.12 |
| Zimbabwe | LNS+WSH vs WSH | CDI | 436 | 408 | −0.03 (−0.06, 0.01) | 0.12 |
| Zimbabwe | SHINE¹ (HIV+) (34), LNS vs SOC | CDI | 66 | 68 | 0.03 (−0.08, 0.15) | 0.07 |
| Zimbabwe | LNS+WSH vs WSH | CDI | 99 | 79 | −0.09 (−0.17, 0.00) | 0.07 |
| **I² = 0.65, Tau² = 0.00** | | | **8130** | **8382** | **−0.02 (−0.04, 0.00)** | |

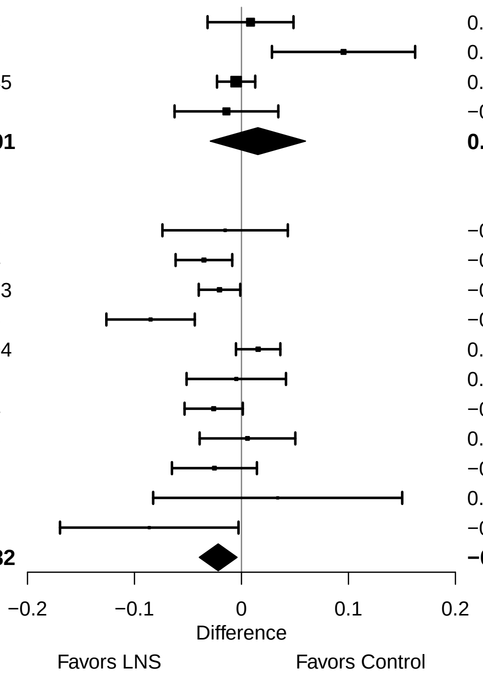

#### Supplemental figure 4C: Language lowest decile prevalence difference

### 4C3: Stratified by Malaria prevalence

**Malaria prevalence**
**(p−diff = 0.289)**

| Country | Trial | Tool | N | N | PD (95% CI) | W |
|---|---|---|---|---|---|---|
| **Malaria prevalence − Less than 10%** | | | | | | |
| Bangladesh | JiVitA−4 (21) | BSID−III | 445 | 143 | −0.02 (−0.07, 0.04) | 0.09 |
| Bangladesh | RDNS (22) | CDI | 1663 | 814 | −0.04 (−0.06, −0.01) | 0.15 |
| Bangladesh | WASH−B (23) | CDI | 1109 | 3353 | −0.02 (−0.04, 0.00) | 0.16 |
| Haiti | HAITI (27) | Total sounds | 150 | 149 | 0.10 (0.03, 0.16) | 0.08 |
| Kenya | WASH−B (28) | EASQ | 1362 | 4745 | 0.00 (−0.02, 0.01) | 0.16 |
| Madagascar | MAHAY (29) | ASQI | 1613 | 1604 | 0.02 (−0.01, 0.04) | 0.16 |
| Zimbabwe | SHINE[1] (HIV−) (33), LNS vs SOC | CDI | 381 | 373 | 0.01 (−0.04, 0.05) | 0.14 |
| Zimbabwe | LNS+WSH vs WSH | CDI | 436 | 408 | −0.03 (−0.06, 0.01) | 0.14 |
| Zimbabwe | SHINE[1] (HIV+) (34), LNS vs SOC | CDI | 66 | 68 | 0.03 (−0.08, 0.15) | 0.07 |
| Zimbabwe | LNS+WSH vs WSH | CDI | 99 | 79 | −0.09 (−0.17, 0.00) | 0.07 |
| **$I^2$ = 0.66, $Tau^2$ = 0.00** | | | **7324** | **11736** | **−0.01 (−0.03, 0.02)** | |
| **Malaria prevalence − At least 10%** | | | | | | |
| Burkina Faso | iLiNS−Zinc (24) | DMC | 746 | 375 | −0.08 (−0.13, −0.04) | 0.20 |
| Ghana | GHANA (25) | | | | | |
| Ghana | iLiNS−DYADG (26) | CDI | 331 | 658 | 0.01 (−0.03, 0.05) | 0.20 |
| Malawi | iLiNS−DYADM (30) | CDI | 215 | 439 | −0.01 (−0.06, 0.03) | 0.18 |
| Malawi | iLiNS−DOSE (31) | CDI | 645 | 221 | 0.00 (−0.05, 0.04) | 0.18 |
| Mali | PROMIS CS (32) | DMC | 927 | 944 | −0.03 (−0.05, 0.00) | 0.24 |
| **$I^2$ = 0.66, $Tau^2$ = 0.00** | | | **2864** | **2637** | **−0.02 (−0.06, 0.01)** | |

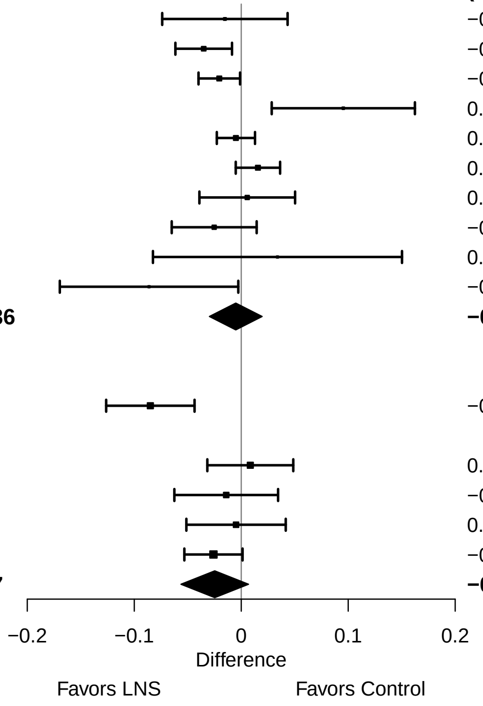

#### Supplemental figure 4C: Language lowest decile prevalence difference

### 4C4: Stratified by Anemia prevalence

**Anemia prevalence**
**(p−diff = 0.762)**

| Country | Trial | Tool | N | N | PD (95% CI) | W |
|---|---|---|---|---|---|---|
| **Anemia prevalence − High** | | | | | | |
| Burkina Faso | iLiNS−Zinc (24) | DMC | 746 | 375 | −0.08 (−0.13, −0.04) | 0.21 |
| Ghana | GHANA (25) | | | | | |
| Haiti | HAITI (27) | Total sounds | 150 | 149 | 0.10 (0.03, 0.16) | 0.14 |
| Malawi | iLiNS−DYADM (30) | CDI | 215 | 439 | −0.01 (−0.06, 0.03) | 0.19 |
| Malawi | iLiNS−DOSE (31) | CDI | 645 | 221 | 0.00 (−0.05, 0.04) | 0.20 |
| Mali | PROMIS CS (32) | DMC | 927 | 944 | −0.03 (−0.05, 0.00) | 0.26 |
| **I² = 0.81, Tau² = 0.00** | | | **2683** | **2128** | **−0.01 (−0.07, 0.04)** | |
| **Anemia prevalence − Moderate** | | | | | | |
| Bangladesh | JiVitA−4 (21) | BSID−III | 445 | 143 | −0.02 (−0.07, 0.04) | 0.08 |
| Bangladesh | RDNS (22) | CDI | 1663 | 814 | −0.04 (−0.06, −0.01) | 0.14 |
| Bangladesh | WASH−B (23) | CDI | 1109 | 3353 | −0.02 (−0.04, 0.00) | 0.15 |
| Ghana | iLiNS−DYADG (26) | CDI | 331 | 658 | 0.01 (−0.03, 0.05) | 0.11 |
| Kenya | WASH−B (28) | EASQ | 1362 | 4745 | 0.00 (−0.02, 0.01) | 0.15 |
| Madagascar | MAHAY (29) | ASQI | 1613 | 1604 | 0.02 (−0.01, 0.04) | 0.15 |
| Zimbabwe | SHINE[1] (HIV−) (33), LNS vs SOC | CDI | 381 | 373 | 0.01 (−0.04, 0.05) | 0.13 |
| Zimbabwe | LNS+WSH vs WSH | CDI | 436 | 408 | −0.03 (−0.06, 0.01) | 0.13 |
| Zimbabwe | SHINE[1] (HIV+) (34), LNS vs SOC | CDI | 66 | 68 | 0.03 (−0.08, 0.15) | 0.07 |
| Zimbabwe | LNS+WSH vs WSH | CDI | 99 | 79 | −0.09 (−0.17, 0.00) | 0.07 |
| **I² = 0.42, Tau² = 0.00** | | | **7505** | **12245** | **−0.01 (−0.02, 0.00)** | |

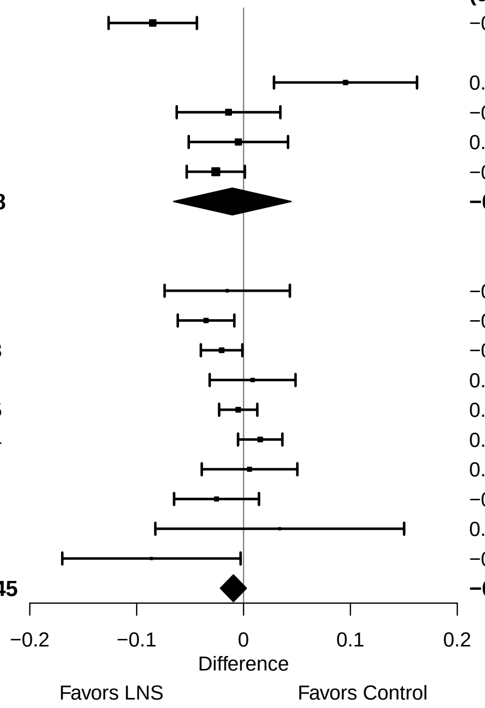

#### Supplemental figure 4C: Language lowest decile prevalence difference

### 4C5: Stratified by Source water quality

**Source water quality**
**(p−diff = 0.748)**

| Country | Trial | Tool | N | N | PD (95% CI) | W |
|---|---|---|---|---|---|---|
| **Source water quality − Improved** | | | | | | |
| Bangladesh | JiVitA−4 (21) | BSID−III | 445 | 143 | −0.02 (−0.07, 0.04) | 0.11 |
| Bangladesh | RDNS (22) | CDI | 1663 | 814 | −0.04 (−0.06, −0.01) | 0.18 |
| Bangladesh | WASH−B (23) | CDI | 548 | 1099 | −0.04 (−0.07, −0.01) | 0.18 |
| Ghana | GHANA (25) | | | | | |
| Ghana | iLiNS−DYADG (26) | CDI | 331 | 658 | 0.01 (−0.03, 0.05) | 0.15 |
| Haiti | HAITI (27) | Total sounds | 150 | 149 | 0.10 (0.03, 0.16) | 0.10 |
| Malawi | iLiNS−DYADM (30) | CDI | 215 | 439 | −0.01 (−0.06, 0.03) | 0.13 |
| Malawi | iLiNS−DOSE (31) | CDI | 645 | 221 | 0.00 (−0.05, 0.04) | 0.14 |
| **I² = 0.65, Tau² = 0.00** | | | **3997** | **3523** | **−0.01 (−0.04, 0.02)** | |
| **Source water quality − Unimproved** | | | | | | |
| Burkina Faso | iLiNS−Zinc (24) | DMC | 746 | 375 | −0.08 (−0.13, −0.04) | 0.17 |
| Kenya | WASH−B (28) | EASQ | 649 | 2073 | 0.00 (−0.03, 0.02) | 0.20 |
| Madagascar | MAHAY (29) | ASQI | 1613 | 1604 | 0.02 (−0.01, 0.04) | 0.22 |
| Mali | PROMIS CS (32) | DMC | 927 | 944 | −0.03 (−0.05, 0.00) | 0.20 |
| Zimbabwe | SHINE (HIV−) (33) | CDI | 381 | 373 | 0.01 (−0.04, 0.05) | 0.16 |
| Zimbabwe | SHINE (HIV+) (34) | CDI | 66 | 68 | 0.03 (−0.08, 0.15) | 0.05 |
| **I² = 0.76, Tau² = 0.00** | | | **4382** | **5437** | **−0.01 (−0.05, 0.02)** | |

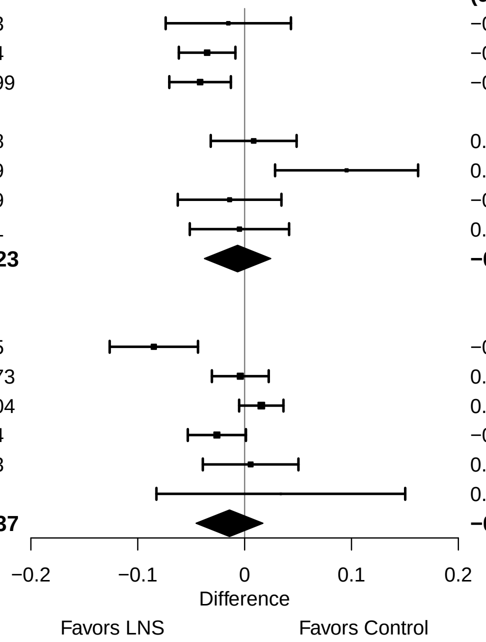

#### Supplemental figure 4C: Language lowest decile prevalence difference

### 4C6: Stratified by Sanitation

**Sanitation**
**(p−diff = 0.989)**

| Country | Trial | Tool | N | N | PD (95% CI) | W |
|---|---|---|---|---|---|---|
| **Sanitation − Improved** | | | | | | |
| Bangladesh | JiVitA−4 (21) | BSID−III | 445 | 143 | −0.02 (−0.07, 0.04) | 0.13 |
| Bangladesh | RDNS (22) | CDI | 1663 | 814 | −0.04 (−0.06, −0.01) | 0.20 |
| Bangladesh | WASH−B (23) | CDI | 548 | 1099 | −0.04 (−0.07, −0.01) | 0.20 |
| Ghana | GHANA (25) | | | | | |
| Ghana | iLiNS−DYADG (26) | CDI | 331 | 658 | 0.01 (−0.03, 0.05) | 0.17 |
| Haiti | HAITI (27) | Total sounds | 150 | 149 | 0.10 (0.03, 0.16) | 0.11 |
| Mali | PROMIS CS (32) | DMC | 927 | 944 | −0.03 (−0.05, 0.00) | 0.20 |
| **I² = 0.70, Tau² = 0.00** | | | **4064** | **3807** | **−0.01 (−0.05, 0.03)** | |
| **Sanitation − Unimproved** | | | | | | |
| Burkina Faso | iLiNS−Zinc (24) | DMC | 746 | 375 | −0.08 (−0.13, −0.04) | 0.15 |
| Kenya | WASH−B (28) | EASQ | 649 | 2073 | 0.00 (−0.03, 0.02) | 0.18 |
| Madagascar | MAHAY (29) | ASQI | 1613 | 1604 | 0.02 (−0.01, 0.04) | 0.20 |
| Malawi | iLiNS−DYADM (30) | CDI | 215 | 439 | −0.01 (−0.06, 0.03) | 0.14 |
| Malawi | iLiNS−DOSE (31) | CDI | 645 | 221 | 0.00 (−0.05, 0.04) | 0.14 |
| Zimbabwe | SHINE (HIV−) (33) | CDI | 381 | 373 | 0.01 (−0.04, 0.05) | 0.14 |
| Zimbabwe | SHINE (HIV+) (34) | CDI | 66 | 68 | 0.03 (−0.08, 0.15) | 0.05 |
| **I² = 0.68, Tau² = 0.00** | | | **4315** | **5153** | **−0.01 (−0.04, 0.02)** | |

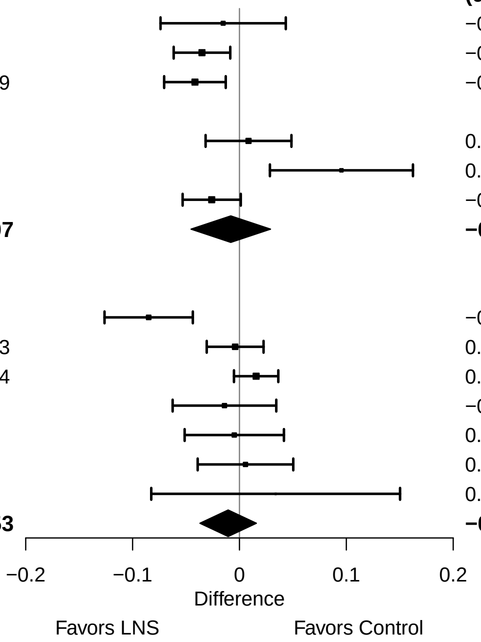

#### Supplemental figure 4C: Language lowest decile prevalence difference

### 4C7: Stratified by Supplement duration

**Supplement duration**
**(p−diff = 0.443)**

| Country | Trial | | Tool | N | N | PD (95% CI) | W |
|---|---|---|---|---|---|---|---|
| **Supplement duration − 12m or less** | | | | | | | |
| Bangladesh | JiVitA−4 (21) | | BSID−III | 445 | 143 | −0.02 (−0.07, 0.04) | 0.09 |
| Burkina Faso | iLiNS−Zinc (24) | | DMC | 746 | 375 | −0.08 (−0.13, −0.04) | 0.12 |
| Ghana | GHANA (25) | | | | | | |
| Ghana | iLiNS−DYADG (26) | | CDI | 331 | 658 | 0.01 (−0.03, 0.05) | 0.12 |
| Haiti | HAITI (27) | | Total sounds | 150 | 149 | 0.10 (0.03, 0.16) | 0.08 |
| Madagascar | MAHAY (29) | | ASQI | 1613 | 1604 | 0.02 (−0.01, 0.04) | 0.16 |
| Malawi | iLiNS−DYADM (30) | | CDI | 215 | 439 | −0.01 (−0.06, 0.03) | 0.11 |
| Malawi | iLiNS−DOSE (31) | | CDI | 645 | 221 | 0.00 (−0.05, 0.04) | 0.11 |
| Zimbabwe | SHINE¹ (HIV−) (33), | LNS vs SOC | CDI | 381 | 373 | 0.01 (−0.04, 0.05) | 0.14 |
| Zimbabwe | | LNS+WSH vs WSH | CDI | 436 | 408 | −0.03 (−0.06, 0.01) | 0.14 |
| Zimbabwe | SHINE¹ (HIV+) (34), | LNS vs SOC | CDI | 66 | 68 | 0.03 (−0.08, 0.15) | 0.08 |
| Zimbabwe | | LNS+WSH vs WSH | CDI | 99 | 79 | −0.09 (−0.17, 0.00) | 0.08 |
| **I² = 0.72, Tau² = 0.00** | | | | **5127** | **4517** | **−0.01 (−0.04, 0.02)** | |
| **Supplement duration − > 12m** | | | | | | | |
| Bangladesh | RDNS (22) | | CDI | 1663 | 814 | −0.04 (−0.06, −0.01) | 0.24 |
| Bangladesh | WASH−B (23) | | CDI | 1109 | 3353 | −0.02 (−0.04, 0.00) | 0.26 |
| Kenya | WASH−B (28) | | EASQ | 1362 | 4745 | 0.00 (−0.02, 0.01) | 0.26 |
| Mali | PROMIS CS (32) | | DMC | 927 | 944 | −0.03 (−0.05, 0.00) | 0.24 |
| **I² = 0.26, Tau² = 0.00** | | | | **5061** | **9856** | **−0.02 (−0.03, −0.01)** | |

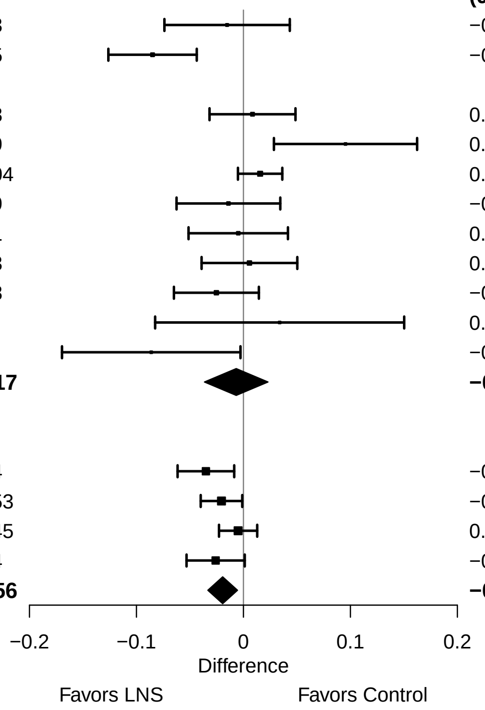

#### Supplemental figure 4C: Language lowest decile prevalence difference

### 4C8: Stratified by Frequency of contact

**Frequency of contact**
**(p−diff = 0.340)**

| Country | Trial | Tool | N | N | PD (95% CI) | W |
|---|---|---|---|---|---|---|
| **Frequency of contact − Monthly** | | | | | | |
| Bangladesh | RDNS (22) | CDI | 1663 | 814 | −0.04 (−0.06, −0.01) | 0.16 |
| Haiti | HAITI (27) | Total sounds | 150 | 149 | 0.10 (0.03, 0.16) | 0.08 |
| Kenya | WASH−B (28) | EASQ | 1362 | 4745 | 0.00 (−0.02, 0.01) | 0.18 |
| Madagascar | MAHAY (29) | ASQI | 1613 | 1604 | 0.02 (−0.01, 0.04) | 0.17 |
| Mali | PROMIS CS (32) | DMC | 927 | 944 | −0.03 (−0.05, 0.00) | 0.16 |
| Zimbabwe | SHINE[1] (HIV−) (33), LNS vs SOC | CDI | 381 | 373 | 0.01 (−0.04, 0.05) | 0.16 |
| Zimbabwe | LNS+WSH vs WSH | CDI | 436 | 408 | −0.03 (−0.06, 0.01) | 0.16 |
| Zimbabwe | SHINE[1] (HIV+) (34), LNS vs SOC | CDI | 66 | 68 | 0.03 (−0.08, 0.15) | 0.08 |
| Zimbabwe | LNS+WSH vs WSH | CDI | 99 | 79 | −0.09 (−0.17, 0.00) | 0.08 |
| **$I^2$ = 0.71, $Tau^2$ = 0.00** | | | **6697** | **9184** | **0.00 (−0.03, 0.03)** | |
| **Frequency of contact − Weekly** | | | | | | |
| Bangladesh | JiVitA−4 (21) | BSID−III | 445 | 143 | −0.02 (−0.07, 0.04) | 0.13 |
| Bangladesh | WASH−B (23) | CDI | 1109 | 3353 | −0.02 (−0.04, 0.00) | 0.23 |
| Burkina Faso | iLiNS−Zinc (24) | DMC | 746 | 375 | −0.08 (−0.13, −0.04) | 0.17 |
| Ghana | GHANA (25) | | | | | |
| Ghana | iLiNS−DYADG (26) | CDI | 331 | 658 | 0.01 (−0.03, 0.05) | 0.17 |
| Malawi | iLiNS−DYADM (30) | CDI | 215 | 439 | −0.01 (−0.06, 0.03) | 0.15 |
| Malawi | iLiNS−DOSE (31) | CDI | 645 | 221 | 0.00 (−0.05, 0.04) | 0.15 |
| **$I^2$ = 0.58, $Tau^2$ = 0.00** | | | **3491** | **5189** | **−0.02 (−0.05, 0.00)** | |

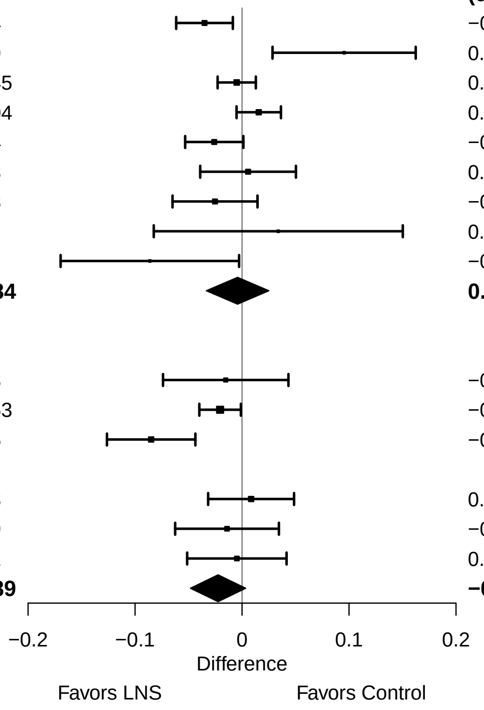

#### Supplemental figure 4C: Language lowest decile prevalence difference

### 4C9: Stratified by Average SQ-LNS compliance

**Average SQ−LNS compliance**
**(p−diff = 0.770)**

| Country | Trial | | Tool | N | N | PD (95% CI) | W |
|---|---|---|---|---|---|---|---|
| **Average SQ−LNS compliance − Low** | | | | | | | |
| Ghana | iLiNS−DYADG (26) | | CDI | 331 | 658 | 0.01 (−0.03, 0.05) | 0.17 |
| Malawi | iLiNS−DYADM (30) | | CDI | 215 | 439 | −0.01 (−0.06, 0.03) | 0.15 |
| Malawi | iLiNS−DOSE (31) | | CDI | 645 | 221 | 0.00 (−0.05, 0.04) | 0.16 |
| Mali | PROMIS CS (32) | | DMC | 927 | 944 | −0.03 (−0.05, 0.00) | 0.21 |
| Zimbabwe | SHINE[1] (HIV−) (33), | LNS vs SOC | CDI | 381 | 373 | 0.01 (−0.04, 0.05) | 0.20 |
| Zimbabwe | | LNS+WSH vs WSH | CDI | 436 | 408 | −0.03 (−0.06, 0.01) | 0.20 |
| Zimbabwe | SHINE[1] (HIV+) (34), | LNS vs SOC | CDI | 66 | 68 | 0.03 (−0.08, 0.15) | 0.11 |
| Zimbabwe | | LNS+WSH vs WSH | CDI | 99 | 79 | −0.09 (−0.17, 0.00) | 0.11 |
| **$I^2$ = 0.00, $Tau^2$ = 0.00** | | | | **3100** | **3190** | **−0.01 (−0.03, 0.00)** | |
| **Average SQ−LNS compliance − High** | | | | | | | |
| Bangladesh | JiVitA−4 (21) | | BSID−III | 445 | 143 | −0.02 (−0.07, 0.04) | 0.12 |
| Bangladesh | RDNS (22) | | CDI | 1663 | 814 | −0.04 (−0.06, −0.01) | 0.20 |
| Bangladesh | WASH−B (23) | | CDI | 1109 | 3353 | −0.02 (−0.04, 0.00) | 0.21 |
| Burkina Faso | iLiNS−Zinc (24) | | DMC | 746 | 375 | −0.08 (−0.13, −0.04) | 0.16 |
| Ghana | GHANA (25) | | | | | | |
| Haiti | HAITI (27) | | Total sounds | 150 | 149 | 0.10 (0.03, 0.16) | 0.10 |
| Kenya | WASH−B (28) | | EASQ | 1362 | 4745 | 0.00 (−0.02, 0.01) | 0.22 |
| **$I^2$ = 0.80, $Tau^2$ = 0.00** | | | | **5475** | **9579** | **−0.01 (−0.06, 0.03)** | |

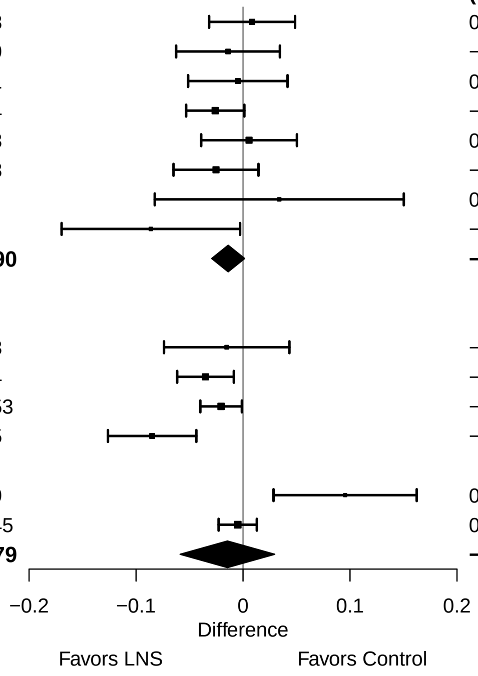

#### Supplemental figure 4D: Mean difference in social-emotional z-score

##### 4D1: Stratified by Geographic region (insufficient comparisons)

#### Supplemental figure 4D: Mean difference in social-emotional z-score

### 4D2: Stratified by Stunting burden

**Stunting burden**
**(p−diff = 0.066)**

| Country | Trial | Tool | N | N | MD (95% CI) | W |
|---|---|---|---|---|---|---|
| **Stunting burden − Less than 35%** | | | | | | |
| Ghana | GHANA (25) | | | | | |
| Ghana | iLiNS−DYADG (26) | PSED | 332 | 657 | 0.01 (−0.12, 0.14) | 0.32 |
| Haiti | HAITI (27) | | | | | |
| Kenya | WASH−B (28) | EASQ | 1362 | 4745 | 0.01 (−0.05, 0.08) | 0.41 |
| Malawi | iLiNS−DYADM (30) | PSED | 215 | 438 | 0.01 (−0.15, 0.18) | 0.28 |
| **I² = 0.00, Tau² = 0.00** | | | **1909** | **5840** | **0.01 (−0.04, 0.07)** | |
| **Stunting burden − More than 35%** | | | | | | |
| Bangladesh | JiVitA−4 (21) | | | | | |
| Bangladesh | RDNS (22) | DMC | 1657 | 815 | 0.06 (−0.04, 0.16) | 0.15 |
| Bangladesh | WASH−B (23) | EASQ | 1066 | 3266 | 0.12 (0.05, 0.20) | 0.16 |
| Burkina Faso | iLiNS−Zinc (24) | DMC | 746 | 375 | 0.36 (0.18, 0.53) | 0.11 |
| Madagascar | MAHAY (29) | ASQI | 1613 | 1604 | −0.07 (−0.26, 0.11) | 0.10 |
| Malawi | iLiNS−DOSE (31) | PSED | 644 | 221 | 0.03 (−0.12, 0.18) | 0.12 |
| Mali | PROMIS CS (32) | DMC | 927 | 944 | 0.15 (−0.02, 0.31) | 0.11 |
| Zimbabwe | SHINE¹ (HIV−) (33), LNS vs SOC | MDAT | 395 | 382 | 0.07 (−0.07, 0.21) | 0.15 |
| Zimbabwe | LNS+WSH vs WSH | MDAT | 446 | 417 | 0.17 (0.02, 0.31) | 0.15 |
| Zimbabwe | SHINE¹ (HIV+) (34), LNS vs SOC | MDAT | 67 | 68 | 0.24 (0.00, 0.48) | 0.10 |
| Zimbabwe | LNS+WSH vs WSH | MDAT | 103 | 83 | 0.44 (0.14, 0.73) | 0.10 |
| **I² = 0.65, Tau² = 0.01** | | | **7664** | **8175** | **0.13 (0.04, 0.23)** | |

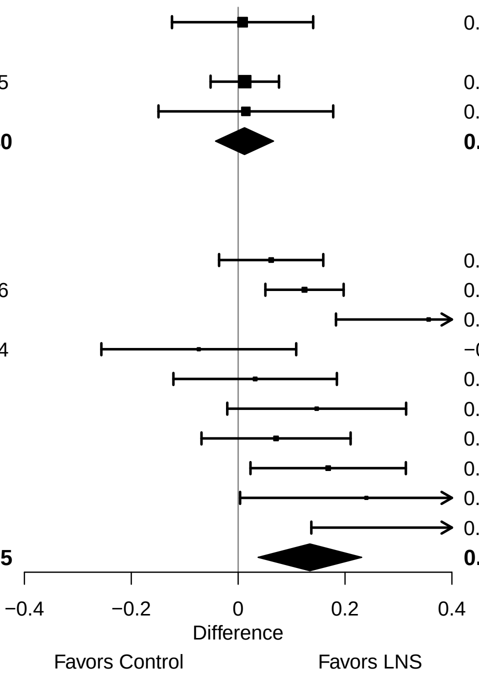

#### Supplemental figure 4D: Mean difference in social-emotional z-score

### 4D3: Stratified by Malaria prevalence

**Malaria prevalence**
**(p−diff = 0.863)**

| Country | Trial | | Tool | N | N | MD (95% CI) | W |
|---|---|---|---|---|---|---|---|
| **Malaria prevalence − Less than 10%** | | | | | | | |
| Bangladesh | JiVitA−4 (21) | | | | | | |
| Bangladesh | RDNS (22) | | DMC | 1657 | 815 | 0.06 (−0.04, 0.16) | 0.18 |
| Bangladesh | WASH−B (23) | | EASQ | 1066 | 3266 | 0.12 (0.05, 0.20) | 0.20 |
| Haiti | HAITI (27) | | | | | | |
| Kenya | WASH−B (28) | | EASQ | 1362 | 4745 | 0.01 (−0.05, 0.08) | 0.20 |
| Madagascar | MAHAY (29) | | ASQI | 1613 | 1604 | −0.07 (−0.26, 0.11) | 0.13 |
| Zimbabwe | SHINE[1] (HIV−) (33), | LNS vs SOC | MDAT | 395 | 382 | 0.07 (−0.07, 0.21) | 0.18 |
| Zimbabwe | | LNS+WSH vs WSH | MDAT | 446 | 417 | 0.17 (0.02, 0.31) | 0.18 |
| Zimbabwe | SHINE[1] (HIV+) (34), | LNS vs SOC | MDAT | 67 | 68 | 0.24 (0.00, 0.48) | 0.12 |
| Zimbabwe | | LNS+WSH vs WSH | MDAT | 103 | 83 | 0.44 (0.14, 0.73) | 0.12 |
| **$I^2$ = 0.70, $Tau^2$ = 0.01** | | | | **6709** | **11380** | **0.09 (−0.01, 0.20)** | |
| **Malaria prevalence − At least 10%** | | | | | | | |
| Burkina Faso | iLiNS−Zinc (24) | | DMC | 746 | 375 | 0.36 (0.18, 0.53) | 0.19 |
| Ghana | GHANA (25) | | | | | | |
| Ghana | iLiNS−DYADG (26) | | PSED | 332 | 657 | 0.01 (−0.12, 0.14) | 0.22 |
| Malawi | iLiNS−DYADM (30) | | PSED | 215 | 438 | 0.01 (−0.15, 0.18) | 0.19 |
| Malawi | iLiNS−DOSE (31) | | PSED | 644 | 221 | 0.03 (−0.12, 0.18) | 0.20 |
| Mali | PROMIS CS (32) | | DMC | 927 | 944 | 0.15 (−0.02, 0.31) | 0.19 |
| **$I^2$ = 0.68, $Tau^2$ = 0.01** | | | | **2864** | **2635** | **0.11 (−0.02, 0.23)** | |

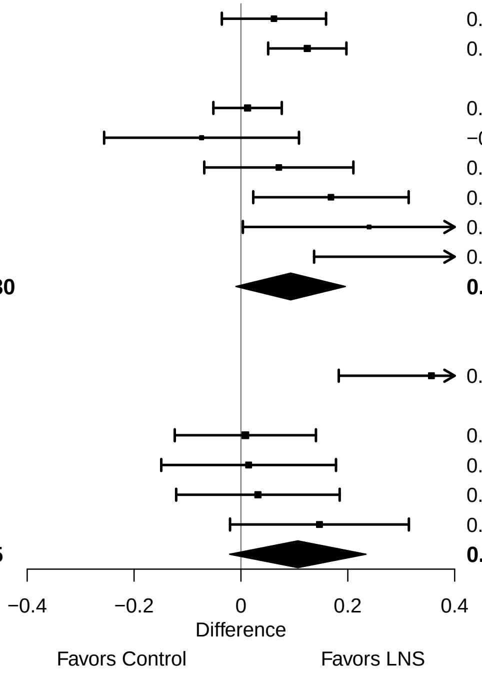

#### Supplemental figure 4D: Mean difference in social-emotional z-score

### 4D4: Stratified by Anemia prevalence

**Anemia prevalence**
**(p−diff = 0.496)**

| Country | Trial | Tool | N | N | MD (95% CI) | W |
|---|---|---|---|---|---|---|
| **Anemia prevalence − High** | | | | | | |
| Burkina Faso | iLiNS−Zinc (24) | DMC | 746 | 375 | 0.36 (0.18, 0.53) | 0.24 |
| Ghana | GHANA (25) | | | | | |
| Haiti | HAITI (27) | | | | | |
| Malawi | iLiNS−DYADM (30) | PSED | 215 | 438 | 0.01 (−0.15, 0.18) | 0.25 |
| Malawi | iLiNS−DOSE (31) | PSED | 644 | 221 | 0.03 (−0.12, 0.18) | 0.26 |
| Mali | PROMIS CS (32) | DMC | 927 | 944 | 0.15 (−0.02, 0.31) | 0.25 |
| **I² = 0.70, Tau² = 0.02** | | | **2532** | **1978** | **0.13 (−0.02, 0.29)** | |
| **Anemia prevalence − Moderate** | | | | | | |
| Bangladesh | JiVitA−4 (21) | | | | | |
| Bangladesh | RDNS (22) | DMC | 1657 | 815 | 0.06 (−0.04, 0.16) | 0.16 |
| Bangladesh | WASH−B (23) | EASQ | 1066 | 3266 | 0.12 (0.05, 0.20) | 0.17 |
| Ghana | iLiNS−DYADG (26) | PSED | 332 | 657 | 0.01 (−0.12, 0.14) | 0.14 |
| Kenya | WASH−B (28) | EASQ | 1362 | 4745 | 0.01 (−0.05, 0.08) | 0.17 |
| Madagascar | MAHAY (29) | ASQI | 1613 | 1604 | −0.07 (−0.26, 0.11) | 0.11 |
| Zimbabwe | SHINE¹ (HIV−) (33), LNS vs SOC | MDAT | 395 | 382 | 0.07 (−0.07, 0.21) | 0.15 |
| Zimbabwe | LNS+WSH vs WSH | MDAT | 446 | 417 | 0.17 (0.02, 0.31) | 0.15 |
| Zimbabwe | SHINE¹ (HIV+) (34), LNS vs SOC | MDAT | 67 | 68 | 0.24 (0.00, 0.48) | 0.10 |
| Zimbabwe | LNS+WSH vs WSH | MDAT | 103 | 83 | 0.44 (0.14, 0.73) | 0.10 |
| **I² = 0.66, Tau² = 0.01** | | | **7041** | **12037** | **0.08 (−0.01, 0.17)** | |

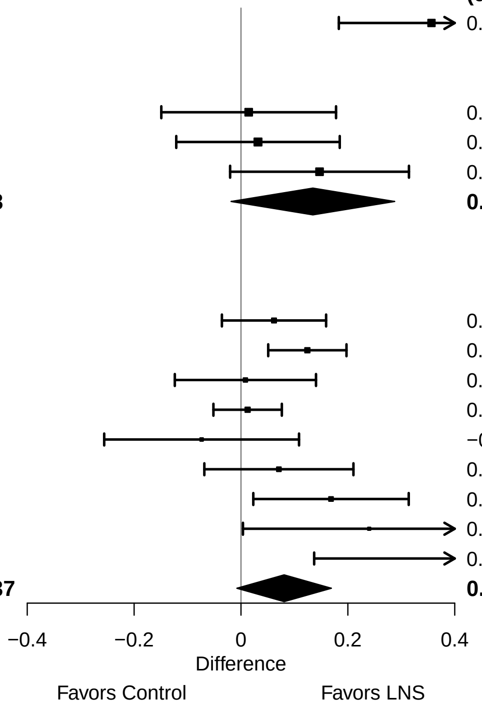

#### Supplemental figure 4D: Mean difference in social-emotional z-score

### 4D5: Stratified by Source water quality

**Source water quality**
**(p−diff = 0.681)**

**Source water quality − Improved**

| **Country** | **Trial** | **Tool** | **N** | **N** | **MD (95% CI)** | **W** |
|---|---|---|---|---|---|---|
| Bangladesh | JiVitA−4 (21) | | | | | |
| Bangladesh | RDNS (22) | DMC | 1657 | 815 | 0.06 (−0.04, 0.16) | 0.23 |
| Bangladesh | WASH−B (23) | EASQ | 528 | 1099 | 0.22 (0.11, 0.33) | 0.22 |
| Ghana | GHANA (25) | | | | | |
| Ghana | iLiNS−DYADG (26) | PSED | 332 | 657 | 0.01 (−0.12, 0.14) | 0.20 |
| Haiti | HAITI (27) | | | | | |
| Malawi | iLiNS−DYADM (30) | PSED | 215 | 438 | 0.01 (−0.15, 0.18) | 0.17 |
| Malawi | iLiNS−DOSE (31) | PSED | 644 | 221 | 0.03 (−0.12, 0.18) | 0.18 |
| **I² = 0.54, Tau² = 0.00** | | | **3376** | **3230** | **0.08 (0.00, 0.16)** | |

**Source water quality − Unimproved**

| **Country** | **Trial** | **Tool** | **N** | **N** | **MD (95% CI)** | **W** |
|---|---|---|---|---|---|---|
| Burkina Faso | iLiNS−Zinc (24) | DMC | 746 | 375 | 0.36 (0.18, 0.53) | 0.16 |
| Kenya | WASH−B (28) | EASQ | 649 | 2073 | −0.03 (−0.12, 0.07) | 0.23 |
| Madagascar | MAHAY (29) | ASQI | 1613 | 1604 | −0.07 (−0.26, 0.11) | 0.15 |
| Mali | PROMIS CS (32) | DMC | 927 | 944 | 0.15 (−0.02, 0.31) | 0.16 |
| Zimbabwe | SHINE (HIV−) (33) | MDAT | 395 | 382 | 0.07 (−0.07, 0.21) | 0.19 |
| Zimbabwe | SHINE (HIV+) (34) | MDAT | 67 | 68 | 0.24 (0.00, 0.48) | 0.12 |
| **I² = 0.74, Tau² = 0.02** | | | **4397** | **5446** | **0.11 (−0.02, 0.24)** | |

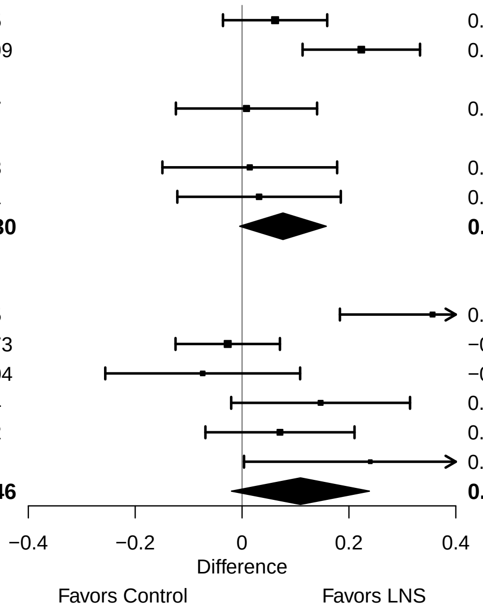

#### Supplemental figure 4D: Mean difference in social-emotional z-score

### 4D6: Stratified by Sanitation

**Sanitation**
**(p−diff = 0.660)**

| Country | Trial | Tool | N | N | MD (95% CI) | W |
|---|---|---|---|---|---|---|
| **Sanitation − Improved** | | | | | | |
| Bangladesh | JiVitA−4 (21) | | | | | |
| Bangladesh | RDNS (22) | DMC | 1657 | 815 | 0.06 (−0.04, 0.16) | 0.28 |
| Bangladesh | WASH−B (23) | EASQ | 528 | 1099 | 0.22 (0.11, 0.33) | 0.27 |
| Ghana | GHANA (25) | | | | | |
| Ghana | iLiNS−DYADG (26) | PSED | 332 | 657 | 0.01 (−0.12, 0.14) | 0.24 |
| Haiti | HAITI (27) | | | | | |
| Mali | PROMIS CS (32) | DMC | 927 | 944 | 0.15 (−0.02, 0.31) | 0.20 |
| **I² = 0.60, Tau² = 0.01** | | | **3444** | **3515** | **0.11 (0.02, 0.20)** | |
| **Sanitation − Unimproved** | | | | | | |
| Burkina Faso | iLiNS−Zinc (24) | DMC | 746 | 375 | 0.36 (0.18, 0.53) | 0.13 |
| Kenya | WASH−B (28) | EASQ | 649 | 2073 | −0.03 (−0.12, 0.07) | 0.19 |
| Madagascar | MAHAY (29) | ASQI | 1613 | 1604 | −0.07 (−0.26, 0.11) | 0.13 |
| Malawi | iLiNS−DYADM (30) | PSED | 215 | 438 | 0.01 (−0.15, 0.18) | 0.14 |
| Malawi | iLiNS−DOSE (31) | PSED | 644 | 221 | 0.03 (−0.12, 0.18) | 0.15 |
| Zimbabwe | SHINE (HIV−) (33) | MDAT | 395 | 382 | 0.07 (−0.07, 0.21) | 0.16 |
| Zimbabwe | SHINE (HIV+) (34) | MDAT | 67 | 68 | 0.24 (0.00, 0.48) | 0.10 |
| **I² = 0.68, Tau² = 0.02** | | | **4329** | **5161** | **0.08 (−0.03, 0.19)** | |

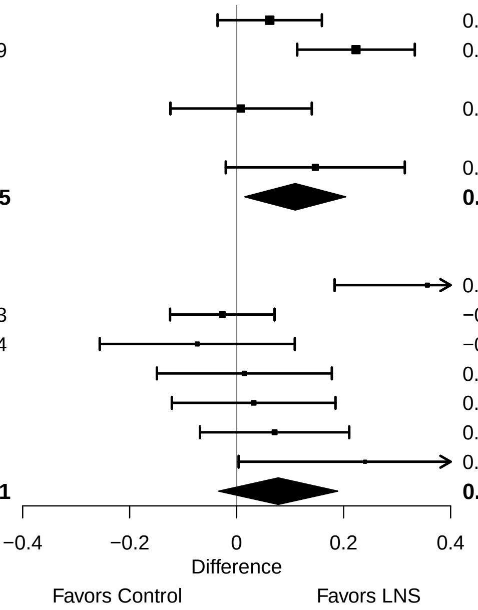

#### Supplemental figure 4D: Mean difference in social-emotional z-score

### 4D7: Stratified by Supplement duration

**Supplement duration**
**(p−diff = 0.702)**

| Country | Trial | Tool | N | N | MD (95% CI) | W |
|---|---|---|---|---|---|---|
| **Supplement duration − 12m or less** | | | | | | |
| Bangladesh | JiVitA−4 (21) | | | | | |
| Burkina Faso | iLiNS−Zinc (24) | DMC | 746 | 375 | 0.36 (0.18, 0.53) | 0.13 |
| Ghana | GHANA (25) | | | | | |
| Ghana | iLiNS−DYADG (26) | PSED | 332 | 657 | 0.01 (−0.12, 0.14) | 0.16 |
| Haiti | HAITI (27) | | | | | |
| Madagascar | MAHAY (29) | ASQI | 1613 | 1604 | −0.07 (−0.26, 0.11) | 0.13 |
| Malawi | iLiNS−DYADM (30) | PSED | 215 | 438 | 0.01 (−0.15, 0.18) | 0.14 |
| Malawi | iLiNS−DOSE (31) | PSED | 644 | 221 | 0.03 (−0.12, 0.18) | 0.15 |
| Zimbabwe | SHINE[1] (HIV−) (33), LNS vs SOC | MDAT | 395 | 382 | 0.07 (−0.07, 0.21) | 0.18 |
| Zimbabwe | LNS+WSH vs WSH | MDAT | 446 | 417 | 0.17 (0.02, 0.31) | 0.18 |
| Zimbabwe | SHINE[1] (HIV+) (34), LNS vs SOC | MDAT | 67 | 68 | 0.24 (0.00, 0.48) | 0.12 |
| Zimbabwe | LNS+WSH vs WSH | MDAT | 103 | 83 | 0.44 (0.14, 0.73) | 0.12 |
| **$I^2$ = 0.73, $Tau^2$ = 0.02** | | | **4561** | **4245** | **0.11 (−0.01, 0.24)** | |
| **Supplement duration − > 12m** | | | | | | |
| Bangladesh | RDNS (22) | DMC | 1657 | 815 | 0.06 (−0.04, 0.16) | 0.25 |
| Bangladesh | WASH−B (23) | EASQ | 1066 | 3266 | 0.12 (0.05, 0.20) | 0.27 |
| Kenya | WASH−B (28) | EASQ | 1362 | 4745 | 0.01 (−0.05, 0.08) | 0.28 |
| Mali | PROMIS CS (32) | DMC | 927 | 944 | 0.15 (−0.02, 0.31) | 0.19 |
| **$I^2$ = 0.50, $Tau^2$ = 0.00** | | | **5012** | **9770** | **0.07 (0.01, 0.13)** | |

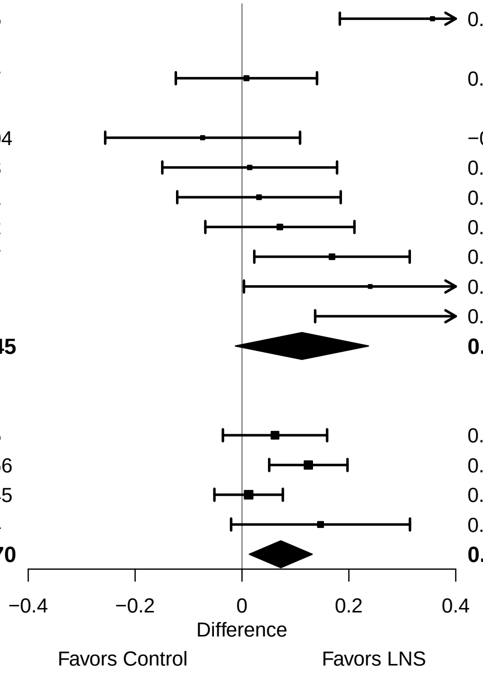

#### Supplemental figure 4D: Mean difference in social-emotional z-score

### 4D8: Stratified by Frequency of contact

**Frequency of contact**
**(p−diff = 0.897)**

| Country | Trial | Tool | N | N | MD (95% CI) | W |
|---|---|---|---|---|---|---|
| **Frequency of contact − Monthly** | | | | | | |
| Bangladesh | RDNS (22) | DMC | 1657 | 815 | 0.06 (−0.04, 0.16) | 0.19 |
| Haiti | HAITI (27) | | | | | |
| Kenya | WASH−B (28) | EASQ | 1362 | 4745 | 0.01 (−0.05, 0.08) | 0.21 |
| Madagascar | MAHAY (29) | ASQI | 1613 | 1604 | −0.07 (−0.26, 0.11) | 0.13 |
| Mali | PROMIS CS (32) | DMC | 927 | 944 | 0.15 (−0.02, 0.31) | 0.14 |
| Zimbabwe | SHINE[1] (HIV−) (33), LNS vs SOC | MDAT | 395 | 382 | 0.07 (−0.07, 0.21) | 0.19 |
| Zimbabwe | LNS+WSH vs WSH | MDAT | 446 | 417 | 0.17 (0.02, 0.31) | 0.19 |
| Zimbabwe | SHINE[1] (HIV+) (34), LNS vs SOC | MDAT | 67 | 68 | 0.24 (0.00, 0.48) | 0.13 |
| Zimbabwe | LNS+WSH vs WSH | MDAT | 103 | 83 | 0.44 (0.14, 0.73) | 0.13 |
| **I² = 0.68, Tau² = 0.01** | | | **6570** | **9058** | **0.09 (−0.01, 0.20)** | |
| **Frequency of contact − Weekly** | | | | | | |
| Bangladesh | JiVitA−4 (21) | | | | | |
| Bangladesh | WASH−B (23) | EASQ | 1066 | 3266 | 0.12 (0.05, 0.20) | 0.26 |
| Burkina Faso | iLiNS−Zinc (24) | DMC | 746 | 375 | 0.36 (0.18, 0.53) | 0.17 |
| Ghana | GHANA (25) | | | | | |
| Ghana | iLiNS−DYADG (26) | PSED | 332 | 657 | 0.01 (−0.12, 0.14) | 0.21 |
| Malawi | iLiNS−DYADM (30) | PSED | 215 | 438 | 0.01 (−0.15, 0.18) | 0.18 |
| Malawi | iLiNS−DOSE (31) | PSED | 644 | 221 | 0.03 (−0.12, 0.18) | 0.19 |
| **I² = 0.68, Tau² = 0.01** | | | **3003** | **4957** | **0.10 (−0.02, 0.23)** | |

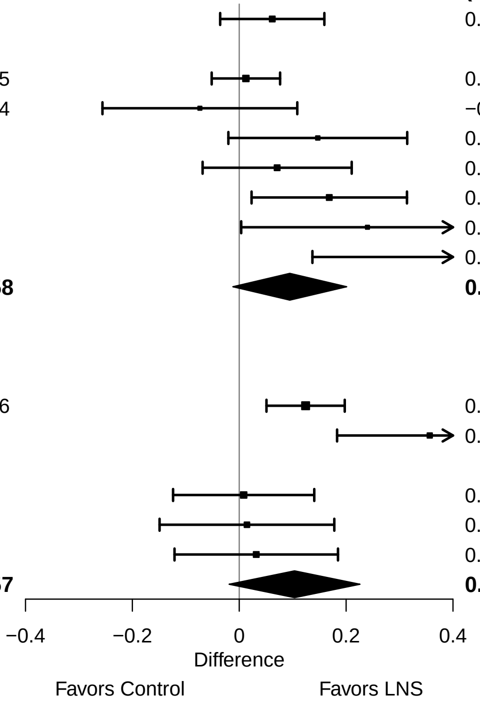

#### Supplemental figure 4D: Mean difference in social-emotional z-score

### 4D9: Stratified by Average SQ-LNS compliance

**Average SQ−LNS compliance**
**(p−diff = 0.863)**

**Average SQ−LNS compliance − Low**

| **Country** | **Trial** | **Tool** | **N** | **N** | **MD (95% CI)** | **W** |
|---|---|---|---|---|---|---|
| Ghana | iLiNS−DYADG (26) | PSED | 332 | 657 | 0.01 (−0.12, 0.14) | 0.18 |
| Malawi | iLiNS−DYADM (30) | PSED | 215 | 438 | 0.01 (−0.15, 0.18) | 0.16 |
| Malawi | iLiNS−DOSE (31) | PSED | 644 | 221 | 0.03 (−0.12, 0.18) | 0.17 |
| Mali | PROMIS CS (32) | DMC | 927 | 944 | 0.15 (−0.02, 0.31) | 0.15 |
| Zimbabwe | SHINE[1] (HIV−) (33), LNS vs SOC | MDAT | 395 | 382 | 0.07 (−0.07, 0.21) | 0.21 |
| Zimbabwe | LNS+WSH vs WSH | MDAT | 446 | 417 | 0.17 (0.02, 0.31) | 0.21 |
| Zimbabwe | SHINE[1] (HIV+) (34), LNS vs SOC | MDAT | 67 | 68 | 0.24 (0.00, 0.48) | 0.13 |
| Zimbabwe | LNS+WSH vs WSH | MDAT | 103 | 83 | 0.44 (0.14, 0.73) | 0.13 |
| **$I^2$ = 0.53, $Tau^2$ = 0.01** | | | **3129** | **3210** | **0.10 (0.01, 0.20)** | |

**Average SQ−LNS compliance − High**

| Country | Trial | Tool | N | N | MD (95% CI) | W |
|---|---|---|---|---|---|---|
| Bangladesh | JiVitA−4 (21) | | | | | |
| Bangladesh | RDNS (22) | DMC | 1657 | 815 | 0.06 (−0.04, 0.16) | 0.26 |
| Bangladesh | WASH−B (23) | EASQ | 1066 | 3266 | 0.12 (0.05, 0.20) | 0.28 |
| Burkina Faso | iLiNS−Zinc (24) | DMC | 746 | 375 | 0.36 (0.18, 0.53) | 0.18 |
| Ghana | GHANA (25) | | | | | |
| Haiti | HAITI (27) | | | | | |
| Kenya | WASH−B (28) | EASQ | 1362 | 4745 | 0.01 (−0.05, 0.08) | 0.29 |
| **$I^2$ = 0.81, $Tau^2$ = 0.02** | | | **4831** | **9201** | **0.12 (−0.02, 0.26)** | |

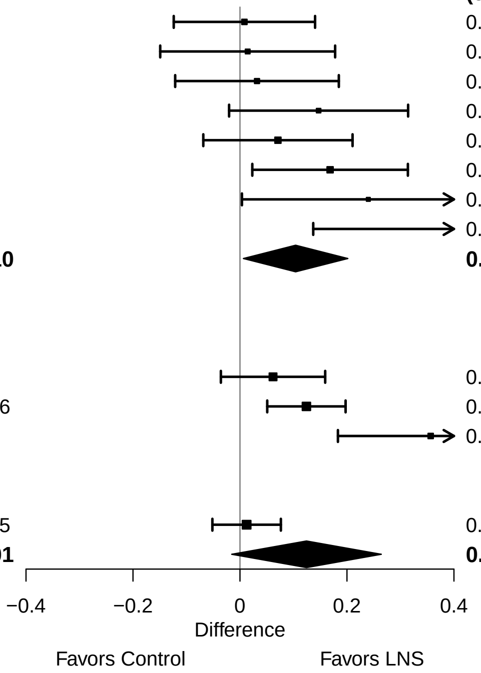

#### Supplemental figure 4E: Social-emotional lowest decile prevalence ratio

##### 4E1: Stratified by Geographic region (insufficient comparisons)

#### Supplemental figure 4E: Social-emotional lowest decile prevalence ratio

### 4E2: Stratified by Stunting burden

**Stunting burden**
**(p−diff = 0.228)**

| Country | Trial | | Tool | N | N | PR (95% CI) | W |
|---|---|---|---|---|---|---|---|
| **Stunting burden − Less than 35%** | | | | | | | |
| Ghana | GHANA (25) | | | | | | |
| Ghana | iLiNS−DYADG (26) | | PSED | 332 | 657 | 0.99 (0.67, 1.47) | 0.31 |
| Haiti | HAITI (27) | | | | | | |
| Kenya | WASH−B (28) | | EASQ | 1362 | 4745 | 0.87 (0.71, 1.08) | 0.44 |
| Malawi | iLiNS−DYADM (30) | | PSED | 215 | 438 | 1.00 (0.61, 1.61) | 0.26 |
| **I² = 0.00, Tau² = 0.00** | | | | **1909** | **5840** | **0.91 (0.77, 1.08)** | |
| **Stunting burden − More than 35%** | | | | | | | |
| Bangladesh | JiVitA−4 (21) | | | | | | |
| Bangladesh | RDNS (22) | | DMC | 1657 | 815 | 0.84 (0.66, 1.07) | 0.16 |
| Bangladesh | WASH−B (23) | | EASQ | 1066 | 3266 | 0.81 (0.64, 1.03) | 0.16 |
| Burkina Faso | iLiNS−Zinc (24) | | DMC | 746 | 375 | 0.41 (0.28, 0.62) | 0.12 |
| Madagascar | MAHAY (29) | | ASQI | 1613 | 1604 | 1.15 (0.75, 1.78) | 0.11 |
| Malawi | iLiNS−DOSE (31) | | PSED | 644 | 221 | 1.08 (0.68, 1.72) | 0.10 |
| Mali | PROMIS CS (32) | | DMC | 927 | 944 | 0.77 (0.56, 1.07) | 0.14 |
| Zimbabwe | SHINE¹ (HIV−) (33), | LNS vs SOC | MDAT | 395 | 382 | 0.86 (0.56, 1.33) | 0.14 |
| Zimbabwe | | LNS+WSH vs WSH | MDAT | 446 | 417 | 0.62 (0.41, 0.93) | 0.14 |
| Zimbabwe | SHINE¹ (HIV+) (34), | LNS vs SOC | MDAT | 67 | 68 | 0.68 (0.22, 2.04) | 0.07 |
| Zimbabwe | | LNS+WSH vs WSH | MDAT | 103 | 83 | 0.33 (0.15, 0.76) | 0.07 |
| **I² = 0.63, Tau² = 0.08** | | | | **7664** | **8175** | **0.75 (0.59, 0.95)** | |

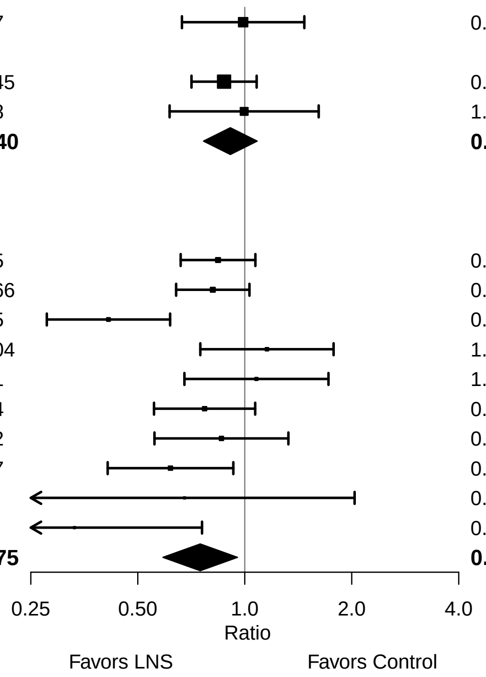

#### Supplemental figure 4E: Social-emotional lowest decile prevalence ratio

### 4E3: Stratified by Malaria prevalence

**Malaria prevalence**
**(p−diff = 0.902)**

| Country | Trial | | Tool | N | N | PR (95% CI) | W |
|---|---|---|---|---|---|---|---|
| **Malaria prevalence − Less than 10%** | | | | | | | |
| Bangladesh | JiVitA−4 (21) | | | | | | |
| Bangladesh | RDNS (22) | | DMC | 1657 | 815 | 0.84 (0.66, 1.07) | 0.20 |
| Bangladesh | WASH−B (23) | | EASQ | 1066 | 3266 | 0.81 (0.64, 1.03) | 0.20 |
| Haiti | HAITI (27) | | | | | | |
| Kenya | WASH−B (28) | | EASQ | 1362 | 4745 | 0.87 (0.71, 1.08) | 0.21 |
| Madagascar | MAHAY (29) | | ASQI | 1613 | 1604 | 1.15 (0.75, 1.78) | 0.14 |
| Zimbabwe | SHINE[1] (HIV−) (33), | LNS vs SOC | MDAT | 395 | 382 | 0.86 (0.56, 1.33) | 0.18 |
| Zimbabwe | | LNS+WSH vs WSH | MDAT | 446 | 417 | 0.62 (0.41, 0.93) | 0.18 |
| Zimbabwe | SHINE[1] (HIV+) (34), | LNS vs SOC | MDAT | 67 | 68 | 0.68 (0.22, 2.04) | 0.08 |
| Zimbabwe | | LNS+WSH vs WSH | MDAT | 103 | 83 | 0.33 (0.15, 0.76) | 0.08 |
| **I² = 0.33, Tau² = 0.03** | | | | **6709** | **11380** | **0.81 (0.67, 0.98)** | |
| **Malaria prevalence − At least 10%** | | | | | | | |
| Burkina Faso | iLiNS−Zinc (24) | | DMC | 746 | 375 | 0.41 (0.28, 0.62) | 0.21 |
| Ghana | GHANA (25) | | | | | | |
| Ghana | iLiNS−DYADG (26) | | PSED | 332 | 657 | 0.99 (0.67, 1.47) | 0.21 |
| Malawi | iLiNS−DYADM (30) | | PSED | 215 | 438 | 1.00 (0.61, 1.61) | 0.17 |
| Malawi | iLiNS−DOSE (31) | | PSED | 644 | 221 | 1.08 (0.68, 1.72) | 0.18 |
| Mali | PROMIS CS (32) | | DMC | 927 | 944 | 0.77 (0.56, 1.07) | 0.24 |
| **I² = 0.71, Tau² = 0.11** | | | | **2864** | **2635** | **0.80 (0.56, 1.12)** | |

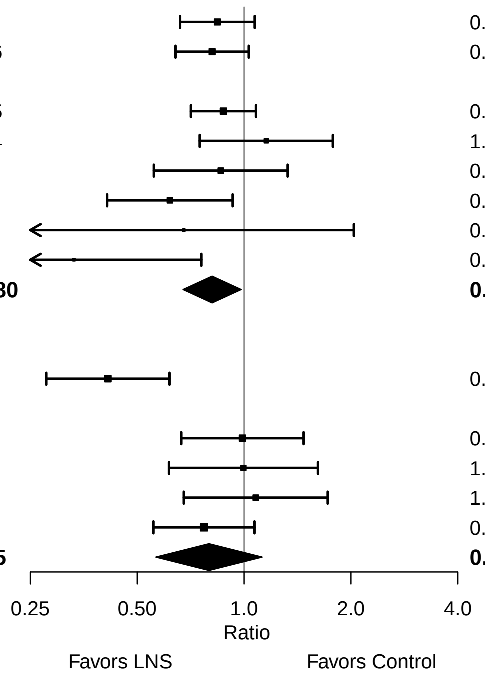

#### Supplemental figure 4E: Social-emotional lowest decile prevalence ratio

### 4E4: Stratified by Anemia prevalence

**Anemia prevalence**
**(p−diff = 0.534)**

| Country | Trial | | Tool | N | N | PR (95% CI) | W |
|---|---|---|---|---|---|---|---|
| **Anemia prevalence − High** | | | | | | | |
| Burkina Faso | iLiNS−Zinc (24) | | DMC | 746 | 375 | 0.41 (0.28, 0.62) | 0.26 |
| Ghana | GHANA (25) | | | | | | |
| Haiti | HAITI (27) | | | | | | |
| Malawi | iLiNS−DYADM (30) | | PSED | 215 | 438 | 1.00 (0.61, 1.61) | 0.22 |
| Malawi | iLiNS−DOSE (31) | | PSED | 644 | 221 | 1.08 (0.68, 1.72) | 0.22 |
| Mali | PROMIS CS (32) | | DMC | 927 | 944 | 0.77 (0.56, 1.07) | 0.30 |
| **I² = 0.75, Tau² = 0.14** | | | | **2532** | **1978** | **0.76 (0.49, 1.15)** | |
| **Anemia prevalence − Moderate** | | | | | | | |
| Bangladesh | JiVitA−4 (21) | | | | | | |
| Bangladesh | RDNS (22) | | DMC | 1657 | 815 | 0.84 (0.66, 1.07) | 0.17 |
| Bangladesh | WASH−B (23) | | EASQ | 1066 | 3266 | 0.81 (0.64, 1.03) | 0.17 |
| Ghana | iLiNS−DYADG (26) | | PSED | 332 | 657 | 0.99 (0.67, 1.47) | 0.13 |
| Kenya | WASH−B (28) | | EASQ | 1362 | 4745 | 0.87 (0.71, 1.08) | 0.18 |
| Madagascar | MAHAY (29) | | ASQI | 1613 | 1604 | 1.15 (0.75, 1.78) | 0.12 |
| Zimbabwe | SHINE¹ (HIV−) (33), | LNS vs SOC | MDAT | 395 | 382 | 0.86 (0.56, 1.33) | 0.15 |
| Zimbabwe | | LNS+WSH vs WSH | MDAT | 446 | 417 | 0.62 (0.41, 0.93) | 0.15 |
| Zimbabwe | SHINE¹ (HIV+) (34), | LNS vs SOC | MDAT | 67 | 68 | 0.68 (0.22, 2.04) | 0.07 |
| Zimbabwe | | LNS+WSH vs WSH | MDAT | 103 | 83 | 0.33 (0.15, 0.76) | 0.07 |
| **I² = 0.27, Tau² = 0.02** | | | | **7041** | **12037** | **0.83 (0.71, 0.98)** | |

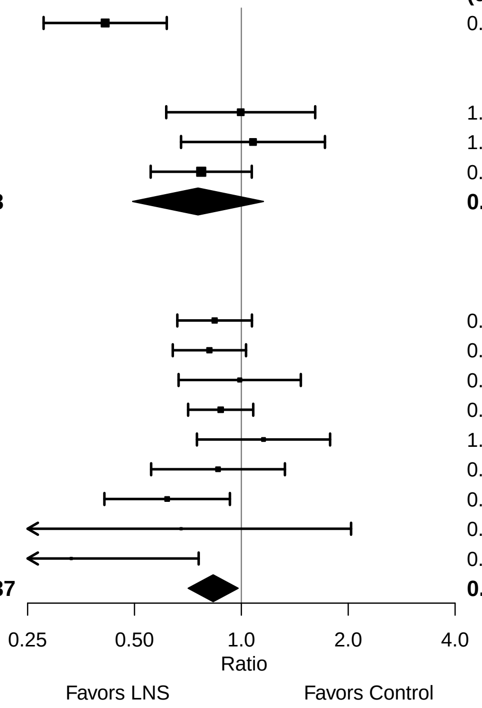

#### Supplemental figure 4E: Social-emotional lowest decile prevalence ratio

### 4E5: Stratified by Source water quality

**Source water quality**
**(p−diff = 0.394)**

| Country | Trial | Tool | N | N | PR (95% CI) | W |
|---|---|---|---|---|---|---|
| **Source water quality − Improved** | | | | | | |
| Bangladesh | JiVitA−4 (21) | | | | | |
| Bangladesh | RDNS (22) | DMC | 1657 | 815 | 0.84 (0.66, 1.07) | 0.28 |
| Bangladesh | WASH−B (23) | EASQ | 528 | 1099 | 0.70 (0.50, 0.99) | 0.22 |
| Ghana | GHANA (25) | | | | | |
| Ghana | iLiNS−DYADG (26) | PSED | 332 | 657 | 0.99 (0.67, 1.47) | 0.19 |
| Haiti | HAITI (27) | | | | | |
| Malawi | iLiNS−DYADM (30) | PSED | 215 | 438 | 1.00 (0.61, 1.61) | 0.15 |
| Malawi | iLiNS−DOSE (31) | PSED | 644 | 221 | 1.08 (0.68, 1.72) | 0.16 |
| **I² = 0.00, Tau² = 0.00** | | | **3376** | **3230** | **0.87 (0.74, 1.02)** | |
| **Source water quality − Unimproved** | | | | | | |
| Burkina Faso | iLiNS−Zinc (24) | DMC | 746 | 375 | 0.41 (0.28, 0.62) | 0.19 |
| Kenya | WASH−B (28) | EASQ | 649 | 2073 | 0.85 (0.60, 1.21) | 0.22 |
| Madagascar | MAHAY (29) | ASQI | 1613 | 1604 | 1.15 (0.75, 1.78) | 0.18 |
| Mali | PROMIS CS (32) | DMC | 927 | 944 | 0.77 (0.56, 1.07) | 0.23 |
| Zimbabwe | SHINE (HIV−) (33) | MDAT | 395 | 382 | 0.86 (0.56, 1.33) | 0.18 |
| Zimbabwe | SHINE (HIV+) (34) | MDAT | | | | |
| **I² = 0.70, Tau² = 0.10** | | | **4330** | **5378** | **0.77 (0.55, 1.06)** | |

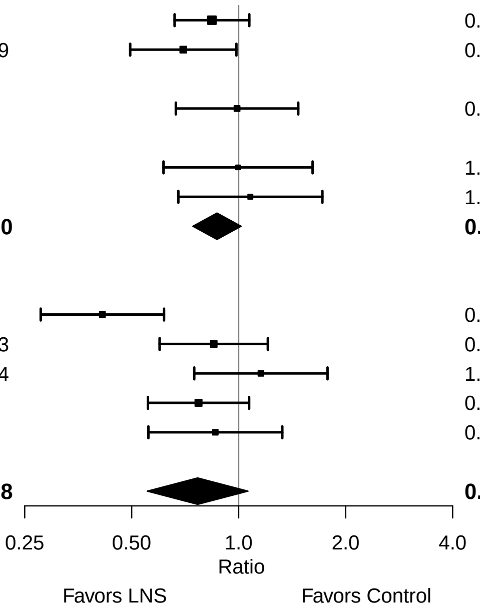

##### Supplemental figure 4E: Social-emotional lowest decile prevalence ratio

#### 4E6: Stratified by Sanitation

**Sanitation**
**(p−diff = 0.888)**

| **Sanitation − Improved** | | | | | |
|---|---|---|---|---|---|---|
| **Country** | **Trial** | **Tool** | **N** | **N** | **PR (95% CI)** | **W** |
| Bangladesh | JiVitA−4 (21) | | | | | |
| Bangladesh | RDNS (22) | DMC | 1657 | 815 | 0.84 (0.66, 1.07) | 0.30 |
| Bangladesh | WASH−B (23) | EASQ | 528 | 1099 | 0.70 (0.50, 0.99) | 0.24 |
| Ghana | GHANA (25) | | | | | |
| Ghana | iLiNS−DYADG (26) | PSED | 332 | 657 | 0.99 (0.67, 1.47) | 0.21 |
| Haiti | HAITI (27) | | | | | |
| Mali | PROMIS CS (32) | DMC | 927 | 944 | 0.77 (0.56, 1.07) | 0.25 |
| **I² = 0.00, Tau² = 0.00** | | | **3444** | **3515** | **0.81 (0.70, 0.95)** | |
| **Sanitation − Unimproved** | | | | | | |
| Burkina Faso | iLiNS−Zinc (24) | DMC | 746 | 375 | 0.41 (0.28, 0.62) | 0.18 |
| Kenya | WASH−B (28) | EASQ | 649 | 2073 | 0.85 (0.60, 1.21) | 0.20 |
| Madagascar | MAHAY (29) | ASQI | 1613 | 1604 | 1.15 (0.75, 1.78) | 0.16 |
| Malawi | iLiNS−DYADM (30) | PSED | 215 | 438 | 1.00 (0.61, 1.61) | 0.14 |
| Malawi | iLiNS−DOSE (31) | PSED | 644 | 221 | 1.08 (0.68, 1.72) | 0.15 |
| Zimbabwe | SHINE (HIV−) (33) | MDAT | 395 | 382 | 0.86 (0.56, 1.33) | 0.16 |
| Zimbabwe | SHINE (HIV+) (34) | MDAT | | | | |
| **I² = 0.68, Tau² = 0.09** | | | **4262** | **5093** | **0.84 (0.62, 1.14)** | |

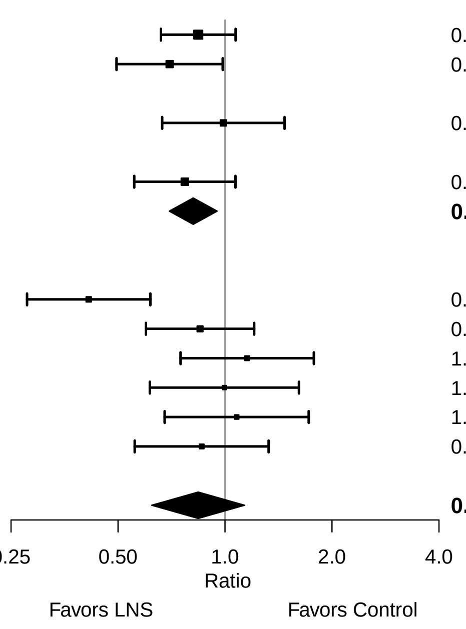

#### Supplemental figure 4E: Social-emotional lowest decile prevalence ratio

### 4E7: Stratified by Supplement duration

**Supplement duration**
**(p−diff = 0.731)**

| Country | Trial | Tool | N | N | PR (95% CI) | W |
|---|---|---|---|---|---|---|
| **Supplement duration − 12m or less** | | | | | | |
| Bangladesh | JiVitA−4 (21) | | | | | |
| Burkina Faso | iLiNS−Zinc (24) | DMC | 746 | 375 | 0.41 (0.28, 0.62) | 0.16 |
| Ghana | GHANA (25) | | | | | |
| Ghana | iLiNS−DYADG (26) | PSED | 332 | 657 | 0.99 (0.67, 1.47) | 0.16 |
| Haiti | HAITI (27) | | | | | |
| Madagascar | MAHAY (29) | ASQI | 1613 | 1604 | 1.15 (0.75, 1.78) | 0.14 |
| Malawi | iLiNS−DYADM (30) | PSED | 215 | 438 | 1.00 (0.61, 1.61) | 0.13 |
| Malawi | iLiNS−DOSE (31) | PSED | 644 | 221 | 1.08 (0.68, 1.72) | 0.13 |
| Zimbabwe | SHINE[1] (HIV−) (33), LNS vs SOC | MDAT | 395 | 382 | 0.86 (0.56, 1.33) | 0.19 |
| Zimbabwe | LNS+WSH vs WSH | MDAT | 446 | 417 | 0.62 (0.41, 0.93) | 0.19 |
| Zimbabwe | SHINE[1] (HIV+) (34), LNS vs SOC | MDAT | 67 | 68 | 0.68 (0.22, 2.04) | 0.09 |
| Zimbabwe | LNS+WSH vs WSH | MDAT | 103 | 83 | 0.33 (0.15, 0.76) | 0.09 |
| **I² = 0.71, Tau² = 0.13** | | | **4561** | **4245** | **0.78 (0.57, 1.06)** | |
| **Supplement duration − > 12m** | | | | | | |
| Bangladesh | RDNS (22) | DMC | 1657 | 815 | 0.84 (0.66, 1.07) | 0.26 |
| Bangladesh | WASH−B (23) | EASQ | 1066 | 3266 | 0.81 (0.64, 1.03) | 0.26 |
| Kenya | WASH−B (28) | EASQ | 1362 | 4745 | 0.87 (0.71, 1.08) | 0.27 |
| Mali | PROMIS CS (32) | DMC | 927 | 944 | 0.77 (0.56, 1.07) | 0.22 |
| **I² = 0.00, Tau² = 0.00** | | | **5012** | **9770** | **0.83 (0.74, 0.94)** | |

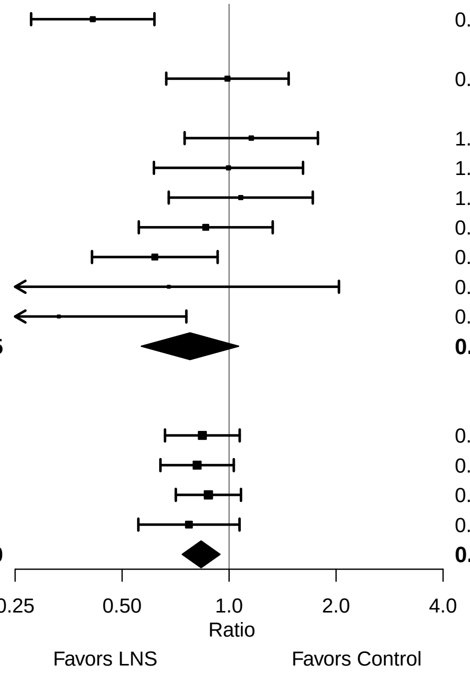

#### Supplemental figure 4E: Social-emotional lowest decile prevalence ratio

### 4E8: Stratified by Frequency of contact

**Frequency of contact**
**(p−diff = 0.991)**

| Country | Trial | Tool | N | N | PR (95% CI) | W |
|---|---|---|---|---|---|---|
| **Frequency of contact − Monthly** | | | | | | |
| Bangladesh | RDNS (22) | DMC | 1657 | 815 | 0.84 (0.66, 1.07) | 0.20 |
| Haiti | HAITI (27) | | | | | |
| Kenya | WASH−B (28) | EASQ | 1362 | 4745 | 0.87 (0.71, 1.08) | 0.21 |
| Madagascar | MAHAY (29) | ASQI | 1613 | 1604 | 1.15 (0.75, 1.78) | 0.14 |
| Mali | PROMIS CS (32) | DMC | 927 | 944 | 0.77 (0.56, 1.07) | 0.17 |
| Zimbabwe | SHINE[1] (HIV−) (33), LNS vs SOC | MDAT | 395 | 382 | 0.86 (0.56, 1.33) | 0.18 |
| Zimbabwe | LNS+WSH vs WSH | MDAT | 446 | 417 | 0.62 (0.41, 0.93) | 0.18 |
| Zimbabwe | SHINE[1] (HIV+) (34), LNS vs SOC | MDAT | 67 | 68 | 0.68 (0.22, 2.04) | 0.09 |
| Zimbabwe | LNS+WSH vs WSH | MDAT | 103 | 83 | 0.33 (0.15, 0.76) | 0.09 |
| **$I^2$ = 0.34, $Tau^2$ = 0.03** | | | **6570** | **9058** | **0.81 (0.66, 0.98)** | |
| **Frequency of contact − Weekly** | | | | | | |
| Bangladesh | JiVitA−4 (21) | | | | | |
| Bangladesh | WASH−B (23) | EASQ | 1066 | 3266 | 0.81 (0.64, 1.03) | 0.27 |
| Burkina Faso | iLiNS−Zinc (24) | DMC | 746 | 375 | 0.41 (0.28, 0.62) | 0.20 |
| Ghana | GHANA (25) | | | | | |
| Ghana | iLiNS−DYADG (26) | PSED | 332 | 657 | 0.99 (0.67, 1.47) | 0.20 |
| Malawi | iLiNS−DYADM (30) | PSED | 215 | 438 | 1.00 (0.61, 1.61) | 0.16 |
| Malawi | iLiNS−DOSE (31) | PSED | 644 | 221 | 1.08 (0.68, 1.72) | 0.17 |
| **$I^2$ = 0.71, $Tau^2$ = 0.11** | | | **3003** | **4957** | **0.81 (0.57, 1.13)** | |

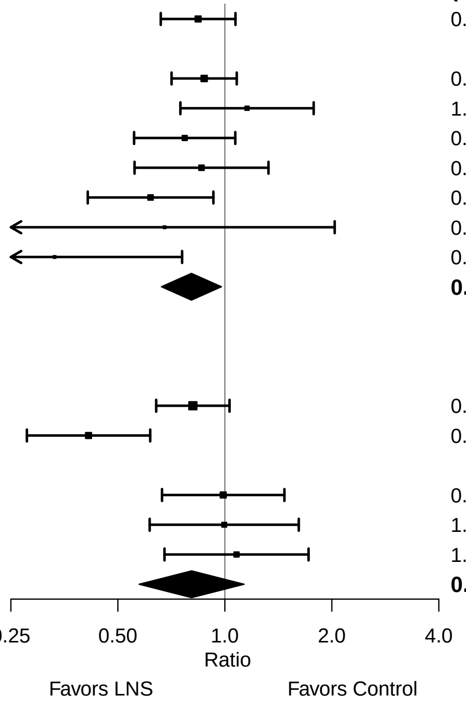

#### Supplemental figure 4E: Social-emotional lowest decile prevalence ratio

### 4E9: Stratified by Average SQ-LNS compliance

**Average SQ−LNS compliance**
**(p−diff = 0.575)**

| **Average SQ−LNS compliance − Low** | | | | | **PR** | |
|---|---|---|---|---|---|---|
| **Country** | **Trial** | **Tool** | **N** | **N** | **(95% CI)** | **W** |
| Ghana | iLiNS−DYADG (26) | PSED | 332 | 657 | 0.99 (0.67, 1.47) | 0.18 |
| Malawi | iLiNS−DYADM (30) | PSED | 215 | 438 | 1.00 (0.61, 1.61) | 0.15 |
| Malawi | iLiNS−DOSE (31) | PSED | 644 | 221 | 1.08 (0.68, 1.72) | 0.15 |
| Mali | PROMIS CS (32) | DMC | 927 | 944 | 0.77 (0.56, 1.07) | 0.21 |
| Zimbabwe | SHINE¹ (HIV−) (33), LNS vs SOC | MDAT | 395 | 382 | 0.86 (0.56, 1.33) | 0.22 |
| Zimbabwe | LNS+WSH vs WSH | MDAT | 446 | 417 | 0.62 (0.41, 0.93) | 0.22 |
| Zimbabwe | SHINE¹ (HIV+) (34), LNS vs SOC | MDAT | 67 | 68 | 0.68 (0.22, 2.04) | 0.10 |
| Zimbabwe | LNS+WSH vs WSH | MDAT | 103 | 83 | 0.33 (0.15, 0.76) | 0.10 |
| **I² = 0.34, Tau² = 0.03** | | | **3129** | **3210** | **0.82 (0.65, 1.02)** | |
| **Average SQ−LNS compliance − High** | | | | | | |
| Bangladesh | JiVitA−4 (21) | | | | | |
| Bangladesh | RDNS (22) | DMC | 1657 | 815 | 0.84 (0.66, 1.07) | 0.26 |
| Bangladesh | WASH−B (23) | EASQ | 1066 | 3266 | 0.81 (0.64, 1.03) | 0.27 |
| Burkina Faso | iLiNS−Zinc (24) | DMC | 746 | 375 | 0.41 (0.28, 0.62) | 0.19 |
| Ghana | GHANA (25) | | | | | |
| Haiti | HAITI (27) | | | | | |
| Kenya | WASH−B (28) | EASQ | 1362 | 4745 | 0.87 (0.71, 1.08) | 0.28 |
| **I² = 0.73, Tau² = 0.09** | | | **4831** | **9201** | **0.73 (0.52, 1.01)** | |

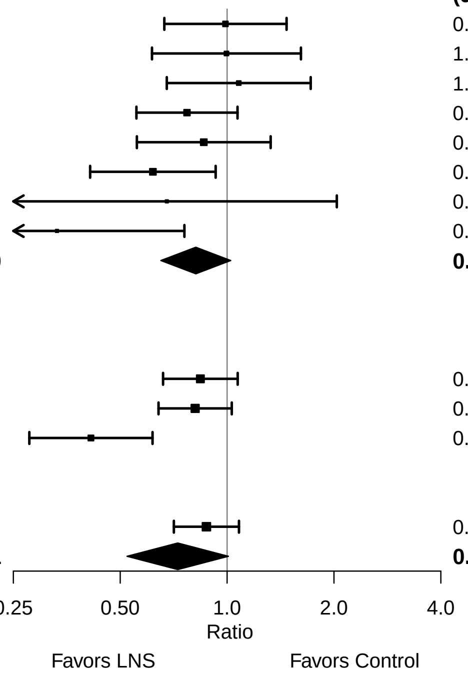

### Supplemental figure 4F: Social-emotional lowest decile prevalence difference

#### 4F1: Stratified by Geographic region (insufficient comparisons)

#### Supplemental figure 4F: Social-emotional lowest decile prevalence difference

### 4F2: Stratified by Stunting burden

**Stunting burden**
**(p−diff = 0.324)**

| Country | Trial | | Tool | N | N | PD (95% CI) | W |
|---|---|---|---|---|---|---|---|
| **Stunting burden − Less than 35%** | | | | | | | |
| Ghana | GHANA (25) | | | | | | |
| Ghana | iLiNS−DYADG (26) | | PSED | 332 | 657 | 0.00 (−0.04, 0.04) | 0.31 |
| Haiti | HAITI (27) | | | | | | |
| Kenya | WASH−B (28) | | EASQ | 1362 | 4745 | −0.01 (−0.03, 0.00) | 0.43 |
| Malawi | iLiNS−DYADM (30) | | PSED | 215 | 438 | 0.00 (−0.05, 0.05) | 0.26 |
| **I² = 0.00, Tau² = 0.00** | | | | **1909** | **5840** | **−0.01 (−0.03, 0.01)** | |
| **Stunting burden − More than 35%** | | | | | | | |
| Bangladesh | JiVitA−4 (21) | | | | | | |
| Bangladesh | RDNS (22) | | DMC | 1657 | 815 | −0.02 (−0.04, 0.01) | 0.14 |
| Bangladesh | WASH−B (23) | | EASQ | 1066 | 3266 | −0.02 (−0.04, 0.00) | 0.15 |
| Burkina Faso | iLiNS−Zinc (24) | | DMC | 746 | 375 | −0.10 (−0.14, −0.06) | 0.11 |
| Madagascar | MAHAY (29) | | ASQI | 1613 | 1604 | 0.01 (−0.01, 0.04) | 0.15 |
| Malawi | iLiNS−DOSE (31) | | PSED | 644 | 221 | 0.01 (−0.04, 0.05) | 0.10 |
| Mali | PROMIS CS (32) | | DMC | 927 | 944 | −0.03 (−0.05, 0.00) | 0.14 |
| Zimbabwe | SHINE¹ (HIV−) (33), | LNS vs SOC | MDAT | 395 | 382 | −0.01 (−0.05, 0.03) | 0.13 |
| Zimbabwe | | LNS+WSH vs WSH | MDAT | 446 | 417 | −0.05 (−0.10, −0.01) | 0.13 |
| Zimbabwe | SHINE¹ (HIV+) (34), | LNS vs SOC | MDAT | 67 | 68 | −0.03 (−0.12, 0.06) | 0.07 |
| Zimbabwe | | LNS+WSH vs WSH | MDAT | 103 | 83 | −0.14 (−0.24, −0.04) | 0.07 |
| **I² = 0.77, Tau² = 0.00** | | | | **7664** | **8175** | **−0.03 (−0.05, 0.00)** | |

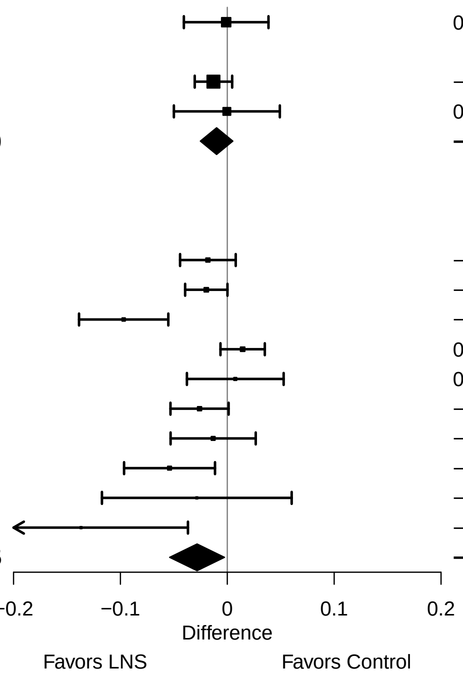

#### Supplemental figure 4F: Social-emotional lowest decile prevalence difference

### 4F3: Stratified by Malaria prevalence

**Malaria prevalence**
**(p−diff = 0.802)**

| Country | Trial | Tool | N | N | PD (95% CI) | W |
|---|---|---|---|---|---|---|
| **Malaria prevalence − Less than 10%** | | | | | | |
| Bangladesh | JiVitA−4 (21) | | | | | |
| Bangladesh | RDNS (22) | DMC | 1657 | 815 | −0.02 (−0.04, 0.01) | 0.18 |
| Bangladesh | WASH−B (23) | EASQ | 1066 | 3266 | −0.02 (−0.04, 0.00) | 0.19 |
| Haiti | HAITI (27) | | | | | |
| Kenya | WASH−B (28) | EASQ | 1362 | 4745 | −0.01 (−0.03, 0.00) | 0.20 |
| Madagascar | MAHAY (29) | ASQI | 1613 | 1604 | 0.01 (−0.01, 0.04) | 0.19 |
| Zimbabwe | SHINE¹ (HIV−) (33), LNS vs SOC | MDAT | 395 | 382 | −0.01 (−0.05, 0.03) | 0.17 |
| Zimbabwe | LNS+WSH vs WSH | MDAT | 446 | 417 | −0.05 (−0.10, −0.01) | 0.17 |
| Zimbabwe | SHINE¹ (HIV+) (34), LNS vs SOC | MDAT | 67 | 68 | −0.03 (−0.12, 0.06) | 0.08 |
| Zimbabwe | LNS+WSH vs WSH | MDAT | 103 | 83 | −0.14 (−0.24, −0.04) | 0.08 |
| **I² = 0.64, Tau² = 0.00** | | | **6709** | **11380** | **−0.02 (−0.04, 0.00)** | |
| **Malaria prevalence − At least 10%** | | | | | | |
| Burkina Faso | iLiNS−Zinc (24) | DMC | 746 | 375 | −0.10 (−0.14, −0.06) | 0.20 |
| Ghana | GHANA (25) | | | | | |
| Ghana | iLiNS−DYADG (26) | PSED | 332 | 657 | 0.00 (−0.04, 0.04) | 0.20 |
| Malawi | iLiNS−DYADM (30) | PSED | 215 | 438 | 0.00 (−0.05, 0.05) | 0.17 |
| Malawi | iLiNS−DOSE (31) | PSED | 644 | 221 | 0.01 (−0.04, 0.05) | 0.18 |
| Mali | PROMIS CS (32) | DMC | 927 | 944 | −0.03 (−0.05, 0.00) | 0.25 |
| **I² = 0.75, Tau² = 0.00** | | | **2864** | **2635** | **−0.02 (−0.06, 0.01)** | |

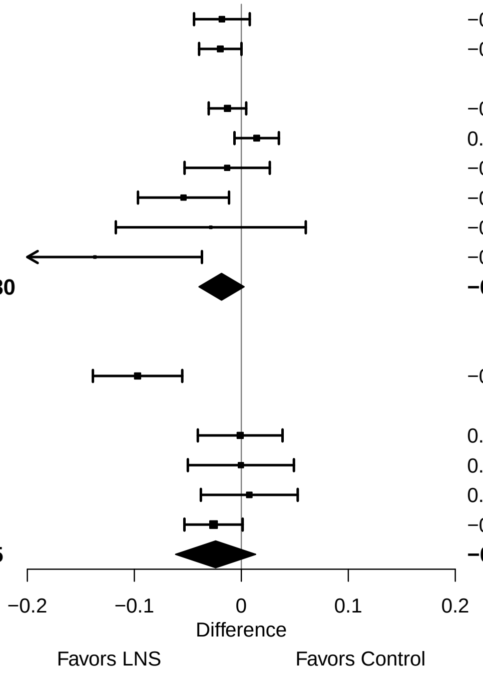

#### Supplemental figure 4F: Social-emotional lowest decile prevalence difference

### 4F4: Stratified by Anemia prevalence

**Anemia prevalence**
**(p−diff = 0.492)**

| Country | Trial | Tool | N | N | PD (95% CI) | W |
|---|---|---|---|---|---|---|
| **Anemia prevalence − High** | | | | | | |
| Burkina Faso | iLiNS−Zinc (24) | DMC | 746 | 375 | −0.10 (−0.14, −0.06) | 0.24 |
| Ghana | GHANA (25) | | | | | |
| Haiti | HAITI (27) | | | | | |
| Malawi | iLiNS−DYADM (30) | PSED | 215 | 438 | 0.00 (−0.05, 0.05) | 0.21 |
| Malawi | iLiNS−DOSE (31) | PSED | 644 | 221 | 0.01 (−0.04, 0.05) | 0.23 |
| Mali | PROMIS CS (32) | DMC | 927 | 944 | −0.03 (−0.05, 0.00) | 0.31 |
| **I² = 0.78, Tau² = 0.00** | | | **2532** | **1978** | **−0.03 (−0.08, 0.02)** | |
| **Anemia prevalence − Moderate** | | | | | | |
| Bangladesh | JiVitA−4 (21) | | | | | |
| Bangladesh | RDNS (22) | DMC | 1657 | 815 | −0.02 (−0.04, 0.01) | 0.15 |
| Bangladesh | WASH−B (23) | EASQ | 1066 | 3266 | −0.02 (−0.04, 0.00) | 0.17 |
| Ghana | iLiNS−DYADG (26) | PSED | 332 | 657 | 0.00 (−0.04, 0.04) | 0.12 |
| Kenya | WASH−B (28) | EASQ | 1362 | 4745 | −0.01 (−0.03, 0.00) | 0.17 |
| Madagascar | MAHAY (29) | ASQI | 1613 | 1604 | 0.01 (−0.01, 0.04) | 0.17 |
| Zimbabwe | SHINE¹ (HIV−) (33), LNS vs SOC | MDAT | 395 | 382 | −0.01 (−0.05, 0.03) | 0.15 |
| Zimbabwe | LNS+WSH vs WSH | MDAT | 446 | 417 | −0.05 (−0.10, −0.01) | 0.15 |
| Zimbabwe | SHINE¹ (HIV+) (34), LNS vs SOC | MDAT | 67 | 68 | −0.03 (−0.12, 0.06) | 0.07 |
| Zimbabwe | LNS+WSH vs WSH | MDAT | 103 | 83 | −0.14 (−0.24, −0.04) | 0.07 |
| **I² = 0.58, Tau² = 0.00** | | | **7041** | **12037** | **−0.02 (−0.03, 0.00)** | |

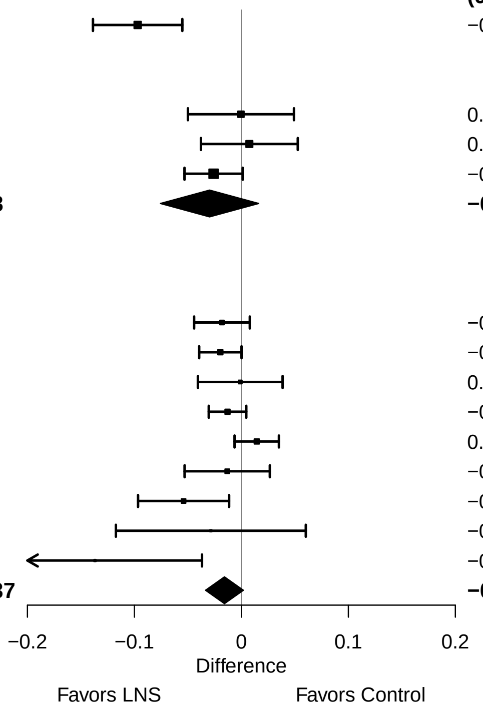

#### Supplemental figure 4F: Social-emotional lowest decile prevalence difference

### 4F5: Stratified by Source water quality

**Source water quality**
**(p−diff = 0.587)**

| Country | Trial | Tool | N | N | PD (95% CI) | W |
|---|---|---|---|---|---|---|
| **Source water quality − Improved** | | | | | | |
| Bangladesh | JiVitA−4 (21) | | | | | |
| Bangladesh | RDNS (22) | DMC | 1657 | 815 | −0.02 (−0.04, 0.01) | 0.25 |
| Bangladesh | WASH−B (23) | EASQ | 528 | 1099 | −0.04 (−0.07, −0.01) | 0.22 |
| Ghana | GHANA (25) | | | | | |
| Ghana | iLiNS−DYADG (26) | PSED | 332 | 657 | 0.00 (−0.04, 0.04) | 0.19 |
| Haiti | HAITI (27) | | | | | |
| Malawi | iLiNS−DYADM (30) | PSED | 215 | 438 | 0.00 (−0.05, 0.05) | 0.16 |
| Malawi | iLiNS−DOSE (31) | PSED | 644 | 221 | 0.01 (−0.04, 0.05) | 0.17 |
| **I² = 0.07, Tau² = 0.00** | | | **3376** | **3230** | **−0.02 (−0.03, 0.00)** | |
| **Source water quality − Unimproved** | | | | | | |
| Burkina Faso | iLiNS−Zinc (24) | DMC | 746 | 375 | −0.10 (−0.14, −0.06) | 0.16 |
| Kenya | WASH−B (28) | EASQ | 649 | 2073 | −0.01 (−0.04, 0.01) | 0.22 |
| Madagascar | MAHAY (29) | ASQI | 1613 | 1604 | 0.01 (−0.01, 0.04) | 0.24 |
| Mali | PROMIS CS (32) | DMC | 927 | 944 | −0.03 (−0.05, 0.00) | 0.21 |
| Zimbabwe | SHINE (HIV−) (33) | MDAT | 395 | 382 | −0.01 (−0.05, 0.03) | 0.17 |
| Zimbabwe | SHINE (HIV+) (34) | MDAT | | | | |
| **I² = 0.83, Tau² = 0.00** | | | **4330** | **5378** | **−0.03 (−0.06, 0.01)** | |

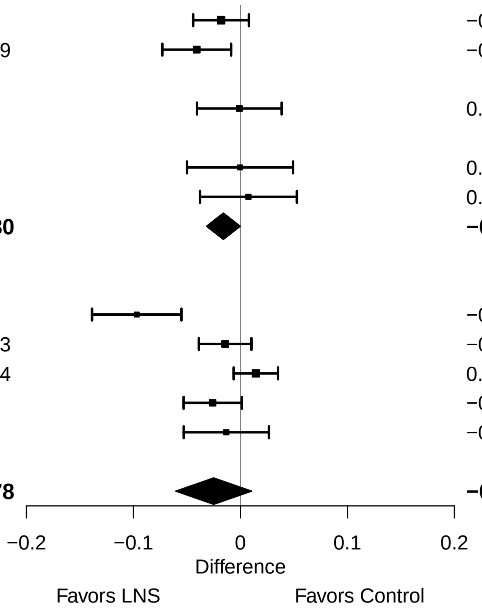

#### Supplemental figure 4F: Social-emotional lowest decile prevalence difference

### 4F6: Stratified by Sanitation

**Sanitation**
**(p−diff = 0.739)**

| Country | Trial | Tool | N | N | PD (95% CI) | W |
|---|---|---|---|---|---|---|
| **Sanitation − Improved** | | | | | | |
| Bangladesh | JiVitA−4 (21) | | | | | |
| Bangladesh | RDNS (22) | DMC | 1657 | 815 | −0.02 (−0.04, 0.01) | 0.27 |
| Bangladesh | WASH−B (23) | EASQ | 528 | 1099 | −0.04 (−0.07, −0.01) | 0.25 |
| Ghana | GHANA (25) | | | | | |
| Ghana | iLiNS−DYADG (26) | PSED | 332 | 657 | 0.00 (−0.04, 0.04) | 0.21 |
| Haiti | HAITI (27) | | | | | |
| Mali | PROMIS CS (32) | DMC | 927 | 944 | −0.03 (−0.05, 0.00) | 0.27 |
| **I² = 0.00, Tau² = 0.00** | | | **3444** | **3515** | **−0.02 (−0.04, −0.01)** | |
| **Sanitation − Unimproved** | | | | | | |
| Burkina Faso | iLiNS−Zinc (24) | DMC | 746 | 375 | −0.10 (−0.14, −0.06) | 0.15 |
| Kenya | WASH−B (28) | EASQ | 649 | 2073 | −0.01 (−0.04, 0.01) | 0.21 |
| Madagascar | MAHAY (29) | ASQI | 1613 | 1604 | 0.01 (−0.01, 0.04) | 0.22 |
| Malawi | iLiNS−DYADM (30) | PSED | 215 | 438 | 0.00 (−0.05, 0.05) | 0.13 |
| Malawi | iLiNS−DOSE (31) | PSED | 644 | 221 | 0.01 (−0.04, 0.05) | 0.14 |
| Zimbabwe | SHINE (HIV−) (33) | MDAT | 395 | 382 | −0.01 (−0.05, 0.03) | 0.16 |
| Zimbabwe | SHINE (HIV+) (34) | MDAT | | | | |
| **I² = 0.78, Tau² = 0.00** | | | **4262** | **5093** | **−0.02 (−0.05, 0.02)** | |

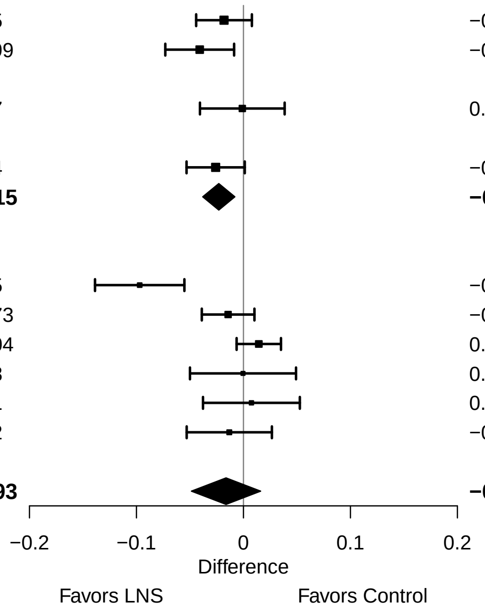

#### Supplemental figure 4F: Social-emotional lowest decile prevalence difference

### 4F7: Stratified by Supplement duration

**Supplement duration**
**(p−diff = 0.815)**

| Country | Trial | Tool | N | N | PD (95% CI) | W |
|---|---|---|---|---|---|---|
| **Supplement duration − 12m or less** | | | | | | |
| Bangladesh | JiVitA−4 (21) | | | | | |
| Burkina Faso | iLiNS−Zinc (24) | DMC | 746 | 375 | −0.10 (−0.14, −0.06) | 0.14 |
| Ghana | GHANA (25) | | | | | |
| Ghana | iLiNS−DYADG (26) | PSED | 332 | 657 | 0.00 (−0.04, 0.04) | 0.15 |
| Haiti | HAITI (27) | | | | | |
| Madagascar | MAHAY (29) | ASQI | 1613 | 1604 | 0.01 (−0.01, 0.04) | 0.20 |
| Malawi | iLiNS−DYADM (30) | PSED | 215 | 438 | 0.00 (−0.05, 0.05) | 0.12 |
| Malawi | iLiNS−DOSE (31) | PSED | 644 | 221 | 0.01 (−0.04, 0.05) | 0.13 |
| Zimbabwe | SHINE¹ (HIV−) (33), LNS vs SOC | MDAT | 395 | 382 | −0.01 (−0.05, 0.03) | 0.17 |
| Zimbabwe | LNS+WSH vs WSH | MDAT | 446 | 417 | −0.05 (−0.10, −0.01) | 0.17 |
| Zimbabwe | SHINE¹ (HIV+) (34), LNS vs SOC | MDAT | 67 | 68 | −0.03 (−0.12, 0.06) | 0.09 |
| Zimbabwe | LNS+WSH vs WSH | MDAT | 103 | 83 | −0.14 (−0.24, −0.04) | 0.09 |
| **I² = 0.80, Tau² = 0.00** | | | **4561** | **4245** | **−0.03 (−0.06, 0.01)** | |
| **Supplement duration − > 12m** | | | | | | |
| Bangladesh | RDNS (22) | DMC | 1657 | 815 | −0.02 (−0.04, 0.01) | 0.24 |
| Bangladesh | WASH−B (23) | EASQ | 1066 | 3266 | −0.02 (−0.04, 0.00) | 0.26 |
| Kenya | WASH−B (28) | EASQ | 1362 | 4745 | −0.01 (−0.03, 0.00) | 0.27 |
| Mali | PROMIS CS (32) | DMC | 927 | 944 | −0.03 (−0.05, 0.00) | 0.23 |
| **I² = 0.00, Tau² = 0.00** | | | **5012** | **9770** | **−0.02 (−0.03, −0.01)** | |

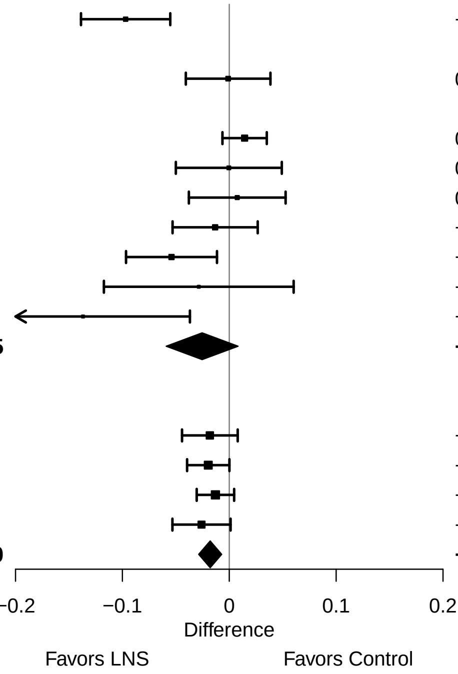

#### Supplemental figure 4F: Social-emotional lowest decile prevalence difference

### 4F8: Stratified by Frequency of contact

**Frequency of contact**
**(p−diff = 0.919)**

| Country | Trial | Tool | N | N | PD (95% CI) | W |
|---|---|---|---|---|---|---|
| **Frequency of contact − Monthly** | | | | | | |
| Bangladesh | RDNS (22) | DMC | 1657 | 815 | −0.02 (−0.04, 0.01) | 0.18 |
| Haiti | HAITI (27) | | | | | |
| Kenya | WASH−B (28) | EASQ | 1362 | 4745 | −0.01 (−0.03, 0.00) | 0.20 |
| Madagascar | MAHAY (29) | ASQI | 1613 | 1604 | 0.01 (−0.01, 0.04) | 0.19 |
| Mali | PROMIS CS (32) | DMC | 927 | 944 | −0.03 (−0.05, 0.00) | 0.18 |
| Zimbabwe | SHINE[1] (HIV−) (33), LNS vs SOC | MDAT | 395 | 382 | −0.01 (−0.05, 0.03) | 0.17 |
| Zimbabwe | LNS+WSH vs WSH | MDAT | 446 | 417 | −0.05 (−0.10, −0.01) | 0.17 |
| Zimbabwe | SHINE[1] (HIV+) (34), LNS vs SOC | MDAT | 67 | 68 | −0.03 (−0.12, 0.06) | 0.08 |
| Zimbabwe | LNS+WSH vs WSH | MDAT | 103 | 83 | −0.14 (−0.24, −0.04) | 0.08 |
| **$I^2$ = 0.65, $Tau^2$ = 0.00** | | | **6570** | **9058** | **−0.02 (−0.04, 0.00)** | |
| **Frequency of contact − Weekly** | | | | | | |
| Bangladesh | JiVitA−4 (21) | | | | | |
| Bangladesh | WASH−B (23) | EASQ | 1066 | 3266 | −0.02 (−0.04, 0.00) | 0.27 |
| Burkina Faso | iLiNS−Zinc (24) | DMC | 746 | 375 | −0.10 (−0.14, −0.06) | 0.19 |
| Ghana | GHANA (25) | | | | | |
| Ghana | iLiNS−DYADG (26) | PSED | 332 | 657 | 0.00 (−0.04, 0.04) | 0.20 |
| Malawi | iLiNS−DYADM (30) | PSED | 215 | 438 | 0.00 (−0.05, 0.05) | 0.16 |
| Malawi | iLiNS−DOSE (31) | PSED | 644 | 221 | 0.01 (−0.04, 0.05) | 0.18 |
| **$I^2$ = 0.75, $Tau^2$ = 0.00** | | | **3003** | **4957** | **−0.02 (−0.06, 0.01)** | |

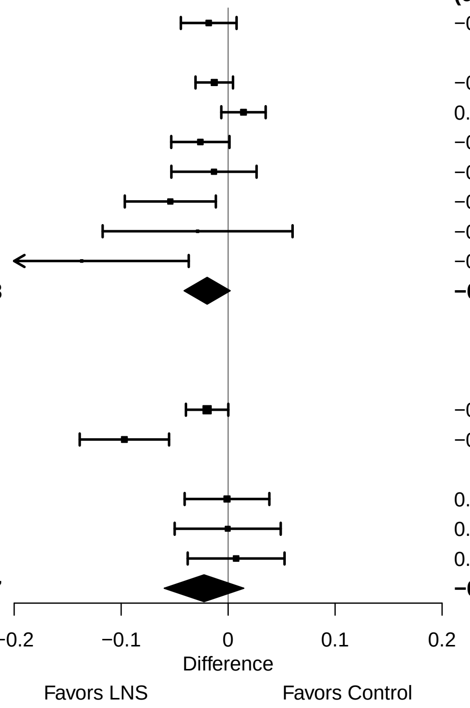

**Supplemental figure 4F: Social-emotional lowest decile prevalence difference**

#### 4F9: Stratified by Average SQ-LNS compliance

**Average SQ−LNS compliance**
**(p−diff = 0.563)**

| Country | Trial | | Tool | N | N | PD (95% CI) | W |
|---|---|---|---|---|---|---|---|
| **Average SQ−LNS compliance − Low** | | | | | | | |
| Ghana | iLiNS−DYADG (26) | | PSED | 332 | 657 | 0.00 (−0.04, 0.04) | 0.17 |
| Malawi | iLiNS−DYADM (30) | | PSED | 215 | 438 | 0.00 (−0.05, 0.05) | 0.14 |
| Malawi | iLiNS−DOSE (31) | | PSED | 644 | 221 | 0.01 (−0.04, 0.05) | 0.16 |
| Mali | PROMIS CS (32) | | DMC | 927 | 944 | −0.03 (−0.05, 0.00) | 0.22 |
| Zimbabwe | SHINE[1] (HIV−) (33), | LNS vs SOC | MDAT | 395 | 382 | −0.01 (−0.05, 0.03) | 0.21 |
| Zimbabwe | | LNS+WSH vs WSH | MDAT | 446 | 417 | −0.05 (−0.10, −0.01) | 0.21 |
| Zimbabwe | SHINE[1] (HIV+) (34), | LNS vs SOC | MDAT | 67 | 68 | −0.03 (−0.12, 0.06) | 0.10 |
| Zimbabwe | | LNS+WSH vs WSH | MDAT | 103 | 83 | −0.14 (−0.24, −0.04) | 0.10 |
| **$I^2$ = 0.35, $Tau^2$ = 0.00** | | | | **3129** | **3210** | **−0.02 (−0.04, 0.00)** | |
| **Average SQ−LNS compliance − High** | | | | | | | |
| Bangladesh | JiVitA−4 (21) | | | | | | |
| Bangladesh | RDNS (22) | | DMC | 1657 | 815 | −0.02 (−0.04, 0.01) | 0.25 |
| Bangladesh | WASH−B (23) | | EASQ | 1066 | 3266 | −0.02 (−0.04, 0.00) | 0.28 |
| Burkina Faso | iLiNS−Zinc (24) | | DMC | 746 | 375 | −0.10 (−0.14, −0.06) | 0.19 |
| Ghana | GHANA (25) | | | | | | |
| Haiti | HAITI (27) | | | | | | |
| Kenya | WASH−B (28) | | EASQ | 1362 | 4745 | −0.01 (−0.03, 0.00) | 0.28 |
| **$I^2$ = 0.78, $Tau^2$ = 0.00** | | | | **4831** | **9201** | **−0.03 (−0.07, 0.00)** | |

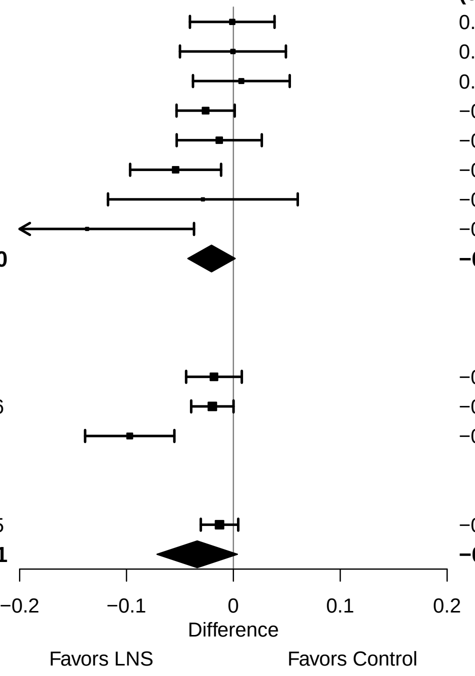

### Supplemental figure 4G: Mean difference in motor z-score

#### 4G1: Stratified by Geographic region

**Geographic region**
**(p−diff = 0.591)**

| Country | Trial | Tool | N | N | MD (95% CI) | W |
|---|---|---|---|---|---|---|
| **Geographic region − SEAR** | | | | | | |
| Bangladesh | JiVitA−4 (21) | BSID−III | 445 | 143 | −0.02 (−0.17, 0.14) | 0.25 |
| Bangladesh | RDNS (22) | DMC | 1556 | 753 | 0.10 (0.03, 0.17) | 0.38 |
| Bangladesh | WASH−B (23) | EASQ | 1074 | 3279 | 0.10 (0.04, 0.17) | 0.38 |
| **I² = 0.05, Tau² = 0.00** | | | **3075** | **4175** | **0.09 (0.04, 0.14)** | |
| **Geographic region − AFR** | | | | | | |
| Burkina Faso | iLiNS−Zinc (24) | DMC | 746 | 375 | 0.40 (0.24, 0.57) | 0.10 |
| Ghana | GHANA (25) | | | | | |
| Ghana | iLiNS−DYADG (26) | KDI | 302 | 601 | 0.03 (−0.11, 0.16) | 0.12 |
| Kenya | WASH−B (28) | EASQ | 1362 | 4745 | 0.01 (−0.05, 0.07) | 0.17 |
| Madagascar | MAHAY (29) | ASQI | 1613 | 1604 | 0.07 (−0.13, 0.27) | 0.08 |
| Malawi | iLiNS−DYADM (30) | KDI | 214 | 436 | 0.03 (−0.13, 0.18) | 0.11 |
| Malawi | iLiNS−DOSE (31) | KDI | 646 | 221 | 0.05 (−0.10, 0.20) | 0.11 |
| Mali | PROMIS CS (32) | DMC | 902 | 921 | 0.13 (−0.04, 0.30) | 0.10 |
| Zimbabwe | SHINE¹ (HIV−) (33), LNS vs SOC | MDAT | 395 | 382 | 0.00 (−0.16, 0.15) | 0.14 |
| Zimbabwe | LNS+WSH vs WSH | MDAT | 446 | 417 | 0.20 (0.05, 0.35) | 0.14 |
| Zimbabwe | SHINE¹ (HIV+) (34), LNS vs SOC | MDAT | 67 | 68 | 0.04 (−0.30, 0.39) | 0.08 |
| Zimbabwe | LNS+WSH vs WSH | MDAT | 103 | 83 | 0.49 (0.23, 0.74) | 0.08 |
| **I² = 0.68, Tau² = 0.01** | | | **6796** | **9853** | **0.11 (0.03, 0.20)** | |

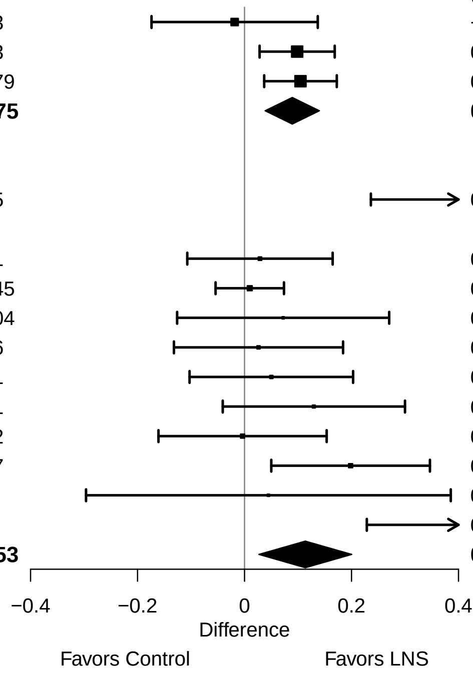

#### Supplemental figure 4G: Mean difference in motor z-score

### 4G2: Stratified by Stunting burden

**Stunting burden**
**(p−diff = 0.045)**

| Country | Trial | Tool | N | N | MD (95% CI) | W |
|---|---|---|---|---|---|---|
| **Stunting burden − Less than 35%** | | | | | | |
| Ghana | GHANA (25) | | | | | |
| Ghana | iLiNS−DYADG (26) | KDI | 302 | 601 | 0.03 (−0.11, 0.16) | 0.30 |
| Haiti | HAITI (27) | | | | | |
| Kenya | WASH−B (28) | EASQ | 1362 | 4745 | 0.01 (−0.05, 0.07) | 0.43 |
| Malawi | iLiNS−DYADM (30) | KDI | 214 | 436 | 0.03 (−0.13, 0.18) | 0.27 |
| **$I^2$ = 0.00, $Tau^2$ = 0.00** | | | **1878** | **5782** | **0.01 (−0.04, 0.07)** | |
| **Stunting burden − More than 35%** | | | | | | |
| Bangladesh | JiVitA−4 (21) | BSID−III | 445 | 143 | −0.02 (−0.17, 0.14) | 0.10 |
| Bangladesh | RDNS (22) | DMC | 1556 | 753 | 0.10 (0.03, 0.17) | 0.16 |
| Bangladesh | WASH−B (23) | EASQ | 1074 | 3279 | 0.10 (0.04, 0.17) | 0.16 |
| Burkina Faso | iLiNS−Zinc (24) | DMC | 746 | 375 | 0.40 (0.24, 0.57) | 0.10 |
| Madagascar | MAHAY (29) | ASQI | 1613 | 1604 | 0.07 (−0.13, 0.27) | 0.08 |
| Malawi | iLiNS−DOSE (31) | KDI | 646 | 221 | 0.05 (−0.10, 0.20) | 0.10 |
| Mali | PROMIS CS (32) | DMC | 902 | 921 | 0.13 (−0.04, 0.30) | 0.09 |
| Zimbabwe | SHINE[1] (HIV−) (33), LNS vs SOC | MDAT | 395 | 382 | 0.00 (−0.16, 0.15) | 0.13 |
| Zimbabwe | LNS+WSH vs WSH | MDAT | 446 | 417 | 0.20 (0.05, 0.35) | 0.13 |
| Zimbabwe | SHINE[1] (HIV+) (34), LNS vs SOC | MDAT | 67 | 68 | 0.04 (−0.30, 0.39) | 0.07 |
| Zimbabwe | LNS+WSH vs WSH | MDAT | 103 | 83 | 0.49 (0.23, 0.74) | 0.07 |
| **$I^2$ = 0.57, $Tau^2$ = 0.01** | | | **7993** | **8246** | **0.13 (0.05, 0.21)** | |

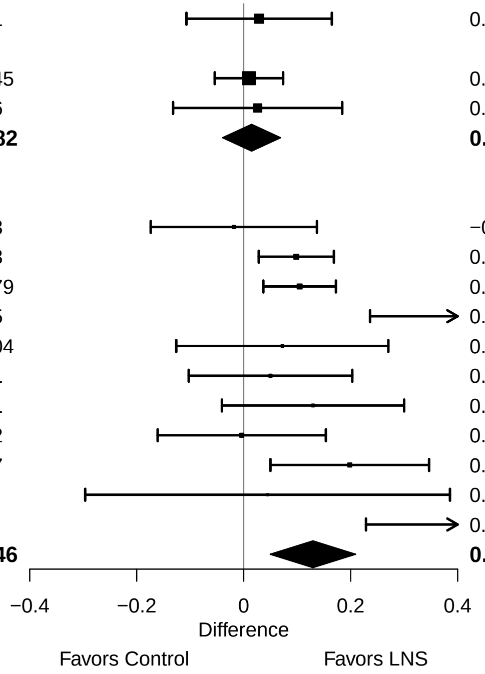

#### Supplemental figure 4G: Mean difference in motor z-score

### 4G3: Stratified by Malaria prevalence

**Malaria prevalence**
**(p−diff = 0.557)**

**Malaria prevalence − Less than 10%**

| Country | Trial | Tool | N | N | MD (95% CI) | W |
|---|---|---|---|---|---|---|
| Bangladesh | JiVitA−4 (21) | BSID−III | 445 | 143 | −0.02 (−0.17, 0.14) | 0.12 |
| Bangladesh | RDNS (22) | DMC | 1556 | 753 | 0.10 (0.03, 0.17) | 0.18 |
| Bangladesh | WASH−B (23) | EASQ | 1074 | 3279 | 0.10 (0.04, 0.17) | 0.18 |
| Haiti | HAITI (27) | | | | | |
| Kenya | WASH−B (28) | EASQ | 1362 | 4745 | 0.01 (−0.05, 0.07) | 0.19 |
| Madagascar | MAHAY (29) | ASQI | 1613 | 1604 | 0.07 (−0.13, 0.27) | 0.09 |
| Zimbabwe | SHINE¹ (HIV−) (33), LNS vs SOC | MDAT | 395 | 382 | 0.00 (−0.16, 0.15) | 0.15 |
| Zimbabwe | LNS+WSH vs WSH | MDAT | 446 | 417 | 0.20 (0.05, 0.35) | 0.15 |
| Zimbabwe | SHINE¹ (HIV+) (34), LNS vs SOC | MDAT | 67 | 68 | 0.04 (−0.30, 0.39) | 0.08 |
| Zimbabwe | LNS+WSH vs WSH | MDAT | 103 | 83 | 0.49 (0.23, 0.74) | 0.08 |
| **I² = 0.47, Tau² = 0.00** | | | **7061** | **11474** | **0.08 (0.02, 0.14)** | |

**Malaria prevalence − At least 10%**

| Country | Trial | Tool | N | N | MD (95% CI) | W |
|---|---|---|---|---|---|---|
| Burkina Faso | iLiNS−Zinc (24) | DMC | 746 | 375 | 0.40 (0.24, 0.57) | 0.19 |
| Ghana | GHANA (25) | | | | | |
| Ghana | iLiNS−DYADG (26) | KDI | 302 | 601 | 0.03 (−0.11, 0.16) | 0.22 |
| Malawi | iLiNS−DYADM (30) | KDI | 214 | 436 | 0.03 (−0.13, 0.18) | 0.20 |
| Malawi | iLiNS−DOSE (31) | KDI | 646 | 221 | 0.05 (−0.10, 0.20) | 0.20 |
| Mali | PROMIS CS (32) | DMC | 902 | 921 | 0.13 (−0.04, 0.30) | 0.18 |
| **I² = 0.73, Tau² = 0.02** | | | **2810** | **2554** | **0.12 (−0.01, 0.26)** | |

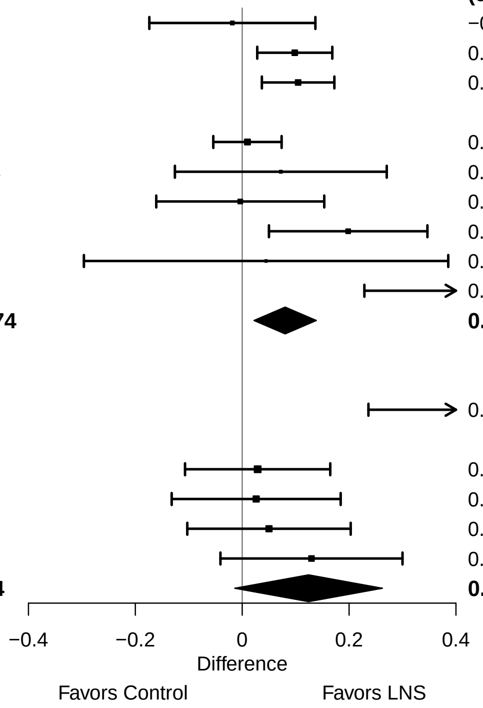

##### Supplemental figure 4G: Mean difference in motor z-score

#### 4G4: Stratified by Anemia prevalence

**Anemia prevalence**
**(p−diff = 0.269)**

**Anemia prevalence − High**

| Country | Trial | Tool | N | N | MD (95% CI) | W |
|---|---|---|---|---|---|---|
| Burkina Faso | iLiNS−Zinc (24) | DMC | 746 | 375 | 0.40 (0.24, 0.57) | 0.25 |
| Ghana | GHANA (25) | | | | | |
| Haiti | HAITI (27) | | | | | |
| Malawi | iLiNS−DYADM (30) | KDI | 214 | 436 | 0.03 (−0.13, 0.18) | 0.25 |
| Malawi | iLiNS−DOSE (31) | KDI | 646 | 221 | 0.05 (−0.10, 0.20) | 0.26 |
| Mali | PROMIS CS (32) | DMC | 902 | 921 | 0.13 (−0.04, 0.30) | 0.24 |
| **I² = 0.77, Tau² = 0.02** | | | **2508** | **1953** | **0.15 (−0.02, 0.32)** | |

**Anemia prevalence − Moderate**

| Country | Trial | Tool | N | N | MD (95% CI) | W |
|---|---|---|---|---|---|---|
| Bangladesh | JiVitA−4 (21) | BSID−III | 445 | 143 | −0.02 (−0.17, 0.14) | 0.10 |
| Bangladesh | RDNS (22) | DMC | 1556 | 753 | 0.10 (0.03, 0.17) | 0.16 |
| Bangladesh | WASH−B (23) | EASQ | 1074 | 3279 | 0.10 (0.04, 0.17) | 0.16 |
| Ghana | iLiNS−DYADG (26) | KDI | 302 | 601 | 0.03 (−0.11, 0.16) | 0.12 |
| Kenya | WASH−B (28) | EASQ | 1362 | 4745 | 0.01 (−0.05, 0.07) | 0.16 |
| Madagascar | MAHAY (29) | ASQI | 1613 | 1604 | 0.07 (−0.13, 0.27) | 0.08 |
| Zimbabwe | SHINE[1] (HIV−) (33), LNS vs SOC | MDAT | 395 | 382 | 0.00 (−0.16, 0.15) | 0.13 |
| Zimbabwe | LNS+WSH vs WSH | MDAT | 446 | 417 | 0.20 (0.05, 0.35) | 0.13 |
| Zimbabwe | SHINE[1] (HIV+) (34), LNS vs SOC | MDAT | 67 | 68 | 0.04 (−0.30, 0.39) | 0.07 |
| Zimbabwe | LNS+WSH vs WSH | MDAT | 103 | 83 | 0.49 (0.23, 0.74) | 0.07 |
| **I² = 0.40, Tau² = 0.00** | | | **7363** | **12075** | **0.07 (0.02, 0.13)** | |

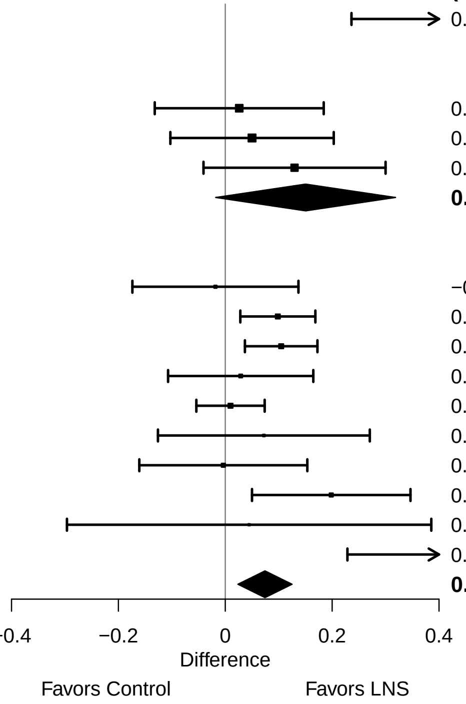

#### Supplemental figure 4G: Mean difference in motor z-score

### 4G5: Stratified by Source water quality

**Source water quality**
**(p−diff = 0.530)**

**Source water quality − Improved**

| Country | Trial | Tool | N | N | MD (95% CI) | W |
|---|---|---|---|---|---|---|
| Bangladesh | JiVitA−4 (21) | BSID−III | 445 | 143 | −0.02 (−0.17, 0.14) | 0.14 |
| Bangladesh | RDNS (22) | DMC | 1556 | 753 | 0.10 (0.03, 0.17) | 0.23 |
| Bangladesh | WASH−B (23) | EASQ | 528 | 1099 | 0.19 (0.09, 0.30) | 0.19 |
| Ghana | GHANA (25) | | | | | |
| Ghana | iLiNS−DYADG (26) | KDI | 302 | 601 | 0.03 (−0.11, 0.16) | 0.16 |
| Haiti | HAITI (27) | | | | | |
| Malawi | iLiNS−DYADM (30) | KDI | 214 | 436 | 0.03 (−0.13, 0.18) | 0.14 |
| Malawi | iLiNS−DOSE (31) | KDI | 646 | 221 | 0.05 (−0.10, 0.20) | 0.14 |
| **I² = 0.32, Tau² = 0.00** | | | **3691** | **3253** | **0.08 (0.02, 0.14)** | |

**Source water quality − Unimproved**

| Country | Trial | Tool | N | N | MD (95% CI) | W |
|---|---|---|---|---|---|---|
| Burkina Faso | iLiNS−Zinc (24) | DMC | 746 | 375 | 0.40 (0.24, 0.57) | 0.18 |
| Kenya | WASH−B (28) | EASQ | 649 | 2073 | 0.03 (−0.06, 0.13) | 0.26 |
| Madagascar | MAHAY (29) | ASQI | 1613 | 1604 | 0.07 (−0.13, 0.27) | 0.14 |
| Mali | PROMIS CS (32) | DMC | 902 | 921 | 0.13 (−0.04, 0.30) | 0.17 |
| Zimbabwe | SHINE (HIV−) (33) | MDAT | 395 | 382 | 0.00 (−0.16, 0.15) | 0.18 |
| Zimbabwe | SHINE (HIV+) (34) | MDAT | 67 | 68 | 0.04 (−0.30, 0.39) | 0.07 |
| **I² = 0.70, Tau² = 0.01** | | | **4372** | **5423** | **0.12 (−0.01, 0.24)** | |

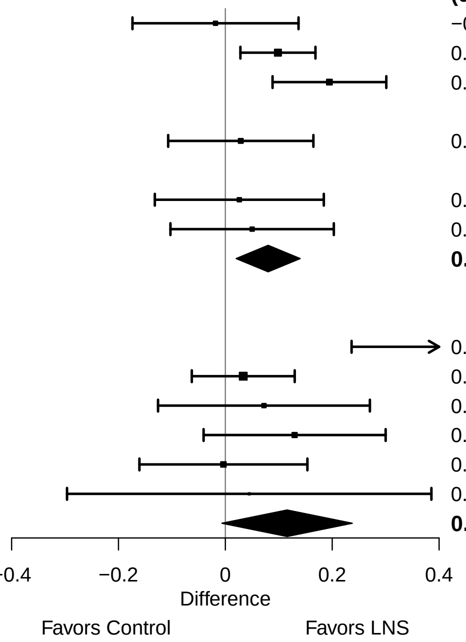

#### Supplemental figure 4G: Mean difference in motor z-score

### 4G6: Stratified by Sanitation

**Sanitation**
**(p−diff = 0.963)**

| Country | Trial | Tool | N | N | MD (95% CI) | W |
|---|---|---|---|---|---|---|
| **Sanitation − Improved** | | | | | | |
| Bangladesh | JiVitA−4 (21) | BSID−III | 445 | 143 | −0.02 (−0.17, 0.14) | 0.17 |
| Bangladesh | RDNS (22) | DMC | 1556 | 753 | 0.10 (0.03, 0.17) | 0.27 |
| Bangladesh | WASH−B (23) | EASQ | 528 | 1099 | 0.19 (0.09, 0.30) | 0.22 |
| Ghana | GHANA (25) | | | | | |
| Ghana | iLiNS−DYADG (26) | KDI | 302 | 601 | 0.03 (−0.11, 0.16) | 0.19 |
| Haiti | HAITI (27) | | | | | |
| Mali | PROMIS CS (32) | DMC | 902 | 921 | 0.13 (−0.04, 0.30) | 0.15 |
| **$I^2$ = 0.38, $Tau^2$ = 0.00** | | | **3733** | **3517** | **0.10 (0.03, 0.17)** | |
| **Sanitation − Unimproved** | | | | | | |
| Burkina Faso | iLiNS−Zinc (24) | DMC | 746 | 375 | 0.40 (0.24, 0.57) | 0.15 |
| Kenya | WASH−B (28) | EASQ | 649 | 2073 | 0.03 (−0.06, 0.13) | 0.22 |
| Madagascar | MAHAY (29) | ASQI | 1613 | 1604 | 0.07 (−0.13, 0.27) | 0.12 |
| Malawi | iLiNS−DYADM (30) | KDI | 214 | 436 | 0.03 (−0.13, 0.18) | 0.15 |
| Malawi | iLiNS−DOSE (31) | KDI | 646 | 221 | 0.05 (−0.10, 0.20) | 0.16 |
| Zimbabwe | SHINE (HIV−) (33) | MDAT | 395 | 382 | 0.00 (−0.16, 0.15) | 0.15 |
| Zimbabwe | SHINE (HIV+) (34) | MDAT | 67 | 68 | 0.04 (−0.30, 0.39) | 0.05 |
| **$I^2$ = 0.65, $Tau^2$ = 0.01** | | | **4330** | **5159** | **0.09 (−0.02, 0.19)** | |

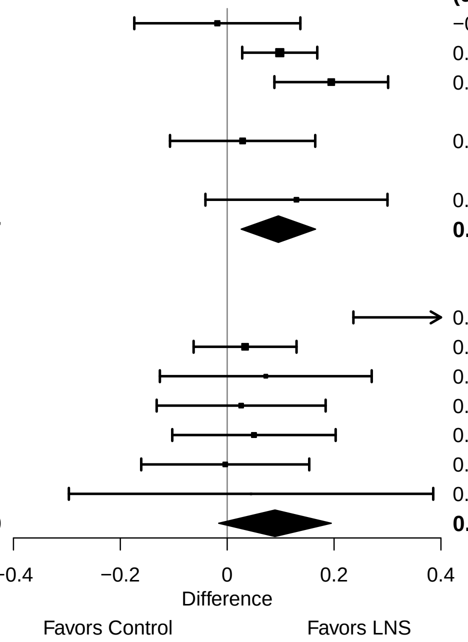

#### Supplemental figure 4G: Mean difference in motor z-score

### 4G7: Stratified by Supplement duration

**Supplement duration**
**(p−diff = 0.621)**

| Country | Trial | Tool | N | N | MD (95% CI) | W |
|---|---|---|---|---|---|---|
| **Supplement duration − 12m or less** | | | | | | |
| Bangladesh | JiVitA−4 (21) | BSID−III | 445 | 143 | −0.02 (−0.17, 0.14) | 0.13 |
| Burkina Faso | iLiNS−Zinc (24) | DMC | 746 | 375 | 0.40 (0.24, 0.57) | 0.12 |
| Ghana | GHANA (25) | | | | | |
| Ghana | iLiNS−DYADG (26) | KDI | 302 | 601 | 0.03 (−0.11, 0.16) | 0.14 |
| Haiti | HAITI (27) | | | | | |
| Madagascar | MAHAY (29) | ASQI | 1613 | 1604 | 0.07 (−0.13, 0.27) | 0.10 |
| Malawi | iLiNS−DYADM (30) | KDI | 214 | 436 | 0.03 (−0.13, 0.18) | 0.13 |
| Malawi | iLiNS−DOSE (31) | KDI | 646 | 221 | 0.05 (−0.10, 0.20) | 0.13 |
| Zimbabwe | SHINE[1] (HIV−) (33), LNS vs SOC | MDAT | 395 | 382 | 0.00 (−0.16, 0.15) | 0.16 |
| Zimbabwe | LNS+WSH vs WSH | MDAT | 446 | 417 | 0.20 (0.05, 0.35) | 0.16 |
| Zimbabwe | SHINE[1] (HIV+) (34), LNS vs SOC | MDAT | 67 | 68 | 0.04 (−0.30, 0.39) | 0.09 |
| Zimbabwe | LNS+WSH vs WSH | MDAT | 103 | 83 | 0.49 (0.23, 0.74) | 0.09 |
| **$I^2$ = 0.67, $Tau^2$ = 0.01** | | | **4977** | **4330** | **0.11 (0.01, 0.22)** | |
| **Supplement duration − > 12m** | | | | | | |
| Bangladesh | RDNS (22) | DMC | 1556 | 753 | 0.10 (0.03, 0.17) | 0.28 |
| Bangladesh | WASH−B (23) | EASQ | 1074 | 3279 | 0.10 (0.04, 0.17) | 0.28 |
| Kenya | WASH−B (28) | EASQ | 1362 | 4745 | 0.01 (−0.05, 0.07) | 0.28 |
| Mali | PROMIS CS (32) | DMC | 902 | 921 | 0.13 (−0.04, 0.30) | 0.17 |
| **$I^2$ = 0.45, $Tau^2$ = 0.00** | | | **4894** | **9698** | **0.07 (0.02, 0.13)** | |

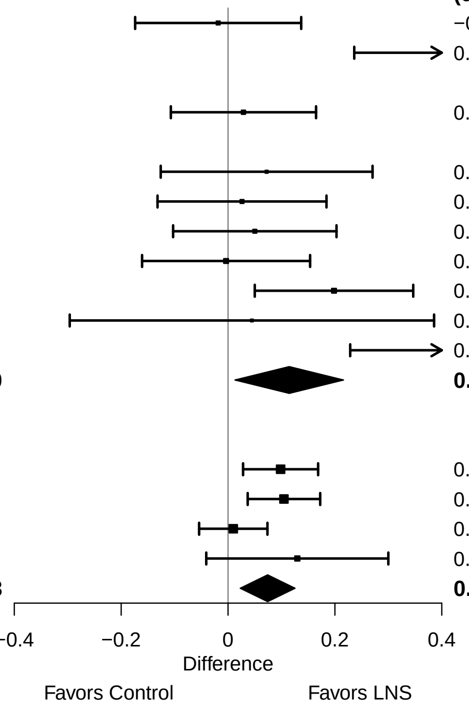

#### Supplemental figure 4G: Mean difference in motor z-score

### 4G8: Stratified by Frequency of contact

**Frequency of contact**
**(p−diff = 0.932)**

| Country | Trial | | Tool | N | N | MD (95% CI) | W |
|---|---|---|---|---|---|---|---|
| **Frequency of contact − Monthly** | | | | | | | |
| Bangladesh | RDNS (22) | | DMC | 1556 | 753 | 0.10 (0.03, 0.17) | 0.23 |
| Haiti | HAITI (27) | | | | | | |
| Kenya | WASH−B (28) | | EASQ | 1362 | 4745 | 0.01 (−0.05, 0.07) | 0.23 |
| Madagascar | MAHAY (29) | | ASQI | 1613 | 1604 | 0.07 (−0.13, 0.27) | 0.12 |
| Mali | PROMIS CS (32) | | DMC | 902 | 921 | 0.13 (−0.04, 0.30) | 0.14 |
| Zimbabwe | SHINE¹ (HIV−) (33), | LNS vs SOC | MDAT | 395 | 382 | 0.00 (−0.16, 0.15) | 0.19 |
| Zimbabwe | | LNS+WSH vs WSH | MDAT | 446 | 417 | 0.20 (0.05, 0.35) | 0.19 |
| Zimbabwe | SHINE¹ (HIV+) (34), | LNS vs SOC | MDAT | 67 | 68 | 0.04 (−0.30, 0.39) | 0.10 |
| Zimbabwe | | LNS+WSH vs WSH | MDAT | 103 | 83 | 0.49 (0.23, 0.74) | 0.10 |
| **$I^2$ = 0.47, $Tau^2$ = 0.00** | | | | **6444** | **8973** | **0.09 (0.03, 0.16)** | |
| **Frequency of contact − Weekly** | | | | | | | |
| Bangladesh | JiVitA−4 (21) | | BSID−III | 445 | 143 | −0.02 (−0.17, 0.14) | 0.15 |
| Bangladesh | WASH−B (23) | | EASQ | 1074 | 3279 | 0.10 (0.04, 0.17) | 0.23 |
| Burkina Faso | iLiNS−Zinc (24) | | DMC | 746 | 375 | 0.40 (0.24, 0.57) | 0.14 |
| Ghana | GHANA (25) | | | | | | |
| Ghana | iLiNS−DYADG (26) | | KDI | 302 | 601 | 0.03 (−0.11, 0.16) | 0.17 |
| Malawi | iLiNS−DYADM (30) | | KDI | 214 | 436 | 0.03 (−0.13, 0.18) | 0.15 |
| Malawi | iLiNS−DOSE (31) | | KDI | 646 | 221 | 0.05 (−0.10, 0.20) | 0.15 |
| **$I^2$ = 0.71, $Tau^2$ = 0.02** | | | | **3427** | **5055** | **0.10 (−0.02, 0.21)** | |

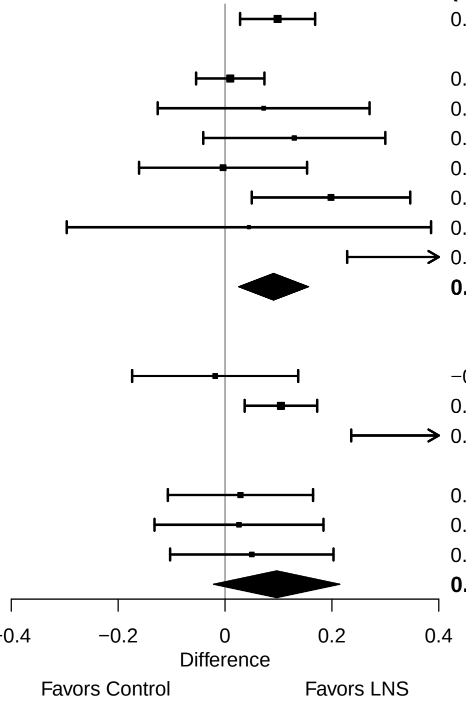

#### Supplemental figure 4G: Mean difference in motor z-score

### 4G9: Stratified by Average SQ-LNS compliance

**Average SQ−LNS compliance**
**(p−diff = 0.898)**

**Average SQ−LNS compliance − Low**

| Country | Trial | Tool | N | N | MD (95% CI) | W |
|---|---|---|---|---|---|---|
| Ghana | iLiNS−DYADG (26) | KDI | 302 | 601 | 0.03 (−0.11, 0.16) | 0.18 |
| Malawi | iLiNS−DYADM (30) | KDI | 214 | 436 | 0.03 (−0.13, 0.18) | 0.16 |
| Malawi | iLiNS−DOSE (31) | KDI | 646 | 221 | 0.05 (−0.10, 0.20) | 0.17 |
| Mali | PROMIS CS (32) | DMC | 902 | 921 | 0.13 (−0.04, 0.30) | 0.15 |
| Zimbabwe | SHINE¹ (HIV−) (33), LNS vs SOC | MDAT | 395 | 382 | 0.00 (−0.16, 0.15) | 0.21 |
| Zimbabwe | LNS+WSH vs WSH | MDAT | 446 | 417 | 0.20 (0.05, 0.35) | 0.21 |
| Zimbabwe | SHINE¹ (HIV+) (34), LNS vs SOC | MDAT | 67 | 68 | 0.04 (−0.30, 0.39) | 0.12 |
| Zimbabwe | LNS+WSH vs WSH | MDAT | 103 | 83 | 0.49 (0.23, 0.74) | 0.12 |
| **I² = 0.14, Tau² = 0.00** | | | **3075** | **3129** | **0.09 (0.02, 0.16)** | |

**Average SQ−LNS compliance − High**

| Country | Trial | Tool | N | N | MD (95% CI) | W |
|---|---|---|---|---|---|---|
| Bangladesh | JiVitA−4 (21) | BSID−III | 445 | 143 | −0.02 (−0.17, 0.14) | 0.16 |
| Bangladesh | RDNS (22) | DMC | 1556 | 753 | 0.10 (0.03, 0.17) | 0.23 |
| Bangladesh | WASH−B (23) | EASQ | 1074 | 3279 | 0.10 (0.04, 0.17) | 0.23 |
| Burkina Faso | iLiNS−Zinc (24) | DMC | 746 | 375 | 0.40 (0.24, 0.57) | 0.15 |
| Ghana | GHANA (25) | | | | | |
| Haiti | HAITI (27) | | | | | |
| Kenya | WASH−B (28) | EASQ | 1362 | 4745 | 0.01 (−0.05, 0.07) | 0.23 |
| **I² = 0.81, Tau² = 0.02** | | | **5183** | **9295** | **0.11 (−0.03, 0.25)** | |

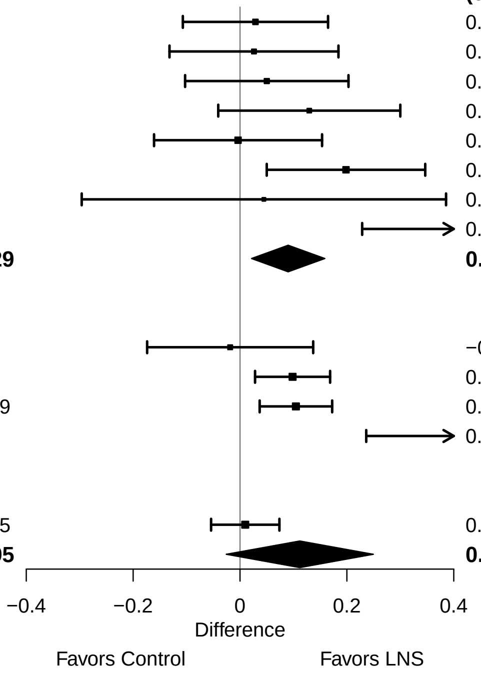

### Supplemental figure 4H: Motor lowest decile prevalence ratio

#### 4H1: Stratified by Geographic region

**Geographic region**
**(p−diff = 0.610)**

| Country | Trial | | Tool | N | N | PR (95% CI) | W |
|---|---|---|---|---|---|---|---|
| **Geographic region − SEAR** | | | | | | | |
| Bangladesh | JiVitA−4 (21) | | BSID−III | 445 | 143 | 1.26 (0.77, 2.07) | 0.17 |
| Bangladesh | RDNS (22) | | DMC | 1556 | 753 | 0.85 (0.67, 1.07) | 0.40 |
| Bangladesh | WASH−B (23) | | EASQ | 1074 | 3279 | 0.82 (0.66, 1.01) | 0.43 |
| **I² = 0.20, Tau² = 0.01** | | | | **3075** | **4175** | **0.88 (0.72, 1.08)** | |
| **Geographic region − AFR** | | | | | | | |
| Burkina Faso | iLiNS−Zinc (24) | | DMC | 746 | 375 | 0.49 (0.36, 0.67) | 0.14 |
| Ghana | GHANA (25) | | | | | | |
| Ghana | iLiNS−DYADG (26) | | KDI | 302 | 601 | 1.13 (0.76, 1.70) | 0.10 |
| Kenya | WASH−B (28) | | EASQ | 1362 | 4745 | 0.86 (0.70, 1.05) | 0.21 |
| Madagascar | MAHAY (29) | | ASQI | 1613 | 1604 | 0.91 (0.58, 1.44) | 0.09 |
| Malawi | iLiNS−DYADM (30) | | KDI | 214 | 436 | 0.84 (0.51, 1.40) | 0.07 |
| Malawi | iLiNS−DOSE (31) | | KDI | 646 | 221 | 1.08 (0.67, 1.71) | 0.08 |
| Mali | PROMIS CS (32) | | DMC | 902 | 921 | 0.81 (0.56, 1.18) | 0.11 |
| Zimbabwe | SHINE¹ (HIV−) (33), | LNS vs SOC | MDAT | 395 | 382 | 1.08 (0.67, 1.73) | 0.14 |
| Zimbabwe | | LNS+WSH vs WSH | MDAT | 446 | 417 | 0.82 (0.54, 1.24) | 0.14 |
| Zimbabwe | SHINE¹ (HIV+) (34), | LNS vs SOC | MDAT | 67 | 68 | 1.16 (0.44, 3.08) | 0.05 |
| Zimbabwe | | LNS+WSH vs WSH | MDAT | 103 | 83 | 0.40 (0.17, 0.95) | 0.05 |
| **I² = 0.48, Tau² = 0.03** | | | | **6796** | **9853** | **0.83 (0.70, 0.98)** | |

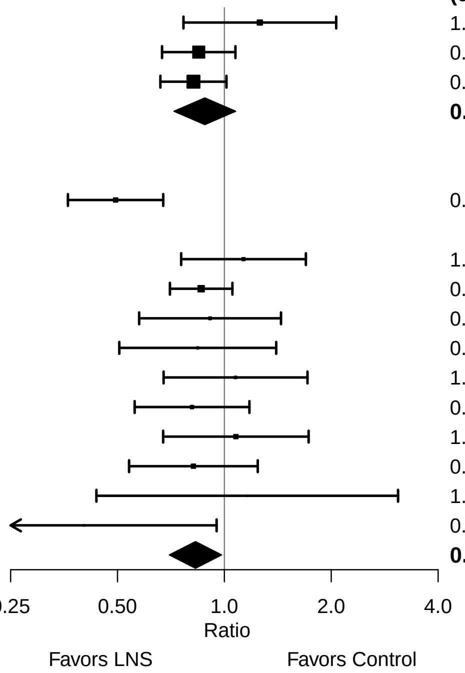

#### Supplemental figure 4H: Motor lowest decile prevalence ratio

### 4H2: Stratified by Stunting burden

**Stunting burden**
**(p−diff = 0.466)**

| Country | Trial | Tool | N | N | PR (95% CI) | W |
|---|---|---|---|---|---|---|
| **Stunting burden − Less than 35%** | | | | | | |
| Ghana | GHANA (25) | | | | | |
| Ghana | iLiNS−DYADG (26) | KDI | 302 | 601 | 1.13 (0.76, 1.70) | 0.27 |
| Haiti | HAITI (27) | | | | | |
| Kenya | WASH−B (28) | EASQ | 1362 | 4745 | 0.86 (0.70, 1.05) | 0.54 |
| Malawi | iLiNS−DYADM (30) | KDI | 214 | 436 | 0.84 (0.51, 1.40) | 0.19 |
| **I² = 0.00, Tau² = 0.00** | | | **1878** | **5782** | **0.90 (0.76, 1.07)** | |
| **Stunting burden − More than 35%** | | | | | | |
| Bangladesh | JiVitA−4 (21) | BSID−III | 445 | 143 | 1.26 (0.77, 2.07) | 0.07 |
| Bangladesh | RDNS (22) | DMC | 1556 | 753 | 0.85 (0.67, 1.07) | 0.17 |
| Bangladesh | WASH−B (23) | EASQ | 1074 | 3279 | 0.82 (0.66, 1.01) | 0.18 |
| Burkina Faso | iLiNS−Zinc (24) | DMC | 746 | 375 | 0.49 (0.36, 0.67) | 0.13 |
| Madagascar | MAHAY (29) | ASQI | 1613 | 1604 | 0.91 (0.58, 1.44) | 0.08 |
| Malawi | iLiNS−DOSE (31) | KDI | 646 | 221 | 1.08 (0.67, 1.71) | 0.08 |
| Mali | PROMIS CS (32) | DMC | 902 | 921 | 0.81 (0.56, 1.18) | 0.11 |
| Zimbabwe | SHINE[1] (HIV−) (33), LNS vs SOC | MDAT | 395 | 382 | 1.08 (0.67, 1.73) | 0.13 |
| Zimbabwe | LNS+WSH vs WSH | MDAT | 446 | 417 | 0.82 (0.54, 1.24) | 0.13 |
| Zimbabwe | SHINE[1] (HIV+) (34), LNS vs SOC | MDAT | 67 | 68 | 1.16 (0.44, 3.08) | 0.05 |
| Zimbabwe | LNS+WSH vs WSH | MDAT | 103 | 83 | 0.40 (0.17, 0.95) | 0.05 |
| **I² = 0.49, Tau² = 0.03** | | | **7993** | **8246** | **0.83 (0.70, 0.98)** | |

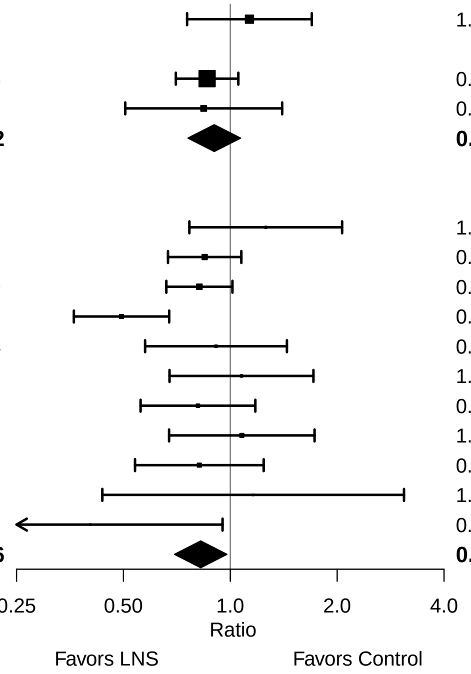

#### Supplemental figure 4H: Motor lowest decile prevalence ratio

#### 4H3: Stratified by Malaria prevalence

**Malaria prevalence**
**(p−diff = 0.491)**

**Malaria prevalence − Less than 10%**

| **Country** | **Trial** | **Tool** | **N** | **N** | **PR (95% CI)** | **W** |
|---|---|---|---|---|---|---|
| Bangladesh | JiVitA−4 (21) | BSID−III | 445 | 143 | 1.26 (0.77, 2.07) | 0.08 |
| Bangladesh | RDNS (22) | DMC | 1556 | 753 | 0.85 (0.67, 1.07) | 0.19 |
| Bangladesh | WASH−B (23) | EASQ | 1074 | 3279 | 0.82 (0.66, 1.01) | 0.21 |
| Haiti | HAITI (27) | | | | | |
| Kenya | WASH−B (28) | EASQ | 1362 | 4745 | 0.86 (0.70, 1.05) | 0.22 |
| Madagascar | MAHAY (29) | ASQI | 1613 | 1604 | 0.91 (0.58, 1.44) | 0.09 |
| Zimbabwe | SHINE[1] (HIV−) (33), LNS vs SOC | MDAT | 395 | 382 | 1.08 (0.67, 1.73) | 0.15 |
| Zimbabwe | LNS+WSH vs WSH | MDAT | 446 | 417 | 0.82 (0.54, 1.24) | 0.15 |
| Zimbabwe | SHINE[1] (HIV+) (34), LNS vs SOC | MDAT | 67 | 68 | 1.16 (0.44, 3.08) | 0.05 |
| Zimbabwe | LNS+WSH vs WSH | MDAT | 103 | 83 | 0.40 (0.17, 0.95) | 0.05 |
| **I² = 0.00, Tau² = 0.00** | | | **7061** | **11474** | **0.87 (0.78, 0.97)** | |

**Malaria prevalence − At least 10%**

| | | | | | | |
|---|---|---|---|---|---|---|
| Burkina Faso | iLiNS−Zinc (24) | DMC | 746 | 375 | 0.49 (0.36, 0.67) | 0.27 |
| Ghana | GHANA (25) | | | | | |
| Ghana | iLiNS−DYADG (26) | KDI | 302 | 601 | 1.13 (0.76, 1.70) | 0.20 |
| Malawi | iLiNS−DYADM (30) | KDI | 214 | 436 | 0.84 (0.51, 1.40) | 0.14 |
| Malawi | iLiNS−DOSE (31) | KDI | 646 | 221 | 1.08 (0.67, 1.71) | 0.16 |
| Mali | PROMIS CS (32) | DMC | 902 | 921 | 0.81 (0.56, 1.18) | 0.22 |
| **I² = 0.71, Tau² = 0.08** | | | **2810** | **2554** | **0.82 (0.60, 1.10)** | |

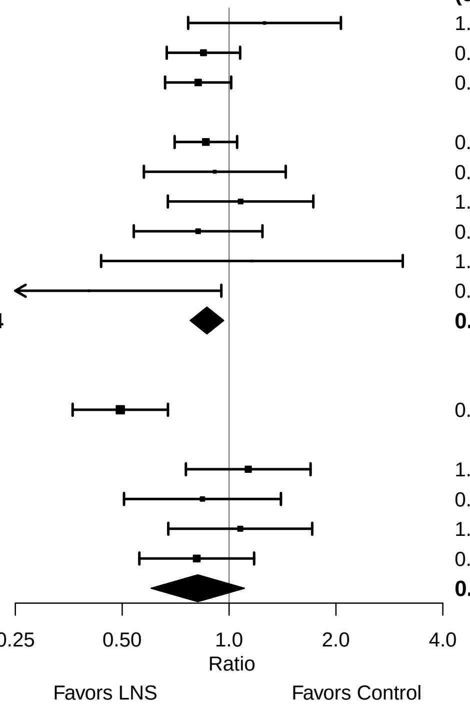

#### Supplemental figure 4H: Motor lowest decile prevalence ratio

### 4H4: Stratified by Anemia prevalence

**Anemia prevalence**
**(p−diff = 0.093)**

| Country | Trial | | Tool | N | N | PR (95% CI) | W |
|---|---|---|---|---|---|---|---|
| **Anemia prevalence − High** | | | | | | | |
| Burkina Faso | iLiNS−Zinc (24) | | DMC | 746 | 375 | 0.49 (0.36, 0.67) | 0.34 |
| Ghana | GHANA (25) | | | | | | |
| Haiti | HAITI (27) | | | | | | |
| Malawi | iLiNS−DYADM (30) | | KDI | 214 | 436 | 0.84 (0.51, 1.40) | 0.18 |
| Malawi | iLiNS−DOSE (31) | | KDI | 646 | 221 | 1.08 (0.67, 1.71) | 0.20 |
| Mali | PROMIS CS (32) | | DMC | 902 | 921 | 0.81 (0.56, 1.18) | 0.28 |
| **I² = 0.68, Tau² = 0.07** | | | | **2508** | **1953** | **0.75 (0.54, 1.05)** | |
| **Anemia prevalence − Moderate** | | | | | | | |
| Bangladesh | JiVitA−4 (21) | | BSID−III | 445 | 143 | 1.26 (0.77, 2.07) | 0.07 |
| Bangladesh | RDNS (22) | | DMC | 1556 | 753 | 0.85 (0.67, 1.07) | 0.18 |
| Bangladesh | WASH−B (23) | | EASQ | 1074 | 3279 | 0.82 (0.66, 1.01) | 0.19 |
| Ghana | iLiNS−DYADG (26) | | KDI | 302 | 601 | 1.13 (0.76, 1.70) | 0.10 |
| Kenya | WASH−B (28) | | EASQ | 1362 | 4745 | 0.86 (0.70, 1.05) | 0.20 |
| Madagascar | MAHAY (29) | | ASQI | 1613 | 1604 | 0.91 (0.58, 1.44) | 0.08 |
| Zimbabwe | SHINE¹ (HIV−) (33), | LNS vs SOC | MDAT | 395 | 382 | 1.08 (0.67, 1.73) | 0.13 |
| Zimbabwe | | LNS+WSH vs WSH | MDAT | 446 | 417 | 0.82 (0.54, 1.24) | 0.13 |
| Zimbabwe | SHINE¹ (HIV+) (34), | LNS vs SOC | MDAT | 67 | 68 | 1.16 (0.44, 3.08) | 0.05 |
| Zimbabwe | | LNS+WSH vs WSH | MDAT | 103 | 83 | 0.40 (0.17, 0.95) | 0.05 |
| **I² = 0.00, Tau² = 0.00** | | | | **7363** | **12075** | **0.88 (0.79, 0.98)** | |

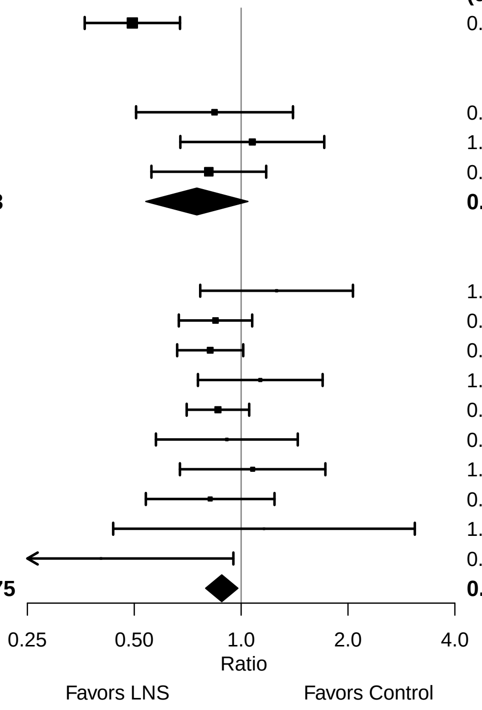

#### Supplemental figure 4H: Motor lowest decile prevalence ratio

### 4H5: Stratified by Source water quality

**Source water quality**
**(p−diff = 0.303)**

| Country | Trial | Tool | N | N | PR (95% CI) | W |
|---|---|---|---|---|---|---|
| **Source water quality − Improved** | | | | | | |
| Bangladesh | JiVitA−4 (21) | BSID−III | 445 | 143 | 1.26 (0.77, 2.07) | 0.12 |
| Bangladesh | RDNS (22) | DMC | 1556 | 753 | 0.85 (0.67, 1.07) | 0.27 |
| Bangladesh | WASH−B (23) | EASQ | 528 | 1099 | 0.81 (0.58, 1.13) | 0.20 |
| Ghana | GHANA (25) | | | | | |
| Ghana | iLiNS−DYADG (26) | KDI | 302 | 601 | 1.13 (0.76, 1.70) | 0.16 |
| Haiti | HAITI (27) | | | | | |
| Malawi | iLiNS−DYADM (30) | KDI | 214 | 436 | 0.84 (0.51, 1.40) | 0.12 |
| Malawi | iLiNS−DOSE (31) | KDI | 646 | 221 | 1.08 (0.67, 1.71) | 0.13 |
| **I² = 0.00, Tau² = 0.00** | | | **3691** | **3253** | **0.93 (0.80, 1.08)** | |
| **Source water quality − Unimproved** | | | | | | |
| Burkina Faso | iLiNS−Zinc (24) | DMC | 746 | 375 | 0.49 (0.36, 0.67) | 0.24 |
| Kenya | WASH−B (28) | EASQ | 649 | 2073 | 0.96 (0.71, 1.31) | 0.24 |
| Madagascar | MAHAY (29) | ASQI | 1613 | 1604 | 0.91 (0.58, 1.44) | 0.15 |
| Mali | PROMIS CS (32) | DMC | 902 | 921 | 0.81 (0.56, 1.18) | 0.19 |
| Zimbabwe | SHINE (HIV−) (33) | MDAT | 395 | 382 | 1.08 (0.67, 1.73) | 0.14 |
| Zimbabwe | SHINE (HIV+) (34) | MDAT | 67 | 68 | 1.16 (0.44, 3.08) | 0.04 |
| **I² = 0.62, Tau² = 0.05** | | | **4372** | **5423** | **0.82 (0.64, 1.05)** | |

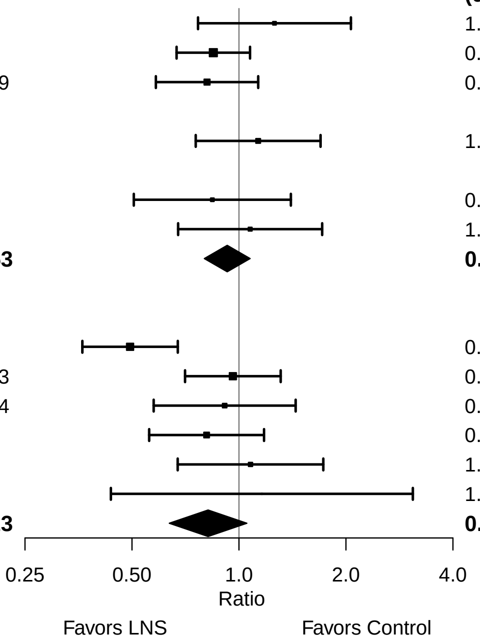

#### Supplemental figure 4H: Motor lowest decile prevalence ratio

### 4H6: Stratified by Sanitation

**Sanitation**
**(p−diff = 0.599)**

| Country | Trial | Tool | N | N | PR (95% CI) | W |
|---|---|---|---|---|---|---|
| **Sanitation − Improved** | | | | | | |
| Bangladesh | JiVitA−4 (21) | BSID−III | 445 | 143 | 1.26 (0.77, 2.07) | 0.13 |
| Bangladesh | RDNS (22) | DMC | 1556 | 753 | 0.85 (0.67, 1.07) | 0.29 |
| Bangladesh | WASH−B (23) | EASQ | 528 | 1099 | 0.81 (0.58, 1.13) | 0.22 |
| Ghana | GHANA (25) | | | | | |
| Ghana | iLiNS−DYADG (26) | KDI | 302 | 601 | 1.13 (0.76, 1.70) | 0.17 |
| Haiti | HAITI (27) | | | | | |
| Mali | PROMIS CS (32) | DMC | 902 | 921 | 0.81 (0.56, 1.18) | 0.19 |
| **I² = 0.00, Tau² = 0.00** | | | **3733** | **3517** | **0.90 (0.77, 1.05)** | |
| **Sanitation − Unimproved** | | | | | | |
| Burkina Faso | iLiNS−Zinc (24) | DMC | 746 | 375 | 0.49 (0.36, 0.67) | 0.22 |
| Kenya | WASH−B (28) | EASQ | 649 | 2073 | 0.96 (0.71, 1.31) | 0.22 |
| Madagascar | MAHAY (29) | ASQI | 1613 | 1604 | 0.91 (0.58, 1.44) | 0.14 |
| Malawi | iLiNS−DYADM (30) | KDI | 214 | 436 | 0.84 (0.51, 1.40) | 0.12 |
| Malawi | iLiNS−DOSE (31) | KDI | 646 | 221 | 1.08 (0.67, 1.71) | 0.13 |
| Zimbabwe | SHINE (HIV−) (33) | MDAT | 395 | 382 | 1.08 (0.67, 1.73) | 0.13 |
| Zimbabwe | SHINE (HIV+) (34) | MDAT | 67 | 68 | 1.16 (0.44, 3.08) | 0.04 |
| **I² = 0.59, Tau² = 0.04** | | | **4330** | **5159** | **0.85 (0.68, 1.08)** | |

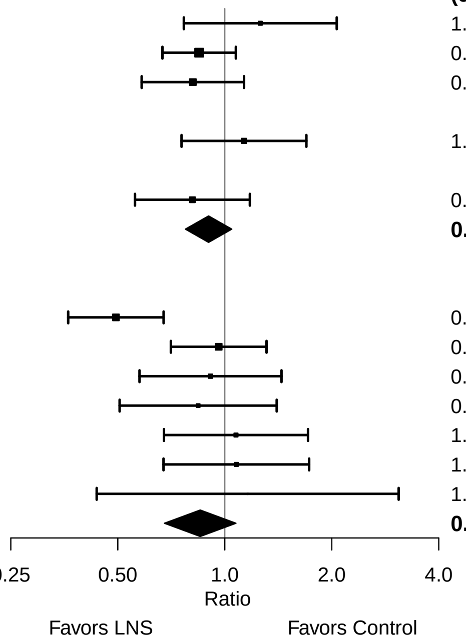

#### Supplemental figure 4H: Motor lowest decile prevalence ratio

### 4H7: Stratified by Supplement duration

**Supplement duration**
**(p−diff = 0.839)**

| Country | Trial | | Tool | N | N | PR (95% CI) | W |
|---|---|---|---|---|---|---|---|
| **Supplement duration − 12m or less** | | | | | | | |
| Bangladesh | JiVitA−4 (21) | | BSID−III | 445 | 143 | 1.26 (0.77, 2.07) | 0.10 |
| Burkina Faso | iLiNS−Zinc (24) | | DMC | 746 | 375 | 0.49 (0.36, 0.67) | 0.19 |
| Ghana | GHANA (25) | | | | | | |
| Ghana | iLiNS−DYADG (26) | | KDI | 302 | 601 | 1.13 (0.76, 1.70) | 0.14 |
| Haiti | HAITI (27) | | | | | | |
| Madagascar | MAHAY (29) | | ASQI | 1613 | 1604 | 0.91 (0.58, 1.44) | 0.11 |
| Malawi | iLiNS−DYADM (30) | | KDI | 214 | 436 | 0.84 (0.51, 1.40) | 0.10 |
| Malawi | iLiNS−DOSE (31) | | KDI | 646 | 221 | 1.08 (0.67, 1.71) | 0.11 |
| Zimbabwe | SHINE[1] (HIV−) (33), | LNS vs SOC | MDAT | 395 | 382 | 1.08 (0.67, 1.73) | 0.18 |
| Zimbabwe | | LNS+WSH vs WSH | MDAT | 446 | 417 | 0.82 (0.54, 1.24) | 0.18 |
| Zimbabwe | SHINE[1] (HIV+) (34), | LNS vs SOC | MDAT | 67 | 68 | 1.16 (0.44, 3.08) | 0.06 |
| Zimbabwe | | LNS+WSH vs WSH | MDAT | 103 | 83 | 0.40 (0.17, 0.95) | 0.06 |
| **$I^2$ = 0.61, $Tau^2$ = 0.05** | | | | **4977** | **4330** | **0.87 (0.69, 1.08)** | |
| **Supplement duration − > 12m** | | | | | | | |
| Bangladesh | RDNS (22) | | DMC | 1556 | 753 | 0.85 (0.67, 1.07) | 0.26 |
| Bangladesh | WASH−B (23) | | EASQ | 1074 | 3279 | 0.82 (0.66, 1.01) | 0.28 |
| Kenya | WASH−B (28) | | EASQ | 1362 | 4745 | 0.86 (0.70, 1.05) | 0.29 |
| Mali | PROMIS CS (32) | | DMC | 902 | 921 | 0.81 (0.56, 1.18) | 0.16 |
| **$I^2$ = 0.00, $Tau^2$ = 0.00** | | | | **4894** | **9698** | **0.84 (0.74, 0.94)** | |

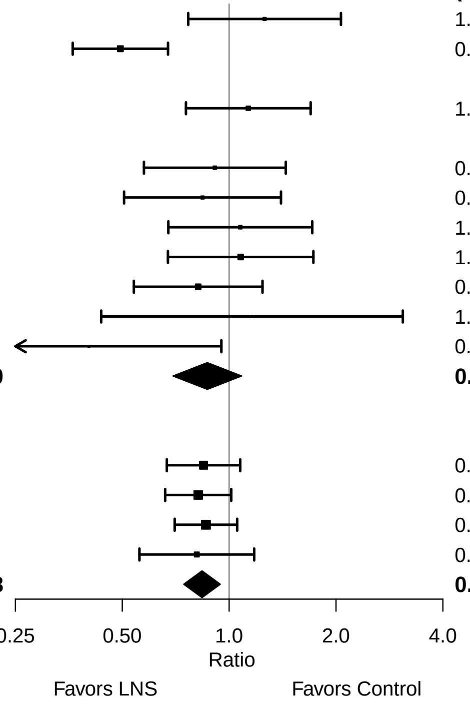

#### Supplemental figure 4H: Motor lowest decile prevalence ratio

#### 4H8: Stratified by Frequency of contact

**Frequency of contact**
**(p−diff = 0.977)**

**Frequency of contact − Monthly**

| Country | Trial | Tool | N | N | PR (95% CI) | W |
|---|---|---|---|---|---|---|
| Bangladesh | RDNS (22) | DMC | 1556 | 753 | 0.85 (0.67, 1.07) | 0.23 |
| Haiti | HAITI (27) | | | | | |
| Kenya | WASH−B (28) | EASQ | 1362 | 4745 | 0.86 (0.70, 1.05) | 0.26 |
| Madagascar | MAHAY (29) | ASQI | 1613 | 1604 | 0.91 (0.58, 1.44) | 0.11 |
| Mali | PROMIS CS (32) | DMC | 902 | 921 | 0.81 (0.56, 1.18) | 0.15 |
| Zimbabwe | SHINE[1] (HIV−) (33), LNS vs SOC | MDAT | 395 | 382 | 1.08 (0.67, 1.73) | 0.18 |
| Zimbabwe | LNS+WSH vs WSH | MDAT | 446 | 417 | 0.82 (0.54, 1.24) | 0.18 |
| Zimbabwe | SHINE[1] (HIV+) (34), LNS vs SOC | MDAT | 67 | 68 | 1.16 (0.44, 3.08) | 0.06 |
| Zimbabwe | LNS+WSH vs WSH | MDAT | 103 | 83 | 0.40 (0.17, 0.95) | 0.06 |
| **$I^2$ = 0.00, $Tau^2$ = 0.00** | | | **6444** | **8973** | **0.86 (0.76, 0.97)** | |

**Frequency of contact − Weekly**

| Country | Trial | Tool | N | N | PR (95% CI) | W |
|---|---|---|---|---|---|---|
| Bangladesh | JiVitA−4 (21) | BSID−III | 445 | 143 | 1.26 (0.77, 2.07) | 0.11 |
| Bangladesh | WASH−B (23) | EASQ | 1074 | 3279 | 0.82 (0.66, 1.01) | 0.29 |
| Burkina Faso | iLiNS−Zinc (24) | DMC | 746 | 375 | 0.49 (0.36, 0.67) | 0.21 |
| Ghana | GHANA (25) | | | | | |
| Ghana | iLiNS−DYADG (26) | KDI | 302 | 601 | 1.13 (0.76, 1.70) | 0.15 |
| Malawi | iLiNS−DYADM (30) | KDI | 214 | 436 | 0.84 (0.51, 1.40) | 0.11 |
| Malawi | iLiNS−DOSE (31) | KDI | 646 | 221 | 1.08 (0.67, 1.71) | 0.12 |
| **$I^2$ = 0.71, $Tau^2$ = 0.08** | | | **3427** | **5055** | **0.87 (0.66, 1.15)** | |

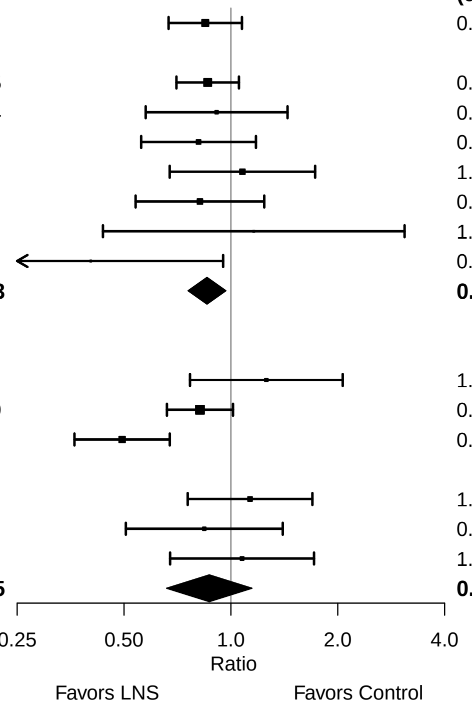

#### Supplemental figure 4H: Motor lowest decile prevalence ratio

### 4H9: Stratified by Average SQ-LNS compliance

**Average SQ−LNS compliance**
**(p−diff = 0.330)**

| Country | Trial | Tool | N | N | PR (95% CI) | W |
|---|---|---|---|---|---|---|
| **Average SQ−LNS compliance − Low** | | | | | | |
| Ghana | iLiNS−DYADG (26) | KDI | 302 | 601 | 1.13 (0.76, 1.70) | 0.18 |
| Malawi | iLiNS−DYADM (30) | KDI | 214 | 436 | 0.84 (0.51, 1.40) | 0.13 |
| Malawi | iLiNS−DOSE (31) | KDI | 646 | 221 | 1.08 (0.67, 1.71) | 0.15 |
| Mali | PROMIS CS (32) | DMC | 902 | 921 | 0.81 (0.56, 1.18) | 0.20 |
| Zimbabwe | SHINE[1] (HIV−) (33), LNS vs SOC | MDAT | 395 | 382 | 1.08 (0.67, 1.73) | 0.24 |
| Zimbabwe | LNS+WSH vs WSH | MDAT | 446 | 417 | 0.82 (0.54, 1.24) | 0.24 |
| Zimbabwe | SHINE[1] (HIV+) (34), LNS vs SOC | MDAT | 67 | 68 | 1.16 (0.44, 3.08) | 0.09 |
| Zimbabwe | LNS+WSH vs WSH | MDAT | 103 | 83 | 0.40 (0.17, 0.95) | 0.09 |
| **$I^2$ = 0.00, $Tau^2$ = 0.00** | | | **3075** | **3129** | **0.92 (0.78, 1.09)** | |
| **Average SQ−LNS compliance − High** | | | | | | |
| Bangladesh | JiVitA−4 (21) | BSID−III | 445 | 143 | 1.26 (0.77, 2.07) | 0.10 |
| Bangladesh | RDNS (22) | DMC | 1556 | 753 | 0.85 (0.67, 1.07) | 0.23 |
| Bangladesh | WASH−B (23) | EASQ | 1074 | 3279 | 0.82 (0.66, 1.01) | 0.24 |
| Burkina Faso | iLiNS−Zinc (24) | DMC | 746 | 375 | 0.49 (0.36, 0.67) | 0.18 |
| Ghana | GHANA (25) | | | | | |
| Haiti | HAITI (27) | | | | | |
| Kenya | WASH−B (28) | EASQ | 1362 | 4745 | 0.86 (0.70, 1.05) | 0.25 |
| **$I^2$ = 0.70, $Tau^2$ = 0.07** | | | **5183** | **9295** | **0.80 (0.61, 1.05)** | |

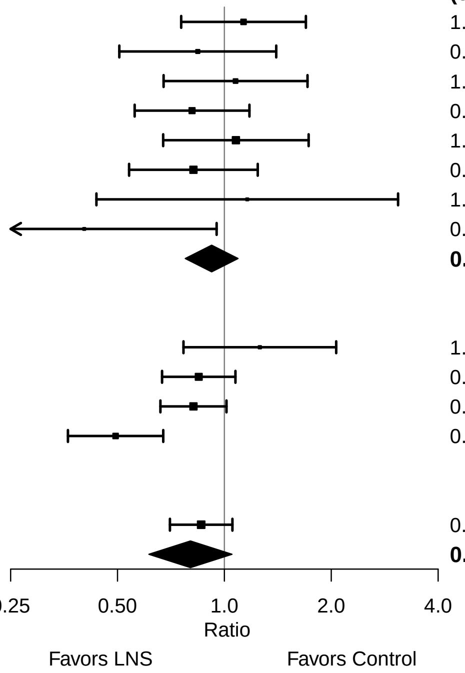

### Supplemental figure 4I: Motor lowest decile prevalence difference

#### 4I1: Stratified by Geographic region

**Geographic region**
**(p−diff = 0.995)**

**Geographic region − SEAR**

| Country | Trial | Tool | N | N | PD (95% CI) | W |
|---|---|---|---|---|---|---|
| Bangladesh | JiVitA−4 (21) | BSID−III | 445 | 143 | 0.02 (−0.03, 0.08) | 0.13 |
| Bangladesh | RDNS (22) | DMC | 1556 | 753 | −0.02 (−0.04, 0.01) | 0.37 |
| Bangladesh | WASH−B (23) | EASQ | 1074 | 3279 | −0.02 (−0.04, 0.00) | 0.50 |
| **I² = 0.00, Tau² = 0.00** | | | **3075** | **4175** | **−0.02 (−0.03, 0.00)** | |
| **Geographic region − AFR** | | | | | | |
| Burkina Faso | iLiNS−Zinc (24) | DMC | 746 | 375 | −0.08 (−0.12, −0.04) | 0.08 |
| Ghana | GHANA (25) | | | | | |
| Ghana | iLiNS−DYADG (26) | KDI | 302 | 601 | 0.01 (−0.03, 0.06) | 0.08 |
| Kenya | WASH−B (28) | EASQ | 1362 | 4745 | −0.01 (−0.03, 0.00) | 0.22 |
| Madagascar | MAHAY (29) | ASQI | 1613 | 1604 | −0.01 (−0.03, 0.01) | 0.19 |
| Malawi | iLiNS−DYADM (30) | KDI | 214 | 436 | −0.02 (−0.06, 0.03) | 0.06 |
| Malawi | iLiNS−DOSE (31) | KDI | 646 | 221 | 0.01 (−0.04, 0.05) | 0.07 |
| Mali | PROMIS CS (32) | DMC | 902 | 921 | −0.02 (−0.05, 0.01) | 0.14 |
| Zimbabwe | SHINE¹ (HIV−) (33), LNS vs SOC | MDAT | 395 | 382 | 0.01 (−0.03, 0.05) | 0.13 |
| Zimbabwe | LNS+WSH vs WSH | MDAT | 446 | 417 | −0.02 (−0.06, 0.02) | 0.13 |
| Zimbabwe | SHINE¹ (HIV+) (34), LNS vs SOC | MDAT | 67 | 68 | 0.02 (−0.09, 0.12) | 0.03 |
| Zimbabwe | LNS+WSH vs WSH | MDAT | 103 | 83 | −0.09 (−0.17, 0.00) | 0.03 |
| **I² = 0.37, Tau² = 0.00** | | | **6796** | **9853** | **−0.02 (−0.03, 0.00)** | |

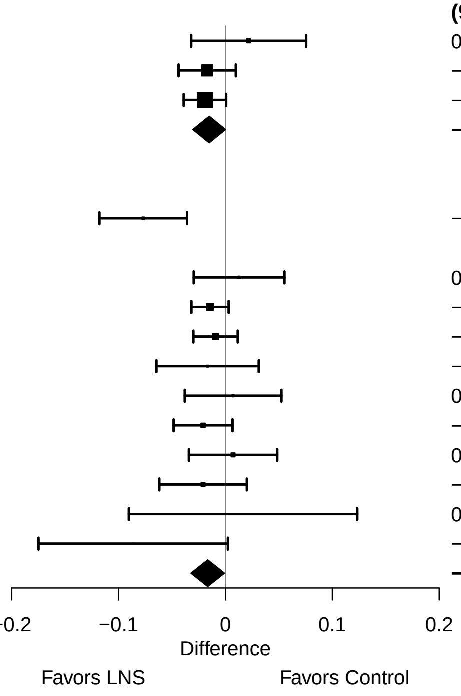

#### Supplemental figure 4I: Motor lowest decile prevalence difference

### 4I2: Stratified by Stunting burden

**Stunting burden**
**(p−diff = 0.537)**

| Country | Trial | Tool | N | N | PD (95% CI) | W |
|---|---|---|---|---|---|---|
| **Stunting burden − Less than 35%** | | | | | | |
| Ghana | GHANA (25) | | | | | |
| Ghana | iLiNS−DYADG (26) | KDI | 302 | 601 | 0.01 (−0.03, 0.06) | 0.21 |
| Haiti | HAITI (27) | | | | | |
| Kenya | WASH−B (28) | EASQ | 1362 | 4745 | −0.01 (−0.03, 0.00) | 0.61 |
| Malawi | iLiNS−DYADM (30) | KDI | 214 | 436 | −0.02 (−0.06, 0.03) | 0.17 |
| **I² = 0.00, Tau² = 0.00** | | | **1878** | **5782** | **−0.01 (−0.03, 0.00)** | |
| **Stunting burden − More than 35%** | | | | | | |
| Bangladesh | JiVitA−4 (21) | BSID−III | 445 | 143 | 0.02 (−0.03, 0.08) | 0.05 |
| Bangladesh | RDNS (22) | DMC | 1556 | 753 | −0.02 (−0.04, 0.01) | 0.14 |
| Bangladesh | WASH−B (23) | EASQ | 1074 | 3279 | −0.02 (−0.04, 0.00) | 0.19 |
| Burkina Faso | iLiNS−Zinc (24) | DMC | 746 | 375 | −0.08 (−0.12, −0.04) | 0.08 |
| Madagascar | MAHAY (29) | ASQI | 1613 | 1604 | −0.01 (−0.03, 0.01) | 0.18 |
| Malawi | iLiNS−DOSE (31) | KDI | 646 | 221 | 0.01 (−0.04, 0.05) | 0.07 |
| Mali | PROMIS CS (32) | DMC | 902 | 921 | −0.02 (−0.05, 0.01) | 0.13 |
| Zimbabwe | SHINE¹ (HIV−) (33), LNS vs SOC | MDAT | 395 | 382 | 0.01 (−0.03, 0.05) | 0.13 |
| Zimbabwe | LNS+WSH vs WSH | MDAT | 446 | 417 | −0.02 (−0.06, 0.02) | 0.13 |
| Zimbabwe | SHINE¹ (HIV+) (34), LNS vs SOC | MDAT | 67 | 68 | 0.02 (−0.09, 0.12) | 0.03 |
| Zimbabwe | LNS+WSH vs WSH | MDAT | 103 | 83 | −0.09 (−0.17, 0.00) | 0.03 |
| **I² = 0.38, Tau² = 0.00** | | | **7993** | **8246** | **−0.02 (−0.03, 0.00)** | |

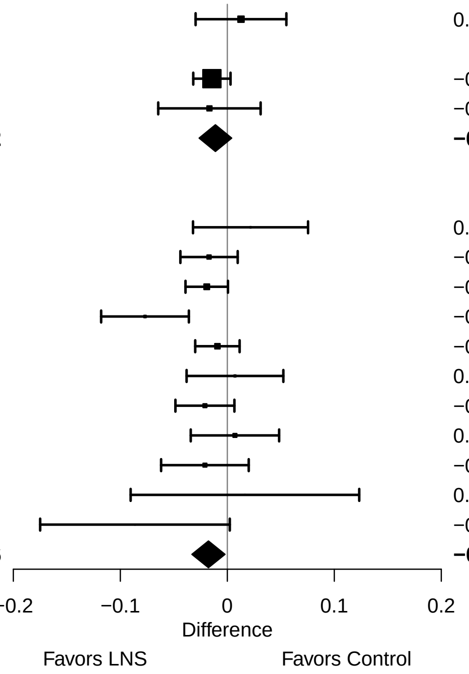

#### Supplemental figure 4I: Motor lowest decile prevalence difference

### 4I3: Stratified by Malaria prevalence

**Malaria prevalence**
**(p−diff = 0.476)**

| Country | Trial | Tool | N | N | PD (95% CI) | W |
|---|---|---|---|---|---|---|
| **Malaria prevalence − Less than 10%** | | | | | | |
| Bangladesh | JiVitA−4 (21) | BSID−III | 445 | 143 | 0.02 (−0.03, 0.08) | 0.05 |
| Bangladesh | RDNS (22) | DMC | 1556 | 753 | −0.02 (−0.04, 0.01) | 0.15 |
| Bangladesh | WASH−B (23) | EASQ | 1074 | 3279 | −0.02 (−0.04, 0.00) | 0.20 |
| Haiti | HAITI (27) | | | | | |
| Kenya | WASH−B (28) | EASQ | 1362 | 4745 | −0.01 (−0.03, 0.00) | 0.23 |
| Madagascar | MAHAY (29) | ASQI | 1613 | 1604 | −0.01 (−0.03, 0.01) | 0.20 |
| Zimbabwe | SHINE¹ (HIV−) (33), LNS vs SOC | MDAT | 395 | 382 | 0.01 (−0.03, 0.05) | 0.13 |
| Zimbabwe | LNS+WSH vs WSH | MDAT | 446 | 417 | −0.02 (−0.06, 0.02) | 0.13 |
| Zimbabwe | SHINE¹ (HIV+) (34), LNS vs SOC | MDAT | 67 | 68 | 0.02 (−0.09, 0.12) | 0.04 |
| Zimbabwe | LNS+WSH vs WSH | MDAT | 103 | 83 | −0.09 (−0.17, 0.00) | 0.04 |
| **$I^2$ = 0.00, $Tau^2$ = 0.00** | | | **7061** | **11474** | **−0.01 (−0.02, 0.00)** | |
| **Malaria prevalence − At least 10%** | | | | | | |
| Burkina Faso | iLiNS−Zinc (24) | DMC | 746 | 375 | −0.08 (−0.12, −0.04) | 0.19 |
| Ghana | GHANA (25) | | | | | |
| Ghana | iLiNS−DYADG (26) | KDI | 302 | 601 | 0.01 (−0.03, 0.06) | 0.18 |
| Malawi | iLiNS−DYADM (30) | KDI | 214 | 436 | −0.02 (−0.06, 0.03) | 0.15 |
| Malawi | iLiNS−DOSE (31) | KDI | 646 | 221 | 0.01 (−0.04, 0.05) | 0.16 |
| Mali | PROMIS CS (32) | DMC | 902 | 921 | −0.02 (−0.05, 0.01) | 0.33 |
| **$I^2$ = 0.64, $Tau^2$ = 0.00** | | | **2810** | **2554** | **−0.02 (−0.05, 0.01)** | |

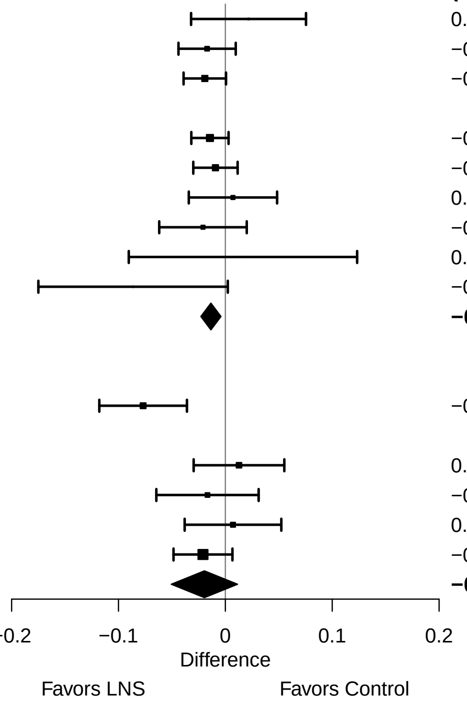

#### Supplemental figure 4I: Motor lowest decile prevalence difference

### 4I4: Stratified by Anemia prevalence

**Anemia prevalence**
**(p−diff = 0.163)**

| Country | Trial | Tool | N | N | PD (95% CI) | W |
|---|---|---|---|---|---|---|
| **Anemia prevalence − High** | | | | | | |
| Burkina Faso | iLiNS−Zinc (24) | DMC | 746 | 375 | −0.08 (−0.12, −0.04) | 0.23 |
| Ghana | GHANA (25) | | | | | |
| Haiti | HAITI (27) | | | | | |
| Malawi | iLiNS−DYADM (30) | KDI | 214 | 436 | −0.02 (−0.06, 0.03) | 0.18 |
| Malawi | iLiNS−DOSE (31) | KDI | 646 | 221 | 0.01 (−0.04, 0.05) | 0.20 |
| Mali | PROMIS CS (32) | DMC | 902 | 921 | −0.02 (−0.05, 0.01) | 0.40 |
| **I² = 0.64, Tau² = 0.00** | | | **2508** | **1953** | **−0.03 (−0.06, 0.01)** | |
| **Anemia prevalence − Moderate** | | | | | | |
| Bangladesh | JiVitA−4 (21) | BSID−III | 445 | 143 | 0.02 (−0.03, 0.08) | 0.05 |
| Bangladesh | RDNS (22) | DMC | 1556 | 753 | −0.02 (−0.04, 0.01) | 0.14 |
| Bangladesh | WASH−B (23) | EASQ | 1074 | 3279 | −0.02 (−0.04, 0.00) | 0.19 |
| Ghana | iLiNS−DYADG (26) | KDI | 302 | 601 | 0.01 (−0.03, 0.06) | 0.07 |
| Kenya | WASH−B (28) | EASQ | 1362 | 4745 | −0.01 (−0.03, 0.00) | 0.21 |
| Madagascar | MAHAY (29) | ASQI | 1613 | 1604 | −0.01 (−0.03, 0.01) | 0.18 |
| Zimbabwe | SHINE¹ (HIV−) (33), LNS vs SOC | MDAT | 395 | 382 | 0.01 (−0.03, 0.05) | 0.12 |
| Zimbabwe | LNS+WSH vs WSH | MDAT | 446 | 417 | −0.02 (−0.06, 0.02) | 0.12 |
| Zimbabwe | SHINE¹ (HIV+) (34), LNS vs SOC | MDAT | 67 | 68 | 0.02 (−0.09, 0.12) | 0.03 |
| Zimbabwe | LNS+WSH vs WSH | MDAT | 103 | 83 | −0.09 (−0.17, 0.00) | 0.03 |
| **I² = 0.00, Tau² = 0.00** | | | **7363** | **12075** | **−0.01 (−0.02, 0.00)** | |

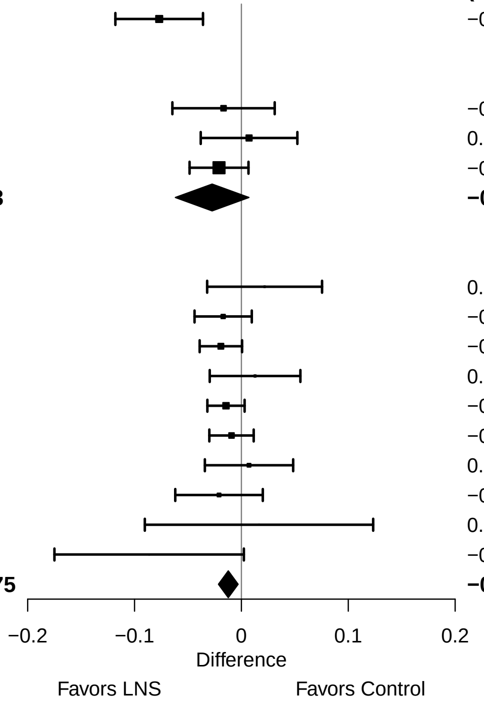

##### Supplemental figure 4I: Motor lowest decile prevalence difference

#### 4I5: Stratified by Source water quality

**Source water quality**
**(p−diff = 0.444)**

| Country | Trial | Tool | N | N | PD (95% CI) | W |
|---|---|---|---|---|---|---|
| **Source water quality − Improved** | | | | | | |
| Bangladesh | JiVitA−4 (21) | BSID−III | 445 | 143 | 0.02 (−0.03, 0.08) | 0.10 |
| Bangladesh | RDNS (22) | DMC | 1556 | 753 | −0.02 (−0.04, 0.01) | 0.27 |
| Bangladesh | WASH−B (23) | EASQ | 528 | 1099 | −0.02 (−0.05, 0.01) | 0.22 |
| Ghana | GHANA (25) | | | | | |
| Ghana | iLiNS−DYADG (26) | KDI | 302 | 601 | 0.01 (−0.03, 0.06) | 0.15 |
| Haiti | HAITI (27) | | | | | |
| Malawi | iLiNS−DYADM (30) | KDI | 214 | 436 | −0.02 (−0.06, 0.03) | 0.12 |
| Malawi | iLiNS−DOSE (31) | KDI | 646 | 221 | 0.01 (−0.04, 0.05) | 0.14 |
| **I² = 0.00, Tau² = 0.00** | | | **3691** | **3253** | **−0.01 (−0.02, 0.01)** | |
| **Source water quality − Unimproved** | | | | | | |
| Burkina Faso | iLiNS−Zinc (24) | DMC | 746 | 375 | −0.08 (−0.12, −0.04) | 0.13 |
| Kenya | WASH−B (28) | EASQ | 649 | 2073 | 0.00 (−0.03, 0.02) | 0.23 |
| Madagascar | MAHAY (29) | ASQI | 1613 | 1604 | −0.01 (−0.03, 0.01) | 0.28 |
| Mali | PROMIS CS (32) | DMC | 902 | 921 | −0.02 (−0.05, 0.01) | 0.21 |
| Zimbabwe | SHINE (HIV−) (33) | MDAT | 395 | 382 | 0.01 (−0.03, 0.05) | 0.13 |
| Zimbabwe | SHINE (HIV+) (34) | MDAT | 67 | 68 | 0.02 (−0.09, 0.12) | 0.03 |
| **I² = 0.56, Tau² = 0.00** | | | **4372** | **5423** | **−0.02 (−0.04, 0.01)** | |

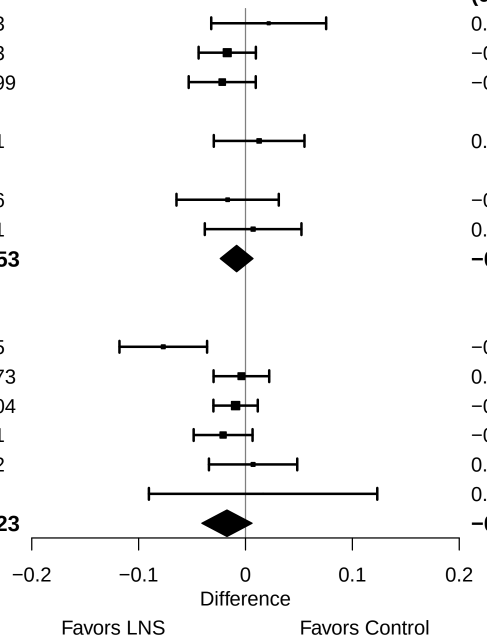

#### Supplemental figure 4I: Motor lowest decile prevalence difference

### 4I6: Stratified by Sanitation

**Sanitation**
**(p−diff = 0.871)**

| Country | Trial | Tool | N | N | PD (95% CI) | W |
|---|---|---|---|---|---|---|
| **Sanitation − Improved** | | | | | | |
| Bangladesh | JiVitA−4 (21) | BSID−III | 445 | 143 | 0.02 (−0.03, 0.08) | 0.10 |
| Bangladesh | RDNS (22) | DMC | 1556 | 753 | −0.02 (−0.04, 0.01) | 0.27 |
| Bangladesh | WASH−B (23) | EASQ | 528 | 1099 | −0.02 (−0.05, 0.01) | 0.22 |
| Ghana | GHANA (25) | | | | | |
| Ghana | iLiNS−DYADG (26) | KDI | 302 | 601 | 0.01 (−0.03, 0.06) | 0.15 |
| Haiti | HAITI (27) | | | | | |
| Mali | PROMIS CS (32) | DMC | 902 | 921 | −0.02 (−0.05, 0.01) | 0.26 |
| **I² = 0.00, Tau² = 0.00** | | | **3733** | **3517** | **−0.01 (−0.03, 0.00)** | |
| **Sanitation − Unimproved** | | | | | | |
| Burkina Faso | iLiNS−Zinc (24) | DMC | 746 | 375 | −0.08 (−0.12, −0.04) | 0.13 |
| Kenya | WASH−B (28) | EASQ | 649 | 2073 | 0.00 (−0.03, 0.02) | 0.23 |
| Madagascar | MAHAY (29) | ASQI | 1613 | 1604 | −0.01 (−0.03, 0.01) | 0.28 |
| Malawi | iLiNS−DYADM (30) | KDI | 214 | 436 | −0.02 (−0.06, 0.03) | 0.10 |
| Malawi | iLiNS−DOSE (31) | KDI | 646 | 221 | 0.01 (−0.04, 0.05) | 0.11 |
| Zimbabwe | SHINE (HIV−) (33) | MDAT | 395 | 382 | 0.01 (−0.03, 0.05) | 0.13 |
| Zimbabwe | SHINE (HIV+) (34) | MDAT | 67 | 68 | 0.02 (−0.09, 0.12) | 0.03 |
| **I² = 0.50, Tau² = 0.00** | | | **4330** | **5159** | **−0.01 (−0.04, 0.01)** | |

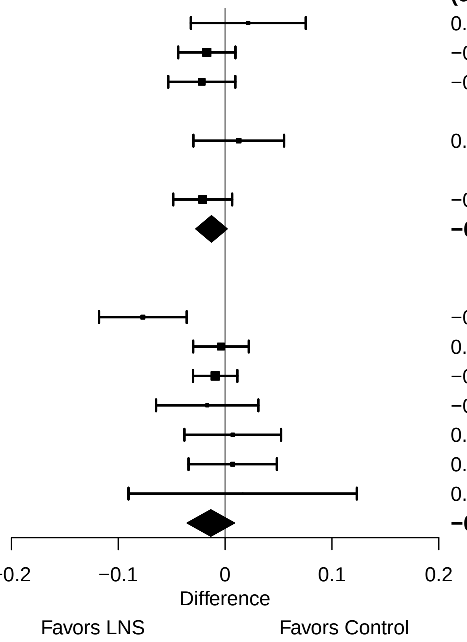

#### Supplemental figure 4I: Motor lowest decile prevalence difference

### 4I7: Stratified by Supplement duration

**Supplement duration**
**(p−diff = 0.553)**

**Supplement duration − 12m or less**

| Country | Trial | Tool | N | N | PD (95% CI) | W |
|---|---|---|---|---|---|---|
| Bangladesh | JiVitA−4 (21) | BSID−III | 445 | 143 | 0.02 (−0.03, 0.08) | 0.07 |
| Burkina Faso | iLiNS−Zinc (24) | DMC | 746 | 375 | −0.08 (−0.12, −0.04) | 0.12 |
| Ghana | GHANA (25) | | | | | |
| Ghana | iLiNS−DYADG (26) | KDI | 302 | 601 | 0.01 (−0.03, 0.06) | 0.11 |
| Haiti | HAITI (27) | | | | | |
| Madagascar | MAHAY (29) | ASQI | 1613 | 1604 | −0.01 (−0.03, 0.01) | 0.27 |
| Malawi | iLiNS−DYADM (30) | KDI | 214 | 436 | −0.02 (−0.06, 0.03) | 0.09 |
| Malawi | iLiNS−DOSE (31) | KDI | 646 | 221 | 0.01 (−0.04, 0.05) | 0.10 |
| Zimbabwe | SHINE[1] (HIV−) (33), LNS vs SOC | MDAT | 395 | 382 | 0.01 (−0.03, 0.05) | 0.19 |
| Zimbabwe | LNS+WSH vs WSH | MDAT | 446 | 417 | −0.02 (−0.06, 0.02) | 0.19 |
| Zimbabwe | SHINE[1] (HIV+) (34), LNS vs SOC | MDAT | 67 | 68 | 0.02 (−0.09, 0.12) | 0.05 |
| Zimbabwe | LNS+WSH vs WSH | MDAT | 103 | 83 | −0.09 (−0.17, 0.00) | 0.05 |
| **I² = 0.51, Tau² = 0.00** | | | **4977** | **4330** | **−0.01 (−0.03, 0.01)** | |

**Supplement duration − > 12m**

| Country | Trial | Tool | N | N | PD (95% CI) | W |
|---|---|---|---|---|---|---|
| Bangladesh | RDNS (22) | DMC | 1556 | 753 | −0.02 (−0.04, 0.01) | 0.21 |
| Bangladesh | WASH−B (23) | EASQ | 1074 | 3279 | −0.02 (−0.04, 0.00) | 0.28 |
| Kenya | WASH−B (28) | EASQ | 1362 | 4745 | −0.01 (−0.03, 0.00) | 0.31 |
| Mali | PROMIS CS (32) | DMC | 902 | 921 | −0.02 (−0.05, 0.01) | 0.20 |
| **I² = 0.00, Tau² = 0.00** | | | **4894** | **9698** | **−0.02 (−0.03, −0.01)** | |

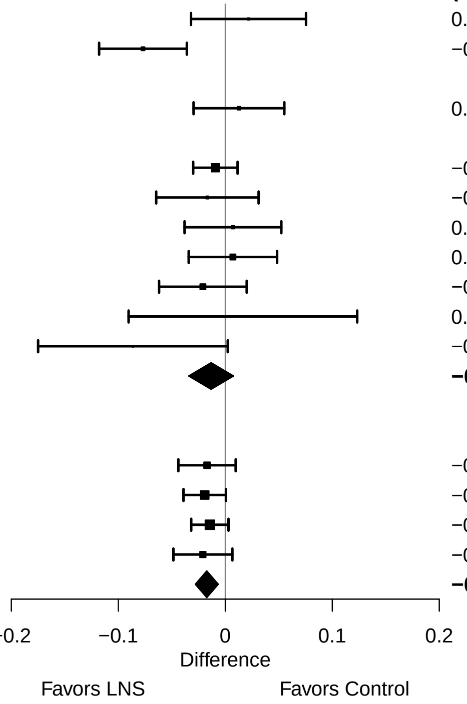

#### Supplemental figure 4I: Motor lowest decile prevalence difference

### 4I8: Stratified by Frequency of contact

**Frequency of contact**
**(p−diff = 0.780)**

| Country | Trial | Tool | N | N | PD (95% CI) | W |
|---|---|---|---|---|---|---|
| **Frequency of contact − Monthly** | | | | | | |
| Bangladesh | RDNS (22) | DMC | 1556 | 753 | −0.02 (−0.04, 0.01) | 0.17 |
| Haiti | HAITI (27) | | | | | |
| Kenya | WASH−B (28) | EASQ | 1362 | 4745 | −0.01 (−0.03, 0.00) | 0.26 |
| Madagascar | MAHAY (29) | ASQI | 1613 | 1604 | −0.01 (−0.03, 0.01) | 0.22 |
| Mali | PROMIS CS (32) | DMC | 902 | 921 | −0.02 (−0.05, 0.01) | 0.16 |
| Zimbabwe | SHINE[1] (HIV−) (33), LNS vs SOC | MDAT | 395 | 382 | 0.01 (−0.03, 0.05) | 0.15 |
| Zimbabwe | LNS+WSH vs WSH | MDAT | 446 | 417 | −0.02 (−0.06, 0.02) | 0.15 |
| Zimbabwe | SHINE[1] (HIV+) (34), LNS vs SOC | MDAT | 67 | 68 | 0.02 (−0.09, 0.12) | 0.04 |
| Zimbabwe | LNS+WSH vs WSH | MDAT | 103 | 83 | −0.09 (−0.17, 0.00) | 0.04 |
| **I² = 0.00, Tau² = 0.00** | | | **6444** | **8973** | **−0.01 (−0.02, 0.00)** | |
| **Frequency of contact − Weekly** | | | | | | |
| Bangladesh | JiVitA−4 (21) | BSID−III | 445 | 143 | 0.02 (−0.03, 0.08) | 0.10 |
| Bangladesh | WASH−B (23) | EASQ | 1074 | 3279 | −0.02 (−0.04, 0.00) | 0.37 |
| Burkina Faso | iLiNS−Zinc (24) | DMC | 746 | 375 | −0.08 (−0.12, −0.04) | 0.15 |
| Ghana | GHANA (25) | | | | | |
| Ghana | iLiNS−DYADG (26) | KDI | 302 | 601 | 0.01 (−0.03, 0.06) | 0.14 |
| Malawi | iLiNS−DYADM (30) | KDI | 214 | 436 | −0.02 (−0.06, 0.03) | 0.12 |
| Malawi | iLiNS−DOSE (31) | KDI | 646 | 221 | 0.01 (−0.04, 0.05) | 0.13 |
| **I² = 0.62, Tau² = 0.00** | | | **3427** | **5055** | **−0.01 (−0.04, 0.01)** | |

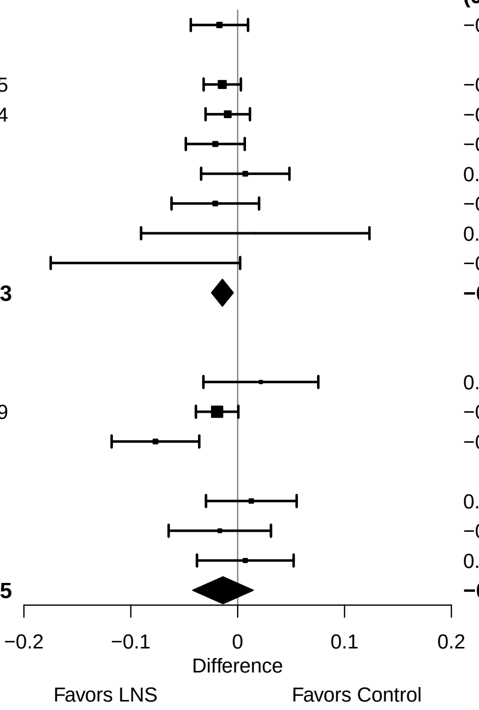

#### Supplemental figure 4I: Motor lowest decile prevalence difference

### 4I9: Stratified by Average SQ-LNS compliance

**Average SQ−LNS compliance**
**(p−diff = 0.342)**

| Country | Trial | Tool | N | N | PD (95% CI) | W |
|---|---|---|---|---|---|---|
| **Average SQ−LNS compliance − Low** | | | | | | |
| Ghana | iLiNS−DYADG (26) | KDI | 302 | 601 | 0.01 (−0.03, 0.06) | 0.15 |
| Malawi | iLiNS−DYADM (30) | KDI | 214 | 436 | −0.02 (−0.06, 0.03) | 0.13 |
| Malawi | iLiNS−DOSE (31) | KDI | 646 | 221 | 0.01 (−0.04, 0.05) | 0.14 |
| Mali | PROMIS CS (32) | DMC | 902 | 921 | −0.02 (−0.05, 0.01) | 0.26 |
| Zimbabwe | SHINE[1] (HIV−) (33), LNS vs SOC | MDAT | 395 | 382 | 0.01 (−0.03, 0.05) | 0.24 |
| Zimbabwe | LNS+WSH vs WSH | MDAT | 446 | 417 | −0.02 (−0.06, 0.02) | 0.24 |
| Zimbabwe | SHINE[1] (HIV+) (34), LNS vs SOC | MDAT | 67 | 68 | 0.02 (−0.09, 0.12) | 0.08 |
| Zimbabwe | LNS+WSH vs WSH | MDAT | 103 | 83 | −0.09 (−0.17, 0.00) | 0.08 |
| **$I^2$ = 0.00, $Tau^2$ = 0.00** | | | **3075** | **3129** | **−0.01 (−0.03, 0.01)** | |
| **Average SQ−LNS compliance − High** | | | | | | |
| Bangladesh | JiVitA−4 (21) | BSID−III | 445 | 143 | 0.02 (−0.03, 0.08) | 0.09 |
| Bangladesh | RDNS (22) | DMC | 1556 | 753 | −0.02 (−0.04, 0.01) | 0.21 |
| Bangladesh | WASH−B (23) | EASQ | 1074 | 3279 | −0.02 (−0.04, 0.00) | 0.27 |
| Burkina Faso | iLiNS−Zinc (24) | DMC | 746 | 375 | −0.08 (−0.12, −0.04) | 0.13 |
| Ghana | GHANA (25) | | | | | |
| Haiti | HAITI (27) | | | | | |
| Kenya | WASH−B (28) | EASQ | 1362 | 4745 | −0.01 (−0.03, 0.00) | 0.29 |
| **$I^2$ = 0.61, $Tau^2$ = 0.00** | | | **5183** | **9295** | **−0.02 (−0.05, 0.01)** | |

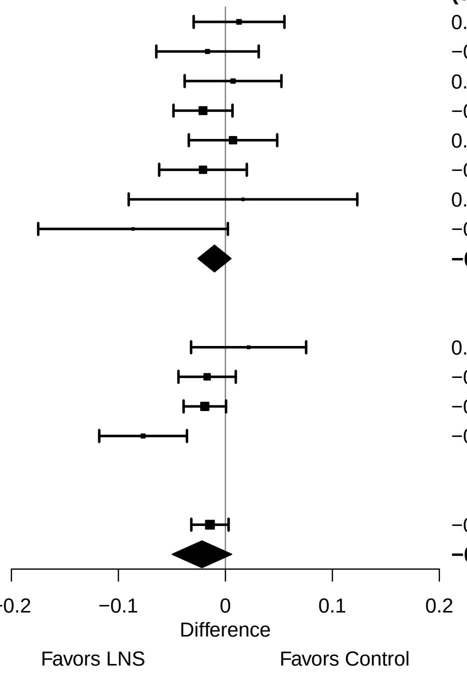

### Supplemental figure 4J: Mean difference in gross motor z-score

#### 4J1: Stratified by Geographic region

**Geographic region**
**(p−diff = 0.929)**

| Country | Trial | Tool | N | N | MD (95% CI) | W |
|---|---|---|---|---|---|---|
| **Geographic region − SEAR** | | | | | | |
| Bangladesh | JiVitA−4 (21) | BSID−III | 445 | 143 | −0.10 (−0.25, 0.05) | 0.22 |
| Bangladesh | RDNS (22) | DMC | 1570 | 758 | 0.08 (0.02, 0.15) | 0.40 |
| Bangladesh | WASH−B (23) | EASQ | 1074 | 3279 | 0.10 (0.04, 0.17) | 0.38 |
| **I² = 0.66, Tau² = 0.01** | | | **3089** | **4180** | **0.05 (−0.07, 0.16)** | |
| **Geographic region − AFR** | | | | | | |
| Burkina Faso | iLiNS−Zinc (24) | DMC | | | | |
| Ghana | GHANA (25) | | | | | |
| Ghana | iLiNS−DYADG (26) | KDI | 302 | 601 | 0.01 (−0.13, 0.15) | 0.12 |
| Kenya | WASH−B (28) | EASQ | 1362 | 4745 | 0.01 (−0.05, 0.07) | 0.19 |
| Madagascar | MAHAY (29) | ASQI | 1613 | 1604 | 0.08 (−0.08, 0.23) | 0.10 |
| Malawi | iLiNS−DYADM (30) | KDI | 214 | 436 | −0.03 (−0.19, 0.13) | 0.10 |
| Malawi | iLiNS−DOSE (31) | KDI | 646 | 221 | 0.00 (−0.15, 0.16) | 0.11 |
| Mali | PROMIS CS (32) | DMC | 940 | 957 | 0.09 (0.00, 0.17) | 0.17 |
| Zimbabwe | SHINE¹ (HIV−) (33), LNS vs SOC | MDAT | 395 | 382 | 0.03 (−0.13, 0.18) | 0.14 |
| Zimbabwe | LNS+WSH vs WSH | MDAT | 446 | 417 | 0.16 (0.00, 0.32) | 0.14 |
| Zimbabwe | SHINE¹ (HIV+) (34), LNS vs SOC | MDAT | 67 | 68 | 0.14 (−0.14, 0.42) | 0.08 |
| Zimbabwe | LNS+WSH vs WSH | MDAT | 103 | 83 | 0.48 (0.24, 0.72) | 0.08 |
| **I² = 0.50, Tau² = 0.01** | | | **6088** | **9514** | **0.06 (−0.01, 0.14)** | |

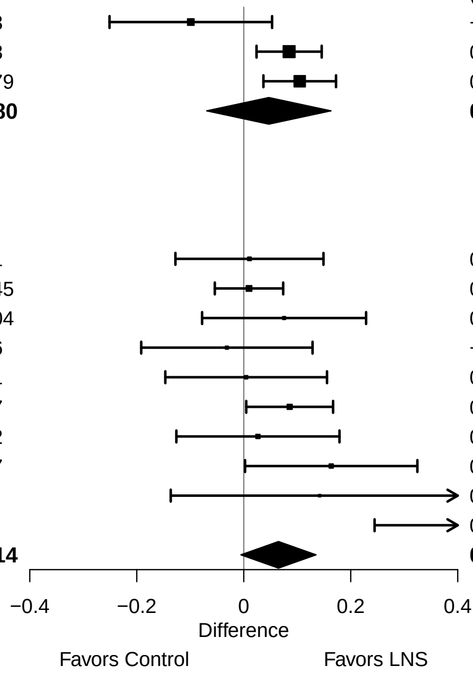

#### Supplemental figure 4J: Mean difference in gross motor z-score

### 4J2: Stratified by Stunting burden

**Stunting burden**
**(p−diff = 0.014)**

| Country | Trial | Tool | N | N | MD (95% CI) | W |
|---|---|---|---|---|---|---|
| **Stunting burden − Less than 35%** | | | | | | |
| Ghana | GHANA (25) | | | | | |
| Ghana | iLiNS−DYADG (26) | KDI | 302 | 601 | 0.01 (−0.13, 0.15) | 0.29 |
| Haiti | HAITI (27) | | | | | |
| Kenya | WASH−B (28) | EASQ | 1362 | 4745 | 0.01 (−0.05, 0.07) | 0.46 |
| Malawi | iLiNS−DYADM (30) | KDI | 214 | 436 | −0.03 (−0.19, 0.13) | 0.25 |
| **$I^2$ = 0.00, $Tau^2$ = 0.00** | | | **1878** | **5782** | **0.01 (−0.05, 0.06)** | |
| **Stunting burden − More than 35%** | | | | | | |
| Bangladesh | JiVitA−4 (21) | BSID−III | 445 | 143 | −0.10 (−0.25, 0.05) | 0.10 |
| Bangladesh | RDNS (22) | DMC | 1570 | 758 | 0.08 (0.02, 0.15) | 0.18 |
| Bangladesh | WASH−B (23) | EASQ | 1074 | 3279 | 0.10 (0.04, 0.17) | 0.17 |
| Burkina Faso | iLiNS−Zinc (24) | DMC | | | | |
| Madagascar | MAHAY (29) | ASQI | 1613 | 1604 | 0.08 (−0.08, 0.23) | 0.10 |
| Malawi | iLiNS−DOSE (31) | KDI | 646 | 221 | 0.00 (−0.15, 0.16) | 0.10 |
| Mali | PROMIS CS (32) | DMC | 940 | 957 | 0.09 (0.00, 0.17) | 0.16 |
| Zimbabwe | SHINE[1] (HIV−) (33), LNS vs SOC | MDAT | 395 | 382 | 0.03 (−0.13, 0.18) | 0.13 |
| Zimbabwe | LNS+WSH vs WSH | MDAT | 446 | 417 | 0.16 (0.00, 0.32) | 0.13 |
| Zimbabwe | SHINE[1] (HIV+) (34), LNS vs SOC | MDAT | 67 | 68 | 0.14 (−0.14, 0.42) | 0.08 |
| Zimbabwe | LNS+WSH vs WSH | MDAT | 103 | 83 | 0.48 (0.24, 0.72) | 0.08 |
| **$I^2$ = 0.51, $Tau^2$ = 0.01** | | | **7299** | **7912** | **0.08 (0.01, 0.16)** | |

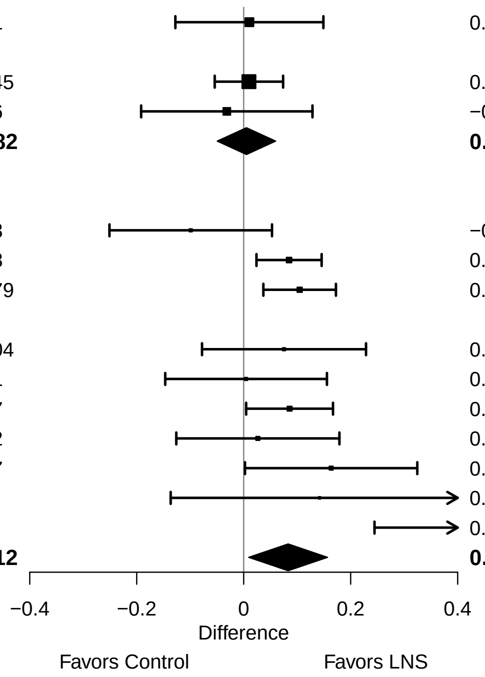

##### Supplemental figure 4J: Mean difference in gross motor z-score

#### 4J3: Stratified by Malaria prevalence

**Malaria prevalence**
**(p−diff = 0.387)**

| Country | Trial | Tool | N | N | MD (95% CI) | W |
|---|---|---|---|---|---|---|
| **Malaria prevalence − Less than 10%** | | | | | | |
| Bangladesh | JiVitA−4 (21) | BSID−III | 445 | 143 | −0.10 (−0.25, 0.05) | 0.11 |
| Bangladesh | RDNS (22) | DMC | 1570 | 758 | 0.08 (0.02, 0.15) | 0.19 |
| Bangladesh | WASH−B (23) | EASQ | 1074 | 3279 | 0.10 (0.04, 0.17) | 0.18 |
| Haiti | HAITI (27) | | | | | |
| Kenya | WASH−B (28) | EASQ | 1362 | 4745 | 0.01 (−0.05, 0.07) | 0.19 |
| Madagascar | MAHAY (29) | ASQI | 1613 | 1604 | 0.08 (−0.08, 0.23) | 0.11 |
| Zimbabwe | SHINE[1] (HIV−) (33), LNS vs SOC | MDAT | 395 | 382 | 0.03 (−0.13, 0.18) | 0.14 |
| Zimbabwe | LNS+WSH vs WSH | MDAT | 446 | 417 | 0.16 (0.00, 0.32) | 0.14 |
| Zimbabwe | SHINE[1] (HIV+) (34), LNS vs SOC | MDAT | 67 | 68 | 0.14 (−0.14, 0.42) | 0.08 |
| Zimbabwe | LNS+WSH vs WSH | MDAT | 103 | 83 | 0.48 (0.24, 0.72) | 0.08 |
| **$I^2$ = 0.66, $Tau^2$ = 0.01** | | | **7075** | **11479** | **0.08 (−0.01, 0.17)** | |
| **Malaria prevalence − At least 10%** | | | | | | |
| Burkina Faso | iLiNS−Zinc (24) | DMC | | | | |
| Ghana | GHANA (25) | | | | | |
| Ghana | iLiNS−DYADG (26) | KDI | 302 | 601 | 0.01 (−0.13, 0.15) | 0.24 |
| Malawi | iLiNS−DYADM (30) | KDI | 214 | 436 | −0.03 (−0.19, 0.13) | 0.20 |
| Malawi | iLiNS−DOSE (31) | KDI | 646 | 221 | 0.00 (−0.15, 0.16) | 0.22 |
| Mali | PROMIS CS (32) | DMC | 940 | 957 | 0.09 (0.00, 0.17) | 0.34 |
| **$I^2$ = 0.00, $Tau^2$ = 0.00** | | | **2102** | **2215** | **0.04 (−0.02, 0.10)** | |

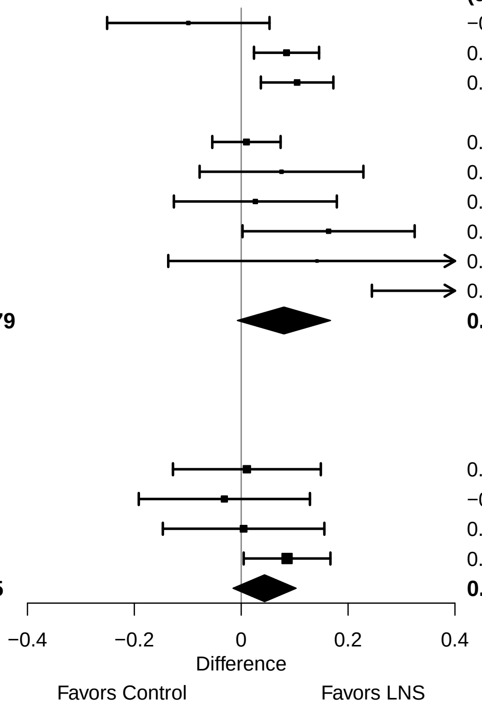

#### Supplemental figure 4J: Mean difference in gross motor z-score

### 4J4: Stratified by Anemia prevalence

**Anemia prevalence**
**(p−diff = 0.558)**

**Anemia prevalence − High**

| **Country** | **Trial** | **Tool** | **N** | **N** | **MD (95% CI)** | **W** |
|---|---|---|---|---|---|---|
| Burkina Faso | iLiNS−Zinc (24) | DMC | | | | |
| Ghana | GHANA (25) | | | | | |
| Haiti | HAITI (27) | | | | | |
| Malawi | iLiNS−DYADM (30) | KDI | 214 | 436 | −0.03 (−0.19, 0.13) | 0.27 |
| Malawi | iLiNS−DOSE (31) | KDI | 646 | 221 | 0.00 (−0.15, 0.16) | 0.28 |
| Mali | PROMIS CS (32) | DMC | 940 | 957 | 0.09 (0.00, 0.17) | 0.45 |
| **I² = 0.04, Tau² = 0.00** | | | **1800** | **1614** | **0.05 (−0.02, 0.12)** | |

**Anemia prevalence − Moderate**

| **Country** | **Trial** | **Tool** | **N** | **N** | **MD (95% CI)** | **W** |
|---|---|---|---|---|---|---|
| Bangladesh | JiVitA−4 (21) | BSID−III | 445 | 143 | −0.10 (−0.25, 0.05) | 0.10 |
| Bangladesh | RDNS (22) | DMC | 1570 | 758 | 0.08 (0.02, 0.15) | 0.17 |
| Bangladesh | WASH−B (23) | EASQ | 1074 | 3279 | 0.10 (0.04, 0.17) | 0.17 |
| Ghana | iLiNS−DYADG (26) | KDI | 302 | 601 | 0.01 (−0.13, 0.15) | 0.11 |
| Kenya | WASH−B (28) | EASQ | 1362 | 4745 | 0.01 (−0.05, 0.07) | 0.17 |
| Madagascar | MAHAY (29) | ASQI | 1613 | 1604 | 0.08 (−0.08, 0.23) | 0.09 |
| Zimbabwe | SHINE¹ (HIV−) (33), LNS vs SOC | MDAT | 395 | 382 | 0.03 (−0.13, 0.18) | 0.13 |
| Zimbabwe | LNS+WSH vs WSH | MDAT | 446 | 417 | 0.16 (0.00, 0.32) | 0.13 |
| Zimbabwe | SHINE¹ (HIV+) (34), LNS vs SOC | MDAT | 67 | 68 | 0.14 (−0.14, 0.42) | 0.07 |
| Zimbabwe | LNS+WSH vs WSH | MDAT | 103 | 83 | 0.48 (0.24, 0.72) | 0.07 |
| **I² = 0.62, Tau² = 0.01** | | | **7377** | **12080** | **0.07 (0.00, 0.15)** | |

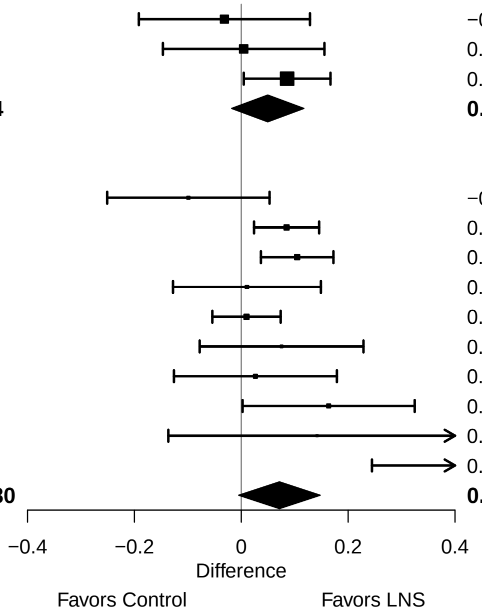

#### Supplemental figure 4J: Mean difference in gross motor z-score

#### 4J5: Stratified by Source water quality

**Source water quality**
**(p−diff = 0.777)**

| Country | Trial | Tool | N | N | MD (95% CI) | W |
|---|---|---|---|---|---|---|
| **Source water quality − Improved** | | | | | | |
| Bangladesh | JiVitA−4 (21) | BSID−III | 445 | 143 | −0.10 (−0.25, 0.05) | 0.12 |
| Bangladesh | RDNS (22) | DMC | 1570 | 758 | 0.08 (0.02, 0.15) | 0.33 |
| Bangladesh | WASH−B (23) | EASQ | 528 | 1099 | 0.19 (0.09, 0.30) | 0.19 |
| Ghana | GHANA (25) | | | | | |
| Ghana | iLiNS−DYADG (26) | KDI | 302 | 601 | 0.01 (−0.13, 0.15) | 0.13 |
| Haiti | HAITI (27) | | | | | |
| Malawi | iLiNS−DYADM (30) | KDI | 214 | 436 | −0.03 (−0.19, 0.13) | 0.11 |
| Malawi | iLiNS−DOSE (31) | KDI | 646 | 221 | 0.00 (−0.15, 0.16) | 0.12 |
| **I² = 0.62, Tau² = 0.01** | | | **3705** | **3258** | **0.04 (−0.04, 0.12)** | |
| **Source water quality − Unimproved** | | | | | | |
| Burkina Faso | iLiNS−Zinc (24) | DMC | | | | |
| Kenya | WASH−B (28) | EASQ | 649 | 2073 | 0.03 (−0.06, 0.13) | 0.29 |
| Madagascar | MAHAY (29) | ASQI | 1613 | 1604 | 0.08 (−0.08, 0.23) | 0.15 |
| Mali | PROMIS CS (32) | DMC | 940 | 957 | 0.09 (0.00, 0.17) | 0.35 |
| Zimbabwe | SHINE (HIV−) (33) | MDAT | 395 | 382 | 0.03 (−0.13, 0.18) | 0.15 |
| Zimbabwe | SHINE (HIV+) (34) | MDAT | 67 | 68 | 0.14 (−0.14, 0.42) | 0.05 |
| **I² = 0.00, Tau² = 0.00** | | | **3664** | **5084** | **0.06 (0.01, 0.12)** | |

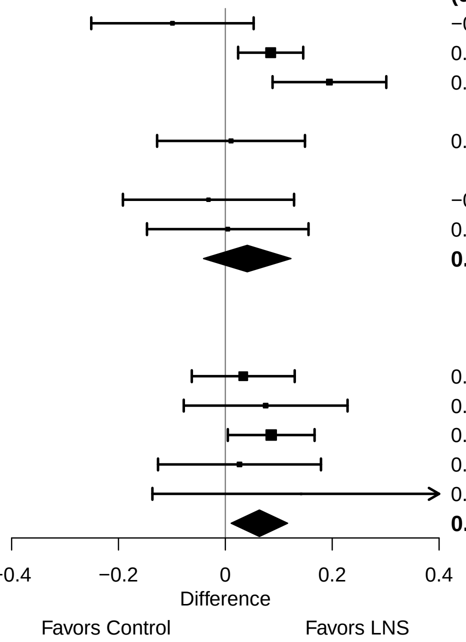

#### Supplemental figure 4J: Mean difference in gross motor z-score

### 4J6: Stratified by Sanitation

**Sanitation**
**(p−diff = 0.276)**

| Country | Trial | Tool | N | N | MD (95% CI) | W |
|---|---|---|---|---|---|---|
| **Sanitation − Improved** | | | | | | |
| Bangladesh | JiVitA−4 (21) | BSID−III | 445 | 143 | −0.10 (−0.25, 0.05) | 0.11 |
| Bangladesh | RDNS (22) | DMC | 1570 | 758 | 0.08 (0.02, 0.15) | 0.32 |
| Bangladesh | WASH−B (23) | EASQ | 528 | 1099 | 0.19 (0.09, 0.30) | 0.19 |
| Ghana | GHANA (25) | | | | | |
| Ghana | iLiNS−DYADG (26) | KDI | 302 | 601 | 0.01 (−0.13, 0.15) | 0.13 |
| Haiti | HAITI (27) | | | | | |
| Mali | PROMIS CS (32) | DMC | 940 | 957 | 0.09 (0.00, 0.17) | 0.25 |
| **I² = 0.63, Tau² = 0.01** | | | **3785** | **3558** | **0.07 (−0.02, 0.16)** | |
| **Sanitation − Unimproved** | | | | | | |
| Burkina Faso | iLiNS−Zinc (24) | DMC | | | | |
| Kenya | WASH−B (28) | EASQ | 649 | 2073 | 0.03 (−0.06, 0.13) | 0.30 |
| Madagascar | MAHAY (29) | ASQI | 1613 | 1604 | 0.08 (−0.08, 0.23) | 0.16 |
| Malawi | iLiNS−DYADM (30) | KDI | 214 | 436 | −0.03 (−0.19, 0.13) | 0.15 |
| Malawi | iLiNS−DOSE (31) | KDI | 646 | 221 | 0.00 (−0.15, 0.16) | 0.16 |
| Zimbabwe | SHINE (HIV−) (33) | MDAT | 395 | 382 | 0.03 (−0.13, 0.18) | 0.16 |
| Zimbabwe | SHINE (HIV+) (34) | MDAT | 67 | 68 | 0.14 (−0.14, 0.42) | 0.06 |
| **I² = 0.00, Tau² = 0.00** | | | **3584** | **4784** | **0.03 (−0.03, 0.09)** | |

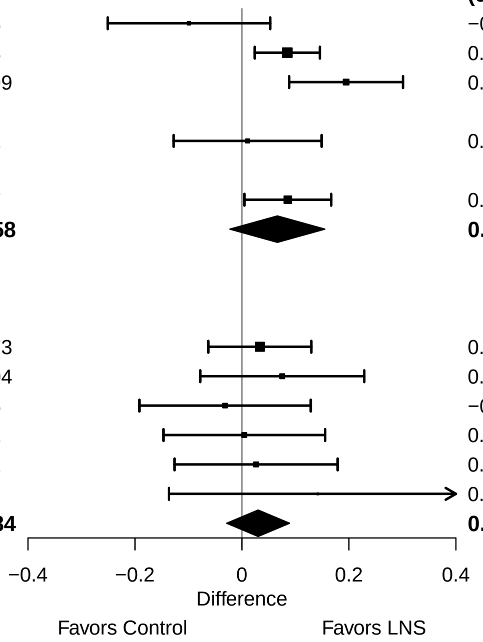

#### Supplemental figure 4J: Mean difference in gross motor z-score

### 4J7: Stratified by Supplement duration

**Supplement duration**
**(p−diff = 0.643)**

| Country | Trial | Tool | N | N | MD (95% CI) | W |
|---|---|---|---|---|---|---|
| **Supplement duration − 12m or less** | | | | | | |
| Bangladesh | JiVitA−4 (21) | BSID−III | 445 | 143 | −0.10 (−0.25, 0.05) | 0.14 |
| Burkina Faso | iLiNS−Zinc (24) | DMC | | | | |
| Ghana | GHANA (25) | | | | | |
| Ghana | iLiNS−DYADG (26) | KDI | 302 | 601 | 0.01 (−0.13, 0.15) | 0.15 |
| Haiti | HAITI (27) | | | | | |
| Madagascar | MAHAY (29) | ASQI | 1613 | 1604 | 0.08 (−0.08, 0.23) | 0.14 |
| Malawi | iLiNS−DYADM (30) | KDI | 214 | 436 | −0.03 (−0.19, 0.13) | 0.13 |
| Malawi | iLiNS−DOSE (31) | KDI | 646 | 221 | 0.00 (−0.15, 0.16) | 0.14 |
| Zimbabwe | SHINE¹ (HIV−) (33), LNS vs SOC | MDAT | 395 | 382 | 0.03 (−0.13, 0.18) | 0.18 |
| Zimbabwe | LNS+WSH vs WSH | MDAT | 446 | 417 | 0.16 (0.00, 0.32) | 0.18 |
| Zimbabwe | SHINE¹ (HIV+) (34), LNS vs SOC | MDAT | 67 | 68 | 0.14 (−0.14, 0.42) | 0.11 |
| Zimbabwe | LNS+WSH vs WSH | MDAT | 103 | 83 | 0.48 (0.24, 0.72) | 0.11 |
| **I² = 0.61, Tau² = 0.01** | | | **4231** | **3955** | **0.05 (−0.05, 0.15)** | |
| **Supplement duration − > 12m** | | | | | | |
| Bangladesh | RDNS (22) | DMC | 1570 | 758 | 0.08 (0.02, 0.15) | 0.26 |
| Bangladesh | WASH−B (23) | EASQ | 1074 | 3279 | 0.10 (0.04, 0.17) | 0.25 |
| Kenya | WASH−B (28) | EASQ | 1362 | 4745 | 0.01 (−0.05, 0.07) | 0.26 |
| Mali | PROMIS CS (32) | DMC | 940 | 957 | 0.09 (0.00, 0.17) | 0.23 |
| **I² = 0.37, Tau² = 0.00** | | | **4946** | **9739** | **0.07 (0.03, 0.11)** | |

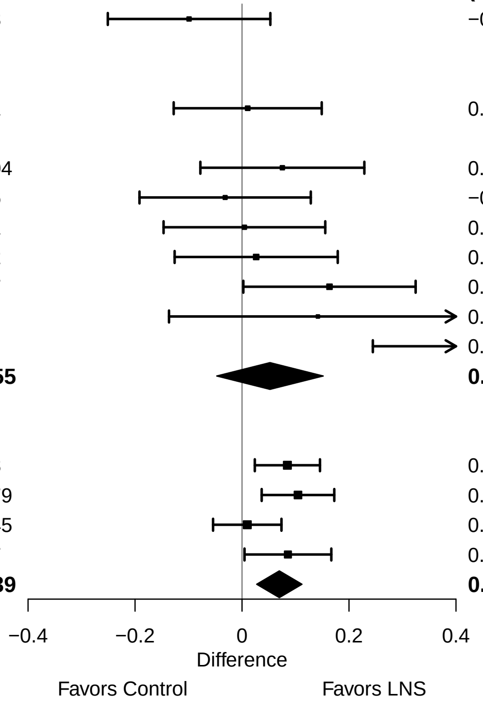

#### Supplemental figure 4J: Mean difference in gross motor z-score

### 4J8: Stratified by Frequency of contact

**Frequency of contact**
**(p−diff = 0.150)**

| Country | Trial | Tool | N | N | MD (95% CI) | W |
|---|---|---|---|---|---|---|
| **Frequency of contact − Monthly** | | | | | | |
| Bangladesh | RDNS (22) | DMC | 1570 | 758 | 0.08 (0.02, 0.15) | 0.22 |
| Haiti | HAITI (27) | | | | | |
| Kenya | WASH−B (28) | EASQ | 1362 | 4745 | 0.01 (−0.05, 0.07) | 0.21 |
| Madagascar | MAHAY (29) | ASQI | 1613 | 1604 | 0.08 (−0.08, 0.23) | 0.12 |
| Mali | PROMIS CS (32) | DMC | 940 | 957 | 0.09 (0.00, 0.17) | 0.19 |
| Zimbabwe | SHINE¹ (HIV−) (33), LNS vs SOC | MDAT | 395 | 382 | 0.03 (−0.13, 0.18) | 0.16 |
| Zimbabwe | LNS+WSH vs WSH | MDAT | 446 | 417 | 0.16 (0.00, 0.32) | 0.16 |
| Zimbabwe | SHINE¹ (HIV+) (34), LNS vs SOC | MDAT | 67 | 68 | 0.14 (−0.14, 0.42) | 0.09 |
| Zimbabwe | LNS+WSH vs WSH | MDAT | 103 | 83 | 0.48 (0.24, 0.72) | 0.09 |
| **I² = 0.59, Tau² = 0.01** | | | **6496** | **9014** | **0.10 (0.02, 0.17)** | |
| **Frequency of contact − Weekly** | | | | | | |
| Bangladesh | JiVitA−4 (21) | BSID−III | 445 | 143 | −0.10 (−0.25, 0.05) | 0.17 |
| Bangladesh | WASH−B (23) | EASQ | 1074 | 3279 | 0.10 (0.04, 0.17) | 0.30 |
| Burkina Faso | iLiNS−Zinc (24) | DMC | | | | |
| Ghana | GHANA (25) | | | | | |
| Ghana | iLiNS−DYADG (26) | KDI | 302 | 601 | 0.01 (−0.13, 0.15) | 0.19 |
| Malawi | iLiNS−DYADM (30) | KDI | 214 | 436 | −0.03 (−0.19, 0.13) | 0.16 |
| Malawi | iLiNS−DOSE (31) | KDI | 646 | 221 | 0.00 (−0.15, 0.16) | 0.17 |
| **I² = 0.49, Tau² = 0.00** | | | **2681** | **4680** | **0.02 (−0.05, 0.09)** | |

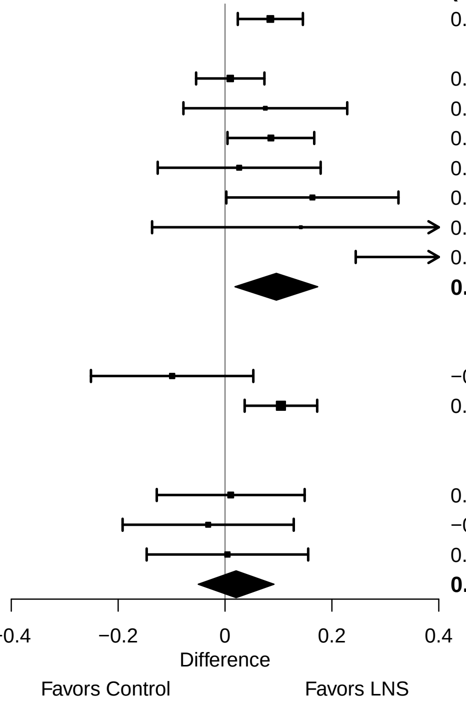

#### Supplemental figure 4J: Mean difference in gross motor z-score

### 4J9: Stratified by Average SQ-LNS compliance

**Average SQ−LNS compliance**
**(p−diff = 0.525)**

| Country | Trial | Tool | N | N | MD (95% CI) | W |
|---|---|---|---|---|---|---|
| **Average SQ−LNS compliance − Low** | | | | | | |
| Ghana | iLiNS−DYADG (26) | KDI | 302 | 601 | 0.01 (−0.13, 0.15) | 0.16 |
| Malawi | iLiNS−DYADM (30) | KDI | 214 | 436 | −0.03 (−0.19, 0.13) | 0.14 |
| Malawi | iLiNS−DOSE (31) | KDI | 646 | 221 | 0.00 (−0.15, 0.16) | 0.15 |
| Mali | PROMIS CS (32) | DMC | 940 | 957 | 0.09 (0.00, 0.17) | 0.23 |
| Zimbabwe | SHINE¹ (HIV−) (33), LNS vs SOC | MDAT | 395 | 382 | 0.03 (−0.13, 0.18) | 0.19 |
| Zimbabwe | LNS+WSH vs WSH | MDAT | 446 | 417 | 0.16 (0.00, 0.32) | 0.19 |
| Zimbabwe | SHINE¹ (HIV+) (34), LNS vs SOC | MDAT | 67 | 68 | 0.14 (−0.14, 0.42) | 0.12 |
| Zimbabwe | LNS+WSH vs WSH | MDAT | 103 | 83 | 0.48 (0.24, 0.72) | 0.12 |
| **I² = 0.56, Tau² = 0.01** | | | **3113** | **3165** | **0.08 (−0.02, 0.18)** | |
| **Average SQ−LNS compliance − High** | | | | | | |
| Bangladesh | JiVitA−4 (21) | BSID−III | 445 | 143 | −0.10 (−0.25, 0.05) | 0.17 |
| Bangladesh | RDNS (22) | DMC | 1570 | 758 | 0.08 (0.02, 0.15) | 0.28 |
| Bangladesh | WASH−B (23) | EASQ | 1074 | 3279 | 0.10 (0.04, 0.17) | 0.27 |
| Burkina Faso | iLiNS−Zinc (24) | DMC | | | | |
| Ghana | GHANA (25) | | | | | |
| Haiti | HAITI (27) | | | | | |
| Kenya | WASH−B (28) | EASQ | 1362 | 4745 | 0.01 (−0.05, 0.07) | 0.28 |
| **I² = 0.66, Tau² = 0.00** | | | **4451** | **8925** | **0.04 (−0.04, 0.12)** | |

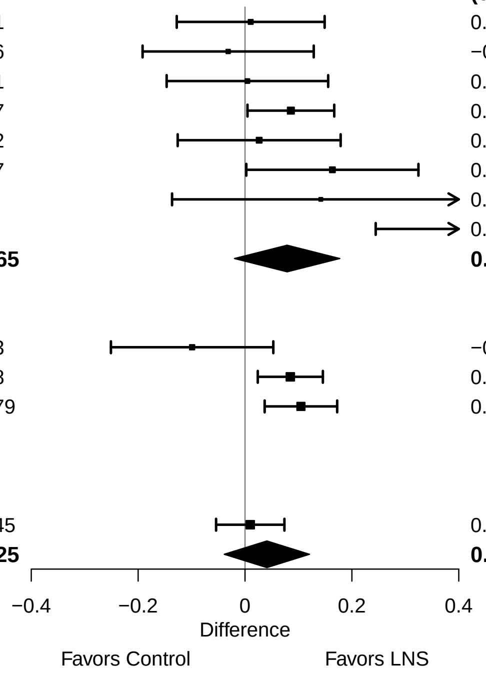

### Supplemental figure 4K: Mean difference in fine motor z-score

#### 4K1: Stratified by Geographic region (insufficient comparisons)

**Supplemental figure 4K: Mean difference in fine motor z-score**

#### 4K2: Stratified by Stunting burden (insufficient comparisons)

#### Supplemental figure 4K: Mean difference in fine motor z-score

### 4K3: Stratified by Malaria prevalence

**Malaria prevalence**
**(p−diff = 0.626)**

| Country | Trial | Tool | N | N | MD (95% CI) | W |
|---|---|---|---|---|---|---|
| **Malaria prevalence − Less than 10%** | | | | | | |
| Bangladesh | JiVitA−4 (21) | BSID−III | 445 | 143 | 0.14 (−0.05, 0.34) | 0.11 |
| Bangladesh | RDNS (22) | DMC | 1641 | 804 | 0.10 (−0.01, 0.20) | 0.35 |
| Bangladesh | WASH−B (23) | EASQ | | | | |
| Haiti | HAITI (27) | | | | | |
| Kenya | WASH−B (28) | EASQ | | | | |
| Madagascar | MAHAY (29) | ASQI | 1613 | 1604 | 0.05 (−0.17, 0.26) | 0.08 |
| Zimbabwe | SHINE¹ (HIV−) (33), LNS vs SOC | MDAT | 395 | 382 | −0.03 (−0.18, 0.11) | 0.38 |
| Zimbabwe | LNS+WSH vs WSH | MDAT | 446 | 417 | 0.17 (0.03, 0.31) | 0.38 |
| Zimbabwe | SHINE¹ (HIV+) (34), LNS vs SOC | MDAT | 67 | 68 | −0.04 (−0.40, 0.31) | 0.08 |
| Zimbabwe | LNS+WSH vs WSH | MDAT | 103 | 83 | 0.31 (0.03, 0.59) | 0.08 |
| **I² = 0.00, Tau² = 0.00** | | | **4710** | **3501** | **0.10 (0.03, 0.16)** | |
| **Malaria prevalence − At least 10%** | | | | | | |
| Burkina Faso | iLiNS−Zinc (24) | DMC | | | | |
| Ghana | GHANA (25) | | | | | |
| Ghana | iLiNS−DYADG (26) | KDI | 302 | 601 | 0.04 (−0.09, 0.17) | 0.36 |
| Malawi | iLiNS−DYADM (30) | KDI | 214 | 436 | 0.08 (−0.08, 0.24) | 0.24 |
| Malawi | iLiNS−DOSE (31) | KDI | 646 | 221 | 0.09 (−0.06, 0.24) | 0.26 |
| Mali | PROMIS CS (32) | DMC | 905 | 924 | 0.11 (−0.11, 0.32) | 0.13 |
| **I² = 0.00, Tau² = 0.00** | | | **2067** | **2182** | **0.07 (−0.01, 0.15)** | |

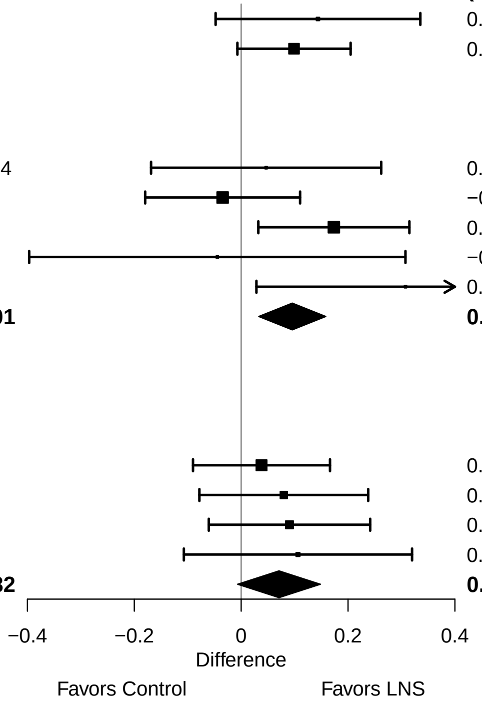

#### Supplemental figure 4K: Mean difference in fine motor z-score

### 4K4: Stratified by Anemia prevalence

**Anemia prevalence**
**(p−diff = 0.928)**

| Country | Trial | Tool | N | N | MD (95% CI) | W |
|---|---|---|---|---|---|---|
| **Anemia prevalence − High** | | | | | | |
| Burkina Faso | iLiNS−Zinc (24) | DMC | | | | |
| Ghana | GHANA (25) | | | | | |
| Haiti | HAITI (27) | | | | | |
| Malawi | iLiNS−DYADM (30) | KDI | 214 | 436 | 0.08 (−0.08, 0.24) | 0.38 |
| Malawi | iLiNS−DOSE (31) | KDI | 646 | 221 | 0.09 (−0.06, 0.24) | 0.41 |
| Mali | PROMIS CS (32) | DMC | 905 | 924 | 0.11 (−0.11, 0.32) | 0.21 |
| **I² = 0.00, Tau² = 0.00** | | | **1765** | **1581** | **0.09 (−0.01, 0.19)** | |
| **Anemia prevalence − Moderate** | | | | | | |
| Bangladesh | JiVitA−4 (21) | BSID−III | 445 | 143 | 0.14 (−0.05, 0.34) | 0.09 |
| Bangladesh | RDNS (22) | DMC | 1641 | 804 | 0.10 (−0.01, 0.20) | 0.28 |
| Bangladesh | WASH−B (23) | EASQ | | | | |
| Ghana | iLiNS−DYADG (26) | KDI | 302 | 601 | 0.04 (−0.09, 0.17) | 0.19 |
| Kenya | WASH−B (28) | EASQ | | | | |
| Madagascar | MAHAY (29) | ASQI | 1613 | 1604 | 0.05 (−0.17, 0.26) | 0.07 |
| Zimbabwe | SHINE¹ (HIV−) (33), LNS vs SOC | MDAT | 395 | 382 | −0.03 (−0.18, 0.11) | 0.31 |
| Zimbabwe | LNS+WSH vs WSH | MDAT | 446 | 417 | 0.17 (0.03, 0.31) | 0.31 |
| Zimbabwe | SHINE¹ (HIV+) (34), LNS vs SOC | MDAT | 67 | 68 | −0.04 (−0.40, 0.31) | 0.06 |
| Zimbabwe | LNS+WSH vs WSH | MDAT | 103 | 83 | 0.31 (0.03, 0.59) | 0.06 |
| **I² = 0.00, Tau² = 0.00** | | | **5012** | **4102** | **0.08 (0.03, 0.14)** | |

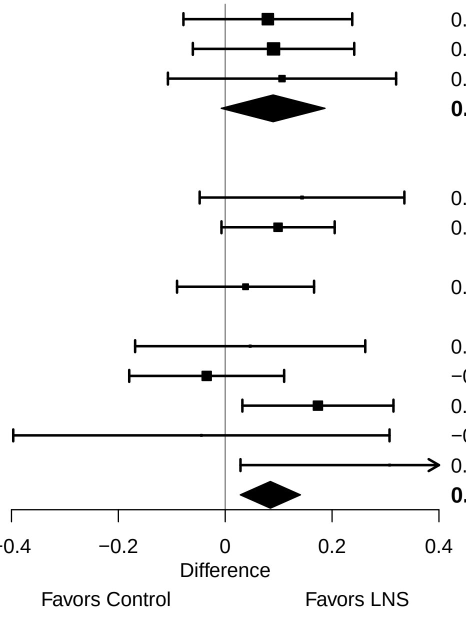

#### Supplemental figure 4K: Mean difference in fine motor z-score

### 4K5: Stratified by Source water quality

**Source water quality**
**(p−diff = 0.234)**

**Source water quality − Improved**

| **Country** | **Trial** | **Tool** | **N** | **N** | **MD (95% CI)** | **W** |
|---|---|---|---|---|---|---|
| Bangladesh | JiVitA−4 (21) | BSID−III | 445 | 143 | 0.14 (−0.05, 0.34) | 0.10 |
| Bangladesh | RDNS (22) | DMC | 1641 | 804 | 0.10 (−0.01, 0.20) | 0.34 |
| Bangladesh | WASH−B (23) | EASQ | | | | |
| Ghana | GHANA (25) | | | | | |
| Ghana | iLiNS−DYADG (26) | KDI | 302 | 601 | 0.04 (−0.09, 0.17) | 0.23 |
| Haiti | HAITI (27) | | | | | |
| Malawi | iLiNS−DYADM (30) | KDI | 214 | 436 | 0.08 (−0.08, 0.24) | 0.15 |
| Malawi | iLiNS−DOSE (31) | KDI | 646 | 221 | 0.09 (−0.06, 0.24) | 0.17 |
| **I² = 0.00, Tau² = 0.00** | | | **3248** | **2205** | **0.09 (0.02, 0.15)** | |

**Source water quality − Unimproved**

| **Country** | **Trial** | **Tool** | **N** | **N** | **MD (95% CI)** | **W** |
|---|---|---|---|---|---|---|
| Burkina Faso | iLiNS−Zinc (24) | DMC | | | | |
| Kenya | WASH−B (28) | EASQ | | | | |
| Madagascar | MAHAY (29) | ASQI | 1613 | 1604 | 0.05 (−0.17, 0.26) | 0.22 |
| Mali | PROMIS CS (32) | DMC | 905 | 924 | 0.11 (−0.11, 0.32) | 0.22 |
| Zimbabwe | SHINE (HIV−) (33) | MDAT | 395 | 382 | −0.03 (−0.18, 0.11) | 0.48 |
| Zimbabwe | SHINE (HIV+) (34) | MDAT | 67 | 68 | −0.04 (−0.40, 0.31) | 0.08 |
| **I² = 0.00, Tau² = 0.00** | | | **2980** | **2978** | **0.01 (−0.09, 0.11)** | |

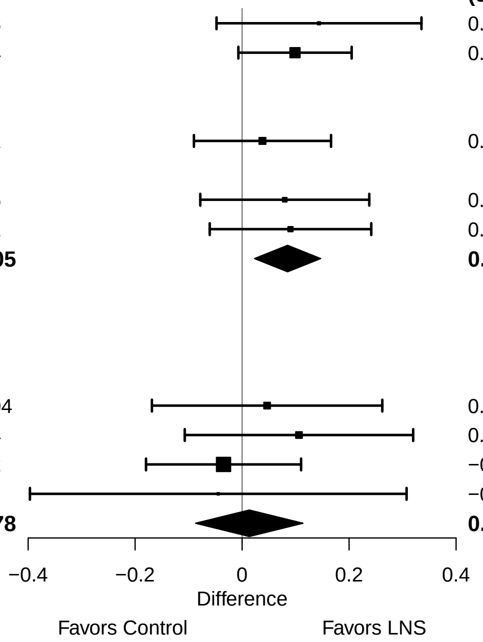

#### Supplemental figure 4K: Mean difference in fine motor z-score

### 4K6: Stratified by Sanitation

**Sanitation**
**(p−diff = 0.364)**

| Country | Trial | Tool | N | N | MD (95% CI) | W |
|---|---|---|---|---|---|---|
| **Sanitation − Improved** | | | | | | |
| Bangladesh | JiVitA−4 (21) | BSID−III | 445 | 143 | 0.14 (−0.05, 0.34) | 0.14 |
| Bangladesh | RDNS (22) | DMC | 1641 | 804 | 0.10 (−0.01, 0.20) | 0.45 |
| Bangladesh | WASH−B (23) | EASQ | | | | |
| Ghana | GHANA (25) | | | | | |
| Ghana | iLiNS−DYADG (26) | KDI | 302 | 601 | 0.04 (−0.09, 0.17) | 0.31 |
| Haiti | HAITI (27) | | | | | |
| Mali | PROMIS CS (32) | DMC | 905 | 924 | 0.11 (−0.11, 0.32) | 0.11 |
| **I² = 0.00, Tau² = 0.00** | | | **3293** | **2472** | **0.09 (0.02, 0.16)** | |
| **Sanitation − Unimproved** | | | | | | |
| Burkina Faso | iLiNS−Zinc (24) | DMC | | | | |
| Kenya | WASH−B (28) | EASQ | | | | |
| Madagascar | MAHAY (29) | ASQI | 1613 | 1604 | 0.05 (−0.17, 0.26) | 0.13 |
| Malawi | iLiNS−DYADM (30) | KDI | 214 | 436 | 0.08 (−0.08, 0.24) | 0.25 |
| Malawi | iLiNS−DOSE (31) | KDI | 646 | 221 | 0.09 (−0.06, 0.24) | 0.27 |
| Zimbabwe | SHINE (HIV−) (33) | MDAT | 395 | 382 | −0.03 (−0.18, 0.11) | 0.30 |
| Zimbabwe | SHINE (HIV+) (34) | MDAT | 67 | 68 | −0.04 (−0.40, 0.31) | 0.05 |
| **I² = 0.00, Tau² = 0.00** | | | **2935** | **2711** | **0.04 (−0.04, 0.12)** | |

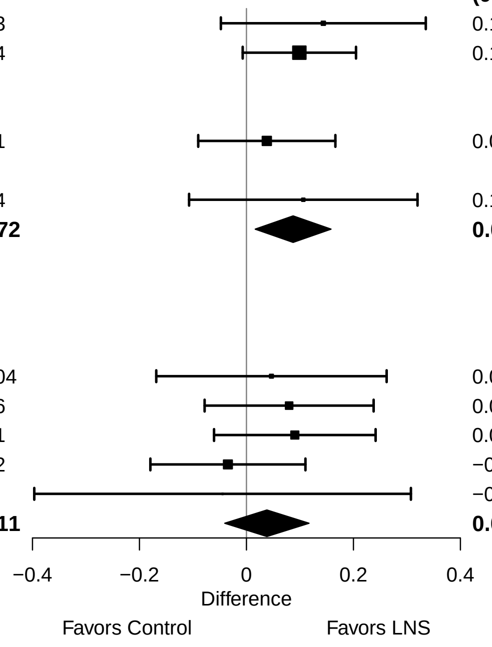

**Supplemental figure 4K: Mean difference in fine motor z-score**

#### 4K7: Stratified by Supplement duration (insufficient comparisons)

#### Supplemental figure 4K: Mean difference in fine motor z-score

### 4K8: Stratified by Frequency of contact

**Frequency of contact**
**(p−diff = 0.791)**

| Country | Trial | Tool | N | N | MD (95% CI) | W |
|---|---|---|---|---|---|---|
| **Frequency of contact − Monthly** | | | | | | |
| Bangladesh | RDNS (22) | DMC | 1641 | 804 | 0.10 (−0.01, 0.20) | 0.36 |
| Haiti | HAITI (27) | | | | | |
| Kenya | WASH−B (28) | EASQ | | | | |
| Madagascar | MAHAY (29) | ASQI | 1613 | 1604 | 0.05 (−0.17, 0.26) | 0.09 |
| Mali | PROMIS CS (32) | DMC | 905 | 924 | 0.11 (−0.11, 0.32) | 0.09 |
| Zimbabwe | SHINE[1] (HIV−) (33), LNS vs SOC | MDAT | 395 | 382 | −0.03 (−0.18, 0.11) | 0.39 |
| Zimbabwe | LNS+WSH vs WSH | MDAT | 446 | 417 | 0.17 (0.03, 0.31) | 0.39 |
| Zimbabwe | SHINE[1] (HIV+) (34), LNS vs SOC | MDAT | 67 | 68 | −0.04 (−0.40, 0.31) | 0.08 |
| Zimbabwe | LNS+WSH vs WSH | MDAT | 103 | 83 | 0.31 (0.03, 0.59) | 0.08 |
| **I² = 0.00, Tau² = 0.00** | | | **5170** | **4282** | **0.09 (0.03, 0.15)** | |
| **Frequency of contact − Weekly** | | | | | | |
| Bangladesh | JiVitA−4 (21) | BSID−III | 445 | 143 | 0.14 (−0.05, 0.34) | 0.16 |
| Bangladesh | WASH−B (23) | EASQ | | | | |
| Burkina Faso | iLiNS−Zinc (24) | DMC | | | | |
| Ghana | GHANA (25) | | | | | |
| Ghana | iLiNS−DYADG (26) | KDI | 302 | 601 | 0.04 (−0.09, 0.17) | 0.35 |
| Malawi | iLiNS−DYADM (30) | KDI | 214 | 436 | 0.08 (−0.08, 0.24) | 0.23 |
| Malawi | iLiNS−DOSE (31) | KDI | 646 | 221 | 0.09 (−0.06, 0.24) | 0.25 |
| **I² = 0.00, Tau² = 0.00** | | | **1607** | **1401** | **0.08 (0.00, 0.15)** | |

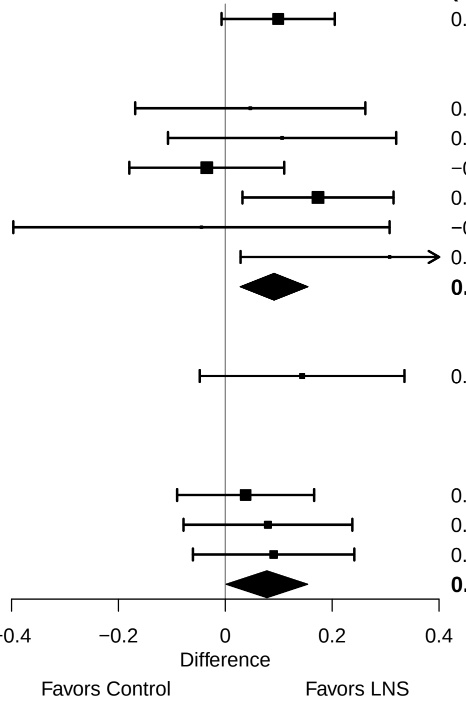

**Supplemental figure 4K: Mean difference in fine motor z-score**

#### 4K9: Stratified by Average SQ-LNS compliance (insufficient comparisons)

### Supplemental figure 4L: Mean difference in executive function z-score

#### 4L1: Stratified by Geographic region (insufficient comparisons)

**Supplemental figure 4L: Mean difference in executive function z-score**

#### 4L2: Stratified by Stunting burden (insufficient comparisons)

#### Supplemental figure 4L: Mean difference in executive function z-score

### 4L3: Stratified by Malaria prevalence

**Malaria prevalence**
**(p−diff = 0.242)**

| Country | Trial | | Tool | N | N | MD (95% CI) | W |
|---|---|---|---|---|---|---|---|
| **Malaria prevalence − Less than 10%** | | | | | | | |
| Bangladesh | JiVitA−4 (21) | | | | | | |
| Bangladesh | RDNS (22) | | A not B task | 533 | 266 | 0.10 (−0.02, 0.22) | 0.20 |
| Bangladesh | WASH−B (23) | | A not B task | 1097 | 3327 | 0.00 (−0.07, 0.07) | 0.53 |
| Haiti | HAITI (27) | | | | | | |
| Kenya | WASH−B (28) | | | | | | |
| Madagascar | MAHAY (29) | | | | | | |
| Zimbabwe | SHINE¹ (HIV−) (33), | LNS vs SOC | A not B task | 370 | 357 | −0.05 (−0.21, 0.10) | 0.21 |
| Zimbabwe | | LNS+WSH vs WSH | A not B task | 417 | 382 | 0.12 (−0.05, 0.29) | 0.21 |
| Zimbabwe | SHINE¹ (HIV+) (34), | LNS vs SOC | A not B task | 63 | 57 | −0.16 (−0.49, 0.16) | 0.06 |
| Zimbabwe | | LNS+WSH vs WSH | A not B task | 94 | 76 | −0.05 (−0.35, 0.25) | 0.06 |
| **I² = 0.06, Tau² = 0.00** | | | | **2574** | **4465** | **0.02 (−0.04, 0.08)** | |
| **Malaria prevalence − At least 10%** | | | | | | | |
| Burkina Faso | iLiNS−Zinc (24) | | | | | | |
| Ghana | GHANA (25) | | | | | | |
| Ghana | iLiNS−DYADG (26) | | A not B task | 290 | 576 | 0.02 (−0.12, 0.17) | 0.44 |
| Malawi | iLiNS−DYADM (30) | | A not B task | 182 | 369 | −0.09 (−0.27, 0.09) | 0.28 |
| Malawi | iLiNS−DOSE (31) | | A not B task | 475 | 164 | −0.11 (−0.29, 0.07) | 0.28 |
| Mali | PROMIS CS (32) | | | | | | |
| **I² = 0.00, Tau² = 0.00** | | | | **947** | **1109** | **−0.04 (−0.14, 0.05)** | |

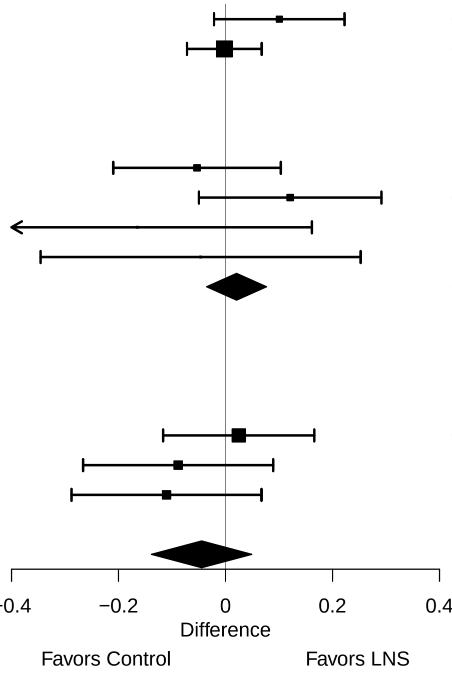

**Supplemental figure 4L: Mean difference in executive function z-score**

#### 4L4: Stratified by Anemia prevalence (insufficient comparisons)

**Supplemental figure 4L: Mean difference in executive function z-score**

#### 4L5: Stratified by Source water quality (insufficient comparisons)

#### Supplemental figure 4L: Mean difference in executive function z-score

### 4L6: Stratified by Sanitation

**Sanitation**
**(p−diff = 0.012)**

| Country | Trial | Tool | N | N | MD (95% CI) | W |
|---|---|---|---|---|---|---|
| **Sanitation − Improved** | | | | | | |
| Bangladesh | JiVitA−4 (21) | | | | | |
| Bangladesh | RDNS (22) | A not B task | 533 | 266 | 0.10 (−0.02, 0.22) | 0.32 |
| Bangladesh | WASH−B (23) | A not B task | 541 | 1099 | 0.05 (−0.05, 0.15) | 0.42 |
| Ghana | GHANA (25) | | | | | |
| Ghana | iLiNS−DYADG (26) | A not B task | 290 | 576 | 0.02 (−0.12, 0.17) | 0.26 |
| Haiti | HAITI (27) | | | | | |
| Mali | PROMIS CS (32) | | | | | |
| **I² = 0.00, Tau² = 0.00** | | | **1364** | **1941** | **0.06 (−0.01, 0.13)** | |
| **Sanitation − Unimproved** | | | | | | |
| Burkina Faso | iLiNS−Zinc (24) | | | | | |
| Kenya | WASH−B (28) | | | | | |
| Madagascar | MAHAY (29) | | | | | |
| Malawi | iLiNS−DYADM (30) | A not B task | 182 | 369 | −0.09 (−0.27, 0.09) | 0.28 |
| Malawi | iLiNS−DOSE (31) | A not B task | 475 | 164 | −0.11 (−0.29, 0.07) | 0.28 |
| Zimbabwe | SHINE (HIV−) (33) | A not B task | 370 | 357 | −0.05 (−0.21, 0.10) | 0.35 |
| Zimbabwe | SHINE (HIV+) (34) | A not B task | 63 | 57 | −0.16 (−0.49, 0.16) | 0.09 |
| **I² = 0.00, Tau² = 0.00** | | | **1090** | **947** | **−0.09 (−0.18, 0.01)** | |

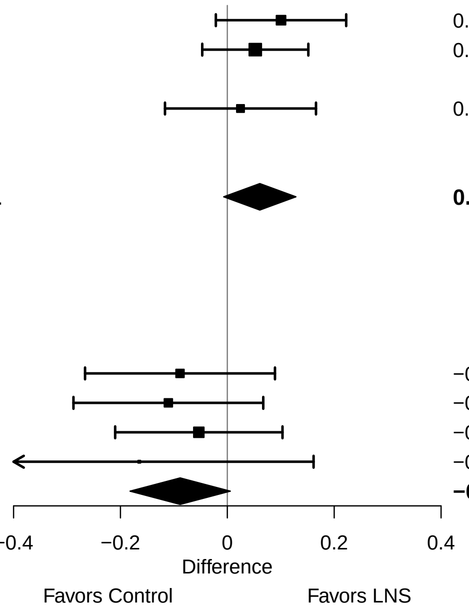

**Supplemental figure 4L: Mean difference in executive function z-score**

#### 4L7: Stratified by Supplement duration (insufficient comparisons)

#### Supplemental figure 4L: Mean difference in executive function z-score

#### 4L8: Stratified by Frequency of contact

**Frequency of contact**
**(p−diff = 0.192)**

| Country | Trial | | Tool | N | N | MD (95% CI) | W |
|---|---|---|---|---|---|---|---|
| **Frequency of contact − Monthly** | | | | | | | |
| Bangladesh | RDNS (22) | | A not B task | 533 | 266 | 0.10 (−0.02, 0.22) | 0.42 |
| Haiti | HAITI (27) | | | | | | |
| Kenya | WASH−B (28) | | | | | | |
| Madagascar | MAHAY (29) | | | | | | |
| Mali | PROMIS CS (32) | | | | | | |
| Zimbabwe | SHINE¹ (HIV−) (33), | LNS vs SOC | A not B task | 370 | 357 | −0.05 (−0.21, 0.10) | 0.45 |
| Zimbabwe | | LNS+WSH vs WSH | A not B task | 417 | 382 | 0.12 (−0.05, 0.29) | 0.45 |
| Zimbabwe | SHINE¹ (HIV+) (34), | LNS vs SOC | A not B task | 63 | 57 | −0.16 (−0.49, 0.16) | 0.13 |
| Zimbabwe | | LNS+WSH vs WSH | A not B task | 94 | 76 | −0.05 (−0.35, 0.25) | 0.13 |
| **$I^2$ = 0.15, $Tau^2$ = 0.00** | | | | **1477** | **1138** | **0.04 (−0.05, 0.13)** | |
| **Frequency of contact − Weekly** | | | | | | | |
| Bangladesh | JiVitA−4 (21) | | | | | | |
| Bangladesh | WASH−B (23) | | A not B task | 1097 | 3327 | 0.00 (−0.07, 0.07) | 0.61 |
| Burkina Faso | iLiNS−Zinc (24) | | | | | | |
| Ghana | GHANA (25) | | | | | | |
| Ghana | iLiNS−DYADG (26) | | A not B task | 290 | 576 | 0.02 (−0.12, 0.17) | 0.17 |
| Malawi | iLiNS−DYADM (30) | | A not B task | 182 | 369 | −0.09 (−0.27, 0.09) | 0.11 |
| Malawi | iLiNS−DOSE (31) | | A not B task | 475 | 164 | −0.11 (−0.29, 0.07) | 0.11 |
| **$I^2$ = 0.00, $Tau^2$ = 0.00** | | | | **2044** | **4436** | **−0.02 (−0.07, 0.04)** | |

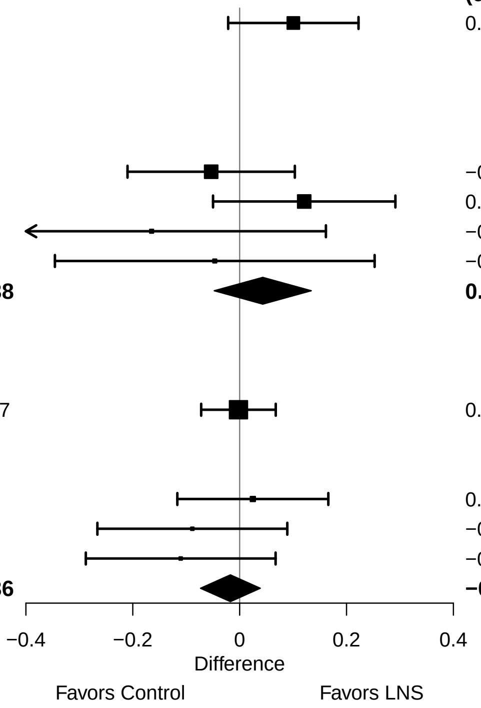

**Supplemental figure 4L: Mean difference in executive function z-score**

#### 4L9: Stratified by Average SQ-LNS compliance (insufficient comparisons)

### Supplemental figure 4M: Executive function lowest decile prevalence ratio

#### 4M1: Stratified by Geographic region (insufficient comparisons)

**Supplemental figure 4M: Executive function lowest decile prevalence ratio**

#### 4M2: Stratified by Stunting burden (insufficient comparisons)

#### Supplemental figure 4M: Executive function lowest decile prevalence ratio

### 4M3: Stratified by Malaria prevalence

**Malaria prevalence**
**(p−diff = 0.990)**

| Country | Trial | Tool | N | N | PR (95% CI) | W |
|---|---|---|---|---|---|---|
| **Malaria prevalence − Less than 10%** | | | | | | |
| Bangladesh | JiVitA−4 (21) | | | | | |
| Bangladesh | RDNS (22) | A not B task | 533 | 266 | 0.88 (0.60, 1.29) | 0.21 |
| Bangladesh | WASH−B (23) | A not B task | 1097 | 3327 | 0.95 (0.77, 1.18) | 0.42 |
| Haiti | HAITI (27) | | | | | |
| Kenya | WASH−B (28) | | | | | |
| Madagascar | MAHAY (29) | | | | | |
| Zimbabwe | SHINE¹ (HIV−) (33), LNS vs SOC | A not B task | 370 | 357 | 0.82 (0.53, 1.26) | 0.28 |
| Zimbabwe | LNS+WSH vs WSH | A not B task | 417 | 382 | 0.78 (0.50, 1.21) | 0.28 |
| Zimbabwe | SHINE¹ (HIV+) (34), LNS vs SOC | A not B task | 63 | 57 | 1.09 (0.37, 3.22) | 0.09 |
| Zimbabwe | LNS+WSH vs WSH | A not B task | 94 | 76 | 2.10 (0.92, 4.82) | 0.09 |
| **$I^2$ = 0.22, $Tau^2$ = 0.02** | | | **2574** | **4465** | **0.94 (0.75, 1.16)** | |
| **Malaria prevalence − At least 10%** | | | | | | |
| Burkina Faso | iLiNS−Zinc (24) | | | | | |
| Ghana | GHANA (25) | | | | | |
| Ghana | iLiNS−DYADG (26) | A not B task | 290 | 576 | 0.78 (0.50, 1.20) | 0.44 |
| Malawi | iLiNS−DYADM (30) | A not B task | 182 | 369 | 0.81 (0.47, 1.41) | 0.30 |
| Malawi | iLiNS−DOSE (31) | A not B task | 475 | 164 | 1.50 (0.82, 2.73) | 0.26 |
| Mali | PROMIS CS (32) | | | | | |
| **$I^2$ = 0.39, $Tau^2$ = 0.05** | | | **947** | **1109** | **0.95 (0.64, 1.41)** | |

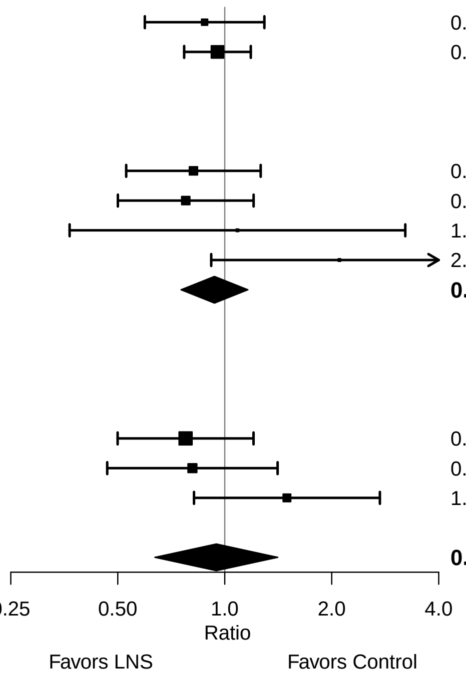

**Supplemental figure 4M: Executive function lowest decile prevalence ratio**

#### 4M4: Stratified by Anemia prevalence (insufficient comparisons)

**Supplemental figure 4M: Executive function lowest decile prevalence ratio**

#### 4M5: Stratified by Source water quality (insufficient comparisons)

#### Supplemental figure 4M: Executive function lowest decile prevalence ratio

### 4M6: Stratified by Sanitation

**Sanitation**
**(p−diff = 0.286)**

| Country | Trial | Tool | N | N | PR (95% CI) | W |
|---|---|---|---|---|---|---|
| **Sanitation − Improved** | | | | | | |
| Bangladesh | JiVitA−4 (21) | | | | | |
| Bangladesh | RDNS (22) | A not B task | 533 | 266 | 0.88 (0.60, 1.29) | 0.28 |
| Bangladesh | WASH−B (23) | A not B task | 541 | 1099 | 0.74 (0.56, 1.00) | 0.50 |
| Ghana | GHANA (25) | | | | | |
| Ghana | iLiNS−DYADG (26) | A not B task | 290 | 576 | 0.78 (0.50, 1.20) | 0.22 |
| Haiti | HAITI (27) | | | | | |
| Mali | PROMIS CS (32) | | | | | |
| **I² = 0.00, Tau² = 0.00** | | | **1364** | **1941** | **0.79 (0.64, 0.97)** | |
| **Sanitation − Unimproved** | | | | | | |
| Burkina Faso | iLiNS−Zinc (24) | | | | | |
| Kenya | WASH−B (28) | | | | | |
| Madagascar | MAHAY (29) | | | | | |
| Malawi | iLiNS−DYADM (30) | A not B task | 182 | 369 | 0.81 (0.47, 1.41) | 0.27 |
| Malawi | iLiNS−DOSE (31) | A not B task | 475 | 164 | 1.50 (0.82, 2.73) | 0.23 |
| Zimbabwe | SHINE (HIV−) (33) | A not B task | 370 | 357 | 0.82 (0.53, 1.26) | 0.43 |
| Zimbabwe | SHINE (HIV+) (34) | A not B task | 63 | 57 | 1.09 (0.37, 3.22) | 0.07 |
| **I² = 0.01, Tau² = 0.00** | | | **1090** | **947** | **0.95 (0.72, 1.27)** | |

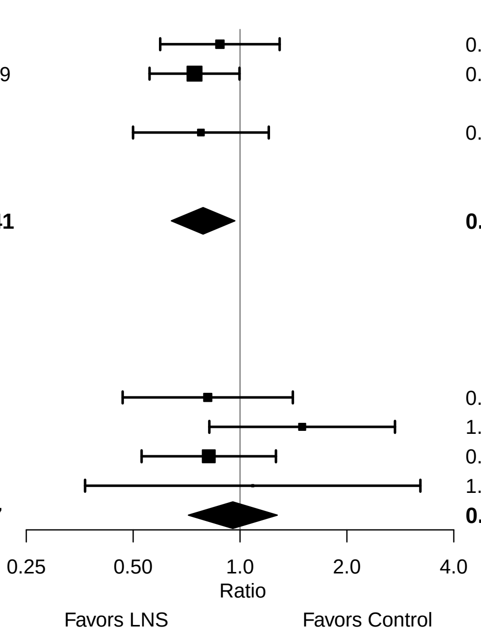

##### Supplemental figure 4M: Executive function lowest decile prevalence ratio

#### 4M7: Stratified by Supplement duration (insufficient comparisons)

#### Supplemental figure 4M: Executive function lowest decile prevalence ratio

### 4M8: Stratified by Frequency of contact

**Frequency of contact**
**(p−diff = 0.724)**

| Country | Trial | Tool | N | N | PR (95% CI) | W |
|---|---|---|---|---|---|---|
| **Frequency of contact − Monthly** | | | | | | |
| Bangladesh | RDNS (22) | A not B task | 533 | 266 | 0.88 (0.60, 1.29) | 0.36 |
| Haiti | HAITI (27) | | | | | |
| Kenya | WASH−B (28) | | | | | |
| Madagascar | MAHAY (29) | | | | | |
| Mali | PROMIS CS (32) | | | | | |
| Zimbabwe | SHINE[1] (HIV−) (33), LNS vs SOC | A not B task | 370 | 357 | 0.82 (0.53, 1.26) | 0.49 |
| Zimbabwe | LNS+WSH vs WSH | A not B task | 417 | 382 | 0.78 (0.50, 1.21) | 0.49 |
| Zimbabwe | SHINE[1] (HIV+) (34), LNS vs SOC | A not B task | 63 | 57 | 1.09 (0.37, 3.22) | 0.15 |
| Zimbabwe | LNS+WSH vs WSH | A not B task | 94 | 76 | 2.10 (0.92, 4.82) | 0.15 |
| **$I^2$ = 0.46, $Tau^2$ = 0.07** | | | **1477** | **1138** | **0.96 (0.66, 1.41)** | |
| **Frequency of contact − Weekly** | | | | | | |
| Bangladesh | JiVitA−4 (21) | | | | | |
| Bangladesh | WASH−B (23) | A not B task | 1097 | 3327 | 0.95 (0.77, 1.18) | 0.52 |
| Burkina Faso | iLiNS−Zinc (24) | | | | | |
| Ghana | GHANA (25) | | | | | |
| Ghana | iLiNS−DYADG (26) | A not B task | 290 | 576 | 0.78 (0.50, 1.20) | 0.21 |
| Malawi | iLiNS−DYADM (30) | A not B task | 182 | 369 | 0.81 (0.47, 1.41) | 0.14 |
| Malawi | iLiNS−DOSE (31) | A not B task | 475 | 164 | 1.50 (0.82, 2.73) | 0.12 |
| **$I^2$ = 0.09, $Tau^2$ = 0.01** | | | **2044** | **4436** | **0.94 (0.77, 1.16)** | |

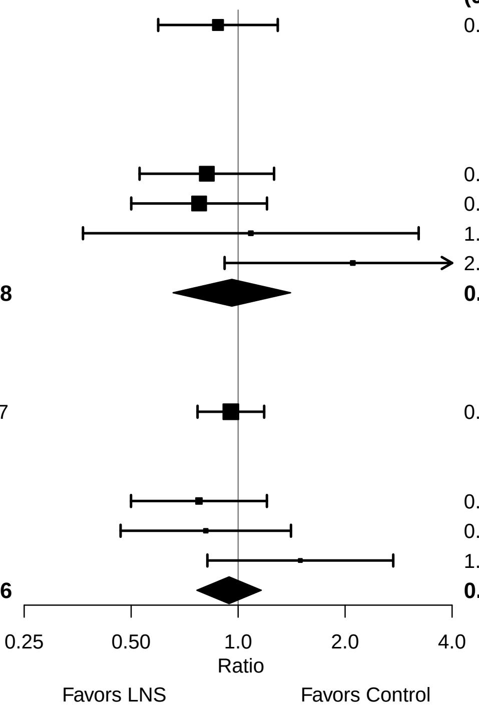

#### Supplemental figure 4M: Executive function lowest decile prevalence ratio

### 4M9: Stratified by Average SQ-LNS compliance (insufficient comparisons)

### Supplemental figure 4N: Executive function lowest decile prevalence difference

#### 4N1: Stratified by Geographic region (insufficient comparisons)

**Supplemental figure 4N: Executive function lowest decile prevalence difference**

#### 4N2: Stratified by Stunting burden (insufficient comparisons)

#### Supplemental figure 4N: Executive function lowest decile prevalence difference

### 4N3: Stratified by Malaria prevalence

**Malaria prevalence**
**(p−diff = 0.865)**

| Country | Trial | | Tool | N | N | PD (95% CI) | W |
|---|---|---|---|---|---|---|---|
| **Malaria prevalence − Less than 10%** | | | | | | | |
| Bangladesh | JiVitA−4 (21) | | | | | | |
| Bangladesh | RDNS (22) | | A not B task | 533 | 266 | −0.01 (−0.06, 0.03) | 0.17 |
| Bangladesh | WASH−B (23) | | A not B task | 1097 | 3327 | 0.00 (−0.02, 0.02) | 0.44 |
| Haiti | HAITI (27) | | | | | | |
| Kenya | WASH−B (28) | | | | | | |
| Madagascar | MAHAY (29) | | | | | | |
| Zimbabwe | SHINE[1] (HIV−) (33), | LNS vs SOC | A not B task | 370 | 357 | −0.02 (−0.06, 0.02) | 0.30 |
| Zimbabwe | | LNS+WSH vs WSH | A not B task | 417 | 382 | −0.03 (−0.07, 0.02) | 0.30 |
| Zimbabwe | SHINE[1] (HIV+) (34), | LNS vs SOC | A not B task | 63 | 57 | 0.01 (−0.10, 0.11) | 0.09 |
| Zimbabwe | | LNS+WSH vs WSH | A not B task | 94 | 76 | 0.07 (−0.02, 0.16) | 0.09 |
| **$I^2$ = 0.15, $Tau^2$ = 0.00** | | | | **2574** | **4465** | **−0.01 (−0.03, 0.01)** | |
| **Malaria prevalence − At least 10%** | | | | | | | |
| Burkina Faso | iLiNS−Zinc (24) | | | | | | |
| Ghana | GHANA (25) | | | | | | |
| Ghana | iLiNS−DYADG (26) | | A not B task | 290 | 576 | −0.02 (−0.07, 0.02) | 0.40 |
| Malawi | iLiNS−DYADM (30) | | A not B task | 182 | 369 | −0.02 (−0.07, 0.03) | 0.28 |
| Malawi | iLiNS−DOSE (31) | | A not B task | 475 | 164 | 0.04 (−0.01, 0.09) | 0.31 |
| Mali | PROMIS CS (32) | | | | | | |
| **$I^2$ = 0.50, $Tau^2$ = 0.00** | | | | **947** | **1109** | **0.00 (−0.04, 0.03)** | |

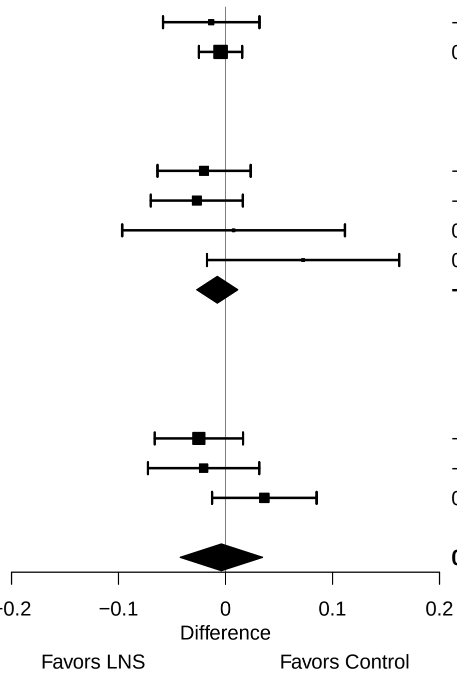

**Supplemental figure 4N: Executive function lowest decile prevalence difference**

#### 4N4: Stratified by Anemia prevalence (insufficient comparisons)

**Supplemental figure 4N: Executive function lowest decile prevalence difference**

#### 4N5: Stratified by Source water quality (insufficient comparisons)

#### Supplemental figure 4N: Executive function lowest decile prevalence difference

### 4N6: Stratified by Sanitation

**Sanitation**
**(p−diff = 0.181)**

**Sanitation − Improved**

| **Country** | **Trial** | **Tool** | **N** | **N** | **PD (95% CI)** | **W** |
|---|---|---|---|---|---|---|
| Bangladesh | JiVitA−4 (21) | | | | | |
| Bangladesh | RDNS (22) | A not B task | 533 | 266 | −0.01 (−0.06, 0.03) | 0.23 |
| Bangladesh | WASH−B (23) | A not B task | 541 | 1099 | −0.03 (−0.06, 0.00) | 0.50 |
| Ghana | GHANA (25) | | | | | |
| Ghana | iLiNS−DYADG (26) | A not B task | 290 | 576 | −0.02 (−0.07, 0.02) | 0.27 |
| Haiti | HAITI (27) | | | | | |
| Mali | PROMIS CS (32) | | | | | |
| **I² = 0.00, Tau² = 0.00** | | | **1364** | **1941** | **−0.03 (−0.05, 0.00)** | |

**Sanitation − Unimproved**

| **Country** | **Trial** | **Tool** | **N** | **N** | **PD (95% CI)** | **W** |
|---|---|---|---|---|---|---|
| Burkina Faso | iLiNS−Zinc (24) | | | | | |
| Kenya | WASH−B (28) | | | | | |
| Madagascar | MAHAY (29) | | | | | |
| Malawi | iLiNS−DYADM (30) | A not B task | 182 | 369 | −0.02 (−0.07, 0.03) | 0.26 |
| Malawi | iLiNS−DOSE (31) | A not B task | 475 | 164 | 0.04 (−0.01, 0.09) | 0.30 |
| Zimbabwe | SHINE (HIV−) (33) | A not B task | 370 | 357 | −0.02 (−0.06, 0.02) | 0.37 |
| Zimbabwe | SHINE (HIV+) (34) | A not B task | 63 | 57 | 0.01 (−0.10, 0.11) | 0.07 |
| **I² = 0.15, Tau² = 0.00** | | | **1090** | **947** | **0.00 (−0.03, 0.03)** | |

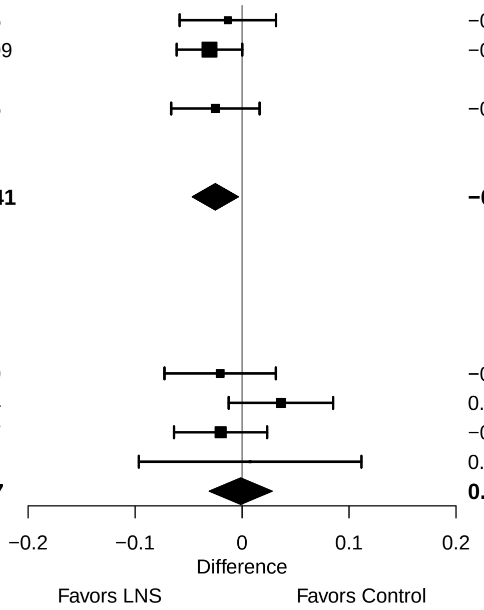

**Supplemental figure 4N: Executive function lowest decile prevalence difference**

#### 4N7: Stratified by Supplement duration (insufficient comparisons)

#### Supplemental figure 4N: Executive function lowest decile prevalence difference

### 4N8: Stratified by Frequency of contact

**Frequency of contact**
**(p−diff = 0.609)**

| Country | Trial | Tool | N | N | PD (95% CI) | W |
|---|---|---|---|---|---|---|
| **Frequency of contact − Monthly** | | | | | | |
| Bangladesh | RDNS (22) | A not B task | 533 | 266 | −0.01 (−0.06, 0.03) | 0.31 |
| Haiti | HAITI (27) | | | | | |
| Kenya | WASH−B (28) | | | | | |
| Madagascar | MAHAY (29) | | | | | |
| Mali | PROMIS CS (32) | | | | | |
| Zimbabwe | SHINE¹ (HIV−) (33), LNS vs SOC | A not B task | 370 | 357 | −0.02 (−0.06, 0.02) | 0.53 |
| Zimbabwe | LNS+WSH vs WSH | A not B task | 417 | 382 | −0.03 (−0.07, 0.02) | 0.53 |
| Zimbabwe | SHINE¹ (HIV+) (34), LNS vs SOC | A not B task | 63 | 57 | 0.01 (−0.10, 0.11) | 0.16 |
| Zimbabwe | LNS+WSH vs WSH | A not B task | 94 | 76 | 0.07 (−0.02, 0.16) | 0.16 |
| **I² = 0.40, Tau² = 0.00** | | | **1477** | **1138** | **−0.01 (−0.04, 0.03)** | |
| **Frequency of contact − Weekly** | | | | | | |
| Bangladesh | JiVitA−4 (21) | | | | | |
| Bangladesh | WASH−B (23) | A not B task | 1097 | 3327 | 0.00 (−0.02, 0.02) | 0.47 |
| Burkina Faso | iLiNS−Zinc (24) | | | | | |
| Ghana | GHANA (25) | | | | | |
| Ghana | iLiNS−DYADG (26) | A not B task | 290 | 576 | −0.02 (−0.07, 0.02) | 0.21 |
| Malawi | iLiNS−DYADM (30) | A not B task | 182 | 369 | −0.02 (−0.07, 0.03) | 0.15 |
| Malawi | iLiNS−DOSE (31) | A not B task | 475 | 164 | 0.04 (−0.01, 0.09) | 0.17 |
| **I² = 0.25, Tau² = 0.00** | | | **2044** | **4436** | **0.00 (−0.03, 0.02)** | |

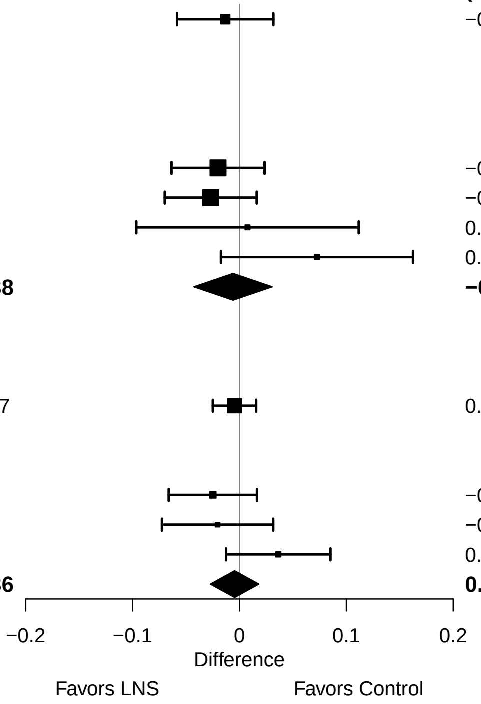

**Supplemental figure 4N: Executive function lowest decile prevalence difference**

#### 4N9: Stratified by Average SQ-LNS compliance (insufficient comparisons)

### Supplemental figure 4O: 12-mo walking without support prevalence ratio

#### 4O1: Stratified by Geographic region

**Geographic region**
**(p−diff = 0.525)**

| Country | Trial | N | N | PR (95% CI) | W |
|---|---|---|---|---|---|
| **Geographic region − SEAR** | | | | | |
| Bangladesh | RDNS (22) | 1628 | 790 | 1.28 (1.07, 1.53) | 0.28 |
| Bangladesh | WASH−B (23) | 497 | 1474 | 1.20 (1.05, 1.38) | 0.32 |
| Bangladesh | JiVitA−4 (21) | 3014 | 1363 | 1.05 (0.99, 1.12) | 0.39 |
| **I² = 0.69, Tau² = 0.01** | | **5139** | **3627** | **1.15 (1.02, 1.29)** | |
| **Geographic region − AFR** | | | | | |
| Burkina Faso | iLiNS−Zinc (24) | | | | |
| Ghana | GHANA (25) | 90 | 87 | 1.67 (1.08, 2.58) | 0.08 |
| Ghana | iLiNS−DYADG (26) | 327 | 663 | 1.15 (1.01, 1.31) | 0.23 |
| Kenya | WASH−B (28) | 447 | 1604 | 0.97 (0.85, 1.10) | 0.23 |
| Madagascar | MAHAY (29) | | | | |
| Malawi | iLiNS−DYADM (30) | 208 | 426 | 1.20 (1.03, 1.40) | 0.22 |
| Malawi | iLiNS−DOSE (31) | 602 | 200 | 1.04 (0.84, 1.30) | 0.17 |
| Mali | PROMIS CS (32) | 118 | 151 | 0.70 (0.44, 1.11) | 0.07 |
| Zimbabwe | SHINE (HIV−) (33) | | | | |
| Zimbabwe | SHINE (HIV+) (34) | | | | |
| **I² = 0.62, Tau² = 0.04** | | **1792** | **3131** | **1.09 (0.91, 1.30)** | |

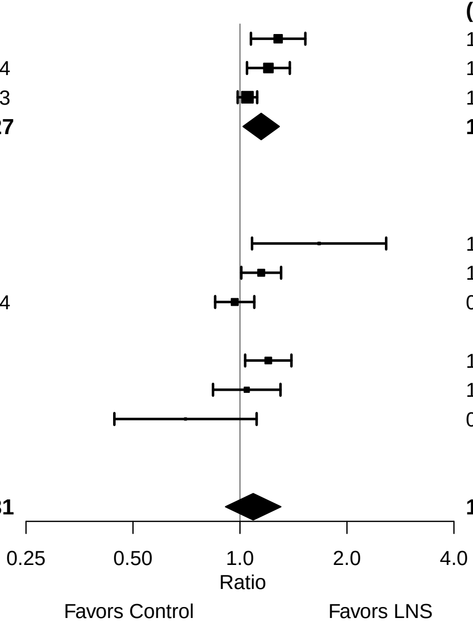

#### Supplemental figure 4O: 12-mo walking without support prevalence ratio

#### 4O2: Stratified by Stunting burden

**Stunting burden**
**(p−diff = 0.711)**

| Country | Trial | N | N | PR (95% CI) | W |
|---|---|---|---|---|---|
| **Stunting burden − Less than 35%** | | | | | |
| Ghana | GHANA (25) | 90 | 87 | 1.67 (1.08, 2.58) | 0.08 |
| Ghana | iLiNS−DYADG (26) | 327 | 663 | 1.15 (1.01, 1.31) | 0.28 |
| Haiti | HAITI (27) | 83 | 79 | 1.25 (0.86, 1.81) | 0.10 |
| Kenya | WASH−B (28) | 447 | 1604 | 0.97 (0.85, 1.10) | 0.28 |
| Malawi | iLiNS−DYADM (30) | 208 | 426 | 1.20 (1.03, 1.40) | 0.26 |
| **I² = 0.59, Tau² = 0.01** | | **1155** | **2859** | **1.15 (1.00, 1.33)** | |
| **Stunting burden − More than 35%** | | | | | |
| Bangladesh | RDNS (22) | 1628 | 790 | 1.28 (1.07, 1.53) | 0.21 |
| Bangladesh | WASH−B (23) | 497 | 1474 | 1.20 (1.05, 1.38) | 0.24 |
| Burkina Faso | iLiNS−Zinc (24) | | | | |
| Madagascar | MAHAY (29) | | | | |
| Malawi | iLiNS−DOSE (31) | 602 | 200 | 1.04 (0.84, 1.30) | 0.17 |
| Mali | PROMIS CS (32) | 118 | 151 | 0.70 (0.44, 1.11) | 0.07 |
| Zimbabwe | SHINE (HIV−) (33) | | | | |
| Zimbabwe | SHINE (HIV+) (34) | | | | |
| Bangladesh | JiVitA−4 (21) | 3014 | 1363 | 1.05 (0.99, 1.12) | 0.31 |
| **I² = 0.60, Tau² = 0.02** | | **5859** | **3978** | **1.09 (0.94, 1.27)** | |

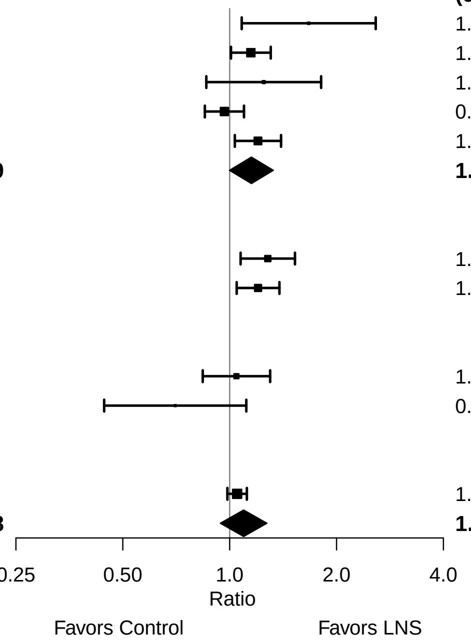

#### Supplemental figure 4O: 12-mo walking without support prevalence ratio

### 4O3: Stratified by Malaria prevalence

**Malaria prevalence**
**(p−diff = 0.768)**

| Country | Trial | N | N | PR (95% CI) | W |
|---|---|---|---|---|---|
| **Malaria prevalence − Less than 10%** | | | | | |
| Bangladesh | RDNS (22) | 1628 | 790 | 1.28 (1.07, 1.53) | 0.19 |
| Bangladesh | WASH−B (23) | 497 | 1474 | 1.20 (1.05, 1.38) | 0.22 |
| Haiti | HAITI (27) | 83 | 79 | 1.25 (0.86, 1.81) | 0.08 |
| Kenya | WASH−B (28) | 447 | 1604 | 0.97 (0.85, 1.10) | 0.23 |
| Madagascar | MAHAY (29) | | | | |
| Zimbabwe | SHINE (HIV−) (33) | | | | |
| Zimbabwe | SHINE (HIV+) (34) | | | | |
| Bangladesh | JiVitA−4 (21) | 3014 | 1363 | 1.05 (0.99, 1.12) | 0.28 |
| **I² = 0.61, Tau² = 0.01** | | **5669** | **5310** | **1.11 (1.00, 1.23)** | |
| **Malaria prevalence − At least 10%** | | | | | |
| Burkina Faso | iLiNS−Zinc (24) | | | | |
| Ghana | GHANA (25) | 90 | 87 | 1.67 (1.08, 2.58) | 0.09 |
| Ghana | iLiNS−DYADG (26) | 327 | 663 | 1.15 (1.01, 1.31) | 0.32 |
| Malawi | iLiNS−DYADM (30) | 208 | 426 | 1.20 (1.03, 1.40) | 0.29 |
| Malawi | iLiNS−DOSE (31) | 602 | 200 | 1.04 (0.84, 1.30) | 0.22 |
| Mali | PROMIS CS (32) | 118 | 151 | 0.70 (0.44, 1.11) | 0.08 |
| **I² = 0.52, Tau² = 0.05** | | **1345** | **1527** | **1.12 (0.90, 1.41)** | |

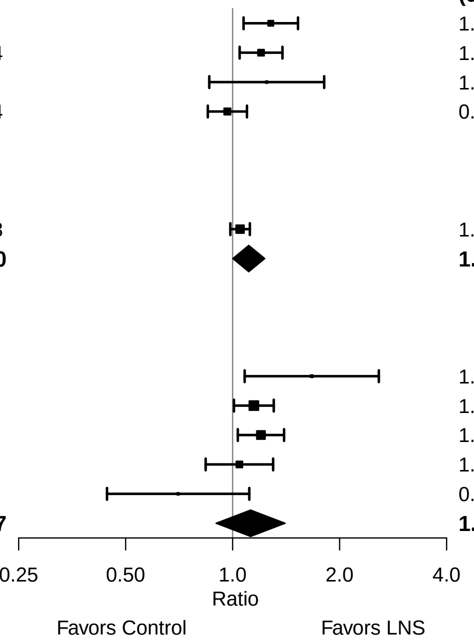

#### Supplemental figure 4O: 12-mo walking without support prevalence ratio

### 4O4: Stratified by Anemia prevalence

**Anemia prevalence**
**(p−diff = 0.720)**

| Country | Trial | N | N | PR (95% CI) | W |
|---|---|---|---|---|---|
| **Anemia prevalence − High** | | | | | |
| Burkina Faso | iLiNS−Zinc (24) | | | | |
| Ghana | GHANA (25) | 90 | 87 | 1.67 (1.08, 2.58) | 0.11 |
| Haiti | HAITI (27) | 83 | 79 | 1.25 (0.86, 1.81) | 0.14 |
| Malawi | iLiNS−DYADM (30) | 208 | 426 | 1.20 (1.03, 1.40) | 0.37 |
| Malawi | iLiNS−DOSE (31) | 602 | 200 | 1.04 (0.84, 1.30) | 0.27 |
| Mali | PROMIS CS (32) | 118 | 151 | 0.70 (0.44, 1.11) | 0.10 |
| **I² = 0.53, Tau² = 0.05** | | **1101** | **943** | **1.14 (0.89, 1.45)** | |
| **Anemia prevalence − Moderate** | | | | | |
| Bangladesh | RDNS (22) | 1628 | 790 | 1.28 (1.07, 1.53) | 0.16 |
| Bangladesh | WASH−B (23) | 497 | 1474 | 1.20 (1.05, 1.38) | 0.19 |
| Ghana | iLiNS−DYADG (26) | 327 | 663 | 1.15 (1.01, 1.31) | 0.20 |
| Kenya | WASH−B (28) | 447 | 1604 | 0.97 (0.85, 1.10) | 0.20 |
| Madagascar | MAHAY (29) | | | | |
| Zimbabwe | SHINE (HIV−) (33) | | | | |
| Zimbabwe | SHINE (HIV+) (34) | | | | |
| Bangladesh | JiVitA−4 (21) | 3014 | 1363 | 1.05 (0.99, 1.12) | 0.24 |
| **I² = 0.62, Tau² = 0.01** | | **5913** | **5894** | **1.11 (1.01, 1.22)** | |

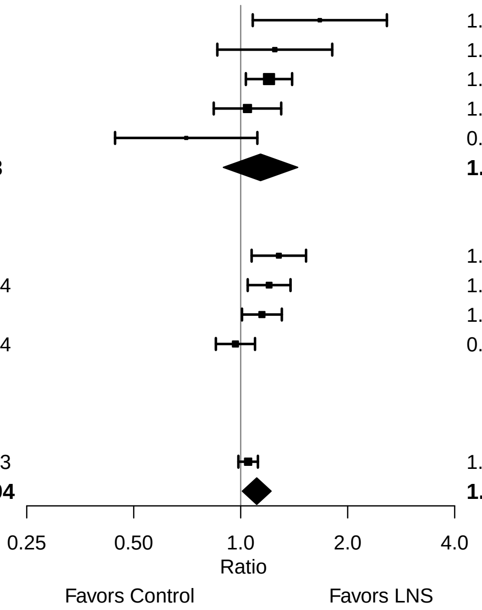

**Supplemental figure 4O: 12-mo walking without support prevalence ratio**

#### 4O5: Stratified by Source water quality (insufficient comparisons)

#### Supplemental figure 4O: 12-mo walking without support prevalence ratio

### 4O6: Stratified by Sanitation

**Sanitation**
**(p−diff = 0.631)**

| Country | Trial | N | N | PR (95% CI) | W |
|---|---|---|---|---|---|
| **Sanitation − Improved** | | | | | |
| Bangladesh | RDNS (22) | 1628 | 790 | 1.28 (1.07, 1.53) | 0.18 |
| Bangladesh | WASH−B (23) | 240 | 484 | 1.19 (0.94, 1.51) | 0.13 |
| Ghana | GHANA (25) | 90 | 87 | 1.67 (1.08, 2.58) | 0.05 |
| Ghana | iLiNS−DYADG (26) | 327 | 663 | 1.15 (1.01, 1.31) | 0.22 |
| Haiti | HAITI (27) | 83 | 79 | 1.25 (0.86, 1.81) | 0.07 |
| Mali | PROMIS CS (32) | 118 | 151 | 0.70 (0.44, 1.11) | 0.05 |
| Bangladesh | JiVitA−4 (21) | 3014 | 1363 | 1.05 (0.99, 1.12) | 0.29 |
| **I² = 0.55, Tau² = 0.02** | | **5500** | **3617** | **1.15 (1.00, 1.33)** | |
| **Sanitation − Unimproved** | | | | | |
| Burkina Faso | iLiNS−Zinc (24) | | | | |
| Kenya | WASH−B (28) | 202 | 706 | 1.00 (0.81, 1.22) | 0.31 |
| Madagascar | MAHAY (29) | | | | |
| Malawi | iLiNS−DYADM (30) | 208 | 426 | 1.20 (1.03, 1.40) | 0.40 |
| Malawi | iLiNS−DOSE (31) | 602 | 200 | 1.04 (0.84, 1.30) | 0.29 |
| Zimbabwe | SHINE (HIV−) (33) | | | | |
| Zimbabwe | SHINE (HIV+) (34) | | | | |
| **I² = 0.18, Tau² = 0.00** | | **1012** | **1332** | **1.10 (0.98, 1.24)** | |

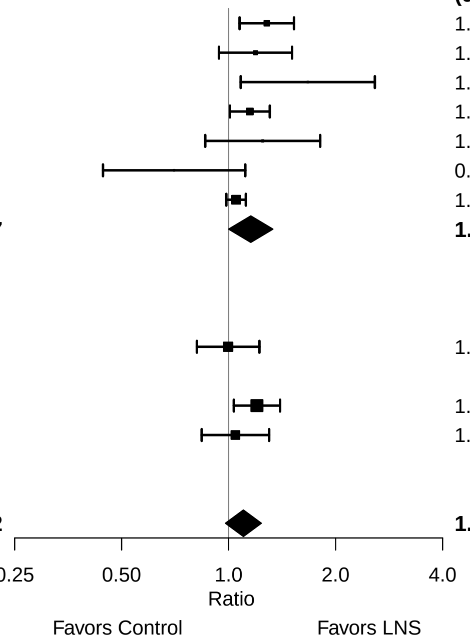

#### Supplemental figure 4O: 12-mo walking without support prevalence ratio

### 4O7: Stratified by Supplement duration

**Supplement duration**
**(p−diff = 0.618)**

| Country | Trial | N | N | PR (95% CI) | W |
|---|---|---|---|---|---|
| **Supplement duration − 12m or less** | | | | | |
| Burkina Faso | iLiNS−Zinc (24) | | | | |
| Ghana | GHANA (25) | 90 | 87 | 1.67 (1.08, 2.58) | 0.06 |
| Ghana | iLiNS−DYADG (26) | 327 | 663 | 1.15 (1.01, 1.31) | 0.22 |
| Haiti | HAITI (27) | 83 | 79 | 1.25 (0.86, 1.81) | 0.08 |
| Madagascar | MAHAY (29) | | | | |
| Malawi | iLiNS−DYADM (30) | 208 | 426 | 1.20 (1.03, 1.40) | 0.21 |
| Malawi | iLiNS−DOSE (31) | 602 | 200 | 1.04 (0.84, 1.30) | 0.15 |
| Zimbabwe | SHINE (HIV−) (33) | | | | |
| Zimbabwe | SHINE (HIV+) (34) | | | | |
| Bangladesh | JiVitA−4 (21) | 3014 | 1363 | 1.05 (0.99, 1.12) | 0.27 |
| **$I^2$ = 0.37, $Tau^2$ = 0.00** | | **4324** | **2818** | **1.13 (1.03, 1.23)** | |
| **Supplement duration − > 12m** | | | | | |
| Bangladesh | RDNS (22) | 1628 | 790 | 1.28 (1.07, 1.53) | 0.27 |
| Bangladesh | WASH−B (23) | 497 | 1474 | 1.20 (1.05, 1.38) | 0.32 |
| Kenya | WASH−B (28) | 447 | 1604 | 0.97 (0.85, 1.10) | 0.33 |
| Mali | PROMIS CS (32) | 118 | 151 | 0.70 (0.44, 1.11) | 0.09 |
| **$I^2$ = 0.75, $Tau^2$ = 0.04** | | **2690** | **4019** | **1.06 (0.85, 1.34)** | |

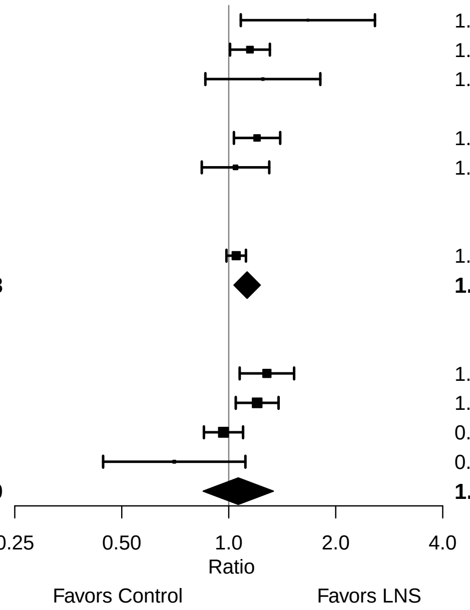

#### Supplemental figure 4O: 12-mo walking without support prevalence ratio

### 4O8: Stratified by Frequency of contact

**Frequency of contact**
**(p−diff = 0.433)**

| Country | Trial | N | N | PR (95% CI) | W |
|---|---|---|---|---|---|
| **Frequency of contact − Monthly** | | | | | |
| Bangladesh | RDNS (22) | 1628 | 790 | 1.28 (1.07, 1.53) | 0.34 |
| Haiti | HAITI (27) | 83 | 79 | 1.25 (0.86, 1.81) | 0.15 |
| Kenya | WASH−B (28) | 447 | 1604 | 0.97 (0.85, 1.10) | 0.41 |
| Madagascar | MAHAY (29) | | | | |
| Mali | PROMIS CS (32) | 118 | 151 | 0.70 (0.44, 1.11) | 0.11 |
| Zimbabwe | SHINE (HIV−) (33) | | | | |
| Zimbabwe | SHINE (HIV+) (34) | | | | |
| **I² = 0.71, Tau² = 0.04** | | **2276** | **2624** | **1.05 (0.83, 1.34)** | |
| **Frequency of contact − Weekly** | | | | | |
| Bangladesh | WASH−B (23) | 497 | 1474 | 1.20 (1.05, 1.38) | 0.19 |
| Burkina Faso | iLiNS−Zinc (24) | | | | |
| Ghana | GHANA (25) | 90 | 87 | 1.67 (1.08, 2.58) | 0.06 |
| Ghana | iLiNS−DYADG (26) | 327 | 663 | 1.15 (1.01, 1.31) | 0.20 |
| Malawi | iLiNS−DYADM (30) | 208 | 426 | 1.20 (1.03, 1.40) | 0.18 |
| Malawi | iLiNS−DOSE (31) | 602 | 200 | 1.04 (0.84, 1.30) | 0.14 |
| Bangladesh | JiVitA−4 (21) | 3014 | 1363 | 1.05 (0.99, 1.12) | 0.24 |
| **I² = 0.46, Tau² = 0.00** | | **4738** | **4213** | **1.14 (1.05, 1.23)** | |

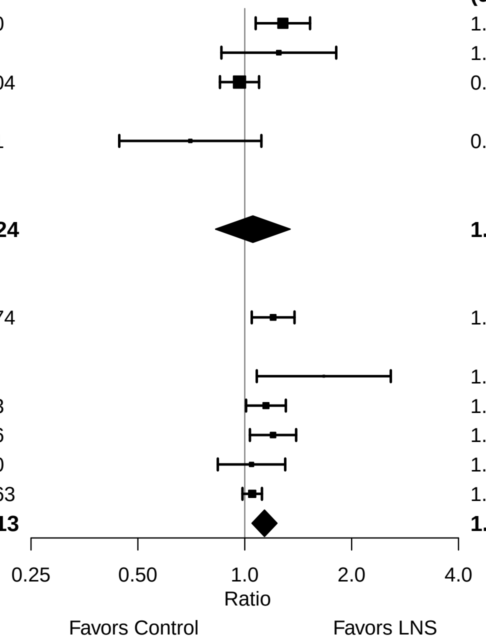

#### Supplemental figure 4O: 12-mo walking without support prevalence ratio

### 4O9: Stratified by Average SQ-LNS compliance

**Average SQ−LNS compliance**
**(p−diff = 0.700)**

| Country | Trial | N | N | PR (95% CI) | W |
|---|---|---|---|---|---|
| **Average SQ−LNS compliance − Low** | | | | | |
| Ghana | iLiNS−DYADG (26) | 327 | 663 | 1.15 (1.01, 1.31) | 0.35 |
| Malawi | iLiNS−DYADM (30) | 208 | 426 | 1.20 (1.03, 1.40) | 0.32 |
| Malawi | iLiNS−DOSE (31) | 602 | 200 | 1.04 (0.84, 1.30) | 0.24 |
| Mali | PROMIS CS (32) | 118 | 151 | 0.70 (0.44, 1.11) | 0.09 |
| Zimbabwe | SHINE (HIV−) (33) | | | | |
| Zimbabwe | SHINE (HIV+) (34) | | | | |
| **I² = 0.43, Tau² = 0.02** | | **1255** | **1440** | **1.08 (0.90, 1.29)** | |
| **Average SQ−LNS compliance − High** | | | | | |
| Bangladesh | RDNS (22) | 1628 | 790 | 1.28 (1.07, 1.53) | 0.18 |
| Bangladesh | WASH−B (23) | 497 | 1474 | 1.20 (1.05, 1.38) | 0.21 |
| Burkina Faso | iLiNS−Zinc (24) | | | | |
| Ghana | GHANA (25) | 90 | 87 | 1.67 (1.08, 2.58) | 0.06 |
| Haiti | HAITI (27) | 83 | 79 | 1.25 (0.86, 1.81) | 0.08 |
| Kenya | WASH−B (28) | 447 | 1604 | 0.97 (0.85, 1.10) | 0.22 |
| Bangladesh | JiVitA−4 (21) | 3014 | 1363 | 1.05 (0.99, 1.12) | 0.26 |
| **I² = 0.64, Tau² = 0.01** | | **5759** | **5397** | **1.15 (1.01, 1.30)** | |

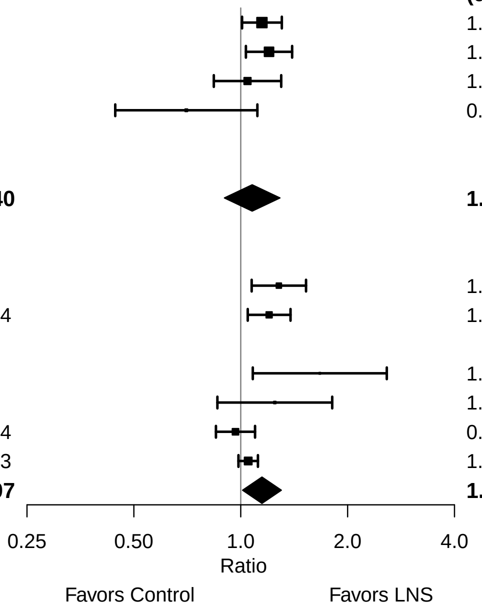

### Supplemental figure 4P: 12-mo walking without support prevalence difference

#### 4P1: Stratified by Geographic region

**Geographic region**
**(p−diff = 0.639)**

| Country | Trial | N | N | PD (95% CI) | W |
|---|---|---|---|---|---|
| **Geographic region − SEAR** | | | | | |
| Bangladesh | RDNS (22) | 1628 | 790 | 0.07 (0.03, 0.11) | 0.34 |
| Bangladesh | WASH−B (23) | 497 | 1474 | 0.05 (0.01, 0.10) | 0.32 |
| Bangladesh | JiVitA−4 (21) | 3014 | 1363 | 0.03 (−0.01, 0.06) | 0.35 |
| **I² = 0.38, Tau² = 0.00** | | **5139** | **3627** | **0.05 (0.02, 0.08)** | |
| **Geographic region − AFR** | | | | | |
| Burkina Faso | iLiNS−Zinc (24) | | | | |
| Ghana | GHANA (25) | 90 | 87 | 0.17 (0.03, 0.31) | 0.10 |
| Ghana | iLiNS−DYADG (26) | 327 | 663 | 0.07 (0.00, 0.14) | 0.20 |
| Kenya | WASH−B (28) | 447 | 1604 | −0.01 (−0.06, 0.04) | 0.22 |
| Madagascar | MAHAY (29) | | | | |
| Malawi | iLiNS−DYADM (30) | 208 | 426 | 0.10 (0.02, 0.18) | 0.17 |
| Malawi | iLiNS−DOSE (31) | 602 | 200 | 0.02 (−0.06, 0.09) | 0.18 |
| Mali | PROMIS CS (32) | 118 | 151 | −0.10 (−0.21, 0.01) | 0.13 |
| Zimbabwe | SHINE (HIV−) (33) | | | | |
| Zimbabwe | SHINE (HIV+) (34) | | | | |
| **I² = 0.69, Tau² = 0.01** | | **1792** | **3131** | **0.04 (−0.03, 0.10)** | |

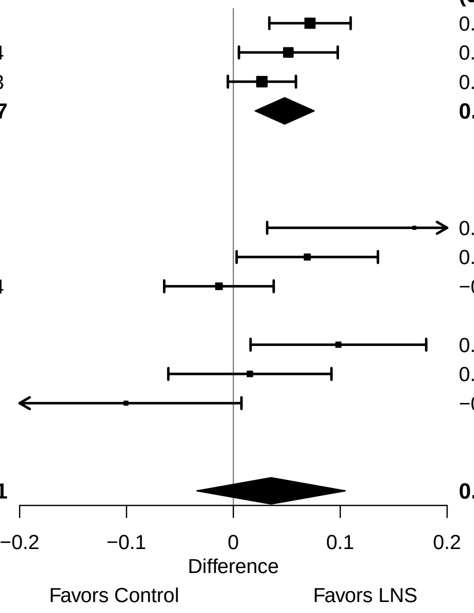

**Supplemental figure 4P: 12-mo walking without support prevalence difference**

#### 4P2: Stratified by Stunting burden

**Stunting burden**
**(p−diff = 0.316)**

| Country | Trial | N | N | PD (95% CI) | W |
|---|---|---|---|---|---|
| **Stunting burden − Less than 35%** | | | | | |
| Ghana | GHANA (25) | 90 | 87 | 0.17 (0.03, 0.31) | 0.12 |
| Ghana | iLiNS−DYADG (26) | 327 | 663 | 0.07 (0.00, 0.14) | 0.26 |
| Haiti | HAITI (27) | 83 | 79 | 0.09 (−0.06, 0.24) | 0.11 |
| Kenya | WASH−B (28) | 447 | 1604 | −0.01 (−0.06, 0.04) | 0.30 |
| Malawi | iLiNS−DYADM (30) | 208 | 426 | 0.10 (0.02, 0.18) | 0.22 |
| **I² = 0.62, Tau² = 0.00** | | **1155** | **2859** | **0.07 (0.01, 0.12)** | |
| **Stunting burden − More than 35%** | | | | | |
| Bangladesh | RDNS (22) | 1628 | 790 | 0.07 (0.03, 0.11) | 0.24 |
| Bangladesh | WASH−B (23) | 497 | 1474 | 0.05 (0.01, 0.10) | 0.22 |
| Burkina Faso | iLiNS−Zinc (24) | | | | |
| Madagascar | MAHAY (29) | | | | |
| Malawi | iLiNS−DOSE (31) | 602 | 200 | 0.02 (−0.06, 0.09) | 0.17 |
| Mali | PROMIS CS (32) | 118 | 151 | −0.10 (−0.21, 0.01) | 0.12 |
| Zimbabwe | SHINE (HIV−) (33) | | | | |
| Zimbabwe | SHINE (HIV+) (34) | | | | |
| Bangladesh | JiVitA−4 (21) | 3014 | 1363 | 0.03 (−0.01, 0.06) | 0.25 |
| **I² = 0.62, Tau² = 0.00** | | **5859** | **3978** | **0.03 (−0.02, 0.08)** | |

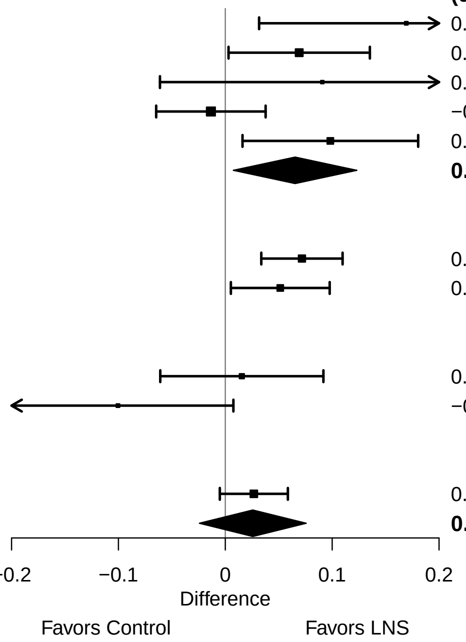

#### Supplemental figure 4P: 12-mo walking without support prevalence difference

#### 4P3: Stratified by Malaria prevalence

**Malaria prevalence**
**(p−diff = 0.814)**

| Country | Trial | N | N | PD (95% CI) | W |
|---|---|---|---|---|---|
| **Malaria prevalence − Less than 10%** | | | | | |
| Bangladesh | RDNS (22) | 1628 | 790 | 0.07 (0.03, 0.11) | 0.24 |
| Bangladesh | WASH−B (23) | 497 | 1474 | 0.05 (0.01, 0.10) | 0.22 |
| Haiti | HAITI (27) | 83 | 79 | 0.09 (−0.06, 0.24) | 0.08 |
| Kenya | WASH−B (28) | 447 | 1604 | −0.01 (−0.06, 0.04) | 0.21 |
| Madagascar | MAHAY (29) | | | | |
| Zimbabwe | SHINE (HIV−) (33) | | | | |
| Zimbabwe | SHINE (HIV+) (34) | | | | |
| Bangladesh | JiVitA−4 (21) | 3014 | 1363 | 0.03 (−0.01, 0.06) | 0.25 |
| **I² = 0.51, Tau² = 0.00** | | **5669** | **5310** | **0.04 (0.01, 0.07)** | |
| **Malaria prevalence − At least 10%** | | | | | |
| Burkina Faso | iLiNS−Zinc (24) | | | | |
| Ghana | GHANA (25) | 90 | 87 | 0.17 (0.03, 0.31) | 0.12 |
| Ghana | iLiNS−DYADG (26) | 327 | 663 | 0.07 (0.00, 0.14) | 0.26 |
| Malawi | iLiNS−DYADM (30) | 208 | 426 | 0.10 (0.02, 0.18) | 0.22 |
| Malawi | iLiNS−DOSE (31) | 602 | 200 | 0.02 (−0.06, 0.09) | 0.23 |
| Mali | PROMIS CS (32) | 118 | 151 | −0.10 (−0.21, 0.01) | 0.17 |
| **I² = 0.69, Tau² = 0.01** | | **1345** | **1527** | **0.05 (−0.04, 0.13)** | |

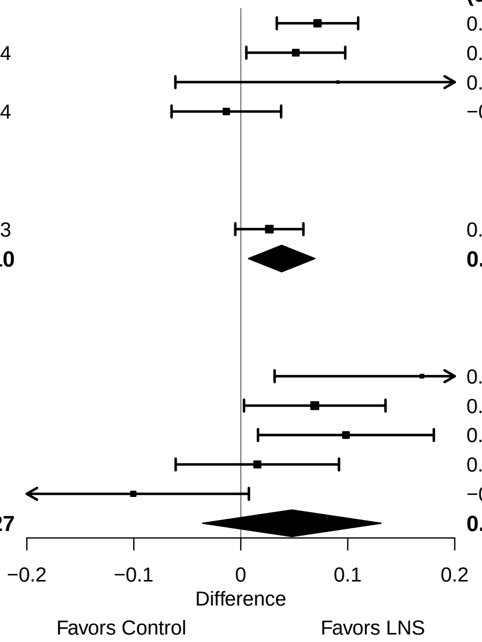

#### Supplemental figure 4P: 12-mo walking without support prevalence difference

### 4P4: Stratified by Anemia prevalence

**Anemia prevalence**
**(p−diff = 0.903)**

| Country | Trial | N | N | PD (95% CI) | W |
|---|---|---|---|---|---|
| **Anemia prevalence − High** | | | | | |
| Burkina Faso | iLiNS−Zinc (24) | | | | |
| Ghana | GHANA (25) | 90 | 87 | 0.17 (0.03, 0.31) | 0.14 |
| Haiti | HAITI (27) | 83 | 79 | 0.09 (−0.06, 0.24) | 0.13 |
| Malawi | iLiNS−DYADM (30) | 208 | 426 | 0.10 (0.02, 0.18) | 0.26 |
| Malawi | iLiNS−DOSE (31) | 602 | 200 | 0.02 (−0.06, 0.09) | 0.27 |
| Mali | PROMIS CS (32) | 118 | 151 | −0.10 (−0.21, 0.01) | 0.20 |
| **I² = 0.68, Tau² = 0.01** | | **1101** | **943** | **0.05 (−0.04, 0.14)** | |
| **Anemia prevalence − Moderate** | | | | | |
| Bangladesh | RDNS (22) | 1628 | 790 | 0.07 (0.03, 0.11) | 0.21 |
| Bangladesh | WASH−B (23) | 497 | 1474 | 0.05 (0.01, 0.10) | 0.20 |
| Ghana | iLiNS−DYADG (26) | 327 | 663 | 0.07 (0.00, 0.14) | 0.17 |
| Kenya | WASH−B (28) | 447 | 1604 | −0.01 (−0.06, 0.04) | 0.19 |
| Madagascar | MAHAY (29) | | | | |
| Zimbabwe | SHINE (HIV−) (33) | | | | |
| Zimbabwe | SHINE (HIV+) (34) | | | | |
| Bangladesh | JiVitA−4 (21) | 3014 | 1363 | 0.03 (−0.01, 0.06) | 0.22 |
| **I² = 0.53, Tau² = 0.00** | | **5913** | **5894** | **0.04 (0.01, 0.07)** | |

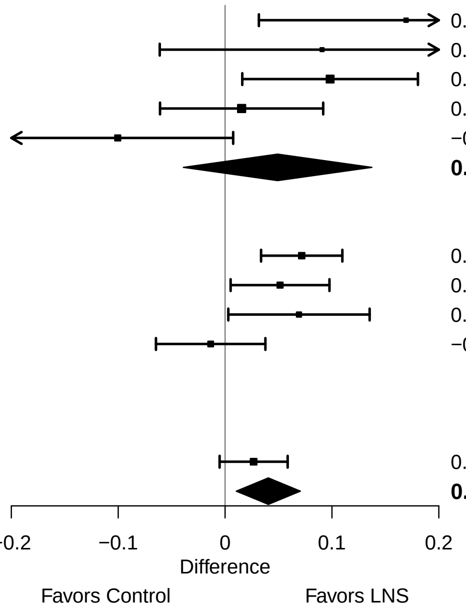

**Supplemental figure 4P: 12-mo walking without support prevalence difference**

#### 4P5: Stratified by Source water quality (insufficient comparisons)

#### Supplemental figure 4P: 12-mo walking without support prevalence difference

### 4P6: Stratified by Sanitation

**Sanitation**
**(p−diff = 0.733)**

| Country | Trial | N | N | PD (95% CI) | W |
|---|---|---|---|---|---|
| **Sanitation − Improved** | | | | | |
| Bangladesh | RDNS (22) | 1628 | 790 | 0.07 (0.03, 0.11) | 0.22 |
| Bangladesh | WASH−B (23) | 240 | 484 | 0.05 (−0.02, 0.12) | 0.16 |
| Ghana | GHANA (25) | 90 | 87 | 0.17 (0.03, 0.31) | 0.07 |
| Ghana | iLiNS−DYADG (26) | 327 | 663 | 0.07 (0.00, 0.14) | 0.16 |
| Haiti | HAITI (27) | 83 | 79 | 0.09 (−0.06, 0.24) | 0.06 |
| Mali | PROMIS CS (32) | 118 | 151 | −0.10 (−0.21, 0.01) | 0.10 |
| Bangladesh | JiVitA−4 (21) | 3014 | 1363 | 0.03 (−0.01, 0.06) | 0.23 |
| **I² = 0.57, Tau² = 0.00** | | **5500** | **3617** | **0.05 (0.00, 0.10)** | |
| **Sanitation − Unimproved** | | | | | |
| Burkina Faso | iLiNS−Zinc (24) | | | | |
| Kenya | WASH−B (28) | 202 | 706 | 0.00 (−0.08, 0.08) | 0.34 |
| Madagascar | MAHAY (29) | | | | |
| Malawi | iLiNS−DYADM (30) | 208 | 426 | 0.10 (0.02, 0.18) | 0.32 |
| Malawi | iLiNS−DOSE (31) | 602 | 200 | 0.02 (−0.06, 0.09) | 0.34 |
| Zimbabwe | SHINE (HIV−) (33) | | | | |
| Zimbabwe | SHINE (HIV+) (34) | | | | |
| **I² = 0.41, Tau² = 0.00** | | **1012** | **1332** | **0.04 (−0.02, 0.10)** | |

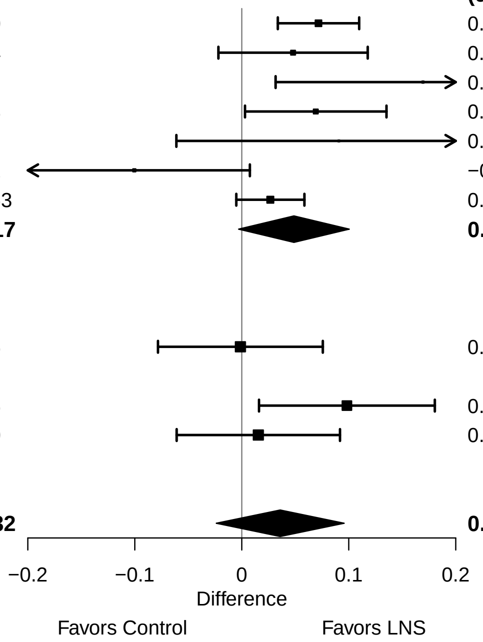

##### Supplemental figure 4P: 12-mo walking without support prevalence difference

#### 4P7: Stratified by Supplement duration

**Supplement duration**
**(p−diff = 0.221)**

**Supplement duration − 12m or less**

| Country | Trial | N | N | PD (95% CI) | W |
|---|---|---|---|---|---|
| Burkina Faso | iLiNS−Zinc (24) | | | | |
| Ghana | GHANA (25) | 90 | 87 | 0.17 (0.03, 0.31) | 0.10 |
| Ghana | iLiNS−DYADG (26) | 327 | 663 | 0.07 (0.00, 0.14) | 0.20 |
| Haiti | HAITI (27) | 83 | 79 | 0.09 (−0.06, 0.24) | 0.08 |
| Madagascar | MAHAY (29) | | | | |
| Malawi | iLiNS−DYADM (30) | 208 | 426 | 0.10 (0.02, 0.18) | 0.17 |
| Malawi | iLiNS−DOSE (31) | 602 | 200 | 0.02 (−0.06, 0.09) | 0.18 |
| Zimbabwe | SHINE (HIV−) (33) | | | | |
| Zimbabwe | SHINE (HIV+) (34) | | | | |
| Bangladesh | JiVitA−4 (21) | 3014 | 1363 | 0.03 (−0.01, 0.06) | 0.27 |
| **I² = 0.33, Tau² = 0.00** | | **4324** | **2818** | **0.06 (0.02, 0.09)** | |

**Supplement duration − > 12m**

| Country | Trial | N | N | PD (95% CI) | W |
|---|---|---|---|---|---|
| Bangladesh | RDNS (22) | 1628 | 790 | 0.07 (0.03, 0.11) | 0.30 |
| Bangladesh | WASH−B (23) | 497 | 1474 | 0.05 (0.01, 0.10) | 0.28 |
| Kenya | WASH−B (28) | 447 | 1604 | −0.01 (−0.06, 0.04) | 0.27 |
| Mali | PROMIS CS (32) | 118 | 151 | −0.10 (−0.21, 0.01) | 0.15 |
| **I² = 0.78, Tau² = 0.00** | | **2690** | **4019** | **0.01 (−0.06, 0.08)** | |

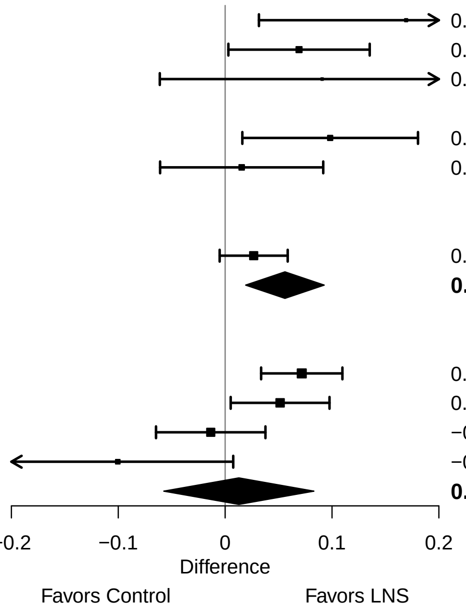

#### Supplemental figure 4P: 12-mo walking without support prevalence difference

### 4P8: Stratified by Frequency of contact

**Frequency of contact**
**(p−diff = 0.235)**

| Country | Trial | N | N | PD (95% CI) | W |
|---|---|---|---|---|---|
| **Frequency of contact − Monthly** | | | | | |
| Bangladesh | RDNS (22) | 1628 | 790 | 0.07 (0.03, 0.11) | 0.37 |
| Haiti | HAITI (27) | 83 | 79 | 0.09 (−0.06, 0.24) | 0.12 |
| Kenya | WASH−B (28) | 447 | 1604 | −0.01 (−0.06, 0.04) | 0.33 |
| Madagascar | MAHAY (29) | | | | |
| Mali | PROMIS CS (32) | 118 | 151 | −0.10 (−0.21, 0.01) | 0.18 |
| Zimbabwe | SHINE (HIV−) (33) | | | | |
| Zimbabwe | SHINE (HIV+) (34) | | | | |
| **I² = 0.78, Tau² = 0.00** | | **2276** | **2624** | **0.01 (−0.07, 0.09)** | |
| **Frequency of contact − Weekly** | | | | | |
| Bangladesh | WASH−B (23) | 497 | 1474 | 0.05 (0.01, 0.10) | 0.21 |
| Burkina Faso | iLiNS−Zinc (24) | | | | |
| Ghana | GHANA (25) | 90 | 87 | 0.17 (0.03, 0.31) | 0.08 |
| Ghana | iLiNS−DYADG (26) | 327 | 663 | 0.07 (0.00, 0.14) | 0.17 |
| Malawi | iLiNS−DYADM (30) | 208 | 426 | 0.10 (0.02, 0.18) | 0.15 |
| Malawi | iLiNS−DOSE (31) | 602 | 200 | 0.02 (−0.06, 0.09) | 0.16 |
| Bangladesh | JiVitA−4 (21) | 3014 | 1363 | 0.03 (−0.01, 0.06) | 0.23 |
| **I² = 0.30, Tau² = 0.00** | | **4738** | **4213** | **0.05 (0.02, 0.09)** | |

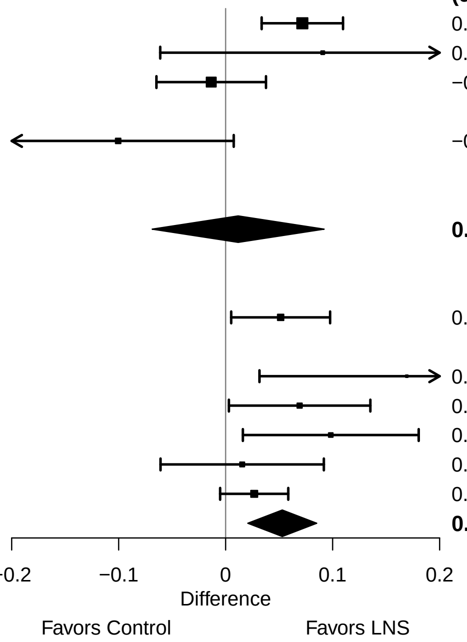

**Supplemental figure 4P: 12-mo walking without support prevalence difference**

#### 4P9: Stratified by Average SQ-LNS compliance

**Average SQ−LNS compliance**
**(p−diff = 0.639)**

| Country | Trial | N | N | PD (95% CI) | W |
|---|---|---|---|---|---|
| **Average SQ−LNS compliance − Low** | | | | | |
| Ghana | iLiNS−DYADG (26) | 327 | 663 | 0.07 (0.00, 0.14) | 0.29 |
| Malawi | iLiNS−DYADM (30) | 208 | 426 | 0.10 (0.02, 0.18) | 0.25 |
| Malawi | iLiNS−DOSE (31) | 602 | 200 | 0.02 (−0.06, 0.09) | 0.27 |
| Mali | PROMIS CS (32) | 118 | 151 | −0.10 (−0.21, 0.01) | 0.19 |
| Zimbabwe | SHINE (HIV−) (33) | | | | |
| Zimbabwe | SHINE (HIV+) (34) | | | | |
| **I² = 0.69, Tau² = 0.01** | | **1255** | **1440** | **0.03 (−0.06, 0.11)** | |
| **Average SQ−LNS compliance − High** | | | | | |
| Bangladesh | RDNS (22) | 1628 | 790 | 0.07 (0.03, 0.11) | 0.22 |
| Bangladesh | WASH−B (23) | 497 | 1474 | 0.05 (0.01, 0.10) | 0.20 |
| Burkina Faso | iLiNS−Zinc (24) | | | | |
| Ghana | GHANA (25) | 90 | 87 | 0.17 (0.03, 0.31) | 0.08 |
| Haiti | HAITI (27) | 83 | 79 | 0.09 (−0.06, 0.24) | 0.07 |
| Kenya | WASH−B (28) | 447 | 1604 | −0.01 (−0.06, 0.04) | 0.20 |
| Bangladesh | JiVitA−4 (21) | 3014 | 1363 | 0.03 (−0.01, 0.06) | 0.23 |
| **I² = 0.57, Tau² = 0.00** | | **5759** | **5397** | **0.05 (0.01, 0.09)** | |

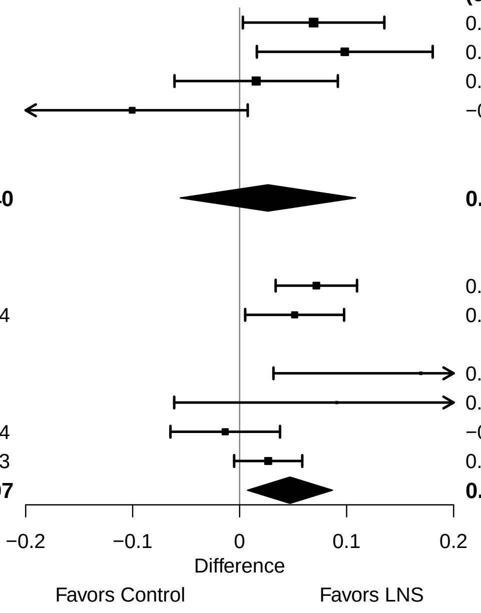

### Supplemental figure 5: Forest plots for effects of SQ-LNS on developmental outcomes stratified by individual-level maternal and child effect modifiers

#### Contents

**Supplemental figure 5A: Mean difference in language z-score** 5
5A1: Stratified by Maternal height 5
5A2: Stratified by Maternal BMI 6
5A3: Stratified by Maternal age 7
5A4: Stratified by Maternal education 8
5A5: Stratified by Maternal depressive symptoms 9
5A6: Stratified by Child sex 10
5A7: Stratified by Child birth order 11
5A8: Stratified by Child baseline stunting 12
5A9: Stratified by Child baseline acute malnutrition 13
5A10: Stratified by Child baseline anemia 14

**Supplemental figure 5B: Language lowest decile prevalence ratio** 15
5B1: Stratified by Maternal height 15
5B2: Stratified by Maternal BMI 16
5B3: Stratified by Maternal age 17
5B4: Stratified by Maternal education 18
5B5: Stratified by Maternal depressive symptoms 19
5B6: Stratified by Child sex 20
5B7: Stratified by Child birth order 21
5B8: Stratified by Child baseline stunting 22
5B9: Stratified by Child baseline acute malnutrition 23
5B10: Stratified by Child baseline anemia 24

**Supplemental figure 5C: Language lowest decile prevalence difference** 25
5C1: Stratified by Maternal height 25
5C2: Stratified by Maternal BMI 26
5C3: Stratified by Maternal age 27
5C4: Stratified by Maternal education 28
5C5: Stratified by Maternal depressive symptoms 29
5C6: Stratified by Child sex 30
5C7: Stratified by Child birth order 31
5C8: Stratified by Child baseline stunting 32
5C9: Stratified by Child baseline acute malnutrition 33
5C10: Stratified by Child baseline anemia 34

**Supplemental figure 5D: Mean difference in social-emotional z-score** 35
5D1: Stratified by Maternal height 35
5D2: Stratified by Maternal BMI 36
5D3: Stratified by Maternal age 37
5D4: Stratified by Maternal education 38

These figures are forest plots showing the individual-level effect modification of intervention effects. Each figure has the estimates of intervention effect stratified within study by individual-level effect modifier category. For dichotomous outcomes analyzed via prevalence ratios, the effect estimate is the prevalence in the LNS group divided by the prevalence in the control group. For dichotomous outcomes analyzed via prevalence differences, the effect estimate is the prevalence in the LNS group minus the prevalence in the control group. The labels on the far left correspond to trial level information. In the middle left and on the right the values indicate the study level effect estimate, confidence interval, and weighting for deriving the pooled estimates is shown by subgroup.

Motor milestone figures show individual trial estimates excluding the JiVitA-4 data because those individual trial results have not yet been published. The pooled estimate in each figure includes the JiVitA-4 data for consistency with all other analyses.

### Supplemental figure 5A: Mean difference in language z-score

#### 5A1: Stratified by Maternal height

**P−for−interaction = 0.857**
**Difference in MDs = −0.01 (−0.09, 0.07)**

| | | | At least 150.1 cm | | | | | | Less than 150.1 cm | | | | | |
|---|---|---|---|---|---|---|---|---|---|---|---|---|---|---|
| Country | Trial | Tool | LNS N | Control N | Control Mean | MD (95% CI) | Fixed W | Random W | LNS N | Control N | Control Mean | MD (95% CI) | Fixed W | Random W |
| Bangladesh | JiVitA−4 (21) | BSID−III | | | | | | | | | | | | |
| Bangladesh | RDNS (22) | CDI | 868 | 431 | −0.10 | 0.25 (0.10, 0.39) | 0.07 | 0.09 | 735 | 358 | −0.13 | 0.07 (−0.06, 0.20) | 0.27 | 0.26 |
| Bangladesh | WASH−B (23) | CDI | 589 | 1812 | 0.02 | 0.08 (−0.01, 0.17) | 0.18 | 0.12 | 515 | 1499 | −0.06 | 0.11 (0.01, 0.21) | 0.46 | 0.33 |
| Burkina Faso | iLiNS−Zinc (24) | DMC | 727 | 365 | −0.25 | 0.38 (0.22, 0.55) | 0.05 | 0.08 | 17 | 10 | 0.06 | −0.14 (−0.97, 0.69) | 0.01 | 0.01 |
| Ghana | GHANA (25) | | | | | | | | | | | | | |
| Ghana | iLiNS−DYADG (26) | CDI | 310 | 612 | 0.01 | 0.00 (−0.13, 0.14) | 0.07 | 0.10 | 16 | 35 | −0.23 | 0.23 (−0.33, 0.79) | 0.02 | 0.03 |
| Haiti | HAITI (27) | Total sounds | | | | | | | | | | | | |
| Kenya | WASH−B (28) | EASQ | 1237 | 4372 | 0.01 | 0.01 (−0.06, 0.08) | 0.32 | 0.13 | 51 | 175 | −0.21 | 0.34 (0.03, 0.65) | 0.05 | 0.08 |
| Madagascar | MAHAY (29) | ASQI | 956 | 986 | 0.06 | −0.07 (−0.27, 0.13) | 0.04 | 0.07 | 553 | 538 | 0.08 | −0.18 (−0.40, 0.04) | 0.10 | 0.14 |
| Malawi | iLiNS−DYADM (30) | CDI | 187 | 378 | 0.02 | 0.01 (−0.17, 0.18) | 0.05 | 0.08 | 28 | 58 | −0.15 | 0.07 (−0.38, 0.51) | 0.02 | 0.04 |
| Malawi | iLiNS−DOSE (31) | CDI | 542 | 185 | −0.05 | 0.06 (−0.11, 0.22) | 0.05 | 0.08 | 102 | 35 | −0.01 | 0.04 (−0.36, 0.44) | 0.03 | 0.05 |
| Mali | PROMIS CS (32) | DMC | 889 | 911 | −0.06 | 0.14 (−0.02, 0.31) | 0.05 | 0.08 | 32 | 27 | −0.27 | 0.39 (−0.15, 0.92) | 0.02 | 0.03 |
| Zimbabwe | SHINE (HIV−) (33) | CDI | 766 | 745 | −0.05 | 0.09 (−0.03, 0.21) | 0.10 | 0.10 | 32 | 21 | −0.12 | −0.08 (−0.67, 0.51) | 0.01 | 0.02 |
| Zimbabwe | SHINE (HIV+) (34) | CDI | 153 | 136 | −0.11 | 0.22 (−0.02, 0.45) | 0.03 | 0.06 | 10 | 6 | −0.39 | 0.69 (−0.31, 1.68) | 0.00 | 0.01 |
| | | | **7224** | **10933** | | **I² = 0.64, Tau² = 0.01** | | | **2091** | **2762** | | **I² = 0.18, Tau² = 0.00** | | |
| **Fixed** | | | | | | **0.08 (0.04, 0.11)** | | | | | | **0.08 (0.01, 0.15)** | | |
| **Random** | | | | | | **0.10 (0.03, 0.18)** | | | | | | **0.08 (−0.01, 0.18)** | | |

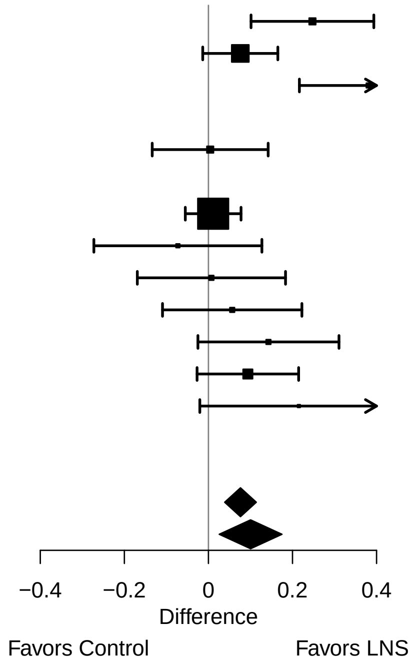

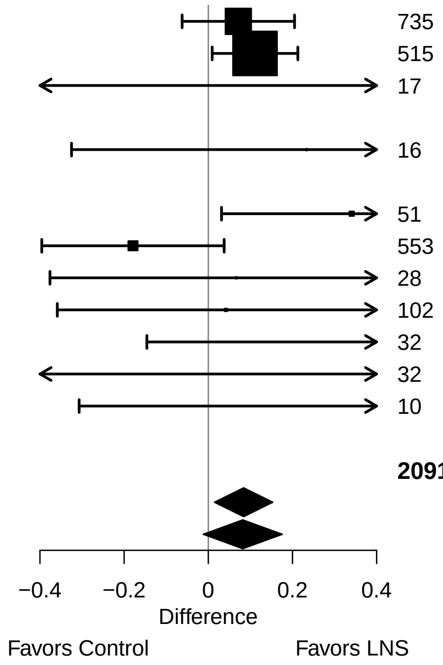

#### Supplemental figure 5A: Mean difference in language z-score

#### 5A2: Stratified by Maternal BMI

**P−for−interaction = 0.194**
**Difference in MDs = −0.04 (−0.11, 0.02)**

| Country | Trial | Tool | LNS N | Control N | Control Mean | At least 20 kg/m² MD (95% CI) | Fixed W | Random W | LNS N | Control N | Control Mean | Less than 20 kg/m² MD (95% CI) | Fixed W | Random W |
|---|---|---|---|---|---|---|---|---|---|---|---|---|---|---|
| Bangladesh | JiVitA−4 (21) | BSID−III | | | | | | | | | | | | |
| Bangladesh | RDNS (22) | CDI | 730 | 334 | −0.13 | 0.26 (0.13, 0.40) | 0.09 | 0.11 | 873 | 455 | −0.11 | 0.09 (−0.02, 0.19) | 0.25 | 0.15 |
| Bangladesh | WASH−B (23) | CDI | 498 | 1529 | 0.06 | 0.11 (0.00, 0.21) | 0.16 | 0.14 | 606 | 1781 | −0.08 | 0.08 (−0.01, 0.18) | 0.31 | 0.15 |
| Burkina Faso | iLiNS−Zinc (24) | DMC | 480 | 225 | −0.16 | 0.32 (0.13, 0.51) | 0.04 | 0.08 | 264 | 150 | −0.37 | 0.43 (0.23, 0.63) | 0.07 | 0.11 |
| Ghana | GHANA (25) | | | | | | | | | | | | | |
| Ghana | iLiNS−DYADG (26) | CDI | 291 | 531 | 0.00 | 0.03 (−0.12, 0.17) | 0.08 | 0.11 | 35 | 116 | −0.04 | −0.10 (−0.48, 0.27) | 0.02 | 0.06 |
| Haiti | HAITI (27) | Total sounds | | | | | | | | | | | | |
| Kenya | WASH−B (28) | EASQ | 1031 | 3535 | 0.01 | 0.01 (−0.06, 0.08) | 0.36 | 0.17 | 257 | 1012 | −0.04 | 0.07 (−0.07, 0.20) | 0.15 | 0.14 |
| Madagascar | MAHAY (29) | ASQI | | | | | | | | | | | | |
| Malawi | iLiNS−DYADM (30) | CDI | 127 | 261 | −0.01 | −0.01 (−0.23, 0.21) | 0.04 | 0.07 | 87 | 175 | −0.01 | 0.07 (−0.17, 0.32) | 0.05 | 0.09 |
| Malawi | iLiNS−DOSE (31) | CDI | 479 | 158 | −0.14 | 0.18 (0.00, 0.35) | 0.05 | 0.08 | 163 | 61 | 0.21 | −0.27 (−0.57, 0.04) | 0.03 | 0.08 |
| Mali | PROMIS CS (32) | DMC | 683 | 664 | −0.07 | 0.18 (0.00, 0.37) | 0.05 | 0.08 | 238 | 274 | −0.07 | 0.06 (−0.13, 0.25) | 0.08 | 0.11 |
| Zimbabwe | SHINE (HIV−) (33) | CDI | 581 | 569 | −0.03 | 0.08 (−0.05, 0.20) | 0.10 | 0.12 | 96 | 93 | −0.13 | 0.21 (−0.08, 0.51) | 0.03 | 0.08 |
| Zimbabwe | SHINE (HIV+) (34) | CDI | 121 | 110 | −0.11 | 0.23 (−0.02, 0.48) | 0.03 | 0.05 | 23 | 22 | −0.51 | 0.37 (−0.21, 0.95) | 0.01 | 0.03 |
| | | | **5021** | **7916** | | **I² = 0.58, Tau² = 0.01** | | | **2642** | **4139** | | **I² = 0.54, Tau² = 0.02** | | |
| **Fixed** | | | | | | **0.09 (0.05, 0.14)** | | | | | | **0.10 (0.04, 0.15)** | | |
| **Random** | | | | | | **0.12 (0.06, 0.19)** | | | | | | **0.10 (−0.01, 0.21)** | | |

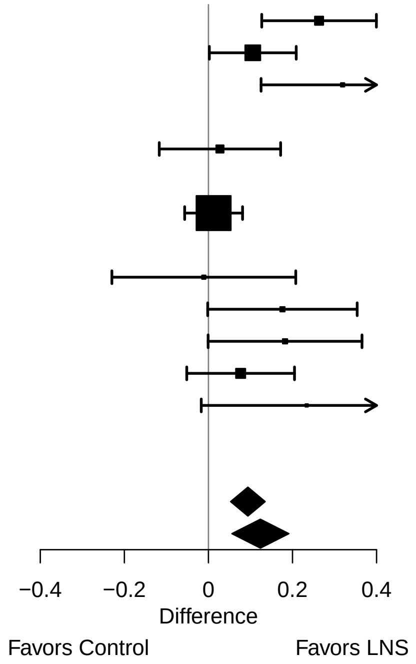

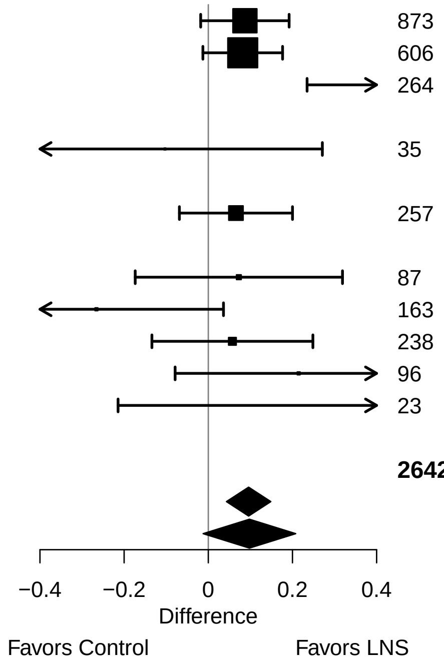

#### Supplemental figure 5A: Mean difference in language z-score

#### 5A3: Stratified by Maternal age

**P−for−interaction = 0.361**
**Difference in MDs = −0.03 (−0.08, 0.03)**

| | | | At least 25 y | | | | | | Less than 25 y | | | | | |
|---|---|---|---|---|---|---|---|---|---|---|---|---|---|---|
| **Country** | **Trial** | **Tool** | **LNS N** | **Control N** | **Control Mean** | **MD (95% CI)** | **Fixed W** | **Random W** | **LNS N** | **Control N** | **Control Mean** | **MD (95% CI)** | **Fixed W** | **Random W** |
| Bangladesh | JiVitA−4 (21) | BSID−III | 200 | 68 | 0.06 | −0.14 (−0.42, 0.14) | 0.03 | 0.06 | 242 | 74 | 0.05 | −0.03 (−0.27, 0.20) | 0.03 | 0.05 |
| Bangladesh | RDNS (22) | CDI | 453 | 220 | −0.22 | 0.24 (0.03, 0.45) | 0.05 | 0.07 | 1210 | 594 | −0.07 | 0.14 (0.04, 0.24) | 0.17 | 0.15 |
| Bangladesh | WASH−B (23) | CDI | 477 | 1475 | −0.04 | 0.05 (−0.08, 0.17) | 0.14 | 0.10 | 627 | 1863 | −0.01 | 0.12 (0.04, 0.20) | 0.27 | 0.18 |
| Burkina Faso | iLiNS−Zinc (24) | DMC | 421 | 232 | −0.29 | 0.47 (0.33, 0.62) | 0.11 | 0.09 | 321 | 141 | −0.16 | 0.20 (−0.09, 0.49) | 0.02 | 0.03 |
| Ghana | GHANA (25) | | | | | | | | | | | | | |
| Ghana | iLiNS−DYADG (26) | CDI | 211 | 405 | 0.05 | 0.11 (−0.07, 0.28) | 0.07 | 0.08 | 120 | 253 | −0.08 | −0.17 (−0.37, 0.03) | 0.04 | 0.06 |
| Haiti | HAITI (27) | Total sounds | 102 | 103 | 0.06 | −0.16 (−0.43, 0.11) | 0.03 | 0.06 | 47 | 45 | 0.05 | −0.03 (−0.44, 0.38) | 0.01 | 0.02 |
| Kenya | WASH−B (28) | EASQ | 798 | 2622 | 0.02 | 0.01 (−0.08, 0.10) | 0.25 | 0.10 | 549 | 2092 | −0.03 | 0.03 (−0.05, 0.12) | 0.24 | 0.18 |
| Madagascar | MAHAY (29) | ASQI | 852 | 901 | 0.03 | −0.09 (−0.28, 0.10) | 0.06 | 0.08 | 759 | 703 | 0.08 | −0.13 (−0.33, 0.06) | 0.04 | 0.07 |
| Malawi | iLiNS−DYADM (30) | CDI | 103 | 225 | 0.02 | 0.09 (−0.15, 0.33) | 0.04 | 0.06 | 112 | 214 | −0.03 | −0.05 (−0.27, 0.17) | 0.04 | 0.06 |
| Malawi | iLiNS−DOSE (31) | CDI | 345 | 124 | −0.05 | 0.01 (−0.19, 0.22) | 0.05 | 0.07 | 288 | 90 | 0.00 | 0.08 (−0.15, 0.31) | 0.03 | 0.05 |
| Mali | PROMIS CS (32) | DMC | 492 | 492 | −0.07 | 0.17 (−0.02, 0.36) | 0.06 | 0.08 | 435 | 452 | −0.07 | 0.12 (−0.07, 0.31) | 0.05 | 0.07 |
| Zimbabwe | SHINE (HIV−) (33) | CDI | 414 | 384 | 0.06 | 0.10 (−0.06, 0.25) | 0.09 | 0.09 | 315 | 310 | −0.18 | 0.10 (−0.08, 0.27) | 0.05 | 0.08 |
| Zimbabwe | SHINE (HIV+) (34) | CDI | 122 | 107 | −0.08 | 0.20 (−0.08, 0.47) | 0.03 | 0.06 | 29 | 26 | −0.22 | 0.25 (−0.32, 0.82) | 0.01 | 0.01 |
| | | | **4990** | **7358** | | **I² = 0.73, Tau² = 0.02** | | | **5054** | **6857** | | **I² = 0.31, Tau² = 0.00** | | |
| **Fixed** | | | | | | **0.09 (0.05, 0.14)** | | | | | | **0.07 (0.03, 0.11)** | | |
| **Random** | | | | | | **0.09 (0.00, 0.18)** | | | | | | **0.05 (0.00, 0.11)** | | |

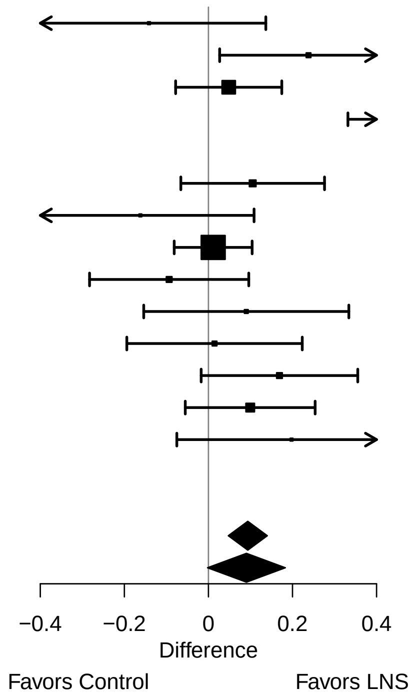

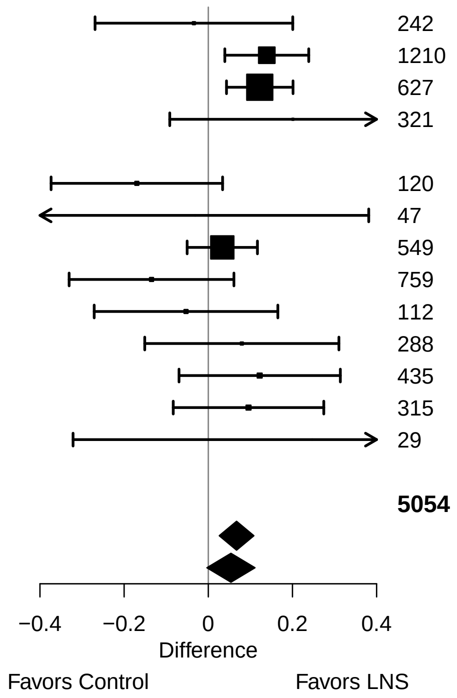

#### Supplemental figure 5A: Mean difference in language z-score

#### 5A4: Stratified by Maternal education

**P−for−interaction = 0.411**
**Difference in MDs = 0.03 (−0.04, 0.09)**

| Country | Trial | Tool | LNS N | Control N | Control Mean | Primary or greater MD (95% CI) | Fixed W | Random W | LNS N | Control N | Control Mean | Incomplete or no formal MD (95% CI) | Fixed W | Random W |
|---|---|---|---|---|---|---|---|---|---|---|---|---|---|---|
| Bangladesh | JiVitA−4 (21) | BSID−III | 292 | 92 | 0.25 | −0.15 (−0.41, 0.11) | 0.02 | 0.06 | 152 | 51 | −0.27 | 0.03 (−0.20, 0.25) | 0.04 | 0.07 |
| Bangladesh | RDNS (22) | CDI | 1245 | 588 | −0.03 | 0.17 (0.07, 0.27) | 0.15 | 0.13 | 418 | 226 | −0.33 | 0.13 (−0.03, 0.30) | 0.08 | 0.10 |
| Bangladesh | WASH−B (23) | CDI | 781 | 2406 | 0.08 | 0.12 (0.04, 0.20) | 0.25 | 0.14 | 328 | 947 | −0.28 | 0.05 (−0.08, 0.18) | 0.12 | 0.12 |
| Burkina Faso | iLiNS−Zinc (24) | DMC | 32 | 9 | 0.06 | 0.10 (−0.48, 0.67) | 0.00 | 0.02 | 712 | 366 | −0.25 | 0.37 (0.21, 0.54) | 0.08 | 0.10 |
| Ghana | GHANA (25) | | | | | | | | | | | | | |
| Ghana | iLiNS−DYADG (26) | CDI | 256 | 519 | 0.01 | −0.03 (−0.17, 0.12) | 0.07 | 0.10 | 75 | 139 | −0.05 | 0.13 (−0.16, 0.43) | 0.03 | 0.05 |
| Haiti | HAITI (27) | Total sounds | 130 | 126 | 0.11 | −0.18 (−0.42, 0.07) | 0.03 | 0.06 | 20 | 23 | −0.13 | 0.17 (−0.40, 0.74) | 0.01 | 0.02 |
| Kenya | WASH−B (28) | EASQ | 658 | 2258 | 0.15 | 0.01 (−0.07, 0.08) | 0.27 | 0.14 | 702 | 2484 | −0.15 | 0.03 (−0.05, 0.11) | 0.34 | 0.16 |
| Madagascar | MAHAY (29) | ASQI | 339 | 416 | 0.43 | −0.25 (−0.49, −0.01) | 0.03 | 0.06 | 1274 | 1188 | −0.08 | −0.04 (−0.22, 0.13) | 0.07 | 0.09 |
| Malawi | iLiNS−DYADM (30) | CDI | 33 | 69 | 0.17 | 0.21 (−0.16, 0.57) | 0.01 | 0.03 | 181 | 367 | −0.03 | −0.02 (−0.20, 0.16) | 0.07 | 0.09 |
| Malawi | iLiNS−DOSE (31) | CDI | 148 | 52 | 0.11 | 0.07 (−0.24, 0.38) | 0.02 | 0.04 | 486 | 162 | −0.07 | 0.04 (−0.14, 0.22) | 0.07 | 0.09 |
| Mali | PROMIS CS (32) | DMC | 100 | 94 | 0.17 | −0.03 (−0.37, 0.30) | 0.01 | 0.04 | 826 | 850 | −0.10 | 0.17 (0.00, 0.33) | 0.08 | 0.10 |
| Zimbabwe | SHINE (HIV−) (33) | CDI | 742 | 715 | −0.04 | 0.11 (−0.01, 0.23) | 0.10 | 0.12 | 27 | 24 | −0.12 | −0.42 (−1.04, 0.20) | 0.01 | 0.01 |
| Zimbabwe | SHINE (HIV+) (34) | CDI | 145 | 126 | −0.09 | 0.22 (−0.01, 0.46) | 0.03 | 0.06 | 9 | 8 | −0.18 | −0.09 (−1.06, 0.88) | 0.00 | 0.01 |
| | | | **4901** | **7470** | | **I² = 0.53, Tau² = 0.01** | | | **5210** | **6835** | | **I² = 0.45, Tau² = 0.01** | | |
| **Fixed** | | | | | | **0.06 (0.02, 0.10)** | | | | | | **0.07 (0.03, 0.12)** | | |
| **Random** | | | | | | **0.04 (−0.04, 0.11)** | | | | | | **0.08 (0.01, 0.15)** | | |

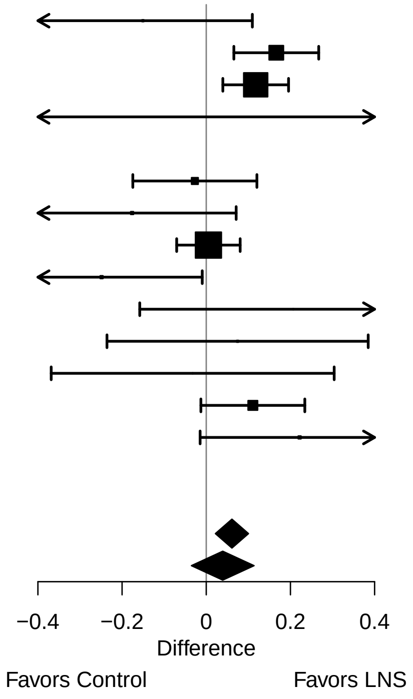

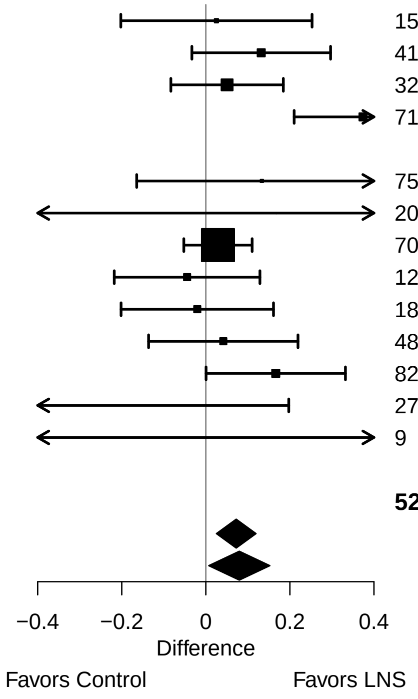

#### Supplemental figure 5A: Mean difference in language z-score

#### 5A5: Stratified by Maternal depressive symptoms

**P−for−interaction = 0.950**
**Difference in MDs = 0.00 (−0.07, 0.08)**

| | | | Less than 75th percentile | | | | | | At least 75th percentile | | | | | |
|---|---|---|---|---|---|---|---|---|---|---|---|---|---|---|
| **Country** | **Trial** | **Tool** | **LNS N** | **Control N** | **Control Mean** | **MD (95% CI)** | **Fixed W** | **Random W** | **LNS N** | **Control N** | **Control Mean** | **MD (95% CI)** | **Fixed W** | **Random W** |
| Bangladesh | JiVitA−4 (21) | BSID−III | | | | | | | | | | | | |
| Bangladesh | RDNS (22) | CDI | 1081 | 471 | −0.06 | 0.18 (0.06, 0.31) | 0.11 | 0.14 | 520 | 288 | −0.21 | 0.11 (−0.03, 0.25) | 0.23 | 0.23 |
| Bangladesh | WASH−B (23) | CDI | 856 | 2410 | 0.01 | 0.11 (0.02, 0.19) | 0.25 | 0.17 | 232 | 859 | −0.12 | 0.02 (−0.13, 0.17) | 0.20 | 0.20 |
| Burkina Faso | iLiNS−Zinc (24) | DMC | | | | | | | | | | | | |
| Ghana | GHANA (25) | | | | | | | | | | | | | |
| Ghana | iLiNS−DYADG (26) | CDI | 241 | 435 | 0.00 | −0.01 (−0.17, 0.15) | 0.07 | 0.12 | 78 | 202 | 0.00 | 0.10 (−0.16, 0.36) | 0.07 | 0.07 |
| Haiti | HAITI (27) | Total sounds | | | | | | | | | | | | |
| Kenya | WASH−B (28) | EASQ | 960 | 3317 | 0.01 | 0.01 (−0.06, 0.08) | 0.37 | 0.18 | 317 | 1156 | 0.11 | 0.02 (−0.11, 0.14) | 0.29 | 0.29 |
| Madagascar | MAHAY (29) | ASQI | 615 | 667 | 0.08 | −0.17 (−0.43, 0.10) | 0.02 | 0.07 | 268 | 242 | 0.03 | −0.01 (−0.28, 0.27) | 0.06 | 0.06 |
| Malawi | iLiNS−DYADM (30) | CDI | 153 | 302 | −0.01 | 0.01 (−0.19, 0.20) | 0.05 | 0.10 | 47 | 112 | 0.00 | 0.11 (−0.25, 0.48) | 0.03 | 0.03 |
| Malawi | iLiNS−DOSE (31) | CDI | | | | | | | | | | | | |
| Mali | PROMIS CS (32) | DMC | | | | | | | | | | | | |
| Zimbabwe | SHINE (HIV−) (33) | CDI | 566 | 552 | −0.02 | 0.06 (−0.07, 0.19) | 0.10 | 0.14 | 190 | 173 | −0.12 | 0.21 (0.00, 0.42) | 0.10 | 0.10 |
| Zimbabwe | SHINE (HIV+) (34) | CDI | 127 | 107 | −0.22 | 0.35 (0.11, 0.60) | 0.03 | 0.08 | 32 | 31 | 0.32 | −0.32 (−0.91, 0.26) | 0.01 | 0.01 |
| | | | **4599** | **8261** | | **I² = 0.57, Tau² = 0.01** | | | **1684** | **3063** | | **I² = 0.00, Tau² = 0.00** | | |
| **Fixed** | | | | | | **0.06 (0.02, 0.11)** | | | | | | **0.06 (−0.01, 0.13)** | | |
| **Random** | | | | | | **0.07 (−0.02, 0.16)** | | | | | | **0.06 (−0.01, 0.13)** | | |

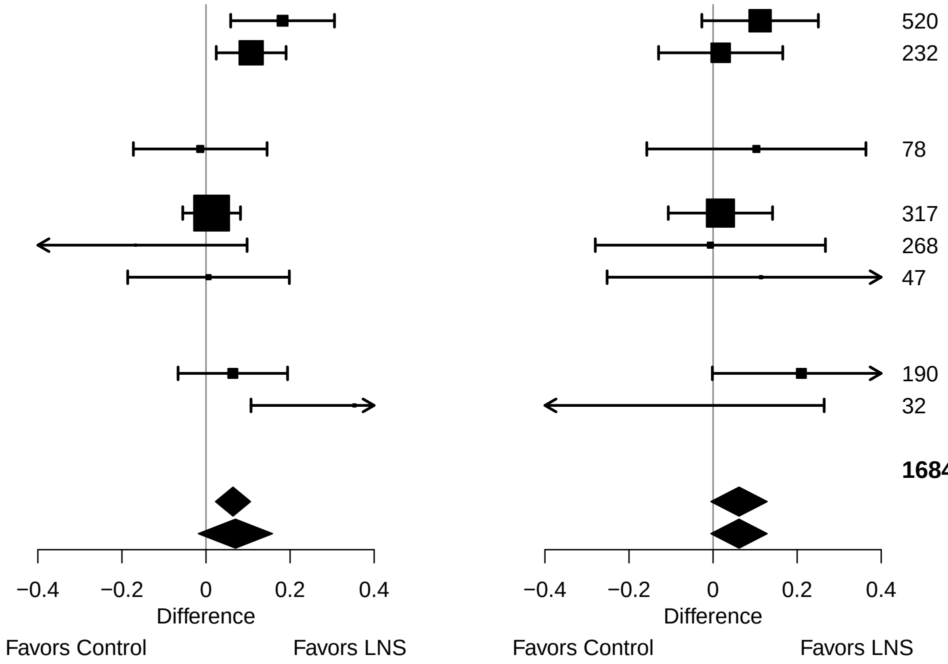

#### Supplemental figure 5A: Mean difference in language z-score

#### 5A6: Stratified by Child sex

**P−for−interaction = 0.424**
**Difference in MDs = 0.02 (−0.03, 0.08)**

| Country | Trial | Tool | Male LNS N | Male Control N | Male Control Mean | Male MD (95% CI) | Male Fixed W | Male Random W | Female LNS N | Female Control N | Female Control Mean | Female MD (95% CI) | Female Fixed W | Female Random W |
|---|---|---|---|---|---|---|---|---|---|---|---|---|---|---|
| Bangladesh | JiVitA−4 (21) | BSID−III | 231 | 70 | 0.04 | −0.05 (−0.35, 0.25) | 0.02 | 0.03 | 214 | 73 | 0.09 | −0.12 (−0.34, 0.10) | 0.04 | 0.07 |
| Bangladesh | RDNS (22) | CDI | 833 | 411 | −0.04 | 0.05 (−0.07, 0.18) | 0.13 | 0.13 | 830 | 403 | −0.18 | 0.28 (0.14, 0.42) | 0.10 | 0.09 |
| Bangladesh | WASH−B (23) | CDI | 545 | 1685 | −0.02 | 0.09 (−0.01, 0.19) | 0.19 | 0.15 | 564 | 1668 | −0.03 | 0.10 (0.01, 0.19) | 0.21 | 0.10 |
| Burkina Faso | iLiNS−Zinc (24) | DMC | 372 | 193 | −0.21 | 0.32 (0.10, 0.55) | 0.04 | 0.05 | 374 | 182 | −0.28 | 0.41 (0.25, 0.58) | 0.07 | 0.08 |
| Ghana | GHANA (25) | | | | | | | | | | | | | |
| Ghana | iLiNS−DYADG (26) | CDI | 164 | 306 | 0.03 | −0.08 (−0.28, 0.11) | 0.05 | 0.06 | 167 | 352 | −0.03 | 0.10 (−0.08, 0.27) | 0.06 | 0.08 |
| Haiti | HAITI (27) | Total sounds | 71 | 60 | 0.04 | −0.08 (−0.43, 0.28) | 0.02 | 0.02 | 79 | 89 | 0.09 | −0.17 (−0.47, 0.13) | 0.02 | 0.05 |
| Kenya | WASH−B (28) | EASQ | 682 | 2260 | 0.00 | −0.01 (−0.10, 0.08) | 0.25 | 0.17 | 680 | 2485 | −0.01 | 0.05 (−0.04, 0.13) | 0.26 | 0.10 |
| Madagascar | MAHAY (29) | ASQI | 795 | 787 | 0.04 | −0.08 (−0.26, 0.10) | 0.06 | 0.07 | 818 | 817 | 0.07 | −0.15 (−0.36, 0.06) | 0.04 | 0.07 |
| Malawi | iLiNS−DYADM (30) | CDI | 105 | 205 | −0.03 | 0.10 (−0.13, 0.33) | 0.04 | 0.05 | 110 | 234 | 0.02 | −0.06 (−0.30, 0.17) | 0.03 | 0.07 |
| Malawi | iLiNS−DOSE (31) | CDI | 320 | 117 | −0.05 | 0.07 (−0.15, 0.28) | 0.04 | 0.06 | 325 | 104 | −0.02 | 0.02 (−0.20, 0.24) | 0.04 | 0.07 |
| Mali | PROMIS CS (32) | DMC | 469 | 514 | −0.08 | 0.17 (0.00, 0.33) | 0.07 | 0.08 | 458 | 430 | −0.07 | 0.13 (−0.08, 0.33) | 0.04 | 0.07 |
| Zimbabwe | SHINE (HIV−) (33) | CDI | 412 | 378 | −0.07 | 0.14 (−0.03, 0.31) | 0.07 | 0.08 | 405 | 403 | −0.01 | 0.03 (−0.14, 0.20) | 0.07 | 0.08 |
| Zimbabwe | SHINE (HIV+) (34) | CDI | 82 | 76 | −0.08 | 0.15 (−0.18, 0.48) | 0.02 | 0.03 | 83 | 71 | −0.19 | 0.35 (0.04, 0.65) | 0.02 | 0.05 |
| | | | **5081** | **7062** | | **I² = 0.26, Tau² = 0.00** | | | **5107** | **7311** | | **I² = 0.69, Tau² = 0.02** | | |
| **Fixed** | | | | | | **0.05 (0.01, 0.10)** | | | | | | **0.09 (0.05, 0.14)** | | |
| **Random** | | | | | | **0.06 (0.00, 0.12)** | | | | | | **0.08 (−0.01, 0.18)** | | |

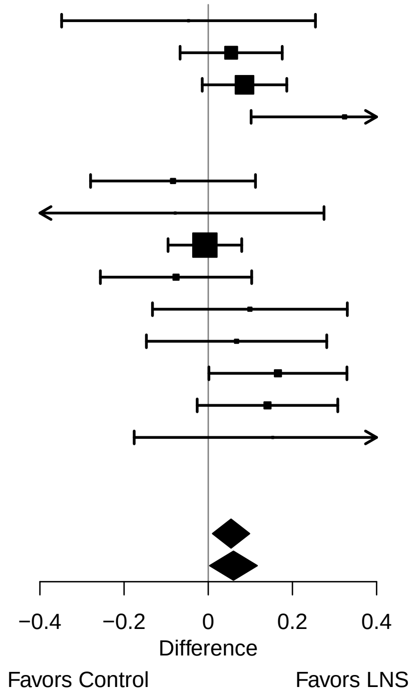

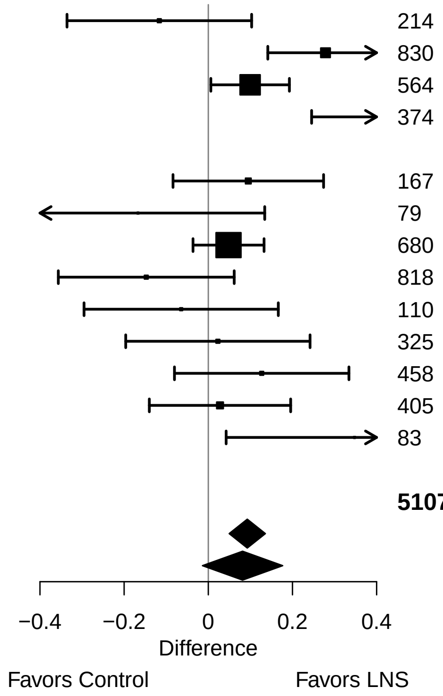

#### Supplemental figure 5A: Mean difference in language z-score

#### 5A7: Stratified by Child birth order

**P−for−interaction = 0.519**
**Difference in MDs = −0.02 (−0.09, 0.04)**

| Country | Trial | Tool | Later born LNS N | Later born Control N | Later born Control Mean | Later born MD (95% CI) | Later born Fixed W | Later born Random W | Firstborn LNS N | Firstborn Control N | Firstborn Control Mean | Firstborn MD (95% CI) | Firstborn Fixed W | Firstborn Random W |
|---|---|---|---|---|---|---|---|---|---|---|---|---|---|---|
| Bangladesh | JiVitA−4 (21) | BSID−III | 111 | 31 | −0.08 | −0.14 (−0.60, 0.32) | 0.01 | 0.02 | 333 | 111 | 0.11 | −0.06 (−0.29, 0.17) | 0.06 | 0.08 |
| Bangladesh | RDNS (22) | CDI | 974 | 507 | −0.21 | 0.24 (0.13, 0.35) | 0.13 | 0.10 | 688 | 307 | 0.06 | 0.04 (−0.12, 0.20) | 0.12 | 0.11 |
| Bangladesh | WASH−B (23) | CDI | 693 | 2187 | −0.07 | 0.08 (−0.02, 0.17) | 0.15 | 0.11 | 395 | 1060 | 0.09 | 0.10 (0.00, 0.21) | 0.27 | 0.13 |
| Burkina Faso | iLiNS−Zinc (24) | DMC | 587 | 290 | −0.25 | 0.41 (0.25, 0.57) | 0.06 | 0.08 | 159 | 85 | −0.21 | 0.22 (−0.14, 0.57) | 0.02 | 0.05 |
| Ghana | GHANA (25) | | | | | | | | | | | | | |
| Ghana | iLiNS−DYADG (26) | CDI | 224 | 445 | 0.04 | 0.05 (−0.11, 0.22) | 0.06 | 0.08 | 107 | 213 | −0.10 | −0.08 (−0.30, 0.13) | 0.07 | 0.09 |
| Haiti | HAITI (27) | Total sounds | 89 | 99 | −0.01 | −0.06 (−0.33, 0.22) | 0.02 | 0.05 | 61 | 50 | 0.22 | −0.27 (−0.67, 0.12) | 0.02 | 0.04 |
| Kenya | WASH−B (28) | EASQ | 1071 | 3767 | 0.00 | 0.01 (−0.06, 0.08) | 0.30 | 0.12 | 289 | 974 | −0.03 | 0.07 (−0.05, 0.19) | 0.22 | 0.12 |
| Madagascar | MAHAY (29) | ASQI | 1162 | 1118 | 0.01 | −0.10 (−0.28, 0.08) | 0.05 | 0.07 | 433 | 465 | 0.16 | −0.14 (−0.38, 0.09) | 0.06 | 0.08 |
| Malawi | iLiNS−DYADM (30) | CDI | 170 | 350 | 0.03 | 0.03 (−0.15, 0.22) | 0.04 | 0.07 | 45 | 87 | −0.14 | −0.04 (−0.37, 0.29) | 0.03 | 0.06 |
| Malawi | iLiNS−DOSE (31) | CDI | 424 | 151 | −0.09 | 0.05 (−0.14, 0.24) | 0.04 | 0.07 | 134 | 41 | 0.18 | 0.03 (−0.31, 0.37) | 0.03 | 0.05 |
| Mali | PROMIS CS (32) | DMC | 770 | 811 | −0.07 | 0.16 (−0.01, 0.34) | 0.05 | 0.08 | 142 | 122 | −0.05 | 0.08 (−0.25, 0.40) | 0.03 | 0.06 |
| Zimbabwe | SHINE (HIV−) (33) | CDI | 587 | 562 | 0.06 | 0.06 (−0.08, 0.20) | 0.08 | 0.09 | 194 | 185 | −0.34 | 0.19 (−0.03, 0.40) | 0.07 | 0.09 |
| Zimbabwe | SHINE (HIV+) (34) | CDI | 144 | 114 | −0.06 | 0.14 (−0.13, 0.41) | 0.02 | 0.05 | 18 | 29 | −0.42 | 0.77 (0.28, 1.27) | 0.01 | 0.03 |
| | | | **7006** | **10432** | | **I² = 0.66, Tau² = 0.01** | | | **2998** | **3729** | | **I² = 0.38, Tau² = 0.02** | | |
| **Fixed** | | | | | | **0.08 (0.04, 0.12)** | | | | | | **0.05 (0.00, 0.11)** | | |
| **Random** | | | | | | **0.09 (0.01, 0.16)** | | | | | | **0.05 (−0.05, 0.14)** | | |

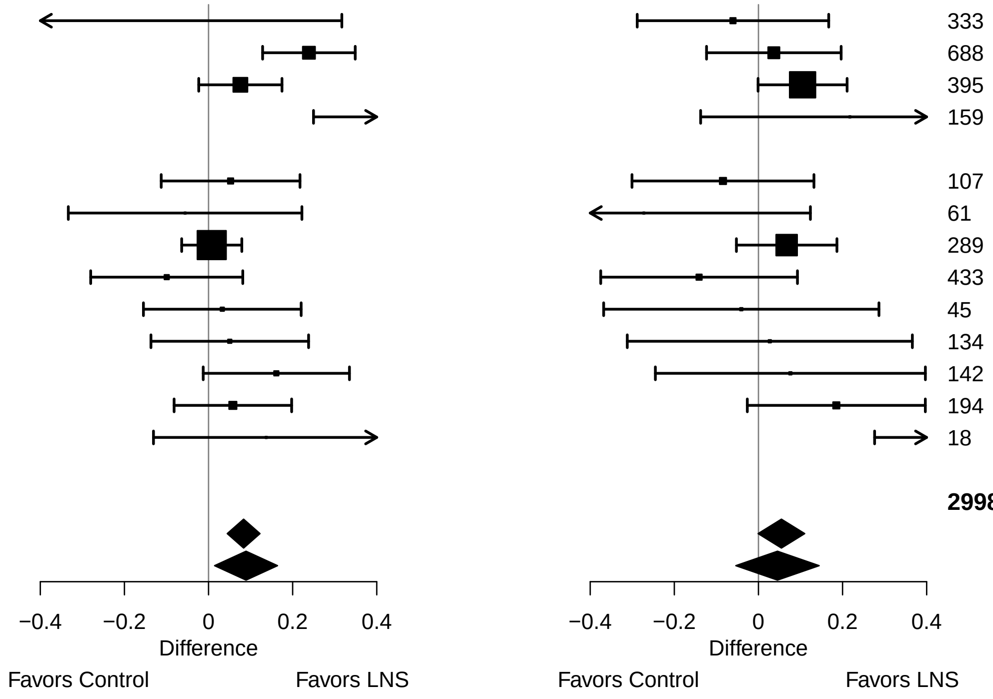

#### Supplemental figure 5A: Mean difference in language z-score

#### 5A8: Stratified by Child baseline stunting

**P−for−interaction = 0.255**
**Difference in MDs = 0.07 (−0.05, 0.20)**

| | | | No | | | | | | Yes | | | | | |
|---|---|---|---|---|---|---|---|---|---|---|---|---|---|---|
| **Country** | **Trial** | **Tool** | **LNS N** | **Control N** | **Control Mean** | **MD (95% CI)** | **Fixed W** | **Random W** | **LNS N** | **Control N** | **Control Mean** | **MD (95% CI)** | **Fixed W** | **Random W** |
| Bangladesh | JiVitA−4 (21) | BSID−III | 327 | 114 | 0.15 | −0.08 (−0.33, 0.17) | 0.04 | 0.08 | 116 | 29 | −0.30 | 0.03 (−0.32, 0.38) | 0.10 | 0.10 |
| Bangladesh | RDNS (22) | CDI | 1246 | 573 | −0.03 | 0.13 (0.04, 0.22) | 0.35 | 0.16 | 355 | 185 | −0.38 | 0.26 (0.01, 0.51) | 0.19 | 0.19 |
| Bangladesh | WASH−B (23) | CDI | | | | | | | | | | | | |
| Burkina Faso | iLiNS−Zinc (24) | DMC | 576 | 294 | −0.18 | 0.37 (0.17, 0.57) | 0.07 | 0.11 | 170 | 80 | −0.47 | 0.37 (0.13, 0.60) | 0.22 | 0.22 |
| Ghana | GHANA (25) | | | | | | | | | | | | | |
| Ghana | iLiNS−DYADG (26) | CDI | 279 | 548 | 0.04 | −0.03 (−0.18, 0.11) | 0.14 | 0.13 | 28 | 63 | −0.28 | 0.21 (−0.26, 0.68) | 0.05 | 0.05 |
| Haiti | HAITI (27) | Total sounds | 137 | 132 | 0.09 | −0.11 (−0.35, 0.13) | 0.05 | 0.09 | 12 | 17 | −0.08 | −0.35 (−1.09, 0.38) | 0.02 | 0.02 |
| Kenya | WASH−B (28) | EASQ | | | | | | | | | | | | |
| Madagascar | MAHAY (29) | ASQI | | | | | | | | | | | | |
| Malawi | iLiNS−DYADM (30) | CDI | 153 | 323 | 0.04 | 0.00 (−0.19, 0.19) | 0.08 | 0.11 | 48 | 97 | −0.25 | 0.24 (−0.12, 0.61) | 0.09 | 0.09 |
| Malawi | iLiNS−DOSE (31) | CDI | 464 | 147 | 0.07 | −0.02 (−0.20, 0.16) | 0.09 | 0.11 | 181 | 74 | −0.24 | 0.15 (−0.12, 0.43) | 0.16 | 0.16 |
| Mali | PROMIS CS (32) | DMC | | | | | | | | | | | | |
| Zimbabwe | SHINE (HIV−) (33) | CDI | 523 | 482 | −0.01 | 0.16 (0.01, 0.30) | 0.14 | 0.13 | 119 | 100 | −0.25 | −0.03 (−0.34, 0.28) | 0.13 | 0.13 |
| Zimbabwe | SHINE (HIV+) (34) | CDI | 112 | 87 | −0.04 | 0.24 (−0.03, 0.51) | 0.04 | 0.08 | 33 | 32 | −0.19 | 0.20 (−0.38, 0.79) | 0.04 | 0.04 |
| | | | **3817** | **2700** | | **I² = 0.60, Tau² = 0.01** | | | **1062** | **677** | | **I² = 0.00, Tau² = 0.00** | | |
| **Fixed** | | | | | | **0.09 (0.03, 0.14)** | | | | | | **0.19 (0.08, 0.30)** | | |
| **Random** | | | | | | **0.07 (−0.03, 0.18)** | | | | | | **0.19 (0.08, 0.30)** | | |

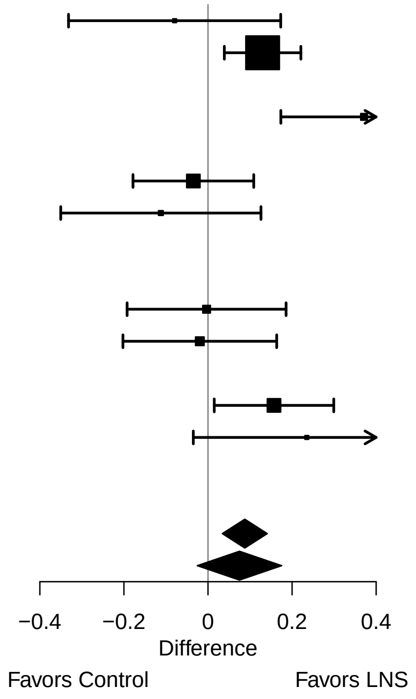

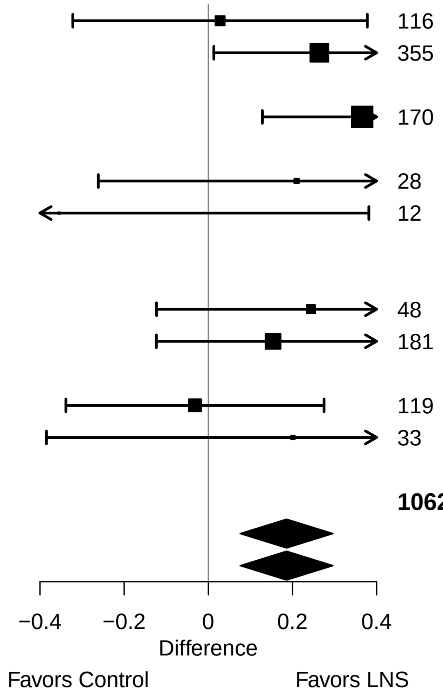

##### Supplemental figure 5A: Mean difference in language z-score

#### 5A9: Stratified by Child baseline acute malnutrition

**P−for−interaction = 0.034**
**Difference in MDs = 0.16 (0.01, 0.31)**

| | | | No | | | | | | Yes | | | | | |
|---|---|---|---|---|---|---|---|---|---|---|---|---|---|---|
| **Country** | **Trial** | **Tool** | **LNS N** | **Control N** | **Control Mean** | **MD (95% CI)** | **Fixed W** | **Random W** | **LNS N** | **Control N** | **Control Mean** | **MD (95% CI)** | **Fixed W** | **Random W** |
| Bangladesh | JiVitA−4 (21) | BSID−III | 359 | 112 | 0.14 | −0.14 (−0.39, 0.12) | 0.04 | 0.08 | 84 | 31 | −0.21 | 0.12 (−0.16, 0.40) | 0.23 | 0.23 |
| Bangladesh | RDNS (22) | CDI | 1477 | 706 | −0.08 | 0.16 (0.08, 0.25) | 0.35 | 0.17 | 124 | 53 | −0.56 | 0.26 (−0.24, 0.75) | 0.08 | 0.08 |
| Bangladesh | WASH−B (23) | CDI | | | | | | | | | | | | |
| Burkina Faso | iLiNS−Zinc (24) | DMC | 556 | 266 | −0.14 | 0.34 (0.17, 0.52) | 0.09 | 0.12 | 190 | 109 | −0.49 | 0.40 (0.18, 0.61) | 0.40 | 0.40 |
| Ghana | GHANA (25) | | | | | | | | | | | | | |
| Ghana | iLiNS−DYADG (26) | CDI | 289 | 555 | 0.01 | −0.03 (−0.17, 0.12) | 0.13 | 0.14 | 19 | 56 | 0.00 | 0.28 (−0.18, 0.75) | 0.09 | 0.09 |
| Haiti | HAITI (27) | Total sounds | | | | | | | | | | | | |
| Kenya | WASH−B (28) | EASQ | | | | | | | | | | | | |
| Madagascar | MAHAY (29) | ASQI | | | | | | | | | | | | |
| Malawi | iLiNS−DYADM (30) | CDI | 187 | 386 | 0.02 | 0.02 (−0.15, 0.20) | 0.09 | 0.12 | 14 | 37 | −0.45 | 0.33 (−0.26, 0.93) | 0.05 | 0.05 |
| Malawi | iLiNS−DOSE (31) | CDI | 608 | 206 | −0.02 | 0.04 (−0.12, 0.20) | 0.11 | 0.13 | 37 | 15 | −0.21 | 0.15 (−0.48, 0.78) | 0.05 | 0.05 |
| Mali | PROMIS CS (32) | DMC | | | | | | | | | | | | |
| Zimbabwe | SHINE (HIV−) (33) | CDI | 608 | 553 | −0.03 | 0.10 (−0.03, 0.23) | 0.15 | 0.15 | 36 | 34 | −0.43 | 0.44 (−0.03, 0.91) | 0.09 | 0.09 |
| Zimbabwe | SHINE (HIV+) (34) | CDI | 135 | 113 | −0.05 | 0.22 (−0.03, 0.47) | 0.04 | 0.09 | 12 | 7 | −0.65 | 0.64 (−0.29, 1.58) | 0.02 | 0.02 |
| | | | **4219** | **2897** | | **I² = 0.61, Tau² = 0.01** | | | **516** | **342** | | **I² = 0.00, Tau² = 0.00** | | |
| **Fixed** | | | | | | **0.11 (0.06, 0.16)** | | | | | | **0.31 (0.17, 0.44)** | | |
| **Random** | | | | | | **0.10 (0.00, 0.19)** | | | | | | **0.31 (0.17, 0.44)** | | |

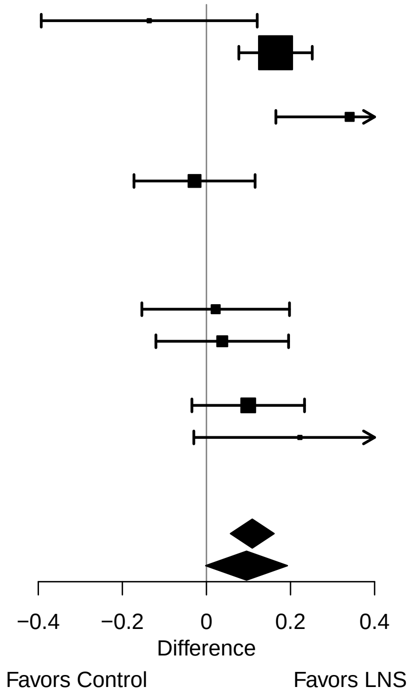

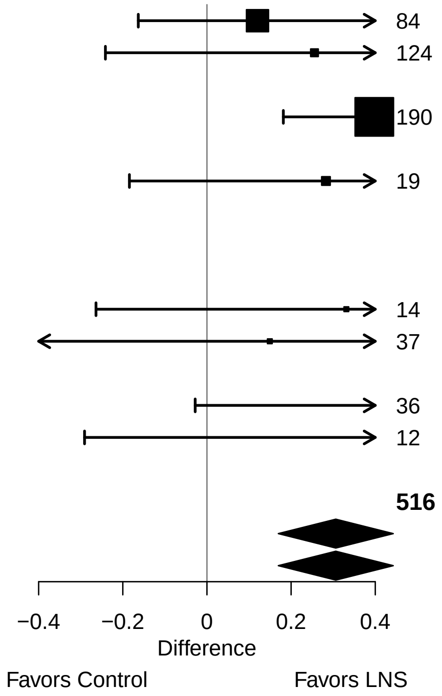

#### Supplemental figure 5A: Mean difference in language z-score

##### 5A10: Stratified by Child baseline anemia

**P−for−interaction = 0.650**
**Difference in MDs = −0.04 (−0.19, 0.12)**

| Country | Trial | Tool | Not anemic LNS N | Not anemic Control N | Not anemic Control Mean | Not anemic MD (95% CI) | Not anemic Fixed W | Not anemic Random W | Anemic LNS N | Anemic Control N | Anemic Control Mean | Anemic MD (95% CI) | Anemic Fixed W | Anemic Random W |
|---|---|---|---|---|---|---|---|---|---|---|---|---|---|---|
| Bangladesh | JiVitA−4 (21) | BSID−III | | | | | | | | | | | | |
| Bangladesh | RDNS (22) | CDI | 212 | 96 | −0.24 | 0.28 (−0.02, 0.58) | 0.14 | 0.18 | 318 | 160 | −0.21 | 0.30 (0.08, 0.51) | 0.17 | 0.19 |
| Bangladesh | WASH−B (23) | CDI | | | | | | | | | | | | |
| Burkina Faso | iLiNS−Zinc (24) | DMC | 59 | 34 | −0.35 | 0.48 (0.08, 0.87) | 0.08 | 0.14 | 687 | 341 | −0.23 | 0.36 (0.19, 0.52) | 0.30 | 0.23 |
| Ghana | GHANA (25) | | | | | | | | | | | | | |
| Ghana | iLiNS−DYADG (26) | CDI | 192 | 351 | 0.05 | −0.10 (−0.27, 0.07) | 0.41 | 0.27 | 90 | 204 | −0.02 | 0.07 (−0.19, 0.32) | 0.12 | 0.17 |
| Haiti | HAITI (27) | Total sounds | | | | | | | | | | | | |
| Kenya | WASH−B (28) | EASQ | | | | | | | | | | | | |
| Madagascar | MAHAY (29) | ASQI | | | | | | | | | | | | |
| Malawi | iLiNS−DYADM (30) | CDI | 66 | 147 | −0.06 | 0.20 (−0.12, 0.51) | 0.13 | 0.18 | 138 | 275 | 0.02 | −0.03 (−0.23, 0.16) | 0.21 | 0.21 |
| Malawi | iLiNS−DOSE (31) | CDI | 224 | 94 | −0.01 | 0.10 (−0.12, 0.33) | 0.24 | 0.23 | 417 | 126 | −0.06 | 0.03 (−0.18, 0.24) | 0.19 | 0.20 |
| Mali | PROMIS CS (32) | DMC | | | | | | | | | | | | |
| Zimbabwe | SHINE (HIV−) (33) | CDI | | | | | | | | | | | | |
| Zimbabwe | SHINE (HIV+) (34) | CDI | | | | | | | | | | | | |
| | | | **753** | **722** | | **I² = 0.61, Tau² = 0.02** | | | **1650** | **1106** | | **I² = 0.69, Tau² = 0.02** | | |
| **Fixed** | | | | | | **0.08 (−0.03, 0.20)** | | | | | | **0.17 (0.08, 0.26)** | | |
| **Random** | | | | | | **0.15 (−0.04, 0.33)** | | | | | | **0.15 (−0.01, 0.30)** | | |

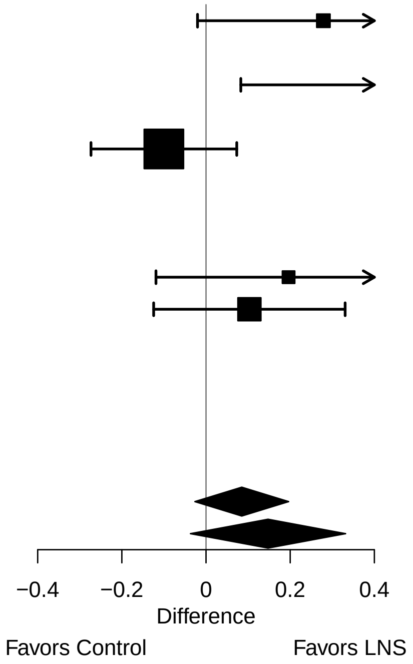

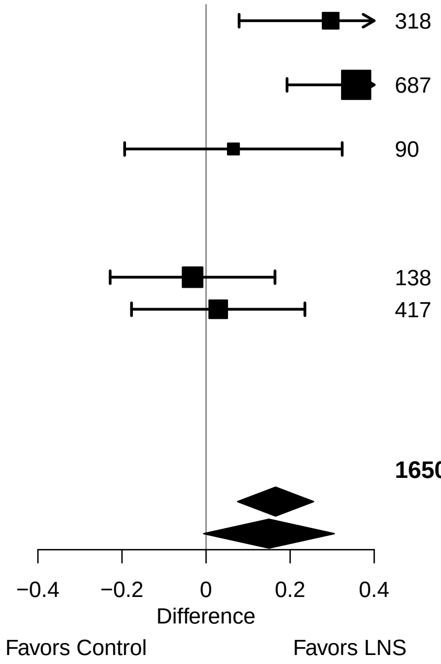

### Supplemental figure 5B: Language lowest decile prevalence ratio

#### 5B1: Stratified by Maternal height

**P−for−interaction = 0.678**
**Ratio of PRs = 1.06 (0.81, 1.39)**

| Country | Trial | Tool | LNS N | Control N | Control Prevalence | At least 150.1 cm PR (95% CI) | Fixed W | Random W | LNS N | Control N | Control Prevalence | Less than 150.1 cm PR (95% CI) | Fixed W | Random W |
|---|---|---|---|---|---|---|---|---|---|---|---|---|---|---|
| Bangladesh | JiVitA−4 (21) | BSID−III | | | | | | | | | | | | |
| Bangladesh | RDNS (22) | CDI | 868 | 431 | 12.5 | 0.58 (0.40, 0.84) | 0.16 | 0.23 | 735 | 358 | 12.0 | 0.93 (0.65, 1.32) | 0.36 | 0.36 |
| Bangladesh | WASH−B (23) | CDI | 589 | 1812 | 10.2 | 0.82 (0.62, 1.08) | 0.27 | 0.28 | 515 | 1499 | 10.8 | 0.77 (0.56, 1.07) | 0.42 | 0.42 |
| Burkina Faso | iLiNS−Zinc (24) | DMC | | | | | | | | | | | | |
| Ghana | GHANA (25) | | | | | | | | | | | | | |
| Ghana | iLiNS−DYADG (26) | CDI | | | | | | | | | | | | |
| Haiti | HAITI (27) | Total sounds | | | | | | | | | | | | |
| Kenya | WASH−B (28) | EASQ | 1237 | 4372 | 9.7 | 0.96 (0.78, 1.18) | 0.50 | 0.33 | 51 | 175 | 15.4 | 0.64 (0.26, 1.58) | 0.05 | 0.05 |
| Madagascar | MAHAY (29) | ASQI | 956 | 986 | 8.7 | 1.18 (0.69, 2.00) | 0.07 | 0.16 | 553 | 538 | 9.9 | 1.14 (0.68, 1.91) | 0.17 | 0.17 |
| Malawi | iLiNS−DYADM (30) | CDI | | | | | | | | | | | | |
| Malawi | iLiNS−DOSE (31) | CDI | | | | | | | | | | | | |
| Mali | PROMIS CS (32) | DMC | | | | | | | | | | | | |
| Zimbabwe | SHINE (HIV−) (33) | CDI | | | | | | | | | | | | |
| Zimbabwe | SHINE (HIV+) (34) | CDI | | | | | | | | | | | | |
| | | | **3650** | **7601** | | **I² = 0.57, Tau² = 0.04** | | | **1854** | **2570** | | **I² = 0.00, Tau² = 0.00** | | |
| **Fixed** | | | | | | **0.86 (0.74, 1.00)** | | | | | | **0.87 (0.70, 1.08)** | | |
| **Random** | | | | | | **0.84 (0.65, 1.10)** | | | | | | **0.87 (0.70, 1.08)** | | |

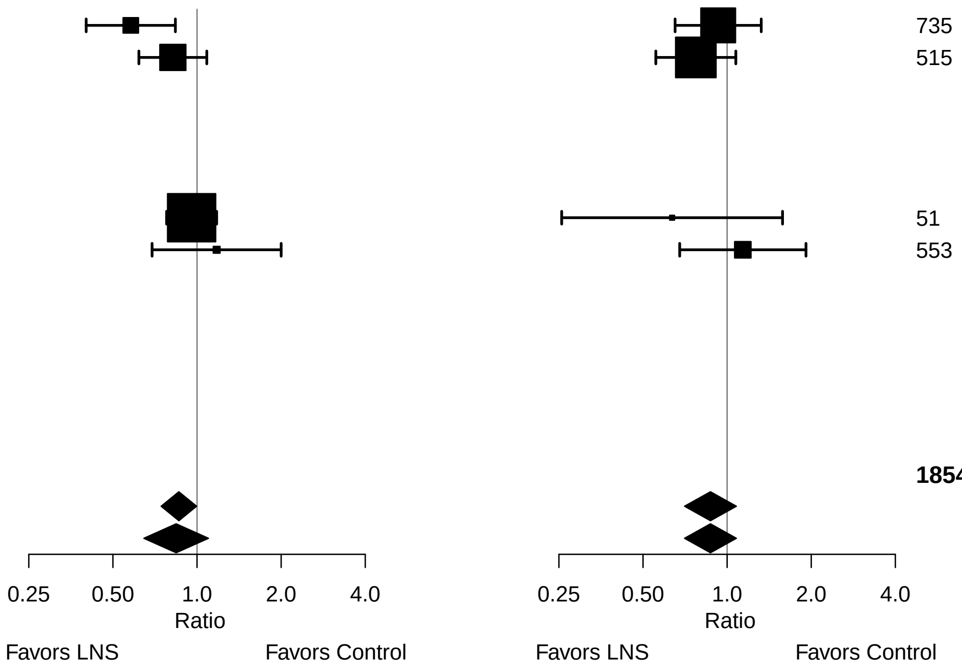

#### Supplemental figure 5B: Language lowest decile prevalence ratio

##### 5B2: Stratified by Maternal BMI

**P−for−interaction = 0.149**
**Ratio of PRs = 1.18 (0.94, 1.47)**

| | | | At least 20 kg/m² | | | | | | Less than 20 kg/m² | | | | | |
|---|---|---|---|---|---|---|---|---|---|---|---|---|---|---|
| Country | Trial | Tool | LNS N | Control N | Control Prevalence | PR (95% CI) | Fixed W | Random W | LNS N | Control N | Control Prevalence | PR (95% CI) | Fixed W | Random W |
| Bangladesh | JiVitA−4 (21) | BSID−III | | | | | | | | | | | | |
| Bangladesh | RDNS (22) | CDI | 730 | 334 | 12.0 | 0.54 (0.36, 0.80) | 0.12 | 0.15 | 873 | 455 | 12.5 | 0.90 (0.66, 1.21) | 0.27 | 0.20 |
| Bangladesh | WASH−B (23) | CDI | 498 | 1529 | 9.5 | 0.63 (0.44, 0.91) | 0.14 | 0.16 | 606 | 1781 | 11.2 | 0.91 (0.70, 1.18) | 0.36 | 0.21 |
| Burkina Faso | iLiNS−Zinc (24) | DMC | 480 | 225 | 12.4 | 0.57 (0.36, 0.91) | 0.09 | 0.12 | 264 | 150 | 20.7 | 0.37 (0.21, 0.63) | 0.09 | 0.15 |
| Ghana | GHANA (25) | | | | | | | | | | | | | |
| Ghana | iLiNS−DYADG (26) | CDI | | | | | | | | | | | | |
| Haiti | HAITI (27) | Total sounds | | | | | | | | | | | | |
| Kenya | WASH−B (28) | EASQ | 1031 | 3535 | 10.0 | 0.94 (0.75, 1.18) | 0.37 | 0.24 | 257 | 1012 | 9.8 | 0.91 (0.61, 1.38) | 0.15 | 0.18 |
| Madagascar | MAHAY (29) | ASQI | | | | | | | | | | | | |
| Malawi | iLiNS−DYADM (30) | CDI | | | | | | | | | | | | |
| Malawi | iLiNS−DOSE (31) | CDI | | | | | | | | | | | | |
| Mali | PROMIS CS (32) | DMC | 683 | 664 | 11.9 | 0.66 (0.45, 0.98) | 0.12 | 0.15 | 238 | 274 | 9.9 | 1.07 (0.67, 1.70) | 0.11 | 0.17 |
| Zimbabwe | SHINE (HIV−) (33) | CDI | 581 | 569 | 10.9 | 0.93 (0.67, 1.31) | 0.16 | 0.18 | 96 | 93 | 15.1 | 0.42 (0.17, 1.00) | 0.03 | 0.09 |
| Zimbabwe | SHINE (HIV+) (34) | CDI | | | | | | | | | | | | |
| | | | **4003** | **6856** | | **I² = 0.54, Tau² = 0.03** | | | **2334** | **3765** | | **I² = 0.62, Tau² = 0.13** | | |
| **Fixed** | | | | | | **0.76 (0.67, 0.87)** | | | | | | **0.83 (0.71, 0.98)** | | |
| **Random** | | | | | | **0.72 (0.59, 0.88)** | | | | | | **0.76 (0.54, 1.06)** | | |

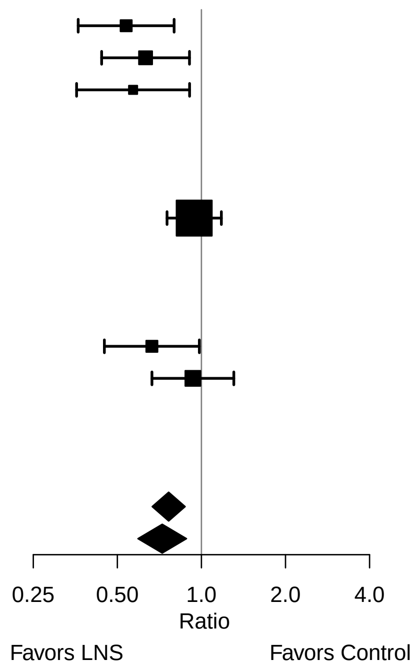

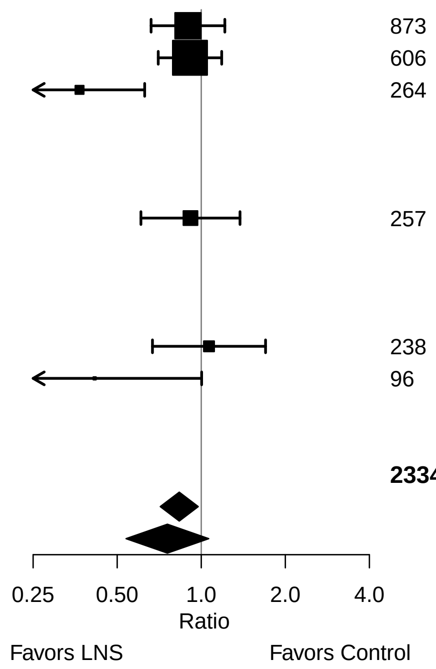

#### Supplemental figure 5B: Language lowest decile prevalence ratio

##### 5B3: Stratified by Maternal age

**P−for−interaction = 0.832**
**Ratio of PRs = 1.02 (0.86, 1.20)**

| Country | Trial | Tool | LNS N | Control N | Control Prevalence | At least 25 y PR (95% CI) | Fixed W | Random W | LNS N | Control N | Control Prevalence | Less than 25 y PR (95% CI) | Fixed W | Random W |
|---|---|---|---|---|---|---|---|---|---|---|---|---|---|---|
| Bangladesh | JiVitA−4 (21) | BSID−III | 200 | 68 | 11.8 | 0.89 (0.48, 1.66) | 0.04 | 0.04 | 242 | 74 | 10.8 | 0.84 (0.38, 1.85) | 0.03 | 0.05 |
| Bangladesh | RDNS (22) | CDI | 453 | 220 | 13.6 | 0.83 (0.57, 1.20) | 0.12 | 0.12 | 1210 | 594 | 12.0 | 0.67 (0.51, 0.88) | 0.21 | 0.13 |
| Bangladesh | WASH−B (23) | CDI | 477 | 1475 | 11.5 | 0.90 (0.66, 1.22) | 0.18 | 0.18 | 627 | 1863 | 9.8 | 0.73 (0.54, 0.99) | 0.17 | 0.13 |
| Burkina Faso | iLiNS−Zinc (24) | DMC | 421 | 232 | 15.1 | 0.52 (0.33, 0.83) | 0.08 | 0.08 | 321 | 141 | 17.0 | 0.38 (0.22, 0.67) | 0.05 | 0.08 |
| Ghana | GHANA (25) | | | | | | | | | | | | | |
| Ghana | iLiNS−DYADG (26) | CDI | 211 | 405 | 10.4 | 0.82 (0.49, 1.39) | 0.06 | 0.06 | 120 | 253 | 8.7 | 1.63 (0.90, 2.95) | 0.05 | 0.07 |
| Haiti | HAITI (27) | Total sounds | | | | | | | | | | | | |
| Kenya | WASH−B (28) | EASQ | 798 | 2622 | 9.9 | 1.01 (0.76, 1.35) | 0.19 | 0.19 | 549 | 2092 | 10.3 | 0.89 (0.66, 1.19) | 0.18 | 0.13 |
| Madagascar | MAHAY (29) | ASQI | 852 | 901 | 9.8 | 1.14 (0.72, 1.82) | 0.08 | 0.08 | 759 | 703 | 8.5 | 1.20 (0.72, 2.01) | 0.06 | 0.09 |
| Malawi | iLiNS−DYADM (30) | CDI | 103 | 225 | 14.7 | 0.66 (0.34, 1.29) | 0.04 | 0.04 | 112 | 214 | 6.5 | 1.36 (0.63, 2.97) | 0.03 | 0.05 |
| Malawi | iLiNS−DOSE (31) | CDI | 345 | 124 | 11.3 | 0.92 (0.52, 1.65) | 0.05 | 0.05 | 288 | 90 | 8.9 | 1.02 (0.48, 2.16) | 0.03 | 0.05 |
| Mali | PROMIS CS (32) | DMC | 492 | 492 | 11.0 | 0.65 (0.42, 1.00) | 0.09 | 0.09 | 435 | 452 | 11.7 | 0.90 (0.61, 1.34) | 0.10 | 0.11 |
| Zimbabwe | SHINE (HIV−) (33) | CDI | 414 | 384 | 9.9 | 0.76 (0.48, 1.19) | 0.08 | 0.08 | 315 | 310 | 12.9 | 0.96 (0.63, 1.47) | 0.09 | 0.10 |
| Zimbabwe | SHINE (HIV+) (34) | CDI | | | | | | | | | | | | |
| | | | **4766** | **7148** | | **I² = 0.00, Tau² = 0.00** | | | **4978** | **6786** | | **I² = 0.50, Tau² = 0.07** | | |
| **Fixed** | | | | | | **0.84 (0.74, 0.95)** | | | | | | **0.83 (0.73, 0.94)** | | |
| **Random** | | | | | | **0.84 (0.74, 0.95)** | | | | | | **0.87 (0.70, 1.07)** | | |

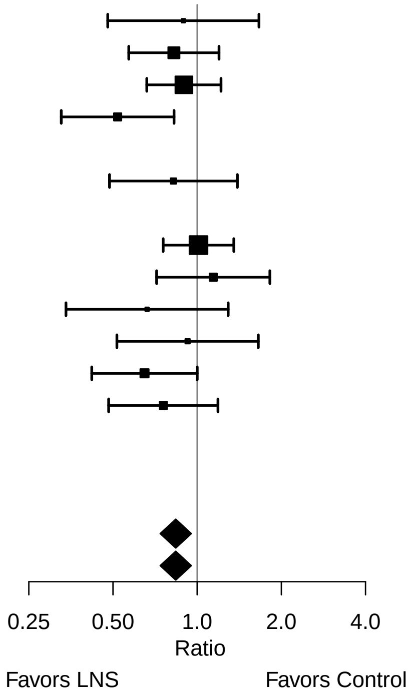

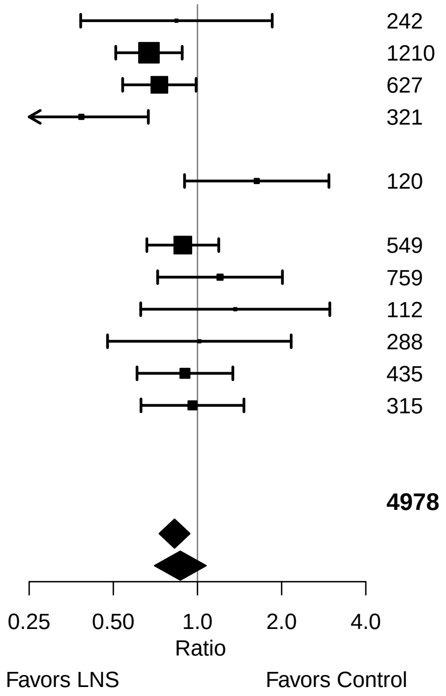

#### Supplemental figure 5B: Language lowest decile prevalence ratio

##### 5B4: Stratified by Maternal education

**P−for−interaction = 0.804**
**Ratio of PRs = 0.97 (0.79, 1.20)**

| Country | Trial | Tool | Primary or greater: LNS N | Control N | Control Prevalence | PR (95% CI) | Fixed W | Random W | Incomplete or no formal: LNS N | Control N | Control Prevalence | PR (95% CI) | Fixed W | Random W |
|---|---|---|---|---|---|---|---|---|---|---|---|---|---|---|
| Bangladesh | JiVitA−4 (21) | BSID−III | 292 | 92 | 8.7 | 0.87 (0.38, 1.98) | 0.03 | 0.08 | 152 | 51 | 15.7 | 0.88 (0.43, 1.81) | 0.04 | 0.04 |
| Bangladesh | RDNS (22) | CDI | 1245 | 588 | 9.9 | 0.67 (0.49, 0.90) | 0.23 | 0.20 | 418 | 226 | 19.0 | 0.83 (0.59, 1.17) | 0.16 | 0.16 |
| Bangladesh | WASH−B (23) | CDI | 781 | 2406 | 8.9 | 0.87 (0.69, 1.09) | 0.40 | 0.22 | 328 | 947 | 14.8 | 0.70 (0.49, 1.00) | 0.15 | 0.15 |
| Burkina Faso | iLiNS−Zinc (24) | DMC | | | | | | | | | | | | |
| Ghana | GHANA (25) | | | | | | | | | | | | | |
| Ghana | iLiNS−DYADG (26) | CDI | 256 | 519 | 9.4 | 0.99 (0.62, 1.58) | 0.10 | 0.16 | 75 | 139 | 10.8 | 1.36 (0.66, 2.81) | 0.04 | 0.04 |
| Haiti | HAITI (27) | Total sounds | | | | | | | | | | | | |
| Kenya | WASH−B (28) | EASQ | 658 | 2258 | 6.9 | 0.84 (0.60, 1.16) | 0.20 | 0.20 | 702 | 2484 | 13.0 | 1.02 (0.81, 1.28) | 0.37 | 0.37 |
| Madagascar | MAHAY (29) | ASQI | 339 | 416 | 3.6 | 2.62 (1.24, 5.52) | 0.04 | 0.10 | 1274 | 1188 | 11.2 | 1.00 (0.63, 1.57) | 0.09 | 0.09 |
| Malawi | iLiNS−DYADM (30) | CDI | | | | | | | | | | | | |
| Malawi | iLiNS−DOSE (31) | CDI | | | | | | | | | | | | |
| Mali | PROMIS CS (32) | DMC | 100 | 94 | 7.4 | 1.21 (0.35, 4.19) | 0.01 | 0.05 | 826 | 850 | 11.8 | 0.74 (0.52, 1.05) | 0.16 | 0.16 |
| Zimbabwe | SHINE (HIV−) (33) | CDI | | | | | | | | | | | | |
| Zimbabwe | SHINE (HIV+) (34) | CDI | | | | | | | | | | | | |
| | | | **3671** | **6373** | | **I² = 0.50, Tau² = 0.08** | | | **3775** | **5885** | | **I² = 0.00, Tau² = 0.00** | | |
| **Fixed** | | | | | | **0.86 (0.74, 0.99)** | | | | | | **0.89 (0.77, 1.02)** | | |
| **Random** | | | | | | **0.94 (0.70, 1.26)** | | | | | | **0.89 (0.77, 1.02)** | | |

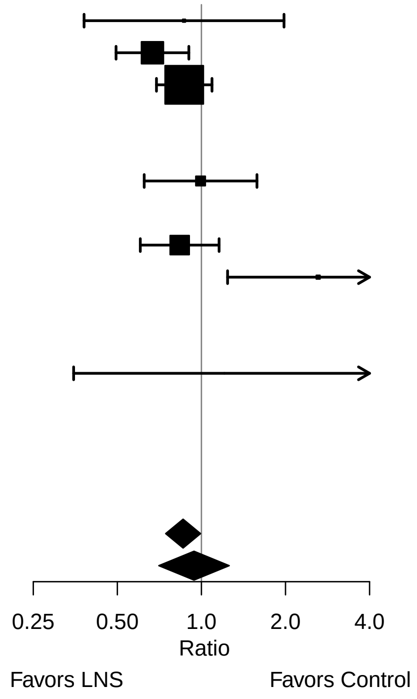

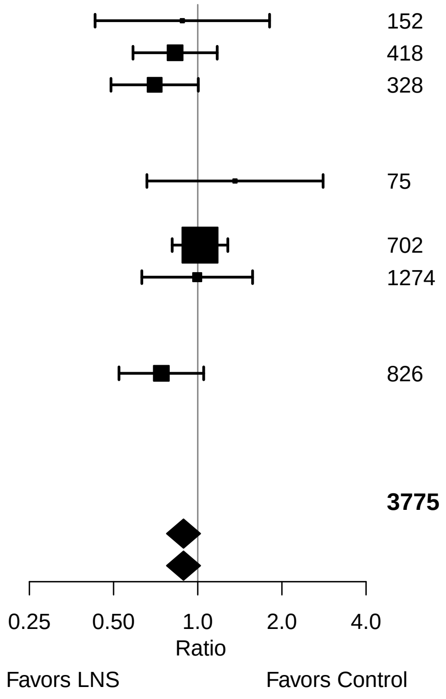

##### Supplemental figure 5B: Language lowest decile prevalence ratio

#### 5B5: Stratified by Maternal depressive symptoms

**P−for−interaction = 0.204**
**Ratio of PRs = 0.86 (0.69, 1.08)**

| | | | Less than 75th percentile | | | | | | At least 75th percentile | | | | | |
|---|---|---|---|---|---|---|---|---|---|---|---|---|---|---|
| **Country** | **Trial** | **Tool** | **LNS N** | **Control N** | **Control Prevalence** | **PR (95% CI)** | **Fixed W** | **Random W** | **LNS N** | **Control N** | **Control Prevalence** | **PR (95% CI)** | **Fixed W** | **Random W** |
| Bangladesh | JiVitA−4 (21) | BSID−III | | | | | | | | | | | | |
| Bangladesh | RDNS (22) | CDI | 1081 | 471 | 11.9 | 0.65 (0.46, 0.92) | 0.14 | 0.15 | 520 | 288 | 13.5 | 0.87 (0.62, 1.21) | 0.28 | 0.28 |
| Bangladesh | WASH−B (23) | CDI | 856 | 2410 | 10.2 | 0.68 (0.52, 0.90) | 0.23 | 0.22 | 232 | 859 | 11.3 | 1.18 (0.80, 1.76) | 0.20 | 0.20 |
| Burkina Faso | iLiNS−Zinc (24) | DMC | | | | | | | | | | | | |
| Ghana | GHANA (25) | | | | | | | | | | | | | |
| Ghana | iLiNS−DYADG (26) | CDI | 241 | 435 | 10.8 | 1.00 (0.64, 1.57) | 0.08 | 0.10 | 78 | 202 | 7.4 | 1.38 (0.61, 3.13) | 0.05 | 0.05 |
| Haiti | HAITI (27) | Total sounds | | | | | | | | | | | | |
| Kenya | WASH−B (28) | EASQ | 960 | 3317 | 9.5 | 0.90 (0.72, 1.12) | 0.34 | 0.29 | 317 | 1156 | 8.7 | 1.06 (0.74, 1.52) | 0.25 | 0.25 |
| Madagascar | MAHAY (29) | ASQI | 615 | 667 | 8.8 | 1.29 (0.70, 2.35) | 0.05 | 0.06 | 268 | 242 | 12.0 | 0.72 (0.37, 1.40) | 0.07 | 0.07 |
| Malawi | iLiNS−DYADM (30) | CDI | 153 | 302 | 9.9 | 0.92 (0.50, 1.68) | 0.05 | 0.06 | 47 | 112 | 12.5 | 0.85 (0.33, 2.23) | 0.03 | 0.03 |
| Malawi | iLiNS−DOSE (31) | CDI | | | | | | | | | | | | |
| Mali | PROMIS CS (32) | DMC | | | | | | | | | | | | |
| Zimbabwe | SHINE (HIV−) (33) | CDI | 566 | 552 | 10.3 | 0.89 (0.60, 1.31) | 0.11 | 0.13 | 190 | 173 | 14.5 | 0.69 (0.41, 1.17) | 0.12 | 0.12 |
| Zimbabwe | SHINE (HIV+) (34) | CDI | | | | | | | | | | | | |
| | | | **4472** | **8154** | | **I² = 0.15, Tau² = 0.01** | | | **1652** | **3032** | | **I² = 0.00, Tau² = 0.00** | | |
| **Fixed** | | | | | | **0.83 (0.73, 0.94)** | | | | | | **0.95 (0.80, 1.14)** | | |
| **Random** | | | | | | **0.83 (0.72, 0.96)** | | | | | | **0.95 (0.80, 1.14)** | | |

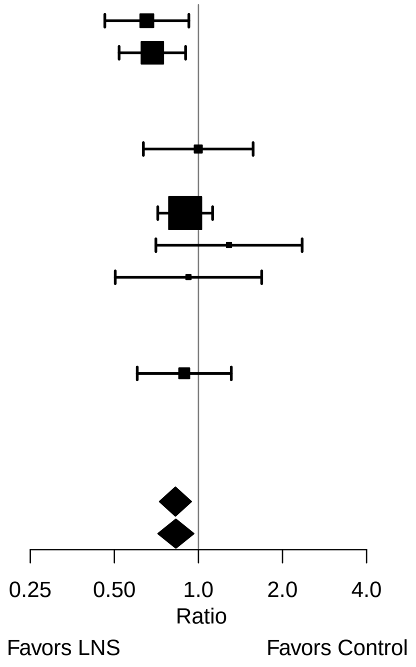

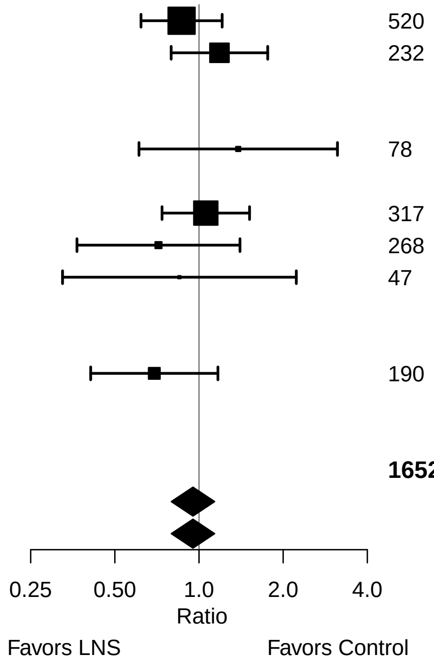

#### Supplemental figure 5B: Language lowest decile prevalence ratio

##### 5B6: Stratified by Child sex

**P−for−interaction = 0.255**
**Ratio of PRs = 0.91 (0.77, 1.07)**

| Country | Trial | Tool | Male LNS N | Male Control N | Male Control Prevalence | Male PR (95% CI) | Male Fixed W | Male Random W | Female LNS N | Female Control N | Female Control Prevalence | Female PR (95% CI) | Female Fixed W | Female Random W |
|---|---|---|---|---|---|---|---|---|---|---|---|---|---|---|
| Bangladesh | JiVitA−4 (21) | BSID−III | 231 | 70 | 14.3 | 0.64 (0.30, 1.34) | 0.03 | 0.04 | 214 | 73 | 8.2 | 1.25 (0.65, 2.42) | 0.04 | 0.05 |
| Bangladesh | RDNS (22) | CDI | 833 | 411 | 10.5 | 1.01 (0.72, 1.42) | 0.13 | 0.12 | 830 | 403 | 14.4 | 0.50 (0.35, 0.73) | 0.12 | 0.11 |
| Bangladesh | WASH−B (23) | CDI | 545 | 1685 | 11.5 | 0.80 (0.60, 1.06) | 0.18 | 0.15 | 564 | 1668 | 9.6 | 0.81 (0.59, 1.12) | 0.17 | 0.13 |
| Burkina Faso | iLiNS−Zinc (24) | DMC | 372 | 193 | 12.4 | 0.48 (0.30, 0.76) | 0.07 | 0.08 | 374 | 182 | 19.2 | 0.44 (0.30, 0.67) | 0.11 | 0.10 |
| Ghana | GHANA (25) | | | | | | | | | | | | | |
| Ghana | iLiNS−DYADG (26) | CDI | 164 | 306 | 8.8 | 1.45 (0.85, 2.48) | 0.05 | 0.06 | 167 | 352 | 10.5 | 0.80 (0.44, 1.43) | 0.05 | 0.07 |
| Haiti | HAITI (27) | Total sounds | | | | | | | | | | | | |
| Kenya | WASH−B (28) | EASQ | 682 | 2260 | 10.7 | 1.03 (0.80, 1.32) | 0.23 | 0.17 | 680 | 2485 | 9.6 | 0.86 (0.62, 1.17) | 0.17 | 0.13 |
| Madagascar | MAHAY (29) | ASQI | 795 | 787 | 8.9 | 1.13 (0.70, 1.84) | 0.06 | 0.07 | 818 | 817 | 9.5 | 1.20 (0.71, 2.04) | 0.06 | 0.08 |
| Malawi | iLiNS−DYADM (30) | CDI | 105 | 205 | 8.8 | 0.76 (0.33, 1.76) | 0.02 | 0.03 | 110 | 234 | 12.4 | 0.95 (0.52, 1.76) | 0.05 | 0.06 |
| Malawi | iLiNS−DOSE (31) | CDI | 320 | 117 | 11.1 | 0.87 (0.47, 1.61) | 0.04 | 0.05 | 325 | 104 | 9.6 | 1.06 (0.54, 2.07) | 0.04 | 0.05 |
| Mali | PROMIS CS (32) | DMC | 469 | 514 | 10.7 | 0.76 (0.51, 1.12) | 0.09 | 0.10 | 458 | 430 | 12.1 | 0.78 (0.52, 1.16) | 0.11 | 0.11 |
| Zimbabwe | SHINE (HIV−) (33) | CDI | 412 | 378 | 11.6 | 0.92 (0.62, 1.37) | 0.09 | 0.10 | 405 | 403 | 9.9 | 0.87 (0.52, 1.46) | 0.06 | 0.08 |
| Zimbabwe | SHINE (HIV+) (34) | CDI | 82 | 76 | 13.2 | 0.74 (0.29, 1.88) | 0.02 | 0.02 | 83 | 71 | 11.3 | 0.64 (0.24, 1.72) | 0.02 | 0.03 |
| | | | **5010** | **7002** | | **I² = 0.28, Tau² = 0.02** | | | **5028** | **7222** | | **I² = 0.43, Tau² = 0.04** | | |
| **Fixed** | | | | | | **0.89 (0.79, 1.00)** | | | | | | **0.76 (0.67, 0.87)** | | |
| **Random** | | | | | | **0.88 (0.76, 1.02)** | | | | | | **0.78 (0.65, 0.93)** | | |

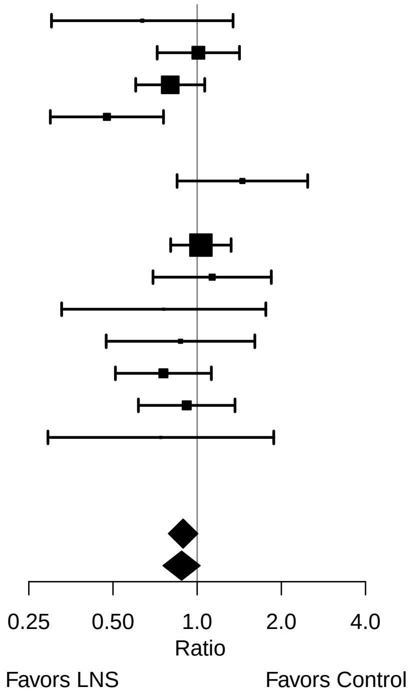

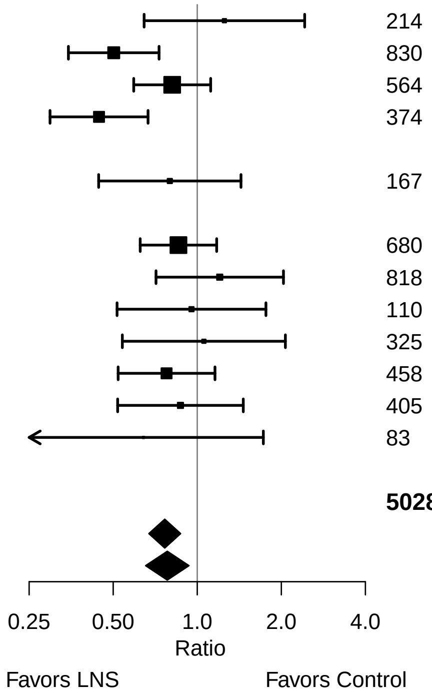

**Supplemental figure 5B: Language lowest decile prevalence ratio**

#### 5B7: Stratified by Child birth order

**P−for−interaction = 0.362**
**Ratio of PRs = 1.10 (0.90, 1.35)**

| Country | Trial | Tool | LNS N | Control N | Control Prevalence | Later born PR (95% CI) | Fixed W | Random W | LNS N | Control N | Control Prevalence | Firstborn PR (95% CI) | Fixed W | Random W |
|---|---|---|---|---|---|---|---|---|---|---|---|---|---|---|
| Bangladesh | JiVitA−4 (21) | BSID−III | | | | | | | | | | | | |
| Bangladesh | RDNS (22) | CDI | 974 | 507 | 14.0 | 0.70 (0.53, 0.92) | 0.17 | 0.15 | 688 | 307 | 9.8 | 0.79 (0.51, 1.22) | 0.16 | 0.15 |
| Bangladesh | WASH−B (23) | CDI | 693 | 2187 | 11.6 | 0.75 (0.57, 0.99) | 0.18 | 0.15 | 395 | 1060 | 8.3 | 0.91 (0.63, 1.32) | 0.23 | 0.17 |
| Burkina Faso | iLiNS−Zinc (24) | DMC | 587 | 290 | 15.5 | 0.47 (0.31, 0.72) | 0.08 | 0.10 | 159 | 85 | 16.5 | 0.42 (0.20, 0.89) | 0.05 | 0.08 |
| Ghana | GHANA (25) | | | | | | | | | | | | | |
| Ghana | iLiNS−DYADG (26) | CDI | 224 | 445 | 10.1 | 0.88 (0.53, 1.46) | 0.05 | 0.08 | 107 | 213 | 8.9 | 1.57 (0.83, 2.97) | 0.08 | 0.10 |
| Haiti | HAITI (27) | Total sounds | | | | | | | | | | | | |
| Kenya | WASH−B (28) | EASQ | 1071 | 3767 | 9.8 | 0.97 (0.77, 1.24) | 0.24 | 0.17 | 289 | 974 | 11.4 | 0.88 (0.57, 1.36) | 0.16 | 0.15 |
| Madagascar | MAHAY (29) | ASQI | 1162 | 1118 | 9.1 | 1.20 (0.75, 1.91) | 0.06 | 0.08 | 433 | 465 | 9.7 | 1.07 (0.61, 1.89) | 0.10 | 0.11 |
| Malawi | iLiNS−DYADM (30) | CDI | 170 | 350 | 11.7 | 0.65 (0.36, 1.19) | 0.04 | 0.06 | 45 | 87 | 6.9 | 2.26 (0.81, 6.31) | 0.03 | 0.05 |
| Malawi | iLiNS−DOSE (31) | CDI | | | | | | | | | | | | |
| Mali | PROMIS CS (32) | DMC | 770 | 811 | 11.2 | 0.75 (0.53, 1.07) | 0.11 | 0.12 | 142 | 122 | 9.0 | 1.09 (0.52, 2.31) | 0.05 | 0.08 |
| Zimbabwe | SHINE (HIV−) (33) | CDI | 587 | 562 | 9.4 | 0.94 (0.62, 1.42) | 0.08 | 0.10 | 194 | 185 | 15.7 | 0.79 (0.49, 1.27) | 0.13 | 0.13 |
| Zimbabwe | SHINE (HIV+) (34) | CDI | | | | | | | | | | | | |
| | | | **6238** | **10037** | | **I² = 0.43, Tau² = 0.03** | | | **2452** | **3498** | | **I² = 0.29, Tau² = 0.06** | | |
| **Fixed** | | | | | | **0.80 (0.71, 0.90)** | | | | | | **0.91 (0.77, 1.09)** | | |
| **Random** | | | | | | **0.79 (0.67, 0.93)** | | | | | | **0.93 (0.73, 1.19)** | | |

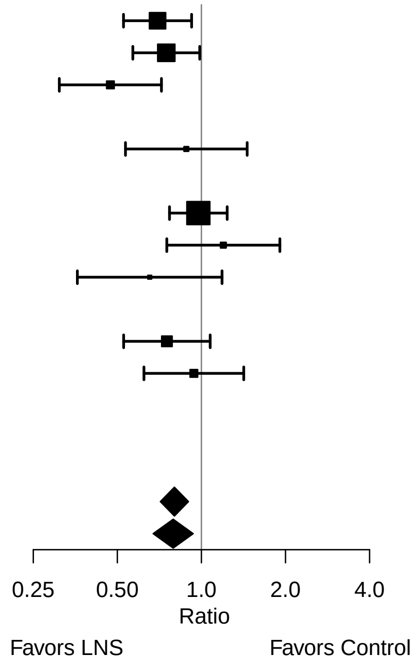

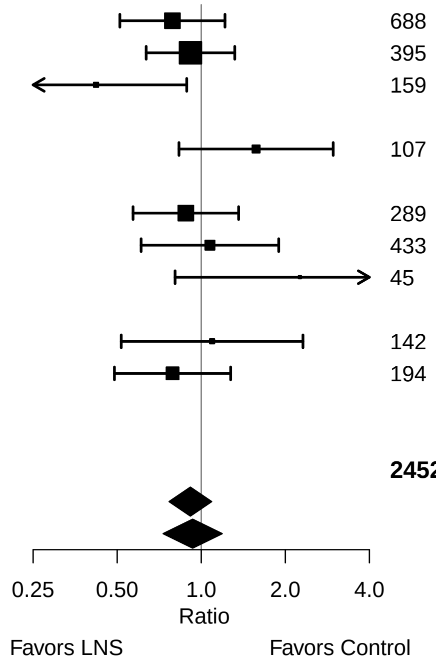

#### Supplemental figure 5B: Language lowest decile prevalence ratio

#### 5B8: Stratified by Child baseline stunting

**P−for−interaction = 0.111**
**Ratio of PRs = 0.79 (0.59, 1.06)**

| Country | Trial | Tool | No: LNS N | No: Control N | No: Control Prevalence | No: PR (95% CI) | No: Fixed W | No: Random W | Yes: LNS N | Yes: Control N | Yes: Control Prevalence | Yes: PR (95% CI) | Yes: Fixed W | Yes: Random W |
|---|---|---|---|---|---|---|---|---|---|---|---|---|---|---|
| Bangladesh | JiVitA−4 (21) | BSID−III | 327 | 114 | 8.8 | 0.91 (0.42, 1.94) | 0.05 | 0.05 | 116 | 29 | 20.7 | 0.67 (0.35, 1.25) | 0.15 | 0.16 |
| Bangladesh | RDNS (22) | CDI | 1246 | 573 | 10.6 | 0.75 (0.58, 0.99) | 0.41 | 0.39 | 355 | 185 | 18.4 | 0.69 (0.46, 1.03) | 0.37 | 0.30 |
| Bangladesh | WASH−B (23) | CDI | | | | | | | | | | | | |
| Burkina Faso | iLiNS−Zinc (24) | DMC | 576 | 294 | 12.6 | 0.55 (0.38, 0.80) | 0.21 | 0.21 | 170 | 80 | 27.5 | 0.30 (0.16, 0.56) | 0.15 | 0.16 |
| Ghana | GHANA (25) | | | | | | | | | | | | | |
| Ghana | iLiNS−DYADG (26) | CDI | | | | | | | | | | | | |
| Haiti | HAITI (27) | Total sounds | | | | | | | | | | | | |
| Kenya | WASH−B (28) | EASQ | | | | | | | | | | | | |
| Madagascar | MAHAY (29) | ASQI | | | | | | | | | | | | |
| Malawi | iLiNS−DYADM (30) | CDI | 153 | 323 | 8.4 | 1.02 (0.54, 1.91) | 0.07 | 0.08 | 48 | 97 | 19.6 | 0.64 (0.27, 1.49) | 0.08 | 0.10 |
| Malawi | iLiNS−DOSE (31) | CDI | 464 | 147 | 8.2 | 1.11 (0.60, 2.05) | 0.08 | 0.08 | 181 | 74 | 14.9 | 0.82 (0.42, 1.60) | 0.13 | 0.14 |
| Mali | PROMIS CS (32) | DMC | | | | | | | | | | | | |
| Zimbabwe | SHINE (HIV−) (33) | CDI | 523 | 482 | 10.0 | 0.77 (0.51, 1.16) | 0.18 | 0.18 | 119 | 100 | 16.0 | 0.74 (0.37, 1.47) | 0.12 | 0.14 |
| Zimbabwe | SHINE (HIV+) (34) | CDI | | | | | | | | | | | | |
| | | | **3289** | **1933** | | **I² = 0.05, Tau² = 0.00** | | | **989** | **565** | | **I² = 0.23, Tau² = 0.03** | | |
| **Fixed** | | | | | | **0.75 (0.63, 0.89)** | | | | | | **0.62 (0.49, 0.79)** | | |
| **Random** | | | | | | **0.76 (0.63, 0.90)** | | | | | | **0.61 (0.46, 0.82)** | | |

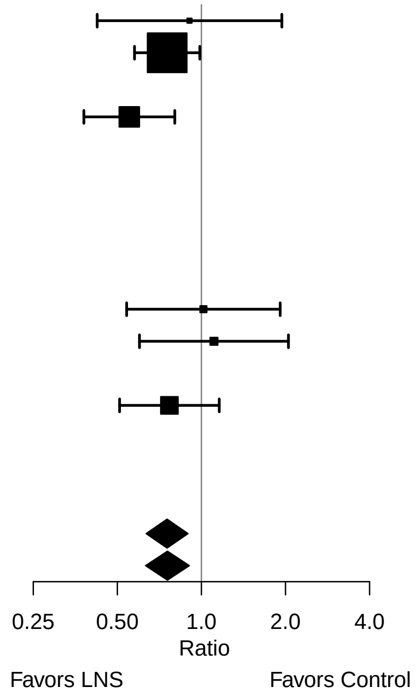

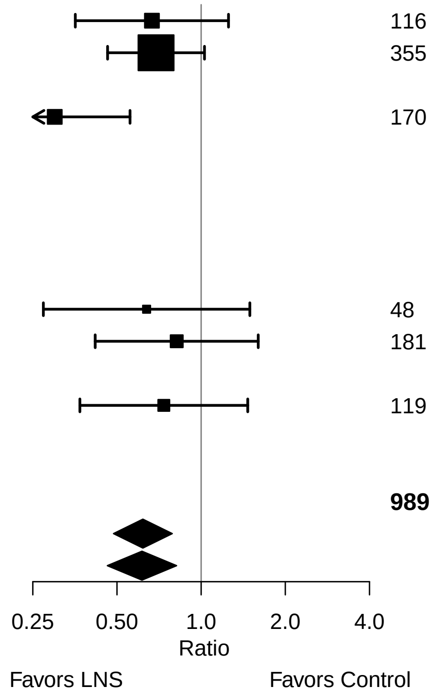

**Supplemental figure 5B: Language lowest decile prevalence ratio**

#### 5B9: Stratified by Child baseline acute malnutrition

**P−for−interaction = 0.588**
**Ratio of PRs = 1.13 (0.73, 1.76)**

| Country | Trial | Tool | No: LNS N | No: Control N | No: Control Prevalence | No: PR (95% CI) | No: Fixed W | No: Random W | Yes: LNS N | Yes: Control N | Yes: Control Prevalence | Yes: PR (95% CI) | Yes: Fixed W | Yes: Random W |
|---|---|---|---|---|---|---|---|---|---|---|---|---|---|---|
| Bangladesh | JiVitA−4 (21) | BSID−III | 359 | 112 | 9.8 | 0.88 (0.43, 1.80) | 0.08 | 0.20 | 84 | 31 | 16.1 | 0.81 (0.33, 2.00) | 0.18 | 0.18 |
| Bangladesh | RDNS (22) | CDI | 1477 | 706 | 11.9 | 0.71 (0.56, 0.90) | 0.73 | 0.48 | 124 | 53 | 20.8 | 0.78 (0.42, 1.44) | 0.39 | 0.39 |
| Bangladesh | WASH−B (23) | CDI | | | | | | | | | | | | |
| Burkina Faso | iLiNS−Zinc (24) | DMC | 556 | 266 | 13.5 | 0.43 (0.27, 0.67) | 0.19 | 0.32 | 190 | 109 | 21.1 | 0.55 (0.31, 0.98) | 0.44 | 0.44 |
| Ghana | GHANA (25) | | | | | | | | | | | | | |
| Ghana | iLiNS−DYADG (26) | CDI | | | | | | | | | | | | |
| Haiti | HAITI (27) | Total sounds | | | | | | | | | | | | |
| Kenya | WASH−B (28) | EASQ | | | | | | | | | | | | |
| Madagascar | MAHAY (29) | ASQI | | | | | | | | | | | | |
| Malawi | iLiNS−DYADM (30) | CDI | | | | | | | | | | | | |
| Malawi | iLiNS−DOSE (31) | CDI | | | | | | | | | | | | |
| Mali | PROMIS CS (32) | DMC | | | | | | | | | | | | |
| Zimbabwe | SHINE (HIV−) (33) | CDI | | | | | | | | | | | | |
| Zimbabwe | SHINE (HIV+) (34) | CDI | | | | | | | | | | | | |
| | | | **2392** | **1084** | | **I² = 0.56, Tau² = 0.07** | | | **398** | **193** | | **I² = 0.00, Tau² = 0.00** | | |
| **Fixed** | | | | | | **0.65 (0.53, 0.80)** | | | | | | **0.67 (0.46, 0.99)** | | |
| **Random** | | | | | | **0.63 (0.43, 0.93)** | | | | | | **0.67 (0.46, 0.99)** | | |

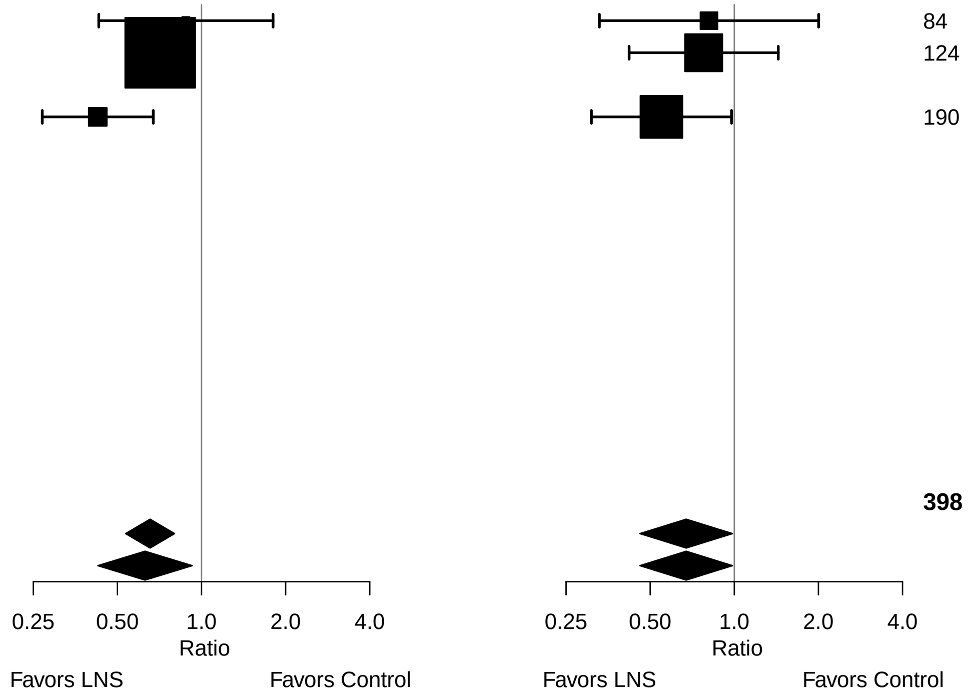

#### Supplemental figure 5B: Language lowest decile prevalence ratio

##### 5B10: Stratified by Child baseline anemia

**P−for−interaction = 0.847**
**Ratio of PRs = 1.05 (0.65, 1.68)**

| Country | Trial | Tool | LNS N | Control N | Control Prevalence | Not anemic PR (95% CI) | Fixed W | Random W | LNS N | Control N | Control Prevalence | Anemic PR (95% CI) | Fixed W | Random W |
|---|---|---|---|---|---|---|---|---|---|---|---|---|---|---|
| Bangladesh | JiVitA−4 (21) | BSID−III | | | | | | | | | | | | |
| Bangladesh | RDNS (22) | CDI | 212 | 96 | 14.6 | 0.71 (0.43, 1.17) | 0.39 | 0.35 | 318 | 160 | 12.5 | 0.70 (0.37, 1.33) | 0.25 | 0.25 |
| Bangladesh | WASH−B (23) | CDI | | | | | | | | | | | | |
| Burkina Faso | iLiNS−Zinc (24) | DMC | | | | | | | | | | | | |
| Ghana | GHANA (25) | | | | | | | | | | | | | |
| Ghana | iLiNS−DYADG (26) | CDI | 192 | 351 | 7.4 | 1.41 (0.81, 2.45) | 0.32 | 0.31 | 90 | 204 | 10.8 | 0.93 (0.44, 1.93) | 0.19 | 0.19 |
| Haiti | HAITI (27) | Total sounds | | | | | | | | | | | | |
| Kenya | WASH−B (28) | EASQ | | | | | | | | | | | | |
| Madagascar | MAHAY (29) | ASQI | | | | | | | | | | | | |
| Malawi | iLiNS−DYADM (30) | CDI | 66 | 147 | 14.3 | 0.53 (0.21, 1.35) | 0.11 | 0.15 | 138 | 275 | 9.1 | 1.04 (0.55, 1.96) | 0.25 | 0.25 |
| Malawi | iLiNS−DOSE (31) | CDI | 224 | 94 | 9.6 | 0.84 (0.39, 1.80) | 0.17 | 0.20 | 417 | 126 | 11.1 | 0.99 (0.56, 1.75) | 0.32 | 0.32 |
| Mali | PROMIS CS (32) | DMC | | | | | | | | | | | | |
| Zimbabwe | SHINE (HIV−) (33) | CDI | | | | | | | | | | | | |
| Zimbabwe | SHINE (HIV+) (34) | CDI | | | | | | | | | | | | |
| | | | **694** | **688** | | **I² = 0.34, Tau² = 0.05** | | | **963** | **765** | | **I² = 0.00, Tau² = 0.00** | | |
| **Fixed** | | | | | | **0.88 (0.64, 1.21)** | | | | | | **0.91 (0.66, 1.25)** | | |
| **Random** | | | | | | **0.87 (0.59, 1.29)** | | | | | | **0.91 (0.66, 1.25)** | | |

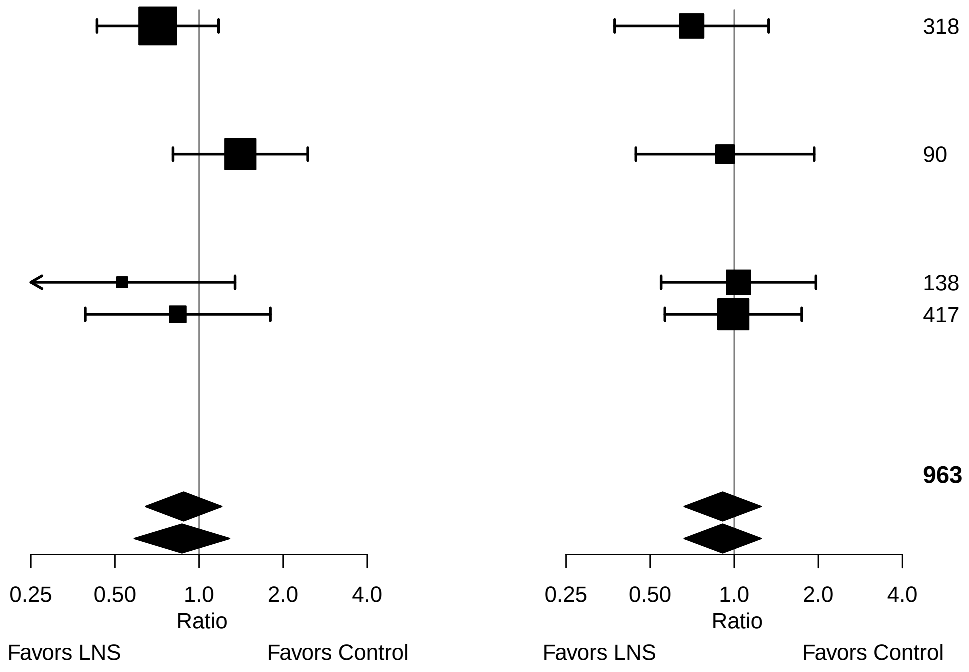

### Supplemental figure 5C: Language lowest decile prevalence difference

#### 5C1: Stratified by Maternal height

**P−for−interaction = 0.813**
**Difference in PDs = 0.00 (−0.02, 0.03)**

| Country | Trial | Tool | At least 150.1 cm: LNS N | Control N | Control Prevalence | PD (95% CI) | Fixed W | Random W | Less than 150.1 cm: LNS N | Control N | Control Prevalence | PD (95% CI) | Fixed W | Random W |
|---|---|---|---|---|---|---|---|---|---|---|---|---|---|---|
| Bangladesh | JiVitA−4 (21) | BSID−III | | | | | | | | | | | | |
| Bangladesh | RDNS (22) | CDI | 868 | 431 | 12.5 | −0.05 (−0.09, −0.02) | 0.14 | 0.23 | 735 | 358 | 12.0 | −0.01 (−0.05, 0.03) | 0.27 | 0.27 |
| Bangladesh | WASH−B (23) | CDI | 589 | 1812 | 10.2 | −0.02 (−0.04, 0.01) | 0.31 | 0.29 | 515 | 1499 | 10.8 | −0.02 (−0.05, 0.00) | 0.53 | 0.53 |
| Burkina Faso | iLiNS−Zinc (24) | DMC | | | | | | | | | | | | |
| Ghana | GHANA (25) | | | | | | | | | | | | | |
| Ghana | iLiNS−DYADG (26) | CDI | | | | | | | | | | | | |
| Haiti | HAITI (27) | Total sounds | | | | | | | | | | | | |
| Kenya | WASH−B (28) | EASQ | 1237 | 4372 | 9.7 | 0.00 (−0.02, 0.02) | 0.48 | 0.32 | 51 | 175 | 15.4 | −0.06 (−0.16, 0.04) | 0.05 | 0.05 |
| Madagascar | MAHAY (29) | ASQI | 956 | 986 | 8.7 | 0.02 (−0.04, 0.07) | 0.07 | 0.16 | 553 | 538 | 9.9 | 0.01 (−0.04, 0.07) | 0.16 | 0.16 |
| Malawi | iLiNS−DYADM (30) | CDI | | | | | | | | | | | | |
| Malawi | iLiNS−DOSE (31) | CDI | | | | | | | | | | | | |
| Mali | PROMIS CS (32) | DMC | | | | | | | | | | | | |
| Zimbabwe | SHINE (HIV−) (33) | CDI | | | | | | | | | | | | |
| Zimbabwe | SHINE (HIV+) (34) | CDI | | | | | | | | | | | | |
| | | | **3650** | **7601** | | **$I^2$ = 0.57, $Tau^2$ = 0.00** | | | **1854** | **2570** | | **$I^2$ = 0.00, $Tau^2$ = 0.00** | | |
| **Fixed** | | | | | | **−0.01 (−0.03, 0.00)** | | | | | | **−0.02 (−0.04, 0.01)** | | |
| **Random** | | | | | | **−0.02 (−0.04, 0.01)** | | | | | | **−0.02 (−0.04, 0.01)** | | |

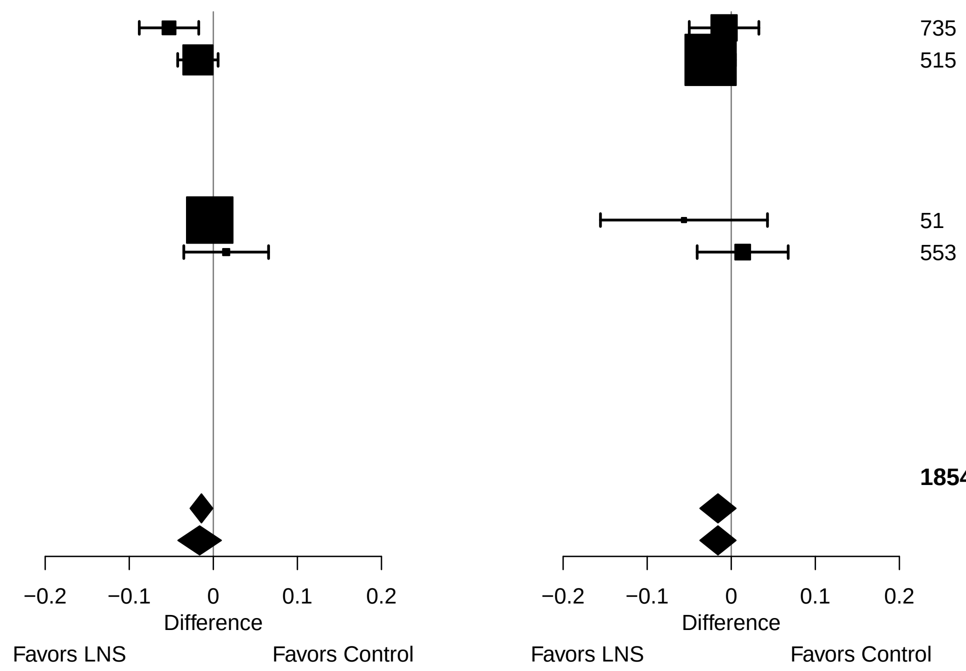

#### Supplemental figure 5C: Language lowest decile prevalence difference

##### 5C2: Stratified by Maternal BMI

**P−for−interaction = 0.303**
**Difference in PDs = 0.01 (−0.01, 0.03)**

| | | | At least 20 kg/m² | | | | | | Less than 20 kg/m² | | | | | |
|---|---|---|---|---|---|---|---|---|---|---|---|---|---|---|
| **Country** | **Trial** | **Tool** | **LNS N** | **Control N** | **Control Prevalence** | **PD (95% CI)** | **Fixed W** | **Random W** | **LNS N** | **Control N** | **Control Prevalence** | **PD (95% CI)** | **Fixed W** | **Random W** |
| Bangladesh | JiVitA−4 (21) | BSID−III | | | | | | | | | | | | |
| Bangladesh | RDNS (22) | CDI | 730 | 334 | 12.0 | −0.06 (−0.09, −0.02) | 0.12 | 0.15 | 873 | 455 | 12.5 | −0.01 (−0.05, 0.02) | 0.21 | 0.19 |
| Bangladesh | WASH−B (23) | CDI | 498 | 1529 | 9.5 | −0.04 (−0.06, −0.01) | 0.27 | 0.23 | 606 | 1781 | 11.2 | −0.01 (−0.04, 0.02) | 0.36 | 0.20 |
| Burkina Faso | iLiNS−Zinc (24) | DMC | 480 | 225 | 12.4 | −0.05 (−0.10, −0.01) | 0.07 | 0.10 | 264 | 150 | 20.7 | −0.13 (−0.19, −0.07) | 0.07 | 0.15 |
| Ghana | GHANA (25) | | | | | | | | | | | | | |
| Ghana | iLiNS−DYADG (26) | CDI | | | | | | | | | | | | |
| Haiti | HAITI (27) | Total sounds | | | | | | | | | | | | |
| Kenya | WASH−B (28) | EASQ | 1031 | 3535 | 10.0 | −0.01 (−0.03, 0.02) | 0.33 | 0.25 | 257 | 1012 | 9.8 | −0.01 (−0.05, 0.03) | 0.19 | 0.18 |
| Madagascar | MAHAY (29) | ASQI | | | | | | | | | | | | |
| Malawi | iLiNS−DYADM (30) | CDI | | | | | | | | | | | | |
| Malawi | iLiNS−DOSE (31) | CDI | | | | | | | | | | | | |
| Mali | PROMIS CS (32) | DMC | 683 | 664 | 11.9 | −0.04 (−0.08, 0.00) | 0.10 | 0.13 | 238 | 274 | 9.9 | 0.01 (−0.04, 0.05) | 0.12 | 0.17 |
| Zimbabwe | SHINE (HIV−) (33) | CDI | 581 | 569 | 10.9 | −0.01 (−0.04, 0.03) | 0.12 | 0.15 | 96 | 93 | 15.1 | −0.09 (−0.17, 0.00) | 0.04 | 0.12 |
| Zimbabwe | SHINE (HIV+) (34) | CDI | | | | | | | | | | | | |
| | | | **4003** | **6856** | | **I² = 0.48, Tau² = 0.00** | | | **2334** | **3765** | | **I² = 0.71, Tau² = 0.00** | | |
| **Fixed** | | | | | | **−0.03 (−0.04, −0.01)** | | | | | | **−0.02 (−0.04, 0.00)** | | |
| **Random** | | | | | | **−0.03 (−0.05, −0.01)** | | | | | | **−0.03 (−0.08, 0.01)** | | |

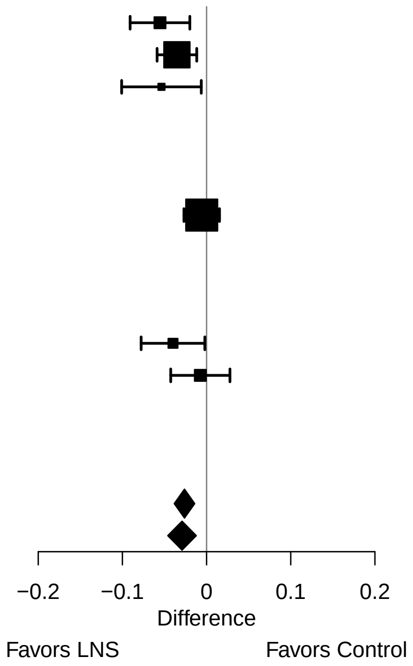

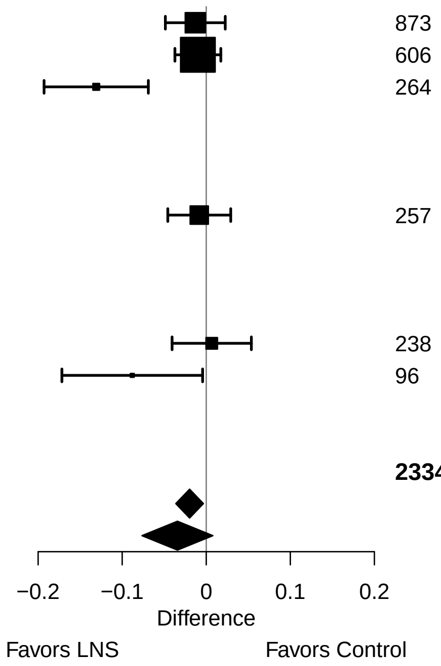

#### Supplemental figure 5C: Language lowest decile prevalence difference

#### 5C3: Stratified by Maternal age

**P−for−interaction = 0.820**
**Difference in PDs = 0.00 (−0.02, 0.02)**

| Country | Trial | Tool | At least 25 y: LNS N | Control N | Control Prevalence | PD (95% CI) | Fixed W | Random W | Less than 25 y: LNS N | Control N | Control Prevalence | PD (95% CI) | Fixed W | Random W |
|---|---|---|---|---|---|---|---|---|---|---|---|---|---|---|
| Bangladesh | JiVitA−4 (21) | BSID−III | 200 | 68 | 11.8 | −0.01 (−0.08, 0.06) | 0.03 | 0.03 | 242 | 74 | 10.8 | −0.02 (−0.10, 0.07) | 0.02 | 0.05 |
| Bangladesh | RDNS (22) | CDI | 453 | 220 | 13.6 | −0.02 (−0.07, 0.02) | 0.08 | 0.08 | 1210 | 594 | 12.0 | −0.04 (−0.07, −0.01) | 0.20 | 0.14 |
| Bangladesh | WASH−B (23) | CDI | 477 | 1475 | 11.5 | −0.01 (−0.04, 0.02) | 0.17 | 0.17 | 627 | 1863 | 9.8 | −0.03 (−0.05, 0.00) | 0.27 | 0.15 |
| Burkina Faso | iLiNS−Zinc (24) | DMC | 421 | 232 | 15.1 | −0.07 (−0.13, −0.02) | 0.06 | 0.06 | 321 | 141 | 17.0 | −0.10 (−0.18, −0.03) | 0.03 | 0.05 |
| Ghana | GHANA (25) | | | | | | | | | | | | | |
| Ghana | iLiNS−DYADG (26) | CDI | 211 | 405 | 10.4 | −0.02 (−0.07, 0.03) | 0.07 | 0.07 | 120 | 253 | 8.7 | 0.05 (−0.01, 0.12) | 0.03 | 0.06 |
| Haiti | HAITI (27) | Total sounds | | | | | | | | | | | | |
| Kenya | WASH−B (28) | EASQ | 798 | 2622 | 9.9 | 0.00 (−0.03, 0.03) | 0.20 | 0.20 | 549 | 2092 | 10.3 | −0.01 (−0.04, 0.02) | 0.19 | 0.14 |
| Madagascar | MAHAY (29) | ASQI | 852 | 901 | 9.8 | 0.01 (−0.03, 0.06) | 0.07 | 0.07 | 759 | 703 | 8.5 | 0.02 (−0.03, 0.07) | 0.06 | 0.09 |
| Malawi | iLiNS−DYADM (30) | CDI | 103 | 225 | 14.7 | −0.05 (−0.13, 0.03) | 0.03 | 0.03 | 112 | 214 | 6.5 | 0.02 (−0.04, 0.08) | 0.04 | 0.07 |
| Malawi | iLiNS−DOSE (31) | CDI | 345 | 124 | 11.3 | −0.01 (−0.07, 0.05) | 0.04 | 0.04 | 288 | 90 | 8.9 | 0.00 (−0.07, 0.07) | 0.03 | 0.06 |
| Mali | PROMIS CS (32) | DMC | 492 | 492 | 11.0 | −0.04 (−0.08, 0.00) | 0.12 | 0.12 | 435 | 452 | 11.7 | −0.01 (−0.06, 0.03) | 0.08 | 0.10 |
| Zimbabwe | SHINE (HIV−) (33) | CDI | 414 | 384 | 9.9 | −0.02 (−0.06, 0.01) | 0.12 | 0.12 | 315 | 310 | 12.9 | −0.01 (−0.06, 0.05) | 0.05 | 0.08 |
| Zimbabwe | SHINE (HIV+) (34) | CDI | | | | | | | | | | | | |
| | | | **4766** | **7148** | | **I² = 0.00, Tau² = 0.00** | | | **4978** | **6786** | | **I² = 0.44, Tau² = 0.00** | | |
| **Fixed** | | | | | | **−0.02 (−0.03, 0.00)** | | | | | | **−0.02 (−0.03, −0.01)** | | |
| **Random** | | | | | | **−0.02 (−0.03, 0.00)** | | | | | | **−0.01 (−0.03, 0.01)** | | |

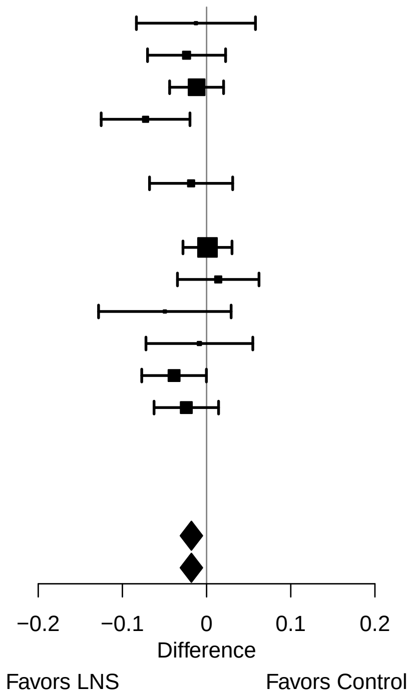

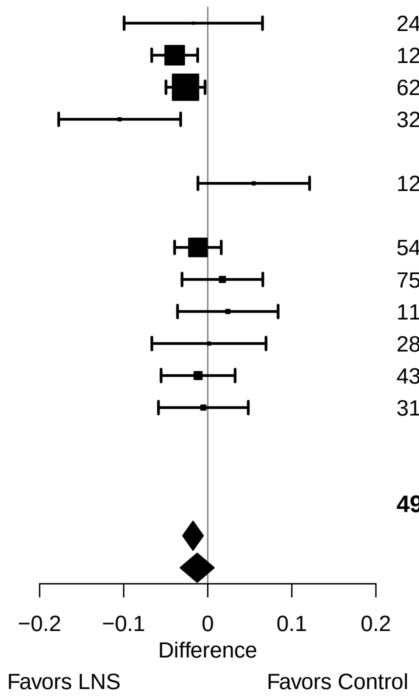

**Supplemental figure 5C: Language lowest decile prevalence difference**

#### 5C4: Stratified by Maternal education

**P−for−interaction = 0.116**
**Difference in PDs = −0.02 (−0.04, 0.00)**

| Country | Trial | Tool | LNS N | Control N | Control Prevalence | Primary or greater PD (95% CI) | Fixed W | Random W | LNS N | Control N | Control Prevalence | Incomplete or no formal PD (95% CI) | Fixed W | Random W |
|---|---|---|---|---|---|---|---|---|---|---|---|---|---|---|
| Bangladesh | JiVitA−4 (21) | BSID−III | 292 | 92 | 8.7 | −0.01 (−0.08, 0.06) | 0.02 | 0.07 | 152 | 51 | 15.7 | −0.02 (−0.13, 0.09) | 0.02 | 0.02 |
| Bangladesh | RDNS (22) | CDI | 1245 | 588 | 9.9 | −0.03 (−0.06, −0.01) | 0.20 | 0.20 | 418 | 226 | 19.0 | −0.03 (−0.10, 0.03) | 0.07 | 0.07 |
| Bangladesh | WASH−B (23) | CDI | 781 | 2406 | 8.9 | −0.01 (−0.03, 0.01) | 0.35 | 0.23 | 328 | 947 | 14.8 | −0.04 (−0.08, 0.00) | 0.18 | 0.18 |
| Burkina Faso | iLiNS−Zinc (24) | DMC | | | | | | | | | | | | |
| Ghana | GHANA (25) | | | | | | | | | | | | | |
| Ghana | iLiNS−DYADG (26) | CDI | 256 | 519 | 9.4 | 0.00 (−0.04, 0.04) | 0.06 | 0.12 | 75 | 139 | 10.8 | 0.04 (−0.05, 0.13) | 0.03 | 0.03 |
| Haiti | HAITI (27) | Total sounds | | | | | | | | | | | | |
| Kenya | WASH−B (28) | EASQ | 658 | 2258 | 6.9 | −0.01 (−0.03, 0.01) | 0.31 | 0.23 | 702 | 2484 | 13.0 | 0.00 (−0.03, 0.03) | 0.32 | 0.32 |
| Madagascar | MAHAY (29) | ASQI | 339 | 416 | 3.6 | 0.06 (0.01, 0.11) | 0.05 | 0.11 | 1274 | 1188 | 11.2 | 0.00 (−0.05, 0.05) | 0.11 | 0.11 |
| Malawi | iLiNS−DYADM (30) | CDI | | | | | | | | | | | | |
| Malawi | iLiNS−DOSE (31) | CDI | | | | | | | | | | | | |
| Mali | PROMIS CS (32) | DMC | 100 | 94 | 7.4 | 0.02 (−0.09, 0.12) | 0.01 | 0.03 | 826 | 850 | 11.8 | −0.03 (−0.06, 0.00) | 0.25 | 0.25 |
| Zimbabwe | SHINE (HIV−) (33) | CDI | | | | | | | | | | | | |
| Zimbabwe | SHINE (HIV+) (34) | CDI | | | | | | | | | | | | |
| | | | **3671** | **6373** | | **I² = 0.50, Tau² = 0.00** | | | **3775** | **5885** | | **I² = 0.00, Tau² = 0.00** | | |
| **Fixed** | | | | | | **−0.01 (−0.02, 0.00)** | | | | | | **−0.02 (−0.03, 0.00)** | | |
| **Random** | | | | | | **−0.01 (−0.03, 0.01)** | | | | | | **−0.02 (−0.03, 0.00)** | | |

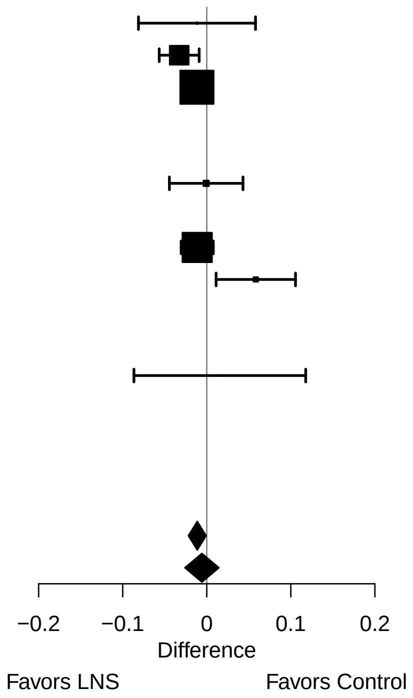

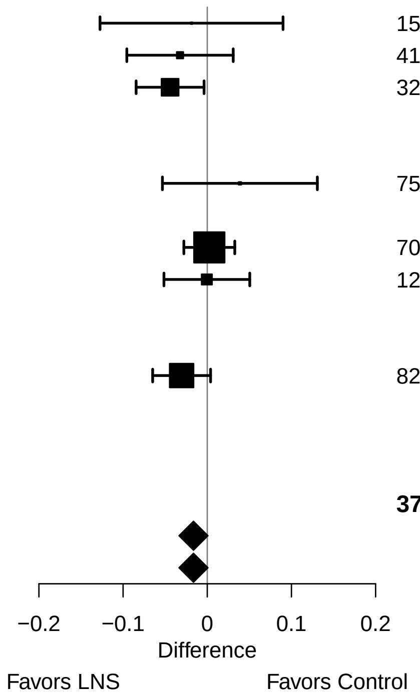

#### Supplemental figure 5C: Language lowest decile prevalence difference

##### 5C5: Stratified by Maternal depressive symptoms

**−for−interaction = 0.461**
**ifference in PDs = −0.01 (−0.03, 0.01)**

| ountry | Trial | Tool | LNS N | Control N | Control Prevalence | Less than 75th percentile PD (95% CI) | Fixed W | Random W | LNS N | Control N | Control Prevalence | At least 75th percentile PD (95% CI) | Fixed W | Random W |
|---|---|---|---|---|---|---|---|---|---|---|---|---|---|---|
| angladesh | JiVitA−4 (21) | BSID−III | | | | | | | | | | | | |
| angladesh | RDNS (22) | CDI | 1081 | 471 | 11.9 | −0.04 (−0.08, −0.01) | 0.11 | 0.11 | 520 | 288 | 13.5 | −0.02 (−0.06, 0.02) | 0.21 | 0.21 |
| angladesh | WASH−B (23) | CDI | 856 | 2410 | 10.2 | −0.03 (−0.05, −0.01) | 0.31 | 0.31 | 232 | 859 | 11.3 | 0.02 (−0.03, 0.07) | 0.15 | 0.15 |
| urkina Faso | iLiNS−Zinc (24) | DMC | | | | | | | | | | | | |
| hana | GHANA (25) | | | | | | | | | | | | | |
| hana | iLiNS−DYADG (26) | CDI | 241 | 435 | 10.8 | 0.00 (−0.05, 0.05) | 0.06 | 0.06 | 78 | 202 | 7.4 | 0.03 (−0.04, 0.10) | 0.07 | 0.07 |
| aiti | HAITI (27) | Total sounds | | | | | | | | | | | | |
| enya | WASH−B (28) | EASQ | 960 | 3317 | 9.5 | −0.01 (−0.03, 0.01) | 0.35 | 0.35 | 317 | 1156 | 8.7 | 0.00 (−0.03, 0.04) | 0.36 | 0.36 |
| ladagascar | MAHAY (29) | ASQI | 615 | 667 | 8.8 | 0.03 (−0.03, 0.08) | 0.04 | 0.04 | 268 | 242 | 12.0 | −0.03 (−0.10, 0.04) | 0.08 | 0.08 |
| lalawi | iLiNS−DYADM (30) | CDI | 153 | 302 | 9.9 | −0.01 (−0.07, 0.05) | 0.04 | 0.04 | 47 | 112 | 12.5 | −0.02 (−0.13, 0.09) | 0.03 | 0.03 |
| lalawi | iLiNS−DOSE (31) | CDI | | | | | | | | | | | | |
| lali | PROMIS CS (32) | DMC | | | | | | | | | | | | |
| imbabwe | SHINE (HIV−) (33) | CDI | 566 | 552 | 10.3 | −0.01 (−0.05, 0.03) | 0.10 | 0.10 | 190 | 173 | 14.5 | −0.04 (−0.11, 0.02) | 0.10 | 0.10 |
| imbabwe | SHINE (HIV+) (34) | CDI | | | | | | | | | | | | |
| | | | **4472** | **8154** | | **I² = 0.12, Tau² = 0.00** | | | **1652** | **3032** | | **I² = 0.00, Tau² = 0.00** | | |
| **ixed** | | | | | | **−0.02 (−0.03, −0.01)** | | | | | | **0.00 (−0.02, 0.02)** | | |
| **andom** | | | | | | **−0.02 (−0.03, −0.01)** | | | | | | **0.00 (−0.02, 0.02)** | | |

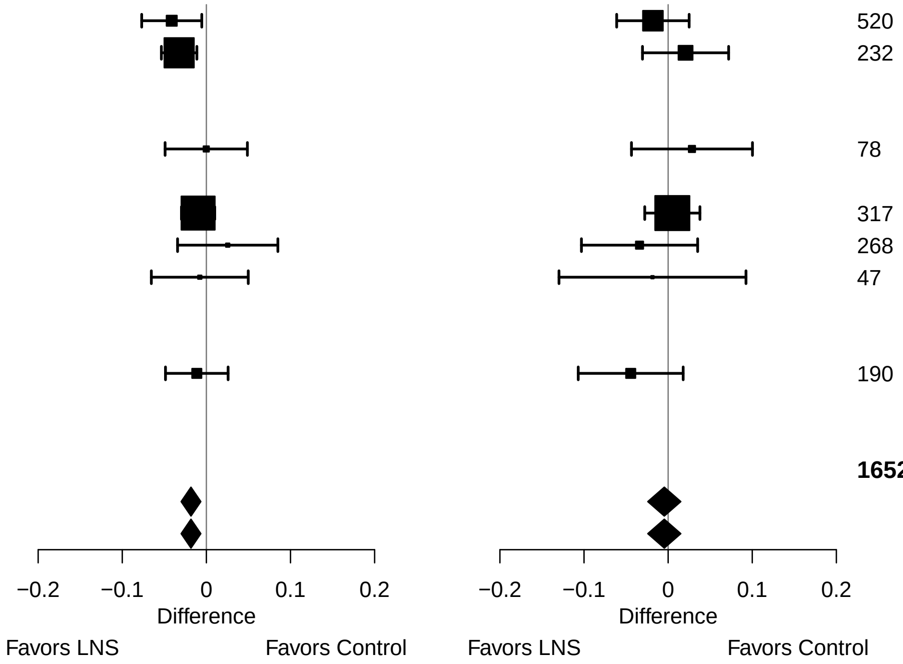

#### Supplemental figure 5C: Language lowest decile prevalence difference

#### 5C6: Stratified by Child sex

**P−for−interaction = 0.199**

**Difference in PDs = −0.01 (−0.03, 0.01)**

| Country | Trial | Tool | LNS N | Control N | Control Prevalence | Male PD (95% CI) | Fixed W | Random W | LNS N | Control N | Control Prevalence | Female PD (95% CI) | Fixed W | Random W |
|---|---|---|---|---|---|---|---|---|---|---|---|---|---|---|
| Bangladesh | JiVitA−4 (21) | BSID−III | 231 | 70 | 14.3 | −0.05 (−0.15, 0.05) | 0.01 | 0.02 | 214 | 73 | 8.2 | 0.02 (−0.04, 0.08) | 0.05 | 0.07 |
| Bangladesh | RDNS (22) | CDI | 833 | 411 | 10.5 | 0.00 (−0.03, 0.04) | 0.11 | 0.11 | 830 | 403 | 14.4 | −0.07 (−0.11, −0.04) | 0.12 | 0.11 |
| Bangladesh | WASH−B (23) | CDI | 545 | 1685 | 11.5 | −0.02 (−0.05, 0.00) | 0.19 | 0.15 | 564 | 1668 | 9.6 | −0.02 (−0.04, 0.01) | 0.22 | 0.13 |
| Burkina Faso | iLiNS−Zinc (24) | DMC | 372 | 193 | 12.4 | −0.07 (−0.10, −0.03) | 0.11 | 0.11 | 374 | 182 | 19.2 | −0.11 (−0.16, −0.05) | 0.05 | 0.08 |
| Ghana | GHANA (25) | | | | | | | | | | | | | |
| Ghana | iLiNS−DYADG (26) | CDI | 164 | 306 | 8.8 | 0.04 (−0.02, 0.10) | 0.04 | 0.06 | 167 | 352 | 10.5 | −0.02 (−0.08, 0.03) | 0.05 | 0.07 |
| Haiti | HAITI (27) | Total sounds | | | | | | | | | | | | |
| Kenya | WASH−B (28) | EASQ | 682 | 2260 | 10.7 | 0.00 (−0.02, 0.03) | 0.19 | 0.15 | 680 | 2485 | 9.6 | −0.01 (−0.04, 0.01) | 0.22 | 0.13 |
| Madagascar | MAHAY (29) | ASQI | 795 | 787 | 8.9 | 0.01 (−0.03, 0.06) | 0.07 | 0.08 | 818 | 817 | 9.5 | 0.02 (−0.04, 0.07) | 0.05 | 0.07 |
| Malawi | iLiNS−DYADM (30) | CDI | 105 | 205 | 8.8 | −0.02 (−0.09, 0.04) | 0.03 | 0.05 | 110 | 234 | 12.4 | −0.01 (−0.08, 0.07) | 0.03 | 0.05 |
| Malawi | iLiNS−DOSE (31) | CDI | 320 | 117 | 11.1 | −0.01 (−0.08, 0.05) | 0.04 | 0.05 | 325 | 104 | 9.6 | 0.01 (−0.06, 0.07) | 0.03 | 0.06 |
| Mali | PROMIS CS (32) | DMC | 469 | 514 | 10.7 | −0.03 (−0.06, 0.01) | 0.11 | 0.11 | 458 | 430 | 12.1 | −0.03 (−0.07, 0.02) | 0.08 | 0.10 |
| Zimbabwe | SHINE (HIV−) (33) | CDI | 412 | 378 | 11.6 | −0.01 (−0.05, 0.03) | 0.07 | 0.08 | 405 | 403 | 9.9 | −0.01 (−0.06, 0.03) | 0.07 | 0.09 |
| Zimbabwe | SHINE (HIV+) (34) | CDI | 82 | 76 | 13.2 | −0.03 (−0.14, 0.07) | 0.01 | 0.02 | 83 | 71 | 11.3 | −0.04 (−0.13, 0.05) | 0.02 | 0.04 |
| | | | **5010** | **7002** | | **I² = 0.30, Tau² = 0.00** | | | **5028** | **7222** | | **I² = 0.52, Tau² = 0.00** | | |
| **Fixed** | | | | | | **−0.01 (−0.03, 0.00)** | | | | | | **−0.02 (−0.04, −0.01)** | | |
| **Random** | | | | | | **−0.01 (−0.03, 0.00)** | | | | | | **−0.02 (−0.04, 0.00)** | | |

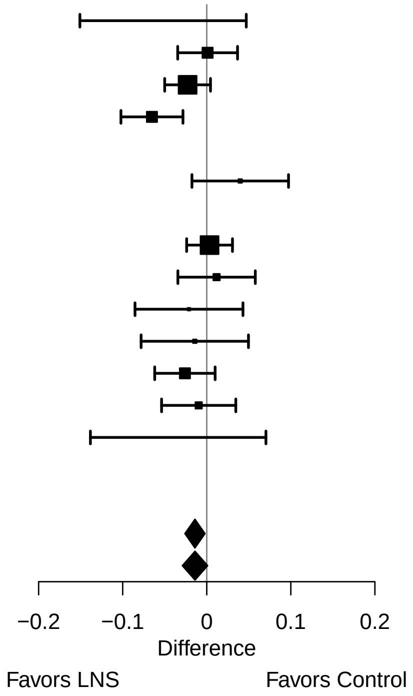

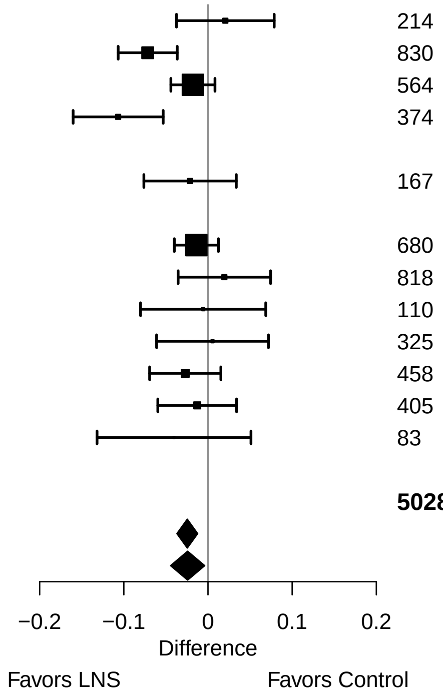

**Supplemental figure 5C: Language lowest decile prevalence difference**

#### 5C7: Stratified by Child birth order

**P−for−interaction = 0.300**
**Difference in PDs = 0.01 (−0.01, 0.03)**

| Country | Trial | Tool | Later born LNS N | Later born Control N | Later born Control Prevalence | Later born PD (95% CI) | Later born Fixed W | Later born Random W | Firstborn LNS N | Firstborn Control N | Firstborn Control Prevalence | Firstborn PD (95% CI) | Firstborn Fixed W | Firstborn Random W |
|---|---|---|---|---|---|---|---|---|---|---|---|---|---|---|
| Bangladesh | JiVitA−4 (21) | BSID−III | | | | | | | | | | | | |
| Bangladesh | RDNS (22) | CDI | 974 | 507 | 14.0 | −0.04 (−0.08, −0.01) | 0.11 | 0.12 | 688 | 307 | 9.8 | −0.02 (−0.06, 0.02) | 0.18 | 0.16 |
| Bangladesh | WASH−B (23) | CDI | 693 | 2187 | 11.6 | −0.03 (−0.05, 0.00) | 0.20 | 0.16 | 395 | 1060 | 8.3 | −0.01 (−0.04, 0.02) | 0.35 | 0.19 |
| Burkina Faso | iLiNS−Zinc (24) | DMC | 587 | 290 | 15.5 | −0.08 (−0.13, −0.03) | 0.05 | 0.08 | 159 | 85 | 16.5 | −0.10 (−0.18, −0.01) | 0.04 | 0.07 |
| Ghana | GHANA (25) | | | | | | | | | | | | | |
| Ghana | iLiNS−DYADG (26) | CDI | 224 | 445 | 10.1 | −0.01 (−0.06, 0.04) | 0.06 | 0.08 | 107 | 213 | 8.9 | 0.05 (−0.02, 0.12) | 0.06 | 0.09 |
| Haiti | HAITI (27) | Total sounds | | | | | | | | | | | | |
| Kenya | WASH−B (28) | EASQ | 1071 | 3767 | 9.8 | 0.00 (−0.03, 0.02) | 0.25 | 0.17 | 289 | 974 | 11.4 | −0.01 (−0.06, 0.03) | 0.14 | 0.14 |
| Madagascar | MAHAY (29) | ASQI | 1162 | 1118 | 9.1 | 0.02 (−0.03, 0.06) | 0.06 | 0.09 | 433 | 465 | 9.7 | 0.01 (−0.05, 0.06) | 0.09 | 0.11 |
| Malawi | iLiNS−DYADM (30) | CDI | 170 | 350 | 11.7 | −0.04 (−0.10, 0.02) | 0.04 | 0.07 | 45 | 87 | 6.9 | 0.09 (−0.02, 0.19) | 0.02 | 0.05 |
| Malawi | iLiNS−DOSE (31) | CDI | | | | | | | | | | | | |
| Mali | PROMIS CS (32) | DMC | 770 | 811 | 11.2 | −0.03 (−0.06, 0.01) | 0.12 | 0.13 | 142 | 122 | 9.0 | 0.01 (−0.06, 0.08) | 0.06 | 0.09 |
| Zimbabwe | SHINE (HIV−) (33) | CDI | 587 | 562 | 9.4 | −0.01 (−0.04, 0.03) | 0.09 | 0.11 | 194 | 185 | 15.7 | −0.03 (−0.10, 0.04) | 0.06 | 0.09 |
| Zimbabwe | SHINE (HIV+) (34) | CDI | | | | | | | | | | | | |
| | | | **6238** | **10037** | | **I² = 0.45, Tau² = 0.00** | | | **2452** | **3498** | | **I² = 0.30, Tau² = 0.00** | | |
| **Fixed** | | | | | | **−0.02 (−0.03, −0.01)** | | | | | | **−0.01 (−0.02, 0.01)** | | |
| **Random** | | | | | | **−0.02 (−0.04, −0.01)** | | | | | | **−0.01 (−0.03, 0.02)** | | |

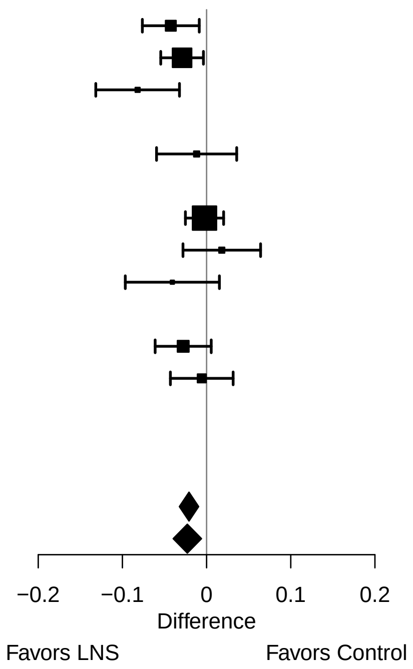

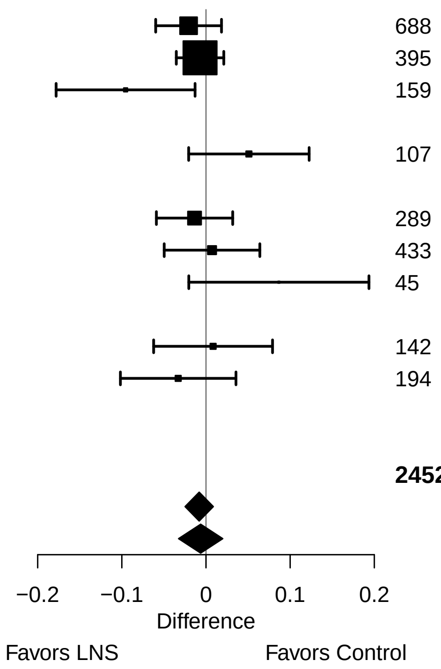

#### Supplemental figure 5C: Language lowest decile prevalence difference

##### 5C8: Stratified by Child baseline stunting

**P−for−interaction = 0.021**
**Difference in PDs = −0.05 (−0.09, −0.01)**

| | | | No | | | | | | Yes | | | | | |
|---|---|---|---|---|---|---|---|---|---|---|---|---|---|---|
| **Country** | **Trial** | **Tool** | **LNS N** | **Control N** | **Control Prevalence** | **PD (95% CI)** | **Fixed W** | **Random W** | **LNS N** | **Control N** | **Control Prevalence** | **PD (95% CI)** | **Fixed W** | **Random W** |
| Bangladesh | JiVitA−4 (21) | BSID−III | 327 | 114 | 8.8 | −0.01 (−0.07, 0.06) | 0.06 | 0.07 | 116 | 29 | 20.7 | −0.07 (−0.18, 0.05) | 0.10 | 0.13 |
| Bangladesh | RDNS (22) | CDI | 1246 | 573 | 10.6 | −0.03 (−0.05, 0.00) | 0.38 | 0.32 | 355 | 185 | 18.4 | −0.06 (−0.12, 0.01) | 0.35 | 0.27 |
| Bangladesh | WASH−B (23) | CDI | | | | | | | | | | | | |
| Burkina Faso | iLiNS−Zinc (24) | DMC | 576 | 294 | 12.6 | −0.06 (−0.09, −0.02) | 0.20 | 0.20 | 170 | 80 | 27.5 | −0.19 (−0.29, −0.10) | 0.15 | 0.16 |
| Ghana | GHANA (25) | | | | | | | | | | | | | |
| Ghana | iLiNS−DYADG (26) | CDI | | | | | | | | | | | | |
| Haiti | HAITI (27) | Total sounds | | | | | | | | | | | | |
| Kenya | WASH−B (28) | EASQ | | | | | | | | | | | | |
| Madagascar | MAHAY (29) | ASQI | | | | | | | | | | | | |
| Malawi | iLiNS−DYADM (30) | CDI | 153 | 323 | 8.4 | 0.00 (−0.05, 0.05) | 0.08 | 0.10 | 48 | 97 | 19.6 | −0.07 (−0.20, 0.06) | 0.08 | 0.10 |
| Malawi | iLiNS−DOSE (31) | CDI | 464 | 147 | 8.2 | 0.01 (−0.04, 0.06) | 0.09 | 0.10 | 181 | 74 | 14.9 | −0.03 (−0.12, 0.06) | 0.17 | 0.18 |
| Mali | PROMIS CS (32) | DMC | | | | | | | | | | | | |
| Zimbabwe | SHINE (HIV−) (33) | CDI | 523 | 482 | 10.0 | −0.02 (−0.06, 0.01) | 0.19 | 0.20 | 119 | 100 | 16.0 | −0.04 (−0.14, 0.05) | 0.15 | 0.17 |
| Zimbabwe | SHINE (HIV+) (34) | CDI | | | | | | | | | | | | |
| | | | **3289** | **1933** | | **I² = 0.15, Tau² = 0.00** | | | **989** | **565** | | **I² = 0.34, Tau² = 0.00** | | |
| **Fixed** | | | | | | **−0.03 (−0.04, −0.01)** | | | | | | **−0.07 (−0.11, −0.04)** | | |
| **Random** | | | | | | **−0.02 (−0.04, −0.01)** | | | | | | **−0.07 (−0.12, −0.03)** | | |

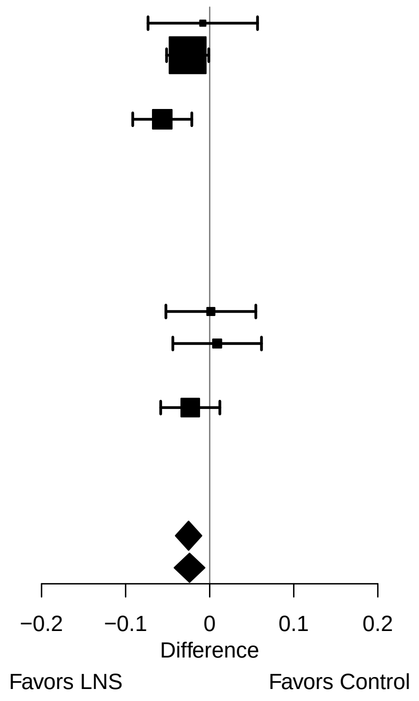

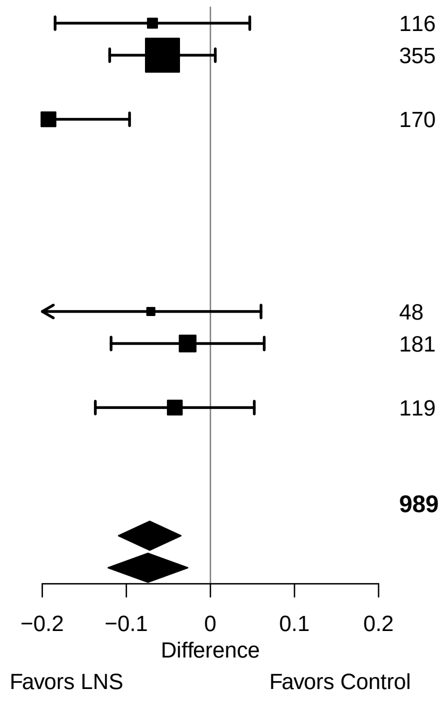

#### Supplemental figure 5C: Language lowest decile prevalence difference

#### 5C9: Stratified by Child baseline acute malnutrition

**P−for−interaction = 0.660**
**Difference in PDs = −0.02 (−0.09, 0.06)**

| Country | Trial | Tool | No: LNS N | No: Control N | No: Control Prevalence | No: PD (95% CI) | No: Fixed W | No: Random W | Yes: LNS N | Yes: Control N | Yes: Control Prevalence | Yes: PD (95% CI) | Yes: Fixed W | Yes: Random W |
|---|---|---|---|---|---|---|---|---|---|---|---|---|---|---|
| Bangladesh | JiVitA−4 (21) | BSID−III | 359 | 112 | 9.8 | −0.01 (−0.08, 0.06) | 0.08 | 0.18 | 84 | 31 | 16.1 | −0.03 (−0.17, 0.11) | 0.23 | 0.23 |
| Bangladesh | RDNS (22) | CDI | 1477 | 706 | 11.9 | −0.03 (−0.06, −0.01) | 0.66 | 0.47 | 124 | 53 | 20.8 | −0.05 (−0.16, 0.07) | 0.33 | 0.33 |
| Bangladesh | WASH−B (23) | CDI | | | | | | | | | | | | |
| Burkina Faso | iLiNS−Zinc (24) | DMC | 556 | 266 | 13.5 | −0.08 (−0.12, −0.04) | 0.26 | 0.35 | 190 | 109 | 21.1 | −0.10 (−0.19, 0.00) | 0.44 | 0.44 |
| Ghana | GHANA (25) | | | | | | | | | | | | | |
| Ghana | iLiNS−DYADG (26) | CDI | | | | | | | | | | | | |
| Haiti | HAITI (27) | Total sounds | | | | | | | | | | | | |
| Kenya | WASH−B (28) | EASQ | | | | | | | | | | | | |
| Madagascar | MAHAY (29) | ASQI | | | | | | | | | | | | |
| Malawi | iLiNS−DYADM (30) | CDI | | | | | | | | | | | | |
| Malawi | iLiNS−DOSE (31) | CDI | | | | | | | | | | | | |
| Mali | PROMIS CS (32) | DMC | | | | | | | | | | | | |
| Zimbabwe | SHINE (HIV−) (33) | CDI | | | | | | | | | | | | |
| Zimbabwe | SHINE (HIV+) (34) | CDI | | | | | | | | | | | | |
| | | | **2392** | **1084** | | **I² = 0.56, Tau² = 0.00** | | | **398** | **193** | | **I² = 0.00, Tau² = 0.00** | | |
| **Fixed** | | | | | | **−0.04 (−0.06, −0.02)** | | | | | | **−0.06 (−0.13, 0.00)** | | |
| **Random** | | | | | | **−0.05 (−0.08, −0.01)** | | | | | | **−0.06 (−0.13, 0.00)** | | |

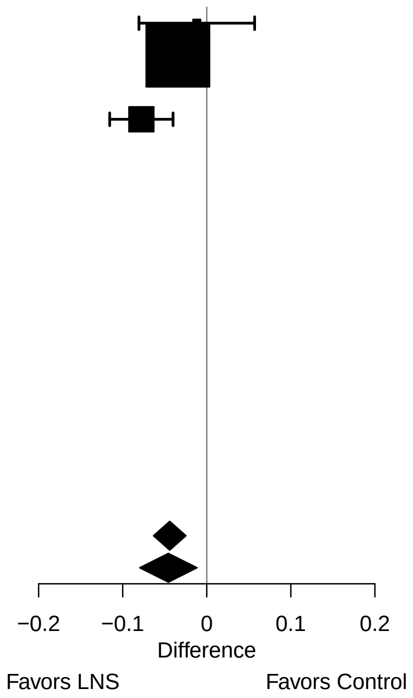

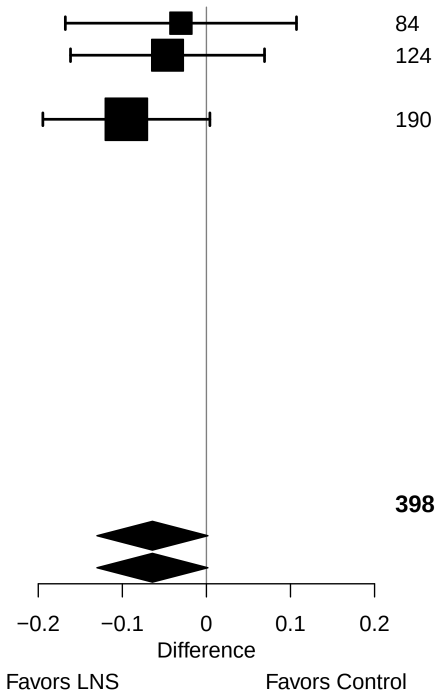

#### Supplemental figure 5C: Language lowest decile prevalence difference

##### 5C10: Stratified by Child baseline anemia

**P−for−interaction = 0.698**
**Difference in PDs = 0.01 (−0.04, 0.06)**

| Country | Trial | Tool | LNS N | Control N | Control Prevalence | Not anemic PD (95% CI) | Fixed W | Random W | LNS N | Control N | Control Prevalence | Anemic PD (95% CI) | Fixed W | Random W |
|---|---|---|---|---|---|---|---|---|---|---|---|---|---|---|
| Bangladesh | JiVitA−4 (21) | BSID−III | | | | | | | | | | | | |
| Bangladesh | RDNS (22) | CDI | 212 | 96 | 14.6 | −0.04 (−0.11, 0.02) | 0.23 | 0.25 | 318 | 160 | 12.5 | −0.04 (−0.11, 0.04) | 0.21 | 0.21 |
| Bangladesh | WASH−B (23) | CDI | | | | | | | | | | | | |
| Burkina Faso | iLiNS−Zinc (24) | DMC | | | | | | | | | | | | |
| Ghana | GHANA (25) | | | | | | | | | | | | | |
| Ghana | iLiNS−DYADG (26) | CDI | 192 | 351 | 7.4 | 0.03 (−0.02, 0.08) | 0.43 | 0.36 | 90 | 204 | 10.8 | −0.01 (−0.08, 0.07) | 0.19 | 0.19 |
| Haiti | HAITI (27) | Total sounds | | | | | | | | | | | | |
| Kenya | WASH−B (28) | EASQ | | | | | | | | | | | | |
| Madagascar | MAHAY (29) | ASQI | | | | | | | | | | | | |
| Malawi | iLiNS−DYADM (30) | CDI | 66 | 147 | 14.3 | −0.07 (−0.16, 0.03) | 0.11 | 0.15 | 138 | 275 | 9.1 | 0.00 (−0.06, 0.06) | 0.32 | 0.32 |
| Malawi | iLiNS−DOSE (31) | CDI | 224 | 94 | 9.6 | −0.02 (−0.08, 0.05) | 0.23 | 0.25 | 417 | 126 | 11.1 | 0.00 (−0.06, 0.06) | 0.28 | 0.28 |
| Mali | PROMIS CS (32) | DMC | | | | | | | | | | | | |
| Zimbabwe | SHINE (HIV−) (33) | CDI | | | | | | | | | | | | |
| Zimbabwe | SHINE (HIV+) (34) | CDI | | | | | | | | | | | | |
| | | | **694** | **688** | | **I² = 0.38, Tau² = 0.00** | | | **963** | **765** | | **I² = 0.00, Tau² = 0.00** | | |
| **Fixed** | | | | | | **−0.01 (−0.04, 0.02)** | | | | | | **−0.01 (−0.04, 0.02)** | | |
| **Random** | | | | | | **−0.01 (−0.05, 0.03)** | | | | | | **−0.01 (−0.04, 0.02)** | | |

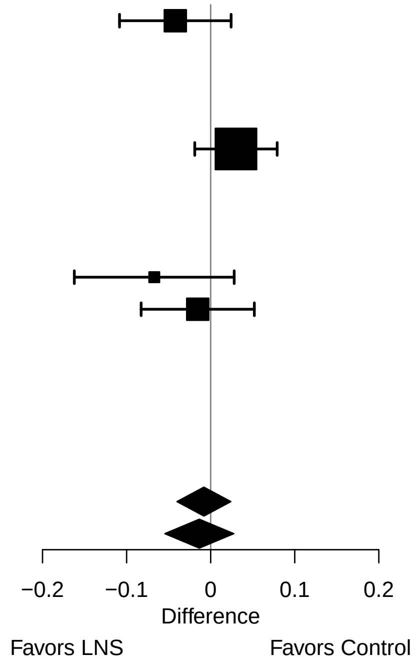

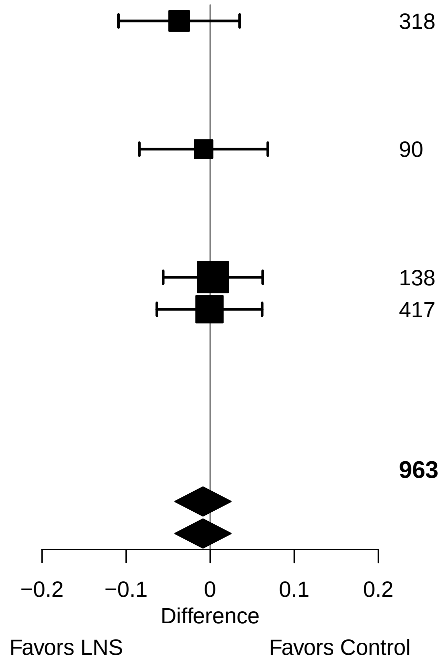

### Supplemental figure 5D: Mean difference in social-emotional z-score

#### 5D1: Stratified by Maternal height

**P−for−interaction = 0.511**
**Difference in MDs = 0.03 (−0.05, 0.10)**

| | | | At least 150.1 cm | | | | | | | Less than 150.1 cm | | | | |
|---|---|---|---|---|---|---|---|---|---|---|---|---|---|---|
| **Country** | **Trial** | **Tool** | **LNS N** | **Control N** | **Control Mean** | **MD (95% CI)** | **Fixed W** | **Random W** | **LNS N** | **Control N** | **Control Mean** | **MD (95% CI)** | **Fixed W** | **Random W** |
| Bangladesh | JiVitA−4 (21) | | | | | | | | | | | | | |
| Bangladesh | RDNS (22) | DMC | 865 | 432 | 0.00 | 0.09 (−0.05, 0.24) | 0.07 | 0.09 | 732 | 358 | −0.08 | 0.04 (−0.06, 0.14) | 0.41 | 0.24 |
| Bangladesh | WASH−B (23) | EASQ | 567 | 1768 | 0.03 | 0.11 (0.01, 0.20) | 0.15 | 0.11 | 494 | 1458 | −0.10 | 0.14 (0.03, 0.24) | 0.36 | 0.24 |
| Burkina Faso | iLiNS−Zinc (24) | DMC | 727 | 365 | −0.23 | 0.36 (0.18, 0.53) | 0.04 | 0.08 | 17 | 10 | −0.30 | 0.32 (−0.34, 0.98) | 0.01 | 0.03 |
| Ghana | GHANA (25) | | | | | | | | | | | | | |
| Ghana | iLiNS−DYADG (26) | PSED | 311 | 611 | 0.00 | 0.00 (−0.14, 0.14) | 0.07 | 0.09 | 16 | 35 | −0.11 | −0.04 (−0.61, 0.54) | 0.01 | 0.04 |
| Haiti | HAITI (27) | | | | | | | | | | | | | |
| Kenya | WASH−B (28) | EASQ | 1237 | 4372 | 0.01 | −0.01 (−0.08, 0.05) | 0.33 | 0.13 | 51 | 175 | −0.15 | 0.49 (0.18, 0.80) | 0.04 | 0.09 |
| Madagascar | MAHAY (29) | ASQI | 956 | 986 | 0.03 | −0.05 (−0.24, 0.15) | 0.03 | 0.07 | 553 | 538 | 0.10 | −0.13 (−0.34, 0.07) | 0.09 | 0.15 |
| Malawi | iLiNS−DYADM (30) | PSED | 187 | 377 | 0.01 | −0.01 (−0.19, 0.16) | 0.04 | 0.08 | 28 | 58 | −0.14 | 0.22 (−0.22, 0.66) | 0.02 | 0.06 |
| Malawi | iLiNS−DOSE (31) | PSED | 541 | 185 | 0.03 | 0.01 (−0.16, 0.17) | 0.05 | 0.08 | 102 | 35 | −0.25 | 0.11 (−0.29, 0.51) | 0.02 | 0.07 |
| Mali | PROMIS CS (32) | DMC | 889 | 911 | −0.06 | 0.14 (−0.02, 0.31) | 0.05 | 0.08 | 32 | 27 | −0.27 | 0.39 (−0.15, 0.92) | 0.01 | 0.04 |
| Zimbabwe | SHINE (HIV−) (33) | MDAT | 788 | 762 | −0.07 | 0.13 (0.03, 0.24) | 0.12 | 0.11 | 34 | 22 | −0.04 | 0.10 (−0.47, 0.67) | 0.01 | 0.04 |
| Zimbabwe | SHINE (HIV+) (34) | MDAT | 158 | 140 | −0.15 | 0.32 (0.12, 0.51) | 0.04 | 0.07 | 10 | 6 | −0.30 | 0.70 (−0.18, 1.59) | 0.00 | 0.02 |
| | | | **7226** | **10909** | | **I² = 0.65, Tau² = 0.01** | | | **2069** | **2722** | | **I² = 0.40, Tau² = 0.01** | | |
| **Fixed** | | | | | | **0.07 (0.03, 0.10)** | | | | | | **0.09 (0.03, 0.16)** | | |
| **Random** | | | | | | **0.09 (0.02, 0.17)** | | | | | | **0.13 (0.01, 0.24)** | | |

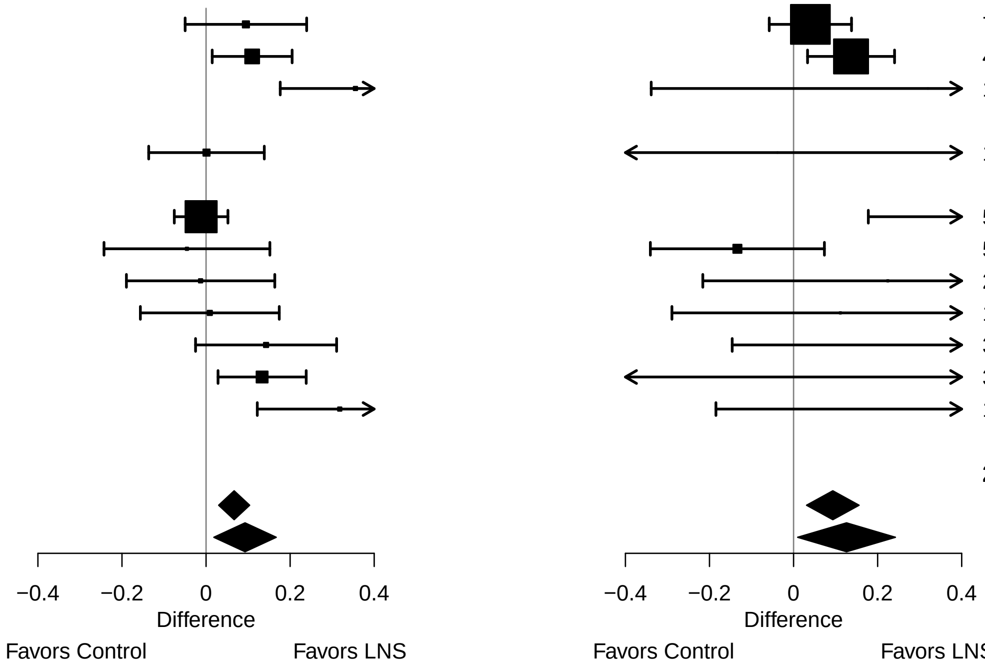

**Supplemental figure 5D: Mean difference in social-emotional z-score**

#### 5D2: Stratified by Maternal BMI

**P−for−interaction = 0.885**
**Difference in MDs = −0.01 (−0.07, 0.06)**

| | | | At least 20 kg/m$^2$ | | | | | | Less than 20 kg/m$^2$ | | | | | |
|---|---|---|---|---|---|---|---|---|---|---|---|---|---|---|
| **Country** | **Trial** | **Tool** | **LNS N** | **Control N** | **Control Mean** | **MD (95% CI)** | **Fixed W** | **Random W** | **LNS N** | **Control N** | **Control Mean** | **MD (95% CI)** | **Fixed W** | **Random W** |
| Bangladesh | JiVitA−4 (21) | | | | | | | | | | | | | |
| Bangladesh | RDNS (22) | DMC | 726 | 335 | 0.05 | 0.05 (−0.09, 0.18) | 0.09 | 0.11 | 871 | 455 | −0.11 | 0.08 (−0.04, 0.20) | 0.22 | 0.17 |
| Bangladesh | WASH−B (23) | EASQ | 479 | 1493 | 0.03 | 0.09 (−0.02, 0.19) | 0.17 | 0.14 | 582 | 1732 | −0.08 | 0.15 (0.04, 0.26) | 0.28 | 0.18 |
| Burkina Faso | iLiNS−Zinc (24) | DMC | 480 | 225 | −0.16 | 0.30 (0.11, 0.50) | 0.04 | 0.07 | 264 | 150 | −0.35 | 0.42 (0.16, 0.69) | 0.05 | 0.07 |
| Ghana | GHANA (25) | | | | | | | | | | | | | |
| Ghana | iLiNS−DYADG (26) | PSED | 292 | 530 | 0.01 | −0.01 (−0.15, 0.14) | 0.08 | 0.10 | 35 | 116 | −0.04 | 0.01 (−0.34, 0.37) | 0.03 | 0.05 |
| Haiti | HAITI (27) | | | | | | | | | | | | | |
| Kenya | WASH−B (28) | EASQ | 1031 | 3535 | 0.01 | 0.01 (−0.06, 0.08) | 0.32 | 0.16 | 257 | 1012 | −0.02 | −0.01 (−0.14, 0.12) | 0.19 | 0.16 |
| Madagascar | MAHAY (29) | ASQI | | | | | | | | | | | | |
| Malawi | iLiNS−DYADM (30) | PSED | 127 | 260 | −0.05 | 0.12 (−0.09, 0.34) | 0.04 | 0.06 | 87 | 175 | 0.05 | −0.16 (−0.41, 0.09) | 0.05 | 0.08 |
| Malawi | iLiNS−DOSE (31) | PSED | 478 | 158 | 0.05 | 0.01 (−0.17, 0.19) | 0.05 | 0.08 | 163 | 61 | −0.20 | 0.05 (−0.24, 0.34) | 0.04 | 0.07 |
| Mali | PROMIS CS (32) | DMC | 683 | 664 | −0.07 | 0.18 (0.00, 0.37) | 0.05 | 0.08 | 238 | 274 | −0.07 | 0.06 (−0.13, 0.25) | 0.09 | 0.11 |
| Zimbabwe | SHINE (HIV−) (33) | MDAT | 601 | 581 | −0.07 | 0.17 (0.06, 0.29) | 0.12 | 0.12 | 100 | 96 | −0.15 | 0.18 (−0.09, 0.44) | 0.05 | 0.07 |
| Zimbabwe | SHINE (HIV+) (34) | MDAT | 124 | 113 | −0.14 | 0.31 (0.09, 0.52) | 0.04 | 0.07 | 25 | 23 | −0.24 | 0.25 (−0.26, 0.76) | 0.01 | 0.03 |
| | | | **5021** | **7894** | | **$I^2$ = 0.52, $Tau^2$ = 0.01** | | | **2622** | **4094** | | **$I^2$ = 0.39, $Tau^2$ = 0.01** | | |
| **Fixed** | | | | | | **0.08 (0.04, 0.12)** | | | | | | **0.09 (0.03, 0.14)** | | |
| **Random** | | | | | | **0.11 (0.04, 0.17)** | | | | | | **0.09 (0.00, 0.17)** | | |

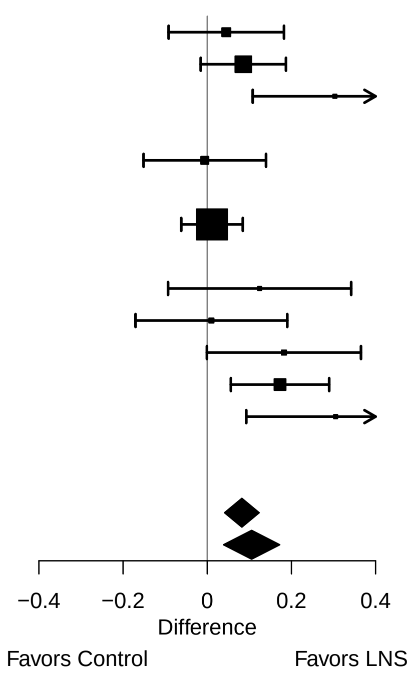

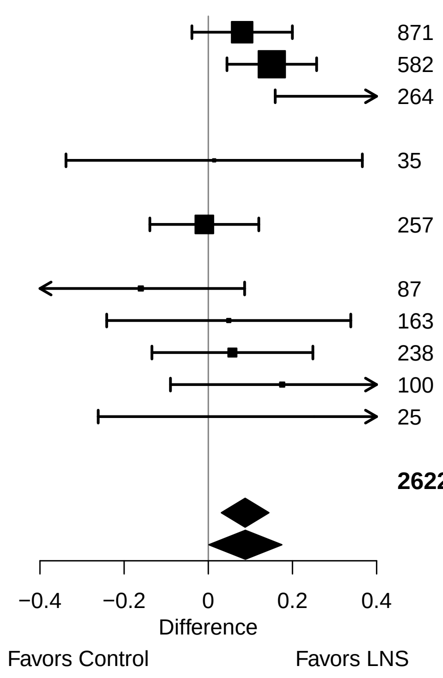

#### Supplemental figure 5D: Mean difference in social-emotional z-score

#### 5D3: Stratified by Maternal age

**P−for−interaction = 0.666**
**Difference in MDs = 0.01 (−0.05, 0.07)**

| Country | Trial | Tool | At least 25 y: LNS N | Control N | Control Mean | MD (95% CI) | Fixed W | Random W | Less than 25 y: LNS N | Control N | Control Mean | MD (95% CI) | Fixed W | Random W |
|---|---|---|---|---|---|---|---|---|---|---|---|---|---|---|
| Bangladesh | JiVitA−4 (21) | | | | | | | | | | | | | |
| Bangladesh | RDNS (22) | DMC | 450 | 221 | −0.08 | 0.04 (−0.17, 0.25) | 0.05 | 0.08 | 1207 | 594 | −0.01 | 0.07 (−0.03, 0.17) | 0.21 | 0.16 |
| Bangladesh | WASH−B (23) | EASQ | 458 | 1442 | −0.01 | −0.02 (−0.13, 0.09) | 0.19 | 0.11 | 603 | 1809 | −0.04 | 0.23 (0.13, 0.32) | 0.21 | 0.17 |
| Burkina Faso | iLiNS−Zinc (24) | DMC | 421 | 232 | −0.30 | 0.43 (0.23, 0.62) | 0.05 | 0.08 | 321 | 141 | −0.12 | 0.23 (0.04, 0.42) | 0.06 | 0.08 |
| Ghana | GHANA (25) | | | | | | | | | | | | | |
| Ghana | iLiNS−DYADG (26) | PSED | 212 | 404 | 0.00 | 0.04 (−0.13, 0.21) | 0.07 | 0.09 | 120 | 253 | −0.01 | −0.05 (−0.25, 0.16) | 0.05 | 0.07 |
| Haiti | HAITI (27) | | | | | | | | | | | | | |
| Kenya | WASH−B (28) | EASQ | 798 | 2622 | 0.02 | −0.02 (−0.10, 0.07) | 0.29 | 0.12 | 549 | 2092 | −0.03 | 0.06 (−0.04, 0.15) | 0.22 | 0.17 |
| Madagascar | MAHAY (29) | ASQI | 852 | 901 | 0.03 | −0.05 (−0.23, 0.14) | 0.06 | 0.09 | 759 | 703 | 0.05 | −0.10 (−0.31, 0.11) | 0.05 | 0.07 |
| Malawi | iLiNS−DYADM (30) | PSED | 103 | 224 | −0.03 | 0.04 (−0.19, 0.27) | 0.04 | 0.08 | 112 | 214 | 0.02 | −0.01 (−0.25, 0.22) | 0.04 | 0.06 |
| Malawi | iLiNS−DOSE (31) | PSED | 346 | 124 | −0.05 | 0.06 (−0.15, 0.27) | 0.05 | 0.08 | 286 | 90 | 0.00 | 0.00 (−0.23, 0.24) | 0.04 | 0.06 |
| Mali | PROMIS CS (32) | DMC | 492 | 492 | −0.07 | 0.17 (−0.02, 0.36) | 0.06 | 0.09 | 435 | 452 | −0.07 | 0.12 (−0.07, 0.31) | 0.05 | 0.08 |
| Zimbabwe | SHINE (HIV−) (33) | MDAT | 427 | 389 | −0.06 | 0.17 (0.03, 0.32) | 0.09 | 0.10 | 326 | 321 | −0.10 | 0.11 (−0.05, 0.28) | 0.07 | 0.09 |
| Zimbabwe | SHINE (HIV+) (34) | MDAT | 126 | 111 | −0.25 | 0.43 (0.22, 0.64) | 0.05 | 0.08 | 29 | 26 | −0.01 | 0.03 (−0.58, 0.65) | 0.01 | 0.01 |
| | | | **4685** | **7162** | | **I² = 0.72, Tau² = 0.02** | | | **4747** | **6695** | | **I² = 0.41, Tau² = 0.00** | | |
| **Fixed** | | | | | | **0.07 (0.02, 0.11)** | | | | | | **0.10 (0.05, 0.14)** | | |
| **Random** | | | | | | **0.11 (0.01, 0.21)** | | | | | | **0.09 (0.02, 0.15)** | | |

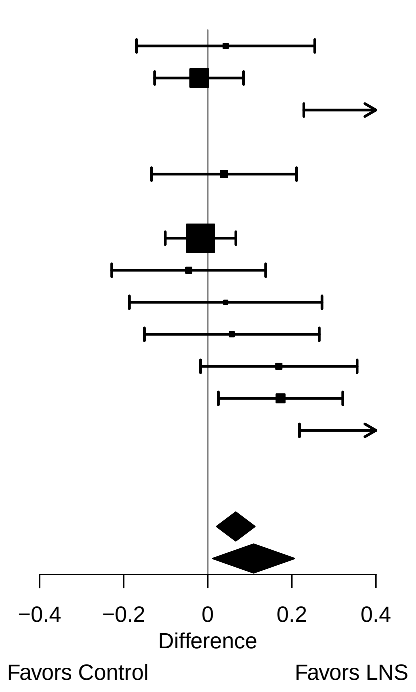

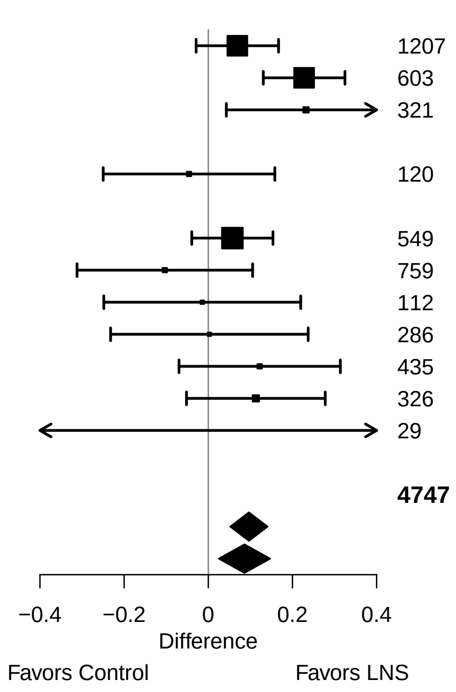

##### Supplemental figure 5D: Mean difference in social-emotional z-score

#### 5D4: Stratified by Maternal education

**P−for−interaction = 0.700**
**Difference in MDs = −0.01 (−0.08, 0.05)**

| Country | Trial | Tool | LNS N | Control N | Control Mean | Primary or greater MD (95% CI) | Fixed W | Random W | LNS N | Control N | Control Mean | Incomplete or no formal MD (95% CI) | Fixed W | Random W |
|---|---|---|---|---|---|---|---|---|---|---|---|---|---|---|
| Bangladesh | JiVitA−4 (21) | | | | | | | | | | | | | |
| Bangladesh | RDNS (22) | DMC | 1241 | 589 | 0.05 | 0.06 (−0.05, 0.16) | 0.15 | 0.12 | 416 | 226 | −0.25 | 0.04 (−0.12, 0.21) | 0.09 | 0.11 |
| Bangladesh | WASH−B (23) | EASQ | 750 | 2339 | 0.03 | 0.17 (0.08, 0.25) | 0.25 | 0.13 | 316 | 927 | −0.18 | 0.03 (−0.09, 0.15) | 0.16 | 0.14 |
| Burkina Faso | iLiNS−Zinc (24) | DMC | 32 | 9 | −0.42 | 0.52 (−0.09, 1.13) | 0.00 | 0.03 | 712 | 366 | −0.23 | 0.36 (0.18, 0.53) | 0.08 | 0.10 |
| Ghana | GHANA (25) | | | | | | | | | | | | | |
| Ghana | iLiNS−DYADG (26) | PSED | 257 | 518 | −0.01 | −0.03 (−0.17, 0.12) | 0.08 | 0.11 | 75 | 139 | 0.02 | 0.13 (−0.18, 0.43) | 0.03 | 0.05 |
| Haiti | HAITI (27) | | | | | | | | | | | | | |
| Kenya | WASH−B (28) | EASQ | 658 | 2258 | 0.10 | 0.04 (−0.04, 0.12) | 0.25 | 0.13 | 702 | 2484 | −0.09 | −0.02 (−0.10, 0.07) | 0.34 | 0.18 |
| Madagascar | MAHAY (29) | ASQI | 339 | 416 | 0.32 | −0.28 (−0.50, −0.07) | 0.04 | 0.09 | 1274 | 1188 | −0.06 | 0.01 (−0.18, 0.19) | 0.07 | 0.09 |
| Malawi | iLiNS−DYADM (30) | PSED | 33 | 69 | 0.01 | 0.24 (−0.16, 0.65) | 0.01 | 0.05 | 181 | 366 | −0.01 | −0.03 (−0.21, 0.15) | 0.07 | 0.10 |
| Malawi | iLiNS−DOSE (31) | PSED | 148 | 52 | −0.02 | −0.06 (−0.38, 0.26) | 0.02 | 0.06 | 485 | 162 | −0.03 | 0.06 (−0.12, 0.24) | 0.07 | 0.10 |
| Mali | PROMIS CS (32) | DMC | 100 | 94 | 0.17 | −0.03 (−0.37, 0.30) | 0.02 | 0.06 | 826 | 850 | −0.10 | 0.17 (0.00, 0.33) | 0.09 | 0.11 |
| Zimbabwe | SHINE (HIV−) (33) | MDAT | 763 | 731 | −0.08 | 0.15 (0.04, 0.26) | 0.15 | 0.12 | 29 | 25 | 0.18 | −0.24 (−0.78, 0.30) | 0.01 | 0.02 |
| Zimbabwe | SHINE (HIV+) (34) | MDAT | 149 | 130 | −0.19 | 0.37 (0.16, 0.57) | 0.04 | 0.09 | 9 | 8 | −0.22 | 0.11 (−0.82, 1.04) | 0.00 | 0.01 |
| | | | **4470** | **7205** | | **I² = 0.68, Tau² = 0.02** | | | **5025** | **6741** | | **I² = 0.47, Tau² = 0.01** | | |
| **Fixed** | | | | | | **0.09 (0.05, 0.13)** | | | | | | **0.05 (0.00, 0.10)** | | |
| **Random** | | | | | | **0.08 (−0.03, 0.19)** | | | | | | **0.07 (−0.01, 0.14)** | | |

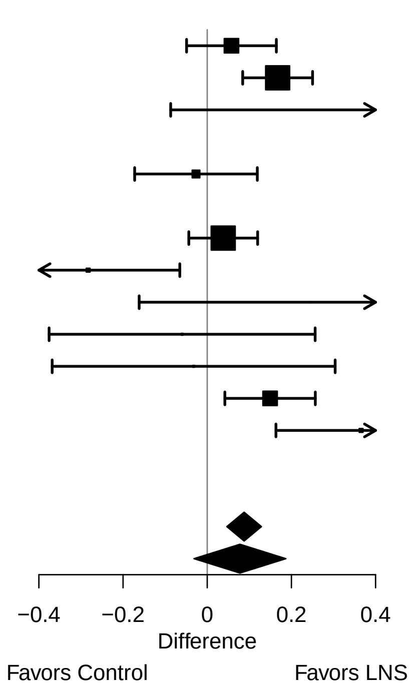

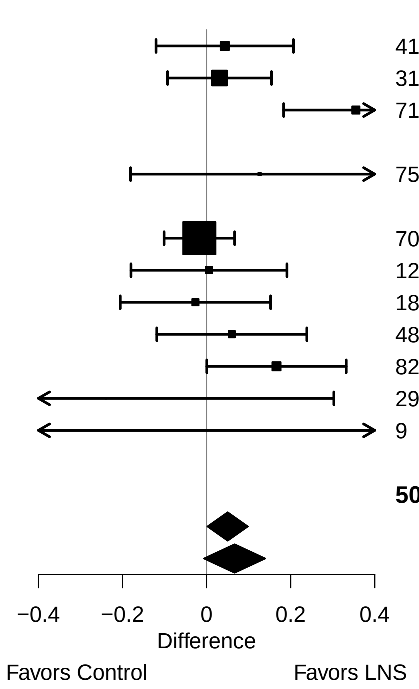

**Supplemental figure 5D: Mean difference in social-emotional z-score**

#### 5D5: Stratified by Maternal depressive symptoms

**P−for−interaction = 0.335**
**Difference in MDs = 0.04 (−0.04, 0.11)**

| | | | Less than 75th percentile | | | | | | At least 75th percentile | | | | | |
|---|---|---|---|---|---|---|---|---|---|---|---|---|---|---|
| Country | Trial | Tool | LNS N | Control N | Control Mean | MD (95% CI) | Fixed W | Random W | LNS N | Control N | Control Mean | MD (95% CI) | Fixed W | Random W |
| Bangladesh | JiVitA−4 (21) | | | | | | | | | | | | | |
| Bangladesh | RDNS (22) | DMC | 1075 | 472 | 0.03 | 0.05 (−0.06, 0.16) | 0.15 | 0.15 | 520 | 288 | −0.11 | 0.05 (−0.12, 0.22) | 0.16 | 0.16 |
| Bangladesh | WASH−B (23) | EASQ | 822 | 2356 | −0.02 | 0.13 (0.04, 0.21) | 0.24 | 0.17 | 223 | 830 | −0.08 | 0.10 (−0.05, 0.24) | 0.22 | 0.22 |
| Burkina Faso | iLiNS−Zinc (24) | DMC | | | | | | | | | | | | |
| Ghana | GHANA (25) | | | | | | | | | | | | | |
| Ghana | iLiNS−DYADG (26) | PSED | 242 | 435 | 0.00 | 0.08 (−0.08, 0.24) | 0.07 | 0.11 | 78 | 201 | −0.05 | −0.14 (−0.40, 0.11) | 0.07 | 0.07 |
| Haiti | HAITI (27) | | | | | | | | | | | | | |
| Kenya | WASH−B (28) | EASQ | 960 | 3317 | 0.00 | −0.01 (−0.08, 0.07) | 0.31 | 0.17 | 317 | 1156 | 0.14 | 0.05 (−0.08, 0.17) | 0.30 | 0.30 |
| Madagascar | MAHAY (29) | ASQI | 615 | 667 | −0.02 | −0.02 (−0.25, 0.22) | 0.03 | 0.08 | 268 | 242 | 0.08 | −0.12 (−0.37, 0.13) | 0.07 | 0.07 |
| Malawi | iLiNS−DYADM (30) | PSED | 153 | 302 | 0.09 | −0.01 (−0.20, 0.18) | 0.05 | 0.09 | 47 | 111 | −0.23 | 0.05 (−0.28, 0.38) | 0.04 | 0.04 |
| Malawi | iLiNS−DOSE (31) | PSED | | | | | | | | | | | | |
| Mali | PROMIS CS (32) | DMC | | | | | | | | | | | | |
| Zimbabwe | SHINE (HIV−) (33) | MDAT | 583 | 560 | −0.07 | 0.19 (0.08, 0.30) | 0.13 | 0.15 | 197 | 180 | −0.08 | 0.01 (−0.20, 0.22) | 0.11 | 0.11 |
| Zimbabwe | SHINE (HIV+) (34) | MDAT | 131 | 110 | −0.17 | 0.38 (0.15, 0.62) | 0.03 | 0.08 | 33 | 32 | −0.06 | 0.01 (−0.49, 0.51) | 0.02 | 0.02 |
| | | | **4581** | **8219** | | **I² = 0.63, Tau² = 0.01** | | | **1683** | **3040** | | **I² = 0.00, Tau² = 0.00** | | |
| **Fixed** | | | | | | **0.08 (0.04, 0.12)** | | | | | | **0.03 (−0.04, 0.10)** | | |
| **Random** | | | | | | **0.09 (0.01, 0.17)** | | | | | | **0.03 (−0.04, 0.10)** | | |

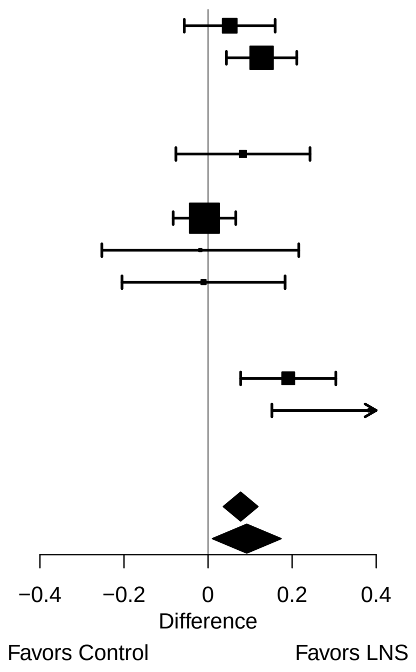

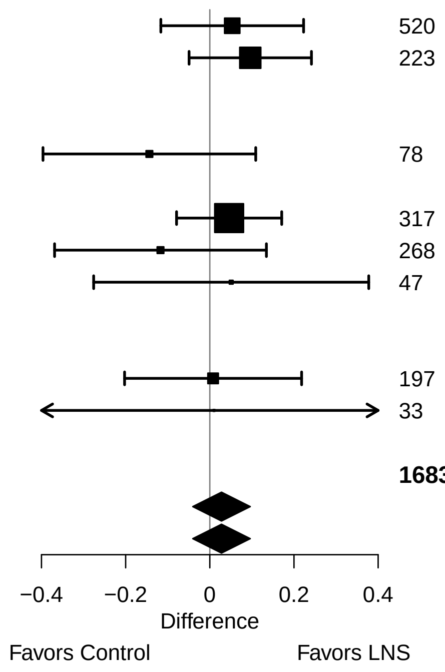

**Supplemental figure 5D: Mean difference in social-emotional z-score**

#### 5D6: Stratified by Child sex

**P−for−interaction = 0.706**
**Difference in MDs = −0.01 (−0.06, 0.04)**

| Country | Trial | Tool | Male LNS N | Male Control N | Male Control Mean | Male MD (95% CI) | Male Fixed W | Male Random W | Female LNS N | Female Control N | Female Control Mean | Female MD (95% CI) | Female Fixed W | Female Random W |
|---|---|---|---|---|---|---|---|---|---|---|---|---|---|---|
| Bangladesh | JiVitA−4 (21) | | | | | | | | | | | | | |
| Bangladesh | RDNS (22) | DMC | 830 | 411 | 0.02 | −0.02 (−0.12, 0.08) | 0.18 | 0.12 | 827 | 404 | −0.08 | 0.15 (0.01, 0.29) | 0.10 | 0.11 |
| Bangladesh | WASH−B (23) | EASQ | 522 | 1635 | −0.04 | 0.16 (0.05, 0.27) | 0.15 | 0.12 | 544 | 1631 | −0.02 | 0.09 (−0.01, 0.19) | 0.22 | 0.12 |
| Burkina Faso | iLiNS−Zinc (24) | DMC | 372 | 193 | −0.22 | 0.34 (0.20, 0.48) | 0.09 | 0.10 | 374 | 182 | −0.25 | 0.38 (0.12, 0.63) | 0.03 | 0.07 |
| Ghana | GHANA (25) | | | | | | | | | | | | | |
| Ghana | iLiNS−DYADG (26) | PSED | 164 | 306 | 0.01 | −0.04 (−0.22, 0.15) | 0.05 | 0.08 | 168 | 351 | −0.02 | 0.05 (−0.14, 0.24) | 0.06 | 0.09 |
| Haiti | HAITI (27) | | | | | | | | | | | | | |
| Kenya | WASH−B (28) | EASQ | 682 | 2260 | 0.00 | 0.00 (−0.08, 0.09) | 0.23 | 0.13 | 680 | 2485 | 0.00 | 0.02 (−0.06, 0.11) | 0.28 | 0.13 |
| Madagascar | MAHAY (29) | ASQI | 795 | 787 | 0.00 | 0.00 (−0.19, 0.19) | 0.05 | 0.08 | 818 | 817 | 0.07 | −0.14 (−0.34, 0.05) | 0.05 | 0.09 |
| Malawi | iLiNS−DYADM (30) | PSED | 105 | 205 | −0.02 | 0.07 (−0.17, 0.31) | 0.03 | 0.06 | 110 | 233 | 0.01 | −0.04 (−0.26, 0.19) | 0.04 | 0.08 |
| Malawi | iLiNS−DOSE (31) | PSED | 320 | 117 | 0.05 | −0.07 (−0.29, 0.15) | 0.04 | 0.07 | 324 | 104 | −0.11 | 0.15 (−0.07, 0.36) | 0.04 | 0.08 |
| Mali | PROMIS CS (32) | DMC | 469 | 514 | −0.08 | 0.17 (0.00, 0.33) | 0.07 | 0.09 | 458 | 430 | −0.07 | 0.13 (−0.08, 0.33) | 0.05 | 0.08 |
| Zimbabwe | SHINE (HIV−) (33) | MDAT | 424 | 388 | −0.10 | 0.20 (0.05, 0.34) | 0.09 | 0.10 | 417 | 411 | −0.02 | 0.05 (−0.09, 0.19) | 0.11 | 0.11 |
| Zimbabwe | SHINE (HIV+) (34) | MDAT | 84 | 79 | −0.12 | 0.24 (−0.04, 0.51) | 0.02 | 0.05 | 86 | 72 | −0.26 | 0.48 (0.16, 0.80) | 0.02 | 0.05 |
| | | | **4767** | **6895** | | **I² = 0.68, Tau² = 0.01** | | | **4806** | **7120** | | **I² = 0.52, Tau² = 0.01** | | |
| **Fixed** | | | | | | **0.08 (0.04, 0.13)** | | | | | | **0.07 (0.03, 0.12)** | | |
| **Random** | | | | | | **0.09 (0.02, 0.17)** | | | | | | **0.10 (0.01, 0.19)** | | |

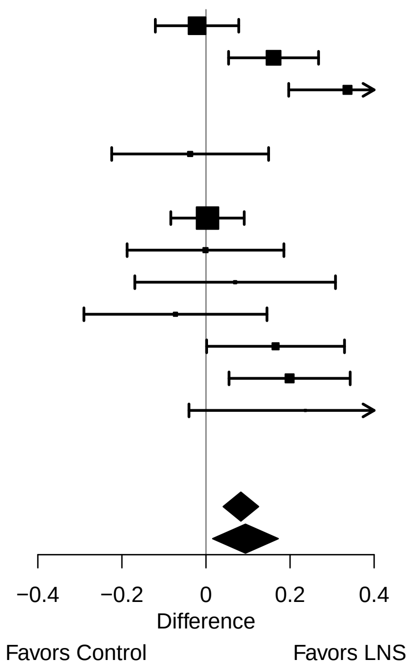

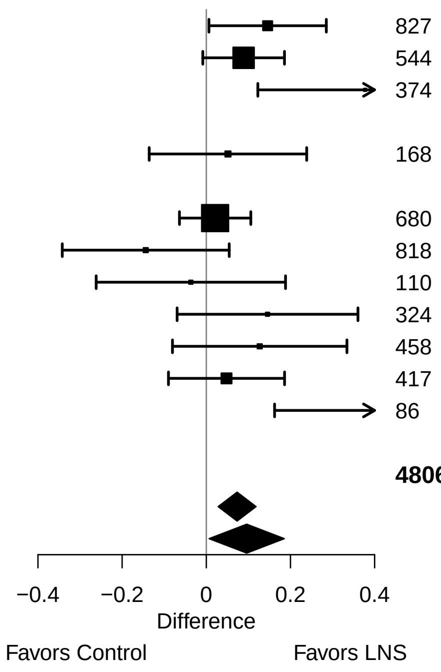

**Supplemental figure 5D: Mean difference in social-emotional z-score**

#### 5D7: Stratified by Child birth order

**P−for−interaction = 0.979**
**Difference in MDs = 0.00 (−0.06, 0.07)**

| Country | Trial | Tool | Later born LNS N | Later born Control N | Later born Control Mean | Later born MD (95% CI) | Later born Fixed W | Later born Random W | Firstborn LNS N | Firstborn Control N | Firstborn Control Mean | Firstborn MD (95% CI) | Firstborn Fixed W | Firstborn Random W |
|---|---|---|---|---|---|---|---|---|---|---|---|---|---|---|
| Bangladesh | JiVitA−4 (21) | | | | | | | | | | | | | |
| Bangladesh | RDNS (22) | DMC | 969 | 508 | −0.05 | 0.08 (−0.04, 0.20) | 0.10 | 0.10 | 687 | 307 | 0.00 | 0.03 (−0.10, 0.16) | 0.20 | 0.16 |
| Bangladesh | WASH−B (23) | EASQ | 669 | 2134 | −0.07 | 0.08 (0.00, 0.17) | 0.20 | 0.12 | 377 | 1031 | 0.07 | 0.14 (0.02, 0.26) | 0.24 | 0.17 |
| Burkina Faso | iLiNS−Zinc (24) | DMC | 587 | 290 | −0.27 | 0.42 (0.25, 0.60) | 0.05 | 0.08 | 159 | 85 | −0.12 | 0.11 (−0.15, 0.38) | 0.05 | 0.07 |
| Ghana | GHANA (25) | | | | | | | | | | | | | |
| Ghana | iLiNS−DYADG (26) | PSED | 225 | 444 | 0.02 | −0.05 (−0.21, 0.12) | 0.05 | 0.09 | 107 | 213 | −0.06 | 0.12 (−0.10, 0.34) | 0.07 | 0.09 |
| Haiti | HAITI (27) | | | | | | | | | | | | | |
| Kenya | WASH−B (28) | EASQ | 1071 | 3767 | 0.00 | −0.01 (−0.08, 0.06) | 0.30 | 0.12 | 289 | 974 | −0.01 | 0.08 (−0.06, 0.21) | 0.18 | 0.15 |
| Madagascar | MAHAY (29) | ASQI | 1162 | 1118 | 0.00 | −0.04 (−0.22, 0.15) | 0.04 | 0.08 | 433 | 465 | 0.09 | −0.14 (−0.38, 0.11) | 0.06 | 0.08 |
| Malawi | iLiNS−DYADM (30) | PSED | 170 | 349 | 0.01 | 0.07 (−0.11, 0.25) | 0.04 | 0.08 | 45 | 87 | −0.07 | −0.18 (−0.53, 0.18) | 0.03 | 0.05 |
| Malawi | iLiNS−DOSE (31) | PSED | 425 | 151 | −0.02 | 0.04 (−0.14, 0.22) | 0.04 | 0.08 | 133 | 41 | 0.05 | −0.02 (−0.39, 0.34) | 0.03 | 0.04 |
| Mali | PROMIS CS (32) | DMC | 770 | 811 | −0.07 | 0.16 (−0.01, 0.34) | 0.05 | 0.08 | 142 | 122 | −0.05 | 0.08 (−0.25, 0.40) | 0.03 | 0.05 |
| Zimbabwe | SHINE (HIV−) (33) | MDAT | 602 | 574 | −0.02 | 0.10 (−0.03, 0.22) | 0.10 | 0.10 | 203 | 191 | −0.20 | 0.19 (0.02, 0.37) | 0.11 | 0.12 |
| Zimbabwe | SHINE (HIV+) (34) | MDAT | 149 | 118 | −0.14 | 0.29 (0.08, 0.49) | 0.04 | 0.07 | 18 | 29 | −0.29 | 0.73 (0.16, 1.31) | 0.01 | 0.02 |
| | | | **6799** | **10264** | | **I² = 0.65, Tau² = 0.01** | | | **2593** | **3545** | | **I² = 0.26, Tau² = 0.01** | | |
| **Fixed** | | | | | | **0.07 (0.03, 0.11)** | | | | | | **0.09 (0.03, 0.14)** | | |
| **Random** | | | | | | **0.10 (0.02, 0.18)** | | | | | | **0.08 (0.00, 0.16)** | | |

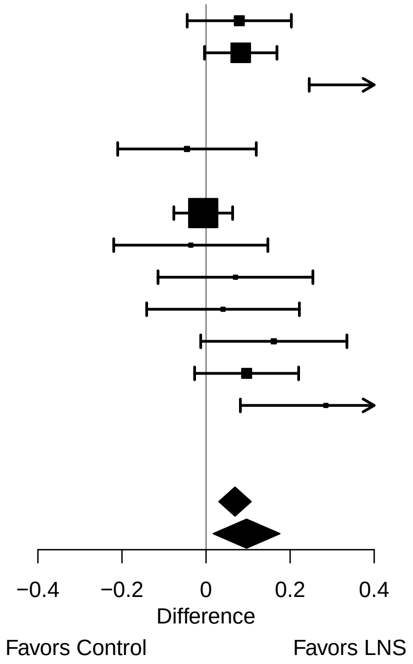

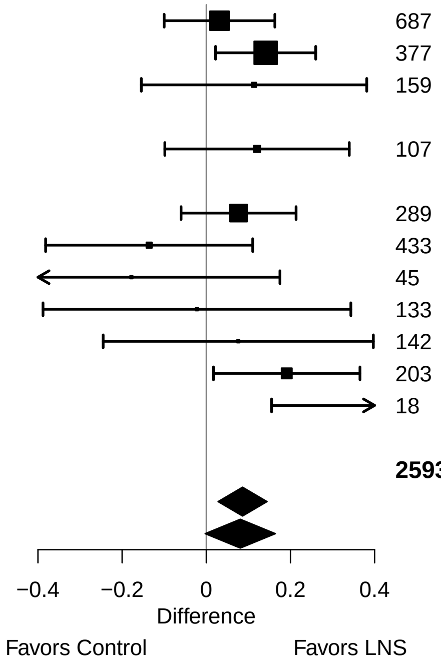

##### Supplemental figure 5D: Mean difference in social-emotional z-score

#### 5D8: Stratified by Child baseline stunting

**P−for−interaction = 0.495**
**Difference in MDs = 0.04 (−0.08, 0.16)**

| Country | Trial | Tool | No: LNS N | No: Control N | No: Control Mean | No: MD (95% CI) | No: Fixed W | No: Random W | Yes: LNS N | Yes: Control N | Yes: Control Mean | Yes: MD (95% CI) | Yes: Fixed W | Yes: Random W |
|---|---|---|---|---|---|---|---|---|---|---|---|---|---|---|
| Bangladesh | JiVitA−4 (21) | | | | | | | | | | | | | |
| Bangladesh | RDNS (22) | DMC | 1241 | 574 | 0.06 | 0.01 (−0.08, 0.10) | 0.36 | 0.19 | 354 | 185 | −0.29 | 0.18 (−0.03, 0.40) | 0.25 | 0.24 |
| Bangladesh | WASH−B (23) | EASQ | | | | | | | | | | | | |
| Burkina Faso | iLiNS−Zinc (24) | DMC | 576 | 294 | −0.16 | 0.35 (0.19, 0.52) | 0.11 | 0.14 | 170 | 80 | −0.50 | 0.37 (0.06, 0.68) | 0.12 | 0.13 |
| Ghana | GHANA (25) | | | | | | | | | | | | | |
| Ghana | iLiNS−DYADG (26) | PSED | 280 | 547 | 0.00 | 0.00 (−0.14, 0.15) | 0.14 | 0.16 | 28 | 63 | 0.02 | 0.09 (−0.32, 0.51) | 0.07 | 0.07 |
| Haiti | HAITI (27) | | | | | | | | | | | | | |
| Kenya | WASH−B (28) | EASQ | | | | | | | | | | | | |
| Madagascar | MAHAY (29) | ASQI | | | | | | | | | | | | |
| Malawi | iLiNS−DYADM (30) | PSED | 153 | 323 | 0.03 | 0.01 (−0.19, 0.20) | 0.08 | 0.13 | 48 | 96 | −0.06 | 0.02 (−0.31, 0.35) | 0.11 | 0.11 |
| Malawi | iLiNS−DOSE (31) | PSED | 464 | 147 | −0.04 | 0.07 (−0.11, 0.26) | 0.09 | 0.13 | 180 | 74 | 0.02 | −0.06 (−0.33, 0.21) | 0.17 | 0.17 |
| Mali | PROMIS CS (32) | DMC | | | | | | | | | | | | |
| Zimbabwe | SHINE (HIV−) (33) | MDAT | 532 | 490 | −0.02 | 0.15 (0.02, 0.28) | 0.17 | 0.16 | 122 | 105 | −0.26 | 0.12 (−0.11, 0.35) | 0.22 | 0.21 |
| Zimbabwe | SHINE (HIV+) (34) | MDAT | 115 | 89 | −0.18 | 0.35 (0.08, 0.62) | 0.04 | 0.09 | 34 | 32 | −0.34 | 0.42 (−0.02, 0.86) | 0.06 | 0.07 |
| | | | **3361** | **2464** | | **I² = 0.68, Tau² = 0.02** | | | **936** | **635** | | **I² = 0.08, Tau² = 0.00** | | |
| **Fixed** | | | | | | **0.09 (0.04, 0.15)** | | | | | | **0.14 (0.03, 0.25)** | | |
| **Random** | | | | | | **0.12 (0.01, 0.23)** | | | | | | **0.14 (0.03, 0.26)** | | |

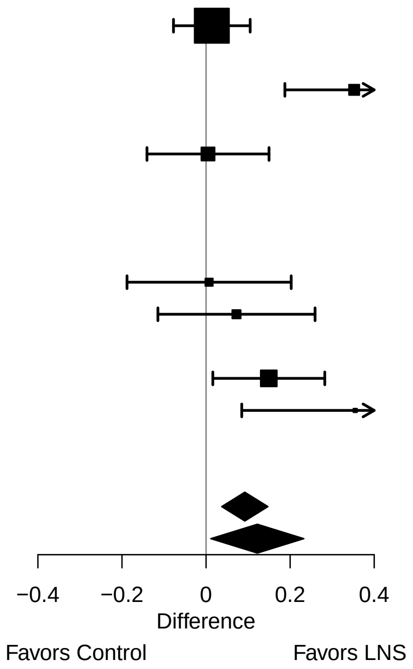

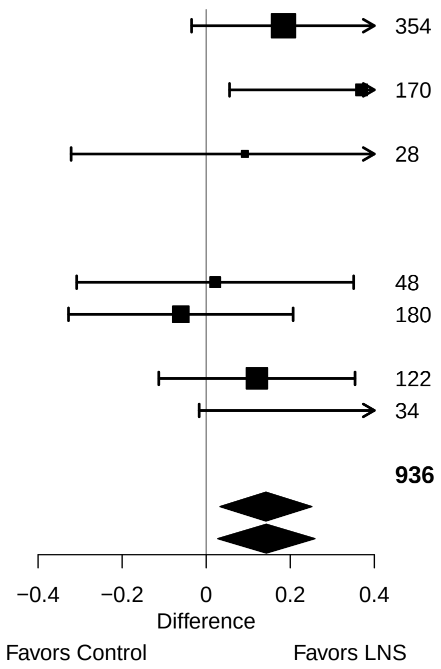

#### Supplemental figure 5D: Mean difference in social-emotional z-score

##### 5D9: Stratified by Child baseline acute malnutrition

**P−for−interaction** = **0.349**
**Difference in MDs =** **0.08 (−0.08, 0.24)**

| Country | Trial | Tool | No: LNS N | No: Control N | No: Control Mean | No: MD (95% CI) | No: Fixed W | No: Random W | Yes: LNS N | Yes: Control N | Yes: Control Mean | Yes: MD (95% CI) | Yes: Fixed W | Yes: Random W |
|---|---|---|---|---|---|---|---|---|---|---|---|---|---|---|
| Bangladesh | JiVitA−4 (21) | | | | | | | | | | | | | |
| Bangladesh | RDNS (22) | DMC | 1471 | 707 | 0.00 | 0.06 (−0.04, 0.16) | 0.30 | 0.17 | 124 | 53 | −0.38 | 0.12 (−0.38, 0.62) | 0.10 | 0.14 |
| Bangladesh | WASH−B (23) | EASQ | | | | | | | | | | | | |
| Burkina Faso | iLiNS−Zinc (24) | DMC | 556 | 266 | −0.15 | 0.31 (0.11, 0.51) | 0.07 | 0.13 | 190 | 109 | −0.44 | 0.46 (0.24, 0.68) | 0.51 | 0.30 |
| Ghana | GHANA (25) | | | | | | | | | | | | | |
| Ghana | iLiNS−DYADG (26) | PSED | 290 | 554 | 0.01 | −0.01 (−0.15, 0.13) | 0.15 | 0.15 | 19 | 56 | −0.10 | 0.26 (−0.26, 0.78) | 0.09 | 0.13 |
| Haiti | HAITI (27) | | | | | | | | | | | | | |
| Kenya | WASH−B (28) | EASQ | | | | | | | | | | | | |
| Madagascar | MAHAY (29) | ASQI | | | | | | | | | | | | |
| Malawi | iLiNS−DYADM (30) | PSED | 187 | 386 | 0.01 | 0.03 (−0.14, 0.21) | 0.10 | 0.14 | 14 | 36 | 0.02 | −0.29 (−0.90, 0.31) | 0.07 | 0.10 |
| Malawi | iLiNS−DOSE (31) | PSED | 607 | 206 | −0.03 | 0.04 (−0.12, 0.20) | 0.12 | 0.14 | 37 | 15 | 0.13 | −0.10 (−0.69, 0.49) | 0.07 | 0.11 |
| Mali | PROMIS CS (32) | DMC | | | | | | | | | | | | |
| Zimbabwe | SHINE (HIV−) (33) | MDAT | 620 | 566 | −0.05 | 0.13 (0.01, 0.25) | 0.20 | 0.16 | 36 | 34 | −0.18 | 0.39 (−0.06, 0.84) | 0.12 | 0.16 |
| Zimbabwe | SHINE (HIV+) (34) | MDAT | 137 | 114 | −0.27 | 0.46 (0.24, 0.68) | 0.06 | 0.12 | 14 | 8 | 0.28 | −0.32 (−1.09, 0.46) | 0.04 | 0.07 |
| | | | **3868** | **2799** | | **I² = 0.68, Tau² = 0.02** | | | **434** | **311** | | **I² = 0.42, Tau² = 0.03** | | |
| **Fixed** | | | | | | **0.10 (0.05, 0.16)** | | | | | | **0.28 (0.12, 0.43)** | | |
| **Random** | | | | | | **0.13 (0.01, 0.25)** | | | | | | **0.18 (−0.04, 0.41)** | | |

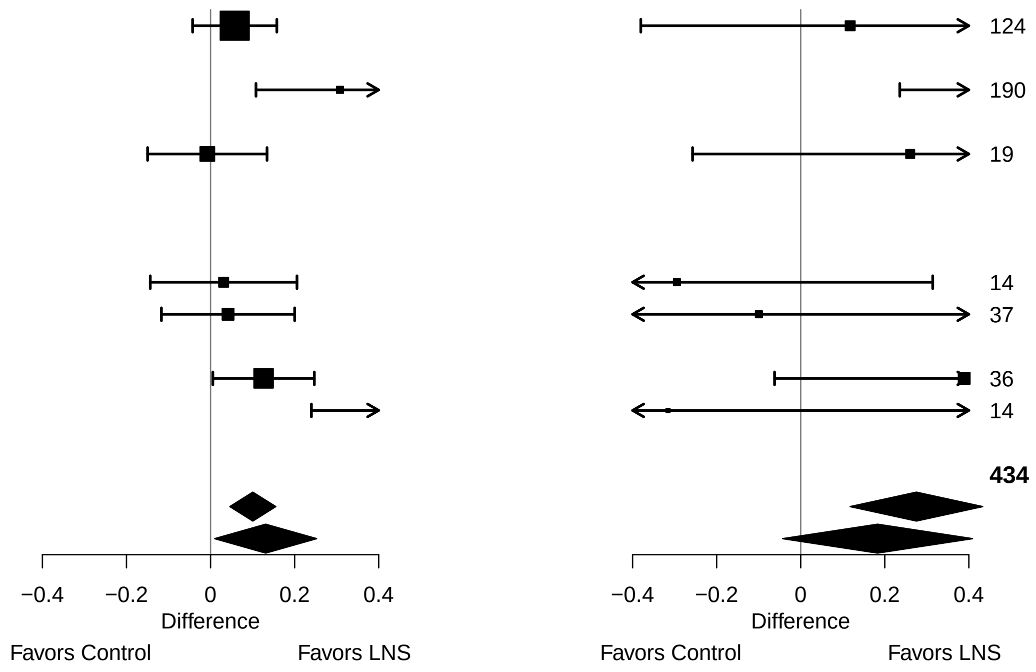

##### Supplemental figure 5D: Mean difference in social-emotional z-score

#### 5D10: Stratified by Child baseline anemia

**P−for−interaction = 0.834**
**Difference in MDs = −0.02 (−0.17, 0.14)**

| Country | Trial | Tool | Not anemic LNS N | Not anemic Control N | Not anemic Control Mean | Not anemic MD (95% CI) | Not anemic Fixed W | Not anemic Random W | Anemic LNS N | Anemic Control N | Anemic Control Mean | Anemic MD (95% CI) | Anemic Fixed W | Anemic Random W |
|---|---|---|---|---|---|---|---|---|---|---|---|---|---|---|
| Bangladesh | JiVitA−4 (21) | | | | | | | | | | | | | |
| Bangladesh | RDNS (22) | DMC | 212 | 96 | −0.11 | 0.15 (−0.11, 0.42) | 0.18 | 0.20 | 316 | 160 | −0.10 | 0.18 (−0.03, 0.39) | 0.19 | 0.20 |
| Bangladesh | WASH−B (23) | EASQ | | | | | | | | | | | | |
| Burkina Faso | iLiNS−Zinc (24) | DMC | 59 | 34 | −0.26 | 0.50 (0.13, 0.87) | 0.09 | 0.13 | 687 | 341 | −0.23 | 0.34 (0.15, 0.54) | 0.23 | 0.21 |
| Ghana | GHANA (25) | | | | | | | | | | | | | |
| Ghana | iLiNS−DYADG (26) | PSED | 193 | 350 | 0.00 | 0.03 (−0.14, 0.21) | 0.38 | 0.28 | 90 | 204 | 0.02 | −0.08 (−0.32, 0.16) | 0.15 | 0.17 |
| Haiti | HAITI (27) | | | | | | | | | | | | | |
| Kenya | WASH−B (28) | EASQ | | | | | | | | | | | | |
| Madagascar | MAHAY (29) | ASQI | | | | | | | | | | | | |
| Malawi | iLiNS−DYADM (30) | PSED | 66 | 147 | 0.05 | 0.02 (−0.27, 0.30) | 0.15 | 0.18 | 138 | 275 | −0.04 | 0.01 (−0.19, 0.22) | 0.21 | 0.20 |
| Malawi | iLiNS−DOSE (31) | PSED | 223 | 94 | 0.05 | −0.02 (−0.26, 0.22) | 0.21 | 0.22 | 417 | 126 | −0.08 | 0.08 (−0.12, 0.28) | 0.22 | 0.21 |
| Mali | PROMIS CS (32) | DMC | | | | | | | | | | | | |
| Zimbabwe | SHINE (HIV−) (33) | MDAT | | | | | | | | | | | | |
| Zimbabwe | SHINE (HIV+) (34) | MDAT | | | | | | | | | | | | |
| | | | **753** | **721** | | **I² = 0.37, Tau² = 0.02** | | | **1648** | **1106** | | **I² = 0.56, Tau² = 0.01** | | |
| **Fixed** | | | | | | **0.08 (−0.03, 0.19)** | | | | | | **0.12 (0.03, 0.22)** | | |
| **Random** | | | | | | **0.10 (−0.06, 0.26)** | | | | | | **0.12 (−0.03, 0.26)** | | |

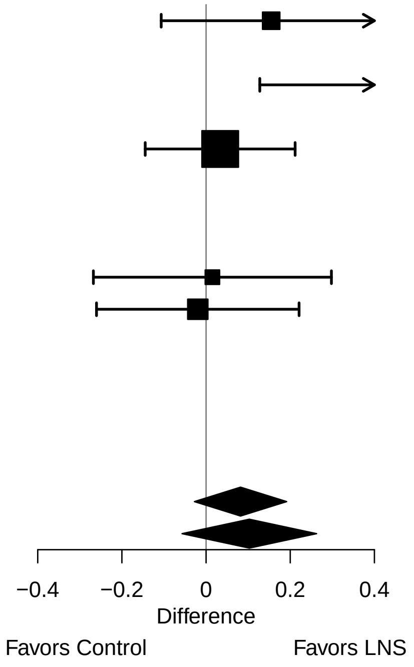

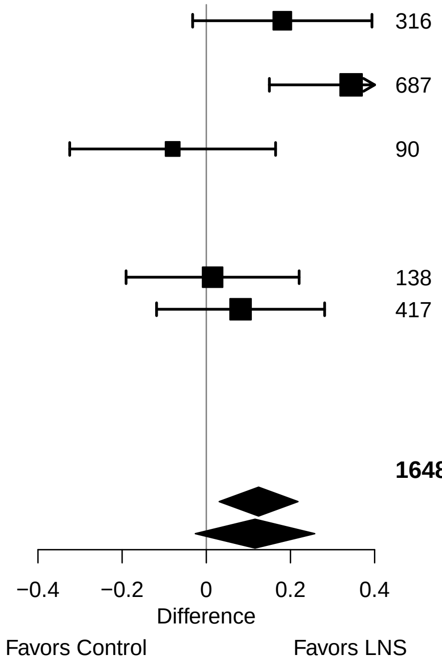

### Supplemental figure 5E: Social-emotional lowest decile prevalence ratio

#### 5E1: Stratified by Maternal height

**P−for−interaction = 0.638**
**Ratio of PRs = 1.06 (0.82, 1.38)**

| Country | Trial | Tool | At least 150.1 cm: LNS N | Control N | Control Prevalence | PR (95% CI) | Fixed W | Random W | Less than 150.1 cm: LNS N | Control N | Control Prevalence | PR (95% CI) | Fixed W | Random W |
|---|---|---|---|---|---|---|---|---|---|---|---|---|---|---|
| Bangladesh | JiVitA−4 (21) | | | | | | | | | | | | | |
| Bangladesh | RDNS (22) | DMC | 865 | 432 | 10.6 | 0.76 (0.53, 1.08) | 0.36 | 0.36 | 732 | 358 | 12.3 | 0.92 (0.68, 1.25) | 0.43 | 0.43 |
| Bangladesh | WASH−B (23) | EASQ | 567 | 1768 | 8.9 | 0.83 (0.60, 1.14) | 0.43 | 0.43 | 494 | 1458 | 12.2 | 0.80 (0.59, 1.07) | 0.46 | 0.46 |
| Burkina Faso | iLiNS−Zinc (24) | DMC | | | | | | | | | | | | |
| Ghana | GHANA (25) | | | | | | | | | | | | | |
| Ghana | iLiNS−DYADG (26) | PSED | | | | | | | | | | | | |
| Haiti | HAITI (27) | | | | | | | | | | | | | |
| Kenya | WASH−B (28) | EASQ | | | | | | | | | | | | |
| Madagascar | MAHAY (29) | ASQI | 956 | 986 | 9.4 | 1.12 (0.70, 1.78) | 0.21 | 0.21 | 553 | 538 | 8.4 | 1.19 (0.66, 2.16) | 0.11 | 0.11 |
| Malawi | iLiNS−DYADM (30) | PSED | | | | | | | | | | | | |
| Malawi | iLiNS−DOSE (31) | PSED | | | | | | | | | | | | |
| Mali | PROMIS CS (32) | DMC | | | | | | | | | | | | |
| Zimbabwe | SHINE (HIV−) (33) | MDAT | | | | | | | | | | | | |
| Zimbabwe | SHINE (HIV+) (34) | MDAT | | | | | | | | | | | | |
| | | | **2388** | **3186** | | **I² = 0.00, Tau² = 0.00** | | | **1779** | **2354** | | **I² = 0.00, Tau² = 0.00** | | |
| **Fixed** | | | | | | **0.86 (0.69, 1.06)** | | | | | | **0.89 (0.73, 1.08)** | | |
| **Random** | | | | | | **0.86 (0.69, 1.06)** | | | | | | **0.89 (0.73, 1.08)** | | |

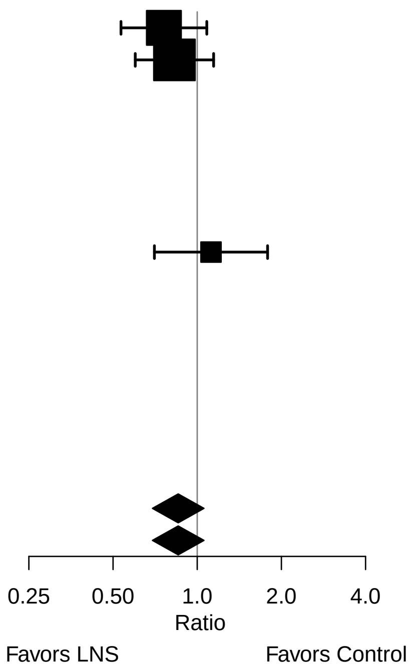

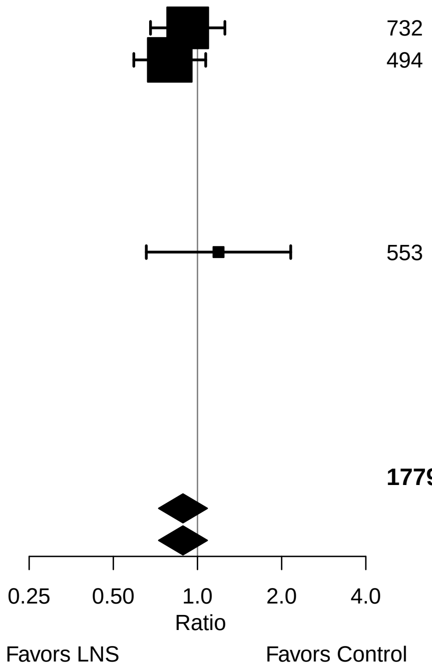

#### Supplemental figure 5E: Social-emotional lowest decile prevalence ratio

### 5E2: Stratified by Maternal BMI

**P−for−interaction = 0.199**
**Ratio of PRs = 1.16 (0.93, 1.45)**

| Country | Trial | Tool | At least 20 kg/m²: LNS N | Control N | Control Prevalence | PR (95% CI) | Fixed W | Random W | Less than 20 kg/m²: LNS N | Control N | Control Prevalence | PR (95% CI) | Fixed W | Random W |
|---|---|---|---|---|---|---|---|---|---|---|---|---|---|---|
| Bangladesh | JiVitA−4 (21) | | | | | | | | | | | | | |
| Bangladesh | RDNS (22) | DMC | 726 | 335 | 9.0 | 0.86 (0.54, 1.38) | 0.09 | 0.09 | 871 | 455 | 13.2 | 0.84 (0.61, 1.16) | 0.25 | 0.17 |
| Bangladesh | WASH−B (23) | EASQ | 479 | 1493 | 9.1 | 0.80 (0.54, 1.18) | 0.13 | 0.13 | 582 | 1732 | 11.5 | 0.82 (0.61, 1.10) | 0.29 | 0.17 |
| Burkina Faso | iLiNS−Zinc (24) | DMC | 480 | 225 | 14.7 | 0.44 (0.27, 0.71) | 0.09 | 0.09 | 264 | 150 | 19.3 | 0.39 (0.20, 0.75) | 0.06 | 0.11 |
| Ghana | GHANA (25) | | | | | | | | | | | | | |
| Ghana | iLiNS−DYADG (26) | PSED | | | | | | | | | | | | |
| Haiti | HAITI (27) | | | | | | | | | | | | | |
| Kenya | WASH−B (28) | EASQ | 1031 | 3535 | 10.0 | 0.85 (0.66, 1.10) | 0.31 | 0.29 | 257 | 1012 | 10.5 | 1.08 (0.73, 1.58) | 0.17 | 0.16 |
| Madagascar | MAHAY (29) | ASQI | | | | | | | | | | | | |
| Malawi | iLiNS−DYADM (30) | PSED | 127 | 260 | 12.7 | 0.74 (0.40, 1.39) | 0.05 | 0.05 | 87 | 175 | 6.9 | 1.68 (0.75, 3.73) | 0.04 | 0.09 |
| Malawi | iLiNS−DOSE (31) | PSED | 478 | 158 | 8.9 | 1.02 (0.57, 1.81) | 0.06 | 0.06 | 163 | 61 | 9.8 | 1.43 (0.61, 3.35) | 0.04 | 0.08 |
| Mali | PROMIS CS (32) | DMC | 683 | 664 | 11.9 | 0.66 (0.45, 0.98) | 0.13 | 0.13 | 238 | 274 | 9.9 | 1.07 (0.67, 1.70) | 0.12 | 0.14 |
| Zimbabwe | SHINE (HIV−) (33) | MDAT | 601 | 581 | 11.5 | 0.74 (0.50, 1.07) | 0.14 | 0.14 | 100 | 96 | 17.7 | 0.45 (0.20, 1.02) | 0.04 | 0.08 |
| Zimbabwe | SHINE (HIV+) (34) | MDAT | | | | | | | | | | | | |
| | | | **4605** | **7251** | | **I² = 0.07, Tau² = 0.00** | | | **2562** | **3955** | | **I² = 0.51, Tau² = 0.12** | | |
| **Fixed** | | | | | | **0.76 (0.66, 0.88)** | | | | | | **0.88 (0.75, 1.03)** | | |
| **Random** | | | | | | **0.76 (0.65, 0.88)** | | | | | | **0.87 (0.64, 1.19)** | | |

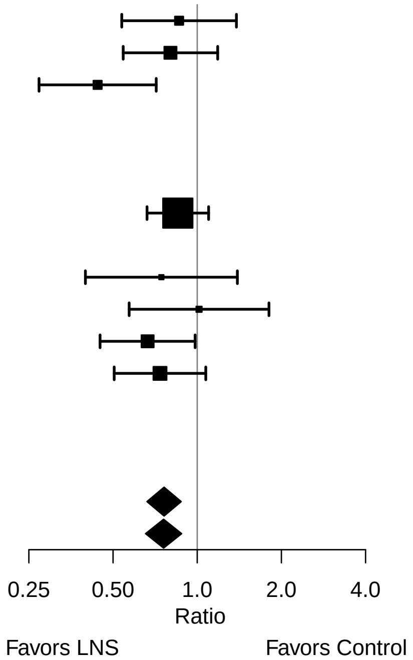

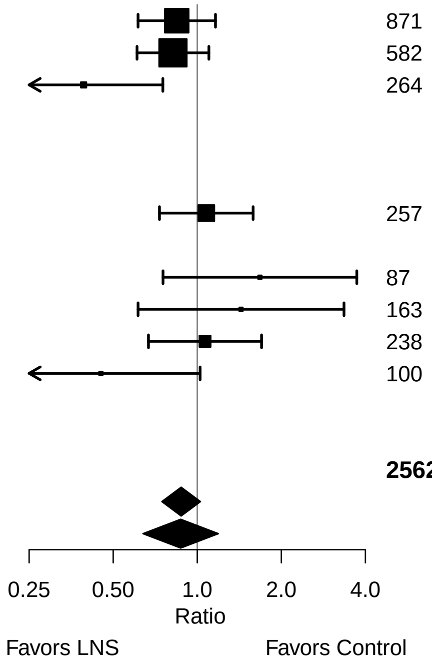

#### Supplemental figure 5E: Social-emotional lowest decile prevalence ratio

##### 5E3: Stratified by Maternal age

**P−for−interaction = 0.546**
**Ratio of PRs = 0.95 (0.79, 1.13)**

| Country | Trial | Tool | LNS N | Control N | Control Prevalence | At least 25 y PR (95% CI) | Fixed W | Random W | LNS N | Control N | Control Prevalence | Less than 25 y PR (95% CI) | Fixed W | Random W |
|---|---|---|---|---|---|---|---|---|---|---|---|---|---|---|
| Bangladesh | JiVitA−4 (21) | | | | | | | | | | | | | |
| Bangladesh | RDNS (22) | DMC | 450 | 221 | 11.8 | 1.06 (0.67, 1.66) | 0.09 | 0.10 | 1207 | 594 | 11.3 | 0.76 (0.58, 0.99) | 0.25 | 0.25 |
| Bangladesh | WASH−B (23) | EASQ | 458 | 1442 | 11.2 | 1.00 (0.73, 1.37) | 0.18 | 0.13 | 603 | 1809 | 10.0 | 0.66 (0.47, 0.94) | 0.15 | 0.15 |
| Burkina Faso | iLiNS−Zinc (24) | DMC | 421 | 232 | 19.8 | 0.36 (0.21, 0.62) | 0.06 | 0.09 | 321 | 141 | 11.3 | 0.58 (0.28, 1.17) | 0.04 | 0.04 |
| Ghana | GHANA (25) | | | | | | | | | | | | | |
| Ghana | iLiNS−DYADG (26) | PSED | 212 | 404 | 10.6 | 1.15 (0.73, 1.82) | 0.08 | 0.10 | 120 | 253 | 9.1 | 0.64 (0.28, 1.45) | 0.03 | 0.03 |
| Haiti | HAITI (27) | | | | | | | | | | | | | |
| Kenya | WASH−B (28) | EASQ | 798 | 2622 | 10.1 | 0.85 (0.65, 1.10) | 0.26 | 0.14 | 549 | 2092 | 10.7 | 0.90 (0.66, 1.24) | 0.18 | 0.18 |
| Madagascar | MAHAY (29) | ASQI | 852 | 901 | 8.8 | 1.23 (0.79, 1.91) | 0.09 | 0.10 | 759 | 703 | 10.0 | 1.06 (0.64, 1.75) | 0.07 | 0.07 |
| Malawi | iLiNS−DYADM (30) | PSED | 103 | 224 | 10.3 | 0.95 (0.47, 1.91) | 0.04 | 0.07 | 112 | 214 | 10.3 | 1.04 (0.54, 2.03) | 0.04 | 0.04 |
| Malawi | iLiNS−DOSE (31) | PSED | 346 | 124 | 10.5 | 1.05 (0.58, 1.90) | 0.05 | 0.08 | 286 | 90 | 6.7 | 1.47 (0.63, 3.43) | 0.03 | 0.03 |
| Mali | PROMIS CS (32) | DMC | 492 | 492 | 11.0 | 0.65 (0.42, 1.00) | 0.09 | 0.10 | 435 | 452 | 11.7 | 0.90 (0.61, 1.34) | 0.12 | 0.12 |
| Zimbabwe | SHINE (HIV−) (33) | MDAT | 427 | 389 | 12.6 | 0.56 (0.34, 0.91) | 0.07 | 0.09 | 326 | 321 | 13.1 | 0.75 (0.48, 1.18) | 0.09 | 0.09 |
| Zimbabwe | SHINE (HIV+) (34) | MDAT | | | | | | | | | | | | |
| | | | **4559** | **7051** | | **I² = 0.56, Tau² = 0.08** | | | **4718** | **6669** | | **I² = 0.00, Tau² = 0.00** | | |
| **Fixed** | | | | | | **0.86 (0.75, 0.98)** | | | | | | **0.81 (0.71, 0.93)** | | |
| **Random** | | | | | | **0.84 (0.67, 1.06)** | | | | | | **0.81 (0.71, 0.93)** | | |

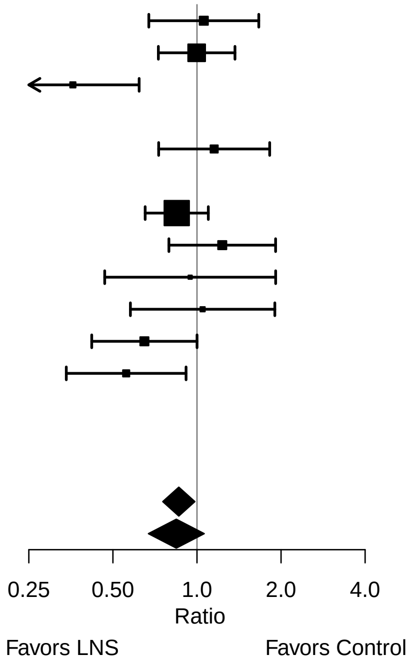

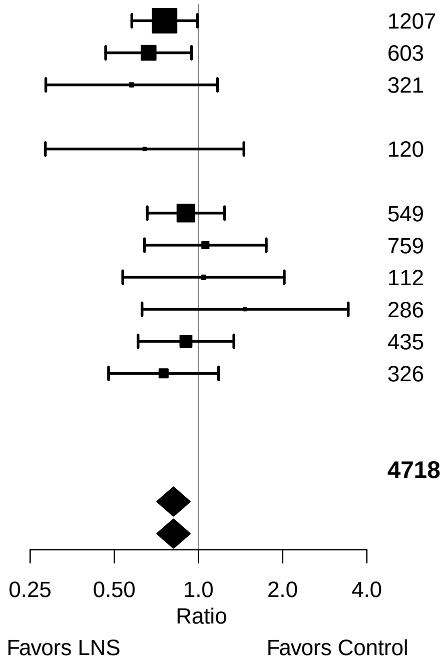

**Supplemental figure 5E: Social-emotional lowest decile prevalence ratio**

#### 5E4: Stratified by Maternal education

**P−for−interaction = 0.638**
**Ratio of PRs = 0.95 (0.76, 1.18)**

| Country | Trial | Tool | LNS N | Control N | Control Prevalence | Primary or greater PR (95% CI) | Fixed W | Random W | LNS N | Control N | Control Prevalence | Incomplete or no formal PR (95% CI) | Fixed W | Random W |
|---|---|---|---|---|---|---|---|---|---|---|---|---|---|---|
| Bangladesh | JiVitA−4 (21) | | | | | | | | | | | | | |
| Bangladesh | RDNS (22) | DMC | 1241 | 589 | 9.3 | 0.77 (0.55, 1.07) | 0.24 | 0.23 | 416 | 226 | 16.8 | 1.00 (0.70, 1.44) | 0.17 | 0.17 |
| Bangladesh | WASH−B (23) | EASQ | 750 | 2339 | 9.1 | 0.79 (0.57, 1.10) | 0.24 | 0.23 | 316 | 927 | 14.1 | 0.83 (0.61, 1.13) | 0.23 | 0.23 |
| Burkina Faso | iLiNS−Zinc (24) | DMC | | | | | | | | | | | | |
| Ghana | GHANA (25) | | | | | | | | | | | | | |
| Ghana | iLiNS−DYADG (26) | PSED | 257 | 518 | 10.0 | 1.05 (0.67, 1.63) | 0.13 | 0.16 | 75 | 139 | 10.1 | 0.79 (0.32, 1.98) | 0.03 | 0.03 |
| Haiti | HAITI (27) | | | | | | | | | | | | | |
| Kenya | WASH−B (28) | EASQ | 658 | 2258 | 7.4 | 0.90 (0.68, 1.21) | 0.31 | 0.26 | 702 | 2484 | 13.0 | 0.87 (0.66, 1.15) | 0.28 | 0.28 |
| Madagascar | MAHAY (29) | ASQI | 339 | 416 | 5.8 | 1.84 (0.98, 3.47) | 0.06 | 0.10 | 1274 | 1188 | 10.5 | 1.02 (0.66, 1.59) | 0.11 | 0.11 |
| Malawi | iLiNS−DYADM (30) | PSED | | | | | | | | | | | | |
| Malawi | iLiNS−DOSE (31) | PSED | | | | | | | | | | | | |
| Mali | PROMIS CS (32) | DMC | 100 | 94 | 7.4 | 1.21 (0.35, 4.19) | 0.02 | 0.03 | 826 | 850 | 11.8 | 0.74 (0.52, 1.05) | 0.18 | 0.18 |
| Zimbabwe | SHINE (HIV−) (33) | MDAT | | | | | | | | | | | | |
| Zimbabwe | SHINE (HIV+) (34) | MDAT | | | | | | | | | | | | |
| | | | **3345** | **6214** | | **I² = 0.29, Tau² = 0.03** | | | **3609** | **5814** | | **I² = 0.00, Tau² = 0.00** | | |
| **Fixed** | | | | | | **0.90 (0.77, 1.06)** | | | | | | **0.87 (0.75, 1.01)** | | |
| **Random** | | | | | | **0.93 (0.75, 1.16)** | | | | | | **0.87 (0.75, 1.01)** | | |

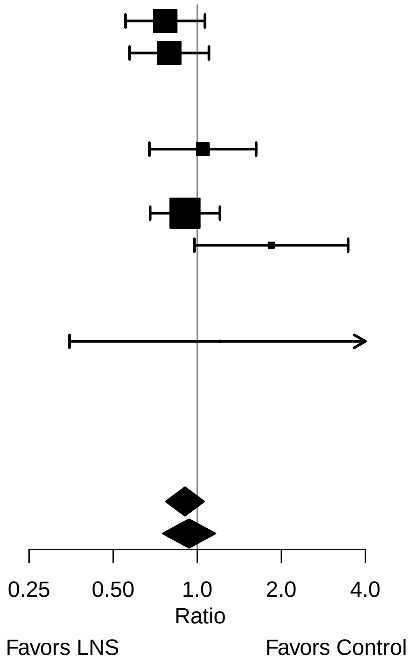

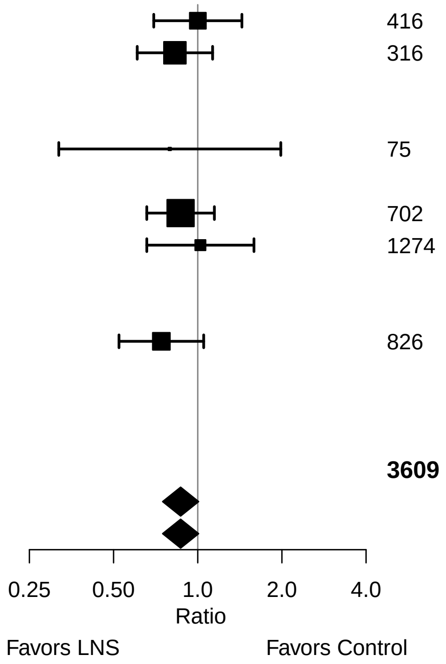

#### Supplemental figure 5E: Social-emotional lowest decile prevalence ratio

#### 5E5: Stratified by Maternal depressive symptoms

**P−for−interaction = 0.165**
**Ratio of PRs = 0.86 (0.69, 1.07)**

| | | | Less than 75th percentile | | | | | | | At least 75th percentile | | | | |
|---|---|---|---|---|---|---|---|---|---|---|---|---|---|---|
| **Country** | **Trial** | **Tool** | **LNS N** | **Control N** | **Control Prevalence** | **PR (95% CI)** | **Fixed W** | **Random W** | **LNS N** | **Control N** | **Control Prevalence** | **PR (95% CI)** | **Fixed W** | **Random W** |
| Bangladesh | JiVitA−4 (21) | | | | | | | | | | | | | |
| Bangladesh | RDNS (22) | DMC | 1075 | 472 | 10.8 | 0.74 (0.56, 0.97) | 0.21 | 0.21 | 520 | 288 | 12.2 | 1.01 (0.69, 1.50) | 0.24 | 0.24 |
| Bangladesh | WASH−B (23) | EASQ | 822 | 2356 | 9.6 | 0.81 (0.61, 1.07) | 0.20 | 0.20 | 223 | 830 | 13.1 | 0.89 (0.60, 1.31) | 0.24 | 0.24 |
| Burkina Faso | iLiNS−Zinc (24) | DMC | | | | | | | | | | | | |
| Ghana | GHANA (25) | | | | | | | | | | | | | |
| Ghana | iLiNS−DYADG (26) | PSED | 242 | 435 | 11.0 | 0.79 (0.48, 1.28) | 0.07 | 0.07 | 78 | 201 | 9.0 | 1.29 (0.60, 2.74) | 0.06 | 0.06 |
| Haiti | HAITI (27) | | | | | | | | | | | | | |
| Kenya | WASH−B (28) | EASQ | 960 | 3317 | 10.0 | 0.94 (0.74, 1.18) | 0.30 | 0.30 | 317 | 1156 | 7.7 | 0.74 (0.44, 1.23) | 0.14 | 0.14 |
| Madagascar | MAHAY (29) | ASQI | 615 | 667 | 10.5 | 0.85 (0.48, 1.51) | 0.05 | 0.05 | 268 | 242 | 9.1 | 1.03 (0.57, 1.85) | 0.10 | 0.10 |
| Malawi | iLiNS−DYADM (30) | PSED | 153 | 302 | 9.9 | 0.72 (0.37, 1.40) | 0.04 | 0.04 | 47 | 111 | 9.0 | 1.89 (0.80, 4.49) | 0.05 | 0.05 |
| Malawi | iLiNS−DOSE (31) | PSED | | | | | | | | | | | | |
| Mali | PROMIS CS (32) | DMC | | | | | | | | | | | | |
| Zimbabwe | SHINE (HIV−) (33) | MDAT | 583 | 560 | 11.1 | 0.70 (0.48, 1.02) | 0.11 | 0.11 | 197 | 180 | 16.7 | 0.64 (0.39, 1.04) | 0.15 | 0.15 |
| Zimbabwe | SHINE (HIV+) (34) | MDAT | 131 | 110 | 13.6 | 0.34 (0.14, 0.82) | 0.02 | 0.02 | 33 | 32 | 15.6 | 0.97 (0.32, 2.92) | 0.03 | 0.03 |
| | | | **4581** | **8219** | | **I² = 0.00, Tau² = 0.00** | | | **1683** | **3040** | | **I² = 0.00, Tau² = 0.00** | | |
| **Fixed** | | | | | | **0.80 (0.70, 0.91)** | | | | | | **0.92 (0.76, 1.11)** | | |
| **Random** | | | | | | **0.80 (0.70, 0.91)** | | | | | | **0.92 (0.76, 1.11)** | | |

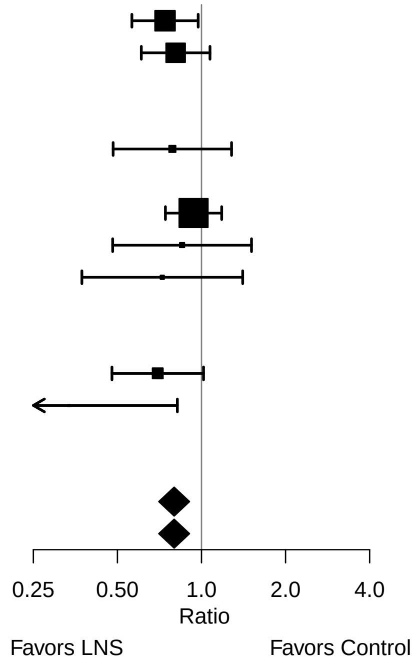

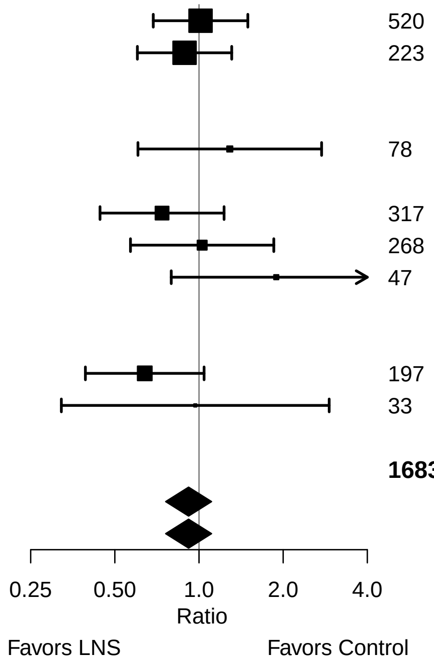

#### Supplemental figure 5E: Social-emotional lowest decile prevalence ratio

### 5E6: Stratified by Child sex

**P−for−interaction = 0.757**
**Ratio of PRs = 1.03 (0.87, 1.21)**

| Country | Trial | Tool | LNS N | Control N | Control Prevalence | Male PR (95% CI) | Fixed W | Random W | LNS N | Control N | Control Prevalence | Female PR (95% CI) | Fixed W | Random W |
|---|---|---|---|---|---|---|---|---|---|---|---|---|---|---|
| Bangladesh | JiVitA−4 (21) | | | | | | | | | | | | | |
| Bangladesh | RDNS (22) | DMC | 830 | 411 | 9.0 | 1.03 (0.73, 1.45) | 0.13 | 0.11 | 827 | 404 | 13.9 | 0.72 (0.51, 0.99) | 0.17 | 0.13 |
| Bangladesh | WASH−B (23) | EASQ | 522 | 1635 | 10.4 | 0.77 (0.56, 1.08) | 0.14 | 0.11 | 544 | 1631 | 10.6 | 0.85 (0.61, 1.19) | 0.16 | 0.13 |
| Burkina Faso | iLiNS−Zinc (24) | DMC | 372 | 193 | 13.5 | 0.44 (0.27, 0.70) | 0.07 | 0.09 | 374 | 182 | 19.8 | 0.39 (0.22, 0.69) | 0.06 | 0.08 |
| Ghana | GHANA (25) | | | | | | | | | | | | | |
| Ghana | iLiNS−DYADG (26) | PSED | 164 | 306 | 7.8 | 1.32 (0.73, 2.39) | 0.04 | 0.08 | 168 | 351 | 12.0 | 0.80 (0.46, 1.37) | 0.06 | 0.08 |
| Haiti | HAITI (27) | | | | | | | | | | | | | |
| Kenya | WASH−B (28) | EASQ | 682 | 2260 | 10.8 | 0.96 (0.76, 1.22) | 0.27 | 0.13 | 680 | 2485 | 9.9 | 0.77 (0.55, 1.09) | 0.16 | 0.13 |
| Madagascar | MAHAY (29) | ASQI | 795 | 787 | 11.1 | 0.92 (0.60, 1.42) | 0.08 | 0.10 | 818 | 817 | 7.6 | 1.48 (0.90, 2.43) | 0.08 | 0.09 |
| Malawi | iLiNS−DYADM (30) | PSED | 105 | 205 | 12.2 | 0.62 (0.29, 1.34) | 0.03 | 0.06 | 110 | 233 | 8.6 | 1.48 (0.78, 2.82) | 0.04 | 0.07 |
| Malawi | iLiNS−DOSE (31) | PSED | 320 | 117 | 6.8 | 1.74 (0.83, 3.61) | 0.03 | 0.06 | 324 | 104 | 12.5 | 0.69 (0.37, 1.28) | 0.05 | 0.07 |
| Mali | PROMIS CS (32) | DMC | 469 | 514 | 10.7 | 0.76 (0.51, 1.12) | 0.10 | 0.10 | 458 | 430 | 12.1 | 0.78 (0.52, 1.16) | 0.12 | 0.11 |
| Zimbabwe | SHINE (HIV−) (33) | MDAT | 424 | 388 | 13.1 | 0.66 (0.43, 1.01) | 0.09 | 0.10 | 417 | 411 | 10.7 | 0.76 (0.48, 1.21) | 0.09 | 0.10 |
| Zimbabwe | SHINE (HIV+) (34) | MDAT | 84 | 79 | 20.3 | 0.35 (0.15, 0.82) | 0.02 | 0.05 | 86 | 72 | 9.7 | 0.60 (0.17, 2.16) | 0.01 | 0.02 |
| | | | **4767** | **6895** | | **I² = 0.55, Tau² = 0.10** | | | **4806** | **7120** | | **I² = 0.40, Tau² = 0.05** | | |
| **Fixed** | | | | | | **0.84 (0.74, 0.95)** | | | | | | **0.80 (0.70, 0.92)** | | |
| **Random** | | | | | | **0.81 (0.64, 1.03)** | | | | | | **0.80 (0.66, 0.98)** | | |

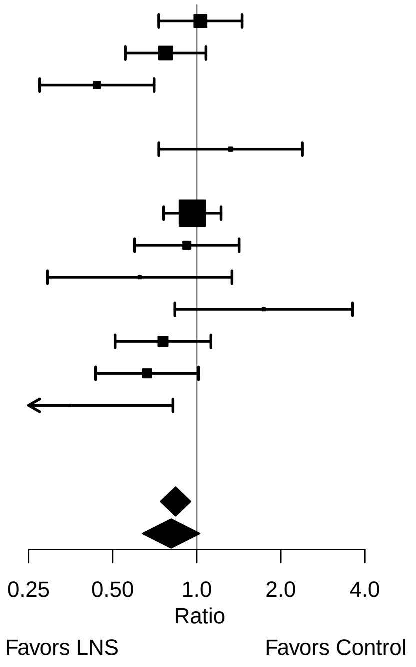

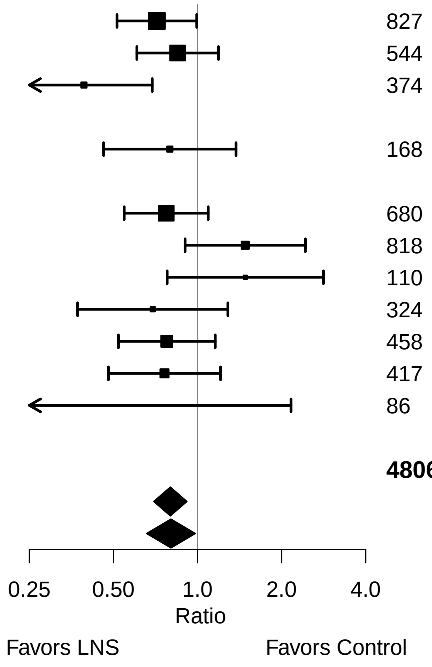

#### Supplemental figure 5E: Social-emotional lowest decile prevalence ratio

##### 5E7: Stratified by Child birth order

**P−for−interaction = 0.920**
**Ratio of PRs = 1.01 (0.83, 1.24)**

| Country | Trial | Tool | LNS N | Control N | Control Prevalence | Later born PR (95% CI) | Fixed W | Random W | LNS N | Control N | Control Prevalence | Firstborn PR (95% CI) | Fixed W | Random W |
|---|---|---|---|---|---|---|---|---|---|---|---|---|---|---|
| Bangladesh | JiVitA−4 (21) | | | | | | | | | | | | | |
| Bangladesh | RDNS (22) | DMC | 969 | 508 | 12.4 | 0.82 (0.59, 1.12) | 0.13 | 0.12 | 687 | 307 | 9.8 | 0.91 (0.65, 1.26) | 0.29 | 0.29 |
| Bangladesh | WASH−B (23) | EASQ | 669 | 2134 | 10.8 | 0.88 (0.67, 1.16) | 0.17 | 0.13 | 377 | 1031 | 9.0 | 0.76 (0.50, 1.18) | 0.17 | 0.17 |
| Burkina Faso | iLiNS−Zinc (24) | DMC | 587 | 290 | 17.9 | 0.34 (0.23, 0.52) | 0.07 | 0.10 | 159 | 85 | 11.8 | 0.80 (0.34, 1.87) | 0.04 | 0.04 |
| Ghana | GHANA (25) | | | | | | | | | | | | | |
| Ghana | iLiNS−DYADG (26) | PSED | 225 | 444 | 9.7 | 1.24 (0.79, 1.95) | 0.06 | 0.10 | 107 | 213 | 10.8 | 0.52 (0.22, 1.24) | 0.04 | 0.04 |
| Haiti | HAITI (27) | | | | | | | | | | | | | |
| Kenya | WASH−B (28) | EASQ | 1071 | 3767 | 10.4 | 0.89 (0.71, 1.11) | 0.26 | 0.14 | 289 | 974 | 10.1 | 0.83 (0.53, 1.29) | 0.16 | 0.16 |
| Madagascar | MAHAY (29) | ASQI | 1162 | 1118 | 8.9 | 1.17 (0.74, 1.84) | 0.06 | 0.10 | 433 | 465 | 10.8 | 1.05 (0.59, 1.88) | 0.09 | 0.09 |
| Malawi | iLiNS−DYADM (30) | PSED | 170 | 349 | 10.9 | 0.81 (0.46, 1.43) | 0.04 | 0.08 | 45 | 87 | 8.0 | 1.93 (0.72, 5.17) | 0.03 | 0.03 |
| Malawi | iLiNS−DOSE (31) | PSED | | | | | | | | | | | | |
| Mali | PROMIS CS (32) | DMC | 770 | 811 | 11.2 | 0.75 (0.53, 1.07) | 0.10 | 0.11 | 142 | 122 | 9.0 | 1.09 (0.52, 2.31) | 0.06 | 0.06 |
| Zimbabwe | SHINE (HIV−) (33) | MDAT | 602 | 574 | 11.5 | 0.74 (0.52, 1.05) | 0.10 | 0.12 | 203 | 191 | 14.7 | 0.60 (0.36, 1.02) | 0.11 | 0.11 |
| Zimbabwe | SHINE (HIV+) (34) | MDAT | | | | | | | | | | | | |
| | | | **6225** | **9995** | | **I² = 0.66, Tau² = 0.09** | | | **2442** | **3475** | | **I² = 0.00, Tau² = 0.00** | | |
| **Fixed** | | | | | | **0.82 (0.73, 0.92)** | | | | | | **0.85 (0.71, 1.01)** | | |
| **Random** | | | | | | **0.80 (0.64, 1.01)** | | | | | | **0.85 (0.71, 1.01)** | | |

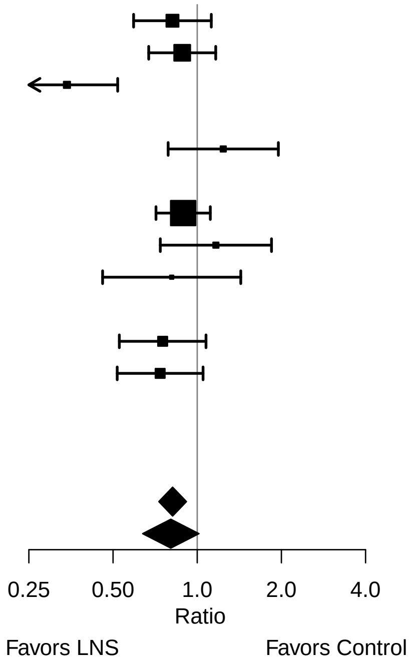

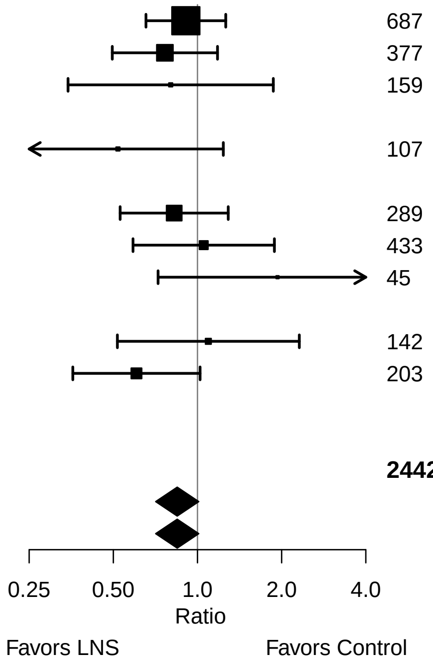

**Supplemental figure 5E: Social-emotional lowest decile prevalence ratio**

#### 5E8: Stratified by Child baseline stunting

**P−for−interaction = 0.728**
**Ratio of PRs = 0.94 (0.67, 1.32)**

| Country | Trial | Tool | No: LNS N | No: Control N | No: Control Prevalence | No: PR (95% CI) | No: Fixed W | No: Random W | Yes: LNS N | Yes: Control N | Yes: Control Prevalence | Yes: PR (95% CI) | Yes: Fixed W | Yes: Random W |
|---|---|---|---|---|---|---|---|---|---|---|---|---|---|---|
| Bangladesh | JiVitA−4 (21) | | | | | | | | | | | | | |
| Bangladesh | RDNS (22) | DMC | 1241 | 574 | 8.9 | 0.95 (0.70, 1.29) | 0.45 | 0.29 | 354 | 185 | 18.9 | 0.67 (0.44, 1.02) | 0.45 | 0.36 |
| Bangladesh | WASH−B (23) | EASQ | | | | | | | | | | | | |
| Burkina Faso | iLiNS−Zinc (24) | DMC | 576 | 294 | 13.3 | 0.41 (0.26, 0.63) | 0.22 | 0.25 | 170 | 80 | 27.5 | 0.43 (0.24, 0.75) | 0.24 | 0.26 |
| Ghana | GHANA (25) | | | | | | | | | | | | | |
| Ghana | iLiNS−DYADG (26) | PSED | | | | | | | | | | | | |
| Haiti | HAITI (27) | | | | | | | | | | | | | |
| Kenya | WASH−B (28) | EASQ | | | | | | | | | | | | |
| Madagascar | MAHAY (29) | ASQI | | | | | | | | | | | | |
| Malawi | iLiNS−DYADM (30) | PSED | | | | | | | | | | | | |
| Malawi | iLiNS−DOSE (31) | PSED | 464 | 147 | 8.8 | 1.10 (0.61, 1.98) | 0.12 | 0.21 | 180 | 74 | 10.8 | 1.08 (0.50, 2.33) | 0.13 | 0.17 |
| Mali | PROMIS CS (32) | DMC | | | | | | | | | | | | |
| Zimbabwe | SHINE (HIV−) (33) | MDAT | 532 | 490 | 10.8 | 0.59 (0.38, 0.91) | 0.22 | 0.25 | 122 | 105 | 14.3 | 0.86 (0.45, 1.65) | 0.18 | 0.21 |
| Zimbabwe | SHINE (HIV+) (34) | MDAT | | | | | | | | | | | | |
| | | | **2813** | **1505** | | **I² = 0.76, Tau² = 0.15** | | | **826** | **444** | | **I² = 0.33, Tau² = 0.05** | | |
| **Fixed** | | | | | | **0.73 (0.59, 0.89)** | | | | | | **0.67 (0.51, 0.89)** | | |
| **Random** | | | | | | **0.70 (0.45, 1.09)** | | | | | | **0.68 (0.48, 0.98)** | | |

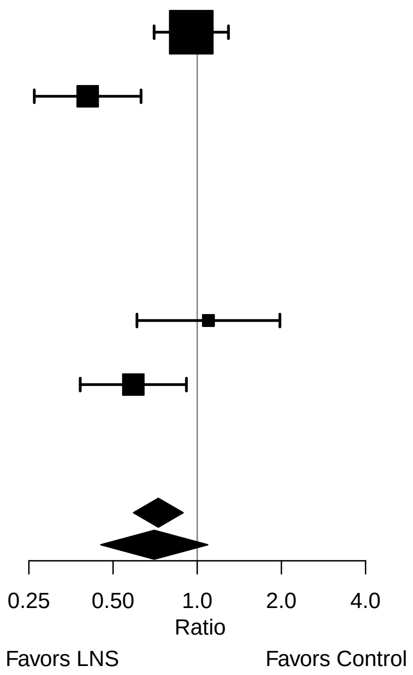

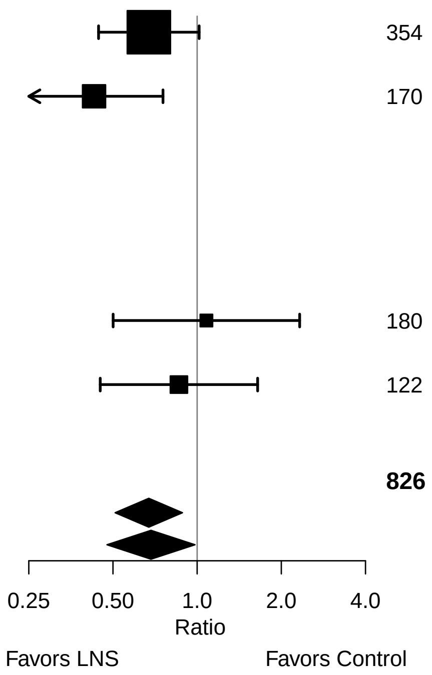

**Supplemental figure 5E: Social-emotional lowest decile prevalence ratio**

#### 5E9: Stratified by Child baseline acute malnutrition (insufficient comparisons)

**Supplemental figure 5E: Social-emotional lowest decile prevalence ratio**

#### 5E10: Stratified by Child baseline anemia

**P−for−interaction = 0.881**
**Ratio of PRs = 1.04 (0.63, 1.70)**

| Country | Trial | Tool | LNS N | Control N | Control Prevalence | Not anemic PR (95% CI) | Fixed W | Random W | LNS N | Control N | Control Prevalence | Anemic PR (95% CI) | Fixed W | Random W |
|---|---|---|---|---|---|---|---|---|---|---|---|---|---|---|
| Bangladesh | JiVitA−4 (21) | | | | | | | | | | | | | |
| Bangladesh | RDNS (22) | DMC | 212 | 96 | 14.6 | 0.49 (0.23, 1.01) | 0.23 | 0.24 | 316 | 160 | 12.5 | 0.76 (0.48, 1.21) | 0.37 | 0.37 |
| Bangladesh | WASH−B (23) | EASQ | | | | | | | | | | | | |
| Burkina Faso | iLiNS−Zinc (24) | DMC | | | | | | | | | | | | |
| Ghana | GHANA (25) | | | | | | | | | | | | | |
| Ghana | iLiNS−DYADG (26) | PSED | 193 | 350 | 10.3 | 1.01 (0.60, 1.69) | 0.46 | 0.41 | 90 | 204 | 9.8 | 0.91 (0.42, 1.98) | 0.13 | 0.13 |
| Haiti | HAITI (27) | | | | | | | | | | | | | |
| Kenya | WASH−B (28) | EASQ | | | | | | | | | | | | |
| Madagascar | MAHAY (29) | ASQI | | | | | | | | | | | | |
| Malawi | iLiNS−DYADM (30) | PSED | 66 | 147 | 8.2 | 0.93 (0.34, 2.53) | 0.12 | 0.14 | 138 | 275 | 10.9 | 1.06 (0.60, 1.88) | 0.25 | 0.25 |
| Malawi | iLiNS−DOSE (31) | PSED | 223 | 94 | 7.4 | 1.32 (0.59, 2.99) | 0.19 | 0.21 | 417 | 126 | 11.1 | 0.93 (0.53, 1.64) | 0.25 | 0.25 |
| Mali | PROMIS CS (32) | DMC | | | | | | | | | | | | |
| Zimbabwe | SHINE (HIV−) (33) | MDAT | | | | | | | | | | | | |
| Zimbabwe | SHINE (HIV+) (34) | MDAT | | | | | | | | | | | | |
| | | | **694** | **687** | | **I² = 0.21, Tau² = 0.04** | | | **961** | **765** | | **I² = 0.00, Tau² = 0.00** | | |
| **Fixed** | | | | | | **0.89 (0.62, 1.26)** | | | | | | **0.89 (0.67, 1.18)** | | |
| **Random** | | | | | | **0.88 (0.59, 1.32)** | | | | | | **0.89 (0.67, 1.18)** | | |

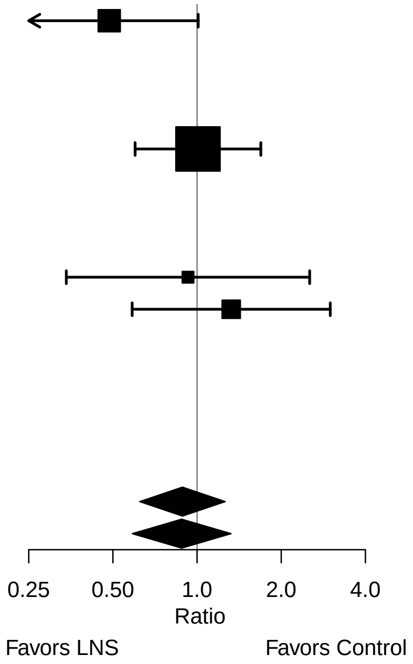

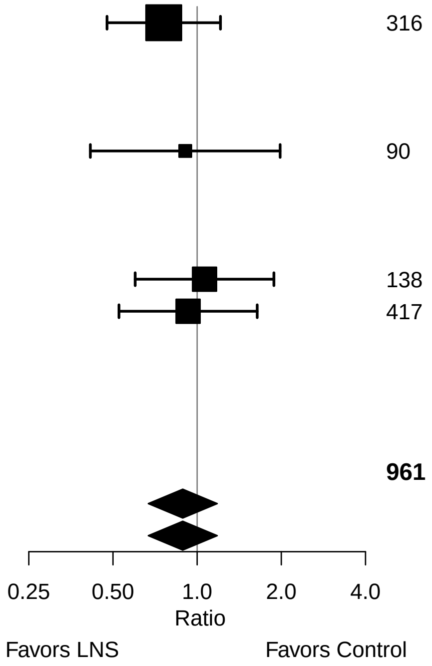

### Supplemental figure 5F: Social-emotional lowest decile prevalence difference

#### 5F1: Stratified by Maternal height

**P−for−interaction = 0.920**
**Difference in PDs = 0.00 (−0.02, 0.03)**

| Country | Trial | Tool | At least 150.1 cm: LNS N | Control N | Control Prevalence | PD (95% CI) | Fixed W | Random W | Less than 150.1 cm: LNS N | Control N | Control Prevalence | PD (95% CI) | Fixed W | Random W |
|---|---|---|---|---|---|---|---|---|---|---|---|---|---|---|
| Bangladesh | JiVitA−4 (21) | | | | | | | | | | | | | |
| Bangladesh | RDNS (22) | DMC | 865 | 432 | 10.6 | −0.03 (−0.06, 0.01) | 0.29 | 0.29 | 732 | 358 | 12.3 | −0.01 (−0.05, 0.03) | 0.36 | 0.36 |
| Bangladesh | WASH−B (23) | EASQ | 567 | 1768 | 8.9 | −0.02 (−0.04, 0.01) | 0.55 | 0.55 | 494 | 1458 | 12.2 | −0.02 (−0.06, 0.01) | 0.49 | 0.49 |
| Burkina Faso | iLiNS−Zinc (24) | DMC | | | | | | | | | | | | |
| Ghana | GHANA (25) | | | | | | | | | | | | | |
| Ghana | iLiNS−DYADG (26) | PSED | | | | | | | | | | | | |
| Haiti | HAITI (27) | | | | | | | | | | | | | |
| Kenya | WASH−B (28) | EASQ | | | | | | | | | | | | |
| Madagascar | MAHAY (29) | ASQI | 956 | 986 | 9.4 | 0.01 (−0.04, 0.06) | 0.16 | 0.16 | 553 | 538 | 8.4 | 0.02 (−0.04, 0.07) | 0.15 | 0.15 |
| Malawi | iLiNS−DYADM (30) | PSED | | | | | | | | | | | | |
| Malawi | iLiNS−DOSE (31) | PSED | | | | | | | | | | | | |
| Mali | PROMIS CS (32) | DMC | | | | | | | | | | | | |
| Zimbabwe | SHINE (HIV−) (33) | MDAT | | | | | | | | | | | | |
| Zimbabwe | SHINE (HIV+) (34) | MDAT | | | | | | | | | | | | |
| | | | **2388** | **3186** | | **I² = 0.00, Tau² = 0.00** | | | **1779** | **2354** | | **I² = 0.00, Tau² = 0.00** | | |
| **Fixed** | | | | | | **−0.01 (−0.03, 0.00)** | | | | | | **−0.01 (−0.03, 0.01)** | | |
| **Random** | | | | | | **−0.01 (−0.03, 0.00)** | | | | | | **−0.01 (−0.03, 0.01)** | | |

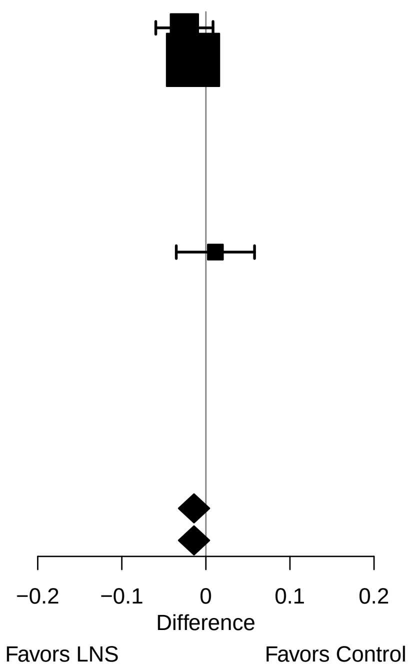

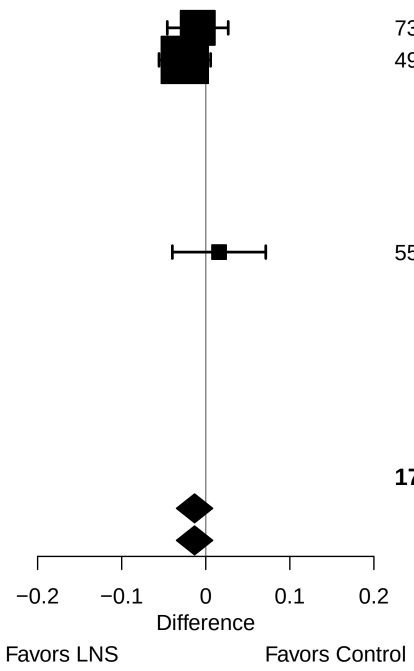

#### Supplemental figure 5F: Social-emotional lowest decile prevalence difference

### 5F2: Stratified by Maternal BMI

**P−for−interaction = 0.455**
**Difference in PDs = 0.01 (−0.01, 0.03)**

| | | | At least 20 kg/m² | | | | | | Less than 20 kg/m² | | | | | |
|---|---|---|---|---|---|---|---|---|---|---|---|---|---|---|
| **Country** | **Trial** | **Tool** | **LNS N** | **Control N** | **Control Prevalence** | **PD (95% CI)** | **Fixed W** | **Random W** | **LNS N** | **Control N** | **Control Prevalence** | **PD (95% CI)** | **Fixed W** | **Random W** |
| Bangladesh | JiVitA−4 (21) | | | | | | | | | | | | | |
| Bangladesh | RDNS (22) | DMC | 726 | 335 | 9.0 | −0.01 (−0.05, 0.03) | 0.10 | 0.10 | 871 | 455 | 13.2 | −0.02 (−0.06, 0.02) | 0.19 | 0.16 |
| Bangladesh | WASH−B (23) | EASQ | 479 | 1493 | 9.1 | −0.02 (−0.05, 0.01) | 0.18 | 0.18 | 582 | 1732 | 11.5 | −0.02 (−0.05, 0.01) | 0.34 | 0.17 |
| Burkina Faso | iLiNS−Zinc (24) | DMC | 480 | 225 | 14.7 | −0.08 (−0.14, −0.02) | 0.04 | 0.04 | 264 | 150 | 19.3 | −0.12 (−0.20, −0.03) | 0.04 | 0.10 |
| Ghana | GHANA (25) | | | | | | | | | | | | | |
| Ghana | iLiNS−DYADG (26) | PSED | | | | | | | | | | | | |
| Haiti | HAITI (27) | | | | | | | | | | | | | |
| Kenya | WASH−B (28) | EASQ | 1031 | 3535 | 10.0 | −0.01 (−0.04, 0.01) | 0.34 | 0.34 | 257 | 1012 | 10.5 | 0.01 (−0.03, 0.05) | 0.16 | 0.15 |
| Madagascar | MAHAY (29) | ASQI | | | | | | | | | | | | |
| Malawi | iLiNS−DYADM (30) | PSED | 127 | 260 | 12.7 | −0.03 (−0.10, 0.04) | 0.04 | 0.04 | 87 | 175 | 6.9 | 0.05 (−0.02, 0.12) | 0.06 | 0.11 |
| Malawi | iLiNS−DOSE (31) | PSED | 478 | 158 | 8.9 | 0.00 (−0.05, 0.05) | 0.06 | 0.06 | 163 | 61 | 9.8 | 0.04 (−0.06, 0.14) | 0.03 | 0.08 |
| Mali | PROMIS CS (32) | DMC | 683 | 664 | 11.9 | −0.04 (−0.08, 0.00) | 0.12 | 0.12 | 238 | 274 | 9.9 | 0.01 (−0.04, 0.05) | 0.14 | 0.15 |
| Zimbabwe | SHINE (HIV−) (33) | MDAT | 601 | 581 | 11.5 | −0.03 (−0.07, 0.01) | 0.12 | 0.12 | 100 | 96 | 17.7 | −0.10 (−0.19, 0.00) | 0.03 | 0.08 |
| Zimbabwe | SHINE (HIV+) (34) | MDAT | | | | | | | | | | | | |
| | | | **4605** | **7251** | | **I² = 0.00, Tau² = 0.00** | | | **2562** | **3955** | | **I² = 0.52, Tau² = 0.00** | | |
| **Fixed** | | | | | | **−0.02 (−0.04, −0.01)** | | | | | | **−0.01 (−0.03, 0.00)** | | |
| **Random** | | | | | | **−0.02 (−0.04, −0.01)** | | | | | | **−0.02 (−0.05, 0.02)** | | |

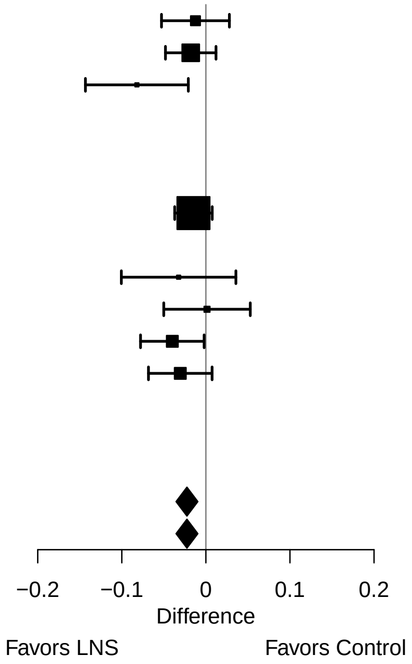

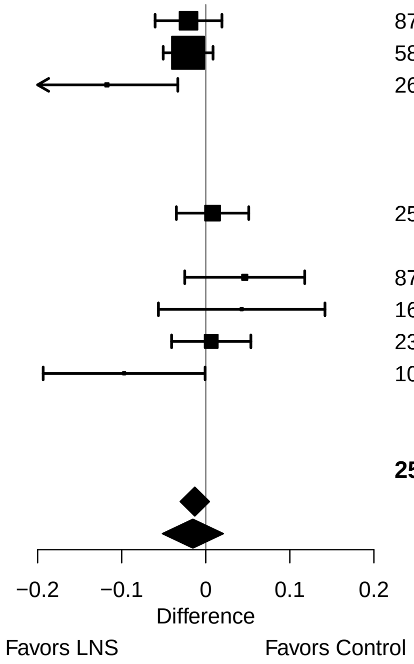

**Supplemental figure 5F: Social-emotional lowest decile prevalence difference**

#### 5F3: Stratified by Maternal age

**P−for−interaction = 0.701**
**Difference in PDs = 0.00 (−0.02, 0.01)**

| Country | Trial | Tool | LNS N | Control N | Control Prevalence | At least 25 y PD (95% CI) | Fixed W | Random W | LNS N | Control N | Control Prevalence | Less than 25 y PD (95% CI) | Fixed W | Random W |
|---|---|---|---|---|---|---|---|---|---|---|---|---|---|---|
| Bangladesh | JiVitA−4 (21) | | | | | | | | | | | | | |
| Bangladesh | RDNS (22) | DMC | 450 | 221 | 11.8 | 0.01 (−0.05, 0.06) | 0.06 | 0.09 | 1207 | 594 | 11.3 | −0.03 (−0.05, 0.00) | 0.22 | 0.22 |
| Bangladesh | WASH−B (23) | EASQ | 458 | 1442 | 11.2 | 0.00 (−0.04, 0.03) | 0.14 | 0.12 | 603 | 1809 | 10.0 | −0.03 (−0.06, −0.01) | 0.24 | 0.24 |
| Burkina Faso | iLiNS−Zinc (24) | DMC | 421 | 232 | 19.8 | −0.13 (−0.19, −0.06) | 0.04 | 0.08 | 321 | 141 | 11.3 | −0.05 (−0.11, 0.02) | 0.04 | 0.04 |
| Ghana | GHANA (25) | | | | | | | | | | | | | |
| Ghana | iLiNS−DYADG (26) | PSED | 212 | 404 | 10.6 | 0.02 (−0.04, 0.07) | 0.06 | 0.09 | 120 | 253 | 9.1 | −0.03 (−0.09, 0.03) | 0.05 | 0.05 |
| Haiti | HAITI (27) | | | | | | | | | | | | | |
| Kenya | WASH−B (28) | EASQ | 798 | 2622 | 10.1 | −0.02 (−0.04, 0.01) | 0.33 | 0.14 | 549 | 2092 | 10.7 | −0.01 (−0.04, 0.02) | 0.17 | 0.17 |
| Madagascar | MAHAY (29) | ASQI | 852 | 901 | 8.8 | 0.02 (−0.02, 0.06) | 0.09 | 0.11 | 759 | 703 | 10.0 | 0.01 (−0.05, 0.06) | 0.06 | 0.06 |
| Malawi | iLiNS−DYADM (30) | PSED | 103 | 224 | 10.3 | −0.01 (−0.08, 0.06) | 0.03 | 0.07 | 112 | 214 | 10.3 | 0.00 (−0.07, 0.07) | 0.03 | 0.03 |
| Malawi | iLiNS−DOSE (31) | PSED | 346 | 124 | 10.5 | 0.00 (−0.06, 0.07) | 0.04 | 0.08 | 286 | 90 | 6.7 | 0.03 (−0.04, 0.10) | 0.04 | 0.04 |
| Mali | PROMIS CS (32) | DMC | 492 | 492 | 11.0 | −0.04 (−0.08, 0.00) | 0.12 | 0.12 | 435 | 452 | 11.7 | −0.01 (−0.06, 0.03) | 0.09 | 0.09 |
| Zimbabwe | SHINE (HIV−) (33) | MDAT | 427 | 389 | 12.6 | −0.06 (−0.10, −0.01) | 0.09 | 0.11 | 326 | 321 | 13.1 | −0.03 (−0.09, 0.02) | 0.06 | 0.06 |
| Zimbabwe | SHINE (HIV+) (34) | MDAT | | | | | | | | | | | | |
| | | | **4559** | **7051** | | **I² = 0.57, Tau² = 0.00** | | | **4718** | **6669** | | **I² = 0.00, Tau² = 0.00** | | |
| **Fixed** | | | | | | **−0.02 (−0.03, 0.00)** | | | | | | **−0.02 (−0.03, −0.01)** | | |
| **Random** | | | | | | **−0.02 (−0.04, 0.01)** | | | | | | **−0.02 (−0.03, −0.01)** | | |

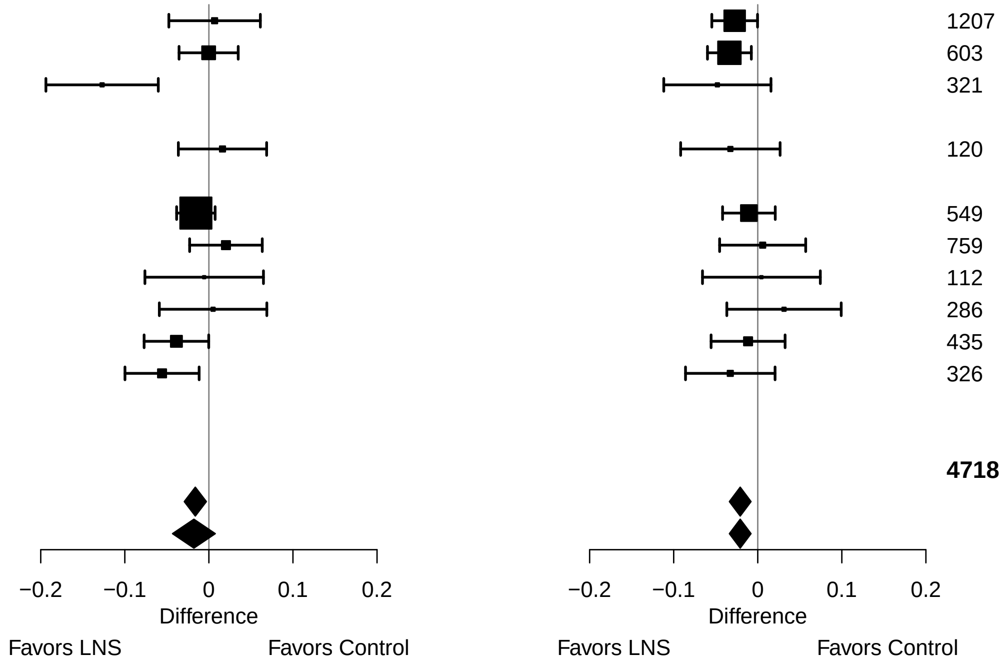

**Supplemental figure 5F: Social-emotional lowest decile prevalence difference**

#### 5F4: Stratified by Maternal education

**P−for−interaction = 0.125**
**Difference in PDs = −0.02 (−0.04, 0.00)**

| Country | Trial | Tool | LNS N | Control N | Control Prevalence | Primary or greater PD (95% CI) | Fixed W | Random W | LNS N | Control N | Control Prevalence | Incomplete or no formal PD (95% CI) | Fixed W | Random W |
|---|---|---|---|---|---|---|---|---|---|---|---|---|---|---|
| Bangladesh | JiVitA−4 (21) | | | | | | | | | | | | | |
| Bangladesh | RDNS (22) | DMC | 1241 | 589 | 9.3 | −0.02 (−0.05, 0.01) | 0.20 | 0.22 | 416 | 226 | 16.8 | 0.00 (−0.06, 0.06) | 0.08 | 0.08 |
| Bangladesh | WASH−B (23) | EASQ | 750 | 2339 | 9.1 | −0.02 (−0.04, 0.01) | 0.26 | 0.25 | 316 | 927 | 14.1 | −0.02 (−0.06, 0.01) | 0.20 | 0.20 |
| Burkina Faso | iLiNS−Zinc (24) | DMC | | | | | | | | | | | | |
| Ghana | GHANA (25) | | | | | | | | | | | | | |
| Ghana | iLiNS−DYADG (26) | PSED | 257 | 518 | 10.0 | 0.00 (−0.04, 0.05) | 0.08 | 0.11 | 75 | 139 | 10.1 | −0.02 (−0.10, 0.06) | 0.04 | 0.04 |
| Haiti | HAITI (27) | | | | | | | | | | | | | |
| Kenya | WASH−B (28) | EASQ | 658 | 2258 | 7.4 | −0.01 (−0.03, 0.01) | 0.40 | 0.30 | 702 | 2484 | 13.0 | −0.02 (−0.05, 0.02) | 0.29 | 0.29 |
| Madagascar | MAHAY (29) | ASQI | 339 | 416 | 5.8 | 0.05 (−0.01, 0.10) | 0.05 | 0.09 | 1274 | 1188 | 10.5 | 0.00 (−0.04, 0.05) | 0.14 | 0.14 |
| Malawi | iLiNS−DYADM (30) | PSED | | | | | | | | | | | | |
| Malawi | iLiNS−DOSE (31) | PSED | | | | | | | | | | | | |
| Mali | PROMIS CS (32) | DMC | 100 | 94 | 7.4 | 0.02 (−0.09, 0.12) | 0.01 | 0.03 | 826 | 850 | 11.8 | −0.03 (−0.06, 0.00) | 0.25 | 0.25 |
| Zimbabwe | SHINE (HIV−) (33) | MDAT | | | | | | | | | | | | |
| Zimbabwe | SHINE (HIV+) (34) | MDAT | | | | | | | | | | | | |
| | | | **3345** | **6214** | | **I² = 0.22, Tau² = 0.00** | | | **3609** | **5814** | | **I² = 0.00, Tau² = 0.00** | | |
| **Fixed** | | | | | | **−0.01 (−0.02, 0.00)** | | | | | | **−0.02 (−0.04, 0.00)** | | |
| **Random** | | | | | | **−0.01 (−0.02, 0.01)** | | | | | | **−0.02 (−0.04, 0.00)** | | |

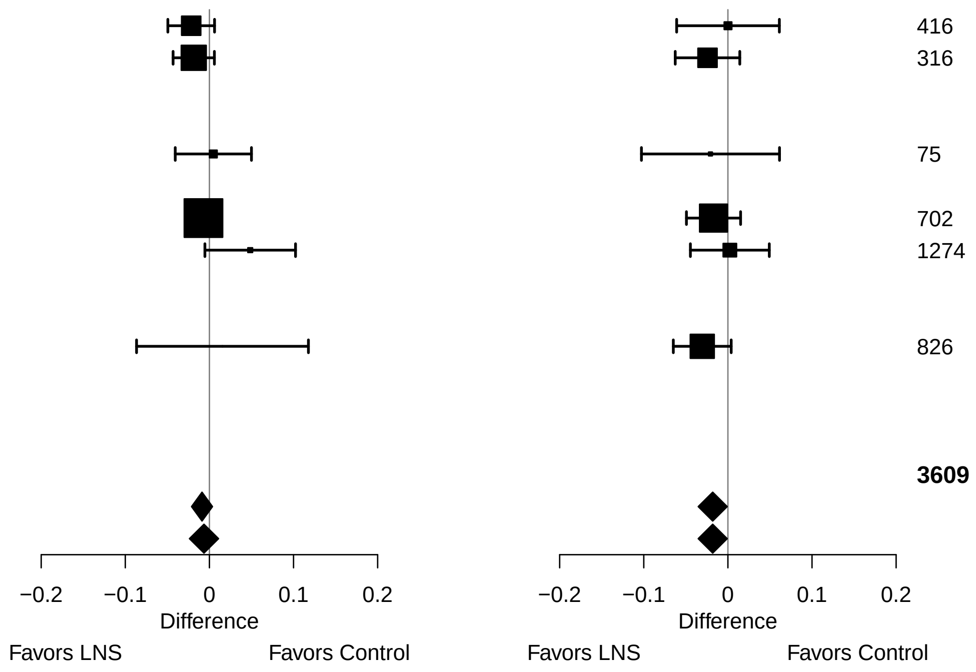

#### Supplemental figure 5F: Social-emotional lowest decile prevalence difference

#### 5F5: Stratified by Maternal depressive symptoms

**P−for−interaction = 0.525**
**Difference in PDs = −0.01 (−0.03, 0.01)**

| Country | Trial | Tool | LNS N | Control N | Control Prevalence | Less than 75th percentile PD (95% CI) | Fixed W | Random W | LNS N | Control N | Control Prevalence | At least 75th percentile PD (95% CI) | Fixed W | Random W |
|---|---|---|---|---|---|---|---|---|---|---|---|---|---|---|
| Bangladesh | JiVitA−4 (21) | | | | | | | | | | | | | |
| Bangladesh | RDNS (22) | DMC | 1075 | 472 | 10.8 | −0.03 (−0.05, 0.00) | 0.20 | 0.20 | 520 | 288 | 12.2 | 0.00 (−0.05, 0.05) | 0.16 | 0.16 |
| Bangladesh | WASH−B (23) | EASQ | 822 | 2356 | 9.6 | −0.02 (−0.04, 0.00) | 0.24 | 0.24 | 223 | 830 | 13.1 | −0.01 (−0.06, 0.03) | 0.16 | 0.16 |
| Burkina Faso | iLiNS−Zinc (24) | DMC | | | | | | | | | | | | |
| Ghana | GHANA (25) | | | | | | | | | | | | | |
| Ghana | iLiNS−DYADG (26) | PSED | 242 | 435 | 11.0 | −0.02 (−0.07, 0.02) | 0.06 | 0.06 | 78 | 201 | 9.0 | 0.03 (−0.05, 0.10) | 0.06 | 0.06 |
| Haiti | HAITI (27) | | | | | | | | | | | | | |
| Kenya | WASH−B (28) | EASQ | 960 | 3317 | 10.0 | −0.01 (−0.03, 0.02) | 0.28 | 0.28 | 317 | 1156 | 7.7 | −0.02 (−0.05, 0.01) | 0.38 | 0.38 |
| Madagascar | MAHAY (29) | ASQI | 615 | 667 | 10.5 | −0.02 (−0.07, 0.04) | 0.04 | 0.04 | 268 | 242 | 9.1 | 0.00 (−0.05, 0.06) | 0.12 | 0.12 |
| Malawi | iLiNS−DYADM (30) | PSED | 153 | 302 | 9.9 | −0.03 (−0.08, 0.03) | 0.04 | 0.04 | 47 | 111 | 9.0 | 0.08 (−0.03, 0.19) | 0.03 | 0.03 |
| Malawi | iLiNS−DOSE (31) | PSED | | | | | | | | | | | | |
| Mali | PROMIS CS (32) | DMC | | | | | | | | | | | | |
| Zimbabwe | SHINE (HIV−) (33) | MDAT | 583 | 560 | 11.1 | −0.03 (−0.07, 0.00) | 0.11 | 0.11 | 197 | 180 | 16.7 | −0.06 (−0.13, 0.01) | 0.08 | 0.08 |
| Zimbabwe | SHINE (HIV+) (34) | MDAT | 131 | 110 | 13.6 | −0.09 (−0.16, −0.02) | 0.03 | 0.03 | 33 | 32 | 15.6 | 0.00 (−0.17, 0.17) | 0.01 | 0.01 |
| | | | **4581** | **8219** | | **I² = 0.00, Tau² = 0.00** | | | **1683** | **3040** | | **I² = 0.00, Tau² = 0.00** | | |
| **Fixed** | | | | | | **−0.02 (−0.03, −0.01)** | | | | | | **−0.01 (−0.03, 0.01)** | | |
| **Random** | | | | | | **−0.02 (−0.03, −0.01)** | | | | | | **−0.01 (−0.03, 0.01)** | | |

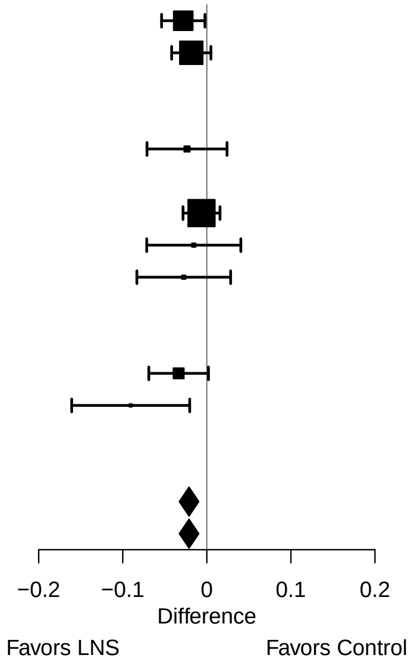

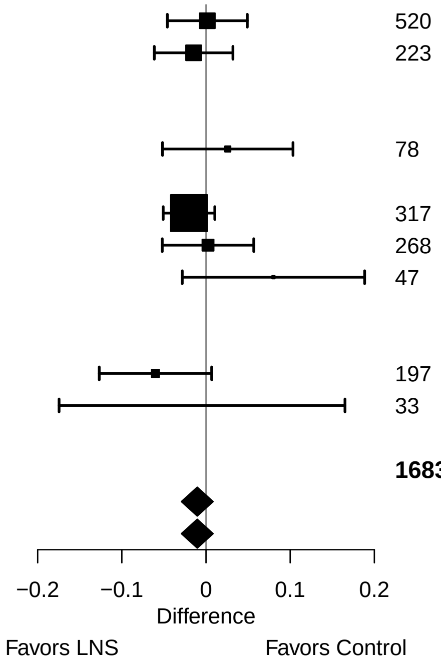

**Supplemental figure 5F: Social-emotional lowest decile prevalence difference**

#### 5F6: Stratified by Child sex

**P−for−interaction = 0.852**
**Difference in PDs = 0.00 (−0.02, 0.02)**

| Country | Trial | Tool | LNS N | Control N | Control Prevalence | Male PD (95% CI) | Fixed W | Random W | LNS N | Control N | Control Prevalence | Female PD (95% CI) | Fixed W | Random W |
|---|---|---|---|---|---|---|---|---|---|---|---|---|---|---|
| Bangladesh | JiVitA−4 (21) | | | | | | | | | | | | | |
| Bangladesh | RDNS (22) | DMC | 830 | 411 | 9.0 | 0.00 (−0.03, 0.03) | 0.15 | 0.11 | 827 | 404 | 13.9 | −0.04 (−0.08, 0.00) | 0.11 | 0.11 |
| Bangladesh | WASH−B (23) | EASQ | 522 | 1635 | 10.4 | −0.02 (−0.05, 0.01) | 0.18 | 0.12 | 544 | 1631 | 10.6 | −0.02 (−0.05, 0.02) | 0.18 | 0.13 |
| Burkina Faso | iLiNS−Zinc (24) | DMC | 372 | 193 | 13.5 | −0.08 (−0.12, −0.03) | 0.08 | 0.10 | 374 | 182 | 19.8 | −0.12 (−0.19, −0.05) | 0.03 | 0.06 |
| Ghana | GHANA (25) | | | | | | | | | | | | | |
| Ghana | iLiNS−DYADG (26) | PSED | 164 | 306 | 7.8 | 0.03 (−0.03, 0.08) | 0.05 | 0.08 | 168 | 351 | 12.0 | −0.02 (−0.08, 0.03) | 0.05 | 0.08 |
| Haiti | HAITI (27) | | | | | | | | | | | | | |
| Kenya | WASH−B (28) | EASQ | 682 | 2260 | 10.8 | 0.00 (−0.03, 0.02) | 0.23 | 0.12 | 680 | 2485 | 9.9 | −0.02 (−0.05, 0.01) | 0.24 | 0.14 |
| Madagascar | MAHAY (29) | ASQI | 795 | 787 | 11.1 | −0.01 (−0.05, 0.04) | 0.07 | 0.09 | 818 | 817 | 7.6 | 0.04 (−0.01, 0.08) | 0.08 | 0.10 |
| Malawi | iLiNS−DYADM (30) | PSED | 105 | 205 | 12.2 | −0.05 (−0.12, 0.03) | 0.03 | 0.06 | 110 | 233 | 8.6 | 0.04 (−0.03, 0.11) | 0.04 | 0.07 |
| Malawi | iLiNS−DOSE (31) | PSED | 320 | 117 | 6.8 | 0.05 (−0.01, 0.12) | 0.03 | 0.07 | 324 | 104 | 12.5 | −0.04 (−0.10, 0.03) | 0.04 | 0.07 |
| Mali | PROMIS CS (32) | DMC | 469 | 514 | 10.7 | −0.03 (−0.06, 0.01) | 0.11 | 0.11 | 458 | 430 | 12.1 | −0.03 (−0.07, 0.02) | 0.10 | 0.11 |
| Zimbabwe | SHINE (HIV−) (33) | MDAT | 424 | 388 | 13.1 | −0.04 (−0.09, 0.00) | 0.06 | 0.09 | 417 | 411 | 10.7 | −0.03 (−0.07, 0.02) | 0.10 | 0.11 |
| Zimbabwe | SHINE (HIV+) (34) | MDAT | 84 | 79 | 20.3 | −0.13 (−0.22, −0.04) | 0.02 | 0.05 | 86 | 72 | 9.7 | −0.04 (−0.14, 0.07) | 0.02 | 0.03 |
| | | | **4767** | **6895** | | **I² = 0.59, Tau² = 0.00** | | | **4806** | **7120** | | **I² = 0.43, Tau² = 0.00** | | |
| **Fixed** | | | | | | **−0.02 (−0.03, −0.01)** | | | | | | **−0.02 (−0.03, −0.01)** | | |
| **Random** | | | | | | **−0.02 (−0.05, 0.00)** | | | | | | **−0.02 (−0.04, 0.00)** | | |

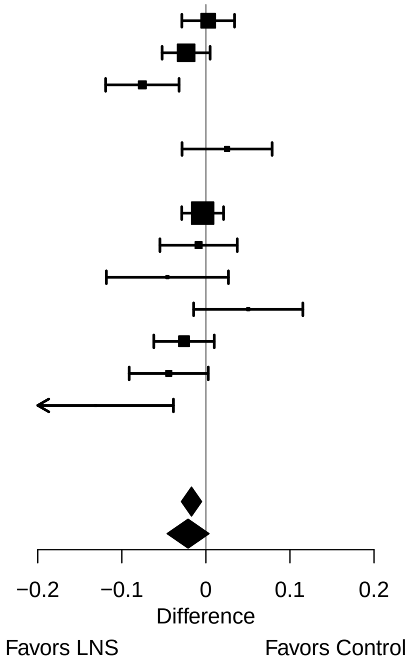

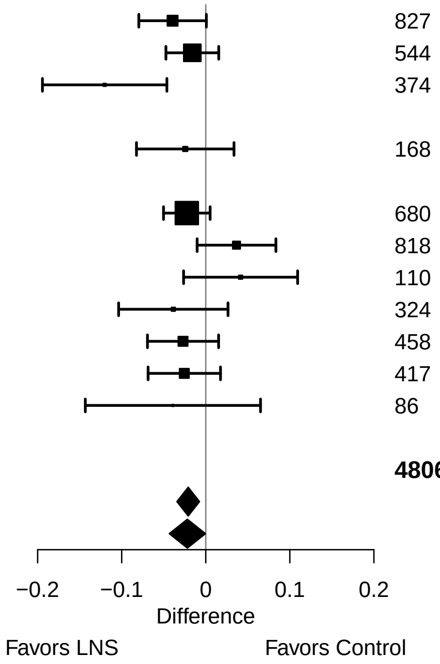

**Supplemental figure 5F: Social-emotional lowest decile prevalence difference**

#### 5F7: Stratified by Child birth order

**P−for−interaction = 0.909**
**Difference in PDs = 0.00 (−0.02, 0.02)**

| Country | Trial | Tool | Later born LNS N | Later born Control N | Later born Control Prevalence | Later born PD (95% CI) | Later born Fixed W | Later born Random W | Firstborn LNS N | Firstborn Control N | Firstborn Control Prevalence | Firstborn PD (95% CI) | Firstborn Fixed W | Firstborn Random W |
|---|---|---|---|---|---|---|---|---|---|---|---|---|---|---|
| Bangladesh | JiVitA−4 (21) | | | | | | | | | | | | | |
| Bangladesh | RDNS (22) | DMC | 969 | 508 | 12.4 | −0.02 (−0.06, 0.01) | 0.09 | 0.11 | 687 | 307 | 9.8 | −0.01 (−0.04, 0.02) | 0.27 | 0.27 |
| Bangladesh | WASH−B (23) | EASQ | 669 | 2134 | 10.8 | −0.01 (−0.04, 0.01) | 0.18 | 0.13 | 377 | 1031 | 9.0 | −0.02 (−0.05, 0.01) | 0.25 | 0.25 |
| Burkina Faso | iLiNS−Zinc (24) | DMC | 587 | 290 | 17.9 | −0.12 (−0.17, −0.07) | 0.05 | 0.09 | 159 | 85 | 11.8 | −0.02 (−0.12, 0.07) | 0.03 | 0.03 |
| Ghana | GHANA (25) | | | | | | | | | | | | | |
| Ghana | iLiNS−DYADG (26) | PSED | 225 | 444 | 9.7 | 0.02 (−0.03, 0.07) | 0.05 | 0.10 | 107 | 213 | 10.8 | −0.05 (−0.12, 0.01) | 0.06 | 0.06 |
| Haiti | HAITI (27) | | | | | | | | | | | | | |
| Kenya | WASH−B (28) | EASQ | 1071 | 3767 | 10.4 | −0.01 (−0.03, 0.01) | 0.29 | 0.14 | 289 | 974 | 10.1 | −0.02 (−0.06, 0.02) | 0.18 | 0.18 |
| Madagascar | MAHAY (29) | ASQI | 1162 | 1118 | 8.9 | 0.01 (−0.03, 0.06) | 0.07 | 0.10 | 433 | 465 | 10.8 | 0.01 (−0.06, 0.07) | 0.06 | 0.06 |
| Malawi | iLiNS−DYADM (30) | PSED | 170 | 349 | 10.9 | −0.02 (−0.08, 0.03) | 0.04 | 0.09 | 45 | 87 | 8.0 | 0.08 (−0.04, 0.19) | 0.02 | 0.02 |
| Malawi | iLiNS−DOSE (31) | PSED | | | | | | | | | | | | |
| Mali | PROMIS CS (32) | DMC | 770 | 811 | 11.2 | −0.03 (−0.06, 0.01) | 0.12 | 0.12 | 142 | 122 | 9.0 | 0.01 (−0.06, 0.08) | 0.05 | 0.05 |
| Zimbabwe | SHINE (HIV−) (33) | MDAT | 602 | 574 | 11.5 | −0.03 (−0.07, 0.00) | 0.11 | 0.12 | 203 | 191 | 14.7 | −0.06 (−0.12, 0.00) | 0.07 | 0.07 |
| Zimbabwe | SHINE (HIV+) (34) | MDAT | | | | | | | | | | | | |
| | | | **6225** | **9995** | | **$I^2$ = 0.61, $Tau^2$ = 0.00** | | | **2442** | **3475** | | **$I^2$ = 0.00, $Tau^2$ = 0.00** | | |
| **Fixed** | | | | | | **−0.02 (−0.03, −0.01)** | | | | | | **−0.02 (−0.03, 0.00)** | | |
| **Random** | | | | | | **−0.02 (−0.05, 0.00)** | | | | | | **−0.02 (−0.03, 0.00)** | | |

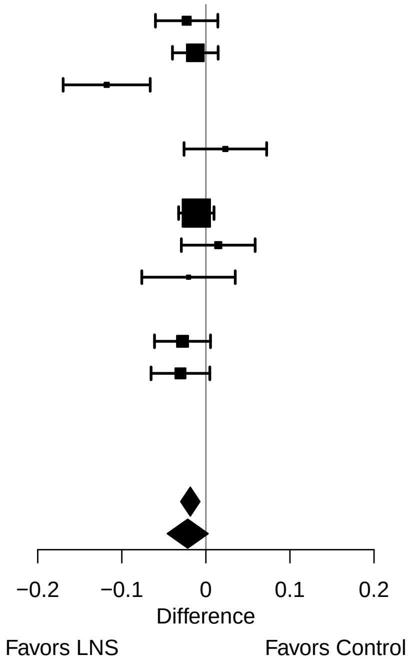

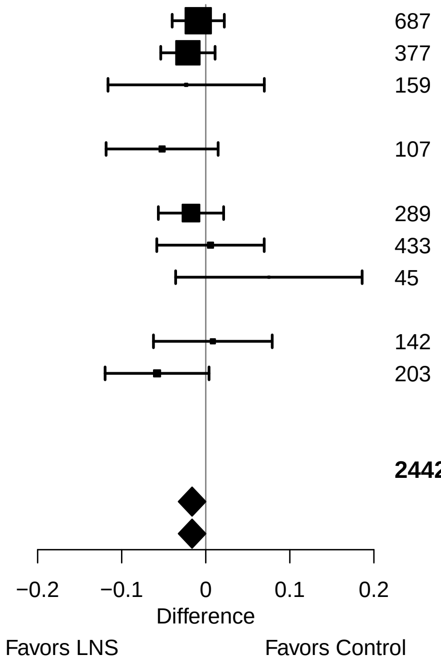

#### Supplemental figure 5F: Social-emotional lowest decile prevalence difference

##### 5F8: Stratified by Child baseline stunting

**P−for−interaction = 0.202**
**Difference in PDs = −0.03 (−0.08, 0.02)**

| Country | Trial | Tool | No: LNS N | No: Control N | No: Control Prevalence | No: PD (95% CI) | No: Fixed W | No: Random W | Yes: LNS N | Yes: Control N | Yes: Control Prevalence | Yes: PD (95% CI) | Yes: Fixed W | Yes: Random W |
|---|---|---|---|---|---|---|---|---|---|---|---|---|---|---|
| Bangladesh | JiVitA−4 (21) | | | | | | | | | | | | | |
| Bangladesh | RDNS (22) | DMC | 1241 | 574 | 8.9 | 0.00 (−0.03, 0.02) | 0.48 | 0.31 | 354 | 185 | 18.9 | −0.06 (−0.13, 0.01) | 0.36 | 0.28 |
| Bangladesh | WASH−B (23) | EASQ | | | | | | | | | | | | |
| Burkina Faso | iLiNS−Zinc (24) | DMC | 576 | 294 | 13.3 | −0.08 (−0.13, −0.03) | 0.14 | 0.22 | 170 | 80 | 27.5 | −0.16 (−0.25, −0.06) | 0.19 | 0.23 |
| Ghana | GHANA (25) | | | | | | | | | | | | | |
| Ghana | iLiNS−DYADG (26) | PSED | | | | | | | | | | | | |
| Haiti | HAITI (27) | | | | | | | | | | | | | |
| Kenya | WASH−B (28) | EASQ | | | | | | | | | | | | |
| Madagascar | MAHAY (29) | ASQI | | | | | | | | | | | | |
| Malawi | iLiNS−DYADM (30) | PSED | | | | | | | | | | | | |
| Malawi | iLiNS−DOSE (31) | PSED | 464 | 147 | 8.8 | 0.01 (−0.05, 0.06) | 0.12 | 0.20 | 180 | 74 | 10.8 | 0.01 (−0.08, 0.09) | 0.23 | 0.24 |
| Mali | PROMIS CS (32) | DMC | | | | | | | | | | | | |
| Zimbabwe | SHINE (HIV−) (33) | MDAT | 532 | 490 | 10.8 | −0.04 (−0.08, −0.01) | 0.26 | 0.27 | 122 | 105 | 14.3 | −0.02 (−0.11, 0.07) | 0.22 | 0.24 |
| Zimbabwe | SHINE (HIV+) (34) | MDAT | | | | | | | | | | | | |
| | | | **2813** | **1505** | | **I² = 0.68, Tau² = 0.00** | | | **826** | **444** | | **I² = 0.59, Tau² = 0.00** | | |
| **Fixed** | | | | | | **−0.02 (−0.04, −0.01)** | | | | | | **−0.05 (−0.10, −0.01)** | | |
| **Random** | | | | | | **−0.03 (−0.07, 0.01)** | | | | | | **−0.06 (−0.13, 0.01)** | | |

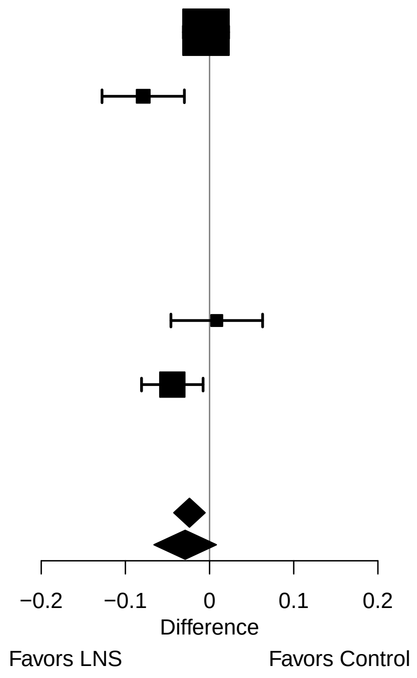

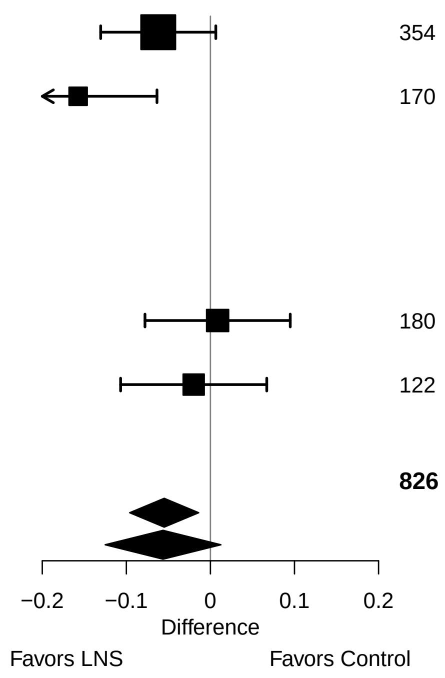

**Supplemental figure 5F: Social-emotional lowest decile prevalence difference**

#### 5F9: Stratified by Child baseline acute malnutrition (insufficient comparisons)

#### Supplemental figure 5F: Social-emotional lowest decile prevalence difference

### 5F10: Stratified by Child baseline anemia

**P−for−interaction = 0.952**
**Difference in PDs = 0.00 (−0.05, 0.05)**

| Country | Trial | Tool | LNS N | Control N | Control Prevalence | Not anemic PD (95% CI) | Fixed W | Random W | LNS N | Control N | Control Prevalence | Anemic PD (95% CI) | Fixed W | Random W |
|---|---|---|---|---|---|---|---|---|---|---|---|---|---|---|
| Bangladesh | JiVitA−4 (21) | | | | | | | | | | | | | |
| Bangladesh | RDNS (22) | DMC | 212 | 96 | 14.6 | −0.08 (−0.16, 0.01) | 0.17 | 0.19 | 316 | 160 | 12.5 | −0.03 (−0.08, 0.02) | 0.35 | 0.35 |
| Bangladesh | WASH−B (23) | EASQ | | | | | | | | | | | | |
| Burkina Faso | iLiNS−Zinc (24) | DMC | | | | | | | | | | | | |
| Ghana | GHANA (25) | | | | | | | | | | | | | |
| Ghana | iLiNS−DYADG (26) | PSED | 193 | 350 | 10.3 | 0.00 (−0.05, 0.05) | 0.40 | 0.37 | 90 | 204 | 9.8 | −0.01 (−0.08, 0.06) | 0.18 | 0.18 |
| Haiti | HAITI (27) | | | | | | | | | | | | | |
| Kenya | WASH−B (28) | EASQ | | | | | | | | | | | | |
| Madagascar | MAHAY (29) | ASQI | | | | | | | | | | | | |
| Malawi | iLiNS−DYADM (30) | PSED | 66 | 147 | 8.2 | −0.01 (−0.08, 0.07) | 0.19 | 0.20 | 138 | 275 | 10.9 | 0.01 (−0.06, 0.07) | 0.23 | 0.23 |
| Malawi | iLiNS−DOSE (31) | PSED | 223 | 94 | 7.4 | 0.02 (−0.05, 0.09) | 0.24 | 0.25 | 417 | 126 | 11.1 | −0.01 (−0.07, 0.05) | 0.25 | 0.25 |
| Mali | PROMIS CS (32) | DMC | | | | | | | | | | | | |
| Zimbabwe | SHINE (HIV−) (33) | MDAT | | | | | | | | | | | | |
| Zimbabwe | SHINE (HIV+) (34) | MDAT | | | | | | | | | | | | |
| | | | **694** | **687** | | **I² = 0.14, Tau² = 0.00** | | | **961** | **765** | | **I² = 0.00, Tau² = 0.00** | | |
| **Fixed** | | | | | | **−0.01 (−0.04, 0.03)** | | | | | | **−0.01 (−0.04, 0.02)** | | |
| **Random** | | | | | | **−0.01 (−0.05, 0.03)** | | | | | | **−0.01 (−0.04, 0.02)** | | |

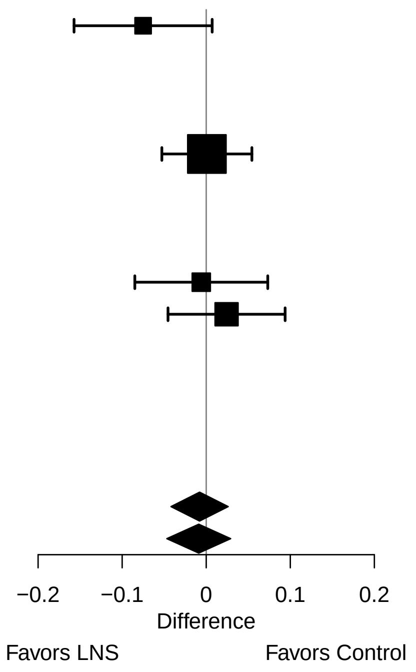

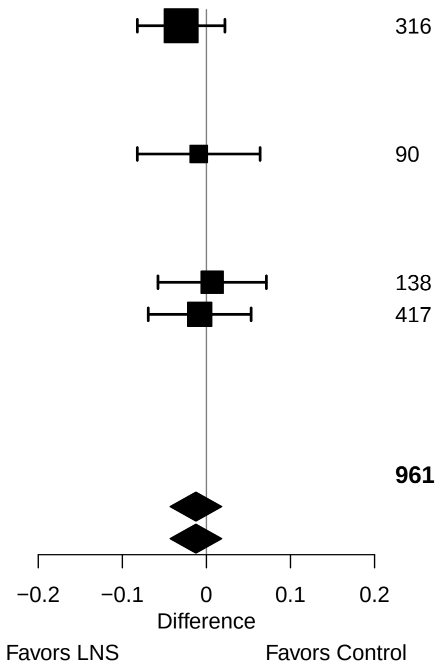

### Supplemental figure 5G: Mean difference in motor z-score

#### 5G1: Stratified by Maternal height

**P−for−interaction = 0.457**
**Difference in MDs = −0.03 (−0.10, 0.05)**

| Country | Trial | Tool | LNS N | Control N | Control Mean | At least 150.1 cm MD (95% CI) | Fixed W | Random W | LNS N | Control N | Control Mean | Less than 150.1 cm MD (95% CI) | Fixed W | Random W |
|---|---|---|---|---|---|---|---|---|---|---|---|---|---|---|
| Bangladesh | JiVitA−4 (21) | BSID−III | | | | | | | | | | | | |
| Bangladesh | RDNS (22) | DMC | 818 | 399 | −0.04 | 0.16 (0.08, 0.24) | 0.20 | 0.12 | 684 | 333 | −0.06 | 0.04 (−0.06, 0.15) | 0.36 | 0.36 |
| Bangladesh | WASH−B (23) | EASQ | 568 | 1773 | 0.04 | 0.09 (−0.02, 0.19) | 0.12 | 0.11 | 501 | 1467 | −0.09 | 0.13 (0.03, 0.23) | 0.42 | 0.42 |
| Burkina Faso | iLiNS−Zinc (24) | DMC | 727 | 365 | −0.25 | 0.40 (0.23, 0.57) | 0.04 | 0.08 | 17 | 10 | −0.63 | 0.39 (−0.48, 1.26) | 0.01 | 0.01 |
| Ghana | GHANA (25) | | | | | | | | | | | | | |
| Ghana | iLiNS−DYADG (26) | KDI | 284 | 557 | 0.02 | 0.01 (−0.13, 0.14) | 0.07 | 0.09 | 13 | 34 | −0.46 | 0.30 (−0.62, 1.23) | 0.00 | 0.00 |
| Haiti | HAITI (27) | | | | | | | | | | | | | |
| Kenya | WASH−B (28) | EASQ | 1237 | 4372 | 0.01 | 0.00 (−0.07, 0.06) | 0.29 | 0.13 | 51 | 175 | −0.21 | 0.27 (−0.01, 0.56) | 0.05 | 0.05 |
| Madagascar | MAHAY (29) | ASQI | 956 | 986 | −0.05 | 0.09 (−0.13, 0.30) | 0.03 | 0.06 | 553 | 538 | 0.06 | 0.03 (−0.19, 0.25) | 0.08 | 0.08 |
| Malawi | iLiNS−DYADM (30) | KDI | 186 | 376 | −0.03 | 0.04 (−0.13, 0.20) | 0.04 | 0.08 | 28 | 57 | 0.16 | −0.04 (−0.49, 0.42) | 0.02 | 0.02 |
| Malawi | iLiNS−DOSE (31) | KDI | 543 | 185 | −0.01 | 0.04 (−0.13, 0.20) | 0.04 | 0.08 | 102 | 35 | −0.17 | 0.10 (−0.30, 0.50) | 0.03 | 0.03 |
| Mali | PROMIS CS (32) | DMC | 865 | 889 | −0.07 | 0.14 (−0.03, 0.31) | 0.04 | 0.08 | 32 | 26 | −0.01 | −0.04 (−0.60, 0.53) | 0.01 | 0.01 |
| Zimbabwe | SHINE (HIV−) (33) | MDAT | 788 | 762 | −0.06 | 0.11 (0.00, 0.22) | 0.10 | 0.11 | 34 | 22 | −0.15 | 0.11 (−0.35, 0.57) | 0.02 | 0.02 |
| Zimbabwe | SHINE (HIV+) (34) | MDAT | 158 | 140 | −0.14 | 0.28 (0.06, 0.51) | 0.02 | 0.06 | 10 | 6 | −0.36 | 0.42 (−0.76, 1.60) | 0.00 | 0.00 |
| | | | **7130** | **10804** | | **$I^2$ = 0.67, $Tau^2$ = 0.01** | | | **2025** | **2703** | | **$I^2$ = 0.00, $Tau^2$ = 0.00** | | |
| **Fixed** | | | | | | **0.09 (0.05, 0.12)** | | | | | | **0.09 (0.03, 0.16)** | | |
| **Random** | | | | | | **0.11 (0.04, 0.18)** | | | | | | **0.09 (0.03, 0.16)** | | |

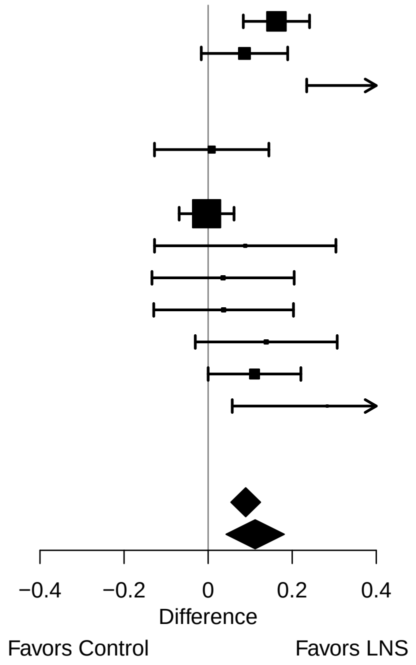

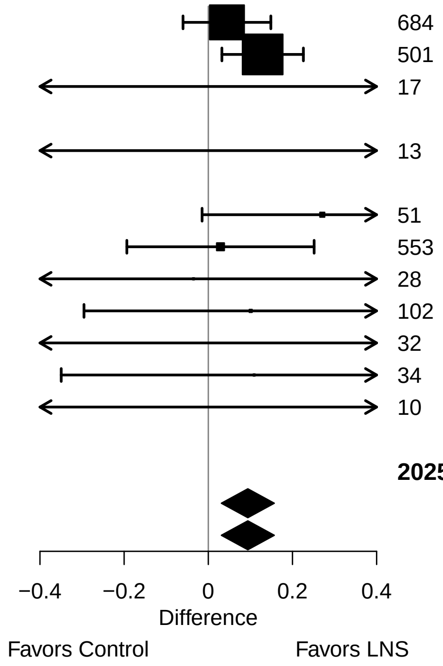

#### Supplemental figure 5G: Mean difference in motor z-score

#### 5G2: Stratified by Maternal BMI

**P−for−interaction = 0.553**
**Difference in MDs = 0.02 (−0.05, 0.09)**

| Country | Trial | Tool | LNS N | Control N | Control Mean | At least 20 kg/m² MD (95% CI) | Fixed W | Random W | LNS N | Control N | Control Mean | Less than 20 kg/m² MD (95% CI) | Fixed W | Random W |
|---|---|---|---|---|---|---|---|---|---|---|---|---|---|---|
| Bangladesh | JiVitA−4 (21) | BSID−III | | | | | | | | | | | | |
| Bangladesh | RDNS (22) | DMC | 689 | 306 | −0.01 | 0.10 (−0.01, 0.20) | 0.13 | 0.12 | 813 | 426 | −0.08 | 0.11 (0.03, 0.20) | 0.36 | 0.20 |
| Bangladesh | WASH−B (23) | EASQ | 478 | 1502 | 0.05 | 0.09 (−0.02, 0.19) | 0.14 | 0.13 | 591 | 1737 | −0.09 | 0.12 (0.03, 0.22) | 0.28 | 0.19 |
| Burkina Faso | iLiNS−Zinc (24) | DMC | 480 | 225 | −0.13 | 0.33 (0.14, 0.52) | 0.05 | 0.08 | 264 | 150 | −0.46 | 0.49 (0.23, 0.75) | 0.04 | 0.08 |
| Ghana | GHANA (25) | | | | | | | | | | | | | |
| Ghana | iLiNS−DYADG (26) | KDI | 262 | 485 | 0.00 | 0.04 (−0.10, 0.19) | 0.07 | 0.10 | 35 | 106 | −0.05 | −0.10 (−0.50, 0.29) | 0.02 | 0.04 |
| Haiti | HAITI (27) | | | | | | | | | | | | | |
| Kenya | WASH−B (28) | EASQ | 1031 | 3535 | 0.01 | −0.02 (−0.09, 0.05) | 0.33 | 0.15 | 257 | 1012 | −0.04 | 0.12 (−0.02, 0.27) | 0.13 | 0.15 |
| Madagascar | MAHAY (29) | ASQI | | | | | | | | | | | | |
| Malawi | iLiNS−DYADM (30) | KDI | 126 | 258 | −0.04 | 0.06 (−0.16, 0.28) | 0.03 | 0.07 | 87 | 175 | 0.05 | −0.03 (−0.24, 0.19) | 0.05 | 0.10 |
| Malawi | iLiNS−DOSE (31) | KDI | 480 | 158 | 0.01 | 0.02 (−0.16, 0.19) | 0.05 | 0.09 | 163 | 61 | −0.16 | 0.13 (−0.19, 0.45) | 0.03 | 0.06 |
| Mali | PROMIS CS (32) | DMC | 665 | 653 | −0.08 | 0.18 (0.00, 0.36) | 0.05 | 0.09 | 232 | 262 | −0.04 | 0.00 (−0.22, 0.22) | 0.05 | 0.10 |
| Zimbabwe | SHINE (HIV−) (33) | MDAT | 601 | 581 | −0.05 | 0.13 (0.01, 0.25) | 0.11 | 0.12 | 100 | 96 | −0.22 | 0.21 (−0.08, 0.51) | 0.03 | 0.07 |
| Zimbabwe | SHINE (HIV+) (34) | MDAT | 124 | 113 | −0.11 | 0.36 (0.13, 0.60) | 0.03 | 0.06 | 25 | 23 | −0.31 | −0.13 (−0.68, 0.42) | 0.01 | 0.02 |
| | | | **4936** | **7816** | | **I² = 0.62, Tau² = 0.01** | | | **2567** | **4048** | | **I² = 0.31, Tau² = 0.01** | | |
| **Fixed** | | | | | | **0.07 (0.03, 0.11)** | | | | | | **0.12 (0.06, 0.17)** | | |
| **Random** | | | | | | **0.11 (0.04, 0.18)** | | | | | | **0.11 (0.03, 0.20)** | | |

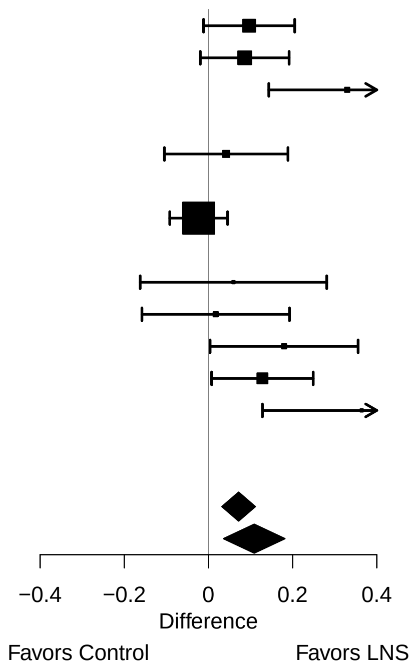

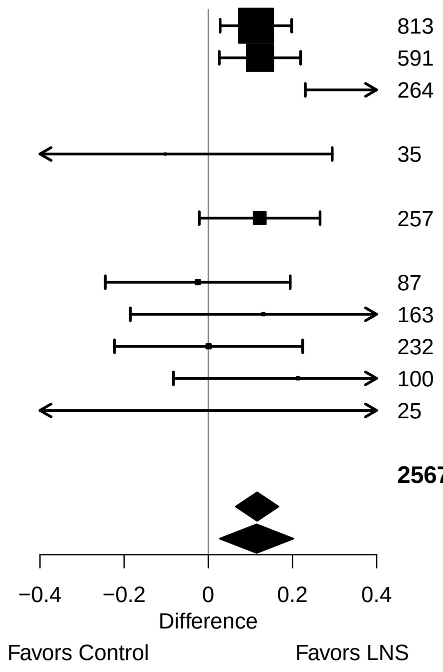

#### Supplemental figure 5G: Mean difference in motor z-score

#### 5G3: Stratified by Maternal age

**P−for−interaction = 0.043**
**Difference in MDs = −0.06 (−0.11, 0.00)**

**At least 25 y**

| Country | Trial | Tool | LNS N | Control N | Control Mean | MD (95% CI) | Fixed W | Random W |
|---|---|---|---|---|---|---|---|---|
| Bangladesh | JiVitA−4 (21) | BSID−III | 200 | 68 | 0.06 | −0.06 (−0.27, 0.15) | 0.04 | 0.07 |
| Bangladesh | RDNS (22) | DMC | 424 | 200 | −0.11 | 0.15 (0.00, 0.30) | 0.08 | 0.09 |
| Bangladesh | WASH−B (23) | EASQ | 466 | 1450 | −0.02 | 0.09 (−0.03, 0.20) | 0.14 | 0.10 |
| Burkina Faso | iLiNS−Zinc (24) | DMC | 421 | 232 | −0.33 | 0.53 (0.36, 0.70) | 0.06 | 0.08 |
| Ghana | GHANA (25) | | | | | | | |
| Ghana | iLiNS−DYADG (26) | KDI | 192 | 378 | −0.01 | 0.11 (−0.06, 0.27) | 0.07 | 0.09 |
| Haiti | HAITI (27) | | | | | | | |
| Kenya | WASH−B (28) | EASQ | 798 | 2622 | 0.04 | −0.02 (−0.10, 0.05) | 0.30 | 0.11 |
| Madagascar | MAHAY (29) | ASQI | 852 | 901 | −0.09 | 0.08 (−0.12, 0.29) | 0.04 | 0.07 |
| Malawi | iLiNS−DYADM (30) | KDI | 102 | 224 | 0.01 | 0.12 (−0.10, 0.34) | 0.04 | 0.07 |
| Malawi | iLiNS−DOSE (31) | KDI | 346 | 124 | −0.08 | 0.09 (−0.12, 0.30) | 0.04 | 0.07 |
| Mali | PROMIS CS (32) | DMC | 480 | 479 | −0.08 | 0.21 (0.04, 0.38) | 0.06 | 0.08 |
| Zimbabwe | SHINE (HIV−) (33) | MDAT | 427 | 389 | −0.03 | 0.18 (0.03, 0.32) | 0.09 | 0.09 |
| Zimbabwe | SHINE (HIV+) (34) | MDAT | 126 | 111 | −0.12 | 0.28 (0.03, 0.52) | 0.03 | 0.06 |
| | | | **4834** | **7178** | | **I² = 0.74, Tau² = 0.02** | | |
| **Fixed** | | | | | | **0.10 (0.06, 0.15)** | | |
| **Random** | | | | | | **0.14 (0.06, 0.23)** | | |

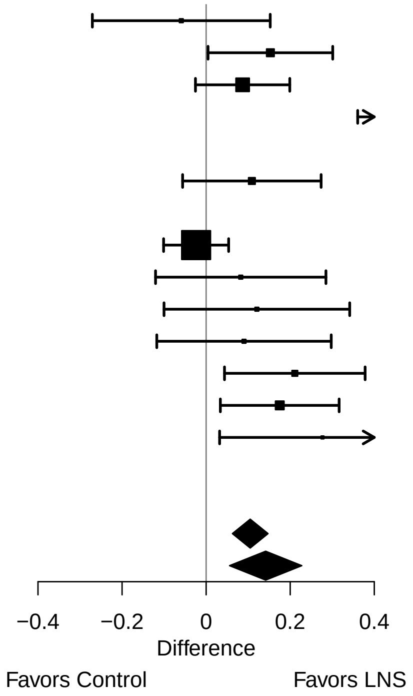

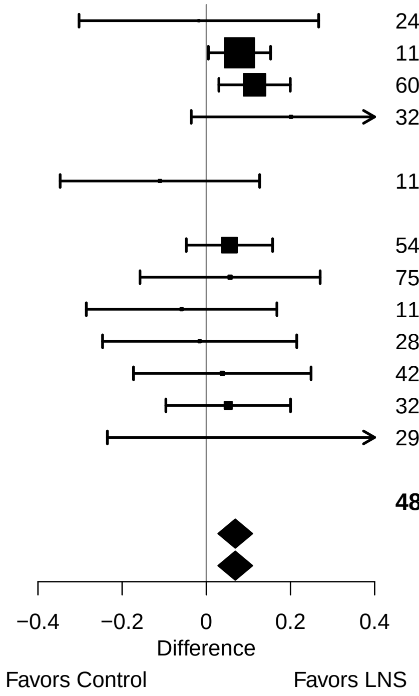

**Less than 25 y**

| Country | Trial | LNS N | Control N | Control Mean | MD (95% CI) | Fixed W | Random W |
|---|---|---|---|---|---|---|---|
| Bangladesh | JiVitA−4 (21) | 242 | 74 | 0.01 | −0.02 (−0.30, 0.27) | 0.02 | 0.02 |
| Bangladesh | RDNS (22) | 1132 | 553 | −0.02 | 0.08 (0.00, 0.15) | 0.31 | 0.31 |
| Bangladesh | WASH−B (23) | 603 | 1815 | −0.03 | 0.11 (0.03, 0.20) | 0.23 | 0.23 |
| Burkina Faso | iLiNS−Zinc (24) | 321 | 141 | −0.14 | 0.20 (−0.04, 0.44) | 0.03 | 0.03 |
| Ghana | GHANA (25) | | | | | | |
| Ghana | iLiNS−DYADG (26) | 110 | 223 | 0.01 | −0.11 (−0.35, 0.13) | 0.03 | 0.03 |
| Haiti | HAITI (27) | | | | | | |
| Kenya | WASH−B (28) | 549 | 2092 | −0.05 | 0.05 (−0.05, 0.16) | 0.16 | 0.16 |
| Madagascar | MAHAY (29) | 759 | 703 | 0.04 | 0.06 (−0.16, 0.27) | 0.04 | 0.04 |
| Malawi | iLiNS−DYADM (30) | 112 | 212 | −0.01 | −0.06 (−0.29, 0.17) | 0.03 | 0.03 |
| Malawi | iLiNS−DOSE (31) | 288 | 90 | 0.04 | −0.02 (−0.25, 0.21) | 0.03 | 0.03 |
| Mali | PROMIS CS (32) | 422 | 442 | −0.05 | 0.04 (−0.17, 0.25) | 0.04 | 0.04 |
| Zimbabwe | SHINE (HIV−) (33) | 326 | 321 | −0.11 | 0.05 (−0.10, 0.20) | 0.08 | 0.08 |
| Zimbabwe | SHINE (HIV+) (34) | 29 | 26 | −0.20 | 0.40 (−0.24, 1.03) | 0.00 | 0.00 |
| | | **4893** | **6692** | | **I² = 0.00, Tau² = 0.00** | | |
| **Fixed** | | | | | **0.07 (0.03, 0.11)** | | |
| **Random** | | | | | **0.07 (0.03, 0.11)** | | |

#### Supplemental figure 5G: Mean difference in motor z-score

##### 5G4: Stratified by Maternal education

**P−for−interaction = 0.015**
**Difference in MDs = 0.09 (0.02, 0.16)**

| | | | Primary or greater | | | | | | | Incomplete or no formal | | | | |
|---|---|---|---|---|---|---|---|---|---|---|---|---|---|---|
| **Country** | **Trial** | **Tool** | **LNS N** | **Control N** | **Control Mean** | **MD (95% CI)** | **Fixed W** | **Random W** | **LNS N** | **Control N** | **Control Mean** | **MD (95% CI)** | **Fixed W** | **Random W** |
| Bangladesh | JiVitA−4 (21) | BSID−III | 292 | 92 | 0.20 | −0.14 (−0.32, 0.05) | 0.04 | 0.08 | 152 | 51 | −0.31 | 0.18 (−0.14, 0.51) | 0.02 | 0.05 |
| Bangladesh | RDNS (22) | DMC | 1166 | 547 | 0.03 | 0.08 (0.02, 0.15) | 0.29 | 0.14 | 390 | 206 | −0.24 | 0.12 (−0.05, 0.28) | 0.09 | 0.11 |
| Bangladesh | WASH−B (23) | EASQ | 755 | 2344 | 0.04 | 0.08 (0.00, 0.17) | 0.21 | 0.14 | 319 | 935 | −0.17 | 0.16 (0.04, 0.29) | 0.16 | 0.13 |
| Burkina Faso | iLiNS−Zinc (24) | DMC | 32 | 9 | −0.22 | 0.41 (−0.18, 1.00) | 0.00 | 0.01 | 712 | 366 | −0.26 | 0.40 (0.23, 0.57) | 0.09 | 0.10 |
| Ghana | GHANA (25) | | | | | | | | | | | | | |
| Ghana | iLiNS−DYADG (26) | KDI | 235 | 467 | 0.02 | −0.02 (−0.18, 0.13) | 0.06 | 0.10 | 67 | 134 | −0.09 | 0.22 (−0.09, 0.53) | 0.03 | 0.05 |
| Haiti | HAITI (27) | | | | | | | | | | | | | |
| Kenya | WASH−B (28) | EASQ | 658 | 2258 | 0.11 | −0.01 (−0.10, 0.08) | 0.17 | 0.13 | 702 | 2484 | −0.11 | 0.02 (−0.07, 0.12) | 0.29 | 0.15 |
| Madagascar | MAHAY (29) | ASQI | 339 | 416 | 0.27 | −0.13 (−0.33, 0.08) | 0.03 | 0.07 | 1274 | 1188 | −0.14 | 0.15 (−0.05, 0.36) | 0.06 | 0.09 |
| Malawi | iLiNS−DYADM (30) | KDI | 33 | 67 | 0.06 | −0.09 (−0.42, 0.25) | 0.01 | 0.04 | 180 | 366 | −0.02 | 0.04 (−0.14, 0.22) | 0.08 | 0.10 |
| Malawi | iLiNS−DOSE (31) | KDI | 148 | 52 | 0.24 | 0.00 (−0.26, 0.27) | 0.02 | 0.05 | 487 | 162 | −0.12 | 0.06 (−0.12, 0.25) | 0.08 | 0.10 |
| Mali | PROMIS CS (32) | DMC | 99 | 93 | 0.06 | 0.09 (−0.28, 0.46) | 0.01 | 0.03 | 802 | 828 | −0.08 | 0.13 (−0.03, 0.30) | 0.09 | 0.11 |
| Zimbabwe | SHINE (HIV−) (33) | MDAT | 763 | 731 | −0.06 | 0.13 (0.02, 0.24) | 0.12 | 0.12 | 29 | 25 | 0.05 | −0.43 (−0.96, 0.09) | 0.01 | 0.02 |
| Zimbabwe | SHINE (HIV+) (34) | MDAT | 149 | 130 | −0.13 | 0.34 (0.13, 0.55) | 0.03 | 0.07 | 9 | 8 | −0.06 | −0.60 (−1.94, 0.74) | 0.00 | 0.00 |
| | | | **4669** | **7206** | | **I² = 0.50, Tau² = 0.01** | | | **5123** | **6753** | | **I² = 0.49, Tau² = 0.01** | | |
| **Fixed** | | | | | | **0.06 (0.02, 0.09)** | | | | | | **0.11 (0.06, 0.16)** | | |
| **Random** | | | | | | **0.05 (−0.03, 0.12)** | | | | | | **0.12 (0.04, 0.21)** | | |

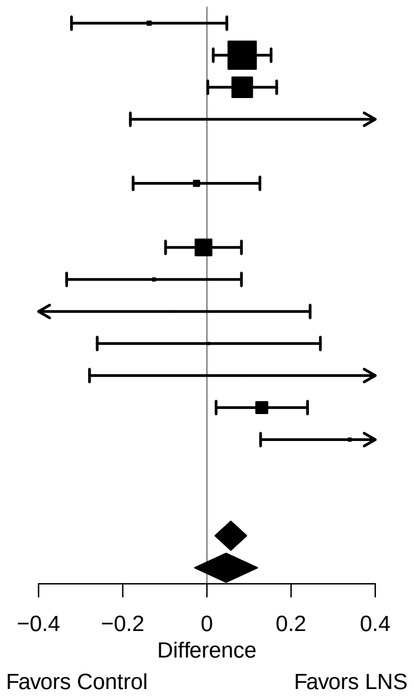

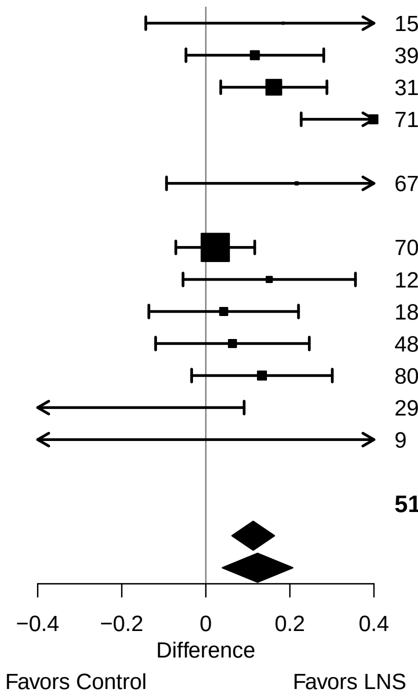

#### Supplemental figure 5G: Mean difference in motor z-score

#### 5G5: Stratified by Maternal depressive symptoms

**P−for−interaction = 0.823**
**Difference in MDs = −0.01 (−0.07, 0.06)**

| | | | Less than 75th percentile | | | | | | At least 75th percentile | | | | | |
|---|---|---|---|---|---|---|---|---|---|---|---|---|---|---|
| **Country** | **Trial** | **Tool** | **LNS N** | **Control N** | **Control Mean** | **MD (95% CI)** | **Fixed W** | **Random W** | **LNS N** | **Control N** | **Control Mean** | **MD (95% CI)** | **Fixed W** | **Random W** |
| Bangladesh | JiVitA−4 (21) | BSID−III | | | | | | | | | | | | |
| Bangladesh | RDNS (22) | DMC | 1008 | 435 | −0.03 | 0.12 (0.02, 0.23) | 0.15 | 0.16 | 489 | 267 | −0.08 | 0.06 (−0.04, 0.16) | 0.33 | 0.32 |
| Bangladesh | WASH−B (23) | EASQ | 832 | 2358 | −0.01 | 0.11 (0.04, 0.19) | 0.30 | 0.20 | 221 | 838 | −0.08 | 0.07 (−0.07, 0.22) | 0.15 | 0.15 |
| Burkina Faso | iLiNS−Zinc (24) | DMC | | | | | | | | | | | | |
| Ghana | GHANA (25) | | | | | | | | | | | | | |
| Ghana | iLiNS−DYADG (26) | KDI | 226 | 393 | 0.00 | −0.05 (−0.21, 0.12) | 0.06 | 0.10 | 67 | 190 | −0.03 | 0.30 (0.04, 0.57) | 0.05 | 0.05 |
| Haiti | HAITI (27) | | | | | | | | | | | | | |
| Kenya | WASH−B (28) | EASQ | 960 | 3317 | 0.04 | −0.02 (−0.09, 0.06) | 0.30 | 0.20 | 317 | 1156 | 0.09 | 0.03 (−0.08, 0.13) | 0.32 | 0.31 |
| Madagascar | MAHAY (29) | ASQI | 615 | 667 | 0.02 | 0.11 (−0.13, 0.34) | 0.03 | 0.06 | 268 | 242 | −0.08 | 0.11 (−0.22, 0.43) | 0.03 | 0.03 |
| Malawi | iLiNS−DYADM (30) | KDI | 152 | 299 | −0.04 | 0.07 (−0.11, 0.26) | 0.04 | 0.09 | 47 | 112 | 0.07 | −0.14 (−0.48, 0.21) | 0.03 | 0.03 |
| Malawi | iLiNS−DOSE (31) | KDI | | | | | | | | | | | | |
| Mali | PROMIS CS (32) | DMC | | | | | | | | | | | | |
| Zimbabwe | SHINE (HIV−) (33) | MDAT | 583 | 560 | −0.04 | 0.08 (−0.05, 0.21) | 0.10 | 0.13 | 197 | 180 | −0.10 | 0.18 (−0.01, 0.38) | 0.09 | 0.09 |
| Zimbabwe | SHINE (HIV+) (34) | MDAT | 131 | 110 | −0.19 | 0.37 (0.10, 0.64) | 0.02 | 0.05 | 33 | 32 | 0.07 | −0.14 (−0.65, 0.37) | 0.01 | 0.01 |
| | | | **4507** | **8139** | | **I² = 0.52, Tau² = 0.00** | | | **1639** | **3017** | | **I² = 0.02, Tau² = 0.00** | | |
| **Fixed** | | | | | | **0.07 (0.03, 0.11)** | | | | | | **0.07 (0.01, 0.12)** | | |
| **Random** | | | | | | **0.08 (0.01, 0.15)** | | | | | | **0.07 (0.01, 0.13)** | | |

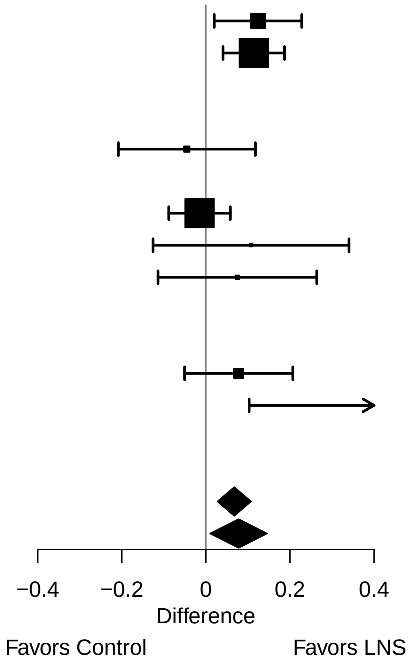

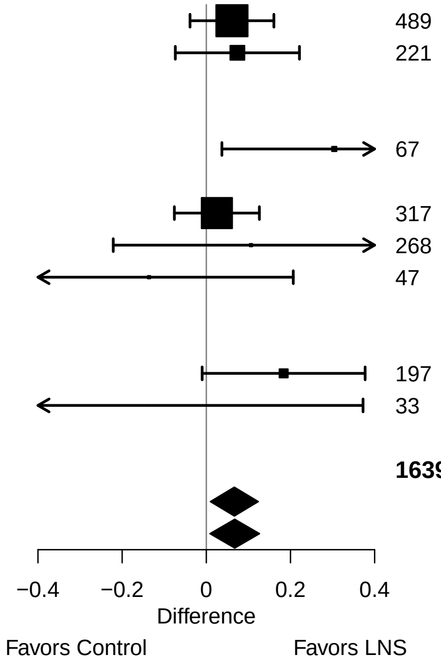

#### Supplemental figure 5G: Mean difference in motor z-score

#### 5G6: Stratified by Child sex

**P−for−interaction = 0.754**
**Difference in MDs = 0.01 (−0.04, 0.06)**

| Country | Trial | Tool | Male LNS N | Male Control N | Male Control Mean | Male MD (95% CI) | Male Fixed W | Male Random W | Female LNS N | Female Control N | Female Control Mean | Female MD (95% CI) | Female Fixed W | Female Random W |
|---|---|---|---|---|---|---|---|---|---|---|---|---|---|---|
| Bangladesh | JiVitA−4 (21) | BSID−III | 231 | 70 | 0.14 | −0.18 (−0.39, 0.03) | 0.04 | 0.06 | 214 | 73 | −0.10 | 0.14 (−0.06, 0.34) | 0.04 | 0.08 |
| Bangladesh | RDNS (22) | DMC | 782 | 386 | −0.02 | 0.05 (−0.04, 0.14) | 0.21 | 0.14 | 774 | 367 | −0.06 | 0.15 (0.06, 0.24) | 0.22 | 0.11 |
| Bangladesh | WASH−B (23) | EASQ | 528 | 1646 | −0.02 | 0.08 (−0.02, 0.18) | 0.17 | 0.13 | 546 | 1633 | −0.03 | 0.13 (0.04, 0.23) | 0.19 | 0.11 |
| Burkina Faso | iLiNS−Zinc (24) | DMC | 372 | 193 | −0.20 | 0.32 (0.13, 0.52) | 0.05 | 0.07 | 374 | 182 | −0.32 | 0.48 (0.29, 0.67) | 0.04 | 0.08 |
| Ghana | GHANA (25) | | | | | | | | | | | | | |
| Ghana | iLiNS−DYADG (26) | KDI | 150 | 280 | −0.05 | 0.14 (−0.04, 0.33) | 0.05 | 0.07 | 152 | 321 | 0.03 | −0.08 (−0.28, 0.12) | 0.04 | 0.08 |
| Haiti | HAITI (27) | | | | | | | | | | | | | |
| Kenya | WASH−B (28) | EASQ | 682 | 2260 | −0.01 | 0.03 (−0.05, 0.12) | 0.22 | 0.14 | 680 | 2485 | 0.00 | −0.01 (−0.09, 0.07) | 0.24 | 0.11 |
| Madagascar | MAHAY (29) | ASQI | 795 | 787 | −0.07 | 0.13 (−0.06, 0.33) | 0.04 | 0.07 | 818 | 817 | −0.01 | 0.01 (−0.20, 0.23) | 0.03 | 0.07 |
| Malawi | iLiNS−DYADM (30) | KDI | 104 | 204 | −0.07 | 0.23 (−0.01, 0.46) | 0.03 | 0.05 | 110 | 232 | 0.05 | −0.16 (−0.37, 0.05) | 0.04 | 0.07 |
| Malawi | iLiNS−DOSE (31) | KDI | 321 | 117 | −0.01 | 0.01 (−0.21, 0.23) | 0.04 | 0.06 | 325 | 104 | −0.07 | 0.09 (−0.12, 0.31) | 0.04 | 0.07 |
| Mali | PROMIS CS (32) | DMC | 456 | 501 | −0.06 | 0.13 (−0.06, 0.31) | 0.05 | 0.07 | 446 | 420 | −0.07 | 0.13 (−0.07, 0.33) | 0.04 | 0.08 |
| Zimbabwe | SHINE (HIV−) (33) | MDAT | 424 | 388 | −0.06 | 0.12 (−0.02, 0.27) | 0.08 | 0.10 | 417 | 411 | −0.04 | 0.08 (−0.08, 0.25) | 0.06 | 0.09 |
| Zimbabwe | SHINE (HIV+) (34) | MDAT | 84 | 79 | −0.11 | 0.22 (−0.08, 0.52) | 0.02 | 0.04 | 86 | 72 | −0.22 | 0.40 (0.08, 0.72) | 0.02 | 0.05 |
| | | | **4929** | **6911** | | **I² = 0.38, Tau² = 0.01** | | | **4942** | **7117** | | **I² = 0.70, Tau² = 0.02** | | |
| **Fixed** | | | | | | **0.08 (0.04, 0.12)** | | | | | | **0.09 (0.05, 0.13)** | | |
| **Random** | | | | | | **0.09 (0.03, 0.16)** | | | | | | **0.11 (0.01, 0.20)** | | |

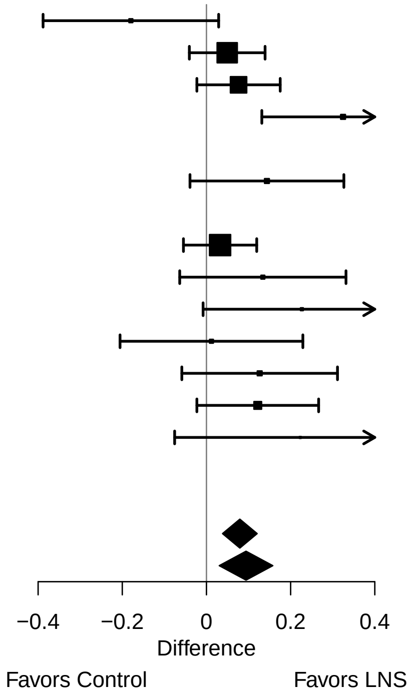

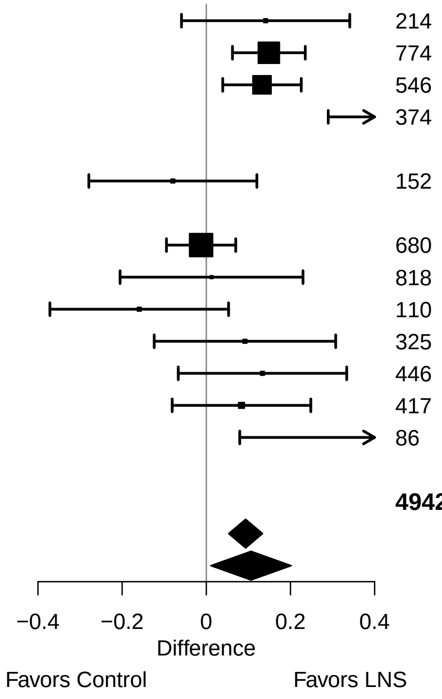

#### Supplemental figure 5G: Mean difference in motor z-score

#### 5G7: Stratified by Child birth order

**P−for−interaction = 0.042**
**Difference in MDs = −0.07 (−0.13, 0.00)**

| | | | | | | Later born | | | | | | Firstborn | | |
|---|---|---|---|---|---|---|---|---|---|---|---|---|---|---|
| | | Tool | LNS | Control | Control | MD | Fixed | Random | LNS | Control | Control | MD | Fixed | Random |
| Country | Trial | | N | N | Mean | (95% CI) | W | W | N | N | Mean | (95% CI) | W | W |
| Bangladesh | JiVitA−4 (21) | BSID−III | 111 | 31 | −0.26 | 0.12 (−0.22, 0.46) | 0.01 | 0.03 | 333 | 111 | 0.08 | −0.04 (−0.21, 0.13) | 0.10 | 0.11 |
| Bangladesh | RDNS (22) | DMC | 909 | 469 | −0.09 | 0.16 (0.06, 0.26) | 0.15 | 0.12 | 646 | 284 | 0.04 | 0.00 (−0.13, 0.13) | 0.19 | 0.16 |
| Bangladesh | WASH−B (23) | EASQ | 675 | 2149 | −0.03 | 0.11 (0.02, 0.19) | 0.19 | 0.12 | 379 | 1028 | 0.02 | 0.07 (−0.04, 0.18) | 0.25 | 0.18 |
| Burkina Faso | iLiNS−Zinc (24) | DMC | 587 | 290 | −0.29 | 0.47 (0.31, 0.63) | 0.06 | 0.08 | 159 | 85 | −0.18 | 0.16 (−0.22, 0.54) | 0.02 | 0.03 |
| Ghana | GHANA (25) | | | | | | | | | | | | | |
| Ghana | iLiNS−DYADG (26) | KDI | 203 | 406 | −0.04 | 0.07 (−0.10, 0.24) | 0.05 | 0.08 | 99 | 195 | 0.06 | −0.05 (−0.27, 0.16) | 0.07 | 0.08 |
| Haiti | HAITI (27) | | | | | | | | | | | | | |
| Kenya | WASH−B (28) | EASQ | 1071 | 3767 | −0.02 | 0.01 (−0.06, 0.08) | 0.28 | 0.13 | 289 | 974 | 0.07 | 0.01 (−0.14, 0.16) | 0.14 | 0.13 |
| Madagascar | MAHAY (29) | ASQI | 1162 | 1118 | −0.09 | 0.08 (−0.12, 0.28) | 0.03 | 0.07 | 433 | 465 | 0.06 | 0.08 (−0.15, 0.31) | 0.06 | 0.07 |
| Malawi | iLiNS−DYADM (30) | KDI | 169 | 349 | −0.02 | 0.03 (−0.15, 0.20) | 0.04 | 0.07 | 45 | 85 | 0.04 | 0.03 (−0.31, 0.37) | 0.03 | 0.04 |
| Malawi | iLiNS−DOSE (31) | KDI | 425 | 151 | −0.07 | 0.11 (−0.08, 0.30) | 0.04 | 0.07 | 134 | 41 | 0.16 | −0.09 (−0.42, 0.25) | 0.03 | 0.04 |
| Mali | PROMIS CS (32) | DMC | 749 | 786 | −0.06 | 0.17 (0.00, 0.35) | 0.04 | 0.08 | 139 | 123 | −0.03 | −0.10 (−0.41, 0.21) | 0.03 | 0.05 |
| Zimbabwe | SHINE (HIV−) (33) | MDAT | 602 | 574 | −0.01 | 0.13 (−0.01, 0.26) | 0.08 | 0.10 | 203 | 191 | −0.17 | 0.04 (−0.16, 0.24) | 0.08 | 0.09 |
| Zimbabwe | SHINE (HIV+) (34) | MDAT | 149 | 118 | −0.10 | 0.22 (−0.02, 0.46) | 0.02 | 0.05 | 18 | 29 | −0.35 | 0.84 (0.30, 1.38) | 0.01 | 0.02 |
| | | | **6812** | **10208** | | **I² = 0.66, Tau² = 0.01** | | | **2877** | **3611** | | **I² = 0.11, Tau² = 0.00** | | |
| **Fixed** | | | | | | **0.11 (0.07, 0.15)** | | | | | | **0.03 (−0.03, 0.08)** | | |
| **Random** | | | | | | **0.13 (0.06, 0.20)** | | | | | | **0.03 (−0.04, 0.10)** | | |

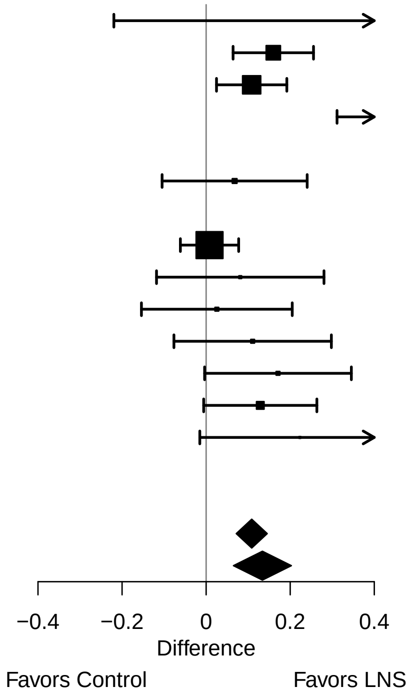

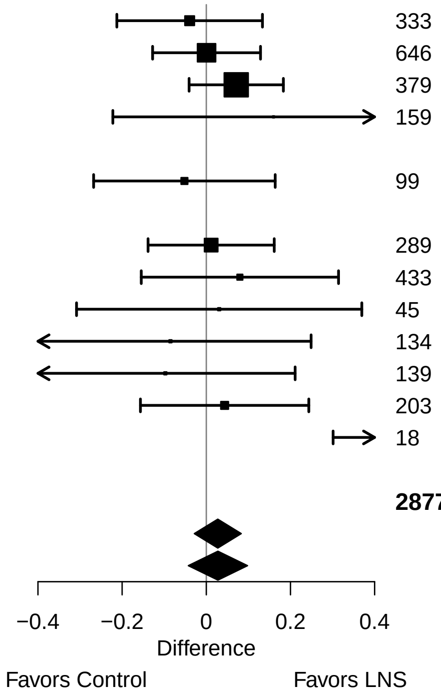

#### Supplemental figure 5G: Mean difference in motor z-score

##### 5G8: Stratified by Child baseline stunting

**P−for−interaction = 0.207**
**Difference in MDs = 0.07 (−0.04, 0.18)**

| Country | Trial | Tool | No: LNS N | No: Control N | No: Control Mean | No: MD (95% CI) | No: Fixed W | No: Random W | Yes: LNS N | Yes: Control N | Yes: Control Mean | Yes: MD (95% CI) | Yes: Fixed W | Yes: Random W |
|---|---|---|---|---|---|---|---|---|---|---|---|---|---|---|
| Bangladesh | JiVitA−4 (21) | BSID−III | 327 | 114 | 0.06 | −0.02 (−0.20, 0.16) | 0.07 | 0.11 | 116 | 29 | −0.14 | 0.03 (−0.24, 0.30) | 0.14 | 0.15 |
| Bangladesh | RDNS (22) | DMC | 1170 | 535 | −0.01 | 0.10 (0.03, 0.17) | 0.44 | 0.19 | 327 | 166 | −0.18 | 0.12 (−0.06, 0.29) | 0.34 | 0.19 |
| Bangladesh | WASH−B (23) | EASQ | | | | | | | | | | | | |
| Burkina Faso | iLiNS−Zinc (24) | DMC | 576 | 294 | −0.08 | 0.33 (0.15, 0.50) | 0.07 | 0.11 | 170 | 80 | −0.90 | 0.69 (0.32, 1.07) | 0.07 | 0.11 |
| Ghana | GHANA (25) | | | | | | | | | | | | | |
| Ghana | iLiNS−DYADG (26) | KDI | 259 | 504 | 0.04 | −0.02 (−0.16, 0.12) | 0.12 | 0.14 | 22 | 57 | −0.36 | 0.36 (−0.28, 1.00) | 0.02 | 0.05 |
| Haiti | HAITI (27) | | | | | | | | | | | | | |
| Kenya | WASH−B (28) | EASQ | | | | | | | | | | | | |
| Madagascar | MAHAY (29) | ASQI | | | | | | | | | | | | |
| Malawi | iLiNS−DYADM (30) | KDI | 152 | 320 | 0.08 | −0.02 (−0.20, 0.15) | 0.08 | 0.12 | 48 | 97 | −0.24 | 0.13 (−0.26, 0.53) | 0.06 | 0.10 |
| Malawi | iLiNS−DOSE (31) | KDI | 465 | 147 | 0.05 | 0.06 (−0.11, 0.23) | 0.08 | 0.12 | 181 | 74 | −0.20 | −0.03 (−0.35, 0.28) | 0.10 | 0.13 |
| Mali | PROMIS CS (32) | DMC | | | | | | | | | | | | |
| Zimbabwe | SHINE (HIV−) (33) | MDAT | 532 | 490 | 0.02 | 0.08 (−0.06, 0.22) | 0.11 | 0.14 | 122 | 105 | −0.25 | 0.08 (−0.15, 0.30) | 0.20 | 0.17 |
| Zimbabwe | SHINE (HIV+) (34) | MDAT | 115 | 89 | −0.05 | 0.27 (0.01, 0.54) | 0.03 | 0.07 | 34 | 32 | −0.42 | 0.51 (0.12, 0.90) | 0.07 | 0.11 |
| | | | **3596** | **2493** | | **$I^2$ = 0.50, $Tau^2$ = 0.01** | | | **1020** | **640** | | **$I^2$ = 0.52, $Tau^2$ = 0.03** | | |
| **Fixed** | | | | | | **0.08 (0.04, 0.13)** | | | | | | **0.16 (0.06, 0.26)** | | |
| **Random** | | | | | | **0.09 (0.00, 0.17)** | | | | | | **0.20 (0.03, 0.37)** | | |

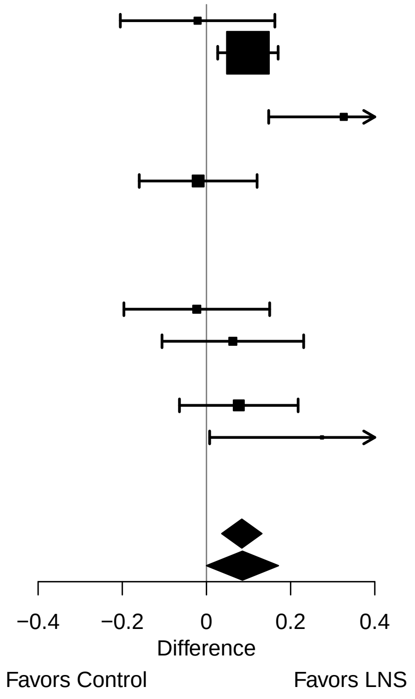

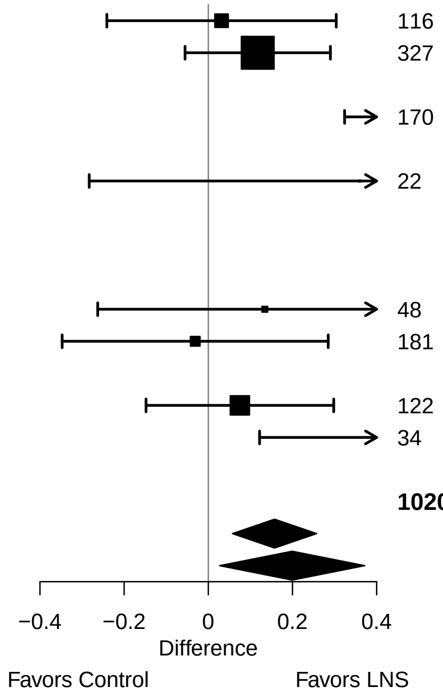

**Supplemental figure 5G: Mean difference in motor z-score**

#### 5G9: Stratified by Child baseline acute malnutrition

**P−for−interaction = 0.131**
**Difference in MDs = 0.12 (−0.04, 0.28)**

| Country | Trial | Tool | No: LNS N | No: Control N | No: Control Mean | No: MD (95% CI) | No: Fixed W | No: Random W | Yes: LNS N | Yes: Control N | Yes: Control Mean | Yes: MD (95% CI) | Yes: Fixed W | Yes: Random W |
|---|---|---|---|---|---|---|---|---|---|---|---|---|---|---|
| Bangladesh | JiVitA−4 (21) | BSID−III | 359 | 112 | 0.10 | −0.06 (−0.24, 0.11) | 0.06 | 0.11 | 84 | 31 | −0.27 | 0.12 (−0.27, 0.51) | 0.16 | 0.16 |
| Bangladesh | RDNS (22) | DMC | 1382 | 653 | −0.03 | 0.11 (0.04, 0.18) | 0.45 | 0.19 | 115 | 49 | −0.29 | 0.11 (−0.25, 0.47) | 0.19 | 0.18 |
| Bangladesh | WASH−B (23) | EASQ | | | | | | | | | | | | |
| Burkina Faso | iLiNS−Zinc (24) | DMC | 556 | 266 | −0.03 | 0.28 (0.11, 0.45) | 0.07 | 0.11 | 190 | 109 | −0.83 | 0.65 (0.36, 0.94) | 0.30 | 0.22 |
| Ghana | GHANA (25) | | | | | | | | | | | | | |
| Ghana | iLiNS−DYADG (26) | KDI | 265 | 509 | 0.02 | 0.04 (−0.11, 0.18) | 0.10 | 0.13 | 17 | 52 | −0.20 | −0.34 (−0.94, 0.27) | 0.07 | 0.09 |
| Haiti | HAITI (27) | | | | | | | | | | | | | |
| Kenya | WASH−B (28) | EASQ | | | | | | | | | | | | |
| Madagascar | MAHAY (29) | ASQI | | | | | | | | | | | | |
| Malawi | iLiNS−DYADM (30) | KDI | 186 | 383 | 0.02 | 0.02 (−0.15, 0.18) | 0.07 | 0.12 | 14 | 37 | −0.25 | 0.06 (−0.75, 0.88) | 0.04 | 0.06 |
| Malawi | iLiNS−DOSE (31) | KDI | 609 | 206 | 0.00 | 0.05 (−0.10, 0.20) | 0.09 | 0.12 | 37 | 15 | −0.49 | −0.09 (−0.96, 0.77) | 0.03 | 0.05 |
| Mali | PROMIS CS (32) | DMC | | | | | | | | | | | | |
| Zimbabwe | SHINE (HIV−) (33) | MDAT | 620 | 566 | −0.03 | 0.07 (−0.06, 0.20) | 0.12 | 0.14 | 36 | 34 | −0.10 | 0.24 (−0.19, 0.67) | 0.13 | 0.15 |
| Zimbabwe | SHINE (HIV+) (34) | MDAT | 137 | 114 | −0.13 | 0.33 (0.10, 0.57) | 0.04 | 0.08 | 14 | 8 | −0.34 | 0.50 (−0.12, 1.12) | 0.07 | 0.09 |
| | | | **4114** | **2809** | | **I² = 0.48, Tau² = 0.01** | | | **507** | **335** | | **I² = 0.47, Tau² = 0.04** | | |
| **Fixed** | | | | | | **0.09 (0.05, 0.14)** | | | | | | **0.28 (0.12, 0.44)** | | |
| **Random** | | | | | | **0.09 (0.01, 0.18)** | | | | | | **0.23 (0.01, 0.44)** | | |

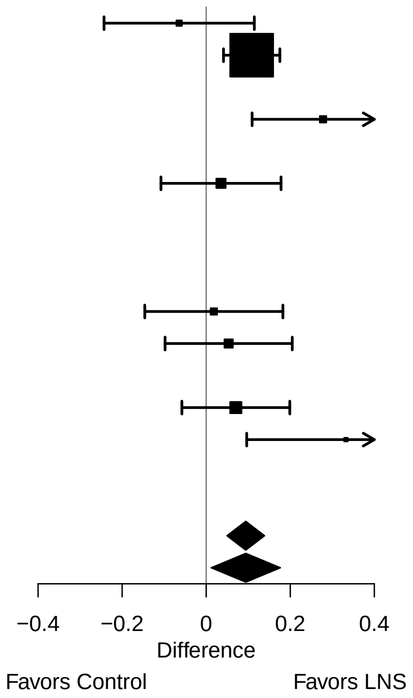

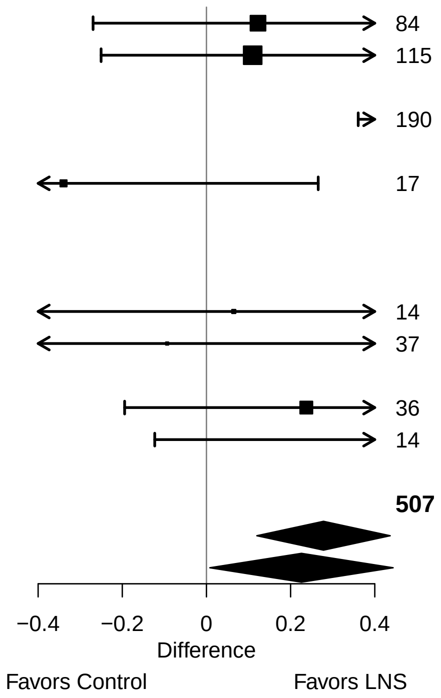

#### Supplemental figure 5G: Mean difference in motor z-score

#### 5G10: Stratified by Child baseline anemia

**P−for−interaction = 0.369**
**Difference in MDs = 0.07 (−0.08, 0.21)**

| Country | Trial | Tool | LNS N | Control N | Control Mean | Not anemic MD (95% CI) | Fixed W | Random W | LNS N | Control N | Control Mean | Anemic MD (95% CI) | Fixed W | Random W |
|---|---|---|---|---|---|---|---|---|---|---|---|---|---|---|
| Bangladesh | JiVitA−4 (21) | BSID−III | | | | | | | | | | | | |
| Bangladesh | RDNS (22) | DMC | 198 | 91 | −0.16 | 0.19 (−0.03, 0.41) | 0.23 | 0.21 | 303 | 151 | −0.06 | 0.12 (0.00, 0.24) | 0.42 | 0.28 |
| Bangladesh | WASH−B (23) | EASQ | | | | | | | | | | | | |
| Burkina Faso | iLiNS−Zinc (24) | DMC | 59 | 34 | −0.25 | 0.51 (0.18, 0.85) | 0.10 | 0.16 | 687 | 341 | −0.26 | 0.39 (0.21, 0.58) | 0.18 | 0.20 |
| Ghana | GHANA (25) | | | | | | | | | | | | | |
| Ghana | iLiNS−DYADG (26) | KDI | 180 | 321 | 0.02 | −0.07 (−0.26, 0.12) | 0.30 | 0.23 | 81 | 189 | −0.06 | 0.20 (−0.04, 0.44) | 0.11 | 0.15 |
| Haiti | HAITI (27) | | | | | | | | | | | | | |
| Kenya | WASH−B (28) | EASQ | | | | | | | | | | | | |
| Madagascar | MAHAY (29) | ASQI | | | | | | | | | | | | |
| Malawi | iLiNS−DYADM (30) | KDI | 65 | 146 | 0.02 | −0.05 (−0.33, 0.22) | 0.15 | 0.19 | 138 | 273 | 0.01 | 0.03 (−0.16, 0.23) | 0.16 | 0.19 |
| Malawi | iLiNS−DOSE (31) | KDI | 224 | 94 | 0.07 | −0.04 (−0.26, 0.18) | 0.23 | 0.21 | 418 | 126 | −0.12 | 0.12 (−0.09, 0.33) | 0.14 | 0.18 |
| Mali | PROMIS CS (32) | DMC | | | | | | | | | | | | |
| Zimbabwe | SHINE (HIV−) (33) | MDAT | | | | | | | | | | | | |
| Zimbabwe | SHINE (HIV+) (34) | MDAT | | | | | | | | | | | | |
| | | | **726** | **686** | | **I² = 0.66, Tau² = 0.04** | | | **1627** | **1080** | | **I² = 0.52, Tau² = 0.01** | | |
| **Fixed** | | | | | | **0.06 (−0.05, 0.16)** | | | | | | **0.16 (0.08, 0.24)** | | |
| **Random** | | | | | | **0.09 (−0.12, 0.30)** | | | | | | **0.17 (0.05, 0.29)** | | |

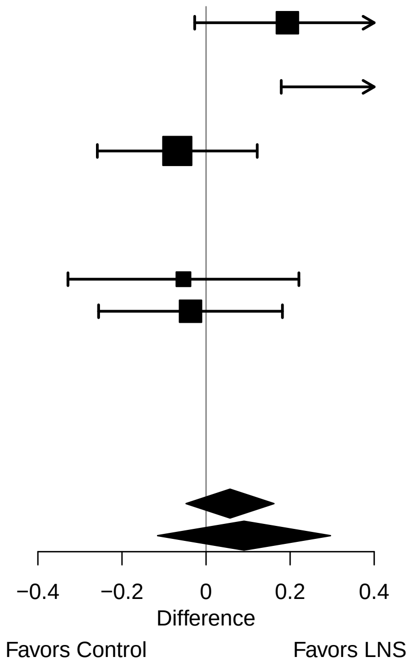

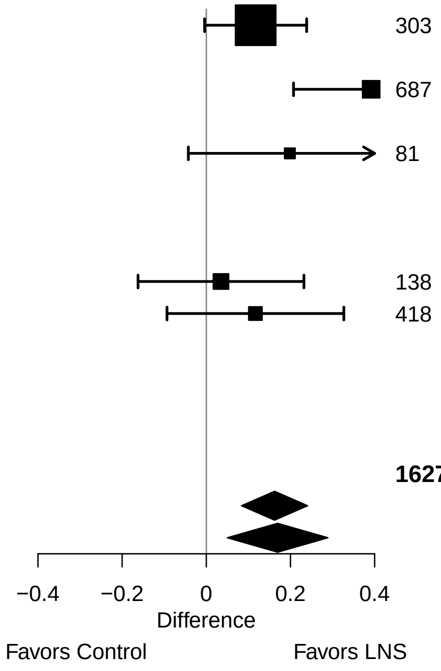

### Supplemental figure 5H: Motor lowest decile prevalence ratio

#### 5H1: Stratified by Maternal height

**P−for−interaction = 0.550**
**Ratio of PRs = 1.09 (0.82, 1.45)**

| Country | Trial | Tool | At least 150.1 cm: LNS N | Control N | Control Prevalence | PR (95% CI) | Fixed W | Random W | Less than 150.1 cm: LNS N | Control N | Control Prevalence | PR (95% CI) | Fixed W | Random W |
|---|---|---|---|---|---|---|---|---|---|---|---|---|---|---|
| Bangladesh | JiVitA−4 (21) | BSID−III | | | | | | | | | | | | |
| Bangladesh | RDNS (22) | DMC | 818 | 399 | 10.3 | 0.79 (0.56, 1.11) | 0.40 | 0.40 | 684 | 333 | 12.9 | 0.85 (0.62, 1.16) | 0.43 | 0.43 |
| Bangladesh | WASH−B (23) | EASQ | 568 | 1773 | 9.4 | 0.73 (0.52, 1.03) | 0.42 | 0.42 | 501 | 1467 | 11.9 | 0.87 (0.64, 1.18) | 0.46 | 0.46 |
| Burkina Faso | iLiNS−Zinc (24) | DMC | | | | | | | | | | | | |
| Ghana | GHANA (25) | | | | | | | | | | | | | |
| Ghana | iLiNS−DYADG (26) | KDI | | | | | | | | | | | | |
| Haiti | HAITI (27) | | | | | | | | | | | | | |
| Kenya | WASH−B (28) | EASQ | | | | | | | | | | | | |
| Madagascar | MAHAY (29) | ASQI | 956 | 986 | 10.2 | 0.95 (0.56, 1.60) | 0.17 | 0.17 | 553 | 538 | 9.1 | 0.93 (0.52, 1.68) | 0.12 | 0.12 |
| Malawi | iLiNS−DYADM (30) | KDI | | | | | | | | | | | | |
| Malawi | iLiNS−DOSE (31) | KDI | | | | | | | | | | | | |
| Mali | PROMIS CS (32) | DMC | | | | | | | | | | | | |
| Zimbabwe | SHINE (HIV−) (33) | MDAT | | | | | | | | | | | | |
| Zimbabwe | SHINE (HIV+) (34) | MDAT | | | | | | | | | | | | |
| | | | **2342** | **3158** | | **I² = 0.00, Tau² = 0.00** | | | **1738** | **2338** | | **I² = 0.00, Tau² = 0.00** | | |
| **Fixed** | | | | | | **0.79 (0.63, 0.98)** | | | | | | **0.87 (0.71, 1.07)** | | |
| **Random** | | | | | | **0.79 (0.63, 0.98)** | | | | | | **0.87 (0.71, 1.07)** | | |

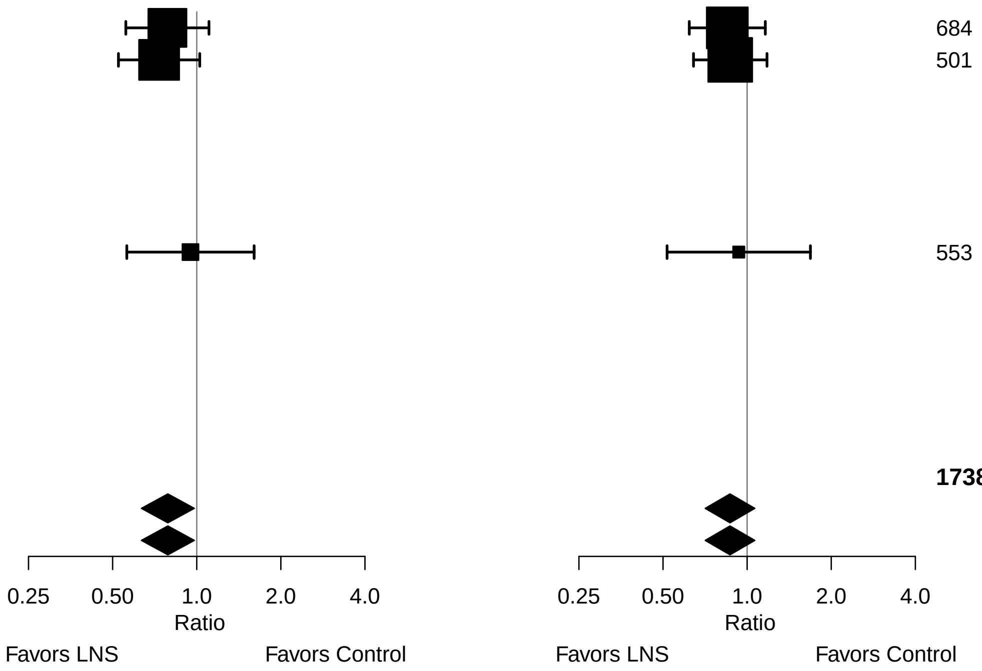

##### Supplemental figure 5H: Motor lowest decile prevalence ratio

#### 5H2: Stratified by Maternal BMI

**P−for−interaction = 0.477**
**Ratio of PRs = 1.08 (0.87, 1.34)**

| Country | Trial | Tool | At least 20 kg/m² LNS N | At least 20 kg/m² Control N | At least 20 kg/m² Control Prevalence | At least 20 kg/m² PR (95% CI) | At least 20 kg/m² Fixed W | At least 20 kg/m² Random W | Less than 20 kg/m² LNS N | Less than 20 kg/m² Control N | Less than 20 kg/m² Control Prevalence | Less than 20 kg/m² PR (95% CI) | Less than 20 kg/m² Fixed W | Less than 20 kg/m² Random W |
|---|---|---|---|---|---|---|---|---|---|---|---|---|---|---|
| Bangladesh | JiVitA−4 (21) | BSID−III | | | | | | | | | | | | |
| Bangladesh | RDNS (22) | DMC | 689 | 306 | 10.1 | 0.77 (0.54, 1.12) | 0.13 | 0.13 | 813 | 426 | 12.4 | 0.86 (0.65, 1.13) | 0.31 | 0.26 |
| Bangladesh | WASH−B (23) | EASQ | 478 | 1502 | 9.7 | 0.75 (0.53, 1.07) | 0.14 | 0.14 | 591 | 1737 | 11.2 | 0.84 (0.63, 1.13) | 0.28 | 0.24 |
| Burkina Faso | iLiNS−Zinc (24) | DMC | 480 | 225 | 12.0 | 0.56 (0.36, 0.86) | 0.09 | 0.09 | 264 | 150 | 20.0 | 0.45 (0.28, 0.75) | 0.10 | 0.11 |
| Ghana | GHANA (25) | | | | | | | | | | | | | |
| Ghana | iLiNS−DYADG (26) | KDI | | | | | | | | | | | | |
| Haiti | HAITI (27) | | | | | | | | | | | | | |
| Kenya | WASH−B (28) | EASQ | 1031 | 3535 | 10.2 | 0.88 (0.70, 1.11) | 0.34 | 0.34 | 257 | 1012 | 9.9 | 0.79 (0.50, 1.24) | 0.11 | 0.13 |
| Madagascar | MAHAY (29) | ASQI | | | | | | | | | | | | |
| Malawi | iLiNS−DYADM (30) | KDI | 126 | 258 | 10.9 | 0.80 (0.41, 1.56) | 0.04 | 0.04 | 87 | 175 | 10.3 | 0.89 (0.40, 1.97) | 0.04 | 0.05 |
| Malawi | iLiNS−DOSE (31) | KDI | 480 | 158 | 8.9 | 1.03 (0.58, 1.84) | 0.05 | 0.05 | 163 | 61 | 11.5 | 1.12 (0.50, 2.51) | 0.04 | 0.05 |
| Mali | PROMIS CS (32) | DMC | 665 | 653 | 11.5 | 0.67 (0.43, 1.04) | 0.09 | 0.09 | 232 | 262 | 10.3 | 1.21 (0.72, 2.04) | 0.09 | 0.10 |
| Zimbabwe | SHINE (HIV−) (33) | MDAT | 601 | 581 | 10.5 | 0.81 (0.55, 1.20) | 0.11 | 0.11 | 100 | 96 | 14.6 | 0.75 (0.35, 1.64) | 0.04 | 0.05 |
| Zimbabwe | SHINE (HIV+) (34) | MDAT | | | | | | | | | | | | |
| | | | **4550** | **7218** | | **I² = 0.00, Tau² = 0.00** | | | **2507** | **3919** | | **I² = 0.17, Tau² = 0.01** | | |
| **Fixed** | | | | | | **0.79 (0.69, 0.90)** | | | | | | **0.83 (0.71, 0.96)** | | |
| **Random** | | | | | | **0.79 (0.69, 0.90)** | | | | | | **0.82 (0.68, 0.99)** | | |

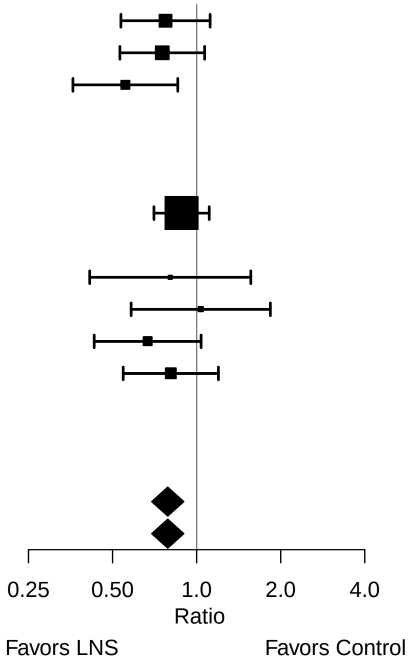

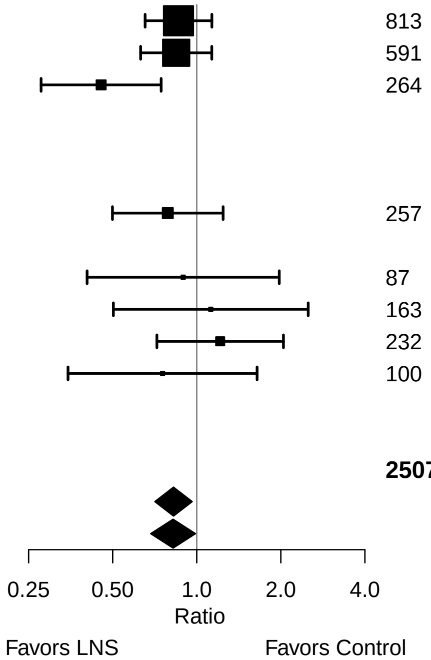

**Supplemental figure 5H: Motor lowest decile prevalence ratio**

#### 5H3: Stratified by Maternal age

**P−for−interaction = 0.466**
**Ratio of PRs = 1.07 (0.89, 1.28)**

| Country | Trial | Tool | At least 25 y LNS N | Control N | Control Prevalence | PR (95% CI) | Fixed W | Random W | Less than 25 y LNS N | Control N | Control Prevalence | PR (95% CI) | Fixed W | Random W |
|---|---|---|---|---|---|---|---|---|---|---|---|---|---|---|
| Bangladesh | JiVitA−4 (21) | BSID−III | 200 | 68 | 8.8 | 1.13 (0.53, 2.42) | 0.03 | 0.04 | 242 | 74 | 6.8 | 1.65 (0.64, 4.29) | 0.02 | 0.02 |
| Bangladesh | RDNS (22) | DMC | 424 | 200 | 13.0 | 0.82 (0.50, 1.34) | 0.07 | 0.08 | 1132 | 553 | 10.5 | 0.86 (0.61, 1.20) | 0.16 | 0.16 |
| Bangladesh | WASH−B (23) | EASQ | 466 | 1450 | 10.6 | 0.89 (0.63, 1.27) | 0.14 | 0.13 | 603 | 1815 | 10.7 | 0.76 (0.57, 1.01) | 0.22 | 0.22 |
| Burkina Faso | iLiNS−Zinc (24) | DMC | 421 | 232 | 17.2 | 0.40 (0.26, 0.61) | 0.10 | 0.10 | 321 | 141 | 11.3 | 0.74 (0.43, 1.28) | 0.06 | 0.06 |
| Ghana | GHANA (25) | | | | | | | | | | | | | |
| Ghana | iLiNS−DYADG (26) | KDI | 192 | 378 | 10.1 | 0.98 (0.58, 1.66) | 0.06 | 0.08 | 110 | 223 | 9.0 | 1.42 (0.75, 2.70) | 0.04 | 0.04 |
| Haiti | HAITI (27) | | | | | | | | | | | | | |
| Kenya | WASH−B (28) | EASQ | 798 | 2622 | 10.1 | 0.86 (0.66, 1.11) | 0.25 | 0.18 | 549 | 2092 | 10.6 | 0.86 (0.62, 1.19) | 0.17 | 0.17 |
| Madagascar | MAHAY (29) | ASQI | 852 | 901 | 11.3 | 0.90 (0.57, 1.43) | 0.08 | 0.09 | 759 | 703 | 9.4 | 0.93 (0.53, 1.61) | 0.06 | 0.06 |
| Malawi | iLiNS−DYADM (30) | KDI | 102 | 224 | 8.9 | 0.88 (0.40, 1.93) | 0.03 | 0.04 | 112 | 212 | 12.3 | 0.80 (0.41, 1.56) | 0.04 | 0.04 |
| Malawi | iLiNS−DOSE (31) | KDI | 346 | 124 | 8.9 | 1.11 (0.58, 2.12) | 0.04 | 0.05 | 288 | 90 | 10.0 | 1.01 (0.50, 2.05) | 0.04 | 0.04 |
| Mali | PROMIS CS (32) | DMC | 480 | 479 | 10.9 | 0.77 (0.52, 1.14) | 0.11 | 0.11 | 422 | 442 | 11.3 | 0.86 (0.51, 1.45) | 0.07 | 0.07 |
| Zimbabwe | SHINE (HIV−) (33) | MDAT | 427 | 389 | 10.5 | 0.87 (0.55, 1.37) | 0.08 | 0.09 | 326 | 321 | 11.5 | 0.96 (0.65, 1.42) | 0.12 | 0.12 |
| Zimbabwe | SHINE (HIV+) (34) | MDAT | | | | | | | | | | | | |
| | | | **4708** | **7067** | | **I² = 0.29, Tau² = 0.02** | | | **4864** | **6666** | | **I² = 0.00, Tau² = 0.00** | | |
| **Fixed** | | | | | | **0.81 (0.71, 0.93)** | | | | | | **0.88 (0.77, 1.00)** | | |
| **Random** | | | | | | **0.82 (0.69, 0.96)** | | | | | | **0.88 (0.77, 1.00)** | | |

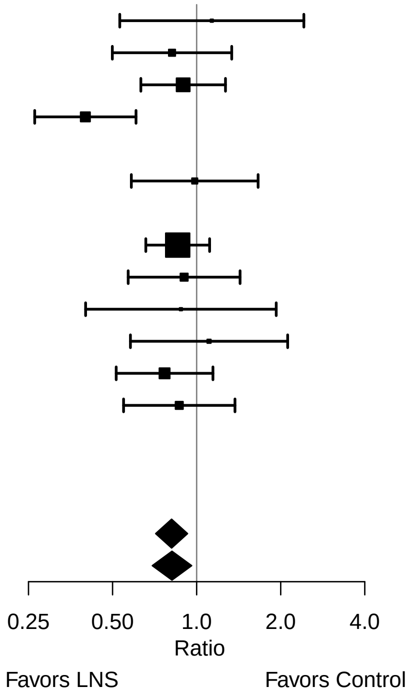

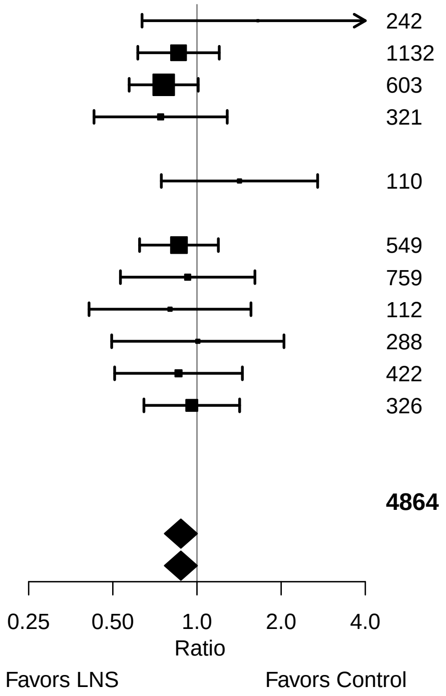

#### Supplemental figure 5H: Motor lowest decile prevalence ratio

#### 5H4: Stratified by Maternal education

**P−for−interaction = 0.225**
**Ratio of PRs = 0.87 (0.69, 1.09)**

| | | | Primary or greater | | | | | | | Incomplete or no formal | | | | |
|---|---|---|---|---|---|---|---|---|---|---|---|---|---|---|
| **Country** | **Trial** | **Tool** | **LNS N** | **Control N** | **Control Prevalence** | **PR (95% CI)** | **Fixed W** | **Random W** | **LNS N** | **Control N** | **Control Prevalence** | **PR (95% CI)** | **Fixed W** | **Random W** |
| Bangladesh | JiVitA−4 (21) | BSID−III | 292 | 92 | 5.4 | 1.76 (0.79, 3.96) | 0.04 | 0.04 | 152 | 51 | 13.7 | 0.91 (0.44, 1.87) | 0.04 | 0.04 |
| Bangladesh | RDNS (22) | DMC | 1166 | 547 | 7.5 | 0.94 (0.65, 1.35) | 0.19 | 0.19 | 390 | 206 | 20.9 | 0.80 (0.55, 1.16) | 0.16 | 0.16 |
| Bangladesh | WASH−B (23) | EASQ | 755 | 2344 | 9.7 | 0.86 (0.66, 1.11) | 0.36 | 0.36 | 319 | 935 | 12.7 | 0.74 (0.50, 1.10) | 0.14 | 0.14 |
| Burkina Faso | iLiNS−Zinc (24) | DMC | | | | | | | | | | | | |
| Ghana | GHANA (25) | | | | | | | | | | | | | |
| Ghana | iLiNS−DYADG (26) | KDI | 235 | 467 | 8.8 | 1.31 (0.83, 2.07) | 0.12 | 0.12 | 67 | 134 | 12.7 | 0.71 (0.29, 1.71) | 0.03 | 0.03 |
| Haiti | HAITI (27) | | | | | | | | | | | | | |
| Kenya | WASH−B (28) | EASQ | 658 | 2258 | 7.5 | 0.81 (0.57, 1.14) | 0.20 | 0.20 | 702 | 2484 | 12.8 | 0.90 (0.70, 1.15) | 0.37 | 0.37 |
| Madagascar | MAHAY (29) | ASQI | 339 | 416 | 5.8 | 1.12 (0.63, 2.02) | 0.07 | 0.07 | 1274 | 1188 | 12.1 | 0.85 (0.53, 1.38) | 0.10 | 0.10 |
| Malawi | iLiNS−DYADM (30) | KDI | | | | | | | | | | | | |
| Malawi | iLiNS−DOSE (31) | KDI | | | | | | | | | | | | |
| Mali | PROMIS CS (32) | DMC | 99 | 93 | 8.6 | 0.94 (0.37, 2.41) | 0.03 | 0.03 | 802 | 828 | 11.4 | 0.80 (0.55, 1.16) | 0.16 | 0.16 |
| Zimbabwe | SHINE (HIV−) (33) | MDAT | | | | | | | | | | | | |
| Zimbabwe | SHINE (HIV+) (34) | MDAT | | | | | | | | | | | | |
| | | | **3544** | **6217** | | **$I^2$ = 0.00, $Tau^2$ = 0.00** | | | **3706** | **5826** | | **$I^2$ = 0.00, $Tau^2$ = 0.00** | | |
| **Fixed** | | | | | | **0.95 (0.81, 1.11)** | | | | | | **0.83 (0.72, 0.97)** | | |
| **Random** | | | | | | **0.95 (0.81, 1.11)** | | | | | | **0.83 (0.72, 0.97)** | | |

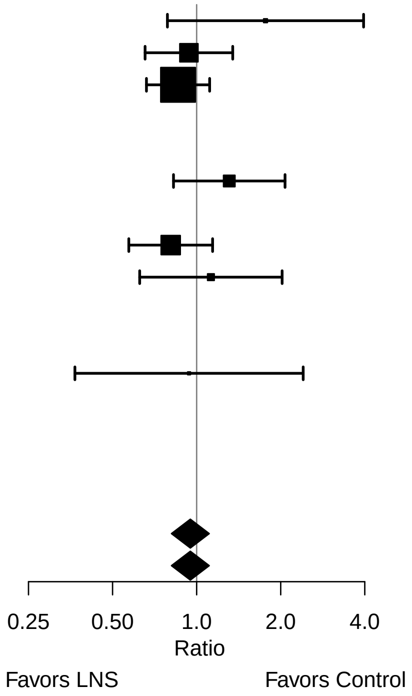

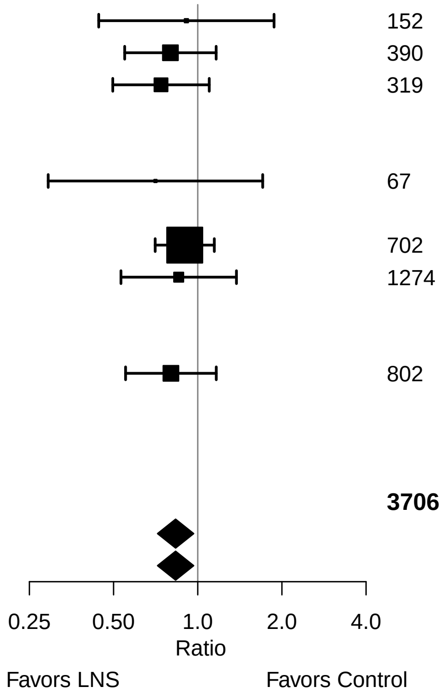

##### Supplemental figure 5H: Motor lowest decile prevalence ratio

#### 5H5: Stratified by Maternal depressive symptoms

**P−for−interaction = 0.649**
**Ratio of PRs = 1.06 (0.83, 1.34)**

| | | | | | | Less than 75th percentile | | | | | | At least 75th percentile | | |
|---|---|---|---|---|---|---|---|---|---|---|---|---|---|---|
| **Country** | **Trial** | **Tool** | **LNS N** | **Control N** | **Control Prevalence** | **PR (95% CI)** | **Fixed W** | **Random W** | **LNS N** | **Control N** | **Control Prevalence** | **PR (95% CI)** | **Fixed W** | **Random W** |
| Bangladesh | JiVitA−4 (21) | BSID−III | | | | | | | | | | | | |
| Bangladesh | RDNS (22) | DMC | 1008 | 435 | 10.8 | 0.80 (0.58, 1.10) | 0.18 | 0.18 | 489 | 267 | 12.0 | 0.82 (0.52, 1.28) | 0.22 | 0.22 |
| Bangladesh | WASH−B (23) | EASQ | 832 | 2358 | 10.3 | 0.79 (0.62, 1.01) | 0.31 | 0.31 | 221 | 838 | 11.3 | 0.88 (0.58, 1.34) | 0.25 | 0.25 |
| Burkina Faso | iLiNS−Zinc (24) | DMC | | | | | | | | | | | | |
| Ghana | GHANA (25) | | | | | | | | | | | | | |
| Ghana | iLiNS−DYADG (26) | KDI | | | | | | | | | | | | |
| Haiti | HAITI (27) | | | | | | | | | | | | | |
| Kenya | WASH−B (28) | EASQ | 960 | 3317 | 8.3 | 0.98 (0.76, 1.26) | 0.29 | 0.29 | 317 | 1156 | 10.3 | 0.77 (0.50, 1.18) | 0.24 | 0.24 |
| Madagascar | MAHAY (29) | ASQI | 615 | 667 | 10.9 | 0.73 (0.39, 1.37) | 0.05 | 0.05 | 268 | 242 | 11.6 | 0.87 (0.40, 1.91) | 0.07 | 0.07 |
| Malawi | iLiNS−DYADM (30) | KDI | 152 | 299 | 11.7 | 0.73 (0.40, 1.34) | 0.05 | 0.05 | 47 | 112 | 8.0 | 1.32 (0.47, 3.74) | 0.04 | 0.04 |
| Malawi | iLiNS−DOSE (31) | KDI | | | | | | | | | | | | |
| Mali | PROMIS CS (32) | DMC | | | | | | | | | | | | |
| Zimbabwe | SHINE (HIV−) (33) | MDAT | 583 | 560 | 8.9 | 1.06 (0.72, 1.55) | 0.12 | 0.12 | 197 | 180 | 16.1 | 0.72 (0.44, 1.19) | 0.18 | 0.18 |
| Zimbabwe | SHINE (HIV+) (34) | MDAT | | | | | | | | | | | | |
| | | | **4150** | **7636** | | **I² = 0.00, Tau² = 0.00** | | | **1539** | **2795** | | **I² = 0.00, Tau² = 0.00** | | |
| **Fixed** | | | | | | **0.87 (0.76, 0.99)** | | | | | | **0.82 (0.67, 1.01)** | | |
| **Random** | | | | | | **0.87 (0.76, 0.99)** | | | | | | **0.82 (0.67, 1.01)** | | |

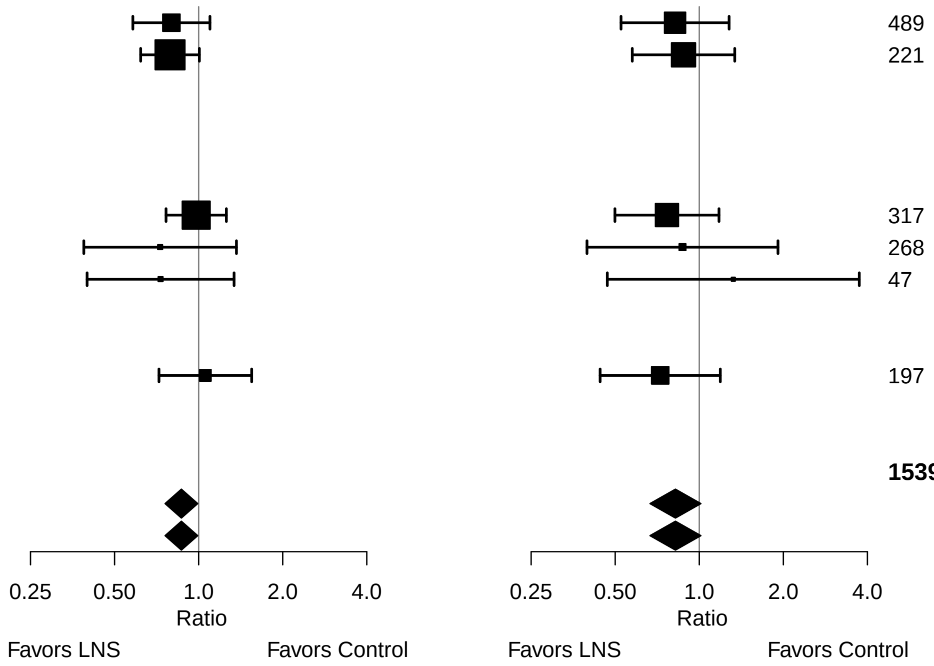

**Supplemental figure 5H: Motor lowest decile prevalence ratio**

#### 5H6: Stratified by Child sex

**P−for−interaction = 0.701**
**Ratio of PRs = 0.97 (0.81, 1.15)**

| Country | Trial | Tool | Male LNS N | Male Control N | Male Control Prevalence | Male PR (95% CI) | Male Fixed W | Male Random W | Female LNS N | Female Control N | Female Control Prevalence | Female PR (95% CI) | Female Fixed W | Female Random W |
|---|---|---|---|---|---|---|---|---|---|---|---|---|---|---|
| Bangladesh | JiVitA−4 (21) | BSID−III | | | | | | | | | | | | |
| Bangladesh | RDNS (22) | DMC | 782 | 386 | 10.4 | 0.96 (0.66, 1.40) | 0.12 | 0.12 | 774 | 367 | 12.0 | 0.74 (0.53, 1.04) | 0.15 | 0.13 |
| Bangladesh | WASH−B (23) | EASQ | 528 | 1646 | 14.1 | 0.90 (0.69, 1.17) | 0.24 | 0.24 | 546 | 1633 | 7.0 | 0.68 (0.46, 1.00) | 0.11 | 0.11 |
| Burkina Faso | iLiNS−Zinc (24) | DMC | 372 | 193 | 14.0 | 0.54 (0.37, 0.78) | 0.12 | 0.12 | 374 | 182 | 16.5 | 0.45 (0.30, 0.68) | 0.10 | 0.11 |
| Ghana | GHANA (25) | | | | | | | | | | | | | |
| Ghana | iLiNS−DYADG (26) | KDI | 150 | 280 | 8.6 | 1.01 (0.53, 1.93) | 0.04 | 0.04 | 152 | 321 | 10.6 | 1.24 (0.74, 2.08) | 0.06 | 0.08 |
| Haiti | HAITI (27) | | | | | | | | | | | | | |
| Kenya | WASH−B (28) | EASQ | 682 | 2260 | 10.0 | 0.89 (0.66, 1.20) | 0.18 | 0.18 | 680 | 2485 | 10.6 | 0.83 (0.64, 1.08) | 0.25 | 0.16 |
| Madagascar | MAHAY (29) | ASQI | 795 | 787 | 11.1 | 0.83 (0.52, 1.33) | 0.07 | 0.07 | 818 | 817 | 9.9 | 1.00 (0.57, 1.75) | 0.05 | 0.07 |
| Malawi | iLiNS−DYADM (30) | KDI | 104 | 204 | 13.2 | 0.44 (0.19, 1.02) | 0.02 | 0.02 | 110 | 232 | 8.2 | 1.44 (0.74, 2.81) | 0.04 | 0.06 |
| Malawi | iLiNS−DOSE (31) | KDI | 321 | 117 | 10.3 | 1.03 (0.55, 1.93) | 0.04 | 0.04 | 325 | 104 | 8.7 | 1.14 (0.56, 2.30) | 0.03 | 0.05 |
| Mali | PROMIS CS (32) | DMC | 456 | 501 | 10.6 | 0.83 (0.51, 1.36) | 0.07 | 0.07 | 446 | 420 | 11.7 | 0.79 (0.50, 1.23) | 0.08 | 0.10 |
| Zimbabwe | SHINE (HIV−) (33) | MDAT | 424 | 388 | 9.8 | 0.92 (0.58, 1.45) | 0.08 | 0.08 | 417 | 411 | 10.9 | 0.94 (0.63, 1.41) | 0.10 | 0.11 |
| Zimbabwe | SHINE (HIV+) (34) | MDAT | 84 | 79 | 13.9 | 0.60 (0.23, 1.53) | 0.02 | 0.02 | 86 | 72 | 11.1 | 0.73 (0.30, 1.78) | 0.02 | 0.04 |
| | | | **4698** | **6841** | | **I² = 0.00, Tau² = 0.00** | | | **4728** | **7044** | | **I² = 0.39, Tau² = 0.04** | | |
| **Fixed** | | | | | | **0.83 (0.73, 0.95)** | | | | | | **0.81 (0.71, 0.92)** | | |
| **Random** | | | | | | **0.83 (0.73, 0.95)** | | | | | | **0.83 (0.69, 0.99)** | | |

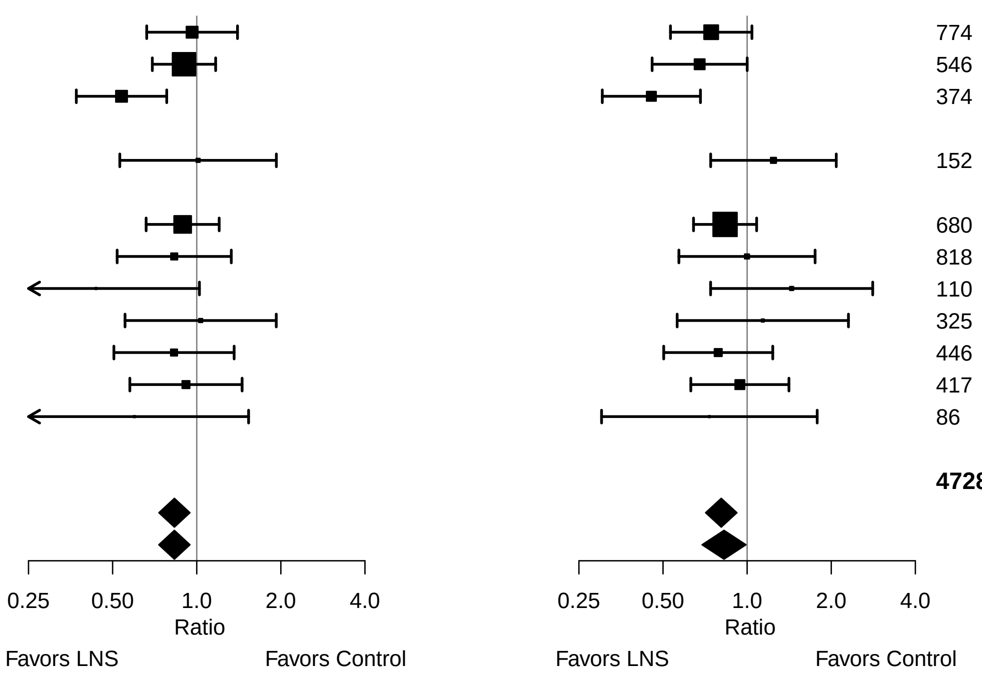

#### Supplemental figure 5H: Motor lowest decile prevalence ratio

#### 5H7: Stratified by Child birth order

**P−for−interaction = 0.015**
**Ratio of PRs = 1.29 (1.05, 1.59)**

| Country | Trial | Tool | LNS N | Control N | Control Prevalence | Later born PR (95% CI) | Fixed W | Random W | LNS N | Control N | Control Prevalence | Firstborn PR (95% CI) | Fixed W | Random W |
|---|---|---|---|---|---|---|---|---|---|---|---|---|---|---|
| Bangladesh | JiVitA−4 (21) | BSID−III | | | | | | | | | | | | |
| Bangladesh | RDNS (22) | DMC | 909 | 469 | 12.4 | 0.73 (0.54, 0.98) | 0.16 | 0.14 | 646 | 284 | 9.2 | 1.10 (0.72, 1.68) | 0.18 | 0.18 |
| Bangladesh | WASH−B (23) | EASQ | 675 | 2149 | 10.5 | 0.77 (0.58, 1.03) | 0.17 | 0.15 | 379 | 1028 | 10.2 | 0.90 (0.64, 1.27) | 0.28 | 0.28 |
| Burkina Faso | iLiNS−Zinc (24) | DMC | 587 | 290 | 16.6 | 0.38 (0.26, 0.56) | 0.09 | 0.12 | 159 | 85 | 10.6 | 1.13 (0.56, 2.29) | 0.06 | 0.06 |
| Ghana | GHANA (25) | | | | | | | | | | | | | |
| Ghana | iLiNS−DYADG (26) | KDI | 203 | 406 | 11.1 | 1.07 (0.67, 1.70) | 0.06 | 0.10 | 99 | 195 | 6.7 | 1.36 (0.60, 3.08) | 0.05 | 0.05 |
| Haiti | HAITI (27) | | | | | | | | | | | | | |
| Kenya | WASH−B (28) | EASQ | 1071 | 3767 | 10.6 | 0.85 (0.68, 1.08) | 0.26 | 0.16 | 289 | 974 | 9.1 | 0.91 (0.56, 1.48) | 0.14 | 0.14 |
| Madagascar | MAHAY (29) | ASQI | 1162 | 1118 | 10.9 | 0.89 (0.56, 1.43) | 0.06 | 0.10 | 433 | 465 | 9.9 | 0.91 (0.49, 1.68) | 0.09 | 0.09 |
| Malawi | iLiNS−DYADM (30) | KDI | | | | | | | | | | | | |
| Malawi | iLiNS−DOSE (31) | KDI | | | | | | | | | | | | |
| Mali | PROMIS CS (32) | DMC | 749 | 786 | 11.1 | 0.77 (0.54, 1.11) | 0.11 | 0.12 | 139 | 123 | 8.9 | 1.21 (0.55, 2.63) | 0.05 | 0.05 |
| Zimbabwe | SHINE (HIV−) (33) | MDAT | 602 | 574 | 9.6 | 0.83 (0.56, 1.25) | 0.09 | 0.11 | 203 | 191 | 14.1 | 1.05 (0.66, 1.67) | 0.15 | 0.15 |
| Zimbabwe | SHINE (HIV+) (34) | MDAT | | | | | | | | | | | | |
| | | | **5958** | **9559** | | **I² = 0.56, Tau² = 0.05** | | | **2347** | **3345** | | **I² = 0.00, Tau² = 0.00** | | |
| **Fixed** | | | | | | **0.76 (0.68, 0.86)** | | | | | | **1.01 (0.84, 1.21)** | | |
| **Random** | | | | | | **0.76 (0.62, 0.92)** | | | | | | **1.01 (0.84, 1.21)** | | |

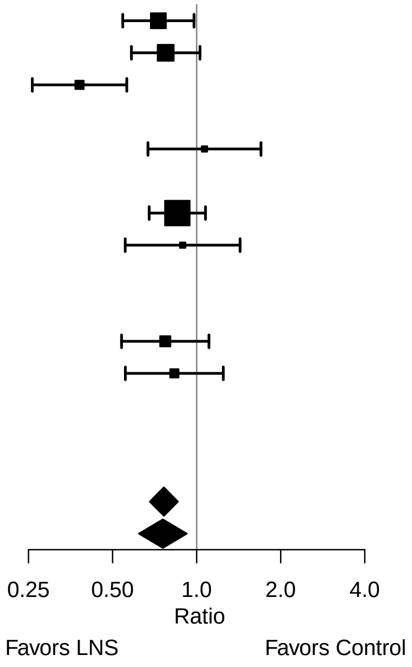

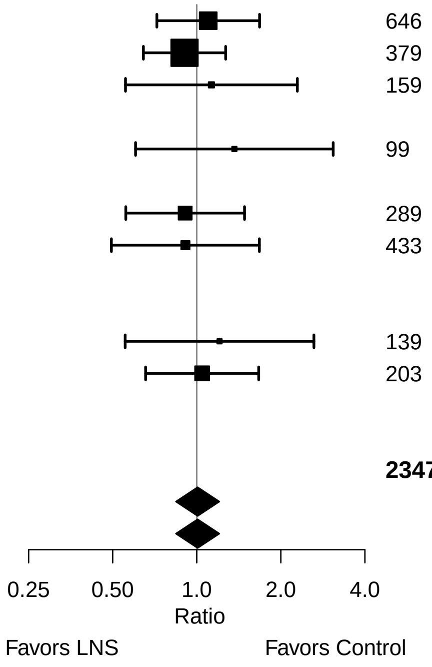

##### Supplemental figure 5H: Motor lowest decile prevalence ratio

#### 5H8: Stratified by Child baseline stunting

**P−for−interaction = 0.870**
**Ratio of PRs = 0.97 (0.72, 1.32)**

| Country | Trial | Tool | No: LNS N | No: Control N | No: Control Prevalence | No: PR (95% CI) | No: Fixed W | No: Random W | Yes: LNS N | Yes: Control N | Yes: Control Prevalence | Yes: PR (95% CI) | Yes: Fixed W | Yes: Random W |
|---|---|---|---|---|---|---|---|---|---|---|---|---|---|---|
| Bangladesh | JiVitA−4 (21) | BSID−III | | | | | | | | | | | | |
| Bangladesh | RDNS (22) | DMC | 1170 | 535 | 10.1 | 0.83 (0.62, 1.11) | 0.40 | 0.25 | 327 | 166 | 15.1 | 0.75 (0.49, 1.15) | 0.35 | 0.30 |
| Bangladesh | WASH−B (23) | EASQ | | | | | | | | | | | | |
| Burkina Faso | iLiNS−Zinc (24) | DMC | 576 | 294 | 11.6 | 0.41 (0.28, 0.58) | 0.26 | 0.23 | 170 | 80 | 28.7 | 0.59 (0.37, 0.95) | 0.29 | 0.27 |
| Ghana | GHANA (25) | | | | | | | | | | | | | |
| Ghana | iLiNS−DYADG (26) | KDI | | | | | | | | | | | | |
| Haiti | HAITI (27) | | | | | | | | | | | | | |
| Kenya | WASH−B (28) | EASQ | | | | | | | | | | | | |
| Madagascar | MAHAY (29) | ASQI | | | | | | | | | | | | |
| Malawi | iLiNS−DYADM (30) | KDI | 152 | 320 | 8.4 | 0.94 (0.49, 1.80) | 0.08 | 0.15 | 48 | 97 | 14.4 | 0.87 (0.36, 2.11) | 0.08 | 0.10 |
| Malawi | iLiNS−DOSE (31) | KDI | 465 | 147 | 8.2 | 0.92 (0.49, 1.73) | 0.08 | 0.16 | 181 | 74 | 12.2 | 1.41 (0.71, 2.81) | 0.13 | 0.16 |
| Mali | PROMIS CS (32) | DMC | | | | | | | | | | | | |
| Zimbabwe | SHINE (HIV−) (33) | MDAT | 532 | 490 | 8.2 | 1.17 (0.76, 1.81) | 0.18 | 0.21 | 122 | 105 | 19.0 | 0.52 (0.27, 0.97) | 0.15 | 0.18 |
| Zimbabwe | SHINE (HIV+) (34) | MDAT | | | | | | | | | | | | |
| | | | **2895** | **1786** | | **I² = 0.76, Tau² = 0.12** | | | **848** | **522** | | **I² = 0.28, Tau² = 0.04** | | |
| **Fixed** | | | | | | **0.75 (0.62, 0.90)** | | | | | | **0.73 (0.57, 0.93)** | | |
| **Random** | | | | | | **0.78 (0.54, 1.14)** | | | | | | **0.74 (0.54, 1.01)** | | |

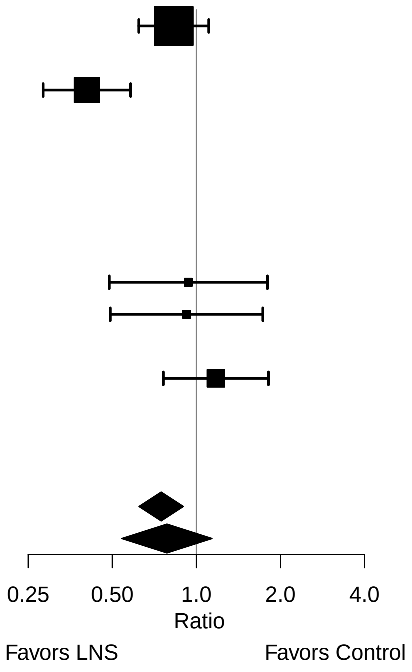

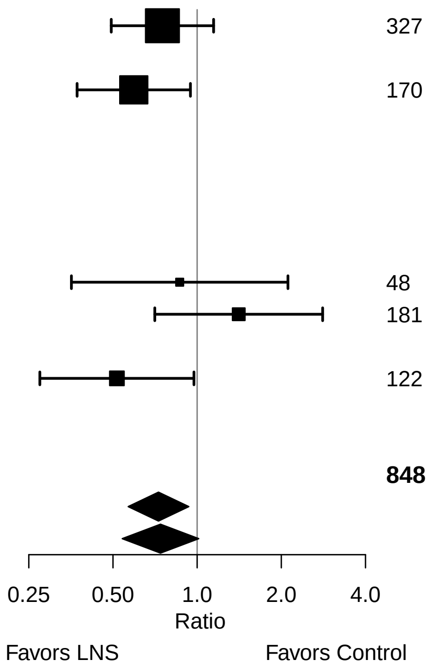

#### Supplemental figure 5H: Motor lowest decile prevalence ratio

### 5H9: Stratified by Child baseline acute malnutrition

**P−for−interaction = 0.612**
**Ratio of PRs = 1.14 (0.69, 1.88)**

| Country | Trial | Tool | No: LNS N | No: Control N | No: Control Prevalence | No: PR (95% CI) | No: Fixed W | No: Random W | Yes: LNS N | Yes: Control N | Yes: Control Prevalence | Yes: PR (95% CI) | Yes: Fixed W | Yes: Random W |
|---|---|---|---|---|---|---|---|---|---|---|---|---|---|---|
| Bangladesh | JiVitA−4 (21) | BSID−III | 359 | 112 | 5.4 | 1.87 (0.86, 4.05) | 0.07 | 0.28 | 84 | 31 | 19.4 | 0.68 (0.28, 1.61) | 0.21 | 0.25 |
| Bangladesh | RDNS (22) | DMC | 1382 | 653 | 11.2 | 0.75 (0.59, 0.96) | 0.70 | 0.37 | 115 | 49 | 12.2 | 1.35 (0.57, 3.20) | 0.21 | 0.25 |
| Bangladesh | WASH−B (23) | EASQ | | | | | | | | | | | | |
| Burkina Faso | iLiNS−Zinc (24) | DMC | 556 | 266 | 10.2 | 0.46 (0.30, 0.71) | 0.23 | 0.35 | 190 | 109 | 27.5 | 0.57 (0.34, 0.97) | 0.58 | 0.50 |
| Ghana | GHANA (25) | | | | | | | | | | | | | |
| Ghana | iLiNS−DYADG (26) | KDI | | | | | | | | | | | | |
| Haiti | HAITI (27) | | | | | | | | | | | | | |
| Kenya | WASH−B (28) | EASQ | | | | | | | | | | | | |
| Madagascar | MAHAY (29) | ASQI | | | | | | | | | | | | |
| Malawi | iLiNS−DYADM (30) | KDI | | | | | | | | | | | | |
| Malawi | iLiNS−DOSE (31) | KDI | | | | | | | | | | | | |
| Mali | PROMIS CS (32) | DMC | | | | | | | | | | | | |
| Zimbabwe | SHINE (HIV−) (33) | MDAT | | | | | | | | | | | | |
| Zimbabwe | SHINE (HIV+) (34) | MDAT | | | | | | | | | | | | |
| | | | **2297** | **1031** | | **$I^2$ = 0.80, $Tau^2$ = 0.40** | | | **389** | **189** | | **$I^2$ = 0.28, $Tau^2$ = 0.05** | | |
| **Fixed** | | | | | | **0.72 (0.58, 0.88)** | | | | | | **0.71 (0.48, 1.06)** | | |
| **Random** | | | | | | **0.82 (0.38, 1.77)** | | | | | | **0.74 (0.45, 1.21)** | | |

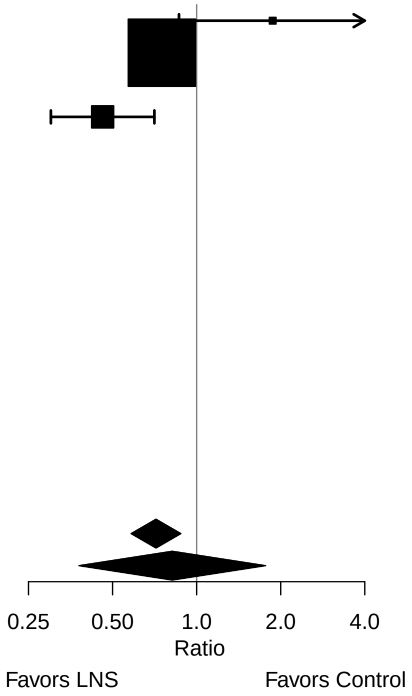

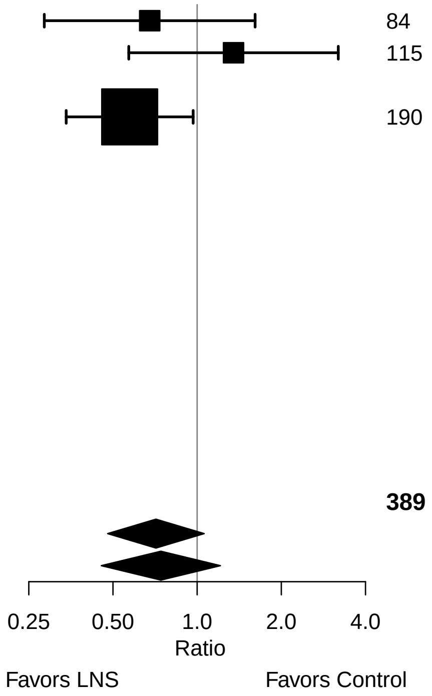

#### Supplemental figure 5H: Motor lowest decile prevalence ratio

##### 5H10: Stratified by Child baseline anemia

**P−for−interaction = 0.237**
**Ratio of PRs = 0.74 (0.46, 1.21)**

| Country | Trial | Tool | Not anemic LNS N | Not anemic Control N | Not anemic Control Prevalence | Not anemic PR (95% CI) | Not anemic Fixed W | Not anemic Random W | Anemic LNS N | Anemic Control N | Anemic Control Prevalence | Anemic PR (95% CI) | Anemic Fixed W | Anemic Random W |
|---|---|---|---|---|---|---|---|---|---|---|---|---|---|---|
| Bangladesh | JiVitA−4 (21) | BSID−III | | | | | | | | | | | | |
| Bangladesh | RDNS (22) | DMC | 198 | 91 | 15.4 | 0.56 (0.30, 1.03) | 0.30 | 0.29 | 303 | 151 | 12.6 | 0.66 (0.38, 1.14) | 0.31 | 0.31 |
| Bangladesh | WASH−B (23) | EASQ | | | | | | | | | | | | |
| Burkina Faso | iLiNS−Zinc (24) | DMC | | | | | | | | | | | | |
| Ghana | GHANA (25) | | | | | | | | | | | | | |
| Ghana | iLiNS−DYADG (26) | KDI | 180 | 321 | 8.7 | 1.59 (0.96, 2.64) | 0.43 | 0.34 | 81 | 189 | 12.7 | 0.58 (0.25, 1.37) | 0.13 | 0.13 |
| Haiti | HAITI (27) | | | | | | | | | | | | | |
| Kenya | WASH−B (28) | EASQ | | | | | | | | | | | | |
| Madagascar | MAHAY (29) | ASQI | | | | | | | | | | | | |
| Malawi | iLiNS−DYADM (30) | KDI | 65 | 146 | 7.5 | 1.23 (0.47, 3.17) | 0.12 | 0.17 | 138 | 273 | 11.7 | 0.80 (0.44, 1.48) | 0.26 | 0.26 |
| Malawi | iLiNS−DOSE (31) | KDI | 224 | 94 | 7.4 | 1.14 (0.50, 2.62) | 0.16 | 0.20 | 418 | 126 | 11.1 | 1.01 (0.58, 1.78) | 0.30 | 0.30 |
| Mali | PROMIS CS (32) | DMC | | | | | | | | | | | | |
| Zimbabwe | SHINE (HIV−) (33) | MDAT | | | | | | | | | | | | |
| Zimbabwe | SHINE (HIV+) (34) | MDAT | | | | | | | | | | | | |
| | | | **667** | **652** | | **I² = 0.56, Tau² = 0.11** | | | **940** | **739** | | **I² = 0.00, Tau² = 0.00** | | |
| **Fixed** | | | | | | **1.07 (0.77, 1.49)** | | | | | | **0.78 (0.57, 1.06)** | | |
| **Random** | | | | | | **1.05 (0.65, 1.70)** | | | | | | **0.78 (0.57, 1.06)** | | |

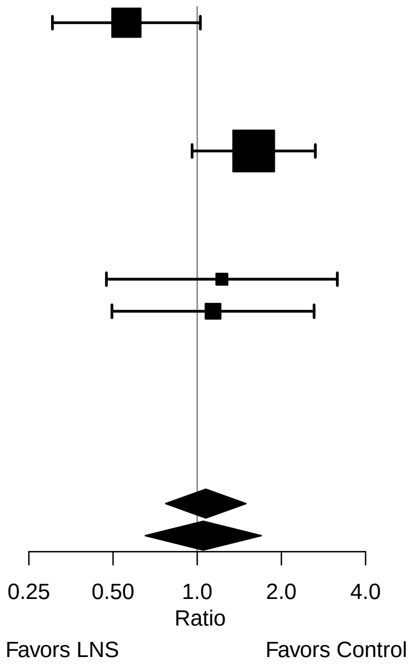

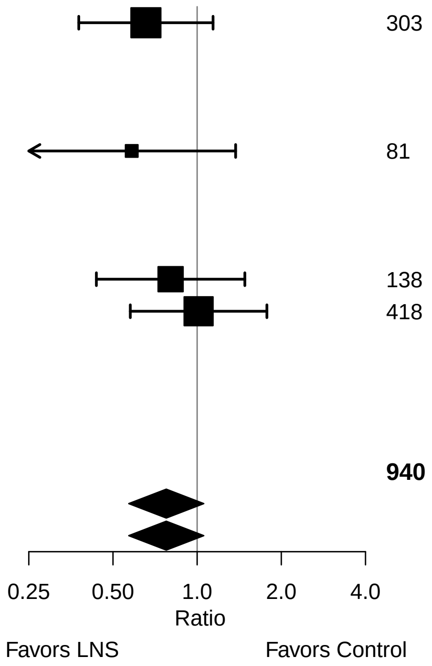

### Supplemental figure 5I: Motor lowest decile prevalence difference

#### 5I1: Stratified by Maternal height

P−for−interaction = 0.752
Difference in PDs = 0.00 (−0.02, 0.03)

| Country | Trial | Tool | LNS N | Control N | Control Prevalence | At least 150.1 cm PD (95% CI) | Fixed W | Random W | LNS N | Control N | Control Prevalence | Less than 150.1 cm PD (95% CI) | Fixed W | Random W |
|---|---|---|---|---|---|---|---|---|---|---|---|---|---|---|
| Bangladesh | JiVitA−4 (21) | BSID−III | | | | | | | | | | | | |
| Bangladesh | RDNS (22) | DMC | 818 | 399 | 10.3 | −0.02 (−0.05, 0.01) | 0.35 | 0.35 | 684 | 333 | 12.9 | −0.02 (−0.06, 0.02) | 0.34 | 0.34 |
| Bangladesh | WASH−B (23) | EASQ | 568 | 1773 | 9.4 | −0.02 (−0.05, 0.00) | 0.53 | 0.53 | 501 | 1467 | 11.9 | −0.02 (−0.05, 0.02) | 0.48 | 0.48 |
| Burkina Faso | iLiNS−Zinc (24) | DMC | | | | | | | | | | | | |
| Ghana | GHANA (25) | | | | | | | | | | | | | |
| Ghana | iLiNS−DYADG (26) | KDI | | | | | | | | | | | | |
| Haiti | HAITI (27) | | | | | | | | | | | | | |
| Kenya | WASH−B (28) | EASQ | | | | | | | | | | | | |
| Madagascar | MAHAY (29) | ASQI | 956 | 986 | 10.2 | −0.01 (−0.06, 0.05) | 0.12 | 0.12 | 553 | 538 | 9.1 | −0.01 (−0.06, 0.05) | 0.18 | 0.18 |
| Malawi | iLiNS−DYADM (30) | KDI | | | | | | | | | | | | |
| Malawi | iLiNS−DOSE (31) | KDI | | | | | | | | | | | | |
| Mali | PROMIS CS (32) | DMC | | | | | | | | | | | | |
| Zimbabwe | SHINE (HIV−) (33) | MDAT | | | | | | | | | | | | |
| Zimbabwe | SHINE (HIV+) (34) | MDAT | | | | | | | | | | | | |
| | | | **2342** | **3158** | | **I² = 0.00, Tau² = 0.00** | | | **1738** | **2338** | | **I² = 0.00, Tau² = 0.00** | | |
| **Fixed** | | | | | | **−0.02 (−0.04, 0.00)** | | | | | | **−0.02 (−0.04, 0.01)** | | |
| **Random** | | | | | | **−0.02 (−0.04, 0.00)** | | | | | | **−0.02 (−0.04, 0.01)** | | |

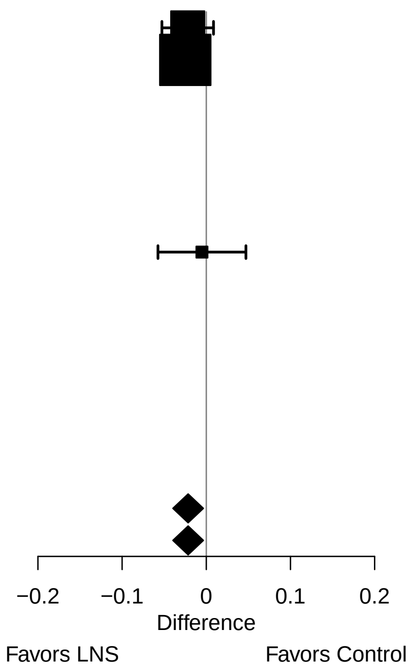

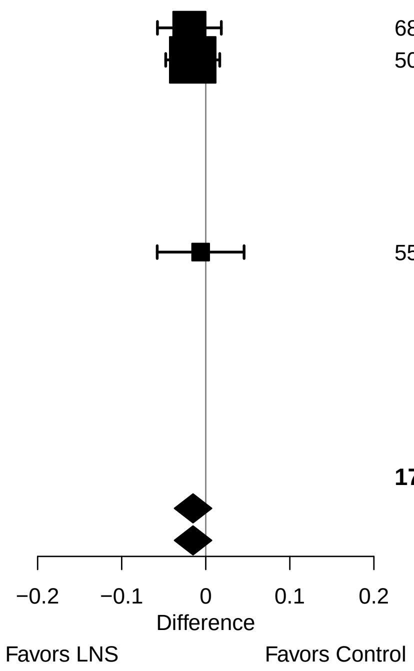

#### Supplemental figure 5I: Motor lowest decile prevalence difference

#### 5I2: Stratified by Maternal BMI

P−for−interaction = 0.721
Difference in PDs = 0.00 (−0.02, 0.03)

| | | | At least 20 kg/m² | | | | | | Less than 20 kg/m² | | | | | |
|---|---|---|---|---|---|---|---|---|---|---|---|---|---|---|
| Country | Trial | Tool | LNS N | Control N | Control Prevalence | PD (95% CI) | Fixed W | Random W | LNS N | Control N | Control Prevalence | PD (95% CI) | Fixed W | Random W |
| Bangladesh | JiVitA−4 (21) | BSID−III | | | | | | | | | | | | |
| Bangladesh | RDNS (22) | DMC | 689 | 306 | 10.1 | −0.02 (−0.06, 0.01) | 0.13 | 0.13 | 813 | 426 | 12.4 | −0.02 (−0.05, 0.01) | 0.26 | 0.24 |
| Bangladesh | WASH−B (23) | EASQ | 478 | 1502 | 9.7 | −0.02 (−0.05, 0.00) | 0.19 | 0.19 | 591 | 1737 | 11.2 | −0.02 (−0.05, 0.01) | 0.33 | 0.28 |
| Burkina Faso | iLiNS−Zinc (24) | DMC | 480 | 225 | 12.0 | −0.05 (−0.09, −0.01) | 0.08 | 0.08 | 264 | 150 | 20.0 | −0.11 (−0.19, −0.03) | 0.05 | 0.06 |
| Ghana | GHANA (25) | | | | | | | | | | | | | |
| Ghana | iLiNS−DYADG (26) | KDI | | | | | | | | | | | | |
| Haiti | HAITI (27) | | | | | | | | | | | | | |
| Kenya | WASH−B (28) | EASQ | 1031 | 3535 | 10.2 | −0.01 (−0.03, 0.01) | 0.31 | 0.31 | 257 | 1012 | 9.9 | −0.02 (−0.06, 0.02) | 0.18 | 0.19 |
| Madagascar | MAHAY (29) | ASQI | | | | | | | | | | | | |
| Malawi | iLiNS−DYADM (30) | KDI | 126 | 258 | 10.9 | −0.02 (−0.09, 0.04) | 0.03 | 0.03 | 87 | 175 | 10.3 | −0.01 (−0.09, 0.07) | 0.05 | 0.06 |
| Malawi | iLiNS−DOSE (31) | KDI | 480 | 158 | 8.9 | 0.00 (−0.05, 0.05) | 0.05 | 0.05 | 163 | 61 | 11.5 | 0.01 (−0.08, 0.11) | 0.03 | 0.04 |
| Mali | PROMIS CS (32) | DMC | 665 | 653 | 11.5 | −0.04 (−0.08, 0.00) | 0.09 | 0.09 | 232 | 262 | 10.3 | 0.02 (−0.04, 0.08) | 0.08 | 0.09 |
| Zimbabwe | SHINE (HIV−) (33) | MDAT | 601 | 581 | 10.5 | −0.02 (−0.06, 0.02) | 0.10 | 0.10 | 100 | 96 | 14.6 | −0.04 (−0.13, 0.06) | 0.03 | 0.04 |
| Zimbabwe | SHINE (HIV+) (34) | MDAT | | | | | | | | | | | | |
| | | | **4550** | **7218** | | **I² = 0.00, Tau² = 0.00** | | | **2507** | **3919** | | **I² = 0.09, Tau² = 0.00** | | |
| **Fixed** | | | | | | **−0.02 (−0.03, −0.01)** | | | | | | **−0.02 (−0.04, 0.00)** | | |
| **Random** | | | | | | **−0.02 (−0.03, −0.01)** | | | | | | **−0.02 (−0.04, 0.00)** | | |

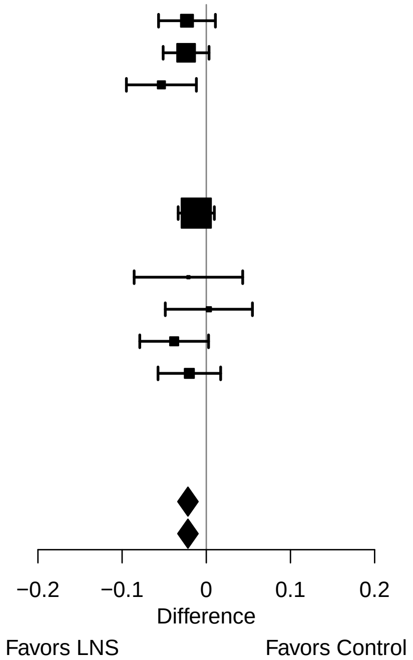

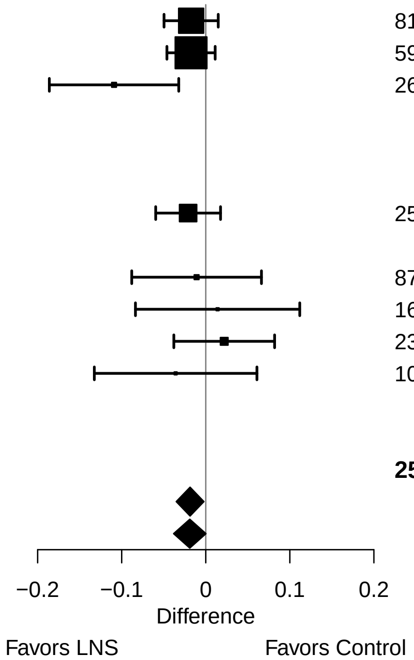

#### Supplemental figure 5I: Motor lowest decile prevalence difference

#### 5I3: Stratified by Maternal age

**P−for−interaction = 0.555**
**Difference in PDs = 0.01 (−0.01, 0.02)**

| | | | At least 25 y | | | | | | Less than 25 y | | | | | |
|---|---|---|---|---|---|---|---|---|---|---|---|---|---|---|
| **Country** | **Trial** | **Tool** | **LNS N** | **Control N** | **Control Prevalence** | **PD (95% CI)** | **Fixed W** | **Random W** | **LNS N** | **Control N** | **Control Prevalence** | **PD (95% CI)** | **Fixed W** | **Random W** |
| Bangladesh | JiVitA−4 (21) | BSID−III | 200 | 68 | 8.8 | 0.01 (−0.06, 0.08) | 0.03 | 0.05 | 242 | 74 | 6.8 | 0.04 (−0.03, 0.12) | 0.03 | 0.03 |
| Bangladesh | RDNS (22) | DMC | 424 | 200 | 13.0 | −0.02 (−0.08, 0.03) | 0.05 | 0.07 | 1132 | 553 | 10.5 | −0.01 (−0.05, 0.02) | 0.15 | 0.15 |
| Bangladesh | WASH−B (23) | EASQ | 466 | 1450 | 10.6 | −0.01 (−0.04, 0.02) | 0.14 | 0.12 | 603 | 1815 | 10.7 | −0.03 (−0.05, 0.00) | 0.28 | 0.28 |
| Burkina Faso | iLiNS−Zinc (24) | DMC | 421 | 232 | 17.2 | −0.10 (−0.14, −0.06) | 0.10 | 0.11 | 321 | 141 | 11.3 | −0.03 (−0.09, 0.03) | 0.05 | 0.05 |
| Ghana | GHANA (25) | | | | | | | | | | | | | |
| Ghana | iLiNS−DYADG (26) | KDI | 192 | 378 | 10.1 | 0.00 (−0.05, 0.05) | 0.06 | 0.08 | 110 | 223 | 9.0 | 0.04 (−0.03, 0.11) | 0.03 | 0.03 |
| Haiti | HAITI (27) | | | | | | | | | | | | | |
| Kenya | WASH−B (28) | EASQ | 798 | 2622 | 10.1 | −0.01 (−0.04, 0.01) | 0.28 | 0.15 | 549 | 2092 | 10.6 | −0.01 (−0.05, 0.02) | 0.18 | 0.18 |
| Madagascar | MAHAY (29) | ASQI | 852 | 901 | 11.3 | −0.01 (−0.06, 0.04) | 0.07 | 0.08 | 759 | 703 | 9.4 | −0.01 (−0.06, 0.04) | 0.07 | 0.07 |
| Malawi | iLiNS−DYADM (30) | KDI | 102 | 224 | 8.9 | −0.01 (−0.08, 0.05) | 0.04 | 0.06 | 112 | 212 | 12.3 | −0.02 (−0.10, 0.05) | 0.03 | 0.03 |
| Malawi | iLiNS−DOSE (31) | KDI | 346 | 124 | 8.9 | 0.01 (−0.05, 0.07) | 0.04 | 0.07 | 288 | 90 | 10.0 | 0.00 (−0.07, 0.07) | 0.03 | 0.03 |
| Mali | PROMIS CS (32) | DMC | 480 | 479 | 10.9 | −0.03 (−0.06, 0.01) | 0.12 | 0.11 | 422 | 442 | 11.3 | −0.02 (−0.07, 0.04) | 0.06 | 0.06 |
| Zimbabwe | SHINE (HIV−) (33) | MDAT | 427 | 389 | 10.5 | −0.01 (−0.06, 0.03) | 0.08 | 0.09 | 326 | 321 | 11.5 | 0.00 (−0.05, 0.04) | 0.08 | 0.08 |
| Zimbabwe | SHINE (HIV+) (34) | MDAT | | | | | | | | | | | | |
| | | | **4708** | **7067** | | **I² = 0.48, Tau² = 0.00** | | | **4864** | **6666** | | **I² = 0.00, Tau² = 0.00** | | |
| **Fixed** | | | | | | **−0.02 (−0.03, −0.01)** | | | | | | **−0.01 (−0.03, 0.00)** | | |
| **Random** | | | | | | **−0.02 (−0.04, 0.00)** | | | | | | **−0.01 (−0.03, 0.00)** | | |

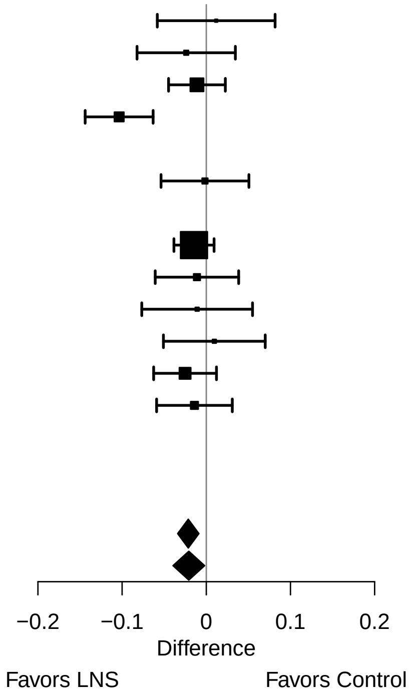

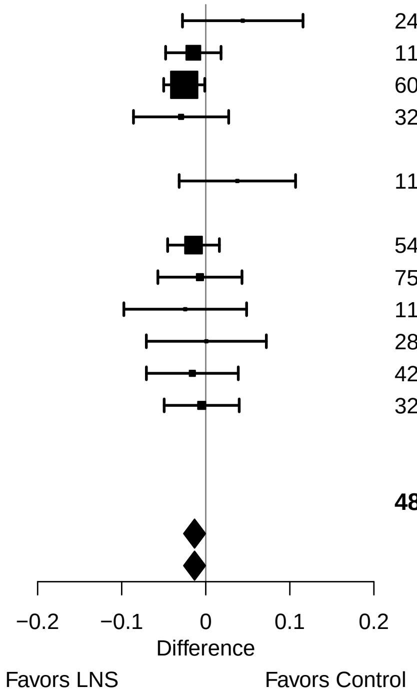

#### Supplemental figure 5I: Motor lowest decile prevalence difference

#### 5I4: Stratified by Maternal education

**P−for−interaction = 0.095**
**Difference in PDs = −0.02 (−0.04, 0.00)**

| Country | Trial | Tool | Primary or greater: LNS N | Control N | Control Prevalence | PD (95% CI) | Fixed W | Random W | Incomplete or no formal: LNS N | Control N | Control Prevalence | PD (95% CI) | Fixed W | Random W |
|---|---|---|---|---|---|---|---|---|---|---|---|---|---|---|
| Bangladesh | JiVitA−4 (21) | BSID−III | 292 | 92 | 5.4 | 0.04 (−0.01, 0.09) | 0.05 | 0.05 | 152 | 51 | 13.7 | −0.01 (−0.11, 0.08) | 0.03 | 0.03 |
| Bangladesh | RDNS (22) | DMC | 1166 | 547 | 7.5 | 0.00 (−0.03, 0.02) | 0.20 | 0.20 | 390 | 206 | 20.9 | −0.04 (−0.11, 0.03) | 0.06 | 0.06 |
| Bangladesh | WASH−B (23) | EASQ | 755 | 2344 | 9.7 | −0.01 (−0.04, 0.01) | 0.28 | 0.28 | 319 | 935 | 12.7 | −0.03 (−0.07, 0.01) | 0.19 | 0.19 |
| Burkina Faso | iLiNS−Zinc (24) | DMC | | | | | | | | | | | | |
| Ghana | GHANA (25) | | | | | | | | | | | | | |
| Ghana | iLiNS−DYADG (26) | KDI | 235 | 467 | 8.8 | 0.03 (−0.02, 0.07) | 0.06 | 0.06 | 67 | 134 | 12.7 | −0.04 (−0.13, 0.06) | 0.03 | 0.03 |
| Haiti | HAITI (27) | | | | | | | | | | | | | |
| Kenya | WASH−B (28) | EASQ | 658 | 2258 | 7.5 | −0.01 (−0.04, 0.01) | 0.28 | 0.28 | 702 | 2484 | 12.8 | −0.01 (−0.04, 0.02) | 0.36 | 0.36 |
| Madagascar | MAHAY (29) | ASQI | 339 | 416 | 5.8 | 0.01 (−0.03, 0.04) | 0.11 | 0.11 | 1274 | 1188 | 12.1 | −0.02 (−0.07, 0.04) | 0.11 | 0.11 |
| Malawi | iLiNS−DYADM (30) | KDI | | | | | | | | | | | | |
| Malawi | iLiNS−DOSE (31) | KDI | | | | | | | | | | | | |
| Mali | PROMIS CS (32) | DMC | 99 | 93 | 8.6 | −0.01 (−0.08, 0.07) | 0.02 | 0.02 | 802 | 828 | 11.4 | −0.02 (−0.06, 0.02) | 0.22 | 0.22 |
| Zimbabwe | SHINE (HIV−) (33) | MDAT | | | | | | | | | | | | |
| Zimbabwe | SHINE (HIV+) (34) | MDAT | | | | | | | | | | | | |
| | | | **3544** | **6217** | | **I² = 0.09, Tau² = 0.00** | | | **3706** | **5826** | | **I² = 0.00, Tau² = 0.00** | | |
| **Fixed** | | | | | | **0.00 (−0.02, 0.01)** | | | | | | **−0.02 (−0.04, 0.00)** | | |
| **Random** | | | | | | **0.00 (−0.02, 0.01)** | | | | | | **−0.02 (−0.04, 0.00)** | | |

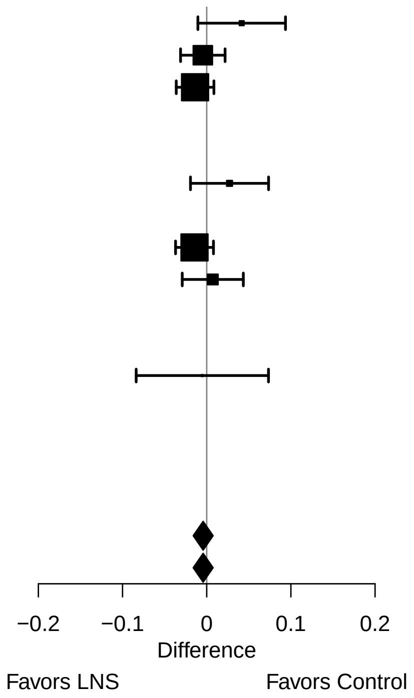

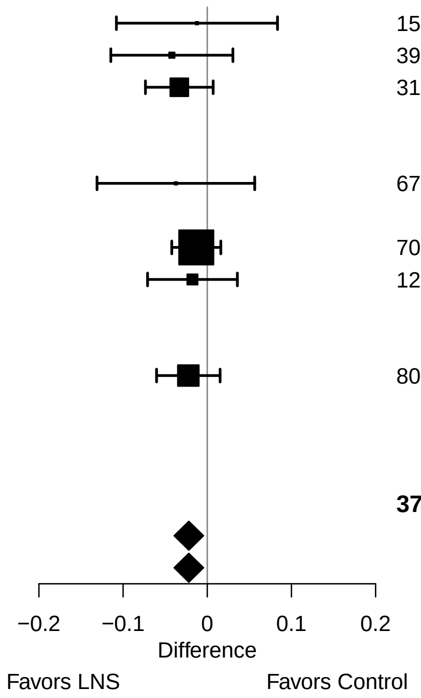

#### Supplemental figure 5I: Motor lowest decile prevalence difference

#### 5I5: Stratified by Maternal depressive symptoms

**P−for−interaction = 0.603**
**Difference in PDs = 0.01 (−0.02, 0.03)**

| **Country** | **Trial** | **Tool** | **Less than 75th percentile: LNS N** | **Control N** | **Control Prevalence** | **PD (95% CI)** | **Fixed W** | **Random W** | **At least 75th percentile: LNS N** | **Control N** | **Control Prevalence** | **PD (95% CI)** | **Fixed W** | **Random W** |
|---|---|---|---|---|---|---|---|---|---|---|---|---|---|---|
| Bangladesh | JiVitA−4 (21) | BSID−III | | | | | | | | | | | | |
| Bangladesh | RDNS (22) | DMC | 1008 | 435 | 10.8 | −0.02 (−0.05, 0.01) | 0.14 | 0.14 | 489 | 267 | 12.0 | −0.02 (−0.07, 0.03) | 0.19 | 0.19 |
| Bangladesh | WASH−B (23) | EASQ | 832 | 2358 | 10.3 | −0.02 (−0.04, 0.00) | 0.33 | 0.33 | 221 | 838 | 11.3 | −0.01 (−0.06, 0.03) | 0.24 | 0.24 |
| Burkina Faso | iLiNS−Zinc (24) | DMC | | | | | | | | | | | | |
| Ghana | GHANA (25) | | | | | | | | | | | | | |
| Ghana | iLiNS−DYADG (26) | KDI | | | | | | | | | | | | |
| Haiti | HAITI (27) | | | | | | | | | | | | | |
| Kenya | WASH−B (28) | EASQ | 960 | 3317 | 8.3 | 0.00 (−0.02, 0.02) | 0.34 | 0.34 | 317 | 1156 | 10.3 | −0.02 (−0.06, 0.01) | 0.36 | 0.36 |
| Madagascar | MAHAY (29) | ASQI | 615 | 667 | 10.9 | −0.03 (−0.09, 0.03) | 0.04 | 0.04 | 268 | 242 | 11.6 | −0.01 (−0.10, 0.07) | 0.06 | 0.06 |
| Malawi | iLiNS−DYADM (30) | KDI | 152 | 299 | 11.7 | −0.03 (−0.09, 0.03) | 0.04 | 0.04 | 47 | 112 | 8.0 | 0.03 (−0.07, 0.12) | 0.05 | 0.05 |
| Malawi | iLiNS−DOSE (31) | KDI | | | | | | | | | | | | |
| Mali | PROMIS CS (32) | DMC | | | | | | | | | | | | |
| Zimbabwe | SHINE (HIV−) (33) | MDAT | 583 | 560 | 8.9 | 0.01 (−0.03, 0.04) | 0.11 | 0.11 | 197 | 180 | 16.1 | −0.04 (−0.11, 0.03) | 0.09 | 0.09 |
| Zimbabwe | SHINE (HIV+) (34) | MDAT | | | | | | | | | | | | |
| | | | **4150** | **7636** | | **I² = 0.00, Tau² = 0.00** | | | **1539** | **2795** | | **I² = 0.00, Tau² = 0.00** | | |
| **Fixed** | | | | | | **−0.01 (−0.02, 0.00)** | | | | | | **−0.02 (−0.04, 0.00)** | | |
| **Random** | | | | | | **−0.01 (−0.02, 0.00)** | | | | | | **−0.02 (−0.04, 0.00)** | | |

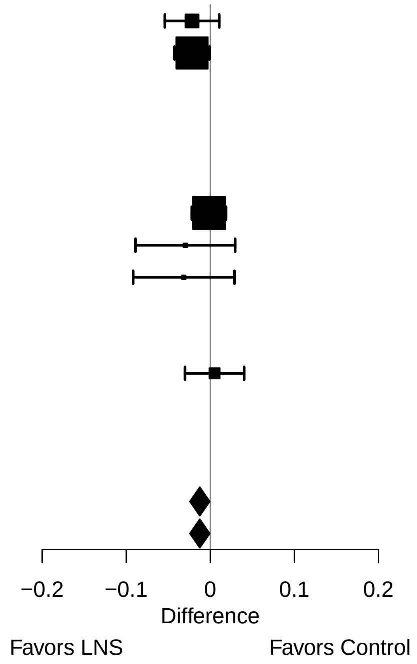

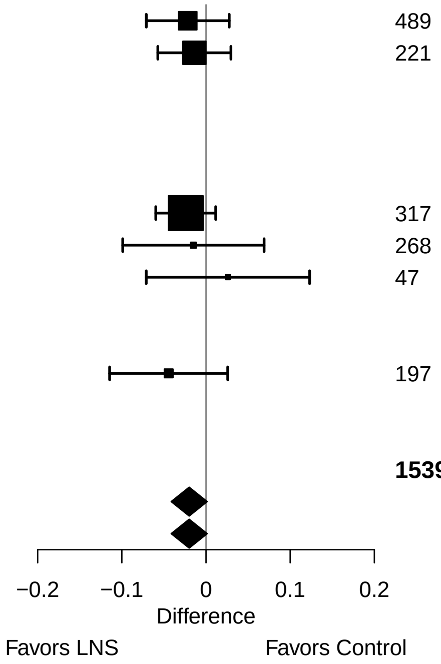

**Supplemental figure 5I: Motor lowest decile prevalence difference**

#### 5I6: Stratified by Child sex

**P−for−interaction = 0.917**
**Difference in PDs = 0.00 (−0.02, 0.02)**

| Country | Trial | Tool | LNS N | Control N | Control Prevalence | Male PD (95% CI) | Fixed W | Random W | LNS N | Control N | Control Prevalence | Female PD (95% CI) | Fixed W | Random W |
|---|---|---|---|---|---|---|---|---|---|---|---|---|---|---|
| Bangladesh | JiVitA−4 (21) | BSID−III | | | | | | | | | | | | |
| Bangladesh | RDNS (22) | DMC | 782 | 386 | 10.4 | 0.00 (−0.04, 0.03) | 0.12 | 0.12 | 774 | 367 | 12.0 | −0.03 (−0.06, 0.00) | 0.12 | 0.12 |
| Bangladesh | WASH−B (23) | EASQ | 528 | 1646 | 14.1 | −0.01 (−0.05, 0.02) | 0.15 | 0.15 | 546 | 1633 | 7.0 | −0.02 (−0.04, 0.00) | 0.30 | 0.16 |
| Burkina Faso | iLiNS−Zinc (24) | DMC | 372 | 193 | 14.0 | −0.06 (−0.10, −0.03) | 0.11 | 0.11 | 374 | 182 | 16.5 | −0.09 (−0.14, −0.04) | 0.06 | 0.09 |
| Ghana | GHANA (25) | | | | | | | | | | | | | |
| Ghana | iLiNS−DYADG (26) | KDI | 150 | 280 | 8.6 | 0.00 (−0.05, 0.06) | 0.06 | 0.06 | 152 | 321 | 10.6 | 0.03 (−0.04, 0.09) | 0.04 | 0.06 |
| Haiti | HAITI (27) | | | | | | | | | | | | | |
| Kenya | WASH−B (28) | EASQ | 682 | 2260 | 10.0 | −0.01 (−0.04, 0.02) | 0.22 | 0.22 | 680 | 2485 | 10.6 | −0.02 (−0.04, 0.01) | 0.22 | 0.15 |
| Madagascar | MAHAY (29) | ASQI | 795 | 787 | 11.1 | −0.02 (−0.07, 0.03) | 0.08 | 0.08 | 818 | 817 | 9.9 | 0.00 (−0.06, 0.06) | 0.04 | 0.07 |
| Malawi | iLiNS−DYADM (30) | KDI | 104 | 204 | 13.2 | −0.07 (−0.15, 0.00) | 0.03 | 0.03 | 110 | 232 | 8.2 | 0.04 (−0.03, 0.10) | 0.03 | 0.06 |
| Malawi | iLiNS−DOSE (31) | KDI | 321 | 117 | 10.3 | 0.00 (−0.06, 0.07) | 0.04 | 0.04 | 325 | 104 | 8.7 | 0.01 (−0.05, 0.08) | 0.03 | 0.06 |
| Mali | PROMIS CS (32) | DMC | 456 | 501 | 10.6 | −0.02 (−0.06, 0.03) | 0.08 | 0.08 | 446 | 420 | 11.7 | −0.02 (−0.07, 0.02) | 0.06 | 0.09 |
| Zimbabwe | SHINE (HIV−) (33) | MDAT | 424 | 388 | 9.8 | −0.01 (−0.05, 0.04) | 0.09 | 0.09 | 417 | 411 | 10.9 | −0.01 (−0.05, 0.04) | 0.07 | 0.10 |
| Zimbabwe | SHINE (HIV+) (34) | MDAT | 84 | 79 | 13.9 | −0.06 (−0.15, 0.04) | 0.02 | 0.02 | 86 | 72 | 11.1 | −0.03 (−0.11, 0.05) | 0.02 | 0.04 |
| | | | **4698** | **6841** | | **I² = 0.03, Tau² = 0.00** | | | **4728** | **7044** | | **I² = 0.36, Tau² = 0.00** | | |
| **Fixed** | | | | | | **−0.02 (−0.03, −0.01)** | | | | | | **−0.02 (−0.03, −0.01)** | | |
| **Random** | | | | | | **−0.02 (−0.03, −0.01)** | | | | | | **−0.02 (−0.04, 0.00)** | | |

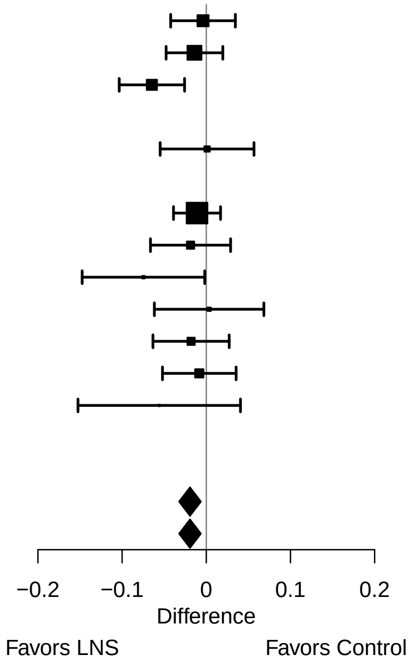

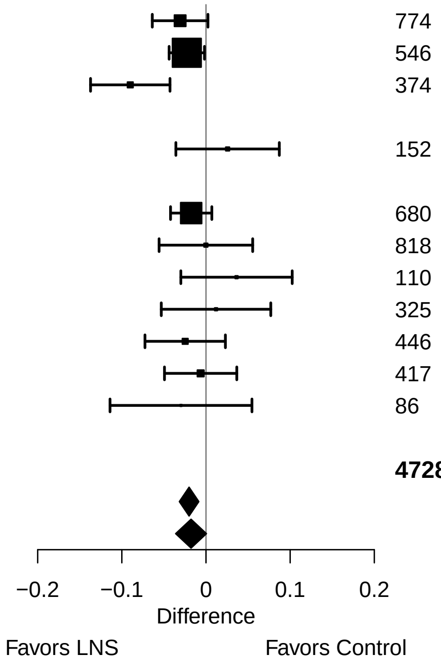

#### Supplemental figure 5I: Motor lowest decile prevalence difference

### 5I7: Stratified by Child birth order

**P−for−interaction = 0.035**
**Difference in PDs = 0.02 (0.00, 0.04)**

| Country | Trial | Tool | LNS N | Control N | Control Prevalence | Later born PD (95% CI) | Fixed W | Random W | LNS N | Control N | Control Prevalence | Firstborn PD (95% CI) | Fixed W | Random W |
|---|---|---|---|---|---|---|---|---|---|---|---|---|---|---|
| Bangladesh | JiVitA−4 (21) | BSID−III | | | | | | | | | | | | |
| Bangladesh | RDNS (22) | DMC | 909 | 469 | 12.4 | −0.03 (−0.06, 0.00) | 0.14 | 0.14 | 646 | 284 | 9.2 | 0.01 (−0.03, 0.05) | 0.19 | 0.19 |
| Bangladesh | WASH−B (23) | EASQ | 675 | 2149 | 10.5 | −0.02 (−0.05, 0.00) | 0.22 | 0.15 | 379 | 1028 | 10.2 | −0.01 (−0.04, 0.02) | 0.30 | 0.30 |
| Burkina Faso | iLiNS−Zinc (24) | DMC | 587 | 290 | 16.6 | −0.10 (−0.14, −0.06) | 0.08 | 0.12 | 159 | 85 | 10.6 | 0.01 (−0.07, 0.09) | 0.05 | 0.05 |
| Ghana | GHANA (25) | | | | | | | | | | | | | |
| Ghana | iLiNS−DYADG (26) | KDI | 203 | 406 | 11.1 | 0.01 (−0.05, 0.06) | 0.04 | 0.09 | 99 | 195 | 6.7 | 0.02 (−0.04, 0.09) | 0.08 | 0.08 |
| Haiti | HAITI (27) | | | | | | | | | | | | | |
| Kenya | WASH−B (28) | EASQ | 1071 | 3767 | 10.6 | −0.02 (−0.04, 0.01) | 0.25 | 0.16 | 289 | 974 | 9.1 | −0.01 (−0.05, 0.03) | 0.18 | 0.18 |
| Madagascar | MAHAY (29) | ASQI | 1162 | 1118 | 10.9 | −0.01 (−0.06, 0.04) | 0.05 | 0.10 | 433 | 465 | 9.9 | −0.01 (−0.07, 0.05) | 0.09 | 0.09 |
| Malawi | iLiNS−DYADM (30) | KDI | | | | | | | | | | | | |
| Malawi | iLiNS−DOSE (31) | KDI | | | | | | | | | | | | |
| Mali | PROMIS CS (32) | DMC | 749 | 786 | 11.1 | −0.03 (−0.06, 0.01) | 0.11 | 0.13 | 139 | 123 | 8.9 | 0.02 (−0.06, 0.10) | 0.05 | 0.05 |
| Zimbabwe | SHINE (HIV−) (33) | MDAT | 602 | 574 | 9.6 | −0.02 (−0.05, 0.02) | 0.10 | 0.13 | 203 | 191 | 14.1 | 0.01 (−0.06, 0.07) | 0.07 | 0.07 |
| Zimbabwe | SHINE (HIV+) (34) | MDAT | | | | | | | | | | | | |
| | | | **5958** | **9559** | | **I² = 0.61, Tau² = 0.00** | | | **2347** | **3345** | | **I² = 0.00, Tau² = 0.00** | | |
| **Fixed** | | | | | | **−0.03 (−0.04, −0.02)** | | | | | | **0.00 (−0.02, 0.02)** | | |
| **Random** | | | | | | **−0.03 (−0.05, −0.01)** | | | | | | **0.00 (−0.02, 0.02)** | | |

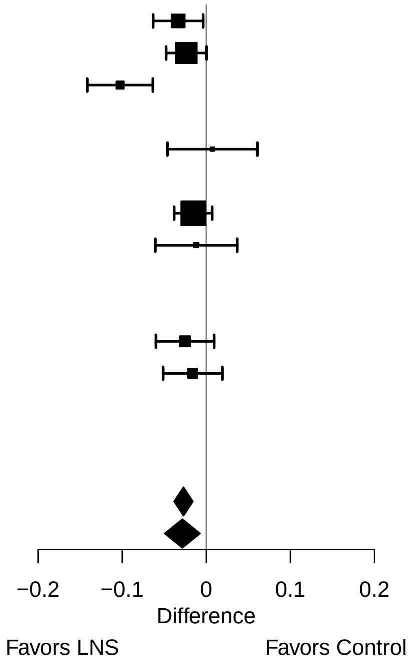

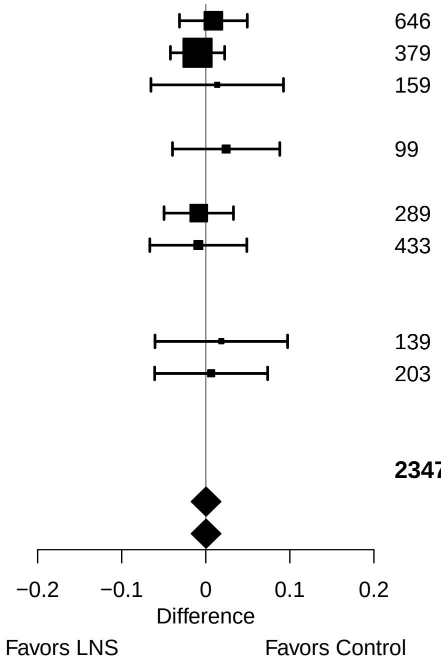

##### Supplemental figure 5I: Motor lowest decile prevalence difference

#### 5I8: Stratified by Child baseline stunting

**P−for−interaction = 0.226**
**Difference in PDs = −0.03 (−0.07, 0.02)**

| Country | Trial | Tool | No: LNS N | No: Control N | No: Control Prevalence | No: PD (95% CI) | No: Fixed W | No: Random W | Yes: LNS N | Yes: Control N | Yes: Control Prevalence | Yes: PD (95% CI) | Yes: Fixed W | Yes: Random W |
|---|---|---|---|---|---|---|---|---|---|---|---|---|---|---|
| Bangladesh | JiVitA−4 (21) | BSID−III | | | | | | | | | | | | |
| Bangladesh | RDNS (22) | DMC | 1170 | 535 | 10.1 | −0.02 (−0.04, 0.01) | 0.32 | 0.24 | 327 | 166 | 15.1 | −0.04 (−0.09, 0.02) | 0.47 | 0.33 |
| Bangladesh | WASH−B (23) | EASQ | | | | | | | | | | | | |
| Burkina Faso | iLiNS−Zinc (24) | DMC | 576 | 294 | 11.6 | −0.07 (−0.09, −0.04) | 0.34 | 0.24 | 170 | 80 | 28.7 | −0.12 (−0.24, 0.00) | 0.10 | 0.14 |
| Ghana | GHANA (25) | | | | | | | | | | | | | |
| Ghana | iLiNS−DYADG (26) | KDI | | | | | | | | | | | | |
| Haiti | HAITI (27) | | | | | | | | | | | | | |
| Kenya | WASH−B (28) | EASQ | | | | | | | | | | | | |
| Madagascar | MAHAY (29) | ASQI | | | | | | | | | | | | |
| Malawi | iLiNS−DYADM (30) | KDI | 152 | 320 | 8.4 | −0.01 (−0.06, 0.05) | 0.08 | 0.15 | 48 | 97 | 14.4 | −0.02 (−0.14, 0.10) | 0.10 | 0.14 |
| Malawi | iLiNS−DOSE (31) | KDI | 465 | 147 | 8.2 | −0.01 (−0.06, 0.04) | 0.09 | 0.16 | 181 | 74 | 12.2 | 0.05 (−0.05, 0.15) | 0.15 | 0.18 |
| Mali | PROMIS CS (32) | DMC | | | | | | | | | | | | |
| Zimbabwe | SHINE (HIV−) (33) | MDAT | 532 | 490 | 8.2 | 0.01 (−0.02, 0.05) | 0.16 | 0.20 | 122 | 105 | 19.0 | −0.09 (−0.18, 0.00) | 0.18 | 0.21 |
| Zimbabwe | SHINE (HIV+) (34) | MDAT | | | | | | | | | | | | |
| | | | **2895** | **1786** | | **I² = 0.75, Tau² = 0.00** | | | **848** | **522** | | **I² = 0.35, Tau² = 0.00** | | |
| **Fixed** | | | | | | **−0.03 (−0.04, −0.01)** | | | | | | **−0.04 (−0.08, 0.00)** | | |
| **Random** | | | | | | **−0.02 (−0.05, 0.01)** | | | | | | **−0.04 (−0.09, 0.01)** | | |

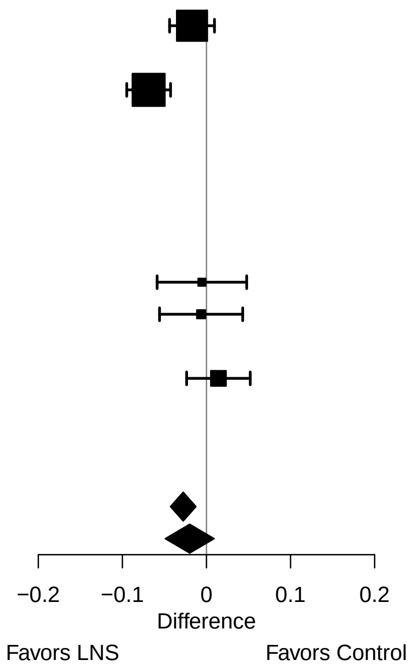

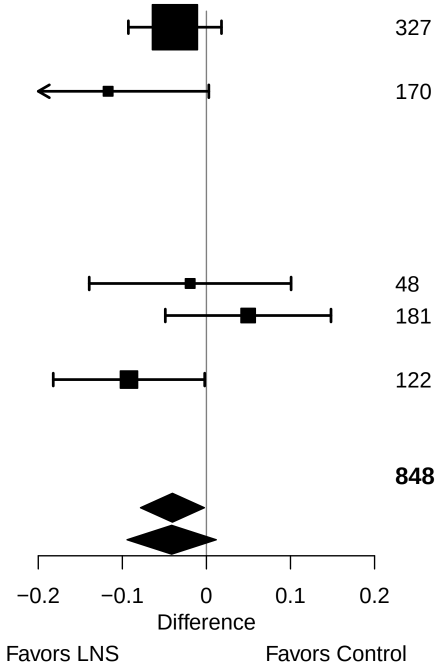

#### Supplemental figure 5I: Motor lowest decile prevalence difference

#### 5I9: Stratified by Child baseline acute malnutrition

**P−for−interaction = 0.540**
**Difference in PDs = −0.02 (−0.09, 0.05)**

| Country | Trial | Tool | No: LNS N | No: Control N | No: Control Prevalence | No: PD (95% CI) | No: Fixed W | No: Random W | Yes: LNS N | Yes: Control N | Yes: Control Prevalence | Yes: PD (95% CI) | Yes: Fixed W | Yes: Random W |
|---|---|---|---|---|---|---|---|---|---|---|---|---|---|---|
| Bangladesh | JiVitA−4 (21) | BSID−III | 359 | 112 | 5.4 | 0.05 (0.00, 0.10) | 0.14 | 0.30 | 84 | 31 | 19.4 | −0.06 (−0.21, 0.09) | 0.22 | 0.27 |
| Bangladesh | RDNS (22) | DMC | 1382 | 653 | 11.2 | −0.03 (−0.05, 0.00) | 0.58 | 0.36 | 115 | 49 | 12.2 | 0.04 (−0.07, 0.16) | 0.37 | 0.36 |
| Bangladesh | WASH−B (23) | EASQ | | | | | | | | | | | | |
| Burkina Faso | iLiNS−Zinc (24) | DMC | 556 | 266 | 10.2 | −0.05 (−0.09, −0.02) | 0.28 | 0.34 | 190 | 109 | 27.5 | −0.12 (−0.23, −0.01) | 0.41 | 0.38 |
| Ghana | GHANA (25) | | | | | | | | | | | | | |
| Ghana | iLiNS−DYADG (26) | KDI | | | | | | | | | | | | |
| Haiti | HAITI (27) | | | | | | | | | | | | | |
| Kenya | WASH−B (28) | EASQ | | | | | | | | | | | | |
| Madagascar | MAHAY (29) | ASQI | | | | | | | | | | | | |
| Malawi | iLiNS−DYADM (30) | KDI | | | | | | | | | | | | |
| Malawi | iLiNS−DOSE (31) | KDI | | | | | | | | | | | | |
| Mali | PROMIS CS (32) | DMC | | | | | | | | | | | | |
| Zimbabwe | SHINE (HIV−) (33) | MDAT | | | | | | | | | | | | |
| Zimbabwe | SHINE (HIV+) (34) | MDAT | | | | | | | | | | | | |
| | | | **2297** | **1031** | | **I² = 0.82, Tau² = 0.00** | | | **389** | **189** | | **I² = 0.50, Tau² = 0.00** | | |
| **Fixed** | | | | | | **−0.03 (−0.04, −0.01)** | | | | | | **−0.05 (−0.12, 0.02)** | | |
| **Random** | | | | | | **−0.01 (−0.07, 0.04)** | | | | | | **−0.05 (−0.14, 0.05)** | | |

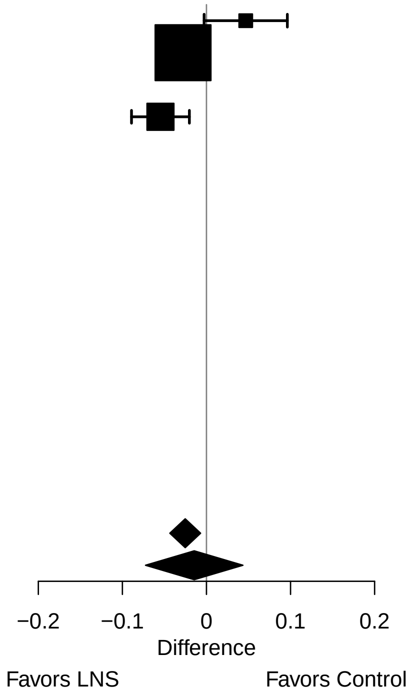

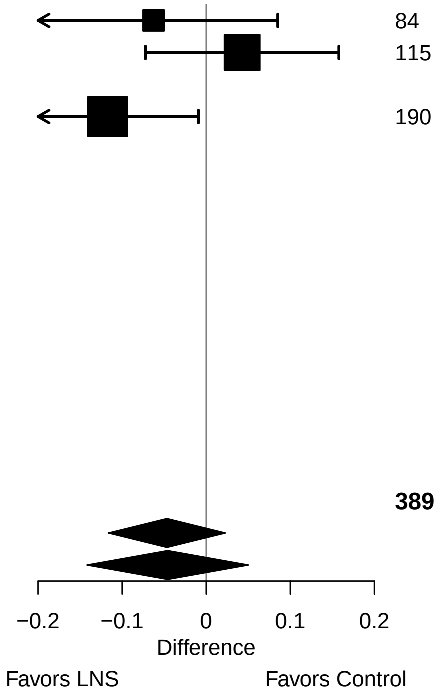

#### Supplemental figure 5I: Motor lowest decile prevalence difference

##### 5I10: Stratified by Child baseline anemia

**P−for−interaction = 0.177**
**Difference in PDs = −0.03 (−0.08, 0.02)**

| Country | Trial | Tool | Not anemic LNS N | Not anemic Control N | Not anemic Control Prevalence | Not anemic PD (95% CI) | Not anemic Fixed W | Not anemic Random W | Anemic LNS N | Anemic Control N | Anemic Control Prevalence | Anemic PD (95% CI) | Anemic Fixed W | Anemic Random W |
|---|---|---|---|---|---|---|---|---|---|---|---|---|---|---|
| Bangladesh | JiVitA−4 (21) | BSID−III | | | | | | | | | | | | |
| Bangladesh | RDNS (22) | DMC | 198 | 91 | 15.4 | −0.07 (−0.14, 0.00) | 0.22 | 0.24 | 303 | 151 | 12.6 | −0.04 (−0.10, 0.01) | 0.34 | 0.34 |
| Bangladesh | WASH−B (23) | EASQ | | | | | | | | | | | | |
| Burkina Faso | iLiNS−Zinc (24) | DMC | | | | | | | | | | | | |
| Ghana | GHANA (25) | | | | | | | | | | | | | |
| Ghana | iLiNS−DYADG (26) | KDI | 180 | 321 | 8.7 | 0.05 (0.00, 0.11) | 0.36 | 0.29 | 81 | 189 | 12.7 | −0.05 (−0.13, 0.03) | 0.15 | 0.15 |
| Haiti | HAITI (27) | | | | | | | | | | | | | |
| Kenya | WASH−B (28) | EASQ | | | | | | | | | | | | |
| Madagascar | MAHAY (29) | ASQI | | | | | | | | | | | | |
| Malawi | iLiNS−DYADM (30) | KDI | 65 | 146 | 7.5 | 0.02 (−0.06, 0.10) | 0.17 | 0.21 | 138 | 273 | 11.7 | −0.02 (−0.09, 0.04) | 0.25 | 0.25 |
| Malawi | iLiNS−DOSE (31) | KDI | 224 | 94 | 7.4 | 0.01 (−0.06, 0.08) | 0.25 | 0.26 | 418 | 126 | 11.1 | 0.00 (−0.06, 0.06) | 0.26 | 0.26 |
| Mali | PROMIS CS (32) | DMC | | | | | | | | | | | | |
| Zimbabwe | SHINE (HIV−) (33) | MDAT | | | | | | | | | | | | |
| Zimbabwe | SHINE (HIV+) (34) | MDAT | | | | | | | | | | | | |
| | | | **667** | **652** | | **I² = 0.55, Tau² = 0.00** | | | **940** | **739** | | **I² = 0.00, Tau² = 0.00** | | |
| **Fixed** | | | | | | **0.01 (−0.02, 0.04)** | | | | | | **−0.03 (−0.06, 0.00)** | | |
| **Random** | | | | | | **0.01 (−0.04, 0.06)** | | | | | | **−0.03 (−0.06, 0.00)** | | |

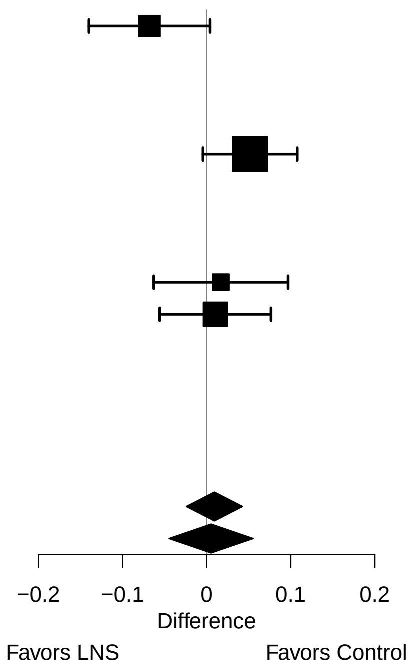

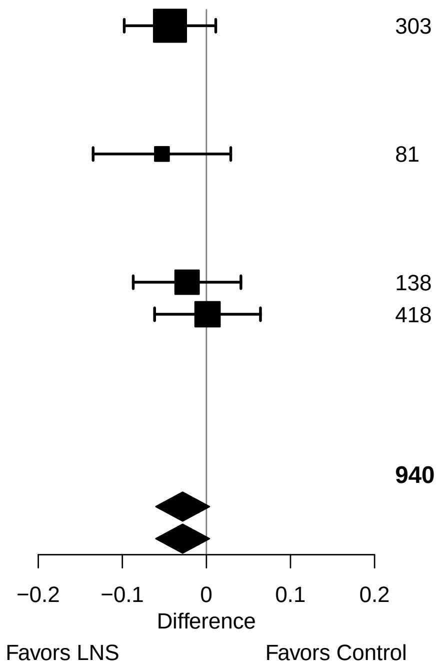

### Supplemental figure 5J: Mean difference in gross motor z-score

#### 5J1: Stratified by Maternal height

**P−for−interaction = 0.787**
**Difference in MDs = −0.01 (−0.09, 0.06)**

| | | | At least 150.1 cm | | | | | | Less than 150.1 cm | | | | | |
|---|---|---|---|---|---|---|---|---|---|---|---|---|---|---|
| Country | Trial | Tool | LNS N | Control N | Control Mean | MD (95% CI) | Fixed W | Random W | LNS N | Control N | Control Mean | MD (95% CI) | Fixed W | Random W |
| Bangladesh | JiVitA−4 (21) | BSID−III | | | | | | | | | | | | |
| Bangladesh | RDNS (22) | DMC | 824 | 399 | −0.04 | 0.15 (0.08, 0.22) | 0.22 | 0.15 | 691 | 337 | −0.04 | 0.03 (−0.08, 0.13) | 0.35 | 0.35 |
| Bangladesh | WASH−B (23) | EASQ | 568 | 1773 | 0.04 | 0.09 (−0.02, 0.19) | 0.10 | 0.12 | 501 | 1467 | −0.09 | 0.13 (0.03, 0.23) | 0.41 | 0.41 |
| Burkina Faso | iLiNS−Zinc (24) | DMC | | | | | | | | | | | | |
| Ghana | GHANA (25) | | | | | | | | | | | | | |
| Ghana | iLiNS−DYADG (26) | KDI | 284 | 557 | 0.02 | −0.02 (−0.16, 0.13) | 0.05 | 0.08 | 13 | 34 | −0.28 | 0.44 (−0.17, 1.05) | 0.01 | 0.01 |
| Haiti | HAITI (27) | | | | | | | | | | | | | |
| Kenya | WASH−B (28) | EASQ | 1237 | 4372 | 0.01 | 0.00 (−0.07, 0.06) | 0.25 | 0.15 | 51 | 175 | −0.21 | 0.27 (−0.01, 0.56) | 0.05 | 0.05 |
| Madagascar | MAHAY (29) | ASQI | 956 | 986 | −0.01 | 0.07 (−0.10, 0.23) | 0.04 | 0.07 | 553 | 538 | −0.01 | 0.08 (−0.12, 0.28) | 0.10 | 0.10 |
| Malawi | iLiNS−DYADM (30) | KDI | 186 | 376 | −0.01 | 0.00 (−0.17, 0.17) | 0.04 | 0.07 | 28 | 57 | 0.19 | −0.23 (−0.67, 0.21) | 0.02 | 0.02 |
| Malawi | iLiNS−DOSE (31) | KDI | 543 | 185 | 0.03 | −0.02 (−0.18, 0.15) | 0.04 | 0.07 | 102 | 35 | −0.16 | 0.09 (−0.28, 0.47) | 0.03 | 0.03 |
| Mali | PROMIS CS (32) | DMC | 902 | 924 | −0.04 | 0.09 (0.00, 0.17) | 0.16 | 0.13 | 32 | 27 | −0.09 | 0.17 (−0.42, 0.76) | 0.01 | 0.01 |
| Zimbabwe | SHINE (HIV−) (33) | MDAT | 788 | 762 | −0.05 | 0.10 (−0.02, 0.21) | 0.08 | 0.11 | 34 | 22 | −0.14 | 0.18 (−0.22, 0.59) | 0.02 | 0.02 |
| Zimbabwe | SHINE (HIV+) (34) | MDAT | 158 | 140 | −0.17 | 0.34 (0.14, 0.54) | 0.03 | 0.05 | 10 | 6 | −0.12 | 0.02 (−0.77, 0.80) | 0.01 | 0.01 |
| | | | **6446** | **10474** | | **I² = 0.56, Tau² = 0.00** | | | **2015** | **2698** | | **I² = 0.00, Tau² = 0.00** | | |
| **Fixed** | | | | | | **0.07 (0.04, 0.11)** | | | | | | **0.09 (0.03, 0.15)** | | |
| **Random** | | | | | | **0.07 (0.02, 0.13)** | | | | | | **0.09 (0.03, 0.15)** | | |

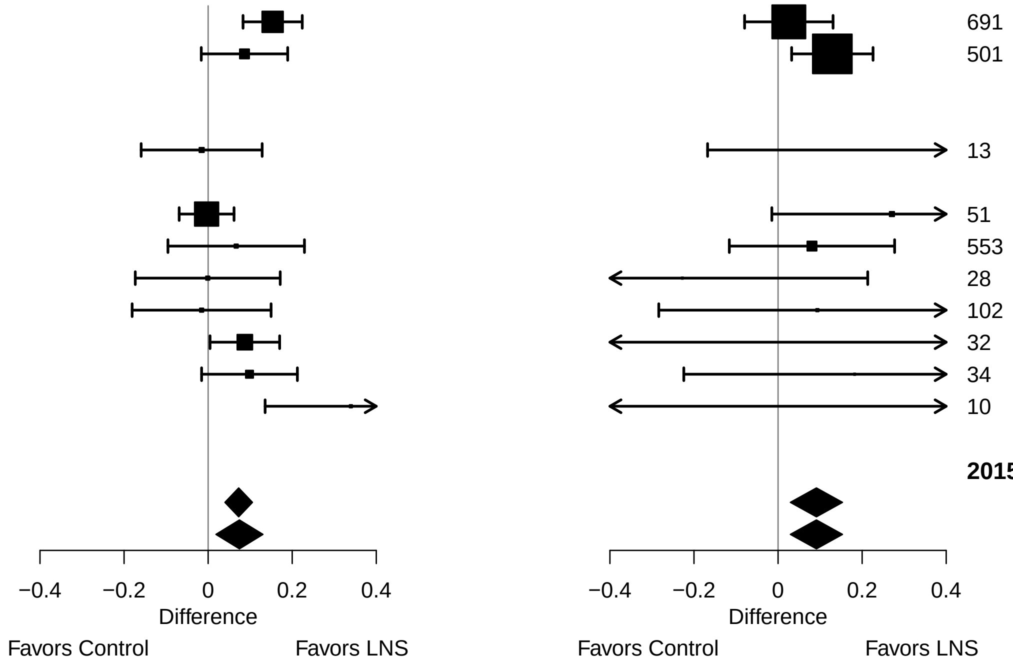

##### Supplemental figure 5J: Mean difference in gross motor z-score

#### 5J2: Stratified by Maternal BMI

**P−for−interaction = 0.642**
**Difference in MDs = 0.01 (−0.05, 0.08)**

| | | | At least 20 kg/m² | | | | | | Less than 20 kg/m² | | | | | |
|---|---|---|---|---|---|---|---|---|---|---|---|---|---|---|
| Country | Trial | Tool | LNS N | Control N | Control Mean | MD (95% CI) | Fixed W | Random W | LNS N | Control N | Control Mean | MD (95% CI) | Fixed W | Random W |
| Bangladesh | JiVitA−4 (21) | BSID−III | | | | | | | | | | | | |
| Bangladesh | RDNS (22) | DMC | 693 | 308 | −0.01 | 0.09 (0.00, 0.18) | 0.17 | 0.15 | 822 | 428 | −0.06 | 0.09 (0.02, 0.17) | 0.40 | 0.40 |
| Bangladesh | WASH−B (23) | EASQ | 478 | 1502 | 0.05 | 0.09 (−0.02, 0.19) | 0.13 | 0.14 | 591 | 1737 | −0.09 | 0.12 (0.03, 0.22) | 0.24 | 0.24 |
| Burkina Faso | iLiNS−Zinc (24) | DMC | | | | | | | | | | | | |
| Ghana | GHANA (25) | | | | | | | | | | | | | |
| Ghana | iLiNS−DYADG (26) | KDI | 262 | 485 | 0.01 | 0.02 (−0.13, 0.17) | 0.06 | 0.09 | 35 | 106 | −0.03 | −0.09 (−0.48, 0.30) | 0.01 | 0.01 |
| Haiti | HAITI (27) | | | | | | | | | | | | | |
| Kenya | WASH−B (28) | EASQ | 1031 | 3535 | 0.01 | −0.02 (−0.09, 0.05) | 0.30 | 0.18 | 257 | 1012 | −0.04 | 0.12 (−0.02, 0.27) | 0.11 | 0.11 |
| Madagascar | MAHAY (29) | ASQI | | | | | | | | | | | | |
| Malawi | iLiNS−DYADM (30) | KDI | 126 | 258 | −0.05 | 0.00 (−0.23, 0.23) | 0.03 | 0.05 | 87 | 175 | 0.12 | −0.08 (−0.28, 0.13) | 0.05 | 0.05 |
| Malawi | iLiNS−DOSE (31) | KDI | 480 | 158 | 0.02 | 0.01 (−0.16, 0.18) | 0.05 | 0.08 | 163 | 61 | −0.06 | −0.01 (−0.33, 0.31) | 0.02 | 0.02 |
| Mali | PROMIS CS (32) | DMC | 693 | 676 | −0.07 | 0.12 (0.02, 0.22) | 0.14 | 0.14 | 241 | 275 | 0.01 | 0.01 (−0.12, 0.15) | 0.12 | 0.12 |
| Zimbabwe | SHINE (HIV−) (33) | MDAT | 601 | 581 | −0.04 | 0.11 (−0.02, 0.24) | 0.09 | 0.11 | 100 | 96 | −0.18 | 0.19 (−0.07, 0.44) | 0.03 | 0.03 |
| Zimbabwe | SHINE (HIV+) (34) | MDAT | 124 | 113 | −0.13 | 0.35 (0.13, 0.58) | 0.03 | 0.05 | 25 | 23 | −0.42 | 0.27 (−0.16, 0.69) | 0.01 | 0.01 |
| | | | **4488** | **7616** | | **I² = 0.49, Tau² = 0.00** | | | **2321** | **3913** | | **I² = 0.00, Tau² = 0.00** | | |
| **Fixed** | | | | | | **0.06 (0.02, 0.10)** | | | | | | **0.09 (0.04, 0.13)** | | |
| **Random** | | | | | | **0.07 (0.01, 0.13)** | | | | | | **0.09 (0.04, 0.13)** | | |

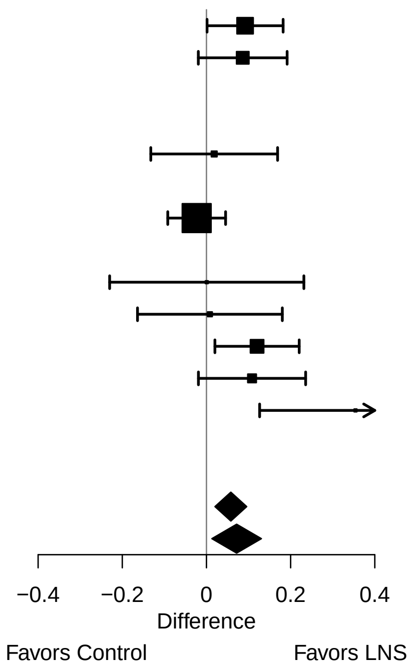

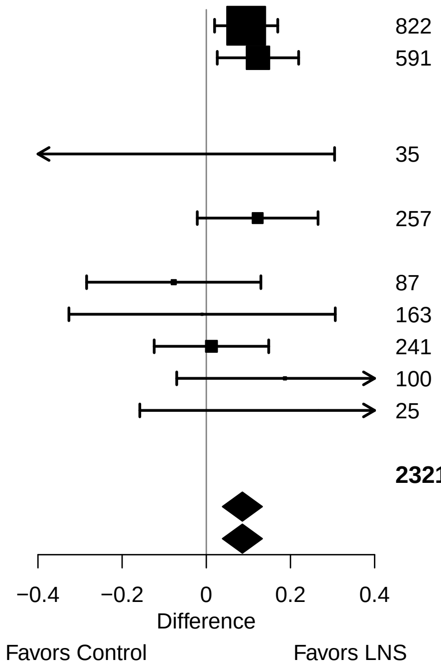

#### Supplemental figure 5J: Mean difference in gross motor z-score

##### 5J3: Stratified by Maternal age

P−for−interaction = 0.610
Difference in MDs = −0.02 (−0.07, 0.04)

| Country | Trial | Tool | At least 25 y LNS N | Control N | Control Mean | MD (95% CI) | Fixed W | Random W | Less than 25 y LNS N | Control N | Control Mean | MD (95% CI) | Fixed W | Random W |
|---|---|---|---|---|---|---|---|---|---|---|---|---|---|---|
| Bangladesh | JiVitA−4 (21) | BSID−III | 200 | 68 | 0.08 | −0.07 (−0.29, 0.14) | 0.04 | 0.06 | 242 | 74 | 0.12 | −0.15 (−0.44, 0.14) | 0.02 | 0.02 |
| Bangladesh | RDNS (22) | DMC | 426 | 202 | −0.12 | 0.12 (−0.03, 0.26) | 0.08 | 0.10 | 1144 | 556 | 0.00 | 0.07 (0.00, 0.15) | 0.30 | 0.26 |
| Bangladesh | WASH−B (23) | EASQ | 466 | 1450 | −0.02 | 0.09 (−0.03, 0.20) | 0.14 | 0.13 | 603 | 1815 | −0.03 | 0.11 (0.03, 0.20) | 0.22 | 0.21 |
| Burkina Faso | iLiNS−Zinc (24) | DMC | | | | | | | | | | | | |
| Ghana | GHANA (25) | | | | | | | | | | | | | |
| Ghana | iLiNS−DYADG (26) | KDI | 192 | 378 | −0.02 | 0.07 (−0.10, 0.24) | 0.06 | 0.08 | 110 | 223 | 0.02 | −0.10 (−0.33, 0.14) | 0.03 | 0.03 |
| Haiti | HAITI (27) | | | | | | | | | | | | | |
| Kenya | WASH−B (28) | EASQ | 798 | 2622 | 0.04 | −0.02 (−0.10, 0.05) | 0.30 | 0.18 | 549 | 2092 | −0.05 | 0.05 (−0.05, 0.16) | 0.15 | 0.15 |
| Madagascar | MAHAY (29) | ASQI | 852 | 901 | −0.09 | 0.09 (−0.08, 0.25) | 0.07 | 0.08 | 759 | 703 | 0.03 | 0.06 (−0.11, 0.23) | 0.06 | 0.06 |
| Malawi | iLiNS−DYADM (30) | KDI | 102 | 224 | 0.06 | −0.05 (−0.28, 0.18) | 0.03 | 0.05 | 112 | 212 | −0.03 | −0.01 (−0.24, 0.21) | 0.03 | 0.04 |
| Malawi | iLiNS−DOSE (31) | KDI | 346 | 124 | −0.06 | 0.06 (−0.15, 0.26) | 0.04 | 0.06 | 288 | 90 | 0.08 | −0.07 (−0.29, 0.16) | 0.03 | 0.04 |
| Mali | PROMIS CS (32) | DMC | 495 | 500 | −0.08 | 0.15 (0.03, 0.27) | 0.12 | 0.12 | 445 | 457 | 0.00 | 0.02 (−0.11, 0.14) | 0.10 | 0.10 |
| Zimbabwe | SHINE (HIV−) (33) | MDAT | 427 | 389 | −0.02 | 0.15 (0.00, 0.30) | 0.08 | 0.10 | 326 | 321 | −0.11 | 0.09 (−0.06, 0.25) | 0.07 | 0.07 |
| Zimbabwe | SHINE (HIV+) (34) | MDAT | 126 | 111 | −0.15 | 0.31 (0.09, 0.54) | 0.04 | 0.05 | 29 | 26 | −0.27 | 0.63 (−0.01, 1.26) | 0.00 | 0.00 |
| | | | **4430** | **6969** | | **$I^2$ = 0.38, $Tau^2$ = 0.00** | | | **4607** | **6569** | | **$I^2$ = 0.06, $Tau^2$ = 0.00** | | |
| **Fixed** | | | | | | **0.06 (0.02, 0.11)** | | | | | | **0.06 (0.02, 0.10)** | | |
| **Random** | | | | | | **0.08 (0.02, 0.13)** | | | | | | **0.06 (0.01, 0.10)** | | |

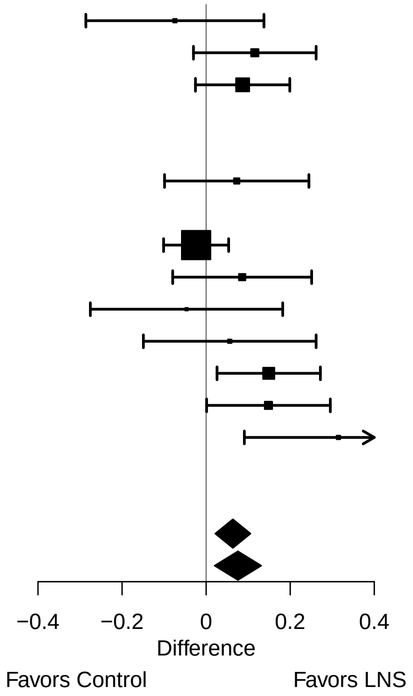

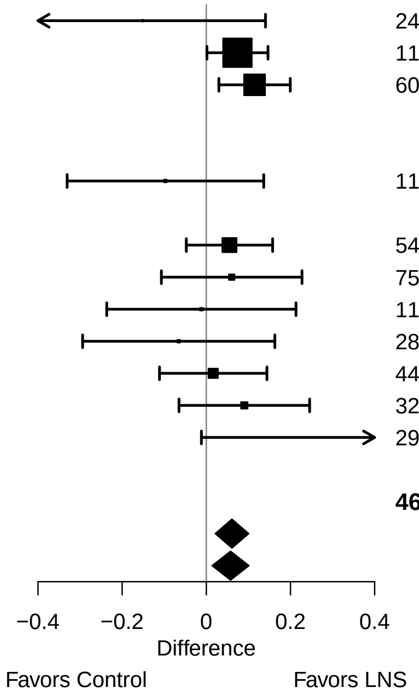

#### Supplemental figure 5J: Mean difference in gross motor z-score

### 5J4: Stratified by Maternal education

**P−for−interaction = 0.104**
**Difference in MDs = 0.06 (−0.01, 0.13)**

| Country | Trial | Tool | Primary or greater: LNS N | Control N | Control Mean | MD (95% CI) | Fixed W | Random W | Incomplete or no formal: LNS N | Control N | Control Mean | MD (95% CI) | Fixed W | Random W |
|---|---|---|---|---|---|---|---|---|---|---|---|---|---|---|
| Bangladesh | JiVitA−4 (21) | BSID−III | 292 | 92 | 0.24 | −0.19 (−0.37, −0.01) | 0.04 | 0.09 | 152 | 51 | −0.19 | 0.06 (−0.29, 0.40) | 0.02 | 0.02 |
| Bangladesh | RDNS (22) | DMC | 1178 | 549 | 0.03 | 0.09 (0.02, 0.15) | 0.31 | 0.14 | 392 | 209 | −0.19 | 0.05 (−0.11, 0.21) | 0.08 | 0.08 |
| Bangladesh | WASH−B (23) | EASQ | 755 | 2344 | 0.04 | 0.08 (0.00, 0.17) | 0.20 | 0.13 | 319 | 935 | −0.17 | 0.16 (0.04, 0.29) | 0.13 | 0.13 |
| Burkina Faso | iLiNS−Zinc (24) | DMC | | | | | | | | | | | | |
| Ghana | GHANA (25) | | | | | | | | | | | | | |
| Ghana | iLiNS−DYADG (26) | KDI | 235 | 467 | −0.01 | 0.00 (−0.15, 0.15) | 0.06 | 0.10 | 67 | 134 | 0.00 | 0.04 (−0.28, 0.36) | 0.02 | 0.02 |
| Haiti | HAITI (27) | | | | | | | | | | | | | |
| Kenya | WASH−B (28) | EASQ | 658 | 2258 | 0.11 | −0.01 (−0.10, 0.08) | 0.16 | 0.13 | 702 | 2484 | −0.11 | 0.02 (−0.07, 0.12) | 0.23 | 0.23 |
| Madagascar | MAHAY (29) | ASQI | 339 | 416 | 0.20 | −0.07 (−0.26, 0.11) | 0.04 | 0.08 | 1274 | 1188 | −0.12 | 0.13 (−0.02, 0.29) | 0.08 | 0.08 |
| Malawi | iLiNS−DYADM (30) | KDI | 33 | 67 | 0.06 | −0.04 (−0.40, 0.31) | 0.01 | 0.04 | 180 | 366 | 0.01 | −0.04 (−0.21, 0.14) | 0.06 | 0.06 |
| Malawi | iLiNS−DOSE (31) | KDI | 148 | 52 | 0.22 | −0.04 (−0.29, 0.20) | 0.02 | 0.06 | 487 | 162 | −0.07 | 0.02 (−0.16, 0.21) | 0.06 | 0.06 |
| Mali | PROMIS CS (32) | DMC | 103 | 96 | 0.11 | −0.04 (−0.34, 0.25) | 0.02 | 0.05 | 836 | 861 | −0.06 | 0.10 (0.02, 0.18) | 0.30 | 0.30 |
| Zimbabwe | SHINE (HIV−) (33) | MDAT | 763 | 731 | −0.05 | 0.12 (0.01, 0.24) | 0.11 | 0.12 | 29 | 25 | −0.02 | −0.26 (−0.74, 0.22) | 0.01 | 0.01 |
| Zimbabwe | SHINE (HIV+) (34) | MDAT | 149 | 130 | −0.16 | 0.37 (0.17, 0.57) | 0.03 | 0.08 | 9 | 8 | −0.12 | 0.04 (−0.57, 0.65) | 0.01 | 0.01 |
| | | | **4653** | **7202** | | **I² = 0.60, Tau² = 0.01** | | | **4447** | **6423** | | **I² = 0.00, Tau² = 0.00** | | |
| **Fixed** | | | | | | **0.06 (0.02, 0.09)** | | | | | | **0.07 (0.02, 0.11)** | | |
| **Random** | | | | | | **0.04 (−0.04, 0.12)** | | | | | | **0.07 (0.02, 0.11)** | | |

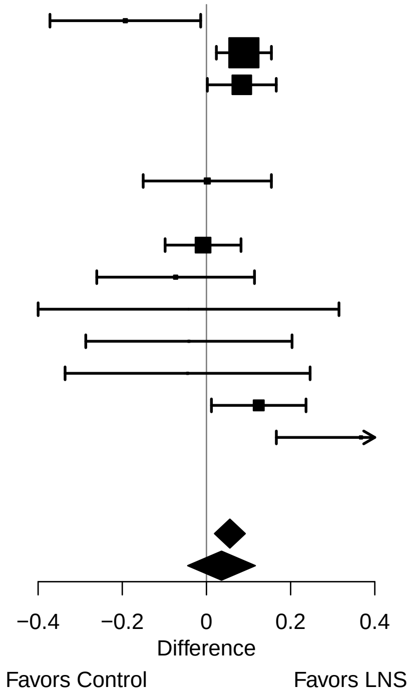

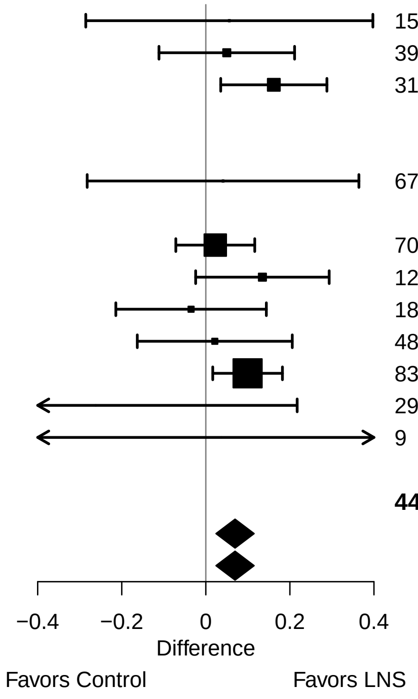

##### Supplemental figure 5J: Mean difference in gross motor z-score

#### 5J5: Stratified by Maternal depressive symptoms

**P−for−interaction = 0.753**
**Difference in MDs = −0.01 (−0.08, 0.06)**

| | | | Less than 75th percentile | | | | | | At least 75th percentile | | | | | |
|---|---|---|---|---|---|---|---|---|---|---|---|---|---|---|
| Country | Trial | Tool | LNS N | Control N | Control Mean | MD (95% CI) | Fixed W | Random W | LNS N | Control N | Control Mean | MD (95% CI) | Fixed W | Random W |
| Bangladesh | JiVitA−4 (21) | BSID−III | | | | | | | | | | | | |
| Bangladesh | RDNS (22) | DMC | 1018 | 436 | −0.01 | 0.10 (0.00, 0.20) | 0.16 | 0.15 | 493 | 270 | −0.07 | 0.05 (−0.06, 0.15) | 0.30 | 0.30 |
| Bangladesh | WASH−B (23) | EASQ | 832 | 2358 | −0.01 | 0.11 (0.04, 0.19) | 0.29 | 0.16 | 221 | 838 | −0.08 | 0.07 (−0.07, 0.22) | 0.15 | 0.15 |
| Burkina Faso | iLiNS−Zinc (24) | DMC | | | | | | | | | | | | |
| Ghana | GHANA (25) | | | | | | | | | | | | | |
| Ghana | iLiNS−DYADG (26) | KDI | 226 | 393 | 0.00 | −0.04 (−0.21, 0.12) | 0.05 | 0.11 | 67 | 190 | −0.03 | 0.16 (−0.10, 0.42) | 0.05 | 0.05 |
| Haiti | HAITI (27) | | | | | | | | | | | | | |
| Kenya | WASH−B (28) | EASQ | 960 | 3317 | 0.04 | −0.02 (−0.09, 0.06) | 0.29 | 0.16 | 317 | 1156 | 0.09 | 0.03 (−0.08, 0.13) | 0.33 | 0.33 |
| Madagascar | MAHAY (29) | ASQI | 615 | 667 | 0.05 | 0.09 (−0.08, 0.27) | 0.05 | 0.11 | 268 | 242 | −0.05 | 0.12 (−0.14, 0.39) | 0.05 | 0.05 |
| Malawi | iLiNS−DYADM (30) | KDI | 152 | 299 | −0.01 | −0.01 (−0.20, 0.18) | 0.04 | 0.10 | 47 | 112 | 0.04 | −0.13 (−0.50, 0.25) | 0.02 | 0.02 |
| Malawi | iLiNS−DOSE (31) | KDI | | | | | | | | | | | | |
| Mali | PROMIS CS (32) | DMC | | | | | | | | | | | | |
| Zimbabwe | SHINE (HIV−) (33) | MDAT | 583 | 560 | −0.06 | 0.08 (−0.05, 0.21) | 0.09 | 0.13 | 197 | 180 | −0.02 | 0.18 (−0.01, 0.37) | 0.09 | 0.09 |
| Zimbabwe | SHINE (HIV+) (34) | MDAT | 131 | 110 | −0.24 | 0.43 (0.19, 0.68) | 0.03 | 0.08 | 33 | 32 | 0.13 | −0.19 (−0.77, 0.40) | 0.01 | 0.01 |
| | | | **4517** | **8140** | | **I² = 0.62, Tau² = 0.01** | | | **1643** | **3020** | | **I² = 0.00, Tau² = 0.00** | | |
| **Fixed** | | | | | | **0.06 (0.03, 0.10)** | | | | | | **0.06 (0.00, 0.12)** | | |
| **Random** | | | | | | **0.08 (−0.01, 0.17)** | | | | | | **0.06 (0.00, 0.12)** | | |

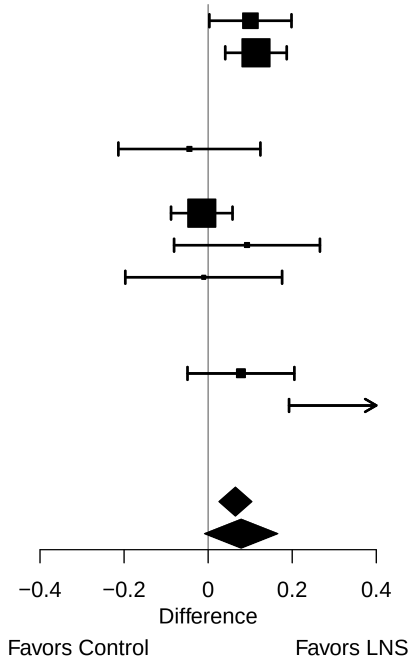

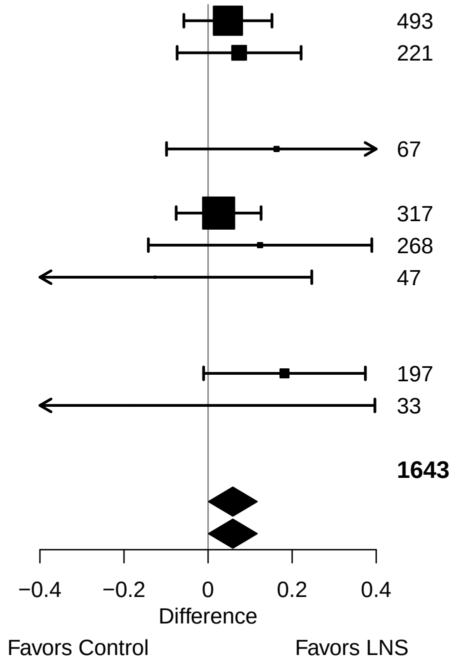

#### Supplemental figure 5J: Mean difference in gross motor z-score

#### 5J6: Stratified by Child sex

**P−for−interaction = 0.155**
**Difference in MDs = −0.04 (−0.09, 0.01)**

| | | | Male | | | | | | Female | | | | | |
|---|---|---|---|---|---|---|---|---|---|---|---|---|---|---|
| **Country** | **Trial** | **Tool** | **LNS N** | **Control N** | **Control Mean** | **MD (95% CI)** | **Fixed W** | **Random W** | **LNS N** | **Control N** | **Control Mean** | **MD (95% CI)** | **Fixed W** | **Random W** |
| Bangladesh | JiVitA−4 (21) | BSID−III | 231 | 70 | 0.19 | −0.24 (−0.44, −0.03) | 0.04 | 0.06 | 214 | 73 | −0.02 | 0.04 (−0.18, 0.25) | 0.03 | 0.07 |
| Bangladesh | RDNS (22) | DMC | 790 | 390 | −0.03 | 0.06 (−0.03, 0.14) | 0.22 | 0.15 | 780 | 368 | −0.04 | 0.11 (0.03, 0.20) | 0.21 | 0.12 |
| Bangladesh | WASH−B (23) | EASQ | 528 | 1646 | −0.02 | 0.08 (−0.02, 0.18) | 0.16 | 0.13 | 546 | 1633 | −0.03 | 0.13 (0.04, 0.23) | 0.18 | 0.12 |
| Burkina Faso | iLiNS−Zinc (24) | DMC | | | | | | | | | | | | |
| Ghana | GHANA (25) | | | | | | | | | | | | | |
| Ghana | iLiNS−DYADG (26) | KDI | 150 | 280 | −0.04 | 0.11 (−0.09, 0.30) | 0.04 | 0.06 | 152 | 321 | 0.03 | −0.08 (−0.28, 0.11) | 0.04 | 0.08 |
| Haiti | HAITI (27) | | | | | | | | | | | | | |
| Kenya | WASH−B (28) | EASQ | 682 | 2260 | −0.01 | 0.03 (−0.05, 0.12) | 0.20 | 0.15 | 680 | 2485 | 0.00 | −0.01 (−0.09, 0.07) | 0.23 | 0.13 |
| Madagascar | MAHAY (29) | ASQI | 795 | 787 | −0.08 | 0.16 (0.00, 0.33) | 0.06 | 0.08 | 818 | 817 | 0.00 | −0.01 (−0.18, 0.16) | 0.06 | 0.09 |
| Malawi | iLiNS−DYADM (30) | KDI | 104 | 204 | −0.05 | 0.17 (−0.07, 0.41) | 0.03 | 0.05 | 110 | 232 | 0.07 | −0.22 (−0.43, −0.01) | 0.03 | 0.07 |
| Malawi | iLiNS−DOSE (31) | KDI | 321 | 117 | −0.01 | 0.02 (−0.19, 0.23) | 0.03 | 0.06 | 325 | 104 | 0.01 | −0.01 (−0.23, 0.20) | 0.03 | 0.07 |
| Mali | PROMIS CS (32) | DMC | 473 | 522 | −0.07 | 0.14 (0.03, 0.25) | 0.13 | 0.13 | 467 | 435 | −0.01 | 0.02 (−0.09, 0.14) | 0.11 | 0.11 |
| Zimbabwe | SHINE (HIV−) (33) | MDAT | 424 | 388 | −0.05 | 0.10 (−0.04, 0.24) | 0.07 | 0.09 | 417 | 411 | −0.04 | 0.10 (−0.07, 0.26) | 0.06 | 0.09 |
| Zimbabwe | SHINE (HIV+) (34) | MDAT | 84 | 79 | −0.11 | 0.22 (−0.06, 0.49) | 0.02 | 0.04 | 86 | 72 | −0.26 | 0.48 (0.16, 0.79) | 0.02 | 0.05 |
| | | | **4582** | **6743** | | **I² = 0.33, Tau² = 0.00** | | | **4595** | **6951** | | **I² = 0.57, Tau² = 0.01** | | |
| **Fixed** | | | | | | **0.07 (0.03, 0.11)** | | | | | | **0.05 (0.01, 0.09)** | | |
| **Random** | | | | | | **0.07 (0.02, 0.13)** | | | | | | **0.04 (−0.04, 0.12)** | | |

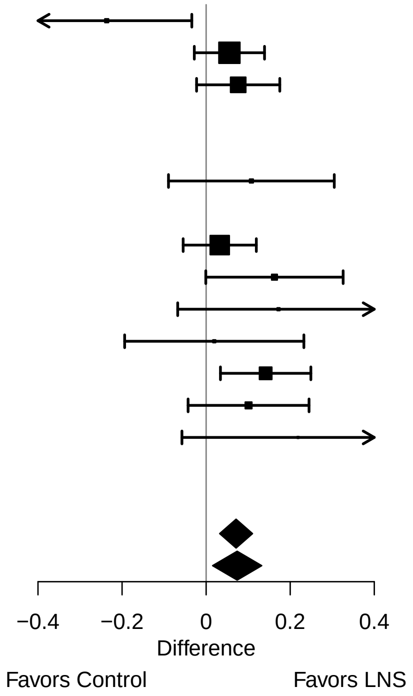

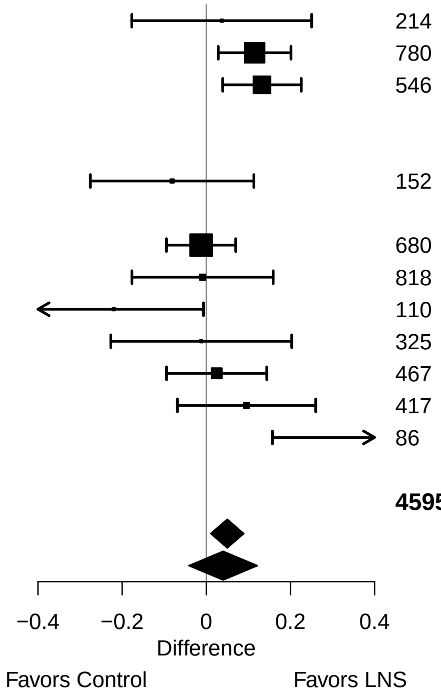

#### Supplemental figure 5J: Mean difference in gross motor z-score

### 5J7: Stratified by Child birth order

**P−for−interaction = 0.145**
**Difference in MDs = −0.05 (−0.12, 0.02)**

| Country | Trial | Tool | Later born LNS N | Later born Control N | Later born Control Mean | Later born MD (95% CI) | Later born Fixed W | Later born Random W | Firstborn LNS N | Firstborn Control N | Firstborn Control Mean | Firstborn MD (95% CI) | Firstborn Fixed W | Firstborn Random W |
|---|---|---|---|---|---|---|---|---|---|---|---|---|---|---|
| Bangladesh | JiVitA−4 (21) | BSID−III | 111 | 31 | −0.15 | 0.03 (−0.36, 0.41) | 0.01 | 0.01 | 333 | 111 | 0.14 | −0.12 (−0.28, 0.04) | 0.11 | 0.12 |
| Bangladesh | RDNS (22) | DMC | 916 | 471 | −0.09 | 0.13 (0.03, 0.23) | 0.13 | 0.14 | 653 | 287 | 0.05 | 0.01 (−0.10, 0.12) | 0.24 | 0.16 |
| Bangladesh | WASH−B (23) | EASQ | 675 | 2149 | −0.03 | 0.11 (0.02, 0.19) | 0.18 | 0.17 | 379 | 1028 | 0.02 | 0.07 (−0.04, 0.18) | 0.22 | 0.15 |
| Burkina Faso | iLiNS−Zinc (24) | DMC | | | | | | | | | | | | |
| Ghana | GHANA (25) | | | | | | | | | | | | | |
| Ghana | iLiNS−DYADG (26) | KDI | 203 | 406 | −0.05 | 0.09 (−0.09, 0.27) | 0.04 | 0.05 | 99 | 195 | 0.10 | −0.16 (−0.37, 0.06) | 0.06 | 0.08 |
| Haiti | HAITI (27) | | | | | | | | | | | | | |
| Kenya | WASH−B (28) | EASQ | 1071 | 3767 | −0.02 | 0.01 (−0.06, 0.08) | 0.26 | 0.22 | 289 | 974 | 0.07 | 0.01 (−0.14, 0.16) | 0.12 | 0.12 |
| Madagascar | MAHAY (29) | ASQI | 1162 | 1118 | −0.08 | 0.07 (−0.08, 0.22) | 0.05 | 0.06 | 433 | 465 | 0.04 | 0.11 (−0.09, 0.30) | 0.07 | 0.09 |
| Malawi | iLiNS−DYADM (30) | KDI | 169 | 349 | 0.01 | −0.04 (−0.23, 0.14) | 0.04 | 0.04 | 45 | 85 | 0.04 | 0.02 (−0.30, 0.34) | 0.03 | 0.05 |
| Malawi | iLiNS−DOSE (31) | KDI | 425 | 151 | −0.06 | 0.07 (−0.12, 0.25) | 0.04 | 0.04 | 134 | 41 | 0.21 | −0.11 (−0.42, 0.21) | 0.03 | 0.05 |
| Mali | PROMIS CS (32) | DMC | 780 | 819 | −0.04 | 0.12 (0.03, 0.21) | 0.16 | 0.16 | 145 | 126 | −0.03 | −0.07 (−0.32, 0.17) | 0.05 | 0.07 |
| Zimbabwe | SHINE (HIV−) (33) | MDAT | 602 | 574 | 0.00 | 0.10 (−0.03, 0.24) | 0.07 | 0.08 | 203 | 191 | −0.15 | 0.08 (−0.13, 0.29) | 0.06 | 0.09 |
| Zimbabwe | SHINE (HIV+) (34) | MDAT | 149 | 118 | −0.14 | 0.29 (0.08, 0.50) | 0.03 | 0.03 | 18 | 29 | −0.30 | 0.73 (0.15, 1.31) | 0.01 | 0.02 |
| | | | **6263** | **9953** | | **I² = 0.18, Tau² = 0.00** | | | **2731** | **3532** | | **I² = 0.30, Tau² = 0.01** | | |
| **Fixed** | | | | | | **0.08 (0.05, 0.12)** | | | | | | **0.01 (−0.04, 0.06)** | | |
| **Random** | | | | | | **0.08 (0.04, 0.12)** | | | | | | **0.01 (−0.07, 0.09)** | | |

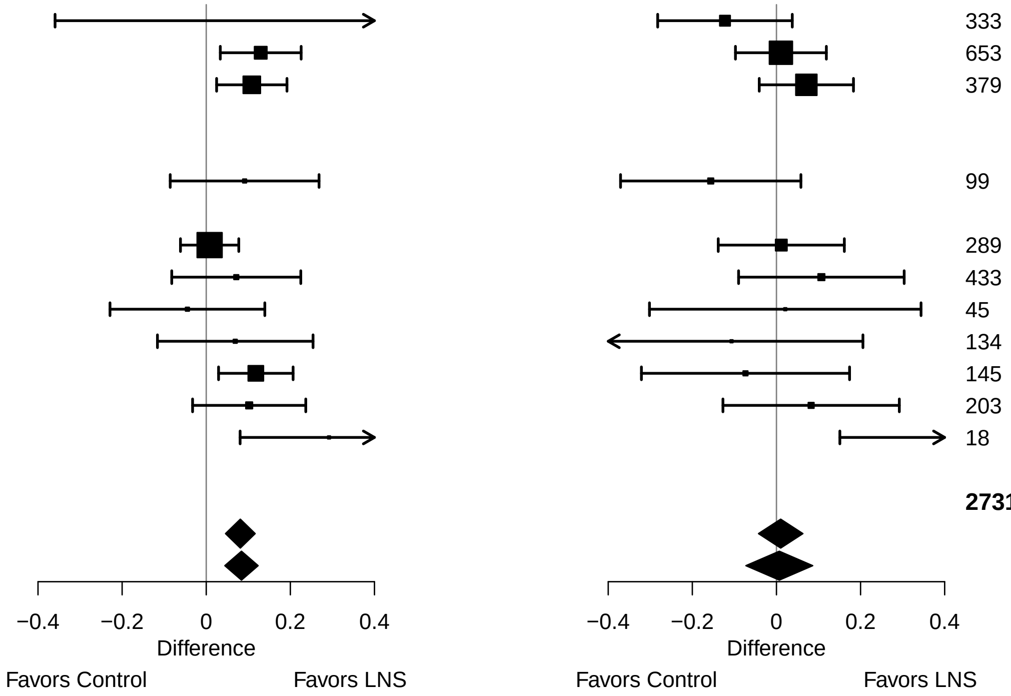

#### Supplemental figure 5J: Mean difference in gross motor z-score

#### 5J8: Stratified by Child baseline stunting

**P−for−interaction = 0.312**
**Difference in MDs = 0.06 (−0.05, 0.17)**

| | | | No | | | | | | Yes | | | | | |
|---|---|---|---|---|---|---|---|---|---|---|---|---|---|---|
| **Country** | **Trial** | **Tool** | **LNS N** | **Control N** | **Control Mean** | **MD (95% CI)** | **Fixed W** | **Random W** | **LNS N** | **Control N** | **Control Mean** | **MD (95% CI)** | **Fixed W** | **Random W** |
| Bangladesh | JiVitA−4 (21) | BSID−III | 327 | 114 | 0.14 | −0.13 (−0.29, 0.04) | 0.07 | 0.13 | 116 | 29 | −0.16 | 0.04 (−0.31, 0.39) | 0.08 | 0.13 |
| Bangladesh | RDNS (22) | DMC | 1180 | 537 | 0.00 | 0.08 (0.02, 0.14) | 0.57 | 0.22 | 331 | 168 | −0.14 | 0.09 (−0.08, 0.26) | 0.37 | 0.21 |
| Bangladesh | WASH−B (23) | EASQ | | | | | | | | | | | | |
| Burkina Faso | iLiNS−Zinc (24) | DMC | | | | | | | | | | | | |
| Ghana | GHANA (25) | | | | | | | | | | | | | |
| Ghana | iLiNS−DYADG (26) | KDI | 259 | 504 | 0.03 | −0.03 (−0.18, 0.11) | 0.09 | 0.15 | 22 | 57 | −0.28 | 0.32 (−0.23, 0.88) | 0.03 | 0.07 |
| Haiti | HAITI (27) | | | | | | | | | | | | | |
| Kenya | WASH−B (28) | EASQ | | | | | | | | | | | | |
| Madagascar | MAHAY (29) | ASQI | | | | | | | | | | | | |
| Malawi | iLiNS−DYADM (30) | KDI | 152 | 320 | 0.09 | −0.06 (−0.23, 0.12) | 0.07 | 0.13 | 48 | 97 | −0.17 | 0.01 (−0.39, 0.42) | 0.06 | 0.11 |
| Malawi | iLiNS−DOSE (31) | KDI | 465 | 147 | 0.13 | −0.04 (−0.20, 0.13) | 0.07 | 0.13 | 181 | 74 | −0.26 | 0.04 (−0.27, 0.35) | 0.11 | 0.14 |
| Mali | PROMIS CS (32) | DMC | | | | | | | | | | | | |
| Zimbabwe | SHINE (HIV−) (33) | MDAT | 532 | 490 | 0.01 | 0.09 (−0.05, 0.24) | 0.10 | 0.15 | 122 | 105 | −0.19 | 0.00 (−0.20, 0.21) | 0.25 | 0.19 |
| Zimbabwe | SHINE (HIV+) (34) | MDAT | 115 | 89 | −0.11 | 0.30 (0.04, 0.55) | 0.03 | 0.08 | 34 | 32 | −0.44 | 0.66 (0.34, 0.99) | 0.10 | 0.14 |
| | | | **3030** | **2201** | | **I² = 0.54, Tau² = 0.01** | | | **854** | **562** | | **I² = 0.54, Tau² = 0.03** | | |
| **Fixed** | | | | | | **0.05 (0.00, 0.09)** | | | | | | **0.12 (0.01, 0.22)** | | |
| **Random** | | | | | | **0.02 (−0.07, 0.11)** | | | | | | **0.15 (−0.03, 0.32)** | | |

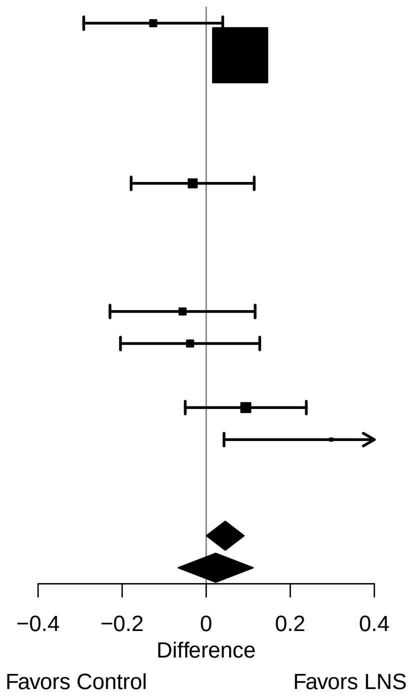

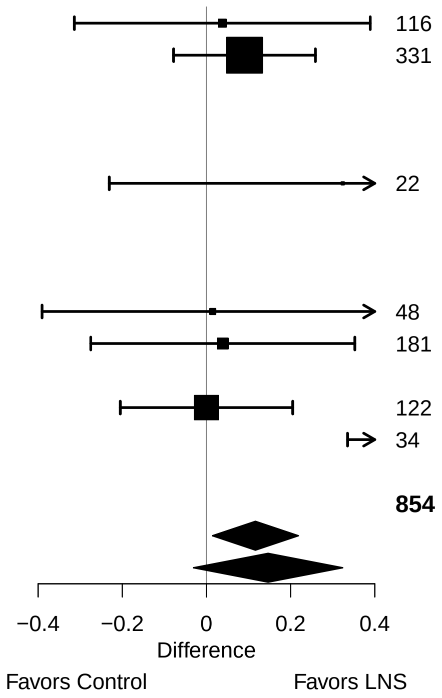

##### Supplemental figure 5J: Mean difference in gross motor z-score

#### 5J9: Stratified by Child baseline acute malnutrition

**P−for−interaction** **= 0.417**
**Difference in MDs =** **0.08 (−0.11, 0.26)**

| | | | No | | | | | | Yes | | | | | |
|---|---|---|---|---|---|---|---|---|---|---|---|---|---|---|
| **Country** | **Trial** | **Tool** | **LNS N** | **Control N** | **Control Mean** | **MD (95% CI)** | **Fixed W** | **Random W** | **LNS N** | **Control N** | **Control Mean** | **MD (95% CI)** | **Fixed W** | **Random W** |
| Bangladesh | JiVitA−4 (21) | BSID−III | 359 | 112 | 0.15 | −0.14 (−0.34, 0.05) | 0.05 | 0.12 | 84 | 31 | −0.16 | 0.04 (−0.31, 0.40) | 0.24 | 0.24 |
| Bangladesh | RDNS (22) | DMC | 1396 | 657 | −0.02 | 0.09 (0.03, 0.15) | 0.54 | 0.18 | 115 | 49 | −0.23 | 0.05 (−0.34, 0.44) | 0.21 | 0.21 |
| Bangladesh | WASH−B (23) | EASQ | | | | | | | | | | | | |
| Burkina Faso | iLiNS−Zinc (24) | DMC | | | | | | | | | | | | |
| Ghana | GHANA (25) | | | | | | | | | | | | | |
| Ghana | iLiNS−DYADG (26) | KDI | 265 | 509 | 0.02 | 0.00 (−0.15, 0.15) | 0.09 | 0.14 | 17 | 52 | −0.17 | −0.08 (−0.50, 0.34) | 0.18 | 0.18 |
| Haiti | HAITI (27) | | | | | | | | | | | | | |
| Kenya | WASH−B (28) | EASQ | | | | | | | | | | | | |
| Madagascar | MAHAY (29) | ASQI | | | | | | | | | | | | |
| Malawi | iLiNS−DYADM (30) | KDI | 186 | 383 | 0.03 | −0.03 (−0.19, 0.14) | 0.07 | 0.14 | 14 | 37 | −0.21 | −0.01 (−0.86, 0.83) | 0.04 | 0.04 |
| Malawi | iLiNS−DOSE (31) | KDI | 609 | 206 | 0.03 | 0.01 (−0.14, 0.16) | 0.08 | 0.14 | 37 | 15 | −0.40 | −0.14 (−0.93, 0.65) | 0.05 | 0.05 |
| Mali | PROMIS CS (32) | DMC | | | | | | | | | | | | |
| Zimbabwe | SHINE (HIV−) (33) | MDAT | 620 | 566 | −0.02 | 0.07 (−0.06, 0.19) | 0.12 | 0.16 | 36 | 34 | −0.11 | 0.34 (−0.07, 0.75) | 0.19 | 0.19 |
| Zimbabwe | SHINE (HIV+) (34) | MDAT | 137 | 114 | −0.17 | 0.38 (0.17, 0.60) | 0.04 | 0.11 | 14 | 8 | −0.54 | 0.55 (−0.03, 1.13) | 0.09 | 0.09 |
| | | | **3572** | **2547** | | **I² = 0.64, Tau² = 0.02** | | | **317** | **226** | | **I² = 0.00, Tau² = 0.00** | | |
| **Fixed** | | | | | | **0.07 (0.02, 0.11)** | | | | | | **0.11 (−0.06, 0.29)** | | |
| **Random** | | | | | | **0.05 (−0.06, 0.16)** | | | | | | **0.11 (−0.06, 0.29)** | | |

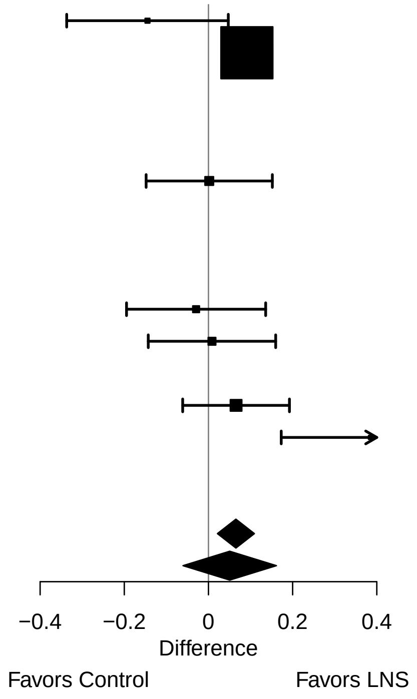

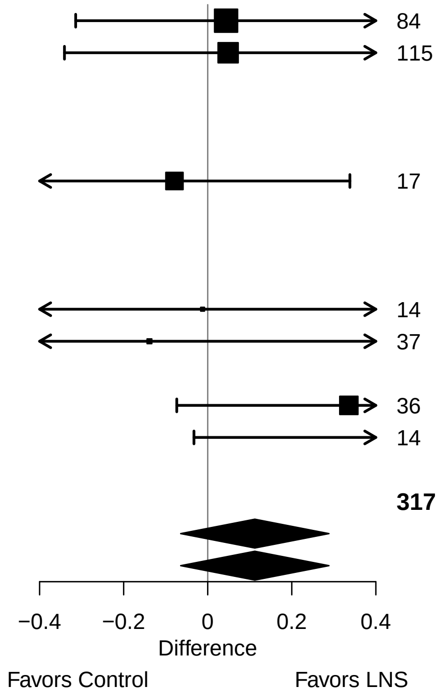

##### Supplemental figure 5J: Mean difference in gross motor z-score

#### 5J10: Stratified by Child baseline anemia

**P−for−interaction = 0.658**
**Difference in MDs = 0.03 (−0.12, 0.19)**

| Country | Trial | Tool | Not anemic: LNS N | Control N | Control Mean | MD (95% CI) | Fixed W | Random W | Anemic: LNS N | Control N | Control Mean | MD (95% CI) | Fixed W | Random W |
|---|---|---|---|---|---|---|---|---|---|---|---|---|---|---|
| Bangladesh | JiVitA−4 (21) | BSID−III | | | | | | | | | | | | |
| Bangladesh | RDNS (22) | DMC | 202 | 91 | −0.16 | 0.19 (−0.03, 0.40) | 0.27 | 0.27 | 304 | 151 | −0.08 | 0.14 (0.02, 0.27) | 0.50 | 0.46 |
| Bangladesh | WASH−B (23) | EASQ | | | | | | | | | | | | |
| Burkina Faso | iLiNS−Zinc (24) | DMC | | | | | | | | | | | | |
| Ghana | GHANA (25) | | | | | | | | | | | | | |
| Ghana | iLiNS−DYADG (26) | KDI | 180 | 321 | 0.03 | −0.04 (−0.23, 0.14) | 0.34 | 0.33 | 81 | 189 | −0.09 | 0.12 (−0.14, 0.38) | 0.12 | 0.13 |
| Haiti | HAITI (27) | | | | | | | | | | | | | |
| Kenya | WASH−B (28) | EASQ | | | | | | | | | | | | |
| Madagascar | MAHAY (29) | ASQI | | | | | | | | | | | | |
| Malawi | iLiNS−DYADM (30) | KDI | 65 | 146 | −0.01 | 0.01 (−0.28, 0.29) | 0.15 | 0.15 | 138 | 273 | 0.04 | −0.07 (−0.27, 0.12) | 0.20 | 0.21 |
| Malawi | iLiNS−DOSE (31) | KDI | 224 | 94 | 0.13 | −0.05 (−0.27, 0.17) | 0.25 | 0.25 | 418 | 126 | −0.10 | 0.06 (−0.15, 0.26) | 0.19 | 0.20 |
| Mali | PROMIS CS (32) | DMC | | | | | | | | | | | | |
| Zimbabwe | SHINE (HIV−) (33) | MDAT | | | | | | | | | | | | |
| Zimbabwe | SHINE (HIV+) (34) | MDAT | | | | | | | | | | | | |
| | | | **671** | **652** | | **I² = 0.08, Tau² = 0.00** | | | **941** | **739** | | **I² = 0.11, Tau² = 0.00** | | |
| **Fixed** | | | | | | **0.02 (−0.09, 0.13)** | | | | | | **0.08 (−0.01, 0.17)** | | |
| **Random** | | | | | | **0.02 (−0.09, 0.14)** | | | | | | **0.08 (−0.02, 0.17)** | | |

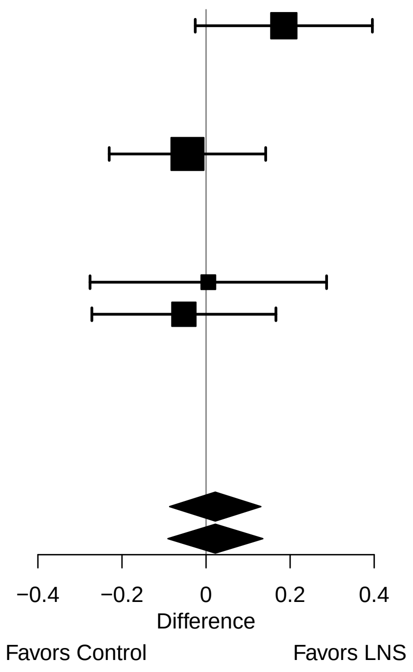

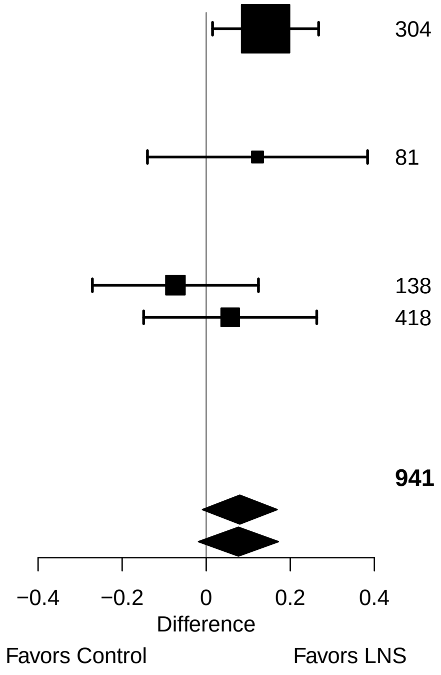

### Supplemental figure 5K: Mean difference in fine motor z-score

#### 5K1: Stratified by Maternal height

**P−for−interaction = 0.069**
**Difference in MDs = −0.10 (−0.20, 0.01)**

| | | | At least 150.1 cm | | | | | | Less than 150.1 cm | | | | | |
|---|---|---|---|---|---|---|---|---|---|---|---|---|---|---|
| Country | Trial | Tool | LNS N | Control N | Control Mean | MD (95% CI) | Fixed W | Random W | LNS N | Control N | Control Mean | MD (95% CI) | Fixed W | Random W |
| Bangladesh | JiVitA−4 (21) | BSID−III | | | | | | | | | | | | |
| Bangladesh | RDNS (22) | DMC | 857 | 430 | −0.05 | 0.16 (0.03, 0.30) | 0.16 | 0.16 | 725 | 350 | −0.06 | 0.05 (−0.07, 0.16) | 0.65 | 0.65 |
| Bangladesh | WASH−B (23) | EASQ | | | | | | | | | | | | |
| Burkina Faso | iLiNS−Zinc (24) | DMC | | | | | | | | | | | | |
| Ghana | GHANA (25) | | | | | | | | | | | | | |
| Ghana | iLiNS−DYADG (26) | KDI | 284 | 557 | 0.03 | 0.03 (−0.10, 0.16) | 0.19 | 0.19 | 13 | 34 | −0.44 | 0.09 (−0.93, 1.11) | 0.01 | 0.01 |
| Haiti | HAITI (27) | | | | | | | | | | | | | |
| Kenya | WASH−B (28) | EASQ | | | | | | | | | | | | |
| Madagascar | MAHAY (29) | ASQI | 956 | 986 | −0.07 | 0.08 (−0.16, 0.32) | 0.05 | 0.05 | 553 | 538 | 0.10 | −0.03 (−0.26, 0.19) | 0.18 | 0.18 |
| Malawi | iLiNS−DYADM (30) | KDI | 186 | 376 | −0.03 | 0.06 (−0.11, 0.23) | 0.10 | 0.10 | 28 | 57 | 0.08 | 0.21 (−0.25, 0.67) | 0.04 | 0.04 |
| Malawi | iLiNS−DOSE (31) | KDI | 543 | 185 | −0.05 | 0.09 (−0.08, 0.25) | 0.11 | 0.11 | 102 | 35 | −0.13 | 0.09 (−0.31, 0.49) | 0.05 | 0.05 |
| Mali | PROMIS CS (32) | DMC | 868 | 892 | −0.06 | 0.12 (−0.09, 0.33) | 0.06 | 0.06 | 32 | 26 | 0.12 | −0.25 (−0.82, 0.33) | 0.03 | 0.03 |
| Zimbabwe | SHINE (HIV−) (33) | MDAT | 788 | 762 | −0.04 | 0.09 (−0.01, 0.19) | 0.27 | 0.27 | 34 | 22 | −0.10 | −0.03 (−0.53, 0.47) | 0.03 | 0.03 |
| Zimbabwe | SHINE (HIV+) (34) | MDAT | 158 | 140 | −0.06 | 0.13 (−0.10, 0.36) | 0.06 | 0.06 | 10 | 6 | −0.49 | 0.71 (−0.66, 2.09) | 0.00 | 0.00 |
| | | | **4640** | **4328** | | **I² = 0.00, Tau² = 0.00** | | | **1497** | **1068** | | **I² = 0.00, Tau² = 0.00** | | |
| **Fixed** | | | | | | **0.09 (0.04, 0.14)** | | | | | | **0.04 (−0.06, 0.13)** | | |
| **Random** | | | | | | **0.09 (0.04, 0.14)** | | | | | | **0.04 (−0.06, 0.13)** | | |

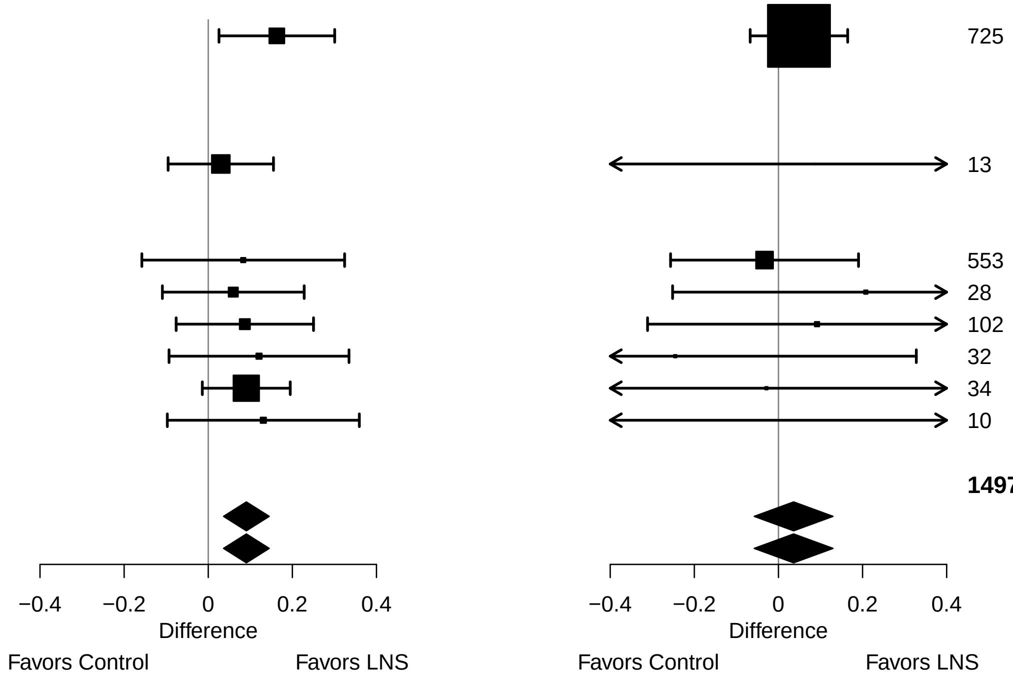

#### Supplemental figure 5K: Mean difference in fine motor z-score

##### 5K2: Stratified by Maternal BMI

**P−for−interaction = 0.767**
**Difference in MDs = −0.02 (−0.12, 0.09)**

| Country | Trial | Tool | At least 20 kg/m²: LNS N | Control N | Control Mean | MD (95% CI) | Fixed W | Random W | Less than 20 kg/m²: LNS N | Control N | Control Mean | MD (95% CI) | Fixed W | Random W |
|---|---|---|---|---|---|---|---|---|---|---|---|---|---|---|
| Bangladesh | JiVitA−4 (21) | BSID−III | | | | | | | | | | | | |
| Bangladesh | RDNS (22) | DMC | 722 | 330 | 0.01 | 0.09 (−0.05, 0.23) | 0.18 | 0.18 | 860 | 450 | −0.11 | 0.12 (0.00, 0.25) | 0.52 | 0.52 |
| Bangladesh | WASH−B (23) | EASQ | | | | | | | | | | | | |
| Burkina Faso | iLiNS−Zinc (24) | DMC | | | | | | | | | | | | |
| Ghana | GHANA (25) | | | | | | | | | | | | | |
| Ghana | iLiNS−DYADG (26) | KDI | 262 | 485 | 0.01 | 0.04 (−0.09, 0.18) | 0.19 | 0.19 | 35 | 106 | −0.04 | −0.02 (−0.40, 0.35) | 0.06 | 0.06 |
| Haiti | HAITI (27) | | | | | | | | | | | | | |
| Kenya | WASH−B (28) | EASQ | | | | | | | | | | | | |
| Madagascar | MAHAY (29) | ASQI | | | | | | | | | | | | |
| Malawi | iLiNS−DYADM (30) | KDI | 126 | 258 | −0.01 | 0.10 (−0.11, 0.31) | 0.08 | 0.08 | 87 | 175 | −0.03 | 0.05 (−0.20, 0.29) | 0.13 | 0.13 |
| Malawi | iLiNS−DOSE (31) | KDI | 480 | 158 | 0.00 | 0.03 (−0.15, 0.20) | 0.12 | 0.12 | 163 | 61 | −0.22 | 0.25 (−0.06, 0.55) | 0.09 | 0.09 |
| Mali | PROMIS CS (32) | DMC | 668 | 655 | −0.05 | 0.15 (−0.07, 0.37) | 0.08 | 0.08 | 232 | 263 | −0.06 | −0.01 (−0.29, 0.28) | 0.10 | 0.10 |
| Zimbabwe | SHINE (HIV−) (33) | MDAT | 601 | 581 | −0.04 | 0.11 (0.00, 0.22) | 0.28 | 0.28 | 100 | 96 | −0.17 | 0.19 (−0.11, 0.49) | 0.09 | 0.09 |
| Zimbabwe | SHINE (HIV+) (34) | MDAT | 124 | 113 | −0.06 | 0.24 (0.00, 0.48) | 0.06 | 0.06 | 25 | 23 | −0.08 | −0.43 (−1.04, 0.17) | 0.02 | 0.02 |
| | | | **2983** | **2580** | | **I² = 0.00, Tau² = 0.00** | | | **1502** | **1174** | | **I² = 0.00, Tau² = 0.00** | | |
| **Fixed** | | | | | | **0.09 (0.03, 0.15)** | | | | | | **0.10 (0.01, 0.19)** | | |
| **Random** | | | | | | **0.09 (0.03, 0.15)** | | | | | | **0.10 (0.01, 0.19)** | | |

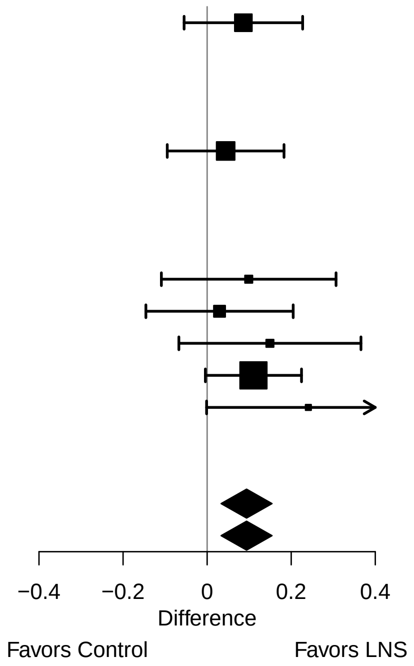

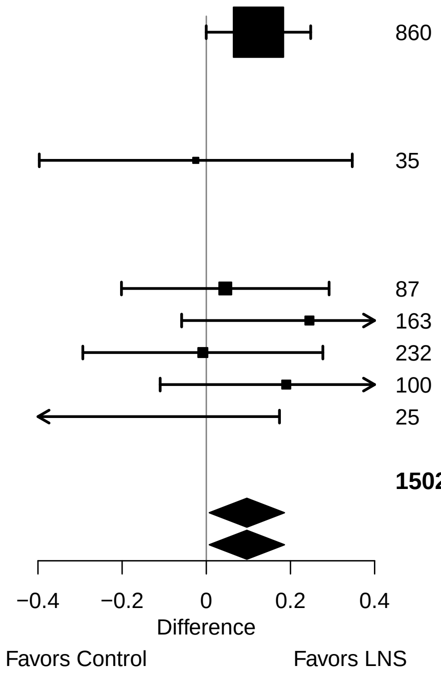

#### Supplemental figure 5K: Mean difference in fine motor z-score

##### 5K3: Stratified by Maternal age

**P−for−interaction = 0.005**
**Difference in MDs = −0.10 (−0.17, −0.03)**

| Country | Trial | Tool | At least 25 y: LNS N | Control N | Control Mean | MD (95% CI) | Fixed W | Random W | Less than 25 y: LNS N | Control N | Control Mean | MD (95% CI) | Fixed W | Random W |
|---|---|---|---|---|---|---|---|---|---|---|---|---|---|---|
| Bangladesh | JiVitA−4 (21) | BSID−III | 200 | 68 | 0.00 | −0.01 (−0.30, 0.28) | 0.05 | 0.05 | 242 | 74 | −0.19 | 0.25 (−0.03, 0.52) | 0.05 | 0.05 |
| Bangladesh | RDNS (22) | DMC | 451 | 218 | −0.06 | 0.17 (−0.01, 0.36) | 0.12 | 0.12 | 1190 | 586 | −0.05 | 0.07 (−0.03, 0.17) | 0.40 | 0.40 |
| Bangladesh | WASH−B (23) | EASQ | | | | | | | | | | | | |
| Burkina Faso | iLiNS−Zinc (24) | DMC | | | | | | | | | | | | |
| Ghana | GHANA (25) | | | | | | | | | | | | | |
| Ghana | iLiNS−DYADG (26) | KDI | 192 | 378 | 0.00 | 0.10 (−0.06, 0.25) | 0.17 | 0.17 | 110 | 223 | 0.00 | −0.06 (−0.28, 0.16) | 0.08 | 0.08 |
| Haiti | HAITI (27) | | | | | | | | | | | | | |
| Kenya | WASH−B (28) | EASQ | | | | | | | | | | | | |
| Madagascar | MAHAY (29) | ASQI | 852 | 901 | −0.07 | 0.05 (−0.17, 0.27) | 0.08 | 0.08 | 759 | 703 | 0.04 | 0.04 (−0.20, 0.27) | 0.07 | 0.07 |
| Malawi | iLiNS−DYADM (30) | KDI | 102 | 224 | −0.06 | 0.29 (0.07, 0.50) | 0.09 | 0.09 | 112 | 212 | 0.01 | −0.11 (−0.35, 0.12) | 0.07 | 0.07 |
| Malawi | iLiNS−DOSE (31) | KDI | 346 | 124 | −0.08 | 0.11 (−0.10, 0.31) | 0.10 | 0.10 | 288 | 90 | −0.02 | 0.05 (−0.18, 0.28) | 0.07 | 0.07 |
| Mali | PROMIS CS (32) | DMC | 482 | 480 | −0.04 | 0.17 (−0.04, 0.38) | 0.09 | 0.09 | 423 | 444 | −0.06 | 0.03 (−0.22, 0.28) | 0.06 | 0.06 |
| Zimbabwe | SHINE (HIV−) (33) | MDAT | 427 | 389 | −0.03 | 0.15 (0.01, 0.28) | 0.22 | 0.22 | 326 | 321 | −0.06 | 0.00 (−0.15, 0.14) | 0.19 | 0.19 |
| Zimbabwe | SHINE (HIV+) (34) | MDAT | 126 | 111 | −0.05 | 0.15 (−0.09, 0.39) | 0.07 | 0.07 | 29 | 26 | −0.05 | 0.00 (−0.65, 0.65) | 0.01 | 0.01 |
| | | | **3178** | **2893** | | **I² = 0.00, Tau² = 0.00** | | | **3479** | **2679** | | **I² = 0.00, Tau² = 0.00** | | |
| **Fixed** | | | | | | **0.14 (0.07, 0.20)** | | | | | | **0.03 (−0.03, 0.10)** | | |
| **Random** | | | | | | **0.14 (0.07, 0.20)** | | | | | | **0.03 (−0.03, 0.10)** | | |

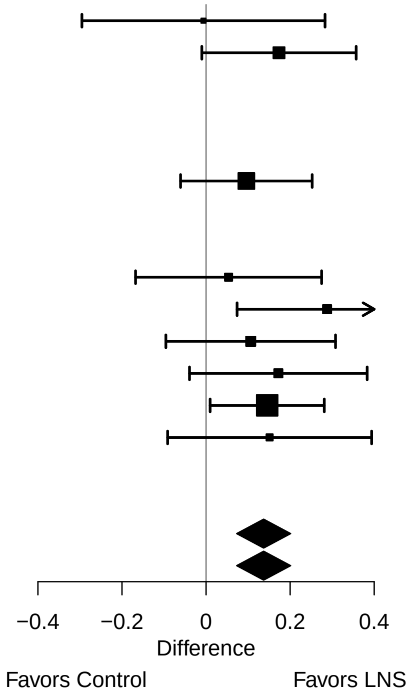

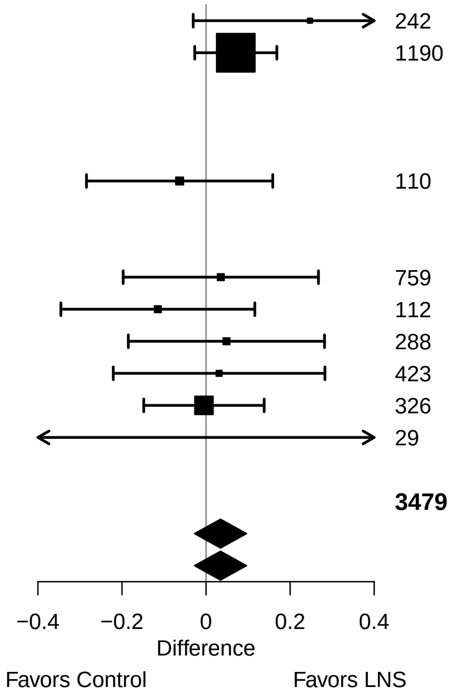

#### Supplemental figure 5K: Mean difference in fine motor z-score

#### 5K4: Stratified by Maternal education

**P−for−interaction = 0.003**
**Difference in MDs = 0.15 (0.05, 0.25)**

| | | | Primary or greater | | | | | | | Incomplete or no formal | | | | | |
|---|---|---|---|---|---|---|---|---|---|---|---|---|---|---|---|
| Country | Trial | Tool | LNS N | Control N | Control Mean | MD (95% CI) | Fixed W | Random W | | LNS N | Control N | Control Mean | MD (95% CI) | Fixed W | Random W |
| Bangladesh | JiVitA−4 (21) | BSID−III | 292 | 92 | 0.05 | 0.04 (−0.17, 0.24) | 0.07 | 0.07 | | 152 | 51 | −0.39 | 0.34 (0.03, 0.64) | 0.06 | 0.09 |
| Bangladesh | RDNS (22) | DMC | 1227 | 582 | 0.03 | 0.06 (−0.03, 0.16) | 0.31 | 0.31 | | 414 | 222 | −0.26 | 0.17 (0.00, 0.35) | 0.19 | 0.16 |
| Bangladesh | WASH−B (23) | EASQ | | | | | | | | | | | | | |
| Burkina Faso | iLiNS−Zinc (24) | DMC | | | | | | | | | | | | | |
| Ghana | GHANA (25) | | | | | | | | | | | | | | |
| Ghana | iLiNS−DYADG (26) | KDI | 235 | 467 | 0.03 | −0.03 (−0.18, 0.11) | 0.14 | 0.14 | | 67 | 134 | −0.12 | 0.29 (0.02, 0.56) | 0.08 | 0.11 |
| Haiti | HAITI (27) | | | | | | | | | | | | | | |
| Kenya | WASH−B (28) | EASQ | | | | | | | | | | | | | |
| Madagascar | MAHAY (29) | ASQI | 339 | 416 | 0.27 | −0.14 (−0.36, 0.08) | 0.06 | 0.06 | | 1274 | 1188 | −0.13 | 0.12 (−0.10, 0.35) | 0.12 | 0.13 |
| Malawi | iLiNS−DYADM (30) | KDI | 33 | 67 | 0.04 | −0.12 (−0.49, 0.25) | 0.02 | 0.02 | | 180 | 366 | −0.03 | 0.11 (−0.06, 0.29) | 0.20 | 0.17 |
| Malawi | iLiNS−DOSE (31) | KDI | 148 | 52 | 0.19 | 0.05 (−0.24, 0.35) | 0.03 | 0.03 | | 487 | 162 | −0.13 | 0.10 (−0.08, 0.27) | 0.19 | 0.16 |
| Mali | PROMIS CS (32) | DMC | 100 | 93 | 0.00 | 0.15 (−0.26, 0.55) | 0.02 | 0.02 | | 804 | 831 | −0.06 | 0.10 (−0.11, 0.31) | 0.13 | 0.14 |
| Zimbabwe | SHINE (HIV−) (33) | MDAT | 763 | 731 | −0.04 | 0.10 (−0.01, 0.20) | 0.28 | 0.28 | | 29 | 25 | 0.13 | −0.49 (−1.04, 0.06) | 0.02 | 0.04 |
| Zimbabwe | SHINE (HIV+) (34) | MDAT | 149 | 130 | −0.06 | 0.18 (−0.03, 0.40) | 0.06 | 0.06 | | 9 | 8 | 0.04 | −0.83 (−2.38, 0.73) | 0.00 | 0.00 |
| | | | **3286** | **2630** | | **$I^2$ = 0.00, $Tau^2$ = 0.00** | | | | **3416** | **2987** | | **$I^2$ = 0.19, $Tau^2$ = 0.01** | | |
| **Fixed** | | | | | | **0.05 (−0.01, 0.10)** | | | | | | | **0.14 (0.06, 0.21)** | | |
| **Random** | | | | | | **0.05 (−0.01, 0.10)** | | | | | | | **0.13 (0.02, 0.24)** | | |

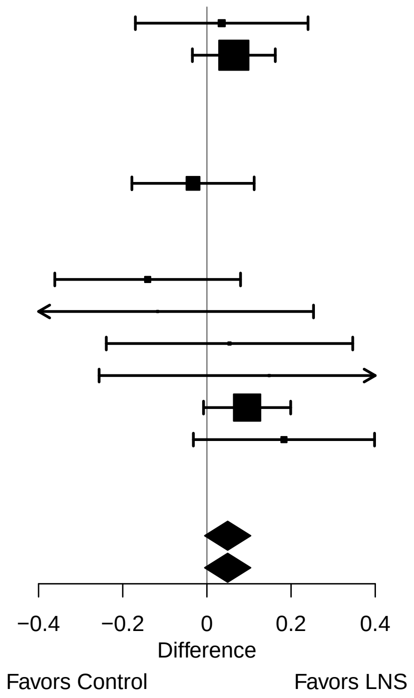

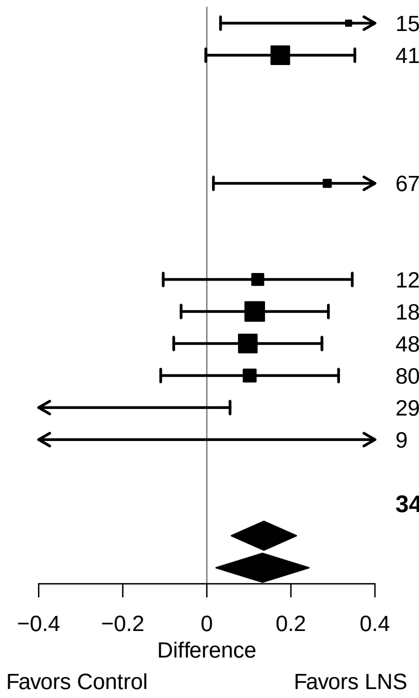

**Supplemental figure 5K: Mean difference in fine motor z-score**

#### 5K5: Stratified by Maternal depressive symptoms

**P−for−interaction = 0.647**
**Difference in MDs = −0.02 (−0.11, 0.07)**

| Country | Trial | Tool | LNS N | Control N | Control Mean | Less than 75th percentile MD (95% CI) | Fixed W | Random W | LNS N | Control N | Control Mean | At least 75th percentile MD (95% CI) | Fixed W | Random W |
|---|---|---|---|---|---|---|---|---|---|---|---|---|---|---|
| Bangladesh | JiVitA−4 (21) | BSID−III | | | | | | | | | | | | |
| Bangladesh | RDNS (22) | DMC | 1067 | 469 | −0.03 | 0.11 (−0.02, 0.23) | 0.27 | 0.27 | 512 | 281 | −0.13 | 0.12 (0.00, 0.24) | 0.53 | 0.47 |
| Bangladesh | WASH−B (23) | EASQ | | | | | | | | | | | | |
| Burkina Faso | iLiNS−Zinc (24) | DMC | | | | | | | | | | | | |
| Ghana | GHANA (25) | | | | | | | | | | | | | |
| Ghana | iLiNS−DYADG (26) | KDI | 226 | 393 | 0.00 | −0.02 (−0.17, 0.13) | 0.19 | 0.19 | 67 | 190 | 0.00 | 0.32 (0.06, 0.57) | 0.11 | 0.13 |
| Haiti | HAITI (27) | | | | | | | | | | | | | |
| Kenya | WASH−B (28) | EASQ | | | | | | | | | | | | |
| Madagascar | MAHAY (29) | ASQI | 615 | 667 | −0.02 | 0.09 (−0.19, 0.37) | 0.05 | 0.05 | 268 | 242 | −0.08 | 0.06 (−0.28, 0.39) | 0.07 | 0.08 |
| Malawi | iLiNS−DYADM (30) | KDI | 152 | 299 | −0.07 | 0.15 (−0.04, 0.35) | 0.12 | 0.12 | 47 | 112 | 0.08 | −0.14 (−0.46, 0.18) | 0.07 | 0.08 |
| Malawi | iLiNS−DOSE (31) | KDI | | | | | | | | | | | | |
| Mali | PROMIS CS (32) | DMC | | | | | | | | | | | | |
| Zimbabwe | SHINE (HIV−) (33) | MDAT | 583 | 560 | 0.00 | 0.06 (−0.06, 0.18) | 0.31 | 0.31 | 197 | 180 | −0.17 | 0.12 (−0.08, 0.31) | 0.19 | 0.21 |
| Zimbabwe | SHINE (HIV+) (34) | MDAT | 131 | 110 | −0.07 | 0.18 (−0.07, 0.44) | 0.06 | 0.06 | 33 | 32 | −0.03 | −0.03 (−0.54, 0.47) | 0.03 | 0.03 |
| | | | **2774** | **2498** | | **I² = 0.00, Tau² = 0.00** | | | **1124** | **1037** | | **I² = 0.05, Tau² = 0.00** | | |
| **Fixed** | | | | | | **0.08 (0.01, 0.14)** | | | | | | **0.11 (0.03, 0.20)** | | |
| **Random** | | | | | | **0.08 (0.01, 0.14)** | | | | | | **0.11 (0.02, 0.21)** | | |

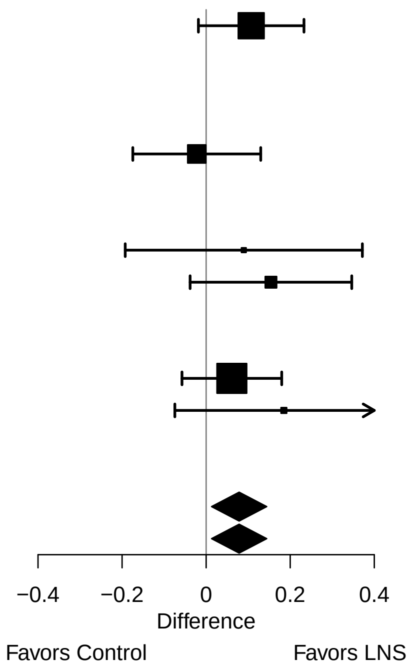

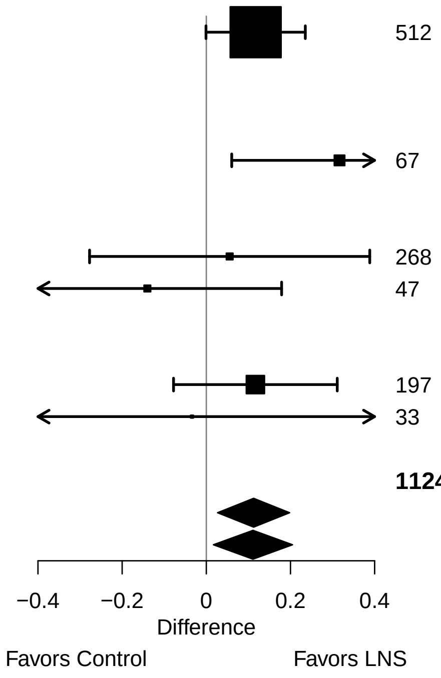

#### Supplemental figure 5K: Mean difference in fine motor z-score

#### 5K6: Stratified by Child sex

**P−for−interaction = 0.162**
**Difference in MDs = 0.05 (−0.02, 0.11)**

| Country | Trial | Tool | Male: LNS N | Male: Control N | Male: Control Mean | Male: MD (95% CI) | Male: Fixed W | Male: Random W | Female: LNS N | Female: Control N | Female: Control Mean | Female: MD (95% CI) | Female: Fixed W | Female: Random W |
|---|---|---|---|---|---|---|---|---|---|---|---|---|---|---|
| Bangladesh | JiVitA−4 (21) | BSID−III | 231 | 70 | −0.01 | 0.02 (−0.26, 0.30) | 0.04 | 0.04 | 214 | 73 | −0.20 | 0.27 (0.07, 0.47) | 0.10 | 0.11 |
| Bangladesh | RDNS (22) | DMC | 819 | 404 | 0.00 | 0.01 (−0.10, 0.11) | 0.33 | 0.33 | 822 | 400 | −0.10 | 0.19 (0.05, 0.33) | 0.21 | 0.17 |
| Bangladesh | WASH−B (23) | EASQ | | | | | | | | | | | | |
| Burkina Faso | iLiNS−Zinc (24) | DMC | | | | | | | | | | | | |
| Ghana | GHANA (25) | | | | | | | | | | | | | |
| Ghana | iLiNS−DYADG (26) | KDI | 150 | 280 | −0.04 | 0.14 (−0.03, 0.31) | 0.12 | 0.12 | 152 | 321 | 0.03 | −0.05 (−0.24, 0.14) | 0.11 | 0.12 |
| Haiti | HAITI (27) | | | | | | | | | | | | | |
| Kenya | WASH−B (28) | EASQ | | | | | | | | | | | | |
| Madagascar | MAHAY (29) | ASQI | 795 | 787 | −0.03 | 0.06 (−0.15, 0.27) | 0.08 | 0.08 | 818 | 817 | −0.02 | 0.03 (−0.21, 0.27) | 0.07 | 0.08 |
| Malawi | iLiNS−DYADM (30) | KDI | 104 | 204 | −0.06 | 0.21 (−0.02, 0.45) | 0.06 | 0.06 | 110 | 232 | 0.01 | −0.05 (−0.26, 0.17) | 0.09 | 0.10 |
| Malawi | iLiNS−DOSE (31) | KDI | 321 | 117 | 0.00 | 0.00 (−0.22, 0.22) | 0.07 | 0.07 | 325 | 104 | −0.14 | 0.19 (−0.02, 0.40) | 0.10 | 0.10 |
| Mali | PROMIS CS (32) | DMC | 457 | 501 | −0.03 | 0.06 (−0.17, 0.29) | 0.07 | 0.07 | 448 | 423 | −0.08 | 0.16 (−0.07, 0.39) | 0.08 | 0.09 |
| Zimbabwe | SHINE (HIV−) (33) | MDAT | 424 | 388 | −0.06 | 0.12 (−0.02, 0.25) | 0.19 | 0.19 | 417 | 411 | −0.01 | 0.03 (−0.12, 0.18) | 0.18 | 0.16 |
| Zimbabwe | SHINE (HIV+) (34) | MDAT | 84 | 79 | −0.07 | 0.15 (−0.19, 0.48) | 0.03 | 0.03 | 86 | 72 | −0.09 | 0.19 (−0.09, 0.47) | 0.05 | 0.06 |
| | | | **3385** | **2830** | | **$I^2$ = 0.00, $Tau^2$ = 0.00** | | | **3392** | **2853** | | **$I^2$ = 0.27, $Tau^2$ = 0.00** | | |
| **Fixed** | | | | | | **0.07 (0.01, 0.13)** | | | | | | **0.11 (0.04, 0.17)** | | |
| **Random** | | | | | | **0.07 (0.01, 0.13)** | | | | | | **0.11 (0.03, 0.18)** | | |

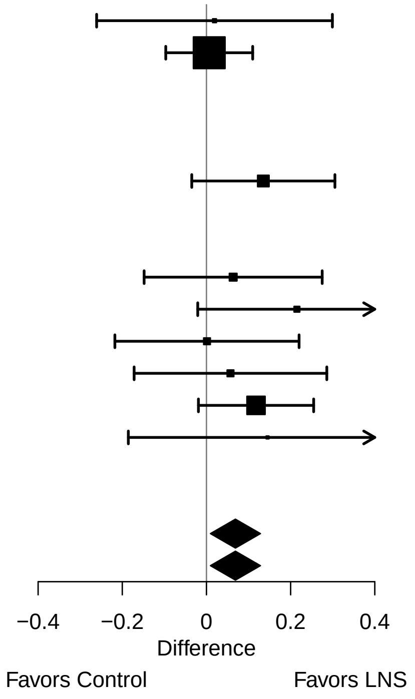

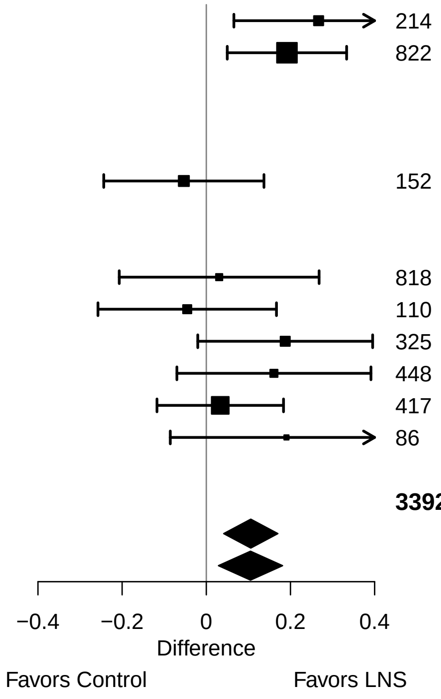

#### Supplemental figure 5K: Mean difference in fine motor z-score

### 5K7: Stratified by Child birth order

**P−for−interaction = 0.062**

**Difference in MDs = −0.08 (−0.17, 0.00)**

| Country | Trial | Tool | LNS N | Control N | Control Mean | Later born MD (95% CI) | Fixed W | Random W | LNS N | Control N | Control Mean | Firstborn MD (95% CI) | Fixed W | Random W |
|---|---|---|---|---|---|---|---|---|---|---|---|---|---|---|
| Bangladesh | JiVitA−4 (21) | BSID−III | 111 | 31 | −0.35 | 0.23 (−0.18, 0.64) | 0.02 | 0.02 | 333 | 111 | −0.05 | 0.14 (−0.07, 0.34) | 0.16 | 0.16 |
| Bangladesh | RDNS (22) | DMC | 961 | 505 | −0.08 | 0.17 (0.05, 0.28) | 0.27 | 0.27 | 679 | 299 | 0.00 | 0.00 (−0.17, 0.16) | 0.24 | 0.24 |
| Bangladesh | WASH−B (23) | EASQ | | | | | | | | | | | | |
| Burkina Faso | iLiNS−Zinc (24) | DMC | | | | | | | | | | | | |
| Ghana | GHANA (25) | | | | | | | | | | | | | |
| Ghana | iLiNS−DYADG (26) | KDI | 203 | 406 | −0.01 | 0.02 (−0.14, 0.18) | 0.13 | 0.13 | 99 | 195 | 0.01 | 0.07 (−0.15, 0.28) | 0.15 | 0.15 |
| Haiti | HAITI (27) | | | | | | | | | | | | | |
| Kenya | WASH−B (28) | EASQ | | | | | | | | | | | | |
| Madagascar | MAHAY (29) | ASQI | 1162 | 1118 | −0.07 | 0.07 (−0.16, 0.29) | 0.07 | 0.07 | 433 | 465 | 0.07 | 0.03 (−0.21, 0.27) | 0.11 | 0.11 |
| Malawi | iLiNS−DYADM (30) | KDI | 169 | 349 | −0.04 | 0.09 (−0.09, 0.27) | 0.10 | 0.10 | 45 | 85 | 0.03 | 0.03 (−0.32, 0.39) | 0.05 | 0.05 |
| Malawi | iLiNS−DOSE (31) | KDI | 425 | 151 | −0.07 | 0.13 (−0.06, 0.31) | 0.10 | 0.10 | 134 | 41 | 0.06 | −0.03 (−0.39, 0.34) | 0.05 | 0.05 |
| Mali | PROMIS CS (32) | DMC | 751 | 789 | −0.05 | 0.14 (−0.09, 0.36) | 0.07 | 0.07 | 140 | 123 | −0.03 | −0.07 (−0.38, 0.24) | 0.07 | 0.07 |
| Zimbabwe | SHINE (HIV−) (33) | MDAT | 602 | 574 | −0.01 | 0.12 (−0.01, 0.25) | 0.20 | 0.20 | 203 | 191 | −0.14 | −0.02 (−0.22, 0.19) | 0.15 | 0.15 |
| Zimbabwe | SHINE (HIV+) (34) | MDAT | 149 | 118 | −0.02 | 0.08 (−0.17, 0.32) | 0.05 | 0.05 | 18 | 29 | −0.27 | 0.65 (0.15, 1.16) | 0.03 | 0.03 |
| | | | **4533** | **4041** | | **I² = 0.00, Tau² = 0.00** | | | **2084** | **1539** | | **I² = 0.00, Tau² = 0.00** | | |
| **Fixed** | | | | | | **0.12 (0.06, 0.17)** | | | | | | **0.04 (−0.04, 0.13)** | | |
| **Random** | | | | | | **0.12 (0.06, 0.17)** | | | | | | **0.04 (−0.04, 0.13)** | | |

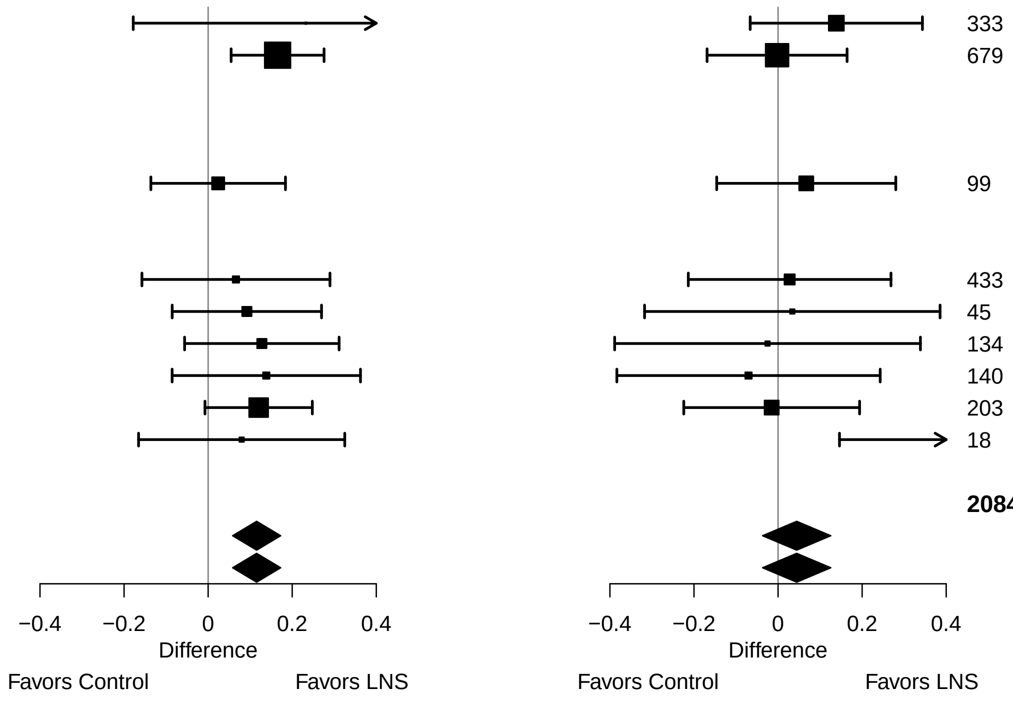

#### Supplemental figure 5K: Mean difference in fine motor z-score

#### 5K8: Stratified by Child baseline stunting

**P−for−interaction = 0.613**
**Difference in MDs = 0.03 (−0.09, 0.15)**

| Country | Trial | Tool | No: LNS N | No: Control N | No: Control Mean | No: MD (95% CI) | No: Fixed W | No: Random W | Yes: LNS N | Yes: Control N | Yes: Control Mean | Yes: MD (95% CI) | Yes: Fixed W | Yes: Random W |
|---|---|---|---|---|---|---|---|---|---|---|---|---|---|---|
| Bangladesh | JiVitA−4 (21) | BSID−III | 327 | 114 | −0.12 | 0.19 (−0.04, 0.42) | 0.06 | 0.06 | 116 | 29 | −0.05 | 0.01 (−0.30, 0.31) | 0.15 | 0.15 |
| Bangladesh | RDNS (22) | DMC | 1231 | 567 | −0.01 | 0.10 (0.00, 0.19) | 0.34 | 0.34 | 348 | 182 | −0.24 | 0.16 (−0.08, 0.40) | 0.24 | 0.24 |
| Bangladesh | WASH−B (23) | EASQ | | | | | | | | | | | | |
| Burkina Faso | iLiNS−Zinc (24) | DMC | | | | | | | | | | | | |
| Ghana | GHANA (25) | | | | | | | | | | | | | |
| Ghana | iLiNS−DYADG (26) | KDI | 259 | 504 | 0.04 | 0.00 (−0.13, 0.14) | 0.17 | 0.17 | 22 | 57 | −0.28 | 0.24 (−0.38, 0.85) | 0.04 | 0.04 |
| Haiti | HAITI (27) | | | | | | | | | | | | | |
| Kenya | WASH−B (28) | EASQ | | | | | | | | | | | | |
| Madagascar | MAHAY (29) | ASQI | | | | | | | | | | | | |
| Malawi | iLiNS−DYADM (30) | KDI | 152 | 320 | 0.06 | 0.02 (−0.16, 0.20) | 0.10 | 0.10 | 48 | 97 | −0.25 | 0.21 (−0.17, 0.59) | 0.10 | 0.10 |
| Malawi | iLiNS−DOSE (31) | KDI | 465 | 147 | −0.06 | 0.15 (−0.01, 0.32) | 0.11 | 0.11 | 181 | 74 | −0.08 | −0.08 (−0.39, 0.23) | 0.15 | 0.15 |
| Mali | PROMIS CS (32) | DMC | | | | | | | | | | | | |
| Zimbabwe | SHINE (HIV−) (33) | MDAT | 532 | 490 | 0.02 | 0.03 (−0.10, 0.16) | 0.18 | 0.18 | 122 | 105 | −0.24 | 0.17 (−0.06, 0.40) | 0.26 | 0.26 |
| Zimbabwe | SHINE (HIV+) (34) | MDAT | 115 | 89 | 0.04 | 0.17 (−0.08, 0.42) | 0.05 | 0.05 | 34 | 32 | −0.25 | 0.15 (−0.29, 0.59) | 0.07 | 0.07 |
| | | | **3081** | **2231** | | **I² = 0.00, Tau² = 0.00** | | | **871** | **576** | | **I² = 0.00, Tau² = 0.00** | | |
| **Fixed** | | | | | | **0.08 (0.02, 0.13)** | | | | | | **0.11 (−0.01, 0.23)** | | |
| **Random** | | | | | | **0.08 (0.02, 0.13)** | | | | | | **0.11 (−0.01, 0.23)** | | |

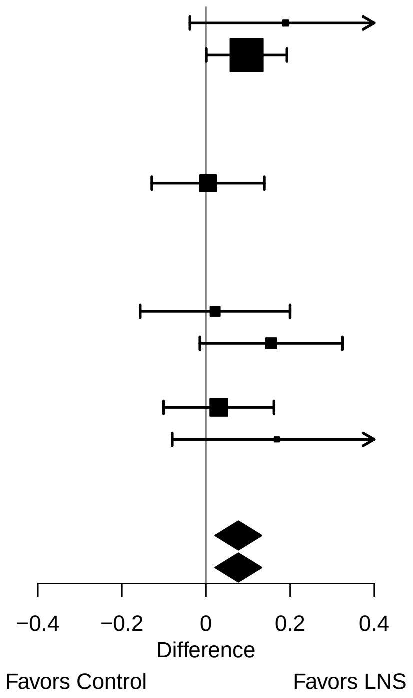

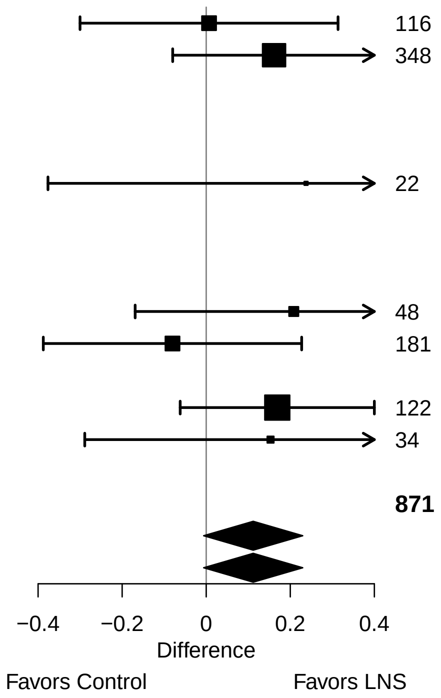

#### Supplemental figure 5K: Mean difference in fine motor z-score

#### 5K9: Stratified by Child baseline acute malnutrition

**P−for−interaction = 0.992**
**Difference in MDs = 0.00 (−0.18, 0.18)**

| | | | No | | | | | | Yes | | | | | |
|---|---|---|---|---|---|---|---|---|---|---|---|---|---|---|
| **Country** | **Trial** | **Tool** | **LNS N** | **Control N** | **Control Mean** | **MD (95% CI)** | **Fixed W** | **Random W** | **LNS N** | **Control N** | **Control Mean** | **MD (95% CI)** | **Fixed W** | **Random W** |
| Bangladesh | JiVitA−4 (21) | BSID−III | 359 | 112 | −0.04 | 0.12 (−0.09, 0.32) | 0.06 | 0.06 | 84 | 31 | −0.36 | 0.21 (−0.14, 0.56) | 0.31 | 0.31 |
| Bangladesh | RDNS (22) | DMC | 1455 | 697 | −0.03 | 0.10 (0.02, 0.18) | 0.38 | 0.38 | 124 | 53 | −0.48 | 0.33 (−0.18, 0.85) | 0.15 | 0.15 |
| Bangladesh | WASH−B (23) | EASQ | | | | | | | | | | | | |
| Burkina Faso | iLiNS−Zinc (24) | DMC | | | | | | | | | | | | |
| Ghana | GHANA (25) | | | | | | | | | | | | | |
| Ghana | iLiNS−DYADG (26) | KDI | 265 | 509 | 0.02 | 0.06 (−0.08, 0.19) | 0.15 | 0.15 | 17 | 52 | −0.14 | −0.44 (−1.15, 0.26) | 0.08 | 0.08 |
| Haiti | HAITI (27) | | | | | | | | | | | | | |
| Kenya | WASH−B (28) | EASQ | | | | | | | | | | | | |
| Madagascar | MAHAY (29) | ASQI | | | | | | | | | | | | |
| Malawi | iLiNS−DYADM (30) | KDI | 186 | 383 | 0.00 | 0.06 (−0.11, 0.23) | 0.09 | 0.09 | 14 | 37 | −0.21 | 0.13 (−0.51, 0.76) | 0.10 | 0.10 |
| Malawi | iLiNS−DOSE (31) | KDI | 609 | 206 | −0.04 | 0.09 (−0.06, 0.24) | 0.11 | 0.11 | 37 | 15 | −0.43 | 0.02 (−0.81, 0.86) | 0.06 | 0.06 |
| Mali | PROMIS CS (32) | DMC | | | | | | | | | | | | |
| Zimbabwe | SHINE (HIV−) (33) | MDAT | 620 | 566 | −0.02 | 0.06 (−0.06, 0.18) | 0.17 | 0.17 | 36 | 34 | −0.05 | 0.02 (−0.40, 0.43) | 0.22 | 0.22 |
| Zimbabwe | SHINE (HIV+) (34) | MDAT | 137 | 114 | −0.04 | 0.17 (−0.06, 0.40) | 0.05 | 0.05 | 14 | 8 | 0.02 | 0.26 (−0.39, 0.91) | 0.09 | 0.09 |
| | | | **3631** | **2587** | | **I² = 0.00, Tau² = 0.00** | | | **326** | **230** | | **I² = 0.00, Tau² = 0.00** | | |
| **Fixed** | | | | | | **0.09 (0.04, 0.14)** | | | | | | **0.12 (−0.07, 0.32)** | | |
| **Random** | | | | | | **0.09 (0.04, 0.14)** | | | | | | **0.12 (−0.07, 0.32)** | | |

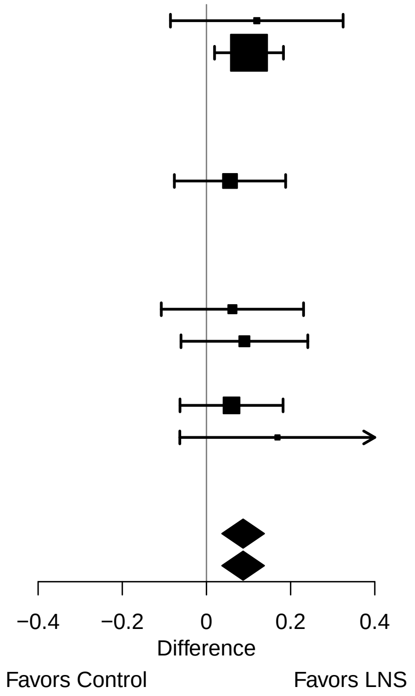

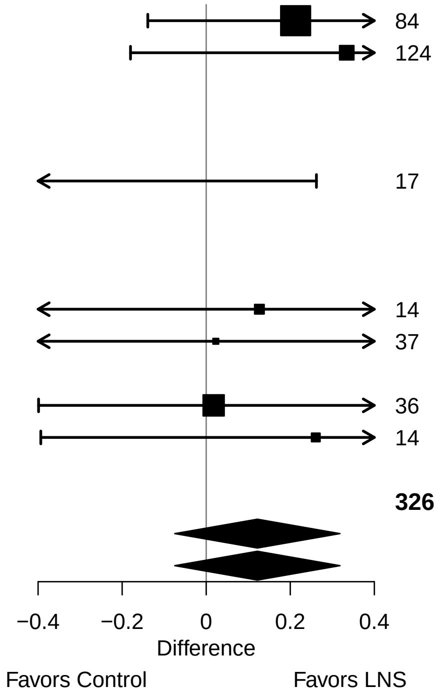

#### Supplemental figure 5K: Mean difference in fine motor z-score

##### 5K10: Stratified by Child baseline anemia

**P−for−interaction = 0.084**
**Difference in MDs = 0.14 (−0.02, 0.29)**

| | | | Not anemic | | | | | | Anemic | | | | | |
|---|---|---|---|---|---|---|---|---|---|---|---|---|---|---|
| **Country** | **Trial** | **Tool** | **LNS N** | **Control N** | **Control Mean** | **MD (95% CI)** | **Fixed W** | **Random W** | **LNS N** | **Control N** | **Control Mean** | **MD (95% CI)** | **Fixed W** | **Random W** |
| Bangladesh | JiVitA−4 (21) | BSID−III | | | | | | | | | | | | |
| Bangladesh | RDNS (22) | DMC | 206 | 96 | −0.10 | 0.15 (−0.08, 0.39) | 0.23 | 0.23 | 315 | 157 | −0.06 | 0.07 (−0.10, 0.25) | 0.32 | 0.32 |
| Bangladesh | WASH−B (23) | EASQ | | | | | | | | | | | | |
| Burkina Faso | iLiNS−Zinc (24) | DMC | | | | | | | | | | | | |
| Ghana | GHANA (25) | | | | | | | | | | | | | |
| Ghana | iLiNS−DYADG (26) | KDI | 180 | 321 | 0.01 | −0.07 (−0.25, 0.11) | 0.38 | 0.38 | 81 | 189 | −0.01 | 0.20 (−0.03, 0.44) | 0.18 | 0.18 |
| Haiti | HAITI (27) | | | | | | | | | | | | | |
| Kenya | WASH−B (28) | EASQ | | | | | | | | | | | | |
| Madagascar | MAHAY (29) | ASQI | | | | | | | | | | | | |
| Malawi | iLiNS−DYADM (30) | KDI | 65 | 146 | 0.04 | −0.12 (−0.39, 0.16) | 0.15 | 0.15 | 138 | 273 | −0.03 | 0.14 (−0.05, 0.34) | 0.26 | 0.26 |
| Malawi | iLiNS−DOSE (31) | KDI | 224 | 94 | −0.01 | 0.00 (−0.22, 0.22) | 0.24 | 0.24 | 418 | 126 | −0.11 | 0.15 (−0.05, 0.36) | 0.24 | 0.24 |
| Mali | PROMIS CS (32) | DMC | | | | | | | | | | | | |
| Zimbabwe | SHINE (HIV−) (33) | MDAT | | | | | | | | | | | | |
| Zimbabwe | SHINE (HIV+) (34) | MDAT | | | | | | | | | | | | |
| | | | **675** | **657** | | **I² = 0.00, Tau² = 0.00** | | | **952** | **745** | | **I² = 0.00, Tau² = 0.00** | | |
| **Fixed** | | | | | | **−0.01 (−0.12, 0.10)** | | | | | | **0.13 (0.03, 0.24)** | | |
| **Random** | | | | | | **−0.01 (−0.12, 0.10)** | | | | | | **0.13 (0.03, 0.24)** | | |

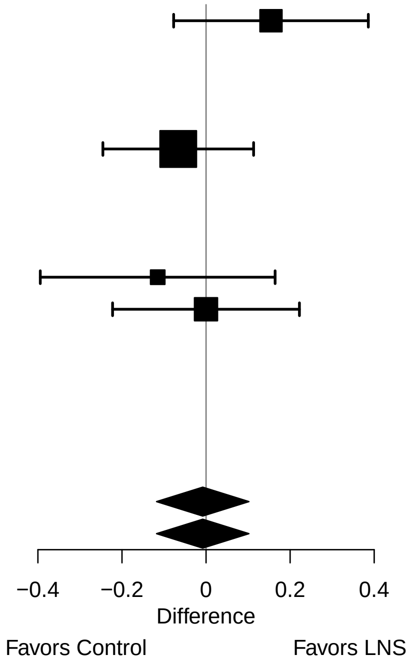

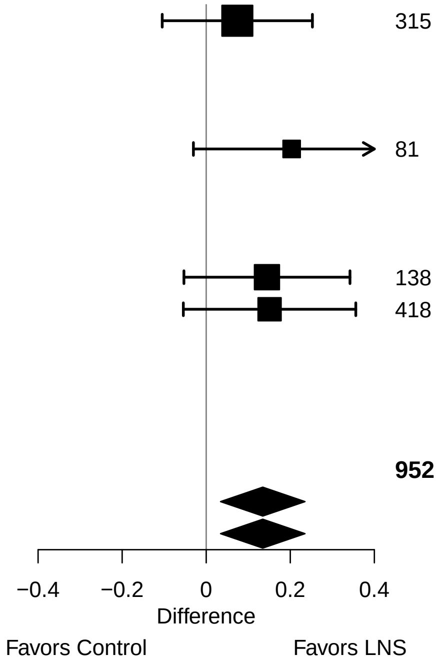

### Supplemental figure 5L: Mean difference in executive function z-score

#### 5L1: Stratified by Maternal height

**P−for−interaction = 0.522**
**Difference in MDs = −0.04 (−0.15, 0.08)**

| Country | Trial | Tool | LNS N | Control N | Control Mean | At least 150.1 cm MD (95% CI) | Fixed W | Random W | LNS N | Control N | Control Mean | Less than 150.1 cm MD (95% CI) | Fixed W | Random W |
|---|---|---|---|---|---|---|---|---|---|---|---|---|---|---|
| Bangladesh | JiVitA−4 (21) | | | | | | | | | | | | | |
| Bangladesh | RDNS (22) | A not B task | 289 | 136 | −0.06 | 0.07 (−0.08, 0.23) | 0.12 | 0.12 | 222 | 120 | −0.11 | 0.16 (−0.08, 0.40) | 0.14 | 0.14 |
| Bangladesh | WASH−B (23) | A not B task | 585 | 1796 | 0.02 | 0.02 (−0.07, 0.12) | 0.33 | 0.33 | 507 | 1489 | 0.00 | −0.04 (−0.15, 0.06) | 0.74 | 0.74 |
| Burkina Faso | iLiNS−Zinc (24) | | | | | | | | | | | | | |
| Ghana | GHANA (25) | | | | | | | | | | | | | |
| Ghana | iLiNS−DYADG (26) | A not B task | 273 | 535 | 0.01 | 0.00 (−0.14, 0.15) | 0.14 | 0.14 | 12 | 31 | −0.19 | 0.25 (−0.44, 0.94) | 0.02 | 0.02 |
| Haiti | HAITI (27) | | | | | | | | | | | | | |
| Kenya | WASH−B (28) | | | | | | | | | | | | | |
| Madagascar | MAHAY (29) | | | | | | | | | | | | | |
| Malawi | iLiNS−DYADM (30) | A not B task | 158 | 315 | 0.06 | −0.10 (−0.29, 0.08) | 0.08 | 0.08 | 24 | 52 | −0.16 | 0.02 (−0.51, 0.56) | 0.03 | 0.03 |
| Malawi | iLiNS−DOSE (31) | A not B task | 398 | 142 | 0.07 | −0.07 (−0.26, 0.12) | 0.08 | 0.08 | 76 | 21 | 0.21 | −0.38 (−0.88, 0.11) | 0.03 | 0.03 |
| Mali | PROMIS CS (32) | | | | | | | | | | | | | |
| Zimbabwe | SHINE (HIV−) (33) | A not B task | 737 | 703 | −0.02 | 0.05 (−0.07, 0.17) | 0.19 | 0.19 | 32 | 22 | −0.09 | −0.07 (−0.54, 0.40) | 0.04 | 0.04 |
| Zimbabwe | SHINE (HIV+) (34) | A not B task | 145 | 124 | 0.05 | −0.08 (−0.31, 0.14) | 0.06 | 0.06 | 10 | 5 | −0.11 | 0.29 (−0.81, 1.39) | 0.01 | 0.01 |
| | | | **2585** | **3751** | | **I² = 0.00, Tau² = 0.00** | | | **883** | **1740** | | **I² = 0.00, Tau² = 0.00** | | |
| **Fixed** | | | | | | **0.01 (−0.05, 0.06)** | | | | | | **−0.02 (−0.11, 0.07)** | | |
| **Random** | | | | | | **0.01 (−0.05, 0.06)** | | | | | | **−0.02 (−0.11, 0.07)** | | |

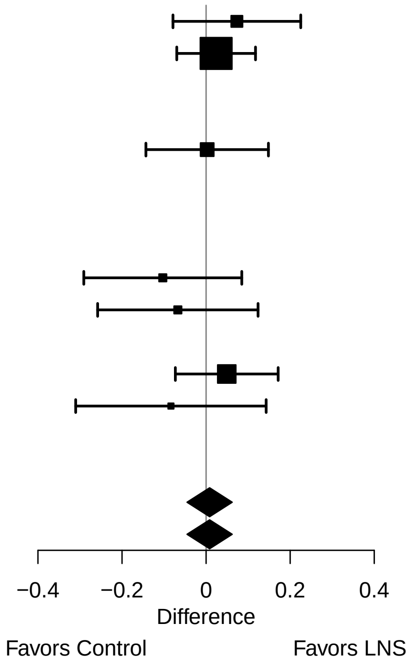

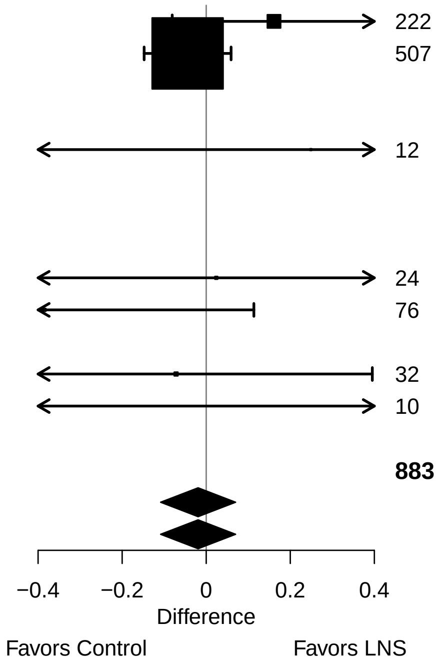

#### Supplemental figure 5L: Mean difference in executive function z-score

##### 5L2: Stratified by Maternal BMI

**P−for−interaction = 0.888**
**Difference in MDs = 0.01 (−0.09, 0.11)**

| | | | At least 20 kg/m² | | | | | | Less than 20 kg/m² | | | | | |
|---|---|---|---|---|---|---|---|---|---|---|---|---|---|---|
| Country | Trial | Tool | LNS N | Control N | Control Mean | MD (95% CI) | Fixed W | Random W | LNS N | Control N | Control Mean | MD (95% CI) | Fixed W | Random W |
| Bangladesh | JiVitA−4 (21) | | | | | | | | | | | | | |
| Bangladesh | RDNS (22) | A not B task | 226 | 103 | −0.03 | 0.09 (−0.13, 0.32) | 0.07 | 0.07 | 285 | 153 | −0.12 | 0.12 (−0.05, 0.29) | 0.18 | 0.18 |
| Bangladesh | WASH−B (23) | A not B task | 495 | 1517 | 0.01 | 0.02 (−0.08, 0.12) | 0.36 | 0.36 | 597 | 1767 | 0.01 | −0.03 (−0.13, 0.06) | 0.60 | 0.60 |
| Burkina Faso | iLiNS−Zinc (24) | | | | | | | | | | | | | |
| Ghana | GHANA (25) | | | | | | | | | | | | | |
| Ghana | iLiNS−DYADG (26) | A not B task | 251 | 464 | 0.01 | −0.01 (−0.16, 0.15) | 0.15 | 0.15 | 34 | 102 | −0.09 | 0.13 (−0.26, 0.53) | 0.03 | 0.03 |
| Haiti | HAITI (27) | | | | | | | | | | | | | |
| Kenya | WASH−B (28) | | | | | | | | | | | | | |
| Madagascar | MAHAY (29) | | | | | | | | | | | | | |
| Malawi | iLiNS−DYADM (30) | A not B task | 108 | 221 | 0.06 | −0.11 (−0.34, 0.11) | 0.07 | 0.07 | 74 | 146 | −0.02 | −0.04 (−0.33, 0.25) | 0.06 | 0.06 |
| Malawi | iLiNS−DOSE (31) | A not B task | 360 | 119 | 0.10 | −0.13 (−0.34, 0.08) | 0.08 | 0.08 | 112 | 44 | 0.05 | −0.06 (−0.41, 0.29) | 0.04 | 0.04 |
| Mali | PROMIS CS (32) | | | | | | | | | | | | | |
| Zimbabwe | SHINE (HIV−) (33) | A not B task | 563 | 539 | 0.00 | 0.03 (−0.10, 0.16) | 0.22 | 0.22 | 93 | 87 | −0.14 | 0.19 (−0.09, 0.47) | 0.07 | 0.07 |
| Zimbabwe | SHINE (HIV+) (34) | A not B task | 115 | 99 | 0.06 | 0.01 (−0.24, 0.27) | 0.05 | 0.05 | 22 | 21 | −0.14 | −0.05 (−0.68, 0.58) | 0.01 | 0.01 |
| | | | **2118** | **3062** | | **I² = 0.00, Tau² = 0.00** | | | **1217** | **2320** | | **I² = 0.00, Tau² = 0.00** | | |
| **Fixed** | | | | | | **0.00 (−0.06, 0.06)** | | | | | | **0.01 (−0.06, 0.09)** | | |
| **Random** | | | | | | **0.00 (−0.06, 0.06)** | | | | | | **0.01 (−0.06, 0.09)** | | |

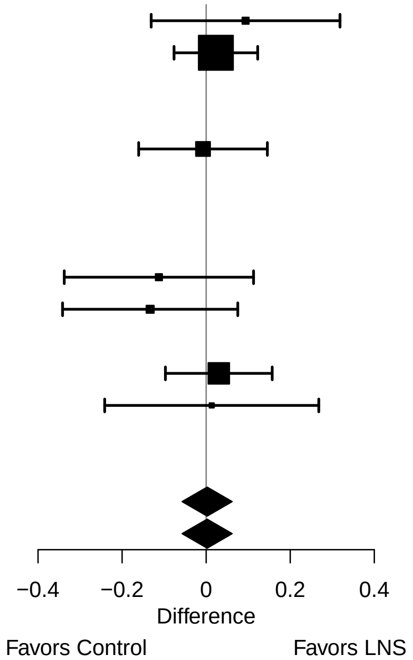

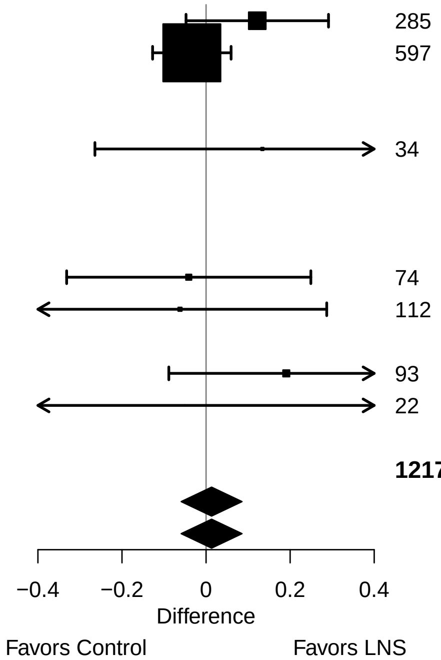

#### Supplemental figure 5L: Mean difference in executive function z-score

##### 5L3: Stratified by Maternal age

**P−for−interaction = 0.470**
**Difference in MDs = 0.04 (−0.06, 0.13)**

| Country | Trial | Tool | At least 25 y LNS N | At least 25 y Control N | At least 25 y Control Mean | At least 25 y MD (95% CI) | At least 25 y Fixed W | At least 25 y Random W | Less than 25 y LNS N | Less than 25 y Control N | Less than 25 y Control Mean | Less than 25 y MD (95% CI) | Less than 25 y Fixed W | Less than 25 y Random W |
|---|---|---|---|---|---|---|---|---|---|---|---|---|---|---|
| Bangladesh | JiVitA−4 (21) | | | | | | | | | | | | | |
| Bangladesh | RDNS (22) | A not B task | 146 | 80 | −0.16 | 0.19 (−0.09, 0.47) | 0.05 | 0.09 | 387 | 186 | −0.03 | 0.06 (−0.07, 0.19) | 0.25 | 0.25 |
| Bangladesh | WASH−B (23) | A not B task | 471 | 1469 | 0.02 | −0.05 (−0.15, 0.05) | 0.42 | 0.26 | 621 | 1844 | −0.01 | 0.04 (−0.07, 0.14) | 0.41 | 0.41 |
| Burkina Faso | iLiNS−Zinc (24) | | | | | | | | | | | | | |
| Ghana | GHANA (25) | | | | | | | | | | | | | |
| Ghana | iLiNS−DYADG (26) | A not B task | 185 | 362 | −0.06 | 0.08 (−0.09, 0.26) | 0.14 | 0.16 | 105 | 214 | 0.08 | −0.08 (−0.31, 0.16) | 0.07 | 0.07 |
| Haiti | HAITI (27) | | | | | | | | | | | | | |
| Kenya | WASH−B (28) | | | | | | | | | | | | | |
| Madagascar | MAHAY (29) | | | | | | | | | | | | | |
| Malawi | iLiNS−DYADM (30) | A not B task | 89 | 190 | 0.06 | −0.26 (−0.51, −0.01) | 0.07 | 0.11 | 93 | 179 | −0.01 | 0.08 (−0.18, 0.33) | 0.06 | 0.06 |
| Malawi | iLiNS−DOSE (31) | A not B task | 253 | 86 | 0.04 | −0.14 (−0.40, 0.11) | 0.07 | 0.10 | 212 | 73 | 0.17 | −0.13 (−0.38, 0.13) | 0.06 | 0.06 |
| Mali | PROMIS CS (32) | | | | | | | | | | | | | |
| Zimbabwe | SHINE (HIV−) (33) | A not B task | 399 | 358 | −0.03 | 0.05 (−0.10, 0.20) | 0.19 | 0.19 | 306 | 300 | −0.02 | 0.04 (−0.15, 0.23) | 0.12 | 0.12 |
| Zimbabwe | SHINE (HIV+) (34) | A not B task | 115 | 97 | 0.05 | −0.07 (−0.36, 0.21) | 0.05 | 0.09 | 27 | 23 | 0.21 | −0.03 (−0.44, 0.38) | 0.02 | 0.02 |
| | | | **1658** | **2642** | | **I² = 0.35, Tau² = 0.01** | | | **1751** | **2819** | | **I² = 0.00, Tau² = 0.00** | | |
| **Fixed** | | | | | | **−0.02 (−0.09, 0.04)** | | | | | | **0.03 (−0.04, 0.09)** | | |
| **Random** | | | | | | **−0.02 (−0.12, 0.07)** | | | | | | **0.03 (−0.04, 0.09)** | | |

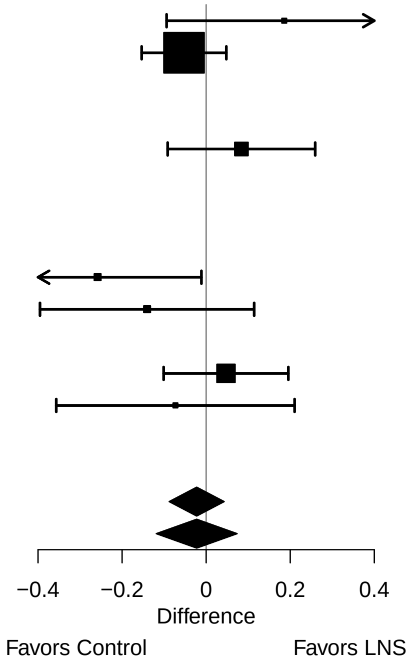

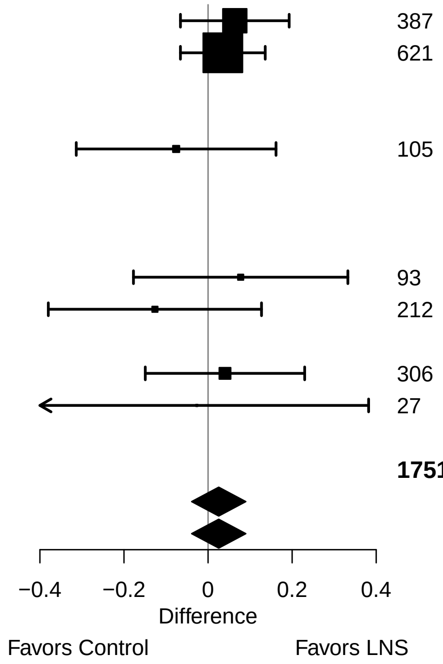

#### Supplemental figure 5L: Mean difference in executive function z-score

#### 5L4: Stratified by Maternal education

P−for−interaction = 0.495
Difference in MDs = 0.04 (−0.08, 0.16)

| Country | Trial | Tool | LNS N | Control N | Control Mean | Primary or greater MD (95% CI) | Fixed W | Random W | LNS N | Control N | Control Mean | Incomplete or no formal MD (95% CI) | Fixed W | Random W |
|---|---|---|---|---|---|---|---|---|---|---|---|---|---|---|
| Bangladesh | JiVitA−4 (21) | | | | | | | | | | | | | |
| Bangladesh | RDNS (22) | A not B task | 394 | 191 | −0.01 | 0.08 (−0.08, 0.25) | 0.11 | 0.11 | 139 | 75 | −0.22 | 0.14 (−0.18, 0.45) | 0.08 | 0.08 |
| Bangladesh | WASH−B (23) | A not B task | 776 | 2386 | 0.03 | −0.02 (−0.10, 0.06) | 0.47 | 0.47 | 321 | 941 | −0.07 | 0.04 (−0.10, 0.18) | 0.42 | 0.42 |
| Burkina Faso | iLiNS−Zinc (24) | | | | | | | | | | | | | |
| Ghana | GHANA (25) | | | | | | | | | | | | | |
| Ghana | iLiNS−DYADG (26) | A not B task | 223 | 449 | 0.03 | 0.01 (−0.15, 0.17) | 0.12 | 0.12 | 67 | 127 | −0.13 | 0.09 (−0.22, 0.39) | 0.09 | 0.09 |
| Haiti | HAITI (27) | | | | | | | | | | | | | |
| Kenya | WASH−B (28) | | | | | | | | | | | | | |
| Madagascar | MAHAY (29) | | | | | | | | | | | | | |
| Malawi | iLiNS−DYADM (30) | A not B task | 26 | 59 | −0.14 | −0.27 (−0.73, 0.18) | 0.01 | 0.01 | 155 | 307 | 0.06 | −0.07 (−0.26, 0.12) | 0.21 | 0.21 |
| Malawi | iLiNS−DOSE (31) | A not B task | 121 | 43 | 0.09 | −0.05 (−0.38, 0.28) | 0.03 | 0.03 | 345 | 116 | 0.10 | −0.17 (−0.38, 0.05) | 0.17 | 0.17 |
| Mali | PROMIS CS (32) | | | | | | | | | | | | | |
| Zimbabwe | SHINE (HIV−) (33) | A not B task | 716 | 679 | −0.02 | 0.06 (−0.06, 0.18) | 0.21 | 0.21 | 27 | 23 | −0.12 | −0.04 (−0.73, 0.65) | 0.02 | 0.02 |
| Zimbabwe | SHINE (HIV+) (34) | A not B task | 137 | 114 | 0.03 | −0.04 (−0.28, 0.21) | 0.05 | 0.05 | 8 | 7 | 0.28 | −0.20 (−1.18, 0.79) | 0.01 | 0.01 |
| | | | **2393** | **3921** | | **I² = 0.00, Tau² = 0.00** | | | **1062** | **1596** | | **I² = 0.00, Tau² = 0.00** | | |
| **Fixed** | | | | | | **0.01 (−0.05, 0.06)** | | | | | | **−0.01 (−0.10, 0.08)** | | |
| **Random** | | | | | | **0.01 (−0.05, 0.06)** | | | | | | **−0.01 (−0.10, 0.08)** | | |

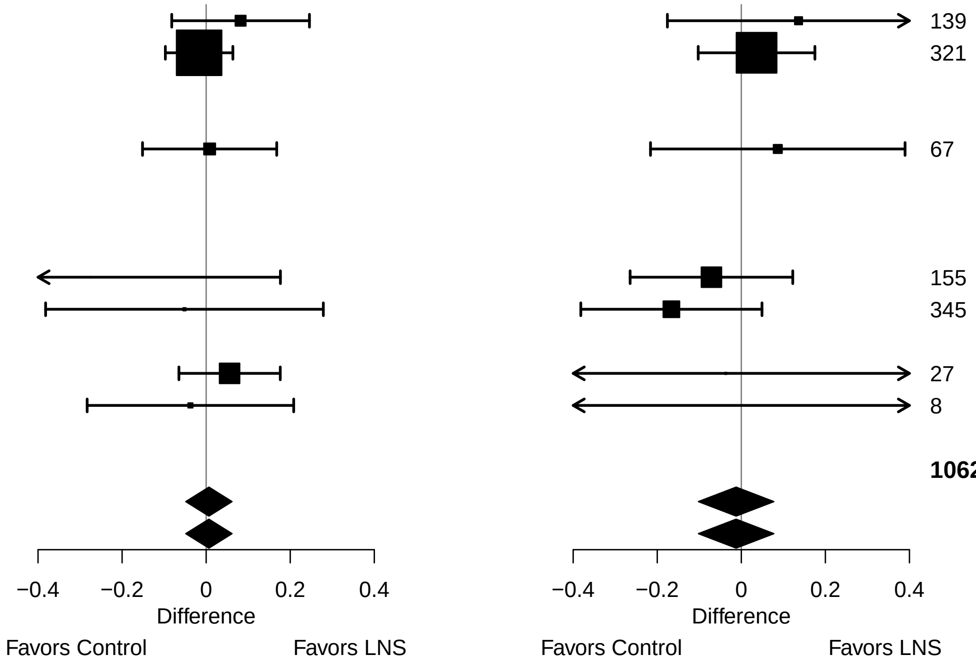

**Supplemental figure 5L: Mean difference in executive function z-score**

#### 5L5: Stratified by Maternal depressive symptoms

**P−for−interaction = 0.332**
**Difference in MDs = 0.06 (−0.06, 0.17)**

| | | | Less than 75th percentile | | | | | | At least 75th percentile | | | | | |
|---|---|---|---|---|---|---|---|---|---|---|---|---|---|---|
| **Country** | **Trial** | **Tool** | **LNS N** | **Control N** | **Control Mean** | **MD (95% CI)** | **Fixed W** | **Random W** | **LNS N** | **Control N** | **Control Mean** | **MD (95% CI)** | **Fixed W** | **Random W** |
| Bangladesh | JiVitA−4 (21) | | | | | | | | | | | | | |
| Bangladesh | RDNS (22) | A not B task | 345 | 150 | −0.06 | 0.12 (−0.05, 0.28) | 0.11 | 0.11 | 166 | 100 | −0.08 | 0.12 (−0.18, 0.41) | 0.11 | 0.11 |
| Bangladesh | WASH−B (23) | A not B task | 847 | 2392 | 0.01 | 0.01 (−0.07, 0.09) | 0.52 | 0.52 | 230 | 852 | −0.03 | −0.07 (−0.23, 0.09) | 0.38 | 0.38 |
| Burkina Faso | iLiNS−Zinc (24) | | | | | | | | | | | | | |
| Ghana | GHANA (25) | | | | | | | | | | | | | |
| Ghana | iLiNS−DYADG (26) | A not B task | 216 | 373 | −0.03 | 0.03 (−0.14, 0.20) | 0.11 | 0.11 | 66 | 185 | 0.09 | 0.01 (−0.26, 0.28) | 0.13 | 0.13 |
| Haiti | HAITI (27) | | | | | | | | | | | | | |
| Kenya | WASH−B (28) | | | | | | | | | | | | | |
| Madagascar | MAHAY (29) | | | | | | | | | | | | | |
| Malawi | iLiNS−DYADM (30) | A not B task | 130 | 254 | 0.03 | −0.07 (−0.29, 0.15) | 0.06 | 0.06 | 42 | 93 | 0.00 | −0.07 (−0.42, 0.28) | 0.08 | 0.08 |
| Malawi | iLiNS−DOSE (31) | A not B task | | | | | | | | | | | | |
| Mali | PROMIS CS (32) | | | | | | | | | | | | | |
| Zimbabwe | SHINE (HIV−) (33) | A not B task | 542 | 520 | −0.06 | 0.05 (−0.09, 0.20) | 0.15 | 0.15 | 186 | 164 | 0.07 | 0.01 (−0.18, 0.21) | 0.25 | 0.25 |
| Zimbabwe | SHINE (HIV+) (34) | A not B task | 122 | 96 | 0.05 | −0.03 (−0.29, 0.22) | 0.05 | 0.05 | 29 | 29 | 0.18 | −0.28 (−0.75, 0.18) | 0.04 | 0.04 |
| | | | **2202** | **3785** | | **I² = 0.00, Tau² = 0.00** | | | **719** | **1423** | | **I² = 0.00, Tau² = 0.00** | | |
| **Fixed** | | | | | | **0.02 (−0.03, 0.08)** | | | | | | **−0.03 (−0.12, 0.07)** | | |
| **Random** | | | | | | **0.02 (−0.03, 0.08)** | | | | | | **−0.03 (−0.12, 0.07)** | | |

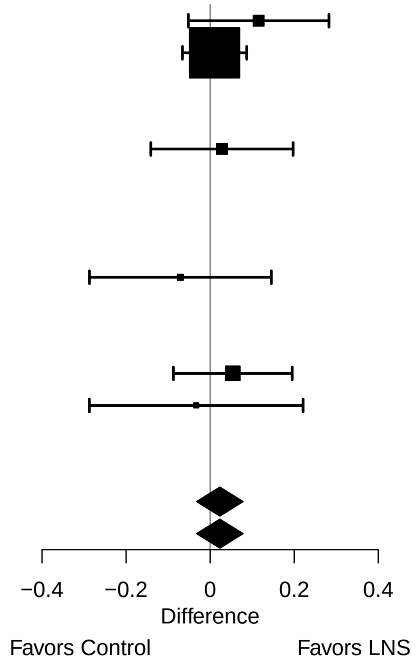

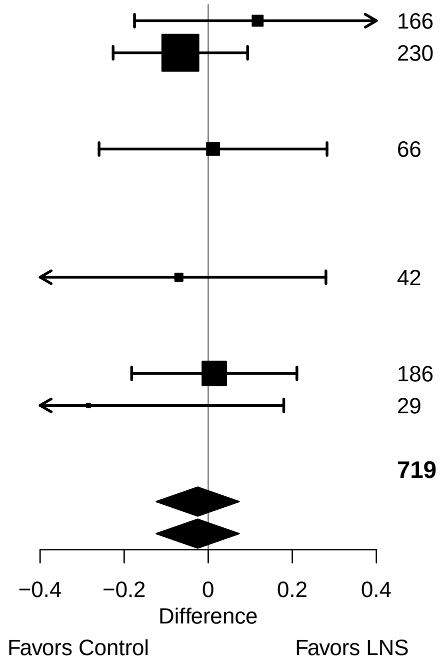

#### Supplemental figure 5L: Mean difference in executive function z-score

#### 5L6: Stratified by Child sex

**P−for−interaction = 0.414**
**Difference in MDs = −0.04 (−0.12, 0.05)**

| Country | Trial | Tool | LNS N | Control N | Control Mean | Male MD (95% CI) | Fixed W | Random W | LNS N | Control N | Control Mean | Female MD (95% CI) | Fixed W | Random W |
|---|---|---|---|---|---|---|---|---|---|---|---|---|---|---|
| Bangladesh | JiVitA−4 (21) | | | | | | | | | | | | | |
| Bangladesh | RDNS (22) | A not B task | 261 | 133 | 0.05 | −0.08 (−0.26, 0.10) | 0.13 | 0.13 | 272 | 133 | −0.19 | 0.28 (0.09, 0.47) | 0.11 | 0.15 |
| Bangladesh | WASH−B (23) | A not B task | 540 | 1671 | 0.00 | 0.02 (−0.08, 0.12) | 0.44 | 0.41 | 557 | 1656 | 0.01 | −0.02 (−0.12, 0.07) | 0.44 | 0.24 |
| Burkina Faso | iLiNS−Zinc (24) | | | | | | | | | | | | | |
| Ghana | GHANA (25) | | | | | | | | | | | | | |
| Ghana | iLiNS−DYADG (26) | A not B task | 145 | 273 | −0.04 | 0.13 (−0.08, 0.34) | 0.10 | 0.10 | 145 | 303 | 0.02 | −0.08 (−0.27, 0.12) | 0.11 | 0.14 |
| Haiti | HAITI (27) | | | | | | | | | | | | | |
| Kenya | WASH−B (28) | | | | | | | | | | | | | |
| Madagascar | MAHAY (29) | | | | | | | | | | | | | |
| Malawi | iLiNS−DYADM (30) | A not B task | 92 | 176 | 0.00 | −0.01 (−0.27, 0.24) | 0.06 | 0.07 | 90 | 193 | 0.05 | −0.16 (−0.41, 0.08) | 0.07 | 0.11 |
| Malawi | iLiNS−DOSE (31) | A not B task | 235 | 93 | 0.12 | −0.16 (−0.41, 0.08) | 0.07 | 0.07 | 240 | 71 | 0.04 | −0.05 (−0.31, 0.21) | 0.06 | 0.10 |
| Mali | PROMIS CS (32) | | | | | | | | | | | | | |
| Zimbabwe | SHINE (HIV−) (33) | A not B task | 399 | 364 | −0.06 | 0.11 (−0.04, 0.27) | 0.17 | 0.18 | 388 | 375 | 0.02 | −0.03 (−0.18, 0.11) | 0.18 | 0.18 |
| Zimbabwe | SHINE (HIV+) (34) | A not B task | 74 | 68 | 0.05 | −0.10 (−0.43, 0.23) | 0.04 | 0.04 | 83 | 65 | 0.06 | −0.09 (−0.42, 0.23) | 0.04 | 0.07 |
| | | | **1746** | **2778** | | **I² = 0.05, Tau² = 0.00** | | | **1775** | **2796** | | **I² = 0.46, Tau² = 0.01** | | |
| **Fixed** | | | | | | **0.01 (−0.05, 0.08)** | | | | | | **−0.01 (−0.07, 0.05)** | | |
| **Random** | | | | | | **0.01 (−0.05, 0.08)** | | | | | | **−0.01 (−0.11, 0.09)** | | |

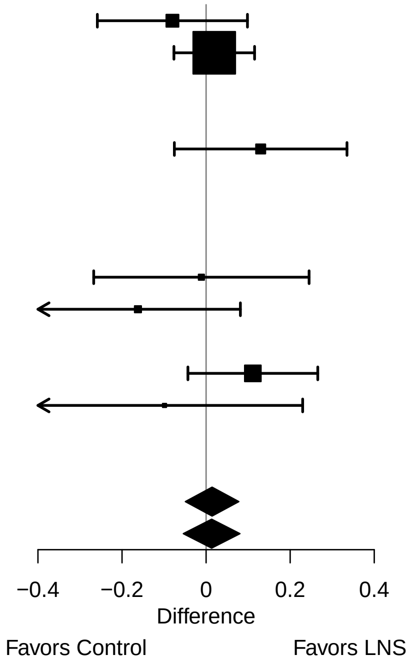

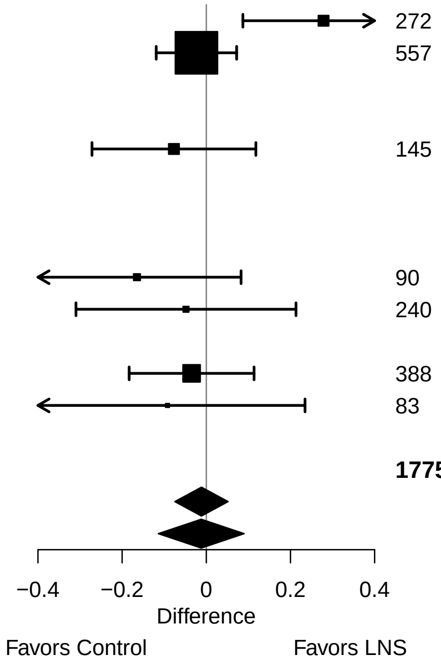

#### Supplemental figure 5L: Mean difference in executive function z-score

#### 5L7: Stratified by Child birth order

**P−for−interaction = 0.285**
**Difference in MDs = 0.05 (−0.04, 0.15)**

| Country | Trial | Tool | Later born LNS N | Later born Control N | Later born Control Mean | Later born MD (95% CI) | Later born Fixed W | Later born Random W | Firstborn LNS N | Firstborn Control N | Firstborn Control Mean | Firstborn MD (95% CI) | Firstborn Fixed W | Firstborn Random W |
|---|---|---|---|---|---|---|---|---|---|---|---|---|---|---|
| Bangladesh | JiVitA−4 (21) | | | | | | | | | | | | | |
| Bangladesh | RDNS (22) | A not B task | 320 | 163 | −0.14 | 0.19 (0.01, 0.38) | 0.09 | 0.11 | 213 | 103 | 0.05 | −0.05 (−0.26, 0.17) | 0.14 | 0.14 |
| Bangladesh | WASH−B (23) | A not B task | 683 | 2174 | 0.02 | −0.05 (−0.13, 0.03) | 0.46 | 0.34 | 394 | 1049 | −0.01 | 0.06 (−0.06, 0.17) | 0.48 | 0.48 |
| Burkina Faso | iLiNS−Zinc (24) | | | | | | | | | | | | | |
| Ghana | GHANA (25) | | | | | | | | | | | | | |
| Ghana | iLiNS−DYADG (26) | A not B task | 195 | 390 | −0.02 | 0.03 (−0.15, 0.20) | 0.10 | 0.13 | 95 | 186 | 0.03 | 0.02 (−0.22, 0.27) | 0.11 | 0.11 |
| Haiti | HAITI (27) | | | | | | | | | | | | | |
| Kenya | WASH−B (28) | | | | | | | | | | | | | |
| Madagascar | MAHAY (29) | | | | | | | | | | | | | |
| Malawi | iLiNS−DYADM (30) | A not B task | 145 | 297 | 0.05 | −0.09 (−0.29, 0.11) | 0.08 | 0.10 | 37 | 70 | −0.09 | −0.07 (−0.47, 0.33) | 0.04 | 0.04 |
| Malawi | iLiNS−DOSE (31) | A not B task | 313 | 109 | 0.05 | −0.10 (−0.32, 0.13) | 0.06 | 0.08 | 97 | 33 | 0.02 | 0.12 (−0.24, 0.48) | 0.05 | 0.05 |
| Mali | PROMIS CS (32) | | | | | | | | | | | | | |
| Zimbabwe | SHINE (HIV−) (33) | A not B task | 568 | 530 | −0.03 | 0.03 (−0.11, 0.17) | 0.16 | 0.17 | 186 | 178 | −0.02 | 0.09 (−0.11, 0.28) | 0.17 | 0.17 |
| Zimbabwe | SHINE (HIV+) (34) | A not B task | 137 | 106 | 0.02 | −0.10 (−0.34, 0.15) | 0.05 | 0.07 | 17 | 23 | 0.31 | −0.04 (−0.64, 0.56) | 0.02 | 0.02 |
| | | | **2361** | **3769** | | **I² = 0.21, Tau² = 0.00** | | | **1039** | **1642** | | **I² = 0.00, Tau² = 0.00** | | |
| **Fixed** | | | | | | **−0.02 (−0.07, 0.04)** | | | | | | **0.04 (−0.04, 0.12)** | | |
| **Random** | | | | | | **−0.01 (−0.08, 0.06)** | | | | | | **0.04 (−0.04, 0.12)** | | |

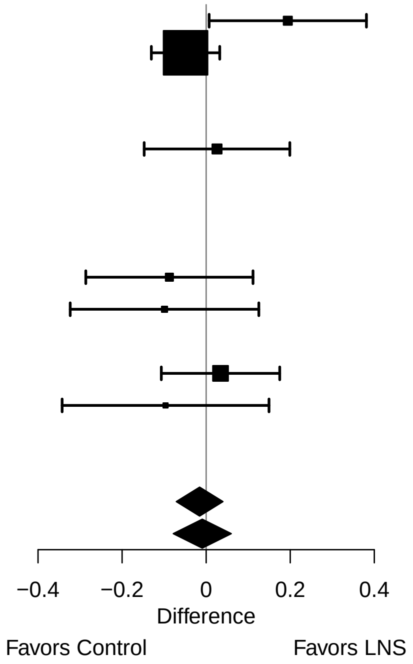

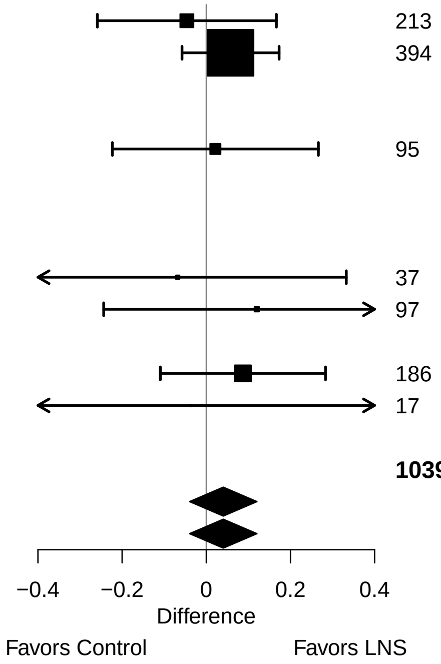

#### Supplemental figure 5L: Mean difference in executive function z-score

#### 5L8: Stratified by Child baseline stunting

**P−for−interaction = 0.702**
**Difference in MDs = −0.03 (−0.20, 0.13)**

| | | | No | | | | | | Yes | | | | | |
|---|---|---|---|---|---|---|---|---|---|---|---|---|---|---|
| **Country** | **Trial** | **Tool** | **LNS N** | **Control N** | **Control Mean** | **MD (95% CI)** | **Fixed W** | **Random W** | **LNS N** | **Control N** | **Control Mean** | **MD (95% CI)** | **Fixed W** | **Random W** |
| Bangladesh | JiVitA−4 (21) | | | | | | | | | | | | | |
| Bangladesh | RDNS (22) | A not B task | 404 | 187 | −0.02 | 0.07 (−0.08, 0.22) | 0.23 | 0.21 | 107 | 62 | −0.22 | 0.24 (−0.12, 0.59) | 0.17 | 0.18 |
| Bangladesh | WASH−B (23) | A not B task | | | | | | | | | | | | |
| Burkina Faso | iLiNS−Zinc (24) | | | | | | | | | | | | | |
| Ghana | GHANA (25) | | | | | | | | | | | | | |
| Ghana | iLiNS−DYADG (26) | A not B task | 248 | 485 | −0.01 | 0.04 (−0.12, 0.19) | 0.21 | 0.20 | 22 | 52 | 0.12 | −0.18 (−0.72, 0.36) | 0.08 | 0.09 |
| Haiti | HAITI (27) | | | | | | | | | | | | | |
| Kenya | WASH−B (28) | | | | | | | | | | | | | |
| Madagascar | MAHAY (29) | | | | | | | | | | | | | |
| Malawi | iLiNS−DYADM (30) | A not B task | 129 | 278 | 0.07 | −0.13 (−0.33, 0.08) | 0.12 | 0.14 | 43 | 76 | −0.06 | −0.10 (−0.49, 0.30) | 0.14 | 0.15 |
| Malawi | iLiNS−DOSE (31) | A not B task | 353 | 111 | 0.14 | −0.22 (−0.43, 0.00) | 0.11 | 0.13 | 122 | 53 | −0.05 | 0.14 (−0.17, 0.46) | 0.22 | 0.21 |
| Mali | PROMIS CS (32) | | | | | | | | | | | | | |
| Zimbabwe | SHINE (HIV−) (33) | A not B task | 497 | 459 | −0.05 | 0.08 (−0.05, 0.22) | 0.28 | 0.23 | 116 | 93 | 0.03 | −0.22 (−0.50, 0.05) | 0.29 | 0.26 |
| Zimbabwe | SHINE (HIV+) (34) | A not B task | 109 | 84 | 0.06 | −0.07 (−0.36, 0.22) | 0.06 | 0.08 | 31 | 27 | 0.02 | −0.21 (−0.67, 0.24) | 0.11 | 0.12 |
| | | | **1740** | **1604** | | **I² = 0.39, Tau² = 0.01** | | | **441** | **363** | | **I² = 0.21, Tau² = 0.01** | | |
| **Fixed** | | | | | | **0.01 (−0.07, 0.08)** | | | | | | **−0.04 (−0.19, 0.11)** | | |
| **Random** | | | | | | **−0.01 (−0.11, 0.08)** | | | | | | **−0.04 (−0.21, 0.12)** | | |

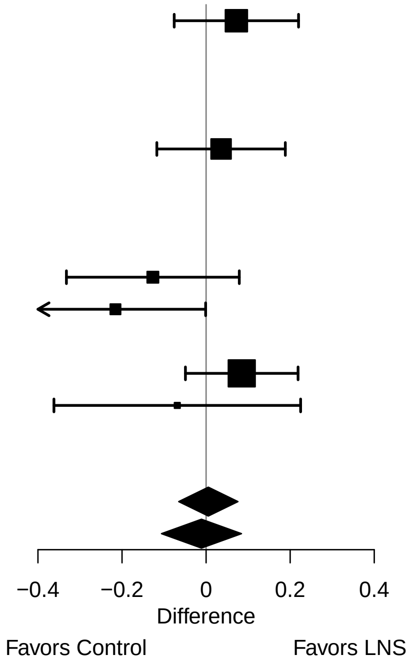

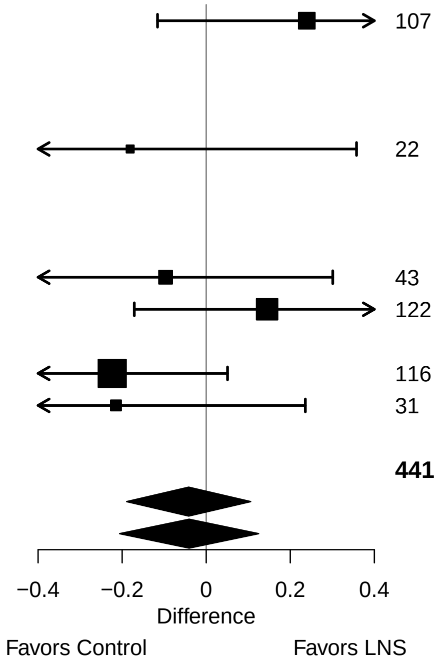

#### Supplemental figure 5L: Mean difference in executive function z-score

#### 5L9: Stratified by Child baseline acute malnutrition

**P−for−interaction = 0.166**
**Difference in MDs = 0.19 (−0.08, 0.47)**

| Country | Trial | Tool | No: LNS N | No: Control N | No: Control Mean | No: MD (95% CI) | No: Fixed W | No: Random W | Yes: LNS N | Yes: Control N | Yes: Control Mean | Yes: MD (95% CI) | Yes: Fixed W | Yes: Random W |
|---|---|---|---|---|---|---|---|---|---|---|---|---|---|---|
| Bangladesh | JiVitA−4 (21) | | | | | | | | | | | | | |
| Bangladesh | RDNS (22) | A not B task | 475 | 238 | −0.04 | 0.10 (−0.02, 0.23) | 0.26 | 0.22 | 36 | 12 | −0.67 | 0.53 (−0.08, 1.15) | 0.18 | 0.18 |
| Bangladesh | WASH−B (23) | A not B task | | | | | | | | | | | | |
| Burkina Faso | iLiNS−Zinc (24) | | | | | | | | | | | | | |
| Ghana | GHANA (25) | | | | | | | | | | | | | |
| Ghana | iLiNS−DYADG (26) | A not B task | 256 | 490 | −0.02 | 0.05 (−0.10, 0.21) | 0.17 | 0.18 | 15 | 47 | 0.18 | −0.41 (−0.97, 0.15) | 0.22 | 0.21 |
| Haiti | HAITI (27) | | | | | | | | | | | | | |
| Kenya | WASH−B (28) | | | | | | | | | | | | | |
| Madagascar | MAHAY (29) | | | | | | | | | | | | | |
| Malawi | iLiNS−DYADM (30) | A not B task | 162 | 332 | 0.04 | −0.14 (−0.33, 0.05) | 0.11 | 0.14 | 10 | 25 | 0.06 | 0.23 (−0.50, 0.95) | 0.13 | 0.14 |
| Malawi | iLiNS−DOSE (31) | A not B task | 453 | 156 | 0.09 | −0.12 (−0.30, 0.06) | 0.12 | 0.15 | 22 | 8 | −0.08 | 0.09 (−0.60, 0.78) | 0.14 | 0.16 |
| Mali | PROMIS CS (32) | | | | | | | | | | | | | |
| Zimbabwe | SHINE (HIV−) (33) | A not B task | 581 | 523 | −0.01 | 0.01 (−0.12, 0.13) | 0.26 | 0.22 | 34 | 33 | −0.33 | 0.28 (−0.21, 0.77) | 0.29 | 0.25 |
| Zimbabwe | SHINE (HIV+) (34) | A not B task | 128 | 103 | 0.12 | −0.18 (−0.43, 0.06) | 0.07 | 0.10 | 14 | 8 | −0.83 | 0.86 (−0.44, 2.16) | 0.04 | 0.05 |
| | | | **2055** | **1842** | | **I² = 0.45, Tau² = 0.01** | | | **131** | **133** | | **I² = 0.27, Tau² = 0.04** | | |
| **Fixed** | | | | | | **−0.01 (−0.07, 0.06)** | | | | | | **0.16 (−0.10, 0.43)** | | |
| **Random** | | | | | | **−0.02 (−0.11, 0.07)** | | | | | | **0.17 (−0.14, 0.48)** | | |

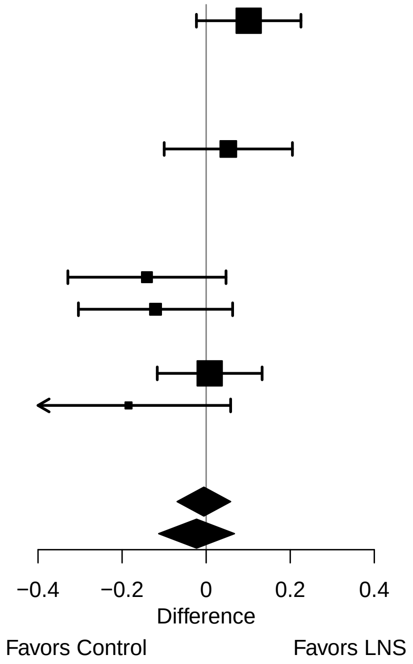

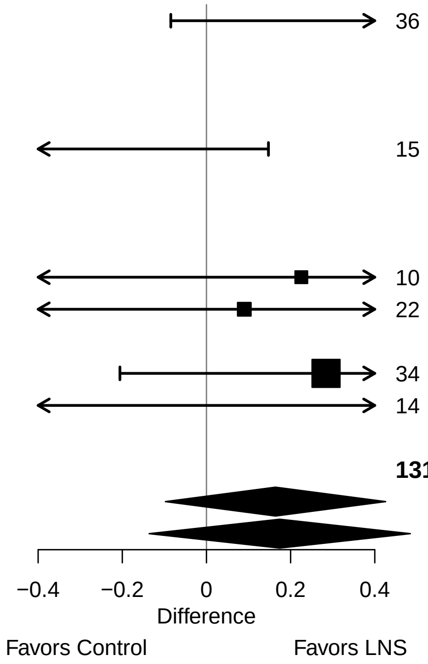

##### Supplemental figure 5L: Mean difference in executive function z-score

#### 5L10: Stratified by Child baseline anemia

**P−for−interaction = 0.567**
**Difference in MDs = 0.05 (−0.12, 0.22)**

| Country | Trial | Tool | Not anemic LNS N | Not anemic Control N | Not anemic Control Mean | Not anemic MD (95% CI) | Not anemic Fixed W | Not anemic Random W | Anemic LNS N | Anemic Control N | Anemic Control Mean | Anemic MD (95% CI) | Anemic Fixed W | Anemic Random W |
|---|---|---|---|---|---|---|---|---|---|---|---|---|---|---|
| Bangladesh | JiVitA−4 (21) | | | | | | | | | | | | | |
| Bangladesh | RDNS (22) | A not B task | 204 | 93 | −0.05 | 0.09 (−0.09, 0.27) | 0.39 | 0.39 | 306 | 153 | −0.10 | 0.15 (−0.04, 0.34) | 0.33 | 0.31 |
| Bangladesh | WASH−B (23) | A not B task | | | | | | | | | | | | |
| Burkina Faso | iLiNS−Zinc (24) | | | | | | | | | | | | | |
| Ghana | GHANA (25) | | | | | | | | | | | | | |
| Ghana | iLiNS−DYADG (26) | A not B task | 172 | 308 | 0.03 | −0.05 (−0.24, 0.14) | 0.33 | 0.33 | 78 | 181 | −0.02 | 0.06 (−0.20, 0.32) | 0.18 | 0.20 |
| Haiti | HAITI (27) | | | | | | | | | | | | | |
| Kenya | WASH−B (28) | | | | | | | | | | | | | |
| Madagascar | MAHAY (29) | | | | | | | | | | | | | |
| Malawi | iLiNS−DYADM (30) | A not B task | 55 | 122 | 0.01 | −0.15 (−0.48, 0.18) | 0.11 | 0.11 | 121 | 234 | 0.04 | −0.06 (−0.28, 0.16) | 0.27 | 0.26 |
| Malawi | iLiNS−DOSE (31) | A not B task | 176 | 70 | 0.05 | −0.06 (−0.33, 0.20) | 0.17 | 0.17 | 296 | 93 | 0.10 | −0.14 (−0.38, 0.10) | 0.22 | 0.23 |
| Mali | PROMIS CS (32) | | | | | | | | | | | | | |
| Zimbabwe | SHINE (HIV−) (33) | A not B task | | | | | | | | | | | | |
| Zimbabwe | SHINE (HIV+) (34) | A not B task | | | | | | | | | | | | |
| | | | **607** | **593** | | **I² = 0.00, Tau² = 0.00** | | | **801** | **661** | | **I² = 0.24, Tau² = 0.00** | | |
| **Fixed** | | | | | | **−0.01 (−0.12, 0.10)** | | | | | | **0.02 (−0.10, 0.13)** | | |
| **Random** | | | | | | **−0.01 (−0.12, 0.10)** | | | | | | **0.01 (−0.12, 0.14)** | | |

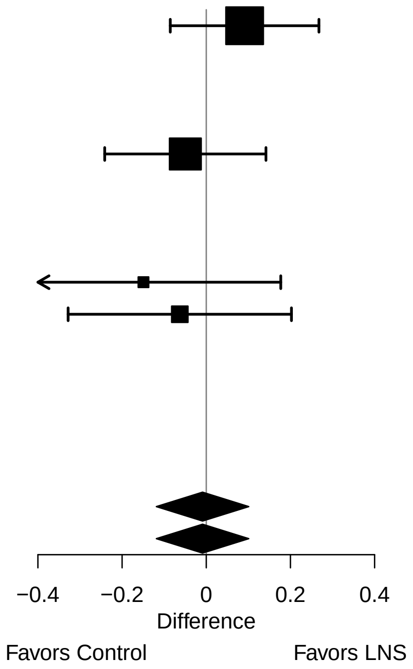

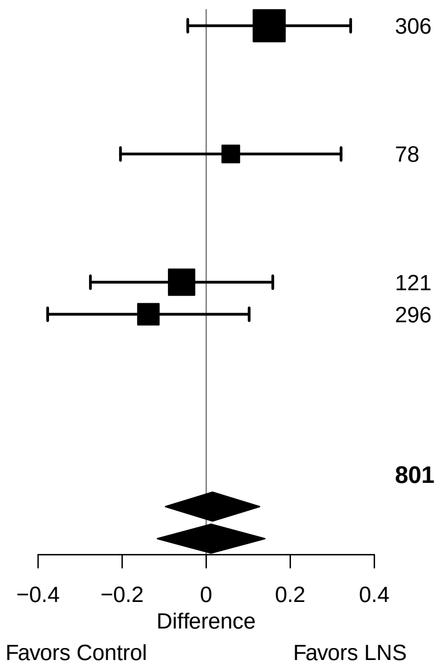

#### Supplemental figure 5M: Executive function lowest decile prevalence ratio

##### 5M1: Stratified by Maternal height (insufficient comparisons)

#### Supplemental figure 5M: Executive function lowest decile prevalence ratio

#### 5M2: Stratified by Maternal BMI

**P−for−interaction = 0.977**
**Ratio of PRs = 1.00 (0.72, 1.40)**

| Country | Trial | Tool | At least 20 kg/m²: LNS N | Control N | Control Prevalence | PR (95% CI) | Fixed W | Random W | Less than 20 kg/m²: LNS N | Control N | Control Prevalence | PR (95% CI) | Fixed W | Random W |
|---|---|---|---|---|---|---|---|---|---|---|---|---|---|---|
| Bangladesh | JiVitA−4 (21) | | | | | | | | | | | | | |
| Bangladesh | RDNS (22) | A not B task | 226 | 103 | 9.7 | 0.87 (0.43, 1.76) | 0.09 | 0.09 | 285 | 153 | 12.4 | 0.85 (0.51, 1.42) | 0.20 | 0.20 |
| Bangladesh | WASH−B (23) | A not B task | 495 | 1517 | 9.6 | 0.92 (0.67, 1.27) | 0.46 | 0.46 | 597 | 1767 | 10.3 | 0.99 (0.75, 1.31) | 0.67 | 0.67 |
| Burkina Faso | iLiNS−Zinc (24) | | | | | | | | | | | | | |
| Ghana | GHANA (25) | | | | | | | | | | | | | |
| Ghana | iLiNS−DYADG (26) | A not B task | | | | | | | | | | | | |
| Haiti | HAITI (27) | | | | | | | | | | | | | |
| Kenya | WASH−B (28) | | | | | | | | | | | | | |
| Madagascar | MAHAY (29) | | | | | | | | | | | | | |
| Malawi | iLiNS−DYADM (30) | A not B task | 108 | 221 | 9.5 | 0.97 (0.48, 2.00) | 0.09 | 0.09 | 74 | 146 | 13.0 | 0.62 (0.26, 1.49) | 0.07 | 0.07 |
| Malawi | iLiNS−DOSE (31) | A not B task | | | | | | | | | | | | |
| Mali | PROMIS CS (32) | | | | | | | | | | | | | |
| Zimbabwe | SHINE (HIV−) (33) | A not B task | 563 | 539 | 11.9 | 0.76 (0.53, 1.09) | 0.36 | 0.36 | 93 | 87 | 11.5 | 0.75 (0.31, 1.80) | 0.07 | 0.07 |
| Zimbabwe | SHINE (HIV+) (34) | A not B task | | | | | | | | | | | | |
| | | | **1392** | **2380** | | **I² = 0.00, Tau² = 0.00** | | | **1049** | **2153** | | **I² = 0.00, Tau² = 0.00** | | |
| **Fixed** | | | | | | **0.86 (0.69, 1.07)** | | | | | | **0.91 (0.73, 1.15)** | | |
| **Random** | | | | | | **0.86 (0.69, 1.07)** | | | | | | **0.91 (0.73, 1.15)** | | |

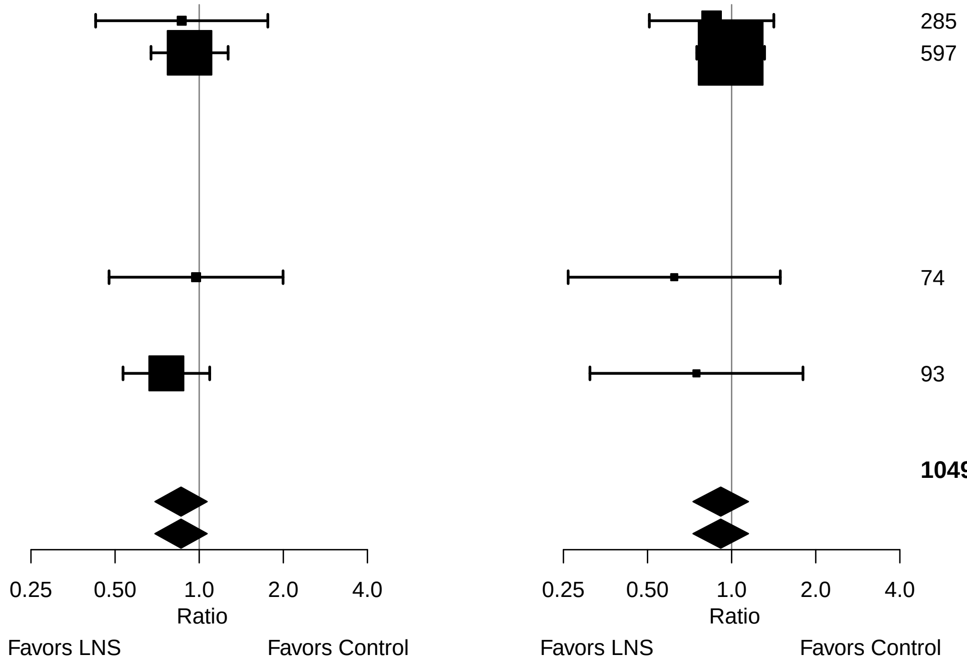

#### Supplemental figure 5M: Executive function lowest decile prevalence ratio

### 5M3: Stratified by Maternal age

**P−for−interaction = 0.088**
**Ratio of PRs = 0.76 (0.55, 1.04)**

| | | | | | | At least 25 y | | | | | | Less than 25 y | | |
|---|---|---|---|---|---|---|---|---|---|---|---|---|---|---|
| **Country** | **Trial** | **Tool** | **LNS N** | **Control N** | **Control Prevalence** | **PR (95% CI)** | **Fixed W** | **Random W** | **LNS N** | **Control N** | **Control Prevalence** | **PR (95% CI)** | **Fixed W** | **Random W** |
| Bangladesh | JiVitA−4 (21) | | | | | | | | | | | | | |
| Bangladesh | RDNS (22) | A not B task | 146 | 80 | 13.8 | 0.75 (0.33, 1.68) | 0.06 | 0.09 | 387 | 186 | 9.7 | 0.96 (0.59, 1.57) | 0.21 | 0.21 |
| Bangladesh | WASH−B (23) | A not B task | 471 | 1469 | 9.7 | 1.21 (0.91, 1.61) | 0.50 | 0.39 | 621 | 1844 | 10.4 | 0.79 (0.56, 1.11) | 0.45 | 0.45 |
| Burkina Faso | iLiNS−Zinc (24) | | | | | | | | | | | | | |
| Ghana | GHANA (25) | | | | | | | | | | | | | |
| Ghana | iLiNS−DYADG (26) | A not B task | 185 | 362 | 12.2 | 0.76 (0.44, 1.29) | 0.14 | 0.18 | 105 | 214 | 9.3 | 0.82 (0.37, 1.79) | 0.08 | 0.08 |
| Haiti | HAITI (27) | | | | | | | | | | | | | |
| Kenya | WASH−B (28) | | | | | | | | | | | | | |
| Madagascar | MAHAY (29) | | | | | | | | | | | | | |
| Malawi | iLiNS−DYADM (30) | A not B task | 89 | 190 | 7.9 | 1.57 (0.75, 3.27) | 0.07 | 0.10 | 93 | 179 | 14.0 | 0.38 (0.15, 0.97) | 0.06 | 0.06 |
| Malawi | iLiNS−DOSE (31) | A not B task | | | | | | | | | | | | |
| Mali | PROMIS CS (32) | | | | | | | | | | | | | |
| Zimbabwe | SHINE (HIV−) (33) | A not B task | 399 | 358 | 11.7 | 0.81 (0.53, 1.25) | 0.22 | 0.24 | 306 | 300 | 11.3 | 0.81 (0.48, 1.35) | 0.20 | 0.20 |
| Zimbabwe | SHINE (HIV+) (34) | A not B task | | | | | | | | | | | | |
| | | | **1290** | **2459** | | **I² = 0.28, Tau² = 0.02** | | | **1512** | **2723** | | **I² = 0.00, Tau² = 0.00** | | |
| **Fixed** | | | | | | **1.02 (0.84, 1.25)** | | | | | | **0.79 (0.63, 1.00)** | | |
| **Random** | | | | | | **1.00 (0.77, 1.28)** | | | | | | **0.79 (0.63, 1.00)** | | |

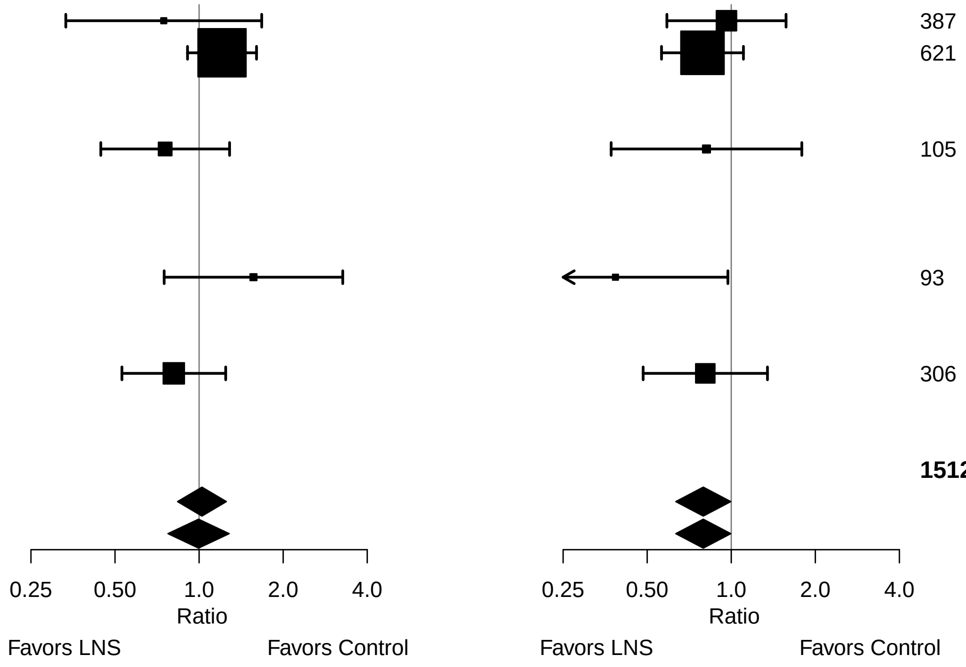

#### Supplemental figure 5M: Executive function lowest decile prevalence ratio

#### 5M4: Stratified by Maternal education

**P−for−interaction = 0.865**
**Ratio of PRs = 1.03 (0.73, 1.46)**

| Country | Trial | Tool | LNS N | Control N | Control Prevalence | Primary or greater PR (95% CI) | Fixed W | Random W | LNS N | Control N | Control Prevalence | Incomplete or no formal PR (95% CI) | Fixed W | Random W |
|---|---|---|---|---|---|---|---|---|---|---|---|---|---|---|
| Bangladesh | JiVitA−4 (21) | | | | | | | | | | | | | |
| Bangladesh | RDNS (22) | A not B task | 394 | 191 | 9.4 | 0.78 (0.44, 1.38) | 0.15 | 0.15 | 139 | 75 | 14.7 | 1.08 (0.61, 1.91) | 0.20 | 0.20 |
| Bangladesh | WASH−B (23) | A not B task | 776 | 2386 | 9.3 | 0.91 (0.69, 1.20) | 0.64 | 0.64 | 321 | 941 | 12.1 | 1.03 (0.73, 1.46) | 0.54 | 0.54 |
| Burkina Faso | iLiNS−Zinc (24) | | | | | | | | | | | | | |
| Ghana | GHANA (25) | | | | | | | | | | | | | |
| Ghana | iLiNS−DYADG (26) | A not B task | 223 | 449 | 10.0 | 0.81 (0.48, 1.36) | 0.18 | 0.18 | 67 | 127 | 15.0 | 0.70 (0.31, 1.58) | 0.10 | 0.10 |
| Haiti | HAITI (27) | | | | | | | | | | | | | |
| Kenya | WASH−B (28) | | | | | | | | | | | | | |
| Madagascar | MAHAY (29) | | | | | | | | | | | | | |
| Malawi | iLiNS−DYADM (30) | A not B task | 26 | 59 | 10.2 | 1.89 (0.63, 5.64) | 0.04 | 0.04 | 155 | 307 | 11.1 | 0.64 (0.33, 1.23) | 0.16 | 0.16 |
| Malawi | iLiNS−DOSE (31) | A not B task | | | | | | | | | | | | |
| Mali | PROMIS CS (32) | | | | | | | | | | | | | |
| Zimbabwe | SHINE (HIV−) (33) | A not B task | | | | | | | | | | | | |
| Zimbabwe | SHINE (HIV+) (34) | A not B task | | | | | | | | | | | | |
| | | | **1419** | **3085** | | **I² = 0.00, Tau² = 0.00** | | | **682** | **1450** | | **I² = 0.00, Tau² = 0.00** | | |
| **Fixed** | | | | | | **0.90 (0.72, 1.12)** | | | | | | **0.93 (0.72, 1.20)** | | |
| **Random** | | | | | | **0.90 (0.72, 1.12)** | | | | | | **0.93 (0.72, 1.20)** | | |

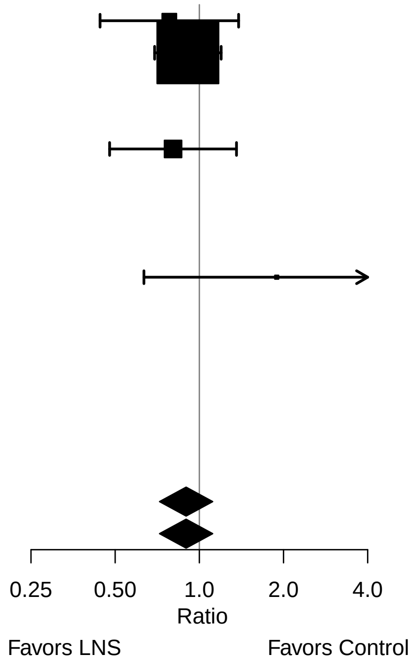

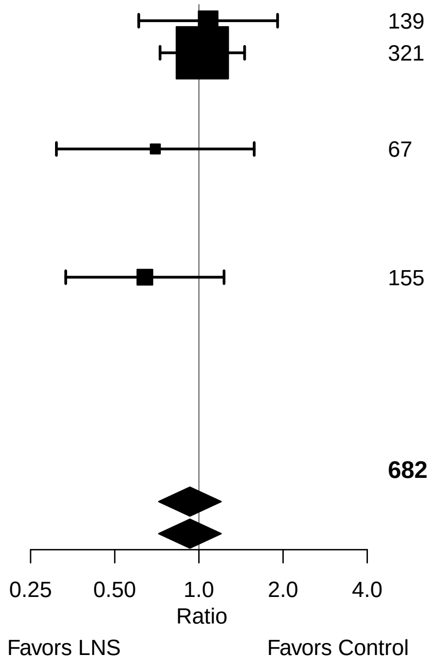

##### Supplemental figure 5M: Executive function lowest decile prevalence ratio

#### 5M5: Stratified by Maternal depressive symptoms

**P−for−interaction = 0.394**
**Ratio of PRs = 1.18 (0.80, 1.75)**

| | | | | | | Less than 75th percentile | | | | | | At least 75th percentile | | |
|---|---|---|---|---|---|---|---|---|---|---|---|---|---|---|
| **Country** | **Trial** | **Tool** | **LNS N** | **Control N** | **Control Prevalence** | **PR (95% CI)** | **Fixed W** | **Random W** | **LNS N** | **Control N** | **Control Prevalence** | **PR (95% CI)** | **Fixed W** | **Random W** |
| Bangladesh | JiVitA−4 (21) | | | | | | | | | | | | | |
| Bangladesh | RDNS (22) | A not B task | 345 | 150 | 10.0 | 0.93 (0.56, 1.55) | 0.14 | 0.14 | 166 | 100 | 12.0 | 0.80 (0.34, 1.89) | 0.15 | 0.17 |
| Bangladesh | WASH−B (23) | A not B task | 847 | 2392 | 9.4 | 0.97 (0.75, 1.26) | 0.56 | 0.56 | 230 | 852 | 12.1 | 0.97 (0.64, 1.48) | 0.62 | 0.57 |
| Burkina Faso | iLiNS−Zinc (24) | | | | | | | | | | | | | |
| Ghana | GHANA (25) | | | | | | | | | | | | | |
| Ghana | iLiNS−DYADG (26) | A not B task | | | | | | | | | | | | |
| Haiti | HAITI (27) | | | | | | | | | | | | | |
| Kenya | WASH−B (28) | | | | | | | | | | | | | |
| Madagascar | MAHAY (29) | | | | | | | | | | | | | |
| Malawi | iLiNS−DYADM (30) | A not B task | | | | | | | | | | | | |
| Malawi | iLiNS−DOSE (31) | A not B task | | | | | | | | | | | | |
| Mali | PROMIS CS (32) | | | | | | | | | | | | | |
| Zimbabwe | SHINE (HIV−) (33) | A not B task | 542 | 520 | 11.5 | 0.90 (0.63, 1.28) | 0.29 | 0.29 | 186 | 164 | 11.6 | 0.51 (0.26, 1.02) | 0.23 | 0.26 |
| Zimbabwe | SHINE (HIV+) (34) | A not B task | | | | | | | | | | | | |
| | | | **1734** | **3062** | | **I² = 0.00, Tau² = 0.00** | | | **582** | **1116** | | **I² = 0.18, Tau² = 0.02** | | |
| **Fixed** | | | | | | **0.94 (0.78, 1.15)** | | | | | | **0.81 (0.58, 1.13)** | | |
| **Random** | | | | | | **0.94 (0.78, 1.15)** | | | | | | **0.79 (0.55, 1.16)** | | |

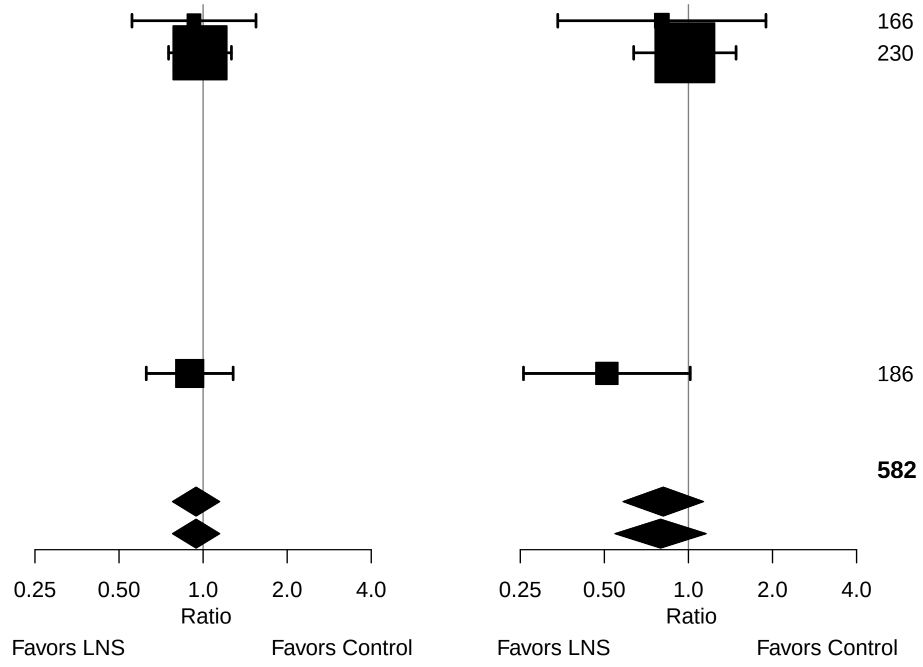

#### Supplemental figure 5M: Executive function lowest decile prevalence ratio

### 5M6: Stratified by Child sex

**P−for−interaction = 0.231**
**Ratio of PRs = 1.19 (0.90, 1.57)**

| Country | Trial | Tool | Male LNS N | Male Control N | Male Control Prevalence | Male PR (95% CI) | Male Fixed W | Male Random W | Female LNS N | Female Control N | Female Control Prevalence | Female PR (95% CI) | Female Fixed W | Female Random W |
|---|---|---|---|---|---|---|---|---|---|---|---|---|---|---|
| Bangladesh | JiVitA−4 (21) | | | | | | | | | | | | | |
| Bangladesh | RDNS (22) | A not B task | 261 | 133 | 6.8 | 1.42 (0.68, 2.94) | 0.09 | 0.14 | 272 | 133 | 15.0 | 0.64 (0.41, 0.98) | 0.16 | 0.16 |
| Bangladesh | WASH−B (23) | A not B task | 540 | 1671 | 9.8 | 0.87 (0.64, 1.20) | 0.51 | 0.31 | 557 | 1656 | 10.5 | 1.03 (0.78, 1.34) | 0.42 | 0.42 |
| Burkina Faso | iLiNS−Zinc (24) | | | | | | | | | | | | | |
| Ghana | GHANA (25) | | | | | | | | | | | | | |
| Ghana | iLiNS−DYADG (26) | A not B task | 145 | 273 | 11.0 | 0.63 (0.32, 1.25) | 0.11 | 0.15 | 145 | 303 | 11.2 | 0.92 (0.52, 1.64) | 0.09 | 0.09 |
| Haiti | HAITI (27) | | | | | | | | | | | | | |
| Kenya | WASH−B (28) | | | | | | | | | | | | | |
| Madagascar | MAHAY (29) | | | | | | | | | | | | | |
| Malawi | iLiNS−DYADM (30) | A not B task | 92 | 176 | 12.5 | 0.61 (0.27, 1.37) | 0.08 | 0.12 | 90 | 193 | 9.3 | 1.07 (0.50, 2.29) | 0.05 | 0.05 |
| Malawi | iLiNS−DOSE (31) | A not B task | 235 | 93 | 7.5 | 1.41 (0.63, 3.15) | 0.08 | 0.12 | 240 | 71 | 7.0 | 1.60 (0.64, 4.00) | 0.04 | 0.04 |
| Mali | PROMIS CS (32) | | | | | | | | | | | | | |
| Zimbabwe | SHINE (HIV−) (33) | A not B task | 399 | 364 | 8.8 | 0.51 (0.28, 0.94) | 0.14 | 0.17 | 388 | 375 | 14.1 | 0.98 (0.68, 1.42) | 0.23 | 0.23 |
| Zimbabwe | SHINE (HIV+) (34) | A not B task | | | | | | | | | | | | |
| | | | **1672** | **2710** | | **I² = 0.33, Tau² = 0.06** | | | **1692** | **2731** | | **I² = 0.00, Tau² = 0.00** | | |
| **Fixed** | | | | | | **0.83 (0.66, 1.04)** | | | | | | **0.95 (0.80, 1.13)** | | |
| **Random** | | | | | | **0.82 (0.60, 1.14)** | | | | | | **0.95 (0.80, 1.13)** | | |

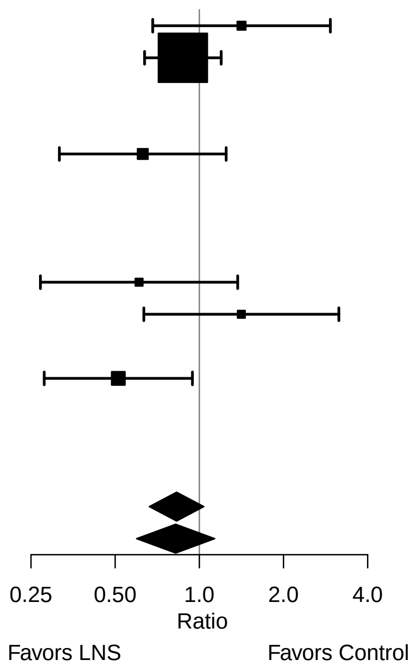

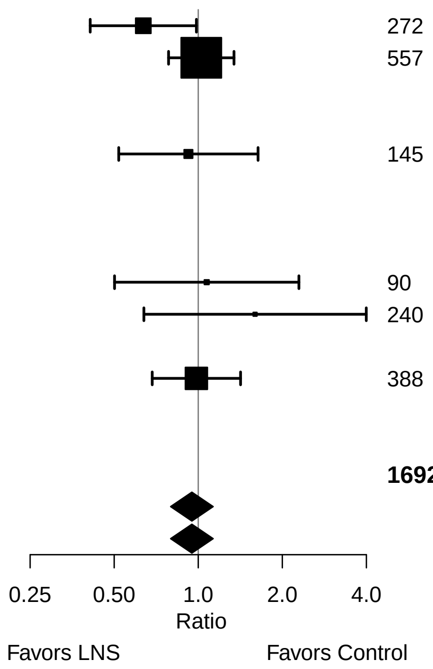

#### Supplemental figure 5M: Executive function lowest decile prevalence ratio

#### 5M7: Stratified by Child birth order

**P−for−interaction = 0.144**
**Ratio of PRs = 0.78 (0.56, 1.09)**

| Country | Trial | Tool | LNS N | Control N | Control Prevalence | Later born PR (95% CI) | Fixed W | Random W | LNS N | Control N | Control Prevalence | Firstborn PR (95% CI) | Fixed W | Random W |
|---|---|---|---|---|---|---|---|---|---|---|---|---|---|---|
| Bangladesh | JiVitA−4 (21) | | | | | | | | | | | | | |
| Bangladesh | RDNS (22) | A not B task | 320 | 163 | 12.9 | 0.68 (0.41, 1.12) | 0.12 | 0.15 | 213 | 103 | 7.8 | 1.39 (0.64, 3.00) | 0.15 | 0.16 |
| Bangladesh | WASH−B (23) | A not B task | 683 | 2174 | 9.5 | 1.14 (0.89, 1.45) | 0.51 | 0.43 | 394 | 1049 | 10.3 | 0.71 (0.47, 1.10) | 0.49 | 0.46 |
| Burkina Faso | iLiNS−Zinc (24) | | | | | | | | | | | | | |
| Ghana | GHANA (25) | | | | | | | | | | | | | |
| Ghana | iLiNS−DYADG (26) | A not B task | 195 | 390 | 10.8 | 0.90 (0.54, 1.51) | 0.11 | 0.14 | 95 | 186 | 11.8 | 0.53 (0.22, 1.27) | 0.12 | 0.13 |
| Haiti | HAITI (27) | | | | | | | | | | | | | |
| Kenya | WASH−B (28) | | | | | | | | | | | | | |
| Madagascar | MAHAY (29) | | | | | | | | | | | | | |
| Malawi | iLiNS−DYADM (30) | A not B task | | | | | | | | | | | | |
| Malawi | iLiNS−DOSE (31) | A not B task | | | | | | | | | | | | |
| Mali | PROMIS CS (32) | | | | | | | | | | | | | |
| Zimbabwe | SHINE (HIV−) (33) | A not B task | 568 | 530 | 11.7 | 0.84 (0.60, 1.19) | 0.26 | 0.28 | 186 | 178 | 11.8 | 0.68 (0.37, 1.25) | 0.24 | 0.25 |
| Zimbabwe | SHINE (HIV+) (34) | A not B task | | | | | | | | | | | | |
| | | | **1766** | **3257** | | **I² = 0.32, Tau² = 0.01** | | | **888** | **1516** | | **I² = 0.06, Tau² = 0.01** | | |
| **Fixed** | | | | | | **0.97 (0.81, 1.15)** | | | | | | **0.75 (0.56, 1.02)** | | |
| **Random** | | | | | | **0.94 (0.76, 1.16)** | | | | | | **0.76 (0.55, 1.04)** | | |

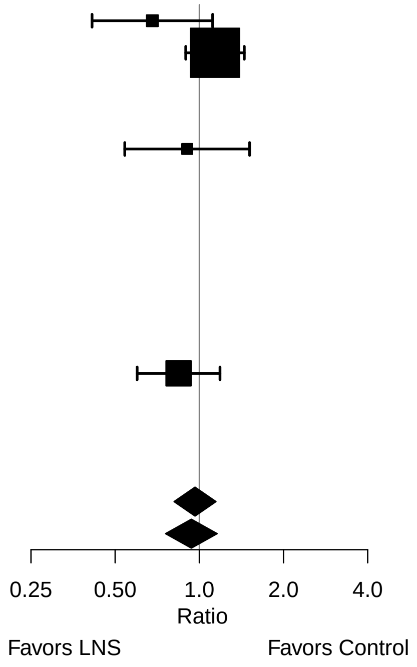

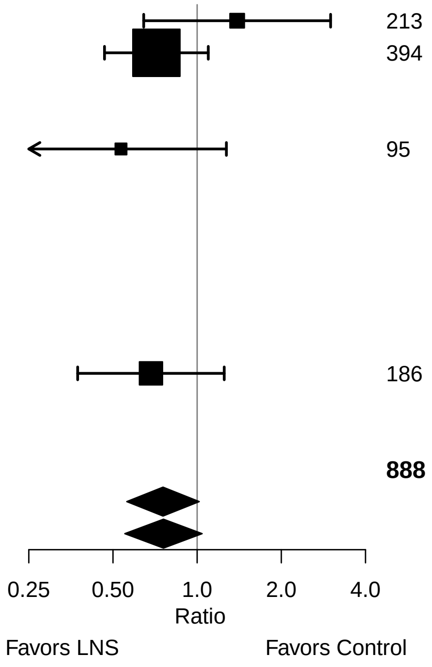

#### Supplemental figure 5M: Executive function lowest decile prevalence ratio

##### 5M8: Stratified by Child baseline stunting

**P−for−interaction = 0.507**
**Ratio of PRs = 1.23 (0.67, 2.28)**

| Country | Trial | Tool | LNS N | Control N | Control Prevalence | No PR (95% CI) | Fixed W | Random W | LNS N | Control N | Control Prevalence | Yes PR (95% CI) | Fixed W | Random W |
|---|---|---|---|---|---|---|---|---|---|---|---|---|---|---|
| Bangladesh | JiVitA−4 (21) | | | | | | | | | | | | | |
| Bangladesh | RDNS (22) | A not B task | 404 | 187 | 10.7 | 0.83 (0.52, 1.34) | 0.36 | 0.36 | 107 | 62 | 11.3 | 0.99 (0.34, 2.90) | 0.24 | 0.24 |
| Bangladesh | WASH−B (23) | A not B task | | | | | | | | | | | | |
| Burkina Faso | iLiNS−Zinc (24) | | | | | | | | | | | | | |
| Ghana | GHANA (25) | | | | | | | | | | | | | |
| Ghana | iLiNS−DYADG (26) | A not B task | | | | | | | | | | | | |
| Haiti | HAITI (27) | | | | | | | | | | | | | |
| Kenya | WASH−B (28) | | | | | | | | | | | | | |
| Madagascar | MAHAY (29) | | | | | | | | | | | | | |
| Malawi | iLiNS−DYADM (30) | A not B task | | | | | | | | | | | | |
| Malawi | iLiNS−DOSE (31) | A not B task | 353 | 111 | 5.4 | 2.10 (0.91, 4.81) | 0.12 | 0.25 | 122 | 53 | 11.3 | 0.87 (0.34, 2.19) | 0.33 | 0.33 |
| Mali | PROMIS CS (32) | | | | | | | | | | | | | |
| Zimbabwe | SHINE (HIV−) (33) | A not B task | 497 | 459 | 11.5 | 0.71 (0.48, 1.06) | 0.53 | 0.39 | 116 | 93 | 9.7 | 1.51 (0.68, 3.38) | 0.43 | 0.43 |
| Zimbabwe | SHINE (HIV+) (34) | A not B task | | | | | | | | | | | | |
| | | | **1254** | **757** | | **I² = 0.62, Tau² = 0.21** | | | **345** | **208** | | **I² = 0.00, Tau² = 0.00** | | |
| **Fixed** | | | | | | **0.86 (0.64, 1.13)** | | | | | | **1.14 (0.67, 1.93)** | | |
| **Random** | | | | | | **0.99 (0.54, 1.81)** | | | | | | **1.14 (0.67, 1.93)** | | |

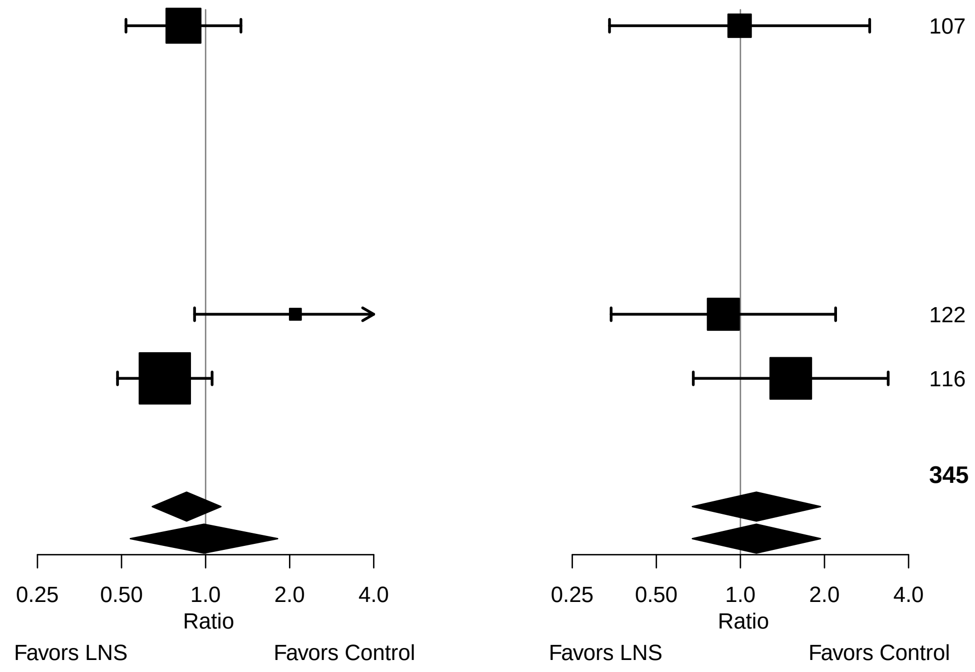

#### Supplemental figure 5M: Executive function lowest decile prevalence ratio

### 5M9: Stratified by Child baseline acute malnutrition (insufficient comparisons)

#### Supplemental figure 5M: Executive function lowest decile prevalence ratio

### 5M10: Stratified by Child baseline anemia

**P−for−interaction = 0.464**
**Ratio of PRs = 0.81 (0.47, 1.41)**

| Country | Trial | Tool | LNS N | Control N | Control Prevalence | Not anemic PR (95% CI) | Fixed W | Random W | LNS N | Control N | Control Prevalence | Anemic PR (95% CI) | Fixed W | Random W |
|---|---|---|---|---|---|---|---|---|---|---|---|---|---|---|
| Bangladesh | JiVitA−4 (21) | | | | | | | | | | | | | |
| Bangladesh | RDNS (22) | A not B task | 204 | 93 | 7.5 | 1.24 (0.65, 2.37) | 0.29 | 0.29 | 306 | 153 | 13.1 | 0.73 (0.40, 1.32) | 0.37 | 0.32 |
| Bangladesh | WASH−B (23) | A not B task | | | | | | | | | | | | |
| Burkina Faso | iLiNS−Zinc (24) | | | | | | | | | | | | | |
| Ghana | GHANA (25) | | | | | | | | | | | | | |
| Ghana | iLiNS−DYADG (26) | A not B task | 172 | 308 | 10.1 | 1.10 (0.64, 1.88) | 0.42 | 0.42 | 78 | 181 | 12.7 | 0.50 (0.20, 1.28) | 0.15 | 0.19 |
| Haiti | HAITI (27) | | | | | | | | | | | | | |
| Kenya | WASH−B (28) | | | | | | | | | | | | | |
| Madagascar | MAHAY (29) | | | | | | | | | | | | | |
| Malawi | iLiNS−DYADM (30) | A not B task | 55 | 122 | 13.1 | 0.69 (0.27, 1.80) | 0.14 | 0.14 | 121 | 234 | 9.4 | 0.97 (0.49, 1.93) | 0.28 | 0.27 |
| Malawi | iLiNS−DOSE (31) | A not B task | 176 | 70 | 8.6 | 1.13 (0.46, 2.74) | 0.16 | 0.16 | 296 | 93 | 6.5 | 1.78 (0.77, 4.11) | 0.19 | 0.22 |
| Mali | PROMIS CS (32) | | | | | | | | | | | | | |
| Zimbabwe | SHINE (HIV−) (33) | A not B task | | | | | | | | | | | | |
| Zimbabwe | SHINE (HIV+) (34) | A not B task | | | | | | | | | | | | |
| | | | **607** | **593** | | **I² = 0.00, Tau² = 0.00** | | | **801** | **661** | | **I² = 0.34, Tau² = 0.09** | | |
| **Fixed** | | | | | | **1.07 (0.76, 1.52)** | | | | | | **0.88 (0.61, 1.27)** | | |
| **Random** | | | | | | **1.07 (0.76, 1.52)** | | | | | | **0.89 (0.55, 1.44)** | | |

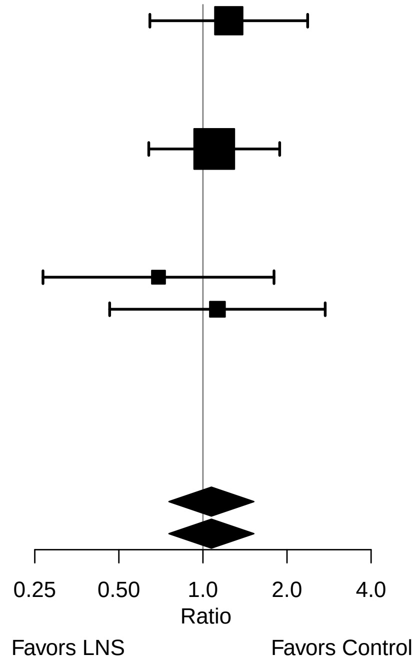

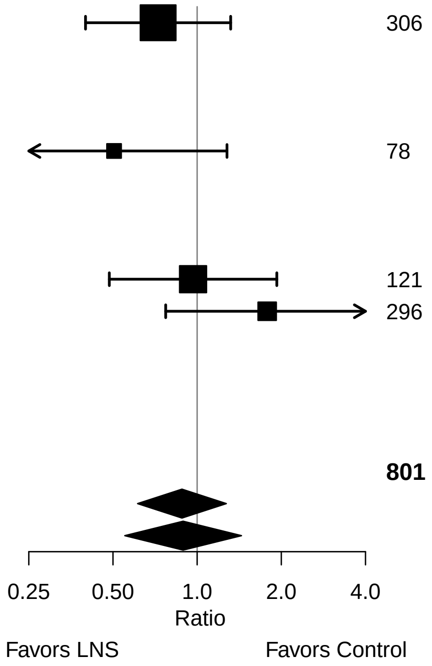

### Supplemental figure 5N: Executive function lowest decile prevalence difference

#### 5N1: Stratified by Maternal height (insufficient comparisons)

#### Supplemental figure 5N: Executive function lowest decile prevalence difference

#### 5N2: Stratified by Maternal BMI

**P−for−interaction = 0.955**
**Difference in PDs = 0.00 (−0.03, 0.03)**

| | | | At least 20 kg/m² | | | | | | Less than 20 kg/m² | | | | | |
|---|---|---|---|---|---|---|---|---|---|---|---|---|---|---|
| Country | Trial | Tool | LNS N | Control N | Control Prevalence | PD (95% CI) | Fixed W | Random W | LNS N | Control N | Control Prevalence | PD (95% CI) | Fixed W | Random W |
| Bangladesh | JiVitA−4 (21) | | | | | | | | | | | | | |
| Bangladesh | RDNS (22) | A not B task | 226 | 103 | 9.7 | −0.01 (−0.08, 0.05) | 0.10 | 0.10 | 285 | 153 | 12.4 | −0.02 (−0.08, 0.04) | 0.16 | 0.16 |
| Bangladesh | WASH−B (23) | A not B task | 495 | 1517 | 9.6 | −0.01 (−0.04, 0.02) | 0.51 | 0.51 | 597 | 1767 | 10.3 | 0.00 (−0.03, 0.03) | 0.70 | 0.70 |
| Burkina Faso | iLiNS−Zinc (24) | | | | | | | | | | | | | |
| Ghana | GHANA (25) | | | | | | | | | | | | | |
| Ghana | iLiNS−DYADG (26) | A not B task | | | | | | | | | | | | |
| Haiti | HAITI (27) | | | | | | | | | | | | | |
| Kenya | WASH−B (28) | | | | | | | | | | | | | |
| Madagascar | MAHAY (29) | | | | | | | | | | | | | |
| Malawi | iLiNS−DYADM (30) | A not B task | 108 | 221 | 9.5 | 0.00 (−0.07, 0.06) | 0.09 | 0.09 | 74 | 146 | 13.0 | −0.05 (−0.14, 0.04) | 0.07 | 0.07 |
| Malawi | iLiNS−DOSE (31) | A not B task | | | | | | | | | | | | |
| Mali | PROMIS CS (32) | | | | | | | | | | | | | |
| Zimbabwe | SHINE (HIV−) (33) | A not B task | 563 | 539 | 11.9 | −0.03 (−0.07, 0.01) | 0.29 | 0.29 | 93 | 87 | 11.5 | −0.03 (−0.12, 0.06) | 0.07 | 0.07 |
| Zimbabwe | SHINE (HIV+) (34) | A not B task | | | | | | | | | | | | |
| | | | **1392** | **2380** | | **I² = 0.00, Tau² = 0.00** | | | **1049** | **2153** | | **I² = 0.00, Tau² = 0.00** | | |
| **Fixed** | | | | | | **−0.01 (−0.03, 0.01)** | | | | | | **−0.01 (−0.03, 0.01)** | | |
| **Random** | | | | | | **−0.01 (−0.03, 0.01)** | | | | | | **−0.01 (−0.03, 0.01)** | | |

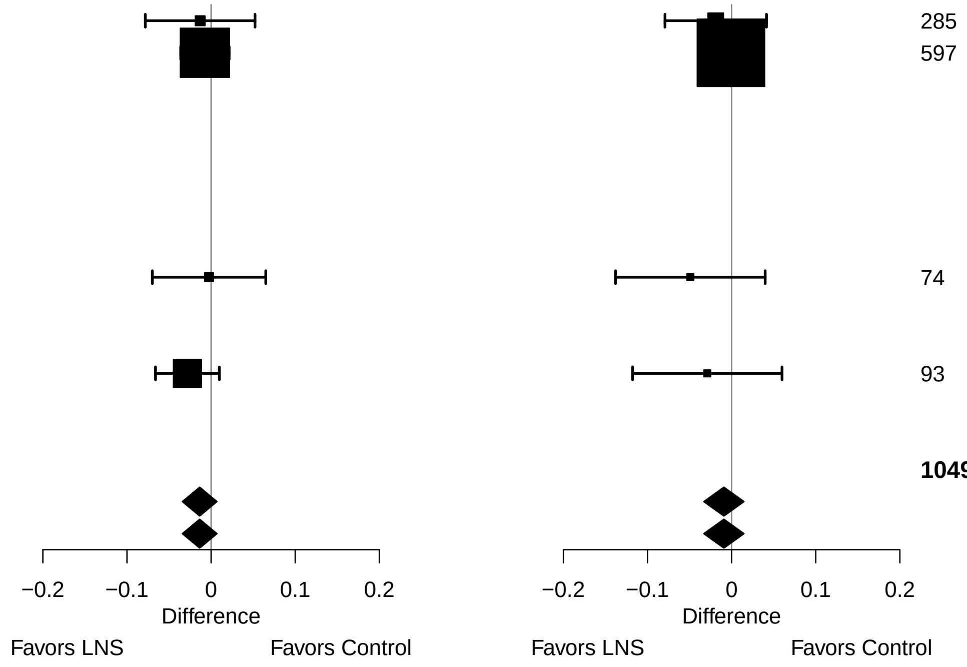

#### Supplemental figure 5N: Executive function lowest decile prevalence difference

##### 5N3: Stratified by Maternal age

**P−for−interaction = 0.066**
**Difference in PDs = −0.03 (−0.06, 0.00)**

| Country | Trial | Tool | LNS N | Control N | Control Prevalence | At least 25 y PD (95% CI) | Fixed W | Random W | LNS N | Control N | Control Prevalence | Less than 25 y PD (95% CI) | Fixed W | Random W |
|---|---|---|---|---|---|---|---|---|---|---|---|---|---|---|
| Bangladesh | JiVitA−4 (21) | | | | | | | | | | | | | |
| Bangladesh | RDNS (22) | A not B task | 146 | 80 | 13.8 | −0.03 (−0.14, 0.07) | 0.05 | 0.06 | 387 | 186 | 9.7 | 0.00 (−0.05, 0.04) | 0.19 | 0.19 |
| Bangladesh | WASH−B (23) | A not B task | 471 | 1469 | 9.7 | 0.02 (−0.01, 0.05) | 0.48 | 0.40 | 621 | 1844 | 10.4 | −0.02 (−0.05, 0.01) | 0.48 | 0.48 |
| Burkina Faso | iLiNS−Zinc (24) | | | | | | | | | | | | | |
| Ghana | GHANA (25) | | | | | | | | | | | | | |
| Ghana | iLiNS−DYADG (26) | A not B task | 185 | 362 | 12.2 | −0.03 (−0.09, 0.03) | 0.16 | 0.18 | 105 | 214 | 9.3 | −0.02 (−0.08, 0.05) | 0.10 | 0.10 |
| Haiti | HAITI (27) | | | | | | | | | | | | | |
| Kenya | WASH−B (28) | | | | | | | | | | | | | |
| Madagascar | MAHAY (29) | | | | | | | | | | | | | |
| Malawi | iLiNS−DYADM (30) | A not B task | 89 | 190 | 7.9 | 0.04 (−0.03, 0.12) | 0.09 | 0.11 | 93 | 179 | 14.0 | −0.09 (−0.16, −0.01) | 0.07 | 0.07 |
| Malawi | iLiNS−DOSE (31) | A not B task | | | | | | | | | | | | |
| Mali | PROMIS CS (32) | | | | | | | | | | | | | |
| Zimbabwe | SHINE (HIV−) (33) | A not B task | 399 | 358 | 11.7 | −0.02 (−0.07, 0.02) | 0.23 | 0.24 | 306 | 300 | 11.3 | −0.02 (−0.07, 0.03) | 0.16 | 0.16 |
| Zimbabwe | SHINE (HIV+) (34) | A not B task | | | | | | | | | | | | |
| | | | **1290** | **2459** | | **I² = 0.25, Tau² = 0.00** | | | **1512** | **2723** | | **I² = 0.00, Tau² = 0.00** | | |
| **Fixed** | | | | | | **0.00 (−0.02, 0.02)** | | | | | | **−0.02 (−0.04, 0.00)** | | |
| **Random** | | | | | | **0.00 (−0.03, 0.03)** | | | | | | **−0.02 (−0.04, 0.00)** | | |

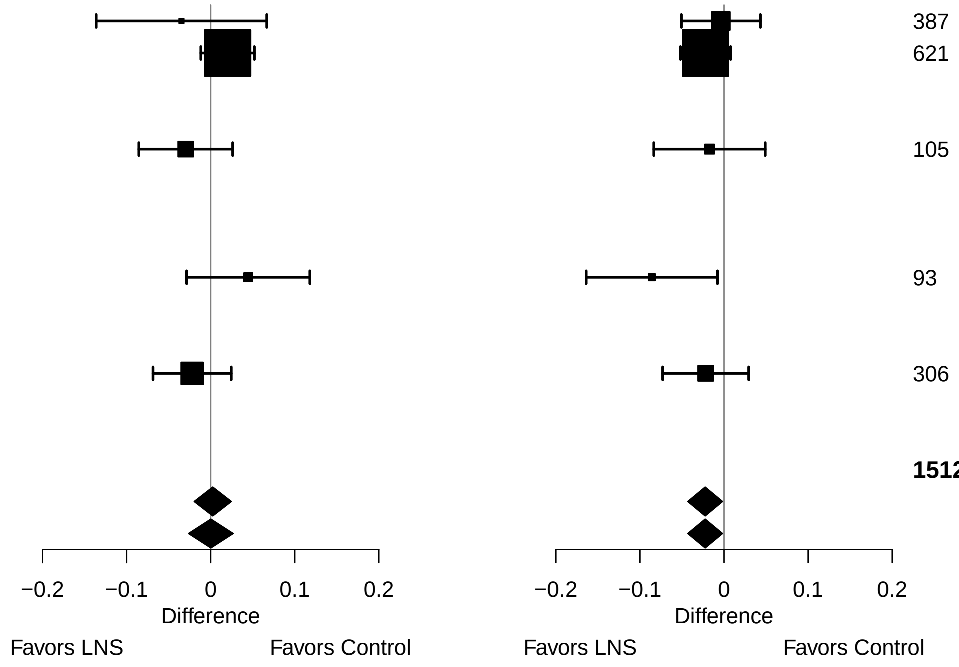

#### Supplemental figure 5N: Executive function lowest decile prevalence difference

##### 5N4: Stratified by Maternal education

**P−for−interaction = 0.909**
**Difference in PDs = 0.00 (−0.04, 0.04)**

| Country | Trial | Tool | Primary or greater: LNS N | Control N | Control Prevalence | PD (95% CI) | Fixed W | Random W | Incomplete or no formal: LNS N | Control N | Control Prevalence | PD (95% CI) | Fixed W | Random W |
|---|---|---|---|---|---|---|---|---|---|---|---|---|---|---|
| Bangladesh | JiVitA−4 (21) | | | | | | | | | | | | | |
| Bangladesh | RDNS (22) | A not B task | 394 | 191 | 9.4 | −0.02 (−0.07, 0.03) | 0.16 | 0.16 | 139 | 75 | 14.7 | 0.01 (−0.07, 0.10) | 0.13 | 0.13 |
| Bangladesh | WASH−B (23) | A not B task | 776 | 2386 | 9.3 | −0.01 (−0.03, 0.02) | 0.65 | 0.65 | 321 | 941 | 12.1 | 0.00 (−0.04, 0.05) | 0.50 | 0.50 |
| Burkina Faso | iLiNS−Zinc (24) | | | | | | | | | | | | | |
| Ghana | GHANA (25) | | | | | | | | | | | | | |
| Ghana | iLiNS−DYADG (26) | A not B task | 223 | 449 | 10.0 | −0.02 (−0.07, 0.03) | 0.17 | 0.17 | 67 | 127 | 15.0 | −0.05 (−0.15, 0.06) | 0.09 | 0.09 |
| Haiti | HAITI (27) | | | | | | | | | | | | | |
| Kenya | WASH−B (28) | | | | | | | | | | | | | |
| Madagascar | MAHAY (29) | | | | | | | | | | | | | |
| Malawi | iLiNS−DYADM (30) | A not B task | 26 | 59 | 10.2 | 0.09 (−0.06, 0.25) | 0.02 | 0.02 | 155 | 307 | 11.1 | −0.04 (−0.10, 0.02) | 0.28 | 0.28 |
| Malawi | iLiNS−DOSE (31) | A not B task | | | | | | | | | | | | |
| Mali | PROMIS CS (32) | | | | | | | | | | | | | |
| Zimbabwe | SHINE (HIV−) (33) | A not B task | | | | | | | | | | | | |
| Zimbabwe | SHINE (HIV+) (34) | A not B task | | | | | | | | | | | | |
| | | | **1419** | **3085** | | **I² = 0.00, Tau² = 0.00** | | | **682** | **1450** | | **I² = 0.00, Tau² = 0.00** | | |
| **Fixed** | | | | | | **−0.01 (−0.03, 0.01)** | | | | | | **−0.01 (−0.04, 0.02)** | | |
| **Random** | | | | | | **−0.01 (−0.03, 0.01)** | | | | | | **−0.01 (−0.04, 0.02)** | | |

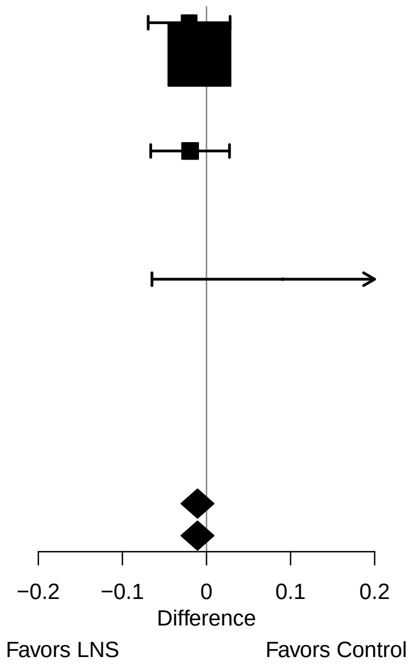

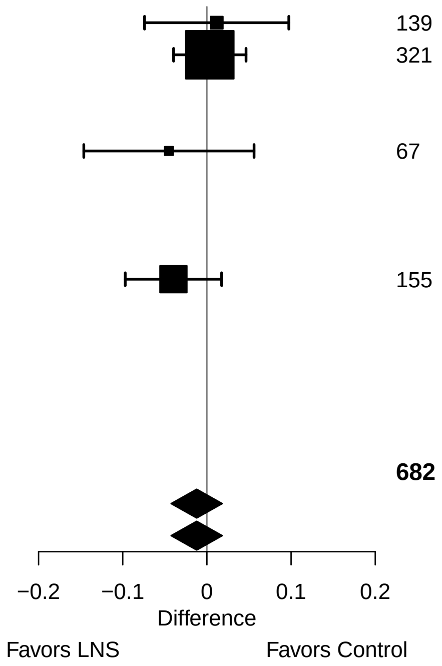

#### Supplemental figure 5N: Executive function lowest decile prevalence difference

##### 5N5: Stratified by Maternal depressive symptoms

**P−for−interaction = 0.401**
**Difference in PDs = 0.02 (−0.02, 0.06)**

| Country | Trial | Tool | LNS N | Control N | Control Prevalence | Less than 75th percentile PD (95% CI) | Fixed W | Random W | LNS N | Control N | Control Prevalence | At least 75th percentile PD (95% CI) | Fixed W | Random W |
|---|---|---|---|---|---|---|---|---|---|---|---|---|---|---|
| Bangladesh | JiVitA−4 (21) | | | | | | | | | | | | | |
| Bangladesh | RDNS (22) | A not B task | 345 | 150 | 10.0 | −0.01 (−0.06, 0.04) | 0.15 | 0.15 | 166 | 100 | 12.0 | −0.02 (−0.12, 0.07) | 0.13 | 0.13 |
| Bangladesh | WASH−B (23) | A not B task | 847 | 2392 | 9.4 | 0.00 (−0.03, 0.02) | 0.62 | 0.62 | 230 | 852 | 12.1 | 0.00 (−0.05, 0.05) | 0.49 | 0.49 |
| Burkina Faso | iLiNS−Zinc (24) | | | | | | | | | | | | | |
| Ghana | GHANA (25) | | | | | | | | | | | | | |
| Ghana | iLiNS−DYADG (26) | A not B task | | | | | | | | | | | | |
| Haiti | HAITI (27) | | | | | | | | | | | | | |
| Kenya | WASH−B (28) | | | | | | | | | | | | | |
| Madagascar | MAHAY (29) | | | | | | | | | | | | | |
| Malawi | iLiNS−DYADM (30) | A not B task | | | | | | | | | | | | |
| Malawi | iLiNS−DOSE (31) | A not B task | | | | | | | | | | | | |
| Mali | PROMIS CS (32) | | | | | | | | | | | | | |
| Zimbabwe | SHINE (HIV−) (33) | A not B task | 542 | 520 | 11.5 | −0.01 (−0.05, 0.03) | 0.23 | 0.23 | 186 | 164 | 11.6 | −0.06 (−0.11, 0.00) | 0.38 | 0.38 |
| Zimbabwe | SHINE (HIV+) (34) | A not B task | | | | | | | | | | | | |
| | | | **1734** | **3062** | | **I² = 0.00, Tau² = 0.00** | | | **582** | **1116** | | **I² = 0.00, Tau² = 0.00** | | |
| **Fixed** | | | | | | **−0.01 (−0.02, 0.01)** | | | | | | **−0.03 (−0.06, 0.01)** | | |
| **Random** | | | | | | **−0.01 (−0.02, 0.01)** | | | | | | **−0.03 (−0.06, 0.01)** | | |

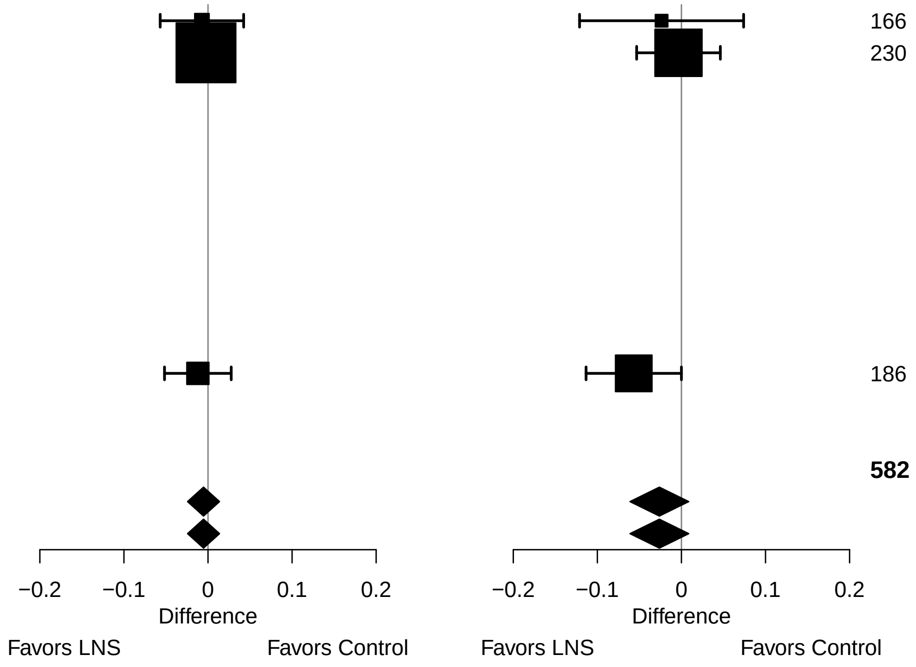

#### Supplemental figure 5N: Executive function lowest decile prevalence difference

#### 5N6: Stratified by Child sex

**P−for−interaction = 0.267**
**Difference in PDs = 0.02 (−0.01, 0.04)**

| Country | Trial | Tool | LNS N | Control N | Control Prevalence | Male PD (95% CI) | Fixed W | Random W | LNS N | Control N | Control Prevalence | Female PD (95% CI) | Fixed W | Random W |
|---|---|---|---|---|---|---|---|---|---|---|---|---|---|---|
| Bangladesh | JiVitA−4 (21) | | | | | | | | | | | | | |
| Bangladesh | RDNS (22) | A not B task | 261 | 133 | 6.8 | 0.03 (−0.03, 0.08) | 0.11 | 0.15 | 272 | 133 | 15.0 | −0.05 (−0.11, 0.00) | 0.14 | 0.15 |
| Bangladesh | WASH−B (23) | A not B task | 540 | 1671 | 9.8 | −0.01 (−0.04, 0.02) | 0.44 | 0.30 | 557 | 1656 | 10.5 | 0.00 (−0.03, 0.03) | 0.47 | 0.43 |
| Burkina Faso | iLiNS−Zinc (24) | | | | | | | | | | | | | |
| Ghana | GHANA (25) | | | | | | | | | | | | | |
| Ghana | iLiNS−DYADG (26) | A not B task | 145 | 273 | 11.0 | −0.04 (−0.10, 0.02) | 0.10 | 0.13 | 145 | 303 | 11.2 | −0.01 (−0.07, 0.05) | 0.10 | 0.11 |
| Haiti | HAITI (27) | | | | | | | | | | | | | |
| Kenya | WASH−B (28) | | | | | | | | | | | | | |
| Madagascar | MAHAY (29) | | | | | | | | | | | | | |
| Malawi | iLiNS−DYADM (30) | A not B task | 92 | 176 | 12.5 | −0.05 (−0.13, 0.03) | 0.06 | 0.09 | 90 | 193 | 9.3 | 0.01 (−0.07, 0.08) | 0.07 | 0.08 |
| Malawi | iLiNS−DOSE (31) | A not B task | 235 | 93 | 7.5 | 0.03 (−0.04, 0.10) | 0.07 | 0.10 | 240 | 71 | 7.0 | 0.04 (−0.04, 0.12) | 0.06 | 0.07 |
| Mali | PROMIS CS (32) | | | | | | | | | | | | | |
| Zimbabwe | SHINE (HIV−) (33) | A not B task | 399 | 364 | 8.8 | −0.04 (−0.08, 0.00) | 0.23 | 0.23 | 388 | 375 | 14.1 | 0.00 (−0.05, 0.05) | 0.15 | 0.16 |
| Zimbabwe | SHINE (HIV+) (34) | A not B task | | | | | | | | | | | | |
| | | | **1672** | **2710** | | **I² = 0.32, Tau² = 0.00** | | | **1692** | **2731** | | **I² = 0.04, Tau² = 0.00** | | |
| **Fixed** | | | | | | **−0.02 (−0.04, 0.00)** | | | | | | **0.00 (−0.02, 0.01)** | | |
| **Random** | | | | | | **−0.02 (−0.04, 0.01)** | | | | | | **−0.01 (−0.03, 0.02)** | | |

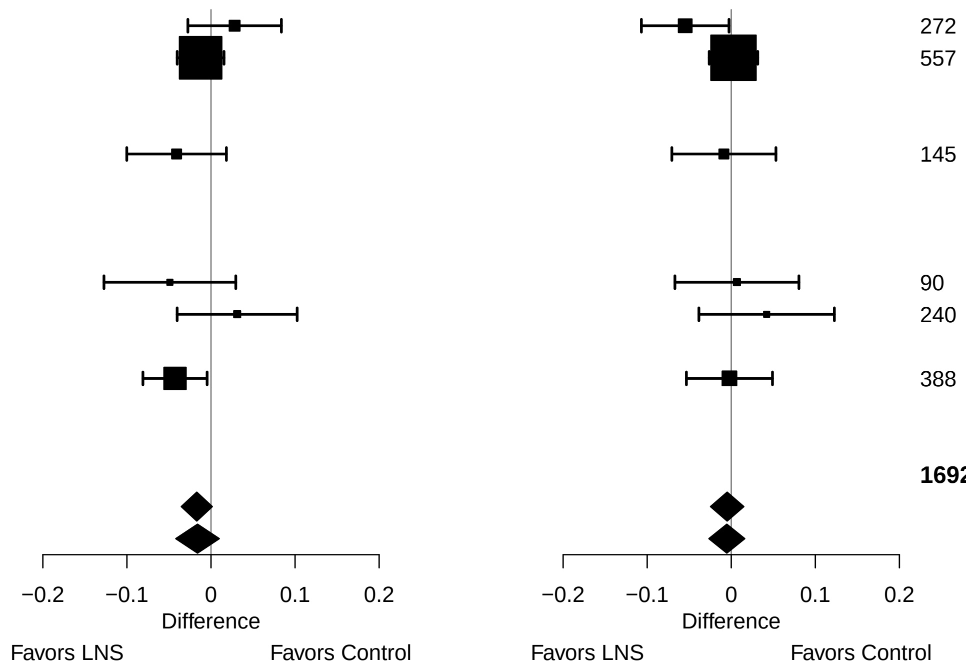

##### Supplemental figure 5N: Executive function lowest decile prevalence difference

#### 5N7: Stratified by Child birth order

**P−for−interaction = 0.116**
**Difference in PDs = −0.02 (−0.06, 0.01)**

| Country | Trial | Tool | LNS N | Control N | Control Prevalence | Later born PD (95% CI) | Fixed W | Random W | LNS N | Control N | Control Prevalence | Firstborn PD (95% CI) | Fixed W | Random W |
|---|---|---|---|---|---|---|---|---|---|---|---|---|---|---|
| Bangladesh | JiVitA−4 (21) | | | | | | | | | | | | | |
| Bangladesh | RDNS (22) | A not B task | 320 | 163 | 12.9 | −0.04 (−0.09, 0.01) | 0.13 | 0.16 | 213 | 103 | 7.8 | 0.03 (−0.04, 0.10) | 0.15 | 0.17 |
| Bangladesh | WASH−B (23) | A not B task | 683 | 2174 | 9.5 | 0.01 (−0.01, 0.04) | 0.52 | 0.43 | 394 | 1049 | 10.3 | −0.03 (−0.06, 0.01) | 0.54 | 0.46 |
| Burkina Faso | iLiNS−Zinc (24) | | | | | | | | | | | | | |
| Ghana | GHANA (25) | | | | | | | | | | | | | |
| Ghana | iLiNS−DYADG (26) | A not B task | 195 | 390 | 10.8 | −0.01 (−0.06, 0.04) | 0.12 | 0.15 | 95 | 186 | 11.8 | −0.06 (−0.13, 0.02) | 0.12 | 0.15 |
| Haiti | HAITI (27) | | | | | | | | | | | | | |
| Kenya | WASH−B (28) | | | | | | | | | | | | | |
| Madagascar | MAHAY (29) | | | | | | | | | | | | | |
| Malawi | iLiNS−DYADM (30) | A not B task | | | | | | | | | | | | |
| Malawi | iLiNS−DOSE (31) | A not B task | | | | | | | | | | | | |
| Mali | PROMIS CS (32) | | | | | | | | | | | | | |
| Zimbabwe | SHINE (HIV−) (33) | A not B task | 568 | 530 | 11.7 | −0.02 (−0.06, 0.02) | 0.24 | 0.26 | 186 | 178 | 11.8 | −0.04 (−0.10, 0.02) | 0.19 | 0.22 |
| Zimbabwe | SHINE (HIV+) (34) | A not B task | | | | | | | | | | | | |
| | | | **1766** | **3257** | | **$I^2$ = 0.32, $Tau^2$ = 0.00** | | | **888** | **1516** | | **$I^2$ = 0.14, $Tau^2$ = 0.00** | | |
| **Fixed** | | | | | | **0.00 (−0.02, 0.01)** | | | | | | **−0.03 (−0.05, 0.00)** | | |
| **Random** | | | | | | **−0.01 (−0.03, 0.02)** | | | | | | **−0.02 (−0.05, 0.01)** | | |

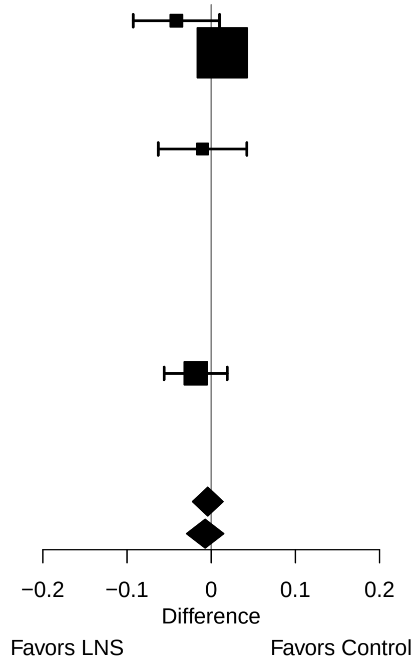

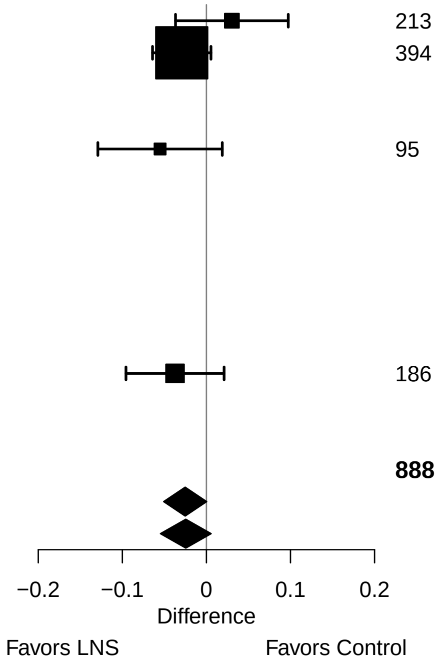

#### Supplemental figure 5N: Executive function lowest decile prevalence difference

#### 5N8: Stratified by Child baseline stunting

**P−for−interaction = 0.577**
**Difference in PDs = 0.02 (−0.04, 0.08)**

| Country | Trial | Tool | LNS N | Control N | Control Prevalence | No PD (95% CI) | Fixed W | Random W | LNS N | Control N | Control Prevalence | Yes PD (95% CI) | Fixed W | Random W |
|---|---|---|---|---|---|---|---|---|---|---|---|---|---|---|
| Bangladesh | JiVitA−4 (21) | | | | | | | | | | | | | |
| Bangladesh | RDNS (22) | A not B task | 404 | 187 | 10.7 | −0.02 (−0.06, 0.03) | 0.33 | 0.34 | 107 | 62 | 11.3 | 0.00 (−0.12, 0.12) | 0.24 | 0.24 |
| Bangladesh | WASH−B (23) | A not B task | | | | | | | | | | | | |
| Burkina Faso | iLiNS−Zinc (24) | | | | | | | | | | | | | |
| Ghana | GHANA (25) | | | | | | | | | | | | | |
| Ghana | iLiNS−DYADG (26) | A not B task | | | | | | | | | | | | |
| Haiti | HAITI (27) | | | | | | | | | | | | | |
| Kenya | WASH−B (28) | | | | | | | | | | | | | |
| Madagascar | MAHAY (29) | | | | | | | | | | | | | |
| Malawi | iLiNS−DYADM (30) | A not B task | | | | | | | | | | | | |
| Malawi | iLiNS−DOSE (31) | A not B task | 353 | 111 | 5.4 | 0.06 (0.00, 0.12) | 0.18 | 0.28 | 122 | 53 | 11.3 | −0.01 (−0.11, 0.08) | 0.36 | 0.36 |
| Mali | PROMIS CS (32) | | | | | | | | | | | | | |
| Zimbabwe | SHINE (HIV−) (33) | A not B task | 497 | 459 | 11.5 | −0.03 (−0.07, 0.01) | 0.49 | 0.38 | 116 | 93 | 9.7 | 0.05 (−0.04, 0.14) | 0.40 | 0.40 |
| Zimbabwe | SHINE (HIV+) (34) | A not B task | | | | | | | | | | | | |
| | | | **1254** | **757** | | **I² = 0.67, Tau² = 0.00** | | | **345** | **208** | | **I² = 0.00, Tau² = 0.00** | | |
| **Fixed** | | | | | | **−0.01 (−0.04, 0.02)** | | | | | | **0.01 (−0.04, 0.07)** | | |
| **Random** | | | | | | **0.00 (−0.06, 0.05)** | | | | | | **0.01 (−0.04, 0.07)** | | |

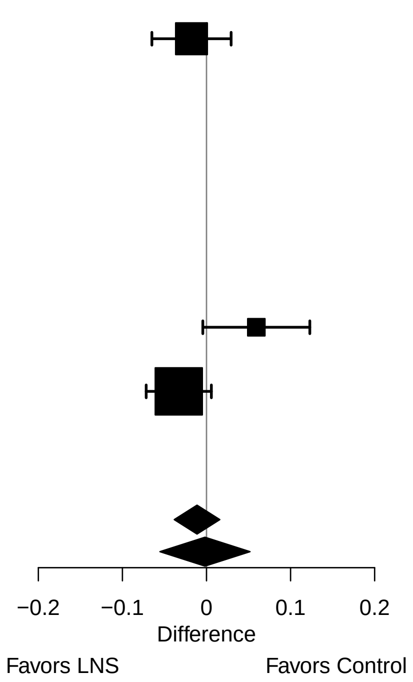

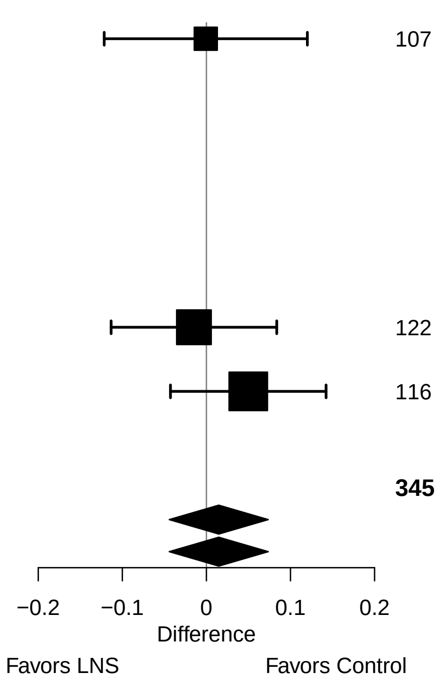

**Supplemental figure 5N: Executive function lowest decile prevalence difference**

#### 5N9: Stratified by Child baseline acute malnutrition (insufficient comparisons)

#### Supplemental figure 5N: Executive function lowest decile prevalence difference

#### 5N10: Stratified by Child baseline anemia

**P−for−interaction = 0.432**
**Difference in PDs = −0.02 (−0.07, 0.03)**

| Country | Trial | Tool | LNS N | Control N | Control Prevalence | Not anemic PD (95% CI) | Fixed W | Random W | LNS N | Control N | Control Prevalence | Anemic PD (95% CI) | Fixed W | Random W |
|---|---|---|---|---|---|---|---|---|---|---|---|---|---|---|
| Bangladesh | JiVitA−4 (21) | | | | | | | | | | | | | |
| Bangladesh | RDNS (22) | A not B task | 204 | 93 | 7.5 | 0.02 (−0.04, 0.07) | 0.38 | 0.38 | 306 | 153 | 13.1 | −0.04 (−0.11, 0.03) | 0.26 | 0.26 |
| Bangladesh | WASH−B (23) | A not B task | | | | | | | | | | | | |
| Burkina Faso | iLiNS−Zinc (24) | | | | | | | | | | | | | |
| Ghana | GHANA (25) | | | | | | | | | | | | | |
| Ghana | iLiNS−DYADG (26) | A not B task | 172 | 308 | 10.1 | 0.01 (−0.05, 0.07) | 0.34 | 0.34 | 78 | 181 | 12.7 | −0.06 (−0.15, 0.02) | 0.19 | 0.21 |
| Haiti | HAITI (27) | | | | | | | | | | | | | |
| Kenya | WASH−B (28) | | | | | | | | | | | | | |
| Madagascar | MAHAY (29) | | | | | | | | | | | | | |
| Malawi | iLiNS−DYADM (30) | A not B task | 55 | 122 | 13.1 | −0.04 (−0.14, 0.06) | 0.10 | 0.10 | 121 | 234 | 9.4 | 0.00 (−0.07, 0.06) | 0.31 | 0.28 |
| Malawi | iLiNS−DOSE (31) | A not B task | 176 | 70 | 8.6 | 0.01 (−0.07, 0.09) | 0.17 | 0.17 | 296 | 93 | 6.5 | 0.05 (−0.02, 0.12) | 0.25 | 0.25 |
| Mali | PROMIS CS (32) | | | | | | | | | | | | | |
| Zimbabwe | SHINE (HIV−) (33) | A not B task | | | | | | | | | | | | |
| Zimbabwe | SHINE (HIV+) (34) | A not B task | | | | | | | | | | | | |
| | | | **607** | **593** | | **I² = 0.00, Tau² = 0.00** | | | **801** | **661** | | **I² = 0.39, Tau² = 0.00** | | |
| **Fixed** | | | | | | **0.01 (−0.03, 0.04)** | | | | | | **−0.01 (−0.04, 0.03)** | | |
| **Random** | | | | | | **0.01 (−0.03, 0.04)** | | | | | | **−0.01 (−0.06, 0.04)** | | |

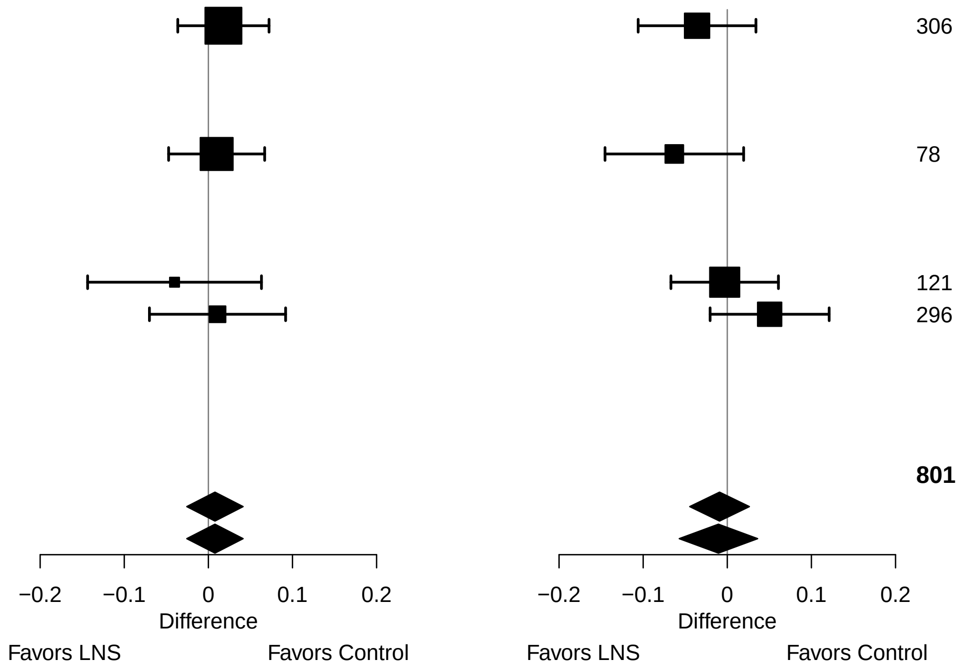

### Supplemental figure 5O: 12-mo walking without support prevalence ratio

#### 5O1: Stratified by Maternal height

**P−for−interaction = 0.624**
**Ratio of PRs = 0.96 (0.79, 1.15)**

| | | At least 150.1 cm | | | | | | Less than 150.1 cm | | | | | |
|---|---|---|---|---|---|---|---|---|---|---|---|---|---|
| Country | Trial | LNS N | Control N | Control Prevalence | PR (95% CI) | Fixed W | Random W | LNS N | Control N | Control Prevalence | PR (95% CI) | Fixed W | Random W |
| Bangladesh | RDNS (22) | 848 | 418 | 74.4 | 1.42 (1.16, 1.73) | 0.15 | 0.16 | 721 | 347 | 76.4 | 1.17 (0.87, 1.56) | 0.27 | 0.27 |
| Bangladesh | WASH−B (23) | 261 | 782 | 73.0 | 1.21 (0.99, 1.47) | 0.16 | 0.16 | 230 | 666 | 76.1 | 1.22 (0.98, 1.52) | 0.46 | 0.46 |
| Burkina Faso | iLiNS−Zinc (24) | | | | | | | | | | | | |
| Ghana | GHANA (25) | | | | | | | | | | | | |
| Ghana | iLiNS−DYADG (26) | 308 | 613 | 52.9 | 1.13 (0.99, 1.29) | 0.34 | 0.34 | 14 | 38 | 63.2 | 1.74 (0.99, 3.09) | 0.07 | 0.07 |
| Haiti | HAITI (27) | | | | | | | | | | | | |
| Kenya | WASH−B (28) | | | | | | | | | | | | |
| Madagascar | MAHAY (29) | | | | | | | | | | | | |
| Malawi | iLiNS−DYADM (30) | 180 | 367 | 50.4 | 1.20 (1.02, 1.40) | 0.24 | 0.24 | 28 | 56 | 53.6 | 1.15 (0.74, 1.80) | 0.12 | 0.12 |
| Malawi | iLiNS−DOSE (31) | 505 | 164 | 66.5 | 1.10 (0.86, 1.40) | 0.10 | 0.10 | 96 | 35 | 62.9 | 0.84 (0.50, 1.42) | 0.08 | 0.08 |
| Mali | PROMIS CS (32) | | | | | | | | | | | | |
| Zimbabwe | SHINE (HIV−) (33) | | | | | | | | | | | | |
| Zimbabwe | SHINE (HIV+) (34) | | | | | | | | | | | | |
| | | **2102** | **2344** | | **I² = 0.00, Tau² = 0.00** | | | **1089** | **1142** | | **I² = 0.00, Tau² = 0.00** | | |
| **Fixed** | | | | | **1.20 (1.11, 1.29)** | | | | | | **1.19 (1.02, 1.39)** | | |
| **Random** | | | | | **1.20 (1.11, 1.29)** | | | | | | **1.19 (1.02, 1.39)** | | |

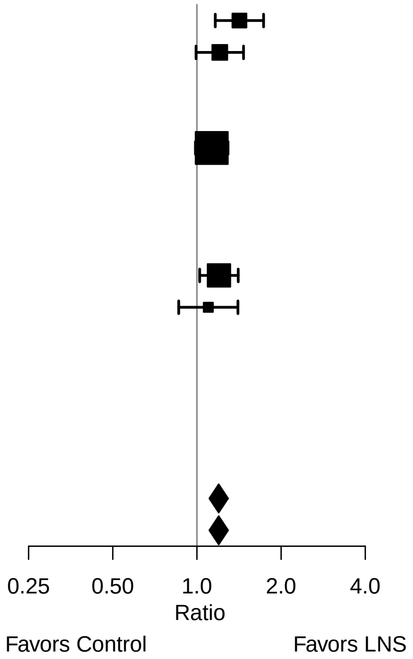

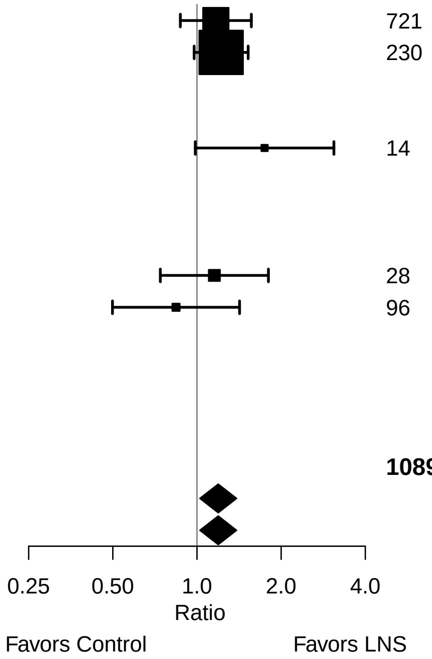

**Supplemental figure 5O: 12-mo walking without support prevalence ratio**

#### 5O2: Stratified by Maternal BMI

**P−for−interaction = 0.343**
**Ratio of PRs = 0.94 (0.82, 1.07)**

| Country | Trial | At least 20 kg/m² LNS N | Control N | Control Prevalence | PR (95% CI) | Fixed W | Random W | Less than 20 kg/m² LNS N | Control N | Control Prevalence | PR (95% CI) | Fixed W | Random W |
|---|---|---|---|---|---|---|---|---|---|---|---|---|---|
| Bangladesh | RDNS (22) | 708 | 327 | 74.6 | 1.57 (1.20, 2.05) | 0.08 | 0.13 | 861 | 438 | 75.8 | 1.08 (0.88, 1.32) | 0.27 | 0.27 |
| Bangladesh | WASH−B (23) | 225 | 637 | 73.0 | 1.30 (1.06, 1.60) | 0.13 | 0.15 | 266 | 810 | 75.7 | 1.13 (0.91, 1.39) | 0.25 | 0.25 |
| Burkina Faso | iLiNS−Zinc (24) | | | | | | | | | | | | |
| Ghana | GHANA (25) | | | | | | | | | | | | |
| Ghana | iLiNS−DYADG (26) | 284 | 535 | 52.3 | 1.15 (1.00, 1.32) | 0.29 | 0.18 | 38 | 116 | 58.6 | 1.08 (0.71, 1.64) | 0.06 | 0.06 |
| Haiti | HAITI (27) | | | | | | | | | | | | |
| Kenya | WASH−B (28) | 334 | 1210 | 58.8 | 0.92 (0.79, 1.07) | 0.25 | 0.17 | 96 | 364 | 61.5 | 1.03 (0.77, 1.38) | 0.13 | 0.13 |
| Madagascar | MAHAY (29) | | | | | | | | | | | | |
| Malawi | iLiNS−DYADM (30) | 121 | 255 | 51.4 | 1.17 (0.96, 1.43) | 0.14 | 0.16 | 86 | 168 | 50.0 | 1.23 (0.98, 1.54) | 0.22 | 0.22 |
| Malawi | iLiNS−DOSE (31) | 451 | 138 | 63.0 | 1.00 (0.78, 1.28) | 0.09 | 0.14 | 148 | 60 | 71.7 | 1.19 (0.75, 1.89) | 0.05 | 0.05 |
| Mali | PROMIS CS (32) | 83 | 98 | 66.3 | 0.68 (0.41, 1.14) | 0.02 | 0.07 | 34 | 52 | 67.3 | 0.81 (0.41, 1.60) | 0.02 | 0.02 |
| Zimbabwe | SHINE (HIV−) (33) | | | | | | | | | | | | |
| Zimbabwe | SHINE (HIV+) (34) | | | | | | | | | | | | |
| | | **2206** | **3200** | | **I² = 0.69, Tau² = 0.04** | | | **1529** | **2008** | | **I² = 0.00, Tau² = 0.00** | | |
| **Fixed** | | | | | **1.11 (1.03, 1.20)** | | | | | | **1.11 (1.00, 1.24)** | | |
| **Random** | | | | | **1.11 (0.94, 1.32)** | | | | | | **1.11 (1.00, 1.24)** | | |

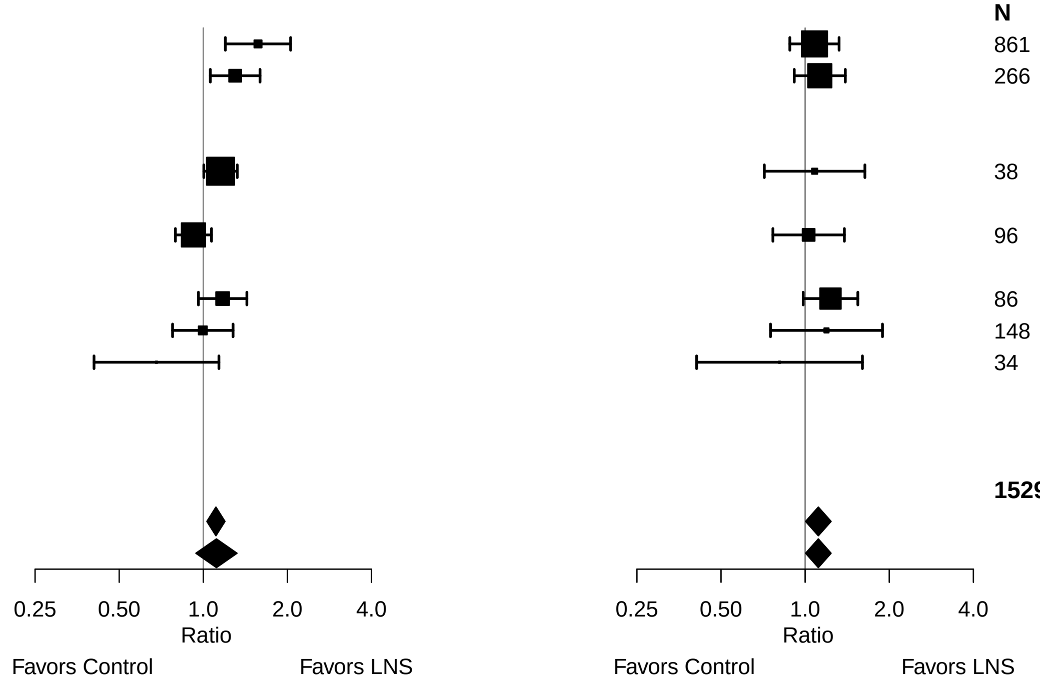

#### Supplemental figure 5O: 12-mo walking without support prevalence ratio

#### 5O3: Stratified by Maternal age

P−for−interaction = 0.394
Ratio of PRs = 1.04 (0.95, 1.13)

| | | At least 25 y | | | | | | Less than 25 y | | | | | |
|---|---|---|---|---|---|---|---|---|---|---|---|---|---|
| Country | Trial | LNS N | Control N | Control Prevalence | PR (95% CI) | Fixed W | Random W | LNS N | Control N | Control Prevalence | PR (95% CI) | Fixed W | Random W |
| Bangladesh | RDNS (22) | 447 | 217 | 76.0 | 1.37 (1.07, 1.76) | 0.11 | 0.13 | 1181 | 573 | 73.8 | 1.25 (1.02, 1.53) | 0.16 | 0.16 |
| Bangladesh | WASH−B (23) | 223 | 676 | 74.6 | 0.95 (0.74, 1.22) | 0.11 | 0.13 | 274 | 791 | 74.6 | 1.41 (1.18, 1.67) | 0.22 | 0.18 |
| Burkina Faso | iLiNS−Zinc (24) | | | | | | | | | | | | |
| Ghana | GHANA (25) | | | | | | | | | | | | |
| Ghana | iLiNS−DYADG (26) | 209 | 405 | 53.8 | 1.08 (0.91, 1.28) | 0.24 | 0.18 | 118 | 258 | 51.9 | 1.27 (1.05, 1.54) | 0.18 | 0.16 |
| Haiti | HAITI (27) | 60 | 57 | 68.4 | 1.58 (1.00, 2.50) | 0.03 | 0.06 | 22 | 22 | 50.0 | 0.73 (0.36, 1.45) | 0.01 | 0.03 |
| Kenya | WASH−B (28) | 243 | 914 | 61.3 | 1.03 (0.87, 1.22) | 0.24 | 0.18 | 197 | 679 | 57.7 | 0.91 (0.76, 1.10) | 0.20 | 0.17 |
| Madagascar | MAHAY (29) | | | | | | | | | | | | |
| Malawi | iLiNS−DYADM (30) | 100 | 213 | 47.9 | 1.06 (0.85, 1.31) | 0.15 | 0.15 | 108 | 213 | 54.5 | 1.36 (1.11, 1.68) | 0.15 | 0.16 |
| Malawi | iLiNS−DOSE (31) | 327 | 111 | 64.9 | 1.11 (0.83, 1.47) | 0.09 | 0.12 | 264 | 83 | 67.5 | 0.99 (0.69, 1.41) | 0.05 | 0.09 |
| Mali | PROMIS CS (32) | 59 | 71 | 62.0 | 0.58 (0.32, 1.06) | 0.02 | 0.04 | 59 | 80 | 70.0 | 0.85 (0.49, 1.45) | 0.02 | 0.05 |
| Zimbabwe | SHINE (HIV−) (33) | | | | | | | | | | | | |
| Zimbabwe | SHINE (HIV+) (34) | | | | | | | | | | | | |
| | | **1668** | **2664** | | **I² = 0.32, Tau² = 0.02** | | | **2223** | **2699** | | **I² = 0.65, Tau² = 0.02** | | |
| **Fixed** | | | | | **1.07 (1.00, 1.14)** | | | | | | **1.12 (1.05, 1.18)** | | |
| **Random** | | | | | **1.08 (0.96, 1.21)** | | | | | | **1.14 (1.01, 1.29)** | | |

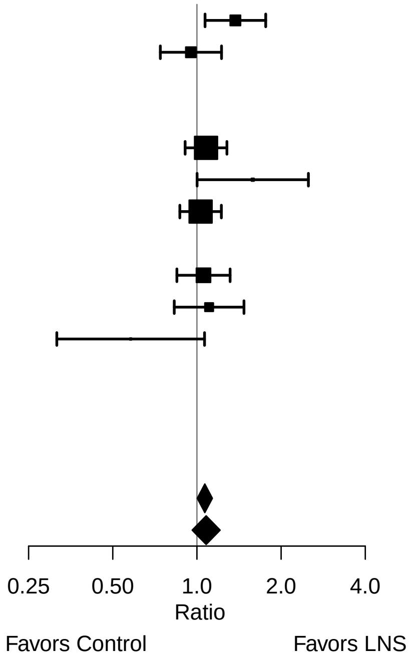

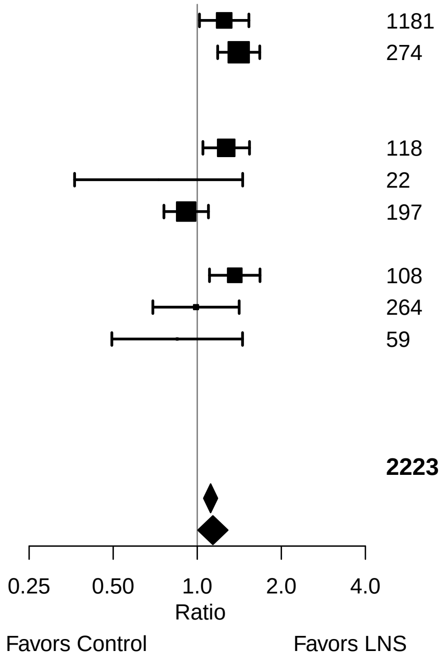

#### Supplemental figure 5O: 12-mo walking without support prevalence ratio

##### 5O4: Stratified by Maternal education

**P−for−interaction = 0.792**
**Ratio of PRs = 0.99 (0.91, 1.08)**

| | | Primary or greater | | | | | | Incomplete or no formal | | | | | |
|---|---|---|---|---|---|---|---|---|---|---|---|---|---|
| Country | Trial | LNS N | Control N | Control Prevalence | PR (95% CI) | Fixed W | Random W | LNS N | Control N | Control Prevalence | PR (95% CI) | Fixed W | Random W |
| Bangladesh | RDNS (22) | 1219 | 574 | 73.9 | 1.32 (1.08, 1.62) | 0.15 | 0.17 | 409 | 216 | 75.9 | 1.14 (0.91, 1.42) | 0.17 | 0.17 |
| Bangladesh | WASH−B (23) | 337 | 1048 | 72.7 | 1.28 (1.11, 1.49) | 0.28 | 0.26 | 160 | 426 | 79.1 | 1.02 (0.76, 1.37) | 0.10 | 0.10 |
| Burkina Faso | iLiNS−Zinc (24) | | | | | | | | | | | | |
| Ghana | GHANA (25) | | | | | | | | | | | | |
| Ghana | iLiNS−DYADG (26) | 257 | 524 | 51.5 | 1.12 (0.97, 1.30) | 0.30 | 0.27 | 70 | 139 | 59.0 | 1.25 (0.93, 1.70) | 0.09 | 0.10 |
| Haiti | HAITI (27) | | | | | | | | | | | | |
| Kenya | WASH−B (28) | 217 | 775 | 59.2 | 1.04 (0.87, 1.24) | 0.19 | 0.20 | 229 | 827 | 60.3 | 0.89 (0.72, 1.10) | 0.20 | 0.20 |
| Madagascar | MAHAY (29) | | | | | | | | | | | | |
| Malawi | iLiNS−DYADM (30) | 32 | 64 | 42.2 | 0.97 (0.67, 1.41) | 0.04 | 0.06 | 175 | 359 | 52.4 | 1.24 (1.05, 1.46) | 0.32 | 0.29 |
| Malawi | iLiNS−DOSE (31) | 129 | 42 | 61.9 | 1.08 (0.70, 1.67) | 0.03 | 0.05 | 462 | 152 | 67.1 | 1.05 (0.81, 1.36) | 0.13 | 0.13 |
| Mali | PROMIS CS (32) | | | | | | | | | | | | |
| Zimbabwe | SHINE (HIV−) (33) | | | | | | | | | | | | |
| Zimbabwe | SHINE (HIV+) (34) | | | | | | | | | | | | |
| | | **2191** | **3027** | | **I² = 0.44, Tau² = 0.00** | | | **1505** | **2119** | | **I² = 0.16, Tau² = 0.00** | | |
| **Fixed** | | | | | **1.10 (1.04, 1.16)** | | | | | | **1.09 (1.02, 1.16)** | | |
| **Random** | | | | | **1.12 (1.04, 1.21)** | | | | | | **1.09 (1.00, 1.18)** | | |

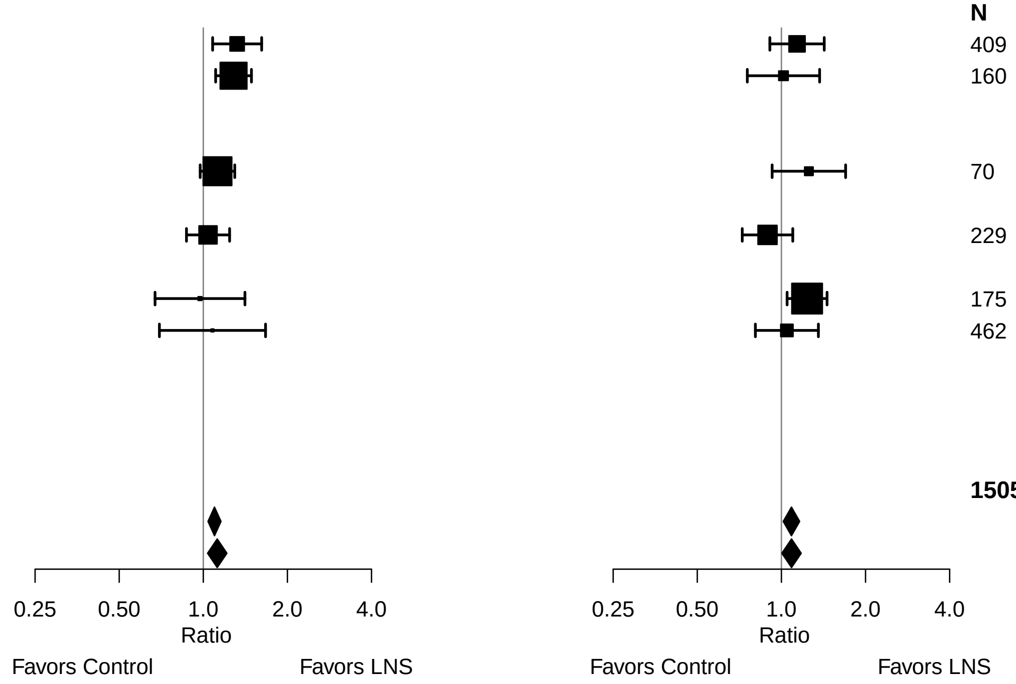

#### Supplemental figure 5O: 12-mo walking without support prevalence ratio

#### 5O5: Stratified by Maternal depressive symptoms

**P−for−interaction = 0.543**
**Ratio of PRs = 0.95 (0.81, 1.12)**

| Country | Trial | Less than 75th percentile: LNS N | Control N | Control Prevalence | PR (95% CI) | Fixed W | Random W | At least 75th percentile: LNS N | Control N | Control Prevalence | PR (95% CI) | Fixed W | Random W |
|---|---|---|---|---|---|---|---|---|---|---|---|---|---|
| Bangladesh | RDNS (22) | 1058 | 460 | 70.9 | 1.11 (0.89, 1.38) | 0.11 | 0.13 | 508 | 279 | 79.6 | 1.67 (1.26, 2.21) | 0.24 | 0.22 |
| Bangladesh | WASH−B (23) | 389 | 1075 | 73.4 | 1.28 (1.08, 1.50) | 0.20 | 0.21 | 108 | 399 | 77.7 | 0.83 (0.53, 1.29) | 0.10 | 0.15 |
| Burkina Faso | iLiNS−Zinc (24) | | | | | | | | | | | | |
| Ghana | GHANA (25) | | | | | | | | | | | | |
| Ghana | iLiNS−DYADG (26) | 242 | 446 | 52.9 | 1.16 (1.00, 1.35) | 0.24 | 0.23 | 75 | 198 | 55.1 | 1.16 (0.89, 1.51) | 0.27 | 0.23 |
| Haiti | HAITI (27) | | | | | | | | | | | | |
| Kenya | WASH−B (28) | 314 | 1098 | 59.6 | 0.98 (0.85, 1.14) | 0.25 | 0.23 | 99 | 373 | 61.1 | 1.01 (0.77, 1.33) | 0.25 | 0.23 |
| Madagascar | MAHAY (29) | | | | | | | | | | | | |
| Malawi | iLiNS−DYADM (30) | 149 | 293 | 48.1 | 1.20 (1.02, 1.42) | 0.20 | 0.20 | 47 | 108 | 60.2 | 1.23 (0.85, 1.78) | 0.14 | 0.17 |
| Malawi | iLiNS−DOSE (31) | | | | | | | | | | | | |
| Mali | PROMIS CS (32) | | | | | | | | | | | | |
| Zimbabwe | SHINE (HIV−) (33) | | | | | | | | | | | | |
| Zimbabwe | SHINE (HIV+) (34) | | | | | | | | | | | | |
| | | **2152** | **3372** | | **I² = 0.34, Tau² = 0.00** | | | **837** | **1357** | | **I² = 0.57, Tau² = 0.04** | | |
| **Fixed** | | | | | **1.14 (1.06, 1.23)** | | | | | | **1.19 (1.04, 1.37)** | | |
| **Random** | | | | | **1.14 (1.04, 1.25)** | | | | | | **1.17 (0.94, 1.46)** | | |

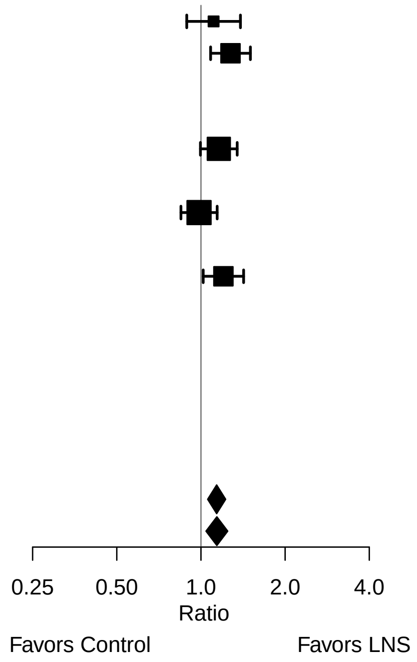

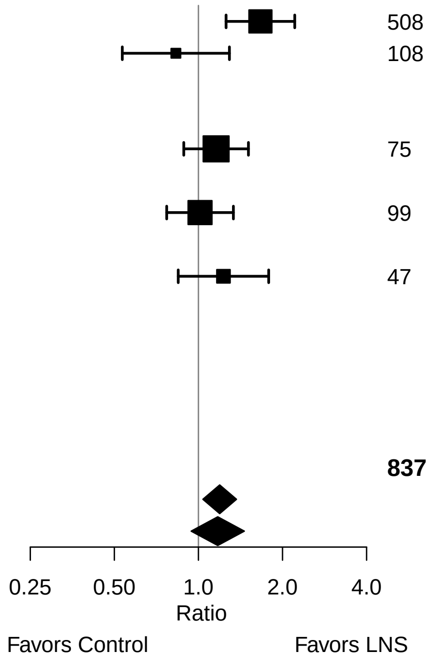

#### Supplemental figure 5O: 12-mo walking without support prevalence ratio

##### 5O6: Stratified by Child sex

**P−for−interaction = 0.952**
**Ratio of PRs = 1.00 (0.92, 1.09)**

| Country | Trial | Male LNS N | Male Control N | Male Control Prevalence | Male PR (95% CI) | Male Fixed W | Male Random W | Female LNS N | Female Control N | Female Control Prevalence | Female PR (95% CI) | Female Fixed W | Female Random W |
|---|---|---|---|---|---|---|---|---|---|---|---|---|---|
| Bangladesh | RDNS (22) | 819 | 396 | 74.5 | 1.17 (0.94, 1.45) | 0.16 | 0.16 | 801 | 394 | 74.4 | 1.40 (1.11, 1.76) | 0.12 | 0.13 |
| Bangladesh | WASH−B (23) | 232 | 692 | 75.9 | 1.27 (1.02, 1.58) | 0.15 | 0.15 | 265 | 782 | 73.4 | 1.15 (0.94, 1.41) | 0.16 | 0.13 |
| Burkina Faso | iLiNS−Zinc (24) | | | | | | | | | | | | |
| Ghana | GHANA (25) | 55 | 40 | 67.5 | 1.29 (0.75, 2.22) | 0.03 | 0.04 | 35 | 47 | 80.9 | 2.24 (1.11, 4.51) | 0.01 | 0.06 |
| Ghana | iLiNS−DYADG (26) | 166 | 321 | 53.6 | 1.16 (0.96, 1.39) | 0.23 | 0.19 | 160 | 341 | 52.8 | 1.14 (0.95, 1.37) | 0.20 | 0.13 |
| Haiti | HAITI (27) | 38 | 29 | 69.0 | 1.19 (0.60, 2.35) | 0.02 | 0.02 | 45 | 50 | 60.0 | 1.33 (0.86, 2.06) | 0.03 | 0.10 |
| Kenya | WASH−B (28) | 231 | 764 | 62.7 | 0.97 (0.78, 1.21) | 0.16 | 0.16 | 216 | 840 | 57.0 | 0.97 (0.81, 1.15) | 0.22 | 0.13 |
| Madagascar | MAHAY (29) | | | | | | | | | | | | |
| Malawi | iLiNS−DYADM (30) | 105 | 198 | 57.6 | 1.48 (1.19, 1.84) | 0.16 | 0.16 | 103 | 228 | 45.6 | 1.00 (0.81, 1.24) | 0.15 | 0.13 |
| Malawi | iLiNS−DOSE (31) | 310 | 111 | 73.9 | 1.40 (0.99, 1.97) | 0.06 | 0.08 | 292 | 89 | 55.1 | 0.79 (0.60, 1.05) | 0.09 | 0.12 |
| Mali | PROMIS CS (32) | 58 | 83 | 71.1 | 1.07 (0.63, 1.83) | 0.03 | 0.04 | 60 | 68 | 60.3 | 0.42 (0.23, 0.75) | 0.02 | 0.08 |
| Zimbabwe | SHINE (HIV−) (33) | | | | | | | | | | | | |
| Zimbabwe | SHINE (HIV+) (34) | | | | | | | | | | | | |
| | | **2014** | **2634** | | **I² = 0.49, Tau² = 0.01** | | | **1977** | **2839** | | **I² = 0.67, Tau² = 0.10** | | |
| **Fixed** | | | | | **1.10 (1.03, 1.17)** | | | | | | **1.09 (1.03, 1.15)** | | |
| **Random** | | | | | **1.15 (1.05, 1.26)** | | | | | | **1.06 (0.86, 1.33)** | | |

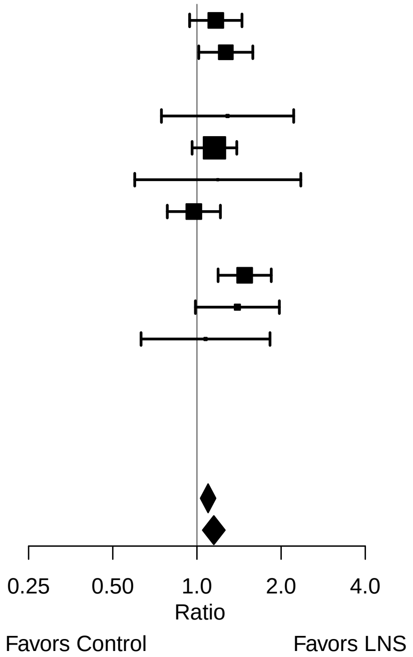

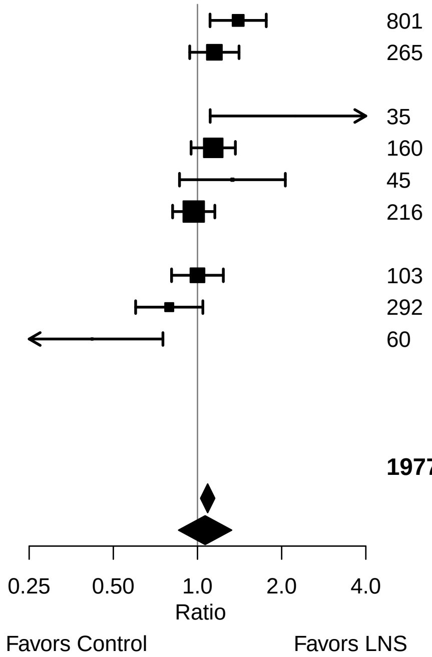

**Supplemental figure 5O: 12-mo walking without support prevalence ratio**

#### 5O7: Stratified by Child birth order

**P−for−interaction = 0.025**
**Ratio of PRs = 0.90 (0.81, 0.99)**

| Country | Trial | Later born LNS N | Later born Control N | Later born Control Prevalence | Later born PR (95% CI) | Later born Fixed W | Later born Random W | Firstborn LNS N | Firstborn Control N | Firstborn Control Prevalence | Firstborn PR (95% CI) | Firstborn Fixed W | Firstborn Random W |
|---|---|---|---|---|---|---|---|---|---|---|---|---|---|
| Bangladesh | RDNS (22) | 953 | 496 | 75.6 | 1.38 (1.13, 1.67) | 0.14 | 0.14 | 673 | 294 | 72.4 | 1.14 (0.90, 1.45) | 0.20 | 0.17 |
| Bangladesh | WASH−B (23) | 314 | 944 | 74.8 | 1.15 (0.94, 1.40) | 0.13 | 0.14 | 157 | 440 | 74.1 | 1.38 (1.09, 1.74) | 0.20 | 0.17 |
| Burkina Faso | iLiNS−Zinc (24) | | | | | | | | | | | | |
| Ghana | GHANA (25) | 54 | 47 | 83.0 | 2.07 (1.00, 4.28) | 0.01 | 0.02 | 36 | 36 | 69.4 | 1.73 (0.97, 3.09) | 0.03 | 0.06 |
| Ghana | iLiNS−DYADG (26) | 222 | 439 | 54.9 | 1.21 (1.03, 1.42) | 0.20 | 0.17 | 105 | 224 | 49.6 | 1.04 (0.83, 1.30) | 0.22 | 0.18 |
| Haiti | HAITI (27) | 57 | 50 | 68.0 | 1.43 (0.87, 2.33) | 0.02 | 0.04 | 26 | 29 | 55.2 | 1.03 (0.58, 1.84) | 0.03 | 0.06 |
| Kenya | WASH−B (28) | 338 | 1296 | 60.2 | 1.02 (0.88, 1.18) | 0.23 | 0.17 | 108 | 306 | 58.2 | 0.80 (0.60, 1.05) | 0.15 | 0.15 |
| Madagascar | MAHAY (29) | | | | | | | | | | | | |
| Malawi | iLiNS−DYADM (30) | 166 | 340 | 50.6 | 1.17 (0.99, 1.39) | 0.18 | 0.16 | 42 | 84 | 52.4 | 1.30 (0.94, 1.80) | 0.11 | 0.12 |
| Malawi | iLiNS−DOSE (31) | 388 | 139 | 67.6 | 1.20 (0.92, 1.58) | 0.07 | 0.10 | 124 | 33 | 57.6 | 0.72 (0.45, 1.16) | 0.05 | 0.08 |
| Mali | PROMIS CS (32) | 103 | 134 | 65.7 | 0.65 (0.39, 1.08) | 0.02 | 0.04 | 14 | 14 | 64.3 | 1.00 (0.39, 2.56) | 0.01 | 0.02 |
| Zimbabwe | SHINE (HIV−) (33) | | | | | | | | | | | | |
| Zimbabwe | SHINE (HIV+) (34) | | | | | | | | | | | | |
| | | **2595** | **3885** | | **I² = 0.37, Tau² = 0.01** | | | **1285** | **1460** | | **I² = 0.46, Tau² = 0.02** | | |
| **Fixed** | | | | | **1.16 (1.09, 1.23)** | | | | | | **1.05 (0.99, 1.11)** | | |
| **Random** | | | | | **1.17 (1.05, 1.30)** | | | | | | **1.08 (0.94, 1.24)** | | |

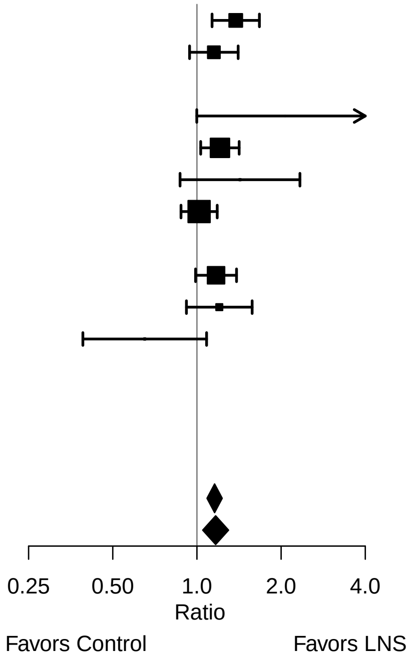

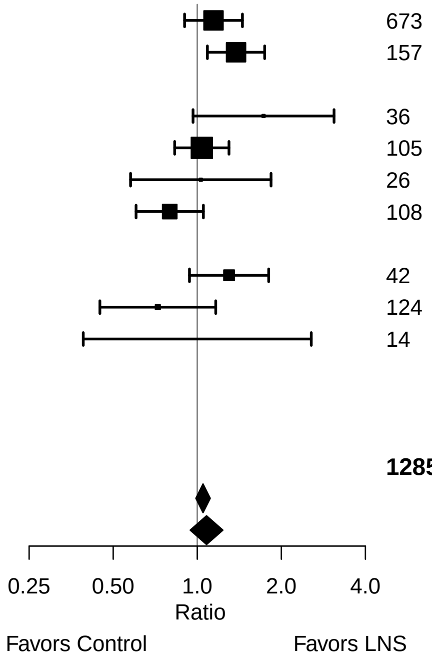

#### Supplemental figure 5O: 12-mo walking without support prevalence ratio

#### 5O8: Stratified by Child baseline stunting

**P−for−interaction = 0.624**
**Ratio of PRs = 1.03 (0.90, 1.18)**

| Country | Trial | No: LNS N | No: Control N | No: Control Prevalence | No: PR (95% CI) | No: Fixed W | No: Random W | Yes: LNS N | Yes: Control N | Yes: Control Prevalence | Yes: PR (95% CI) | Yes: Fixed W | Yes: Random W |
|---|---|---|---|---|---|---|---|---|---|---|---|---|---|
| Bangladesh | RDNS (22) | 1219 | 559 | 72.1 | 1.23 (1.02, 1.47) | 0.22 | 0.24 | 344 | 179 | 80.4 | 1.46 (0.96, 2.22) | 0.34 | 0.33 |
| Bangladesh | WASH−B (23) | | | | | | | | | | | | |
| Burkina Faso | iLiNS−Zinc (24) | | | | | | | | | | | | |
| Ghana | GHANA (25) | | | | | | | | | | | | |
| Ghana | iLiNS−DYADG (26) | 277 | 557 | 52.1 | 1.14 (1.00, 1.31) | 0.37 | 0.32 | 28 | 62 | 69.4 | 1.17 (0.63, 2.17) | 0.15 | 0.16 |
| Haiti | HAITI (27) | | | | | | | | | | | | |
| Kenya | WASH−B (28) | | | | | | | | | | | | |
| Madagascar | MAHAY (29) | | | | | | | | | | | | |
| Malawi | iLiNS−DYADM (30) | 148 | 317 | 47.6 | 1.21 (1.03, 1.42) | 0.27 | 0.27 | 50 | 93 | 63.4 | 1.09 (0.71, 1.69) | 0.32 | 0.32 |
| Malawi | iLiNS−DOSE (31) | 424 | 133 | 57.1 | 0.90 (0.71, 1.13) | 0.13 | 0.17 | 178 | 67 | 82.1 | 1.69 (0.97, 2.96) | 0.19 | 0.19 |
| Mali | PROMIS CS (32) | | | | | | | | | | | | |
| Zimbabwe | SHINE (HIV−) (33) | | | | | | | | | | | | |
| Zimbabwe | SHINE (HIV+) (34) | | | | | | | | | | | | |
| | | **2068** | **1566** | | **$I^2$ = 0.45, $Tau^2$ = 0.01** | | | **600** | **401** | | **$I^2$ = 0.11, $Tau^2$ = 0.00** | | |
| **Fixed** | | | | | **1.09 (1.03, 1.15)** | | | | | | **1.11 (0.98, 1.26)** | | |
| **Random** | | | | | **1.11 (1.01, 1.22)** | | | | | | **1.13 (0.98, 1.31)** | | |

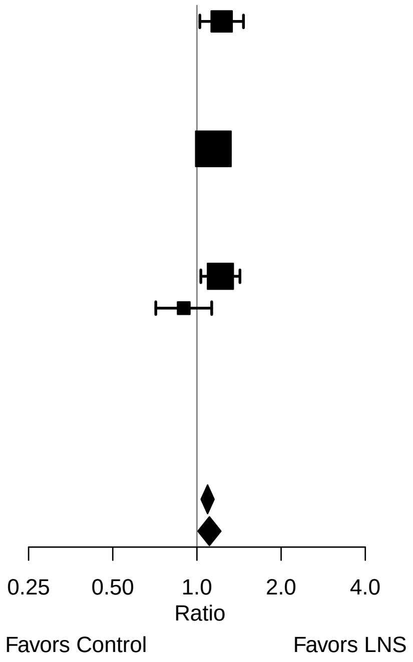

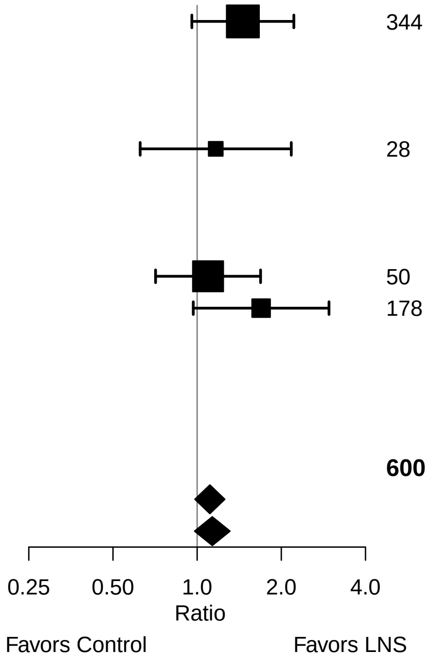

**Supplemental figure 5O: 12-mo walking without support prevalence ratio**

#### 5O9: Stratified by Child baseline acute malnutrition

**P−for−interaction = 0.133**
**Ratio of PRs = 1.13 (0.96, 1.31)**

**No**

| Country | Trial | LNS N | Control N | Control Prevalence | PR (95% CI) | Fixed W | Random W |
|---|---|---|---|---|---|---|---|
| Bangladesh | RDNS (22) | 1447 | 689 | 73.0 | 1.26 (1.05, 1.51) | 0.25 | 0.27 |
| Bangladesh | WASH−B (23) | | | | | | |
| Burkina Faso | iLiNS−Zinc (24) | | | | | | |
| Ghana | GHANA (25) | | | | | | |
| Ghana | iLiNS−DYADG (26) | 284 | 564 | 50.7 | 1.09 (0.95, 1.25) | 0.43 | 0.41 |
| Haiti | HAITI (27) | | | | | | |
| Kenya | WASH−B (28) | | | | | | |
| Madagascar | MAHAY (29) | | | | | | |
| Malawi | iLiNS−DYADM (30) | 184 | 374 | 50.3 | 1.17 (1.00, 1.37) | 0.32 | 0.32 |
| Malawi | iLiNS−DOSE (31) | | | | | | |
| Mali | PROMIS CS (32) | | | | | | |
| Zimbabwe | SHINE (HIV−) (33) | | | | | | |
| Zimbabwe | SHINE (HIV+) (34) | | | | | | |
| | | **1915** | **1627** | | **I² = 0.36, Tau² = 0.00** | | |
| **Fixed** | | | | | **1.08 (1.03, 1.14)** | | |
| **Random** | | | | | **1.10 (1.02, 1.19)** | | |

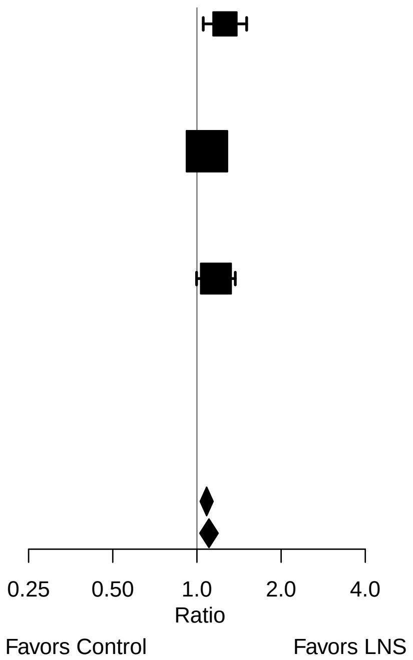

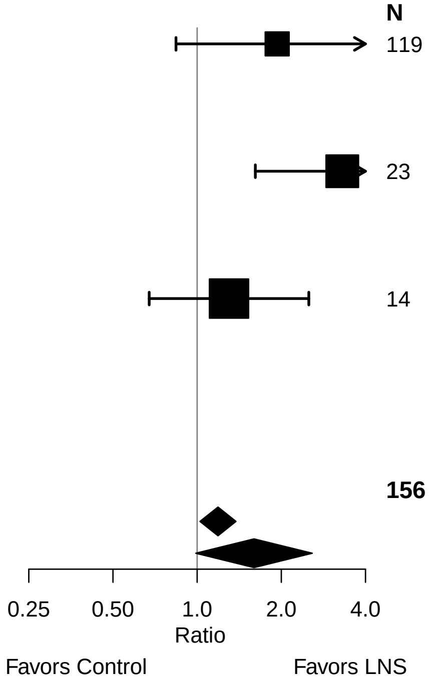

**Yes**

| Country | Trial | LNS N | Control N | Control Prevalence | PR (95% CI) | Fixed W | Random W |
|---|---|---|---|---|---|---|---|
| Bangladesh | RDNS (22) | 119 | 50 | 90.0 | 1.93 (0.84, 4.45) | 0.25 | 0.29 |
| Ghana | iLiNS−DYADG (26) | 23 | 57 | 84.2 | 3.30 (1.61, 6.76) | 0.34 | 0.34 |
| Malawi | iLiNS−DYADM (30) | 14 | 39 | 61.5 | 1.30 (0.67, 2.51) | 0.41 | 0.37 |
| | | **156** | **146** | | **I² = 0.70, Tau² = 0.15** | | |
| **Fixed** | | | | | **1.19 (1.02, 1.38)** | | |
| **Random** | | | | | **1.60 (0.99, 2.58)** | | |

#### Supplemental figure 5O: 12-mo walking without support prevalence ratio

#### 5O10: Stratified by Child baseline anemia

**P−for−interaction = 0.526**
**Ratio of PRs = 0.94 (0.78, 1.14)**

| Country | Trial | Not anemic: LNS N | Not anemic: Control N | Not anemic: Control Prevalence | Not anemic: PR (95% CI) | Not anemic: Fixed W | Not anemic: Random W | Anemic: LNS N | Anemic: Control N | Anemic: Control Prevalence | Anemic: PR (95% CI) | Anemic: Fixed W | Anemic: Random W |
|---|---|---|---|---|---|---|---|---|---|---|---|---|---|
| Bangladesh | RDNS (22) | 211 | 95 | 71.6 | 1.20 (0.87, 1.65) | 0.14 | 0.14 | 310 | 157 | 74.5 | 1.20 (0.84, 1.72) | 0.13 | 0.16 |
| Bangladesh | WASH−B (23) | | | | | | | | | | | | |
| Burkina Faso | iLiNS−Zinc (24) | | | | | | | | | | | | |
| Ghana | GHANA (25) | | | | | | | | | | | | |
| Ghana | iLiNS−DYADG (26) | 185 | 358 | 51.4 | 1.22 (1.04, 1.44) | 0.55 | 0.55 | 91 | 201 | 55.2 | 0.93 (0.70, 1.24) | 0.20 | 0.23 |
| Haiti | HAITI (27) | | | | | | | | | | | | |
| Kenya | WASH−B (28) | | | | | | | | | | | | |
| Madagascar | MAHAY (29) | | | | | | | | | | | | |
| Malawi | iLiNS−DYADM (30) | 63 | 143 | 45.5 | 1.05 (0.81, 1.36) | 0.20 | 0.20 | 135 | 266 | 54.1 | 1.28 (1.05, 1.55) | 0.45 | 0.38 |
| Malawi | iLiNS−DOSE (31) | 205 | 84 | 67.9 | 1.20 (0.84, 1.71) | 0.11 | 0.11 | 393 | 116 | 63.8 | 0.96 (0.73, 1.27) | 0.22 | 0.24 |
| Mali | PROMIS CS (32) | | | | | | | | | | | | |
| Zimbabwe | SHINE (HIV−) (33) | | | | | | | | | | | | |
| Zimbabwe | SHINE (HIV+) (34) | | | | | | | | | | | | |
| | | **664** | **680** | | **I² = 0.00, Tau² = 0.00** | | | **929** | **740** | | **I² = 0.35, Tau² = 0.01** | | |
| **Fixed** | | | | | **1.18 (1.05, 1.33)** | | | | | | **1.12 (0.98, 1.27)** | | |
| **Random** | | | | | **1.18 (1.05, 1.33)** | | | | | | **1.10 (0.94, 1.29)** | | |

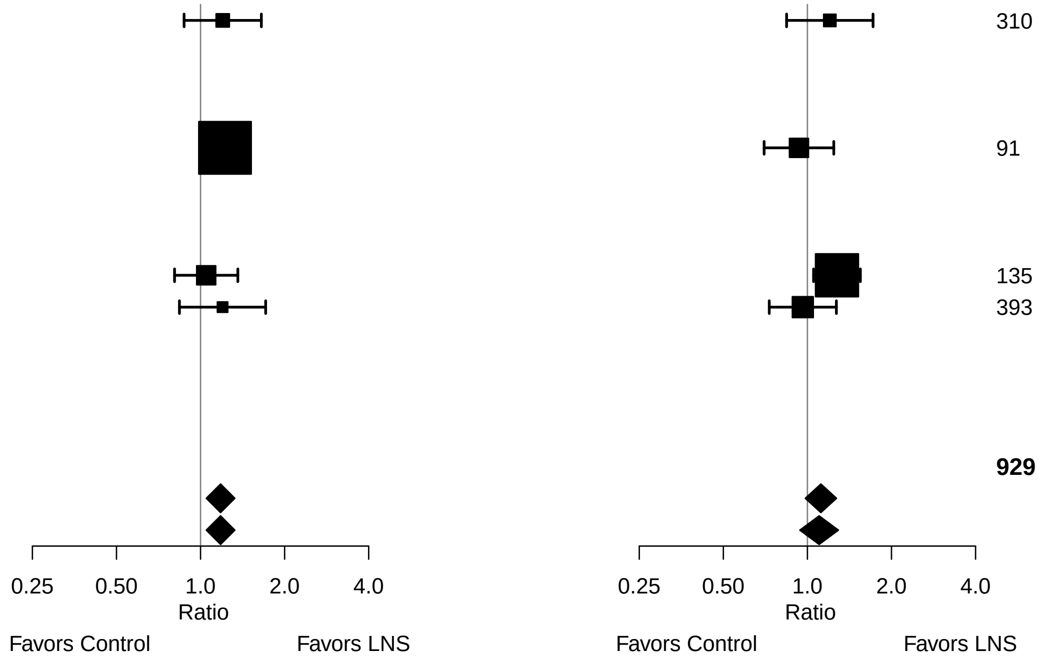

### Supplemental figure 5P: 12-mo walking without support prevalence difference

#### 5P1: Stratified by Maternal height

**P−for−interaction = 0.192**
**Difference in PDs = −0.04 (−0.09, 0.02)**

| Country | Trial | At least 150.1 cm: LNS N | Control N | Control Prevalence | PD (95% CI) | Fixed W | Random W | Less than 150.1 cm: LNS N | Control N | Control Prevalence | PD (95% CI) | Fixed W | Random W |
|---|---|---|---|---|---|---|---|---|---|---|---|---|---|
| Bangladesh | RDNS (22) | 848 | 418 | 74.4 | 0.11 (0.05, 0.16) | 0.28 | 0.28 | 721 | 347 | 76.4 | 0.04 (−0.03, 0.11) | 0.38 | 0.38 |
| Bangladesh | WASH−B (23) | 261 | 782 | 73.0 | 0.06 (0.00, 0.12) | 0.26 | 0.26 | 230 | 666 | 76.1 | 0.05 (−0.01, 0.11) | 0.51 | 0.51 |
| Burkina Faso | iLiNS−Zinc (24) | | | | | | | | | | | | |
| Ghana | GHANA (25) | | | | | | | | | | | | |
| Ghana | iLiNS−DYADG (26) | 308 | 613 | 52.9 | 0.06 (−0.01, 0.13) | 0.20 | 0.20 | 14 | 38 | 63.2 | 0.27 (−0.03, 0.58) | 0.02 | 0.02 |
| Haiti | HAITI (27) | | | | | | | | | | | | |
| Kenya | WASH−B (28) | | | | | | | | | | | | |
| Madagascar | MAHAY (29) | | | | | | | | | | | | |
| Malawi | iLiNS−DYADM (30) | 180 | 367 | 50.4 | 0.10 (0.01, 0.19) | 0.12 | 0.12 | 28 | 56 | 53.6 | 0.07 (−0.16, 0.30) | 0.04 | 0.04 |
| Malawi | iLiNS−DOSE (31) | 505 | 164 | 66.5 | 0.03 (−0.05, 0.12) | 0.13 | 0.13 | 96 | 35 | 62.9 | −0.06 (−0.24, 0.12) | 0.06 | 0.06 |
| Mali | PROMIS CS (32) | | | | | | | | | | | | |
| Zimbabwe | SHINE (HIV−) (33) | | | | | | | | | | | | |
| Zimbabwe | SHINE (HIV+) (34) | | | | | | | | | | | | |
| | | **2102** | **2344** | | **I² = 0.00, Tau² = 0.00** | | | **1089** | **1142** | | **I² = 0.00, Tau² = 0.00** | | |
| **Fixed** | | | | | **0.07 (0.04, 0.10)** | | | | | | **0.05 (0.00, 0.09)** | | |
| **Random** | | | | | **0.07 (0.04, 0.10)** | | | | | | **0.05 (0.00, 0.09)** | | |

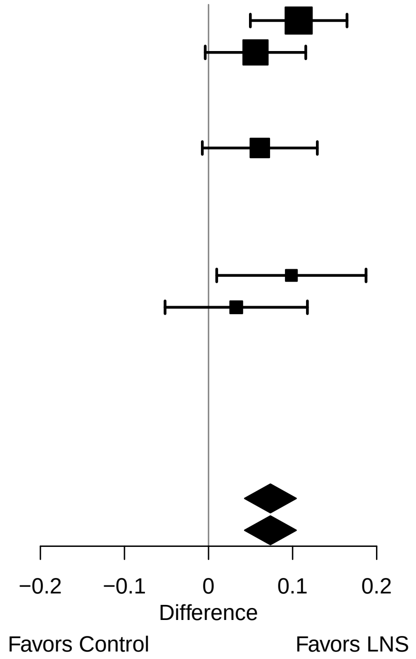

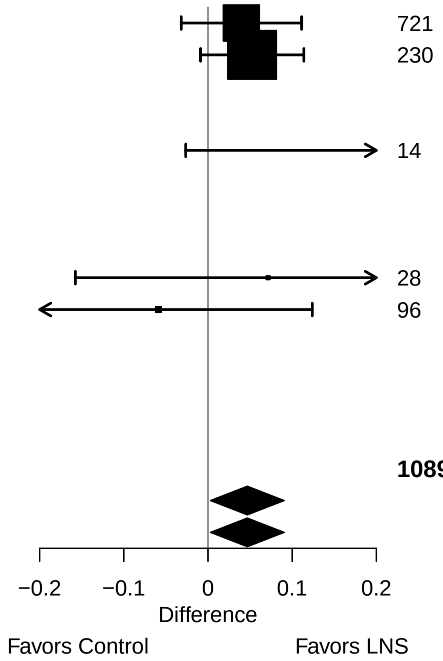

#### Supplemental figure 5P: 12-mo walking without support prevalence difference

### 5P2: Stratified by Maternal BMI

**P−for−interaction = 0.058**
**Difference in PDs = −0.05 (−0.09, 0.00)**

| | | At least 20 kg/m² | | | | | | Less than 20 kg/m² | | | | | |
|---|---|---|---|---|---|---|---|---|---|---|---|---|---|
| **Country** | **Trial** | **LNS N** | **Control N** | **Control Prevalence** | **PD (95% CI)** | **Fixed W** | **Random W** | **LNS N** | **Control N** | **Control Prevalence** | **PD (95% CI)** | **Fixed W** | **Random W** |
| Bangladesh | RDNS (22) | 708 | 327 | 74.6 | 0.14 (0.07, 0.22) | 0.15 | 0.15 | 861 | 438 | 75.8 | 0.02 (−0.03, 0.07) | 0.41 | 0.41 |
| Bangladesh | WASH−B (23) | 225 | 637 | 73.0 | 0.08 (0.01, 0.15) | 0.20 | 0.16 | 266 | 810 | 75.7 | 0.03 (−0.02, 0.09) | 0.33 | 0.33 |
| Burkina Faso | iLiNS−Zinc (24) | | | | | | | | | | | | |
| Ghana | GHANA (25) | | | | | | | | | | | | |
| Ghana | iLiNS−DYADG (26) | 284 | 535 | 52.3 | 0.07 (0.00, 0.14) | 0.17 | 0.16 | 38 | 116 | 58.6 | 0.03 (−0.15, 0.22) | 0.03 | 0.03 |
| Haiti | HAITI (27) | | | | | | | | | | | | |
| Kenya | WASH−B (28) | 334 | 1210 | 58.8 | −0.03 (−0.09, 0.02) | 0.26 | 0.17 | 96 | 364 | 61.5 | 0.01 (−0.10, 0.13) | 0.08 | 0.08 |
| Madagascar | MAHAY (29) | | | | | | | | | | | | |
| Malawi | iLiNS−DYADM (30) | 121 | 255 | 51.4 | 0.08 (−0.02, 0.19) | 0.07 | 0.12 | 86 | 168 | 50.0 | 0.12 (−0.01, 0.25) | 0.06 | 0.06 |
| Malawi | iLiNS−DOSE (31) | 451 | 138 | 63.0 | 0.00 (−0.09, 0.09) | 0.10 | 0.14 | 148 | 60 | 71.7 | 0.05 (−0.09, 0.20) | 0.05 | 0.05 |
| Mali | PROMIS CS (32) | 83 | 98 | 66.3 | −0.11 (−0.25, 0.03) | 0.04 | 0.10 | 34 | 52 | 67.3 | −0.06 (−0.26, 0.13) | 0.03 | 0.03 |
| Zimbabwe | SHINE (HIV−) (33) | | | | | | | | | | | | |
| Zimbabwe | SHINE (HIV+) (34) | | | | | | | | | | | | |
| | | **2206** | **3200** | | **I² = 0.72, Tau² = 0.00** | | | **1529** | **2008** | | **I² = 0.00, Tau² = 0.00** | | |
| **Fixed** | | | | | **0.04 (0.01, 0.07)** | | | | | | **0.03 (0.00, 0.06)** | | |
| **Random** | | | | | **0.04 (−0.02, 0.10)** | | | | | | **0.03 (0.00, 0.06)** | | |

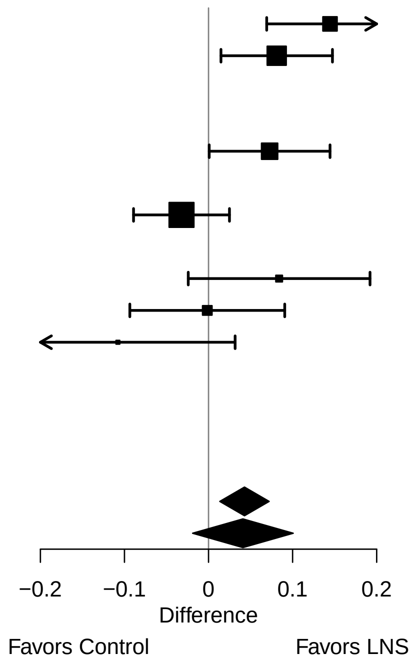

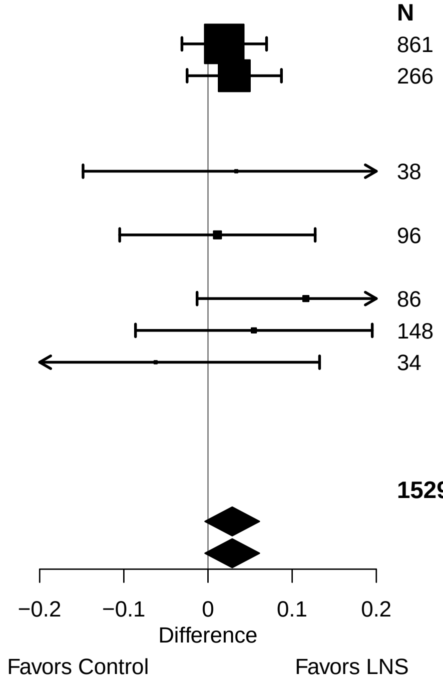

#### Supplemental figure 5P: 12-mo walking without support prevalence difference

### 5P3: Stratified by Maternal age

**P−for−interaction = 0.315**
**Difference in PDs = 0.02 (−0.02, 0.06)**

| | | At least 25 y | | | | | | Less than 25 y | | | | | |
|---|---|---|---|---|---|---|---|---|---|---|---|---|---|
| **Country** | **Trial** | **LNS N** | **Control N** | **Control Prevalence** | **PD (95% CI)** | **Fixed W** | **Random W** | **LNS N** | **Control N** | **Control Prevalence** | **PD (95% CI)** | **Fixed W** | **Random W** |
| Bangladesh | RDNS (22) | 447 | 217 | 76.0 | 0.09 (0.02, 0.16) | 0.19 | 0.17 | 1181 | 573 | 73.8 | 0.07 (0.01, 0.12) | 0.28 | 0.18 |
| Bangladesh | WASH−B (23) | 223 | 676 | 74.6 | −0.01 (−0.07, 0.05) | 0.25 | 0.18 | 274 | 791 | 74.6 | 0.10 (0.05, 0.16) | 0.30 | 0.19 |
| Burkina Faso | iLiNS−Zinc (24) | | | | | | | | | | | | |
| Ghana | GHANA (25) | | | | | | | | | | | | |
| Ghana | iLiNS−DYADG (26) | 209 | 405 | 53.8 | 0.04 (−0.05, 0.12) | 0.14 | 0.15 | 118 | 258 | 51.9 | 0.13 (0.02, 0.24) | 0.08 | 0.12 |
| Haiti | HAITI (27) | 60 | 57 | 68.4 | 0.18 (0.01, 0.36) | 0.03 | 0.06 | 22 | 22 | 50.0 | −0.14 (−0.43, 0.16) | 0.01 | 0.03 |
| Kenya | WASH−B (28) | 243 | 914 | 61.3 | 0.01 (−0.06, 0.08) | 0.21 | 0.17 | 197 | 679 | 57.7 | −0.04 (−0.11, 0.04) | 0.17 | 0.16 |
| Madagascar | MAHAY (29) | | | | | | | | | | | | |
| Malawi | iLiNS−DYADM (30) | 100 | 213 | 47.9 | 0.03 (−0.09, 0.15) | 0.07 | 0.10 | 108 | 213 | 54.5 | 0.16 (0.05, 0.28) | 0.07 | 0.11 |
| Malawi | iLiNS−DOSE (31) | 327 | 111 | 64.9 | 0.04 (−0.07, 0.14) | 0.09 | 0.11 | 264 | 83 | 67.5 | 0.00 (−0.12, 0.11) | 0.07 | 0.11 |
| Mali | PROMIS CS (32) | 59 | 71 | 62.0 | −0.16 (−0.32, 0.00) | 0.03 | 0.06 | 59 | 80 | 70.0 | −0.05 (−0.19, 0.10) | 0.04 | 0.09 |
| Zimbabwe | SHINE (HIV−) (33) | | | | | | | | | | | | |
| Zimbabwe | SHINE (HIV+) (34) | | | | | | | | | | | | |
| | | **1668** | **2664** | | **I² = 0.38, Tau² = 0.00** | | | **2223** | **2699** | | **I² = 0.61, Tau² = 0.00** | | |
| **Fixed** | | | | | **0.03 (0.00, 0.05)** | | | | | | **0.05 (0.02, 0.07)** | | |
| **Random** | | | | | **0.03 (−0.02, 0.07)** | | | | | | **0.05 (0.00, 0.10)** | | |

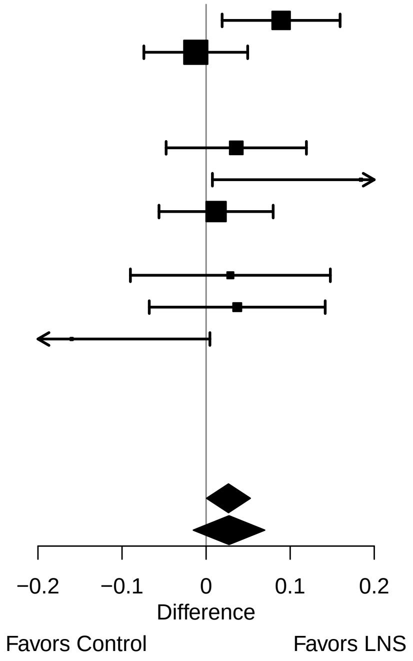

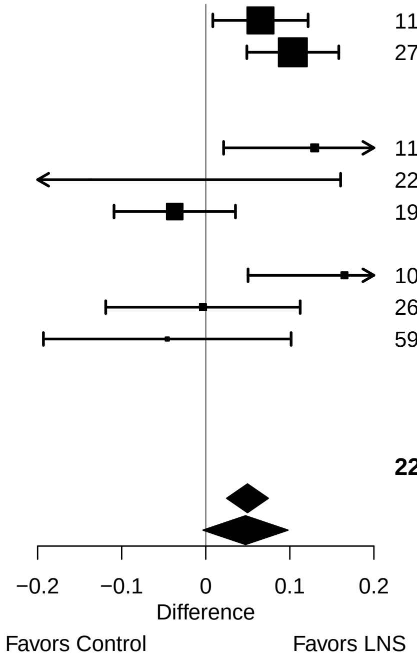

#### Supplemental figure 5P: 12-mo walking without support prevalence difference

#### 5P4: Stratified by Maternal education

**P−for−interaction = 0.291**
**Difference in PDs = −0.02 (−0.06, 0.02)**

| | | Primary or greater | | | | | | Incomplete or no formal | | | | | |
|---|---|---|---|---|---|---|---|---|---|---|---|---|---|
| **Country** | **Trial** | **LNS N** | **Control N** | **Control Prevalence** | **PD (95% CI)** | **Fixed W** | **Random W** | **LNS N** | **Control N** | **Control Prevalence** | **PD (95% CI)** | **Fixed W** | **Random W** |
| Bangladesh | RDNS (22) | 1219 | 574 | 73.9 | 0.08 (0.03, 0.14) | 0.27 | 0.27 | 409 | 216 | 75.9 | 0.03 (−0.02, 0.09) | 0.29 | 0.24 |
| Bangladesh | WASH−B (23) | 337 | 1048 | 72.7 | 0.08 (0.03, 0.13) | 0.37 | 0.37 | 160 | 426 | 79.1 | 0.00 (−0.06, 0.07) | 0.24 | 0.22 |
| Burkina Faso | iLiNS−Zinc (24) | | | | | | | | | | | | |
| Ghana | GHANA (25) | | | | | | | | | | | | |
| Ghana | iLiNS−DYADG (26) | 257 | 524 | 51.5 | 0.06 (−0.01, 0.13) | 0.16 | 0.16 | 70 | 139 | 59.0 | 0.10 (−0.04, 0.25) | 0.05 | 0.07 |
| Haiti | HAITI (27) | | | | | | | | | | | | |
| Kenya | WASH−B (28) | 217 | 775 | 59.2 | 0.02 (−0.06, 0.09) | 0.15 | 0.15 | 229 | 827 | 60.3 | −0.04 (−0.12, 0.03) | 0.17 | 0.18 |
| Madagascar | MAHAY (29) | | | | | | | | | | | | |
| Malawi | iLiNS−DYADM (30) | 32 | 64 | 42.2 | −0.02 (−0.23, 0.20) | 0.02 | 0.02 | 175 | 359 | 52.4 | 0.11 (0.02, 0.20) | 0.12 | 0.14 |
| Malawi | iLiNS−DOSE (31) | 129 | 42 | 61.9 | 0.03 (−0.14, 0.20) | 0.03 | 0.03 | 462 | 152 | 67.1 | 0.02 (−0.07, 0.10) | 0.13 | 0.15 |
| Mali | PROMIS CS (32) | | | | | | | | | | | | |
| Zimbabwe | SHINE (HIV−) (33) | | | | | | | | | | | | |
| Zimbabwe | SHINE (HIV+) (34) | | | | | | | | | | | | |
| | | **2191** | **3027** | | **I² = 0.00, Tau² = 0.00** | | | **1505** | **2119** | | **I² = 0.30, Tau² = 0.00** | | |
| **Fixed** | | | | | **0.05 (0.03, 0.07)** | | | | | | **0.03 (0.00, 0.05)** | | |
| **Random** | | | | | **0.05 (0.03, 0.07)** | | | | | | **0.03 (−0.01, 0.06)** | | |

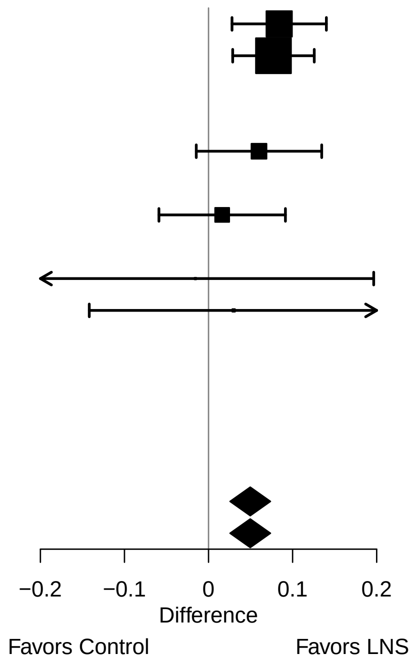

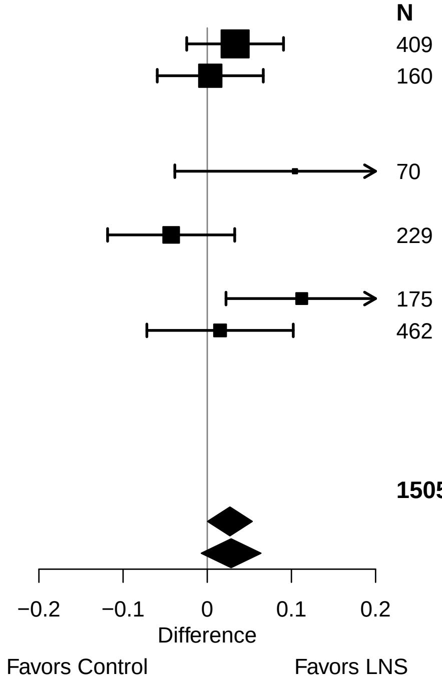

#### Supplemental figure 5P: 12-mo walking without support prevalence difference

##### 5P5: Stratified by Maternal depressive symptoms

**P−for−interaction = 0.606**
**Difference in PDs = −0.01 (−0.06, 0.04)**

| | | Less than 75th percentile | | | | | | At least 75th percentile | | | | | |
|---|---|---|---|---|---|---|---|---|---|---|---|---|---|
| **Country** | **Trial** | **LNS N** | **Control N** | **Control Prevalence** | **PD (95% CI)** | **Fixed W** | **Random W** | **LNS N** | **Control N** | **Control Prevalence** | **PD (95% CI)** | **Fixed W** | **Random W** |
| Bangladesh | RDNS (22) | 1058 | 460 | 70.9 | 0.03 (−0.03, 0.10) | 0.20 | 0.21 | 508 | 279 | 79.6 | 0.14 (0.07, 0.20) | 0.42 | 0.29 |
| Bangladesh | WASH−B (23) | 389 | 1075 | 73.4 | 0.07 (0.02, 0.12) | 0.33 | 0.28 | 108 | 399 | 77.7 | −0.04 (−0.12, 0.05) | 0.26 | 0.25 |
| Burkina Faso | iLiNS−Zinc (24) | | | | | | | | | | | | |
| Ghana | GHANA (25) | | | | | | | | | | | | |
| Ghana | iLiNS−DYADG (26) | 242 | 446 | 52.9 | 0.07 (0.00, 0.15) | 0.14 | 0.16 | 75 | 198 | 55.1 | 0.07 (−0.06, 0.20) | 0.10 | 0.16 |
| Haiti | HAITI (27) | | | | | | | | | | | | |
| Kenya | WASH−B (28) | 314 | 1098 | 59.6 | −0.01 (−0.07, 0.05) | 0.24 | 0.23 | 99 | 373 | 61.1 | 0.01 (−0.10, 0.11) | 0.16 | 0.20 |
| Madagascar | MAHAY (29) | | | | | | | | | | | | |
| Malawi | iLiNS−DYADM (30) | 149 | 293 | 48.1 | 0.11 (0.01, 0.20) | 0.09 | 0.12 | 47 | 108 | 60.2 | 0.09 (−0.08, 0.26) | 0.06 | 0.11 |
| Malawi | iLiNS−DOSE (31) | | | | | | | | | | | | |
| Mali | PROMIS CS (32) | | | | | | | | | | | | |
| Zimbabwe | SHINE (HIV−) (33) | | | | | | | | | | | | |
| Zimbabwe | SHINE (HIV+) (34) | | | | | | | | | | | | |
| | | **2152** | **3372** | | **I² = 0.35, Tau² = 0.00** | | | **837** | **1357** | | **I² = 0.65, Tau² = 0.00** | | |
| **Fixed** | | | | | **0.05 (0.02, 0.08)** | | | | | | **0.06 (0.02, 0.10)** | | |
| **Random** | | | | | **0.05 (0.01, 0.09)** | | | | | | **0.05 (−0.01, 0.12)** | | |

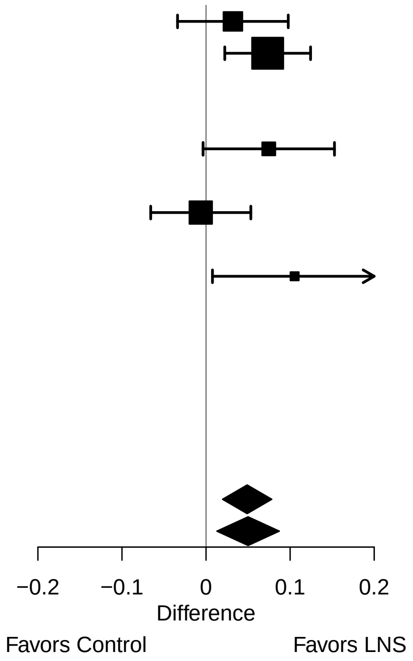

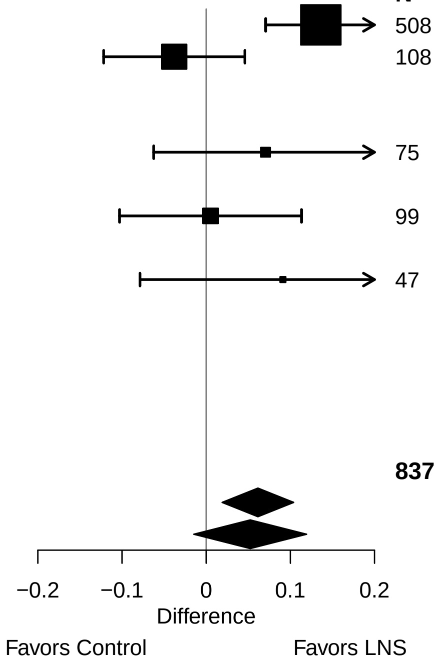

**Supplemental figure 5P: 12-mo walking without support prevalence difference**

#### 5P6: Stratified by Child sex

**P−for−interaction = 0.816**
**Difference in PDs = 0.00 (−0.03, 0.04)**

| Country | Trial | Male LNS N | Male Control N | Male Control Prevalence | Male PD (95% CI) | Male Fixed W | Male Random W | Female LNS N | Female Control N | Female Control Prevalence | Female PD (95% CI) | Female Fixed W | Female Random W |
|---|---|---|---|---|---|---|---|---|---|---|---|---|---|
| Bangladesh | RDNS (22) | 819 | 396 | 74.5 | 0.04 (−0.02, 0.10) | 0.28 | 0.21 | 801 | 394 | 74.4 | 0.10 (0.04, 0.17) | 0.22 | 0.14 |
| Bangladesh | WASH−B (23) | 232 | 692 | 75.9 | 0.06 (0.00, 0.13) | 0.23 | 0.19 | 265 | 782 | 73.4 | 0.04 (−0.02, 0.10) | 0.27 | 0.14 |
| Burkina Faso | iLiNS−Zinc (24) | | | | | | | | | | | | |
| Ghana | GHANA (25) | 55 | 40 | 67.5 | 0.09 (−0.11, 0.29) | 0.02 | 0.04 | 35 | 47 | 80.9 | 0.24 (0.04, 0.43) | 0.02 | 0.08 |
| Ghana | iLiNS−DYADG (26) | 166 | 321 | 53.6 | 0.07 (−0.02, 0.17) | 0.11 | 0.12 | 160 | 341 | 52.8 | 0.07 (−0.03, 0.16) | 0.11 | 0.12 |
| Haiti | HAITI (27) | 38 | 29 | 69.0 | 0.06 (−0.17, 0.29) | 0.02 | 0.03 | 45 | 50 | 60.0 | 0.13 (−0.07, 0.33) | 0.02 | 0.07 |
| Kenya | WASH−B (28) | 231 | 764 | 62.7 | −0.01 (−0.09, 0.07) | 0.15 | 0.15 | 216 | 840 | 57.0 | −0.01 (−0.09, 0.06) | 0.18 | 0.13 |
| Madagascar | MAHAY (29) | | | | | | | | | | | | |
| Malawi | iLiNS−DYADM (30) | 105 | 198 | 57.6 | 0.20 (0.09, 0.32) | 0.07 | 0.09 | 103 | 228 | 45.6 | 0.00 (−0.12, 0.12) | 0.07 | 0.11 |
| Malawi | iLiNS−DOSE (31) | 310 | 111 | 73.9 | 0.10 (0.00, 0.21) | 0.09 | 0.11 | 292 | 89 | 55.1 | −0.09 (−0.21, 0.02) | 0.07 | 0.11 |
| Mali | PROMIS CS (32) | 58 | 83 | 71.1 | 0.02 (−0.14, 0.18) | 0.04 | 0.06 | 60 | 68 | 60.3 | −0.23 (−0.38, −0.08) | 0.04 | 0.10 |
| Zimbabwe | SHINE (HIV−) (33) | | | | | | | | | | | | |
| Zimbabwe | SHINE (HIV+) (34) | | | | | | | | | | | | |
| | | **2014** | **2634** | | **$I^2$ = 0.41, $Tau^2$ = 0.00** | | | **1977** | **2839** | | **$I^2$ = 0.70, $Tau^2$ = 0.01** | | |
| **Fixed** | | | | | **0.04 (0.02, 0.07)** | | | | | | **0.04 (0.01, 0.06)** | | |
| **Random** | | | | | **0.05 (0.02, 0.09)** | | | | | | **0.03 (−0.05, 0.10)** | | |

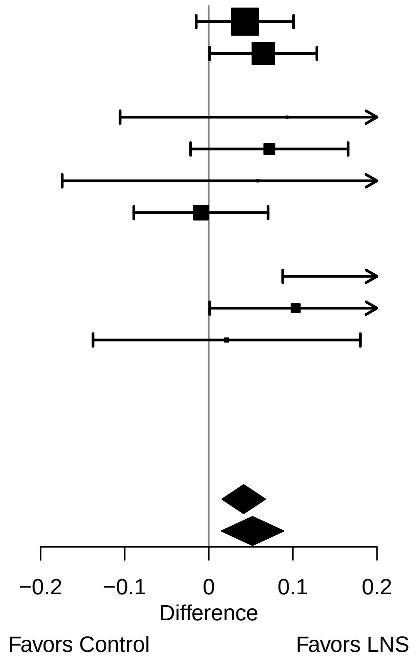

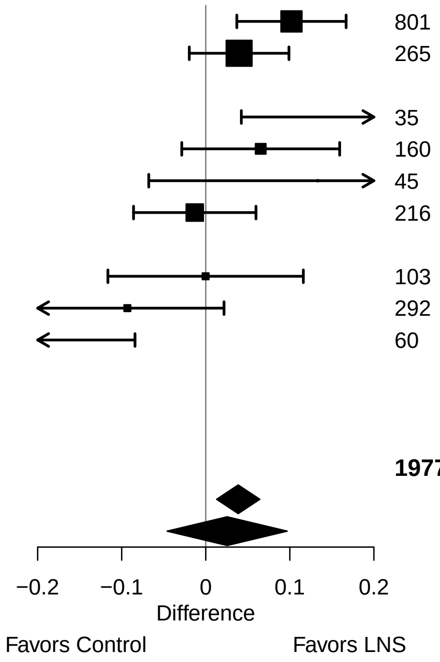

**Supplemental figure 5P: 12-mo walking without support prevalence difference**

#### 5P7: Stratified by Child birth order

**P−for−interaction = 0.055**
**Difference in PDs = −0.04 (−0.08, 0.00)**

Later born

| Country | Trial | LNS N | Control N | Control Prevalence | PD (95% CI) | Fixed W | Random W |
|---|---|---|---|---|---|---|---|
| Bangladesh | RDNS (22) | 953 | 496 | 75.6 | 0.09 (0.04, 0.14) | 0.25 | 0.17 |
| Bangladesh | WASH−B (23) | 314 | 944 | 74.8 | 0.04 (−0.02, 0.09) | 0.22 | 0.17 |
| Burkina Faso | iLiNS−Zinc (24) | | | | | | |
| Ghana | GHANA (25) | 54 | 47 | 83.0 | 0.18 (0.01, 0.35) | 0.02 | 0.05 |
| Ghana | iLiNS−DYADG (26) | 222 | 439 | 54.9 | 0.09 (0.01, 0.17) | 0.11 | 0.13 |
| Haiti | HAITI (27) | 57 | 50 | 68.0 | 0.14 (−0.05, 0.32) | 0.02 | 0.04 |
| Kenya | WASH−B (28) | 338 | 1296 | 60.2 | 0.01 (−0.05, 0.07) | 0.19 | 0.16 |
| Madagascar | MAHAY (29) | | | | | | |
| Malawi | iLiNS−DYADM (30) | 166 | 340 | 50.6 | 0.08 (−0.01, 0.18) | 0.08 | 0.11 |
| Malawi | iLiNS−DOSE (31) | 388 | 139 | 67.6 | 0.07 (−0.03, 0.16) | 0.08 | 0.11 |
| Mali | PROMIS CS (32) | 103 | 134 | 65.7 | −0.12 (−0.25, 0.01) | 0.04 | 0.07 |
| Zimbabwe | SHINE (HIV−) (33) | | | | | | |
| Zimbabwe | SHINE (HIV+) (34) | | | | | | |
| | | **2595** | **3885** | | **I² = 0.42, Tau² = 0.00** | | |
| **Fixed** | | | | | **0.06 (0.03, 0.08)** | | |
| **Random** | | | | | **0.06 (0.02, 0.10)** | | |

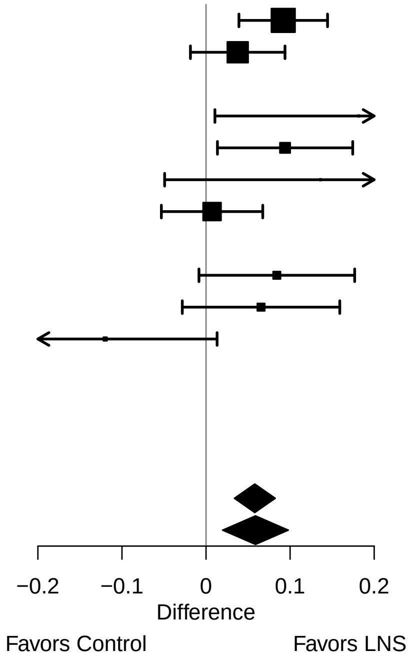

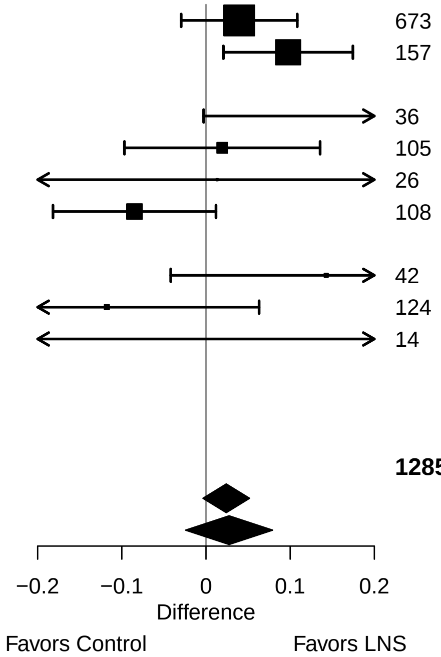

Firstborn

| Country | Trial | LNS N | Control N | Control Prevalence | PD (95% CI) | Fixed W | Random W |
|---|---|---|---|---|---|---|---|
| Bangladesh | RDNS (22) | 673 | 294 | 72.4 | 0.04 (−0.03, 0.11) | 0.32 | 0.22 |
| Bangladesh | WASH−B (23) | 157 | 440 | 74.1 | 0.10 (0.02, 0.17) | 0.26 | 0.20 |
| Burkina Faso | iLiNS−Zinc (24) | | | | | | |
| Ghana | GHANA (25) | 36 | 36 | 69.4 | 0.22 (0.00, 0.45) | 0.03 | 0.06 |
| Ghana | iLiNS−DYADG (26) | 105 | 224 | 49.6 | 0.02 (−0.10, 0.14) | 0.11 | 0.14 |
| Haiti | HAITI (27) | 26 | 29 | 55.2 | 0.01 (−0.26, 0.28) | 0.02 | 0.04 |
| Kenya | WASH−B (28) | 108 | 306 | 58.2 | −0.08 (−0.18, 0.01) | 0.16 | 0.17 |
| Madagascar | MAHAY (29) | | | | | | |
| Malawi | iLiNS−DYADM (30) | 42 | 84 | 52.4 | 0.14 (−0.04, 0.33) | 0.04 | 0.08 |
| Malawi | iLiNS−DOSE (31) | 124 | 33 | 57.6 | −0.12 (−0.30, 0.06) | 0.05 | 0.08 |
| Mali | PROMIS CS (32) | 14 | 14 | 64.3 | 0.00 (−0.34, 0.34) | 0.01 | 0.03 |
| Zimbabwe | SHINE (HIV−) (33) | | | | | | |
| Zimbabwe | SHINE (HIV+) (34) | | | | | | |
| | | **1285** | **1460** | | **I² = 0.43, Tau² = 0.00** | | |
| **Fixed** | | | | | **0.02 (0.00, 0.05)** | | |
| **Random** | | | | | **0.03 (−0.02, 0.08)** | | |

**Supplemental figure 5P: 12-mo walking without support prevalence difference**

#### 5P8: Stratified by Child baseline stunting

**P−for−interaction = 0.517**
**Difference in PDs = 0.02 (−0.03, 0.07)**

| | | No | | | | | | Yes | | | | | |
|---|---|---|---|---|---|---|---|---|---|---|---|---|---|
| **Country** | **Trial** | **LNS N** | **Control N** | **Control Prevalence** | **PD (95% CI)** | **Fixed W** | **Random W** | **LNS N** | **Control N** | **Control Prevalence** | **PD (95% CI)** | **Fixed W** | **Random W** |
| Bangladesh | RDNS (22) | 1219 | 559 | 72.1 | 0.06 (0.01, 0.12) | 0.46 | 0.35 | 344 | 179 | 80.4 | 0.09 (0.00, 0.18) | 0.51 | 0.51 |
| Bangladesh | WASH−B (23) | | | | | | | | | | | | |
| Burkina Faso | iLiNS−Zinc (24) | | | | | | | | | | | | |
| Ghana | GHANA (25) | | | | | | | | | | | | |
| Ghana | iLiNS−DYADG (26) | 277 | 557 | 52.1 | 0.07 (0.00, 0.14) | 0.26 | 0.27 | 28 | 62 | 69.4 | 0.05 (−0.16, 0.26) | 0.09 | 0.09 |
| Haiti | HAITI (27) | | | | | | | | | | | | |
| Kenya | WASH−B (28) | | | | | | | | | | | | |
| Madagascar | MAHAY (29) | | | | | | | | | | | | |
| Malawi | iLiNS−DYADM (30) | 148 | 317 | 47.6 | 0.11 (0.01, 0.21) | 0.14 | 0.19 | 50 | 93 | 63.4 | 0.03 (−0.13, 0.20) | 0.14 | 0.14 |
| Malawi | iLiNS−DOSE (31) | 424 | 133 | 57.1 | −0.04 (−0.14, 0.05) | 0.15 | 0.19 | 178 | 67 | 82.1 | 0.12 (0.00, 0.25) | 0.26 | 0.26 |
| Mali | PROMIS CS (32) | | | | | | | | | | | | |
| Zimbabwe | SHINE (HIV−) (33) | | | | | | | | | | | | |
| Zimbabwe | SHINE (HIV+) (34) | | | | | | | | | | | | |
| | | **2068** | **1566** | | **I² = 0.38, Tau² = 0.00** | | | **600** | **401** | | **I² = 0.00, Tau² = 0.00** | | |
| **Fixed** | | | | | **0.05 (0.02, 0.07)** | | | | | | **0.06 (0.01, 0.10)** | | |
| **Random** | | | | | **0.05 (0.01, 0.09)** | | | | | | **0.06 (0.01, 0.10)** | | |

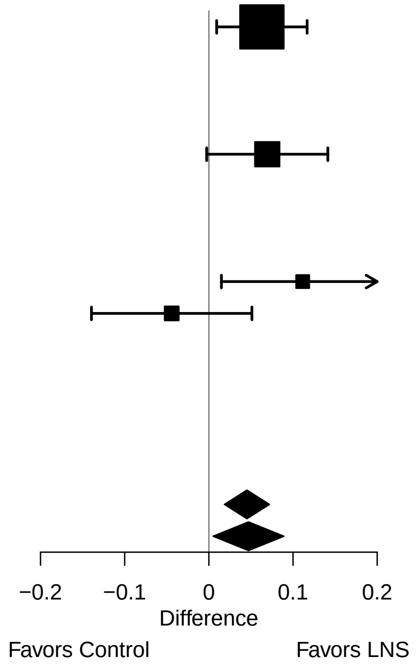

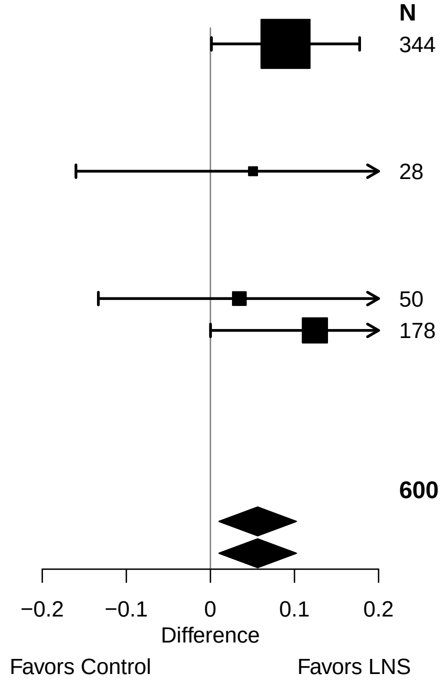

**Supplemental figure 5P: 12-mo walking without support prevalence difference**

#### 5P9: Stratified by Child baseline acute malnutrition

**P−for−interaction = 0.170**
**Difference in PDs = 0.04 (−0.02, 0.11)**

| | | No | | | | | | Yes | | | | | |
|---|---|---|---|---|---|---|---|---|---|---|---|---|---|
| **Country** | **Trial** | **LNS N** | **Control N** | **Control Prevalence** | **PD (95% CI)** | **Fixed W** | **Random W** | **LNS N** | **Control N** | **Control Prevalence** | **PD (95% CI)** | **Fixed W** | **Random W** |
| Bangladesh | RDNS (22) | 1447 | 689 | 73.0 | 0.07 (0.02, 0.12) | 0.54 | 0.54 | 119 | 50 | 90.0 | 0.09 (−0.01, 0.20) | 0.73 | 0.48 |
| Bangladesh | WASH−B (23) | | | | | | | | | | | | |
| Burkina Faso | iLiNS−Zinc (24) | | | | | | | | | | | | |
| Ghana | GHANA (25) | | | | | | | | | | | | |
| Ghana | iLiNS−DYADG (26) | 284 | 564 | 50.7 | 0.04 (−0.03, 0.11) | 0.28 | 0.28 | 23 | 57 | 84.2 | 0.36 (0.16, 0.56) | 0.19 | 0.32 |
| Haiti | HAITI (27) | | | | | | | | | | | | |
| Kenya | WASH−B (28) | | | | | | | | | | | | |
| Madagascar | MAHAY (29) | | | | | | | | | | | | |
| Malawi | iLiNS−DYADM (30) | 184 | 374 | 50.3 | 0.08 (0.00, 0.17) | 0.18 | 0.18 | 14 | 39 | 61.5 | 0.12 (−0.19, 0.42) | 0.08 | 0.20 |
| Malawi | iLiNS−DOSE (31) | | | | | | | | | | | | |
| Mali | PROMIS CS (32) | | | | | | | | | | | | |
| Zimbabwe | SHINE (HIV−) (33) | | | | | | | | | | | | |
| Zimbabwe | SHINE (HIV+) (34) | | | | | | | | | | | | |
| | | **1915** | **1627** | | **I² = 0.00, Tau² = 0.00** | | | **156** | **146** | | **I² = 0.64, Tau² = 0.01** | | |
| **Fixed** | | | | | **0.05 (0.02, 0.07)** | | | | | | **0.09 (0.03, 0.15)** | | |
| **Random** | | | | | **0.05 (0.02, 0.07)** | | | | | | **0.14 (0.00, 0.27)** | | |

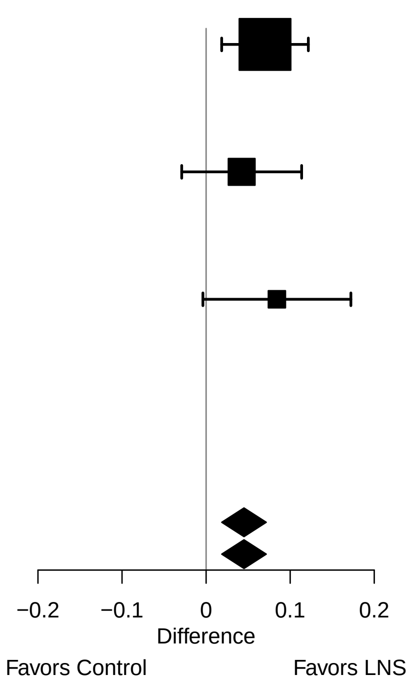

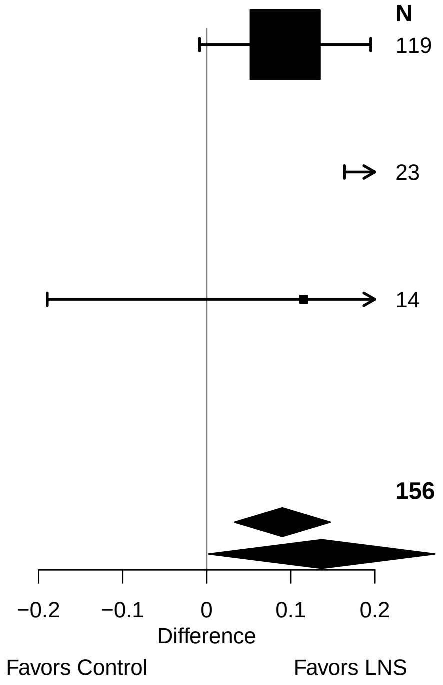

#### Supplemental figure 5P: 12-mo walking without support prevalence difference

### 5P10: Stratified by Child baseline anemia

**P−for−interaction = 0.330**
**Difference in PDs = −0.04 (−0.12, 0.04)**

| | | Not anemic | | | | | | Anemic | | | | | |
|---|---|---|---|---|---|---|---|---|---|---|---|---|---|
| **Country** | **Trial** | **LNS N** | **Control N** | **Control Prevalence** | **PD (95% CI)** | **Fixed W** | **Random W** | **LNS N** | **Control N** | **Control Prevalence** | **PD (95% CI)** | **Fixed W** | **Random W** |
| Bangladesh | RDNS (22) | 211 | 95 | 71.6 | 0.06 (−0.04, 0.15) | 0.31 | 0.31 | 310 | 157 | 74.5 | 0.05 (−0.04, 0.15) | 0.29 | 0.27 |
| Bangladesh | WASH−B (23) | | | | | | | | | | | | |
| Burkina Faso | iLiNS−Zinc (24) | | | | | | | | | | | | |
| Ghana | GHANA (25) | | | | | | | | | | | | |
| Ghana | iLiNS−DYADG (26) | 185 | 358 | 51.4 | 0.11 (0.02, 0.20) | 0.37 | 0.37 | 91 | 201 | 55.2 | −0.03 (−0.15, 0.09) | 0.18 | 0.20 |
| Haiti | HAITI (27) | | | | | | | | | | | | |
| Kenya | WASH−B (28) | | | | | | | | | | | | |
| Madagascar | MAHAY (29) | | | | | | | | | | | | |
| Malawi | iLiNS−DYADM (30) | 63 | 143 | 45.5 | 0.03 (−0.12, 0.17) | 0.13 | 0.13 | 135 | 266 | 54.1 | 0.13 (0.02, 0.23) | 0.25 | 0.25 |
| Malawi | iLiNS−DOSE (31) | 205 | 84 | 67.9 | 0.06 (−0.06, 0.19) | 0.19 | 0.19 | 393 | 116 | 63.8 | −0.01 (−0.11, 0.09) | 0.28 | 0.27 |
| Mali | PROMIS CS (32) | | | | | | | | | | | | |
| Zimbabwe | SHINE (HIV−) (33) | | | | | | | | | | | | |
| Zimbabwe | SHINE (HIV+) (34) | | | | | | | | | | | | |
| | | **664** | **680** | | **I² = 0.00, Tau² = 0.00** | | | **929** | **740** | | **I² = 0.41, Tau² = 0.00** | | |
| **Fixed** | | | | | **0.07 (0.02, 0.13)** | | | | | | **0.04 (−0.01, 0.09)** | | |
| **Random** | | | | | **0.07 (0.02, 0.13)** | | | | | | **0.04 (−0.03, 0.11)** | | |

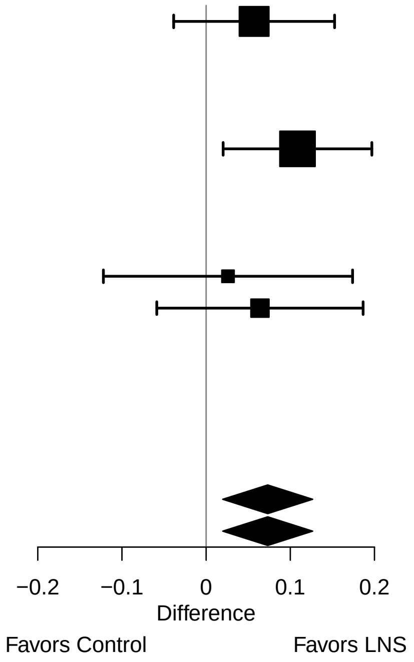

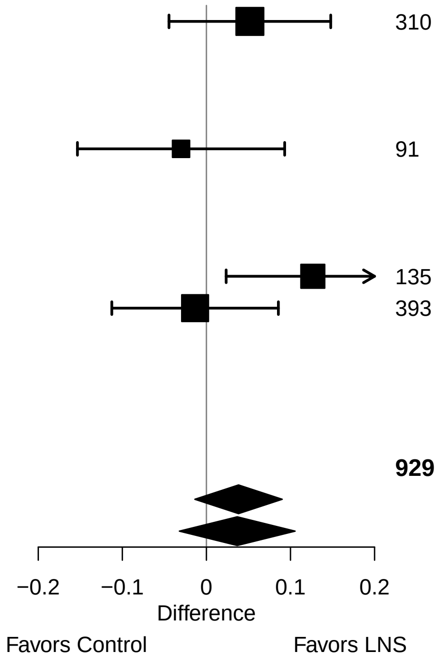

### Supplemental figure 6: Forest plots for effects of SQ-LNS on developmental outcomes stratified by all individual-level household effect modifiers

These figures are forest plots showing the individual-level effect modification of intervention effects. Each figure has the estimates of intervention effect stratified within study by individual-level effect modifier category. For dichotomous outcomes analyzed via prevalence ratios, the effect estimate is the prevalence in the LNS group divided by the prevalence in the control group. For dichotomous outcomes analyzed via prevalence differences, the effect estimate is the prevalence in the LNS group minus the prevalence in the control group. The labels on the far left correspond to trial level information. In the middle left and on the right the values indicate the study level effect estimate, confidence interval, and weighting for deriving the pooled estimates is shown by subgroup.

Motor milestone figures show individual trial estimates excluding the JiVitA-4 data because those individual trial results have not yet been published. The pooled estimate in each figure includes the JiVitA-4 data, for consistency with all other analyses.

### Supplemental figure 6A: Mean difference in language z-score

#### 6A1: Stratified by Household socio-economic status

**P−for−interaction = 0.001**
**Difference in MDs = 0.10 (0.04, 0.15)**

**At least median**

| Country | Trial | Tool | LNS N | Control N | Control Mean | MD (95% CI) | Fixed W | Random W |
|---|---|---|---|---|---|---|---|---|
| Bangladesh | JiVitA−4 (21) | BSID−III | 236 | 77 | 0.31 | −0.19 (−0.46, 0.08) | 0.02 | 0.05 |
| Bangladesh | RDNS (22) | CDI | 834 | 407 | 0.02 | 0.17 (0.01, 0.32) | 0.07 | 0.09 |
| Bangladesh | WASH−B (23) | CDI | 568 | 1703 | 0.15 | 0.07 (−0.02, 0.16) | 0.21 | 0.13 |
| Burkina Faso | iLiNS−Zinc (24) | DMC | 459 | 163 | −0.12 | 0.24 (0.08, 0.41) | 0.06 | 0.09 |
| Ghana | GHANA (25) | | | | | | | |
| Ghana | iLiNS−DYADG (26) | CDI | 152 | 341 | 0.07 | 0.08 (−0.11, 0.27) | 0.05 | 0.08 |
| Haiti | HAITI (27) | Total sounds | 76 | 60 | −0.13 | 0.14 (−0.19, 0.46) | 0.02 | 0.04 |
| Kenya | WASH−B (28) | EASQ | 778 | 2706 | 0.09 | −0.05 (−0.13, 0.02) | 0.31 | 0.14 |
| Madagascar | MAHAY (29) | ASQI | 741 | 845 | 0.32 | −0.15 (−0.34, 0.04) | 0.05 | 0.08 |
| Malawi | iLiNS−DYADM (30) | CDI | 118 | 230 | 0.09 | −0.06 (−0.28, 0.16) | 0.04 | 0.06 |
| Malawi | iLiNS−DOSE (31) | CDI | 278 | 94 | 0.04 | −0.01 (−0.25, 0.23) | 0.03 | 0.06 |
| Mali | PROMIS CS (32) | DMC | 494 | 450 | 0.03 | 0.05 (−0.14, 0.25) | 0.04 | 0.07 |
| Zimbabwe | SHINE (HIV−) (33) | CDI | 396 | 351 | 0.03 | 0.06 (−0.09, 0.21) | 0.08 | 0.10 |
| Zimbabwe | SHINE (HIV+) (34) | CDI | 86 | 77 | −0.10 | 0.32 (−0.01, 0.65) | 0.02 | 0.04 |
| | | | **5216** | **7504** | | **I² = 0.53, Tau² = 0.01** | | |
| **Fixed** | | | | | | **0.03 (−0.01, 0.07)** | | |
| **Random** | | | | | | **0.05 (−0.03, 0.12)** | | |

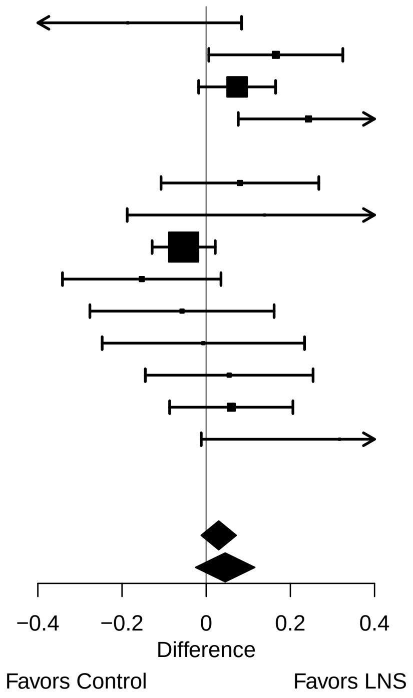

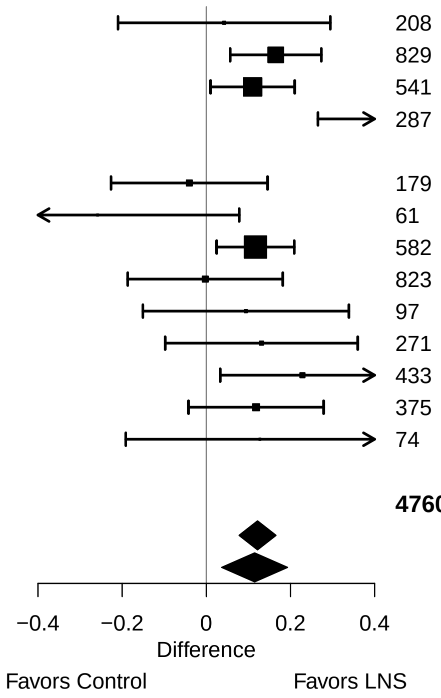

**Less than median**

| LNS N | Control N | Control Mean | MD (95% CI) | Fixed W | Random W |
|---|---|---|---|---|---|
| 208 | 66 | −0.23 | 0.04 (−0.21, 0.29) | 0.03 | 0.06 |
| 829 | 407 | −0.24 | 0.17 (0.06, 0.27) | 0.16 | 0.11 |
| 541 | 1650 | −0.20 | 0.11 (0.01, 0.21) | 0.19 | 0.11 |
| 287 | 212 | −0.34 | 0.47 (0.27, 0.67) | 0.05 | 0.07 |
| 179 | 315 | −0.08 | −0.04 (−0.23, 0.15) | 0.06 | 0.08 |
| 61 | 72 | 0.09 | −0.26 (−0.59, 0.08) | 0.02 | 0.04 |
| 582 | 2036 | −0.13 | 0.12 (0.02, 0.21) | 0.23 | 0.12 |
| 823 | 724 | −0.25 | 0.00 (−0.19, 0.18) | 0.06 | 0.08 |
| 97 | 208 | −0.11 | 0.09 (−0.15, 0.34) | 0.03 | 0.06 |
| 271 | 96 | −0.12 | 0.13 (−0.10, 0.36) | 0.04 | 0.06 |
| 433 | 494 | −0.17 | 0.23 (0.03, 0.42) | 0.05 | 0.07 |
| 375 | 383 | −0.12 | 0.12 (−0.04, 0.28) | 0.07 | 0.09 |
| 74 | 64 | −0.14 | 0.13 (−0.19, 0.45) | 0.02 | 0.04 |
| **4760** | **6727** | | **I² = 0.48, Tau² = 0.01** | | |
| | | | **0.12 (0.08, 0.17)** | | |
| | | | **0.11 (0.04, 0.19)** | | |

#### Supplemental figure 6A: Mean difference in language z-score

#### 6A2: Stratified by Household food insecurity

**P−for−interaction = 0.627**
**Difference in MDs = 0.02 (−0.05, 0.09)**

| Country | Trial | Tool | LNS N | Control N | Control Mean | Mild to secure MD (95% CI) | Fixed W | Random W | LNS N | Control N | Control Mean | Moderate to severe MD (95% CI) | Fixed W | Random W |
|---|---|---|---|---|---|---|---|---|---|---|---|---|---|---|
| Bangladesh | JiVitA−4 (21) | BSID−III | 329 | 111 | 0.16 | −0.15 (−0.37, 0.08) | 0.03 | 0.08 | 115 | 32 | −0.28 | 0.16 (−0.12, 0.44) | 0.05 | 0.06 |
| Bangladesh | RDNS (22) | CDI | 1058 | 493 | −0.02 | 0.14 (0.01, 0.27) | 0.09 | 0.11 | 605 | 321 | −0.25 | 0.19 (0.05, 0.32) | 0.20 | 0.16 |
| Bangladesh | WASH−B (23) | CDI | 883 | 2608 | 0.03 | 0.11 (0.03, 0.19) | 0.24 | 0.12 | 226 | 745 | −0.22 | 0.00 (−0.16, 0.17) | 0.13 | 0.12 |
| Burkina Faso | iLiNS−Zinc (24) | DMC | 389 | 162 | −0.22 | 0.38 (0.17, 0.58) | 0.03 | 0.09 | 357 | 213 | −0.27 | 0.35 (0.18, 0.52) | 0.13 | 0.12 |
| Ghana | GHANA (25) | | | | | | | | | | | | | |
| Ghana | iLiNS−DYADG (26) | CDI | 238 | 453 | 0.00 | 0.00 (−0.15, 0.16) | 0.06 | 0.10 | 92 | 202 | 0.00 | 0.02 (−0.24, 0.28) | 0.05 | 0.07 |
| Haiti | HAITI (27) | Total sounds | | | | | | | | | | | | |
| Kenya | WASH−B (28) | EASQ | 1214 | 4244 | 0.01 | 0.02 (−0.04, 0.08) | 0.38 | 0.12 | 146 | 487 | −0.11 | 0.01 (−0.18, 0.20) | 0.10 | 0.11 |
| Madagascar | MAHAY (29) | ASQI | 940 | 892 | 0.11 | −0.15 (−0.37, 0.07) | 0.03 | 0.08 | 336 | 389 | −0.15 | −0.02 (−0.26, 0.22) | 0.06 | 0.07 |
| Malawi | iLiNS−DYADM (30) | CDI | 68 | 118 | 0.10 | −0.25 (−0.55, 0.06) | 0.02 | 0.06 | 146 | 315 | −0.03 | 0.11 (−0.08, 0.31) | 0.10 | 0.10 |
| Malawi | iLiNS−DOSE (31) | CDI | 133 | 51 | 0.17 | −0.07 (−0.42, 0.27) | 0.01 | 0.05 | 403 | 133 | −0.07 | 0.07 (−0.12, 0.26) | 0.10 | 0.10 |
| Mali | PROMIS CS (32) | DMC | | | | | | | | | | | | |
| Zimbabwe | SHINE (HIV−) (33) | CDI | 612 | 576 | −0.04 | 0.14 (0.01, 0.26) | 0.09 | 0.11 | 136 | 148 | −0.14 | −0.07 (−0.34, 0.20) | 0.05 | 0.06 |
| Zimbabwe | SHINE (HIV+) (34) | CDI | 114 | 103 | −0.06 | 0.28 (0.01, 0.56) | 0.02 | 0.07 | 41 | 35 | −0.22 | −0.02 (−0.44, 0.41) | 0.02 | 0.03 |
| | | | **5978** | **9811** | | **I² = 0.66, Tau² = 0.02** | | | **2603** | **3020** | | **I² = 0.35, Tau² = 0.00** | | |
| **Fixed** | | | | | | **0.06 (0.03, 0.10)** | | | | | | **0.10 (0.04, 0.17)** | | |
| **Random** | | | | | | **0.05 (−0.05, 0.16)** | | | | | | **0.10 (0.02, 0.17)** | | |

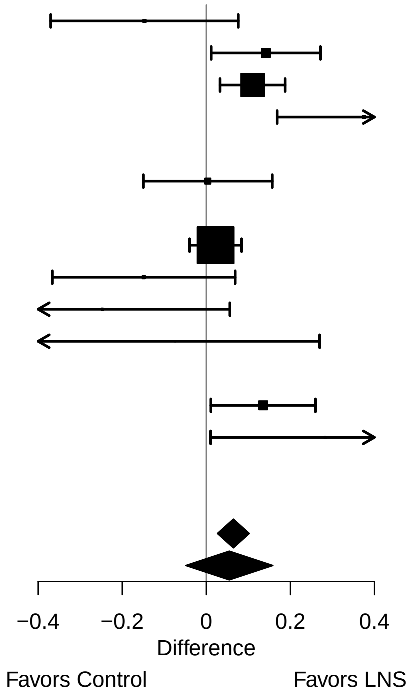

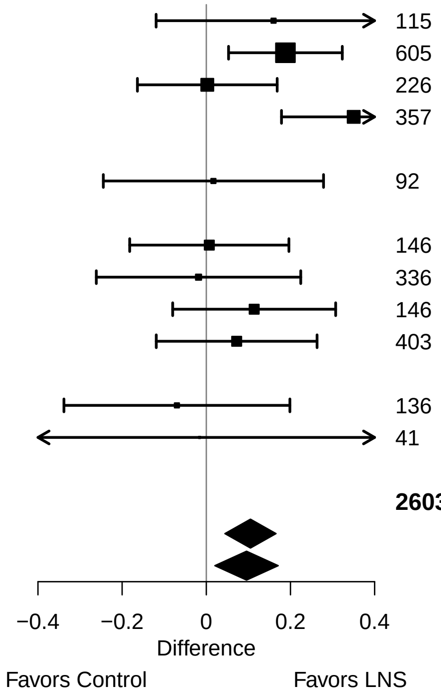

#### Supplemental figure 6A: Mean difference in language z-score

#### 6A3: Stratified by Household source water quality

**P−for−interaction = 0.988**
**Difference in MDs = 0.00 (−0.11, 0.11)**

| Country | Trial | Tool | LNS N | Control N | Control Mean | Improved MD (95% CI) | Fixed W | Random W | LNS N | Control N | Control Mean | Unimproved MD (95% CI) | Fixed W | Random W |
|---|---|---|---|---|---|---|---|---|---|---|---|---|---|---|
| Bangladesh | JiVitA−4 (21) | BSID−III | | | | | | | | | | | | |
| Bangladesh | RDNS (22) | CDI | | | | | | | | | | | | |
| Bangladesh | WASH−B (23) | CDI | 480 | 981 | −0.08 | 0.18 (0.07, 0.30) | 0.26 | 0.24 | 67 | 117 | −0.22 | 0.09 (−0.23, 0.41) | 0.06 | 0.10 |
| Burkina Faso | iLiNS−Zinc (24) | DMC | 220 | 82 | −0.06 | 0.30 (0.08, 0.52) | 0.07 | 0.08 | 526 | 293 | −0.30 | 0.37 (0.20, 0.54) | 0.23 | 0.18 |
| Ghana | GHANA (25) | | | | | | | | | | | | | |
| Ghana | iLiNS−DYADG (26) | CDI | | | | | | | | | | | | |
| Haiti | HAITI (27) | Total sounds | | | | | | | | | | | | |
| Kenya | WASH−B (28) | EASQ | 413 | 1419 | 0.01 | 0.01 (−0.12, 0.15) | 0.20 | 0.19 | 236 | 649 | −0.03 | 0.12 (−0.03, 0.27) | 0.28 | 0.19 |
| Madagascar | MAHAY (29) | ASQI | 405 | 430 | −0.05 | −0.07 (−0.42, 0.29) | 0.03 | 0.03 | 1134 | 1133 | 0.10 | −0.14 (−0.32, 0.05) | 0.18 | 0.16 |
| Malawi | iLiNS−DYADM (30) | CDI | 195 | 401 | 0.01 | 0.00 (−0.17, 0.17) | 0.12 | 0.12 | 20 | 36 | −0.11 | 0.14 (−0.38, 0.66) | 0.02 | 0.05 |
| Malawi | iLiNS−DOSE (31) | CDI | 514 | 177 | −0.04 | 0.07 (−0.10, 0.24) | 0.12 | 0.12 | 44 | 15 | 0.06 | −0.16 (−0.72, 0.40) | 0.02 | 0.04 |
| Mali | PROMIS CS (32) | DMC | 528 | 552 | −0.04 | 0.12 (−0.09, 0.33) | 0.08 | 0.09 | 357 | 378 | −0.15 | 0.17 (−0.05, 0.39) | 0.13 | 0.14 |
| Zimbabwe | SHINE (HIV−) (33) | CDI | 220 | 212 | 0.00 | 0.12 (−0.07, 0.31) | 0.10 | 0.10 | 125 | 122 | −0.10 | −0.13 (−0.42, 0.16) | 0.07 | 0.11 |
| Zimbabwe | SHINE (HIV+) (34) | CDI | 36 | 41 | −0.18 | 0.12 (−0.33, 0.57) | 0.02 | 0.02 | 25 | 25 | −0.16 | −0.20 (−0.86, 0.46) | 0.01 | 0.03 |
| | | | **3011** | **4295** | | **I² = 0.13, Tau² = 0.00** | | | **2534** | **2768** | | **I² = 0.62, Tau² = 0.02** | | |
| **Fixed** | | | | | | **0.10 (0.04, 0.16)** | | | | | | **0.11 (0.03, 0.18)** | | |
| **Random** | | | | | | **0.10 (0.04, 0.17)** | | | | | | **0.08 (−0.05, 0.20)** | | |

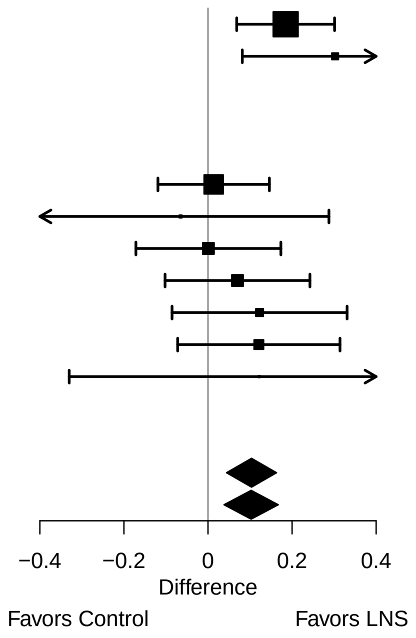

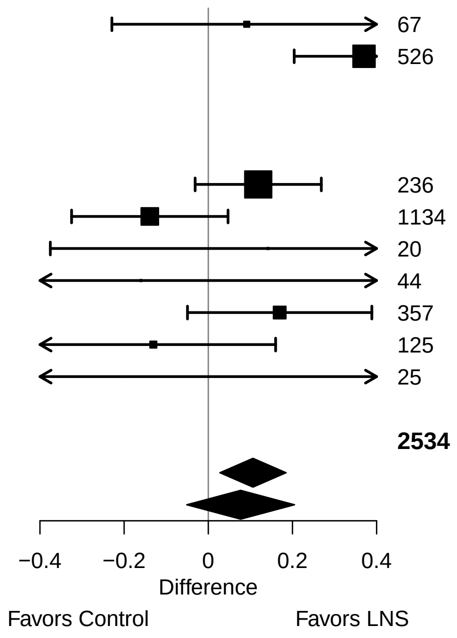

**Supplemental figure 6A: Mean difference in language z-score**

#### 6A4: Stratified by Household sanitation

**P−for−interaction = 0.218**
**Difference in MDs = −0.06 (−0.16, 0.04)**

| | | | Improved | | | | | | Unimproved | | | | | |
|---|---|---|---|---|---|---|---|---|---|---|---|---|---|---|
| **Country** | **Trial** | **Tool** | **LNS N** | **Control N** | **Control Mean** | **MD (95% CI)** | **Fixed W** | **Random W** | **LNS N** | **Control N** | **Control Mean** | **MD (95% CI)** | **Fixed W** | **Random W** |
| Bangladesh | JiVitA−4 (21) | BSID−III | 353 | 112 | 0.12 | −0.10 (−0.30, 0.10) | 0.08 | 0.11 | 91 | 31 | −0.16 | −0.03 (−0.45, 0.39) | 0.02 | 0.03 |
| Bangladesh | RDNS (22) | CDI | 1179 | 572 | −0.06 | 0.18 (0.07, 0.29) | 0.27 | 0.18 | 482 | 241 | −0.22 | 0.13 (0.00, 0.26) | 0.19 | 0.17 |
| Bangladesh | WASH−B (23) | CDI | 492 | 998 | −0.07 | 0.18 (0.06, 0.29) | 0.24 | 0.18 | 29 | 53 | −0.36 | 0.19 (−0.25, 0.64) | 0.02 | 0.03 |
| Burkina Faso | iLiNS−Zinc (24) | DMC | 16 | 11 | −0.45 | 0.77 (0.19, 1.34) | 0.01 | 0.02 | 730 | 364 | −0.24 | 0.36 (0.19, 0.52) | 0.12 | 0.13 |
| Ghana | GHANA (25) | | | | | | | | | | | | | |
| Ghana | iLiNS−DYADG (26) | CDI | 320 | 640 | −0.01 | 0.02 (−0.12, 0.15) | 0.18 | 0.16 | 10 | 16 | 0.30 | −0.40 (−1.24, 0.43) | 0.00 | 0.01 |
| Haiti | HAITI (27) | Total sounds | | | | | | | | | | | | |
| Kenya | WASH−B (28) | EASQ | 94 | 337 | 0.12 | 0.13 (−0.09, 0.36) | 0.06 | 0.10 | 555 | 1732 | −0.02 | 0.04 (−0.07, 0.15) | 0.26 | 0.19 |
| Madagascar | MAHAY (29) | ASQI | | | | | | | | | | | | |
| Malawi | iLiNS−DYADM (30) | CDI | 19 | 42 | 0.19 | −0.04 (−0.54, 0.46) | 0.01 | 0.03 | 196 | 395 | −0.02 | 0.02 (−0.15, 0.19) | 0.11 | 0.12 |
| Malawi | iLiNS−DOSE (31) | CDI | 12 | 9 | 0.32 | −0.23 (−1.09, 0.62) | 0.00 | 0.01 | 546 | 183 | −0.05 | 0.07 (−0.10, 0.24) | 0.12 | 0.13 |
| Mali | PROMIS CS (32) | DMC | 687 | 678 | −0.05 | 0.15 (−0.04, 0.34) | 0.09 | 0.11 | 211 | 242 | −0.10 | 0.11 (−0.12, 0.33) | 0.07 | 0.08 |
| Zimbabwe | SHINE (HIV−) (33) | CDI | 127 | 106 | −0.03 | 0.14 (−0.12, 0.40) | 0.05 | 0.08 | 217 | 229 | −0.04 | −0.02 (−0.22, 0.19) | 0.08 | 0.10 |
| Zimbabwe | SHINE (HIV+) (34) | CDI | 24 | 17 | −0.06 | −0.10 (−0.70, 0.50) | 0.01 | 0.02 | 37 | 49 | −0.21 | 0.01 (−0.44, 0.47) | 0.02 | 0.02 |
| | | | **3323** | **3522** | | **I² = 0.36, Tau² = 0.01** | | | **3104** | **3535** | | **I² = 0.34, Tau² = 0.00** | | |
| **Fixed** | | | | | | **0.12 (0.06, 0.17)** | | | | | | **0.10 (0.04, 0.15)** | | |
| **Random** | | | | | | **0.11 (0.02, 0.20)** | | | | | | **0.10 (0.02, 0.17)** | | |

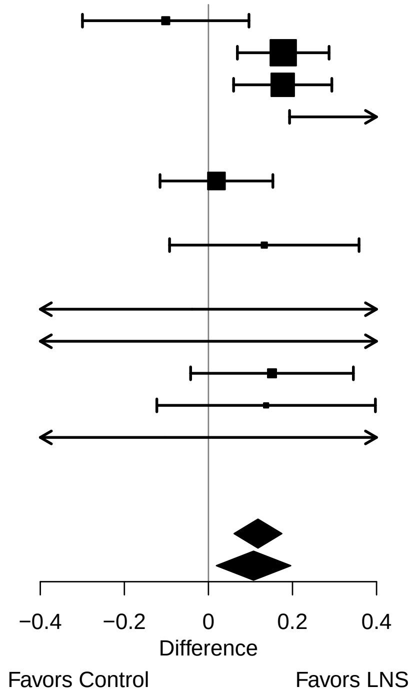

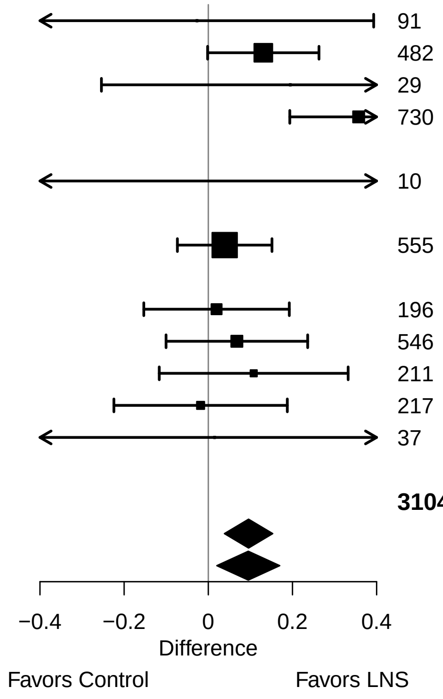

#### Supplemental figure 6A: Mean difference in language z-score

#### 6A5: Stratified by Home environment

**P−for−interaction = 0.251**
**Difference in MDs = 0.04 (−0.03, 0.11)**

| Country | Trial | Tool | At least median: LNS N | Control N | Control Mean | MD (95% CI) | Fixed W | Random W | Less than median: LNS N | Control N | Control Mean | MD (95% CI) | Fixed W | Random W |
|---|---|---|---|---|---|---|---|---|---|---|---|---|---|---|
| Bangladesh | JiVitA−4 (21) | BSID−III | | | | | | | | | | | | |
| Bangladesh | RDNS (22) | CDI | 1192 | 581 | 0.07 | 0.16 (0.05, 0.26) | 0.17 | 0.16 | 468 | 230 | −0.56 | 0.18 (−0.04, 0.40) | 0.06 | 0.11 |
| Bangladesh | WASH−B (23) | CDI | 630 | 1652 | 0.23 | 0.11 (0.03, 0.20) | 0.24 | 0.18 | 479 | 1700 | −0.27 | −0.02 (−0.13, 0.09) | 0.25 | 0.16 |
| Burkina Faso | iLiNS−Zinc (24) | DMC | 473 | 199 | 0.11 | 0.17 (0.02, 0.33) | 0.07 | 0.11 | 272 | 175 | −0.64 | 0.49 (0.29, 0.69) | 0.08 | 0.12 |
| Ghana | GHANA (25) | | | | | | | | | | | | | |
| Ghana | iLiNS−DYADG (26) | CDI | 213 | 410 | 0.18 | 0.01 (−0.15, 0.17) | 0.07 | 0.11 | 118 | 248 | −0.31 | −0.03 (−0.24, 0.19) | 0.07 | 0.12 |
| Haiti | HAITI (27) | Total sounds | | | | | | | | | | | | |
| Kenya | WASH−B (28) | EASQ | 809 | 2682 | 0.20 | −0.02 (−0.10, 0.06) | 0.30 | 0.18 | 552 | 2060 | −0.27 | 0.05 (−0.04, 0.14) | 0.37 | 0.17 |
| Madagascar | MAHAY (29) | ASQI | 843 | 839 | 0.21 | −0.16 (−0.36, 0.03) | 0.04 | 0.08 | 770 | 764 | −0.12 | −0.06 (−0.26, 0.15) | 0.07 | 0.12 |
| Malawi | iLiNS−DYADM (30) | CDI | 137 | 290 | 0.20 | −0.01 (−0.21, 0.19) | 0.05 | 0.09 | 78 | 149 | −0.40 | 0.10 (−0.16, 0.36) | 0.04 | 0.10 |
| Malawi | iLiNS−DOSE (31) | CDI | 375 | 135 | 0.15 | 0.01 (−0.17, 0.19) | 0.05 | 0.09 | 269 | 86 | −0.32 | 0.13 (−0.13, 0.38) | 0.05 | 0.10 |
| Mali | PROMIS CS (32) | DMC | | | | | | | | | | | | |
| Zimbabwe | SHINE (HIV−) (33) | CDI | | | | | | | | | | | | |
| Zimbabwe | SHINE (HIV+) (34) | CDI | | | | | | | | | | | | |
| | | | **4672** | **6788** | | **$I^2$ = 0.59, $Tau^2$ = 0.01** | | | **3006** | **5412** | | **$I^2$ = 0.70, $Tau^2$ = 0.02** | | |
| **Fixed** | | | | | | **0.05 (0.01, 0.10)** | | | | | | **0.07 (0.01, 0.12)** | | |
| **Random** | | | | | | **0.05 (−0.03, 0.12)** | | | | | | **0.10 (−0.02, 0.22)** | | |

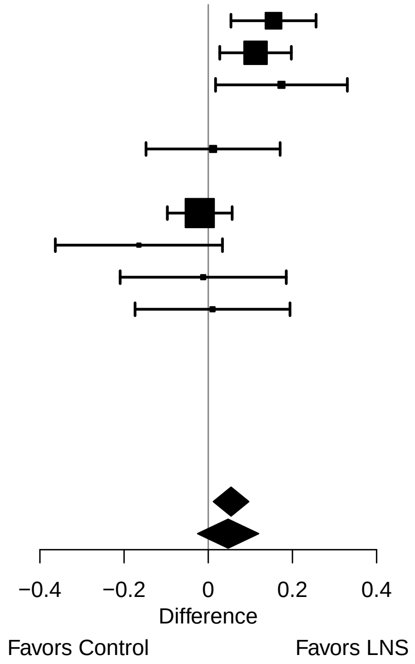

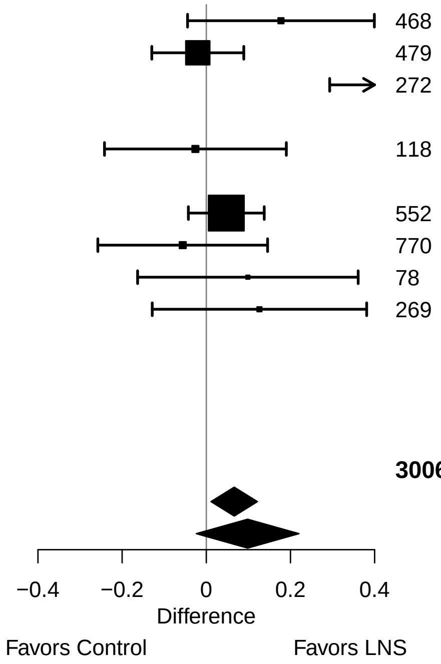

#### Supplemental figure 6A: Mean difference in language z-score

#### 6A6: Stratified by Season at the time of assessment

**P−for−interaction = 0.979**
**Difference in MDs = 0.00 (−0.07, 0.07)**

| Country | Trial | Tool | LNS N | Control N | Control Mean | Dry MD (95% CI) | Fixed W | Random W | LNS N | Control N | Control Mean | Rainy MD (95% CI) | Fixed W | Random W |
|---|---|---|---|---|---|---|---|---|---|---|---|---|---|---|
| Bangladesh | JiVitA−4 (21) | BSID−III | 251 | 70 | 0.11 | −0.08 (−0.34, 0.19) | 0.03 | 0.06 | 194 | 73 | 0.02 | −0.10 (−0.36, 0.15) | 0.03 | 0.06 |
| Bangladesh | RDNS (22) | CDI | 739 | 359 | −0.09 | 0.17 (0.03, 0.31) | 0.12 | 0.12 | 924 | 455 | −0.13 | 0.16 (0.03, 0.29) | 0.12 | 0.14 |
| Bangladesh | WASH−B (23) | CDI | 629 | 1944 | 0.04 | 0.06 (−0.03, 0.15) | 0.29 | 0.16 | 480 | 1409 | −0.11 | 0.14 (0.02, 0.25) | 0.18 | 0.17 |
| Burkina Faso | iLiNS−Zinc (24) | DMC | 514 | 274 | −0.25 | 0.42 (0.18, 0.66) | 0.04 | 0.07 | 232 | 101 | −0.24 | 0.25 (0.03, 0.47) | 0.05 | 0.08 |
| Ghana | GHANA (25) | | | | | | | | | | | | | |
| Ghana | iLiNS−DYADG (26) | CDI | 194 | 388 | −0.03 | 0.10 (−0.07, 0.28) | 0.08 | 0.10 | 137 | 270 | 0.04 | −0.13 (−0.33, 0.08) | 0.05 | 0.09 |
| Haiti | HAITI (27) | Total sounds | | | | | | | | | | | | |
| Kenya | WASH−B (28) | EASQ | 315 | 1059 | −0.18 | 0.07 (−0.05, 0.20) | 0.15 | 0.13 | 1046 | 3682 | 0.05 | 0.01 (−0.06, 0.08) | 0.44 | 0.22 |
| Madagascar | MAHAY (29) | ASQI | | | | | | | | | | | | |
| Malawi | iLiNS−DYADM (30) | CDI | 153 | 311 | 0.05 | −0.01 (−0.21, 0.18) | 0.06 | 0.09 | 62 | 127 | −0.14 | 0.09 (−0.20, 0.39) | 0.02 | 0.05 |
| Malawi | iLiNS−DOSE (31) | CDI | 394 | 118 | 0.02 | 0.00 (−0.21, 0.21) | 0.05 | 0.08 | 244 | 100 | −0.08 | 0.09 (−0.14, 0.32) | 0.04 | 0.07 |
| Mali | PROMIS CS (32) | DMC | | | | | | | | | | | | |
| Zimbabwe | SHINE (HIV−) (33) | CDI | 582 | 555 | −0.09 | 0.04 (−0.09, 0.17) | 0.13 | 0.13 | 235 | 226 | 0.07 | 0.20 (−0.02, 0.41) | 0.05 | 0.08 |
| Zimbabwe | SHINE (HIV+) (34) | CDI | 117 | 106 | −0.18 | 0.25 (−0.02, 0.53) | 0.03 | 0.06 | 48 | 41 | 0.00 | 0.23 (−0.17, 0.62) | 0.01 | 0.03 |
| | | | **3888** | **5184** | | **I² = 0.38, Tau² = 0.01** | | | **3602** | **6484** | | **I² = 0.43, Tau² = 0.00** | | |
| **Fixed** | | | | | | **0.09 (0.04, 0.13)** | | | | | | **0.07 (0.02, 0.12)** | | |
| **Random** | | | | | | **0.09 (0.02, 0.17)** | | | | | | **0.09 (0.01, 0.16)** | | |

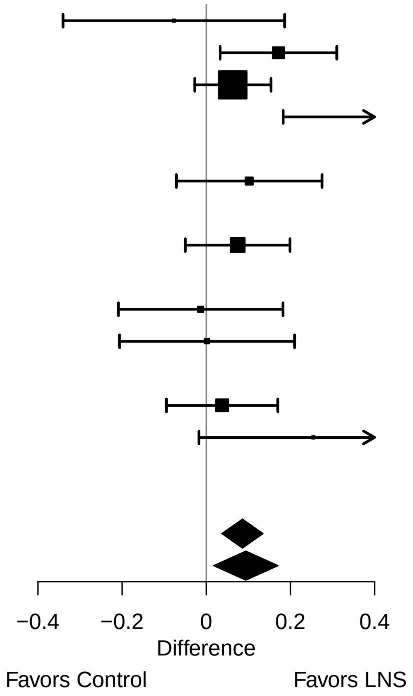

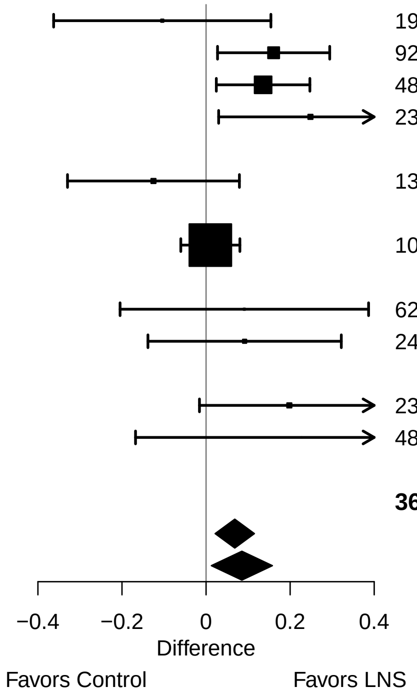

### Supplemental figure 6B: Language lowest decile prevalence ratio

#### 6B1: Stratified by Household socio-economic status

**P−for−interaction = 0.227**
**Ratio of PRs = 0.90 (0.75, 1.07)**

| | | | At least median | | | | | | Less than median | | | | | |
|---|---|---|---|---|---|---|---|---|---|---|---|---|---|---|
| **Country** | **Trial** | **Tool** | **LNS N** | **Control N** | **Control Prevalence** | **PR (95% CI)** | **Fixed W** | **Random W** | **LNS N** | **Control N** | **Control Prevalence** | **PR (95% CI)** | **Fixed W** | **Random W** |
| Bangladesh | JiVitA−4 (21) | BSID−III | 236 | 77 | 7.8 | 1.03 (0.43, 2.46) | 0.02 | 0.03 | 208 | 66 | 15.2 | 0.76 (0.40, 1.44) | 0.03 | 0.03 |
| Bangladesh | RDNS (22) | CDI | 834 | 407 | 9.3 | 0.65 (0.43, 1.00) | 0.10 | 0.11 | 829 | 407 | 15.5 | 0.76 (0.57, 1.00) | 0.17 | 0.17 |
| Bangladesh | WASH−B (23) | CDI | 568 | 1703 | 7.8 | 0.91 (0.66, 1.26) | 0.18 | 0.16 | 541 | 1650 | 13.4 | 0.75 (0.55, 1.01) | 0.15 | 0.15 |
| Burkina Faso | iLiNS−Zinc (24) | DMC | 459 | 163 | 15.3 | 0.50 (0.32, 0.78) | 0.09 | 0.10 | 287 | 212 | 16.0 | 0.41 (0.24, 0.72) | 0.04 | 0.04 |
| Ghana | GHANA (25) | | | | | | | | | | | | | |
| Ghana | iLiNS−DYADG (26) | CDI | 152 | 341 | 7.9 | 1.00 (0.52, 1.91) | 0.04 | 0.06 | 179 | 315 | 11.7 | 1.09 (0.67, 1.78) | 0.06 | 0.06 |
| Haiti | HAITI (27) | Total sounds | | | | | | | | | | | | |
| Kenya | WASH−B (28) | EASQ | 778 | 2706 | 8.5 | 1.09 (0.84, 1.41) | 0.28 | 0.20 | 582 | 2036 | 12.2 | 0.83 (0.64, 1.07) | 0.21 | 0.21 |
| Madagascar | MAHAY (29) | ASQI | 741 | 845 | 5.0 | 1.36 (0.72, 2.56) | 0.05 | 0.06 | 823 | 724 | 14.2 | 1.01 (0.64, 1.59) | 0.07 | 0.07 |
| Malawi | iLiNS−DYADM (30) | CDI | 118 | 230 | 9.1 | 0.74 (0.34, 1.63) | 0.03 | 0.04 | 97 | 208 | 12.5 | 0.99 (0.52, 1.88) | 0.03 | 0.03 |
| Malawi | iLiNS−DOSE (31) | CDI | 278 | 94 | 9.6 | 1.09 (0.54, 2.22) | 0.04 | 0.05 | 271 | 96 | 11.5 | 0.87 (0.45, 1.68) | 0.03 | 0.03 |
| Mali | PROMIS CS (32) | DMC | 494 | 450 | 9.6 | 0.85 (0.52, 1.39) | 0.08 | 0.09 | 433 | 494 | 13.0 | 0.73 (0.48, 1.12) | 0.08 | 0.08 |
| Zimbabwe | SHINE (HIV−) (33) | CDI | 396 | 351 | 10.3 | 0.81 (0.49, 1.34) | 0.07 | 0.09 | 375 | 383 | 12.0 | 0.91 (0.64, 1.29) | 0.12 | 0.12 |
| Zimbabwe | SHINE (HIV+) (34) | CDI | 86 | 77 | 15.6 | 0.45 (0.16, 1.24) | 0.02 | 0.02 | 74 | 64 | 9.4 | 1.15 (0.41, 3.22) | 0.01 | 0.01 |
| | | | **5140** | **7444** | | **I² = 0.27, Tau² = 0.02** | | | **4699** | **6655** | | **I² = 0.00, Tau² = 0.00** | | |
| **Fixed** | | | | | | **0.88 (0.76, 1.00)** | | | | | | **0.81 (0.72, 0.91)** | | |
| **Random** | | | | | | **0.86 (0.73, 1.01)** | | | | | | **0.81 (0.72, 0.91)** | | |

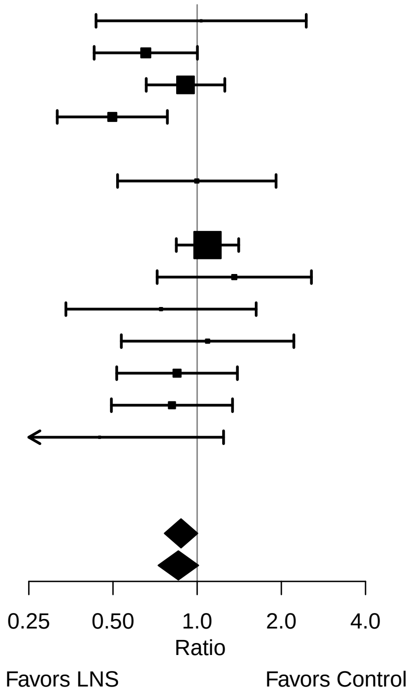

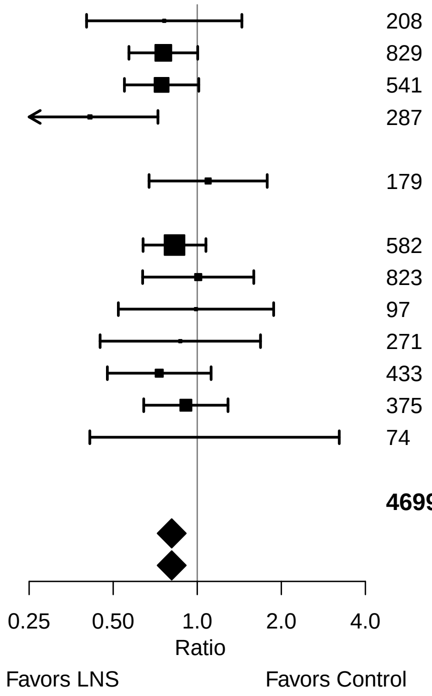

**Supplemental figure 6B: Language lowest decile prevalence ratio**

#### 6B2: Stratified by Household food insecurity

**P−for−interaction = 0.772**
**Ratio of PRs = 1.03 (0.85, 1.25)**

| Country | Trial | Tool | LNS N | Control N | Control Prevalence | Mild to secure PR (95% CI) | Fixed W | Random W | LNS N | Control N | Control Prevalence | Moderate to severe PR (95% CI) | Fixed W | Random W |
|---|---|---|---|---|---|---|---|---|---|---|---|---|---|---|
| Bangladesh | JiVitA−4 (21) | BSID−III | | | | | | | | | | | | |
| Bangladesh | RDNS (22) | CDI | 1058 | 493 | 9.9 | 0.74 (0.54, 1.03) | 0.12 | 0.13 | 605 | 321 | 16.2 | 0.71 (0.53, 0.97) | 0.27 | 0.19 |
| Bangladesh | WASH−B (23) | CDI | 883 | 2608 | 9.6 | 0.80 (0.63, 1.02) | 0.22 | 0.15 | 226 | 745 | 13.7 | 0.84 (0.54, 1.30) | 0.13 | 0.14 |
| Burkina Faso | iLiNS−Zinc (24) | DMC | 389 | 162 | 15.4 | 0.42 (0.28, 0.63) | 0.07 | 0.12 | 357 | 213 | 16.0 | 0.51 (0.35, 0.74) | 0.18 | 0.16 |
| Ghana | GHANA (25) | | | | | | | | | | | | | |
| Ghana | iLiNS−DYADG (26) | CDI | 238 | 453 | 8.4 | 1.45 (0.92, 2.29) | 0.06 | 0.11 | 92 | 202 | 12.9 | 0.51 (0.22, 1.19) | 0.03 | 0.05 |
| Haiti | HAITI (27) | Total sounds | | | | | | | | | | | | |
| Kenya | WASH−B (28) | EASQ | 1214 | 4244 | 10.0 | 0.90 (0.74, 1.09) | 0.34 | 0.16 | 146 | 487 | 11.3 | 1.33 (0.84, 2.12) | 0.12 | 0.13 |
| Madagascar | MAHAY (29) | ASQI | 940 | 892 | 8.0 | 1.40 (0.82, 2.41) | 0.04 | 0.10 | 336 | 389 | 12.6 | 0.92 (0.49, 1.74) | 0.06 | 0.08 |
| Malawi | iLiNS−DYADM (30) | CDI | 68 | 118 | 9.3 | 1.42 (0.62, 3.25) | 0.02 | 0.06 | 146 | 315 | 10.5 | 0.72 (0.37, 1.38) | 0.06 | 0.08 |
| Malawi | iLiNS−DOSE (31) | CDI | 133 | 51 | 9.8 | 0.77 (0.28, 2.13) | 0.01 | 0.05 | 403 | 133 | 10.5 | 1.01 (0.57, 1.79) | 0.08 | 0.10 |
| Mali | PROMIS CS (32) | DMC | | | | | | | | | | | | |
| Zimbabwe | SHINE (HIV−) (33) | CDI | 612 | 576 | 10.6 | 0.82 (0.59, 1.14) | 0.12 | 0.13 | 136 | 148 | 14.2 | 1.04 (0.54, 1.98) | 0.06 | 0.08 |
| Zimbabwe | SHINE (HIV+) (34) | CDI | | | | | | | | | | | | |
| | | | **5535** | **9597** | | **I² = 0.65, Tau² = 0.10** | | | **2447** | **2953** | | **I² = 0.40, Tau² = 0.04** | | |
| **Fixed** | | | | | | **0.85 (0.76, 0.95)** | | | | | | **0.78 (0.67, 0.91)** | | |
| **Random** | | | | | | **0.87 (0.68, 1.13)** | | | | | | **0.80 (0.65, 0.99)** | | |

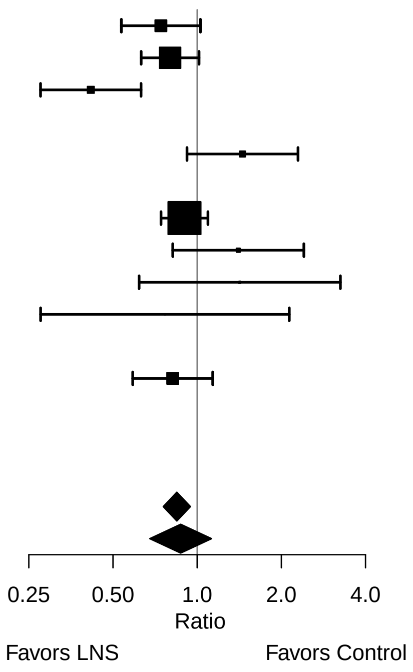

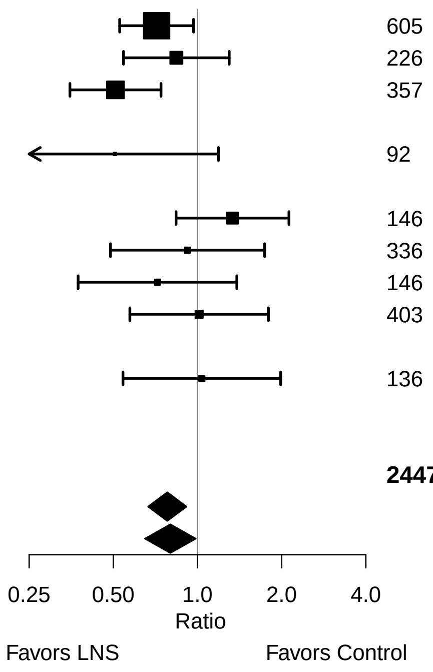

#### Supplemental figure 6B: Language lowest decile prevalence ratio

##### 6B3: Stratified by Household source water quality

**P−for−interaction = 0.707**
**Ratio of PRs = 0.95 (0.73, 1.24)**

| Country | Trial | Tool | LNS N | Control N | Control Prevalence | Improved PR (95% CI) | Fixed W | Random W | LNS N | Control N | Control Prevalence | Unimproved PR (95% CI) | Fixed W | Random W |
|---|---|---|---|---|---|---|---|---|---|---|---|---|---|---|
| Bangladesh | JiVitA−4 (21) | BSID−III | | | | | | | | | | | | |
| Bangladesh | RDNS (22) | CDI | | | | | | | | | | | | |
| Bangladesh | WASH−B (23) | CDI | 480 | 981 | 10.9 | 0.65 (0.45, 0.93) | 0.26 | 0.22 | 67 | 117 | 16.2 | 0.55 (0.21, 1.46) | 0.04 | 0.07 |
| Burkina Faso | iLiNS−Zinc (24) | DMC | 220 | 82 | 12.2 | 0.45 (0.26, 0.77) | 0.11 | 0.14 | 526 | 293 | 16.7 | 0.48 (0.33, 0.69) | 0.27 | 0.22 |
| Ghana | GHANA (25) | | | | | | | | | | | | | |
| Ghana | iLiNS−DYADG (26) | CDI | | | | | | | | | | | | |
| Haiti | HAITI (27) | Total sounds | | | | | | | | | | | | |
| Kenya | WASH−B (28) | EASQ | 413 | 1419 | 10.4 | 1.01 (0.70, 1.44) | 0.26 | 0.22 | 236 | 649 | 10.5 | 0.89 (0.61, 1.29) | 0.26 | 0.22 |
| Madagascar | MAHAY (29) | ASQI | 405 | 430 | 12.3 | 1.18 (0.60, 2.31) | 0.07 | 0.11 | 1134 | 1133 | 8.1 | 1.17 (0.73, 1.90) | 0.16 | 0.18 |
| Malawi | iLiNS−DYADM (30) | CDI | | | | | | | | | | | | |
| Malawi | iLiNS−DOSE (31) | CDI | | | | | | | | | | | | |
| Mali | PROMIS CS (32) | DMC | 528 | 552 | 9.8 | 0.85 (0.55, 1.31) | 0.18 | 0.18 | 357 | 378 | 14.0 | 0.70 (0.44, 1.11) | 0.17 | 0.18 |
| Zimbabwe | SHINE (HIV−) (33) | CDI | 220 | 212 | 10.4 | 0.92 (0.53, 1.59) | 0.11 | 0.14 | 125 | 122 | 12.3 | 1.17 (0.62, 2.22) | 0.09 | 0.13 |
| Zimbabwe | SHINE (HIV+) (34) | CDI | | | | | | | | | | | | |
| | | | **2266** | **3676** | | **I² = 0.43, Tau² = 0.05** | | | **2445** | **2692** | | **I² = 0.59, Tau² = 0.07** | | |
| **Fixed** | | | | | | **0.80 (0.66, 0.96)** | | | | | | **0.76 (0.62, 0.92)** | | |
| **Random** | | | | | | **0.80 (0.61, 1.03)** | | | | | | **0.78 (0.58, 1.06)** | | |

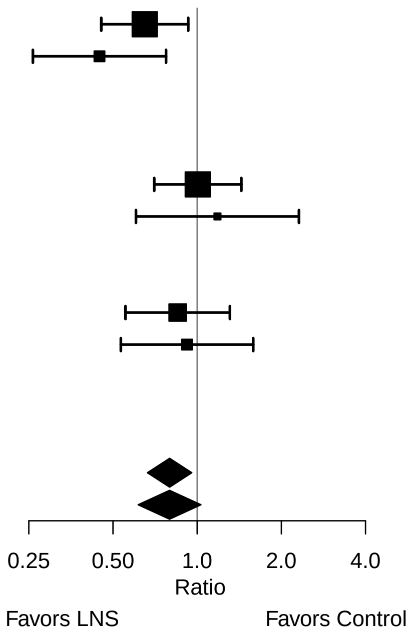

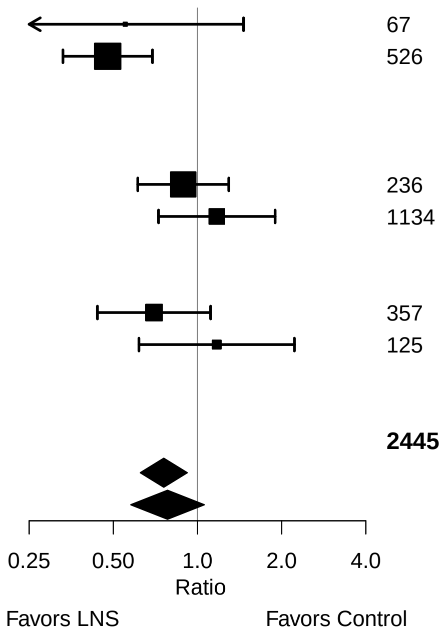

#### Supplemental figure 6B: Language lowest decile prevalence ratio

#### 6B4: Stratified by Household sanitation

**P−for−interaction = 0.821**
**Ratio of PRs = 1.04 (0.72, 1.50)**

| Country | Trial | Tool | LNS N | Control N | Control Prevalence | Improved PR (95% CI) | Fixed W | Random W | LNS N | Control N | Control Prevalence | Unimproved PR (95% CI) | Fixed W | Random W |
|---|---|---|---|---|---|---|---|---|---|---|---|---|---|---|
| Bangladesh | JiVitA−4 (21) | BSID−III | | | | | | | | | | | | |
| Bangladesh | RDNS (22) | CDI | 1179 | 572 | 10.8 | 0.73 (0.54, 0.98) | 0.57 | 0.57 | 482 | 241 | 16.2 | 0.71 (0.48, 1.03) | 0.47 | 0.41 |
| Bangladesh | WASH−B (23) | CDI | | | | | | | | | | | | |
| Burkina Faso | iLiNS−Zinc (24) | DMC | | | | | | | | | | | | |
| Ghana | GHANA (25) | | | | | | | | | | | | | |
| Ghana | iLiNS−DYADG (26) | CDI | | | | | | | | | | | | |
| Haiti | HAITI (27) | Total sounds | | | | | | | | | | | | |
| Kenya | WASH−B (28) | EASQ | | | | | | | | | | | | |
| Madagascar | MAHAY (29) | ASQI | | | | | | | | | | | | |
| Malawi | iLiNS−DYADM (30) | CDI | | | | | | | | | | | | |
| Malawi | iLiNS−DOSE (31) | CDI | | | | | | | | | | | | |
| Mali | PROMIS CS (32) | DMC | 687 | 678 | 10.5 | 0.83 (0.57, 1.21) | 0.36 | 0.36 | 211 | 242 | 12.4 | 0.69 (0.37, 1.27) | 0.18 | 0.24 |
| Zimbabwe | SHINE (HIV−) (33) | CDI | 127 | 106 | 9.4 | 0.58 (0.26, 1.33) | 0.07 | 0.07 | 217 | 229 | 11.8 | 1.21 (0.78, 1.89) | 0.35 | 0.35 |
| Zimbabwe | SHINE (HIV+) (34) | CDI | | | | | | | | | | | | |
| | | | **1993** | **1356** | | **I² = 0.00, Tau² = 0.00** | | | **910** | **712** | | **I² = 0.48, Tau² = 0.05** | | |
| **Fixed** | | | | | | **0.75 (0.60, 0.94)** | | | | | | **0.85 (0.65, 1.10)** | | |
| **Random** | | | | | | **0.75 (0.60, 0.94)** | | | | | | **0.85 (0.59, 1.22)** | | |

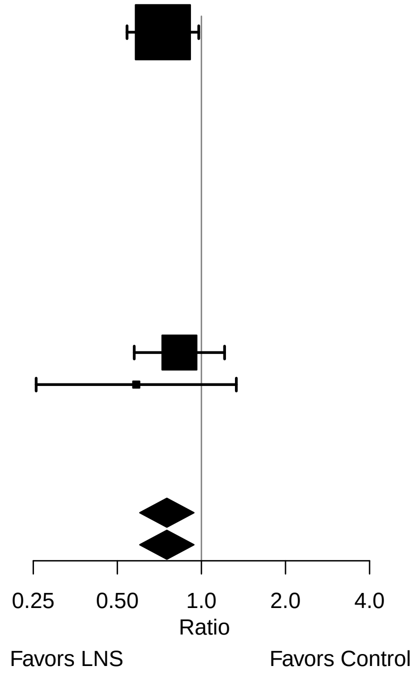

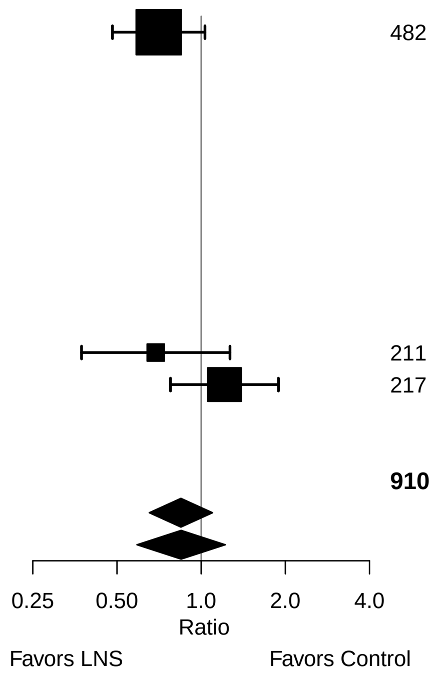

#### Supplemental figure 6B: Language lowest decile prevalence ratio

##### 6B5: Stratified by Home environment

**P−for−interaction = 0.592**
**Ratio of PRs = 0.95 (0.78, 1.16)**

| Country | Trial | Tool | At least median: LNS N | Control N | Control Prevalence | PR (95% CI) | Fixed W | Random W | Less than median: LNS N | Control N | Control Prevalence | PR (95% CI) | Fixed W | Random W |
|---|---|---|---|---|---|---|---|---|---|---|---|---|---|---|
| Bangladesh | JiVitA−4 (21) | BSID−III | | | | | | | | | | | | |
| Bangladesh | RDNS (22) | CDI | 1192 | 581 | 9.0 | 0.60 (0.42, 0.85) | 0.18 | 0.18 | 468 | 230 | 20.4 | 0.88 (0.58, 1.32) | 0.12 | 0.13 |
| Bangladesh | WASH−B (23) | CDI | 630 | 1652 | 5.9 | 0.73 (0.51, 1.05) | 0.16 | 0.16 | 479 | 1700 | 15.1 | 0.93 (0.71, 1.21) | 0.27 | 0.17 |
| Burkina Faso | iLiNS−Zinc (24) | DMC | 473 | 199 | 5.5 | 0.92 (0.60, 1.41) | 0.12 | 0.13 | 272 | 175 | 27.4 | 0.40 (0.26, 0.63) | 0.10 | 0.12 |
| Ghana | GHANA (25) | | | | | | | | | | | | | |
| Ghana | iLiNS−DYADG (26) | CDI | 213 | 410 | 6.3 | 1.04 (0.55, 1.94) | 0.05 | 0.07 | 118 | 248 | 15.3 | 1.16 (0.71, 1.89) | 0.08 | 0.11 |
| Haiti | HAITI (27) | Total sounds | | | | | | | | | | | | |
| Kenya | WASH−B (28) | EASQ | 809 | 2682 | 6.9 | 1.11 (0.86, 1.44) | 0.32 | 0.26 | 552 | 2060 | 14.3 | 0.87 (0.66, 1.15) | 0.25 | 0.17 |
| Madagascar | MAHAY (29) | ASQI | 843 | 839 | 7.7 | 1.07 (0.67, 1.71) | 0.10 | 0.11 | 770 | 764 | 10.9 | 1.24 (0.72, 2.16) | 0.06 | 0.10 |
| Malawi | iLiNS−DYADM (30) | CDI | 137 | 290 | 6.6 | 1.00 (0.47, 2.16) | 0.04 | 0.05 | 78 | 149 | 18.8 | 0.75 (0.40, 1.43) | 0.05 | 0.08 |
| Malawi | iLiNS−DOSE (31) | CDI | 375 | 135 | 5.9 | 0.81 (0.36, 1.82) | 0.03 | 0.04 | 269 | 86 | 17.4 | 0.98 (0.58, 1.67) | 0.07 | 0.10 |
| Mali | PROMIS CS (32) | DMC | | | | | | | | | | | | |
| Zimbabwe | SHINE (HIV−) (33) | CDI | | | | | | | | | | | | |
| Zimbabwe | SHINE (HIV+) (34) | CDI | | | | | | | | | | | | |
| | | | **4672** | **6788** | | **I² = 0.30, Tau² = 0.01** | | | **3006** | **5412** | | **I² = 0.53, Tau² = 0.06** | | |
| **Fixed** | | | | | | **0.89 (0.77, 1.03)** | | | | | | **0.86 (0.75, 0.99)** | | |
| **Random** | | | | | | **0.88 (0.74, 1.05)** | | | | | | **0.86 (0.68, 1.08)** | | |

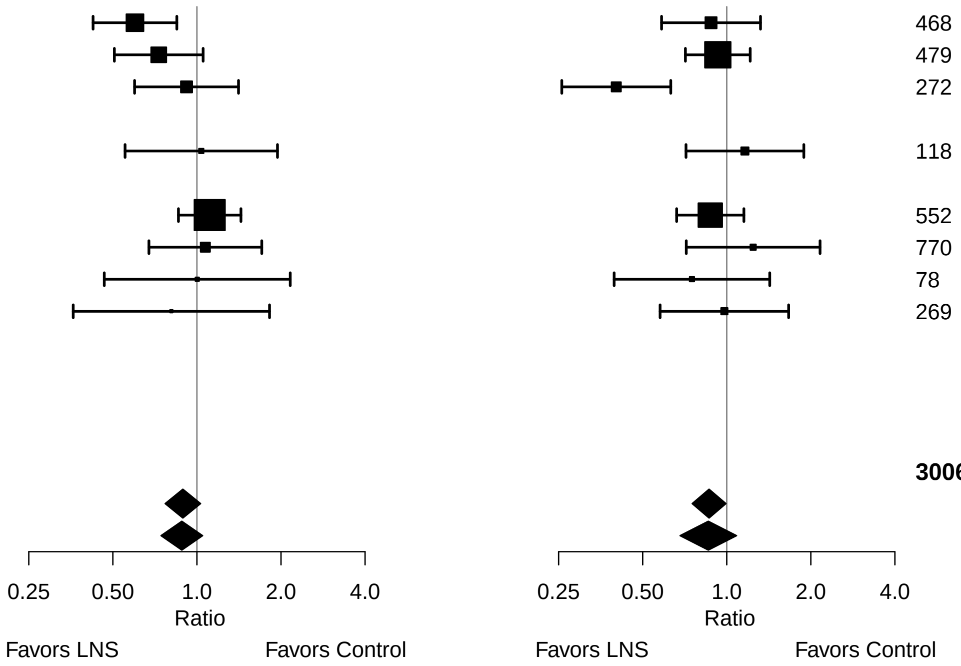

**Supplemental figure 6B: Language lowest decile prevalence ratio**

#### 6B6: Stratified by Season at the time of assessment

**P−for−interaction = 0.524**
**Ratio of PRs = 0.93 (0.76, 1.15)**

| Country | Trial | Tool | Dry: LNS N | Dry: Control N | Dry: Control Prevalence | Dry: PR (95% CI) | Dry: Fixed W | Dry: Random W | Rainy: LNS N | Rainy: Control N | Rainy: Control Prevalence | Rainy: PR (95% CI) | Rainy: Fixed W | Rainy: Random W |
|---|---|---|---|---|---|---|---|---|---|---|---|---|---|---|
| Bangladesh | JiVitA−4 (21) | BSID−III | 251 | 70 | 11.4 | 0.59 (0.26, 1.38) | 0.03 | 0.04 | 194 | 73 | 11.0 | 1.22 (0.62, 2.41) | 0.05 | 0.08 |
| Bangladesh | RDNS (22) | CDI | 739 | 359 | 12.0 | 0.84 (0.61, 1.15) | 0.18 | 0.16 | 924 | 455 | 12.7 | 0.63 (0.45, 0.87) | 0.20 | 0.18 |
| Bangladesh | WASH−B (23) | CDI | 629 | 1944 | 9.4 | 0.92 (0.70, 1.21) | 0.24 | 0.19 | 480 | 1409 | 12.1 | 0.69 (0.49, 0.96) | 0.19 | 0.17 |
| Burkina Faso | iLiNS−Zinc (24) | DMC | 514 | 274 | 16.1 | 0.46 (0.31, 0.69) | 0.11 | 0.12 | 232 | 101 | 14.9 | 0.46 (0.19, 1.16) | 0.02 | 0.05 |
| Ghana | GHANA (25) | | | | | | | | | | | | | |
| Ghana | iLiNS−DYADG (26) | CDI | 194 | 388 | 10.1 | 0.77 (0.44, 1.36) | 0.06 | 0.07 | 137 | 270 | 9.3 | 1.58 (0.91, 2.74) | 0.07 | 0.11 |
| Haiti | HAITI (27) | Total sounds | | | | | | | | | | | | |
| Kenya | WASH−B (28) | EASQ | 315 | 1059 | 13.5 | 1.01 (0.69, 1.48) | 0.13 | 0.13 | 1046 | 3682 | 9.2 | 0.91 (0.72, 1.15) | 0.38 | 0.21 |
| Madagascar | MAHAY (29) | ASQI | | | | | | | | | | | | |
| Malawi | iLiNS−DYADM (30) | CDI | 153 | 311 | 10.3 | 0.95 (0.53, 1.71) | 0.05 | 0.07 | 62 | 127 | 11.8 | 0.68 (0.26, 1.79) | 0.02 | 0.05 |
| Malawi | iLiNS−DOSE (31) | CDI | 394 | 118 | 12.7 | 0.72 (0.41, 1.27) | 0.06 | 0.07 | 244 | 100 | 8.0 | 1.38 (0.65, 2.94) | 0.04 | 0.07 |
| Mali | PROMIS CS (32) | DMC | | | | | | | | | | | | |
| Zimbabwe | SHINE (HIV−) (33) | CDI | 582 | 555 | 11.4 | 0.92 (0.65, 1.30) | 0.15 | 0.15 | 235 | 226 | 9.3 | 0.82 (0.40, 1.69) | 0.04 | 0.08 |
| Zimbabwe | SHINE (HIV+) (34) | CDI | | | | | | | | | | | | |
| | | | **3771** | **5078** | | **I² = 0.28, Tau² = 0.02** | | | **3554** | **6443** | | **I² = 0.45, Tau² = 0.05** | | |
| **Fixed** | | | | | | **0.82 (0.72, 0.94)** | | | | | | **0.83 (0.72, 0.96)** | | |
| **Random** | | | | | | **0.81 (0.69, 0.95)** | | | | | | **0.86 (0.68, 1.09)** | | |

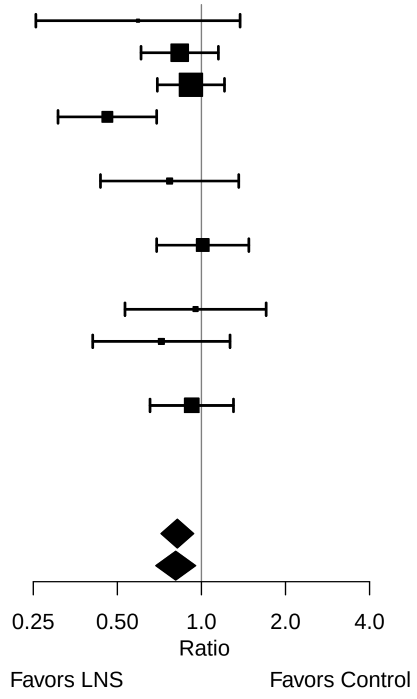

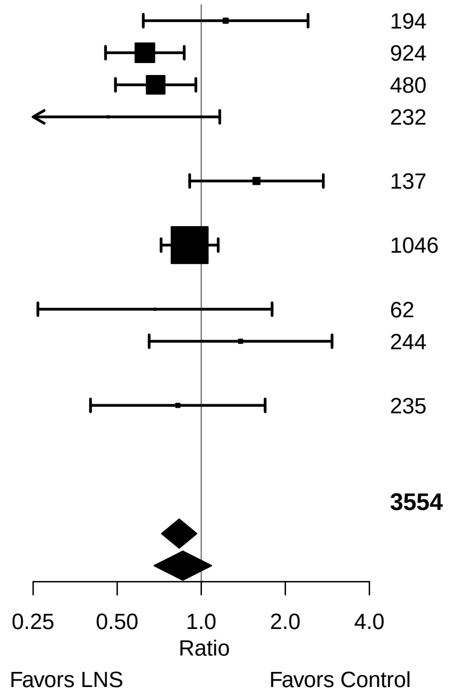

### Supplemental figure 6C: Language lowest decile prevalence difference

#### 6C1: Stratified by Household socio-economic status

**P−for−interaction = 0.078**
**Difference in PDs = −0.02 (−0.03, 0.00)**

| Country | Trial | Tool | At least median: LNS N | At least median: Control N | At least median: Control Prevalence | At least median: PD (95% CI) | At least median: Fixed W | At least median: Random W | Less than median: LNS N | Less than median: Control N | Less than median: Control Prevalence | Less than median: PD (95% CI) | Less than median: Fixed W | Less than median: Random W |
|---|---|---|---|---|---|---|---|---|---|---|---|---|---|---|
| Bangladesh | JiVitA−4 (21) | BSID−III | 236 | 77 | 7.8 | 0.00 (−0.07, 0.07) | 0.03 | 0.04 | 208 | 66 | 15.2 | −0.04 (−0.13, 0.05) | 0.02 | 0.03 |
| Bangladesh | RDNS (22) | CDI | 834 | 407 | 9.3 | −0.03 (−0.07, 0.00) | 0.11 | 0.12 | 829 | 407 | 15.5 | −0.04 (−0.08, 0.00) | 0.12 | 0.12 |
| Bangladesh | WASH−B (23) | CDI | 568 | 1703 | 7.8 | −0.01 (−0.03, 0.02) | 0.24 | 0.17 | 541 | 1650 | 13.4 | −0.03 (−0.07, 0.00) | 0.17 | 0.16 |
| Burkina Faso | iLiNS−Zinc (24) | DMC | 459 | 163 | 15.3 | −0.08 (−0.13, −0.02) | 0.05 | 0.06 | 287 | 212 | 16.0 | −0.09 (−0.14, −0.05) | 0.08 | 0.08 |
| Ghana | GHANA (25) | | | | | | | | | | | | | |
| Ghana | iLiNS−DYADG (26) | CDI | 152 | 341 | 7.9 | 0.00 (−0.05, 0.05) | 0.05 | 0.07 | 179 | 315 | 11.7 | 0.01 (−0.05, 0.07) | 0.05 | 0.06 |
| Haiti | HAITI (27) | Total sounds | | | | | | | | | | | | |
| Kenya | WASH−B (28) | EASQ | 778 | 2706 | 8.5 | 0.01 (−0.02, 0.03) | 0.23 | 0.16 | 582 | 2036 | 12.2 | −0.02 (−0.05, 0.01) | 0.24 | 0.21 |
| Madagascar | MAHAY (29) | ASQI | 741 | 845 | 5.0 | 0.02 (−0.02, 0.05) | 0.09 | 0.10 | 823 | 724 | 14.2 | 0.00 (−0.06, 0.07) | 0.04 | 0.05 |
| Malawi | iLiNS−DYADM (30) | CDI | 118 | 230 | 9.1 | −0.02 (−0.08, 0.04) | 0.03 | 0.05 | 97 | 208 | 12.5 | 0.00 (−0.08, 0.08) | 0.03 | 0.03 |
| Malawi | iLiNS−DOSE (31) | CDI | 278 | 94 | 9.6 | 0.01 (−0.06, 0.08) | 0.03 | 0.04 | 271 | 96 | 11.5 | −0.01 (−0.09, 0.06) | 0.04 | 0.04 |
| Mali | PROMIS CS (32) | DMC | 494 | 450 | 9.6 | −0.01 (−0.06, 0.03) | 0.07 | 0.09 | 433 | 494 | 13.0 | −0.03 (−0.08, 0.01) | 0.09 | 0.09 |
| Zimbabwe | SHINE (HIV−) (33) | CDI | 396 | 351 | 10.3 | −0.02 (−0.06, 0.03) | 0.06 | 0.08 | 375 | 383 | 12.0 | −0.01 (−0.05, 0.03) | 0.11 | 0.12 |
| Zimbabwe | SHINE (HIV+) (34) | CDI | 86 | 77 | 15.6 | −0.09 (−0.19, 0.02) | 0.01 | 0.02 | 74 | 64 | 9.4 | 0.01 (−0.09, 0.12) | 0.02 | 0.02 |
| | | | **5140** | **7444** | | **I² = 0.28, Tau² = 0.00** | | | **4699** | **6655** | | **I² = 0.10, Tau² = 0.00** | | |
| **Fixed** | | | | | | **−0.01 (−0.02, 0.00)** | | | | | | **−0.03 (−0.04, −0.01)** | | |
| **Random** | | | | | | **−0.01 (−0.03, 0.00)** | | | | | | **−0.03 (−0.04, −0.01)** | | |

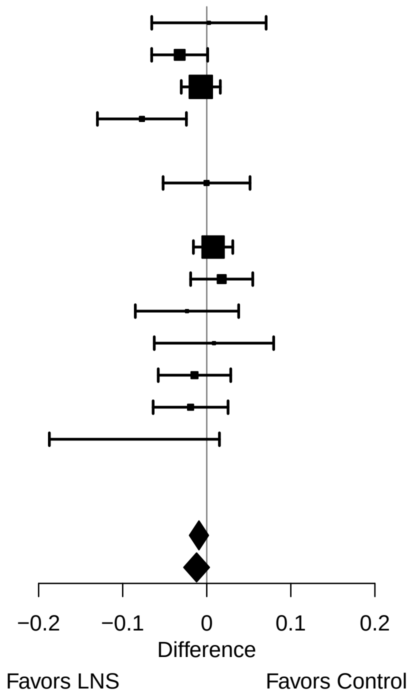

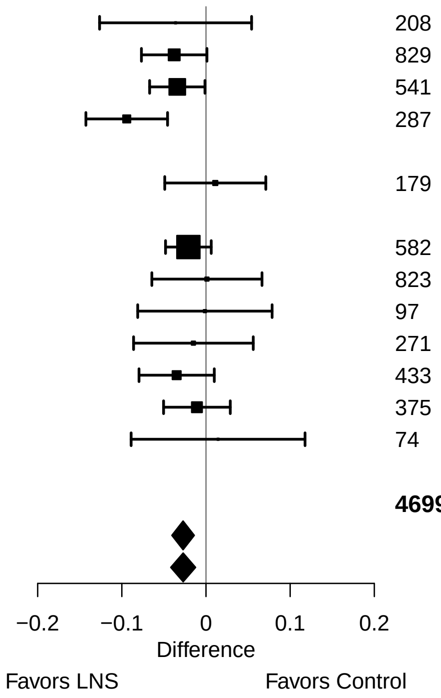

#### Supplemental figure 6C: Language lowest decile prevalence difference

#### 6C2: Stratified by Household food insecurity

**P−for−interaction = 0.269**
**Difference in PDs = −0.01 (−0.04, 0.01)**

| Country | Trial | Tool | LNS N | Control N | Control Prevalence | Mild to secure PD (95% CI) | Fixed W | Random W | LNS N | Control N | Control Prevalence | Moderate to severe PD (95% CI) | Fixed W | Random W |
|---|---|---|---|---|---|---|---|---|---|---|---|---|---|---|
| Bangladesh | JiVitA−4 (21) | BSID−III | | | | | | | | | | | | |
| Bangladesh | RDNS (22) | CDI | 1058 | 493 | 9.9 | −0.03 (−0.06, 0.00) | 0.12 | 0.14 | 605 | 321 | 16.2 | −0.05 (−0.09, 0.00) | 0.21 | 0.18 |
| Bangladesh | WASH−B (23) | CDI | 883 | 2608 | 9.6 | −0.02 (−0.04, 0.00) | 0.27 | 0.15 | 226 | 745 | 13.7 | −0.02 (−0.07, 0.03) | 0.14 | 0.14 |
| Burkina Faso | iLiNS−Zinc (24) | DMC | 389 | 162 | 15.4 | −0.09 (−0.13, −0.05) | 0.06 | 0.12 | 357 | 213 | 16.0 | −0.08 (−0.12, −0.03) | 0.18 | 0.16 |
| Ghana | GHANA (25) | | | | | | | | | | | | | |
| Ghana | iLiNS−DYADG (26) | CDI | 238 | 453 | 8.4 | 0.04 (−0.01, 0.08) | 0.05 | 0.11 | 92 | 202 | 12.9 | −0.06 (−0.14, 0.01) | 0.06 | 0.08 |
| Haiti | HAITI (27) | Total sounds | | | | | | | | | | | | |
| Kenya | WASH−B (28) | EASQ | 1214 | 4244 | 10.0 | −0.01 (−0.03, 0.01) | 0.33 | 0.15 | 146 | 487 | 11.3 | 0.04 (−0.03, 0.10) | 0.08 | 0.10 |
| Madagascar | MAHAY (29) | ASQI | 940 | 892 | 8.0 | 0.03 (−0.02, 0.08) | 0.04 | 0.10 | 336 | 389 | 12.6 | −0.01 (−0.09, 0.07) | 0.06 | 0.08 |
| Malawi | iLiNS−DYADM (30) | CDI | 68 | 118 | 9.3 | 0.04 (−0.05, 0.13) | 0.01 | 0.05 | 146 | 315 | 10.5 | −0.03 (−0.09, 0.03) | 0.11 | 0.12 |
| Malawi | iLiNS−DOSE (31) | CDI | 133 | 51 | 9.8 | −0.02 (−0.11, 0.07) | 0.01 | 0.05 | 403 | 133 | 10.5 | 0.00 (−0.06, 0.06) | 0.10 | 0.11 |
| Mali | PROMIS CS (32) | DMC | | | | | | | | | | | | |
| Zimbabwe | SHINE (HIV−) (33) | CDI | 612 | 576 | 10.6 | −0.02 (−0.05, 0.01) | 0.10 | 0.13 | 136 | 148 | 14.2 | 0.01 (−0.09, 0.10) | 0.04 | 0.05 |
| Zimbabwe | SHINE (HIV+) (34) | CDI | | | | | | | | | | | | |
| | | | **5535** | **9597** | | **$I^2$ = 0.66, $Tau^2$ = 0.00** | | | **2447** | **2953** | | **$I^2$ = 0.32, $Tau^2$ = 0.00** | | |
| **Fixed** | | | | | | **−0.02 (−0.03, −0.01)** | | | | | | **−0.03 (−0.05, −0.01)** | | |
| **Random** | | | | | | **−0.01 (−0.04, 0.01)** | | | | | | **−0.03 (−0.05, −0.01)** | | |

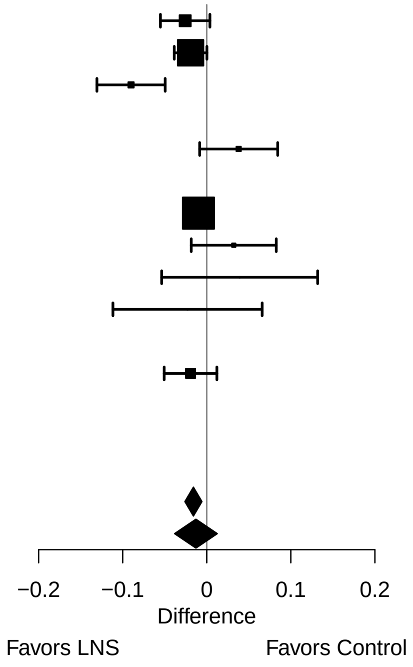

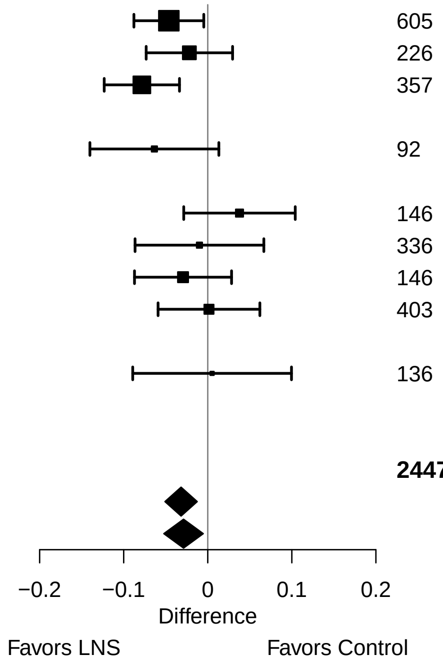

#### Supplemental figure 6C: Language lowest decile prevalence difference

##### 6C3: Stratified by Household source water quality

**P−for−interaction = 0.321**
**Difference in PDs = −0.01 (−0.04, 0.01)**

| Country | Trial | Tool | LNS N | Control N | Control Prevalence | Improved PD (95% CI) | Fixed W | Random W | LNS N | Control N | Control Prevalence | Unimproved PD (95% CI) | Fixed W | Random W |
|---|---|---|---|---|---|---|---|---|---|---|---|---|---|---|
| Bangladesh | JiVitA−4 (21) | BSID−III | | | | | | | | | | | | |
| Bangladesh | RDNS (22) | CDI | | | | | | | | | | | | |
| Bangladesh | WASH−B (23) | CDI | 480 | 981 | 10.9 | −0.04 (−0.07, −0.01) | 0.36 | 0.28 | 67 | 117 | 16.2 | −0.07 (−0.19, 0.05) | 0.03 | 0.07 |
| Burkina Faso | iLiNS−Zinc (24) | DMC | 220 | 82 | 12.2 | −0.07 (−0.12, −0.02) | 0.11 | 0.14 | 526 | 293 | 16.7 | −0.09 (−0.13, −0.05) | 0.25 | 0.21 |
| Ghana | GHANA (25) | | | | | | | | | | | | | |
| Ghana | iLiNS−DYADG (26) | CDI | | | | | | | | | | | | |
| Haiti | HAITI (27) | Total sounds | | | | | | | | | | | | |
| Kenya | WASH−B (28) | EASQ | 413 | 1419 | 10.4 | 0.00 (−0.04, 0.04) | 0.21 | 0.21 | 236 | 649 | 10.5 | −0.01 (−0.05, 0.02) | 0.30 | 0.22 |
| Madagascar | MAHAY (29) | ASQI | 405 | 430 | 12.3 | 0.02 (−0.07, 0.11) | 0.03 | 0.05 | 1134 | 1133 | 8.1 | 0.01 (−0.03, 0.06) | 0.23 | 0.21 |
| Malawi | iLiNS−DYADM (30) | CDI | | | | | | | | | | | | |
| Malawi | iLiNS−DOSE (31) | CDI | | | | | | | | | | | | |
| Mali | PROMIS CS (32) | DMC | 528 | 552 | 9.8 | −0.01 (−0.05, 0.02) | 0.19 | 0.20 | 357 | 378 | 14.0 | −0.04 (−0.10, 0.01) | 0.14 | 0.18 |
| Zimbabwe | SHINE (HIV−) (33) | CDI | 220 | 212 | 10.4 | −0.01 (−0.06, 0.05) | 0.10 | 0.12 | 125 | 122 | 12.3 | 0.02 (−0.06, 0.11) | 0.06 | 0.11 |
| Zimbabwe | SHINE (HIV+) (34) | CDI | | | | | | | | | | | | |
| | | | **2266** | **3676** | | **I² = 0.29, Tau² = 0.00** | | | **2445** | **2692** | | **I² = 0.67, Tau² = 0.00** | | |
| **Fixed** | | | | | | **−0.02 (−0.04, −0.01)** | | | | | | **−0.03 (−0.05, −0.01)** | | |
| **Random** | | | | | | **−0.02 (−0.04, 0.00)** | | | | | | **−0.03 (−0.06, 0.01)** | | |

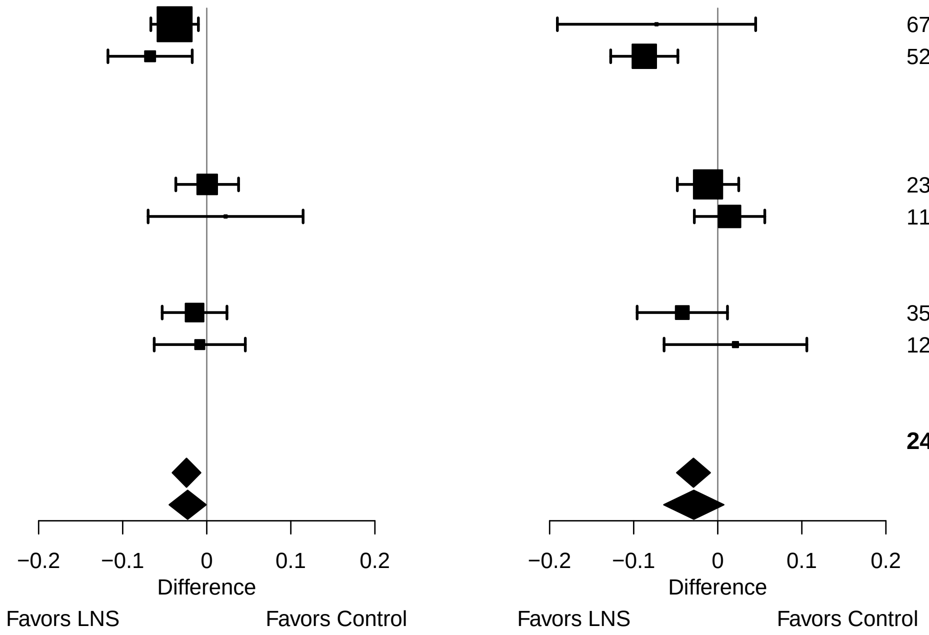

#### Supplemental figure 6C: Language lowest decile prevalence difference

#### 6C4: Stratified by Household sanitation

**P−for−interaction = 0.848**
**Difference in PDs = 0.00 (−0.04, 0.04)**

| Country | Trial | Tool | LNS N | Control N | Control Prevalence | Improved PD (95% CI) | Fixed W | Random W | LNS N | Control N | Control Prevalence | Unimproved PD (95% CI) | Fixed W | Random W |
|---|---|---|---|---|---|---|---|---|---|---|---|---|---|---|
| Bangladesh | JiVitA−4 (21) | BSID−III | | | | | | | | | | | | |
| Bangladesh | RDNS (22) | CDI | 1179 | 572 | 10.8 | −0.03 (−0.06, 0.00) | 0.57 | 0.57 | 482 | 241 | 16.2 | −0.05 (−0.10, 0.00) | 0.39 | 0.37 |
| Bangladesh | WASH−B (23) | CDI | | | | | | | | | | | | |
| Burkina Faso | iLiNS−Zinc (24) | DMC | | | | | | | | | | | | |
| Ghana | GHANA (25) | | | | | | | | | | | | | |
| Ghana | iLiNS−DYADG (26) | CDI | | | | | | | | | | | | |
| Haiti | HAITI (27) | Total sounds | | | | | | | | | | | | |
| Kenya | WASH−B (28) | EASQ | | | | | | | | | | | | |
| Madagascar | MAHAY (29) | ASQI | | | | | | | | | | | | |
| Malawi | iLiNS−DYADM (30) | CDI | | | | | | | | | | | | |
| Malawi | iLiNS−DOSE (31) | CDI | | | | | | | | | | | | |
| Mali | PROMIS CS (32) | DMC | 687 | 678 | 10.5 | −0.02 (−0.05, 0.02) | 0.33 | 0.33 | 211 | 242 | 12.4 | −0.04 (−0.10, 0.02) | 0.28 | 0.31 |
| Zimbabwe | SHINE (HIV−) (33) | CDI | 127 | 106 | 9.4 | −0.04 (−0.10, 0.02) | 0.10 | 0.10 | 217 | 229 | 11.8 | 0.02 (−0.03, 0.08) | 0.32 | 0.33 |
| Zimbabwe | SHINE (HIV+) (34) | CDI | | | | | | | | | | | | |
| | | | **1993** | **1356** | | **I² = 0.00, Tau² = 0.00** | | | **910** | **712** | | **I² = 0.46, Tau² = 0.00** | | |
| **Fixed** | | | | | | **−0.03 (−0.05, −0.01)** | | | | | | **−0.02 (−0.05, 0.01)** | | |
| **Random** | | | | | | **−0.03 (−0.05, −0.01)** | | | | | | **−0.02 (−0.07, 0.02)** | | |

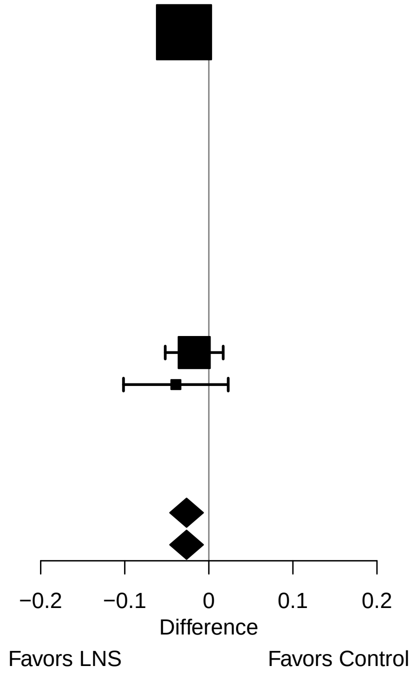

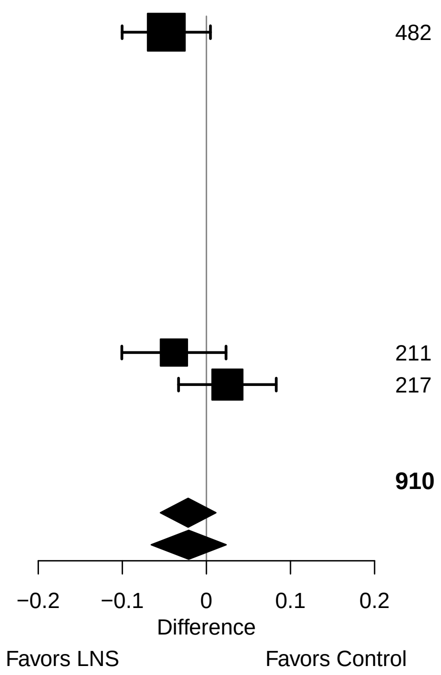

#### Supplemental figure 6C: Language lowest decile prevalence difference

#### 6C5: Stratified by Home environment

**P−for−interaction = 0.364**
**Difference in PDs = −0.01 (−0.03, 0.01)**

| Country | Trial | Tool | At least median: LNS N | Control N | Control Prevalence | PD (95% CI) | Fixed W | Random W | Less than median: LNS N | Control N | Control Prevalence | PD (95% CI) | Fixed W | Random W |
|---|---|---|---|---|---|---|---|---|---|---|---|---|---|---|
| Bangladesh | JiVitA−4 (21) | BSID−III | | | | | | | | | | | | |
| Bangladesh | RDNS (22) | CDI | 1192 | 581 | 9.0 | −0.04 (−0.06, −0.01) | 0.14 | 0.15 | 468 | 230 | 20.4 | −0.02 (−0.10, 0.05) | 0.07 | 0.11 |
| Bangladesh | WASH−B (23) | CDI | 630 | 1652 | 5.9 | −0.02 (−0.03, 0.00) | 0.30 | 0.22 | 479 | 1700 | 15.1 | −0.01 (−0.05, 0.03) | 0.29 | 0.18 |
| Burkina Faso | iLiNS−Zinc (24) | DMC | 473 | 199 | 5.5 | 0.00 (−0.03, 0.02) | 0.16 | 0.16 | 272 | 175 | 27.4 | −0.16 (−0.25, −0.08) | 0.06 | 0.10 |
| Ghana | GHANA (25) | | | | | | | | | | | | | |
| Ghana | iLiNS−DYADG (26) | CDI | 213 | 410 | 6.3 | 0.00 (−0.04, 0.04) | 0.05 | 0.07 | 118 | 248 | 15.3 | 0.02 (−0.06, 0.11) | 0.06 | 0.11 |
| Haiti | HAITI (27) | Total sounds | | | | | | | | | | | | |
| Kenya | WASH−B (28) | EASQ | 809 | 2682 | 6.9 | 0.01 (−0.01, 0.03) | 0.22 | 0.19 | 552 | 2060 | 14.3 | −0.02 (−0.05, 0.02) | 0.34 | 0.18 |
| Madagascar | MAHAY (29) | ASQI | 843 | 839 | 7.7 | 0.01 (−0.03, 0.04) | 0.06 | 0.08 | 770 | 764 | 10.9 | 0.03 (−0.04, 0.09) | 0.10 | 0.13 |
| Malawi | iLiNS−DYADM (30) | CDI | 137 | 290 | 6.6 | 0.00 (−0.05, 0.05) | 0.03 | 0.05 | 78 | 149 | 18.8 | −0.05 (−0.15, 0.06) | 0.04 | 0.08 |
| Malawi | iLiNS−DOSE (31) | CDI | 375 | 135 | 5.9 | −0.01 (−0.05, 0.03) | 0.04 | 0.07 | 269 | 86 | 17.4 | 0.00 (−0.10, 0.09) | 0.05 | 0.10 |
| Mali | PROMIS CS (32) | DMC | | | | | | | | | | | | |
| Zimbabwe | SHINE (HIV−) (33) | CDI | | | | | | | | | | | | |
| Zimbabwe | SHINE (HIV+) (34) | CDI | | | | | | | | | | | | |
| | | | **4672** | **6788** | | **I² = 0.24, Tau² = 0.00** | | | **3006** | **5412** | | **I² = 0.51, Tau² = 0.00** | | |
| **Fixed** | | | | | | **−0.01 (−0.02, 0.00)** | | | | | | **−0.02 (−0.04, 0.00)** | | |
| **Random** | | | | | | **−0.01 (−0.02, 0.00)** | | | | | | **−0.02 (−0.06, 0.02)** | | |

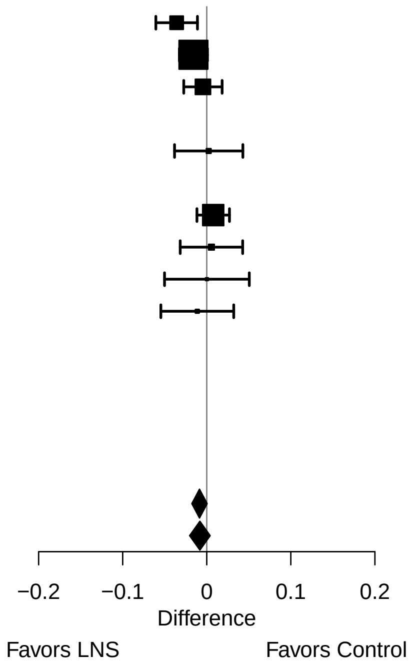

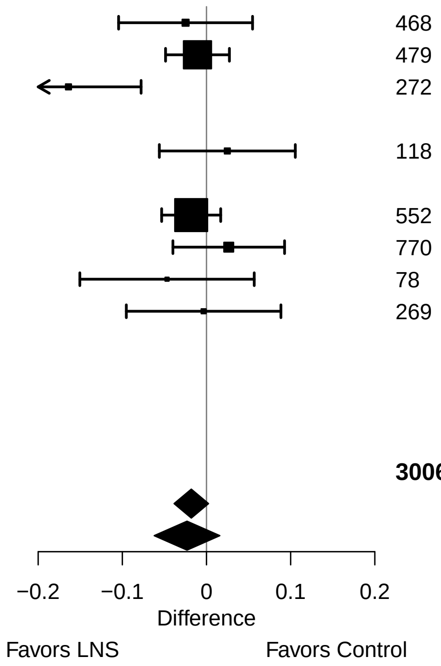

#### Supplemental figure 6C: Language lowest decile prevalence difference

#### 6C6: Stratified by Season at the time of assessment

**P−for−interaction = 0.556**
**Difference in PDs = −0.01 (−0.03, 0.01)**

| Country | Trial | Tool | Dry LNS N | Dry Control N | Dry Control Prevalence | Dry PD (95% CI) | Dry Fixed W | Dry Random W | Rainy LNS N | Rainy Control N | Rainy Control Prevalence | Rainy PD (95% CI) | Rainy Fixed W | Rainy Random W |
|---|---|---|---|---|---|---|---|---|---|---|---|---|---|---|
| Bangladesh | JiVitA−4 (21) | BSID−III | 251 | 70 | 11.4 | −0.05 (−0.13, 0.04) | 0.02 | 0.04 | 194 | 73 | 11.0 | 0.02 (−0.06, 0.10) | 0.03 | 0.07 |
| Bangladesh | RDNS (22) | CDI | 739 | 359 | 12.0 | −0.02 (−0.05, 0.02) | 0.16 | 0.15 | 924 | 455 | 12.7 | −0.05 (−0.08, −0.01) | 0.15 | 0.17 |
| Bangladesh | WASH−B (23) | CDI | 629 | 1944 | 9.4 | −0.01 (−0.03, 0.02) | 0.33 | 0.22 | 480 | 1409 | 12.1 | −0.04 (−0.07, −0.01) | 0.21 | 0.19 |
| Burkina Faso | iLiNS−Zinc (24) | DMC | 514 | 274 | 16.1 | −0.09 (−0.13, −0.04) | 0.10 | 0.12 | 232 | 101 | 14.9 | −0.08 (−0.19, 0.04) | 0.01 | 0.04 |
| Ghana | GHANA (25) | | | | | | | | | | | | | |
| Ghana | iLiNS−DYADG (26) | CDI | 194 | 388 | 10.1 | −0.02 (−0.07, 0.03) | 0.08 | 0.10 | 137 | 270 | 9.3 | 0.05 (−0.01, 0.12) | 0.04 | 0.09 |
| Haiti | HAITI (27) | Total sounds | | | | | | | | | | | | |
| Kenya | WASH−B (28) | EASQ | 315 | 1059 | 13.5 | 0.00 (−0.05, 0.05) | 0.07 | 0.09 | 1046 | 3682 | 9.2 | −0.01 (−0.03, 0.01) | 0.45 | 0.22 |
| Madagascar | MAHAY (29) | ASQI | | | | | | | | | | | | |
| Malawi | iLiNS−DYADM (30) | CDI | 153 | 311 | 10.3 | 0.00 (−0.06, 0.05) | 0.06 | 0.08 | 62 | 127 | 11.8 | −0.04 (−0.13, 0.06) | 0.02 | 0.05 |
| Malawi | iLiNS−DOSE (31) | CDI | 394 | 118 | 12.7 | −0.04 (−0.10, 0.03) | 0.05 | 0.07 | 244 | 100 | 8.0 | 0.03 (−0.04, 0.10) | 0.04 | 0.08 |
| Mali | PROMIS CS (32) | DMC | | | | | | | | | | | | |
| Zimbabwe | SHINE (HIV−) (33) | CDI | 582 | 555 | 11.4 | −0.01 (−0.05, 0.03) | 0.14 | 0.14 | 235 | 226 | 9.3 | −0.02 (−0.08, 0.04) | 0.05 | 0.10 |
| Zimbabwe | SHINE (HIV+) (34) | CDI | | | | | | | | | | | | |
| | | | **3771** | **5078** | | **$I^2$ = 0.32, $Tau^2$ = 0.00** | | | **3554** | **6443** | | **$I^2$ = 0.44, $Tau^2$ = 0.00** | | |
| **Fixed** | | | | | | **−0.02 (−0.03, −0.01)** | | | | | | **−0.02 (−0.03, 0.00)** | | |
| **Random** | | | | | | **−0.02 (−0.04, 0.00)** | | | | | | **−0.01 (−0.04, 0.01)** | | |

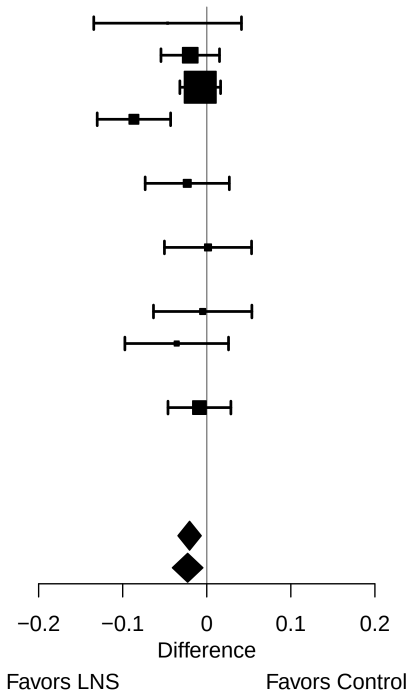

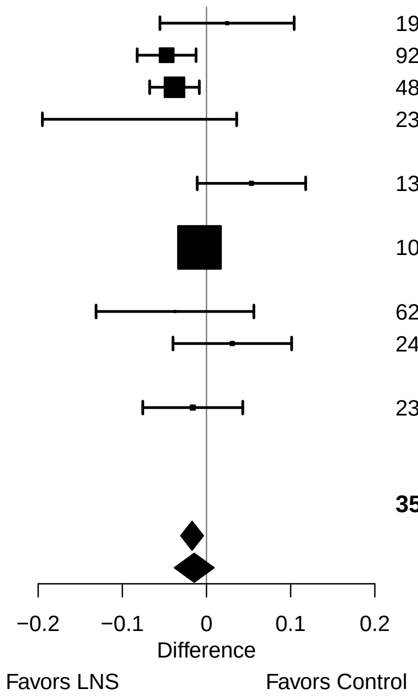

### Supplemental figure 6D: Mean difference in social-emotional z-score

#### 6D1: Stratified by Household socio-economic status

**P−for−interaction = 0.982**
**Difference in MDs = 0.00 (−0.06, 0.06)**

| | | | At least median | | | | | | Less than median | | | | | |
|---|---|---|---|---|---|---|---|---|---|---|---|---|---|---|
| **Country** | **Trial** | **Tool** | **LNS N** | **Control N** | **Control Mean** | **MD (95% CI)** | **Fixed W** | **Random W** | **LNS N** | **Control N** | **Control Mean** | **MD (95% CI)** | **Fixed W** | **Random W** |
| Bangladesh | JiVitA−4 (21) | | | | | | | | | | | | | |
| Bangladesh | RDNS (22) | DMC | 830 | 407 | 0.00 | 0.09 (−0.06, 0.23) | 0.09 | 0.11 | 827 | 408 | −0.07 | 0.04 (−0.09, 0.16) | 0.14 | 0.14 |
| Bangladesh | WASH−B (23) | EASQ | 545 | 1659 | 0.11 | 0.09 (−0.01, 0.19) | 0.19 | 0.12 | 521 | 1607 | −0.17 | 0.16 (0.05, 0.26) | 0.20 | 0.17 |
| Burkina Faso | iLiNS−Zinc (24) | DMC | 459 | 163 | −0.25 | 0.41 (0.14, 0.69) | 0.03 | 0.06 | 287 | 212 | −0.22 | 0.28 (0.05, 0.51) | 0.04 | 0.05 |
| Ghana | GHANA (25) | | | | | | | | | | | | | |
| Ghana | iLiNS−DYADG (26) | PSED | 153 | 341 | 0.00 | 0.00 (−0.19, 0.19) | 0.05 | 0.09 | 179 | 314 | 0.00 | 0.01 (−0.17, 0.20) | 0.06 | 0.08 |
| Haiti | HAITI (27) | | | | | | | | | | | | | |
| Kenya | WASH−B (28) | EASQ | 778 | 2706 | 0.06 | 0.01 (−0.07, 0.08) | 0.32 | 0.13 | 582 | 2036 | −0.08 | 0.02 (−0.07, 0.11) | 0.25 | 0.20 |
| Madagascar | MAHAY (29) | ASQI | 741 | 845 | 0.20 | −0.06 (−0.24, 0.12) | 0.06 | 0.09 | 823 | 724 | −0.16 | −0.04 (−0.26, 0.18) | 0.04 | 0.05 |
| Malawi | iLiNS−DYADM (30) | PSED | 118 | 230 | 0.00 | 0.08 (−0.15, 0.31) | 0.04 | 0.07 | 97 | 207 | −0.02 | −0.06 (−0.29, 0.18) | 0.04 | 0.05 |
| Malawi | iLiNS−DOSE (31) | PSED | 278 | 94 | −0.02 | 0.00 (−0.23, 0.22) | 0.04 | 0.07 | 270 | 96 | 0.01 | 0.06 (−0.18, 0.30) | 0.04 | 0.05 |
| Mali | PROMIS CS (32) | DMC | 494 | 450 | 0.03 | 0.05 (−0.14, 0.25) | 0.05 | 0.08 | 433 | 494 | −0.17 | 0.23 (0.03, 0.42) | 0.05 | 0.07 |
| Zimbabwe | SHINE (HIV−) (33) | MDAT | 406 | 359 | −0.07 | 0.22 (0.08, 0.35) | 0.10 | 0.11 | 389 | 391 | −0.08 | 0.07 (−0.07, 0.21) | 0.11 | 0.11 |
| Zimbabwe | SHINE (HIV+) (34) | MDAT | 91 | 78 | −0.23 | 0.45 (0.18, 0.72) | 0.03 | 0.06 | 74 | 67 | −0.07 | 0.19 (−0.12, 0.49) | 0.02 | 0.03 |
| | | | **4893** | **7332** | | **I² = 0.57, Tau² = 0.01** | | | **4482** | **6556** | | **I² = 0.24, Tau² = 0.00** | | |
| **Fixed** | | | | | | **0.07 (0.03, 0.12)** | | | | | | **0.08 (0.03, 0.12)** | | |
| **Random** | | | | | | **0.10 (0.01, 0.19)** | | | | | | **0.08 (0.02, 0.13)** | | |

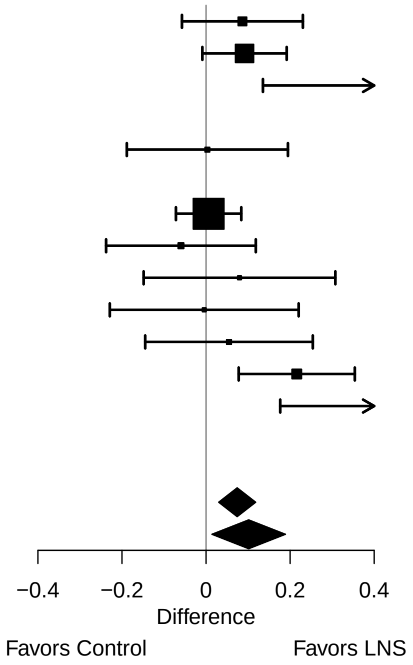

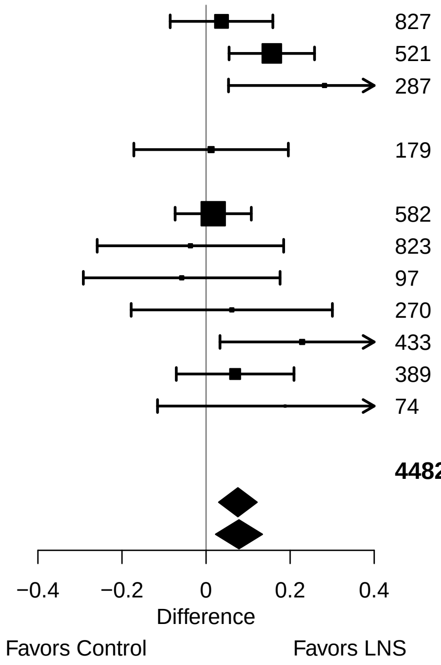

#### Supplemental figure 6D: Mean difference in social-emotional z-score

#### 6D2: Stratified by Household food insecurity

**P−for−interaction = 0.347**
**Difference in MDs = −0.03 (−0.10, 0.04)**

| Country | Trial | Tool | Mild to secure: LNS N | Control N | Control Mean | MD (95% CI) | Fixed W | Random W | Moderate to severe: LNS N | Control N | Control Mean | MD (95% CI) | Fixed W | Random W |
|---|---|---|---|---|---|---|---|---|---|---|---|---|---|---|
| Bangladesh | JiVitA−4 (21) | | | | | | | | | | | | | |
| Bangladesh | RDNS (22) | DMC | 1054 | 493 | 0.02 | 0.07 (−0.04, 0.18) | 0.12 | 0.12 | 603 | 322 | −0.11 | 0.04 (−0.08, 0.16) | 0.23 | 0.17 |
| Bangladesh | WASH−B (23) | EASQ | 853 | 2539 | 0.01 | 0.13 (0.05, 0.21) | 0.25 | 0.13 | 213 | 727 | −0.17 | 0.08 (−0.07, 0.24) | 0.14 | 0.13 |
| Burkina Faso | iLiNS−Zinc (24) | DMC | 389 | 162 | −0.22 | 0.36 (0.09, 0.64) | 0.02 | 0.08 | 357 | 213 | −0.25 | 0.34 (0.17, 0.51) | 0.12 | 0.12 |
| Ghana | GHANA (25) | | | | | | | | | | | | | |
| Ghana | iLiNS−DYADG (26) | PSED | 239 | 452 | 0.03 | 0.00 (−0.15, 0.15) | 0.06 | 0.11 | 92 | 202 | −0.08 | 0.02 (−0.23, 0.28) | 0.05 | 0.07 |
| Haiti | HAITI (27) | | | | | | | | | | | | | |
| Kenya | WASH−B (28) | EASQ | 1214 | 4244 | 0.00 | 0.02 (−0.05, 0.08) | 0.36 | 0.13 | 146 | 487 | −0.07 | −0.03 (−0.22, 0.15) | 0.10 | 0.11 |
| Madagascar | MAHAY (29) | ASQI | 940 | 892 | 0.06 | −0.08 (−0.29, 0.13) | 0.03 | 0.09 | 336 | 389 | −0.11 | −0.05 (−0.28, 0.18) | 0.07 | 0.08 |
| Malawi | iLiNS−DYADM (30) | PSED | 68 | 118 | 0.06 | −0.11 (−0.43, 0.20) | 0.02 | 0.07 | 146 | 314 | −0.04 | 0.08 (−0.12, 0.27) | 0.10 | 0.10 |
| Malawi | iLiNS−DOSE (31) | PSED | 134 | 51 | 0.03 | −0.11 (−0.43, 0.20) | 0.01 | 0.06 | 402 | 133 | −0.04 | 0.10 (−0.09, 0.30) | 0.09 | 0.10 |
| Mali | PROMIS CS (32) | DMC | | | | | | | | | | | | |
| Zimbabwe | SHINE (HIV−) (33) | MDAT | 631 | 586 | −0.09 | 0.18 (0.06, 0.30) | 0.10 | 0.12 | 140 | 154 | −0.02 | −0.06 (−0.27, 0.15) | 0.08 | 0.09 |
| Zimbabwe | SHINE (HIV+) (34) | MDAT | 119 | 105 | −0.20 | 0.46 (0.23, 0.69) | 0.03 | 0.09 | 41 | 37 | −0.10 | 0.01 (−0.41, 0.44) | 0.02 | 0.03 |
| | | | **5641** | **9642** | | **I² = 0.69, Tau² = 0.02** | | | **2476** | **2978** | | **I² = 0.36, Tau² = 0.00** | | |
| **Fixed** | | | | | | **0.08 (0.04, 0.12)** | | | | | | **0.07 (0.01, 0.13)** | | |
| **Random** | | | | | | **0.09 (−0.02, 0.21)** | | | | | | **0.07 (−0.01, 0.14)** | | |

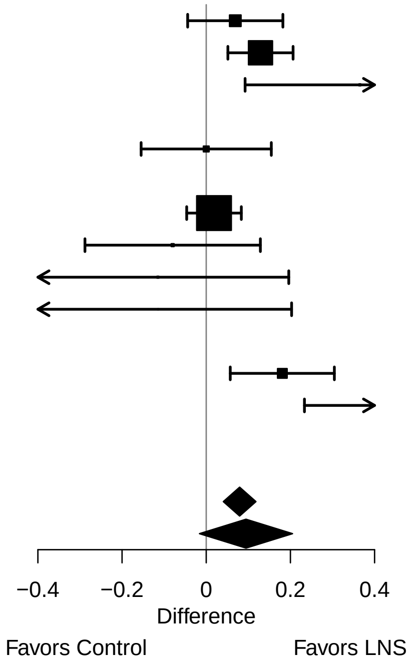

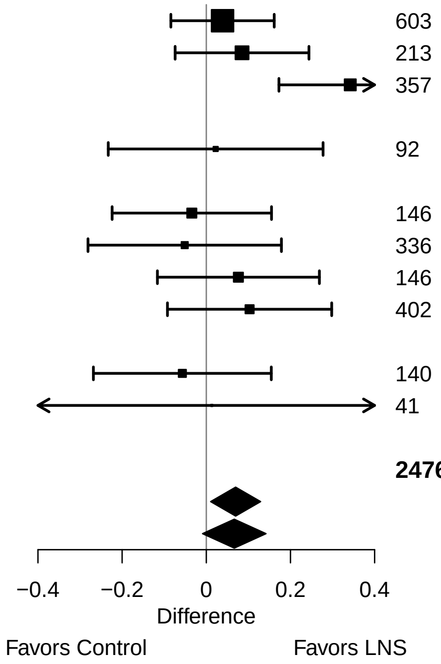

#### Supplemental figure 6D: Mean difference in social-emotional z-score

#### 6D3: Stratified by Household source water quality

**P−for−interaction = 0.939**
**Difference in MDs = 0.00 (−0.09, 0.10)**

| Country | Trial | Tool | LNS N | Control N | Control Mean | Improved MD (95% CI) | Fixed W | Random W | LNS N | Control N | Control Mean | Unimproved MD (95% CI) | Fixed W | Random W |
|---|---|---|---|---|---|---|---|---|---|---|---|---|---|---|
| Bangladesh | JiVitA−4 (21) | | | | | | | | | | | | | |
| Bangladesh | RDNS (22) | DMC | | | | | | | | | | | | |
| Bangladesh | WASH−B (23) | EASQ | 461 | 981 | −0.17 | 0.22 (0.11, 0.34) | 0.25 | 0.17 | 66 | 117 | −0.33 | 0.26 (−0.02, 0.55) | 0.07 | 0.10 |
| Burkina Faso | iLiNS−Zinc (24) | DMC | 220 | 82 | −0.11 | 0.32 (0.10, 0.54) | 0.07 | 0.09 | 526 | 293 | −0.27 | 0.35 (0.17, 0.54) | 0.16 | 0.16 |
| Ghana | GHANA (25) | | | | | | | | | | | | | |
| Ghana | iLiNS−DYADG (26) | PSED | | | | | | | | | | | | |
| Haiti | HAITI (27) | | | | | | | | | | | | | |
| Kenya | WASH−B (28) | EASQ | 413 | 1419 | 0.02 | −0.04 (−0.16, 0.09) | 0.21 | 0.16 | 236 | 649 | 0.04 | −0.01 (−0.15, 0.13) | 0.29 | 0.20 |
| Madagascar | MAHAY (29) | ASQI | 405 | 430 | −0.04 | −0.05 (−0.41, 0.31) | 0.03 | 0.05 | 1134 | 1133 | 0.06 | −0.09 (−0.28, 0.10) | 0.16 | 0.16 |
| Malawi | iLiNS−DYADM (30) | PSED | 195 | 400 | 0.01 | 0.01 (−0.15, 0.18) | 0.12 | 0.13 | 20 | 36 | −0.26 | 0.08 (−0.56, 0.72) | 0.01 | 0.03 |
| Malawi | iLiNS−DOSE (31) | PSED | 514 | 177 | 0.00 | 0.03 (−0.14, 0.21) | 0.11 | 0.12 | 44 | 15 | −0.06 | −0.06 (−0.58, 0.46) | 0.02 | 0.04 |
| Mali | PROMIS CS (32) | DMC | 528 | 552 | −0.04 | 0.12 (−0.09, 0.33) | 0.08 | 0.10 | 357 | 378 | −0.15 | 0.17 (−0.05, 0.39) | 0.12 | 0.13 |
| Zimbabwe | SHINE (HIV−) (33) | MDAT | 228 | 220 | −0.04 | 0.11 (−0.08, 0.30) | 0.09 | 0.11 | 131 | 122 | −0.13 | 0.09 (−0.12, 0.30) | 0.12 | 0.14 |
| Zimbabwe | SHINE (HIV+) (34) | MDAT | 37 | 41 | −0.25 | 0.30 (0.00, 0.60) | 0.04 | 0.06 | 25 | 25 | 0.05 | −0.07 (−0.48, 0.34) | 0.03 | 0.06 |
| | | | **3001** | **4302** | | **I² = 0.53, Tau² = 0.01** | | | **2539** | **2768** | | **I² = 0.51, Tau² = 0.01** | | |
| **Fixed** | | | | | | **0.11 (0.05, 0.16)** | | | | | | **0.09 (0.01, 0.16)** | | |
| **Random** | | | | | | **0.11 (0.02, 0.20)** | | | | | | **0.10 (−0.01, 0.20)** | | |

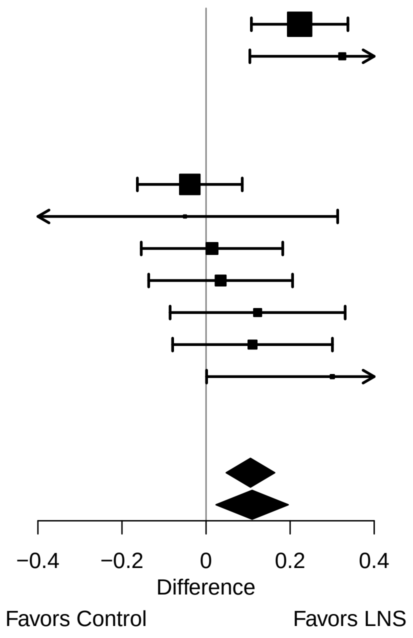

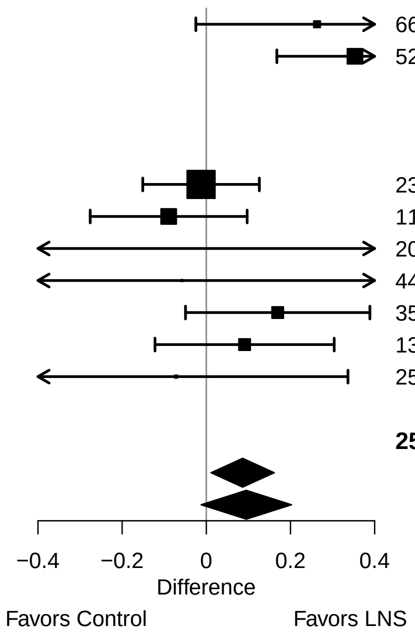

##### Supplemental figure 6D: Mean difference in social-emotional z-score

#### 6D4: Stratified by Household sanitation

**P−for−interaction = 0.550**
**Difference in MDs = −0.03 (−0.14, 0.07)**

| Country | Trial | Tool | LNS N | Control N | Control Mean | Improved MD (95% CI) | Fixed W | Random W | LNS N | Control N | Control Mean | Unimproved MD (95% CI) | Fixed W | Random W |
|---|---|---|---|---|---|---|---|---|---|---|---|---|---|---|
| Bangladesh | JiVitA−4 (21) | | | | | | | | | | | | | |
| Bangladesh | RDNS (22) | DMC | 1174 | 573 | −0.01 | 0.08 (−0.02, 0.19) | 0.30 | 0.14 | 481 | 241 | −0.09 | 0.01 (−0.16, 0.19) | 0.11 | 0.12 |
| Bangladesh | WASH−B (23) | EASQ | 474 | 998 | −0.17 | 0.23 (0.12, 0.34) | 0.27 | 0.14 | 27 | 53 | −0.35 | 0.17 (−0.28, 0.61) | 0.02 | 0.03 |
| Burkina Faso | iLiNS−Zinc (24) | DMC | 16 | 11 | −0.47 | 0.81 (0.30, 1.33) | 0.01 | 0.06 | 730 | 364 | −0.23 | 0.34 (0.17, 0.51) | 0.12 | 0.13 |
| Ghana | GHANA (25) | | | | | | | | | | | | | |
| Ghana | iLiNS−DYADG (26) | PSED | 321 | 639 | 0.00 | 0.00 (−0.14, 0.13) | 0.18 | 0.14 | 10 | 16 | −0.10 | 0.25 (−0.70, 1.20) | 0.00 | 0.01 |
| Haiti | HAITI (27) | | | | | | | | | | | | | |
| Kenya | WASH−B (28) | EASQ | 94 | 337 | 0.11 | −0.03 (−0.29, 0.22) | 0.05 | 0.11 | 555 | 1732 | 0.01 | −0.02 (−0.13, 0.08) | 0.30 | 0.21 |
| Madagascar | MAHAY (29) | ASQI | | | | | | | | | | | | |
| Malawi | iLiNS−DYADM (30) | PSED | 19 | 42 | 0.35 | −0.53 (−1.04, −0.03) | 0.01 | 0.07 | 196 | 394 | −0.05 | 0.07 (−0.10, 0.25) | 0.12 | 0.13 |
| Malawi | iLiNS−DOSE (31) | PSED | 12 | 9 | −0.09 | 0.37 (−0.35, 1.10) | 0.01 | 0.04 | 546 | 183 | 0.00 | 0.02 (−0.15, 0.18) | 0.12 | 0.13 |
| Mali | PROMIS CS (32) | DMC | 687 | 678 | −0.05 | 0.15 (−0.04, 0.34) | 0.09 | 0.12 | 211 | 242 | −0.10 | 0.11 (−0.12, 0.33) | 0.07 | 0.09 |
| Zimbabwe | SHINE (HIV−) (33) | MDAT | 133 | 108 | −0.06 | 0.07 (−0.16, 0.30) | 0.06 | 0.12 | 225 | 235 | −0.08 | 0.12 (−0.06, 0.30) | 0.11 | 0.12 |
| Zimbabwe | SHINE (HIV+) (34) | MDAT | 25 | 17 | −0.22 | 0.28 (−0.26, 0.82) | 0.01 | 0.06 | 37 | 49 | −0.11 | 0.10 (−0.23, 0.43) | 0.03 | 0.04 |
| | | | **2955** | **3412** | | **$I^2 = 0.61$, $Tau^2 = 0.05$** | | | **3018** | **3509** | | **$I^2 = 0.38$, $Tau^2 = 0.00$** | | |
| **Fixed** | | | | | | **0.11 (0.05, 0.17)** | | | | | | **0.07 (0.01, 0.13)** | | |
| **Random** | | | | | | **0.12 (−0.06, 0.29)** | | | | | | **0.09 (0.01, 0.16)** | | |

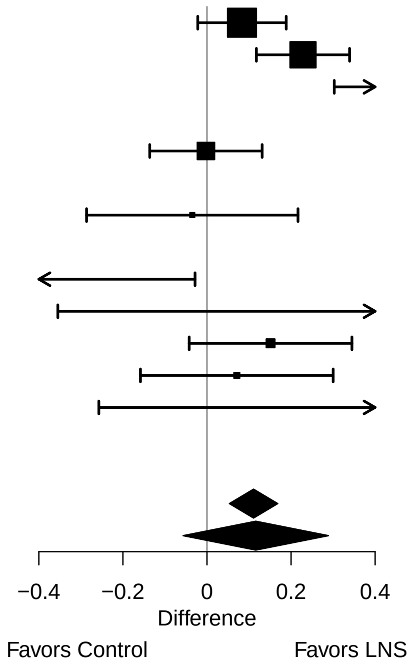

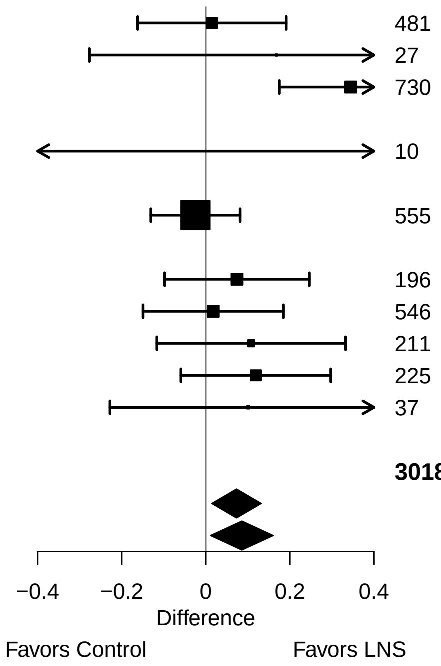

#### Supplemental figure 6D: Mean difference in social-emotional z-score

#### 6D5: Stratified by Home environment

**P−for−interaction = 0.363**
**Difference in MDs = 0.03 (−0.04, 0.10)**

| | | | At least median | | | | | | | Less than median | | | | |
|---|---|---|---|---|---|---|---|---|---|---|---|---|---|---|
| **Country** | **Trial** | **Tool** | **LNS N** | **Control N** | **Control Mean** | **MD (95% CI)** | **Fixed W** | **Random W** | **LNS N** | **Control N** | **Control Mean** | **MD (95% CI)** | **Fixed W** | **Random W** |
| Bangladesh | JiVitA−4 (21) | | | | | | | | | | | | | |
| Bangladesh | RDNS (22) | DMC | 1187 | 582 | 0.12 | 0.00 (−0.08, 0.09) | 0.23 | 0.17 | 467 | 230 | −0.39 | 0.20 (0.00, 0.40) | 0.08 | 0.10 |
| Bangladesh | WASH−B (23) | EASQ | 603 | 1599 | 0.15 | 0.13 (0.03, 0.23) | 0.18 | 0.16 | 463 | 1666 | −0.20 | 0.05 (−0.05, 0.16) | 0.30 | 0.25 |
| Burkina Faso | iLiNS−Zinc (24) | DMC | 473 | 199 | 0.00 | 0.28 (0.13, 0.43) | 0.08 | 0.12 | 272 | 175 | −0.50 | 0.35 (0.07, 0.63) | 0.04 | 0.06 |
| Ghana | GHANA (25) | | | | | | | | | | | | | |
| Ghana | iLiNS−DYADG (26) | PSED | 213 | 410 | −0.05 | −0.02 (−0.18, 0.14) | 0.07 | 0.11 | 119 | 247 | 0.08 | 0.07 (−0.15, 0.29) | 0.06 | 0.09 |
| Haiti | HAITI (27) | | | | | | | | | | | | | |
| Kenya | WASH−B (28) | EASQ | 809 | 2682 | 0.17 | −0.02 (−0.10, 0.06) | 0.29 | 0.18 | 552 | 2060 | −0.22 | 0.04 (−0.06, 0.13) | 0.35 | 0.27 |
| Madagascar | MAHAY (29) | ASQI | 843 | 839 | 0.15 | −0.08 (−0.28, 0.11) | 0.05 | 0.09 | 770 | 764 | −0.09 | −0.07 (−0.28, 0.15) | 0.07 | 0.09 |
| Malawi | iLiNS−DYADM (30) | PSED | 137 | 290 | −0.13 | 0.04 (−0.17, 0.24) | 0.04 | 0.09 | 78 | 148 | 0.24 | −0.05 (−0.32, 0.22) | 0.04 | 0.06 |
| Malawi | iLiNS−DOSE (31) | PSED | 376 | 135 | −0.21 | 0.03 (−0.16, 0.22) | 0.05 | 0.09 | 268 | 86 | 0.26 | 0.01 (−0.23, 0.25) | 0.05 | 0.08 |
| Mali | PROMIS CS (32) | DMC | | | | | | | | | | | | |
| Zimbabwe | SHINE (HIV−) (33) | MDAT | | | | | | | | | | | | |
| Zimbabwe | SHINE (HIV+) (34) | MDAT | | | | | | | | | | | | |
| | | | **4641** | **6736** | | **I² = 0.61, Tau² = 0.01** | | | **2989** | **5376** | | **I² = 0.16, Tau² = 0.00** | | |
| **Fixed** | | | | | | **0.04 (0.00, 0.08)** | | | | | | **0.06 (0.00, 0.11)** | | |
| **Random** | | | | | | **0.05 (−0.03, 0.12)** | | | | | | **0.06 (−0.01, 0.13)** | | |

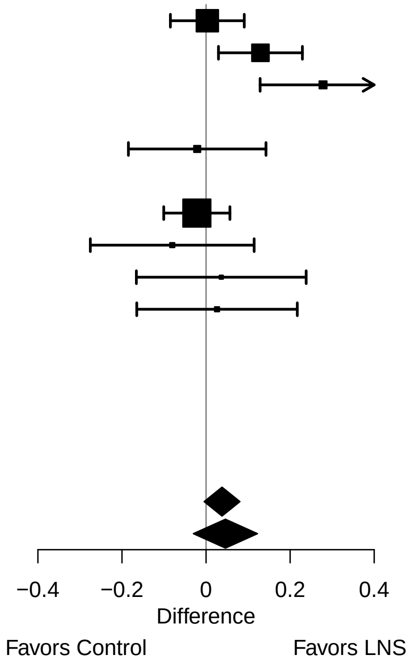

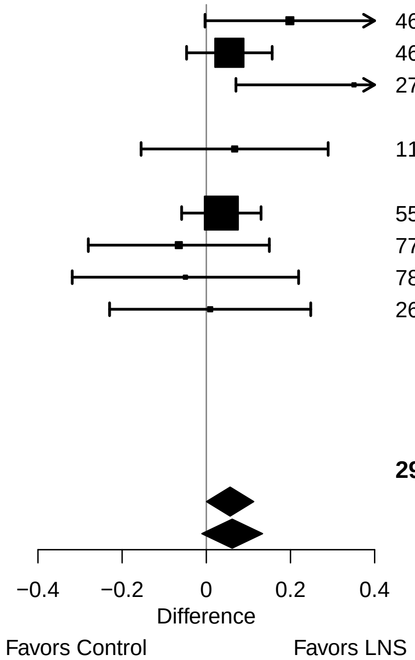

##### Supplemental figure 6D: Mean difference in social-emotional z-score

#### 6D6: Stratified by Season at the time of assessment

**P−for−interaction = 0.734**
**Difference in MDs = −0.01 (−0.08, 0.06)**

| Country | Trial | Tool | Dry LNS N | Dry Control N | Dry Control Mean | Dry MD (95% CI) | Dry Fixed W | Dry Random W | Rainy LNS N | Rainy Control N | Rainy Control Mean | Rainy MD (95% CI) | Rainy Fixed W | Rainy Random W |
|---|---|---|---|---|---|---|---|---|---|---|---|---|---|---|
| Bangladesh | JiVitA−4 (21) | | | | | | | | | | | | | |
| Bangladesh | RDNS (22) | DMC | 737 | 359 | −0.03 | 0.06 (−0.08, 0.21) | 0.10 | 0.12 | 920 | 456 | −0.04 | 0.06 (−0.05, 0.17) | 0.19 | 0.14 |
| Bangladesh | WASH−B (23) | EASQ | 604 | 1891 | −0.02 | 0.09 (0.00, 0.17) | 0.30 | 0.22 | 462 | 1375 | −0.04 | 0.17 (0.06, 0.29) | 0.16 | 0.14 |
| Burkina Faso | iLiNS−Zinc (24) | DMC | 514 | 274 | −0.25 | 0.36 (0.14, 0.57) | 0.05 | 0.06 | 232 | 101 | −0.21 | 0.35 (0.04, 0.67) | 0.02 | 0.08 |
| Ghana | GHANA (25) | | | | | | | | | | | | | |
| Ghana | iLiNS−DYADG (26) | PSED | 195 | 387 | 0.03 | 0.01 (−0.17, 0.19) | 0.07 | 0.09 | 137 | 270 | −0.05 | 0.01 (−0.19, 0.20) | 0.06 | 0.12 |
| Haiti | HAITI (27) | | | | | | | | | | | | | |
| Kenya | WASH−B (28) | EASQ | 315 | 1059 | −0.24 | 0.09 (−0.02, 0.20) | 0.18 | 0.17 | 1046 | 3682 | 0.07 | 0.00 (−0.08, 0.07) | 0.42 | 0.15 |
| Madagascar | MAHAY (29) | ASQI | | | | | | | | | | | | |
| Malawi | iLiNS−DYADM (30) | PSED | 153 | 310 | −0.10 | 0.07 (−0.13, 0.27) | 0.05 | 0.07 | 62 | 127 | 0.25 | −0.14 (−0.41, 0.14) | 0.03 | 0.09 |
| Malawi | iLiNS−DOSE (31) | PSED | 394 | 118 | −0.16 | 0.17 (−0.03, 0.37) | 0.05 | 0.07 | 243 | 100 | 0.14 | −0.14 (−0.37, 0.09) | 0.04 | 0.10 |
| Mali | PROMIS CS (32) | DMC | | | | | | | | | | | | |
| Zimbabwe | SHINE (HIV−) (33) | MDAT | 603 | 568 | −0.04 | 0.09 (−0.02, 0.20) | 0.17 | 0.16 | 238 | 231 | −0.11 | 0.21 (0.02, 0.41) | 0.06 | 0.11 |
| Zimbabwe | SHINE (HIV+) (34) | MDAT | 121 | 110 | −0.13 | 0.30 (0.05, 0.56) | 0.03 | 0.05 | 49 | 41 | −0.36 | 0.51 (0.13, 0.89) | 0.02 | 0.06 |
| | | | **3636** | **5076** | | **I² = 0.18, Tau² = 0.00** | | | **3389** | **6383** | | **I² = 0.64, Tau² = 0.02** | | |
| **Fixed** | | | | | | **0.10 (0.06, 0.15)** | | | | | | **0.06 (0.01, 0.10)** | | |
| **Random** | | | | | | **0.11 (0.05, 0.17)** | | | | | | **0.09 (−0.03, 0.21)** | | |

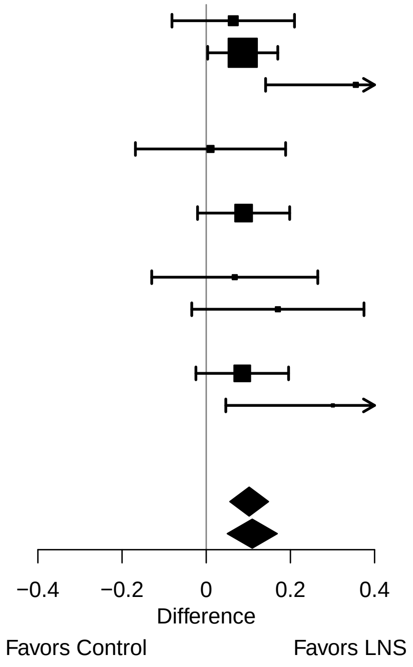

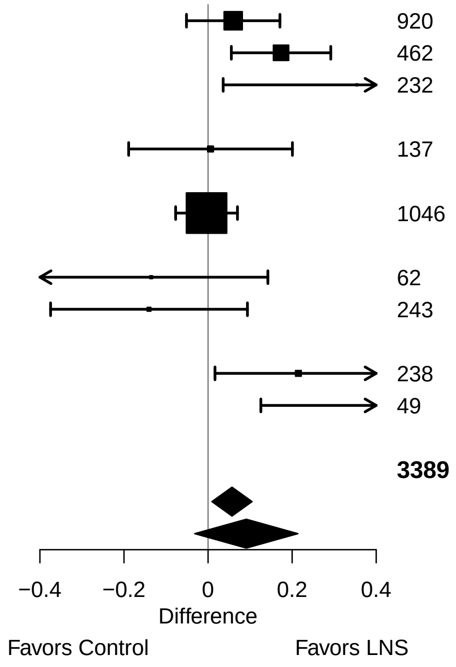

### Supplemental figure 6E: Social-emotional lowest decile prevalence ratio

#### 6E1: Stratified by Household socio-economic status

**P−for−interaction = 0.655**
**Ratio of PRs = 0.96 (0.79, 1.16)**

| | | | At least median | | | | | | Less than median | | | | | |
|---|---|---|---|---|---|---|---|---|---|---|---|---|---|---|
| **Country** | **Trial** | **Tool** | **LNS N** | **Control N** | **Control Prevalence** | **PR (95% CI)** | **Fixed W** | **Random W** | **LNS N** | **Control N** | **Control Prevalence** | **PR (95% CI)** | **Fixed W** | **Random W** |
| Bangladesh | JiVitA−4 (21) | | | | | | | | | | | | | |
| Bangladesh | RDNS (22) | DMC | 830 | 407 | 10.1 | 0.77 (0.52, 1.13) | 0.14 | 0.11 | 827 | 408 | 12.7 | 0.90 (0.67, 1.22) | 0.17 | 0.17 |
| Bangladesh | WASH−B (23) | EASQ | 545 | 1659 | 7.4 | 1.02 (0.67, 1.57) | 0.12 | 0.10 | 521 | 1607 | 13.8 | 0.70 (0.53, 0.93) | 0.19 | 0.19 |
| Burkina Faso | iLiNS−Zinc (24) | DMC | 459 | 163 | 14.7 | 0.31 (0.17, 0.56) | 0.06 | 0.09 | 287 | 212 | 17.9 | 0.58 (0.35, 0.98) | 0.06 | 0.06 |
| Ghana | GHANA (25) | | | | | | | | | | | | | |
| Ghana | iLiNS−DYADG (26) | PSED | 153 | 341 | 8.5 | 1.61 (0.95, 2.74) | 0.08 | 0.09 | 179 | 314 | 11.8 | 0.57 (0.30, 1.06) | 0.04 | 0.04 |
| Haiti | HAITI (27) | | | | | | | | | | | | | |
| Kenya | WASH−B (28) | EASQ | 778 | 2706 | 9.2 | 0.80 (0.59, 1.09) | 0.22 | 0.11 | 582 | 2036 | 11.8 | 0.96 (0.74, 1.23) | 0.24 | 0.24 |
| Madagascar | MAHAY (29) | ASQI | 741 | 845 | 5.7 | 1.38 (0.78, 2.44) | 0.07 | 0.09 | 823 | 724 | 13.1 | 0.98 (0.62, 1.54) | 0.07 | 0.07 |
| Malawi | iLiNS−DYADM (30) | PSED | 118 | 230 | 10.4 | 0.97 (0.51, 1.88) | 0.05 | 0.08 | 97 | 207 | 10.1 | 1.02 (0.50, 2.07) | 0.03 | 0.03 |
| Malawi | iLiNS−DOSE (31) | PSED | 278 | 94 | 6.4 | 1.75 (0.75, 4.06) | 0.03 | 0.06 | 270 | 96 | 11.5 | 0.81 (0.41, 1.58) | 0.03 | 0.03 |
| Mali | PROMIS CS (32) | DMC | 494 | 450 | 9.6 | 0.85 (0.52, 1.39) | 0.09 | 0.10 | 433 | 494 | 13.0 | 0.73 (0.48, 1.12) | 0.08 | 0.08 |
| Zimbabwe | SHINE (HIV−) (33) | MDAT | 406 | 359 | 13.1 | 0.58 (0.38, 0.88) | 0.12 | 0.10 | 389 | 391 | 11.8 | 0.76 (0.48, 1.23) | 0.07 | 0.07 |
| Zimbabwe | SHINE (HIV+) (34) | MDAT | 91 | 78 | 16.7 | 0.40 (0.17, 0.93) | 0.03 | 0.06 | 74 | 67 | 10.4 | 0.65 (0.21, 2.01) | 0.01 | 0.01 |
| | | | **4893** | **7332** | | **I² = 0.67, Tau² = 0.18** | | | **4482** | **6556** | | **I² = 0.00, Tau² = 0.00** | | |
| **Fixed** | | | | | | **0.83 (0.71, 0.95)** | | | | | | **0.81 (0.72, 0.92)** | | |
| **Random** | | | | | | **0.84 (0.62, 1.13)** | | | | | | **0.81 (0.72, 0.92)** | | |

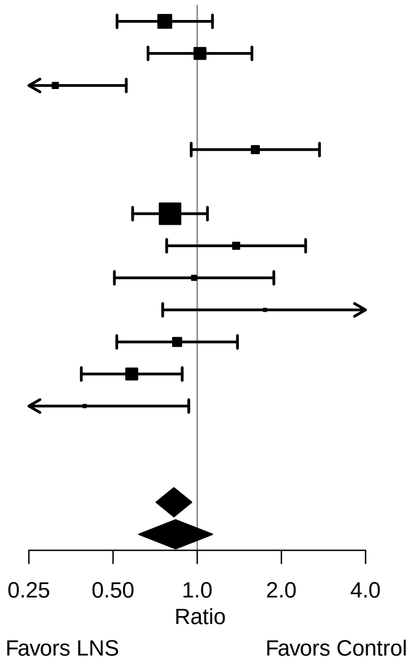

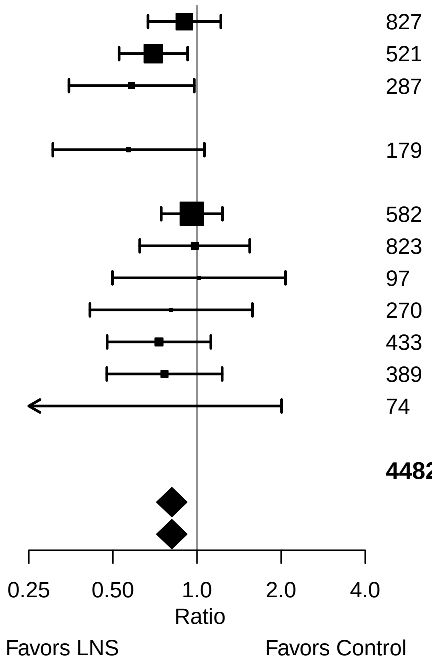

**Supplemental figure 6E: Social-emotional lowest decile prevalence ratio**

#### 6E2: Stratified by Household food insecurity

**P−for−interaction = 0.325**
**Ratio of PRs = 1.11 (0.90, 1.37)**

| Country | Trial | Tool | LNS N | Control N | Control Prevalence | Mild to secure PR (95% CI) | Fixed W | Random W | LNS N | Control N | Control Prevalence | Moderate to severe PR (95% CI) | Fixed W | Random W |
|---|---|---|---|---|---|---|---|---|---|---|---|---|---|---|
| Bangladesh | JiVitA−4 (21) | | | | | | | | | | | | | |
| Bangladesh | RDNS (22) | DMC | 1054 | 493 | 10.1 | 0.77 (0.57, 1.04) | 0.18 | 0.16 | 603 | 322 | 13.4 | 0.96 (0.69, 1.32) | 0.25 | 0.17 |
| Bangladesh | WASH−B (23) | EASQ | 853 | 2539 | 9.6 | 0.76 (0.56, 1.02) | 0.19 | 0.16 | 213 | 727 | 13.8 | 0.99 (0.66, 1.48) | 0.16 | 0.15 |
| Burkina Faso | iLiNS−Zinc (24) | DMC | 389 | 162 | 14.8 | 0.43 (0.22, 0.85) | 0.04 | 0.08 | 357 | 213 | 17.8 | 0.41 (0.28, 0.59) | 0.20 | 0.16 |
| Ghana | GHANA (25) | | | | | | | | | | | | | |
| Ghana | iLiNS−DYADG (26) | PSED | 239 | 452 | 8.6 | 1.07 (0.65, 1.76) | 0.07 | 0.11 | 92 | 202 | 12.9 | 0.93 (0.48, 1.80) | 0.06 | 0.09 |
| Haiti | HAITI (27) | | | | | | | | | | | | | |
| Kenya | WASH−B (28) | EASQ | 1214 | 4244 | 10.2 | 0.83 (0.65, 1.05) | 0.30 | 0.18 | 146 | 487 | 11.1 | 1.24 (0.73, 2.08) | 0.10 | 0.12 |
| Madagascar | MAHAY (29) | ASQI | 940 | 892 | 8.4 | 1.13 (0.65, 1.95) | 0.06 | 0.10 | 336 | 389 | 10.8 | 1.16 (0.70, 1.92) | 0.10 | 0.12 |
| Malawi | iLiNS−DYADM (30) | PSED | 68 | 118 | 8.5 | 1.91 (0.86, 4.26) | 0.03 | 0.06 | 146 | 314 | 11.1 | 0.68 (0.35, 1.29) | 0.06 | 0.10 |
| Malawi | iLiNS−DOSE (31) | PSED | | | | | | | | | | | | |
| Mali | PROMIS CS (32) | DMC | | | | | | | | | | | | |
| Zimbabwe | SHINE (HIV−) (33) | MDAT | 631 | 586 | 12.1 | 0.68 (0.48, 0.97) | 0.14 | 0.15 | 140 | 154 | 13.6 | 0.68 (0.36, 1.29) | 0.06 | 0.10 |
| Zimbabwe | SHINE (HIV+) (34) | MDAT | | | | | | | | | | | | |
| | | | **5388** | **9486** | | **I² = 0.39, Tau² = 0.06** | | | **2033** | **2808** | | **I² = 0.66, Tau² = 0.08** | | |
| **Fixed** | | | | | | **0.81 (0.71, 0.92)** | | | | | | **0.81 (0.69, 0.95)** | | |
| **Random** | | | | | | **0.83 (0.66, 1.05)** | | | | | | **0.83 (0.64, 1.08)** | | |

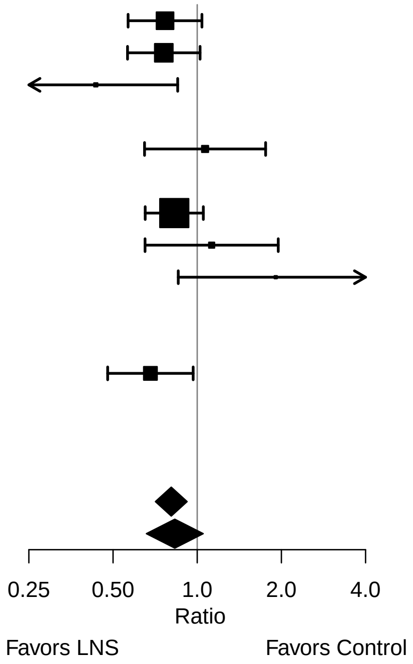

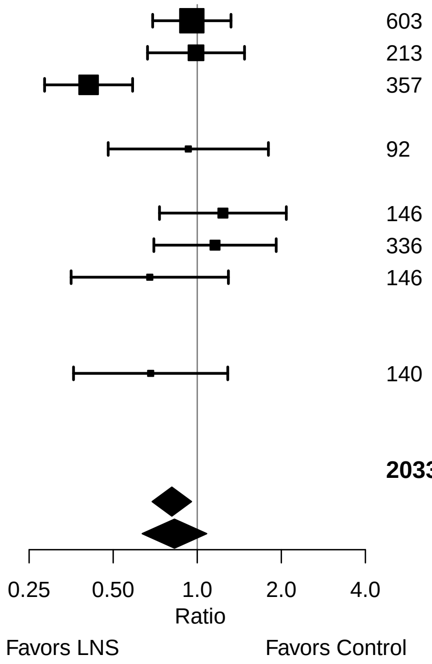

#### Supplemental figure 6E: Social-emotional lowest decile prevalence ratio

##### 6E3: Stratified by Household source water quality

**P−for−interaction = 0.534**
**Ratio of PRs = 1.10 (0.81, 1.49)**

| Country | Trial | Tool | Improved LNS N | Improved Control N | Improved Control Prevalence | Improved PR (95% CI) | Improved Fixed W | Improved Random W | Unimproved LNS N | Unimproved Control N | Unimproved Control Prevalence | Unimproved PR (95% CI) | Unimproved Fixed W | Unimproved Random W |
|---|---|---|---|---|---|---|---|---|---|---|---|---|---|---|
| Bangladesh | JiVitA−4 (21) | | | | | | | | | | | | | |
| Bangladesh | RDNS (22) | DMC | | | | | | | | | | | | |
| Bangladesh | WASH−B (23) | EASQ | 461 | 981 | 12.9 | 0.70 (0.48, 1.03) | 0.29 | 0.27 | 66 | 117 | 18.8 | 0.64 (0.31, 1.34) | 0.09 | 0.12 |
| Burkina Faso | iLiNS−Zinc (24) | DMC | 220 | 82 | 13.4 | 0.34 (0.15, 0.75) | 0.07 | 0.08 | 526 | 293 | 17.4 | 0.45 (0.29, 0.69) | 0.25 | 0.20 |
| Ghana | GHANA (25) | | | | | | | | | | | | | |
| Ghana | iLiNS−DYADG (26) | PSED | | | | | | | | | | | | |
| Haiti | HAITI (27) | | | | | | | | | | | | | |
| Kenya | WASH−B (28) | EASQ | 413 | 1419 | 10.2 | 0.73 (0.47, 1.14) | 0.22 | 0.21 | 236 | 649 | 8.2 | 1.14 (0.70, 1.87) | 0.19 | 0.18 |
| Madagascar | MAHAY (29) | ASQI | 405 | 430 | 11.9 | 1.04 (0.56, 1.93) | 0.11 | 0.12 | 1134 | 1133 | 8.1 | 1.24 (0.75, 2.06) | 0.18 | 0.18 |
| Malawi | iLiNS−DYADM (30) | PSED | | | | | | | | | | | | |
| Malawi | iLiNS−DOSE (31) | PSED | | | | | | | | | | | | |
| Mali | PROMIS CS (32) | DMC | 528 | 552 | 9.8 | 0.85 (0.55, 1.31) | 0.22 | 0.22 | 357 | 378 | 14.0 | 0.70 (0.44, 1.11) | 0.22 | 0.19 |
| Zimbabwe | SHINE (HIV−) (33) | MDAT | 228 | 220 | 9.1 | 0.87 (0.45, 1.68) | 0.09 | 0.11 | 131 | 122 | 11.5 | 0.67 (0.31, 1.43) | 0.08 | 0.12 |
| Zimbabwe | SHINE (HIV+) (34) | MDAT | | | | | | | | | | | | |
| | | | **2255** | **3684** | | **I² = 0.10, Tau² = 0.01** | | | **2450** | **2692** | | **I² = 0.59, Tau² = 0.09** | | |
| **Fixed** | | | | | | **0.75 (0.61, 0.92)** | | | | | | **0.76 (0.61, 0.94)** | | |
| **Random** | | | | | | **0.75 (0.60, 0.94)** | | | | | | **0.76 (0.55, 1.06)** | | |

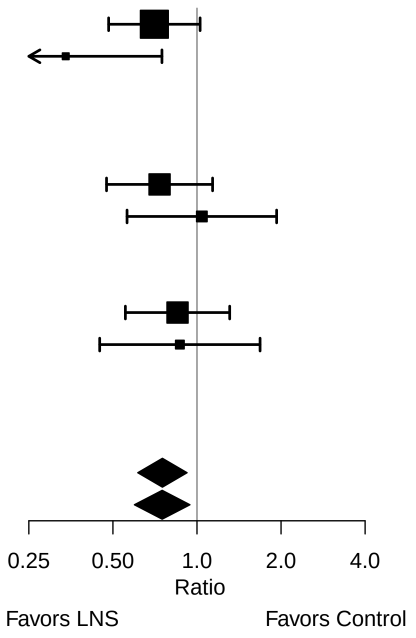

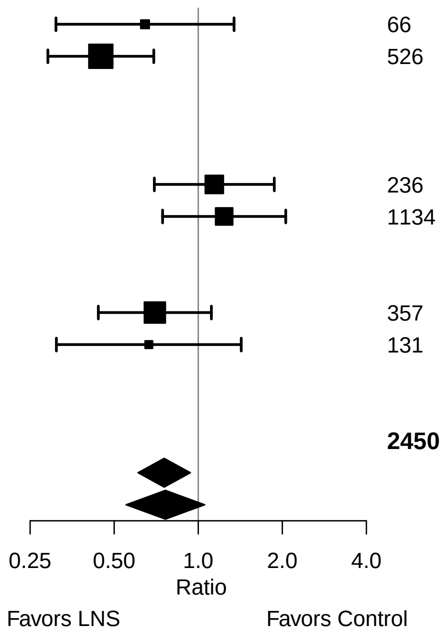

#### Supplemental figure 6E: Social-emotional lowest decile prevalence ratio

### 6E4: Stratified by Household sanitation

**P−for−interaction = 0.304**
**Ratio of PRs = 0.84 (0.60, 1.17)**

| Country | Trial | Tool | LNS N | Control N | Control Prevalence | Improved PR (95% CI) | Fixed W | Random W | LNS N | Control N | Control Prevalence | Unimproved PR (95% CI) | Fixed W | Random W |
|---|---|---|---|---|---|---|---|---|---|---|---|---|---|---|
| Bangladesh | JiVitA−4 (21) | | | | | | | | | | | | | |
| Bangladesh | RDNS (22) | DMC | 1174 | 573 | 10.1 | 0.85 (0.64, 1.13) | 0.56 | 0.56 | 481 | 241 | 14.5 | 0.82 (0.55, 1.21) | 0.35 | 0.35 |
| Bangladesh | WASH−B (23) | EASQ | | | | | | | | | | | | |
| Burkina Faso | iLiNS−Zinc (24) | DMC | | | | | | | | | | | | |
| Ghana | GHANA (25) | | | | | | | | | | | | | |
| Ghana | iLiNS−DYADG (26) | PSED | | | | | | | | | | | | |
| Haiti | HAITI (27) | | | | | | | | | | | | | |
| Kenya | WASH−B (28) | EASQ | 94 | 337 | 6.8 | 0.78 (0.31, 1.97) | 0.05 | 0.05 | 555 | 1732 | 10.1 | 0.86 (0.58, 1.26) | 0.36 | 0.36 |
| Madagascar | MAHAY (29) | ASQI | | | | | | | | | | | | |
| Malawi | iLiNS−DYADM (30) | PSED | | | | | | | | | | | | |
| Malawi | iLiNS−DOSE (31) | PSED | | | | | | | | | | | | |
| Mali | PROMIS CS (32) | DMC | 687 | 678 | 10.5 | 0.83 (0.57, 1.21) | 0.32 | 0.32 | 211 | 242 | 12.4 | 0.69 (0.37, 1.27) | 0.14 | 0.14 |
| Zimbabwe | SHINE (HIV−) (33) | MDAT | 133 | 108 | 6.5 | 1.62 (0.73, 3.61) | 0.07 | 0.07 | 225 | 235 | 11.5 | 0.54 (0.29, 1.01) | 0.14 | 0.14 |
| Zimbabwe | SHINE (HIV+) (34) | MDAT | | | | | | | | | | | | |
| | | | **2088** | **1696** | | **I² = 0.00, Tau² = 0.00** | | | **1472** | **2450** | | **I² = 0.00, Tau² = 0.00** | | |
| **Fixed** | | | | | | **0.88 (0.71, 1.09)** | | | | | | **0.76 (0.61, 0.97)** | | |
| **Random** | | | | | | **0.88 (0.71, 1.09)** | | | | | | **0.76 (0.61, 0.97)** | | |

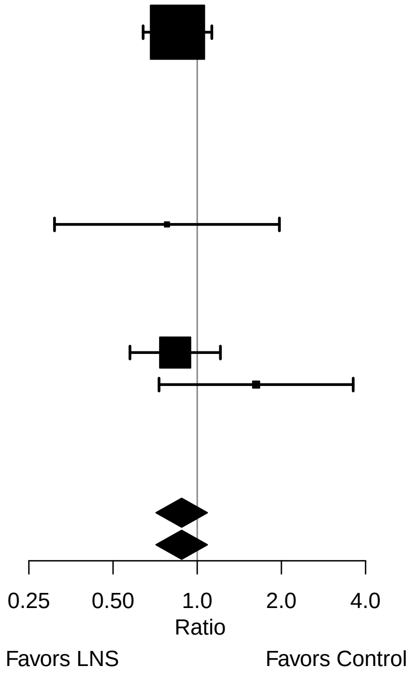

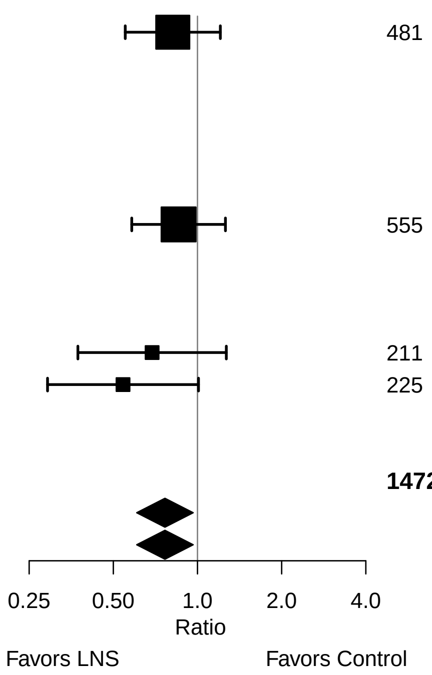

#### Supplemental figure 6E: Social-emotional lowest decile prevalence ratio

#### 6E5: Stratified by Home environment

**P−for−interaction = 0.863**
**Ratio of PRs = 1.02 (0.84, 1.23)**

| Country | Trial | Tool | At least median: LNS N | Control N | Control Prevalence | PR (95% CI) | Fixed W | Random W | Less than median: LNS N | Control N | Control Prevalence | PR (95% CI) | Fixed W | Random W |
|---|---|---|---|---|---|---|---|---|---|---|---|---|---|---|
| Bangladesh | JiVitA−4 (21) | | | | | | | | | | | | | |
| Bangladesh | RDNS (22) | DMC | 1187 | 582 | 8.4 | 0.87 (0.65, 1.16) | 0.26 | 0.23 | 467 | 230 | 18.3 | 0.84 (0.57, 1.24) | 0.13 | 0.16 |
| Bangladesh | WASH−B (23) | EASQ | 603 | 1599 | 6.9 | 0.88 (0.63, 1.25) | 0.17 | 0.17 | 463 | 1666 | 13.9 | 0.84 (0.64, 1.09) | 0.29 | 0.21 |
| Burkina Faso | iLiNS−Zinc (24) | DMC | 473 | 199 | 8.0 | 0.45 (0.25, 0.81) | 0.06 | 0.07 | 272 | 175 | 26.3 | 0.48 (0.31, 0.73) | 0.11 | 0.14 |
| Ghana | GHANA (25) | | | | | | | | | | | | | |
| Ghana | iLiNS−DYADG (26) | PSED | 213 | 410 | 10.5 | 1.07 (0.67, 1.72) | 0.09 | 0.10 | 119 | 247 | 9.3 | 0.81 (0.39, 1.70) | 0.04 | 0.07 |
| Haiti | HAITI (27) | | | | | | | | | | | | | |
| Kenya | WASH−B (28) | EASQ | 809 | 2682 | 7.3 | 0.88 (0.64, 1.21) | 0.20 | 0.19 | 552 | 2060 | 14.2 | 0.90 (0.70, 1.16) | 0.31 | 0.21 |
| Madagascar | MAHAY (29) | ASQI | 843 | 839 | 7.6 | 1.01 (0.61, 1.67) | 0.08 | 0.09 | 770 | 764 | 11.1 | 1.26 (0.77, 2.05) | 0.08 | 0.12 |
| Malawi | iLiNS−DYADM (30) | PSED | 137 | 290 | 12.4 | 0.82 (0.46, 1.47) | 0.06 | 0.07 | 78 | 148 | 6.1 | 1.69 (0.68, 4.20) | 0.02 | 0.05 |
| Malawi | iLiNS−DOSE (31) | PSED | 376 | 135 | 11.1 | 1.24 (0.73, 2.14) | 0.07 | 0.08 | 268 | 86 | 7.0 | 0.75 (0.30, 1.89) | 0.02 | 0.05 |
| Mali | PROMIS CS (32) | DMC | | | | | | | | | | | | |
| Zimbabwe | SHINE (HIV−) (33) | MDAT | | | | | | | | | | | | |
| Zimbabwe | SHINE (HIV+) (34) | MDAT | | | | | | | | | | | | |
| | | | **4641** | **6736** | | **I² = 0.08, Tau² = 0.01** | | | **2989** | **5376** | | **I² = 0.41, Tau² = 0.04** | | |
| **Fixed** | | | | | | **0.89 (0.77, 1.02)** | | | | | | **0.85 (0.74, 0.98)** | | |
| **Random** | | | | | | **0.89 (0.76, 1.04)** | | | | | | **0.85 (0.68, 1.06)** | | |

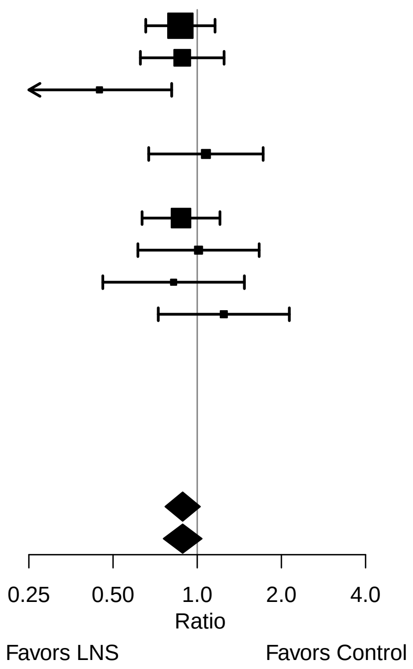

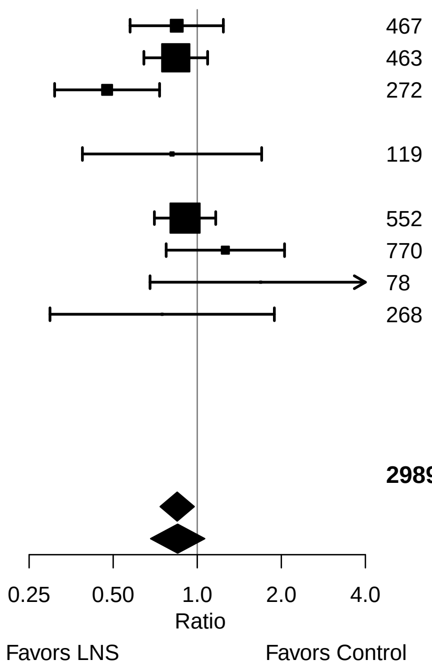

#### Supplemental figure 6E: Social-emotional lowest decile prevalence ratio

#### 6E6: Stratified by Season at the time of assessment

**P−for−interaction = 0.584**
**Ratio of PRs = 1.06 (0.85, 1.33)**

| Country | Trial | Tool | Dry LNS N | Dry Control N | Dry Control Prevalence | Dry PR (95% CI) | Dry Fixed W | Dry Random W | Rainy LNS N | Rainy Control N | Rainy Control Prevalence | Rainy PR (95% CI) | Rainy Fixed W | Rainy Random W |
|---|---|---|---|---|---|---|---|---|---|---|---|---|---|---|
| Bangladesh | JiVitA−4 (21) | | | | | | | | | | | | | |
| Bangladesh | RDNS (22) | DMC | 737 | 359 | 12.8 | 0.77 (0.55, 1.08) | 0.21 | 0.21 | 920 | 456 | 10.3 | 0.91 (0.67, 1.23) | 0.25 | 0.22 |
| Bangladesh | WASH−B (23) | EASQ | 604 | 1891 | 11.3 | 0.83 (0.61, 1.13) | 0.25 | 0.24 | 462 | 1375 | 9.4 | 0.78 (0.54, 1.14) | 0.17 | 0.19 |
| Burkina Faso | iLiNS−Zinc (24) | DMC | 514 | 274 | 15.7 | 0.43 (0.26, 0.72) | 0.09 | 0.10 | 232 | 101 | 18.8 | 0.37 (0.16, 0.83) | 0.04 | 0.08 |
| Ghana | GHANA (25) | | | | | | | | | | | | | |
| Ghana | iLiNS−DYADG (26) | PSED | 195 | 387 | 10.6 | 0.92 (0.55, 1.54) | 0.09 | 0.10 | 137 | 270 | 9.3 | 1.10 (0.59, 2.05) | 0.06 | 0.11 |
| Haiti | HAITI (27) | | | | | | | | | | | | | |
| Kenya | WASH−B (28) | EASQ | 315 | 1059 | 15.2 | 0.71 (0.49, 1.04) | 0.17 | 0.17 | 1046 | 3682 | 8.9 | 0.94 (0.74, 1.21) | 0.39 | 0.25 |
| Madagascar | MAHAY (29) | ASQI | | | | | | | | | | | | |
| Malawi | iLiNS−DYADM (30) | PSED | | | | | | | | | | | | |
| Malawi | iLiNS−DOSE (31) | PSED | | | | | | | | | | | | |
| Mali | PROMIS CS (32) | DMC | | | | | | | | | | | | |
| Zimbabwe | SHINE (HIV−) (33) | MDAT | 603 | 568 | 11.1 | 0.79 (0.56, 1.13) | 0.19 | 0.19 | 238 | 231 | 13.9 | 0.55 (0.33, 0.92) | 0.09 | 0.14 |
| Zimbabwe | SHINE (HIV+) (34) | MDAT | | | | | | | | | | | | |
| | | | **2968** | **4538** | | **I² = 0.11, Tau² = 0.01** | | | **3035** | **6115** | | **I² = 0.42, Tau² = 0.06** | | |
| **Fixed** | | | | | | **0.75 (0.64, 0.88)** | | | | | | **0.84 (0.72, 0.98)** | | |
| **Random** | | | | | | **0.75 (0.63, 0.89)** | | | | | | **0.79 (0.61, 1.03)** | | |

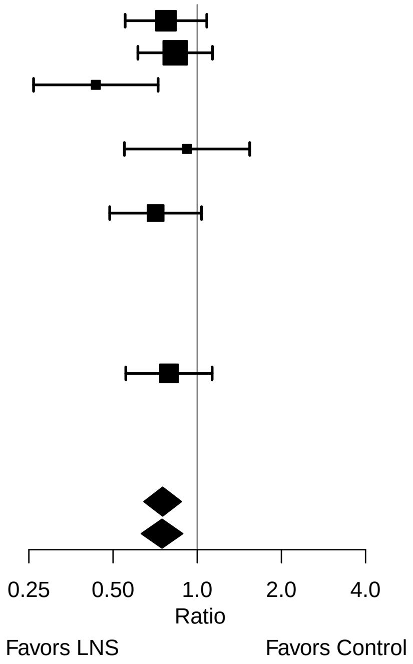

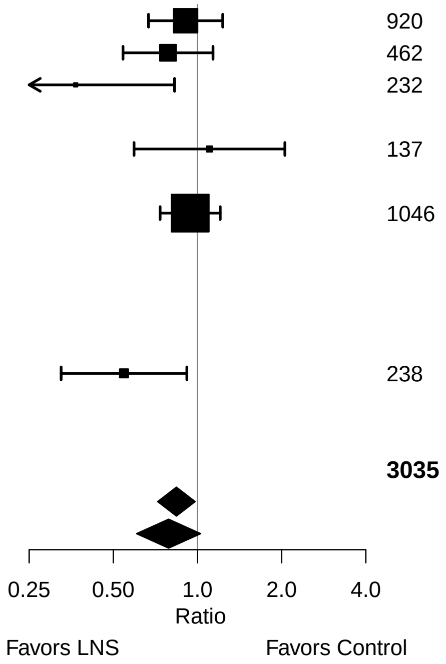

### Supplemental figure 6F: Social-emotional lowest decile prevalence difference

#### 6F1: Stratified by Household socio-economic status

**P−for−interaction = 0.160**
**Difference in PDs = −0.01 (−0.03, 0.00)**

| Country | Trial | Tool | At least median: LNS N | Control N | Control Prevalence | PD (95% CI) | Fixed W | Random W | Less than median: LNS N | Control N | Control Prevalence | PD (95% CI) | Fixed W | Random W |
|---|---|---|---|---|---|---|---|---|---|---|---|---|---|---|
| Bangladesh | JiVitA−4 (21) | | | | | | | | | | | | | |
| Bangladesh | RDNS (22) | DMC | 830 | 407 | 10.1 | −0.02 (−0.06, 0.01) | 0.11 | 0.11 | 827 | 408 | 12.7 | −0.01 (−0.05, 0.02) | 0.14 | 0.14 |
| Bangladesh | WASH−B (23) | EASQ | 545 | 1659 | 7.4 | 0.00 (−0.03, 0.03) | 0.15 | 0.11 | 521 | 1607 | 13.8 | −0.04 (−0.07, −0.01) | 0.21 | 0.21 |
| Burkina Faso | iLiNS−Zinc (24) | DMC | 459 | 163 | 14.7 | −0.10 (−0.16, −0.04) | 0.04 | 0.08 | 287 | 212 | 17.9 | −0.07 (−0.15, 0.00) | 0.04 | 0.04 |
| Ghana | GHANA (25) | | | | | | | | | | | | | |
| Ghana | iLiNS−DYADG (26) | PSED | 153 | 341 | 8.5 | 0.05 (−0.01, 0.11) | 0.05 | 0.08 | 179 | 314 | 11.8 | −0.05 (−0.11, 0.00) | 0.06 | 0.06 |
| Haiti | HAITI (27) | | | | | | | | | | | | | |
| Kenya | WASH−B (28) | EASQ | 778 | 2706 | 9.2 | −0.02 (−0.04, 0.01) | 0.28 | 0.12 | 582 | 2036 | 11.8 | 0.00 (−0.03, 0.02) | 0.23 | 0.23 |
| Madagascar | MAHAY (29) | ASQI | 741 | 845 | 5.7 | 0.02 (−0.02, 0.06) | 0.10 | 0.10 | 823 | 724 | 13.1 | 0.00 (−0.06, 0.06) | 0.05 | 0.05 |
| Malawi | iLiNS−DYADM (30) | PSED | 118 | 230 | 10.4 | 0.00 (−0.07, 0.07) | 0.03 | 0.07 | 97 | 207 | 10.1 | 0.00 (−0.07, 0.07) | 0.04 | 0.04 |
| Malawi | iLiNS−DOSE (31) | PSED | 278 | 94 | 6.4 | 0.05 (−0.02, 0.12) | 0.03 | 0.07 | 270 | 96 | 11.5 | −0.02 (−0.09, 0.05) | 0.04 | 0.04 |
| Mali | PROMIS CS (32) | DMC | 494 | 450 | 9.6 | −0.01 (−0.06, 0.03) | 0.08 | 0.10 | 433 | 494 | 13.0 | −0.03 (−0.08, 0.01) | 0.10 | 0.10 |
| Zimbabwe | SHINE (HIV−) (33) | MDAT | 406 | 359 | 13.1 | −0.05 (−0.10, −0.01) | 0.09 | 0.10 | 389 | 391 | 11.8 | −0.03 (−0.08, 0.02) | 0.08 | 0.08 |
| Zimbabwe | SHINE (HIV+) (34) | MDAT | 91 | 78 | 16.7 | −0.10 (−0.19, −0.01) | 0.02 | 0.06 | 74 | 67 | 10.4 | −0.04 (−0.13, 0.06) | 0.02 | 0.02 |
| | | | **4893** | **7332** | | **I² = 0.64, Tau² = 0.00** | | | **4482** | **6556** | | **I² = 0.00, Tau² = 0.00** | | |
| **Fixed** | | | | | | **−0.01 (−0.03, 0.00)** | | | | | | **−0.02 (−0.04, −0.01)** | | |
| **Random** | | | | | | **−0.02 (−0.04, 0.01)** | | | | | | **−0.02 (−0.04, −0.01)** | | |

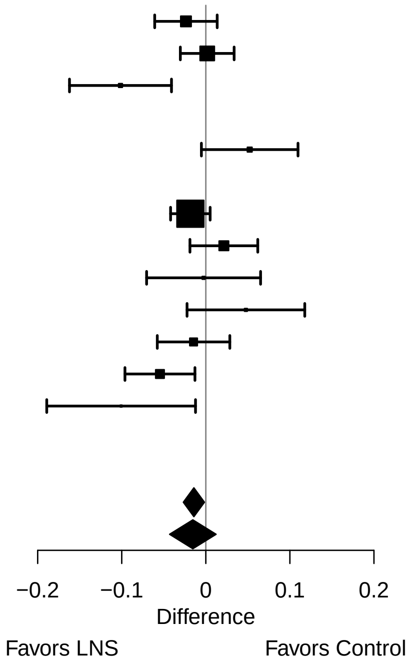

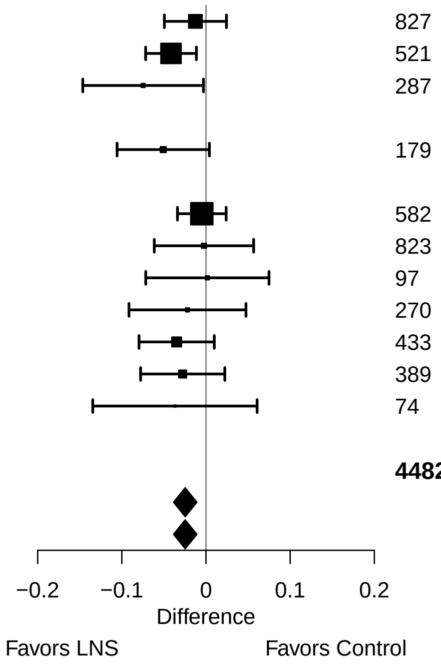

#### Supplemental figure 6F: Social-emotional lowest decile prevalence difference

#### 6F2: Stratified by Household food insecurity

**P−for−interaction = 0.599**
**Difference in PDs = 0.01 (−0.02, 0.03)**

| Country | Trial | Tool | LNS N | Control N | Control Prevalence | Mild to secure PD (95% CI) | Fixed W | Random W | LNS N | Control N | Control Prevalence | Moderate to severe PD (95% CI) | Fixed W | Random W |
|---|---|---|---|---|---|---|---|---|---|---|---|---|---|---|
| Bangladesh | JiVitA−4 (21) | | | | | | | | | | | | | |
| Bangladesh | RDNS (22) | DMC | 1054 | 493 | 10.1 | −0.02 (−0.05, 0.00) | 0.17 | 0.16 | 603 | 322 | 13.4 | −0.01 (−0.05, 0.04) | 0.22 | 0.16 |
| Bangladesh | WASH−B (23) | EASQ | 853 | 2539 | 9.6 | −0.02 (−0.05, 0.00) | 0.25 | 0.18 | 213 | 727 | 13.8 | 0.00 (−0.06, 0.05) | 0.13 | 0.13 |
| Burkina Faso | iLiNS−Zinc (24) | DMC | 389 | 162 | 14.8 | −0.08 (−0.16, −0.01) | 0.02 | 0.06 | 357 | 213 | 17.8 | −0.11 (−0.15, −0.06) | 0.20 | 0.16 |
| Ghana | GHANA (25) | | | | | | | | | | | | | |
| Ghana | iLiNS−DYADG (26) | PSED | 239 | 452 | 8.6 | 0.01 (−0.04, 0.05) | 0.07 | 0.12 | 92 | 202 | 12.9 | −0.01 (−0.09, 0.07) | 0.06 | 0.09 |
| Haiti | HAITI (27) | | | | | | | | | | | | | |
| Kenya | WASH−B (28) | EASQ | 1214 | 4244 | 10.2 | −0.02 (−0.04, 0.00) | 0.31 | 0.19 | 146 | 487 | 11.1 | 0.03 (−0.04, 0.09) | 0.09 | 0.11 |
| Madagascar | MAHAY (29) | ASQI | 940 | 892 | 8.4 | 0.01 (−0.04, 0.06) | 0.06 | 0.11 | 336 | 389 | 10.8 | 0.02 (−0.04, 0.08) | 0.11 | 0.12 |
| Malawi | iLiNS−DYADM (30) | PSED | 68 | 118 | 8.5 | 0.08 (−0.02, 0.17) | 0.02 | 0.04 | 146 | 314 | 11.1 | −0.04 (−0.10, 0.02) | 0.11 | 0.13 |
| Malawi | iLiNS−DOSE (31) | PSED | | | | | | | | | | | | |
| Mali | PROMIS CS (32) | DMC | | | | | | | | | | | | |
| Zimbabwe | SHINE (HIV−) (33) | MDAT | 631 | 586 | 12.1 | −0.04 (−0.08, 0.00) | 0.10 | 0.14 | 140 | 154 | 13.6 | −0.04 (−0.11, 0.03) | 0.08 | 0.10 |
| Zimbabwe | SHINE (HIV+) (34) | MDAT | | | | | | | | | | | | |
| | | | **5388** | **9486** | | **I² = 0.35, Tau² = 0.00** | | | **2033** | **2808** | | **I² = 0.62, Tau² = 0.00** | | |
| **Fixed** | | | | | | **−0.02 (−0.03, −0.01)** | | | | | | **−0.03 (−0.05, −0.01)** | | |
| **Random** | | | | | | **−0.02 (−0.04, 0.01)** | | | | | | **−0.02 (−0.05, 0.01)** | | |

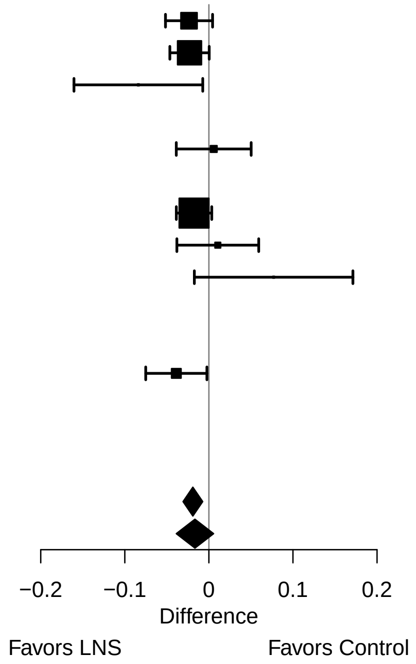

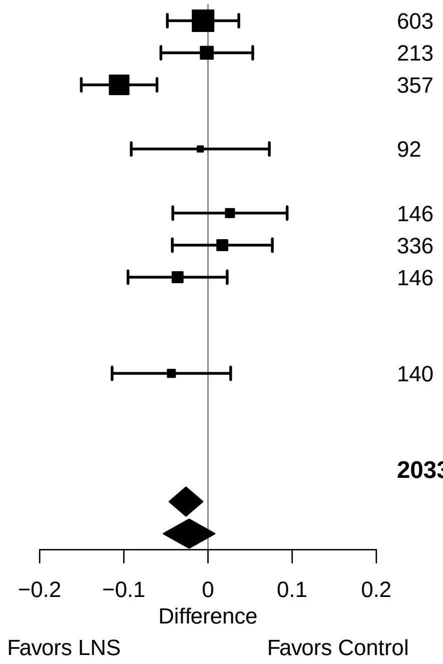

#### Supplemental figure 6F: Social-emotional lowest decile prevalence difference

#### 6F3: Stratified by Household source water quality

**P−for−interaction = 0.782**
**Difference in PDs = 0.00 (−0.02, 0.03)**

| Country | Trial | Tool | LNS N | Control N | Control Prevalence | Improved PD (95% CI) | Fixed W | Random W | LNS N | Control N | Control Prevalence | Unimproved PD (95% CI) | Fixed W | Random W |
|---|---|---|---|---|---|---|---|---|---|---|---|---|---|---|
| Bangladesh | JiVitA−4 (21) | | | | | | | | | | | | | |
| Bangladesh | RDNS (22) | DMC | | | | | | | | | | | | |
| Bangladesh | WASH−B (23) | EASQ | 461 | 981 | 12.9 | −0.04 (−0.08, 0.00) | 0.24 | 0.24 | 66 | 117 | 18.8 | −0.07 (−0.17, 0.04) | 0.05 | 0.09 |
| Burkina Faso | iLiNS−Zinc (24) | DMC | 220 | 82 | 13.4 | −0.09 (−0.17, −0.01) | 0.06 | 0.06 | 526 | 293 | 17.4 | −0.10 (−0.15, −0.05) | 0.20 | 0.19 |
| Ghana | GHANA (25) | | | | | | | | | | | | | |
| Ghana | iLiNS−DYADG (26) | PSED | | | | | | | | | | | | |
| Haiti | HAITI (27) | | | | | | | | | | | | | |
| Kenya | WASH−B (28) | EASQ | 413 | 1419 | 10.2 | −0.03 (−0.06, 0.01) | 0.29 | 0.29 | 236 | 649 | 8.2 | 0.01 (−0.03, 0.06) | 0.25 | 0.20 |
| Madagascar | MAHAY (29) | ASQI | 405 | 430 | 11.9 | 0.00 (−0.07, 0.08) | 0.06 | 0.06 | 1134 | 1133 | 8.1 | 0.02 (−0.03, 0.07) | 0.23 | 0.20 |
| Malawi | iLiNS−DYADM (30) | PSED | | | | | | | | | | | | |
| Malawi | iLiNS−DOSE (31) | PSED | | | | | | | | | | | | |
| Mali | PROMIS CS (32) | DMC | 528 | 552 | 9.8 | −0.01 (−0.05, 0.02) | 0.24 | 0.24 | 357 | 378 | 14.0 | −0.04 (−0.10, 0.01) | 0.17 | 0.18 |
| Zimbabwe | SHINE (HIV−) (33) | MDAT | 228 | 220 | 9.1 | −0.01 (−0.07, 0.04) | 0.11 | 0.11 | 131 | 122 | 11.5 | −0.04 (−0.11, 0.03) | 0.10 | 0.14 |
| Zimbabwe | SHINE (HIV+) (34) | MDAT | | | | | | | | | | | | |
| | | | **2255** | **3684** | | **I² = 0.00, Tau² = 0.00** | | | **2450** | **2692** | | **I² = 0.67, Tau² = 0.00** | | |
| **Fixed** | | | | | | **−0.03 (−0.05, −0.01)** | | | | | | **−0.03 (−0.05, 0.00)** | | |
| **Random** | | | | | | **−0.03 (−0.05, −0.01)** | | | | | | **−0.03 (−0.07, 0.01)** | | |

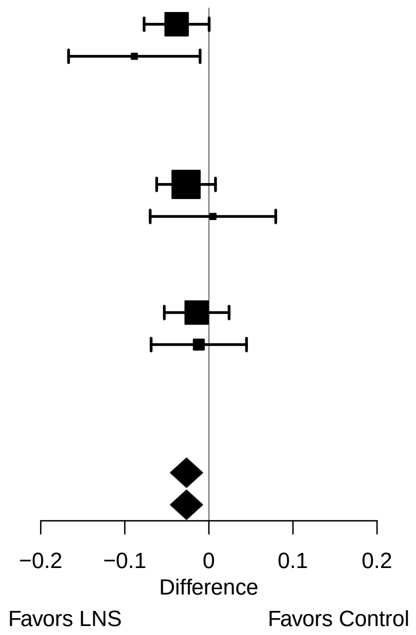

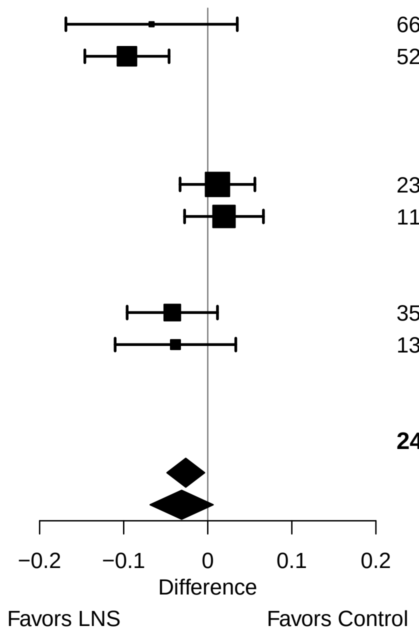

#### Supplemental figure 6F: Social-emotional lowest decile prevalence difference

#### 6F4: Stratified by Household sanitation

**P−for−interaction = 0.189**
**Difference in PDs = −0.02 (−0.05, 0.01)**

| Country | Trial | Tool | LNS N | Control N | Control Prevalence | Improved PD (95% CI) | Fixed W | Random W | LNS N | Control N | Control Prevalence | Unimproved PD (95% CI) | Fixed W | Random W |
|---|---|---|---|---|---|---|---|---|---|---|---|---|---|---|
| Bangladesh | JiVitA−4 (21) | | | | | | | | | | | | | |
| Bangladesh | RDNS (22) | DMC | 1174 | 573 | 10.1 | −0.02 (−0.04, 0.01) | 0.48 | 0.48 | 481 | 241 | 14.5 | −0.03 (−0.08, 0.03) | 0.20 | 0.20 |
| Bangladesh | WASH−B (23) | EASQ | | | | | | | | | | | | |
| Burkina Faso | iLiNS−Zinc (24) | DMC | | | | | | | | | | | | |
| Ghana | GHANA (25) | | | | | | | | | | | | | |
| Ghana | iLiNS−DYADG (26) | PSED | | | | | | | | | | | | |
| Haiti | HAITI (27) | | | | | | | | | | | | | |
| Kenya | WASH−B (28) | EASQ | 94 | 337 | 6.8 | −0.02 (−0.07, 0.04) | 0.13 | 0.13 | 555 | 1732 | 10.1 | −0.01 (−0.05, 0.02) | 0.46 | 0.46 |
| Madagascar | MAHAY (29) | ASQI | | | | | | | | | | | | |
| Malawi | iLiNS−DYADM (30) | PSED | | | | | | | | | | | | |
| Malawi | iLiNS−DOSE (31) | PSED | | | | | | | | | | | | |
| Mali | PROMIS CS (32) | DMC | 687 | 678 | 10.5 | −0.02 (−0.05, 0.02) | 0.30 | 0.30 | 211 | 242 | 12.4 | −0.04 (−0.10, 0.02) | 0.14 | 0.14 |
| Zimbabwe | SHINE (HIV−) (33) | MDAT | 133 | 108 | 6.5 | 0.04 (−0.02, 0.10) | 0.09 | 0.09 | 225 | 235 | 11.5 | −0.05 (−0.11, 0.00) | 0.19 | 0.19 |
| Zimbabwe | SHINE (HIV+) (34) | MDAT | | | | | | | | | | | | |
| | | | **2088** | **1696** | | **I² = 0.00, Tau² = 0.00** | | | **1472** | **2450** | | **I² = 0.00, Tau² = 0.00** | | |
| **Fixed** | | | | | | **−0.01 (−0.03, 0.01)** | | | | | | **−0.03 (−0.05, 0.00)** | | |
| **Random** | | | | | | **−0.01 (−0.03, 0.01)** | | | | | | **−0.03 (−0.05, 0.00)** | | |

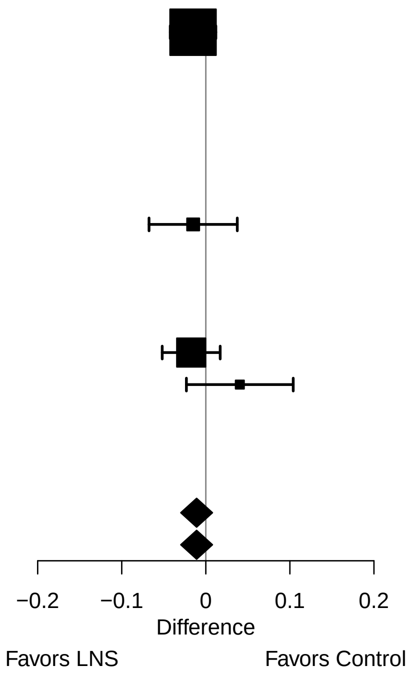

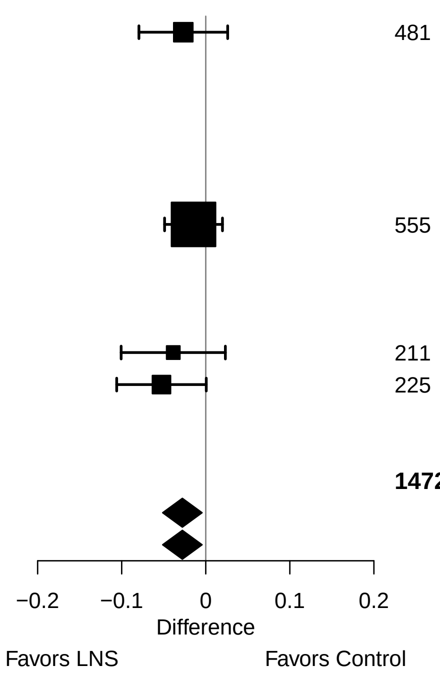

#### Supplemental figure 6F: Social-emotional lowest decile prevalence difference

### 6F5: Stratified by Home environment

**P−for−interaction = 0.545**
**Difference in PDs = −0.01 (−0.03, 0.01)**

| Country | Trial | Tool | LNS N | Control N | Control Prevalence | At least median PD (95% CI) | Fixed W | Random W | LNS N | Control N | Control Prevalence | Less than median PD (95% CI) | Fixed W | Random W |
|---|---|---|---|---|---|---|---|---|---|---|---|---|---|---|
| Bangladesh | JiVitA−4 (21) | | | | | | | | | | | | | |
| Bangladesh | RDNS (22) | DMC | 1187 | 582 | 8.4 | −0.01 (−0.03, 0.01) | 0.23 | 0.23 | 467 | 230 | 18.3 | −0.03 (−0.10, 0.04) | 0.07 | 0.11 |
| Bangladesh | WASH−B (23) | EASQ | 603 | 1599 | 6.9 | −0.01 (−0.03, 0.01) | 0.24 | 0.24 | 463 | 1666 | 13.9 | −0.02 (−0.05, 0.01) | 0.29 | 0.17 |
| Burkina Faso | iLiNS−Zinc (24) | DMC | 473 | 199 | 8.0 | −0.04 (−0.08, −0.01) | 0.08 | 0.08 | 272 | 175 | 26.3 | −0.14 (−0.22, −0.06) | 0.05 | 0.10 |
| Ghana | GHANA (25) | | | | | | | | | | | | | |
| Ghana | iLiNS−DYADG (26) | PSED | 213 | 410 | 10.5 | 0.01 (−0.04, 0.06) | 0.04 | 0.04 | 119 | 247 | 9.3 | −0.02 (−0.08, 0.04) | 0.08 | 0.12 |
| Haiti | HAITI (27) | | | | | | | | | | | | | |
| Kenya | WASH−B (28) | EASQ | 809 | 2682 | 7.3 | −0.01 (−0.03, 0.01) | 0.26 | 0.26 | 552 | 2060 | 14.2 | −0.01 (−0.05, 0.02) | 0.28 | 0.16 |
| Madagascar | MAHAY (29) | ASQI | 843 | 839 | 7.6 | 0.00 (−0.04, 0.04) | 0.08 | 0.08 | 770 | 764 | 11.1 | 0.03 (−0.03, 0.09) | 0.08 | 0.12 |
| Malawi | iLiNS−DYADM (30) | PSED | 137 | 290 | 12.4 | −0.02 (−0.09, 0.04) | 0.03 | 0.03 | 78 | 148 | 6.1 | 0.04 (−0.03, 0.11) | 0.06 | 0.10 |
| Malawi | iLiNS−DOSE (31) | PSED | 376 | 135 | 11.1 | 0.03 (−0.04, 0.09) | 0.03 | 0.03 | 268 | 86 | 7.0 | −0.02 (−0.07, 0.04) | 0.10 | 0.13 |
| Mali | PROMIS CS (32) | DMC | | | | | | | | | | | | |
| Zimbabwe | SHINE (HIV−) (33) | MDAT | | | | | | | | | | | | |
| Zimbabwe | SHINE (HIV+) (34) | MDAT | | | | | | | | | | | | |
| | | | **4641** | **6736** | | **I² = 0.00, Tau² = 0.00** | | | **2989** | **5376** | | **I² = 0.51, Tau² = 0.00** | | |
| **Fixed** | | | | | | **−0.01 (−0.02, 0.00)** | | | | | | **−0.02 (−0.03, 0.00)** | | |
| **Random** | | | | | | **−0.01 (−0.02, 0.00)** | | | | | | **−0.02 (−0.05, 0.01)** | | |

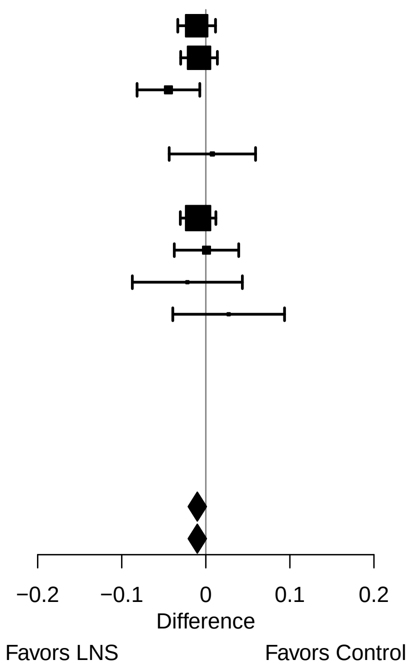

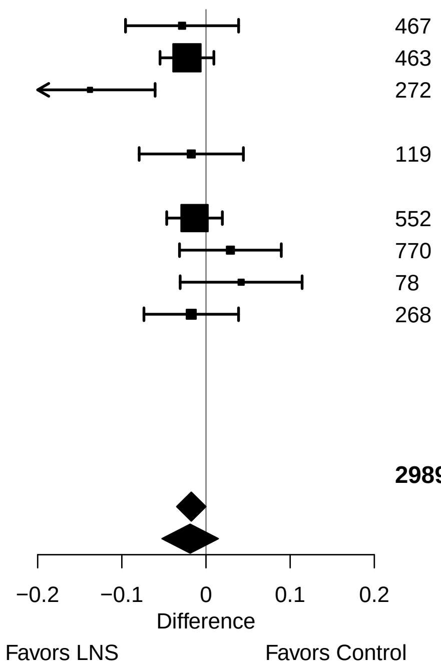

#### Supplemental figure 6F: Social-emotional lowest decile prevalence difference

### 6F6: Stratified by Season at the time of assessment

**P−for−interaction = 0.493**
**Difference in PDs = 0.01 (−0.01, 0.03)**

| Country | Trial | Tool | LNS N | Control N | Control Prevalence | Dry PD (95% CI) | Fixed W | Random W | LNS N | Control N | Control Prevalence | Rainy PD (95% CI) | Fixed W | Random W |
|---|---|---|---|---|---|---|---|---|---|---|---|---|---|---|
| Bangladesh | JiVitA−4 (21) | | | | | | | | | | | | | |
| Bangladesh | RDNS (22) | DMC | 737 | 359 | 12.8 | −0.03 (−0.07, 0.01) | 0.18 | 0.18 | 920 | 456 | 10.3 | −0.01 (−0.04, 0.02) | 0.21 | 0.22 |
| Bangladesh | WASH−B (23) | EASQ | 604 | 1891 | 11.3 | −0.02 (−0.05, 0.01) | 0.29 | 0.27 | 462 | 1375 | 9.4 | −0.02 (−0.05, 0.01) | 0.23 | 0.23 |
| Burkina Faso | iLiNS−Zinc (24) | DMC | 514 | 274 | 15.7 | −0.09 (−0.15, −0.03) | 0.07 | 0.08 | 232 | 101 | 18.8 | −0.12 (−0.24, 0.00) | 0.01 | 0.04 |
| Ghana | GHANA (25) | | | | | | | | | | | | | |
| Ghana | iLiNS−DYADG (26) | PSED | 195 | 387 | 10.6 | −0.01 (−0.06, 0.04) | 0.10 | 0.11 | 137 | 270 | 9.3 | 0.01 (−0.05, 0.07) | 0.05 | 0.11 |
| Haiti | HAITI (27) | | | | | | | | | | | | | |
| Kenya | WASH−B (28) | EASQ | 315 | 1059 | 15.2 | −0.04 (−0.09, 0.00) | 0.14 | 0.15 | 1046 | 3682 | 8.9 | −0.01 (−0.03, 0.02) | 0.43 | 0.27 |
| Madagascar | MAHAY (29) | ASQI | | | | | | | | | | | | |
| Malawi | iLiNS−DYADM (30) | PSED | | | | | | | | | | | | |
| Malawi | iLiNS−DOSE (31) | PSED | | | | | | | | | | | | |
| Mali | PROMIS CS (32) | DMC | | | | | | | | | | | | |
| Zimbabwe | SHINE (HIV−) (33) | MDAT | 603 | 568 | 11.1 | −0.02 (−0.06, 0.01) | 0.22 | 0.21 | 238 | 231 | 13.9 | −0.06 (−0.12, −0.01) | 0.06 | 0.12 |
| Zimbabwe | SHINE (HIV+) (34) | MDAT | | | | | | | | | | | | |
| | | | **2968** | **4538** | | **I² = 0.06, Tau² = 0.00** | | | **3035** | **6115** | | **I² = 0.33, Tau² = 0.00** | | |
| **Fixed** | | | | | | **−0.03 (−0.05, −0.01)** | | | | | | **−0.01 (−0.03, 0.00)** | | |
| **Random** | | | | | | **−0.03 (−0.05, −0.01)** | | | | | | **−0.02 (−0.04, 0.00)** | | |

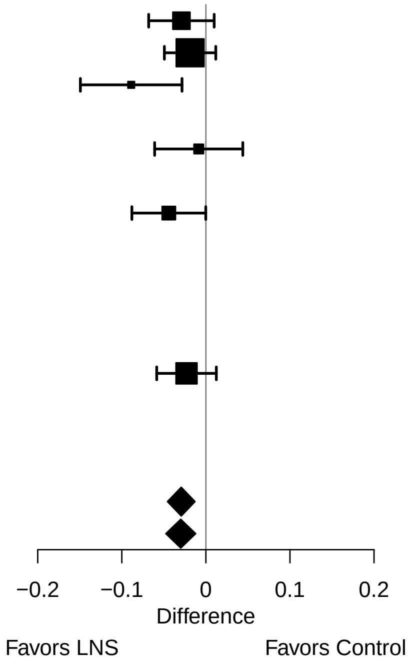

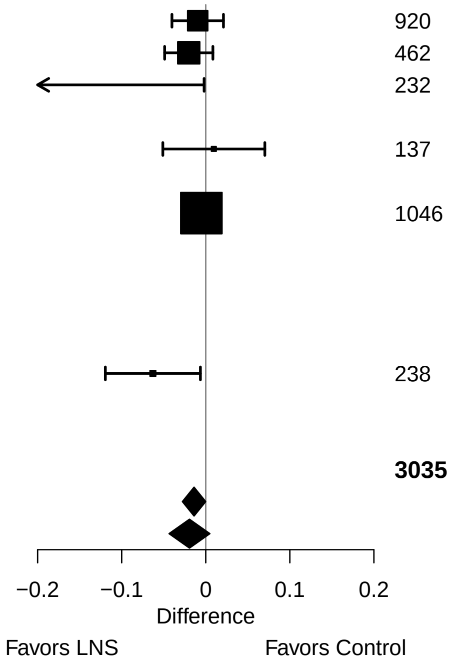

### Supplemental figure 6G: Mean difference in motor z-score

#### 6G1: Stratified by Household socio-economic status

**P−for−interaction = 0.077**
**Difference in MDs = 0.05 (−0.01, 0.11)**

| Country | Trial | Tool | At least median: LNS N | Control N | Control Mean | MD (95% CI) | Fixed W | Random W | Less than median: LNS N | Control N | Control Mean | MD (95% CI) | Fixed W | Random W |
|---|---|---|---|---|---|---|---|---|---|---|---|---|---|---|
| Bangladesh | JiVitA−4 (21) | BSID−III | 236 | 77 | 0.21 | −0.09 (−0.29, 0.11) | 0.04 | 0.07 | 208 | 66 | −0.21 | 0.07 (−0.19, 0.32) | 0.03 | 0.03 |
| Bangladesh | RDNS (22) | DMC | 783 | 381 | 0.04 | 0.09 (−0.01, 0.19) | 0.16 | 0.13 | 773 | 372 | −0.13 | 0.11 (0.01, 0.21) | 0.20 | 0.20 |
| Bangladesh | WASH−B (23) | EASQ | 546 | 1663 | 0.12 | 0.05 (−0.04, 0.14) | 0.20 | 0.14 | 528 | 1616 | −0.17 | 0.16 (0.07, 0.26) | 0.23 | 0.23 |
| Burkina Faso | iLiNS−Zinc (24) | DMC | 459 | 163 | −0.16 | 0.36 (0.15, 0.58) | 0.03 | 0.06 | 287 | 212 | −0.34 | 0.38 (0.15, 0.60) | 0.04 | 0.04 |
| Ghana | GHANA (25) | | | | | | | | | | | | | |
| Ghana | iLiNS−DYADG (26) | KDI | 143 | 311 | 0.01 | 0.09 (−0.11, 0.29) | 0.04 | 0.07 | 159 | 288 | −0.02 | −0.03 (−0.22, 0.16) | 0.06 | 0.06 |
| Haiti | HAITI (27) | | | | | | | | | | | | | |
| Kenya | WASH−B (28) | EASQ | 778 | 2706 | 0.07 | −0.03 (−0.10, 0.04) | 0.30 | 0.15 | 582 | 2036 | −0.10 | 0.07 (−0.04, 0.17) | 0.19 | 0.19 |
| Madagascar | MAHAY (29) | ASQI | 741 | 845 | 0.14 | 0.07 (−0.10, 0.24) | 0.05 | 0.08 | 823 | 724 | −0.25 | 0.13 (−0.12, 0.38) | 0.03 | 0.03 |
| Malawi | iLiNS−DYADM (30) | KDI | 118 | 227 | −0.02 | 0.02 (−0.17, 0.22) | 0.04 | 0.07 | 96 | 208 | 0.01 | 0.03 (−0.22, 0.29) | 0.03 | 0.03 |
| Malawi | iLiNS−DOSE (31) | KDI | 278 | 94 | 0.11 | 0.05 (−0.18, 0.27) | 0.03 | 0.06 | 271 | 96 | −0.15 | 0.07 (−0.16, 0.31) | 0.04 | 0.04 |
| Mali | PROMIS CS (32) | DMC | 479 | 445 | −0.03 | 0.15 (−0.06, 0.37) | 0.03 | 0.06 | 423 | 476 | −0.09 | 0.10 (−0.09, 0.28) | 0.06 | 0.06 |
| Zimbabwe | SHINE (HIV−) (33) | MDAT | 406 | 359 | −0.04 | 0.13 (−0.02, 0.28) | 0.07 | 0.09 | 389 | 391 | −0.08 | 0.11 (−0.03, 0.26) | 0.09 | 0.09 |
| Zimbabwe | SHINE (HIV+) (34) | MDAT | 91 | 78 | −0.11 | 0.30 (−0.01, 0.61) | 0.02 | 0.03 | 74 | 67 | −0.18 | 0.26 (−0.07, 0.60) | 0.02 | 0.02 |
| | | | **5058** | **7349** | | **I² = 0.48, Tau² = 0.01** | | | **4613** | **6552** | | **I² = 0.00, Tau² = 0.00** | | |
| **Fixed** | | | | | | **0.05 (0.01, 0.09)** | | | | | | **0.11 (0.07, 0.16)** | | |
| **Random** | | | | | | **0.08 (0.01, 0.14)** | | | | | | **0.11 (0.07, 0.16)** | | |

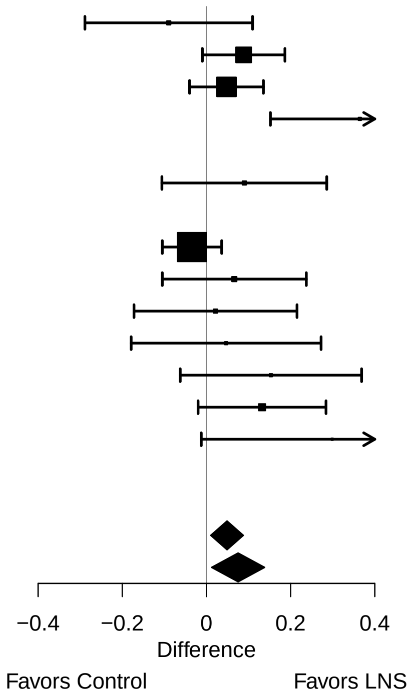

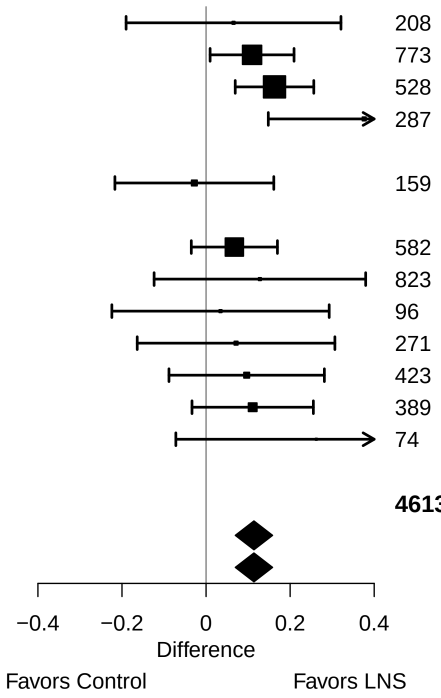

#### Supplemental figure 6G: Mean difference in motor z-score

#### 6G2: Stratified by Household food insecurity

**P−for−interaction = 0.898**
**Difference in MDs = 0.00 (−0.07, 0.08)**

| Country | Trial | Tool | LNS N | Control N | Control Mean | Mild to secure MD (95% CI) | Fixed W | Random W | LNS N | Control N | Control Mean | Moderate to severe MD (95% CI) | Fixed W | Random W |
|---|---|---|---|---|---|---|---|---|---|---|---|---|---|---|
| Bangladesh | JiVitA−4 (21) | BSID−III | 329 | 111 | 0.07 | −0.04 (−0.22, 0.14) | 0.04 | 0.09 | 115 | 32 | −0.15 | 0.06 (−0.29, 0.42) | 0.03 | 0.03 |
| Bangladesh | RDNS (22) | DMC | 987 | 456 | 0.05 | 0.04 (−0.06, 0.15) | 0.12 | 0.12 | 569 | 297 | −0.18 | 0.18 (0.07, 0.28) | 0.32 | 0.32 |
| Bangladesh | WASH−B (23) | EASQ | 853 | 2550 | 0.01 | 0.11 (0.05, 0.18) | 0.29 | 0.13 | 221 | 729 | −0.14 | 0.05 (−0.11, 0.21) | 0.14 | 0.14 |
| Burkina Faso | iLiNS−Zinc (24) | DMC | 389 | 162 | −0.30 | 0.47 (0.22, 0.73) | 0.02 | 0.06 | 357 | 213 | −0.23 | 0.34 (0.11, 0.56) | 0.07 | 0.07 |
| Ghana | GHANA (25) | | | | | | | | | | | | | |
| Ghana | iLiNS−DYADG (26) | KDI | 217 | 412 | 0.02 | 0.07 (−0.08, 0.22) | 0.06 | 0.10 | 84 | 186 | −0.05 | −0.09 (−0.38, 0.20) | 0.04 | 0.04 |
| Haiti | HAITI (27) | | | | | | | | | | | | | |
| Kenya | WASH−B (28) | EASQ | 1214 | 4244 | 0.01 | 0.01 (−0.06, 0.08) | 0.30 | 0.13 | 146 | 487 | −0.11 | 0.01 (−0.19, 0.21) | 0.09 | 0.09 |
| Madagascar | MAHAY (29) | ASQI | 940 | 892 | 0.05 | 0.04 (−0.16, 0.24) | 0.03 | 0.08 | 336 | 389 | −0.27 | 0.12 (−0.23, 0.48) | 0.03 | 0.03 |
| Malawi | iLiNS−DYADM (30) | KDI | 67 | 116 | −0.01 | −0.02 (−0.31, 0.26) | 0.02 | 0.05 | 146 | 314 | 0.00 | 0.04 (−0.15, 0.23) | 0.10 | 0.10 |
| Malawi | iLiNS−DOSE (31) | KDI | 134 | 51 | 0.11 | −0.01 (−0.31, 0.28) | 0.02 | 0.05 | 403 | 133 | −0.06 | 0.09 (−0.12, 0.29) | 0.09 | 0.09 |
| Mali | PROMIS CS (32) | DMC | | | | | | | | | | | | |
| Zimbabwe | SHINE (HIV−) (33) | MDAT | 631 | 586 | −0.04 | 0.12 (0.00, 0.25) | 0.08 | 0.11 | 140 | 154 | −0.14 | 0.11 (−0.09, 0.31) | 0.09 | 0.09 |
| Zimbabwe | SHINE (HIV+) (34) | MDAT | 119 | 105 | −0.15 | 0.36 (0.12, 0.59) | 0.02 | 0.07 | 41 | 37 | −0.14 | 0.07 (−0.44, 0.58) | 0.01 | 0.01 |
| | | | **5880** | **9685** | | **I² = 0.57, Tau² = 0.01** | | | **2558** | **2971** | | **I² = 0.00, Tau² = 0.00** | | |
| **Fixed** | | | | | | **0.07 (0.04, 0.11)** | | | | | | **0.11 (0.05, 0.17)** | | |
| **Random** | | | | | | **0.09 (0.01, 0.18)** | | | | | | **0.11 (0.05, 0.17)** | | |

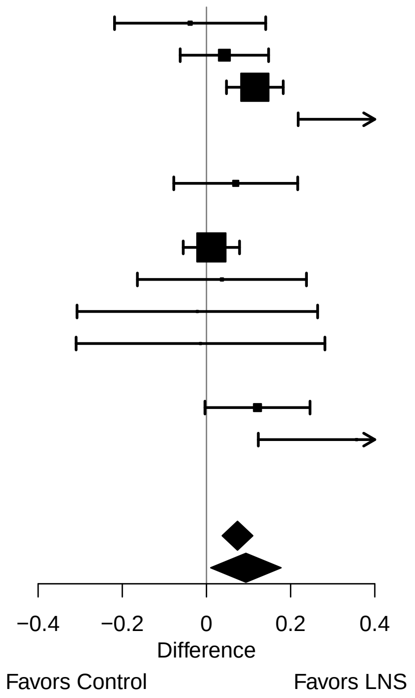

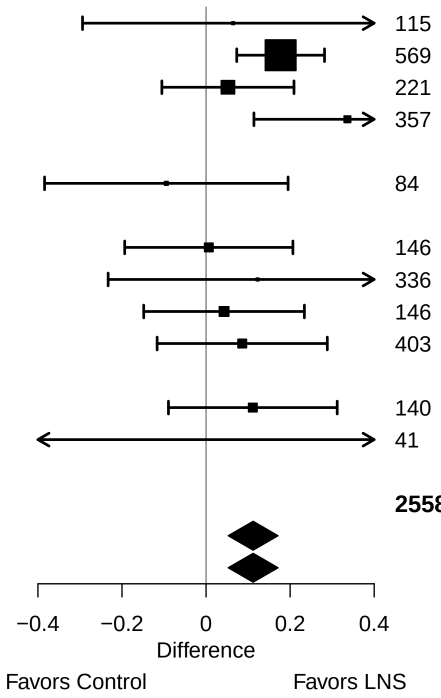

#### Supplemental figure 6G: Mean difference in motor z-score

##### 6G3: Stratified by Household source water quality

**P−for−interaction = 0.514**
**Difference in MDs = 0.04 (−0.07, 0.14)**

| Country | Trial | Tool | Improved LNS N | Improved Control N | Improved Control Mean | Improved MD (95% CI) | Improved Fixed W | Improved Random W | Unimproved LNS N | Unimproved Control N | Unimproved Control Mean | Unimproved MD (95% CI) | Unimproved Fixed W | Unimproved Random W |
|---|---|---|---|---|---|---|---|---|---|---|---|---|---|---|
| Bangladesh | JiVitA−4 (21) | BSID−III | | | | | | | | | | | | |
| Bangladesh | RDNS (22) | DMC | | | | | | | | | | | | |
| Bangladesh | WASH−B (23) | EASQ | 460 | 981 | −0.09 | 0.20 (0.08, 0.31) | 0.25 | 0.21 | 67 | 117 | −0.12 | 0.20 (−0.08, 0.48) | 0.08 | 0.10 |
| Burkina Faso | iLiNS−Zinc (24) | DMC | 220 | 82 | −0.07 | 0.32 (0.07, 0.57) | 0.05 | 0.07 | 526 | 293 | −0.32 | 0.41 (0.25, 0.57) | 0.26 | 0.21 |
| Ghana | GHANA (25) | | | | | | | | | | | | | |
| Ghana | iLiNS−DYADG (26) | KDI | | | | | | | | | | | | |
| Haiti | HAITI (27) | | | | | | | | | | | | | |
| Kenya | WASH−B (28) | EASQ | 413 | 1419 | −0.01 | 0.02 (−0.10, 0.14) | 0.25 | 0.21 | 236 | 649 | 0.05 | 0.04 (−0.12, 0.21) | 0.23 | 0.20 |
| Madagascar | MAHAY (29) | ASQI | 405 | 430 | −0.10 | 0.08 (−0.26, 0.41) | 0.03 | 0.04 | 1134 | 1133 | −0.01 | 0.08 (−0.13, 0.29) | 0.14 | 0.15 |
| Malawi | iLiNS−DYADM (30) | KDI | 194 | 398 | 0.02 | −0.01 (−0.17, 0.16) | 0.13 | 0.13 | 20 | 36 | −0.16 | 0.32 (−0.24, 0.87) | 0.02 | 0.03 |
| Malawi | iLiNS−DOSE (31) | KDI | 515 | 177 | 0.00 | 0.06 (−0.11, 0.23) | 0.12 | 0.13 | 44 | 15 | −0.26 | 0.21 (−0.41, 0.84) | 0.02 | 0.02 |
| Mali | PROMIS CS (32) | DMC | 514 | 539 | −0.04 | 0.16 (−0.07, 0.38) | 0.07 | 0.08 | 347 | 368 | −0.12 | 0.07 (−0.15, 0.28) | 0.14 | 0.15 |
| Zimbabwe | SHINE (HIV−) (33) | MDAT | 228 | 220 | 0.11 | −0.04 (−0.24, 0.16) | 0.08 | 0.10 | 131 | 122 | −0.11 | 0.10 (−0.18, 0.38) | 0.08 | 0.10 |
| Zimbabwe | SHINE (HIV+) (34) | MDAT | 37 | 41 | −0.01 | 0.04 (−0.34, 0.42) | 0.02 | 0.03 | 25 | 25 | −0.25 | −0.10 (−0.70, 0.50) | 0.02 | 0.03 |
| | | | **2986** | **4287** | | **I² = 0.28, Tau² = 0.00** | | | **2530** | **2758** | | **I² = 0.42, Tau² = 0.01** | | |
| **Fixed** | | | | | | **0.09 (0.03, 0.15)** | | | | | | **0.17 (0.09, 0.25)** | | |
| **Random** | | | | | | **0.09 (0.02, 0.16)** | | | | | | **0.16 (0.06, 0.26)** | | |

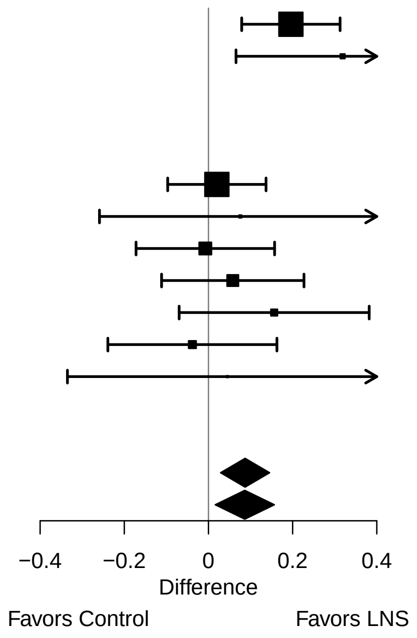

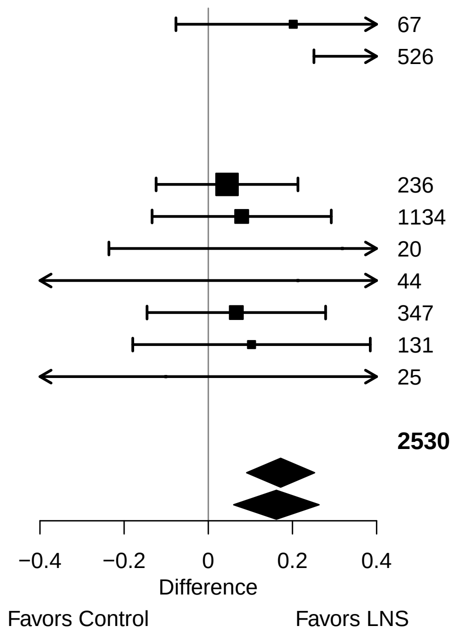

#### Supplemental figure 6G: Mean difference in motor z-score

#### 6G4: Stratified by Household sanitation

**P−for−interaction = 0.193**
**Difference in MDs = −0.07 (−0.17, 0.03)**

| Country | Trial | Tool | Improved LNS N | Improved Control N | Improved Control Mean | Improved MD (95% CI) | Improved Fixed W | Improved Random W | Unimproved LNS N | Unimproved Control N | Unimproved Control Mean | Unimproved MD (95% CI) | Unimproved Fixed W | Unimproved Random W |
|---|---|---|---|---|---|---|---|---|---|---|---|---|---|---|
| Bangladesh | JiVitA−4 (21) | BSID−III | 353 | 112 | 0.08 | −0.02 (−0.18, 0.14) | 0.09 | 0.11 | 91 | 31 | −0.20 | −0.02 (−0.56, 0.52) | 0.01 | 0.02 |
| Bangladesh | RDNS (22) | DMC | 1103 | 531 | −0.01 | 0.11 (0.03, 0.18) | 0.41 | 0.30 | 451 | 222 | −0.11 | 0.08 (−0.05, 0.21) | 0.18 | 0.16 |
| Bangladesh | WASH−B (23) | EASQ | 472 | 998 | −0.09 | 0.21 (0.10, 0.32) | 0.20 | 0.20 | 29 | 53 | −0.20 | 0.18 (−0.39, 0.74) | 0.01 | 0.02 |
| Burkina Faso | iLiNS−Zinc (24) | DMC | 16 | 11 | −0.37 | 0.74 (−0.01, 1.49) | 0.00 | 0.01 | 730 | 364 | −0.26 | 0.39 (0.22, 0.56) | 0.11 | 0.12 |
| Ghana | GHANA (25) | | | | | | | | | | | | | |
| Ghana | iLiNS−DYADG (26) | KDI | 292 | 586 | −0.01 | 0.03 (−0.11, 0.16) | 0.12 | 0.14 | 9 | 13 | 0.07 | 0.02 (−0.74, 0.79) | 0.01 | 0.01 |
| Haiti | HAITI (27) | | | | | | | | | | | | | |
| Kenya | WASH−B (28) | EASQ | 94 | 337 | 0.12 | 0.09 (−0.13, 0.30) | 0.05 | 0.07 | 555 | 1732 | −0.01 | 0.02 (−0.08, 0.13) | 0.31 | 0.20 |
| Madagascar | MAHAY (29) | ASQI | | | | | | | | | | | | |
| Malawi | iLiNS−DYADM (30) | KDI | 19 | 41 | −0.13 | 0.27 (−0.16, 0.69) | 0.01 | 0.02 | 195 | 393 | 0.02 | 0.00 (−0.17, 0.16) | 0.11 | 0.13 |
| Malawi | iLiNS−DOSE (31) | KDI | 12 | 9 | 0.31 | 0.19 (−0.44, 0.83) | 0.01 | 0.01 | 547 | 183 | −0.04 | 0.08 (−0.09, 0.24) | 0.11 | 0.13 |
| Mali | PROMIS CS (32) | DMC | 665 | 658 | −0.08 | 0.17 (−0.03, 0.37) | 0.06 | 0.08 | 210 | 241 | 0.00 | −0.01 (−0.26, 0.25) | 0.05 | 0.07 |
| Zimbabwe | SHINE (HIV−) (33) | MDAT | 133 | 108 | 0.05 | 0.08 (−0.17, 0.34) | 0.04 | 0.05 | 225 | 235 | 0.02 | −0.03 (−0.22, 0.16) | 0.09 | 0.11 |
| Zimbabwe | SHINE (HIV+) (34) | MDAT | 25 | 17 | −0.02 | −0.22 (−0.78, 0.34) | 0.01 | 0.01 | 37 | 49 | −0.13 | 0.09 (−0.28, 0.46) | 0.02 | 0.04 |
| | | | **3184** | **3408** | | **I² = 0.17, Tau² = 0.00** | | | **3079** | **3516** | | **I² = 0.42, Tau² = 0.01** | | |
| **Fixed** | | | | | | **0.11 (0.06, 0.16)** | | | | | | **0.07 (0.02, 0.13)** | | |
| **Random** | | | | | | **0.11 (0.05, 0.17)** | | | | | | **0.08 (0.00, 0.16)** | | |

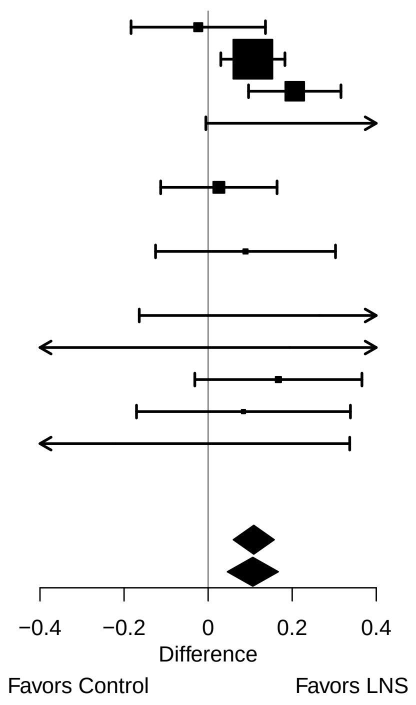

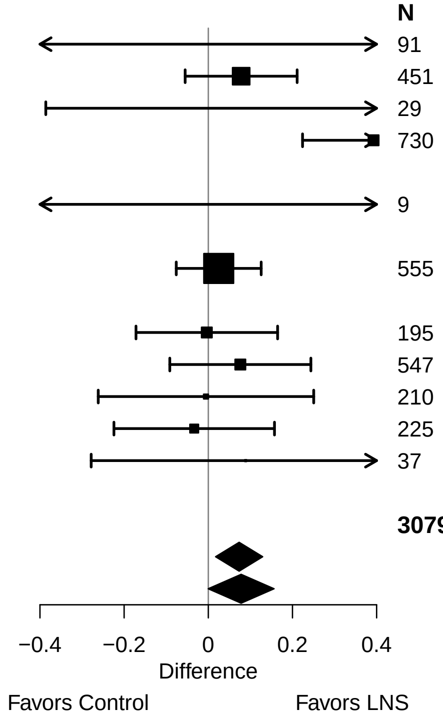

#### Supplemental figure 6G: Mean difference in motor z-score

#### 6G5: Stratified by Home environment

P−for−interaction = 0.565
Difference in MDs = −0.02 (−0.09, 0.05)

| Country | Trial | Tool | At least median: LNS N | Control N | Control Mean | MD (95% CI) | Fixed W | Random W | Less than median: LNS N | Control N | Control Mean | MD (95% CI) | Fixed W | Random W |
|---|---|---|---|---|---|---|---|---|---|---|---|---|---|---|
| Bangladesh | JiVitA−4 (21) | BSID−III | | | | | | | | | | | | |
| Bangladesh | RDNS (22) | DMC | 1115 | 548 | 0.07 | 0.08 (0.02, 0.14) | 0.38 | 0.34 | 439 | 203 | −0.35 | 0.17 (−0.01, 0.34) | 0.10 | 0.13 |
| Bangladesh | WASH−B (23) | EASQ | 605 | 1614 | 0.13 | 0.12 (0.02, 0.21) | 0.16 | 0.17 | 469 | 1664 | −0.17 | 0.04 (−0.06, 0.14) | 0.33 | 0.21 |
| Burkina Faso | iLiNS−Zinc (24) | DMC | 473 | 199 | 0.11 | 0.24 (0.07, 0.41) | 0.05 | 0.06 | 272 | 175 | −0.67 | 0.45 (0.18, 0.73) | 0.04 | 0.08 |
| Ghana | GHANA (25) | | | | | | | | | | | | | |
| Ghana | iLiNS−DYADG (26) | KDI | 191 | 377 | 0.10 | 0.05 (−0.10, 0.21) | 0.06 | 0.07 | 111 | 224 | −0.18 | −0.02 (−0.27, 0.24) | 0.05 | 0.09 |
| Haiti | HAITI (27) | | | | | | | | | | | | | |
| Kenya | WASH−B (28) | EASQ | 809 | 2682 | 0.15 | 0.01 (−0.07, 0.09) | 0.23 | 0.23 | 552 | 2060 | −0.20 | −0.01 (−0.11, 0.09) | 0.32 | 0.21 |
| Madagascar | MAHAY (29) | ASQI | 843 | 839 | 0.05 | 0.11 (−0.12, 0.34) | 0.03 | 0.03 | 770 | 764 | −0.13 | 0.03 (−0.17, 0.24) | 0.08 | 0.12 |
| Malawi | iLiNS−DYADM (30) | KDI | 136 | 287 | 0.11 | 0.02 (−0.15, 0.20) | 0.05 | 0.05 | 78 | 149 | −0.22 | 0.05 (−0.25, 0.36) | 0.03 | 0.07 |
| Malawi | iLiNS−DOSE (31) | KDI | 376 | 135 | 0.03 | 0.06 (−0.13, 0.25) | 0.04 | 0.05 | 269 | 86 | −0.14 | 0.04 (−0.21, 0.29) | 0.05 | 0.09 |
| Mali | PROMIS CS (32) | DMC | | | | | | | | | | | | |
| Zimbabwe | SHINE (HIV−) (33) | MDAT | | | | | | | | | | | | |
| Zimbabwe | SHINE (HIV+) (34) | MDAT | | | | | | | | | | | | |
| | | | **4548** | **6681** | | **I² = 0.10, Tau² = 0.00** | | | **2960** | **5325** | | **I² = 0.39, Tau² = 0.01** | | |
| **Fixed** | | | | | | **0.07 (0.04, 0.11)** | | | | | | **0.05 (−0.01, 0.11)** | | |
| **Random** | | | | | | **0.07 (0.03, 0.12)** | | | | | | **0.07 (−0.02, 0.16)** | | |

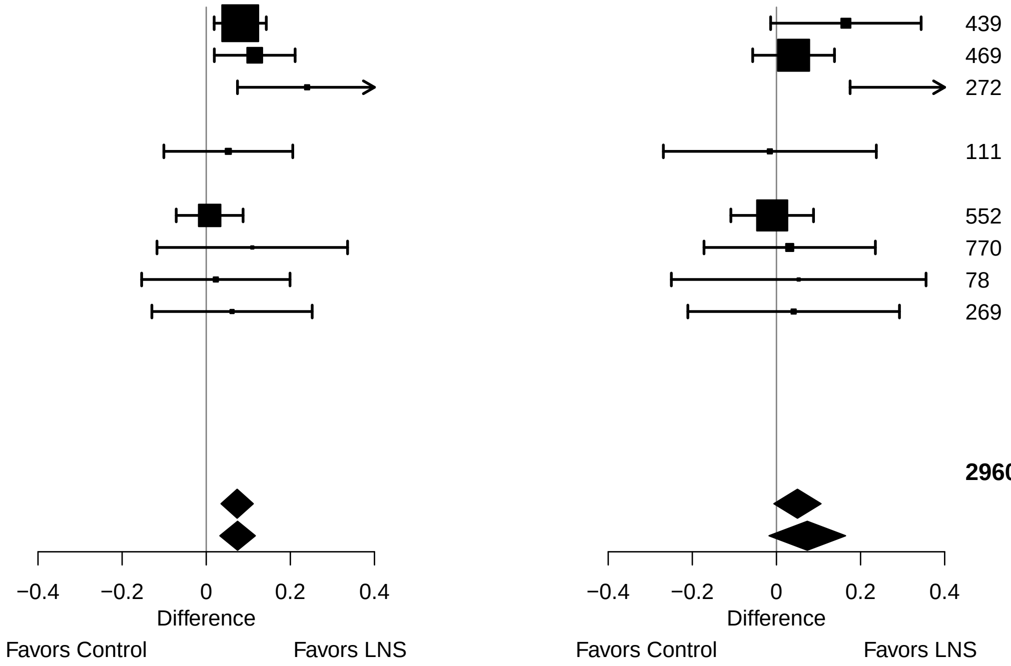

#### Supplemental figure 6G: Mean difference in motor z-score

#### 6G6: Stratified by Season at the time of assessment

**P−for−interaction = 0.827**
**Difference in MDs = −0.01 (−0.07, 0.05)**

| Country | Trial | Tool | Dry LNS N | Dry Control N | Dry Control Mean | Dry MD (95% CI) | Dry Fixed W | Dry Random W | Rainy LNS N | Rainy Control N | Rainy Control Mean | Rainy MD (95% CI) | Rainy Fixed W | Rainy Random W |
|---|---|---|---|---|---|---|---|---|---|---|---|---|---|---|
| Bangladesh | JiVitA−4 (21) | BSID−III | 251 | 70 | 0.26 | −0.03 (−0.22, 0.17) | 0.05 | 0.08 | 194 | 73 | −0.22 | −0.09 (−0.38, 0.20) | 0.02 | 0.05 |
| Bangladesh | RDNS (22) | DMC | 698 | 326 | −0.02 | 0.09 (−0.01, 0.18) | 0.20 | 0.13 | 858 | 427 | −0.06 | 0.11 (0.02, 0.20) | 0.25 | 0.18 |
| Bangladesh | WASH−B (23) | EASQ | 608 | 1896 | 0.04 | 0.08 (−0.01, 0.17) | 0.24 | 0.14 | 466 | 1383 | −0.11 | 0.14 (0.04, 0.24) | 0.19 | 0.17 |
| Burkina Faso | iLiNS−Zinc (24) | DMC | 514 | 274 | −0.31 | 0.42 (0.21, 0.63) | 0.04 | 0.08 | 232 | 101 | −0.13 | 0.34 (0.15, 0.54) | 0.05 | 0.09 |
| Ghana | GHANA (25) | | | | | | | | | | | | | |
| Ghana | iLiNS−DYADG (26) | KDI | 177 | 356 | 0.02 | −0.02 (−0.20, 0.15) | 0.06 | 0.09 | 125 | 245 | −0.04 | 0.11 (−0.12, 0.33) | 0.04 | 0.08 |
| Haiti | HAITI (27) | | | | | | | | | | | | | |
| Kenya | WASH−B (28) | EASQ | 315 | 1059 | −0.30 | 0.09 (−0.01, 0.20) | 0.17 | 0.13 | 1046 | 3682 | 0.08 | −0.01 (−0.09, 0.07) | 0.33 | 0.20 |
| Madagascar | MAHAY (29) | ASQI | | | | | | | | | | | | |
| Malawi | iLiNS−DYADM (30) | KDI | 153 | 308 | 0.02 | 0.04 (−0.15, 0.23) | 0.05 | 0.08 | 61 | 127 | −0.06 | −0.01 (−0.30, 0.28) | 0.02 | 0.05 |
| Malawi | iLiNS−DOSE (31) | KDI | 395 | 118 | −0.09 | 0.01 (−0.19, 0.21) | 0.04 | 0.08 | 244 | 100 | 0.02 | 0.14 (−0.11, 0.38) | 0.03 | 0.07 |
| Mali | PROMIS CS (32) | DMC | | | | | | | | | | | | |
| Zimbabwe | SHINE (HIV−) (33) | MDAT | 603 | 568 | −0.08 | 0.14 (0.02, 0.25) | 0.13 | 0.12 | 238 | 231 | 0.03 | 0.02 (−0.18, 0.23) | 0.05 | 0.08 |
| Zimbabwe | SHINE (HIV+) (34) | MDAT | 121 | 110 | −0.19 | 0.30 (0.05, 0.55) | 0.03 | 0.06 | 49 | 41 | −0.09 | 0.32 (−0.07, 0.71) | 0.01 | 0.03 |
| | | | **3835** | **5085** | | **$I^2$ = 0.46, $Tau^2$ = 0.01** | | | **3513** | **6410** | | **$I^2$ = 0.49, $Tau^2$ = 0.01** | | |
| **Fixed** | | | | | | **0.09 (0.05, 0.14)** | | | | | | **0.08 (0.04, 0.12)** | | |
| **Random** | | | | | | **0.10 (0.02, 0.18)** | | | | | | **0.10 (0.02, 0.17)** | | |

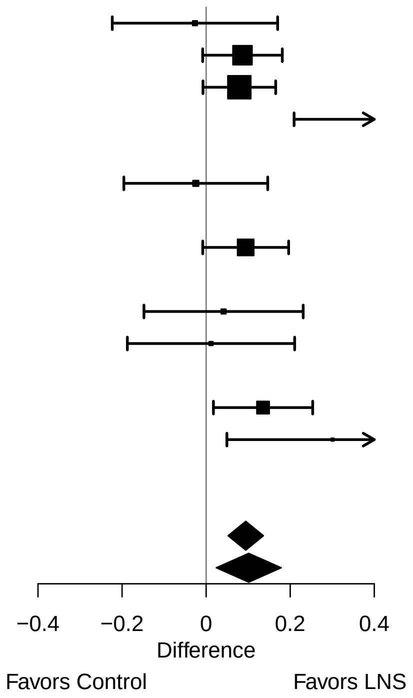

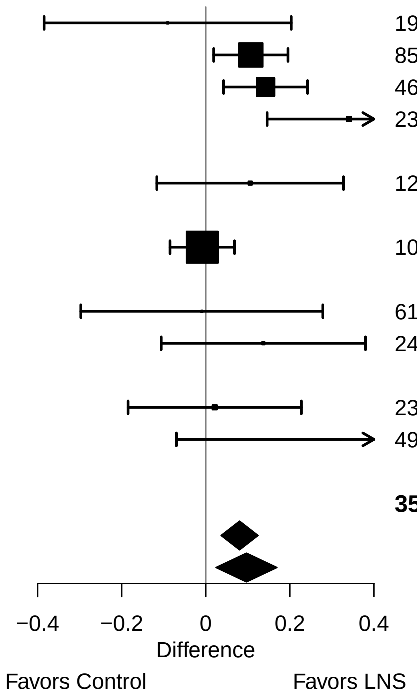

### Supplemental figure 6H: Motor lowest decile prevalence ratio

#### 6H1: Stratified by Household socio-economic status

**P−for−interaction = 0.878**
**Ratio of PRs = 0.99 (0.83, 1.17)**

| Country | Trial | Tool | At least median: LNS N | Control N | Control Prevalence | PR (95% CI) | Fixed W | Random W | Less than median: LNS N | Control N | Control Prevalence | PR (95% CI) | Fixed W | Random W |
|---|---|---|---|---|---|---|---|---|---|---|---|---|---|---|
| Bangladesh | JiVitA−4 (21) | BSID−III | | | | | | | | | | | | |
| Bangladesh | RDNS (22) | DMC | 783 | 381 | 7.1 | 1.08 (0.71, 1.65) | 0.10 | 0.11 | 773 | 372 | 15.3 | 0.73 (0.56, 0.97) | 0.19 | 0.19 |
| Bangladesh | WASH−B (23) | EASQ | 546 | 1663 | 7.6 | 0.79 (0.57, 1.09) | 0.17 | 0.15 | 528 | 1616 | 13.6 | 0.83 (0.63, 1.11) | 0.18 | 0.18 |
| Burkina Faso | iLiNS−Zinc (24) | DMC | 459 | 163 | 16.0 | 0.38 (0.24, 0.62) | 0.08 | 0.09 | 287 | 212 | 14.6 | 0.67 (0.39, 1.13) | 0.05 | 0.05 |
| Ghana | GHANA (25) | | | | | | | | | | | | | |
| Ghana | iLiNS−DYADG (26) | KDI | 143 | 311 | 10.0 | 0.98 (0.54, 1.79) | 0.05 | 0.07 | 159 | 288 | 9.0 | 1.32 (0.76, 2.31) | 0.05 | 0.05 |
| Haiti | HAITI (27) | | | | | | | | | | | | | |
| Kenya | WASH−B (28) | EASQ | 778 | 2706 | 9.3 | 0.84 (0.65, 1.08) | 0.28 | 0.19 | 582 | 2036 | 11.6 | 0.89 (0.69, 1.14) | 0.23 | 0.23 |
| Madagascar | MAHAY (29) | ASQI | 741 | 845 | 6.2 | 0.90 (0.50, 1.62) | 0.05 | 0.07 | 823 | 724 | 15.7 | 0.84 (0.52, 1.36) | 0.06 | 0.06 |
| Malawi | iLiNS−DYADM (30) | KDI | 118 | 227 | 8.8 | 0.96 (0.47, 1.99) | 0.03 | 0.05 | 96 | 208 | 12.5 | 0.75 (0.37, 1.54) | 0.03 | 0.03 |
| Malawi | iLiNS−DOSE (31) | KDI | 278 | 94 | 6.4 | 1.35 (0.57, 3.21) | 0.02 | 0.04 | 271 | 96 | 12.5 | 0.83 (0.44, 1.56) | 0.04 | 0.04 |
| Mali | PROMIS CS (32) | DMC | 479 | 445 | 11.0 | 0.83 (0.52, 1.33) | 0.08 | 0.10 | 423 | 476 | 11.1 | 0.79 (0.51, 1.20) | 0.08 | 0.08 |
| Zimbabwe | SHINE (HIV−) (33) | MDAT | 406 | 359 | 10.6 | 0.93 (0.59, 1.46) | 0.09 | 0.10 | 389 | 391 | 11.3 | 0.87 (0.58, 1.30) | 0.09 | 0.09 |
| Zimbabwe | SHINE (HIV+) (34) | MDAT | 91 | 78 | 11.5 | 0.67 (0.27, 1.62) | 0.02 | 0.03 | 74 | 67 | 13.4 | 0.70 (0.27, 1.82) | 0.02 | 0.02 |
| | | | **4822** | **7272** | | **I² = 0.29, Tau² = 0.03** | | | **4405** | **6486** | | **I² = 0.00, Tau² = 0.00** | | |
| **Fixed** | | | | | | **0.83 (0.72, 0.95)** | | | | | | **0.83 (0.74, 0.93)** | | |
| **Random** | | | | | | **0.83 (0.69, 0.99)** | | | | | | **0.83 (0.74, 0.93)** | | |

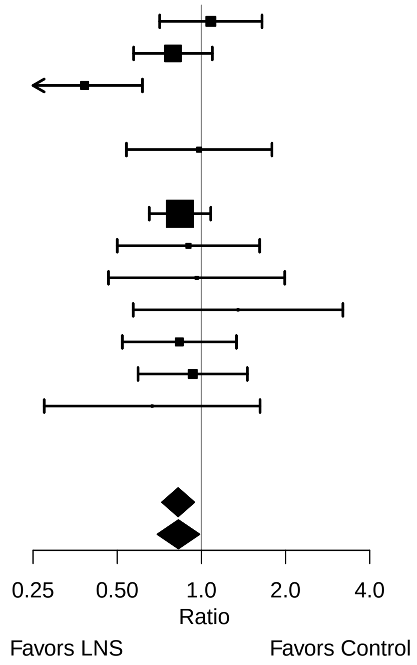

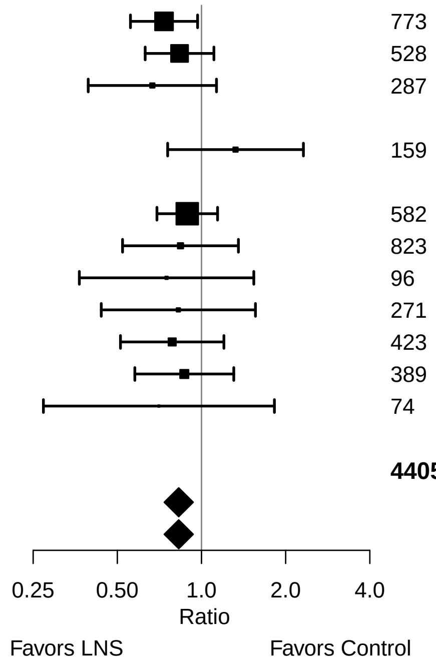

#### Supplemental figure 6H: Motor lowest decile prevalence ratio

##### 6H2: Stratified by Household food insecurity

**P−for−interaction = 0.594**
**Ratio of PRs = 0.94 (0.75, 1.18)**

| Country | Trial | Tool | Mild to secure: LNS N | Control N | Control Prevalence | PR (95% CI) | Fixed W | Random W | Moderate to severe: LNS N | Control N | Control Prevalence | PR (95% CI) | Fixed W | Random W |
|---|---|---|---|---|---|---|---|---|---|---|---|---|---|---|
| Bangladesh | JiVitA−4 (21) | BSID−III | | | | | | | | | | | | |
| Bangladesh | RDNS (22) | DMC | 987 | 456 | 7.2 | 1.15 (0.79, 1.66) | 0.11 | 0.14 | 569 | 297 | 17.2 | 0.67 (0.47, 0.94) | 0.27 | 0.26 |
| Bangladesh | WASH−B (23) | EASQ | 853 | 2550 | 10.0 | 0.79 (0.63, 0.99) | 0.27 | 0.19 | 221 | 729 | 12.8 | 0.92 (0.62, 1.37) | 0.20 | 0.19 |
| Burkina Faso | iLiNS−Zinc (24) | DMC | 389 | 162 | 15.4 | 0.43 (0.28, 0.66) | 0.08 | 0.12 | 357 | 213 | 15.0 | 0.56 (0.34, 0.93) | 0.12 | 0.13 |
| Ghana | GHANA (25) | | | | | | | | | | | | | |
| Ghana | iLiNS−DYADG (26) | KDI | 217 | 412 | 9.2 | 1.00 (0.60, 1.67) | 0.05 | 0.09 | 84 | 186 | 10.2 | 1.52 (0.79, 2.92) | 0.07 | 0.08 |
| Haiti | HAITI (27) | | | | | | | | | | | | | |
| Kenya | WASH−B (28) | EASQ | 1214 | 4244 | 10.0 | 0.88 (0.71, 1.08) | 0.31 | 0.19 | 146 | 487 | 13.6 | 0.76 (0.45, 1.29) | 0.11 | 0.12 |
| Madagascar | MAHAY (29) | ASQI | 940 | 892 | 9.0 | 0.90 (0.53, 1.53) | 0.05 | 0.09 | 336 | 389 | 15.4 | 0.93 (0.50, 1.73) | 0.08 | 0.08 |
| Malawi | iLiNS−DYADM (30) | KDI | 67 | 116 | 11.2 | 1.07 (0.47, 2.44) | 0.02 | 0.05 | 146 | 314 | 10.2 | 0.74 (0.38, 1.43) | 0.07 | 0.08 |
| Malawi | iLiNS−DOSE (31) | KDI | | | | | | | | | | | | |
| Mali | PROMIS CS (32) | DMC | | | | | | | | | | | | |
| Zimbabwe | SHINE (HIV−) (33) | MDAT | 631 | 586 | 10.4 | 0.96 (0.66, 1.39) | 0.10 | 0.13 | 140 | 154 | 13.0 | 0.72 (0.35, 1.44) | 0.06 | 0.07 |
| Zimbabwe | SHINE (HIV+) (34) | MDAT | | | | | | | | | | | | |
| | | | **5298** | **9418** | | **I² = 0.50, Tau² = 0.04** | | | **1999** | **2769** | | **I² = 0.07, Tau² = 0.00** | | |
| **Fixed** | | | | | | **0.85 (0.75, 0.95)** | | | | | | **0.78 (0.65, 0.93)** | | |
| **Random** | | | | | | **0.85 (0.69, 1.04)** | | | | | | **0.78 (0.65, 0.94)** | | |

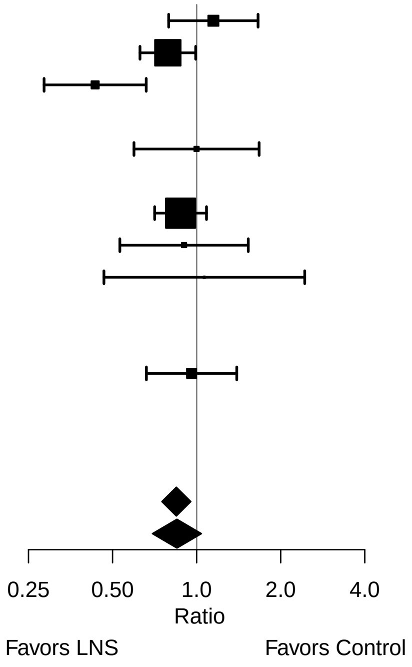

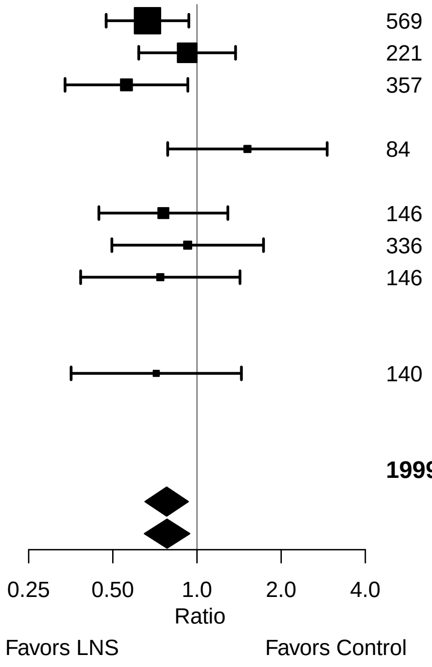

#### Supplemental figure 6H: Motor lowest decile prevalence ratio

#### 6H3: Stratified by Household source water quality

**P−for−interaction = 0.723**
**Ratio of PRs = 1.06 (0.78, 1.42)**

| Country | Trial | Tool | LNS N | Control N | Control Prevalence | Improved PR (95% CI) | Fixed W | Random W | LNS N | Control N | Control Prevalence | Unimproved PR (95% CI) | Fixed W | Random W |
|---|---|---|---|---|---|---|---|---|---|---|---|---|---|---|
| Bangladesh | JiVitA−4 (21) | BSID−III | | | | | | | | | | | | |
| Bangladesh | RDNS (22) | DMC | | | | | | | | | | | | |
| Bangladesh | WASH−B (23) | EASQ | 460 | 981 | 11.6 | 0.84 (0.61, 1.17) | 0.33 | 0.31 | 67 | 117 | 12.0 | 0.62 (0.23, 1.71) | 0.04 | 0.05 |
| Burkina Faso | iLiNS−Zinc (24) | DMC | 220 | 82 | 12.2 | 0.41 (0.20, 0.85) | 0.07 | 0.07 | 526 | 293 | 16.0 | 0.53 (0.39, 0.72) | 0.43 | 0.35 |
| Ghana | GHANA (25) | | | | | | | | | | | | | |
| Ghana | iLiNS−DYADG (26) | KDI | | | | | | | | | | | | |
| Haiti | HAITI (27) | | | | | | | | | | | | | |
| Kenya | WASH−B (28) | EASQ | 413 | 1419 | 10.2 | 1.02 (0.71, 1.47) | 0.27 | 0.26 | 236 | 649 | 9.2 | 0.87 (0.55, 1.37) | 0.19 | 0.21 |
| Madagascar | MAHAY (29) | ASQI | 405 | 430 | 14.0 | 0.87 (0.44, 1.70) | 0.08 | 0.09 | 1134 | 1133 | 9.4 | 0.92 (0.54, 1.58) | 0.14 | 0.16 |
| Malawi | iLiNS−DYADM (30) | KDI | | | | | | | | | | | | |
| Malawi | iLiNS−DOSE (31) | KDI | | | | | | | | | | | | |
| Mali | PROMIS CS (32) | DMC | 514 | 539 | 10.2 | 0.69 (0.42, 1.11) | 0.15 | 0.16 | 347 | 368 | 12.8 | 0.90 (0.52, 1.56) | 0.13 | 0.15 |
| Zimbabwe | SHINE (HIV−) (33) | MDAT | 228 | 220 | 9.5 | 0.87 (0.49, 1.55) | 0.11 | 0.12 | 131 | 122 | 9.8 | 1.16 (0.52, 2.59) | 0.06 | 0.08 |
| Zimbabwe | SHINE (HIV+) (34) | MDAT | | | | | | | | | | | | |
| | | | **2240** | **3671** | | **I² = 0.08, Tau² = 0.01** | | | **2441** | **2682** | | **I² = 0.32, Tau² = 0.02** | | |
| **Fixed** | | | | | | **0.82 (0.68, 0.99)** | | | | | | **0.72 (0.59, 0.88)** | | |
| **Random** | | | | | | **0.82 (0.67, 1.00)** | | | | | | **0.75 (0.59, 0.95)** | | |

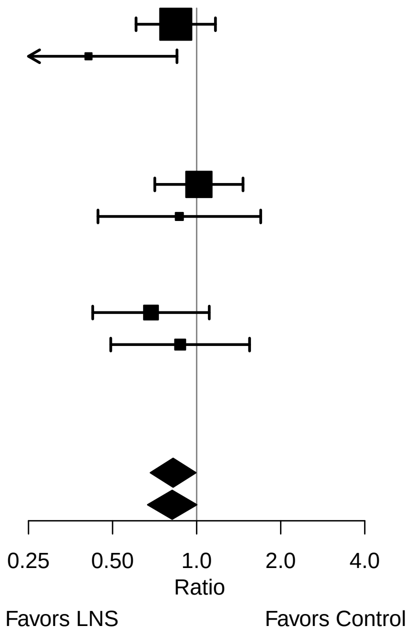

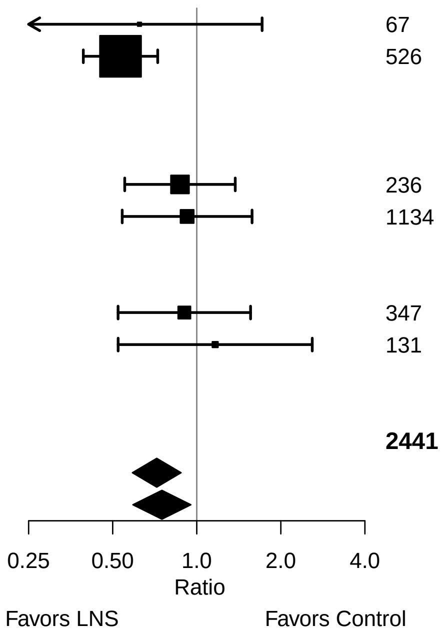

#### Supplemental figure 6H: Motor lowest decile prevalence ratio

##### 6H4: Stratified by Household sanitation

**P−for−interaction = 0.228**
**Ratio of PRs = 1.24 (0.87, 1.76)**

| Country | Trial | Tool | LNS N | Control N | Control Prevalence | Improved PR (95% CI) | Fixed W | Random W | LNS N | Control N | Control Prevalence | Unimproved PR (95% CI) | Fixed W | Random W |
|---|---|---|---|---|---|---|---|---|---|---|---|---|---|---|
| Bangladesh | JiVitA−4 (21) | BSID−III | | | | | | | | | | | | |
| Bangladesh | RDNS (22) | DMC | 1103 | 531 | 10.4 | 0.81 (0.59, 1.12) | 0.52 | 0.52 | 451 | 222 | 13.1 | 0.92 (0.58, 1.45) | 0.22 | 0.22 |
| Bangladesh | WASH−B (23) | EASQ | | | | | | | | | | | | |
| Burkina Faso | iLiNS−Zinc (24) | DMC | | | | | | | | | | | | |
| Ghana | GHANA (25) | | | | | | | | | | | | | |
| Ghana | iLiNS−DYADG (26) | KDI | | | | | | | | | | | | |
| Haiti | HAITI (27) | | | | | | | | | | | | | |
| Kenya | WASH−B (28) | EASQ | 94 | 337 | 9.2 | 0.81 (0.35, 1.89) | 0.07 | 0.07 | 555 | 1732 | 10.0 | 0.99 (0.72, 1.34) | 0.49 | 0.49 |
| Madagascar | MAHAY (29) | ASQI | | | | | | | | | | | | |
| Malawi | iLiNS−DYADM (30) | KDI | | | | | | | | | | | | |
| Malawi | iLiNS−DOSE (31) | KDI | | | | | | | | | | | | |
| Mali | PROMIS CS (32) | DMC | 665 | 658 | 10.8 | 0.81 (0.53, 1.23) | 0.31 | 0.31 | 210 | 241 | 10.8 | 0.93 (0.54, 1.60) | 0.16 | 0.16 |
| Zimbabwe | SHINE (HIV−) (33) | MDAT | 133 | 108 | 10.2 | 0.66 (0.32, 1.39) | 0.10 | 0.10 | 225 | 235 | 9.4 | 1.19 (0.66, 2.15) | 0.13 | 0.13 |
| Zimbabwe | SHINE (HIV+) (34) | MDAT | | | | | | | | | | | | |
| | | | **1995** | **1634** | | **I² = 0.00, Tau² = 0.00** | | | **1441** | **2430** | | **I² = 0.00, Tau² = 0.00** | | |
| **Fixed** | | | | | | **0.80 (0.63, 1.00)** | | | | | | **0.98 (0.79, 1.22)** | | |
| **Random** | | | | | | **0.80 (0.63, 1.00)** | | | | | | **0.98 (0.79, 1.22)** | | |

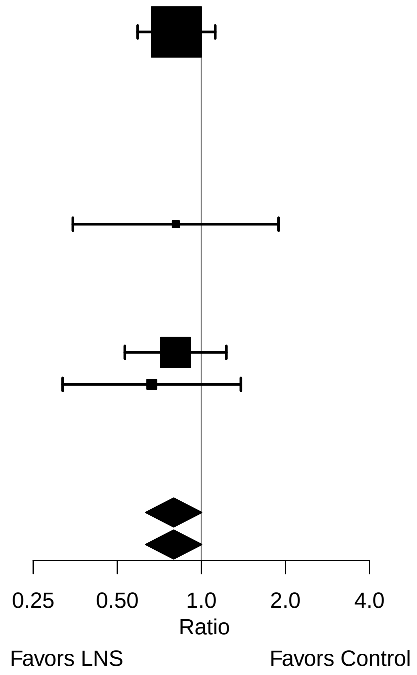

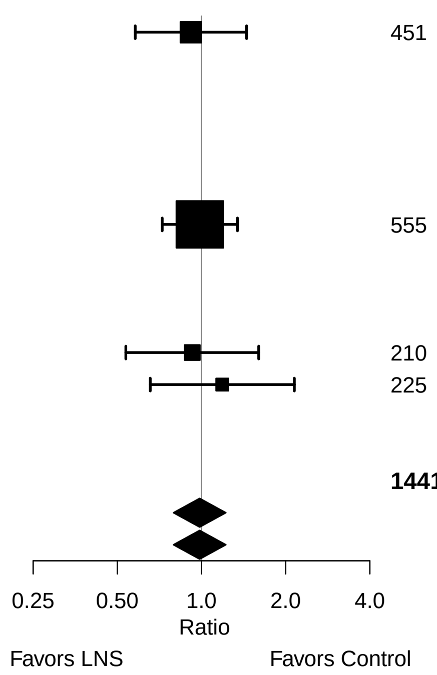

#### Supplemental figure 6H: Motor lowest decile prevalence ratio

#### 6H5: Stratified by Home environment

**P−for−interaction = 0.766**
**Ratio of PRs = 1.03 (0.84, 1.27)**

| Country | Trial | Tool | At least median: LNS N | Control N | Control Prevalence | PR (95% CI) | Fixed W | Random W | Less than median: LNS N | Control N | Control Prevalence | PR (95% CI) | Fixed W | Random W |
|---|---|---|---|---|---|---|---|---|---|---|---|---|---|---|
| Bangladesh | JiVitA−4 (21) | BSID−III | | | | | | | | | | | | |
| Bangladesh | RDNS (22) | DMC | 1115 | 548 | 8.0 | 0.75 (0.54, 1.04) | 0.24 | 0.23 | 439 | 203 | 19.2 | 0.94 (0.67, 1.32) | 0.16 | 0.16 |
| Bangladesh | WASH−B (23) | EASQ | 605 | 1614 | 7.4 | 0.67 (0.44, 1.00) | 0.15 | 0.15 | 469 | 1664 | 13.6 | 0.98 (0.75, 1.29) | 0.25 | 0.23 |
| Burkina Faso | iLiNS−Zinc (24) | DMC | 473 | 199 | 6.5 | 0.58 (0.34, 1.00) | 0.09 | 0.09 | 272 | 175 | 25.1 | 0.56 (0.37, 0.83) | 0.11 | 0.12 |
| Ghana | GHANA (25) | | | | | | | | | | | | | |
| Ghana | iLiNS−DYADG (26) | KDI | 191 | 377 | 6.6 | 1.18 (0.64, 2.19) | 0.07 | 0.07 | 111 | 224 | 14.7 | 1.10 (0.65, 1.87) | 0.07 | 0.08 |
| Haiti | HAITI (27) | | | | | | | | | | | | | |
| Kenya | WASH−B (28) | EASQ | 809 | 2682 | 7.2 | 1.04 (0.78, 1.40) | 0.29 | 0.28 | 552 | 2060 | 14.4 | 0.76 (0.58, 0.99) | 0.25 | 0.23 |
| Madagascar | MAHAY (29) | ASQI | 843 | 839 | 9.3 | 0.78 (0.41, 1.46) | 0.06 | 0.07 | 770 | 764 | 11.8 | 1.03 (0.64, 1.65) | 0.08 | 0.09 |
| Malawi | iLiNS−DYADM (30) | KDI | 136 | 287 | 7.7 | 0.96 (0.47, 1.97) | 0.05 | 0.05 | 78 | 149 | 16.1 | 0.72 (0.35, 1.46) | 0.04 | 0.04 |
| Malawi | iLiNS−DOSE (31) | KDI | 376 | 135 | 8.1 | 1.08 (0.56, 2.07) | 0.06 | 0.06 | 269 | 86 | 11.6 | 1.06 (0.54, 2.05) | 0.04 | 0.05 |
| Mali | PROMIS CS (32) | DMC | | | | | | | | | | | | |
| Zimbabwe | SHINE (HIV−) (33) | MDAT | | | | | | | | | | | | |
| Zimbabwe | SHINE (HIV+) (34) | MDAT | | | | | | | | | | | | |
| | | | **4548** | **6681** | | **$I^2$ = 0.07, $Tau^2$ = 0.00** | | | **2960** | **5325** | | **$I^2$ = 0.17, $Tau^2$ = 0.01** | | |
| **Fixed** | | | | | | **0.85 (0.72, 0.99)** | | | | | | **0.86 (0.75, 0.99)** | | |
| **Random** | | | | | | **0.84 (0.72, 1.00)** | | | | | | **0.86 (0.74, 1.00)** | | |

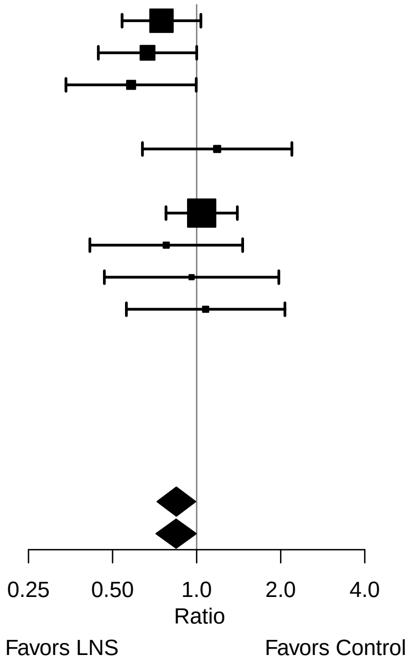

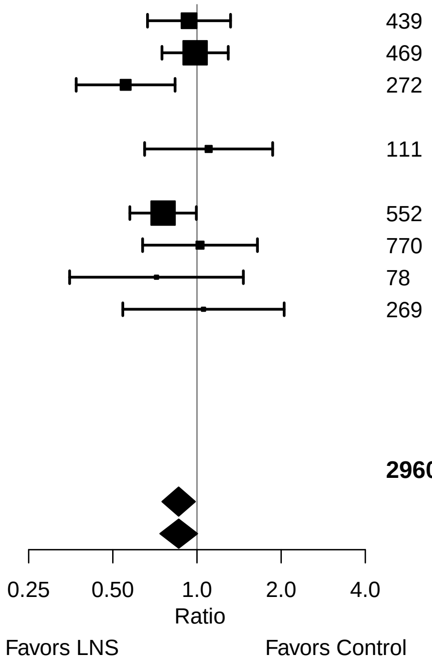

#### Supplemental figure 6H: Motor lowest decile prevalence ratio

### 6H6: Stratified by Season at the time of assessment

**P−for−interaction = 0.839**
**Ratio of PRs = 1.02 (0.83, 1.26)**

| Country | Trial | Tool | Dry LNS N | Dry Control N | Dry Control Prevalence | Dry PR (95% CI) | Dry Fixed W | Dry Random W | Rainy LNS N | Rainy Control N | Rainy Control Prevalence | Rainy PR (95% CI) | Rainy Fixed W | Rainy Random W |
|---|---|---|---|---|---|---|---|---|---|---|---|---|---|---|
| Bangladesh | JiVitA−4 (21) | BSID−III | | | | | | | | | | | | |
| Bangladesh | RDNS (22) | DMC | 698 | 326 | 11.7 | 0.87 (0.62, 1.22) | 0.14 | 0.15 | 858 | 427 | 10.8 | 0.82 (0.58, 1.17) | 0.20 | 0.20 |
| Bangladesh | WASH−B (23) | EASQ | 608 | 1896 | 10.0 | 0.91 (0.70, 1.18) | 0.23 | 0.19 | 466 | 1383 | 11.4 | 0.71 (0.50, 1.01) | 0.20 | 0.20 |
| Burkina Faso | iLiNS−Zinc (24) | DMC | 514 | 274 | 16.8 | 0.52 (0.37, 0.74) | 0.13 | 0.14 | 232 | 101 | 10.9 | 0.44 (0.21, 0.89) | 0.05 | 0.05 |
| Ghana | GHANA (25) | | | | | | | | | | | | | |
| Ghana | iLiNS−DYADG (26) | KDI | 177 | 356 | 9.0 | 1.19 (0.70, 2.05) | 0.06 | 0.08 | 125 | 245 | 10.6 | 1.06 (0.57, 1.95) | 0.07 | 0.07 |
| Haiti | HAITI (27) | | | | | | | | | | | | | |
| Kenya | WASH−B (28) | EASQ | 315 | 1059 | 17.4 | 0.71 (0.54, 0.93) | 0.22 | 0.19 | 1046 | 3682 | 8.3 | 0.93 (0.70, 1.23) | 0.32 | 0.32 |
| Madagascar | MAHAY (29) | ASQI | | | | | | | | | | | | |
| Malawi | iLiNS−DYADM (30) | KDI | 153 | 308 | 10.1 | 0.84 (0.46, 1.57) | 0.04 | 0.06 | 61 | 127 | 11.8 | 0.83 (0.34, 2.04) | 0.03 | 0.03 |
| Malawi | iLiNS−DOSE (31) | KDI | 395 | 118 | 10.2 | 1.07 (0.58, 1.96) | 0.04 | 0.06 | 244 | 100 | 9.0 | 1.00 (0.48, 2.10) | 0.05 | 0.05 |
| Mali | PROMIS CS (32) | DMC | | | | | | | | | | | | |
| Zimbabwe | SHINE (HIV−) (33) | MDAT | 603 | 568 | 10.9 | 0.84 (0.58, 1.19) | 0.13 | 0.14 | 238 | 231 | 9.1 | 1.20 (0.70, 2.07) | 0.08 | 0.08 |
| Zimbabwe | SHINE (HIV+) (34) | MDAT | | | | | | | | | | | | |
| | | | **3463** | **4905** | | **I² = 0.35, Tau² = 0.02** | | | **3270** | **6296** | | **I² = 0.00, Tau² = 0.00** | | |
| **Fixed** | | | | | | **0.80 (0.71, 0.91)** | | | | | | **0.85 (0.73, 1.00)** | | |
| **Random** | | | | | | **0.81 (0.69, 0.96)** | | | | | | **0.85 (0.73, 1.00)** | | |

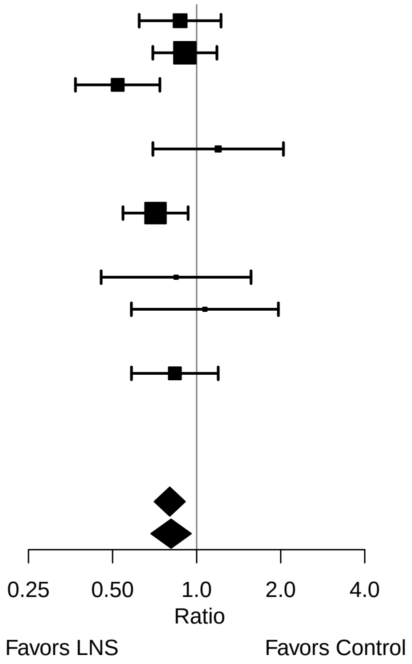

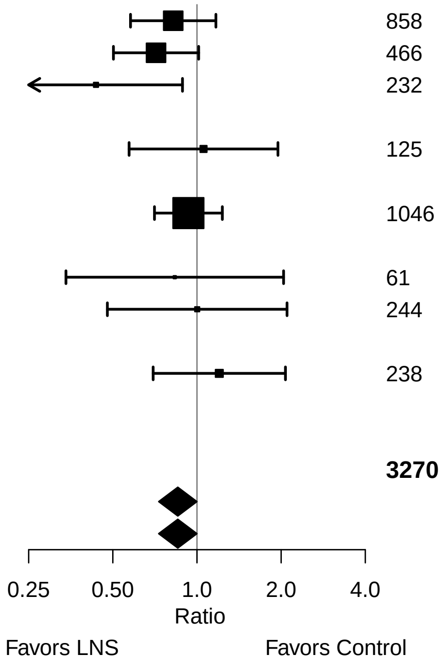

### Supplemental figure 6I: Motor lowest decile prevalence difference

#### 6I1: Stratified by Household socio-economic status

**P−for−interaction = 0.325**
**Difference in PDs = −0.01 (−0.03, 0.01)**

| Country | Trial | Tool | At least median: LNS N | Control N | Control Prevalence | PD (95% CI) | Fixed W | Random W | Less than median: LNS N | Control N | Control Prevalence | PD (95% CI) | Fixed W | Random W |
|---|---|---|---|---|---|---|---|---|---|---|---|---|---|---|
| Bangladesh | JiVitA−4 (21) | BSID−III | | | | | | | | | | | | |
| Bangladesh | RDNS (22) | DMC | 783 | 381 | 7.1 | 0.01 (−0.03, 0.04) | 0.12 | 0.13 | 773 | 372 | 15.3 | −0.04 (−0.08, 0.00) | 0.14 | 0.14 |
| Bangladesh | WASH−B (23) | EASQ | 546 | 1663 | 7.6 | −0.02 (−0.04, 0.00) | 0.27 | 0.19 | 528 | 1616 | 13.6 | −0.02 (−0.06, 0.01) | 0.17 | 0.17 |
| Burkina Faso | iLiNS−Zinc (24) | DMC | 459 | 163 | 16.0 | −0.10 (−0.15, −0.04) | 0.04 | 0.05 | 287 | 212 | 14.6 | −0.05 (−0.11, 0.01) | 0.05 | 0.05 |
| Ghana | GHANA (25) | | | | | | | | | | | | | |
| Ghana | iLiNS−DYADG (26) | KDI | 143 | 311 | 10.0 | 0.00 (−0.06, 0.06) | 0.03 | 0.05 | 159 | 288 | 9.0 | 0.03 (−0.03, 0.09) | 0.05 | 0.05 |
| Haiti | HAITI (27) | | | | | | | | | | | | | |
| Kenya | WASH−B (28) | EASQ | 778 | 2706 | 9.3 | −0.02 (−0.04, 0.01) | 0.26 | 0.19 | 582 | 2036 | 11.6 | −0.01 (−0.04, 0.01) | 0.26 | 0.26 |
| Madagascar | MAHAY (29) | ASQI | 741 | 845 | 6.2 | −0.01 (−0.04, 0.03) | 0.10 | 0.12 | 823 | 724 | 15.7 | −0.03 (−0.09, 0.04) | 0.04 | 0.04 |
| Malawi | iLiNS−DYADM (30) | KDI | 118 | 227 | 8.8 | 0.00 (−0.07, 0.06) | 0.03 | 0.05 | 96 | 208 | 12.5 | −0.03 (−0.11, 0.05) | 0.03 | 0.03 |
| Malawi | iLiNS−DOSE (31) | KDI | 278 | 94 | 6.4 | 0.02 (−0.04, 0.09) | 0.03 | 0.04 | 271 | 96 | 12.5 | −0.02 (−0.09, 0.05) | 0.04 | 0.04 |
| Mali | PROMIS CS (32) | DMC | 479 | 445 | 11.0 | −0.02 (−0.07, 0.03) | 0.05 | 0.07 | 423 | 476 | 11.1 | −0.02 (−0.06, 0.02) | 0.11 | 0.11 |
| Zimbabwe | SHINE (HIV−) (33) | MDAT | 406 | 359 | 10.6 | −0.01 (−0.05, 0.04) | 0.05 | 0.08 | 389 | 391 | 11.3 | −0.01 (−0.06, 0.03) | 0.10 | 0.10 |
| Zimbabwe | SHINE (HIV+) (34) | MDAT | 91 | 78 | 11.5 | −0.04 (−0.12, 0.04) | 0.02 | 0.03 | 74 | 67 | 13.4 | −0.04 (−0.14, 0.06) | 0.02 | 0.02 |
| | | | **4822** | **7272** | | **I² = 0.20, Tau² = 0.00** | | | **4405** | **6486** | | **I² = 0.00, Tau² = 0.00** | | |
| **Fixed** | | | | | | **−0.01 (−0.02, 0.00)** | | | | | | **−0.02 (−0.03, −0.01)** | | |
| **Random** | | | | | | **−0.01 (−0.03, 0.00)** | | | | | | **−0.02 (−0.03, −0.01)** | | |

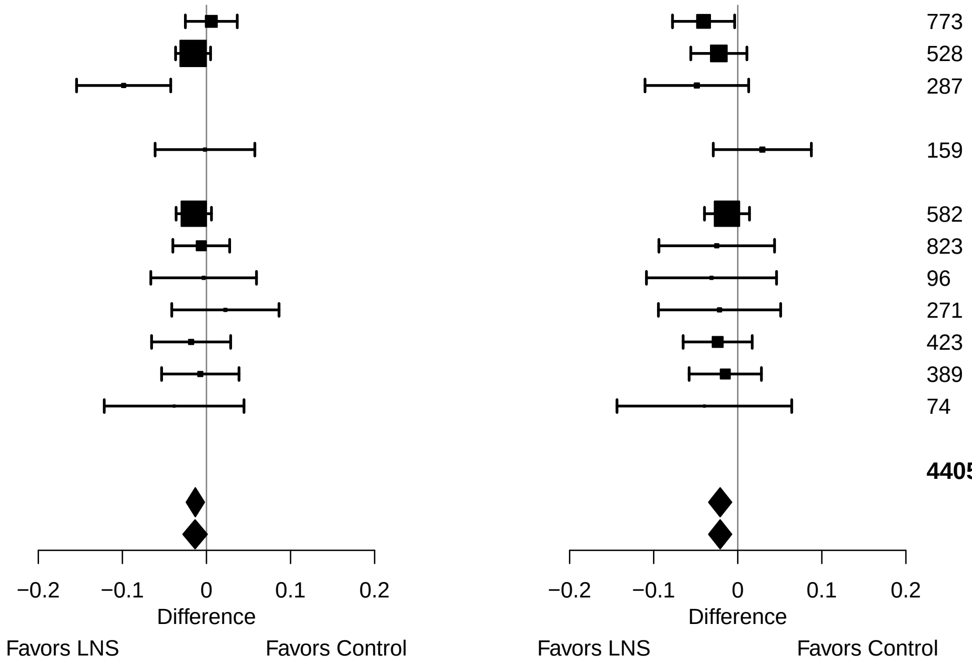

#### Supplemental figure 6I: Motor lowest decile prevalence difference

#### 6I2: Stratified by Household food insecurity

**P−for−interaction = 0.291**
**Difference in PDs = −0.01 (−0.04, 0.01)**

| Country | Trial | Tool | LNS N | Control N | Control Prevalence | Mild to secure PD (95% CI) | Fixed W | Random W | LNS N | Control N | Control Prevalence | Moderate to severe PD (95% CI) | Fixed W | Random W |
|---|---|---|---|---|---|---|---|---|---|---|---|---|---|---|
| Bangladesh | JiVitA−4 (21) | BSID−III | | | | | | | | | | | | |
| Bangladesh | RDNS (22) | DMC | 987 | 456 | 7.2 | 0.01 (−0.02, 0.04) | 0.14 | 0.15 | 569 | 297 | 17.2 | −0.06 (−0.11, −0.01) | 0.19 | 0.19 |
| Bangladesh | WASH−B (23) | EASQ | 853 | 2550 | 10.0 | −0.02 (−0.04, 0.00) | 0.31 | 0.18 | 221 | 729 | 12.8 | −0.01 (−0.06, 0.04) | 0.20 | 0.20 |
| Burkina Faso | iLiNS−Zinc (24) | DMC | 389 | 162 | 15.4 | −0.09 (−0.13, −0.05) | 0.07 | 0.12 | 357 | 213 | 15.0 | −0.07 (−0.13, 0.00) | 0.12 | 0.12 |
| Ghana | GHANA (25) | | | | | | | | | | | | | |
| Ghana | iLiNS−DYADG (26) | KDI | 217 | 412 | 9.2 | 0.00 (−0.05, 0.05) | 0.05 | 0.10 | 84 | 186 | 10.2 | 0.05 (−0.03, 0.14) | 0.07 | 0.07 |
| Haiti | HAITI (27) | | | | | | | | | | | | | |
| Kenya | WASH−B (28) | EASQ | 1214 | 4244 | 10.0 | −0.01 (−0.03, 0.01) | 0.29 | 0.18 | 146 | 487 | 13.6 | −0.03 (−0.09, 0.02) | 0.14 | 0.14 |
| Madagascar | MAHAY (29) | ASQI | 940 | 892 | 9.0 | −0.01 (−0.05, 0.04) | 0.05 | 0.11 | 336 | 389 | 15.4 | −0.01 (−0.10, 0.08) | 0.06 | 0.06 |
| Malawi | iLiNS−DYADM (30) | KDI | 67 | 116 | 11.2 | 0.01 (−0.09, 0.10) | 0.01 | 0.04 | 146 | 314 | 10.2 | −0.03 (−0.08, 0.03) | 0.14 | 0.14 |
| Malawi | iLiNS−DOSE (31) | KDI | | | | | | | | | | | | |
| Mali | PROMIS CS (32) | DMC | | | | | | | | | | | | |
| Zimbabwe | SHINE (HIV−) (33) | MDAT | 631 | 586 | 10.4 | 0.00 (−0.04, 0.03) | 0.08 | 0.12 | 140 | 154 | 13.0 | −0.04 (−0.11, 0.04) | 0.08 | 0.08 |
| Zimbabwe | SHINE (HIV+) (34) | MDAT | | | | | | | | | | | | |
| | | | **5298** | **9418** | | **I² = 0.58, Tau² = 0.00** | | | **1999** | **2769** | | **I² = 0.02, Tau² = 0.00** | | |
| **Fixed** | | | | | | **−0.02 (−0.03, 0.00)** | | | | | | **−0.03 (−0.05, −0.01)** | | |
| **Random** | | | | | | **−0.02 (−0.04, 0.01)** | | | | | | **−0.03 (−0.05, −0.01)** | | |

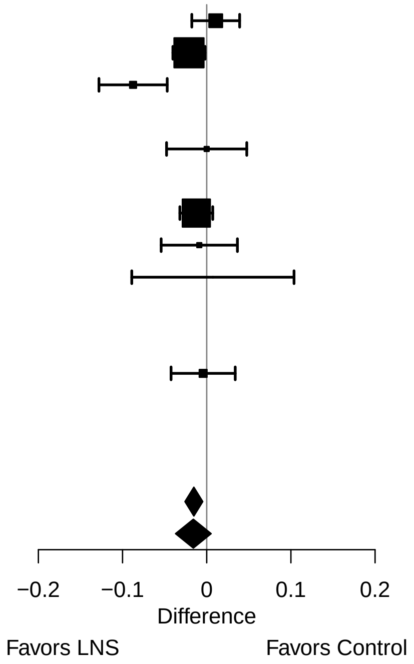

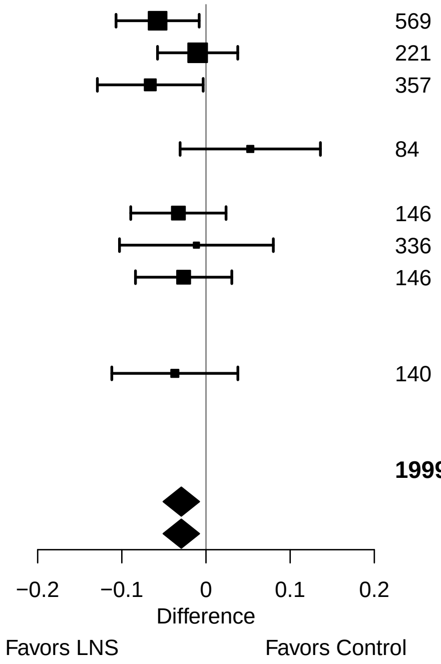

#### Supplemental figure 6I: Motor lowest decile prevalence difference

#### 6I3: Stratified by Household source water quality

**P−for−interaction = 0.795**
**Difference in PDs = 0.00 (−0.02, 0.03)**

| Country | Trial | Tool | LNS N | Control N | Control Prevalence | Improved PD (95% CI) | Fixed W | Random W | LNS N | Control N | Control Prevalence | Unimproved PD (95% CI) | Fixed W | Random W |
|---|---|---|---|---|---|---|---|---|---|---|---|---|---|---|
| Bangladesh | JiVitA−4 (21) | BSID−III | | | | | | | | | | | | |
| Bangladesh | RDNS (22) | DMC | | | | | | | | | | | | |
| Bangladesh | WASH−B (23) | EASQ | 460 | 981 | 11.6 | −0.02 (−0.05, 0.02) | 0.29 | 0.29 | 67 | 117 | 12.0 | −0.05 (−0.13, 0.04) | 0.06 | 0.08 |
| Burkina Faso | iLiNS−Zinc (24) | DMC | 220 | 82 | 12.2 | −0.07 (−0.14, 0.00) | 0.07 | 0.07 | 526 | 293 | 16.0 | −0.07 (−0.11, −0.04) | 0.32 | 0.26 |
| Ghana | GHANA (25) | | | | | | | | | | | | | |
| Ghana | iLiNS−DYADG (26) | KDI | | | | | | | | | | | | |
| Haiti | HAITI (27) | | | | | | | | | | | | | |
| Kenya | WASH−B (28) | EASQ | 413 | 1419 | 10.2 | 0.00 (−0.04, 0.04) | 0.24 | 0.24 | 236 | 649 | 9.2 | −0.01 (−0.05, 0.03) | 0.29 | 0.25 |
| Madagascar | MAHAY (29) | ASQI | 405 | 430 | 14.0 | −0.02 (−0.10, 0.07) | 0.05 | 0.05 | 1134 | 1133 | 9.4 | −0.01 (−0.06, 0.04) | 0.18 | 0.19 |
| Malawi | iLiNS−DYADM (30) | KDI | | | | | | | | | | | | |
| Malawi | iLiNS−DOSE (31) | KDI | | | | | | | | | | | | |
| Mali | PROMIS CS (32) | DMC | 514 | 539 | 10.2 | −0.03 (−0.07, 0.01) | 0.22 | 0.22 | 347 | 368 | 12.8 | −0.01 (−0.08, 0.05) | 0.10 | 0.13 |
| Zimbabwe | SHINE (HIV−) (33) | MDAT | 228 | 220 | 9.5 | −0.01 (−0.06, 0.04) | 0.13 | 0.13 | 131 | 122 | 9.8 | 0.02 (−0.07, 0.10) | 0.06 | 0.08 |
| Zimbabwe | SHINE (HIV+) (34) | MDAT | | | | | | | | | | | | |
| | | | **2240** | **3671** | | **I² = 0.00, Tau² = 0.00** | | | **2441** | **2682** | | **I² = 0.45, Tau² = 0.00** | | |
| **Fixed** | | | | | | **−0.02 (−0.04, 0.00)** | | | | | | **−0.03 (−0.05, −0.01)** | | |
| **Random** | | | | | | **−0.02 (−0.04, 0.00)** | | | | | | **−0.03 (−0.05, 0.00)** | | |

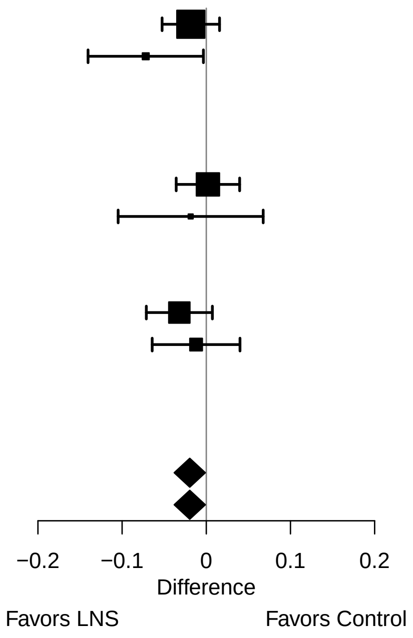

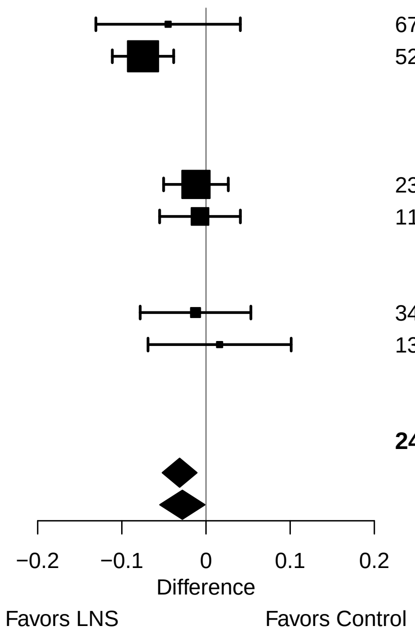

#### Supplemental figure 6I: Motor lowest decile prevalence difference

#### 6I4: Stratified by Household sanitation

**P−for−interaction = 0.308**
**Difference in PDs = 0.02 (−0.02, 0.05)**

| Country | Trial | Tool | Improved LNS N | Improved Control N | Improved Control Prevalence | Improved PD (95% CI) | Improved Fixed W | Improved Random W | Unimproved LNS N | Unimproved Control N | Unimproved Control Prevalence | Unimproved PD (95% CI) | Unimproved Fixed W | Unimproved Random W |
|---|---|---|---|---|---|---|---|---|---|---|---|---|---|---|
| Bangladesh | JiVitA−4 (21) | BSID−III | | | | | | | | | | | | |
| Bangladesh | RDNS (22) | DMC | 1103 | 531 | 10.4 | −0.02 (−0.05, 0.01) | 0.50 | 0.50 | 451 | 222 | 13.1 | −0.01 (−0.07, 0.05) | 0.15 | 0.15 |
| Bangladesh | WASH−B (23) | EASQ | | | | | | | | | | | | |
| Burkina Faso | iLiNS−Zinc (24) | DMC | | | | | | | | | | | | |
| Ghana | GHANA (25) | | | | | | | | | | | | | |
| Ghana | iLiNS−DYADG (26) | KDI | | | | | | | | | | | | |
| Haiti | HAITI (27) | | | | | | | | | | | | | |
| Kenya | WASH−B (28) | EASQ | 94 | 337 | 9.2 | −0.02 (−0.08, 0.05) | 0.11 | 0.11 | 555 | 1732 | 10.0 | 0.00 (−0.03, 0.03) | 0.55 | 0.55 |
| Madagascar | MAHAY (29) | ASQI | | | | | | | | | | | | |
| Malawi | iLiNS−DYADM (30) | KDI | | | | | | | | | | | | |
| Malawi | iLiNS−DOSE (31) | KDI | | | | | | | | | | | | |
| Mali | PROMIS CS (32) | DMC | 665 | 658 | 10.8 | −0.02 (−0.06, 0.02) | 0.29 | 0.29 | 210 | 241 | 10.8 | −0.01 (−0.06, 0.05) | 0.16 | 0.16 |
| Zimbabwe | SHINE (HIV−) (33) | MDAT | 133 | 108 | 10.2 | −0.03 (−0.10, 0.03) | 0.10 | 0.10 | 225 | 235 | 9.4 | 0.02 (−0.04, 0.08) | 0.14 | 0.14 |
| Zimbabwe | SHINE (HIV+) (34) | MDAT | | | | | | | | | | | | |
| | | | **1995** | **1634** | | **I² = 0.00, Tau² = 0.00** | | | **1441** | **2430** | | **I² = 0.00, Tau² = 0.00** | | |
| **Fixed** | | | | | | **−0.02 (−0.04, 0.00)** | | | | | | **0.00 (−0.02, 0.02)** | | |
| **Random** | | | | | | **−0.02 (−0.04, 0.00)** | | | | | | **0.00 (−0.02, 0.02)** | | |

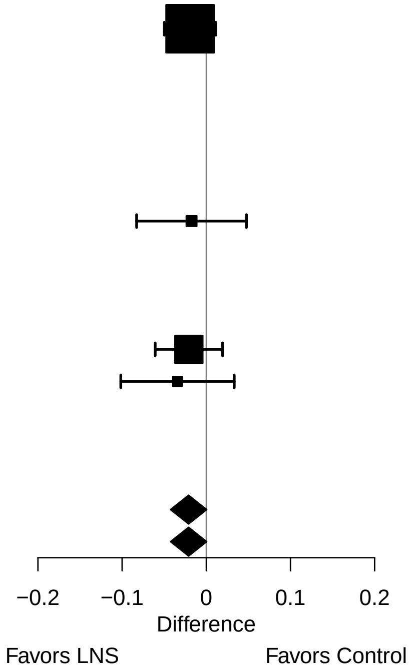

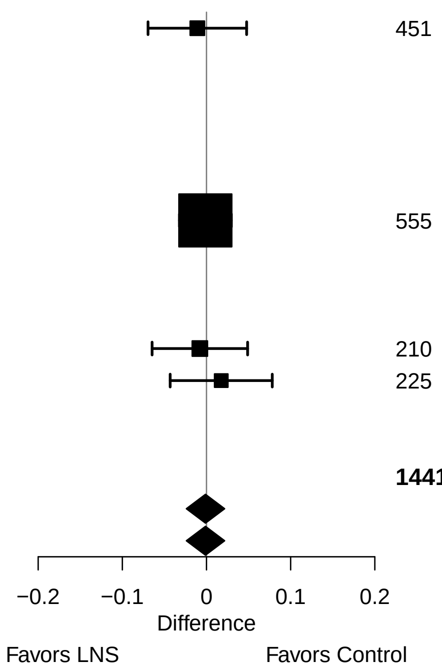

#### Supplemental figure 6I: Motor lowest decile prevalence difference

##### 6I5: Stratified by Home environment

**P−for−interaction = 0.562**
**Difference in PDs = −0.01 (−0.03, 0.01)**

| Country | Trial | Tool | LNS N | Control N | Control Prevalence | At least median PD (95% CI) | Fixed W | Random W | LNS N | Control N | Control Prevalence | Less than median PD (95% CI) | Fixed W | Random W |
|---|---|---|---|---|---|---|---|---|---|---|---|---|---|---|
| Bangladesh | JiVitA−4 (21) | BSID−III | | | | | | | | | | | | |
| Bangladesh | RDNS (22) | DMC | 1115 | 548 | 8.0 | −0.02 (−0.04, 0.00) | 0.21 | 0.21 | 439 | 203 | 19.2 | −0.01 (−0.08, 0.05) | 0.09 | 0.11 |
| Bangladesh | WASH−B (23) | EASQ | 605 | 1614 | 7.4 | −0.02 (−0.05, 0.00) | 0.23 | 0.23 | 469 | 1664 | 13.6 | 0.00 (−0.04, 0.03) | 0.27 | 0.24 |
| Burkina Faso | iLiNS−Zinc (24) | DMC | 473 | 199 | 6.5 | −0.03 (−0.06, 0.00) | 0.14 | 0.14 | 272 | 175 | 25.1 | −0.11 (−0.20, −0.03) | 0.05 | 0.07 |
| Ghana | GHANA (25) | | | | | | | | | | | | | |
| Ghana | iLiNS−DYADG (26) | KDI | 191 | 377 | 6.6 | 0.01 (−0.03, 0.06) | 0.06 | 0.06 | 111 | 224 | 14.7 | 0.01 (−0.07, 0.10) | 0.05 | 0.07 |
| Haiti | HAITI (27) | | | | | | | | | | | | | |
| Kenya | WASH−B (28) | EASQ | 809 | 2682 | 7.2 | 0.00 (−0.02, 0.03) | 0.24 | 0.24 | 552 | 2060 | 14.4 | −0.03 (−0.07, 0.00) | 0.33 | 0.27 |
| Madagascar | MAHAY (29) | ASQI | 843 | 839 | 9.3 | −0.02 (−0.07, 0.03) | 0.05 | 0.05 | 770 | 764 | 11.8 | 0.00 (−0.05, 0.06) | 0.11 | 0.13 |
| Malawi | iLiNS−DYADM (30) | KDI | 136 | 287 | 7.7 | 0.00 (−0.06, 0.05) | 0.04 | 0.04 | 78 | 149 | 16.1 | −0.05 (−0.14, 0.05) | 0.04 | 0.05 |
| Malawi | iLiNS−DOSE (31) | KDI | 376 | 135 | 8.1 | 0.01 (−0.05, 0.06) | 0.04 | 0.04 | 269 | 86 | 11.6 | 0.01 (−0.07, 0.09) | 0.06 | 0.07 |
| Mali | PROMIS CS (32) | DMC | | | | | | | | | | | | |
| Zimbabwe | SHINE (HIV−) (33) | MDAT | | | | | | | | | | | | |
| Zimbabwe | SHINE (HIV+) (34) | MDAT | | | | | | | | | | | | |
| | | | **4548** | **6681** | | **I² = 0.00, Tau² = 0.00** | | | **2960** | **5325** | | **I² = 0.17, Tau² = 0.00** | | |
| **Fixed** | | | | | | **−0.01 (−0.02, 0.00)** | | | | | | **−0.02 (−0.04, 0.00)** | | |
| **Random** | | | | | | **−0.01 (−0.02, 0.00)** | | | | | | **−0.02 (−0.04, 0.00)** | | |

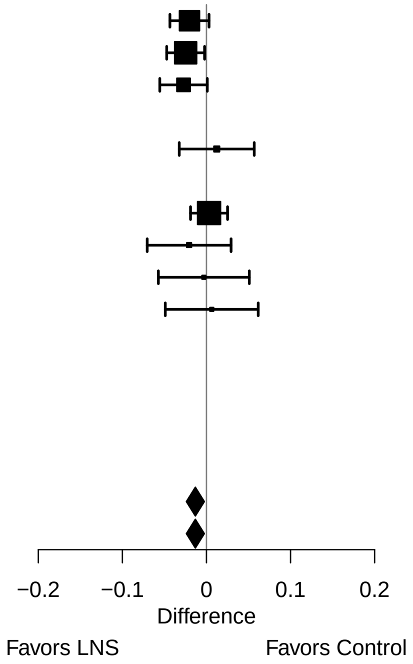

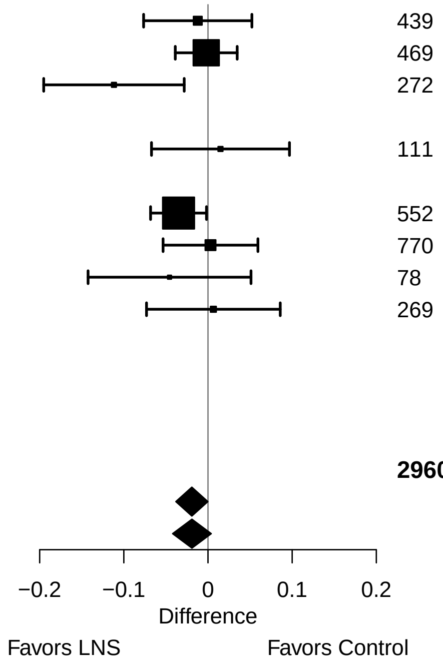

#### Supplemental figure 6I: Motor lowest decile prevalence difference

#### 6I6: Stratified by Season at the time of assessment

P−for−interaction = 0.586
Difference in PDs = 0.01 (−0.02, 0.03)

| Country | Trial | Tool | LNS N | Control N | Control Prevalence | Dry PD (95% CI) | Fixed W | Random W | LNS N | Control N | Control Prevalence | Rainy PD (95% CI) | Fixed W | Random W |
|---|---|---|---|---|---|---|---|---|---|---|---|---|---|---|
| Bangladesh | JiVitA−4 (21) | BSID−III | | | | | | | | | | | | |
| Bangladesh | RDNS (22) | DMC | 698 | 326 | 11.7 | −0.01 (−0.05, 0.02) | 0.14 | 0.15 | 858 | 427 | 10.8 | −0.02 (−0.05, 0.02) | 0.16 | 0.16 |
| Bangladesh | WASH−B (23) | EASQ | 608 | 1896 | 10.0 | −0.01 (−0.03, 0.02) | 0.33 | 0.20 | 466 | 1383 | 11.4 | −0.03 (−0.06, 0.00) | 0.21 | 0.21 |
| Burkina Faso | iLiNS−Zinc (24) | DMC | 514 | 274 | 16.8 | −0.08 (−0.13, −0.03) | 0.09 | 0.11 | 232 | 101 | 10.9 | −0.06 (−0.12, −0.01) | 0.06 | 0.06 |
| Ghana | GHANA (25) | | | | | | | | | | | | | |
| Ghana | iLiNS−DYADG (26) | KDI | 177 | 356 | 9.0 | 0.02 (−0.04, 0.07) | 0.07 | 0.10 | 125 | 245 | 10.6 | 0.01 (−0.06, 0.07) | 0.04 | 0.04 |
| Haiti | HAITI (27) | | | | | | | | | | | | | |
| Kenya | WASH−B (28) | EASQ | 315 | 1059 | 17.4 | −0.05 (−0.09, −0.01) | 0.11 | 0.13 | 1046 | 3682 | 8.3 | −0.01 (−0.03, 0.02) | 0.40 | 0.40 |
| Madagascar | MAHAY (29) | ASQI | | | | | | | | | | | | |
| Malawi | iLiNS−DYADM (30) | KDI | 153 | 308 | 10.1 | −0.02 (−0.07, 0.04) | 0.06 | 0.09 | 61 | 127 | 11.8 | −0.02 (−0.12, 0.08) | 0.02 | 0.02 |
| Malawi | iLiNS−DOSE (31) | KDI | 395 | 118 | 10.2 | 0.01 (−0.06, 0.07) | 0.05 | 0.08 | 244 | 100 | 9.0 | 0.00 (−0.07, 0.07) | 0.04 | 0.04 |
| Mali | PROMIS CS (32) | DMC | | | | | | | | | | | | |
| Zimbabwe | SHINE (HIV−) (33) | MDAT | 603 | 568 | 10.9 | −0.02 (−0.05, 0.02) | 0.15 | 0.15 | 238 | 231 | 9.1 | 0.02 (−0.04, 0.07) | 0.07 | 0.07 |
| Zimbabwe | SHINE (HIV+) (34) | MDAT | | | | | | | | | | | | |
| | | | **3463** | **4905** | | **I² = 0.40, Tau² = 0.00** | | | **3270** | **6296** | | **I² = 0.00, Tau² = 0.00** | | |
| **Fixed** | | | | | | **−0.02 (−0.03, −0.01)** | | | | | | **−0.01 (−0.03, 0.00)** | | |
| **Random** | | | | | | **−0.02 (−0.04, 0.00)** | | | | | | **−0.01 (−0.03, 0.00)** | | |

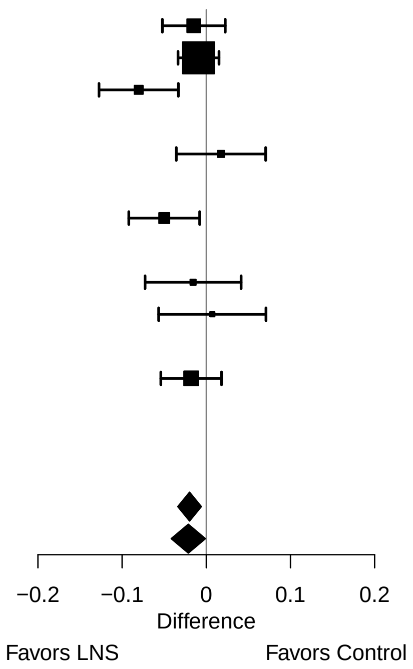

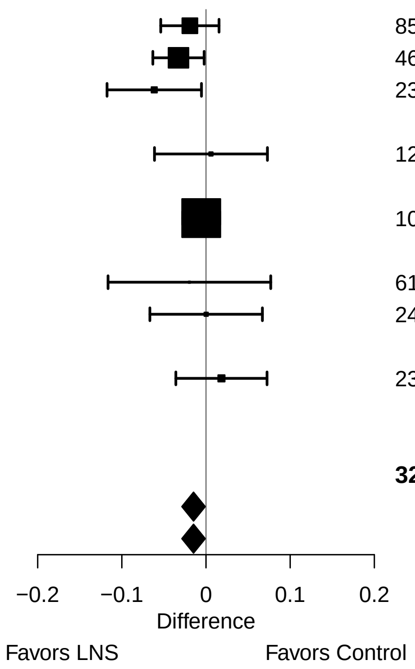

### Supplemental figure 6J: Mean difference in gross motor z-score

#### 6J1: Stratified by Household socio-economic status

**P−for−interaction = 0.091**
**Difference in MDs = 0.05 (−0.01, 0.10)**

| | | | At least median | | | | | | Less than median | | | | | |
|---|---|---|---|---|---|---|---|---|---|---|---|---|---|---|
| Country | Trial | Tool | LNS N | Control N | Control Mean | MD (95% CI) | Fixed W | Random W | LNS N | Control N | Control Mean | MD (95% CI) | Fixed W | Random W |
| Bangladesh | JiVitA−4 (21) | BSID−III | 236 | 77 | 0.26 | −0.15 (−0.35, 0.05) | 0.04 | 0.06 | 208 | 66 | −0.12 | −0.04 (−0.27, 0.19) | 0.03 | 0.03 |
| Bangladesh | RDNS (22) | DMC | 788 | 382 | 0.02 | 0.09 (0.00, 0.18) | 0.18 | 0.15 | 782 | 376 | −0.09 | 0.08 (−0.01, 0.18) | 0.19 | 0.19 |
| Bangladesh | WASH−B (23) | EASQ | 546 | 1663 | 0.12 | 0.05 (−0.04, 0.14) | 0.18 | 0.15 | 528 | 1616 | −0.17 | 0.16 (0.07, 0.26) | 0.20 | 0.20 |
| Burkina Faso | iLiNS−Zinc (24) | DMC | | | | | | | | | | | | |
| Ghana | GHANA (25) | | | | | | | | | | | | | |
| Ghana | iLiNS−DYADG (26) | KDI | 143 | 311 | 0.02 | 0.09 (−0.12, 0.29) | 0.03 | 0.06 | 159 | 288 | −0.04 | −0.05 (−0.24, 0.14) | 0.05 | 0.05 |
| Haiti | HAITI (27) | | | | | | | | | | | | | |
| Kenya | WASH−B (28) | EASQ | 778 | 2706 | 0.07 | −0.03 (−0.10, 0.04) | 0.29 | 0.16 | 582 | 2036 | −0.10 | 0.07 (−0.04, 0.17) | 0.16 | 0.16 |
| Madagascar | MAHAY (29) | ASQI | 741 | 845 | 0.08 | 0.07 (−0.09, 0.23) | 0.06 | 0.08 | 823 | 724 | −0.18 | 0.11 (−0.08, 0.29) | 0.05 | 0.05 |
| Malawi | iLiNS−DYADM (30) | KDI | 118 | 227 | 0.02 | −0.05 (−0.25, 0.14) | 0.04 | 0.07 | 96 | 208 | 0.01 | −0.01 (−0.27, 0.26) | 0.02 | 0.02 |
| Malawi | iLiNS−DOSE (31) | KDI | 278 | 94 | 0.16 | −0.05 (−0.27, 0.17) | 0.03 | 0.05 | 271 | 96 | −0.15 | 0.10 (−0.13, 0.32) | 0.03 | 0.03 |
| Mali | PROMIS CS (32) | DMC | 499 | 457 | −0.03 | 0.10 (−0.04, 0.23) | 0.08 | 0.10 | 441 | 500 | −0.06 | 0.07 (−0.04, 0.17) | 0.16 | 0.16 |
| Zimbabwe | SHINE (HIV−) (33) | MDAT | 406 | 359 | −0.01 | 0.12 (−0.03, 0.27) | 0.06 | 0.09 | 389 | 391 | −0.11 | 0.11 (−0.04, 0.26) | 0.08 | 0.08 |
| Zimbabwe | SHINE (HIV+) (34) | MDAT | 91 | 78 | −0.17 | 0.38 (0.07, 0.70) | 0.01 | 0.03 | 74 | 67 | −0.16 | 0.25 (−0.05, 0.55) | 0.02 | 0.02 |
| | | | **4624** | **7199** | | **I² = 0.41, Tau² = 0.00** | | | **4353** | **6368** | | **I² = 0.00, Tau² = 0.00** | | |
| **Fixed** | | | | | | **0.03 (0.00, 0.07)** | | | | | | **0.09 (0.05, 0.13)** | | |
| **Random** | | | | | | **0.04 (−0.02, 0.10)** | | | | | | **0.09 (0.05, 0.13)** | | |

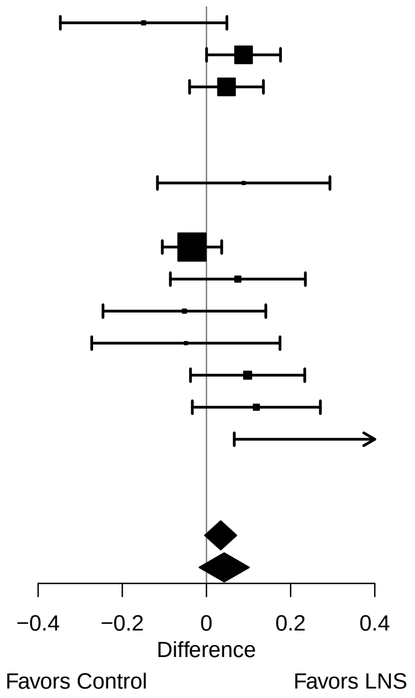

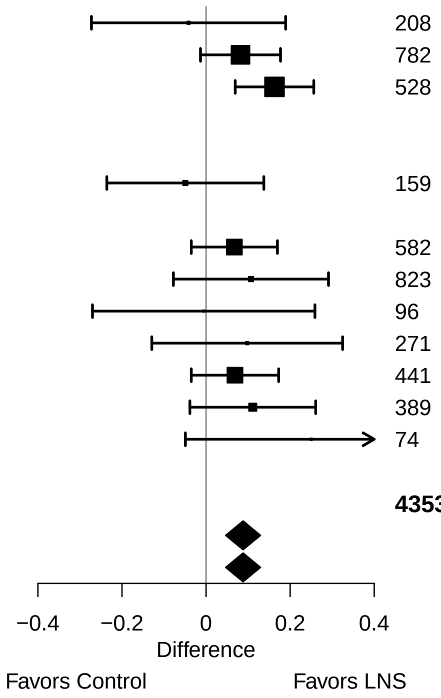

#### Supplemental figure 6J: Mean difference in gross motor z-score

#### 6J2: Stratified by Household food insecurity

**P−for−interaction = 0.951**
**Difference in MDs = 0.00 (−0.07, 0.08)**

| | | | Mild to secure | | | | | | Moderate to severe | | | | | |
|---|---|---|---|---|---|---|---|---|---|---|---|---|---|---|
| **Country** | **Trial** | **Tool** | **LNS N** | **Control N** | **Control Mean** | **MD (95% CI)** | **Fixed W** | **Random W** | **LNS N** | **Control N** | **Control Mean** | **MD (95% CI)** | **Fixed W** | **Random W** |
| Bangladesh | JiVitA−4 (21) | BSID−III | 329 | 111 | 0.12 | −0.12 (−0.29, 0.05) | 0.04 | 0.09 | 115 | 32 | −0.06 | −0.02 (−0.37, 0.33) | 0.03 | 0.03 |
| Bangladesh | RDNS (22) | DMC | 994 | 459 | 0.04 | 0.05 (−0.04, 0.14) | 0.16 | 0.14 | 576 | 299 | −0.14 | 0.14 (0.03, 0.24) | 0.33 | 0.33 |
| Bangladesh | WASH−B (23) | EASQ | 853 | 2550 | 0.01 | 0.11 (0.05, 0.18) | 0.28 | 0.15 | 221 | 729 | −0.14 | 0.05 (−0.11, 0.21) | 0.14 | 0.14 |
| Burkina Faso | iLiNS−Zinc (24) | DMC | | | | | | | | | | | | |
| Ghana | GHANA (25) | | | | | | | | | | | | | |
| Ghana | iLiNS−DYADG (26) | KDI | 217 | 412 | −0.01 | 0.04 (−0.12, 0.21) | 0.05 | 0.09 | 84 | 186 | 0.00 | −0.06 (−0.31, 0.19) | 0.05 | 0.05 |
| Haiti | HAITI (27) | | | | | | | | | | | | | |
| Kenya | WASH−B (28) | EASQ | 1214 | 4244 | 0.01 | 0.01 (−0.06, 0.08) | 0.29 | 0.15 | 146 | 487 | −0.11 | 0.01 (−0.19, 0.21) | 0.09 | 0.09 |
| Madagascar | MAHAY (29) | ASQI | 940 | 892 | 0.04 | 0.04 (−0.12, 0.21) | 0.05 | 0.09 | 336 | 389 | −0.19 | 0.08 (−0.15, 0.32) | 0.06 | 0.06 |
| Malawi | iLiNS−DYADM (30) | KDI | 67 | 116 | 0.05 | −0.13 (−0.39, 0.14) | 0.02 | 0.06 | 146 | 314 | 0.01 | 0.00 (−0.20, 0.20) | 0.09 | 0.09 |
| Malawi | iLiNS−DOSE (31) | KDI | 134 | 51 | 0.18 | −0.08 (−0.36, 0.20) | 0.02 | 0.05 | 403 | 133 | −0.08 | 0.08 (−0.12, 0.28) | 0.09 | 0.09 |
| Mali | PROMIS CS (32) | DMC | | | | | | | | | | | | |
| Zimbabwe | SHINE (HIV−) (33) | MDAT | 631 | 586 | −0.05 | 0.12 (−0.01, 0.25) | 0.08 | 0.11 | 140 | 154 | −0.09 | 0.05 (−0.14, 0.24) | 0.10 | 0.10 |
| Zimbabwe | SHINE (HIV+) (34) | MDAT | 119 | 105 | −0.18 | 0.39 (0.15, 0.63) | 0.02 | 0.07 | 41 | 37 | −0.12 | 0.13 (−0.21, 0.47) | 0.03 | 0.03 |
| | | | **5498** | **9526** | | **I² = 0.55, Tau² = 0.01** | | | **2208** | **2760** | | **I² = 0.00, Tau² = 0.00** | | |
| **Fixed** | | | | | | **0.06 (0.02, 0.09)** | | | | | | **0.07 (0.01, 0.13)** | | |
| **Random** | | | | | | **0.05 (−0.03, 0.13)** | | | | | | **0.07 (0.01, 0.13)** | | |

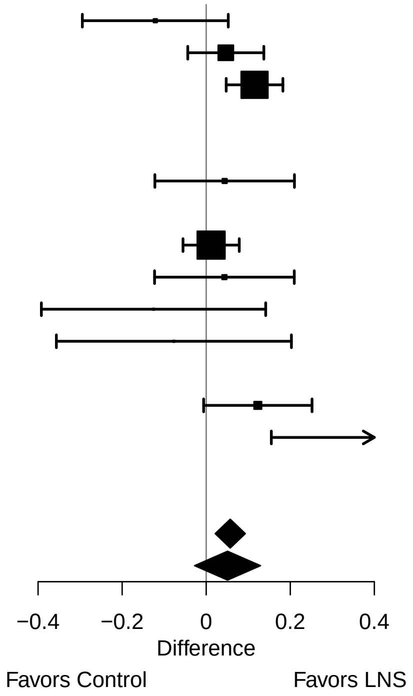

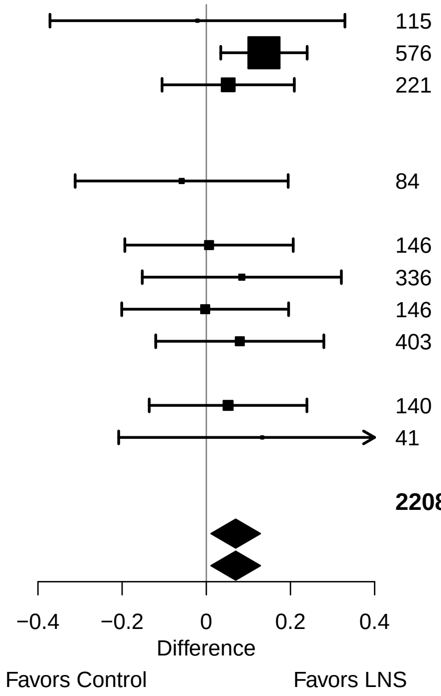

#### Supplemental figure 6J: Mean difference in gross motor z-score

#### 6J3: Stratified by Household source water quality

**P−for−interaction = 0.883**
**Difference in MDs = −0.01 (−0.12, 0.10)**

| Country | Trial | Tool | LNS N | Control N | Control Mean | Improved MD (95% CI) | Fixed W | Random W | LNS N | Control N | Control Mean | Unimproved MD (95% CI) | Fixed W | Random W |
|---|---|---|---|---|---|---|---|---|---|---|---|---|---|---|
| Bangladesh | JiVitA−4 (21) | BSID−III | | | | | | | | | | | | |
| Bangladesh | RDNS (22) | DMC | | | | | | | | | | | | |
| Bangladesh | WASH−B (23) | EASQ | 460 | 981 | −0.09 | 0.20 (0.08, 0.31) | 0.21 | 0.19 | 67 | 117 | −0.12 | 0.20 (−0.08, 0.48) | 0.09 | 0.09 |
| Burkina Faso | iLiNS−Zinc (24) | DMC | | | | | | | | | | | | |
| Ghana | GHANA (25) | | | | | | | | | | | | | |
| Ghana | iLiNS−DYADG (26) | KDI | | | | | | | | | | | | |
| Haiti | HAITI (27) | | | | | | | | | | | | | |
| Kenya | WASH−B (28) | EASQ | 413 | 1419 | −0.01 | 0.02 (−0.10, 0.14) | 0.21 | 0.19 | 236 | 649 | 0.05 | 0.04 (−0.12, 0.21) | 0.24 | 0.24 |
| Madagascar | MAHAY (29) | ASQI | 405 | 430 | −0.04 | 0.10 (−0.14, 0.35) | 0.05 | 0.06 | 1134 | 1133 | −0.04 | 0.07 (−0.10, 0.24) | 0.23 | 0.23 |
| Malawi | iLiNS−DYADM (30) | KDI | 194 | 398 | 0.03 | −0.07 (−0.23, 0.10) | 0.10 | 0.12 | 20 | 36 | −0.08 | 0.23 (−0.31, 0.77) | 0.02 | 0.02 |
| Malawi | iLiNS−DOSE (31) | KDI | 515 | 177 | 0.01 | 0.03 (−0.14, 0.20) | 0.11 | 0.12 | 44 | 15 | −0.08 | 0.08 (−0.53, 0.69) | 0.02 | 0.02 |
| Mali | PROMIS CS (32) | DMC | 539 | 561 | −0.05 | 0.15 (0.04, 0.27) | 0.22 | 0.20 | 359 | 382 | −0.04 | 0.02 (−0.14, 0.18) | 0.27 | 0.27 |
| Zimbabwe | SHINE (HIV−) (33) | MDAT | 228 | 220 | 0.09 | 0.01 (−0.19, 0.20) | 0.08 | 0.09 | 131 | 122 | −0.10 | 0.09 (−0.18, 0.36) | 0.10 | 0.10 |
| Zimbabwe | SHINE (HIV+) (34) | MDAT | 37 | 41 | −0.02 | 0.08 (−0.28, 0.44) | 0.02 | 0.03 | 25 | 25 | −0.25 | 0.12 (−0.31, 0.55) | 0.04 | 0.04 |
| | | | **2791** | **4227** | | **I² = 0.31, Tau² = 0.00** | | | **2016** | **2479** | | **I² = 0.00, Tau² = 0.00** | | |
| **Fixed** | | | | | | **0.08 (0.03, 0.14)** | | | | | | **0.07 (−0.01, 0.15)** | | |
| **Random** | | | | | | **0.08 (0.01, 0.14)** | | | | | | **0.07 (−0.01, 0.15)** | | |

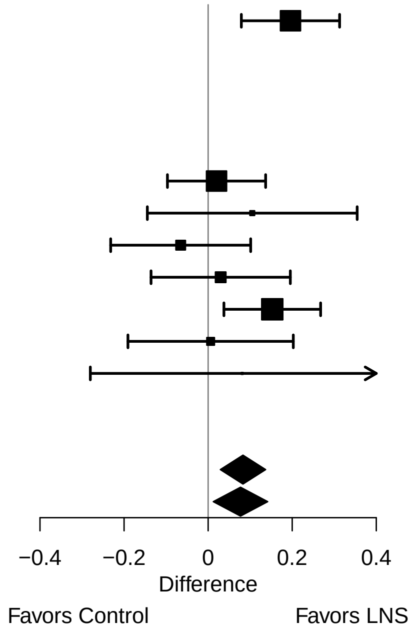

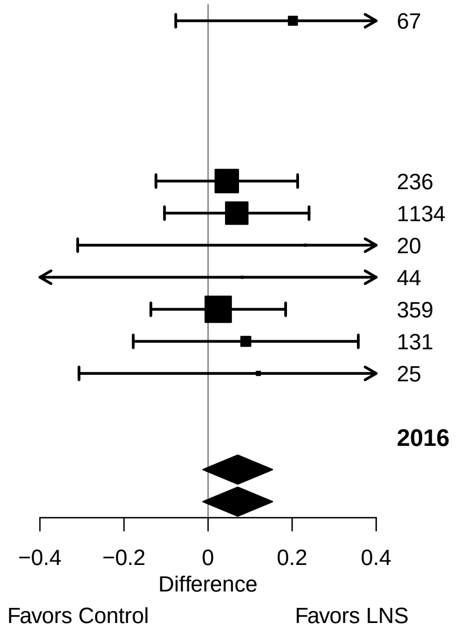

**Supplemental figure 6J: Mean difference in gross motor z-score**

#### 6J4: Stratified by Household sanitation

**P−for−interaction = 0.943**
**Difference in MDs = 0.00 (−0.10, 0.09)**

| | | | | | | Improved | | | | | | Unimproved | | |
|---|---|---|---|---|---|---|---|---|---|---|---|---|---|---|
| **Country** | **Trial** | **Tool** | **LNS N** | **Control N** | **Control Mean** | **MD (95% CI)** | **Fixed W** | **Random W** | **LNS N** | **Control N** | **Control Mean** | **MD (95% CI)** | **Fixed W** | **Random W** |
| Bangladesh | JiVitA−4 (21) | BSID−III | 353 | 112 | 0.14 | −0.11 (−0.27, 0.06) | 0.08 | 0.10 | 91 | 31 | −0.12 | −0.10 (−0.61, 0.40) | 0.01 | 0.01 |
| Bangladesh | RDNS (22) | DMC | 1116 | 532 | −0.02 | 0.10 (0.02, 0.18) | 0.34 | 0.26 | 452 | 226 | −0.07 | 0.04 (−0.07, 0.15) | 0.24 | 0.24 |
| Bangladesh | WASH−B (23) | EASQ | 472 | 998 | −0.09 | 0.21 (0.10, 0.32) | 0.17 | 0.17 | 29 | 53 | −0.20 | 0.18 (−0.39, 0.74) | 0.01 | 0.01 |
| Burkina Faso | iLiNS−Zinc (24) | DMC | | | | | | | | | | | | |
| Ghana | GHANA (25) | | | | | | | | | | | | | |
| Ghana | iLiNS−DYADG (26) | KDI | 292 | 586 | 0.00 | 0.00 (−0.14, 0.14) | 0.10 | 0.12 | 9 | 13 | −0.13 | 0.54 (−0.21, 1.28) | 0.01 | 0.01 |
| Haiti | HAITI (27) | | | | | | | | | | | | | |
| Kenya | WASH−B (28) | EASQ | 94 | 337 | 0.12 | 0.09 (−0.13, 0.30) | 0.04 | 0.06 | 555 | 1732 | −0.01 | 0.02 (−0.08, 0.13) | 0.29 | 0.29 |
| Madagascar | MAHAY (29) | ASQI | | | | | | | | | | | | |
| Malawi | iLiNS−DYADM (30) | KDI | 19 | 41 | −0.09 | 0.12 (−0.29, 0.53) | 0.01 | 0.02 | 195 | 393 | 0.03 | −0.06 (−0.23, 0.11) | 0.10 | 0.10 |
| Malawi | iLiNS−DOSE (31) | KDI | 12 | 9 | 0.43 | −0.01 (−0.57, 0.56) | 0.01 | 0.01 | 547 | 183 | −0.02 | 0.05 (−0.12, 0.21) | 0.11 | 0.11 |
| Mali | PROMIS CS (32) | DMC | 695 | 688 | −0.04 | 0.05 (−0.05, 0.16) | 0.20 | 0.19 | 216 | 244 | −0.04 | 0.18 (0.02, 0.34) | 0.12 | 0.12 |
| Zimbabwe | SHINE (HIV−) (33) | MDAT | 133 | 108 | 0.09 | 0.05 (−0.21, 0.30) | 0.03 | 0.05 | 225 | 235 | −0.01 | 0.02 (−0.17, 0.21) | 0.08 | 0.08 |
| Zimbabwe | SHINE (HIV+) (34) | MDAT | 25 | 17 | 0.02 | −0.04 (−0.49, 0.41) | 0.01 | 0.02 | 37 | 49 | −0.15 | 0.14 (−0.20, 0.48) | 0.03 | 0.03 |
| | | | **3211** | **3428** | | **I² = 0.28, Tau² = 0.00** | | | **2356** | **3159** | | **I² = 0.00, Tau² = 0.00** | | |
| **Fixed** | | | | | | **0.08 (0.03, 0.12)** | | | | | | **0.05 (−0.01, 0.10)** | | |
| **Random** | | | | | | **0.07 (0.01, 0.13)** | | | | | | **0.05 (−0.01, 0.10)** | | |

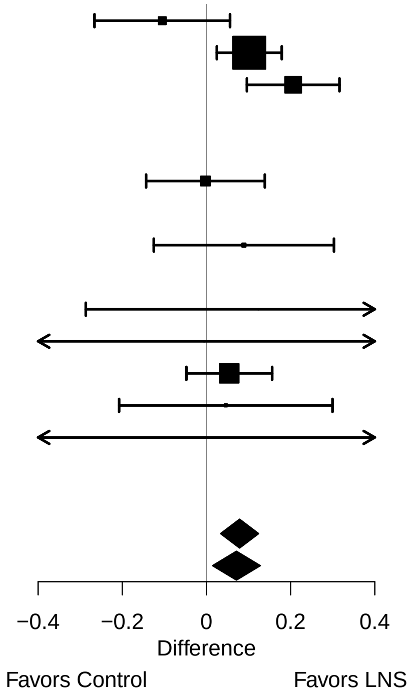

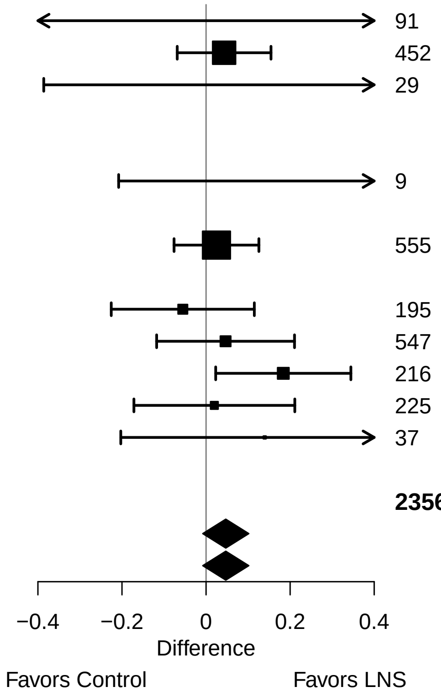

#### Supplemental figure 6J: Mean difference in gross motor z-score

##### 6J5: Stratified by Home environment

**P−for−interaction = 0.300**
**Difference in MDs = −0.04 (−0.10, 0.03)**

| | | | At least median | | | | | | Less than median | | | | | |
|---|---|---|---|---|---|---|---|---|---|---|---|---|---|---|
| Country | Trial | Tool | LNS N | Control N | Control Mean | MD (95% CI) | Fixed W | Random W | LNS N | Control N | Control Mean | MD (95% CI) | Fixed W | Random W |
| Bangladesh | JiVitA−4 (21) | BSID−III | | | | | | | | | | | | |
| Bangladesh | RDNS (22) | DMC | 1126 | 551 | 0.06 | 0.07 (0.01, 0.12) | 0.47 | 0.47 | 442 | 205 | −0.30 | 0.14 (−0.02, 0.31) | 0.11 | 0.11 |
| Bangladesh | WASH−B (23) | EASQ | 605 | 1614 | 0.13 | 0.12 (0.02, 0.21) | 0.15 | 0.15 | 469 | 1664 | −0.17 | 0.04 (−0.06, 0.14) | 0.32 | 0.32 |
| Burkina Faso | iLiNS−Zinc (24) | DMC | | | | | | | | | | | | |
| Ghana | GHANA (25) | | | | | | | | | | | | | |
| Ghana | iLiNS−DYADG (26) | KDI | 191 | 377 | 0.07 | 0.06 (−0.11, 0.22) | 0.05 | 0.05 | 111 | 224 | −0.12 | −0.07 (−0.31, 0.17) | 0.05 | 0.05 |
| Haiti | HAITI (27) | | | | | | | | | | | | | |
| Kenya | WASH−B (28) | EASQ | 809 | 2682 | 0.15 | 0.01 (−0.07, 0.09) | 0.22 | 0.22 | 552 | 2060 | −0.20 | −0.01 (−0.11, 0.09) | 0.31 | 0.31 |
| Madagascar | MAHAY (29) | ASQI | 843 | 839 | 0.03 | 0.11 (−0.07, 0.30) | 0.04 | 0.04 | 770 | 764 | −0.11 | 0.03 (−0.12, 0.19) | 0.12 | 0.12 |
| Malawi | iLiNS−DYADM (30) | KDI | 136 | 287 | 0.14 | −0.03 (−0.21, 0.15) | 0.04 | 0.04 | 78 | 149 | −0.23 | −0.01 (−0.32, 0.30) | 0.03 | 0.03 |
| Malawi | iLiNS−DOSE (31) | KDI | 376 | 135 | 0.04 | 0.03 (−0.16, 0.22) | 0.04 | 0.04 | 269 | 86 | −0.07 | −0.03 (−0.27, 0.22) | 0.05 | 0.05 |
| Mali | PROMIS CS (32) | DMC | | | | | | | | | | | | |
| Zimbabwe | SHINE (HIV−) (33) | MDAT | | | | | | | | | | | | |
| Zimbabwe | SHINE (HIV+) (34) | MDAT | | | | | | | | | | | | |
| | | | **4086** | **6485** | | **I² = 0.00, Tau² = 0.00** | | | **2691** | **5152** | | **I² = 0.00, Tau² = 0.00** | | |
| **Fixed** | | | | | | **0.06 (0.02, 0.09)** | | | | | | **0.02 (−0.03, 0.08)** | | |
| **Random** | | | | | | **0.06 (0.02, 0.09)** | | | | | | **0.02 (−0.03, 0.08)** | | |

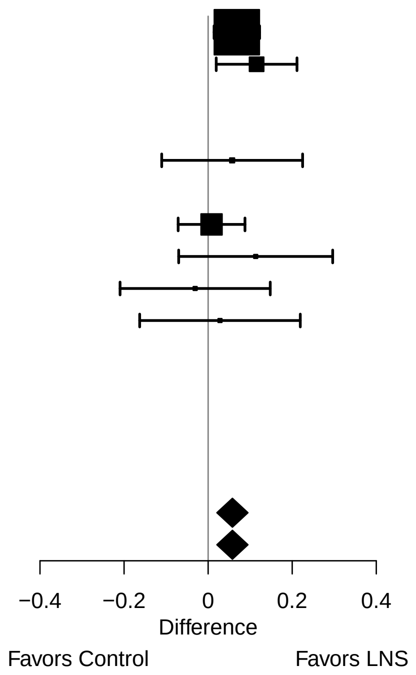

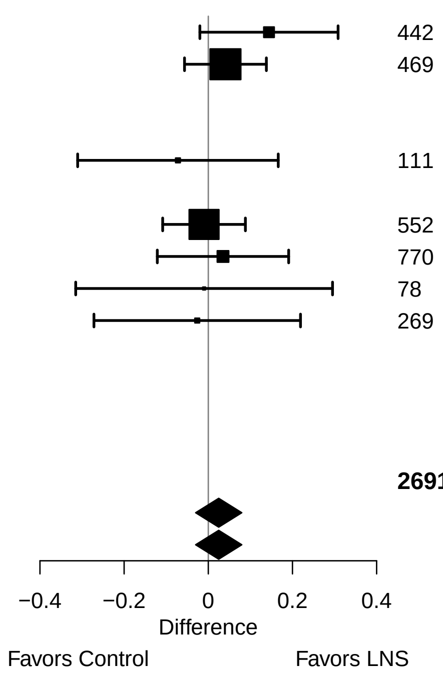

#### Supplemental figure 6J: Mean difference in gross motor z-score

#### 6J6: Stratified by Season at the time of assessment

**P−for−interaction = 0.958**
**Difference in MDs = 0.00 (−0.07, 0.06)**

| Country | Trial | Tool | Dry LNS N | Dry Control N | Dry Control Mean | Dry MD (95% CI) | Dry Fixed W | Dry Random W | Rainy LNS N | Rainy Control N | Rainy Control Mean | Rainy MD (95% CI) | Rainy Fixed W | Rainy Random W |
|---|---|---|---|---|---|---|---|---|---|---|---|---|---|---|
| Bangladesh | JiVitA−4 (21) | BSID−III | 251 | 70 | 0.32 | −0.10 (−0.30, 0.11) | 0.04 | 0.07 | 194 | 73 | −0.14 | −0.18 (−0.43, 0.06) | 0.03 | 0.07 |
| Bangladesh | RDNS (22) | DMC | 703 | 330 | −0.03 | 0.08 (−0.01, 0.17) | 0.21 | 0.16 | 867 | 428 | −0.04 | 0.09 (0.00, 0.17) | 0.28 | 0.21 |
| Bangladesh | WASH−B (23) | EASQ | 608 | 1896 | 0.04 | 0.08 (−0.01, 0.17) | 0.24 | 0.17 | 466 | 1383 | −0.11 | 0.14 (0.04, 0.24) | 0.20 | 0.19 |
| Burkina Faso | iLiNS−Zinc (24) | DMC | | | | | | | | | | | | |
| Ghana | GHANA (25) | | | | | | | | | | | | | |
| Ghana | iLiNS−DYADG (26) | KDI | 177 | 356 | −0.05 | −0.03 (−0.20, 0.15) | 0.06 | 0.09 | 125 | 245 | 0.07 | 0.06 (−0.16, 0.29) | 0.04 | 0.08 |
| Haiti | HAITI (27) | | | | | | | | | | | | | |
| Kenya | WASH−B (28) | EASQ | 315 | 1059 | −0.30 | 0.09 (−0.01, 0.20) | 0.17 | 0.15 | 1046 | 3682 | 0.08 | −0.01 (−0.09, 0.07) | 0.33 | 0.22 |
| Madagascar | MAHAY (29) | ASQI | | | | | | | | | | | | |
| Malawi | iLiNS−DYADM (30) | KDI | 153 | 308 | 0.04 | −0.02 (−0.21, 0.18) | 0.05 | 0.08 | 61 | 127 | −0.05 | −0.06 (−0.34, 0.21) | 0.03 | 0.06 |
| Malawi | iLiNS−DOSE (31) | KDI | 395 | 118 | −0.02 | −0.07 (−0.27, 0.13) | 0.05 | 0.07 | 244 | 100 | 0.02 | 0.12 (−0.11, 0.36) | 0.04 | 0.07 |
| Mali | PROMIS CS (32) | DMC | | | | | | | | | | | | |
| Zimbabwe | SHINE (HIV−) (33) | MDAT | 603 | 568 | −0.07 | 0.12 (0.00, 0.23) | 0.13 | 0.14 | 238 | 231 | 0.01 | 0.05 (−0.17, 0.28) | 0.04 | 0.08 |
| Zimbabwe | SHINE (HIV+) (34) | MDAT | 121 | 110 | −0.17 | 0.31 (0.10, 0.52) | 0.04 | 0.07 | 49 | 41 | −0.21 | 0.43 (0.01, 0.85) | 0.01 | 0.03 |
| | | | **3326** | **4815** | | **I² = 0.34, Tau² = 0.00** | | | **3290** | **6310** | | **I² = 0.42, Tau² = 0.01** | | |
| **Fixed** | | | | | | **0.07 (0.03, 0.11)** | | | | | | **0.06 (0.01, 0.10)** | | |
| **Random** | | | | | | **0.06 (0.00, 0.13)** | | | | | | **0.06 (−0.02, 0.13)** | | |

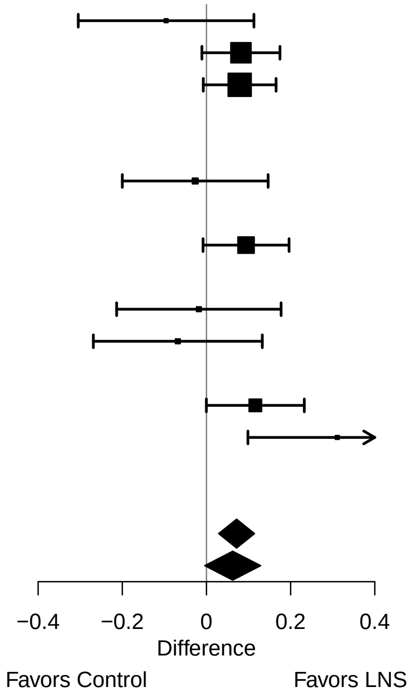

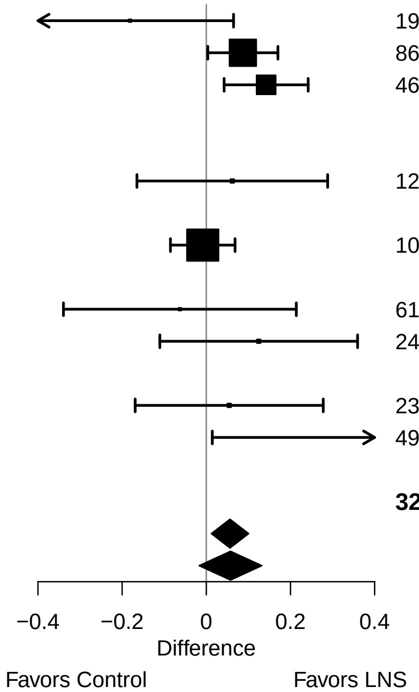

### Supplemental figure 6K: Mean difference in fine motor z-score

#### 6K1: Stratified by Household socio-economic status

**P−for−interaction = 0.967**
**Difference in MDs = 0.00 (−0.08, 0.08)**

| | | | At least median | | | | | | Less than median | | | | | |
|---|---|---|---|---|---|---|---|---|---|---|---|---|---|---|
| **Country** | **Trial** | **Tool** | **LNS N** | **Control N** | **Control Mean** | **MD (95% CI)** | **Fixed W** | **Random W** | **LNS N** | **Control N** | **Control Mean** | **MD (95% CI)** | **Fixed W** | **Random W** |
| Bangladesh | JiVitA−4 (21) | BSID−III | 236 | 77 | 0.03 | 0.07 (−0.18, 0.32) | 0.06 | 0.06 | 208 | 66 | −0.27 | 0.23 (−0.03, 0.50) | 0.06 | 0.06 |
| Bangladesh | RDNS (22) | DMC | 827 | 403 | 0.05 | 0.08 (−0.06, 0.22) | 0.20 | 0.20 | 814 | 401 | −0.16 | 0.12 (0.01, 0.23) | 0.32 | 0.32 |
| Bangladesh | WASH−B (23) | EASQ | | | | | | | | | | | | |
| Burkina Faso | iLiNS−Zinc (24) | DMC | | | | | | | | | | | | |
| Ghana | GHANA (25) | | | | | | | | | | | | | |
| Ghana | iLiNS−DYADG (26) | KDI | 143 | 311 | 0.00 | 0.06 (−0.11, 0.24) | 0.13 | 0.13 | 159 | 288 | 0.01 | 0.00 (−0.18, 0.19) | 0.12 | 0.12 |
| Haiti | HAITI (27) | | | | | | | | | | | | | |
| Kenya | WASH−B (28) | EASQ | | | | | | | | | | | | |
| Madagascar | MAHAY (29) | ASQI | 741 | 845 | 0.16 | 0.04 (−0.14, 0.21) | 0.13 | 0.13 | 823 | 724 | −0.25 | 0.11 (−0.17, 0.39) | 0.05 | 0.05 |
| Malawi | iLiNS−DYADM (30) | KDI | 118 | 227 | −0.06 | 0.11 (−0.10, 0.31) | 0.09 | 0.09 | 96 | 208 | 0.02 | 0.05 (−0.19, 0.29) | 0.07 | 0.07 |
| Malawi | iLiNS−DOSE (31) | KDI | 278 | 94 | 0.03 | 0.14 (−0.07, 0.36) | 0.09 | 0.09 | 271 | 96 | −0.11 | 0.03 (−0.22, 0.28) | 0.06 | 0.06 |
| Mali | PROMIS CS (32) | DMC | 482 | 448 | −0.02 | 0.13 (−0.12, 0.38) | 0.06 | 0.06 | 423 | 476 | −0.08 | 0.08 (−0.15, 0.31) | 0.07 | 0.07 |
| Zimbabwe | SHINE (HIV−) (33) | MDAT | 406 | 359 | −0.07 | 0.10 (−0.05, 0.25) | 0.18 | 0.18 | 389 | 391 | −0.02 | 0.08 (−0.06, 0.21) | 0.22 | 0.22 |
| Zimbabwe | SHINE (HIV+) (34) | MDAT | 91 | 78 | −0.01 | 0.12 (−0.17, 0.41) | 0.05 | 0.05 | 74 | 67 | −0.14 | 0.18 (−0.18, 0.54) | 0.03 | 0.03 |
| | | | **3322** | **2842** | | **I² = 0.00, Tau² = 0.00** | | | **3257** | **2717** | | **I² = 0.00, Tau² = 0.00** | | |
| **Fixed** | | | | | | **0.09 (0.02, 0.15)** | | | | | | **0.09 (0.03, 0.15)** | | |
| **Random** | | | | | | **0.09 (0.02, 0.15)** | | | | | | **0.09 (0.03, 0.15)** | | |

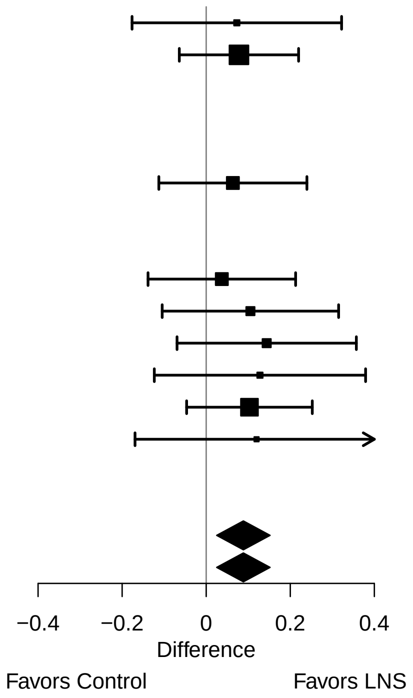

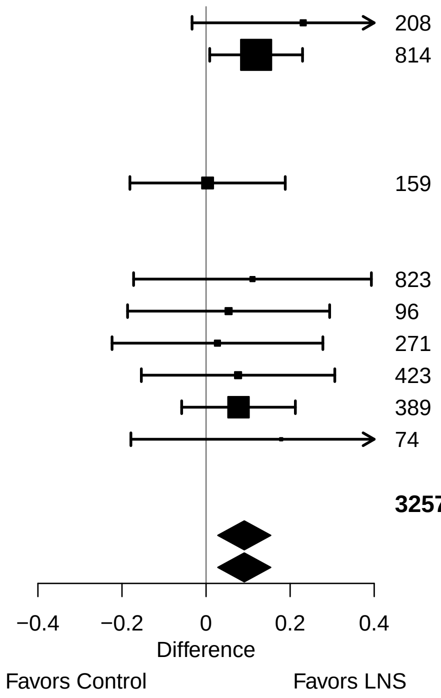

#### Supplemental figure 6K: Mean difference in fine motor z-score

#### 6K2: Stratified by Household food insecurity

**P−for−interaction = 0.352**
**Difference in MDs = 0.05 (−0.05, 0.15)**

| Country | Trial | Tool | LNS N | Control N | Control Mean | Mild to secure MD (95% CI) | Fixed W | Random W | LNS N | Control N | Control Mean | Moderate to severe MD (95% CI) | Fixed W | Random W |
|---|---|---|---|---|---|---|---|---|---|---|---|---|---|---|
| Bangladesh | JiVitA−4 (21) | BSID−III | 329 | 111 | −0.06 | 0.14 (−0.09, 0.36) | 0.07 | 0.07 | 115 | 32 | −0.26 | 0.19 (−0.10, 0.48) | 0.07 | 0.07 |
| Bangladesh | RDNS (22) | DMC | 1045 | 486 | 0.04 | 0.04 (−0.10, 0.17) | 0.20 | 0.20 | 596 | 318 | −0.19 | 0.19 (0.06, 0.31) | 0.37 | 0.37 |
| Bangladesh | WASH−B (23) | EASQ | | | | | | | | | | | | |
| Burkina Faso | iLiNS−Zinc (24) | DMC | | | | | | | | | | | | |
| Ghana | GHANA (25) | | | | | | | | | | | | | |
| Ghana | iLiNS−DYADG (26) | KDI | 217 | 412 | 0.04 | 0.07 (−0.06, 0.20) | 0.22 | 0.22 | 84 | 186 | −0.07 | −0.09 (−0.38, 0.21) | 0.07 | 0.07 |
| Haiti | HAITI (27) | | | | | | | | | | | | | |
| Kenya | WASH−B (28) | EASQ | | | | | | | | | | | | |
| Madagascar | MAHAY (29) | ASQI | 940 | 892 | 0.05 | 0.02 (−0.20, 0.24) | 0.08 | 0.08 | 336 | 389 | −0.26 | 0.12 (−0.28, 0.53) | 0.04 | 0.04 |
| Malawi | iLiNS−DYADM (30) | KDI | 67 | 116 | −0.08 | 0.11 (−0.21, 0.43) | 0.04 | 0.04 | 146 | 314 | 0.00 | 0.07 (−0.11, 0.26) | 0.18 | 0.18 |
| Malawi | iLiNS−DOSE (31) | KDI | 134 | 51 | −0.01 | 0.06 (−0.26, 0.38) | 0.04 | 0.04 | 403 | 133 | −0.02 | 0.08 (−0.12, 0.27) | 0.15 | 0.15 |
| Mali | PROMIS CS (32) | DMC | | | | | | | | | | | | |
| Zimbabwe | SHINE (HIV−) (33) | MDAT | 631 | 586 | −0.02 | 0.08 (−0.03, 0.20) | 0.29 | 0.29 | 140 | 154 | −0.14 | 0.14 (−0.09, 0.37) | 0.11 | 0.11 |
| Zimbabwe | SHINE (HIV+) (34) | MDAT | 119 | 105 | −0.07 | 0.19 (−0.05, 0.42) | 0.07 | 0.07 | 41 | 37 | −0.11 | 0.03 (−0.54, 0.60) | 0.02 | 0.02 |
| | | | **3482** | **2759** | | **I² = 0.00, Tau² = 0.00** | | | **1861** | **1563** | | **I² = 0.00, Tau² = 0.00** | | |
| **Fixed** | | | | | | **0.08 (0.02, 0.14)** | | | | | | **0.12 (0.04, 0.20)** | | |
| **Random** | | | | | | **0.08 (0.02, 0.14)** | | | | | | **0.12 (0.04, 0.20)** | | |

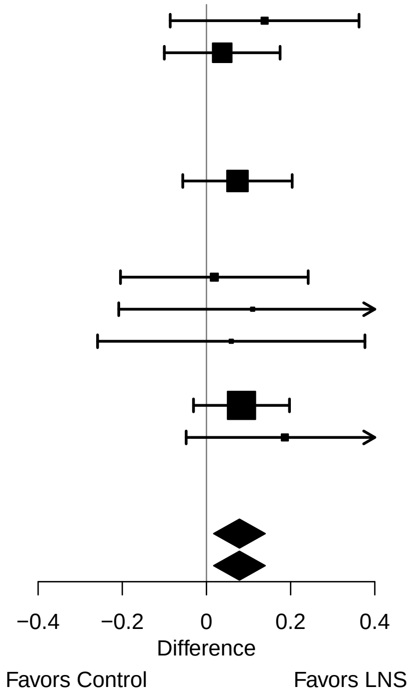

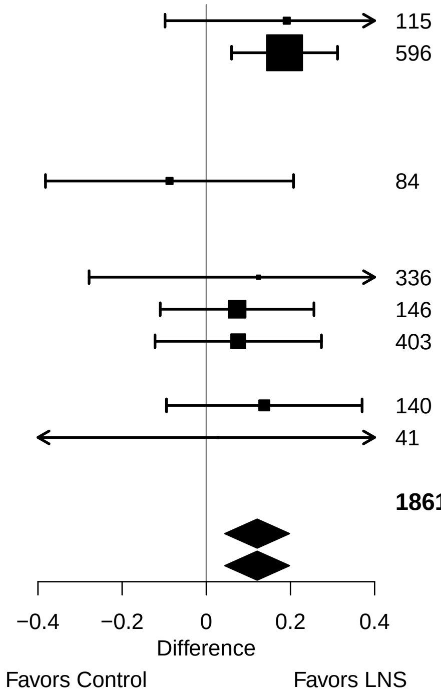

#### Supplemental figure 6K: Mean difference in fine motor z-score

##### 6K3: Stratified by Household source water quality

**P−for−interaction = 0.410**
**Difference in MDs = 0.07 (−0.10, 0.25)**

| Country | Trial | Tool | LNS N | Control N | Control Mean | Improved MD (95% CI) | Fixed W | Random W | LNS N | Control N | Control Mean | Unimproved MD (95% CI) | Fixed W | Random W |
|---|---|---|---|---|---|---|---|---|---|---|---|---|---|---|
| Bangladesh | JiVitA−4 (21) | BSID−III | | | | | | | | | | | | |
| Bangladesh | RDNS (22) | DMC | | | | | | | | | | | | |
| Bangladesh | WASH−B (23) | EASQ | | | | | | | | | | | | |
| Burkina Faso | iLiNS−Zinc (24) | DMC | | | | | | | | | | | | |
| Ghana | GHANA (25) | | | | | | | | | | | | | |
| Ghana | iLiNS−DYADG (26) | KDI | | | | | | | | | | | | |
| Haiti | HAITI (27) | | | | | | | | | | | | | |
| Kenya | WASH−B (28) | EASQ | | | | | | | | | | | | |
| Madagascar | MAHAY (29) | ASQI | 405 | 430 | −0.14 | 0.02 (−0.37, 0.41) | 0.05 | 0.05 | 1134 | 1133 | 0.02 | 0.07 (−0.16, 0.29) | 0.34 | 0.34 |
| Malawi | iLiNS−DYADM (30) | KDI | 194 | 398 | 0.00 | 0.05 (−0.11, 0.22) | 0.30 | 0.30 | 20 | 36 | −0.22 | 0.34 (−0.23, 0.91) | 0.05 | 0.05 |
| Malawi | iLiNS−DOSE (31) | KDI | 515 | 177 | −0.01 | 0.08 (−0.09, 0.25) | 0.28 | 0.28 | 44 | 15 | −0.38 | 0.29 (−0.30, 0.89) | 0.05 | 0.05 |
| Mali | PROMIS CS (32) | DMC | 516 | 540 | −0.02 | 0.09 (−0.19, 0.37) | 0.10 | 0.10 | 348 | 370 | −0.13 | 0.06 (−0.17, 0.29) | 0.30 | 0.30 |
| Zimbabwe | SHINE (HIV−) (33) | MDAT | 228 | 220 | 0.10 | −0.09 (−0.28, 0.11) | 0.22 | 0.22 | 131 | 122 | −0.09 | 0.09 (−0.19, 0.37) | 0.22 | 0.22 |
| Zimbabwe | SHINE (HIV+) (34) | MDAT | 37 | 41 | 0.01 | −0.01 (−0.40, 0.38) | 0.05 | 0.05 | 25 | 25 | −0.16 | −0.21 (−0.85, 0.42) | 0.04 | 0.04 |
| | | | **1895** | **1806** | | **I² = 0.00, Tau² = 0.00** | | | **1702** | **1701** | | **I² = 0.00, Tau² = 0.00** | | |
| **Fixed** | | | | | | **0.03 (−0.06, 0.12)** | | | | | | **0.08 (−0.05, 0.21)** | | |
| **Random** | | | | | | **0.03 (−0.06, 0.12)** | | | | | | **0.08 (−0.05, 0.21)** | | |

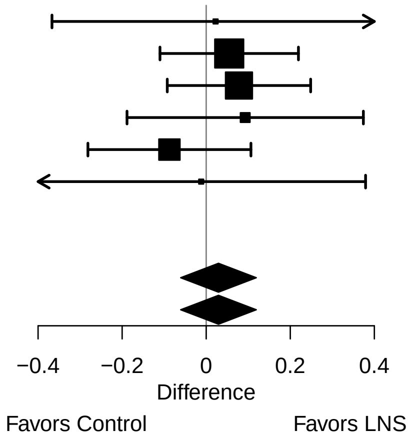

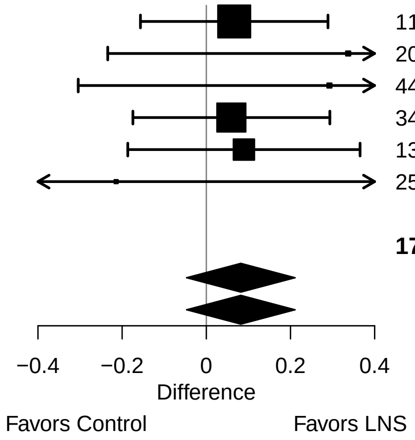

#### Supplemental figure 6K: Mean difference in fine motor z-score

#### 6K4: Stratified by Household sanitation

**P−for−interaction = 0.172**
**Difference in MDs = −0.09 (−0.23, 0.04)**

| Country | Trial | Tool | Improved LNS N | Improved Control N | Improved Control Mean | Improved MD (95% CI) | Improved Fixed W | Improved Random W | Unimproved LNS N | Unimproved Control N | Unimproved Control Mean | Unimproved MD (95% CI) | Unimproved Fixed W | Unimproved Random W |
|---|---|---|---|---|---|---|---|---|---|---|---|---|---|---|
| Bangladesh | JiVitA−4 (21) | BSID−III | 353 | 112 | −0.07 | 0.14 (−0.06, 0.35) | 0.09 | 0.09 | 91 | 31 | −0.26 | 0.13 (−0.32, 0.59) | 0.03 | 0.03 |
| Bangladesh | RDNS (22) | DMC | 1161 | 568 | −0.01 | 0.09 (0.00, 0.17) | 0.50 | 0.50 | 478 | 235 | −0.16 | 0.13 (−0.08, 0.34) | 0.16 | 0.16 |
| Bangladesh | WASH−B (23) | EASQ | | | | | | | | | | | | |
| Burkina Faso | iLiNS−Zinc (24) | DMC | | | | | | | | | | | | |
| Ghana | GHANA (25) | | | | | | | | | | | | | |
| Ghana | iLiNS−DYADG (26) | KDI | 292 | 586 | 0.00 | 0.05 (−0.08, 0.17) | 0.23 | 0.23 | 9 | 13 | 0.22 | −0.47 (−1.23, 0.30) | 0.01 | 0.01 |
| Haiti | HAITI (27) | | | | | | | | | | | | | |
| Kenya | WASH−B (28) | EASQ | | | | | | | | | | | | |
| Madagascar | MAHAY (29) | ASQI | | | | | | | | | | | | |
| Malawi | iLiNS−DYADM (30) | KDI | 19 | 41 | −0.15 | 0.36 (−0.14, 0.87) | 0.02 | 0.02 | 195 | 393 | −0.01 | 0.05 (−0.12, 0.22) | 0.25 | 0.25 |
| Malawi | iLiNS−DOSE (31) | KDI | 12 | 9 | 0.08 | 0.35 (−0.29, 0.99) | 0.01 | 0.01 | 547 | 183 | −0.05 | 0.09 (−0.08, 0.26) | 0.25 | 0.25 |
| Mali | PROMIS CS (32) | DMC | 668 | 660 | −0.07 | 0.18 (−0.06, 0.43) | 0.07 | 0.07 | 210 | 242 | 0.02 | −0.15 (−0.48, 0.18) | 0.06 | 0.06 |
| Zimbabwe | SHINE (HIV−) (33) | MDAT | 133 | 108 | 0.00 | 0.09 (−0.14, 0.32) | 0.07 | 0.07 | 225 | 235 | 0.04 | −0.09 (−0.28, 0.11) | 0.19 | 0.19 |
| Zimbabwe | SHINE (HIV+) (34) | MDAT | 25 | 17 | −0.05 | −0.24 (−0.83, 0.35) | 0.01 | 0.01 | 37 | 49 | −0.06 | 0.00 (−0.39, 0.39) | 0.05 | 0.05 |
| | | | **2663** | **2101** | | **I² = 0.00, Tau² = 0.00** | | | **1792** | **1381** | | **I² = 0.00, Tau² = 0.00** | | |
| **Fixed** | | | | | | **0.09 (0.03, 0.15)** | | | | | | **0.03 (−0.06, 0.11)** | | |
| **Random** | | | | | | **0.09 (0.03, 0.15)** | | | | | | **0.03 (−0.06, 0.11)** | | |

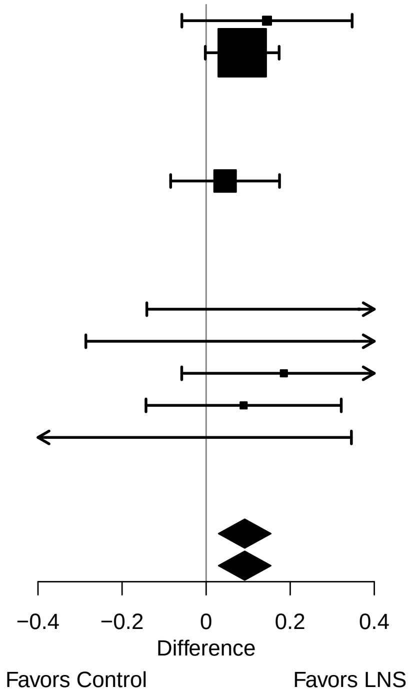

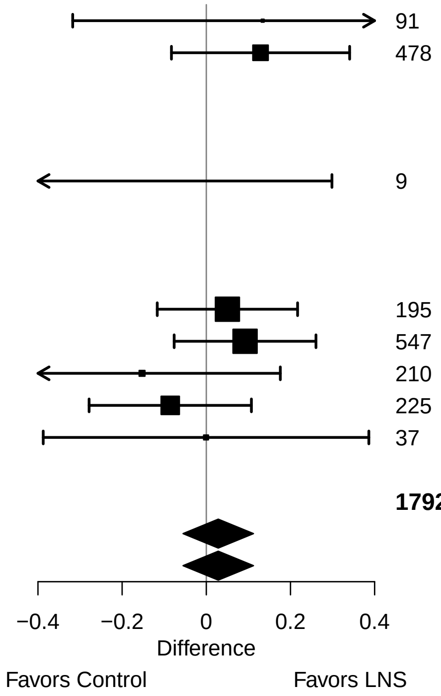

#### Supplemental figure 6K: Mean difference in fine motor z-score

#### 6K5: Stratified by Home environment

**P−for−interaction = 0.860**
**Difference in MDs = 0.01 (−0.11, 0.13)**

| Country | Trial | Tool | LNS N | Control N | Control Mean | At least median MD (95% CI) | Fixed W | Random W | LNS N | Control N | Control Mean | Less than median MD (95% CI) | Fixed W | Random W |
|---|---|---|---|---|---|---|---|---|---|---|---|---|---|---|
| Bangladesh | JiVitA−4 (21) | BSID−III | | | | | | | | | | | | |
| Bangladesh | RDNS (22) | DMC | 1177 | 576 | 0.09 | 0.06 (−0.04, 0.15) | 0.48 | 0.48 | 461 | 225 | −0.40 | 0.20 (−0.05, 0.44) | 0.21 | 0.21 |
| Bangladesh | WASH−B (23) | EASQ | | | | | | | | | | | | |
| Burkina Faso | iLiNS−Zinc (24) | DMC | | | | | | | | | | | | |
| Ghana | GHANA (25) | | | | | | | | | | | | | |
| Ghana | iLiNS−DYADG (26) | KDI | 191 | 377 | 0.09 | 0.04 (−0.10, 0.19) | 0.21 | 0.21 | 111 | 224 | −0.15 | 0.03 (−0.22, 0.27) | 0.21 | 0.21 |
| Haiti | HAITI (27) | | | | | | | | | | | | | |
| Kenya | WASH−B (28) | EASQ | | | | | | | | | | | | |
| Madagascar | MAHAY (29) | ASQI | 843 | 839 | 0.05 | 0.07 (−0.16, 0.30) | 0.08 | 0.08 | 770 | 764 | −0.10 | 0.02 (−0.22, 0.26) | 0.22 | 0.22 |
| Malawi | iLiNS−DYADM (30) | KDI | 136 | 287 | 0.05 | 0.08 (−0.10, 0.26) | 0.12 | 0.12 | 78 | 149 | −0.16 | 0.09 (−0.20, 0.39) | 0.15 | 0.15 |
| Malawi | iLiNS−DOSE (31) | KDI | 376 | 135 | 0.01 | 0.09 (−0.10, 0.27) | 0.12 | 0.12 | 269 | 86 | −0.18 | 0.10 (−0.15, 0.35) | 0.20 | 0.20 |
| Mali | PROMIS CS (32) | DMC | | | | | | | | | | | | |
| Zimbabwe | SHINE (HIV−) (33) | MDAT | | | | | | | | | | | | |
| Zimbabwe | SHINE (HIV+) (34) | MDAT | | | | | | | | | | | | |
| | | | **2723** | **2214** | | **I² = 0.00, Tau² = 0.00** | | | **1689** | **1448** | | **I² = 0.00, Tau² = 0.00** | | |
| **Fixed** | | | | | | **0.06 (0.00, 0.13)** | | | | | | **0.09 (−0.03, 0.20)** | | |
| **Random** | | | | | | **0.06 (0.00, 0.13)** | | | | | | **0.09 (−0.03, 0.20)** | | |

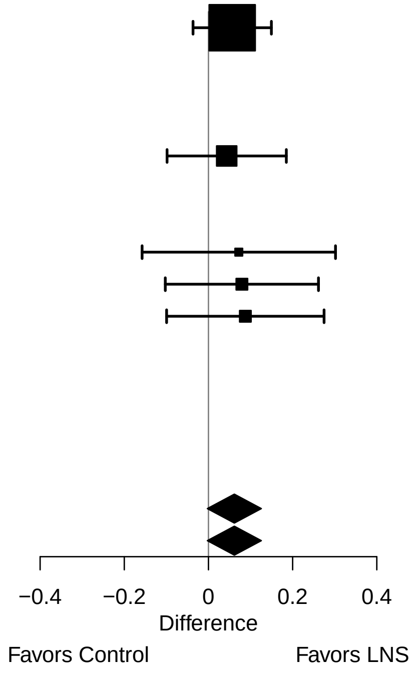

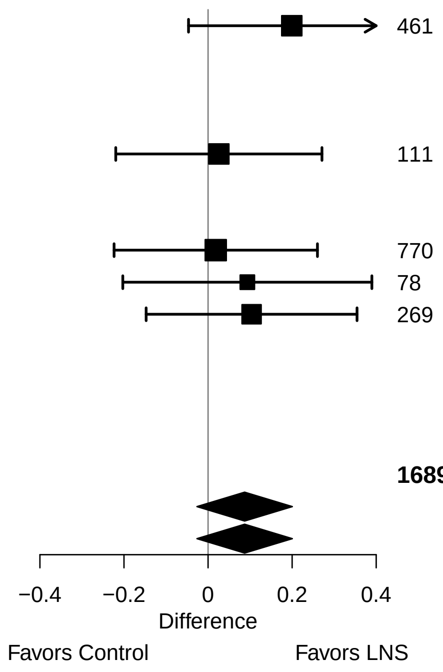

#### Supplemental figure 6K: Mean difference in fine motor z-score

#### 6K6: Stratified by Season at the time of assessment

**P−for−interaction = 0.993**
**Difference in MDs = 0.00 (−0.09, 0.09)**

| Country | Trial | Tool | Dry LNS N | Dry Control N | Dry Control Mean | Dry MD (95% CI) | Dry Fixed W | Dry Random W | Rainy LNS N | Rainy Control N | Rainy Control Mean | Rainy MD (95% CI) | Rainy Fixed W | Rainy Random W |
|---|---|---|---|---|---|---|---|---|---|---|---|---|---|---|
| Bangladesh | JiVitA−4 (21) | BSID−III | 251 | 70 | 0.06 | 0.11 (−0.09, 0.31) | 0.09 | 0.09 | 194 | 73 | −0.27 | 0.13 (−0.22, 0.48) | 0.05 | 0.05 |
| Bangladesh | RDNS (22) | DMC | 731 | 352 | −0.01 | 0.07 (−0.03, 0.18) | 0.31 | 0.31 | 910 | 452 | −0.08 | 0.12 (−0.02, 0.26) | 0.33 | 0.33 |
| Bangladesh | WASH−B (23) | EASQ | | | | | | | | | | | | |
| Burkina Faso | iLiNS−Zinc (24) | DMC | | | | | | | | | | | | |
| Ghana | GHANA (25) | | | | | | | | | | | | | |
| Ghana | iLiNS−DYADG (26) | KDI | 177 | 356 | 0.08 | 0.00 (−0.17, 0.16) | 0.12 | 0.12 | 125 | 245 | −0.12 | 0.10 (−0.10, 0.30) | 0.16 | 0.16 |
| Haiti | HAITI (27) | | | | | | | | | | | | | |
| Kenya | WASH−B (28) | EASQ | | | | | | | | | | | | |
| Madagascar | MAHAY (29) | ASQI | | | | | | | | | | | | |
| Malawi | iLiNS−DYADM (30) | KDI | 153 | 308 | −0.01 | 0.09 (−0.10, 0.28) | 0.10 | 0.10 | 61 | 127 | −0.06 | 0.06 (−0.24, 0.35) | 0.08 | 0.08 |
| Malawi | iLiNS−DOSE (31) | KDI | 395 | 118 | −0.14 | 0.10 (−0.09, 0.30) | 0.09 | 0.09 | 244 | 100 | 0.02 | 0.11 (−0.14, 0.36) | 0.11 | 0.11 |
| Mali | PROMIS CS (32) | DMC | | | | | | | | | | | | |
| Zimbabwe | SHINE (HIV−) (33) | MDAT | 603 | 568 | −0.06 | 0.11 (0.00, 0.23) | 0.25 | 0.25 | 238 | 231 | 0.04 | −0.02 (−0.21, 0.16) | 0.20 | 0.20 |
| Zimbabwe | SHINE (HIV+) (34) | MDAT | 121 | 110 | −0.14 | 0.20 (−0.08, 0.48) | 0.04 | 0.04 | 49 | 41 | 0.08 | 0.08 (−0.22, 0.37) | 0.07 | 0.07 |
| | | | **2431** | **1882** | | **I² = 0.00, Tau² = 0.00** | | | **1821** | **1269** | | **I² = 0.00, Tau² = 0.00** | | |
| **Fixed** | | | | | | **0.09 (0.03, 0.14)** | | | | | | **0.08 (0.00, 0.16)** | | |
| **Random** | | | | | | **0.09 (0.03, 0.14)** | | | | | | **0.08 (0.00, 0.16)** | | |

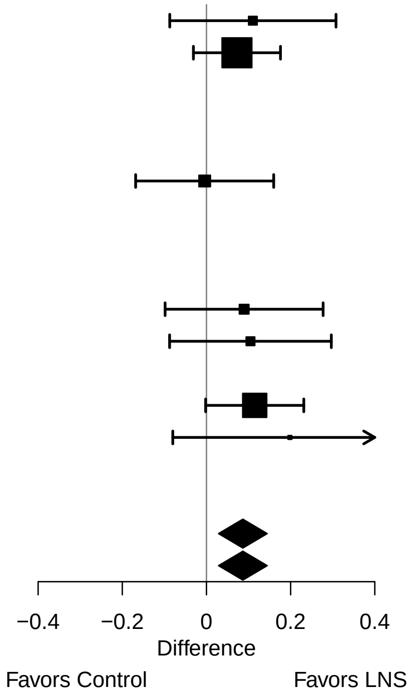

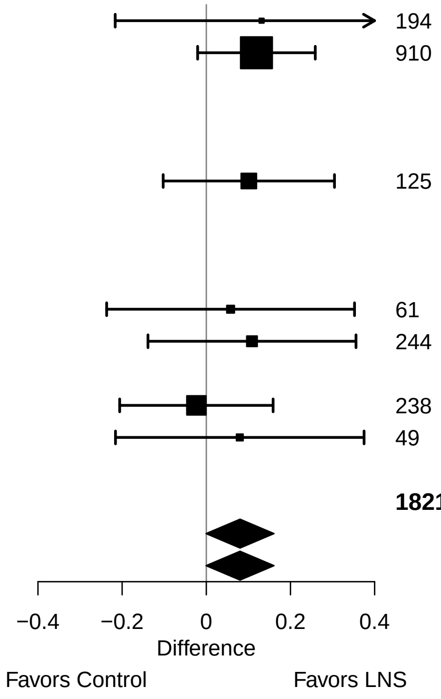

### Supplemental figure 6L: Mean difference in executive function z-score

#### 6L1: Stratified by Household socio-economic status

**P−for−interaction = 0.033**
**Difference in MDs = 0.09 (0.01, 0.18)**

| | | | At least median | | | | | | | Less than median | | | | |
|---|---|---|---|---|---|---|---|---|---|---|---|---|---|---|
| **Country** | **Trial** | **Tool** | **LNS N** | **Control N** | **Control Mean** | **MD (95% CI)** | **Fixed W** | **Random W** | **LNS N** | **Control N** | **Control Mean** | **MD (95% CI)** | **Fixed W** | **Random W** |
| Bangladesh | JiVitA−4 (21) | | | | | | | | | | | | | |
| Bangladesh | RDNS (22) | A not B task | 269 | 133 | −0.01 | 0.07 (−0.11, 0.25) | 0.12 | 0.12 | 264 | 133 | −0.13 | 0.13 (−0.06, 0.32) | 0.11 | 0.11 |
| Bangladesh | WASH−B (23) | A not B task | 566 | 1691 | 0.04 | −0.05 (−0.15, 0.05) | 0.43 | 0.43 | 531 | 1636 | −0.04 | 0.05 (−0.04, 0.14) | 0.49 | 0.49 |
| Burkina Faso | iLiNS−Zinc (24) | | | | | | | | | | | | | |
| Ghana | GHANA (25) | | | | | | | | | | | | | |
| Ghana | iLiNS−DYADG (26) | A not B task | 140 | 297 | −0.04 | 0.08 (−0.12, 0.28) | 0.10 | 0.10 | 150 | 277 | 0.02 | −0.03 (−0.23, 0.17) | 0.10 | 0.10 |
| Haiti | HAITI (27) | | | | | | | | | | | | | |
| Kenya | WASH−B (28) | | | | | | | | | | | | | |
| Madagascar | MAHAY (29) | | | | | | | | | | | | | |
| Malawi | iLiNS−DYADM (30) | A not B task | 102 | 192 | 0.00 | −0.13 (−0.37, 0.11) | 0.07 | 0.07 | 80 | 176 | 0.05 | −0.03 (−0.29, 0.23) | 0.06 | 0.06 |
| Malawi | iLiNS−DOSE (31) | A not B task | 203 | 72 | 0.06 | −0.02 (−0.28, 0.24) | 0.06 | 0.06 | 199 | 69 | 0.03 | −0.06 (−0.35, 0.22) | 0.05 | 0.05 |
| Mali | PROMIS CS (32) | | | | | | | | | | | | | |
| Zimbabwe | SHINE (HIV−) (33) | A not B task | 377 | 340 | −0.02 | −0.05 (−0.20, 0.10) | 0.17 | 0.17 | 365 | 352 | −0.03 | 0.13 (−0.03, 0.29) | 0.15 | 0.15 |
| Zimbabwe | SHINE (HIV+) (34) | A not B task | 88 | 72 | 0.13 | −0.21 (−0.49, 0.07) | 0.05 | 0.05 | 64 | 56 | −0.04 | 0.13 (−0.22, 0.48) | 0.03 | 0.03 |
| | | | **1745** | **2797** | | **$I^2$ = 0.00, $Tau^2$ = 0.00** | | | **1653** | **2699** | | **$I^2$ = 0.00, $Tau^2$ = 0.00** | | |
| **Fixed** | | | | | | **−0.04 (−0.10, 0.03)** | | | | | | **0.06 (−0.01, 0.12)** | | |
| **Random** | | | | | | **−0.04 (−0.10, 0.03)** | | | | | | **0.06 (−0.01, 0.12)** | | |

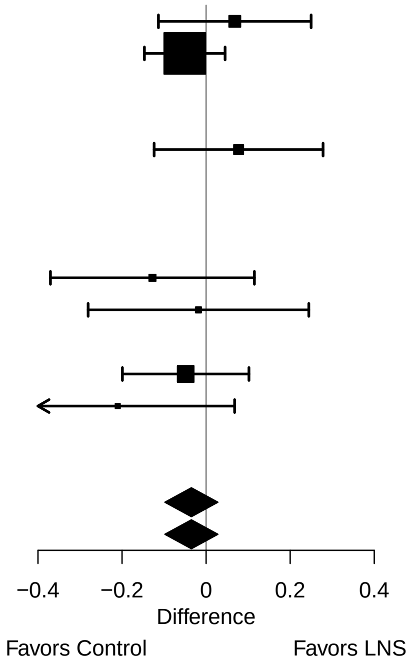

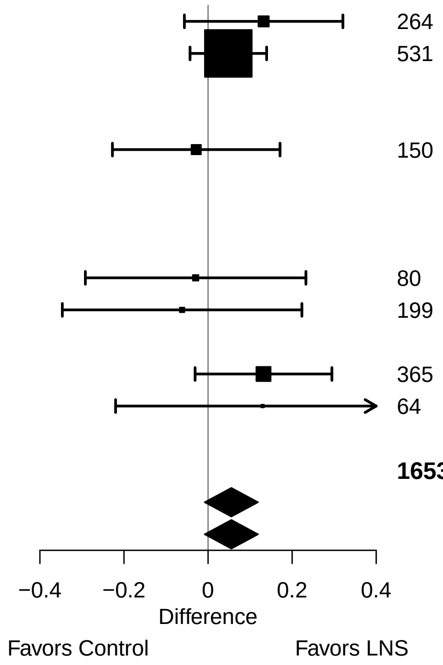

#### Supplemental figure 6L: Mean difference in executive function z-score

##### 6L2: Stratified by Household food insecurity

**P−for−interaction = 0.984**
**Difference in MDs = 0.00 (−0.10, 0.10)**

| Country | Trial | Tool | LNS N | Control N | Control Mean | Mild to secure MD (95% CI) | Fixed W | Random W | LNS N | Control N | Control Mean | Moderate to severe MD (95% CI) | Fixed W | Random W |
|---|---|---|---|---|---|---|---|---|---|---|---|---|---|---|
| Bangladesh | JiVitA−4 (21) | | | | | | | | | | | | | |
| Bangladesh | RDNS (22) | A not B task | 334 | 164 | 0.10 | −0.05 (−0.20, 0.11) | 0.13 | 0.13 | 199 | 102 | −0.34 | 0.34 (0.13, 0.54) | 0.16 | 0.16 |
| Bangladesh | WASH−B (23) | A not B task | 876 | 2590 | 0.00 | 0.02 (−0.06, 0.10) | 0.50 | 0.50 | 221 | 737 | 0.01 | −0.08 (−0.22, 0.06) | 0.33 | 0.21 |
| Burkina Faso | iLiNS−Zinc (24) | | | | | | | | | | | | | |
| Ghana | GHANA (25) | | | | | | | | | | | | | |
| Ghana | iLiNS−DYADG (26) | A not B task | 209 | 401 | 0.01 | −0.03 (−0.19, 0.14) | 0.11 | 0.11 | 80 | 173 | −0.06 | 0.14 (−0.13, 0.40) | 0.10 | 0.13 |
| Haiti | HAITI (27) | | | | | | | | | | | | | |
| Kenya | WASH−B (28) | | | | | | | | | | | | | |
| Madagascar | MAHAY (29) | | | | | | | | | | | | | |
| Malawi | iLiNS−DYADM (30) | A not B task | 55 | 99 | −0.09 | −0.12 (−0.46, 0.22) | 0.03 | 0.03 | 126 | 264 | 0.07 | −0.07 (−0.28, 0.15) | 0.15 | 0.16 |
| Malawi | iLiNS−DOSE (31) | A not B task | 103 | 37 | 0.11 | −0.07 (−0.44, 0.29) | 0.02 | 0.02 | 294 | 99 | 0.02 | −0.02 (−0.24, 0.21) | 0.12 | 0.15 |
| Mali | PROMIS CS (32) | | | | | | | | | | | | | |
| Zimbabwe | SHINE (HIV−) (33) | A not B task | 591 | 547 | −0.04 | 0.08 (−0.05, 0.21) | 0.17 | 0.17 | 131 | 137 | −0.01 | −0.13 (−0.38, 0.12) | 0.11 | 0.13 |
| Zimbabwe | SHINE (HIV+) (34) | A not B task | 112 | 94 | 0.03 | −0.04 (−0.32, 0.25) | 0.04 | 0.04 | 36 | 31 | 0.08 | −0.07 (−0.50, 0.36) | 0.04 | 0.07 |
| | | | **2280** | **3932** | | **I² = 0.00, Tau² = 0.00** | | | **1087** | **1543** | | **I² = 0.58, Tau² = 0.01** | | |
| **Fixed** | | | | | | **0.01 (−0.05, 0.06)** | | | | | | **0.01 (−0.07, 0.09)** | | |
| **Random** | | | | | | **0.01 (−0.05, 0.06)** | | | | | | **0.02 (−0.11, 0.15)** | | |

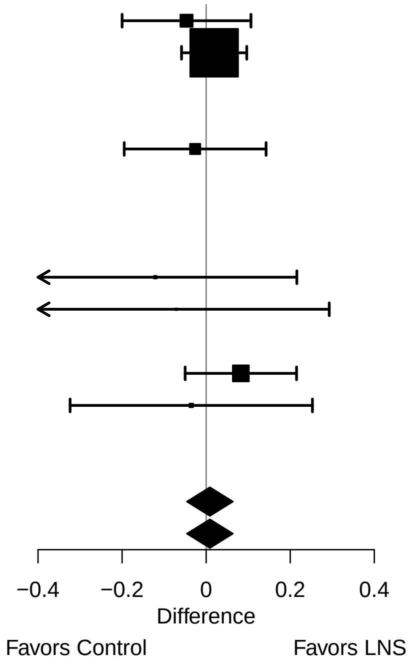

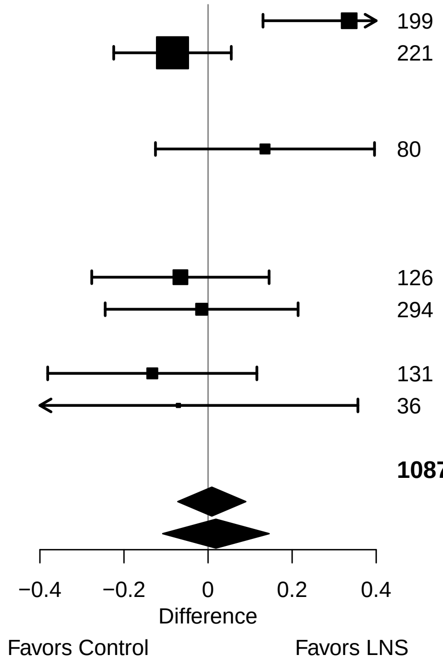

#### Supplemental figure 6L: Mean difference in executive function z-score

#### 6L3: Stratified by Household source water quality

**P−for−interaction = 0.700**
**Difference in MDs = 0.04 (−0.16, 0.24)**

| Country | Trial | Tool | LNS N | Control N | Control Mean | Improved MD (95% CI) | Fixed W | Random W | LNS N | Control N | Control Mean | Unimproved MD (95% CI) | Fixed W | Random W |
|---|---|---|---|---|---|---|---|---|---|---|---|---|---|---|
| Bangladesh | JiVitA−4 (21) | | | | | | | | | | | | | |
| Bangladesh | RDNS (22) | A not B task | | | | | | | | | | | | |
| Bangladesh | WASH−B (23) | A not B task | 474 | 981 | −0.06 | 0.05 (−0.06, 0.15) | 0.51 | 0.51 | 66 | 117 | −0.13 | 0.10 (−0.21, 0.41) | 0.33 | 0.33 |
| Burkina Faso | iLiNS−Zinc (24) | | | | | | | | | | | | | |
| Ghana | GHANA (25) | | | | | | | | | | | | | |
| Ghana | iLiNS−DYADG (26) | A not B task | | | | | | | | | | | | |
| Haiti | HAITI (27) | | | | | | | | | | | | | |
| Kenya | WASH−B (28) | | | | | | | | | | | | | |
| Madagascar | MAHAY (29) | | | | | | | | | | | | | |
| Malawi | iLiNS−DYADM (30) | A not B task | 168 | 335 | 0.02 | −0.09 (−0.28, 0.09) | 0.17 | 0.17 | 14 | 32 | 0.03 | 0.04 (−0.65, 0.73) | 0.07 | 0.07 |
| Malawi | iLiNS−DOSE (31) | A not B task | 380 | 131 | 0.07 | −0.06 (−0.26, 0.14) | 0.15 | 0.15 | 30 | 11 | −0.19 | 0.11 (−0.60, 0.81) | 0.06 | 0.06 |
| Mali | PROMIS CS (32) | | | | | | | | | | | | | |
| Zimbabwe | SHINE (HIV−) (33) | A not B task | 215 | 212 | 0.02 | −0.06 (−0.27, 0.14) | 0.14 | 0.14 | 121 | 108 | −0.01 | −0.02 (−0.29, 0.25) | 0.43 | 0.43 |
| Zimbabwe | SHINE (HIV+) (34) | A not B task | 35 | 38 | 0.03 | −0.08 (−0.53, 0.38) | 0.03 | 0.03 | 23 | 17 | 0.22 | −0.36 (−0.90, 0.18) | 0.11 | 0.11 |
| | | | **1272** | **1697** | | **I² = 0.00, Tau² = 0.00** | | | **254** | **285** | | **I² = 0.00, Tau² = 0.00** | | |
| **Fixed** | | | | | | **−0.01 (−0.09, 0.06)** | | | | | | **−0.01 (−0.18, 0.17)** | | |
| **Random** | | | | | | **−0.01 (−0.09, 0.06)** | | | | | | **−0.01 (−0.18, 0.17)** | | |

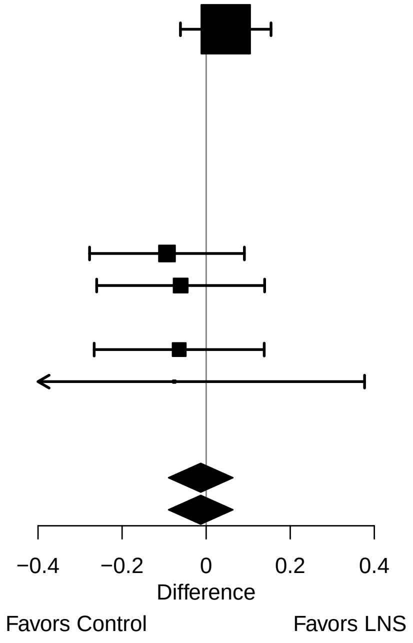

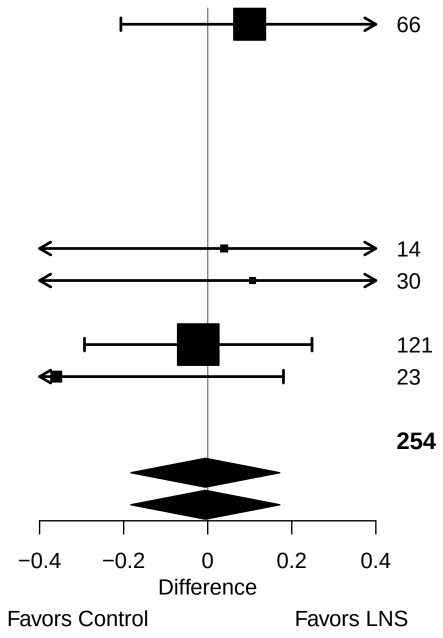

#### Supplemental figure 6L: Mean difference in executive function z-score

#### 6L4: Stratified by Household sanitation

**P−for−interaction = 0.947**
**Difference in MDs = 0.01 (−0.18, 0.19)**

| Country | Trial | Tool | LNS N | Control N | Control Mean | Improved MD (95% CI) | Fixed W | Random W | LNS N | Control N | Control Mean | Unimproved MD (95% CI) | Fixed W | Random W |
|---|---|---|---|---|---|---|---|---|---|---|---|---|---|---|
| Bangladesh | JiVitA−4 (21) | | | | | | | | | | | | | |
| Bangladesh | RDNS (22) | A not B task | 370 | 192 | −0.06 | 0.12 (−0.04, 0.27) | 0.21 | 0.22 | 163 | 74 | −0.09 | 0.06 (−0.18, 0.31) | 0.15 | 0.17 |
| Bangladesh | WASH−B (23) | A not B task | 487 | 998 | −0.04 | 0.04 (−0.07, 0.14) | 0.45 | 0.44 | 27 | 53 | −0.46 | 0.35 (−0.25, 0.94) | 0.03 | 0.05 |
| Burkina Faso | iLiNS−Zinc (24) | | | | | | | | | | | | | |
| Ghana | GHANA (25) | | | | | | | | | | | | | |
| Ghana | iLiNS−DYADG (26) | A not B task | 281 | 561 | −0.02 | 0.04 (−0.10, 0.19) | 0.25 | 0.25 | 8 | 13 | 0.47 | −0.73 (−1.49, 0.02) | 0.02 | 0.03 |
| Haiti | HAITI (27) | | | | | | | | | | | | | |
| Kenya | WASH−B (28) | | | | | | | | | | | | | |
| Madagascar | MAHAY (29) | | | | | | | | | | | | | |
| Malawi | iLiNS−DYADM (30) | A not B task | 18 | 31 | −0.13 | 0.26 (−0.34, 0.87) | 0.01 | 0.01 | 164 | 336 | 0.04 | −0.12 (−0.30, 0.07) | 0.26 | 0.22 |
| Malawi | iLiNS−DOSE (31) | A not B task | 9 | 7 | 0.62 | −0.83 (−1.94, 0.28) | 0.00 | 0.00 | 401 | 135 | 0.02 | −0.01 (−0.21, 0.18) | 0.24 | 0.21 |
| Mali | PROMIS CS (32) | | | | | | | | | | | | | |
| Zimbabwe | SHINE (HIV−) (33) | A not B task | 121 | 102 | 0.13 | −0.18 (−0.47, 0.11) | 0.06 | 0.06 | 214 | 219 | −0.05 | 0.00 (−0.18, 0.19) | 0.25 | 0.22 |
| Zimbabwe | SHINE (HIV+) (34) | A not B task | 23 | 15 | −0.17 | 0.06 (−0.62, 0.75) | 0.01 | 0.01 | 35 | 40 | 0.18 | −0.25 (−0.63, 0.13) | 0.06 | 0.10 |
| | | | **1309** | **1906** | | **$I^2$ = 0.01, $Tau^2$ = 0.00** | | | **1012** | **870** | | **$I^2$ = 0.23, $Tau^2$ = 0.01** | | |
| **Fixed** | | | | | | **0.04 (−0.03, 0.11)** | | | | | | **−0.04 (−0.14, 0.05)** | | |
| **Random** | | | | | | **0.04 (−0.03, 0.12)** | | | | | | **−0.05 (−0.19, 0.09)** | | |

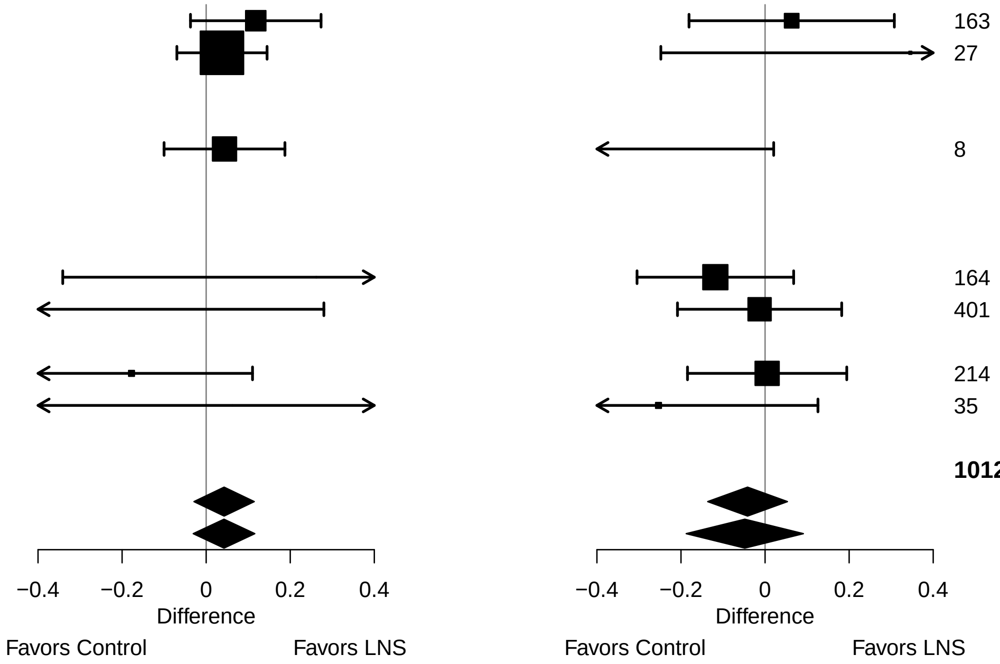

#### Supplemental figure 6L: Mean difference in executive function z-score

##### 6L5: Stratified by Home environment

**P−for−interaction = 0.142**
**Difference in MDs = −0.08 (−0.18, 0.03)**

| Country | Trial | Tool | At least median: LNS N | Control N | Control Mean | MD (95% CI) | Fixed W | Random W | Less than median: LNS N | Control N | Control Mean | MD (95% CI) | Fixed W | Random W |
|---|---|---|---|---|---|---|---|---|---|---|---|---|---|---|
| Bangladesh | JiVitA−4 (21) | | | | | | | | | | | | | |
| Bangladesh | RDNS (22) | A not B task | 385 | 195 | −0.06 | 0.15 (0.02, 0.29) | 0.21 | 0.23 | 146 | 70 | −0.08 | −0.05 (−0.32, 0.21) | 0.09 | 0.09 |
| Bangladesh | WASH−B (23) | A not B task | 623 | 1638 | 0.08 | −0.01 (−0.09, 0.08) | 0.50 | 0.37 | 474 | 1688 | −0.08 | −0.03 (−0.12, 0.07) | 0.66 | 0.66 |
| Burkina Faso | iLiNS−Zinc (24) | | | | | | | | | | | | | |
| Ghana | GHANA (25) | | | | | | | | | | | | | |
| Ghana | iLiNS−DYADG (26) | A not B task | 185 | 363 | −0.04 | 0.11 (−0.07, 0.29) | 0.12 | 0.16 | 105 | 213 | 0.05 | −0.12 (−0.35, 0.11) | 0.12 | 0.12 |
| Haiti | HAITI (27) | | | | | | | | | | | | | |
| Kenya | WASH−B (28) | | | | | | | | | | | | | |
| Madagascar | MAHAY (29) | | | | | | | | | | | | | |
| Malawi | iLiNS−DYADM (30) | A not B task | 120 | 254 | 0.02 | −0.06 (−0.27, 0.15) | 0.09 | 0.13 | 62 | 115 | 0.06 | −0.14 (−0.48, 0.19) | 0.06 | 0.06 |
| Malawi | iLiNS−DOSE (31) | A not B task | 291 | 99 | 0.05 | −0.07 (−0.30, 0.16) | 0.08 | 0.11 | 183 | 65 | 0.14 | −0.18 (−0.46, 0.11) | 0.08 | 0.08 |
| Mali | PROMIS CS (32) | | | | | | | | | | | | | |
| Zimbabwe | SHINE (HIV−) (33) | A not B task | | | | | | | | | | | | |
| Zimbabwe | SHINE (HIV+) (34) | A not B task | | | | | | | | | | | | |
| | | | **1604** | **2549** | | **I² = 0.32, Tau² = 0.00** | | | **970** | **2151** | | **I² = 0.00, Tau² = 0.00** | | |
| **Fixed** | | | | | | **0.03 (−0.03, 0.09)** | | | | | | **−0.06 (−0.14, 0.02)** | | |
| **Random** | | | | | | **0.04 (−0.05, 0.12)** | | | | | | **−0.06 (−0.14, 0.02)** | | |

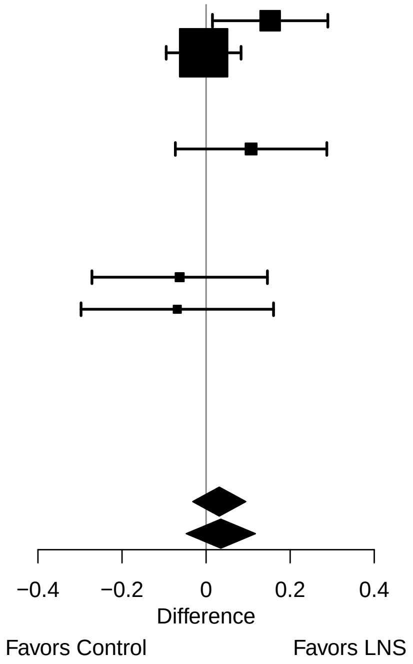

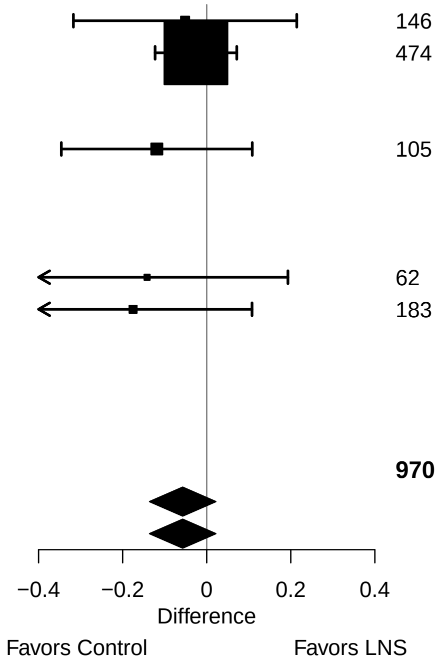

#### Supplemental figure 6L: Mean difference in executive function z-score

##### 6L6: Stratified by Season at the time of assessment

**P−for−interaction = 0.766**
**Difference in MDs = 0.01 (−0.08, 0.11)**

| Country | Trial | Tool | LNS N | Control N | Control Mean | Dry MD (95% CI) | Fixed W | Random W | LNS N | Control N | Control Mean | Rainy MD (95% CI) | Fixed W | Random W |
|---|---|---|---|---|---|---|---|---|---|---|---|---|---|---|
| Bangladesh | JiVitA−4 (21) | | | | | | | | | | | | | |
| Bangladesh | RDNS (22) | A not B task | 242 | 120 | −0.06 | 0.05 (−0.15, 0.26) | 0.09 | 0.09 | 291 | 146 | −0.07 | 0.14 (−0.04, 0.32) | 0.16 | 0.18 |
| Bangladesh | WASH−B (23) | A not B task | 620 | 1921 | −0.01 | 0.02 (−0.08, 0.12) | 0.38 | 0.38 | 477 | 1406 | 0.01 | −0.03 (−0.14, 0.07) | 0.46 | 0.26 |
| Burkina Faso | iLiNS−Zinc (24) | | | | | | | | | | | | | |
| Ghana | GHANA (25) | | | | | | | | | | | | | |
| Ghana | iLiNS−DYADG (26) | A not B task | 172 | 346 | 0.02 | −0.07 (−0.26, 0.12) | 0.10 | 0.10 | 118 | 230 | −0.04 | 0.16 (−0.05, 0.37) | 0.11 | 0.15 |
| Haiti | HAITI (27) | | | | | | | | | | | | | |
| Kenya | WASH−B (28) | | | | | | | | | | | | | |
| Madagascar | MAHAY (29) | | | | | | | | | | | | | |
| Malawi | iLiNS−DYADM (30) | A not B task | 130 | 261 | −0.07 | −0.01 (−0.22, 0.20) | 0.08 | 0.08 | 52 | 107 | 0.29 | −0.28 (−0.60, 0.03) | 0.05 | 0.09 |
| Malawi | iLiNS−DOSE (31) | A not B task | 295 | 89 | 0.21 | −0.25 (−0.48, −0.02) | 0.07 | 0.07 | 176 | 73 | −0.06 | 0.05 (−0.24, 0.33) | 0.06 | 0.10 |
| Mali | PROMIS CS (32) | | | | | | | | | | | | | |
| Zimbabwe | SHINE (HIV−) (33) | A not B task | 563 | 519 | −0.05 | 0.03 (−0.10, 0.16) | 0.21 | 0.21 | 224 | 220 | 0.04 | 0.06 (−0.12, 0.25) | 0.15 | 0.17 |
| Zimbabwe | SHINE (HIV+) (34) | A not B task | 112 | 95 | 0.05 | −0.02 (−0.26, 0.21) | 0.07 | 0.07 | 45 | 38 | 0.06 | −0.28 (−0.79, 0.24) | 0.02 | 0.04 |
| | | | **2134** | **3351** | | **I² = 0.00, Tau² = 0.00** | | | **1383** | **2220** | | **I² = 0.38, Tau² = 0.01** | | |
| **Fixed** | | | | | | **−0.01 (−0.07, 0.05)** | | | | | | **0.02 (−0.05, 0.09)** | | |
| **Random** | | | | | | **−0.01 (−0.07, 0.05)** | | | | | | **0.02 (−0.09, 0.13)** | | |

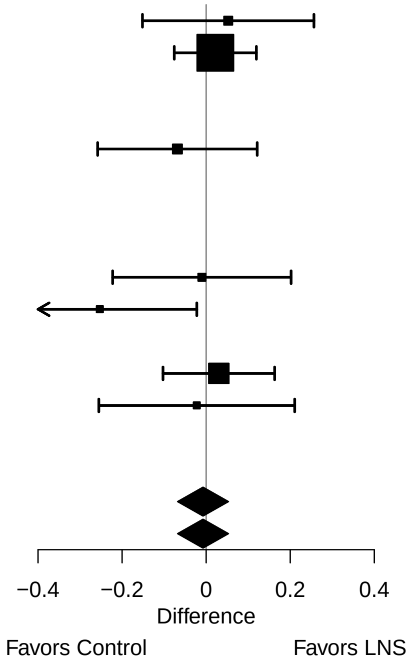

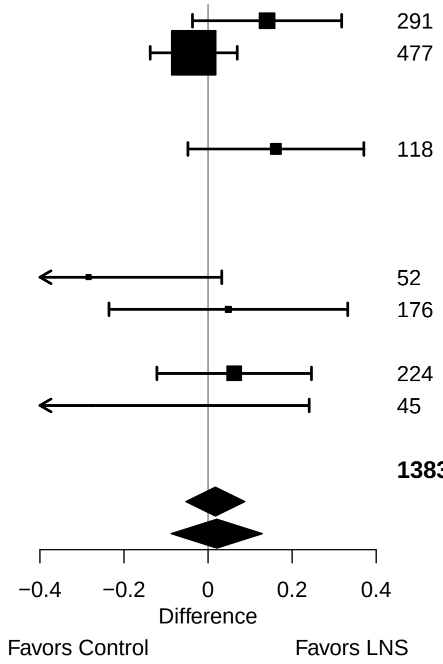

### Supplemental figure 6M: Executive function lowest decile prevalence ratio

#### 6M1: Stratified by Household socio-economic status

**P−for−interaction = 0.389**
**Ratio of PRs = 0.88 (0.66, 1.17)**

| | | | At least median | | | | | | | Less than median | | | | |
|---|---|---|---|---|---|---|---|---|---|---|---|---|---|---|
| **Country** | **Trial** | **Tool** | **LNS N** | **Control N** | **Control Prevalence** | **PR (95% CI)** | **Fixed W** | **Random W** | **LNS N** | **Control N** | **Control Prevalence** | **PR (95% CI)** | **Fixed W** | **Random W** |
| Bangladesh | JiVitA−4 (21) | | | | | | | | | | | | | |
| Bangladesh | RDNS (22) | A not B task | 269 | 133 | 9.8 | 0.80 (0.42, 1.52) | 0.10 | 0.12 | 264 | 133 | 12.0 | 0.94 (0.58, 1.53) | 0.18 | 0.18 |
| Bangladesh | WASH−B (23) | A not B task | 566 | 1691 | 8.8 | 1.17 (0.87, 1.57) | 0.50 | 0.46 | 531 | 1636 | 11.6 | 0.78 (0.58, 1.05) | 0.48 | 0.48 |
| Burkina Faso | iLiNS−Zinc (24) | | | | | | | | | | | | | |
| Ghana | GHANA (25) | | | | | | | | | | | | | |
| Ghana | iLiNS−DYADG (26) | A not B task | 140 | 297 | 13.1 | 0.60 (0.32, 1.13) | 0.11 | 0.12 | 150 | 277 | 9.0 | 1.03 (0.55, 1.93) | 0.11 | 0.11 |
| Haiti | HAITI (27) | | | | | | | | | | | | | |
| Kenya | WASH−B (28) | | | | | | | | | | | | | |
| Madagascar | MAHAY (29) | | | | | | | | | | | | | |
| Malawi | iLiNS−DYADM (30) | A not B task | | | | | | | | | | | | |
| Malawi | iLiNS−DOSE (31) | A not B task | 203 | 72 | 8.3 | 1.00 (0.41, 2.45) | 0.05 | 0.06 | 199 | 69 | 7.2 | 1.73 (0.69, 4.35) | 0.05 | 0.05 |
| Mali | PROMIS CS (32) | | | | | | | | | | | | | |
| Zimbabwe | SHINE (HIV−) (33) | A not B task | 377 | 340 | 12.1 | 0.84 (0.55, 1.28) | 0.24 | 0.25 | 365 | 352 | 11.1 | 0.77 (0.48, 1.22) | 0.19 | 0.19 |
| Zimbabwe | SHINE (HIV+) (34) | A not B task | | | | | | | | | | | | |
| | | | **1555** | **2533** | | **I² = 0.13, Tau² = 0.01** | | | **1509** | **2467** | | **I² = 0.00, Tau² = 0.00** | | |
| **Fixed** | | | | | | **0.96 (0.78, 1.18)** | | | | | | **0.86 (0.70, 1.06)** | | |
| **Random** | | | | | | **0.94 (0.75, 1.18)** | | | | | | **0.86 (0.70, 1.06)** | | |

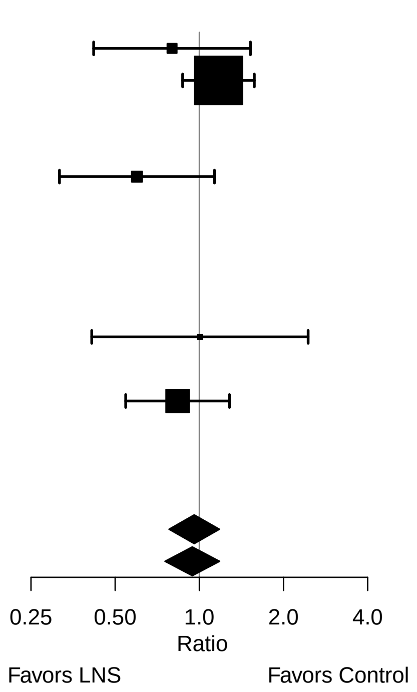

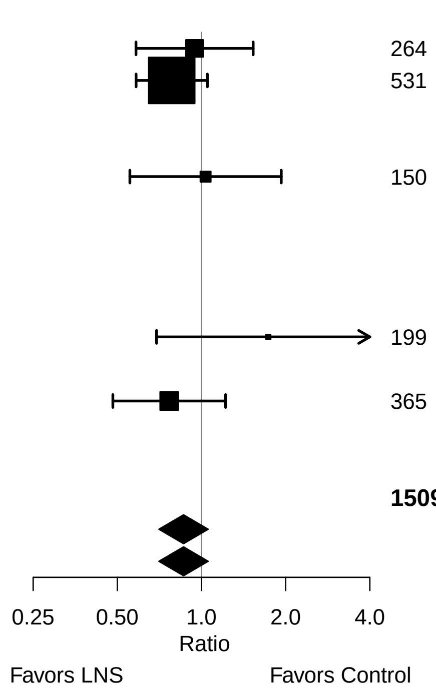

#### Supplemental figure 6M: Executive function lowest decile prevalence ratio

##### 6M2: Stratified by Household food insecurity

**P−for−interaction = 0.640**
**Ratio of PRs = 1.08 (0.79, 1.47)**

| Country | Trial | Tool | LNS N | Control N | Control Prevalence | Mild to secure PR (95% CI) | Fixed W | Random W | LNS N | Control N | Control Prevalence | Moderate to severe PR (95% CI) | Fixed W | Random W |
|---|---|---|---|---|---|---|---|---|---|---|---|---|---|---|
| Bangladesh | JiVitA−4 (21) | | | | | | | | | | | | | |
| Bangladesh | RDNS (22) | A not B task | 334 | 164 | 7.3 | 1.10 (0.60, 2.02) | 0.08 | 0.08 | 199 | 102 | 16.7 | 0.72 (0.45, 1.18) | 0.26 | 0.26 |
| Bangladesh | WASH−B (23) | A not B task | 876 | 2590 | 10.3 | 0.92 (0.72, 1.17) | 0.52 | 0.52 | 221 | 737 | 9.5 | 1.10 (0.72, 1.66) | 0.36 | 0.36 |
| Burkina Faso | iLiNS−Zinc (24) | | | | | | | | | | | | | |
| Ghana | GHANA (25) | | | | | | | | | | | | | |
| Ghana | iLiNS−DYADG (26) | A not B task | 209 | 401 | 12.0 | 0.72 (0.43, 1.20) | 0.12 | 0.12 | 80 | 173 | 9.2 | 0.95 (0.41, 2.21) | 0.09 | 0.09 |
| Haiti | HAITI (27) | | | | | | | | | | | | | |
| Kenya | WASH−B (28) | | | | | | | | | | | | | |
| Madagascar | MAHAY (29) | | | | | | | | | | | | | |
| Malawi | iLiNS−DYADM (30) | A not B task | 55 | 99 | 13.1 | 0.83 (0.33, 2.06) | 0.04 | 0.04 | 126 | 264 | 10.2 | 0.78 (0.39, 1.55) | 0.13 | 0.13 |
| Malawi | iLiNS−DOSE (31) | A not B task | | | | | | | | | | | | |
| Mali | PROMIS CS (32) | | | | | | | | | | | | | |
| Zimbabwe | SHINE (HIV−) (33) | A not B task | 591 | 547 | 11.5 | 0.78 (0.54, 1.11) | 0.24 | 0.24 | 131 | 137 | 11.7 | 0.98 (0.53, 1.83) | 0.16 | 0.16 |
| Zimbabwe | SHINE (HIV+) (34) | A not B task | | | | | | | | | | | | |
| | | | **2065** | **3801** | | **I² = 0.00, Tau² = 0.00** | | | **757** | **1413** | | **I² = 0.00, Tau² = 0.00** | | |
| **Fixed** | | | | | | **0.87 (0.73, 1.04)** | | | | | | **0.91 (0.71, 1.17)** | | |
| **Random** | | | | | | **0.87 (0.73, 1.04)** | | | | | | **0.91 (0.71, 1.17)** | | |

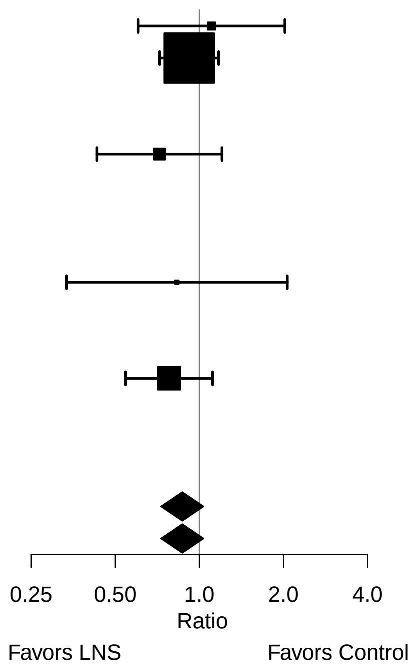

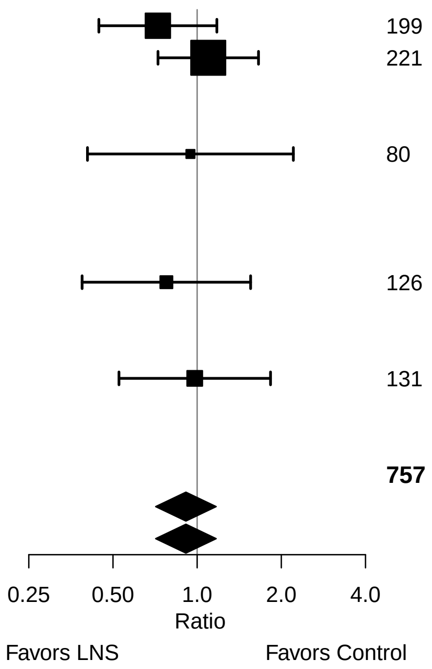

**Supplemental figure 6M: Executive function lowest decile prevalence ratio**

#### 6M3: Stratified by Household source water quality (insufficient comparisons)

**Supplemental figure 6M: Executive function lowest decile prevalence ratio**

#### 6M4: Stratified by Household sanitation (insufficient comparisons)

#### Supplemental figure 6M: Executive function lowest decile prevalence ratio

##### 6M5: Stratified by Home environment

**P−for−interaction = 0.287**
**Ratio of PRs = 1.18 (0.87, 1.59)**

| | | | At least median | | | | | | Less than median | | | | | |
|---|---|---|---|---|---|---|---|---|---|---|---|---|---|---|
| **Country** | **Trial** | **Tool** | **LNS N** | **Control N** | **Control Prevalence** | **PR (95% CI)** | **Fixed W** | **Random W** | **LNS N** | **Control N** | **Control Prevalence** | **PR (95% CI)** | **Fixed W** | **Random W** |
| Bangladesh | JiVitA−4 (21) | | | | | | | | | | | | | |
| Bangladesh | RDNS (22) | A not B task | 385 | 195 | 11.3 | 0.67 (0.41, 1.08) | 0.22 | 0.22 | 146 | 70 | 10.0 | 1.51 (0.78, 2.91) | 0.11 | 0.11 |
| Bangladesh | WASH−B (23) | A not B task | 623 | 1638 | 7.6 | 0.98 (0.72, 1.33) | 0.53 | 0.53 | 474 | 1688 | 12.6 | 1.00 (0.78, 1.29) | 0.73 | 0.73 |
| Burkina Faso | iLiNS−Zinc (24) | | | | | | | | | | | | | |
| Ghana | GHANA (25) | | | | | | | | | | | | | |
| Ghana | iLiNS−DYADG (26) | A not B task | 185 | 363 | 11.8 | 0.68 (0.39, 1.20) | 0.16 | 0.16 | 105 | 213 | 9.9 | 0.97 (0.47, 1.98) | 0.09 | 0.09 |
| Haiti | HAITI (27) | | | | | | | | | | | | | |
| Kenya | WASH−B (28) | | | | | | | | | | | | | |
| Madagascar | MAHAY (29) | | | | | | | | | | | | | |
| Malawi | iLiNS−DYADM (30) | A not B task | 120 | 254 | 8.7 | 0.87 (0.41, 1.82) | 0.09 | 0.09 | 62 | 115 | 15.7 | 0.72 (0.32, 1.63) | 0.07 | 0.07 |
| Malawi | iLiNS−DOSE (31) | A not B task | | | | | | | | | | | | |
| Mali | PROMIS CS (32) | | | | | | | | | | | | | |
| Zimbabwe | SHINE (HIV−) (33) | A not B task | | | | | | | | | | | | |
| Zimbabwe | SHINE (HIV+) (34) | A not B task | | | | | | | | | | | | |
| | | | **1313** | **2450** | | **I² = 0.00, Tau² = 0.00** | | | **787** | **2086** | | **I² = 0.00, Tau² = 0.00** | | |
| **Fixed** | | | | | | **0.84 (0.67, 1.05)** | | | | | | **1.02 (0.82, 1.27)** | | |
| **Random** | | | | | | **0.84 (0.67, 1.05)** | | | | | | **1.02 (0.82, 1.27)** | | |

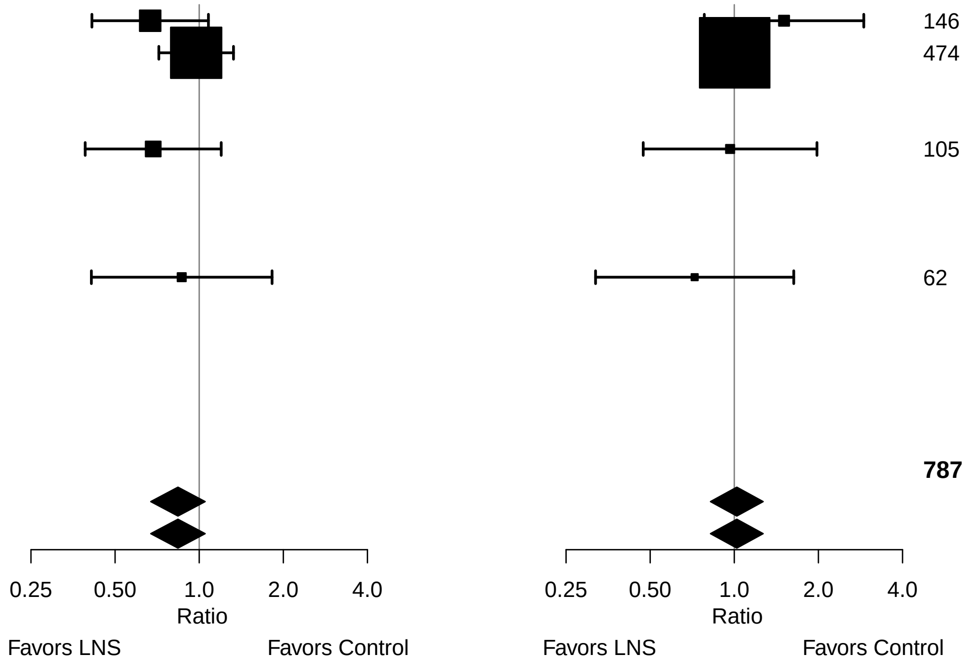

#### Supplemental figure 6M: Executive function lowest decile prevalence ratio

### 6M6: Stratified by Season at the time of assessment

**P−for−interaction = 0.819**
**Ratio of PRs = 1.04 (0.75, 1.44)**

| Country | Trial | Tool | LNS N | Control N | Control Prevalence | Dry PR (95% CI) | Fixed W | Random W | LNS N | Control N | Control Prevalence | Rainy PR (95% CI) | Fixed W | Random W |
|---|---|---|---|---|---|---|---|---|---|---|---|---|---|---|
| Bangladesh | JiVitA−4 (21) | | | | | | | | | | | | | |
| Bangladesh | RDNS (22) | A not B task | 242 | 120 | 11.7 | 0.81 (0.42, 1.57) | 0.10 | 0.10 | 291 | 146 | 10.3 | 0.94 (0.56, 1.57) | 0.23 | 0.23 |
| Bangladesh | WASH−B (23) | A not B task | 620 | 1921 | 10.2 | 0.95 (0.72, 1.26) | 0.52 | 0.52 | 477 | 1406 | 10.1 | 0.95 (0.69, 1.32) | 0.59 | 0.59 |
| Burkina Faso | iLiNS−Zinc (24) | | | | | | | | | | | | | |
| Ghana | GHANA (25) | | | | | | | | | | | | | |
| Ghana | iLiNS−DYADG (26) | A not B task | | | | | | | | | | | | |
| Haiti | HAITI (27) | | | | | | | | | | | | | |
| Kenya | WASH−B (28) | | | | | | | | | | | | | |
| Madagascar | MAHAY (29) | | | | | | | | | | | | | |
| Malawi | iLiNS−DYADM (30) | A not B task | | | | | | | | | | | | |
| Malawi | iLiNS−DOSE (31) | A not B task | | | | | | | | | | | | |
| Mali | PROMIS CS (32) | | | | | | | | | | | | | |
| Zimbabwe | SHINE (HIV−) (33) | A not B task | 563 | 519 | 12.5 | 0.78 (0.56, 1.09) | 0.38 | 0.38 | 224 | 220 | 9.1 | 0.83 (0.46, 1.52) | 0.17 | 0.17 |
| Zimbabwe | SHINE (HIV+) (34) | A not B task | | | | | | | | | | | | |
| | | | **1425** | **2560** | | **I² = 0.00, Tau² = 0.00** | | | **992** | **1772** | | **I² = 0.00, Tau² = 0.00** | | |
| **Fixed** | | | | | | **0.87 (0.71, 1.07)** | | | | | | **0.93 (0.72, 1.19)** | | |
| **Random** | | | | | | **0.87 (0.71, 1.07)** | | | | | | **0.93 (0.72, 1.19)** | | |

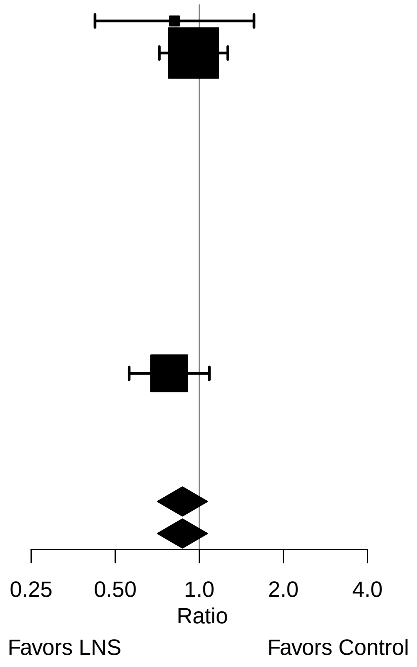

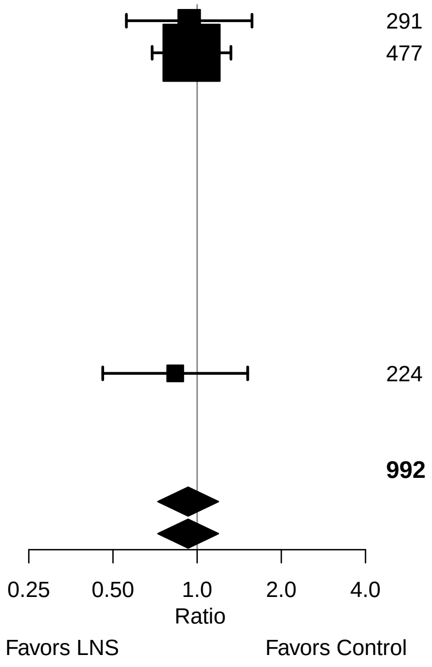

### Supplemental figure 6N: Executive function lowest decile prevalence difference

#### 6N1: Stratified by Household socio-economic status

**P−for−interaction = 0.517**
**Difference in PDs = −0.01 (−0.04, 0.02)**

| Country | Trial | Tool | At least median: LNS N | Control N | Control Prevalence | PD (95% CI) | Fixed W | Random W | Less than median: LNS N | Control N | Control Prevalence | PD (95% CI) | Fixed W | Random W |
|---|---|---|---|---|---|---|---|---|---|---|---|---|---|---|
| Bangladesh | JiVitA−4 (21) | | | | | | | | | | | | | |
| Bangladesh | RDNS (22) | A not B task | 269 | 133 | 9.8 | −0.02 (−0.08, 0.04) | 0.13 | 0.15 | 264 | 133 | 12.0 | −0.01 (−0.06, 0.05) | 0.12 | 0.12 |
| Bangladesh | WASH−B (23) | A not B task | 566 | 1691 | 8.8 | 0.01 (−0.01, 0.04) | 0.50 | 0.45 | 531 | 1636 | 11.6 | −0.03 (−0.05, 0.00) | 0.51 | 0.51 |
| Burkina Faso | iLiNS−Zinc (24) | | | | | | | | | | | | | |
| Ghana | GHANA (25) | | | | | | | | | | | | | |
| Ghana | iLiNS−DYADG (26) | A not B task | 140 | 297 | 13.1 | −0.05 (−0.12, 0.01) | 0.10 | 0.12 | 150 | 277 | 9.0 | 0.00 (−0.05, 0.06) | 0.12 | 0.12 |
| Haiti | HAITI (27) | | | | | | | | | | | | | |
| Kenya | WASH−B (28) | | | | | | | | | | | | | |
| Madagascar | MAHAY (29) | | | | | | | | | | | | | |
| Malawi | iLiNS−DYADM (30) | A not B task | | | | | | | | | | | | |
| Malawi | iLiNS−DOSE (31) | A not B task | 203 | 72 | 8.3 | 0.00 (−0.07, 0.08) | 0.08 | 0.09 | 199 | 69 | 7.2 | 0.05 (−0.03, 0.14) | 0.05 | 0.05 |
| Mali | PROMIS CS (32) | | | | | | | | | | | | | |
| Zimbabwe | SHINE (HIV−) (33) | A not B task | 377 | 340 | 12.1 | −0.02 (−0.07, 0.03) | 0.19 | 0.20 | 365 | 352 | 11.1 | −0.03 (−0.07, 0.02) | 0.20 | 0.20 |
| Zimbabwe | SHINE (HIV+) (34) | A not B task | | | | | | | | | | | | |
| | | | **1555** | **2533** | | **I² = 0.13, Tau² = 0.00** | | | **1509** | **2467** | | **I² = 0.00, Tau² = 0.00** | | |
| **Fixed** | | | | | | **0.00 (−0.02, 0.02)** | | | | | | **−0.02 (−0.04, 0.00)** | | |
| **Random** | | | | | | **−0.01 (−0.03, 0.02)** | | | | | | **−0.02 (−0.04, 0.00)** | | |

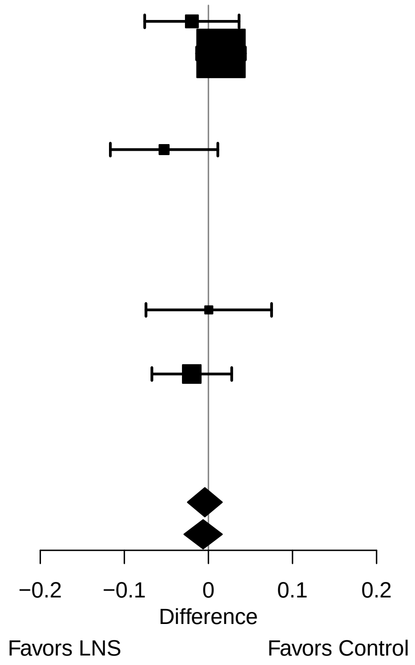

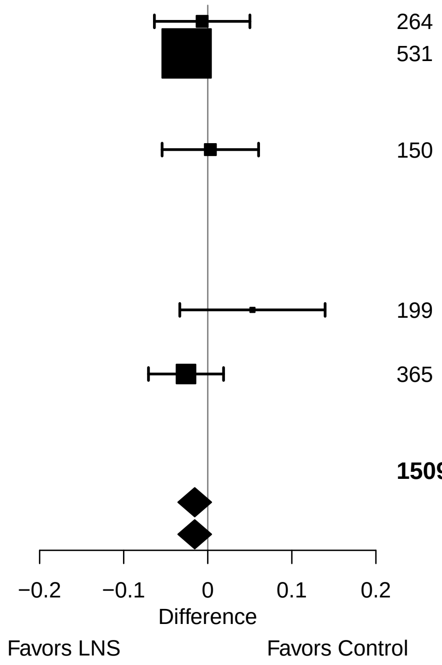

#### Supplemental figure 6N: Executive function lowest decile prevalence difference

##### 6N2: Stratified by Household food insecurity

**P−for−interaction = 0.586**
**Difference in PDs = 0.01 (−0.03, 0.04)**

| Country | Trial | Tool | LNS N | Control N | Control Prevalence | Mild to secure PD (95% CI) | Fixed W | Random W | LNS N | Control N | Control Prevalence | Moderate to severe PD (95% CI) | Fixed W | Random W |
|---|---|---|---|---|---|---|---|---|---|---|---|---|---|---|
| Bangladesh | JiVitA−4 (21) | | | | | | | | | | | | | |
| Bangladesh | RDNS (22) | A not B task | 334 | 164 | 7.3 | 0.01 (−0.04, 0.05) | 0.14 | 0.14 | 199 | 102 | 16.7 | −0.05 (−0.11, 0.02) | 0.16 | 0.16 |
| Bangladesh | WASH−B (23) | A not B task | 876 | 2590 | 10.3 | −0.01 (−0.03, 0.02) | 0.53 | 0.53 | 221 | 737 | 9.5 | 0.01 (−0.03, 0.05) | 0.40 | 0.40 |
| Burkina Faso | iLiNS−Zinc (24) | | | | | | | | | | | | | |
| Ghana | GHANA (25) | | | | | | | | | | | | | |
| Ghana | iLiNS−DYADG (26) | A not B task | 209 | 401 | 12.0 | −0.03 (−0.09, 0.02) | 0.11 | 0.11 | 80 | 173 | 9.2 | 0.00 (−0.08, 0.07) | 0.12 | 0.12 |
| Haiti | HAITI (27) | | | | | | | | | | | | | |
| Kenya | WASH−B (28) | | | | | | | | | | | | | |
| Madagascar | MAHAY (29) | | | | | | | | | | | | | |
| Malawi | iLiNS−DYADM (30) | A not B task | 55 | 99 | 13.1 | −0.02 (−0.13, 0.09) | 0.02 | 0.02 | 126 | 264 | 10.2 | −0.02 (−0.09, 0.04) | 0.18 | 0.18 |
| Malawi | iLiNS−DOSE (31) | A not B task | | | | | | | | | | | | |
| Mali | PROMIS CS (32) | | | | | | | | | | | | | |
| Zimbabwe | SHINE (HIV−) (33) | A not B task | 591 | 547 | 11.5 | −0.03 (−0.06, 0.01) | 0.20 | 0.20 | 131 | 137 | 11.7 | 0.00 (−0.07, 0.07) | 0.14 | 0.14 |
| Zimbabwe | SHINE (HIV+) (34) | A not B task | | | | | | | | | | | | |
| | | | **2065** | **3801** | | **I² = 0.00, Tau² = 0.00** | | | **757** | **1413** | | **I² = 0.00, Tau² = 0.00** | | |
| **Fixed** | | | | | | **−0.01 (−0.03, 0.00)** | | | | | | **−0.01 (−0.04, 0.02)** | | |
| **Random** | | | | | | **−0.01 (−0.03, 0.00)** | | | | | | **−0.01 (−0.04, 0.02)** | | |

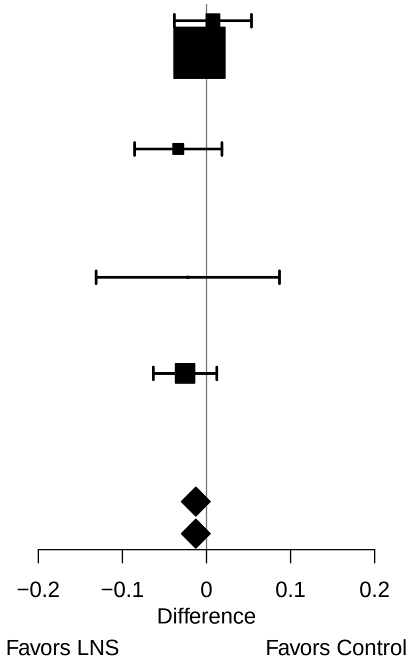

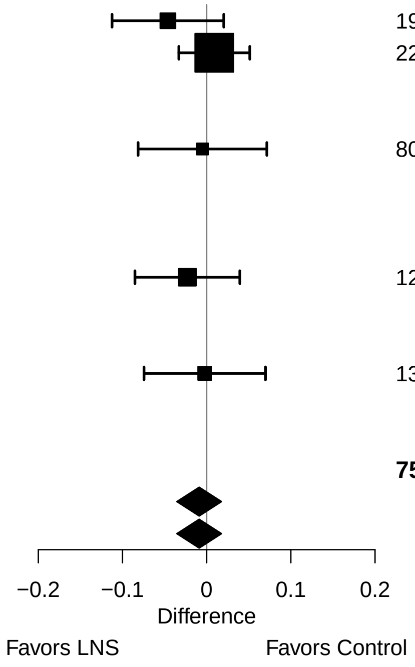

**Supplemental figure 6N: Executive function lowest decile prevalence difference**

#### 6N3: Stratified by Household source water quality (insufficient comparisons)

**Supplemental figure 6N: Executive function lowest decile prevalence difference**

#### 6N4: Stratified by Household sanitation (insufficient comparisons)

#### Supplemental figure 6N: Executive function lowest decile prevalence difference

#### 6N5: Stratified by Home environment

**P−for−interaction = 0.441**
**Difference in PDs = 0.01 (−0.02, 0.05)**

| Country | Trial | Tool | LNS N | Control N | Control Prevalence | At least median PD (95% CI) | Fixed W | Random W | LNS N | Control N | Control Prevalence | Less than median PD (95% CI) | Fixed W | Random W |
|---|---|---|---|---|---|---|---|---|---|---|---|---|---|---|
| Bangladesh | JiVitA−4 (21) | | | | | | | | | | | | | |
| Bangladesh | RDNS (22) | A not B task | 385 | 195 | 11.3 | −0.04 (−0.08, 0.01) | 0.17 | 0.17 | 146 | 70 | 10.0 | 0.05 (−0.02, 0.13) | 0.12 | 0.12 |
| Bangladesh | WASH−B (23) | A not B task | 623 | 1638 | 7.6 | 0.00 (−0.02, 0.02) | 0.63 | 0.63 | 474 | 1688 | 12.6 | 0.00 (−0.03, 0.03) | 0.67 | 0.67 |
| Burkina Faso | iLiNS−Zinc (24) | | | | | | | | | | | | | |
| Ghana | GHANA (25) | | | | | | | | | | | | | |
| Ghana | iLiNS−DYADG (26) | A not B task | 185 | 363 | 11.8 | −0.04 (−0.09, 0.02) | 0.11 | 0.11 | 105 | 213 | 9.9 | 0.00 (−0.07, 0.07) | 0.14 | 0.14 |
| Haiti | HAITI (27) | | | | | | | | | | | | | |
| Kenya | WASH−B (28) | | | | | | | | | | | | | |
| Madagascar | MAHAY (29) | | | | | | | | | | | | | |
| Malawi | iLiNS−DYADM (30) | A not B task | 120 | 254 | 8.7 | −0.01 (−0.07, 0.05) | 0.09 | 0.09 | 62 | 115 | 15.7 | −0.04 (−0.15, 0.06) | 0.06 | 0.06 |
| Malawi | iLiNS−DOSE (31) | A not B task | | | | | | | | | | | | |
| Mali | PROMIS CS (32) | | | | | | | | | | | | | |
| Zimbabwe | SHINE (HIV−) (33) | A not B task | | | | | | | | | | | | |
| Zimbabwe | SHINE (HIV+) (34) | A not B task | | | | | | | | | | | | |
| | | | **1313** | **2450** | | **I² = 0.00, Tau² = 0.00** | | | **787** | **2086** | | **I² = 0.00, Tau² = 0.00** | | |
| **Fixed** | | | | | | **−0.01 (−0.03, 0.01)** | | | | | | **0.00 (−0.02, 0.03)** | | |
| **Random** | | | | | | **−0.01 (−0.03, 0.01)** | | | | | | **0.00 (−0.02, 0.03)** | | |

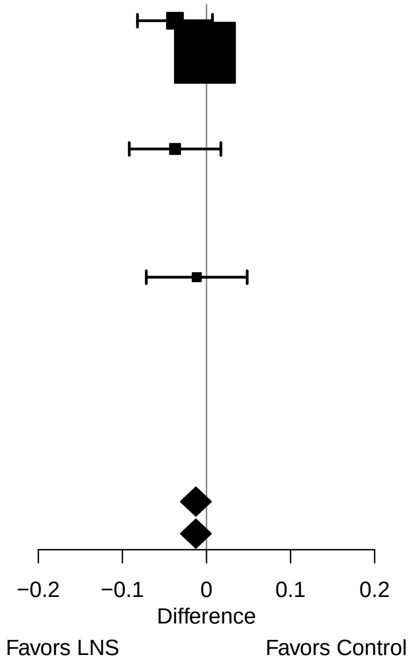

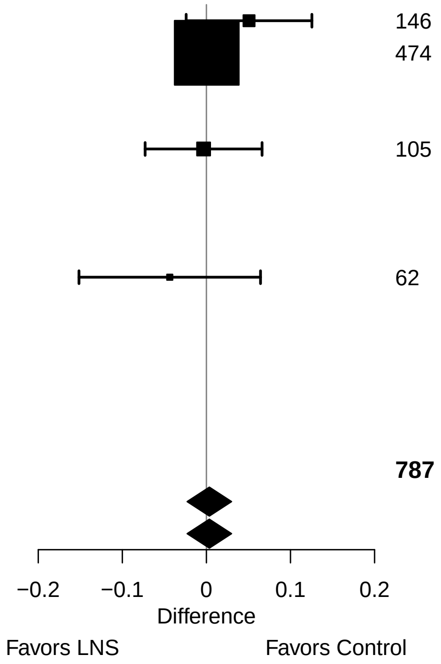

#### Supplemental figure 6N: Executive function lowest decile prevalence difference

#### 6N6: Stratified by Season at the time of assessment

**P−for−interaction = 0.752**
**Difference in PDs = 0.01 (−0.03, 0.04)**

| Country | Trial | Tool | Dry: LNS N | Dry: Control N | Dry: Control Prevalence | Dry: PD (95% CI) | Dry: Fixed W | Dry: Random W | Rainy: LNS N | Rainy: Control N | Rainy: Control Prevalence | Rainy: PD (95% CI) | Rainy: Fixed W | Rainy: Random W |
|---|---|---|---|---|---|---|---|---|---|---|---|---|---|---|
| Bangladesh | JiVitA−4 (21) | | | | | | | | | | | | | |
| Bangladesh | RDNS (22) | A not B task | 242 | 120 | 11.7 | −0.02 (−0.09, 0.05) | 0.09 | 0.09 | 291 | 146 | 10.3 | −0.01 (−0.06, 0.05) | 0.21 | 0.21 |
| Bangladesh | WASH−B (23) | A not B task | 620 | 1921 | 10.2 | 0.00 (−0.03, 0.02) | 0.60 | 0.60 | 477 | 1406 | 10.1 | 0.00 (−0.04, 0.03) | 0.56 | 0.56 |
| Burkina Faso | iLiNS−Zinc (24) | | | | | | | | | | | | | |
| Ghana | GHANA (25) | | | | | | | | | | | | | |
| Ghana | iLiNS−DYADG (26) | A not B task | | | | | | | | | | | | |
| Haiti | HAITI (27) | | | | | | | | | | | | | |
| Kenya | WASH−B (28) | | | | | | | | | | | | | |
| Madagascar | MAHAY (29) | | | | | | | | | | | | | |
| Malawi | iLiNS−DYADM (30) | A not B task | | | | | | | | | | | | |
| Malawi | iLiNS−DOSE (31) | A not B task | | | | | | | | | | | | |
| Mali | PROMIS CS (32) | | | | | | | | | | | | | |
| Zimbabwe | SHINE (HIV−) (33) | A not B task | 563 | 519 | 12.5 | −0.03 (−0.07, 0.01) | 0.32 | 0.32 | 224 | 220 | 9.1 | −0.02 (−0.06, 0.03) | 0.23 | 0.23 |
| Zimbabwe | SHINE (HIV+) (34) | A not B task | | | | | | | | | | | | |
| | | | **1425** | **2560** | | **I² = 0.00, Tau² = 0.00** | | | **992** | **1772** | | **I² = 0.00, Tau² = 0.00** | | |
| **Fixed** | | | | | | **−0.01 (−0.03, 0.01)** | | | | | | **−0.01 (−0.03, 0.02)** | | |
| **Random** | | | | | | **−0.01 (−0.03, 0.01)** | | | | | | **−0.01 (−0.03, 0.02)** | | |

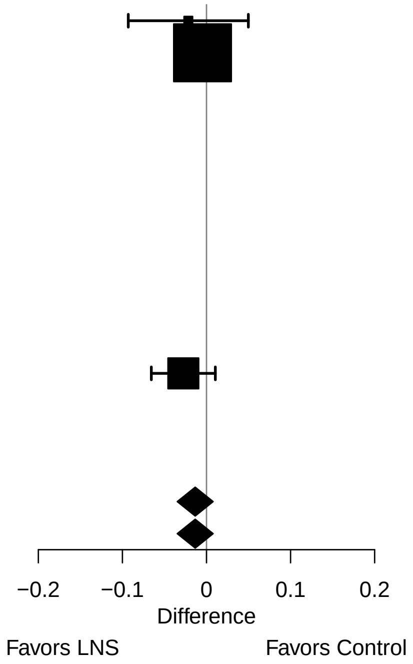

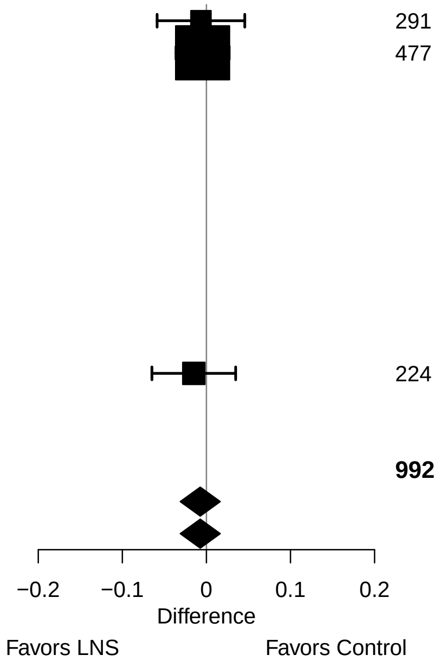

### Supplemental figure 6O: 12-mo walking without support prevalence ratio

#### 6O1: Stratified by Household socio-economic status

**P−for−interaction = 0.721**
**Ratio of PRs = 1.01 (0.94, 1.10)**

**At least median**

| Country | Trial | LNS N | Control N | Control Prevalence | PR (95% CI) | Fixed W | Random W |
|---|---|---|---|---|---|---|---|
| Bangladesh | RDNS (22) | 811 | 396 | 72.0 | 1.22 (1.01, 1.48) | 0.17 | 0.17 |
| Bangladesh | WASH−B (23) | 247 | 735 | 70.3 | 1.28 (1.06, 1.56) | 0.16 | 0.16 |
| Burkina Faso | iLiNS−Zinc (24) | | | | | | |
| Ghana | GHANA (25) | | | | | | |
| Ghana | iLiNS−DYADG (26) | 151 | 349 | 49.9 | 1.11 (0.93, 1.32) | 0.20 | 0.18 |
| Haiti | HAITI (27) | 44 | 31 | 54.8 | 0.91 (0.54, 1.53) | 0.02 | 0.04 |
| Kenya | WASH−B (28) | 240 | 913 | 59.3 | 0.95 (0.81, 1.12) | 0.23 | 0.19 |
| Madagascar | MAHAY (29) | | | | | | |
| Malawi | iLiNS−DYADM (30) | 117 | 227 | 49.8 | 1.24 (1.03, 1.50) | 0.17 | 0.17 |
| Malawi | iLiNS−DOSE (31) | 247 | 81 | 74.1 | 1.67 (1.13, 2.48) | 0.04 | 0.07 |
| Mali | PROMIS CS (32) | 67 | 76 | 73.7 | 0.91 (0.47, 1.76) | 0.01 | 0.03 |
| Zimbabwe | SHINE (HIV−) (33) | | | | | | |
| Zimbabwe | SHINE (HIV+) (34) | | | | | | |
| | | **1924** | **2808** | | **I² = 0.50, Tau² = 0.01** | | |
| **Fixed** | | | | | **1.08 (1.03, 1.14)** | | |
| **Random** | | | | | **1.13 (1.02, 1.25)** | | |

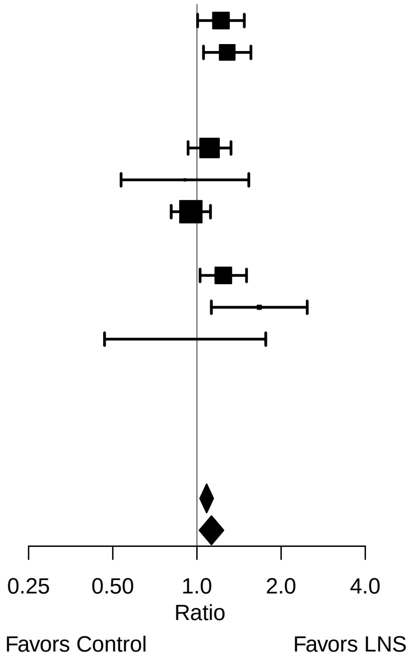

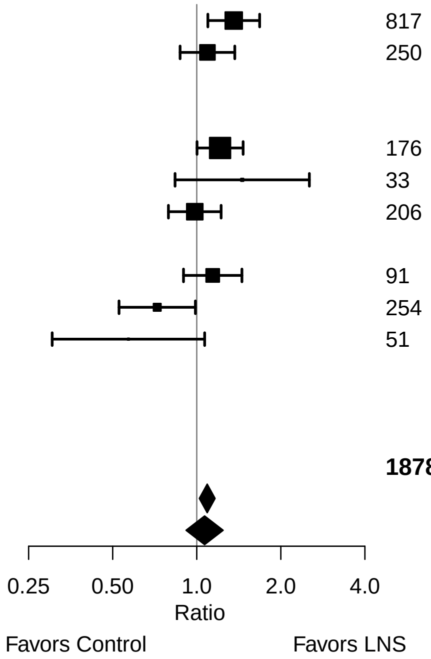

**Less than median**

| Country | Trial | LNS N | Control N | Control Prevalence | PR (95% CI) | Fixed W | Random W |
|---|---|---|---|---|---|---|---|
| Bangladesh | RDNS (22) | 817 | 394 | 76.9 | 1.36 (1.10, 1.68) | 0.18 | 0.15 |
| Bangladesh | WASH−B (23) | 250 | 739 | 78.8 | 1.09 (0.87, 1.37) | 0.16 | 0.15 |
| Burkina Faso | iLiNS−Zinc (24) | | | | | | |
| Ghana | GHANA (25) | | | | | | |
| Ghana | iLiNS−DYADG (26) | 176 | 313 | 56.9 | 1.21 (1.00, 1.47) | 0.22 | 0.16 |
| Haiti | HAITI (27) | 33 | 42 | 66.7 | 1.45 (0.84, 2.53) | 0.03 | 0.06 |
| Kenya | WASH−B (28) | 206 | 689 | 60.5 | 0.98 (0.79, 1.22) | 0.17 | 0.15 |
| Madagascar | MAHAY (29) | | | | | | |
| Malawi | iLiNS−DYADM (30) | 91 | 197 | 52.8 | 1.14 (0.90, 1.45) | 0.14 | 0.14 |
| Malawi | iLiNS−DOSE (31) | 254 | 88 | 59.1 | 0.72 (0.53, 0.99) | 0.08 | 0.12 |
| Mali | PROMIS CS (32) | 51 | 75 | 58.7 | 0.57 (0.30, 1.07) | 0.02 | 0.05 |
| Zimbabwe | SHINE (HIV−) (33) | | | | | | |
| Zimbabwe | SHINE (HIV+) (34) | | | | | | |
| | | **1878** | **2537** | | **I² = 0.56, Tau² = 0.04** | | |
| **Fixed** | | | | | **1.09 (1.02, 1.16)** | | |
| **Random** | | | | | **1.07 (0.92, 1.24)** | | |

#### Supplemental figure 6O: 12-mo walking without support prevalence ratio

#### 6O2: Stratified by Household food insecurity

**P−for−interaction = 0.812**
**Ratio of PRs = 0.99 (0.90, 1.08)**

**Mild to secure**

| Country | Trial | LNS N | Control N | Control Prevalence | PR (95% CI) | Fixed W | Random W |
|---|---|---|---|---|---|---|---|
| Bangladesh | RDNS (22) | 1034 | 483 | 72.9 | 1.19 (0.97, 1.48) | 0.13 | 0.15 |
| Bangladesh | WASH−B (23) | 398 | 1144 | 73.3 | 1.27 (1.08, 1.48) | 0.23 | 0.23 |
| Burkina Faso | iLiNS−Zinc (24) | | | | | | |
| Ghana | GHANA (25) | | | | | | |
| Ghana | iLiNS−DYADG (26) | 230 | 454 | 52.4 | 1.16 (1.00, 1.35) | 0.25 | 0.24 |
| Haiti | HAITI (27) | | | | | | |
| Kenya | WASH−B (28) | 400 | 1448 | 59.0 | 0.96 (0.84, 1.11) | 0.31 | 0.26 |
| Madagascar | MAHAY (29) | | | | | | |
| Malawi | iLiNS−DYADM (30) | 64 | 112 | 56.2 | 1.21 (0.89, 1.66) | 0.06 | 0.08 |
| Malawi | iLiNS−DOSE (31) | 120 | 48 | 70.8 | 1.43 (0.88, 2.33) | 0.02 | 0.04 |
| Mali | PROMIS CS (32) | | | | | | |
| Zimbabwe | SHINE (HIV−) (33) | | | | | | |
| Zimbabwe | SHINE (HIV+) (34) | | | | | | |
| | | **2246** | **3689** | | **I² = 0.45, Tau² = 0.00** | | |
| **Fixed** | | | | | **1.09 (1.03, 1.14)** | | |
| **Random** | | | | | **1.12 (1.03, 1.21)** | | |

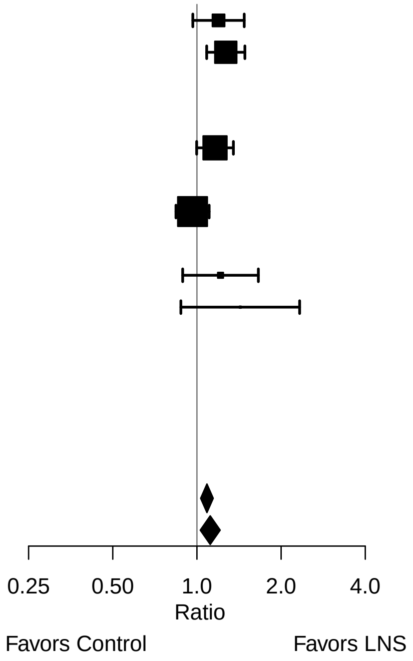

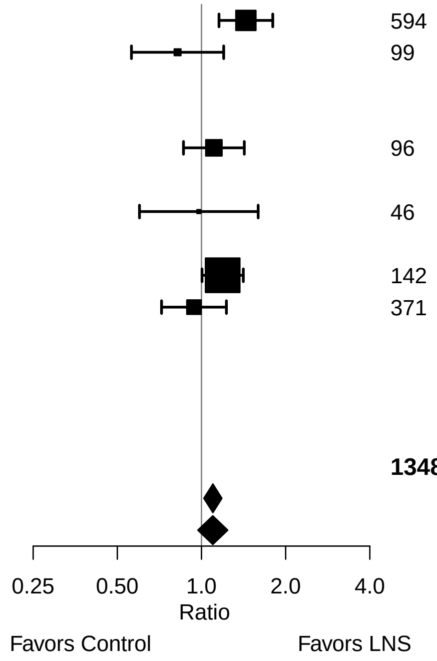

**Moderate to severe**

| Country | Trial | LNS N | Control N | Control Prevalence | PR (95% CI) | Fixed W | Random W |
|---|---|---|---|---|---|---|---|
| Bangladesh | RDNS (22) | 594 | 307 | 76.9 | 1.44 (1.16, 1.80) | 0.21 | 0.21 |
| Bangladesh | WASH−B (23) | 99 | 330 | 79.1 | 0.82 (0.56, 1.20) | 0.07 | 0.11 |
| Burkina Faso | iLiNS−Zinc (24) | | | | | | |
| Ghana | GHANA (25) | | | | | | |
| Ghana | iLiNS−DYADG (26) | 96 | 206 | 54.9 | 1.11 (0.86, 1.42) | 0.17 | 0.18 |
| Haiti | HAITI (27) | | | | | | |
| Kenya | WASH−B (28) | 46 | 153 | 66.7 | 0.98 (0.60, 1.59) | 0.04 | 0.07 |
| Madagascar | MAHAY (29) | | | | | | |
| Malawi | iLiNS−DYADM (30) | 142 | 307 | 48.5 | 1.19 (1.00, 1.41) | 0.36 | 0.26 |
| Malawi | iLiNS−DOSE (31) | 371 | 118 | 61.9 | 0.94 (0.72, 1.23) | 0.15 | 0.17 |
| Mali | PROMIS CS (32) | | | | | | |
| Zimbabwe | SHINE (HIV−) (33) | | | | | | |
| Zimbabwe | SHINE (HIV+) (34) | | | | | | |
| | | **1348** | **1421** | | **I² = 0.46, Tau² = 0.01** | | |
| **Fixed** | | | | | **1.10 (1.02, 1.18)** | | |
| **Random** | | | | | **1.10 (0.97, 1.24)** | | |

#### Supplemental figure 6O: 12-mo walking without support prevalence ratio

### 6O3: Stratified by Household source water quality

**P−for−interaction = 0.190**
**Ratio of PRs = 1.17 (0.92, 1.48)**

| Country | Trial | Improved LNS N | Improved Control N | Improved Control Prevalence | Improved PR (95% CI) | Improved Fixed W | Improved Random W | Unimproved LNS N | Unimproved Control N | Unimproved Control Prevalence | Unimproved PR (95% CI) | Unimproved Fixed W | Unimproved Random W |
|---|---|---|---|---|---|---|---|---|---|---|---|---|---|
| Bangladesh | RDNS (22) | | | | | | | | | | | | |
| Bangladesh | WASH−B (23) | 201 | 428 | 74.5 | 1.17 (0.91, 1.51) | 0.17 | 0.17 | 38 | 56 | 76.8 | 1.36 (0.75, 2.48) | 0.12 | 0.18 |
| Burkina Faso | iLiNS−Zinc (24) | | | | | | | | | | | | |
| Ghana | GHANA (25) | | | | | | | | | | | | |
| Ghana | iLiNS−DYADG (26) | | | | | | | | | | | | |
| Haiti | HAITI (27) | | | | | | | | | | | | |
| Kenya | WASH−B (28) | 118 | 469 | 58.2 | 0.89 (0.68, 1.16) | 0.16 | 0.16 | 84 | 234 | 60.3 | 1.17 (0.89, 1.53) | 0.57 | 0.34 |
| Madagascar | MAHAY (29) | | | | | | | | | | | | |
| Malawi | iLiNS−DYADM (30) | 190 | 395 | 50.9 | 1.17 (1.00, 1.37) | 0.44 | 0.44 | 18 | 28 | 53.6 | 1.56 (0.95, 2.54) | 0.17 | 0.22 |
| Malawi | iLiNS−DOSE (31) | 470 | 157 | 66.2 | 1.09 (0.85, 1.40) | 0.18 | 0.18 | 42 | 15 | 60.0 | 0.95 (0.46, 1.98) | 0.08 | 0.14 |
| Mali | PROMIS CS (32) | 65 | 86 | 68.6 | 0.88 (0.56, 1.40) | 0.05 | 0.05 | 48 | 63 | 61.9 | 0.49 (0.22, 1.12) | 0.06 | 0.12 |
| Zimbabwe | SHINE (HIV−) (33) | | | | | | | | | | | | |
| Zimbabwe | SHINE (HIV+) (34) | | | | | | | | | | | | |
| | | **1044** | **1535** | | **I² = 0.01, Tau² = 0.00** | | | **230** | **396** | | **I² = 0.34, Tau² = 0.07** | | |
| **Fixed** | | | | | **1.09 (0.98, 1.21)** | | | | | | **1.17 (0.95, 1.43)** | | |
| **Random** | | | | | **1.09 (0.98, 1.21)** | | | | | | **1.12 (0.80, 1.57)** | | |

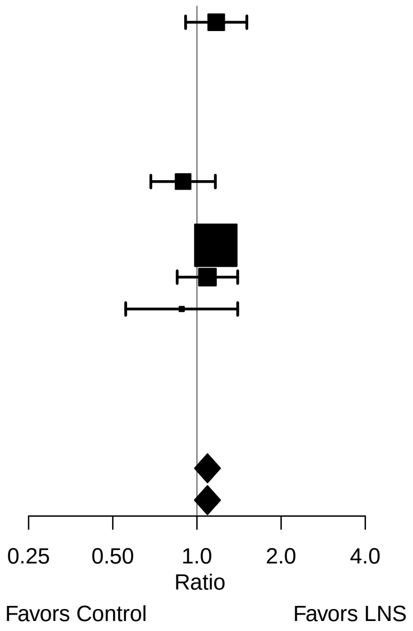

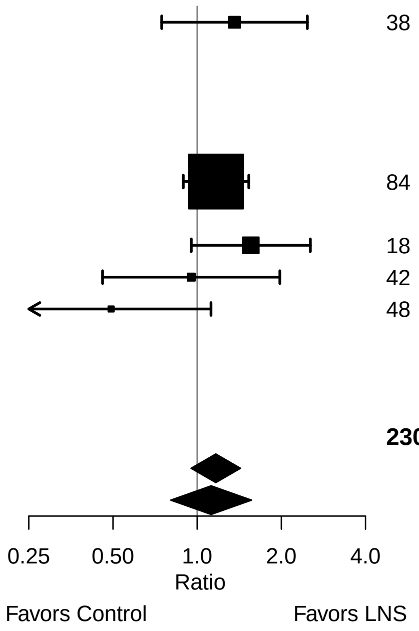

#### Supplemental figure 6O: 12-mo walking without support prevalence ratio

##### 6O4: Stratified by Household sanitation

**P−for−interaction = 0.336**
**Ratio of PRs = 0.94 (0.84, 1.06)**

| | | Improved | | | | | | Unimproved | | | | | |
|---|---|---|---|---|---|---|---|---|---|---|---|---|---|
| **Country** | **Trial** | **LNS N** | **Control N** | **Control Prevalence** | **PR (95% CI)** | **Fixed W** | **Random W** | **LNS N** | **Control N** | **Control Prevalence** | **PR (95% CI)** | **Fixed W** | **Random W** |
| Bangladesh | RDNS (22) | 1149 | 563 | 73.4 | 1.27 (1.02, 1.59) | 0.60 | 0.40 | 477 | 226 | 77.0 | 1.29 (1.00, 1.67) | 0.20 | 0.25 |
| Bangladesh | WASH−B (23) | | | | | | | | | | | | |
| Burkina Faso | iLiNS−Zinc (24) | | | | | | | | | | | | |
| Ghana | GHANA (25) | | | | | | | | | | | | |
| Ghana | iLiNS−DYADG (26) | | | | | | | | | | | | |
| Haiti | HAITI (27) | | | | | | | | | | | | |
| Kenya | WASH−B (28) | 35 | 106 | 58.5 | 1.17 (0.72, 1.89) | 0.13 | 0.19 | 167 | 598 | 59.0 | 0.96 (0.77, 1.20) | 0.27 | 0.30 |
| Madagascar | MAHAY (29) | | | | | | | | | | | | |
| Malawi | iLiNS−DYADM (30) | 19 | 41 | 43.9 | 0.94 (0.57, 1.56) | 0.11 | 0.18 | 189 | 382 | 51.8 | 1.23 (1.05, 1.44) | 0.52 | 0.43 |
| Malawi | iLiNS−DOSE (31) | | | | | | | | | | | | |
| Mali | PROMIS CS (32) | 87 | 108 | 63.0 | 0.65 (0.42, 1.00) | 0.16 | 0.22 | 27 | 38 | 73.7 | 0.84 (0.28, 2.57) | 0.01 | 0.02 |
| Zimbabwe | SHINE (HIV−) (33) | | | | | | | | | | | | |
| Zimbabwe | SHINE (HIV+) (34) | | | | | | | | | | | | |
| | | **1290** | **818** | | **I² = 0.49, Tau² = 0.03** | | | **860** | **1244** | | **I² = 0.51, Tau² = 0.01** | | |
| **Fixed** | | | | | **1.07 (1.00, 1.15)** | | | | | | **1.07 (0.99, 1.16)** | | |
| **Random** | | | | | **1.04 (0.84, 1.28)** | | | | | | **1.09 (0.96, 1.24)** | | |

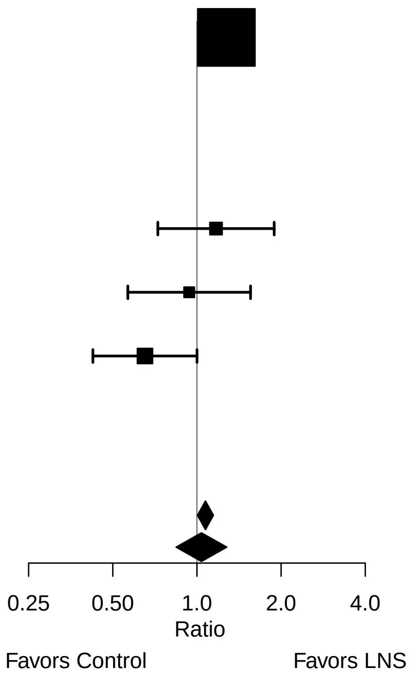

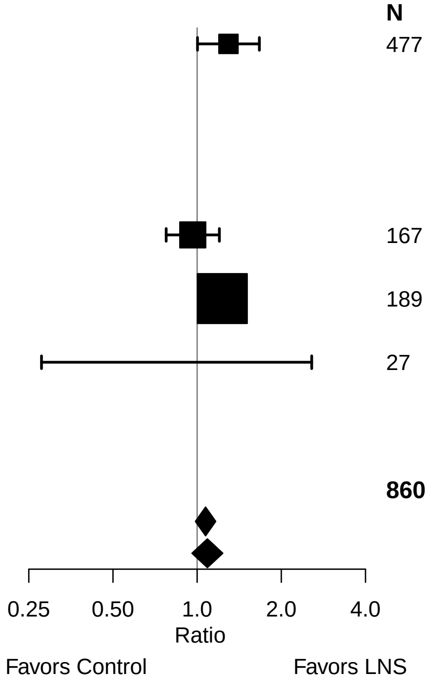

#### Supplemental figure 6O: 12-mo walking without support prevalence ratio

##### 6O5: Stratified by Home environment

**P−for−interaction = 0.458**
**Ratio of PRs = 0.95 (0.84, 1.08)**

| | | At least median | | | | | | | Less than median | | | | |
|---|---|---|---|---|---|---|---|---|---|---|---|---|---|
| **Country** | **Trial** | **LNS N** | **Control N** | **Control Prevalence** | **PR (95% CI)** | **Fixed W** | **Random W** | **LNS N** | **Control N** | **Control Prevalence** | **PR (95% CI)** | **Fixed W** | **Random W** |
| Bangladesh | RDNS (22) | 1158 | 560 | 74.5 | 1.34 (1.09, 1.64) | 0.13 | 0.15 | 448 | 217 | 74.7 | 1.18 (0.90, 1.55) | 0.15 | 0.15 |
| Bangladesh | WASH−B (23) | 243 | 689 | 73.9 | 1.32 (1.10, 1.58) | 0.17 | 0.18 | 231 | 717 | 75.2 | 1.12 (0.90, 1.38) | 0.24 | 0.24 |
| Burkina Faso | iLiNS−Zinc (24) | | | | | | | | | | | | |
| Ghana | GHANA (25) | | | | | | | | | | | | |
| Ghana | iLiNS−DYADG (26) | 198 | 373 | 50.4 | 1.16 (0.99, 1.36) | 0.23 | 0.20 | 110 | 233 | 57.5 | 1.07 (0.83, 1.38) | 0.17 | 0.17 |
| Haiti | HAITI (27) | | | | | | | | | | | | |
| Kenya | WASH−B (28) | 248 | 834 | 55.6 | 0.93 (0.79, 1.09) | 0.23 | 0.20 | 164 | 608 | 65.5 | 1.09 (0.87, 1.38) | 0.20 | 0.20 |
| Madagascar | MAHAY (29) | | | | | | | | | | | | |
| Malawi | iLiNS−DYADM (30) | 130 | 278 | 49.3 | 1.17 (0.97, 1.40) | 0.17 | 0.17 | 72 | 136 | 55.1 | 1.30 (0.99, 1.70) | 0.15 | 0.15 |
| Malawi | iLiNS−DOSE (31) | 306 | 109 | 65.1 | 1.15 (0.86, 1.54) | 0.07 | 0.10 | 224 | 67 | 59.7 | 0.84 (0.60, 1.19) | 0.09 | 0.09 |
| Mali | PROMIS CS (32) | | | | | | | | | | | | |
| Zimbabwe | SHINE (HIV−) (33) | | | | | | | | | | | | |
| Zimbabwe | SHINE (HIV+) (34) | | | | | | | | | | | | |
| | | **2283** | **2843** | | **I² = 0.56, Tau² = 0.01** | | | **1249** | **1978** | | **I² = 0.00, Tau² = 0.00** | | |
| **Fixed** | | | | | **1.15 (1.07, 1.24)** | | | | | | **1.11 (1.00, 1.23)** | | |
| **Random** | | | | | **1.16 (1.04, 1.30)** | | | | | | **1.11 (1.00, 1.23)** | | |

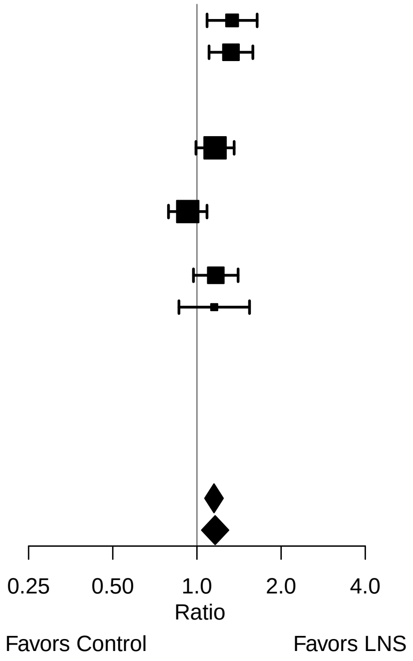

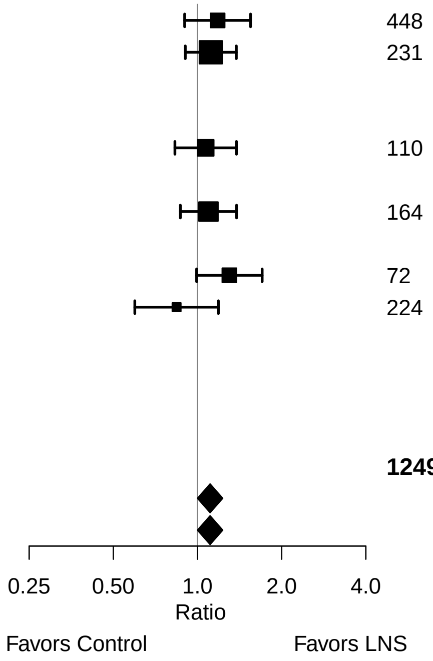

#### Supplemental figure 6O: 12-mo walking without support prevalence ratio

#### 6O6: Stratified by Season at the time of assessment

**P−for−interaction = 0.845**
**Ratio of PRs = 0.99 (0.88, 1.11)**

| Country | Trial | Dry LNS N | Dry Control N | Dry Control Prevalence | Dry PR (95% CI) | Dry Fixed W | Dry Random W | Rainy LNS N | Rainy Control N | Rainy Control Prevalence | Rainy PR (95% CI) | Rainy Fixed W | Rainy Random W |
|---|---|---|---|---|---|---|---|---|---|---|---|---|---|
| Bangladesh | RDNS (22) | 899 | 430 | 77.4 | 1.28 (1.04, 1.59) | 0.15 | 0.16 | 729 | 360 | 70.8 | 1.28 (1.03, 1.60) | 0.14 | 0.14 |
| Bangladesh | WASH−B (23) | 267 | 774 | 72.6 | 1.19 (0.99, 1.43) | 0.19 | 0.18 | 230 | 700 | 76.7 | 1.21 (0.99, 1.49) | 0.17 | 0.17 |
| Burkina Faso | iLiNS−Zinc (24) | | | | | | | | | | | | |
| Ghana | GHANA (25) | 53 | 51 | 78.4 | 2.01 (1.10, 3.69) | 0.02 | 0.03 | 37 | 36 | 69.4 | 1.33 (0.71, 2.49) | 0.02 | 0.02 |
| Ghana | iLiNS−DYADG (26) | 163 | 320 | 48.4 | 1.15 (0.98, 1.36) | 0.25 | 0.21 | 164 | 343 | 57.4 | 1.13 (0.93, 1.38) | 0.17 | 0.17 |
| Haiti | HAITI (27) | | | | | | | | | | | | |
| Kenya | WASH−B (28) | 131 | 461 | 62.5 | 0.92 (0.69, 1.22) | 0.08 | 0.11 | 316 | 1143 | 58.6 | 0.99 (0.86, 1.13) | 0.36 | 0.36 |
| Madagascar | MAHAY (29) | | | | | | | | | | | | |
| Malawi | iLiNS−DYADM (30) | 150 | 311 | 53.7 | 1.24 (1.03, 1.49) | 0.20 | 0.19 | 58 | 115 | 44.3 | 1.12 (0.86, 1.45) | 0.10 | 0.10 |
| Malawi | iLiNS−DOSE (31) | 437 | 152 | 65.8 | 1.04 (0.81, 1.34) | 0.10 | 0.13 | 165 | 48 | 64.6 | 1.04 (0.68, 1.61) | 0.04 | 0.04 |
| Mali | PROMIS CS (32) | | | | | | | | | | | | |
| Zimbabwe | SHINE (HIV−) (33) | | | | | | | | | | | | |
| Zimbabwe | SHINE (HIV+) (34) | | | | | | | | | | | | |
| | | **2100** | **2499** | | **I² = 0.23, Tau² = 0.01** | | | **1699** | **2745** | | **I² = 0.00, Tau² = 0.00** | | |
| **Fixed** | | | | | **1.17 (1.08, 1.27)** | | | | | | **1.11 (1.02, 1.20)** | | |
| **Random** | | | | | **1.17 (1.05, 1.31)** | | | | | | **1.11 (1.02, 1.20)** | | |

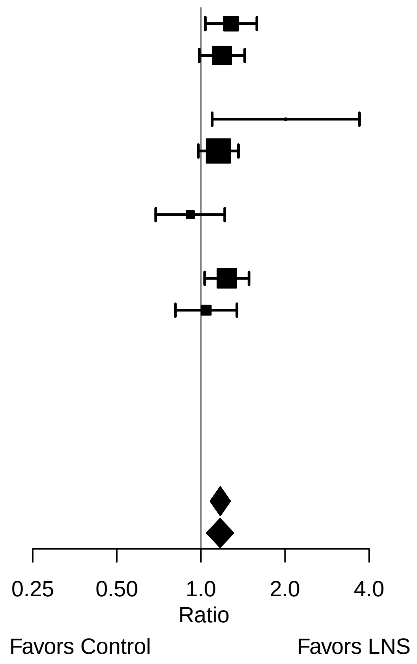

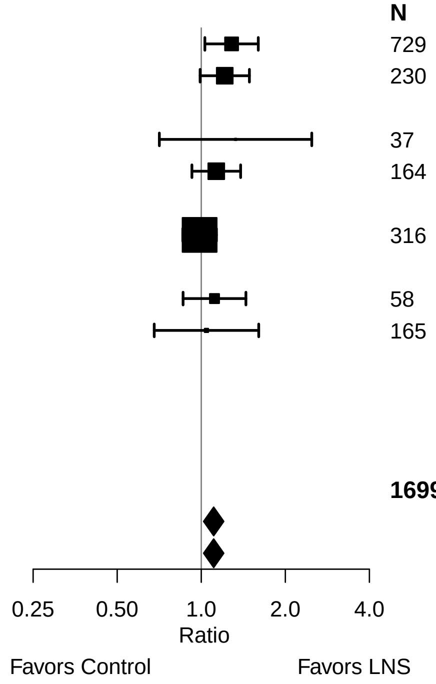

### Supplemental figure 6P: 12-mo walking without support prevalence difference

#### 6P1: Stratified by Household socio-economic status

**P−for−interaction = 0.521**
**Difference in PDs = −0.01 (−0.05, 0.02)**

| Country | Trial | At least median: LNS N | Control N | Control Prevalence | PD (95% CI) | Fixed W | Random W | Less than median: LNS N | Control N | Control Prevalence | PD (95% CI) | Fixed W | Random W |
|---|---|---|---|---|---|---|---|---|---|---|---|---|---|
| Bangladesh | RDNS (22) | 811 | 396 | 72.0 | 0.06 (0.00, 0.12) | 0.28 | 0.21 | 817 | 394 | 76.9 | 0.08 (0.03, 0.13) | 0.31 | 0.18 |
| Bangladesh | WASH−B (23) | 247 | 735 | 70.3 | 0.08 (0.01, 0.15) | 0.19 | 0.18 | 250 | 739 | 78.8 | 0.02 (−0.03, 0.07) | 0.32 | 0.18 |
| Burkina Faso | iLiNS−Zinc (24) | | | | | | | | | | | | |
| Ghana | GHANA (25) | | | | | | | | | | | | |
| Ghana | iLiNS−DYADG (26) | 151 | 349 | 49.9 | 0.05 (−0.04, 0.15) | 0.10 | 0.13 | 176 | 313 | 56.9 | 0.09 (0.00, 0.18) | 0.10 | 0.14 |
| Haiti | HAITI (27) | 44 | 31 | 54.8 | −0.04 (−0.27, 0.19) | 0.02 | 0.03 | 33 | 42 | 66.7 | 0.15 (−0.07, 0.38) | 0.02 | 0.05 |
| Kenya | WASH−B (28) | 240 | 913 | 59.3 | −0.02 (−0.08, 0.04) | 0.24 | 0.19 | 206 | 689 | 60.5 | −0.01 (−0.09, 0.08) | 0.12 | 0.15 |
| Madagascar | MAHAY (29) | | | | | | | | | | | | |
| Malawi | iLiNS−DYADM (30) | 117 | 227 | 49.8 | 0.12 (0.01, 0.23) | 0.08 | 0.11 | 91 | 197 | 52.8 | 0.07 (−0.06, 0.19) | 0.05 | 0.11 |
| Malawi | iLiNS−DOSE (31) | 247 | 81 | 74.1 | 0.17 (0.05, 0.30) | 0.06 | 0.09 | 254 | 88 | 59.1 | −0.11 (−0.23, 0.00) | 0.06 | 0.12 |
| Mali | PROMIS CS (32) | 67 | 76 | 73.7 | −0.02 (−0.19, 0.14) | 0.03 | 0.06 | 51 | 75 | 58.7 | −0.18 (−0.36, 0.00) | 0.03 | 0.07 |
| Zimbabwe | SHINE (HIV−) (33) | | | | | | | | | | | | |
| Zimbabwe | SHINE (HIV+) (34) | | | | | | | | | | | | |
| | | **1924** | **2808** | | **I² = 0.45, Tau² = 0.00** | | | **1878** | **2537** | | **I² = 0.59, Tau² = 0.00** | | |
| **Fixed** | | | | | **0.04 (0.02, 0.07)** | | | | | | **0.03 (0.01, 0.06)** | | |
| **Random** | | | | | **0.05 (0.01, 0.09)** | | | | | | **0.02 (−0.03, 0.08)** | | |

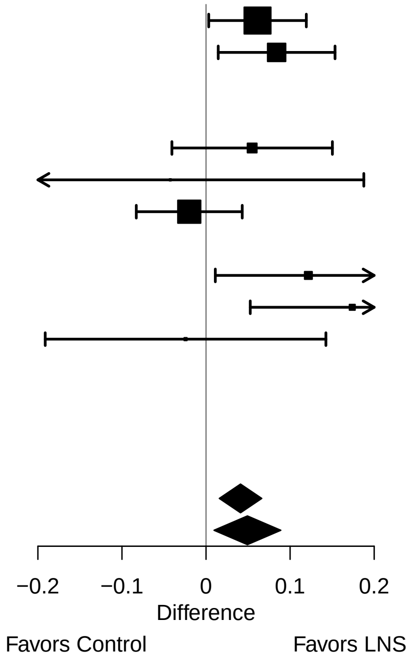

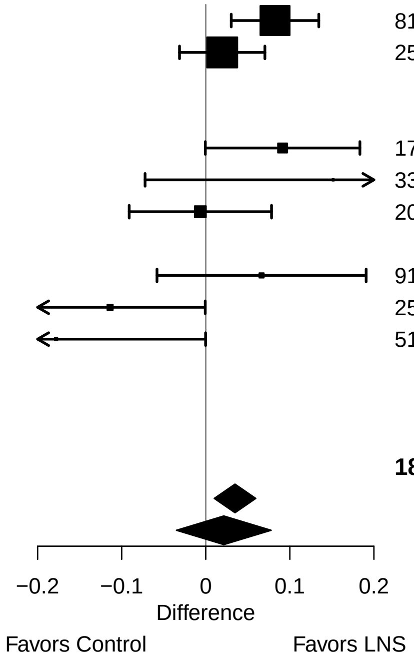

#### Supplemental figure 6P: 12-mo walking without support prevalence difference

#### 6P2: Stratified by Household food insecurity

P−for−interaction = 0.547
Difference in PDs = −0.01 (−0.05, 0.03)

| Country | Trial | Mild to secure: LNS N | Control N | Control Prevalence | PD (95% CI) | Fixed W | Random W | Moderate to severe: LNS N | Control N | Control Prevalence | PD (95% CI) | Fixed W | Random W |
|---|---|---|---|---|---|---|---|---|---|---|---|---|---|
| Bangladesh | RDNS (22) | 1034 | 483 | 72.9 | 0.05 (−0.01, 0.11) | 0.22 | 0.22 | 594 | 307 | 76.9 | 0.10 (0.04, 0.16) | 0.37 | 0.27 |
| Bangladesh | WASH−B (23) | 398 | 1144 | 73.3 | 0.07 (0.02, 0.12) | 0.32 | 0.29 | 99 | 330 | 79.1 | −0.04 (−0.11, 0.03) | 0.26 | 0.23 |
| Burkina Faso | iLiNS−Zinc (24) | | | | | | | | | | | | |
| Ghana | GHANA (25) | | | | | | | | | | | | |
| Ghana | iLiNS−DYADG (26) | 230 | 454 | 52.4 | 0.08 (0.00, 0.16) | 0.13 | 0.15 | 96 | 206 | 54.9 | 0.05 (−0.07, 0.17) | 0.08 | 0.12 |
| Haiti | HAITI (27) | | | | | | | | | | | | |
| Kenya | WASH−B (28) | 400 | 1448 | 59.0 | −0.01 (−0.07, 0.04) | 0.27 | 0.26 | 46 | 153 | 66.7 | −0.01 (−0.17, 0.15) | 0.05 | 0.08 |
| Madagascar | MAHAY (29) | | | | | | | | | | | | |
| Malawi | iLiNS−DYADM (30) | 64 | 112 | 56.2 | 0.09 (−0.06, 0.25) | 0.03 | 0.05 | 142 | 307 | 48.5 | 0.10 (0.00, 0.20) | 0.12 | 0.15 |
| Malawi | iLiNS−DOSE (31) | 120 | 48 | 70.8 | 0.12 (−0.04, 0.29) | 0.03 | 0.04 | 371 | 118 | 61.9 | −0.02 (−0.12, 0.08) | 0.12 | 0.15 |
| Mali | PROMIS CS (32) | | | | | | | | | | | | |
| Zimbabwe | SHINE (HIV−) (33) | | | | | | | | | | | | |
| Zimbabwe | SHINE (HIV+) (34) | | | | | | | | | | | | |
| | | **2246** | **3689** | | **I² = 0.29, Tau² = 0.00** | | | **1348** | **1421** | | **I² = 0.53, Tau² = 0.00** | | |
| **Fixed** | | | | | **0.04 (0.02, 0.06)** | | | | | | **0.04 (0.01, 0.07)** | | |
| **Random** | | | | | **0.04 (0.01, 0.07)** | | | | | | **0.03 (−0.01, 0.08)** | | |

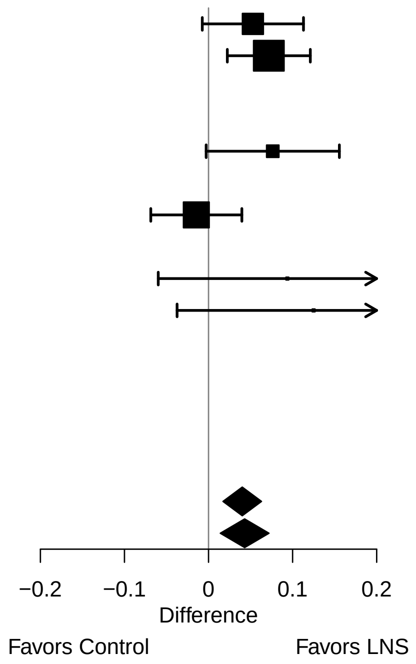

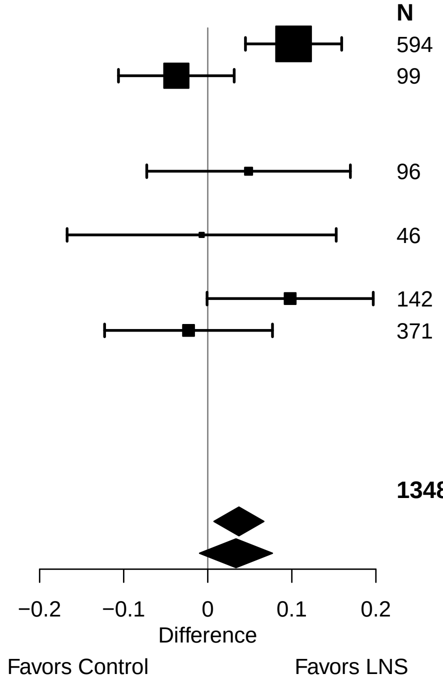

#### Supplemental figure 6P: 12-mo walking without support prevalence difference

#### 6P3: Stratified by Household source water quality

**P−for−interaction = 0.476**
**Difference in PDs = 0.03 (−0.06, 0.13)**

| Country | Trial | Improved LNS N | Improved Control N | Improved Control Prevalence | Improved PD (95% CI) | Improved Fixed W | Improved Random W | Unimproved LNS N | Unimproved Control N | Unimproved Control Prevalence | Unimproved PD (95% CI) | Unimproved Fixed W | Unimproved Random W |
|---|---|---|---|---|---|---|---|---|---|---|---|---|---|
| Bangladesh | RDNS (22) | | | | | | | | | | | | |
| Bangladesh | WASH−B (23) | 201 | 428 | 74.5 | 0.04 (−0.03, 0.11) | 0.32 | 0.30 | 38 | 56 | 76.8 | 0.08 (−0.07, 0.23) | 0.27 | 0.26 |
| Burkina Faso | iLiNS−Zinc (24) | | | | | | | | | | | | |
| Ghana | GHANA (25) | | | | | | | | | | | | |
| Ghana | iLiNS−DYADG (26) | | | | | | | | | | | | |
| Haiti | HAITI (27) | | | | | | | | | | | | |
| Kenya | WASH−B (28) | 118 | 469 | 58.2 | −0.05 (−0.15, 0.06) | 0.16 | 0.17 | 84 | 234 | 60.3 | 0.07 (−0.05, 0.19) | 0.45 | 0.30 |
| Madagascar | MAHAY (29) | | | | | | | | | | | | |
| Malawi | iLiNS−DYADM (30) | 190 | 395 | 50.9 | 0.08 (0.00, 0.17) | 0.22 | 0.22 | 18 | 28 | 53.6 | 0.26 (−0.03, 0.55) | 0.07 | 0.13 |
| Malawi | iLiNS−DOSE (31) | 470 | 157 | 66.2 | 0.03 (−0.06, 0.12) | 0.21 | 0.21 | 42 | 15 | 60.0 | −0.02 (−0.31, 0.27) | 0.07 | 0.13 |
| Mali | PROMIS CS (32) | 65 | 86 | 68.6 | −0.04 (−0.17, 0.10) | 0.09 | 0.10 | 48 | 63 | 61.9 | −0.19 (−0.41, 0.03) | 0.13 | 0.19 |
| Zimbabwe | SHINE (HIV−) (33) | | | | | | | | | | | | |
| Zimbabwe | SHINE (HIV+) (34) | | | | | | | | | | | | |
| | | **1044** | **1535** | | **I² = 0.14, Tau² = 0.00** | | | **230** | **396** | | **I² = 0.45, Tau² = 0.01** | | |
| **Fixed** | | | | | **0.03 (−0.01, 0.07)** | | | | | | **0.05 (−0.03, 0.12)** | | |
| **Random** | | | | | **0.03 (−0.02, 0.07)** | | | | | | **0.04 (−0.09, 0.17)** | | |

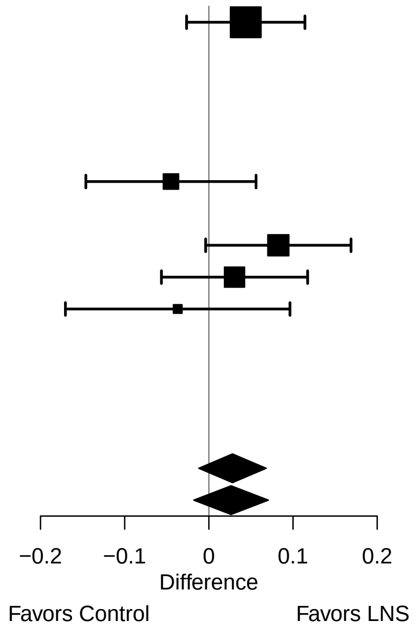

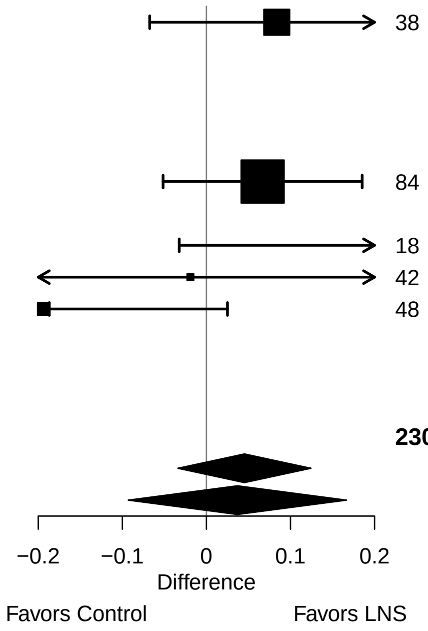

#### Supplemental figure 6P: 12-mo walking without support prevalence difference

#### 6P4: Stratified by Household sanitation

**P−for−interaction = 0.417**
**Difference in PDs = −0.02 (−0.07, 0.03)**

| Country | Trial | Improved LNS N | Improved Control N | Improved Control Prevalence | Improved PD (95% CI) | Improved Fixed W | Improved Random W | Unimproved LNS N | Unimproved Control N | Unimproved Control Prevalence | Unimproved PD (95% CI) | Unimproved Fixed W | Unimproved Random W |
|---|---|---|---|---|---|---|---|---|---|---|---|---|---|
| Bangladesh | RDNS (22) | 1149 | 563 | 73.4 | 0.07 (0.01, 0.13) | 0.72 | 0.48 | 477 | 226 | 77.0 | 0.07 (0.00, 0.13) | 0.48 | 0.39 |
| Bangladesh | WASH−B (23) | | | | | | | | | | | | |
| Burkina Faso | iLiNS−Zinc (24) | | | | | | | | | | | | |
| Ghana | GHANA (25) | | | | | | | | | | | | |
| Ghana | iLiNS−DYADG (26) | | | | | | | | | | | | |
| Haiti | HAITI (27) | | | | | | | | | | | | |
| Kenya | WASH−B (28) | 35 | 106 | 58.5 | 0.07 (−0.15, 0.29) | 0.06 | 0.13 | 167 | 598 | 59.0 | −0.01 (−0.10, 0.07) | 0.25 | 0.28 |
| Madagascar | MAHAY (29) | | | | | | | | | | | | |
| Malawi | iLiNS−DYADM (30) | 19 | 41 | 43.9 | −0.03 (−0.31, 0.24) | 0.04 | 0.09 | 189 | 382 | 51.8 | 0.11 (0.02, 0.20) | 0.25 | 0.28 |
| Malawi | iLiNS−DOSE (31) | | | | | | | | | | | | |
| Mali | PROMIS CS (32) | 87 | 108 | 63.0 | −0.13 (−0.25, −0.01) | 0.19 | 0.30 | 27 | 38 | 73.7 | −0.04 (−0.30, 0.22) | 0.03 | 0.05 |
| Zimbabwe | SHINE (HIV−) (33) | | | | | | | | | | | | |
| Zimbabwe | SHINE (HIV+) (34) | | | | | | | | | | | | |
| | | **1290** | **818** | | **I² = 0.55, Tau² = 0.00** | | | **860** | **1244** | | **I² = 0.46, Tau² = 0.00** | | |
| **Fixed** | | | | | **0.04 (0.00, 0.07)** | | | | | | **0.04 (0.00, 0.07)** | | |
| **Random** | | | | | **0.02 (−0.06, 0.09)** | | | | | | **0.03 (−0.02, 0.09)** | | |

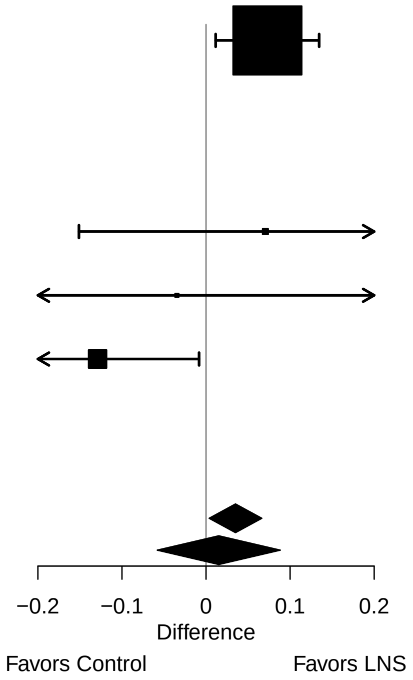

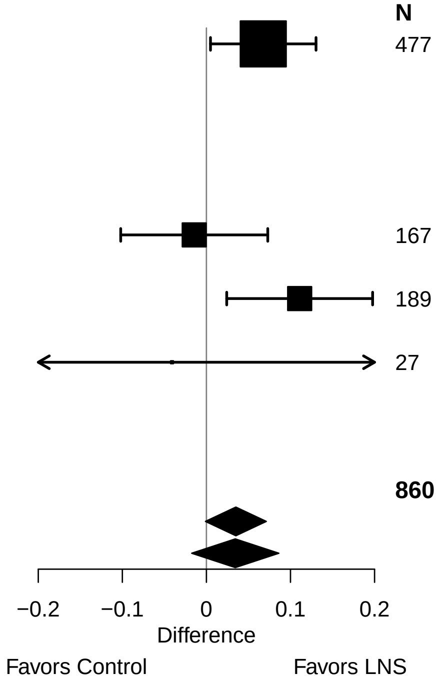

#### Supplemental figure 6P: 12-mo walking without support prevalence difference

##### 6P5: Stratified by Home environment

**P−for−interaction = 0.302**
**Difference in PDs = −0.02 (−0.07, 0.02)**

| Country | Trial | At least median: LNS N | Control N | Control Prevalence | PD (95% CI) | Fixed W | Random W | Less than median: LNS N | Control N | Control Prevalence | PD (95% CI) | Fixed W | Random W |
|---|---|---|---|---|---|---|---|---|---|---|---|---|---|
| Bangladesh | RDNS (22) | 1158 | 560 | 74.5 | 0.09 (0.03, 0.14) | 0.26 | 0.22 | 448 | 217 | 74.7 | 0.05 (−0.03, 0.12) | 0.23 | 0.23 |
| Bangladesh | WASH−B (23) | 243 | 689 | 73.9 | 0.08 (0.03, 0.14) | 0.26 | 0.22 | 231 | 717 | 75.2 | 0.03 (−0.03, 0.09) | 0.38 | 0.38 |
| Burkina Faso | iLiNS−Zinc (24) | | | | | | | | | | | | |
| Ghana | GHANA (25) | | | | | | | | | | | | |
| Ghana | iLiNS−DYADG (26) | 198 | 373 | 50.4 | 0.08 (−0.01, 0.17) | 0.12 | 0.14 | 110 | 233 | 57.5 | 0.03 (−0.08, 0.14) | 0.10 | 0.10 |
| Haiti | HAITI (27) | | | | | | | | | | | | |
| Kenya | WASH−B (28) | 248 | 834 | 55.6 | −0.03 (−0.10, 0.03) | 0.20 | 0.20 | 164 | 608 | 65.5 | 0.03 (−0.05, 0.12) | 0.17 | 0.17 |
| Madagascar | MAHAY (29) | | | | | | | | | | | | |
| Malawi | iLiNS−DYADM (30) | 130 | 278 | 49.3 | 0.09 (−0.02, 0.19) | 0.08 | 0.11 | 72 | 136 | 55.1 | 0.13 (−0.01, 0.28) | 0.06 | 0.06 |
| Malawi | iLiNS−DOSE (31) | 306 | 109 | 65.1 | 0.05 (−0.05, 0.16) | 0.08 | 0.10 | 224 | 67 | 59.7 | −0.06 (−0.19, 0.07) | 0.07 | 0.07 |
| Mali | PROMIS CS (32) | | | | | | | | | | | | |
| Zimbabwe | SHINE (HIV−) (33) | | | | | | | | | | | | |
| Zimbabwe | SHINE (HIV+) (34) | | | | | | | | | | | | |
| | | **2283** | **2843** | | **I² = 0.47, Tau² = 0.00** | | | **1249** | **1978** | | **I² = 0.00, Tau² = 0.00** | | |
| **Fixed** | | | | | **0.06 (0.03, 0.09)** | | | | | | **0.03 (0.00, 0.07)** | | |
| **Random** | | | | | **0.06 (0.02, 0.10)** | | | | | | **0.03 (0.00, 0.07)** | | |

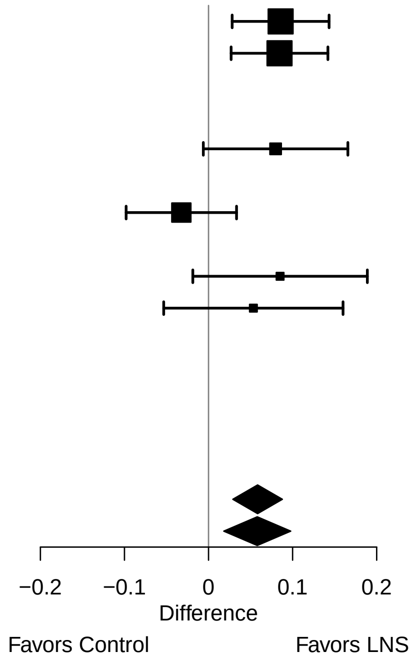

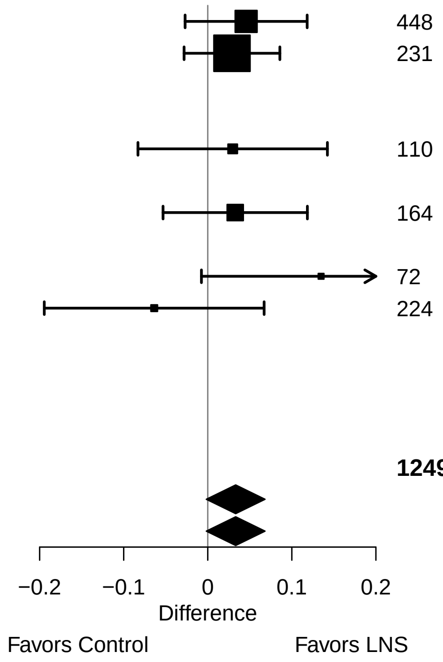

#### Supplemental figure 6P: 12-mo walking without support prevalence difference

#### 6P6: Stratified by Season at the time of assessment

**P−for−interaction = 0.935**
**Difference in PDs = 0.00 (−0.04, 0.05)**

| Country | Trial | Dry LNS N | Dry Control N | Dry Control Prevalence | Dry PD (95% CI) | Dry Fixed W | Dry Random W | Rainy LNS N | Rainy Control N | Rainy Control Prevalence | Rainy PD (95% CI) | Rainy Fixed W | Rainy Random W |
|---|---|---|---|---|---|---|---|---|---|---|---|---|---|
| Bangladesh | RDNS (22) | 899 | 430 | 77.4 | 0.06 (0.01, 0.11) | 0.34 | 0.23 | 729 | 360 | 70.8 | 0.08 (0.01, 0.15) | 0.19 | 0.19 |
| Bangladesh | WASH−B (23) | 267 | 774 | 72.6 | 0.05 (−0.01, 0.11) | 0.25 | 0.21 | 230 | 700 | 76.7 | 0.05 (0.00, 0.10) | 0.31 | 0.31 |
| Burkina Faso | iLiNS−Zinc (24) | | | | | | | | | | | | |
| Ghana | GHANA (25) | 53 | 51 | 78.4 | 0.22 (0.04, 0.40) | 0.03 | 0.05 | 37 | 36 | 69.4 | 0.10 (−0.12, 0.32) | 0.02 | 0.02 |
| Ghana | iLiNS−DYADG (26) | 163 | 320 | 48.4 | 0.08 (−0.01, 0.17) | 0.10 | 0.13 | 164 | 343 | 57.4 | 0.06 (−0.04, 0.15) | 0.11 | 0.11 |
| Haiti | HAITI (27) | | | | | | | | | | | | |
| Kenya | WASH−B (28) | 131 | 461 | 62.5 | −0.03 (−0.13, 0.07) | 0.09 | 0.12 | 316 | 1143 | 58.6 | −0.01 (−0.06, 0.05) | 0.29 | 0.29 |
| Madagascar | MAHAY (29) | | | | | | | | | | | | |
| Malawi | iLiNS−DYADM (30) | 150 | 311 | 53.7 | 0.11 (0.01, 0.21) | 0.09 | 0.12 | 58 | 115 | 44.3 | 0.06 (−0.09, 0.22) | 0.04 | 0.04 |
| Malawi | iLiNS−DOSE (31) | 437 | 152 | 65.8 | 0.01 (−0.07, 0.10) | 0.11 | 0.14 | 165 | 48 | 64.6 | 0.02 (−0.14, 0.17) | 0.04 | 0.04 |
| Mali | PROMIS CS (32) | | | | | | | | | | | | |
| Zimbabwe | SHINE (HIV−) (33) | | | | | | | | | | | | |
| Zimbabwe | SHINE (HIV+) (34) | | | | | | | | | | | | |
| | | **2100** | **2499** | | **$I^2$ = 0.31, $Tau^2$ = 0.00** | | | **1699** | **2745** | | **$I^2$ = 0.00, $Tau^2$ = 0.00** | | |
| **Fixed** | | | | | **0.06 (0.03, 0.09)** | | | | | | **0.04 (0.01, 0.07)** | | |
| **Random** | | | | | **0.06 (0.01, 0.10)** | | | | | | **0.04 (0.01, 0.07)** | | |

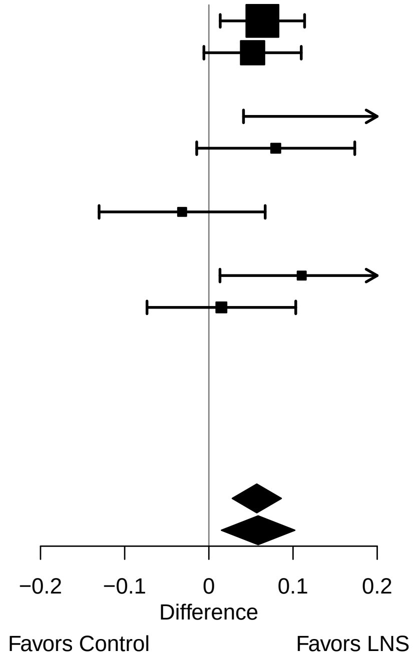

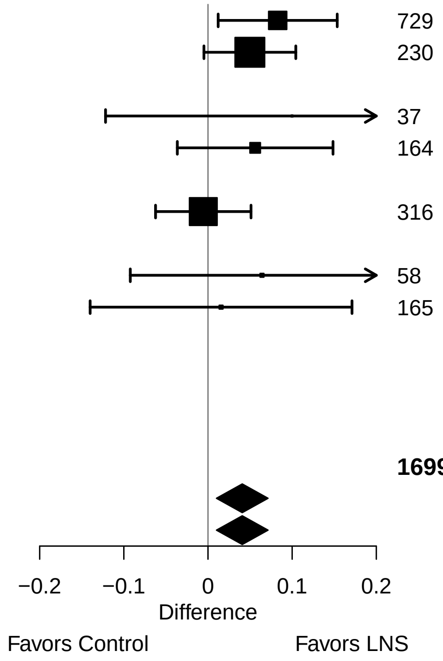
